# The Catatonia Quick Screen (CQS): A Rapid Screening Tool for Catatonia in Adult and Pediatric Populations

**DOI:** 10.1101/2024.11.26.24317940

**Authors:** James Luccarelli, Mark Kalinich, Jo Ellen Wilson, Jinyuan Liu, D. Catherine Fuchs, Andrew Francis, Stephan Heckers, Gregory Fricchione, Joshua Ryan Smith

**Author notes:** Corresponding Author: James Luccarelli, MD, DPhil, Address: Massachusetts General Hospital, 32 Fruit Street, Yawkey 6A, Boston MA 02114. **Declaration of Interest:** JL receives funding from Harvard Medical School Dupont Warren Fellowship and Livingston Awards, the Rappaport Foundation, the American Academy of Child and Adolescent Psychiatry, and the Foundation for Prader-Willi Research. He has received equity and consulting fees from Revival Therapeutics, Inc. MK has received compensation from Watershed Informatics and equity from Watershed Informatics and Revival Therapeutics, Inc. JW receives support from the Department of Veterans Affairs, Geriatric, Research, Education and Clinical Center (GRECC) at the Tennessee Valley Healthcare System in Nashville, TN. GF has received equity from Revival Therapeutics, IP royalties from Adya Health and is on the scientific advisory board of Being Health. SH has received funding from NIH. JRS receives funding from the National Institute of Child and Human Development, Axial, and Roche. All other authors declare no conflicts of interest. **Data Availability:** The data that support the findings of this study are available on request from the corresponding author, JL. The data are not publicly available due to privacy restrictions.

## Abstract

**Introduction:** Catatonia is a neuropsychiatric disorder marked by significant disturbances in motor, cognitive, and affective functioning and that is frequently under-diagnosed. To enhance clinical detection of catatonia, this study aimed to develop a rapid, sensitive Catatonia Quick Screen (CQS) using a reduced set of catatonic signs to facilitate screening in adult and pediatric patients.

**Methods:** Data were derived from two retrospective cohorts totaling 446 patients (254 adults, 192 children) who screened positive for catatonia using the Bush Francis Catatonia Screening Instrument (BFCSI). Sensitivity analyses were performed for all combinations of BFCSI signs, with sensitivity defined as the proportion of patients identified by each subset relative to the full BFCSI. The CQS was developed by selecting signs from the BFCSI based on sensitivity, ease of assessment, and relevance to diverse catatonia presentations.

**Results:** Screening for the presence of any one of four signs—excitement, mutism, staring, or posturing—using the CQS yielded a theoretical sensitivity of 97% (95% CI: 95 to 98%) relative to the full BFCSI (which requires two signs out of 14). The CQS demonstrated 97% sensitivity across both pediatric and adult subsets.

**Conclusion:** The Catatonia Quick Screen provides a rapid screening alternative to the BFCSI with high sensitivity, potentially improving early detection of catatonia in clinical settings. Future prospective studies are necessary to validate the CQS’s sensitivity and to determine its specificity in clinical populations.

**Significant Outcomes:**

- The Catatonia Quick Scale (CQS) is a newly developed rapid screen for catatonia.
- Assessment for any one of four signs—excitement, mutism, staring, and posturing—has 97% sensitivity for detecting catatonia relative to the BFCSI.
- This screening is sensitive in both adult and pediatric populations.

**Limitations:**

- The study relies on retrospective data.
- Only patients who initially screened positive for catatonia were included, so the specificity of the CQS could not be assessed.
- Most of the sample has a primary psychiatric diagnosis so the generalizability to patients with primary medical disorders is unclear.

## Introduction

Catatonia is a neuropsychiatric disorder associated with wide-ranging disturbances of cognition, affect, and motor function.^1,2^ While initially described in adult patients with schizophrenia, catatonia is now recognized to occur in a wide range of psychiatric, neurodevelopmental, neurologic, and medical illnesses and in patients of all ages.^3–10^ In the general hospital setting, catatonia is particularly common in the intensive care unit (ICU)^11,12^ and is associated with a high morbidity and mortality.^11,13,14^ When promptly identified, however, catatonia is highly responsive to pharmacologic therapies and electroconvulsive therapy,^1,15–17^ highlighting the urgent need for sensitive and easily-implemented screening tools.

Catatonia is assessed using a combination of physical exam and interview, with diagnostic criteria that have changed over time and vary in different classification systems.^18^ Presently, under the *Diagnostic and Statistical Manual of Mental Disorders, Fifth Edition, Text Revision (DSM-5-TR)*, at least three of 12 specified signs are required to make the diagnosis of catatonia,^19^ while *the International Classification of Diseases, Eleventh Revision (ICD-11)* requires 3 signs from a different list of specified signs.^20^ To operationalize the identification and grading of catatonia, numerous clinical rating scales have been developed, of which the Bush Francis Catatonia Rating Scale (BFCRS) is the most commonly cited in the literature.^21^ The BFCRS is a 23-item rating scale;^22^ the 14 initial items of the BFCRS are assessed for in the Bush Francis Catatonia Screening Instrument (BFCSI), with at least two of these 14 items required to be present for the remainder of the exam to occur. While the BFCRS has demonstrated high inter-rater reliability in research studies,^22–24^ clinical studies have identified numerous knowledge gaps among active clinicians in the appropriate use of the scale, with 31% of items misidentified on a standardized video.^25^ Moreover, in clinical practice the majority of catatonia cases go unrecognized in the general hospital,^26^ and there may be particular difficulties in identifying catatonia in individuals with baseline impairments in communication such as in neurodevelopmental disorders.^7,27,28^ As a result, there is an urgent clinical need to enhance the sensitivity of catatonia diagnosis.

Existing catatonia instruments, except for the 14-item BFCSI, are designed to be diagnostic tools, and with that comes the requirement to fully assess for all putative catatonic signs. This necessarily means that such instruments must be relatively long, which increased the complexity of administration and makes their use most appropriate in cases where there is high suspicion for catatonia. A parallel tool would be a *screening instrument*,^29^ which is a scale designed to increase identification of potential cases but is not definitively diagnostic itself;^30^ a positive screen result can then trigger a more comprehensive assessment. An example of such a tool in broad psychiatric use is the Patient Health Questionnaire-2 (PHQ-2), a depression screener consisting of the first two items of the nine-item Patient Health Questionnaire depression module (PHQ-9), which is followed by the larger scale only if positive.^31^

An optimal screening instrument would be brief (to enable broad-scale use), utilize easily defined items (to be operationalizable by non-specialist staff), and be highly sensitive for the condition in question (to minimize false-negative results), but high specificity (few false positives) is of lesser importance as a screen will trigger a more in-depth assessment. No existing instrument meets these criteria as the existing BFCSI requires the assessment of 14 items (meaning that even this screening exam is time-consuming) and includes signs that involve idiosyncratic terminology or easily-confused findings (e.g. stereotypies vs. mannerisms; verbigeration vs. echolalia).

The goal of this study is to describe a rapid screen for catatonia that can be easily applied across care settings. The theoretical sensitivity of this new screen will be assessed using a large retrospective cohort of adult and pediatric patients who screened positive for catatonia using the BFCSI.

## Methods

### Clinical Cohort

Clinical data in this study derive from two existing retrospective cohort studies: a single-site cohort of 339 adults and children^32^ and a multi-site cohort of 143 children.^33^ All patients in both studies were screened for catatonia using the BFCRS. Of these, a 446 (254 adults and 192 children) screened positive for catatonia using the BFCSI by having two or more signs from among the 14 included items and were included in this sample. The presence or absence of each of the 14 signs for these 446 patients were then extracted and used for subsequent analysis.

Demographics for these patients were extracted from the electronic health record. The primary diagnosis for each patient was defined as the first listed diagnosis in the patient’s hospital discharge summary. This study was reviewed by the Institutional Review Boards of Vanderbilt University Medical Center and Mass General Brigham and deemed exempt with a waiver of informed consent.

### Combinations of Signs

To develop an abbreviated screen for catatonia, we assessed for the sensitivity of every possible combination of signs within the BFCSI relative to the full instrument (which requires two positive signs from among 14). As all patients in the cohort screened positive using the BFCSI, this means that sensitivity was defined as the proportion of the 446 patients who displayed *at least one catatonic sign* from among the group of signs assessed by each putative screener. To reduce the number of items screened, the presence of any individual catatonic sign was deemed sufficient to screen positive for catatonia (compared to two positive items on the BFCSI). Thus, for a theoretical 2-item screener assessing only for excitement and immobility/stupor, any patient who displayed either of those two signs would screen positive using that screener, whereas patients with any number of the remaining 12 BFCSI signs except for excitement or immobility/stupor would be a negative screen using the 2-item screener, and would be counted as a false negative.

As a further example, if a patient demonstrated four BFCSI features—immobility/stupor, muteness, rigidity, and waxy flexibility—then if any one of these signs was included in a combination of questions, the patient would be considered positive in that combination. Thus, the single sign “muteness” would be positive for this patient, as would the group of three signs “excitement, muteness, withdrawal,” but the group of five “excitement, staring, posturing, grimacing, verbigeration” would not identify the patient as they did not have any of those five features.

Sensitivities were calculated for each individual BFCSI sign, for each possible pair of signs, each possible group of three signs, etc. up to every possible combination of 13 BFCSI signs. For the 14 BFCSI signs, there are 91 possible combinations of two signs, 364 combinations of three signs, 1,001 combinations of four signs, and 2,002 combinations of five signs.

### Statistical Analysis

The pairwise correlation between each item of the BFCSI was assessed using the Kendall Rank Correlation Coefficient. Due to the large number of pairwise comparisons, no correction was made for multiple testing. 95% confidence intervals for sensitivity of each putative screener (groups of 1, 2, 3…13 questions) were generated using 10,000 bootstrapped samples. Statistical analysis was performed using Python (Version 3.12).

### Screener Generation

From among all possible question combinations, the Catatonia Quick Screen (CQS) was generated by the authors, with the presence of any one of these four signs considered a positive screen for catatonia triggering a more detailed examination. The goal was to maximize sensitivity of the CQS while also considering symptom clustering within different catatonia presentations,^18^ practical ease of administration of the screener, and relative reliability of identification of BFCSI items in prior studies.^25^

## Results

Demographics of the clinical cohort of 446 patients who screened positive for catatonia are listed in Table 1. In total there were 230 female patients (51.6%). Median age was 21, with 192 patients aged 18 years or younger and 254 older than 18 years. Full demographic and diagnostic information for the cohort is given in Table 1.

**Table 1:**
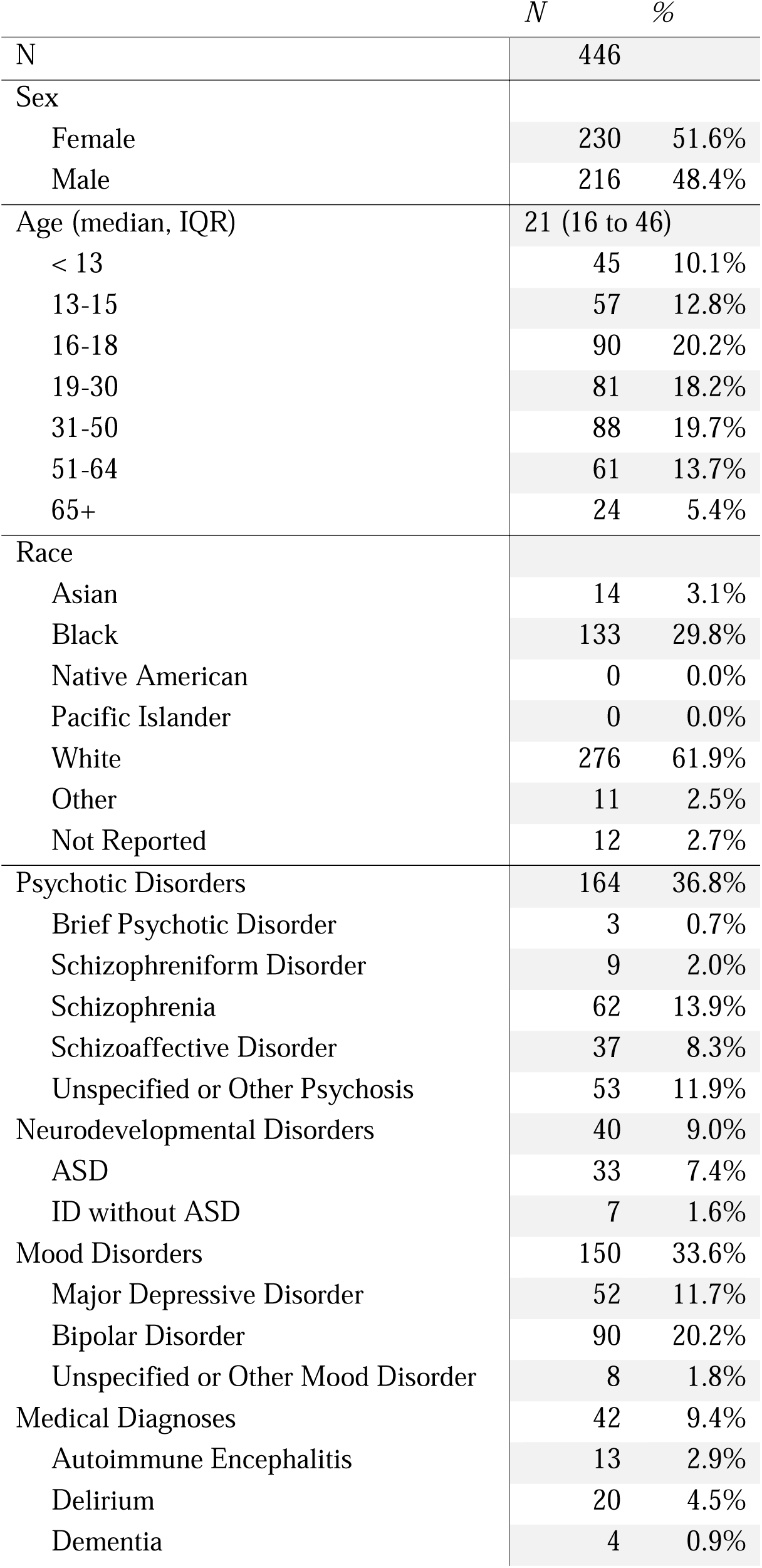
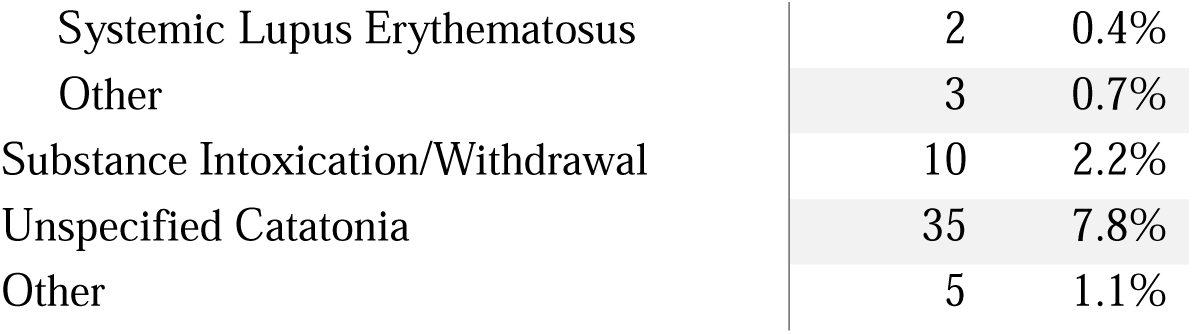
Demographic and diagnostic information for the validation cohort.

|  | *N* | % |
| --- | --- | --- |
| **N** | **446** |  |
| **Sex** |  |  |
| Female | 230 | 51.6% |
| Male | 216 | 48.4% |
| **Age (median, IQR)** | **21 (16 to 46)** |  |
| < 13 | 45 | 10.1% |
| 13-15 | 57 | 12.8% |
| 16-18 | 90 | 20.2% |
| 19-30 | 81 | 18.2% |
| 31-50 | 88 | 19.7% |
| 51-64 | 61 | 13.7% |
| 65+ | 24 | 5.4% |
| **Race** |  |  |
| Asian | 14 | 3.1% |
| Black | 133 | 29.8% |
| Native American | 0 | 0.0% |
| Pacific Islander | 0 | 0.0% |
| White | 276 | 61.9% |
| Other | 11 | 2.5% |
| Not Reported | 12 | 2.7% |
| **Psychotic Disorders** | **164** | **36.8%** |
| Brief Psychotic Disorder | 3 | 0.7% |
| Schizophreniform Disorder | 9 | 2.0% |
| Schizophrenia | 62 | 13.9% |
| Schizoaffective Disorder | 37 | 8.3% |
| Unspecified or Other Psychosis | 53 | 11.9% |
| **Neurodevelopmental Disorders** | **40** | **9.0%** |
| ASD | 33 | 7.4% |
| ID without ASD | 7 | 1.6% |
| **Mood Disorders** | **150** | **33.6%** |
| Major Depressive Disorder | 52 | 11.7% |
| Bipolar Disorder | 90 | 20.2% |
| Unspecified or Other Mood Disorder | 8 | 1.8% |
| **Medical Diagnoses** | **42** | **9.4%** |
| Autoimmune Encephalitis | 13 | 2.9% |
| Delirium | 20 | 4.5% |
| Dementia | 4 | 0.9% |
| Systemic Lupus Erythematosus | 2 | 0.4% |
| Other | 3 | 0.7% |
| Substance Intoxication/Withdrawal | 10 | 2.2% |
| Unspecified Catatonia | 35 | 7.8% |
| Other | 5 | 1.1% |

Patients displayed a mean of 5.5±2.3 and a median of 5 (IQR: 4 to 7) signs using the BFCSI. All patients displayed at least two signs (as required for cohort membership), while the maximum number of signs was 13. Among individual BFCSI signs, the most common was staring, which was present in 78.0% of patients. The least common was mannerisms, which was present in 17.5% of patients. Overall symptom prevalence was similar for the total sample, the adult subset, and the pediatric subset (Figure 1). Catatonic signs were not randomly distributed among patients, with certain signs showing strong correlation (Figure 2). For instance, immobility/stupor and mutism were positively correlated (Kendall Rank Correlation Coefficient = 0.44), while excitement and immobility/stupor were negatively correlated (Kendall Rank Correlation Coefficient = −0.58). Negativism was weakly correlated with other catatonic signs (highest correlation = 0.15).

**Figure 1:**
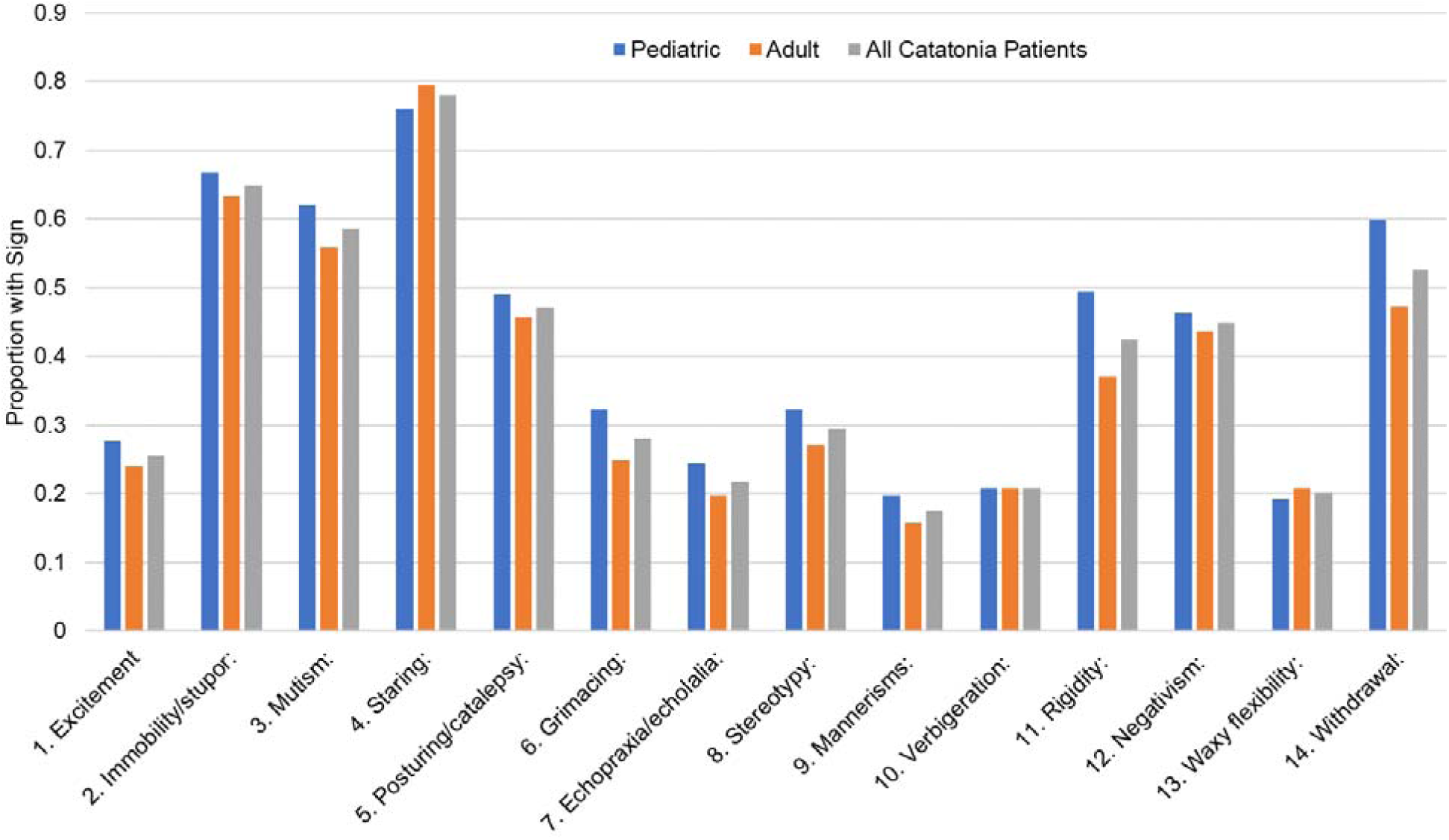
Proportion of the overall sample, pediatric sample, and adult sample expressing each sign of the BFCSI.

**Figure 2:**
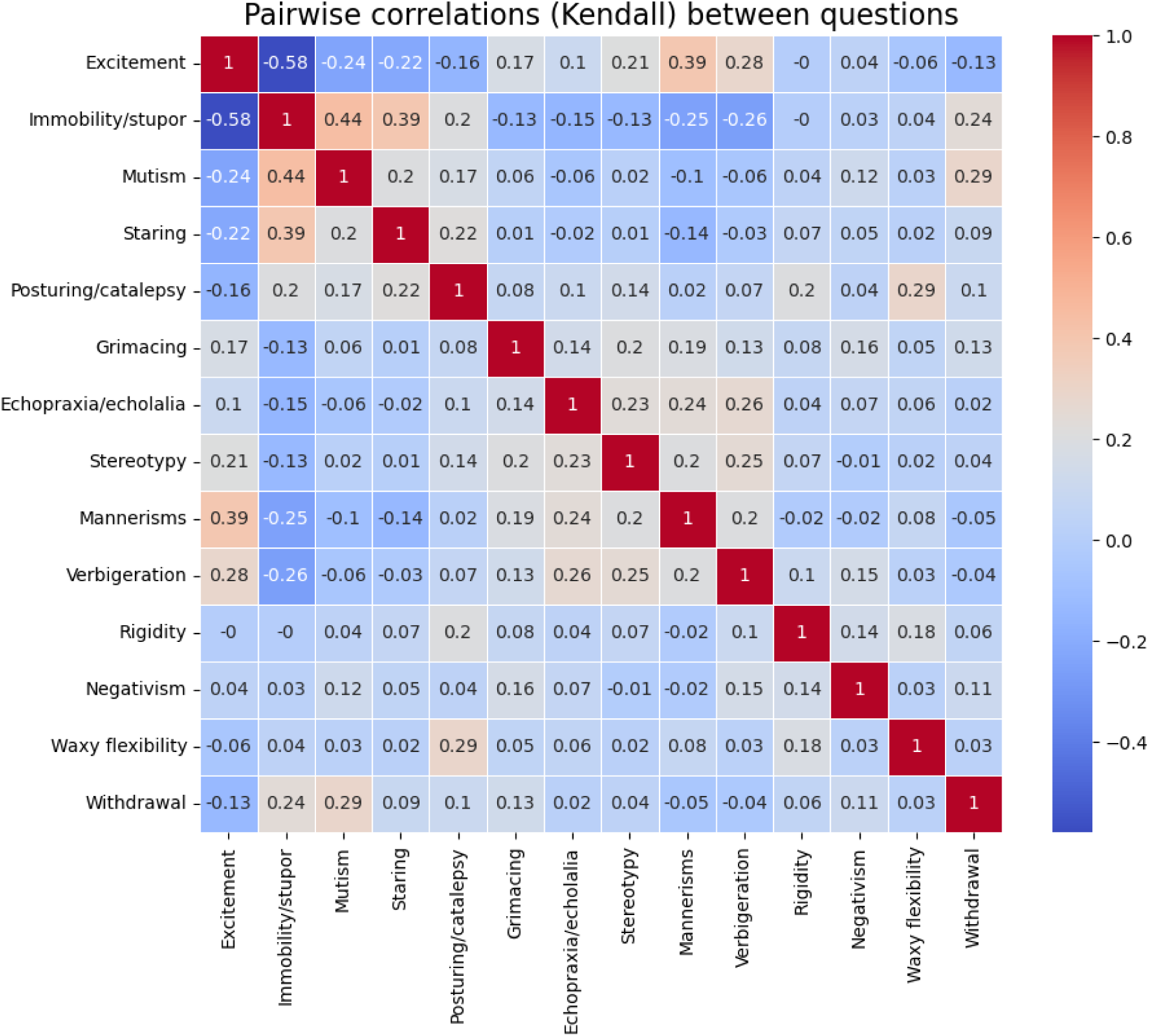
Pairwise correlation between each element of the BFCSI.

To explore the theoretical sensitivity of different combinations of catatonic signs compared to the 14-item BFCSI, every possible combination of signs was computed, with the presence of a single sign within the group considered a positive screen for that combination. Sensitivity was defined as the proportion of patients screening positive with each combination relative to the number screening positive using the full BFCSI. The best combinations of signs yielded a sensitivity of 78% (95% CI: 74% to 82%) for one sign, 88% (95% CI: 85% to 91%) for two signs, 95% (95% CI: 93% to 97%) for three signs, 97% (95% CI: 96% to 99%) for four signs, 99% (95% CI: 98% to 100%) for five signs, and 100% sensitivity for combinations of 6 or more signs (Figure 3). A list of theoretical sensitivities for each possible combination of signs is given in Supplementary Tables S1-S13.

**Figure 3:**
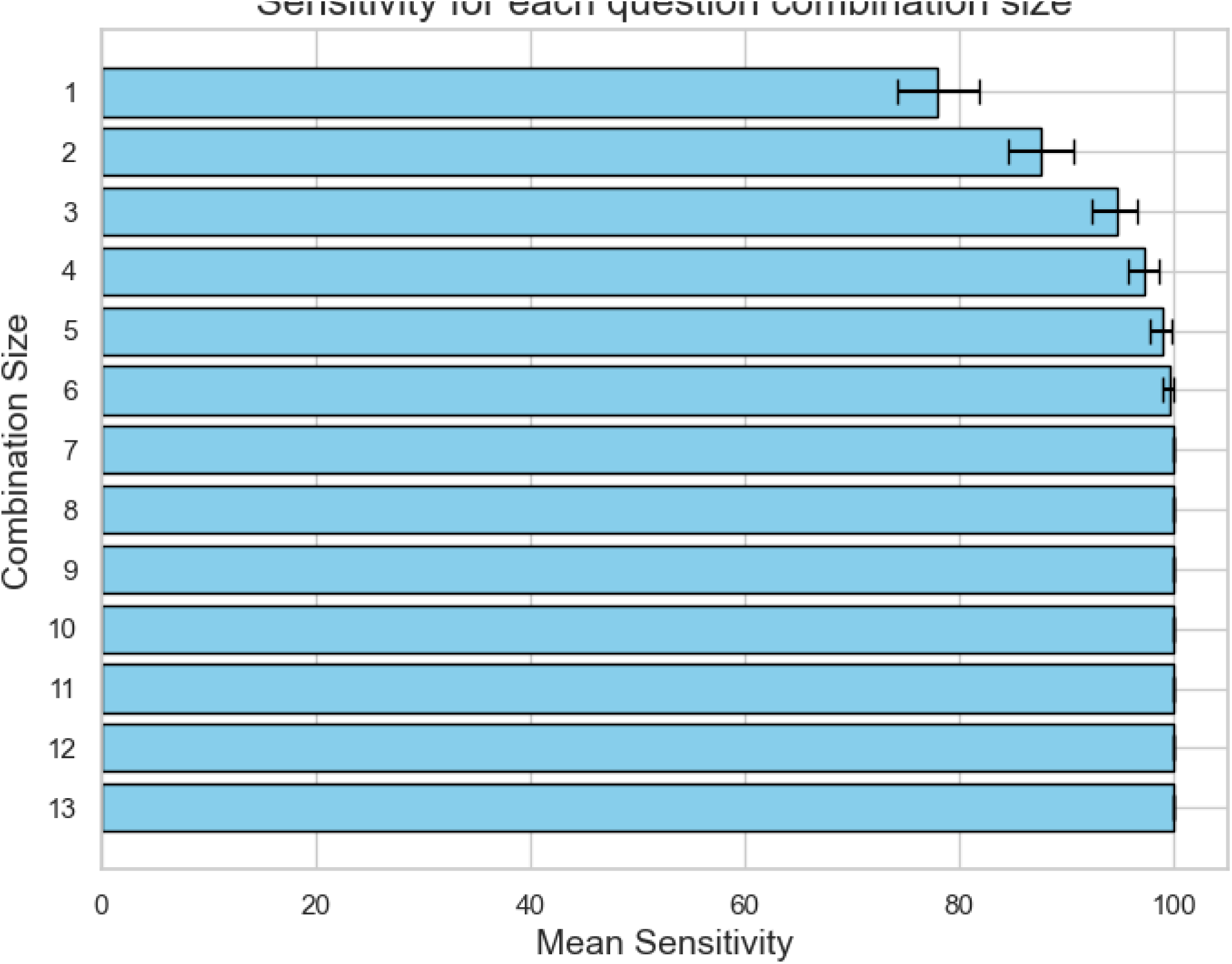
Sensitivity of catatonia screen vs. number of questions asked for the most sensitive question combination. The 95% confidence interval derived from bootstrapping analysis is displayed.

Using these data regarding combinations of signs, the Catatonia Quick Screen (CQS), which combines four signs, is proposed as a screening scale for catatonia (Figure 4). The CQS consists of assessment for four catatonic signs: excitement, mutism, staring, and posturing (BFCSI items 1, 3, 4, and 5). The presence of any one of these four signs is defined as a positive screen for catatonia, which should trigger a full assessment using a catatonia diagnostic instrument. This 4-item CQS has a theoretical sensitivity of 97% (95% CI: 95% to 98%) relative to the full BFCSI, with 97% sensitivity (95% CI: 95% to 99%) in the adult subset and 97% (95% CI: 94% to 99%) in the pediatric subsets of patients. Among the 52 patients with a primary medical or substance diagnosis, theoretical sensitivity of the CQS is 98% (95% CI: 94% to 100%).

**Figure 4:**
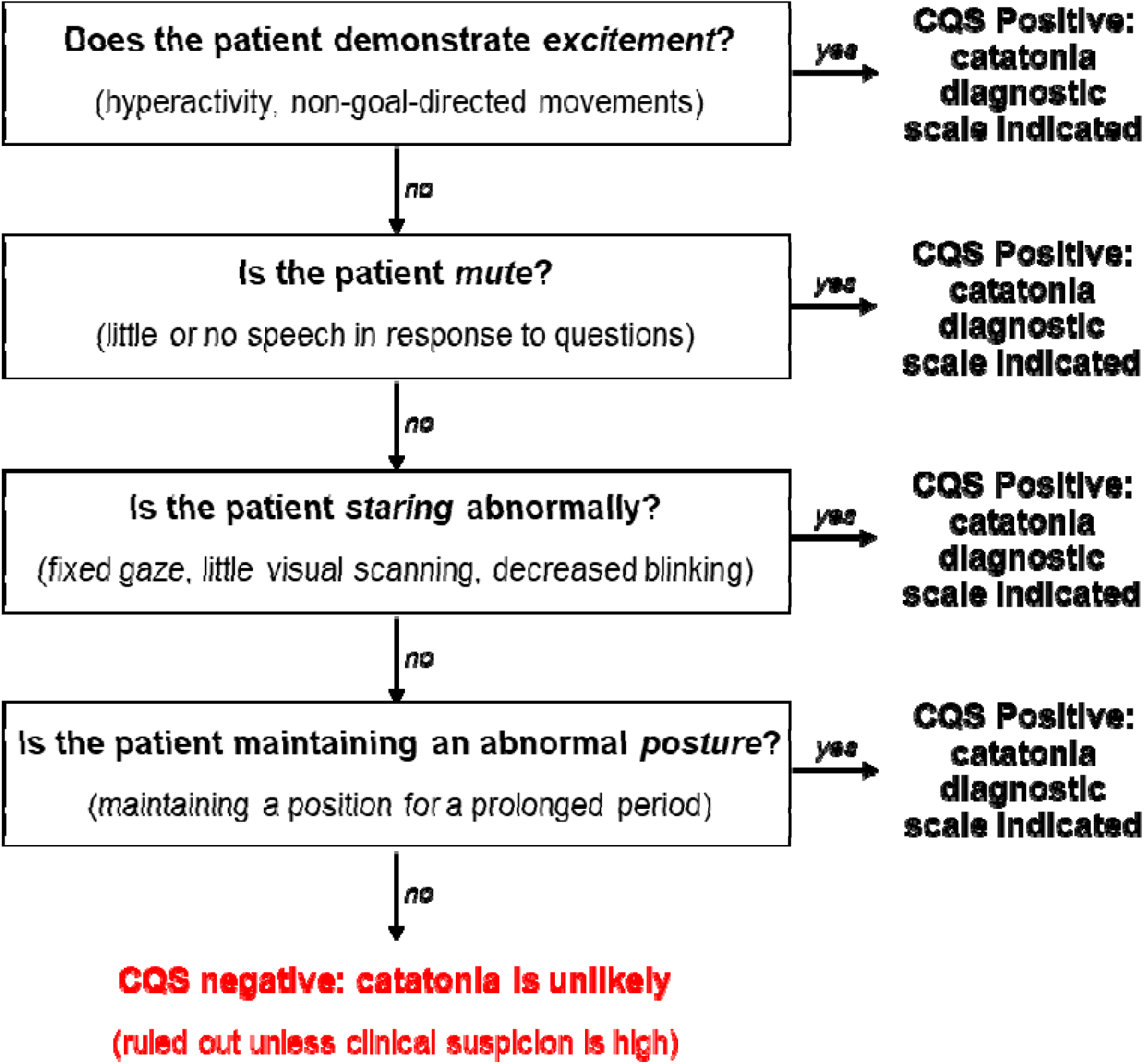
Assessment for catatonia using the Catatonia Quick Screen (CQS). A positive response to any of the four questions is a positive screen, which should be followed up by a complete catatonia diagnostic scale. A negative response to all four questions makes catatonia unlikely and rules out the diagnosis unless there remains high clinical suspicion for catatonia, in which case further assessment can be performed.

## Discussion

Compared to the BFCSI, which requires two catatonic signs from a list of 14 possible signs, assessment of just four signs (excitement, mutism, staring, and posturing), with only one of the four required to screen as positive, has a theoretical 97% sensitivity for detecting putative cases of catatonia, with high sensitivity in both adult and pediatric populations. The CQS has multiple strengths as a potential screening tool. The four items of the CQS are all based on observed signs, without the requirement for a physical exam. A study by Wortzel *et al.* assessed practitioner accuracy in interpretation of the signs in the BFCRS by theoretical assessment (multiple choice questions) and practice assessment using video assessments of standardized patients.^25^ Each of the four items in the CQS demonstrated high recognition in standardized video assessment (from a low of 51.9% for staring to a high of 85.1% for mutism).^25^ Moreover, the signs themselves are easily described, do not require esoteric vocabulary terms not in common use outside of psychiatry, and can be assessed in patients who are hyperactive or hypoactive. They also assess for elements catatonia described in most rating scales: abnormal psychomotor activity, increased psychomotor activity, and decreased psychomotor activity.^18^ Completion of the CQS can be accomplished in less than 60 seconds and, if necessary, via telemedicine examination or from across the room, which further increases its value as a screening tool in both the inpatient and outpatient settings.^34–36^

As with other screening instruments, a positive score on the CQS would not itself be diagnostic for catatonia, as numerous other conditions including delirium,^37^ akinetic mutism,^38^ non-convulsive status epilepticus,^39^ acquired brain injury,^40^ and neuroleptic malignant syndrome^41^ would all be expected to score positive on the CQS. The optimal diagnosis of catatonia remains an area of active research, and there is no clear optimal diagnostic instrument once catatonia is suspected, but a positive CQS should trigger more detailed assessment using an appropriate rating scale for the particular patient (for instance, the Pediatric Catatonia Rating Scale for pediatric patients,^42^ or the Kanner Catatonia Rating Scale for those with neurodevelopmental disorders^43^).

The potential utility of the CQS is strengthened by the clinical cohort used to derive the measure, particularly by its large multi-site nature and its inclusion of patients across the age span. There are, however, numerous limitations of this study. First, all clinical data is derived from retrospective cohorts, so any errors in identifying the BFCSI signs at the time of initial diagnosis would bias the results of the study. This may bias the cohort towards more severe cases of catatonia or those with more obvious combinations of signs, and so may not reflect all catatonia cases. To conclusively determine the utility of this screener, a prospective validation study would need to be conducted, ideally across multiple sites and clinical raters. As only cases screening positive for catatonia using the BFCSI were included, we are unable to comment on the specificity of this screener. A validation study would ideally also measure the number of false positives resulting from the CQS, which would permit an understanding of the positive and negative predictive values of the measure and may help identify other combinations of signs that may have greater sensitivity or specificity. Finally, the four questions in the CQS were determined by the author group, and many other possible sign combinations would have equivalent sensitivity. Among four question combinations, 59 different combinations (from among the 1,001 possible combinations of 14 questions) would have sensitivities ≥95% (Supplementary Table 1), as would 564 different combinations of five questions. The CQS questions may thus not be the optimal question combination overall nor for specific subgroups of patients.

## Conclusion

High sensitivity for detecting putative cases of catatonia can be achieved using far fewer questions than the existing BFCSI. The presence of any one of four catatonic signs—excitement, mutism, staring, or posturing—has a theoretical sensitivity of 97% (95% CI: 95% to 98%) relative to the BFCSI based on a multisite retrospective cohort of 446 pediatric and adult patients who screened positive for catatonia, making this a rapid screener for catatonia that may enhance recognition of this neuropsychiatric disorder. Prospective research is needed to validate this Catatonia Quick Screen and to assess for the sensitivity and specificity of the measure.

## Data Availability

The data that support the findings of this study are available on request from the corresponding author, JL. The data are not publicly available due to privacy restrictions.

## Notes

**Funding:** This work was supported by the National Institute of Mental Health (T32MH112485; JL), National Institute of Child and Human Development (1P50HD103537; JRS). The sponsors had no role in study design, writing of the report, or data collection, analysis, or interpretation.

### Funding Statement

This work was supported by the National Institute of Mental Health (T32MH112485; JL), National Institute of Child and Human Development (1P50HD103537; JRS). The sponsors had no role in study design, writing of the report, or data collection, analysis, or interpretation.

### Author Declarations

This study was reviewed by the Institutional Review Boards of Vanderbilt University Medical Center and Mass General Brigham and deemed exempt with a waiver of informed consent.

