## Supplemental Tables 1-13 for "The Catatonia Quick Screen (CQS): A Rapid Screening Tool for Catatonia in Adult and Pediatric Populations"

James Luccarelli, MD, DPhil^1,2^

Mark Kalinich, MD, PhD^1,3^

Jo Ellen Wilson, MD, PhD, MPH^4,5^

Jinyuan Liu, PhD^6^

D. Catherine Fuchs, MD^4^

Andrew Francis, MD, PhD^7^

Stephan Heckers, MD, MSc^4^

Gregory Fricchione, MD^1,2^

Joshua Ryan Smith, MD^3,8^

^1^Harvard Medical School, Boston, MA, USA

^2^Department of Psychiatry, Massachusetts General Hospital, Boston, MA, USA

^3^Department of Psychiatry, Beth Israel Deaconess Medical Center, Boston, MA, USA

^4^Department of Psychiatry and Behavioral Sciences, Vanderbilt University Medical Center at Village of Vanderbilt, Nashville, Tennessee, USA

^5^Department of Veterans’ Affairs Geriatric Research, Education and Clinical Center, Tennessee Valley Healthcare System, Nashville, TN, USA

^6^Department of Biostatistics; Vanderbilt University, Nashville, TN, USA

^7^Department of Psychiatry and Behavioral Health, Penn State Medical School, Hershey, PA, USA

^8^Vanderbilt Kennedy Center, Vanderbilt University, Nashville, TN, USA

**Corresponding Author**

James Luccarelli, MD, DPhil

Address: Massachusetts General Hospital, 32 Fruit Street, Yawkey 6A, Boston MA 02114

Table S1: sensitivity (relative to the full BFCSI) of all single questions, and the 95% confidence interval derived from bootstrapping, sorted by decreasing sensitivity.

|  |  |  | 95% CI | |
| --- | --- | --- | --- | --- |
| # of Qs | Combination | Sensitivity | Lower | Upper |
| 1 | Staring | 78 | 74.2 | 81.8 |
| 1 | Immobility/stupor | 64.8 | 60.3 | 69.3 |
| 1 | Mutism | 58.5 | 53.8 | 63.2 |
| 1 | Withdrawal | 52.7 | 48.2 | 57.4 |
| 1 | Posturing/catalepsy | 47.1 | 42.4 | 51.6 |
| 1 | Negativism | 44.8 | 40.1 | 49.6 |
| 1 | Rigidity | 42.4 | 37.9 | 47.1 |
| 1 | Stereotypy | 29.4 | 25.1 | 33.6 |
| 1 | Grimacing | 28 | 24 | 32.3 |
| 1 | Excitement | 25.6 | 21.5 | 29.6 |
| 1 | Echopraxia/echolalia | 21.7 | 17.9 | 25.6 |
| 1 | Verbigeration | 20.8 | 17 | 24.7 |
| 1 | Waxy flexibility | 20.2 | 16.6 | 24 |
| 1 | Mannerisms | 17.5 | 14.1 | 21.1 |

Table S2: sensitivity (relative to the full BFCSI) of all two question combinations, and the 95% confidence interval derived from bootstrapping, sorted by decreasing sensitivity.

|  |  |  | 95% CI | |
| --- | --- | --- | --- | --- |
| # of Qs | Combination | Sensitivity | Lower | Upper |
| 2 | Excitement,Staring | 87.7 | 84.5 | 90.6 |
| 2 | Staring,Withdrawal | 87.7 | 84.5 | 90.6 |
| 2 | Mutism,Staring | 86.8 | 83.4 | 89.9 |
| 2 | Staring,Negativism | 86.8 | 83.6 | 89.9 |
| 2 | Excitement,Immobility/stupor | 85.9 | 82.5 | 89 |
| 2 | Staring,Rigidity | 85.9 | 82.5 | 89 |
| 2 | Immobility/stupor,Staring | 84.5 | 81.2 | 87.7 |
| 2 | Staring,Stereotypy | 84.3 | 80.9 | 87.7 |
| 2 | Staring,Grimacing | 84.1 | 80.7 | 87.4 |
| 2 | Staring,Mannerisms | 84.1 | 80.7 | 87.4 |
| 2 | Staring,Posturing/catalepsy | 83.9 | 80.5 | 87.2 |
| 2 | Staring,Echopraxia/echolalia | 83.2 | 79.8 | 86.5 |
| 2 | Staring,Verbigeration | 83.2 | 79.6 | 86.5 |
| 2 | Staring,Waxy flexibility | 82 | 78.3 | 85.4 |
| 2 | Immobility/stupor,Negativism | 79.8 | 76 | 83.6 |
| 2 | Immobility/stupor,Rigidity | 79.8 | 76 | 83.4 |
| 2 | Immobility/stupor,Stereotypy | 78 | 74.2 | 81.8 |
| 2 | Immobility/stupor,Withdrawal | 77.5 | 73.8 | 81.4 |
| 2 | Immobility/stupor,Grimacing | 77.4 | 73.5 | 81.2 |
| 2 | Immobility/stupor,Verbigeration | 77.1 | 73.1 | 81.2 |
| 2 | Immobility/stupor,Posturing/catalepsy | 76.7 | 72.6 | 80.5 |
| 2 | Immobility/stupor,Mannerisms | 75.5 | 71.5 | 79.4 |
| 2 | Immobility/stupor,Echopraxia/echolalia | 75.3 | 71.3 | 79.4 |
| 2 | Immobility/stupor,Mutism | 75.1 | 71.1 | 79.1 |
| 2 | Mutism,Rigidity | 75.1 | 71.1 | 79.1 |
| 2 | Excitement,Mutism | 74.2 | 70.2 | 78.3 |
| 2 | Mutism,Negativism | 74.2 | 70.2 | 78.3 |
| 2 | Mutism,Posturing/catalepsy | 74 | 70 | 78 |
| 2 | Mutism,Withdrawal | 73.3 | 69.1 | 77.4 |
| 2 | Posturing/catalepsy,Withdrawal | 72.5 | 68.4 | 76.5 |
| 2 | Rigidity,Withdrawal | 71.3 | 67 | 75.3 |
| 2 | Immobility/stupor,Waxy flexibility | 71.1 | 66.8 | 75.1 |
| 2 | Negativism,Withdrawal | 71.1 | 66.8 | 75.3 |
| 2 | Mutism,Stereotypy | 70.1 | 65.9 | 74.2 |
| 2 | Posturing/catalepsy,Negativism | 69.7 | 65.5 | 74 |
| 2 | Mutism,Echopraxia/echolalia | 68.9 | 64.6 | 73.1 |
| 2 | Mutism,Grimacing | 68.8 | 64.6 | 73.1 |
| 2 | Mutism,Verbigeration | 68.4 | 64.1 | 72.6 |
| 2 | Excitement,Withdrawal | 67.7 | 63.2 | 72 |
| 2 | Mutism,Mannerisms | 67.7 | 63.2 | 72 |
| 2 | Mutism,Waxy flexibility | 66.4 | 61.9 | 70.6 |
| 2 | Stereotypy,Withdrawal | 65.8 | 61.4 | 70.2 |
| 2 | Rigidity,Negativism | 64.8 | 60.3 | 69.1 |
| 2 | Posturing/catalepsy,Rigidity | 64.6 | 60.3 | 69.1 |
| 2 | Excitement,Posturing/catalepsy | 64.1 | 59.9 | 68.4 |
| 2 | Verbigeration,Withdrawal | 63.4 | 59 | 67.9 |
| 2 | Grimacing,Withdrawal | 63 | 58.5 | 67.5 |
| 2 | Echopraxia/echolalia,Withdrawal | 62.5 | 58.1 | 66.8 |
| 2 | Mannerisms,Withdrawal | 61.9 | 57.4 | 66.4 |
| 2 | Waxy flexibility,Withdrawal | 61.7 | 57.2 | 66.1 |
| 2 | Stereotypy,Negativism | 61.2 | 56.5 | 65.7 |
| 2 | Posturing/catalepsy,Grimacing | 60.1 | 55.4 | 64.6 |
| 2 | Posturing/catalepsy,Stereotypy | 59.4 | 54.7 | 63.9 |
| 2 | Excitement,Negativism | 58.1 | 53.4 | 62.6 |
| 2 | Stereotypy,Rigidity | 57.6 | 53.1 | 62.1 |
| 2 | Excitement,Rigidity | 57.2 | 52.5 | 61.7 |
| 2 | Grimacing,Negativism | 56.7 | 52 | 61.4 |
| 2 | Grimacing,Rigidity | 56.7 | 52.2 | 61.2 |
| 2 | Posturing/catalepsy,Verbigeration | 56.7 | 52 | 61.4 |
| 2 | Posturing/catalepsy,Echopraxia/echolalia | 56.5 | 51.8 | 61 |
| 2 | Posturing/catalepsy,Mannerisms | 56.1 | 51.6 | 60.5 |
| 2 | Echopraxia/echolalia,Negativism | 55.4 | 50.7 | 60.1 |
| 2 | Negativism,Waxy flexibility | 55.4 | 50.7 | 60.1 |
| 2 | Mannerisms,Negativism | 54.9 | 50.2 | 59.6 |
| 2 | Echopraxia/echolalia,Rigidity | 54 | 49.3 | 58.7 |
| 2 | Verbigeration,Negativism | 53.4 | 48.7 | 58.1 |
| 2 | Mannerisms,Rigidity | 52.9 | 48.2 | 57.6 |
| 2 | Verbigeration,Rigidity | 52.5 | 47.8 | 57 |
| 2 | Posturing/catalepsy,Waxy flexibility | 52 | 47.3 | 56.7 |
| 2 | Rigidity,Waxy flexibility | 50.5 | 46 | 55.2 |
| 2 | Grimacing,Stereotypy | 45.1 | 40.4 | 49.8 |
| 2 | Excitement,Stereotypy | 43.3 | 38.8 | 48 |
| 2 | Stereotypy,Waxy flexibility | 43.3 | 38.6 | 48 |
| 2 | Excitement,Grimacing | 43 | 38.3 | 47.5 |
| 2 | Excitement,Waxy flexibility | 41.7 | 37.2 | 46.4 |
| 2 | Grimacing,Waxy flexibility | 41.7 | 37.2 | 46.4 |
| 2 | Grimacing,Echopraxia/echolalia | 41 | 36.5 | 45.7 |
| 2 | Grimacing,Verbigeration | 40.6 | 36.1 | 45.3 |
| 2 | Echopraxia/echolalia,Stereotypy | 40.3 | 35.9 | 45.1 |
| 2 | Excitement,Echopraxia/echolalia | 39.9 | 35.4 | 44.4 |
| 2 | Stereotypy,Verbigeration | 39.5 | 35.2 | 44.2 |
| 2 | Stereotypy,Mannerisms | 38.3 | 33.9 | 42.8 |
| 2 | Grimacing,Mannerisms | 37.4 | 33.2 | 41.9 |
| 2 | Echopraxia/echolalia,Waxy flexibility | 36.5 | 32.3 | 41 |
| 2 | Verbigeration,Waxy flexibility | 36.3 | 31.8 | 40.8 |
| 2 | Excitement,Verbigeration | 36.1 | 31.6 | 40.6 |
| 2 | Echopraxia/echolalia,Verbigeration | 33.6 | 29.4 | 38.1 |
| 2 | Mannerisms,Waxy flexibility | 32.9 | 28.5 | 37.4 |
| 2 | Excitement,Mannerisms | 32.1 | 27.8 | 36.3 |
| 2 | Echopraxia/echolalia,Mannerisms | 31.6 | 27.4 | 36.1 |
| 2 | Mannerisms,Verbigeration | 31.6 | 27.4 | 36.1 |

Table S3: sensitivity (relative to the full BFCSI) of all three question combinations, and the 95% confidence interval derived from bootstrapping, sorted by decreasing sensitivity.

|  |  |  | 95% CI | |
| --- | --- | --- | --- | --- |
| # of Qs | Combination | Sensitivity | Lower | Upper |
| 3 | Excitement,Mutism,Staring | 94.6 | 92.4 | 96.6 |
| 3 | Excitement,Immobility/stupor,Staring | 93.7 | 91.5 | 96 |
| 3 | Excitement,Staring,Withdrawal | 93.5 | 91 | 95.7 |
| 3 | Excitement,Staring,Negativism | 93.1 | 90.6 | 95.3 |
| 3 | Excitement,Staring,Rigidity | 93.1 | 90.6 | 95.3 |
| 3 | Excitement,Immobility/stupor,Rigidity | 92.8 | 90.4 | 95.1 |
| 3 | Staring,Negativism,Withdrawal | 92.8 | 90.4 | 95.1 |
| 3 | Staring,Rigidity,Withdrawal | 92.4 | 89.9 | 94.6 |
| 3 | Excitement,Staring,Posturing/catalepsy | 92.2 | 89.7 | 94.6 |
| 3 | Mutism,Staring,Negativism | 91.9 | 89.2 | 94.4 |
| 3 | Staring,Mannerisms,Withdrawal | 91.7 | 89 | 94.2 |
| 3 | Excitement,Immobility/stupor,Negativism | 91.5 | 88.8 | 93.9 |
| 3 | Mutism,Staring,Mannerisms | 91.5 | 88.8 | 93.9 |
| 3 | Mutism,Staring,Withdrawal | 91.5 | 88.8 | 93.9 |
| 3 | Mutism,Staring,Rigidity | 91.3 | 88.6 | 93.7 |
| 3 | Staring,Mannerisms,Negativism | 91.3 | 88.6 | 93.7 |
| 3 | Staring,Rigidity,Negativism | 91.2 | 88.6 | 93.7 |
| 3 | Excitement,Immobility/stupor,Posturing/catalepsy | 91.1 | 88.3 | 93.5 |
| 3 | Excitement,Staring,Waxy flexibility | 91 | 88.3 | 93.5 |
| 3 | Staring,Posturing/catalepsy,Withdrawal | 91 | 88.3 | 93.5 |
| 3 | Staring,Stereotypy,Negativism | 91 | 88.3 | 93.5 |
| 3 | Excitement,Immobility/stupor,Withdrawal | 90.8 | 88.1 | 93.3 |
| 3 | Excitement,Staring,Grimacing | 90.8 | 88.1 | 93.5 |
| 3 | Mutism,Staring,Stereotypy | 90.8 | 88.1 | 93.5 |
| 3 | Mutism,Staring,Verbigeration | 90.8 | 88.1 | 93.3 |
| 3 | Staring,Mannerisms,Rigidity | 90.8 | 88.1 | 93.5 |
| 3 | Excitement,Staring,Echopraxia/echolalia | 90.6 | 87.9 | 93.3 |
| 3 | Immobility/stupor,Staring,Negativism | 90.6 | 87.9 | 93.3 |
| 3 | Mutism,Staring,Echopraxia/echolalia | 90.6 | 87.9 | 93.3 |
| 3 | Staring,Grimacing,Withdrawal | 90.6 | 87.9 | 93.3 |
| 3 | Staring,Stereotypy,Withdrawal | 90.6 | 87.9 | 93.3 |
| 3 | Excitement,Immobility/stupor,Mutism | 90.4 | 87.4 | 93 |
| 3 | Mutism,Staring,Posturing/catalepsy | 90.4 | 87.4 | 93 |
| 3 | Excitement,Staring,Stereotypy | 90.3 | 87.4 | 92.8 |
| 3 | Staring,Posturing/catalepsy,Negativism | 90.3 | 87.4 | 93 |
| 3 | Staring,Stereotypy,Rigidity | 90.3 | 87.7 | 93 |
| 3 | Immobility/stupor,Staring,Mannerisms | 90.1 | 87.2 | 92.8 |
| 3 | Staring,Echopraxia/echolalia,Negativism | 90.1 | 87.2 | 92.8 |
| 3 | Immobility/stupor,Staring,Rigidity | 89.9 | 87 | 92.6 |
| 3 | Mutism,Staring,Grimacing | 89.9 | 87 | 92.6 |
| 3 | Staring,Echopraxia/echolalia,Withdrawal | 89.9 | 87 | 92.6 |
| 3 | Staring,Verbigeration,Withdrawal | 89.9 | 87 | 92.6 |
| 3 | Staring,Waxy flexibility,Withdrawal | 89.9 | 87 | 92.6 |
| 3 | Excitement,Immobility/stupor,Echopraxia/echolalia | 89.7 | 87 | 92.4 |
| 3 | Excitement,Staring,Verbigeration | 89.7 | 86.8 | 92.4 |
| 3 | Immobility/stupor,Staring,Withdrawal | 89.7 | 86.8 | 92.4 |
| 3 | Staring,Grimacing,Rigidity | 89.7 | 86.8 | 92.4 |
| 3 | Excitement,Immobility/stupor,Grimacing | 89.5 | 86.5 | 92.2 |
| 3 | Staring,Echopraxia/echolalia,Rigidity | 89.5 | 86.5 | 92.2 |
| 3 | Staring,Verbigeration,Negativism | 89.5 | 86.5 | 92.2 |
| 3 | Excitement,Immobility/stupor,Verbigeration | 89.3 | 86.3 | 92.2 |
| 3 | Staring,Negativism,Waxy flexibility | 89.3 | 86.3 | 92.2 |
| 3 | Staring,Verbigeration,Rigidity | 89.3 | 86.3 | 92.2 |
| 3 | Excitement,Immobility/stupor,Stereotypy | 89.2 | 86.3 | 92.2 |
| 3 | Excitement,Staring,Mannerisms | 89.2 | 86.3 | 91.9 |
| 3 | Immobility/stupor,Staring,Stereotypy | 89.2 | 86.3 | 92.2 |
| 3 | Immobility/stupor,Staring,Verbigeration | 89.2 | 86.3 | 92.2 |
| 3 | Mutism,Staring,Waxy flexibility | 89 | 86.1 | 91.9 |
| 3 | Staring,Grimacing,Negativism | 89 | 85.9 | 91.9 |
| 3 | Excitement,Immobility/stupor,Waxy flexibility | 88.8 | 85.9 | 91.5 |
| 3 | Staring,Posturing/catalepsy,Rigidity | 88.8 | 85.9 | 91.7 |
| 3 | Immobility/stupor,Staring,Echopraxia/echolalia | 88.6 | 85.4 | 91.5 |
| 3 | Staring,Grimacing,Mannerisms | 88.6 | 85.7 | 91.5 |
| 3 | Staring,Grimacing,Stereotypy | 88.6 | 85.7 | 91.5 |
| 3 | Staring,Posturing/catalepsy,Mannerisms | 88.6 | 85.7 | 91.5 |
| 3 | Immobility/stupor,Staring,Grimacing | 88.3 | 85.2 | 91.3 |
| 3 | Immobility/stupor,Stereotypy,Negativism | 88.3 | 85.2 | 91.3 |
| 3 | Staring,Posturing/catalepsy,Stereotypy | 88.3 | 85.2 | 91.3 |
| 3 | Immobility/stupor,Rigidity,Negativism | 88.1 | 85 | 91 |
| 3 | Immobility/stupor,Staring,Posturing/catalepsy | 87.9 | 84.8 | 90.8 |
| 3 | Staring,Grimacing,Echopraxia/echolalia | 87.9 | 84.8 | 90.8 |
| 3 | Staring,Grimacing,Verbigeration | 87.9 | 84.8 | 90.8 |
| 3 | Staring,Rigidity,Waxy flexibility | 87.9 | 84.8 | 90.8 |
| 3 | Staring,Stereotypy,Mannerisms | 87.9 | 84.8 | 90.8 |
| 3 | Immobility/stupor,Mannerisms,Rigidity | 87.7 | 84.5 | 90.6 |
| 3 | Immobility/stupor,Rigidity,Withdrawal | 87.7 | 84.5 | 90.6 |
| 3 | Staring,Posturing/catalepsy,Echopraxia/echolalia | 87.7 | 84.5 | 90.8 |
| 3 | Staring,Posturing/catalepsy,Grimacing | 87.7 | 84.5 | 90.6 |
| 3 | Immobility/stupor,Mutism,Staring | 87.6 | 84.5 | 90.6 |
| 3 | Staring,Posturing/catalepsy,Verbigeration | 87.6 | 84.5 | 90.6 |
| 3 | Staring,Stereotypy,Verbigeration | 87.6 | 84.5 | 90.6 |
| 3 | Excitement,Immobility/stupor,Mannerisms | 87.5 | 84.3 | 90.4 |
| 3 | Staring,Echopraxia/echolalia,Stereotypy | 87.4 | 84.3 | 90.4 |
| 3 | Immobility/stupor,Posturing/catalepsy,Negativism | 87.2 | 84.1 | 90.4 |
| 3 | Staring,Echopraxia/echolalia,Mannerisms | 87.2 | 84.1 | 90.4 |
| 3 | Staring,Stereotypy,Waxy flexibility | 87.2 | 84.1 | 90.1 |
| 3 | Immobility/stupor,Mannerisms,Negativism | 87 | 83.9 | 90.1 |
| 3 | Staring,Mannerisms,Verbigeration | 87 | 83.9 | 90.1 |
| 3 | Staring,Mannerisms,Waxy flexibility | 86.9 | 83.9 | 90.1 |
| 3 | Immobility/stupor,Staring,Waxy flexibility | 86.8 | 83.6 | 89.9 |
| 3 | Staring,Grimacing,Waxy flexibility | 86.8 | 83.6 | 89.9 |
| 3 | Immobility/stupor,Verbigeration,Negativism | 86.6 | 83.4 | 89.7 |
| 3 | Immobility/stupor,Negativism,Withdrawal | 86.5 | 83.4 | 89.7 |
| 3 | Immobility/stupor,Stereotypy,Rigidity | 86.5 | 83.2 | 89.7 |
| 3 | Staring,Verbigeration,Waxy flexibility | 86.5 | 83.2 | 89.7 |
| 3 | Immobility/stupor,Grimacing,Rigidity | 86.3 | 83 | 89.5 |
| 3 | Immobility/stupor,Verbigeration,Rigidity | 86.1 | 82.7 | 89.2 |
| 3 | Staring,Echopraxia/echolalia,Verbigeration | 86.1 | 82.7 | 89.2 |
| 3 | Staring,Echopraxia/echolalia,Waxy flexibility | 85.9 | 82.7 | 89 |
| 3 | Excitement,Mutism,Rigidity | 85.4 | 82.1 | 88.6 |
| 3 | Immobility/stupor,Echopraxia/echolalia,Negativism | 85.4 | 82.1 | 88.6 |
| 3 | Immobility/stupor,Echopraxia/echolalia,Rigidity | 85.4 | 82.1 | 88.6 |
| 3 | Immobility/stupor,Mutism,Rigidity | 85.4 | 82.1 | 88.8 |
| 3 | Excitement,Mutism,Posturing/catalepsy | 85.2 | 81.8 | 88.6 |
| 3 | Immobility/stupor,Grimacing,Negativism | 85 | 81.6 | 88.1 |
| 3 | Immobility/stupor,Posturing/catalepsy,Withdrawal | 85 | 81.6 | 88.1 |
| 3 | Immobility/stupor,Stereotypy,Withdrawal | 85 | 81.6 | 88.1 |
| 3 | Staring,Posturing/catalepsy,Waxy flexibility | 85 | 81.6 | 88.3 |
| 3 | Immobility/stupor,Mutism,Negativism | 84.8 | 81.4 | 88.1 |
| 3 | Immobility/stupor,Verbigeration,Withdrawal | 84.8 | 81.4 | 88.1 |
| 3 | Mutism,Rigidity,Negativism | 84.6 | 81.2 | 87.9 |
| 3 | Immobility/stupor,Mannerisms,Withdrawal | 84.5 | 81.2 | 87.9 |
| 3 | Immobility/stupor,Negativism,Waxy flexibility | 84.5 | 81.2 | 87.7 |
| 3 | Immobility/stupor,Grimacing,Stereotypy | 84.3 | 80.9 | 87.7 |
| 3 | Immobility/stupor,Posturing/catalepsy,Rigidity | 84.3 | 80.7 | 87.7 |
| 3 | Immobility/stupor,Grimacing,Withdrawal | 84.1 | 80.5 | 87.4 |
| 3 | Immobility/stupor,Mutism,Verbigeration | 84.1 | 80.7 | 87.4 |
| 3 | Immobility/stupor,Posturing/catalepsy,Grimacing | 84.1 | 80.7 | 87.4 |
| 3 | Immobility/stupor,Stereotypy,Verbigeration | 84.1 | 80.7 | 87.4 |
| 3 | Excitement,Mutism,Withdrawal | 83.9 | 80.3 | 87.2 |
| 3 | Immobility/stupor,Grimacing,Verbigeration | 83.9 | 80.5 | 87.2 |
| 3 | Immobility/stupor,Posturing/catalepsy,Stereotypy | 83.8 | 80.3 | 87.2 |
| 3 | Mutism,Rigidity,Withdrawal | 83.8 | 80.5 | 87.2 |
| 3 | Immobility/stupor,Echopraxia/echolalia,Withdrawal | 83.6 | 80 | 87 |
| 3 | Immobility/stupor,Posturing/catalepsy,Mannerisms | 83.6 | 80.3 | 87 |
| 3 | Mutism,Posturing/catalepsy,Negativism | 83.6 | 80.3 | 87 |
| 3 | Immobility/stupor,Mutism,Mannerisms | 83.4 | 79.8 | 86.8 |
| 3 | Immobility/stupor,Mutism,Stereotypy | 83.4 | 80 | 86.8 |
| 3 | Immobility/stupor,Posturing/catalepsy,Verbigeration | 83.4 | 79.8 | 86.8 |
| 3 | Excitement,Mutism,Negativism | 83.2 | 79.6 | 86.5 |
| 3 | Immobility/stupor,Echopraxia/echolalia,Stereotypy | 83.2 | 79.6 | 86.5 |
| 3 | Immobility/stupor,Grimacing,Mannerisms | 83.2 | 79.6 | 86.5 |
| 3 | Immobility/stupor,Mutism,Echopraxia/echolalia | 83.2 | 79.6 | 86.5 |
| 3 | Immobility/stupor,Mutism,Posturing/catalepsy | 83.2 | 79.6 | 86.6 |
| 3 | Immobility/stupor,Stereotypy,Mannerisms | 83.2 | 79.6 | 86.5 |
| 3 | Posturing/catalepsy,Negativism,Withdrawal | 83.2 | 79.6 | 86.5 |
| 3 | Excitement,Posturing/catalepsy,Withdrawal | 83 | 79.4 | 86.3 |
| 3 | Immobility/stupor,Mutism,Withdrawal | 83 | 79.4 | 86.3 |
| 3 | Mutism,Posturing/catalepsy,Withdrawal | 83 | 79.4 | 86.3 |
| 3 | Immobility/stupor,Grimacing,Echopraxia/echolalia | 82.9 | 79.4 | 86.3 |
| 3 | Immobility/stupor,Mutism,Grimacing | 82.7 | 79.1 | 86.1 |
| 3 | Immobility/stupor,Posturing/catalepsy,Echopraxia/echolalia | 82.7 | 79.1 | 86.3 |
| 3 | Immobility/stupor,Mannerisms,Verbigeration | 82.5 | 78.9 | 85.9 |
| 3 | Mutism,Posturing/catalepsy,Rigidity | 82.5 | 78.9 | 85.9 |
| 3 | Immobility/stupor,Rigidity,Waxy flexibility | 82.3 | 78.7 | 85.7 |
| 3 | Mutism,Negativism,Withdrawal | 82.3 | 78.7 | 85.7 |
| 3 | Immobility/stupor,Echopraxia/echolalia,Verbigeration | 82.1 | 78.5 | 85.7 |
| 3 | Immobility/stupor,Stereotypy,Waxy flexibility | 81.9 | 78.3 | 85.4 |
| 3 | Immobility/stupor,Waxy flexibility,Withdrawal | 81.8 | 78.3 | 85.4 |
| 3 | Mutism,Stereotypy,Negativism | 81.8 | 78.3 | 85.4 |
| 3 | Immobility/stupor,Echopraxia/echolalia,Mannerisms | 81.6 | 78 | 85.2 |
| 3 | Mutism,Mannerisms,Rigidity | 81.6 | 78 | 85.2 |
| 3 | Mutism,Stereotypy,Rigidity | 81.6 | 77.8 | 85.2 |
| 3 | Rigidity,Negativism,Withdrawal | 81.6 | 78 | 85.2 |
| 3 | Immobility/stupor,Grimacing,Waxy flexibility | 81.2 | 77.6 | 84.8 |
| 3 | Posturing/catalepsy,Rigidity,Withdrawal | 81.2 | 77.6 | 84.8 |
| 3 | Mutism,Verbigeration,Rigidity | 81 | 77.1 | 84.5 |
| 3 | Mutism,Echopraxia/echolalia,Rigidity | 80.7 | 76.9 | 84.3 |
| 3 | Immobility/stupor,Verbigeration,Waxy flexibility | 80.5 | 76.9 | 84.3 |
| 3 | Mutism,Grimacing,Rigidity | 80.5 | 76.9 | 84.1 |
| 3 | Mutism,Mannerisms,Negativism | 80.5 | 76.7 | 84.1 |
| 3 | Excitement,Rigidity,Withdrawal | 80.3 | 76.5 | 83.9 |
| 3 | Immobility/stupor,Mutism,Waxy flexibility | 80.1 | 76.2 | 83.6 |
| 3 | Mutism,Echopraxia/echolalia,Negativism | 80.1 | 76.2 | 83.6 |
| 3 | Mutism,Posturing/catalepsy,Verbigeration | 80 | 76.2 | 83.6 |
| 3 | Mutism,Stereotypy,Withdrawal | 80 | 76.2 | 83.6 |
| 3 | Immobility/stupor,Echopraxia/echolalia,Waxy flexibility | 79.8 | 76 | 83.4 |
| 3 | Mutism,Posturing/catalepsy,Echopraxia/echolalia | 79.8 | 76 | 83.4 |
| 3 | Mutism,Posturing/catalepsy,Stereotypy | 79.8 | 76 | 83.4 |
| 3 | Stereotypy,Negativism,Withdrawal | 79.8 | 76 | 83.4 |
| 3 | Excitement,Mutism,Stereotypy | 79.6 | 75.8 | 83.2 |
| 3 | Excitement,Mutism,Waxy flexibility | 79.6 | 75.8 | 83.2 |
| 3 | Mutism,Posturing/catalepsy,Mannerisms | 79.6 | 75.8 | 83.2 |
| 3 | Mutism,Verbigeration,Negativism | 79.6 | 75.8 | 83.2 |
| 3 | Immobility/stupor,Mannerisms,Waxy flexibility | 79.4 | 75.6 | 83 |
| 3 | Mutism,Negativism,Waxy flexibility | 79.4 | 75.6 | 83.2 |
| 3 | Mutism,Verbigeration,Withdrawal | 79.4 | 75.6 | 83.2 |
| 3 | Mutism,Mannerisms,Withdrawal | 79.3 | 75.6 | 83 |
| 3 | Excitement,Mutism,Echopraxia/echolalia | 79.2 | 75.3 | 83 |
| 3 | Mutism,Grimacing,Withdrawal | 79.2 | 75.3 | 83 |
| 3 | Mutism,Rigidity,Waxy flexibility | 79.2 | 75.3 | 83 |
| 3 | Mutism,Waxy flexibility,Withdrawal | 79.2 | 75.3 | 83 |
| 3 | Stereotypy,Rigidity,Withdrawal | 79.2 | 75.3 | 83 |
| 3 | Mutism,Posturing/catalepsy,Grimacing | 79.1 | 75.3 | 82.7 |
| 3 | Excitement,Mutism,Grimacing | 78.9 | 75.1 | 82.5 |
| 3 | Immobility/stupor,Posturing/catalepsy,Waxy flexibility | 78.9 | 75.1 | 82.7 |
| 3 | Mutism,Echopraxia/echolalia,Withdrawal | 78.9 | 75.1 | 82.7 |
| 3 | Posturing/catalepsy,Rigidity,Negativism | 78.9 | 75.1 | 82.7 |
| 3 | Excitement,Negativism,Withdrawal | 78.7 | 74.9 | 82.3 |
| 3 | Mutism,Grimacing,Negativism | 78.7 | 74.9 | 82.3 |
| 3 | Posturing/catalepsy,Mannerisms,Withdrawal | 78.7 | 74.9 | 82.5 |
| 3 | Posturing/catalepsy,Stereotypy,Withdrawal | 78.7 | 74.7 | 82.5 |
| 3 | Excitement,Posturing/catalepsy,Negativism | 78.5 | 74.7 | 82.3 |
| 3 | Posturing/catalepsy,Grimacing,Withdrawal | 78.3 | 74.2 | 82.1 |
| 3 | Posturing/catalepsy,Verbigeration,Withdrawal | 77.8 | 74 | 81.6 |
| 3 | Verbigeration,Rigidity,Withdrawal | 77.4 | 73.3 | 81.2 |
| 3 | Posturing/catalepsy,Echopraxia/echolalia,Withdrawal | 77.3 | 73.3 | 81.2 |
| 3 | Mannerisms,Rigidity,Withdrawal | 77.2 | 73.1 | 80.9 |
| 3 | Mutism,Posturing/catalepsy,Waxy flexibility | 77.2 | 73.1 | 80.9 |
| 3 | Excitement,Mutism,Verbigeration | 77.1 | 73.3 | 80.9 |
| 3 | Posturing/catalepsy,Stereotypy,Negativism | 77.1 | 73.1 | 80.9 |
| 3 | Echopraxia/echolalia,Rigidity,Withdrawal | 76.9 | 72.9 | 80.7 |
| 3 | Mannerisms,Negativism,Withdrawal | 76.9 | 73.1 | 80.7 |
| 3 | Echopraxia/echolalia,Negativism,Withdrawal | 76.7 | 72.9 | 80.5 |
| 3 | Verbigeration,Negativism,Withdrawal | 76.7 | 72.9 | 80.5 |
| 3 | Grimacing,Rigidity,Withdrawal | 76.2 | 72.2 | 80.3 |
| 3 | Negativism,Waxy flexibility,Withdrawal | 76.2 | 72.4 | 80 |
| 3 | Excitement,Mutism,Mannerisms | 76 | 72 | 79.8 |
| 3 | Excitement,Posturing/catalepsy,Rigidity | 76 | 72 | 79.8 |
| 3 | Mutism,Grimacing,Stereotypy | 75.8 | 71.7 | 79.8 |
| 3 | Mutism,Stereotypy,Waxy flexibility | 75.8 | 71.7 | 79.8 |
| 3 | Posturing/catalepsy,Grimacing,Negativism | 75.8 | 71.7 | 79.6 |
| 3 | Grimacing,Negativism,Withdrawal | 75.6 | 71.5 | 79.4 |
| 3 | Mutism,Echopraxia/echolalia,Stereotypy | 75.5 | 71.5 | 79.4 |
| 3 | Mutism,Grimacing,Verbigeration | 75.3 | 71.3 | 79.1 |
| 3 | Mutism,Stereotypy,Mannerisms | 75.3 | 71.3 | 79.4 |
| 3 | Mutism,Stereotypy,Verbigeration | 75.3 | 71.3 | 79.1 |
| 3 | Posturing/catalepsy,Mannerisms,Negativism | 75.3 | 71.3 | 79.4 |
| 3 | Posturing/catalepsy,Waxy flexibility,Withdrawal | 75.3 | 71.3 | 79.4 |
| 3 | Rigidity,Waxy flexibility,Withdrawal | 75.3 | 71.3 | 79.4 |
| 3 | Mutism,Echopraxia/echolalia,Waxy flexibility | 74.9 | 70.9 | 78.9 |
| 3 | Mutism,Echopraxia/echolalia,Mannerisms | 74.7 | 70.6 | 78.7 |
| 3 | Mutism,Grimacing,Echopraxia/echolalia | 74.7 | 70.6 | 78.7 |
| 3 | Posturing/catalepsy,Echopraxia/echolalia,Negativism | 74.7 | 70.6 | 78.7 |
| 3 | Stereotypy,Rigidity,Negativism | 74.6 | 70.6 | 78.7 |
| 3 | Mutism,Grimacing,Mannerisms | 74.5 | 70.2 | 78.5 |
| 3 | Posturing/catalepsy,Verbigeration,Negativism | 74.5 | 70.4 | 78.5 |
| 3 | Excitement,Waxy flexibility,Withdrawal | 74.4 | 70.4 | 78.5 |
| 3 | Excitement,Stereotypy,Withdrawal | 74.2 | 70.2 | 78.3 |
| 3 | Mutism,Grimacing,Waxy flexibility | 74.2 | 70.2 | 78.3 |
| 3 | Excitement,Echopraxia/echolalia,Withdrawal | 74 | 70 | 78 |
| 3 | Mutism,Verbigeration,Waxy flexibility | 74 | 70 | 78 |
| 3 | Mutism,Echopraxia/echolalia,Verbigeration | 73.8 | 69.7 | 77.8 |
| 3 | Mutism,Mannerisms,Waxy flexibility | 73.5 | 69.3 | 77.6 |
| 3 | Mutism,Mannerisms,Verbigeration | 73.1 | 69.1 | 77.1 |
| 3 | Excitement,Rigidity,Negativism | 72.9 | 68.6 | 76.9 |
| 3 | Posturing/catalepsy,Negativism,Waxy flexibility | 72.9 | 68.6 | 76.9 |
| 3 | Excitement,Verbigeration,Withdrawal | 72.4 | 68.4 | 76.5 |
| 3 | Grimacing,Stereotypy,Withdrawal | 72.4 | 68.4 | 76.5 |
| 3 | Stereotypy,Waxy flexibility,Withdrawal | 72.4 | 68.4 | 76.5 |
| 3 | Excitement,Grimacing,Withdrawal | 72.2 | 67.9 | 76.2 |
| 3 | Mannerisms,Rigidity,Negativism | 72.2 | 67.9 | 76.2 |
| 3 | Posturing/catalepsy,Grimacing,Rigidity | 72.2 | 67.9 | 76.2 |
| 3 | Grimacing,Rigidity,Negativism | 72 | 67.9 | 76 |
| 3 | Posturing/catalepsy,Mannerisms,Rigidity | 71.8 | 67.5 | 76 |
| 3 | Posturing/catalepsy,Stereotypy,Rigidity | 71.8 | 67.5 | 76 |
| 3 | Echopraxia/echolalia,Stereotypy,Withdrawal | 71.5 | 67.3 | 75.6 |
| 3 | Excitement,Posturing/catalepsy,Grimacing | 71.5 | 67.3 | 75.8 |
| 3 | Posturing/catalepsy,Echopraxia/echolalia,Rigidity | 71 | 66.8 | 75.3 |
| 3 | Posturing/catalepsy,Verbigeration,Rigidity | 70.9 | 66.6 | 75.1 |
| 3 | Stereotypy,Mannerisms,Withdrawal | 70.9 | 66.6 | 75.1 |
| 3 | Stereotypy,Verbigeration,Withdrawal | 70.9 | 66.6 | 75.1 |
| 3 | Echopraxia/echolalia,Rigidity,Negativism | 70.8 | 66.6 | 75.1 |
| 3 | Excitement,Mannerisms,Withdrawal | 70.4 | 65.9 | 74.7 |
| 3 | Verbigeration,Rigidity,Negativism | 70.4 | 66.1 | 74.4 |
| 3 | Grimacing,Echopraxia/echolalia,Withdrawal | 70.2 | 65.9 | 74.4 |
| 3 | Grimacing,Verbigeration,Withdrawal | 70.2 | 65.9 | 74.4 |
| 3 | Rigidity,Negativism,Waxy flexibility | 70.2 | 65.9 | 74.4 |
| 3 | Verbigeration,Waxy flexibility,Withdrawal | 70 | 65.7 | 74.2 |
| 3 | Echopraxia/echolalia,Waxy flexibility,Withdrawal | 69.7 | 65.5 | 74 |
| 3 | Excitement,Posturing/catalepsy,Echopraxia/echolalia | 69.5 | 65.2 | 73.8 |
| 3 | Grimacing,Waxy flexibility,Withdrawal | 69.3 | 65 | 73.5 |
| 3 | Grimacing,Mannerisms,Withdrawal | 69.2 | 65 | 73.5 |
| 3 | Mannerisms,Verbigeration,Withdrawal | 69.1 | 64.8 | 73.3 |
| 3 | Echopraxia/echolalia,Verbigeration,Withdrawal | 69 | 64.8 | 73.1 |
| 3 | Excitement,Posturing/catalepsy,Stereotypy | 69 | 64.8 | 73.3 |
| 3 | Mannerisms,Waxy flexibility,Withdrawal | 68.9 | 64.3 | 73.1 |
| 3 | Echopraxia/echolalia,Mannerisms,Withdrawal | 68.6 | 64.3 | 72.9 |
| 3 | Excitement,Posturing/catalepsy,Verbigeration | 68.2 | 63.9 | 72.4 |
| 3 | Stereotypy,Negativism,Waxy flexibility | 68.2 | 63.7 | 72.2 |
| 3 | Posturing/catalepsy,Grimacing,Stereotypy | 67.7 | 63.5 | 72 |
| 3 | Posturing/catalepsy,Rigidity,Waxy flexibility | 67.7 | 63.5 | 72 |
| 3 | Excitement,Posturing/catalepsy,Waxy flexibility | 67.5 | 63 | 71.7 |
| 3 | Excitement,Stereotypy,Negativism | 67.5 | 63 | 71.7 |
| 3 | Grimacing,Stereotypy,Negativism | 67 | 62.6 | 71.3 |
| 3 | Posturing/catalepsy,Grimacing,Verbigeration | 67 | 62.6 | 71.5 |
| 3 | Posturing/catalepsy,Grimacing,Echopraxia/echolalia | 66.6 | 62.3 | 70.9 |
| 3 | Excitement,Posturing/catalepsy,Mannerisms | 66.4 | 61.9 | 70.9 |
| 3 | Grimacing,Stereotypy,Rigidity | 66.4 | 61.9 | 70.6 |
| 3 | Stereotypy,Mannerisms,Negativism | 66.4 | 62.1 | 70.6 |
| 3 | Excitement,Stereotypy,Rigidity | 66.3 | 61.9 | 70.6 |
| 3 | Excitement,Grimacing,Rigidity | 66.2 | 61.7 | 70.6 |
| 3 | Echopraxia/echolalia,Stereotypy,Negativism | 66.1 | 61.9 | 70.6 |
| 3 | Posturing/catalepsy,Grimacing,Mannerisms | 65.9 | 61.4 | 70.4 |
| 3 | Excitement,Grimacing,Negativism | 65.7 | 61.2 | 70.2 |
| 3 | Excitement,Negativism,Waxy flexibility | 65.7 | 61.2 | 70.2 |
| 3 | Posturing/catalepsy,Stereotypy,Verbigeration | 65.5 | 61 | 69.7 |
| 3 | Stereotypy,Verbigeration,Negativism | 65.5 | 61 | 69.7 |
| 3 | Excitement,Echopraxia/echolalia,Rigidity | 65.2 | 60.8 | 69.7 |
| 3 | Posturing/catalepsy,Echopraxia/echolalia,Stereotypy | 65 | 60.5 | 69.3 |
| 3 | Excitement,Echopraxia/echolalia,Negativism | 64.8 | 60.3 | 69.3 |
| 3 | Grimacing,Echopraxia/echolalia,Rigidity | 64.4 | 59.9 | 68.8 |
| 3 | Grimacing,Negativism,Waxy flexibility | 64.3 | 59.9 | 68.6 |
| 3 | Echopraxia/echolalia,Stereotypy,Rigidity | 63.9 | 59.4 | 68.4 |
| 3 | Grimacing,Echopraxia/echolalia,Negativism | 63.9 | 59.4 | 68.4 |
| 3 | Grimacing,Verbigeration,Rigidity | 63.9 | 59.4 | 68.4 |
| 3 | Posturing/catalepsy,Grimacing,Waxy flexibility | 63.9 | 59.4 | 68.4 |
| 3 | Posturing/catalepsy,Stereotypy,Mannerisms | 63.9 | 59.4 | 68.2 |
| 3 | Excitement,Rigidity,Waxy flexibility | 63.7 | 59.2 | 68.2 |
| 3 | Stereotypy,Rigidity,Waxy flexibility | 63.7 | 59.2 | 68.2 |
| 3 | Posturing/catalepsy,Stereotypy,Waxy flexibility | 63.5 | 59 | 67.9 |
| 3 | Grimacing,Mannerisms,Negativism | 63.4 | 59 | 67.9 |
| 3 | Mannerisms,Negativism,Waxy flexibility | 63.4 | 59 | 67.7 |
| 3 | Stereotypy,Verbigeration,Rigidity | 63.3 | 58.7 | 67.7 |
| 3 | Grimacing,Mannerisms,Rigidity | 63.2 | 58.7 | 67.7 |
| 3 | Grimacing,Rigidity,Waxy flexibility | 63.2 | 58.7 | 67.7 |
| 3 | Echopraxia/echolalia,Negativism,Waxy flexibility | 63 | 58.5 | 67.5 |
| 3 | Grimacing,Verbigeration,Negativism | 63 | 58.5 | 67.5 |
| 3 | Stereotypy,Mannerisms,Rigidity | 63 | 58.5 | 67.5 |
| 3 | Posturing/catalepsy,Echopraxia/echolalia,Verbigeration | 62.8 | 58.3 | 67 |
| 3 | Posturing/catalepsy,Mannerisms,Verbigeration | 62.6 | 58.1 | 67 |
| 3 | Posturing/catalepsy,Echopraxia/echolalia,Mannerisms | 62.5 | 58.1 | 67 |
| 3 | Echopraxia/echolalia,Mannerisms,Negativism | 62.1 | 57.6 | 66.6 |
| 3 | Excitement,Verbigeration,Rigidity | 62.1 | 57.6 | 66.6 |
| 3 | Excitement,Verbigeration,Negativism | 61.5 | 57 | 66.1 |
| 3 | Verbigeration,Negativism,Waxy flexibility | 61.4 | 57 | 65.9 |
| 3 | Excitement,Mannerisms,Negativism | 61.2 | 56.7 | 65.7 |
| 3 | Posturing/catalepsy,Verbigeration,Waxy flexibility | 60.8 | 56.3 | 65.2 |
| 3 | Echopraxia/echolalia,Verbigeration,Negativism | 60.6 | 56.1 | 65 |
| 3 | Posturing/catalepsy,Echopraxia/echolalia,Waxy flexibility | 60.6 | 56.1 | 65 |
| 3 | Excitement,Mannerisms,Rigidity | 60.5 | 56.1 | 65 |
| 3 | Echopraxia/echolalia,Mannerisms,Rigidity | 60.1 | 55.6 | 64.6 |
| 3 | Echopraxia/echolalia,Rigidity,Waxy flexibility | 60.1 | 55.6 | 64.6 |
| 3 | Mannerisms,Verbigeration,Negativism | 59.9 | 55.2 | 64.3 |
| 3 | Posturing/catalepsy,Mannerisms,Waxy flexibility | 59.9 | 55.2 | 64.3 |
| 3 | Echopraxia/echolalia,Verbigeration,Rigidity | 59.8 | 55.2 | 64.3 |
| 3 | Verbigeration,Rigidity,Waxy flexibility | 59.4 | 54.9 | 63.9 |
| 3 | Mannerisms,Rigidity,Waxy flexibility | 59 | 54.3 | 63.7 |
| 3 | Mannerisms,Verbigeration,Rigidity | 59 | 54.3 | 63.5 |
| 3 | Grimacing,Stereotypy,Waxy flexibility | 55.6 | 50.9 | 60.1 |
| 3 | Excitement,Grimacing,Waxy flexibility | 55.2 | 50.4 | 59.6 |
| 3 | Excitement,Stereotypy,Waxy flexibility | 55.2 | 50.7 | 59.6 |
| 3 | Excitement,Grimacing,Stereotypy | 53.6 | 48.9 | 58.3 |
| 3 | Excitement,Grimacing,Echopraxia/echolalia | 52.7 | 48.2 | 57.4 |
| 3 | Grimacing,Echopraxia/echolalia,Stereotypy | 52.7 | 48.2 | 57.4 |
| 3 | Grimacing,Echopraxia/echolalia,Waxy flexibility | 52.3 | 47.5 | 57 |
| 3 | Echopraxia/echolalia,Stereotypy,Waxy flexibility | 52 | 47.3 | 56.7 |
| 3 | Excitement,Echopraxia/echolalia,Waxy flexibility | 52 | 47.3 | 56.5 |
| 3 | Grimacing,Stereotypy,Verbigeration | 51.8 | 47.1 | 56.5 |
| 3 | Grimacing,Verbigeration,Waxy flexibility | 51.8 | 47.1 | 56.5 |
| 3 | Stereotypy,Verbigeration,Waxy flexibility | 51.1 | 46.6 | 55.8 |
| 3 | Excitement,Echopraxia/echolalia,Stereotypy | 50.9 | 46.2 | 55.6 |
| 3 | Grimacing,Stereotypy,Mannerisms | 50.7 | 46 | 55.2 |
| 3 | Excitement,Grimacing,Verbigeration | 50.3 | 45.7 | 54.7 |
| 3 | Stereotypy,Mannerisms,Waxy flexibility | 50.2 | 45.5 | 54.9 |
| 3 | Excitement,Verbigeration,Waxy flexibility | 50 | 45.3 | 54.7 |
| 3 | Grimacing,Echopraxia/echolalia,Verbigeration | 49.3 | 44.6 | 54 |
| 3 | Grimacing,Mannerisms,Waxy flexibility | 49.3 | 44.8 | 54 |
| 3 | Excitement,Stereotypy,Verbigeration | 48.9 | 44.4 | 53.4 |
| 3 | Grimacing,Echopraxia/echolalia,Mannerisms | 47.3 | 42.6 | 51.8 |
| 3 | Excitement,Stereotypy,Mannerisms | 46.9 | 42.4 | 51.3 |
| 3 | Excitement,Grimacing,Mannerisms | 46.8 | 42.2 | 51.6 |
| 3 | Echopraxia/echolalia,Stereotypy,Verbigeration | 46.6 | 41.9 | 51.3 |
| 3 | Grimacing,Mannerisms,Verbigeration | 46.6 | 41.9 | 51.3 |
| 3 | Echopraxia/echolalia,Stereotypy,Mannerisms | 46.4 | 41.7 | 51.1 |
| 3 | Excitement,Echopraxia/echolalia,Verbigeration | 46.2 | 41.5 | 50.9 |
| 3 | Excitement,Mannerisms,Waxy flexibility | 46.2 | 41.7 | 50.9 |
| 3 | Echopraxia/echolalia,Verbigeration,Waxy flexibility | 45.9 | 41.3 | 50.7 |
| 3 | Stereotypy,Mannerisms,Verbigeration | 45.8 | 41 | 50.4 |
| 3 | Mannerisms,Verbigeration,Waxy flexibility | 44.4 | 39.7 | 49.1 |
| 3 | Echopraxia/echolalia,Mannerisms,Waxy flexibility | 44.2 | 39.7 | 48.9 |
| 3 | Excitement,Echopraxia/echolalia,Mannerisms | 42.8 | 38.3 | 47.3 |
| 3 | Excitement,Mannerisms,Verbigeration | 41.2 | 36.8 | 46 |
| 3 | Echopraxia/echolalia,Mannerisms,Verbigeration | 40.6 | 36.1 | 45.1 |

Table S4: sensitivity (relative to the full BFCSI) of all four question combinations, and the 95% confidence interval derived from bootstrapping, sorted by decreasing sensitivity.

|  |  |  | 95% CI | |
| --- | --- | --- | --- | --- |
| # of Qs | Combination | Sensitivity | Lower | Upper |
| 4 | Excitement,Mutism,Staring,Negativism | 97.3 | 95.7 | 98.7 |
| 4 | Excitement,Mutism,Staring,Rigidity | 97.1 | 95.5 | 98.4 |
| 4 | Excitement,Immobility/stupor,Staring,Rigidity | 96.9 | 95.1 | 98.4 |
| 4 | Excitement,Mutism,Staring,Posturing/catalepsy | 96.9 | 95.1 | 98.4 |
| 4 | Excitement,Staring,Rigidity,Withdrawal | 96.9 | 95.3 | 98.4 |
| 4 | Excitement,Immobility/stupor,Staring,Negativism | 96.6 | 94.8 | 98.2 |
| 4 | Excitement,Mutism,Staring,Echopraxia/echolalia | 96.4 | 94.6 | 98.0 |
| 4 | Excitement,Mutism,Staring,Withdrawal | 96.4 | 94.6 | 98.0 |
| 4 | Excitement,Mutism,Staring,Waxy flexibility | 96.2 | 94.4 | 97.8 |
| 4 | Excitement,Staring,Negativism,Withdrawal | 96.2 | 94.4 | 98.0 |
| 4 | Excitement,Mutism,Staring,Stereotypy | 96.0 | 93.9 | 97.8 |
| 4 | Staring,Mannerisms,Negativism,Withdrawal | 95.8 | 93.7 | 97.5 |
| 4 | Excitement,Immobility/stupor,Rigidity,Withdrawal | 95.7 | 93.7 | 97.5 |
| 4 | Excitement,Immobility/stupor,Staring,Posturing/catalepsy | 95.7 | 93.7 | 97.5 |
| 4 | Excitement,Staring,Posturing/catalepsy,Negativism | 95.7 | 93.7 | 97.5 |
| 4 | Excitement,Staring,Posturing/catalepsy,Withdrawal | 95.7 | 93.7 | 97.5 |
| 4 | Excitement,Staring,Rigidity,Negativism | 95.7 | 93.7 | 97.5 |
| 4 | Staring,Rigidity,Negativism,Withdrawal | 95.7 | 93.9 | 97.5 |
| 4 | Excitement,Immobility/stupor,Mutism,Rigidity | 95.5 | 93.5 | 97.3 |
| 4 | Excitement,Immobility/stupor,Mutism,Staring | 95.5 | 93.5 | 97.3 |
| 4 | Excitement,Immobility/stupor,Rigidity,Negativism | 95.5 | 93.5 | 97.3 |
| 4 | Excitement,Immobility/stupor,Staring,Echopraxia/echolalia | 95.5 | 93.5 | 97.3 |
| 4 | Excitement,Immobility/stupor,Staring,Withdrawal | 95.5 | 93.5 | 97.3 |
| 4 | Excitement,Mutism,Staring,Grimacing | 95.5 | 93.5 | 97.3 |
| 4 | Excitement,Mutism,Staring,Verbigeration | 95.5 | 93.5 | 97.3 |
| 4 | Mutism,Staring,Mannerisms,Negativism | 95.5 | 93.5 | 97.3 |
| 4 | Excitement,Immobility/stupor,Staring,Verbigeration | 95.3 | 93.3 | 97.1 |
| 4 | Excitement,Immobility/stupor,Staring,Waxy flexibility | 95.3 | 93.3 | 97.1 |
| 4 | Excitement,Mutism,Staring,Mannerisms | 95.3 | 93.3 | 97.1 |
| 4 | Excitement,Staring,Posturing/catalepsy,Rigidity | 95.3 | 93.3 | 97.1 |
| 4 | Excitement,Staring,Waxy flexibility,Withdrawal | 95.3 | 93.3 | 97.1 |
| 4 | Staring,Mannerisms,Rigidity,Withdrawal | 95.3 | 93.3 | 97.1 |
| 4 | Excitement,Immobility/stupor,Echopraxia/echolalia,Rigidity | 95.1 | 93.0 | 97.1 |
| 4 | Excitement,Immobility/stupor,Posturing/catalepsy,Negativism | 95.1 | 93.0 | 97.1 |
| 4 | Excitement,Staring,Echopraxia/echolalia,Withdrawal | 95.1 | 93.0 | 96.9 |
| 4 | Excitement,Staring,Negativism,Waxy flexibility | 95.1 | 93.0 | 97.1 |
| 4 | Mutism,Staring,Mannerisms,Rigidity | 95.1 | 92.8 | 97.1 |
| 4 | Mutism,Staring,Negativism,Withdrawal | 95.1 | 93.0 | 96.9 |
| 4 | Staring,Mannerisms,Rigidity,Negativism | 95.1 | 93.0 | 97.1 |
| 4 | Staring,Posturing/catalepsy,Negativism,Withdrawal | 95.1 | 93.0 | 97.1 |
| 4 | Excitement,Immobility/stupor,Posturing/catalepsy,Rigidity | 94.9 | 92.6 | 96.9 |
| 4 | Excitement,Immobility/stupor,Staring,Mannerisms | 94.9 | 92.8 | 96.9 |
| 4 | Excitement,Immobility/stupor,Staring,Stereotypy | 94.9 | 92.8 | 96.9 |
| 4 | Excitement,Staring,Echopraxia/echolalia,Negativism | 94.9 | 92.6 | 96.9 |
| 4 | Excitement,Staring,Grimacing,Rigidity | 94.9 | 92.8 | 96.9 |
| 4 | Excitement,Staring,Stereotypy,Rigidity | 94.9 | 92.6 | 96.9 |
| 4 | Mutism,Staring,Mannerisms,Withdrawal | 94.9 | 92.6 | 96.9 |
| 4 | Excitement,Staring,Echopraxia/echolalia,Rigidity | 94.8 | 92.8 | 96.9 |
| 4 | Excitement,Staring,Grimacing,Withdrawal | 94.8 | 92.6 | 96.9 |
| 4 | Excitement,Staring,Rigidity,Waxy flexibility | 94.8 | 92.6 | 96.9 |
| 4 | Immobility/stupor,Staring,Mannerisms,Negativism | 94.8 | 92.8 | 96.9 |
| 4 | Mutism,Staring,Stereotypy,Negativism | 94.8 | 92.6 | 96.9 |
| 4 | Staring,Stereotypy,Negativism,Withdrawal | 94.8 | 92.8 | 96.9 |
| 4 | Excitement,Immobility/stupor,Staring,Grimacing | 94.6 | 92.4 | 96.6 |
| 4 | Excitement,Staring,Mannerisms,Withdrawal | 94.6 | 92.4 | 96.6 |
| 4 | Excitement,Staring,Stereotypy,Withdrawal | 94.6 | 92.4 | 96.6 |
| 4 | Mutism,Staring,Rigidity,Negativism | 94.6 | 92.4 | 96.6 |
| 4 | Mutism,Staring,Rigidity,Withdrawal | 94.6 | 92.4 | 96.6 |
| 4 | Staring,Stereotypy,Rigidity,Withdrawal | 94.6 | 92.4 | 96.6 |
| 4 | Excitement,Immobility/stupor,Grimacing,Rigidity | 94.4 | 92.2 | 96.4 |
| 4 | Excitement,Immobility/stupor,Verbigeration,Rigidity | 94.4 | 92.2 | 96.4 |
| 4 | Excitement,Staring,Mannerisms,Rigidity | 94.4 | 92.2 | 96.4 |
| 4 | Excitement,Staring,Stereotypy,Negativism | 94.4 | 92.2 | 96.4 |
| 4 | Immobility/stupor,Staring,Mannerisms,Rigidity | 94.4 | 92.2 | 96.4 |
| 4 | Mutism,Staring,Echopraxia/echolalia,Negativism | 94.4 | 92.2 | 96.4 |
| 4 | Mutism,Staring,Posturing/catalepsy,Negativism | 94.4 | 92.2 | 96.4 |
| 4 | Mutism,Staring,Posturing/catalepsy,Withdrawal | 94.4 | 92.2 | 96.4 |
| 4 | Mutism,Staring,Verbigeration,Negativism | 94.4 | 92.2 | 96.4 |
| 4 | Mutism,Staring,Verbigeration,Rigidity | 94.4 | 92.2 | 96.4 |
| 4 | Staring,Echopraxia/echolalia,Negativism,Withdrawal | 94.4 | 92.2 | 96.4 |
| 4 | Staring,Stereotypy,Rigidity,Negativism | 94.4 | 92.2 | 96.4 |
| 4 | Staring,Verbigeration,Negativism,Withdrawal | 94.4 | 92.2 | 96.4 |
| 4 | Excitement,Immobility/stupor,Mutism,Posturing/catalepsy | 94.2 | 91.9 | 96.2 |
| 4 | Excitement,Immobility/stupor,Posturing/catalepsy,Withdrawal | 94.2 | 91.9 | 96.2 |
| 4 | Excitement,Immobility/stupor,Stereotypy,Rigidity | 94.2 | 91.9 | 96.2 |
| 4 | Excitement,Staring,Verbigeration,Rigidity | 94.2 | 91.9 | 96.2 |
| 4 | Excitement,Staring,Verbigeration,Withdrawal | 94.2 | 91.9 | 96.2 |
| 4 | Immobility/stupor,Staring,Negativism,Withdrawal | 94.2 | 91.9 | 96.2 |
| 4 | Immobility/stupor,Staring,Stereotypy,Negativism | 94.2 | 91.9 | 96.2 |
| 4 | Mutism,Staring,Echopraxia/echolalia,Rigidity | 94.2 | 91.9 | 96.2 |
| 4 | Mutism,Staring,Stereotypy,Rigidity | 94.2 | 91.9 | 96.2 |
| 4 | Staring,Posturing/catalepsy,Mannerisms,Withdrawal | 94.2 | 91.9 | 96.2 |
| 4 | Excitement,Immobility/stupor,Negativism,Withdrawal | 94.0 | 91.7 | 96.0 |
| 4 | Excitement,Staring,Posturing/catalepsy,Grimacing | 94.0 | 91.7 | 96.0 |
| 4 | Immobility/stupor,Mannerisms,Rigidity,Negativism | 94.0 | 91.7 | 96.0 |
| 4 | Mutism,Staring,Posturing/catalepsy,Mannerisms | 94.0 | 91.7 | 96.2 |
| 4 | Staring,Grimacing,Mannerisms,Withdrawal | 94.0 | 91.7 | 96.0 |
| 4 | Staring,Negativism,Waxy flexibility,Withdrawal | 94.0 | 91.7 | 96.2 |
| 4 | Excitement,Immobility/stupor,Mannerisms,Rigidity | 93.9 | 91.7 | 96.0 |
| 4 | Excitement,Immobility/stupor,Rigidity,Waxy flexibility | 93.9 | 91.7 | 96.2 |
| 4 | Excitement,Staring,Verbigeration,Negativism | 93.9 | 91.7 | 96.2 |
| 4 | Immobility/stupor,Staring,Rigidity,Negativism | 93.9 | 91.7 | 96.0 |
| 4 | Staring,Echopraxia/echolalia,Rigidity,Withdrawal | 93.9 | 91.7 | 96.0 |
| 4 | Staring,Grimacing,Rigidity,Withdrawal | 93.9 | 91.7 | 96.2 |
| 4 | Staring,Posturing/catalepsy,Mannerisms,Negativism | 93.9 | 91.7 | 96.2 |
| 4 | Staring,Posturing/catalepsy,Rigidity,Withdrawal | 93.9 | 91.7 | 96.0 |
| 4 | Staring,Verbigeration,Rigidity,Withdrawal | 93.9 | 91.7 | 96.0 |
| 4 | Excitement,Immobility/stupor,Mutism,Negativism | 93.7 | 91.3 | 96.0 |
| 4 | Excitement,Staring,Mannerisms,Negativism | 93.7 | 91.5 | 96.0 |
| 4 | Excitement,Staring,Posturing/catalepsy,Echopraxia/echolalia | 93.7 | 91.5 | 96.0 |
| 4 | Immobility/stupor,Staring,Rigidity,Withdrawal | 93.7 | 91.5 | 96.0 |
| 4 | Mutism,Staring,Echopraxia/echolalia,Mannerisms | 93.7 | 91.3 | 96.0 |
| 4 | Mutism,Staring,Posturing/catalepsy,Echopraxia/echolalia | 93.7 | 91.5 | 96.0 |
| 4 | Mutism,Staring,Stereotypy,Mannerisms | 93.7 | 91.3 | 96.0 |
| 4 | Mutism,Staring,Stereotypy,Withdrawal | 93.7 | 91.3 | 96.0 |
| 4 | Staring,Echopraxia/echolalia,Rigidity,Negativism | 93.7 | 91.3 | 96.0 |
| 4 | Staring,Grimacing,Negativism,Withdrawal | 93.7 | 91.3 | 96.0 |
| 4 | Staring,Stereotypy,Mannerisms,Negativism | 93.7 | 91.5 | 96.0 |
| 4 | Staring,Stereotypy,Mannerisms,Withdrawal | 93.7 | 91.5 | 95.7 |
| 4 | Excitement,Immobility/stupor,Echopraxia/echolalia,Negativism | 93.5 | 91.0 | 95.7 |
| 4 | Excitement,Immobility/stupor,Echopraxia/echolalia,Withdrawal | 93.5 | 91.0 | 95.7 |
| 4 | Excitement,Immobility/stupor,Negativism,Waxy flexibility | 93.5 | 91.0 | 95.7 |
| 4 | Excitement,Immobility/stupor,Stereotypy,Negativism | 93.5 | 91.0 | 95.7 |
| 4 | Excitement,Staring,Grimacing,Negativism | 93.5 | 91.0 | 95.7 |
| 4 | Excitement,Staring,Posturing/catalepsy,Mannerisms | 93.5 | 91.0 | 95.7 |
| 4 | Excitement,Staring,Posturing/catalepsy,Verbigeration | 93.5 | 91.0 | 95.7 |
| 4 | Immobility/stupor,Staring,Mannerisms,Withdrawal | 93.5 | 91.0 | 95.7 |
| 4 | Mutism,Staring,Echopraxia/echolalia,Withdrawal | 93.5 | 91.0 | 95.7 |
| 4 | Mutism,Staring,Grimacing,Mannerisms | 93.5 | 91.0 | 95.7 |
| 4 | Mutism,Staring,Grimacing,Verbigeration | 93.5 | 91.0 | 95.7 |
| 4 | Mutism,Staring,Grimacing,Withdrawal | 93.5 | 91.0 | 95.7 |
| 4 | Mutism,Staring,Posturing/catalepsy,Stereotypy | 93.5 | 91.0 | 95.7 |
| 4 | Mutism,Staring,Posturing/catalepsy,Verbigeration | 93.5 | 91.0 | 95.7 |
| 4 | Mutism,Staring,Stereotypy,Verbigeration | 93.5 | 91.0 | 95.7 |
| 4 | Mutism,Staring,Verbigeration,Withdrawal | 93.5 | 91.0 | 95.7 |
| 4 | Staring,Echopraxia/echolalia,Mannerisms,Negativism | 93.5 | 91.0 | 95.7 |
| 4 | Staring,Grimacing,Mannerisms,Rigidity | 93.5 | 91.0 | 95.7 |
| 4 | Staring,Posturing/catalepsy,Stereotypy,Negativism | 93.5 | 91.0 | 95.7 |
| 4 | Excitement,Immobility/stupor,Mutism,Withdrawal | 93.3 | 90.8 | 95.5 |
| 4 | Excitement,Immobility/stupor,Posturing/catalepsy,Echopraxia/echolalia | 93.3 | 90.8 | 95.5 |
| 4 | Excitement,Immobility/stupor,Posturing/catalepsy,Grimacing | 93.3 | 90.8 | 95.5 |
| 4 | Excitement,Immobility/stupor,Verbigeration,Negativism | 93.3 | 90.8 | 95.5 |
| 4 | Excitement,Staring,Grimacing,Waxy flexibility | 93.3 | 90.8 | 95.5 |
| 4 | Excitement,Staring,Posturing/catalepsy,Stereotypy | 93.3 | 90.8 | 95.5 |
| 4 | Immobility/stupor,Staring,Echopraxia/echolalia,Negativism | 93.3 | 90.8 | 95.5 |
| 4 | Immobility/stupor,Staring,Stereotypy,Rigidity | 93.3 | 90.8 | 95.5 |
| 4 | Immobility/stupor,Staring,Verbigeration,Negativism | 93.3 | 90.8 | 95.5 |
| 4 | Immobility/stupor,Staring,Verbigeration,Rigidity | 93.3 | 90.8 | 95.5 |
| 4 | Mutism,Staring,Echopraxia/echolalia,Stereotypy | 93.3 | 90.8 | 95.5 |
| 4 | Mutism,Staring,Grimacing,Negativism | 93.3 | 90.8 | 95.5 |
| 4 | Mutism,Staring,Grimacing,Rigidity | 93.3 | 90.8 | 95.5 |
| 4 | Mutism,Staring,Grimacing,Stereotypy | 93.3 | 90.8 | 95.5 |
| 4 | Mutism,Staring,Mannerisms,Verbigeration | 93.3 | 90.8 | 95.5 |
| 4 | Mutism,Staring,Negativism,Waxy flexibility | 93.3 | 91.0 | 95.5 |
| 4 | Mutism,Staring,Posturing/catalepsy,Rigidity | 93.3 | 90.8 | 95.5 |
| 4 | Staring,Grimacing,Stereotypy,Withdrawal | 93.3 | 90.8 | 95.5 |
| 4 | Staring,Mannerisms,Negativism,Waxy flexibility | 93.3 | 90.8 | 95.5 |
| 4 | Staring,Mannerisms,Waxy flexibility,Withdrawal | 93.3 | 90.8 | 95.5 |
| 4 | Staring,Rigidity,Waxy flexibility,Withdrawal | 93.3 | 90.8 | 95.5 |
| 4 | Staring,Verbigeration,Rigidity,Negativism | 93.3 | 90.8 | 95.5 |
| 4 | Excitement,Immobility/stupor,Mutism,Echopraxia/echolalia | 93.1 | 90.6 | 95.3 |
| 4 | Staring,Posturing/catalepsy,Mannerisms,Rigidity | 93.1 | 90.6 | 95.3 |
| 4 | Staring,Stereotypy,Mannerisms,Rigidity | 93.1 | 90.6 | 95.3 |
| 4 | Excitement,Staring,Grimacing,Echopraxia/echolalia | 93.0 | 90.6 | 95.3 |
| 4 | Excitement,Staring,Posturing/catalepsy,Waxy flexibility | 93.0 | 90.6 | 95.3 |
| 4 | Immobility/stupor,Staring,Posturing/catalepsy,Negativism | 93.0 | 90.6 | 95.3 |
| 4 | Immobility/stupor,Stereotypy,Rigidity,Negativism | 93.0 | 90.6 | 95.3 |
| 4 | Mutism,Staring,Grimacing,Echopraxia/echolalia | 93.0 | 90.6 | 95.3 |
| 4 | Mutism,Staring,Waxy flexibility,Withdrawal | 93.0 | 90.6 | 95.3 |
| 4 | Staring,Echopraxia/echolalia,Mannerisms,Withdrawal | 93.0 | 90.6 | 95.3 |
| 4 | Staring,Grimacing,Stereotypy,Rigidity | 93.0 | 90.6 | 95.3 |
| 4 | Staring,Posturing/catalepsy,Rigidity,Negativism | 93.0 | 90.6 | 95.3 |
| 4 | Staring,Stereotypy,Verbigeration,Rigidity | 92.9 | 90.4 | 95.1 |
| 4 | Excitement,Immobility/stupor,Grimacing,Negativism | 92.8 | 90.4 | 95.1 |
| 4 | Excitement,Immobility/stupor,Mutism,Waxy flexibility | 92.8 | 90.4 | 95.1 |
| 4 | Excitement,Immobility/stupor,Posturing/catalepsy,Verbigeration | 92.8 | 90.4 | 95.1 |
| 4 | Excitement,Immobility/stupor,Waxy flexibility,Withdrawal | 92.8 | 90.4 | 95.1 |
| 4 | Excitement,Staring,Echopraxia/echolalia,Waxy flexibility | 92.8 | 90.4 | 95.1 |
| 4 | Excitement,Staring,Stereotypy,Waxy flexibility | 92.8 | 90.4 | 95.1 |
| 4 | Excitement,Staring,Verbigeration,Waxy flexibility | 92.8 | 90.4 | 95.1 |
| 4 | Immobility/stupor,Staring,Echopraxia/echolalia,Rigidity | 92.8 | 90.4 | 95.3 |
| 4 | Mutism,Staring,Echopraxia/echolalia,Verbigeration | 92.8 | 90.4 | 95.1 |
| 4 | Mutism,Staring,Mannerisms,Waxy flexibility | 92.8 | 90.4 | 95.1 |
| 4 | Staring,Echopraxia/echolalia,Stereotypy,Negativism | 92.8 | 90.4 | 95.3 |
| 4 | Staring,Posturing/catalepsy,Echopraxia/echolalia,Negativism | 92.8 | 90.4 | 95.1 |
| 4 | Staring,Posturing/catalepsy,Echopraxia/echolalia,Withdrawal | 92.8 | 90.4 | 95.1 |
| 4 | Staring,Posturing/catalepsy,Grimacing,Withdrawal | 92.8 | 90.4 | 95.1 |
| 4 | Staring,Posturing/catalepsy,Stereotypy,Withdrawal | 92.8 | 90.4 | 95.1 |
| 4 | Staring,Stereotypy,Verbigeration,Negativism | 92.8 | 90.4 | 95.1 |
| 4 | Excitement,Immobility/stupor,Mutism,Stereotypy | 92.6 | 90.1 | 94.8 |
| 4 | Excitement,Immobility/stupor,Mutism,Verbigeration | 92.6 | 90.1 | 94.8 |
| 4 | Excitement,Immobility/stupor,Stereotypy,Withdrawal | 92.6 | 90.1 | 94.8 |
| 4 | Excitement,Immobility/stupor,Verbigeration,Withdrawal | 92.6 | 90.1 | 94.8 |
| 4 | Excitement,Staring,Grimacing,Stereotypy | 92.6 | 90.1 | 94.8 |
| 4 | Immobility/stupor,Rigidity,Negativism,Withdrawal | 92.6 | 90.1 | 94.8 |
| 4 | Immobility/stupor,Staring,Mannerisms,Verbigeration | 92.6 | 90.1 | 94.8 |
| 4 | Immobility/stupor,Staring,Stereotypy,Mannerisms | 92.6 | 90.1 | 94.8 |
| 4 | Mutism,Staring,Stereotypy,Waxy flexibility | 92.6 | 90.1 | 95.1 |
| 4 | Staring,Grimacing,Echopraxia/echolalia,Withdrawal | 92.6 | 90.1 | 94.8 |
| 4 | Staring,Grimacing,Rigidity,Negativism | 92.6 | 90.1 | 94.8 |
| 4 | Staring,Grimacing,Verbigeration,Rigidity | 92.6 | 90.1 | 95.1 |
| 4 | Staring,Grimacing,Verbigeration,Withdrawal | 92.6 | 90.1 | 94.8 |
| 4 | Staring,Mannerisms,Verbigeration,Withdrawal | 92.6 | 90.1 | 94.8 |
| 4 | Staring,Rigidity,Negativism,Waxy flexibility | 92.6 | 90.1 | 94.8 |
| 4 | Staring,Stereotypy,Negativism,Waxy flexibility | 92.6 | 90.1 | 94.8 |
| 4 | Excitement,Immobility/stupor,Mutism,Grimacing | 92.4 | 89.9 | 94.8 |
| 4 | Excitement,Staring,Echopraxia/echolalia,Stereotypy | 92.4 | 89.9 | 94.8 |
| 4 | Immobility/stupor,Mannerisms,Rigidity,Withdrawal | 92.4 | 89.9 | 94.6 |
| 4 | Immobility/stupor,Mutism,Staring,Mannerisms | 92.4 | 89.9 | 94.6 |
| 4 | Immobility/stupor,Staring,Echopraxia/echolalia,Mannerisms | 92.4 | 89.9 | 94.6 |
| 4 | Immobility/stupor,Staring,Grimacing,Mannerisms | 92.4 | 89.9 | 94.8 |
| 4 | Immobility/stupor,Staring,Grimacing,Negativism | 92.4 | 89.9 | 94.6 |
| 4 | Immobility/stupor,Staring,Grimacing,Rigidity | 92.4 | 89.9 | 94.8 |
| 4 | Immobility/stupor,Staring,Posturing/catalepsy,Mannerisms | 92.4 | 89.7 | 94.8 |
| 4 | Immobility/stupor,Staring,Posturing/catalepsy,Withdrawal | 92.4 | 89.7 | 94.6 |
| 4 | Immobility/stupor,Staring,Stereotypy,Verbigeration | 92.4 | 89.9 | 94.8 |
| 4 | Immobility/stupor,Verbigeration,Rigidity,Negativism | 92.4 | 89.9 | 94.8 |
| 4 | Mutism,Staring,Rigidity,Waxy flexibility | 92.4 | 89.9 | 94.8 |
| 4 | Mutism,Staring,Verbigeration,Waxy flexibility | 92.4 | 89.9 | 94.8 |
| 4 | Staring,Echopraxia/echolalia,Mannerisms,Rigidity | 92.4 | 89.9 | 94.8 |
| 4 | Staring,Echopraxia/echolalia,Stereotypy,Withdrawal | 92.4 | 89.9 | 94.6 |
| 4 | Staring,Grimacing,Mannerisms,Negativism | 92.4 | 89.9 | 94.6 |
| 4 | Staring,Mannerisms,Verbigeration,Negativism | 92.4 | 89.9 | 94.6 |
| 4 | Staring,Mannerisms,Verbigeration,Rigidity | 92.4 | 89.9 | 94.6 |
| 4 | Staring,Posturing/catalepsy,Verbigeration,Negativism | 92.4 | 89.9 | 94.8 |
| 4 | Staring,Posturing/catalepsy,Verbigeration,Withdrawal | 92.4 | 89.9 | 94.6 |
| 4 | Staring,Stereotypy,Verbigeration,Withdrawal | 92.4 | 89.9 | 94.8 |
| 4 | Staring,Stereotypy,Waxy flexibility,Withdrawal | 92.3 | 89.7 | 94.6 |
| 4 | Excitement,Immobility/stupor,Mannerisms,Negativism | 92.2 | 89.7 | 94.6 |
| 4 | Excitement,Immobility/stupor,Posturing/catalepsy,Mannerisms | 92.2 | 89.7 | 94.6 |
| 4 | Excitement,Staring,Grimacing,Mannerisms | 92.2 | 89.7 | 94.6 |
| 4 | Excitement,Staring,Mannerisms,Waxy flexibility | 92.2 | 89.7 | 94.6 |
| 4 | Immobility/stupor,Mutism,Staring,Negativism | 92.2 | 89.5 | 94.6 |
| 4 | Immobility/stupor,Staring,Grimacing,Verbigeration | 92.2 | 89.7 | 94.6 |
| 4 | Immobility/stupor,Staring,Stereotypy,Withdrawal | 92.2 | 89.7 | 94.6 |
| 4 | Immobility/stupor,Stereotypy,Negativism,Withdrawal | 92.2 | 89.7 | 94.6 |
| 4 | Mutism,Staring,Echopraxia/echolalia,Waxy flexibility | 92.2 | 89.7 | 94.6 |
| 4 | Staring,Mannerisms,Rigidity,Waxy flexibility | 92.2 | 89.7 | 94.6 |
| 4 | Excitement,Immobility/stupor,Grimacing,Withdrawal | 92.1 | 89.7 | 94.6 |
| 4 | Mutism,Staring,Posturing/catalepsy,Grimacing | 92.1 | 89.7 | 94.6 |
| 4 | Staring,Grimacing,Echopraxia/echolalia,Rigidity | 92.1 | 89.5 | 94.6 |
| 4 | Staring,Grimacing,Stereotypy,Negativism | 92.1 | 89.5 | 94.6 |
| 4 | Staring,Posturing/catalepsy,Stereotypy,Rigidity | 92.1 | 89.5 | 94.6 |
| 4 | Excitement,Immobility/stupor,Echopraxia/echolalia,Waxy flexibility | 91.9 | 89.2 | 94.4 |
| 4 | Excitement,Immobility/stupor,Grimacing,Echopraxia/echolalia | 91.9 | 89.2 | 94.4 |
| 4 | Excitement,Immobility/stupor,Grimacing,Waxy flexibility | 91.9 | 89.2 | 94.4 |
| 4 | Excitement,Immobility/stupor,Mannerisms,Withdrawal | 91.9 | 89.2 | 94.4 |
| 4 | Excitement,Immobility/stupor,Posturing/catalepsy,Stereotypy | 91.9 | 89.2 | 94.4 |
| 4 | Excitement,Immobility/stupor,Posturing/catalepsy,Waxy flexibility | 91.9 | 89.5 | 94.4 |
| 4 | Excitement,Staring,Grimacing,Verbigeration | 91.9 | 89.2 | 94.4 |
| 4 | Immobility/stupor,Mutism,Staring,Rigidity | 91.9 | 89.2 | 94.4 |
| 4 | Immobility/stupor,Posturing/catalepsy,Mannerisms,Negativism | 91.9 | 89.2 | 94.4 |
| 4 | Immobility/stupor,Staring,Echopraxia/echolalia,Withdrawal | 91.9 | 89.2 | 94.4 |
| 4 | Immobility/stupor,Staring,Grimacing,Stereotypy | 91.9 | 89.2 | 94.4 |
| 4 | Immobility/stupor,Staring,Negativism,Waxy flexibility | 91.9 | 89.2 | 94.4 |
| 4 | Immobility/stupor,Staring,Verbigeration,Withdrawal | 91.9 | 89.2 | 94.4 |
| 4 | Staring,Echopraxia/echolalia,Stereotypy,Rigidity | 91.9 | 89.2 | 94.4 |
| 4 | Staring,Posturing/catalepsy,Grimacing,Negativism | 91.9 | 89.5 | 94.4 |
| 4 | Excitement,Immobility/stupor,Echopraxia/echolalia,Verbigeration | 91.7 | 89.0 | 94.2 |
| 4 | Excitement,Staring,Stereotypy,Verbigeration | 91.7 | 89.0 | 94.2 |
| 4 | Immobility/stupor,Mutism,Mannerisms,Rigidity | 91.7 | 89.0 | 94.2 |
| 4 | Immobility/stupor,Mutism,Staring,Verbigeration | 91.7 | 89.0 | 94.2 |
| 4 | Immobility/stupor,Mutism,Staring,Withdrawal | 91.7 | 89.0 | 94.2 |
| 4 | Immobility/stupor,Posturing/catalepsy,Negativism,Withdrawal | 91.7 | 89.2 | 94.2 |
| 4 | Immobility/stupor,Posturing/catalepsy,Stereotypy,Negativism | 91.7 | 89.0 | 94.2 |
| 4 | Immobility/stupor,Staring,Echopraxia/echolalia,Stereotypy | 91.7 | 89.0 | 94.2 |
| 4 | Immobility/stupor,Staring,Grimacing,Withdrawal | 91.7 | 89.0 | 94.2 |
| 4 | Immobility/stupor,Staring,Posturing/catalepsy,Stereotypy | 91.7 | 89.0 | 94.2 |
| 4 | Immobility/stupor,Stereotypy,Mannerisms,Negativism | 91.7 | 89.0 | 94.2 |
| 4 | Immobility/stupor,Stereotypy,Rigidity,Withdrawal | 91.7 | 89.0 | 94.2 |
| 4 | Staring,Echopraxia/echolalia,Negativism,Waxy flexibility | 91.7 | 89.0 | 94.2 |
| 4 | Staring,Echopraxia/echolalia,Verbigeration,Negativism | 91.7 | 89.0 | 94.2 |
| 4 | Staring,Echopraxia/echolalia,Waxy flexibility,Withdrawal | 91.7 | 89.0 | 94.2 |
| 4 | Staring,Grimacing,Echopraxia/echolalia,Negativism | 91.7 | 89.0 | 94.2 |
| 4 | Staring,Grimacing,Verbigeration,Negativism | 91.7 | 89.0 | 94.2 |
| 4 | Staring,Grimacing,Waxy flexibility,Withdrawal | 91.7 | 89.0 | 94.2 |
| 4 | Staring,Posturing/catalepsy,Echopraxia/echolalia,Rigidity | 91.7 | 89.0 | 94.2 |
| 4 | Staring,Posturing/catalepsy,Verbigeration,Rigidity | 91.7 | 89.0 | 94.2 |
| 4 | Staring,Posturing/catalepsy,Waxy flexibility,Withdrawal | 91.7 | 89.0 | 94.2 |
| 4 | Staring,Verbigeration,Waxy flexibility,Withdrawal | 91.7 | 89.0 | 94.2 |
| 4 | Excitement,Immobility/stupor,Echopraxia/echolalia,Stereotypy | 91.5 | 88.8 | 93.9 |
| 4 | Excitement,Immobility/stupor,Stereotypy,Waxy flexibility | 91.5 | 88.8 | 93.9 |
| 4 | Excitement,Immobility/stupor,Verbigeration,Waxy flexibility | 91.5 | 88.8 | 93.9 |
| 4 | Excitement,Staring,Echopraxia/echolalia,Verbigeration | 91.5 | 88.8 | 93.9 |
| 4 | Immobility/stupor,Mannerisms,Negativism,Withdrawal | 91.5 | 88.8 | 93.9 |
| 4 | Immobility/stupor,Posturing/catalepsy,Rigidity,Negativism | 91.5 | 88.8 | 93.9 |
| 4 | Immobility/stupor,Staring,Grimacing,Echopraxia/echolalia | 91.5 | 88.8 | 93.9 |
| 4 | Immobility/stupor,Staring,Mannerisms,Waxy flexibility | 91.5 | 88.8 | 93.9 |
| 4 | Immobility/stupor,Staring,Posturing/catalepsy,Echopraxia/echolalia | 91.5 | 88.8 | 93.9 |
| 4 | Immobility/stupor,Staring,Posturing/catalepsy,Rigidity | 91.5 | 88.8 | 93.9 |
| 4 | Immobility/stupor,Staring,Posturing/catalepsy,Verbigeration | 91.5 | 88.8 | 93.9 |
| 4 | Immobility/stupor,Stereotypy,Verbigeration,Negativism | 91.5 | 88.8 | 93.9 |
| 4 | Mutism,Staring,Grimacing,Waxy flexibility | 91.5 | 88.8 | 93.9 |
| 4 | Staring,Posturing/catalepsy,Grimacing,Mannerisms | 91.5 | 88.8 | 93.9 |
| 4 | Staring,Posturing/catalepsy,Grimacing,Rigidity | 91.5 | 88.8 | 93.9 |
| 4 | Staring,Stereotypy,Rigidity,Waxy flexibility | 91.5 | 88.8 | 93.9 |
| 4 | Staring,Verbigeration,Negativism,Waxy flexibility | 91.5 | 88.8 | 93.9 |
| 4 | Excitement,Immobility/stupor,Grimacing,Verbigeration | 91.3 | 88.6 | 93.7 |
| 4 | Excitement,Immobility/stupor,Mutism,Mannerisms | 91.3 | 88.6 | 93.7 |
| 4 | Excitement,Staring,Stereotypy,Mannerisms | 91.3 | 88.6 | 93.7 |
| 4 | Immobility/stupor,Echopraxia/echolalia,Rigidity,Negativism | 91.3 | 88.6 | 93.7 |
| 4 | Immobility/stupor,Grimacing,Mannerisms,Rigidity | 91.3 | 88.6 | 93.7 |
| 4 | Immobility/stupor,Grimacing,Rigidity,Negativism | 91.3 | 88.6 | 93.7 |
| 4 | Immobility/stupor,Mutism,Rigidity,Negativism | 91.3 | 88.6 | 93.7 |
| 4 | Immobility/stupor,Mutism,Staring,Echopraxia/echolalia | 91.3 | 88.6 | 93.7 |
| 4 | Immobility/stupor,Staring,Echopraxia/echolalia,Verbigeration | 91.3 | 88.6 | 93.7 |
| 4 | Immobility/stupor,Staring,Waxy flexibility,Withdrawal | 91.3 | 88.6 | 93.7 |
| 4 | Staring,Echopraxia/echolalia,Verbigeration,Rigidity | 91.3 | 88.6 | 93.7 |
| 4 | Staring,Grimacing,Rigidity,Waxy flexibility | 91.3 | 88.6 | 93.9 |
| 4 | Staring,Grimacing,Stereotypy,Mannerisms | 91.3 | 88.6 | 93.7 |
| 4 | Excitement,Immobility/stupor,Stereotypy,Verbigeration | 91.2 | 88.6 | 93.7 |
| 4 | Excitement,Staring,Echopraxia/echolalia,Mannerisms | 91.2 | 88.6 | 93.7 |
| 4 | Immobility/stupor,Mutism,Staring,Stereotypy | 91.2 | 88.6 | 93.7 |
| 4 | Mutism,Staring,Posturing/catalepsy,Waxy flexibility | 91.2 | 88.6 | 93.7 |
| 4 | Excitement,Immobility/stupor,Grimacing,Stereotypy | 91.0 | 88.3 | 93.5 |
| 4 | Excitement,Mutism,Posturing/catalepsy,Rigidity | 91.0 | 88.3 | 93.5 |
| 4 | Excitement,Staring,Mannerisms,Verbigeration | 91.0 | 88.3 | 93.7 |
| 4 | Immobility/stupor,Echopraxia/echolalia,Rigidity,Withdrawal | 91.0 | 88.3 | 93.5 |
| 4 | Immobility/stupor,Posturing/catalepsy,Verbigeration,Negativism | 91.0 | 88.3 | 93.7 |
| 4 | Immobility/stupor,Staring,Rigidity,Waxy flexibility | 91.0 | 88.3 | 93.7 |
| 4 | Immobility/stupor,Staring,Stereotypy,Waxy flexibility | 91.0 | 88.3 | 93.7 |
| 4 | Immobility/stupor,Verbigeration,Negativism,Withdrawal | 91.0 | 88.3 | 93.5 |
| 4 | Immobility/stupor,Verbigeration,Rigidity,Withdrawal | 91.0 | 88.3 | 93.7 |
| 4 | Staring,Echopraxia/echolalia,Verbigeration,Withdrawal | 91.0 | 88.3 | 93.7 |
| 4 | Staring,Grimacing,Echopraxia/echolalia,Mannerisms | 91.0 | 88.3 | 93.5 |
| 4 | Staring,Grimacing,Echopraxia/echolalia,Stereotypy | 91.0 | 88.3 | 93.5 |
| 4 | Staring,Grimacing,Stereotypy,Verbigeration | 91.0 | 88.3 | 93.5 |
| 4 | Staring,Posturing/catalepsy,Grimacing,Stereotypy | 91.0 | 88.3 | 93.5 |
| 4 | Staring,Posturing/catalepsy,Negativism,Waxy flexibility | 91.0 | 88.3 | 93.5 |
| 4 | Staring,Posturing/catalepsy,Stereotypy,Mannerisms | 91.0 | 88.3 | 93.5 |
| 4 | Immobility/stupor,Echopraxia/echolalia,Stereotypy,Negativism | 90.8 | 87.9 | 93.5 |
| 4 | Immobility/stupor,Mutism,Staring,Posturing/catalepsy | 90.8 | 88.1 | 93.3 |
| 4 | Immobility/stupor,Staring,Verbigeration,Waxy flexibility | 90.8 | 88.1 | 93.5 |
| 4 | Immobility/stupor,Stereotypy,Negativism,Waxy flexibility | 90.8 | 88.1 | 93.5 |
| 4 | Staring,Grimacing,Mannerisms,Waxy flexibility | 90.8 | 88.1 | 93.3 |
| 4 | Staring,Grimacing,Negativism,Waxy flexibility | 90.8 | 88.1 | 93.5 |
| 4 | Staring,Posturing/catalepsy,Grimacing,Verbigeration | 90.8 | 88.1 | 93.5 |
| 4 | Staring,Posturing/catalepsy,Mannerisms,Verbigeration | 90.8 | 88.1 | 93.5 |
| 4 | Staring,Posturing/catalepsy,Stereotypy,Verbigeration | 90.8 | 88.1 | 93.5 |
| 4 | Staring,Verbigeration,Rigidity,Waxy flexibility | 90.8 | 88.1 | 93.5 |
| 4 | Excitement,Immobility/stupor,Grimacing,Mannerisms | 90.6 | 87.9 | 93.0 |
| 4 | Excitement,Mutism,Rigidity,Negativism | 90.6 | 87.9 | 93.3 |
| 4 | Immobility/stupor,Grimacing,Rigidity,Withdrawal | 90.6 | 87.7 | 93.0 |
| 4 | Immobility/stupor,Grimacing,Stereotypy,Negativism | 90.6 | 87.7 | 93.3 |
| 4 | Immobility/stupor,Grimacing,Stereotypy,Rigidity | 90.6 | 87.7 | 93.3 |
| 4 | Immobility/stupor,Mannerisms,Verbigeration,Negativism | 90.6 | 87.9 | 93.3 |
| 4 | Immobility/stupor,Mutism,Mannerisms,Negativism | 90.6 | 87.9 | 93.0 |
| 4 | Immobility/stupor,Mutism,Verbigeration,Rigidity | 90.6 | 87.9 | 93.3 |
| 4 | Immobility/stupor,Staring,Posturing/catalepsy,Grimacing | 90.6 | 87.7 | 93.3 |
| 4 | Staring,Posturing/catalepsy,Echopraxia/echolalia,Mannerisms | 90.6 | 87.9 | 93.3 |
| 4 | Staring,Posturing/catalepsy,Echopraxia/echolalia,Stereotypy | 90.6 | 87.7 | 93.3 |
| 4 | Staring,Posturing/catalepsy,Grimacing,Echopraxia/echolalia | 90.6 | 87.9 | 93.0 |
| 4 | Excitement,Mutism,Rigidity,Withdrawal | 90.5 | 87.7 | 93.3 |
| 4 | Excitement,Immobility/stupor,Mannerisms,Verbigeration | 90.4 | 87.7 | 93.0 |
| 4 | Excitement,Mutism,Posturing/catalepsy,Withdrawal | 90.4 | 87.4 | 93.0 |
| 4 | Immobility/stupor,Grimacing,Verbigeration,Rigidity | 90.4 | 87.4 | 93.0 |
| 4 | Immobility/stupor,Mutism,Stereotypy,Negativism | 90.4 | 87.4 | 93.0 |
| 4 | Immobility/stupor,Posturing/catalepsy,Echopraxia/echolalia,Negativism | 90.4 | 87.4 | 93.0 |
| 4 | Immobility/stupor,Posturing/catalepsy,Grimacing,Negativism | 90.4 | 87.7 | 93.0 |
| 4 | Immobility/stupor,Stereotypy,Verbigeration,Rigidity | 90.4 | 87.7 | 93.0 |
| 4 | Staring,Echopraxia/echolalia,Rigidity,Waxy flexibility | 90.4 | 87.7 | 93.0 |
| 4 | Staring,Echopraxia/echolalia,Stereotypy,Mannerisms | 90.4 | 87.7 | 93.0 |
| 4 | Staring,Grimacing,Mannerisms,Verbigeration | 90.4 | 87.4 | 93.0 |
| 4 | Staring,Grimacing,Stereotypy,Waxy flexibility | 90.4 | 87.4 | 93.0 |
| 4 | Immobility/stupor,Echopraxia/echolalia,Mannerisms,Negativism | 90.3 | 87.4 | 93.0 |
| 4 | Immobility/stupor,Echopraxia/echolalia,Negativism,Withdrawal | 90.3 | 87.4 | 93.0 |
| 4 | Immobility/stupor,Mutism,Rigidity,Withdrawal | 90.3 | 87.7 | 93.0 |
| 4 | Immobility/stupor,Mutism,Staring,Grimacing | 90.3 | 87.4 | 93.0 |
| 4 | Immobility/stupor,Posturing/catalepsy,Mannerisms,Rigidity | 90.3 | 87.4 | 93.0 |
| 4 | Immobility/stupor,Stereotypy,Mannerisms,Rigidity | 90.3 | 87.4 | 93.0 |
| 4 | Excitement,Immobility/stupor,Mannerisms,Waxy flexibility | 90.2 | 87.4 | 92.8 |
| 4 | Immobility/stupor,Mutism,Stereotypy,Rigidity | 90.2 | 87.2 | 92.8 |
| 4 | Excitement,Immobility/stupor,Echopraxia/echolalia,Mannerisms | 90.1 | 87.2 | 92.8 |
| 4 | Excitement,Mutism,Posturing/catalepsy,Negativism | 90.1 | 87.2 | 92.8 |
| 4 | Immobility/stupor,Mannerisms,Negativism,Waxy flexibility | 90.1 | 87.2 | 92.8 |
| 4 | Immobility/stupor,Mannerisms,Verbigeration,Rigidity | 90.1 | 87.2 | 92.8 |
| 4 | Immobility/stupor,Mutism,Posturing/catalepsy,Negativism | 90.1 | 87.4 | 92.8 |
| 4 | Immobility/stupor,Posturing/catalepsy,Rigidity,Withdrawal | 90.1 | 87.2 | 92.8 |
| 4 | Immobility/stupor,Rigidity,Negativism,Waxy flexibility | 90.1 | 87.2 | 92.8 |
| 4 | Immobility/stupor,Staring,Echopraxia/echolalia,Waxy flexibility | 90.1 | 87.2 | 92.8 |
| 4 | Staring,Grimacing,Verbigeration,Waxy flexibility | 90.1 | 87.2 | 92.8 |
| 4 | Staring,Stereotypy,Mannerisms,Waxy flexibility | 90.1 | 87.2 | 92.8 |
| 4 | Staring,Stereotypy,Verbigeration,Waxy flexibility | 90.1 | 87.2 | 92.8 |
| 4 | Excitement,Immobility/stupor,Stereotypy,Mannerisms | 89.9 | 87.0 | 92.6 |
| 4 | Immobility/stupor,Echopraxia/echolalia,Mannerisms,Rigidity | 89.9 | 87.0 | 92.6 |
| 4 | Immobility/stupor,Grimacing,Mannerisms,Negativism | 89.9 | 87.0 | 92.6 |
| 4 | Immobility/stupor,Grimacing,Verbigeration,Negativism | 89.9 | 87.0 | 92.6 |
| 4 | Immobility/stupor,Mutism,Staring,Waxy flexibility | 89.9 | 87.0 | 92.6 |
| 4 | Immobility/stupor,Staring,Grimacing,Waxy flexibility | 89.9 | 87.0 | 92.6 |
| 4 | Staring,Grimacing,Echopraxia/echolalia,Verbigeration | 89.9 | 87.0 | 92.6 |
| 4 | Staring,Stereotypy,Mannerisms,Verbigeration | 89.9 | 87.0 | 92.6 |
| 4 | Immobility/stupor,Echopraxia/echolalia,Stereotypy,Rigidity | 89.7 | 86.8 | 92.4 |
| 4 | Immobility/stupor,Echopraxia/echolalia,Verbigeration,Negativism | 89.7 | 86.8 | 92.4 |
| 4 | Immobility/stupor,Grimacing,Echopraxia/echolalia,Rigidity | 89.7 | 86.8 | 92.4 |
| 4 | Immobility/stupor,Mutism,Echopraxia/echolalia,Rigidity | 89.7 | 86.8 | 92.4 |
| 4 | Immobility/stupor,Mutism,Verbigeration,Negativism | 89.7 | 86.8 | 92.4 |
| 4 | Immobility/stupor,Posturing/catalepsy,Mannerisms,Withdrawal | 89.7 | 86.8 | 92.4 |
| 4 | Mutism,Rigidity,Negativism,Withdrawal | 89.7 | 86.8 | 92.4 |
| 4 | Staring,Echopraxia/echolalia,Stereotypy,Waxy flexibility | 89.7 | 86.8 | 92.4 |
| 4 | Staring,Grimacing,Echopraxia/echolalia,Waxy flexibility | 89.7 | 86.8 | 92.4 |
| 4 | Staring,Mannerisms,Verbigeration,Waxy flexibility | 89.7 | 86.8 | 92.4 |
| 4 | Staring,Posturing/catalepsy,Echopraxia/echolalia,Verbigeration | 89.7 | 86.8 | 92.4 |
| 4 | Immobility/stupor,Negativism,Waxy flexibility,Withdrawal | 89.5 | 86.5 | 92.2 |
| 4 | Mutism,Mannerisms,Rigidity,Negativism | 89.5 | 86.5 | 92.2 |
| 4 | Immobility/stupor,Posturing/catalepsy,Stereotypy,Rigidity | 89.4 | 86.5 | 92.2 |
| 4 | Staring,Echopraxia/echolalia,Mannerisms,Waxy flexibility | 89.4 | 86.5 | 92.2 |
| 4 | Immobility/stupor,Posturing/catalepsy,Grimacing,Rigidity | 89.3 | 86.3 | 91.9 |
| 4 | Immobility/stupor,Posturing/catalepsy,Grimacing,Withdrawal | 89.3 | 86.3 | 92.2 |
| 4 | Staring,Posturing/catalepsy,Rigidity,Waxy flexibility | 89.3 | 86.3 | 91.9 |
| 4 | Staring,Posturing/catalepsy,Stereotypy,Waxy flexibility | 89.3 | 86.3 | 92.2 |
| 4 | Immobility/stupor,Grimacing,Negativism,Withdrawal | 89.2 | 86.3 | 91.9 |
| 4 | Immobility/stupor,Mutism,Grimacing,Rigidity | 89.2 | 86.3 | 91.9 |
| 4 | Immobility/stupor,Mutism,Negativism,Withdrawal | 89.2 | 86.3 | 91.9 |
| 4 | Staring,Posturing/catalepsy,Mannerisms,Waxy flexibility | 89.2 | 86.3 | 91.9 |
| 4 | Immobility/stupor,Echopraxia/echolalia,Verbigeration,Rigidity | 89.0 | 86.1 | 91.9 |
| 4 | Immobility/stupor,Grimacing,Echopraxia/echolalia,Negativism | 89.0 | 86.1 | 91.7 |
| 4 | Immobility/stupor,Grimacing,Stereotypy,Withdrawal | 89.0 | 86.1 | 91.7 |
| 4 | Immobility/stupor,Mannerisms,Rigidity,Waxy flexibility | 89.0 | 86.1 | 91.9 |
| 4 | Immobility/stupor,Mutism,Echopraxia/echolalia,Negativism | 89.0 | 86.1 | 91.7 |
| 4 | Immobility/stupor,Posturing/catalepsy,Stereotypy,Withdrawal | 89.0 | 86.1 | 91.9 |
| 4 | Immobility/stupor,Posturing/catalepsy,Verbigeration,Withdrawal | 89.0 | 86.1 | 91.7 |
| 4 | Immobility/stupor,Rigidity,Waxy flexibility,Withdrawal | 89.0 | 86.1 | 91.9 |
| 4 | Mutism,Posturing/catalepsy,Negativism,Withdrawal | 89.0 | 86.1 | 91.9 |
| 4 | Mutism,Stereotypy,Rigidity,Negativism | 89.0 | 86.1 | 91.9 |
| 4 | Staring,Echopraxia/echolalia,Stereotypy,Verbigeration | 89.0 | 85.9 | 91.9 |
| 4 | Excitement,Posturing/catalepsy,Rigidity,Withdrawal | 88.8 | 85.9 | 91.7 |
| 4 | Immobility/stupor,Echopraxia/echolalia,Negativism,Waxy flexibility | 88.8 | 85.9 | 91.5 |
| 4 | Immobility/stupor,Mutism,Mannerisms,Withdrawal | 88.8 | 85.9 | 91.7 |
| 4 | Immobility/stupor,Mutism,Posturing/catalepsy,Withdrawal | 88.8 | 85.7 | 91.7 |
| 4 | Immobility/stupor,Posturing/catalepsy,Negativism,Waxy flexibility | 88.8 | 85.7 | 91.7 |
| 4 | Immobility/stupor,Posturing/catalepsy,Verbigeration,Rigidity | 88.8 | 85.7 | 91.7 |
| 4 | Immobility/stupor,Staring,Posturing/catalepsy,Waxy flexibility | 88.8 | 85.9 | 91.7 |
| 4 | Immobility/stupor,Verbigeration,Negativism,Waxy flexibility | 88.8 | 85.7 | 91.7 |
| 4 | Mutism,Posturing/catalepsy,Rigidity,Negativism | 88.8 | 85.9 | 91.7 |
| 4 | Staring,Echopraxia/echolalia,Mannerisms,Verbigeration | 88.8 | 85.9 | 91.7 |
| 4 | Immobility/stupor,Echopraxia/echolalia,Stereotypy,Withdrawal | 88.6 | 85.4 | 91.5 |
| 4 | Immobility/stupor,Mutism,Posturing/catalepsy,Mannerisms | 88.6 | 85.4 | 91.5 |
| 4 | Immobility/stupor,Mutism,Posturing/catalepsy,Verbigeration | 88.6 | 85.4 | 91.5 |
| 4 | Immobility/stupor,Mutism,Verbigeration,Withdrawal | 88.6 | 85.7 | 91.5 |
| 4 | Immobility/stupor,Posturing/catalepsy,Echopraxia/echolalia,Rigidity | 88.6 | 85.7 | 91.5 |
| 4 | Immobility/stupor,Posturing/catalepsy,Echopraxia/echolalia,Withdrawal | 88.6 | 85.7 | 91.3 |
| 4 | Immobility/stupor,Stereotypy,Mannerisms,Withdrawal | 88.6 | 85.4 | 91.5 |
| 4 | Mutism,Verbigeration,Rigidity,Negativism | 88.6 | 85.4 | 91.5 |
| 4 | Staring,Echopraxia/echolalia,Verbigeration,Waxy flexibility | 88.6 | 85.4 | 91.5 |
| 4 | Staring,Posturing/catalepsy,Verbigeration,Waxy flexibility | 88.6 | 85.4 | 91.5 |
| 4 | Excitement,Posturing/catalepsy,Negativism,Withdrawal | 88.5 | 85.7 | 91.5 |
| 4 | Immobility/stupor,Grimacing,Mannerisms,Withdrawal | 88.5 | 85.7 | 91.5 |
| 4 | Immobility/stupor,Mutism,Negativism,Waxy flexibility | 88.5 | 85.4 | 91.5 |
| 4 | Immobility/stupor,Stereotypy,Verbigeration,Withdrawal | 88.5 | 85.4 | 91.5 |
| 4 | Immobility/stupor,Mutism,Grimacing,Verbigeration | 88.4 | 85.2 | 91.3 |
| 4 | Immobility/stupor,Posturing/catalepsy,Grimacing,Mannerisms | 88.4 | 85.2 | 91.3 |
| 4 | Staring,Posturing/catalepsy,Echopraxia/echolalia,Waxy flexibility | 88.4 | 85.2 | 91.3 |
| 4 | Excitement,Mutism,Echopraxia/echolalia,Rigidity | 88.3 | 85.2 | 91.3 |
| 4 | Excitement,Mutism,Negativism,Withdrawal | 88.3 | 85.4 | 91.3 |
| 4 | Excitement,Mutism,Rigidity,Waxy flexibility | 88.3 | 85.2 | 91.3 |
| 4 | Immobility/stupor,Echopraxia/echolalia,Mannerisms,Withdrawal | 88.3 | 85.4 | 91.3 |
| 4 | Immobility/stupor,Grimacing,Rigidity,Waxy flexibility | 88.3 | 85.2 | 91.3 |
| 4 | Immobility/stupor,Mannerisms,Verbigeration,Withdrawal | 88.3 | 85.2 | 91.3 |
| 4 | Immobility/stupor,Mutism,Echopraxia/echolalia,Mannerisms | 88.3 | 85.2 | 91.3 |
| 4 | Immobility/stupor,Mutism,Posturing/catalepsy,Rigidity | 88.3 | 85.2 | 91.3 |
| 4 | Immobility/stupor,Mutism,Stereotypy,Withdrawal | 88.3 | 85.2 | 91.3 |
| 4 | Immobility/stupor,Posturing/catalepsy,Grimacing,Verbigeration | 88.3 | 85.4 | 91.3 |
| 4 | Immobility/stupor,Stereotypy,Rigidity,Waxy flexibility | 88.3 | 85.2 | 91.3 |
| 4 | Posturing/catalepsy,Rigidity,Negativism,Withdrawal | 88.3 | 85.2 | 91.3 |
| 4 | Staring,Posturing/catalepsy,Grimacing,Waxy flexibility | 88.3 | 85.2 | 91.3 |
| 4 | Excitement,Mutism,Stereotypy,Rigidity | 88.1 | 85.2 | 91.0 |
| 4 | Immobility/stupor,Grimacing,Echopraxia/echolalia,Withdrawal | 88.1 | 85.0 | 91.0 |
| 4 | Immobility/stupor,Grimacing,Negativism,Waxy flexibility | 88.1 | 85.0 | 91.0 |
| 4 | Immobility/stupor,Grimacing,Verbigeration,Withdrawal | 88.1 | 85.0 | 91.0 |
| 4 | Immobility/stupor,Mutism,Echopraxia/echolalia,Withdrawal | 88.1 | 85.0 | 91.0 |
| 4 | Immobility/stupor,Mutism,Grimacing,Negativism | 88.1 | 85.0 | 91.0 |
| 4 | Immobility/stupor,Mutism,Grimacing,Withdrawal | 88.1 | 85.0 | 91.0 |
| 4 | Immobility/stupor,Mutism,Posturing/catalepsy,Echopraxia/echolalia | 88.1 | 85.0 | 91.0 |
| 4 | Immobility/stupor,Mutism,Stereotypy,Verbigeration | 88.1 | 85.2 | 91.0 |
| 4 | Immobility/stupor,Posturing/catalepsy,Grimacing,Stereotypy | 88.1 | 85.0 | 91.0 |
| 4 | Mutism,Mannerisms,Rigidity,Withdrawal | 88.1 | 85.0 | 91.0 |
| 4 | Excitement,Mutism,Grimacing,Rigidity | 87.9 | 84.8 | 90.8 |
| 4 | Immobility/stupor,Grimacing,Echopraxia/echolalia,Stereotypy | 87.9 | 84.8 | 90.8 |
| 4 | Immobility/stupor,Grimacing,Stereotypy,Verbigeration | 87.9 | 84.8 | 90.8 |
| 4 | Immobility/stupor,Mutism,Grimacing,Mannerisms | 87.9 | 84.8 | 90.8 |
| 4 | Immobility/stupor,Mutism,Mannerisms,Verbigeration | 87.9 | 84.8 | 90.8 |
| 4 | Immobility/stupor,Mutism,Posturing/catalepsy,Grimacing | 87.9 | 84.8 | 90.8 |
| 4 | Immobility/stupor,Mutism,Posturing/catalepsy,Stereotypy | 87.9 | 84.8 | 90.8 |
| 4 | Immobility/stupor,Posturing/catalepsy,Stereotypy,Verbigeration | 87.9 | 85.0 | 90.8 |
| 4 | Mutism,Echopraxia/echolalia,Rigidity,Negativism | 87.9 | 84.8 | 90.8 |
| 4 | Mutism,Posturing/catalepsy,Rigidity,Withdrawal | 87.9 | 84.8 | 90.8 |
| 4 | Immobility/stupor,Echopraxia/echolalia,Verbigeration,Withdrawal | 87.7 | 84.5 | 90.6 |
| 4 | Immobility/stupor,Mutism,Grimacing,Stereotypy | 87.7 | 84.5 | 90.6 |
| 4 | Immobility/stupor,Mutism,Stereotypy,Mannerisms | 87.7 | 84.5 | 90.6 |
| 4 | Immobility/stupor,Posturing/catalepsy,Grimacing,Echopraxia/echolalia | 87.7 | 84.5 | 90.6 |
| 4 | Mutism,Posturing/catalepsy,Mannerisms,Negativism | 87.7 | 84.5 | 90.6 |
| 4 | Mutism,Posturing/catalepsy,Mannerisms,Rigidity | 87.7 | 84.5 | 90.6 |
| 4 | Mutism,Stereotypy,Rigidity,Withdrawal | 87.7 | 84.5 | 90.6 |
| 4 | Excitement,Mutism,Posturing/catalepsy,Echopraxia/echolalia | 87.6 | 84.5 | 90.6 |
| 4 | Excitement,Mutism,Waxy flexibility,Withdrawal | 87.6 | 84.5 | 90.6 |
| 4 | Excitement,Mutism,Verbigeration,Rigidity | 87.5 | 84.3 | 90.4 |
| 4 | Immobility/stupor,Posturing/catalepsy,Echopraxia/echolalia,Stereotypy | 87.5 | 84.3 | 90.4 |
| 4 | Mutism,Posturing/catalepsy,Stereotypy,Negativism | 87.5 | 84.3 | 90.4 |
| 4 | Immobility/stupor,Grimacing,Stereotypy,Mannerisms | 87.4 | 84.3 | 90.4 |
| 4 | Immobility/stupor,Mutism,Echopraxia/echolalia,Stereotypy | 87.4 | 84.3 | 90.4 |
| 4 | Immobility/stupor,Mutism,Echopraxia/echolalia,Verbigeration | 87.4 | 84.3 | 90.6 |
| 4 | Immobility/stupor,Mutism,Rigidity,Waxy flexibility | 87.4 | 84.3 | 90.4 |
| 4 | Immobility/stupor,Stereotypy,Waxy flexibility,Withdrawal | 87.4 | 84.3 | 90.4 |
| 4 | Immobility/stupor,Verbigeration,Rigidity,Waxy flexibility | 87.4 | 84.3 | 90.4 |
| 4 | Posturing/catalepsy,Stereotypy,Negativism,Withdrawal | 87.4 | 84.3 | 90.4 |
| 4 | Immobility/stupor,Echopraxia/echolalia,Rigidity,Waxy flexibility | 87.2 | 84.1 | 90.4 |
| 4 | Immobility/stupor,Mannerisms,Waxy flexibility,Withdrawal | 87.2 | 84.1 | 90.1 |
| 4 | Immobility/stupor,Mutism,Grimacing,Echopraxia/echolalia | 87.2 | 84.1 | 90.1 |
| 4 | Immobility/stupor,Posturing/catalepsy,Echopraxia/echolalia,Mannerisms | 87.2 | 84.1 | 90.4 |
| 4 | Immobility/stupor,Posturing/catalepsy,Mannerisms,Verbigeration | 87.2 | 84.1 | 90.4 |
| 4 | Immobility/stupor,Posturing/catalepsy,Stereotypy,Mannerisms | 87.2 | 84.1 | 90.1 |
| 4 | Mutism,Posturing/catalepsy,Echopraxia/echolalia,Negativism | 87.2 | 84.1 | 90.1 |
| 4 | Mutism,Posturing/catalepsy,Verbigeration,Negativism | 87.2 | 84.1 | 90.1 |
| 4 | Mutism,Stereotypy,Negativism,Withdrawal | 87.2 | 84.1 | 90.4 |
| 4 | Mutism,Verbigeration,Rigidity,Withdrawal | 87.2 | 84.1 | 90.4 |
| 4 | Excitement,Mutism,Posturing/catalepsy,Grimacing | 87.0 | 83.9 | 90.1 |
| 4 | Excitement,Mutism,Posturing/catalepsy,Waxy flexibility | 87.0 | 83.9 | 90.1 |
| 4 | Immobility/stupor,Grimacing,Echopraxia/echolalia,Mannerisms | 87.0 | 83.9 | 90.1 |
| 4 | Immobility/stupor,Grimacing,Echopraxia/echolalia,Verbigeration | 87.0 | 83.9 | 90.1 |
| 4 | Immobility/stupor,Stereotypy,Mannerisms,Verbigeration | 87.0 | 83.9 | 89.9 |
| 4 | Immobility/stupor,Verbigeration,Waxy flexibility,Withdrawal | 87.0 | 83.9 | 90.1 |
| 4 | Mutism,Grimacing,Rigidity,Negativism | 87.0 | 83.9 | 90.1 |
| 4 | Mutism,Grimacing,Rigidity,Withdrawal | 87.0 | 83.9 | 89.9 |
| 4 | Mutism,Posturing/catalepsy,Mannerisms,Withdrawal | 87.0 | 83.9 | 90.1 |
| 4 | Mutism,Posturing/catalepsy,Verbigeration,Rigidity | 87.0 | 83.9 | 89.9 |
| 4 | Mutism,Rigidity,Negativism,Waxy flexibility | 87.0 | 83.9 | 89.9 |
| 4 | Mutism,Rigidity,Waxy flexibility,Withdrawal | 87.0 | 83.9 | 90.1 |
| 4 | Posturing/catalepsy,Mannerisms,Negativism,Withdrawal | 87.0 | 83.9 | 89.9 |
| 4 | Stereotypy,Rigidity,Negativism,Withdrawal | 87.0 | 83.9 | 90.1 |
| 4 | Excitement,Mutism,Posturing/catalepsy,Stereotypy | 86.8 | 83.4 | 89.9 |
| 4 | Excitement,Mutism,Posturing/catalepsy,Verbigeration | 86.8 | 83.6 | 89.9 |
| 4 | Excitement,Mutism,Stereotypy,Negativism | 86.8 | 83.4 | 89.9 |
| 4 | Immobility/stupor,Echopraxia/echolalia,Stereotypy,Mannerisms | 86.8 | 83.6 | 89.7 |
| 4 | Immobility/stupor,Echopraxia/echolalia,Stereotypy,Verbigeration | 86.8 | 83.6 | 89.7 |
| 4 | Immobility/stupor,Echopraxia/echolalia,Waxy flexibility,Withdrawal | 86.8 | 83.6 | 89.9 |
| 4 | Immobility/stupor,Grimacing,Mannerisms,Verbigeration | 86.8 | 83.6 | 89.7 |
| 4 | Immobility/stupor,Grimacing,Stereotypy,Waxy flexibility | 86.8 | 83.6 | 89.7 |
| 4 | Immobility/stupor,Mutism,Echopraxia/echolalia,Waxy flexibility | 86.8 | 83.6 | 89.9 |
| 4 | Immobility/stupor,Mutism,Waxy flexibility,Withdrawal | 86.8 | 83.6 | 89.9 |
| 4 | Immobility/stupor,Posturing/catalepsy,Echopraxia/echolalia,Verbigeration | 86.8 | 83.6 | 89.9 |
| 4 | Mutism,Echopraxia/echolalia,Rigidity,Withdrawal | 86.8 | 83.6 | 89.9 |
| 4 | Mutism,Mannerisms,Negativism,Withdrawal | 86.8 | 83.4 | 89.9 |
| 4 | Immobility/stupor,Mutism,Verbigeration,Waxy flexibility | 86.7 | 83.4 | 89.9 |
| 4 | Excitement,Mutism,Stereotypy,Withdrawal | 86.6 | 83.2 | 89.7 |
| 4 | Immobility/stupor,Grimacing,Waxy flexibility,Withdrawal | 86.6 | 83.2 | 89.7 |
| 4 | Immobility/stupor,Mutism,Stereotypy,Waxy flexibility | 86.6 | 83.4 | 89.7 |
| 4 | Mutism,Posturing/catalepsy,Echopraxia/echolalia,Rigidity | 86.6 | 83.2 | 89.7 |
| 4 | Mutism,Posturing/catalepsy,Stereotypy,Rigidity | 86.5 | 83.4 | 89.7 |
| 4 | Excitement,Mutism,Mannerisms,Rigidity | 86.3 | 83.0 | 89.5 |
| 4 | Excitement,Mutism,Negativism,Waxy flexibility | 86.3 | 83.0 | 89.5 |
| 4 | Excitement,Rigidity,Negativism,Withdrawal | 86.3 | 83.0 | 89.5 |
| 4 | Immobility/stupor,Echopraxia/echolalia,Stereotypy,Waxy flexibility | 86.3 | 83.2 | 89.5 |
| 4 | Immobility/stupor,Grimacing,Echopraxia/echolalia,Waxy flexibility | 86.3 | 83.2 | 89.5 |
| 4 | Immobility/stupor,Mutism,Mannerisms,Waxy flexibility | 86.3 | 83.0 | 89.5 |
| 4 | Immobility/stupor,Posturing/catalepsy,Waxy flexibility,Withdrawal | 86.3 | 83.0 | 89.5 |
| 4 | Immobility/stupor,Stereotypy,Verbigeration,Waxy flexibility | 86.3 | 83.0 | 89.5 |
| 4 | Mutism,Posturing/catalepsy,Grimacing,Negativism | 86.3 | 83.2 | 89.5 |
| 4 | Mutism,Posturing/catalepsy,Grimacing,Withdrawal | 86.3 | 83.0 | 89.5 |
| 4 | Mutism,Posturing/catalepsy,Verbigeration,Withdrawal | 86.3 | 83.2 | 89.5 |
| 4 | Posturing/catalepsy,Verbigeration,Negativism,Withdrawal | 86.3 | 83.0 | 89.5 |
| 4 | Excitement,Mutism,Echopraxia/echolalia,Negativism | 86.1 | 82.7 | 89.2 |
| 4 | Excitement,Mutism,Echopraxia/echolalia,Withdrawal | 86.1 | 82.7 | 89.2 |
| 4 | Excitement,Mutism,Posturing/catalepsy,Mannerisms | 86.1 | 82.7 | 89.2 |
| 4 | Immobility/stupor,Grimacing,Mannerisms,Waxy flexibility | 86.1 | 82.7 | 89.2 |
| 4 | Immobility/stupor,Grimacing,Verbigeration,Waxy flexibility | 86.1 | 82.7 | 89.2 |
| 4 | Mannerisms,Rigidity,Negativism,Withdrawal | 86.1 | 82.7 | 89.2 |
| 4 | Mutism,Posturing/catalepsy,Echopraxia/echolalia,Withdrawal | 86.1 | 82.7 | 89.2 |
| 4 | Mutism,Posturing/catalepsy,Stereotypy,Withdrawal | 86.1 | 82.7 | 89.2 |
| 4 | Mutism,Verbigeration,Negativism,Withdrawal | 86.1 | 82.7 | 89.2 |
| 4 | Immobility/stupor,Mutism,Grimacing,Waxy flexibility | 85.9 | 82.5 | 89.0 |
| 4 | Immobility/stupor,Stereotypy,Mannerisms,Waxy flexibility | 85.9 | 82.5 | 89.0 |
| 4 | Mutism,Echopraxia/echolalia,Negativism,Withdrawal | 85.9 | 82.7 | 89.0 |
| 4 | Mutism,Negativism,Waxy flexibility,Withdrawal | 85.9 | 82.5 | 89.0 |
| 4 | Mutism,Posturing/catalepsy,Grimacing,Rigidity | 85.9 | 82.5 | 89.0 |
| 4 | Posturing/catalepsy,Echopraxia/echolalia,Negativism,Withdrawal | 85.9 | 82.5 | 89.0 |
| 4 | Posturing/catalepsy,Mannerisms,Rigidity,Withdrawal | 85.9 | 82.7 | 89.0 |
| 4 | Immobility/stupor,Echopraxia/echolalia,Mannerisms,Verbigeration | 85.7 | 82.3 | 88.8 |
| 4 | Immobility/stupor,Posturing/catalepsy,Grimacing,Waxy flexibility | 85.7 | 82.3 | 88.8 |
| 4 | Immobility/stupor,Posturing/catalepsy,Stereotypy,Waxy flexibility | 85.7 | 82.3 | 88.8 |
| 4 | Posturing/catalepsy,Grimacing,Negativism,Withdrawal | 85.7 | 82.3 | 88.8 |
| 4 | Excitement,Mutism,Grimacing,Withdrawal | 85.6 | 82.3 | 88.8 |
| 4 | Excitement,Posturing/catalepsy,Echopraxia/echolalia,Withdrawal | 85.6 | 82.3 | 88.8 |
| 4 | Immobility/stupor,Posturing/catalepsy,Rigidity,Waxy flexibility | 85.6 | 82.3 | 88.8 |
| 4 | Excitement,Mutism,Verbigeration,Withdrawal | 85.5 | 82.3 | 88.6 |
| 4 | Excitement,Posturing/catalepsy,Grimacing,Withdrawal | 85.4 | 82.3 | 88.6 |
| 4 | Mutism,Posturing/catalepsy,Negativism,Waxy flexibility | 85.4 | 82.1 | 88.6 |
| 4 | Mutism,Stereotypy,Mannerisms,Negativism | 85.4 | 82.1 | 88.6 |
| 4 | Verbigeration,Rigidity,Negativism,Withdrawal | 85.4 | 82.1 | 88.6 |
| 4 | Echopraxia/echolalia,Rigidity,Negativism,Withdrawal | 85.2 | 81.8 | 88.6 |
| 4 | Excitement,Mutism,Grimacing,Negativism | 85.2 | 81.8 | 88.3 |
| 4 | Excitement,Mutism,Verbigeration,Negativism | 85.2 | 81.8 | 88.3 |
| 4 | Immobility/stupor,Mutism,Posturing/catalepsy,Waxy flexibility | 85.2 | 81.8 | 88.3 |
| 4 | Mutism,Grimacing,Mannerisms,Rigidity | 85.2 | 81.8 | 88.6 |
| 4 | Mutism,Grimacing,Negativism,Withdrawal | 85.2 | 81.8 | 88.3 |
| 4 | Mutism,Grimacing,Verbigeration,Rigidity | 85.2 | 81.8 | 88.6 |
| 4 | Mutism,Posturing/catalepsy,Waxy flexibility,Withdrawal | 85.2 | 81.8 | 88.3 |
| 4 | Mutism,Stereotypy,Verbigeration,Rigidity | 85.2 | 81.8 | 88.3 |
| 4 | Excitement,Posturing/catalepsy,Rigidity,Negativism | 85.0 | 81.6 | 88.3 |
| 4 | Excitement,Posturing/catalepsy,Verbigeration,Withdrawal | 85.0 | 81.6 | 88.1 |
| 4 | Excitement,Posturing/catalepsy,Waxy flexibility,Withdrawal | 85.0 | 81.6 | 88.3 |
| 4 | Mutism,Grimacing,Stereotypy,Rigidity | 85.0 | 81.6 | 88.1 |
| 4 | Mutism,Stereotypy,Mannerisms,Rigidity | 85.0 | 81.6 | 88.1 |
| 4 | Mutism,Stereotypy,Negativism,Waxy flexibility | 85.0 | 81.6 | 88.1 |
| 4 | Mutism,Stereotypy,Rigidity,Waxy flexibility | 85.0 | 81.4 | 88.3 |
| 4 | Mutism,Stereotypy,Verbigeration,Negativism | 85.0 | 81.6 | 88.1 |
| 4 | Posturing/catalepsy,Stereotypy,Rigidity,Withdrawal | 85.0 | 81.6 | 88.3 |
| 4 | Excitement,Posturing/catalepsy,Stereotypy,Withdrawal | 84.9 | 81.6 | 88.1 |
| 4 | Excitement,Posturing/catalepsy,Mannerisms,Withdrawal | 84.8 | 81.4 | 88.1 |
| 4 | Immobility/stupor,Posturing/catalepsy,Verbigeration,Waxy flexibility | 84.8 | 81.4 | 88.1 |
| 4 | Mutism,Echopraxia/echolalia,Mannerisms,Rigidity | 84.8 | 81.4 | 88.1 |
| 4 | Mutism,Echopraxia/echolalia,Stereotypy,Rigidity | 84.8 | 81.4 | 87.9 |
| 4 | Immobility/stupor,Echopraxia/echolalia,Verbigeration,Waxy flexibility | 84.7 | 81.2 | 87.9 |
| 4 | Immobility/stupor,Mannerisms,Verbigeration,Waxy flexibility | 84.7 | 81.4 | 87.9 |
| 4 | Immobility/stupor,Posturing/catalepsy,Mannerisms,Waxy flexibility | 84.7 | 81.4 | 87.9 |
| 4 | Mutism,Echopraxia/echolalia,Mannerisms,Negativism | 84.7 | 81.4 | 87.9 |
| 4 | Mutism,Echopraxia/echolalia,Stereotypy,Negativism | 84.7 | 81.4 | 87.9 |
| 4 | Posturing/catalepsy,Verbigeration,Rigidity,Withdrawal | 84.7 | 81.2 | 88.1 |
| 4 | Excitement,Mutism,Mannerisms,Withdrawal | 84.5 | 81.2 | 87.9 |
| 4 | Excitement,Stereotypy,Rigidity,Withdrawal | 84.5 | 81.2 | 87.7 |
| 4 | Immobility/stupor,Echopraxia/echolalia,Mannerisms,Waxy flexibility | 84.5 | 80.9 | 87.9 |
| 4 | Mutism,Mannerisms,Rigidity,Waxy flexibility | 84.5 | 81.2 | 87.9 |
| 4 | Mutism,Posturing/catalepsy,Rigidity,Waxy flexibility | 84.5 | 81.2 | 87.9 |
| 4 | Posturing/catalepsy,Echopraxia/echolalia,Rigidity,Withdrawal | 84.5 | 81.2 | 87.9 |
| 4 | Posturing/catalepsy,Negativism,Waxy flexibility,Withdrawal | 84.5 | 80.9 | 87.7 |
| 4 | Grimacing,Rigidity,Negativism,Withdrawal | 84.3 | 80.9 | 87.7 |
| 4 | Immobility/stupor,Posturing/catalepsy,Echopraxia/echolalia,Waxy flexibility | 84.3 | 80.7 | 87.7 |
| 4 | Mutism,Mannerisms,Negativism,Waxy flexibility | 84.3 | 80.9 | 87.4 |
| 4 | Mutism,Stereotypy,Waxy flexibility,Withdrawal | 84.3 | 80.9 | 87.7 |
| 4 | Posturing/catalepsy,Grimacing,Rigidity,Withdrawal | 84.3 | 80.9 | 87.7 |
| 4 | Excitement,Echopraxia/echolalia,Rigidity,Withdrawal | 84.1 | 80.5 | 87.4 |
| 4 | Mutism,Grimacing,Echopraxia/echolalia,Rigidity | 84.1 | 80.7 | 87.4 |
| 4 | Mutism,Grimacing,Rigidity,Waxy flexibility | 84.1 | 80.7 | 87.2 |
| 4 | Mutism,Grimacing,Stereotypy,Negativism | 84.1 | 80.5 | 87.4 |
| 4 | Mutism,Mannerisms,Verbigeration,Rigidity | 84.1 | 80.5 | 87.4 |
| 4 | Mutism,Verbigeration,Rigidity,Waxy flexibility | 84.1 | 80.5 | 87.4 |
| 4 | Posturing/catalepsy,Mannerisms,Rigidity,Negativism | 84.1 | 80.7 | 87.4 |
| 4 | Excitement,Mutism,Mannerisms,Negativism | 83.9 | 80.5 | 87.2 |
| 4 | Excitement,Mutism,Stereotypy,Waxy flexibility | 83.9 | 80.3 | 87.2 |
| 4 | Mutism,Echopraxia/echolalia,Rigidity,Waxy flexibility | 83.9 | 80.5 | 87.0 |
| 4 | Posturing/catalepsy,Stereotypy,Rigidity,Negativism | 83.9 | 80.5 | 87.2 |
| 4 | Rigidity,Negativism,Waxy flexibility,Withdrawal | 83.9 | 80.3 | 87.2 |
| 4 | Mutism,Echopraxia/echolalia,Verbigeration,Rigidity | 83.8 | 80.3 | 87.2 |
| 4 | Excitement,Rigidity,Waxy flexibility,Withdrawal | 83.7 | 80.0 | 87.0 |
| 4 | Mutism,Echopraxia/echolalia,Negativism,Waxy flexibility | 83.6 | 80.0 | 87.0 |
| 4 | Mutism,Grimacing,Mannerisms,Negativism | 83.6 | 80.0 | 87.0 |
| 4 | Mutism,Mannerisms,Waxy flexibility,Withdrawal | 83.6 | 80.0 | 87.0 |
| 4 | Mutism,Posturing/catalepsy,Grimacing,Verbigeration | 83.6 | 80.0 | 87.0 |
| 4 | Mutism,Posturing/catalepsy,Stereotypy,Verbigeration | 83.6 | 80.3 | 87.0 |
| 4 | Excitement,Mutism,Echopraxia/echolalia,Waxy flexibility | 83.4 | 79.8 | 86.8 |
| 4 | Excitement,Stereotypy,Negativism,Withdrawal | 83.4 | 79.8 | 86.8 |
| 4 | Excitement,Verbigeration,Rigidity,Withdrawal | 83.4 | 79.8 | 86.8 |
| 4 | Mutism,Echopraxia/echolalia,Stereotypy,Withdrawal | 83.4 | 79.8 | 86.8 |
| 4 | Mutism,Echopraxia/echolalia,Verbigeration,Negativism | 83.4 | 79.8 | 86.8 |
| 4 | Mutism,Echopraxia/echolalia,Waxy flexibility,Withdrawal | 83.4 | 79.8 | 86.8 |
| 4 | Mutism,Grimacing,Stereotypy,Withdrawal | 83.4 | 79.8 | 86.8 |
| 4 | Mutism,Posturing/catalepsy,Echopraxia/echolalia,Mannerisms | 83.4 | 79.8 | 86.8 |
| 4 | Mutism,Stereotypy,Mannerisms,Withdrawal | 83.4 | 79.8 | 86.8 |
| 4 | Mutism,Verbigeration,Waxy flexibility,Withdrawal | 83.4 | 79.8 | 86.8 |
| 4 | Excitement,Mutism,Grimacing,Waxy flexibility | 83.2 | 79.8 | 86.5 |
| 4 | Mutism,Grimacing,Mannerisms,Withdrawal | 83.2 | 79.8 | 86.5 |
| 4 | Mutism,Grimacing,Verbigeration,Negativism | 83.2 | 79.6 | 86.5 |
| 4 | Mutism,Mannerisms,Verbigeration,Negativism | 83.2 | 79.6 | 86.5 |
| 4 | Mutism,Posturing/catalepsy,Echopraxia/echolalia,Stereotypy | 83.2 | 79.6 | 86.5 |
| 4 | Mutism,Posturing/catalepsy,Echopraxia/echolalia,Verbigeration | 83.2 | 79.6 | 86.5 |
| 4 | Mutism,Posturing/catalepsy,Grimacing,Mannerisms | 83.2 | 79.6 | 86.5 |
| 4 | Mutism,Posturing/catalepsy,Grimacing,Stereotypy | 83.2 | 79.6 | 86.5 |
| 4 | Mutism,Stereotypy,Verbigeration,Withdrawal | 83.2 | 79.8 | 86.5 |
| 4 | Posturing/catalepsy,Rigidity,Waxy flexibility,Withdrawal | 83.2 | 79.8 | 86.5 |
| 4 | Echopraxia/echolalia,Stereotypy,Negativism,Withdrawal | 83.1 | 79.6 | 86.5 |
| 4 | Mutism,Echopraxia/echolalia,Mannerisms,Withdrawal | 83.0 | 79.4 | 86.3 |
| 4 | Mutism,Grimacing,Verbigeration,Withdrawal | 83.0 | 79.4 | 86.3 |
| 4 | Mutism,Grimacing,Waxy flexibility,Withdrawal | 83.0 | 79.4 | 86.5 |
| 4 | Mutism,Posturing/catalepsy,Mannerisms,Verbigeration | 83.0 | 79.4 | 86.3 |
| 4 | Mutism,Verbigeration,Negativism,Waxy flexibility | 83.0 | 79.4 | 86.3 |
| 4 | Posturing/catalepsy,Grimacing,Stereotypy,Withdrawal | 83.0 | 79.4 | 86.3 |
| 4 | Stereotypy,Negativism,Waxy flexibility,Withdrawal | 83.0 | 79.6 | 86.3 |
| 4 | Mutism,Grimacing,Echopraxia/echolalia,Negativism | 82.9 | 79.4 | 86.3 |
| 4 | Mutism,Posturing/catalepsy,Grimacing,Echopraxia/echolalia | 82.9 | 79.4 | 86.3 |
| 4 | Stereotypy,Mannerisms,Negativism,Withdrawal | 82.9 | 79.4 | 86.3 |
| 4 | Posturing/catalepsy,Grimacing,Mannerisms,Withdrawal | 82.8 | 79.1 | 86.3 |
| 4 | Excitement,Grimacing,Rigidity,Withdrawal | 82.7 | 79.1 | 86.1 |
| 4 | Mutism,Posturing/catalepsy,Stereotypy,Mannerisms | 82.7 | 79.1 | 86.1 |
| 4 | Posturing/catalepsy,Verbigeration,Rigidity,Negativism | 82.7 | 79.1 | 86.3 |
| 4 | Stereotypy,Verbigeration,Negativism,Withdrawal | 82.7 | 79.1 | 86.1 |
| 4 | Echopraxia/echolalia,Stereotypy,Rigidity,Withdrawal | 82.5 | 78.9 | 86.1 |
| 4 | Excitement,Echopraxia/echolalia,Negativism,Withdrawal | 82.5 | 78.9 | 85.9 |
| 4 | Excitement,Mutism,Echopraxia/echolalia,Stereotypy | 82.5 | 78.9 | 85.9 |
| 4 | Mutism,Grimacing,Echopraxia/echolalia,Withdrawal | 82.5 | 78.9 | 86.1 |
| 4 | Mutism,Grimacing,Negativism,Waxy flexibility | 82.5 | 78.9 | 85.9 |
| 4 | Mutism,Mannerisms,Verbigeration,Withdrawal | 82.5 | 78.9 | 86.1 |
| 4 | Posturing/catalepsy,Grimacing,Rigidity,Negativism | 82.5 | 78.9 | 85.9 |
| 4 | Posturing/catalepsy,Stereotypy,Mannerisms,Withdrawal | 82.5 | 78.9 | 86.1 |
| 4 | Stereotypy,Mannerisms,Rigidity,Withdrawal | 82.5 | 78.9 | 85.9 |
| 4 | Stereotypy,Rigidity,Waxy flexibility,Withdrawal | 82.5 | 78.9 | 85.9 |
| 4 | Grimacing,Stereotypy,Negativism,Withdrawal | 82.3 | 78.7 | 85.9 |
| 4 | Grimacing,Stereotypy,Rigidity,Withdrawal | 82.3 | 78.7 | 85.9 |
| 4 | Mutism,Posturing/catalepsy,Stereotypy,Waxy flexibility | 82.3 | 78.7 | 85.7 |
| 4 | Mutism,Posturing/catalepsy,Verbigeration,Waxy flexibility | 82.3 | 78.7 | 85.9 |
| 4 | Posturing/catalepsy,Echopraxia/echolalia,Rigidity,Negativism | 82.3 | 78.7 | 85.7 |
| 4 | Excitement,Mannerisms,Rigidity,Withdrawal | 82.1 | 78.5 | 85.7 |
| 4 | Excitement,Negativism,Waxy flexibility,Withdrawal | 82.1 | 78.5 | 85.7 |
| 4 | Mutism,Echopraxia/echolalia,Verbigeration,Withdrawal | 82.1 | 78.3 | 85.7 |
| 4 | Mutism,Posturing/catalepsy,Echopraxia/echolalia,Waxy flexibility | 82.1 | 78.5 | 85.7 |
| 4 | Posturing/catalepsy,Grimacing,Echopraxia/echolalia,Withdrawal | 82.1 | 78.5 | 85.7 |
| 4 | Posturing/catalepsy,Mannerisms,Verbigeration,Withdrawal | 82.1 | 78.5 | 85.7 |
| 4 | Stereotypy,Verbigeration,Rigidity,Withdrawal | 82.1 | 78.5 | 85.4 |
| 4 | Excitement,Mutism,Grimacing,Stereotypy | 82.0 | 78.5 | 85.4 |
| 4 | Excitement,Posturing/catalepsy,Grimacing,Negativism | 82.0 | 78.5 | 85.7 |
| 4 | Excitement,Mutism,Grimacing,Echopraxia/echolalia | 81.9 | 78.3 | 85.4 |
| 4 | Excitement,Mutism,Verbigeration,Waxy flexibility | 81.9 | 78.3 | 85.4 |
| 4 | Posturing/catalepsy,Echopraxia/echolalia,Mannerisms,Withdrawal | 81.8 | 78.3 | 85.2 |
| 4 | Posturing/catalepsy,Echopraxia/echolalia,Stereotypy,Withdrawal | 81.8 | 78.0 | 85.4 |
| 4 | Posturing/catalepsy,Stereotypy,Verbigeration,Withdrawal | 81.8 | 78.3 | 85.4 |
| 4 | Excitement,Posturing/catalepsy,Stereotypy,Negativism | 81.6 | 77.8 | 85.2 |
| 4 | Mutism,Posturing/catalepsy,Grimacing,Waxy flexibility | 81.6 | 78.0 | 85.2 |
| 4 | Mutism,Posturing/catalepsy,Mannerisms,Waxy flexibility | 81.6 | 78.0 | 85.2 |
| 4 | Posturing/catalepsy,Grimacing,Verbigeration,Withdrawal | 81.6 | 78.0 | 85.2 |
| 4 | Excitement,Mutism,Stereotypy,Verbigeration | 81.4 | 77.8 | 85.0 |
| 4 | Excitement,Verbigeration,Negativism,Withdrawal | 81.4 | 77.8 | 85.0 |
| 4 | Posturing/catalepsy,Stereotypy,Waxy flexibility,Withdrawal | 81.4 | 77.8 | 85.0 |
| 4 | Excitement,Mutism,Grimacing,Verbigeration | 81.2 | 77.4 | 84.8 |
| 4 | Excitement,Mutism,Mannerisms,Waxy flexibility | 81.2 | 77.6 | 84.8 |
| 4 | Excitement,Posturing/catalepsy,Echopraxia/echolalia,Negativism | 81.2 | 77.4 | 84.8 |
| 4 | Mannerisms,Negativism,Waxy flexibility,Withdrawal | 81.2 | 77.6 | 84.8 |
| 4 | Excitement,Grimacing,Negativism,Withdrawal | 81.1 | 77.6 | 84.8 |
| 4 | Echopraxia/echolalia,Mannerisms,Negativism,Withdrawal | 81.0 | 77.4 | 84.5 |
| 4 | Grimacing,Mannerisms,Rigidity,Withdrawal | 81.0 | 77.4 | 84.5 |
| 4 | Posturing/catalepsy,Mannerisms,Waxy flexibility,Withdrawal | 81.0 | 77.1 | 84.5 |
| 4 | Verbigeration,Rigidity,Waxy flexibility,Withdrawal | 81.0 | 77.4 | 84.5 |
| 4 | Grimacing,Verbigeration,Rigidity,Withdrawal | 80.9 | 77.1 | 84.5 |
| 4 | Mannerisms,Verbigeration,Rigidity,Withdrawal | 80.9 | 77.1 | 84.5 |
| 4 | Posturing/catalepsy,Echopraxia/echolalia,Verbigeration,Withdrawal | 80.9 | 77.1 | 84.5 |
| 4 | Echopraxia/echolalia,Mannerisms,Rigidity,Withdrawal | 80.7 | 76.9 | 84.3 |
| 4 | Echopraxia/echolalia,Negativism,Waxy flexibility,Withdrawal | 80.7 | 76.9 | 84.3 |
| 4 | Excitement,Mutism,Echopraxia/echolalia,Verbigeration | 80.7 | 77.1 | 84.3 |
| 4 | Excitement,Mutism,Stereotypy,Mannerisms | 80.7 | 76.9 | 84.3 |
| 4 | Grimacing,Echopraxia/echolalia,Rigidity,Withdrawal | 80.7 | 77.1 | 84.3 |
| 4 | Posturing/catalepsy,Rigidity,Negativism,Waxy flexibility | 80.7 | 76.9 | 84.3 |
| 4 | Echopraxia/echolalia,Verbigeration,Rigidity,Withdrawal | 80.5 | 76.7 | 84.1 |
| 4 | Excitement,Posturing/catalepsy,Verbigeration,Negativism | 80.5 | 76.9 | 84.1 |
| 4 | Mannerisms,Rigidity,Waxy flexibility,Withdrawal | 80.5 | 76.7 | 84.1 |
| 4 | Mannerisms,Verbigeration,Negativism,Withdrawal | 80.5 | 76.7 | 84.1 |
| 4 | Posturing/catalepsy,Grimacing,Stereotypy,Negativism | 80.5 | 76.7 | 84.1 |
| 4 | Echopraxia/echolalia,Rigidity,Waxy flexibility,Withdrawal | 80.3 | 76.5 | 83.9 |
| 4 | Excitement,Mannerisms,Negativism,Withdrawal | 80.3 | 76.5 | 83.9 |
| 4 | Excitement,Posturing/catalepsy,Negativism,Waxy flexibility | 80.3 | 76.5 | 83.9 |
| 4 | Grimacing,Mannerisms,Negativism,Withdrawal | 80.3 | 76.5 | 83.9 |
| 4 | Mutism,Echopraxia/echolalia,Stereotypy,Waxy flexibility | 80.3 | 76.5 | 83.9 |
| 4 | Posturing/catalepsy,Grimacing,Waxy flexibility,Withdrawal | 80.3 | 76.5 | 83.9 |
| 4 | Posturing/catalepsy,Verbigeration,Waxy flexibility,Withdrawal | 80.3 | 76.5 | 83.9 |
| 4 | Verbigeration,Negativism,Waxy flexibility,Withdrawal | 80.3 | 76.5 | 83.9 |
| 4 | Excitement,Posturing/catalepsy,Grimacing,Rigidity | 80.1 | 76.2 | 83.6 |
| 4 | Posturing/catalepsy,Stereotypy,Verbigeration,Negativism | 80.1 | 76.2 | 83.6 |
| 4 | Echopraxia/echolalia,Verbigeration,Negativism,Withdrawal | 80.0 | 76.2 | 83.6 |
| 4 | Excitement,Posturing/catalepsy,Mannerisms,Negativism | 79.9 | 76.0 | 83.4 |
| 4 | Excitement,Mutism,Echopraxia/echolalia,Mannerisms | 79.8 | 76.0 | 83.4 |
| 4 | Excitement,Mutism,Grimacing,Mannerisms | 79.8 | 76.0 | 83.4 |
| 4 | Excitement,Stereotypy,Waxy flexibility,Withdrawal | 79.8 | 76.2 | 83.4 |
| 4 | Grimacing,Echopraxia/echolalia,Negativism,Withdrawal | 79.8 | 76.0 | 83.6 |
| 4 | Grimacing,Rigidity,Waxy flexibility,Withdrawal | 79.8 | 76.0 | 83.4 |
| 4 | Grimacing,Verbigeration,Negativism,Withdrawal | 79.8 | 76.0 | 83.4 |
| 4 | Mutism,Grimacing,Stereotypy,Waxy flexibility | 79.8 | 76.0 | 83.4 |
| 4 | Mutism,Stereotypy,Mannerisms,Waxy flexibility | 79.8 | 76.0 | 83.4 |
| 4 | Posturing/catalepsy,Echopraxia/echolalia,Waxy flexibility,Withdrawal | 79.8 | 76.0 | 83.6 |
| 4 | Posturing/catalepsy,Grimacing,Mannerisms,Negativism | 79.8 | 76.0 | 83.4 |
| 4 | Posturing/catalepsy,Stereotypy,Mannerisms,Negativism | 79.8 | 76.0 | 83.4 |
| 4 | Excitement,Posturing/catalepsy,Echopraxia/echolalia,Rigidity | 79.6 | 75.8 | 83.2 |
| 4 | Mutism,Grimacing,Echopraxia/echolalia,Stereotypy | 79.6 | 75.8 | 83.4 |
| 4 | Mutism,Grimacing,Stereotypy,Verbigeration | 79.6 | 75.8 | 83.2 |
| 4 | Mutism,Stereotypy,Verbigeration,Waxy flexibility | 79.6 | 75.8 | 83.2 |
| 4 | Posturing/catalepsy,Echopraxia/echolalia,Stereotypy,Negativism | 79.6 | 75.8 | 83.2 |
| 4 | Posturing/catalepsy,Grimacing,Verbigeration,Negativism | 79.6 | 75.8 | 83.2 |
| 4 | Mutism,Echopraxia/echolalia,Stereotypy,Mannerisms | 79.4 | 75.6 | 83.0 |
| 4 | Mutism,Grimacing,Echopraxia/echolalia,Waxy flexibility | 79.4 | 75.6 | 83.0 |
| 4 | Mutism,Grimacing,Verbigeration,Waxy flexibility | 79.4 | 75.6 | 83.2 |
| 4 | Posturing/catalepsy,Grimacing,Echopraxia/echolalia,Negativism | 79.4 | 75.6 | 83.2 |
| 4 | Posturing/catalepsy,Stereotypy,Negativism,Waxy flexibility | 79.4 | 75.6 | 83.2 |
| 4 | Excitement,Echopraxia/echolalia,Waxy flexibility,Withdrawal | 79.3 | 75.6 | 83.0 |
| 4 | Mutism,Echopraxia/echolalia,Mannerisms,Waxy flexibility | 79.2 | 75.3 | 83.0 |
| 4 | Grimacing,Negativism,Waxy flexibility,Withdrawal | 79.1 | 75.3 | 83.0 |
| 4 | Excitement,Posturing/catalepsy,Verbigeration,Rigidity | 78.9 | 75.1 | 82.5 |
| 4 | Mutism,Grimacing,Echopraxia/echolalia,Verbigeration | 78.9 | 75.1 | 82.7 |
| 4 | Mutism,Grimacing,Mannerisms,Waxy flexibility | 78.9 | 75.1 | 82.5 |
| 4 | Mutism,Grimacing,Stereotypy,Mannerisms | 78.9 | 75.1 | 82.5 |
| 4 | Excitement,Stereotypy,Rigidity,Negativism | 78.7 | 74.9 | 82.5 |
| 4 | Mutism,Grimacing,Echopraxia/echolalia,Mannerisms | 78.7 | 74.9 | 82.5 |
| 4 | Posturing/catalepsy,Echopraxia/echolalia,Mannerisms,Negativism | 78.7 | 74.9 | 82.5 |
| 4 | Stereotypy,Rigidity,Negativism,Waxy flexibility | 78.7 | 74.9 | 82.3 |
| 4 | Excitement,Mutism,Mannerisms,Verbigeration | 78.5 | 74.7 | 82.3 |
| 4 | Excitement,Posturing/catalepsy,Stereotypy,Rigidity | 78.5 | 74.7 | 82.3 |
| 4 | Mutism,Echopraxia/echolalia,Stereotypy,Verbigeration | 78.5 | 74.7 | 82.3 |
| 4 | Mutism,Stereotypy,Mannerisms,Verbigeration | 78.5 | 74.7 | 82.3 |
| 4 | Stereotypy,Mannerisms,Rigidity,Negativism | 78.5 | 74.7 | 82.3 |
| 4 | Mutism,Echopraxia/echolalia,Verbigeration,Waxy flexibility | 78.4 | 74.7 | 82.1 |
| 4 | Excitement,Posturing/catalepsy,Rigidity,Waxy flexibility | 78.3 | 74.2 | 82.1 |
| 4 | Posturing/catalepsy,Grimacing,Negativism,Waxy flexibility | 78.3 | 74.4 | 82.1 |
| 4 | Excitement,Echopraxia/echolalia,Stereotypy,Withdrawal | 78.2 | 74.2 | 82.1 |
| 4 | Grimacing,Stereotypy,Rigidity,Negativism | 78.2 | 74.4 | 82.1 |
| 4 | Excitement,Verbigeration,Waxy flexibility,Withdrawal | 78.0 | 74.2 | 81.8 |
| 4 | Mutism,Grimacing,Mannerisms,Verbigeration | 78.0 | 74.0 | 81.8 |
| 4 | Posturing/catalepsy,Mannerisms,Verbigeration,Negativism | 78.0 | 74.2 | 81.8 |
| 4 | Stereotypy,Verbigeration,Rigidity,Negativism | 78.0 | 74.2 | 81.8 |
| 4 | Excitement,Grimacing,Rigidity,Negativism | 77.8 | 74.0 | 81.6 |
| 4 | Excitement,Posturing/catalepsy,Mannerisms,Rigidity | 77.8 | 73.8 | 81.6 |
| 4 | Mutism,Mannerisms,Verbigeration,Waxy flexibility | 77.8 | 74.0 | 81.6 |
| 4 | Posturing/catalepsy,Echopraxia/echolalia,Verbigeration,Negativism | 77.8 | 74.0 | 81.6 |
| 4 | Posturing/catalepsy,Mannerisms,Negativism,Waxy flexibility | 77.8 | 73.8 | 81.6 |
| 4 | Echopraxia/echolalia,Stereotypy,Rigidity,Negativism | 77.6 | 73.5 | 81.6 |
| 4 | Excitement,Grimacing,Waxy flexibility,Withdrawal | 77.6 | 73.8 | 81.4 |
| 4 | Excitement,Grimacing,Echopraxia/echolalia,Withdrawal | 77.3 | 73.3 | 81.2 |
| 4 | Excitement,Grimacing,Stereotypy,Withdrawal | 77.3 | 73.3 | 81.2 |
| 4 | Mutism,Echopraxia/echolalia,Mannerisms,Verbigeration | 77.3 | 73.3 | 81.2 |
| 4 | Posturing/catalepsy,Grimacing,Verbigeration,Rigidity | 77.2 | 73.3 | 80.9 |
| 4 | Echopraxia/echolalia,Stereotypy,Waxy flexibility,Withdrawal | 77.1 | 73.1 | 80.9 |
| 4 | Excitement,Echopraxia/echolalia,Rigidity,Negativism | 77.1 | 73.1 | 80.9 |
| 4 | Excitement,Stereotypy,Verbigeration,Withdrawal | 77.1 | 73.1 | 80.9 |
| 4 | Grimacing,Echopraxia/echolalia,Stereotypy,Withdrawal | 77.1 | 73.1 | 80.9 |
| 4 | Grimacing,Mannerisms,Rigidity,Negativism | 77.1 | 73.1 | 80.9 |
| 4 | Grimacing,Stereotypy,Waxy flexibility,Withdrawal | 77.1 | 73.1 | 80.9 |
| 4 | Posturing/catalepsy,Echopraxia/echolalia,Negativism,Waxy flexibility | 77.1 | 73.1 | 80.9 |
| 4 | Excitement,Echopraxia/echolalia,Verbigeration,Withdrawal | 76.9 | 72.9 | 80.7 |
| 4 | Excitement,Rigidity,Negativism,Waxy flexibility | 76.9 | 72.9 | 80.7 |
| 4 | Posturing/catalepsy,Grimacing,Mannerisms,Rigidity | 76.9 | 72.9 | 80.7 |
| 4 | Posturing/catalepsy,Verbigeration,Negativism,Waxy flexibility | 76.9 | 72.9 | 80.9 |
| 4 | Posturing/catalepsy,Grimacing,Echopraxia/echolalia,Rigidity | 76.7 | 72.6 | 80.5 |
| 4 | Posturing/catalepsy,Grimacing,Stereotypy,Rigidity | 76.7 | 72.6 | 80.5 |
| 4 | Excitement,Mannerisms,Waxy flexibility,Withdrawal | 76.5 | 72.4 | 80.3 |
| 4 | Stereotypy,Mannerisms,Waxy flexibility,Withdrawal | 76.4 | 72.4 | 80.3 |
| 4 | Excitement,Grimacing,Verbigeration,Withdrawal | 76.2 | 72.2 | 80.0 |
| 4 | Grimacing,Echopraxia/echolalia,Rigidity,Negativism | 76.2 | 72.2 | 80.0 |
| 4 | Grimacing,Rigidity,Negativism,Waxy flexibility | 76.2 | 72.2 | 80.0 |
| 4 | Grimacing,Verbigeration,Rigidity,Negativism | 76.2 | 72.2 | 80.0 |
| 4 | Mannerisms,Rigidity,Negativism,Waxy flexibility | 76.2 | 72.0 | 80.0 |
| 4 | Stereotypy,Verbigeration,Waxy flexibility,Withdrawal | 76.2 | 72.2 | 80.0 |
| 4 | Echopraxia/echolalia,Mannerisms,Rigidity,Negativism | 76.0 | 72.0 | 79.8 |
| 4 | Excitement,Stereotypy,Mannerisms,Withdrawal | 76.0 | 72.0 | 79.8 |
| 4 | Grimacing,Echopraxia/echolalia,Waxy flexibility,Withdrawal | 76.0 | 72.0 | 79.8 |
| 4 | Grimacing,Stereotypy,Mannerisms,Withdrawal | 76.0 | 72.0 | 80.0 |
| 4 | Grimacing,Stereotypy,Verbigeration,Withdrawal | 76.0 | 72.0 | 79.8 |
| 4 | Posturing/catalepsy,Stereotypy,Verbigeration,Rigidity | 76.0 | 72.0 | 80.0 |
| 4 | Excitement,Posturing/catalepsy,Grimacing,Echopraxia/echolalia | 75.8 | 71.7 | 79.6 |
| 4 | Posturing/catalepsy,Echopraxia/echolalia,Stereotypy,Rigidity | 75.8 | 71.7 | 79.8 |
| 4 | Posturing/catalepsy,Echopraxia/echolalia,Mannerisms,Rigidity | 75.5 | 71.5 | 79.4 |
| 4 | Excitement,Echopraxia/echolalia,Mannerisms,Withdrawal | 75.4 | 71.3 | 79.4 |
| 4 | Excitement,Verbigeration,Rigidity,Negativism | 75.4 | 71.3 | 79.1 |
| 4 | Echopraxia/echolalia,Stereotypy,Mannerisms,Withdrawal | 75.3 | 71.3 | 79.4 |
| 4 | Posturing/catalepsy,Mannerisms,Verbigeration,Rigidity | 75.3 | 71.3 | 79.1 |
| 4 | Posturing/catalepsy,Stereotypy,Mannerisms,Rigidity | 75.3 | 71.3 | 79.1 |
| 4 | Excitement,Mannerisms,Rigidity,Negativism | 75.1 | 71.1 | 79.1 |
| 4 | Mannerisms,Verbigeration,Rigidity,Negativism | 75.1 | 70.9 | 79.1 |
| 4 | Posturing/catalepsy,Echopraxia/echolalia,Verbigeration,Rigidity | 75.1 | 71.1 | 79.1 |
| 4 | Posturing/catalepsy,Grimacing,Rigidity,Waxy flexibility | 75.1 | 71.1 | 79.1 |
| 4 | Echopraxia/echolalia,Rigidity,Negativism,Waxy flexibility | 74.9 | 70.9 | 78.9 |
| 4 | Echopraxia/echolalia,Stereotypy,Verbigeration,Withdrawal | 74.9 | 70.9 | 78.7 |
| 4 | Excitement,Mannerisms,Verbigeration,Withdrawal | 74.9 | 70.9 | 78.7 |
| 4 | Grimacing,Echopraxia/echolalia,Verbigeration,Withdrawal | 74.9 | 70.9 | 78.9 |
| 4 | Grimacing,Verbigeration,Waxy flexibility,Withdrawal | 74.9 | 70.9 | 78.9 |
| 4 | Grimacing,Mannerisms,Waxy flexibility,Withdrawal | 74.8 | 70.6 | 78.9 |
| 4 | Excitement,Posturing/catalepsy,Grimacing,Verbigeration | 74.7 | 70.6 | 78.7 |
| 4 | Stereotypy,Mannerisms,Verbigeration,Withdrawal | 74.7 | 70.6 | 78.7 |
| 4 | Verbigeration,Rigidity,Negativism,Waxy flexibility | 74.7 | 70.6 | 78.7 |
| 4 | Echopraxia/echolalia,Verbigeration,Waxy flexibility,Withdrawal | 74.6 | 70.6 | 78.7 |
| 4 | Excitement,Posturing/catalepsy,Grimacing,Stereotypy | 74.5 | 70.4 | 78.5 |
| 4 | Grimacing,Echopraxia/echolalia,Mannerisms,Withdrawal | 74.5 | 70.2 | 78.5 |
| 4 | Posturing/catalepsy,Stereotypy,Rigidity,Waxy flexibility | 74.5 | 70.4 | 78.5 |
| 4 | Echopraxia/echolalia,Mannerisms,Waxy flexibility,Withdrawal | 74.4 | 70.4 | 78.5 |
| 4 | Echopraxia/echolalia,Verbigeration,Rigidity,Negativism | 74.4 | 70.4 | 78.3 |
| 4 | Excitement,Grimacing,Mannerisms,Withdrawal | 74.4 | 70.4 | 78.5 |
| 4 | Grimacing,Mannerisms,Verbigeration,Withdrawal | 74.4 | 70.4 | 78.5 |
| 4 | Mannerisms,Verbigeration,Waxy flexibility,Withdrawal | 74.4 | 70.4 | 78.5 |
| 4 | Excitement,Posturing/catalepsy,Grimacing,Waxy flexibility | 74.2 | 70.0 | 78.3 |
| 4 | Posturing/catalepsy,Mannerisms,Rigidity,Waxy flexibility | 74.0 | 69.7 | 78.0 |
| 4 | Posturing/catalepsy,Echopraxia/echolalia,Rigidity,Waxy flexibility | 73.8 | 69.7 | 77.8 |
| 4 | Posturing/catalepsy,Verbigeration,Rigidity,Waxy flexibility | 73.5 | 69.3 | 77.6 |
| 4 | Excitement,Posturing/catalepsy,Grimacing,Mannerisms | 73.3 | 69.1 | 77.1 |
| 4 | Echopraxia/echolalia,Mannerisms,Verbigeration,Withdrawal | 73.1 | 68.8 | 77.4 |
| 4 | Excitement,Stereotypy,Negativism,Waxy flexibility | 73.1 | 69.1 | 77.1 |
| 4 | Grimacing,Stereotypy,Negativism,Waxy flexibility | 72.6 | 68.6 | 76.7 |
| 4 | Stereotypy,Mannerisms,Negativism,Waxy flexibility | 72.6 | 68.6 | 76.7 |
| 4 | Excitement,Posturing/catalepsy,Echopraxia/echolalia,Stereotypy | 72.4 | 68.2 | 76.5 |
| 4 | Excitement,Posturing/catalepsy,Echopraxia/echolalia,Waxy flexibility | 72.4 | 68.2 | 76.5 |
| 4 | Excitement,Posturing/catalepsy,Stereotypy,Verbigeration | 72.4 | 68.2 | 76.7 |
| 4 | Posturing/catalepsy,Grimacing,Stereotypy,Verbigeration | 72.4 | 68.2 | 76.7 |
| 4 | Excitement,Grimacing,Stereotypy,Rigidity | 72.2 | 68.2 | 76.5 |
| 4 | Excitement,Posturing/catalepsy,Echopraxia/echolalia,Verbigeration | 72.2 | 67.9 | 76.2 |
| 4 | Excitement,Grimacing,Echopraxia/echolalia,Rigidity | 72.0 | 67.7 | 76.0 |
| 4 | Excitement,Posturing/catalepsy,Stereotypy,Waxy flexibility | 72.0 | 67.7 | 76.2 |
| 4 | Posturing/catalepsy,Grimacing,Echopraxia/echolalia,Stereotypy | 72.0 | 67.7 | 76.0 |
| 4 | Excitement,Stereotypy,Rigidity,Waxy flexibility | 71.8 | 67.5 | 76.0 |
| 4 | Echopraxia/echolalia,Stereotypy,Negativism,Waxy flexibility | 71.7 | 67.5 | 75.8 |
| 4 | Excitement,Grimacing,Negativism,Waxy flexibility | 71.7 | 67.5 | 75.8 |
| 4 | Excitement,Grimacing,Stereotypy,Negativism | 71.6 | 67.5 | 75.8 |
| 4 | Excitement,Posturing/catalepsy,Verbigeration,Waxy flexibility | 71.6 | 67.3 | 75.8 |
| 4 | Excitement,Grimacing,Rigidity,Waxy flexibility | 71.5 | 67.3 | 75.6 |
| 4 | Grimacing,Stereotypy,Rigidity,Waxy flexibility | 71.5 | 67.3 | 75.6 |
| 4 | Grimacing,Stereotypy,Mannerisms,Negativism | 71.4 | 67.0 | 75.6 |
| 4 | Excitement,Echopraxia/echolalia,Stereotypy,Negativism | 71.3 | 67.0 | 75.6 |
| 4 | Posturing/catalepsy,Grimacing,Echopraxia/echolalia,Verbigeration | 71.3 | 67.0 | 75.3 |
| 4 | Stereotypy,Verbigeration,Negativism,Waxy flexibility | 71.3 | 67.0 | 75.6 |
| 4 | Grimacing,Echopraxia/echolalia,Stereotypy,Negativism | 71.1 | 66.8 | 75.3 |
| 4 | Posturing/catalepsy,Grimacing,Stereotypy,Mannerisms | 71.1 | 67.0 | 75.3 |
| 4 | Excitement,Echopraxia/echolalia,Stereotypy,Rigidity | 70.9 | 66.6 | 74.9 |
| 4 | Excitement,Posturing/catalepsy,Echopraxia/echolalia,Mannerisms | 70.9 | 66.6 | 75.1 |
| 4 | Posturing/catalepsy,Grimacing,Echopraxia/echolalia,Mannerisms | 70.9 | 66.6 | 75.1 |
| 4 | Posturing/catalepsy,Grimacing,Mannerisms,Verbigeration | 70.9 | 66.6 | 74.9 |
| 4 | Posturing/catalepsy,Grimacing,Stereotypy,Waxy flexibility | 70.8 | 66.6 | 74.9 |
| 4 | Echopraxia/echolalia,Stereotypy,Mannerisms,Negativism | 70.7 | 66.4 | 74.9 |
| 4 | Excitement,Grimacing,Echopraxia/echolalia,Negativism | 70.6 | 66.4 | 74.9 |
| 4 | Excitement,Echopraxia/echolalia,Negativism,Waxy flexibility | 70.4 | 66.1 | 74.7 |
| 4 | Excitement,Posturing/catalepsy,Stereotypy,Mannerisms | 70.4 | 66.1 | 74.7 |
| 4 | Grimacing,Echopraxia/echolalia,Stereotypy,Rigidity | 70.4 | 65.9 | 74.7 |
| 4 | Grimacing,Stereotypy,Verbigeration,Negativism | 70.4 | 66.1 | 74.7 |
| 4 | Grimacing,Stereotypy,Verbigeration,Rigidity | 70.4 | 66.1 | 74.7 |
| 4 | Posturing/catalepsy,Grimacing,Verbigeration,Waxy flexibility | 70.4 | 66.1 | 74.7 |
| 4 | Excitement,Grimacing,Verbigeration,Rigidity | 70.2 | 65.9 | 74.4 |
| 4 | Excitement,Posturing/catalepsy,Mannerisms,Verbigeration | 70.2 | 65.9 | 74.4 |
| 4 | Excitement,Stereotypy,Verbigeration,Negativism | 70.2 | 65.9 | 74.4 |
| 4 | Grimacing,Echopraxia/echolalia,Negativism,Waxy flexibility | 70.2 | 65.9 | 74.4 |
| 4 | Grimacing,Stereotypy,Mannerisms,Rigidity | 70.2 | 65.9 | 74.4 |
| 4 | Excitement,Echopraxia/echolalia,Rigidity,Waxy flexibility | 69.9 | 65.7 | 74.0 |
| 4 | Grimacing,Mannerisms,Negativism,Waxy flexibility | 69.9 | 65.7 | 74.2 |
| 4 | Posturing/catalepsy,Grimacing,Echopraxia/echolalia,Waxy flexibility | 69.9 | 65.7 | 74.2 |
| 4 | Grimacing,Echopraxia/echolalia,Rigidity,Waxy flexibility | 69.7 | 65.5 | 73.8 |
| 4 | Excitement,Posturing/catalepsy,Mannerisms,Waxy flexibility | 69.5 | 65.2 | 73.8 |
| 4 | Grimacing,Verbigeration,Rigidity,Waxy flexibility | 69.5 | 65.2 | 73.8 |
| 4 | Excitement,Stereotypy,Mannerisms,Negativism | 69.3 | 64.8 | 73.5 |
| 4 | Excitement,Stereotypy,Verbigeration,Rigidity | 69.3 | 65.0 | 73.5 |
| 4 | Stereotypy,Mannerisms,Verbigeration,Negativism | 69.3 | 65.0 | 73.5 |
| 4 | Echopraxia/echolalia,Stereotypy,Rigidity,Waxy flexibility | 69.1 | 64.8 | 73.3 |
| 4 | Grimacing,Verbigeration,Negativism,Waxy flexibility | 69.0 | 64.6 | 73.3 |
| 4 | Posturing/catalepsy,Grimacing,Mannerisms,Waxy flexibility | 69.0 | 64.8 | 73.3 |
| 4 | Posturing/catalepsy,Echopraxia/echolalia,Stereotypy,Verbigeration | 68.9 | 64.6 | 73.1 |
| 4 | Posturing/catalepsy,Stereotypy,Verbigeration,Waxy flexibility | 68.9 | 64.6 | 73.1 |
| 4 | Echopraxia/echolalia,Stereotypy,Verbigeration,Negativism | 68.8 | 64.3 | 73.1 |
| 4 | Grimacing,Echopraxia/echolalia,Mannerisms,Negativism | 68.8 | 64.3 | 73.1 |
| 4 | Grimacing,Echopraxia/echolalia,Verbigeration,Rigidity | 68.8 | 64.6 | 73.1 |
| 4 | Echopraxia/echolalia,Mannerisms,Negativism,Waxy flexibility | 68.6 | 64.3 | 72.9 |
| 4 | Excitement,Grimacing,Verbigeration,Negativism | 68.6 | 64.3 | 72.9 |
| 4 | Grimacing,Echopraxia/echolalia,Mannerisms,Rigidity | 68.6 | 64.3 | 72.9 |
| 4 | Posturing/catalepsy,Echopraxia/echolalia,Stereotypy,Waxy flexibility | 68.6 | 64.3 | 72.9 |
| 4 | Posturing/catalepsy,Stereotypy,Mannerisms,Verbigeration | 68.6 | 64.3 | 72.9 |
| 4 | Stereotypy,Verbigeration,Rigidity,Waxy flexibility | 68.6 | 64.3 | 72.9 |
| 4 | Excitement,Grimacing,Mannerisms,Rigidity | 68.4 | 64.1 | 72.6 |
| 4 | Excitement,Mannerisms,Negativism,Waxy flexibility | 68.4 | 63.9 | 72.6 |
| 4 | Excitement,Verbigeration,Negativism,Waxy flexibility | 68.4 | 64.1 | 72.6 |
| 4 | Excitement,Verbigeration,Rigidity,Waxy flexibility | 68.4 | 64.1 | 72.9 |
| 4 | Grimacing,Mannerisms,Rigidity,Waxy flexibility | 68.4 | 64.1 | 72.6 |
| 4 | Posturing/catalepsy,Echopraxia/echolalia,Stereotypy,Mannerisms | 68.4 | 64.1 | 72.6 |
| 4 | Stereotypy,Mannerisms,Rigidity,Waxy flexibility | 68.4 | 64.1 | 72.6 |
| 4 | Excitement,Stereotypy,Mannerisms,Rigidity | 68.2 | 63.9 | 72.4 |
| 4 | Grimacing,Echopraxia/echolalia,Verbigeration,Negativism | 68.1 | 63.7 | 72.4 |
| 4 | Excitement,Echopraxia/echolalia,Verbigeration,Rigidity | 68.0 | 63.7 | 72.2 |
| 4 | Excitement,Grimacing,Mannerisms,Negativism | 67.9 | 63.5 | 72.2 |
| 4 | Grimacing,Mannerisms,Verbigeration,Rigidity | 67.7 | 63.2 | 72.0 |
| 4 | Echopraxia/echolalia,Stereotypy,Mannerisms,Rigidity | 67.5 | 63.2 | 71.7 |
| 4 | Posturing/catalepsy,Stereotypy,Mannerisms,Waxy flexibility | 67.3 | 63.0 | 71.5 |
| 4 | Echopraxia/echolalia,Stereotypy,Verbigeration,Rigidity | 67.2 | 62.8 | 71.5 |
| 4 | Posturing/catalepsy,Echopraxia/echolalia,Mannerisms,Verbigeration | 67.1 | 62.6 | 71.3 |
| 4 | Grimacing,Mannerisms,Verbigeration,Negativism | 67.0 | 62.6 | 71.3 |
| 4 | Echopraxia/echolalia,Verbigeration,Negativism,Waxy flexibility | 66.8 | 62.3 | 71.1 |
| 4 | Excitement,Echopraxia/echolalia,Mannerisms,Rigidity | 66.8 | 62.3 | 71.1 |
| 4 | Excitement,Echopraxia/echolalia,Verbigeration,Negativism | 66.8 | 62.3 | 71.1 |
| 4 | Mannerisms,Verbigeration,Negativism,Waxy flexibility | 66.8 | 62.3 | 71.1 |
| 4 | Stereotypy,Mannerisms,Verbigeration,Rigidity | 66.8 | 62.6 | 71.1 |
| 4 | Excitement,Echopraxia/echolalia,Mannerisms,Negativism | 66.6 | 62.1 | 70.9 |
| 4 | Posturing/catalepsy,Echopraxia/echolalia,Verbigeration,Waxy flexibility | 66.4 | 62.1 | 70.6 |
| 4 | Posturing/catalepsy,Mannerisms,Verbigeration,Waxy flexibility | 66.2 | 61.9 | 70.4 |
| 4 | Excitement,Mannerisms,Rigidity,Waxy flexibility | 66.1 | 61.7 | 70.4 |
| 4 | Posturing/catalepsy,Echopraxia/echolalia,Mannerisms,Waxy flexibility | 65.9 | 61.4 | 70.2 |
| 4 | Echopraxia/echolalia,Mannerisms,Rigidity,Waxy flexibility | 65.3 | 61.0 | 69.7 |
| 4 | Echopraxia/echolalia,Verbigeration,Rigidity,Waxy flexibility | 65.3 | 60.8 | 69.5 |
| 4 | Excitement,Mannerisms,Verbigeration,Rigidity | 65.1 | 60.5 | 69.3 |
| 4 | Echopraxia/echolalia,Mannerisms,Verbigeration,Negativism | 65.0 | 60.5 | 69.3 |
| 4 | Mannerisms,Verbigeration,Rigidity,Waxy flexibility | 64.6 | 60.1 | 69.1 |
| 4 | Excitement,Mannerisms,Verbigeration,Negativism | 64.3 | 59.9 | 68.8 |
| 4 | Echopraxia/echolalia,Mannerisms,Verbigeration,Rigidity | 64.1 | 59.6 | 68.6 |
| 4 | Excitement,Grimacing,Stereotypy,Waxy flexibility | 63.5 | 59.0 | 67.9 |
| 4 | Excitement,Grimacing,Echopraxia/echolalia,Waxy flexibility | 62.6 | 57.8 | 67.0 |
| 4 | Grimacing,Echopraxia/echolalia,Stereotypy,Waxy flexibility | 62.1 | 57.6 | 66.6 |
| 4 | Grimacing,Stereotypy,Verbigeration,Waxy flexibility | 61.5 | 57.0 | 65.9 |
| 4 | Excitement,Echopraxia/echolalia,Stereotypy,Waxy flexibility | 61.0 | 56.5 | 65.5 |
| 4 | Excitement,Grimacing,Verbigeration,Waxy flexibility | 60.7 | 56.3 | 65.2 |
| 4 | Grimacing,Stereotypy,Mannerisms,Waxy flexibility | 60.5 | 55.8 | 65.0 |
| 4 | Excitement,Stereotypy,Verbigeration,Waxy flexibility | 59.9 | 55.2 | 64.3 |
| 4 | Excitement,Grimacing,Echopraxia/echolalia,Stereotypy | 59.2 | 54.7 | 63.7 |
| 4 | Grimacing,Echopraxia/echolalia,Verbigeration,Waxy flexibility | 59.0 | 54.5 | 63.5 |
| 4 | Excitement,Grimacing,Mannerisms,Waxy flexibility | 58.1 | 53.6 | 62.6 |
| 4 | Excitement,Stereotypy,Mannerisms,Waxy flexibility | 57.9 | 53.4 | 62.3 |
| 4 | Excitement,Grimacing,Stereotypy,Verbigeration | 57.8 | 53.4 | 62.3 |
| 4 | Grimacing,Echopraxia/echolalia,Mannerisms,Waxy flexibility | 57.6 | 52.9 | 62.3 |
| 4 | Excitement,Grimacing,Echopraxia/echolalia,Verbigeration | 57.2 | 52.7 | 61.7 |
| 4 | Grimacing,Echopraxia/echolalia,Stereotypy,Verbigeration | 57.2 | 52.7 | 61.7 |
| 4 | Excitement,Echopraxia/echolalia,Verbigeration,Waxy flexibility | 57.1 | 52.5 | 61.7 |
| 4 | Echopraxia/echolalia,Stereotypy,Mannerisms,Waxy flexibility | 57.0 | 52.5 | 61.4 |
| 4 | Grimacing,Echopraxia/echolalia,Stereotypy,Mannerisms | 57.0 | 52.2 | 61.4 |
| 4 | Grimacing,Mannerisms,Verbigeration,Waxy flexibility | 57.0 | 52.2 | 61.7 |
| 4 | Echopraxia/echolalia,Stereotypy,Verbigeration,Waxy flexibility | 56.7 | 52.0 | 61.2 |
| 4 | Stereotypy,Mannerisms,Verbigeration,Waxy flexibility | 56.3 | 51.8 | 60.8 |
| 4 | Excitement,Grimacing,Stereotypy,Mannerisms | 56.1 | 51.6 | 60.8 |
| 4 | Grimacing,Stereotypy,Mannerisms,Verbigeration | 55.9 | 51.1 | 60.3 |
| 4 | Excitement,Echopraxia/echolalia,Stereotypy,Verbigeration | 54.7 | 50.2 | 59.2 |
| 4 | Excitement,Grimacing,Echopraxia/echolalia,Mannerisms | 54.7 | 50.2 | 59.4 |
| 4 | Excitement,Echopraxia/echolalia,Mannerisms,Waxy flexibility | 54.5 | 49.8 | 59.0 |
| 4 | Excitement,Mannerisms,Verbigeration,Waxy flexibility | 53.6 | 48.9 | 58.3 |
| 4 | Grimacing,Echopraxia/echolalia,Mannerisms,Verbigeration | 53.6 | 49.1 | 58.3 |
| 4 | Excitement,Echopraxia/echolalia,Stereotypy,Mannerisms | 53.4 | 48.7 | 57.8 |
| 4 | Excitement,Grimacing,Mannerisms,Verbigeration | 53.4 | 48.7 | 58.1 |
| 4 | Excitement,Stereotypy,Mannerisms,Verbigeration | 52.3 | 47.5 | 57.0 |
| 4 | Echopraxia/echolalia,Mannerisms,Verbigeration,Waxy flexibility | 51.6 | 46.9 | 56.1 |
| 4 | Echopraxia/echolalia,Stereotypy,Mannerisms,Verbigeration | 51.2 | 46.4 | 55.8 |
| 4 | Excitement,Echopraxia/echolalia,Mannerisms,Verbigeration | 48.9 | 44.4 | 53.6 |

Table S5: sensitivity (relative to the full BFCSI) of all five question combinations, and the 95% confidence interval derived from bootstrapping, sorted by decreasing sensitivity.

|  |  |  | 95% CI | |
| --- | --- | --- | --- | --- |
| # of Qs | Combination | Sensitivity | Lower | Upper |
| 5 | Excitement,Mutism,Staring,Posturing/catalepsy,Negativism | 98.9 | 97.8 | 99.8 |
| 5 | Excitement,Mutism,Staring,Echopraxia/echolalia,Negativism | 98.4 | 97.3 | 99.6 |
| 5 | Excitement,Mutism,Staring,Echopraxia/echolalia,Rigidity | 98.4 | 97.1 | 99.6 |
| 5 | Excitement,Mutism,Staring,Posturing/catalepsy,Rigidity | 98.4 | 97.1 | 99.6 |
| 5 | Excitement,Mutism,Staring,Rigidity,Negativism | 98.4 | 97.1 | 99.6 |
| 5 | Excitement,Mutism,Staring,Rigidity,Withdrawal | 98.4 | 97.1 | 99.6 |
| 5 | Excitement,Immobility/stupor,Staring,Posturing/catalepsy,Negativism | 98.2 | 96.9 | 99.3 |
| 5 | Excitement,Immobility/stupor,Staring,Rigidity,Negativism | 98.2 | 96.9 | 99.3 |
| 5 | Excitement,Immobility/stupor,Staring,Rigidity,Withdrawal | 98.2 | 96.9 | 99.3 |
| 5 | Excitement,Mutism,Staring,Negativism,Waxy flexibility | 98.2 | 96.9 | 99.3 |
| 5 | Excitement,Mutism,Staring,Negativism,Withdrawal | 98.2 | 96.9 | 99.3 |
| 5 | Excitement,Mutism,Staring,Posturing/catalepsy,Echopraxia/echolalia | 98.2 | 96.9 | 99.3 |
| 5 | Excitement,Mutism,Staring,Posturing/catalepsy,Withdrawal | 98.2 | 96.9 | 99.3 |
| 5 | Excitement,Immobility/stupor,Staring,Echopraxia/echolalia,Rigidity | 98.0 | 96.6 | 99.1 |
| 5 | Excitement,Immobility/stupor,Staring,Verbigeration,Rigidity | 98.0 | 96.6 | 99.1 |
| 5 | Excitement,Mutism,Staring,Rigidity,Waxy flexibility | 98.0 | 96.6 | 99.1 |
| 5 | Excitement,Mutism,Staring,Stereotypy,Negativism | 98.0 | 96.6 | 99.1 |
| 5 | Excitement,Mutism,Staring,Stereotypy,Rigidity | 98.0 | 96.6 | 99.1 |
| 5 | Excitement,Mutism,Staring,Verbigeration,Negativism | 98.0 | 96.6 | 99.1 |
| 5 | Excitement,Mutism,Staring,Verbigeration,Rigidity | 98.0 | 96.6 | 99.1 |
| 5 | Excitement,Staring,Posturing/catalepsy,Rigidity,Withdrawal | 98.0 | 96.6 | 99.1 |
| 5 | Excitement,Staring,Rigidity,Negativism,Withdrawal | 98.0 | 96.6 | 99.1 |
| 5 | Staring,Mannerisms,Rigidity,Negativism,Withdrawal | 98.0 | 96.6 | 99.1 |
| 5 | Excitement,Immobility/stupor,Mutism,Staring,Rigidity | 97.8 | 96.2 | 99.1 |
| 5 | Excitement,Immobility/stupor,Staring,Echopraxia/echolalia,Negativism | 97.8 | 96.2 | 99.1 |
| 5 | Excitement,Immobility/stupor,Staring,Mannerisms,Rigidity | 97.8 | 96.4 | 99.1 |
| 5 | Excitement,Immobility/stupor,Staring,Posturing/catalepsy,Rigidity | 97.8 | 96.2 | 99.1 |
| 5 | Excitement,Immobility/stupor,Staring,Rigidity,Waxy flexibility | 97.8 | 96.2 | 99.1 |
| 5 | Excitement,Mutism,Staring,Grimacing,Rigidity | 97.8 | 96.2 | 99.1 |
| 5 | Excitement,Mutism,Staring,Mannerisms,Rigidity | 97.8 | 96.4 | 99.1 |
| 5 | Excitement,Staring,Mannerisms,Rigidity,Withdrawal | 97.8 | 96.4 | 99.1 |
| 5 | Excitement,Staring,Posturing/catalepsy,Negativism,Withdrawal | 97.8 | 96.4 | 99.1 |
| 5 | Excitement,Mutism,Staring,Echopraxia/echolalia,Withdrawal | 97.7 | 96.2 | 99.1 |
| 5 | Excitement,Staring,Echopraxia/echolalia,Rigidity,Withdrawal | 97.7 | 96.2 | 98.9 |
| 5 | Excitement,Staring,Rigidity,Waxy flexibility,Withdrawal | 97.7 | 96.2 | 99.1 |
| 5 | Excitement,Staring,Stereotypy,Rigidity,Withdrawal | 97.7 | 96.4 | 99.1 |
| 5 | Excitement,Staring,Grimacing,Rigidity,Withdrawal | 97.6 | 96.0 | 98.9 |
| 5 | Excitement,Immobility/stupor,Echopraxia/echolalia,Rigidity,Withdrawal | 97.5 | 96.0 | 98.9 |
| 5 | Excitement,Immobility/stupor,Mutism,Staring,Negativism | 97.5 | 96.0 | 98.9 |
| 5 | Excitement,Immobility/stupor,Staring,Negativism,Waxy flexibility | 97.5 | 96.0 | 98.9 |
| 5 | Excitement,Immobility/stupor,Staring,Negativism,Withdrawal | 97.5 | 96.0 | 98.9 |
| 5 | Excitement,Immobility/stupor,Staring,Stereotypy,Rigidity | 97.5 | 96.0 | 98.9 |
| 5 | Excitement,Immobility/stupor,Staring,Verbigeration,Negativism | 97.5 | 96.0 | 98.9 |
| 5 | Excitement,Mutism,Staring,Echopraxia/echolalia,Waxy flexibility | 97.5 | 96.0 | 98.9 |
| 5 | Excitement,Mutism,Staring,Mannerisms,Negativism | 97.5 | 96.0 | 98.9 |
| 5 | Excitement,Mutism,Staring,Posturing/catalepsy,Mannerisms | 97.5 | 96.0 | 98.9 |
| 5 | Excitement,Mutism,Staring,Posturing/catalepsy,Stereotypy | 97.5 | 96.0 | 98.9 |
| 5 | Excitement,Mutism,Staring,Posturing/catalepsy,Verbigeration | 97.5 | 96.0 | 98.9 |
| 5 | Excitement,Mutism,Staring,Posturing/catalepsy,Waxy flexibility | 97.5 | 96.0 | 98.9 |
| 5 | Excitement,Mutism,Staring,Waxy flexibility,Withdrawal | 97.5 | 96.0 | 98.9 |
| 5 | Immobility/stupor,Staring,Mannerisms,Rigidity,Negativism | 97.5 | 96.0 | 98.9 |
| 5 | Mutism,Staring,Mannerisms,Negativism,Withdrawal | 97.5 | 96.0 | 98.9 |
| 5 | Mutism,Staring,Mannerisms,Rigidity,Negativism | 97.5 | 96.0 | 98.9 |
| 5 | Excitement,Immobility/stupor,Mutism,Rigidity,Negativism | 97.3 | 95.7 | 98.7 |
| 5 | Excitement,Immobility/stupor,Mutism,Staring,Posturing/catalepsy | 97.3 | 95.7 | 98.7 |
| 5 | Excitement,Immobility/stupor,Staring,Grimacing,Rigidity | 97.3 | 95.7 | 98.7 |
| 5 | Excitement,Immobility/stupor,Staring,Stereotypy,Negativism | 97.3 | 95.7 | 98.7 |
| 5 | Excitement,Mutism,Staring,Echopraxia/echolalia,Stereotypy | 97.3 | 95.7 | 98.7 |
| 5 | Excitement,Mutism,Staring,Grimacing,Negativism | 97.3 | 95.7 | 98.7 |
| 5 | Excitement,Mutism,Staring,Posturing/catalepsy,Grimacing | 97.3 | 95.7 | 98.7 |
| 5 | Excitement,Mutism,Staring,Stereotypy,Waxy flexibility | 97.3 | 95.7 | 98.7 |
| 5 | Excitement,Mutism,Staring,Stereotypy,Withdrawal | 97.3 | 95.7 | 98.7 |
| 5 | Excitement,Staring,Echopraxia/echolalia,Negativism,Withdrawal | 97.3 | 95.7 | 98.7 |
| 5 | Excitement,Staring,Verbigeration,Rigidity,Withdrawal | 97.3 | 95.7 | 98.7 |
| 5 | Staring,Posturing/catalepsy,Mannerisms,Negativism,Withdrawal | 97.3 | 95.7 | 98.7 |
| 5 | Excitement,Immobility/stupor,Mutism,Echopraxia/echolalia,Rigidity | 97.1 | 95.5 | 98.4 |
| 5 | Excitement,Immobility/stupor,Mutism,Rigidity,Withdrawal | 97.1 | 95.5 | 98.7 |
| 5 | Excitement,Immobility/stupor,Mutism,Staring,Echopraxia/echolalia | 97.1 | 95.5 | 98.4 |
| 5 | Excitement,Immobility/stupor,Mutism,Staring,Waxy flexibility | 97.1 | 95.5 | 98.7 |
| 5 | Excitement,Immobility/stupor,Posturing/catalepsy,Rigidity,Negativism | 97.1 | 95.5 | 98.4 |
| 5 | Excitement,Immobility/stupor,Posturing/catalepsy,Rigidity,Withdrawal | 97.1 | 95.5 | 98.7 |
| 5 | Excitement,Immobility/stupor,Rigidity,Negativism,Withdrawal | 97.1 | 95.5 | 98.7 |
| 5 | Excitement,Immobility/stupor,Staring,Echopraxia/echolalia,Withdrawal | 97.1 | 95.5 | 98.4 |
| 5 | Excitement,Immobility/stupor,Staring,Mannerisms,Negativism | 97.1 | 95.5 | 98.4 |
| 5 | Excitement,Immobility/stupor,Staring,Posturing/catalepsy,Echopraxia/echolalia | 97.1 | 95.5 | 98.4 |
| 5 | Excitement,Immobility/stupor,Staring,Posturing/catalepsy,Withdrawal | 97.1 | 95.5 | 98.4 |
| 5 | Excitement,Mutism,Staring,Grimacing,Echopraxia/echolalia | 97.1 | 95.5 | 98.4 |
| 5 | Excitement,Mutism,Staring,Grimacing,Withdrawal | 97.1 | 95.5 | 98.4 |
| 5 | Excitement,Staring,Negativism,Waxy flexibility,Withdrawal | 97.1 | 95.5 | 98.4 |
| 5 | Excitement,Staring,Posturing/catalepsy,Rigidity,Negativism | 97.1 | 95.5 | 98.4 |
| 5 | Excitement,Staring,Stereotypy,Negativism,Withdrawal | 97.1 | 95.5 | 98.7 |
| 5 | Mutism,Staring,Echopraxia/echolalia,Mannerisms,Negativism | 97.1 | 95.5 | 98.7 |
| 5 | Mutism,Staring,Mannerisms,Rigidity,Withdrawal | 97.1 | 95.5 | 98.4 |
| 5 | Mutism,Staring,Posturing/catalepsy,Mannerisms,Negativism | 97.1 | 95.5 | 98.4 |
| 5 | Mutism,Staring,Posturing/catalepsy,Negativism,Withdrawal | 97.1 | 95.5 | 98.4 |
| 5 | Mutism,Staring,Stereotypy,Mannerisms,Negativism | 97.1 | 95.5 | 98.4 |
| 5 | Staring,Stereotypy,Mannerisms,Negativism,Withdrawal | 97.1 | 95.5 | 98.7 |
| 5 | Staring,Stereotypy,Rigidity,Negativism,Withdrawal | 97.1 | 95.5 | 98.7 |
| 5 | Excitement,Immobility/stupor,Echopraxia/echolalia,Rigidity,Negativism | 96.9 | 95.3 | 98.4 |
| 5 | Excitement,Immobility/stupor,Mutism,Posturing/catalepsy,Rigidity | 96.9 | 95.1 | 98.4 |
| 5 | Excitement,Immobility/stupor,Mutism,Verbigeration,Rigidity | 96.9 | 95.1 | 98.4 |
| 5 | Excitement,Immobility/stupor,Posturing/catalepsy,Negativism,Withdrawal | 96.9 | 95.1 | 98.4 |
| 5 | Excitement,Immobility/stupor,Staring,Posturing/catalepsy,Mannerisms | 96.9 | 95.3 | 98.4 |
| 5 | Excitement,Immobility/stupor,Staring,Posturing/catalepsy,Verbigeration | 96.9 | 95.3 | 98.4 |
| 5 | Excitement,Mutism,Staring,Grimacing,Waxy flexibility | 96.9 | 95.1 | 98.4 |
| 5 | Excitement,Mutism,Staring,Mannerisms,Withdrawal | 96.9 | 95.1 | 98.4 |
| 5 | Excitement,Mutism,Staring,Verbigeration,Waxy flexibility | 96.9 | 95.1 | 98.4 |
| 5 | Excitement,Mutism,Staring,Verbigeration,Withdrawal | 96.9 | 95.1 | 98.4 |
| 5 | Excitement,Staring,Echopraxia/echolalia,Rigidity,Negativism | 96.9 | 95.1 | 98.4 |
| 5 | Excitement,Staring,Mannerisms,Negativism,Withdrawal | 96.9 | 95.1 | 98.4 |
| 5 | Excitement,Staring,Posturing/catalepsy,Mannerisms,Withdrawal | 96.9 | 95.1 | 98.4 |
| 5 | Excitement,Staring,Rigidity,Negativism,Waxy flexibility | 96.9 | 95.1 | 98.4 |
| 5 | Excitement,Staring,Stereotypy,Rigidity,Negativism | 96.9 | 95.1 | 98.4 |
| 5 | Excitement,Staring,Verbigeration,Negativism,Withdrawal | 96.9 | 95.1 | 98.4 |
| 5 | Immobility/stupor,Staring,Mannerisms,Negativism,Withdrawal | 96.9 | 95.1 | 98.4 |
| 5 | Mutism,Staring,Posturing/catalepsy,Mannerisms,Withdrawal | 96.9 | 95.1 | 98.4 |
| 5 | Staring,Echopraxia/echolalia,Mannerisms,Negativism,Withdrawal | 96.9 | 95.1 | 98.4 |
| 5 | Staring,Echopraxia/echolalia,Rigidity,Negativism,Withdrawal | 96.9 | 95.1 | 98.4 |
| 5 | Staring,Stereotypy,Mannerisms,Rigidity,Withdrawal | 96.9 | 95.1 | 98.4 |
| 5 | Staring,Verbigeration,Rigidity,Negativism,Withdrawal | 96.9 | 95.1 | 98.4 |
| 5 | Excitement,Staring,Posturing/catalepsy,Echopraxia/echolalia,Withdrawal | 96.8 | 95.1 | 98.4 |
| 5 | Mutism,Staring,Rigidity,Negativism,Withdrawal | 96.8 | 95.1 | 98.4 |
| 5 | Immobility/stupor,Staring,Stereotypy,Mannerisms,Negativism | 96.7 | 95.1 | 98.2 |
| 5 | Staring,Posturing/catalepsy,Mannerisms,Rigidity,Withdrawal | 96.7 | 94.8 | 98.2 |
| 5 | Excitement,Immobility/stupor,Mannerisms,Rigidity,Withdrawal | 96.6 | 94.8 | 98.2 |
| 5 | Excitement,Immobility/stupor,Mutism,Posturing/catalepsy,Negativism | 96.6 | 94.8 | 98.2 |
| 5 | Excitement,Immobility/stupor,Mutism,Staring,Withdrawal | 96.6 | 94.8 | 98.2 |
| 5 | Excitement,Immobility/stupor,Staring,Echopraxia/echolalia,Waxy flexibility | 96.6 | 94.8 | 98.2 |
| 5 | Excitement,Immobility/stupor,Staring,Grimacing,Negativism | 96.6 | 94.8 | 98.2 |
| 5 | Excitement,Immobility/stupor,Staring,Verbigeration,Waxy flexibility | 96.6 | 94.8 | 98.2 |
| 5 | Excitement,Immobility/stupor,Staring,Waxy flexibility,Withdrawal | 96.6 | 94.8 | 98.2 |
| 5 | Excitement,Immobility/stupor,Stereotypy,Rigidity,Negativism | 96.6 | 94.8 | 98.2 |
| 5 | Excitement,Immobility/stupor,Stereotypy,Rigidity,Withdrawal | 96.6 | 94.8 | 98.2 |
| 5 | Excitement,Immobility/stupor,Verbigeration,Rigidity,Negativism | 96.6 | 94.8 | 98.2 |
| 5 | Excitement,Immobility/stupor,Verbigeration,Rigidity,Withdrawal | 96.6 | 94.8 | 98.2 |
| 5 | Excitement,Mutism,Staring,Echopraxia/echolalia,Verbigeration | 96.6 | 94.8 | 98.2 |
| 5 | Excitement,Mutism,Staring,Grimacing,Stereotypy | 96.6 | 94.8 | 98.2 |
| 5 | Excitement,Mutism,Staring,Mannerisms,Waxy flexibility | 96.6 | 94.8 | 98.2 |
| 5 | Excitement,Mutism,Staring,Stereotypy,Verbigeration | 96.6 | 94.8 | 98.2 |
| 5 | Excitement,Staring,Posturing/catalepsy,Echopraxia/echolalia,Negativism | 96.6 | 94.8 | 98.2 |
| 5 | Excitement,Staring,Posturing/catalepsy,Grimacing,Withdrawal | 96.6 | 94.8 | 98.2 |
| 5 | Immobility/stupor,Staring,Rigidity,Negativism,Withdrawal | 96.6 | 94.8 | 98.2 |
| 5 | Mutism,Staring,Echopraxia/echolalia,Rigidity,Negativism | 96.6 | 94.8 | 98.2 |
| 5 | Mutism,Staring,Posturing/catalepsy,Mannerisms,Rigidity | 96.6 | 94.8 | 98.2 |
| 5 | Mutism,Staring,Posturing/catalepsy,Stereotypy,Negativism | 96.6 | 94.8 | 98.2 |
| 5 | Mutism,Staring,Stereotypy,Negativism,Withdrawal | 96.6 | 94.8 | 98.2 |
| 5 | Mutism,Staring,Stereotypy,Rigidity,Negativism | 96.6 | 94.8 | 98.2 |
| 5 | Mutism,Staring,Verbigeration,Rigidity,Negativism | 96.6 | 94.8 | 98.2 |
| 5 | Staring,Grimacing,Mannerisms,Rigidity,Withdrawal | 96.6 | 94.8 | 98.2 |
| 5 | Staring,Mannerisms,Negativism,Waxy flexibility,Withdrawal | 96.6 | 94.8 | 98.2 |
| 5 | Staring,Posturing/catalepsy,Rigidity,Negativism,Withdrawal | 96.6 | 94.8 | 98.2 |
| 5 | Staring,Stereotypy,Mannerisms,Rigidity,Negativism | 96.6 | 94.8 | 98.2 |
| 5 | Excitement,Immobility/stupor,Grimacing,Rigidity,Negativism | 96.4 | 94.6 | 98.0 |
| 5 | Excitement,Immobility/stupor,Grimacing,Rigidity,Withdrawal | 96.4 | 94.6 | 98.0 |
| 5 | Excitement,Immobility/stupor,Mutism,Grimacing,Rigidity | 96.4 | 94.6 | 98.0 |
| 5 | Excitement,Immobility/stupor,Mutism,Rigidity,Waxy flexibility | 96.4 | 94.6 | 98.0 |
| 5 | Excitement,Immobility/stupor,Mutism,Staring,Stereotypy | 96.4 | 94.6 | 98.0 |
| 5 | Excitement,Immobility/stupor,Mutism,Staring,Verbigeration | 96.4 | 94.6 | 98.0 |
| 5 | Excitement,Immobility/stupor,Mutism,Stereotypy,Rigidity | 96.4 | 94.6 | 98.0 |
| 5 | Excitement,Immobility/stupor,Posturing/catalepsy,Echopraxia/echolalia,Rigidity | 96.4 | 94.6 | 98.0 |
| 5 | Excitement,Immobility/stupor,Rigidity,Waxy flexibility,Withdrawal | 96.4 | 94.6 | 98.0 |
| 5 | Excitement,Immobility/stupor,Staring,Mannerisms,Withdrawal | 96.4 | 94.6 | 98.0 |
| 5 | Excitement,Immobility/stupor,Staring,Posturing/catalepsy,Grimacing | 96.4 | 94.6 | 98.0 |
| 5 | Excitement,Immobility/stupor,Staring,Posturing/catalepsy,Waxy flexibility | 96.4 | 94.6 | 98.0 |
| 5 | Excitement,Mutism,Staring,Echopraxia/echolalia,Mannerisms | 96.4 | 94.6 | 98.0 |
| 5 | Excitement,Mutism,Staring,Grimacing,Verbigeration | 96.4 | 94.6 | 98.0 |
| 5 | Excitement,Staring,Echopraxia/echolalia,Waxy flexibility,Withdrawal | 96.4 | 94.6 | 98.0 |
| 5 | Excitement,Staring,Grimacing,Negativism,Withdrawal | 96.4 | 94.6 | 98.0 |
| 5 | Excitement,Staring,Mannerisms,Rigidity,Negativism | 96.4 | 94.6 | 98.0 |
| 5 | Excitement,Staring,Posturing/catalepsy,Echopraxia/echolalia,Rigidity | 96.4 | 94.6 | 98.0 |
| 5 | Excitement,Staring,Posturing/catalepsy,Grimacing,Rigidity | 96.4 | 94.6 | 98.0 |
| 5 | Excitement,Staring,Posturing/catalepsy,Mannerisms,Negativism | 96.4 | 94.6 | 98.0 |
| 5 | Excitement,Staring,Posturing/catalepsy,Mannerisms,Rigidity | 96.4 | 94.6 | 98.0 |
| 5 | Excitement,Staring,Posturing/catalepsy,Verbigeration,Negativism | 96.4 | 94.6 | 98.0 |
| 5 | Excitement,Staring,Posturing/catalepsy,Waxy flexibility,Withdrawal | 96.4 | 94.6 | 98.0 |
| 5 | Excitement,Staring,Verbigeration,Rigidity,Negativism | 96.4 | 94.6 | 98.0 |
| 5 | Immobility/stupor,Mannerisms,Rigidity,Negativism,Withdrawal | 96.4 | 94.6 | 98.0 |
| 5 | Immobility/stupor,Staring,Echopraxia/echolalia,Mannerisms,Negativism | 96.4 | 94.6 | 98.0 |
| 5 | Immobility/stupor,Staring,Mannerisms,Rigidity,Withdrawal | 96.4 | 94.6 | 98.0 |
| 5 | Immobility/stupor,Staring,Posturing/catalepsy,Mannerisms,Negativism | 96.4 | 94.6 | 98.0 |
| 5 | Immobility/stupor,Staring,Stereotypy,Rigidity,Negativism | 96.4 | 94.6 | 98.0 |
| 5 | Mutism,Staring,Echopraxia/echolalia,Mannerisms,Rigidity | 96.4 | 94.6 | 98.0 |
| 5 | Mutism,Staring,Echopraxia/echolalia,Negativism,Withdrawal | 96.4 | 94.6 | 98.0 |
| 5 | Mutism,Staring,Grimacing,Mannerisms,Rigidity | 96.4 | 94.6 | 98.0 |
| 5 | Mutism,Staring,Mannerisms,Negativism,Waxy flexibility | 96.4 | 94.6 | 98.0 |
| 5 | Mutism,Staring,Mannerisms,Verbigeration,Negativism | 96.4 | 94.6 | 98.0 |
| 5 | Mutism,Staring,Mannerisms,Verbigeration,Rigidity | 96.4 | 94.6 | 98.0 |
| 5 | Mutism,Staring,Posturing/catalepsy,Echopraxia/echolalia,Negativism | 96.4 | 94.6 | 98.0 |
| 5 | Mutism,Staring,Stereotypy,Mannerisms,Rigidity | 96.4 | 94.6 | 98.0 |
| 5 | Mutism,Staring,Stereotypy,Rigidity,Withdrawal | 96.4 | 94.6 | 98.0 |
| 5 | Mutism,Staring,Stereotypy,Verbigeration,Negativism | 96.4 | 94.6 | 98.0 |
| 5 | Mutism,Staring,Stereotypy,Verbigeration,Rigidity | 96.4 | 94.6 | 98.0 |
| 5 | Mutism,Staring,Verbigeration,Negativism,Withdrawal | 96.4 | 94.6 | 98.0 |
| 5 | Staring,Echopraxia/echolalia,Mannerisms,Rigidity,Negativism | 96.4 | 94.6 | 98.0 |
| 5 | Staring,Grimacing,Mannerisms,Negativism,Withdrawal | 96.4 | 94.6 | 98.0 |
| 5 | Staring,Mannerisms,Verbigeration,Negativism,Withdrawal | 96.4 | 94.6 | 98.0 |
| 5 | Staring,Posturing/catalepsy,Mannerisms,Rigidity,Negativism | 96.4 | 94.6 | 98.0 |
| 5 | Excitement,Immobility/stupor,Grimacing,Echopraxia/echolalia,Rigidity | 96.2 | 94.4 | 97.8 |
| 5 | Excitement,Immobility/stupor,Mannerisms,Rigidity,Negativism | 96.2 | 94.4 | 98.0 |
| 5 | Excitement,Immobility/stupor,Mutism,Mannerisms,Rigidity | 96.2 | 94.4 | 98.0 |
| 5 | Excitement,Immobility/stupor,Mutism,Staring,Mannerisms | 96.2 | 94.2 | 97.8 |
| 5 | Excitement,Immobility/stupor,Posturing/catalepsy,Grimacing,Rigidity | 96.2 | 94.4 | 97.8 |
| 5 | Excitement,Immobility/stupor,Posturing/catalepsy,Verbigeration,Negativism | 96.2 | 94.4 | 97.8 |
| 5 | Excitement,Immobility/stupor,Posturing/catalepsy,Verbigeration,Rigidity | 96.2 | 94.4 | 98.0 |
| 5 | Excitement,Immobility/stupor,Rigidity,Negativism,Waxy flexibility | 96.2 | 94.4 | 97.8 |
| 5 | Excitement,Immobility/stupor,Staring,Echopraxia/echolalia,Stereotypy | 96.2 | 94.4 | 97.8 |
| 5 | Excitement,Immobility/stupor,Staring,Echopraxia/echolalia,Verbigeration | 96.2 | 94.4 | 98.0 |
| 5 | Excitement,Immobility/stupor,Staring,Grimacing,Echopraxia/echolalia | 96.2 | 94.4 | 98.0 |
| 5 | Excitement,Immobility/stupor,Staring,Mannerisms,Verbigeration | 96.2 | 94.4 | 97.8 |
| 5 | Excitement,Immobility/stupor,Staring,Mannerisms,Waxy flexibility | 96.2 | 94.4 | 97.8 |
| 5 | Excitement,Immobility/stupor,Staring,Posturing/catalepsy,Stereotypy | 96.2 | 94.4 | 98.0 |
| 5 | Excitement,Immobility/stupor,Staring,Stereotypy,Waxy flexibility | 96.2 | 94.4 | 97.8 |
| 5 | Excitement,Immobility/stupor,Staring,Stereotypy,Withdrawal | 96.2 | 94.4 | 97.8 |
| 5 | Excitement,Immobility/stupor,Staring,Verbigeration,Withdrawal | 96.2 | 94.4 | 98.0 |
| 5 | Excitement,Mutism,Staring,Stereotypy,Mannerisms | 96.2 | 94.4 | 97.8 |
| 5 | Excitement,Staring,Echopraxia/echolalia,Negativism,Waxy flexibility | 96.2 | 94.4 | 98.0 |
| 5 | Excitement,Staring,Grimacing,Echopraxia/echolalia,Rigidity | 96.2 | 94.2 | 98.0 |
| 5 | Excitement,Staring,Grimacing,Echopraxia/echolalia,Withdrawal | 96.2 | 94.4 | 97.8 |
| 5 | Excitement,Staring,Grimacing,Rigidity,Waxy flexibility | 96.2 | 94.4 | 97.8 |
| 5 | Excitement,Staring,Grimacing,Stereotypy,Rigidity | 96.2 | 94.4 | 98.0 |
| 5 | Excitement,Staring,Mannerisms,Waxy flexibility,Withdrawal | 96.2 | 94.4 | 97.8 |
| 5 | Excitement,Staring,Posturing/catalepsy,Negativism,Waxy flexibility | 96.2 | 94.4 | 97.8 |
| 5 | Excitement,Staring,Posturing/catalepsy,Stereotypy,Negativism | 96.2 | 94.4 | 98.0 |
| 5 | Excitement,Staring,Posturing/catalepsy,Verbigeration,Rigidity | 96.2 | 94.4 | 97.8 |
| 5 | Excitement,Staring,Posturing/catalepsy,Verbigeration,Withdrawal | 96.2 | 94.4 | 97.8 |
| 5 | Immobility/stupor,Staring,Posturing/catalepsy,Negativism,Withdrawal | 96.2 | 94.4 | 97.8 |
| 5 | Mutism,Staring,Echopraxia/echolalia,Rigidity,Withdrawal | 96.2 | 94.4 | 98.0 |
| 5 | Mutism,Staring,Echopraxia/echolalia,Stereotypy,Negativism | 96.2 | 94.4 | 97.8 |
| 5 | Mutism,Staring,Grimacing,Mannerisms,Negativism | 96.2 | 94.4 | 97.8 |
| 5 | Mutism,Staring,Grimacing,Mannerisms,Withdrawal | 96.2 | 94.4 | 98.0 |
| 5 | Mutism,Staring,Grimacing,Verbigeration,Rigidity | 96.2 | 94.4 | 98.0 |
| 5 | Mutism,Staring,Posturing/catalepsy,Verbigeration,Negativism | 96.2 | 94.4 | 97.8 |
| 5 | Mutism,Staring,Stereotypy,Mannerisms,Withdrawal | 96.2 | 94.4 | 97.8 |
| 5 | Mutism,Staring,Verbigeration,Rigidity,Withdrawal | 96.2 | 94.4 | 98.0 |
| 5 | Staring,Mannerisms,Rigidity,Negativism,Waxy flexibility | 96.2 | 94.4 | 97.8 |
| 5 | Staring,Posturing/catalepsy,Echopraxia/echolalia,Negativism,Withdrawal | 96.2 | 94.4 | 97.8 |
| 5 | Staring,Posturing/catalepsy,Stereotypy,Negativism,Withdrawal | 96.2 | 94.4 | 98.0 |
| 5 | Staring,Rigidity,Negativism,Waxy flexibility,Withdrawal | 96.2 | 94.4 | 97.8 |
| 5 | Staring,Stereotypy,Verbigeration,Negativism,Withdrawal | 96.2 | 94.4 | 97.8 |
| 5 | Excitement,Immobility/stupor,Echopraxia/echolalia,Negativism,Withdrawal | 96.0 | 94.2 | 97.8 |
| 5 | Excitement,Immobility/stupor,Echopraxia/echolalia,Verbigeration,Rigidity | 96.0 | 93.9 | 97.8 |
| 5 | Excitement,Immobility/stupor,Mutism,Posturing/catalepsy,Withdrawal | 96.0 | 93.9 | 97.8 |
| 5 | Excitement,Immobility/stupor,Mutism,Staring,Grimacing | 96.0 | 93.9 | 97.8 |
| 5 | Excitement,Immobility/stupor,Posturing/catalepsy,Echopraxia/echolalia,Negativism | 96.0 | 94.2 | 97.8 |
| 5 | Excitement,Immobility/stupor,Posturing/catalepsy,Grimacing,Negativism | 96.0 | 93.9 | 97.8 |
| 5 | Excitement,Immobility/stupor,Staring,Grimacing,Waxy flexibility | 96.0 | 93.9 | 97.8 |
| 5 | Excitement,Immobility/stupor,Staring,Stereotypy,Verbigeration | 96.0 | 93.9 | 97.8 |
| 5 | Excitement,Mutism,Staring,Grimacing,Mannerisms | 96.0 | 93.9 | 97.8 |
| 5 | Excitement,Mutism,Staring,Mannerisms,Verbigeration | 96.0 | 93.9 | 97.8 |
| 5 | Excitement,Staring,Grimacing,Mannerisms,Rigidity | 96.0 | 93.9 | 97.8 |
| 5 | Excitement,Staring,Grimacing,Rigidity,Negativism | 96.0 | 94.2 | 97.8 |
| 5 | Excitement,Staring,Grimacing,Waxy flexibility,Withdrawal | 96.0 | 94.2 | 97.8 |
| 5 | Excitement,Staring,Posturing/catalepsy,Stereotypy,Rigidity | 96.0 | 93.9 | 97.8 |
| 5 | Excitement,Staring,Posturing/catalepsy,Stereotypy,Withdrawal | 96.0 | 94.2 | 97.8 |
| 5 | Excitement,Staring,Stereotypy,Rigidity,Waxy flexibility | 96.0 | 94.2 | 97.8 |
| 5 | Excitement,Staring,Stereotypy,Waxy flexibility,Withdrawal | 96.0 | 93.9 | 97.8 |
| 5 | Immobility/stupor,Mutism,Mannerisms,Rigidity,Negativism | 96.0 | 94.2 | 97.8 |
| 5 | Immobility/stupor,Posturing/catalepsy,Mannerisms,Rigidity,Negativism | 96.0 | 93.9 | 97.8 |
| 5 | Immobility/stupor,Staring,Echopraxia/echolalia,Rigidity,Negativism | 96.0 | 93.9 | 97.8 |
| 5 | Immobility/stupor,Staring,Mannerisms,Verbigeration,Negativism | 96.0 | 94.2 | 97.8 |
| 5 | Immobility/stupor,Staring,Mannerisms,Verbigeration,Rigidity | 96.0 | 93.9 | 97.8 |
| 5 | Immobility/stupor,Staring,Posturing/catalepsy,Stereotypy,Negativism | 96.0 | 94.2 | 97.8 |
| 5 | Immobility/stupor,Staring,Stereotypy,Negativism,Withdrawal | 96.0 | 93.9 | 97.8 |
| 5 | Immobility/stupor,Staring,Stereotypy,Verbigeration,Negativism | 96.0 | 93.9 | 97.8 |
| 5 | Immobility/stupor,Staring,Verbigeration,Rigidity,Negativism | 96.0 | 93.9 | 97.5 |
| 5 | Mutism,Staring,Echopraxia/echolalia,Mannerisms,Withdrawal | 96.0 | 93.9 | 97.8 |
| 5 | Mutism,Staring,Echopraxia/echolalia,Verbigeration,Negativism | 96.0 | 94.2 | 97.8 |
| 5 | Mutism,Staring,Posturing/catalepsy,Echopraxia/echolalia,Rigidity | 96.0 | 94.2 | 97.8 |
| 5 | Mutism,Staring,Posturing/catalepsy,Echopraxia/echolalia,Withdrawal | 96.0 | 94.2 | 97.8 |
| 5 | Mutism,Staring,Posturing/catalepsy,Rigidity,Withdrawal | 96.0 | 94.2 | 97.8 |
| 5 | Mutism,Staring,Posturing/catalepsy,Verbigeration,Rigidity | 96.0 | 94.2 | 97.8 |
| 5 | Staring,Echopraxia/echolalia,Mannerisms,Rigidity,Withdrawal | 96.0 | 94.2 | 97.8 |
| 5 | Staring,Echopraxia/echolalia,Stereotypy,Negativism,Withdrawal | 96.0 | 93.9 | 97.8 |
| 5 | Staring,Grimacing,Rigidity,Negativism,Withdrawal | 96.0 | 94.2 | 97.8 |
| 5 | Staring,Grimacing,Stereotypy,Rigidity,Withdrawal | 96.0 | 93.9 | 97.8 |
| 5 | Staring,Mannerisms,Rigidity,Waxy flexibility,Withdrawal | 96.0 | 94.2 | 97.8 |
| 5 | Staring,Mannerisms,Verbigeration,Rigidity,Withdrawal | 96.0 | 93.9 | 97.8 |
| 5 | Staring,Posturing/catalepsy,Verbigeration,Negativism,Withdrawal | 96.0 | 94.2 | 97.8 |
| 5 | Staring,Stereotypy,Verbigeration,Rigidity,Negativism | 96.0 | 94.2 | 97.8 |
| 5 | Staring,Stereotypy,Verbigeration,Rigidity,Withdrawal | 96.0 | 94.2 | 97.8 |
| 5 | Excitement,Immobility/stupor,Staring,Echopraxia/echolalia,Mannerisms | 95.9 | 93.9 | 97.8 |
| 5 | Excitement,Immobility/stupor,Staring,Grimacing,Withdrawal | 95.9 | 93.9 | 97.8 |
| 5 | Excitement,Staring,Echopraxia/echolalia,Stereotypy,Withdrawal | 95.9 | 93.9 | 97.8 |
| 5 | Excitement,Staring,Posturing/catalepsy,Grimacing,Negativism | 95.9 | 93.9 | 97.8 |
| 5 | Excitement,Staring,Verbigeration,Negativism,Waxy flexibility | 95.8 | 93.7 | 97.5 |
| 5 | Excitement,Staring,Verbigeration,Waxy flexibility,Withdrawal | 95.8 | 93.7 | 97.5 |
| 5 | Immobility/stupor,Mutism,Staring,Mannerisms,Rigidity | 95.8 | 93.7 | 97.5 |
| 5 | Immobility/stupor,Staring,Mannerisms,Negativism,Waxy flexibility | 95.8 | 93.9 | 97.5 |
| 5 | Immobility/stupor,Stereotypy,Mannerisms,Rigidity,Negativism | 95.8 | 93.7 | 97.5 |
| 5 | Staring,Grimacing,Stereotypy,Negativism,Withdrawal | 95.8 | 93.7 | 97.5 |
| 5 | Staring,Posturing/catalepsy,Grimacing,Negativism,Withdrawal | 95.8 | 93.7 | 97.5 |
| 5 | Excitement,Immobility/stupor,Echopraxia/echolalia,Rigidity,Waxy flexibility | 95.7 | 93.7 | 97.5 |
| 5 | Excitement,Immobility/stupor,Echopraxia/echolalia,Stereotypy,Rigidity | 95.7 | 93.7 | 97.5 |
| 5 | Excitement,Immobility/stupor,Mutism,Posturing/catalepsy,Echopraxia/echolalia | 95.7 | 93.7 | 97.5 |
| 5 | Excitement,Immobility/stupor,Posturing/catalepsy,Echopraxia/echolalia,Withdrawal | 95.7 | 93.7 | 97.5 |
| 5 | Excitement,Immobility/stupor,Posturing/catalepsy,Mannerisms,Rigidity | 95.7 | 93.7 | 97.5 |
| 5 | Excitement,Immobility/stupor,Staring,Grimacing,Verbigeration | 95.7 | 93.7 | 97.5 |
| 5 | Excitement,Immobility/stupor,Stereotypy,Negativism,Withdrawal | 95.7 | 93.7 | 97.5 |
| 5 | Excitement,Staring,Echopraxia/echolalia,Mannerisms,Withdrawal | 95.7 | 93.7 | 97.5 |
| 5 | Excitement,Staring,Echopraxia/echolalia,Rigidity,Waxy flexibility | 95.7 | 93.7 | 97.5 |
| 5 | Excitement,Staring,Echopraxia/echolalia,Stereotypy,Negativism | 95.7 | 93.7 | 97.5 |
| 5 | Excitement,Staring,Echopraxia/echolalia,Stereotypy,Rigidity | 95.7 | 93.7 | 97.5 |
| 5 | Excitement,Staring,Grimacing,Mannerisms,Withdrawal | 95.7 | 93.7 | 97.5 |
| 5 | Excitement,Staring,Grimacing,Stereotypy,Withdrawal | 95.7 | 93.7 | 97.5 |
| 5 | Excitement,Staring,Grimacing,Verbigeration,Rigidity | 95.7 | 93.7 | 97.5 |
| 5 | Excitement,Staring,Mannerisms,Negativism,Waxy flexibility | 95.7 | 93.7 | 97.5 |
| 5 | Excitement,Staring,Mannerisms,Rigidity,Waxy flexibility | 95.7 | 93.7 | 97.5 |
| 5 | Excitement,Staring,Stereotypy,Negativism,Waxy flexibility | 95.7 | 93.7 | 97.5 |
| 5 | Excitement,Staring,Stereotypy,Verbigeration,Rigidity | 95.7 | 93.7 | 97.5 |
| 5 | Excitement,Staring,Verbigeration,Rigidity,Waxy flexibility | 95.7 | 93.7 | 97.5 |
| 5 | Immobility/stupor,Mutism,Staring,Mannerisms,Negativism | 95.7 | 93.7 | 97.5 |
| 5 | Immobility/stupor,Staring,Echopraxia/echolalia,Negativism,Withdrawal | 95.7 | 93.7 | 97.5 |
| 5 | Immobility/stupor,Staring,Grimacing,Mannerisms,Rigidity | 95.7 | 93.7 | 97.5 |
| 5 | Immobility/stupor,Staring,Stereotypy,Mannerisms,Rigidity | 95.7 | 93.7 | 97.5 |
| 5 | Immobility/stupor,Staring,Stereotypy,Verbigeration,Rigidity | 95.7 | 93.7 | 97.5 |
| 5 | Immobility/stupor,Staring,Verbigeration,Negativism,Withdrawal | 95.7 | 93.7 | 97.5 |
| 5 | Immobility/stupor,Stereotypy,Rigidity,Negativism,Withdrawal | 95.7 | 93.7 | 97.5 |
| 5 | Mutism,Staring,Echopraxia/echolalia,Stereotypy,Rigidity | 95.7 | 93.7 | 97.5 |
| 5 | Mutism,Staring,Echopraxia/echolalia,Verbigeration,Rigidity | 95.7 | 93.7 | 97.5 |
| 5 | Mutism,Staring,Grimacing,Negativism,Withdrawal | 95.7 | 93.7 | 97.5 |
| 5 | Mutism,Staring,Grimacing,Rigidity,Withdrawal | 95.7 | 93.7 | 97.5 |
| 5 | Mutism,Staring,Grimacing,Stereotypy,Negativism | 95.7 | 93.7 | 97.5 |
| 5 | Mutism,Staring,Grimacing,Stereotypy,Rigidity | 95.7 | 93.7 | 97.5 |
| 5 | Mutism,Staring,Grimacing,Verbigeration,Negativism | 95.7 | 93.7 | 97.5 |
| 5 | Mutism,Staring,Mannerisms,Rigidity,Waxy flexibility | 95.7 | 93.7 | 97.5 |
| 5 | Mutism,Staring,Mannerisms,Waxy flexibility,Withdrawal | 95.7 | 93.7 | 97.5 |
| 5 | Mutism,Staring,Negativism,Waxy flexibility,Withdrawal | 95.7 | 93.7 | 97.5 |
| 5 | Mutism,Staring,Posturing/catalepsy,Echopraxia/echolalia,Mannerisms | 95.7 | 93.7 | 97.5 |
| 5 | Mutism,Staring,Posturing/catalepsy,Rigidity,Negativism | 95.7 | 93.7 | 97.5 |
| 5 | Mutism,Staring,Posturing/catalepsy,Stereotypy,Rigidity | 95.7 | 93.7 | 97.5 |
| 5 | Mutism,Staring,Posturing/catalepsy,Stereotypy,Withdrawal | 95.7 | 93.7 | 97.5 |
| 5 | Mutism,Staring,Stereotypy,Negativism,Waxy flexibility | 95.7 | 93.7 | 97.5 |
| 5 | Staring,Echopraxia/echolalia,Stereotypy,Rigidity,Withdrawal | 95.7 | 93.7 | 97.5 |
| 5 | Staring,Grimacing,Stereotypy,Mannerisms,Withdrawal | 95.7 | 93.7 | 97.5 |
| 5 | Staring,Mannerisms,Verbigeration,Rigidity,Negativism | 95.7 | 93.7 | 97.5 |
| 5 | Staring,Posturing/catalepsy,Grimacing,Mannerisms,Withdrawal | 95.7 | 93.7 | 97.5 |
| 5 | Excitement,Immobility/stupor,Grimacing,Verbigeration,Rigidity | 95.5 | 93.5 | 97.3 |
| 5 | Excitement,Immobility/stupor,Mutism,Negativism,Waxy flexibility | 95.5 | 93.5 | 97.3 |
| 5 | Excitement,Immobility/stupor,Mutism,Posturing/catalepsy,Grimacing | 95.5 | 93.5 | 97.3 |
| 5 | Excitement,Immobility/stupor,Posturing/catalepsy,Mannerisms,Negativism | 95.5 | 93.5 | 97.3 |
| 5 | Excitement,Immobility/stupor,Posturing/catalepsy,Negativism,Waxy flexibility | 95.5 | 93.5 | 97.3 |
| 5 | Excitement,Immobility/stupor,Posturing/catalepsy,Stereotypy,Negativism | 95.5 | 93.5 | 97.3 |
| 5 | Excitement,Immobility/stupor,Posturing/catalepsy,Stereotypy,Rigidity | 95.5 | 93.5 | 97.3 |
| 5 | Excitement,Immobility/stupor,Staring,Grimacing,Mannerisms | 95.5 | 93.5 | 97.3 |
| 5 | Excitement,Immobility/stupor,Staring,Grimacing,Stereotypy | 95.5 | 93.5 | 97.3 |
| 5 | Excitement,Immobility/stupor,Staring,Stereotypy,Mannerisms | 95.5 | 93.5 | 97.3 |
| 5 | Excitement,Immobility/stupor,Stereotypy,Verbigeration,Rigidity | 95.5 | 93.5 | 97.3 |
| 5 | Excitement,Immobility/stupor,Verbigeration,Negativism,Withdrawal | 95.5 | 93.5 | 97.3 |
| 5 | Excitement,Staring,Grimacing,Verbigeration,Withdrawal | 95.5 | 93.5 | 97.3 |
| 5 | Excitement,Staring,Posturing/catalepsy,Rigidity,Waxy flexibility | 95.5 | 93.5 | 97.3 |
| 5 | Excitement,Staring,Stereotypy,Mannerisms,Rigidity | 95.5 | 93.5 | 97.3 |
| 5 | Excitement,Staring,Stereotypy,Mannerisms,Withdrawal | 95.5 | 93.5 | 97.3 |
| 5 | Immobility/stupor,Grimacing,Mannerisms,Rigidity,Negativism | 95.5 | 93.5 | 97.3 |
| 5 | Immobility/stupor,Mannerisms,Verbigeration,Rigidity,Negativism | 95.5 | 93.5 | 97.3 |
| 5 | Immobility/stupor,Staring,Echopraxia/echolalia,Mannerisms,Rigidity | 95.5 | 93.5 | 97.3 |
| 5 | Immobility/stupor,Staring,Echopraxia/echolalia,Stereotypy,Negativism | 95.5 | 93.5 | 97.3 |
| 5 | Immobility/stupor,Staring,Grimacing,Mannerisms,Negativism | 95.5 | 93.5 | 97.3 |
| 5 | Immobility/stupor,Staring,Posturing/catalepsy,Mannerisms,Rigidity | 95.5 | 93.5 | 97.3 |
| 5 | Immobility/stupor,Staring,Stereotypy,Rigidity,Withdrawal | 95.5 | 93.5 | 97.3 |
| 5 | Immobility/stupor,Stereotypy,Verbigeration,Rigidity,Negativism | 95.5 | 93.5 | 97.3 |
| 5 | Mutism,Staring,Echopraxia/echolalia,Stereotypy,Mannerisms | 95.5 | 93.5 | 97.3 |
| 5 | Mutism,Staring,Grimacing,Echopraxia/echolalia,Mannerisms | 95.5 | 93.5 | 97.3 |
| 5 | Mutism,Staring,Grimacing,Echopraxia/echolalia,Negativism | 95.5 | 93.5 | 97.3 |
| 5 | Mutism,Staring,Grimacing,Echopraxia/echolalia,Rigidity | 95.5 | 93.5 | 97.3 |
| 5 | Mutism,Staring,Grimacing,Stereotypy,Verbigeration | 95.5 | 93.5 | 97.3 |
| 5 | Mutism,Staring,Grimacing,Stereotypy,Withdrawal | 95.5 | 93.5 | 97.3 |
| 5 | Mutism,Staring,Mannerisms,Verbigeration,Withdrawal | 95.5 | 93.5 | 97.3 |
| 5 | Mutism,Staring,Posturing/catalepsy,Echopraxia/echolalia,Stereotypy | 95.5 | 93.5 | 97.3 |
| 5 | Mutism,Staring,Posturing/catalepsy,Grimacing,Withdrawal | 95.5 | 93.5 | 97.3 |
| 5 | Mutism,Staring,Posturing/catalepsy,Mannerisms,Verbigeration | 95.5 | 93.5 | 97.3 |
| 5 | Mutism,Staring,Posturing/catalepsy,Stereotypy,Mannerisms | 95.5 | 93.5 | 97.3 |
| 5 | Mutism,Staring,Posturing/catalepsy,Stereotypy,Verbigeration | 95.5 | 93.5 | 97.3 |
| 5 | Mutism,Staring,Posturing/catalepsy,Verbigeration,Withdrawal | 95.5 | 93.5 | 97.3 |
| 5 | Staring,Grimacing,Mannerisms,Rigidity,Negativism | 95.5 | 93.5 | 97.3 |
| 5 | Staring,Grimacing,Verbigeration,Rigidity,Withdrawal | 95.5 | 93.5 | 97.3 |
| 5 | Staring,Posturing/catalepsy,Stereotypy,Mannerisms,Negativism | 95.5 | 93.5 | 97.3 |
| 5 | Staring,Posturing/catalepsy,Stereotypy,Rigidity,Negativism | 95.5 | 93.5 | 97.3 |
| 5 | Staring,Posturing/catalepsy,Stereotypy,Rigidity,Withdrawal | 95.5 | 93.5 | 97.3 |
| 5 | Staring,Stereotypy,Negativism,Waxy flexibility,Withdrawal | 95.5 | 93.5 | 97.3 |
| 5 | Excitement,Immobility/stupor,Echopraxia/echolalia,Mannerisms,Rigidity | 95.3 | 93.3 | 97.1 |
| 5 | Excitement,Immobility/stupor,Grimacing,Mannerisms,Rigidity | 95.3 | 93.3 | 97.1 |
| 5 | Excitement,Immobility/stupor,Grimacing,Rigidity,Waxy flexibility | 95.3 | 93.3 | 97.1 |
| 5 | Excitement,Immobility/stupor,Grimacing,Stereotypy,Rigidity | 95.3 | 93.3 | 97.1 |
| 5 | Excitement,Immobility/stupor,Mannerisms,Verbigeration,Rigidity | 95.3 | 93.3 | 97.1 |
| 5 | Excitement,Immobility/stupor,Mutism,Negativism,Withdrawal | 95.3 | 93.3 | 97.1 |
| 5 | Excitement,Immobility/stupor,Mutism,Posturing/catalepsy,Verbigeration | 95.3 | 93.3 | 97.1 |
| 5 | Excitement,Immobility/stupor,Mutism,Stereotypy,Negativism | 95.3 | 93.3 | 97.1 |
| 5 | Excitement,Immobility/stupor,Mutism,Verbigeration,Negativism | 95.3 | 93.3 | 97.1 |
| 5 | Excitement,Immobility/stupor,Posturing/catalepsy,Grimacing,Withdrawal | 95.3 | 93.3 | 97.1 |
| 5 | Excitement,Immobility/stupor,Posturing/catalepsy,Rigidity,Waxy flexibility | 95.3 | 93.3 | 97.1 |
| 5 | Excitement,Immobility/stupor,Posturing/catalepsy,Verbigeration,Withdrawal | 95.3 | 93.3 | 97.1 |
| 5 | Excitement,Immobility/stupor,Stereotypy,Rigidity,Waxy flexibility | 95.3 | 93.3 | 97.1 |
| 5 | Excitement,Immobility/stupor,Verbigeration,Rigidity,Waxy flexibility | 95.3 | 93.3 | 97.1 |
| 5 | Excitement,Staring,Echopraxia/echolalia,Mannerisms,Negativism | 95.3 | 93.3 | 97.1 |
| 5 | Excitement,Staring,Echopraxia/echolalia,Mannerisms,Rigidity | 95.3 | 93.3 | 97.1 |
| 5 | Excitement,Staring,Echopraxia/echolalia,Verbigeration,Rigidity | 95.3 | 93.3 | 97.1 |
| 5 | Excitement,Staring,Mannerisms,Verbigeration,Rigidity | 95.3 | 93.3 | 97.1 |
| 5 | Excitement,Staring,Mannerisms,Verbigeration,Withdrawal | 95.3 | 93.3 | 97.1 |
| 5 | Excitement,Staring,Posturing/catalepsy,Grimacing,Echopraxia/echolalia | 95.3 | 93.3 | 97.1 |
| 5 | Excitement,Staring,Stereotypy,Verbigeration,Negativism | 95.3 | 93.3 | 97.1 |
| 5 | Excitement,Staring,Stereotypy,Verbigeration,Withdrawal | 95.3 | 93.3 | 97.1 |
| 5 | Immobility/stupor,Echopraxia/echolalia,Mannerisms,Rigidity,Negativism | 95.3 | 93.3 | 97.1 |
| 5 | Immobility/stupor,Staring,Echopraxia/echolalia,Rigidity,Withdrawal | 95.3 | 93.3 | 97.1 |
| 5 | Immobility/stupor,Staring,Grimacing,Verbigeration,Rigidity | 95.3 | 93.3 | 97.1 |
| 5 | Immobility/stupor,Staring,Posturing/catalepsy,Echopraxia/echolalia,Negativism | 95.3 | 93.3 | 97.1 |
| 5 | Immobility/stupor,Staring,Posturing/catalepsy,Mannerisms,Withdrawal | 95.3 | 93.3 | 97.1 |
| 5 | Immobility/stupor,Staring,Verbigeration,Rigidity,Withdrawal | 95.3 | 93.3 | 97.1 |
| 5 | Immobility/stupor,Verbigeration,Rigidity,Negativism,Withdrawal | 95.3 | 93.3 | 97.1 |
| 5 | Mutism,Staring,Echopraxia/echolalia,Negativism,Waxy flexibility | 95.3 | 93.3 | 97.1 |
| 5 | Mutism,Staring,Echopraxia/echolalia,Stereotypy,Withdrawal | 95.3 | 93.3 | 97.1 |
| 5 | Mutism,Staring,Grimacing,Echopraxia/echolalia,Stereotypy | 95.3 | 93.3 | 97.1 |
| 5 | Mutism,Staring,Grimacing,Echopraxia/echolalia,Withdrawal | 95.3 | 93.3 | 97.1 |
| 5 | Mutism,Staring,Grimacing,Rigidity,Negativism | 95.3 | 93.3 | 97.1 |
| 5 | Mutism,Staring,Grimacing,Stereotypy,Mannerisms | 95.3 | 93.3 | 97.1 |
| 5 | Mutism,Staring,Grimacing,Verbigeration,Withdrawal | 95.3 | 93.3 | 97.1 |
| 5 | Mutism,Staring,Posturing/catalepsy,Echopraxia/echolalia,Verbigeration | 95.3 | 93.3 | 97.1 |
| 5 | Mutism,Staring,Posturing/catalepsy,Grimacing,Mannerisms | 95.3 | 93.3 | 97.1 |
| 5 | Mutism,Staring,Posturing/catalepsy,Grimacing,Negativism | 95.3 | 93.3 | 97.1 |
| 5 | Mutism,Staring,Rigidity,Negativism,Waxy flexibility | 95.3 | 93.3 | 97.1 |
| 5 | Mutism,Staring,Rigidity,Waxy flexibility,Withdrawal | 95.3 | 93.3 | 97.1 |
| 5 | Mutism,Staring,Stereotypy,Verbigeration,Withdrawal | 95.3 | 93.3 | 97.1 |
| 5 | Mutism,Staring,Verbigeration,Negativism,Waxy flexibility | 95.3 | 93.3 | 97.1 |
| 5 | Staring,Echopraxia/echolalia,Negativism,Waxy flexibility,Withdrawal | 95.3 | 93.3 | 97.1 |
| 5 | Staring,Echopraxia/echolalia,Stereotypy,Mannerisms,Negativism | 95.3 | 93.3 | 97.1 |
| 5 | Staring,Echopraxia/echolalia,Stereotypy,Rigidity,Negativism | 95.3 | 93.3 | 97.1 |
| 5 | Staring,Grimacing,Echopraxia/echolalia,Mannerisms,Withdrawal | 95.3 | 93.3 | 97.1 |
| 5 | Staring,Grimacing,Echopraxia/echolalia,Negativism,Withdrawal | 95.3 | 93.3 | 97.1 |
| 5 | Staring,Grimacing,Echopraxia/echolalia,Rigidity,Withdrawal | 95.3 | 93.3 | 97.1 |
| 5 | Staring,Grimacing,Stereotypy,Mannerisms,Rigidity | 95.3 | 93.3 | 97.1 |
| 5 | Staring,Grimacing,Verbigeration,Negativism,Withdrawal | 95.3 | 93.3 | 97.1 |
| 5 | Staring,Posturing/catalepsy,Echopraxia/echolalia,Mannerisms,Negativism | 95.3 | 93.3 | 97.1 |
| 5 | Staring,Posturing/catalepsy,Echopraxia/echolalia,Rigidity,Withdrawal | 95.3 | 93.3 | 97.1 |
| 5 | Staring,Posturing/catalepsy,Negativism,Waxy flexibility,Withdrawal | 95.3 | 93.3 | 97.1 |
| 5 | Staring,Posturing/catalepsy,Stereotypy,Mannerisms,Withdrawal | 95.3 | 93.3 | 97.1 |
| 5 | Staring,Stereotypy,Rigidity,Waxy flexibility,Withdrawal | 95.3 | 93.3 | 97.1 |
| 5 | Staring,Verbigeration,Negativism,Waxy flexibility,Withdrawal | 95.3 | 93.3 | 97.1 |
| 5 | Excitement,Immobility/stupor,Echopraxia/echolalia,Negativism,Waxy flexibility | 95.1 | 93.0 | 97.1 |
| 5 | Excitement,Immobility/stupor,Mutism,Echopraxia/echolalia,Negativism | 95.1 | 92.8 | 97.1 |
| 5 | Excitement,Immobility/stupor,Mutism,Echopraxia/echolalia,Withdrawal | 95.1 | 93.0 | 96.9 |
| 5 | Excitement,Immobility/stupor,Mutism,Waxy flexibility,Withdrawal | 95.1 | 93.0 | 97.1 |
| 5 | Excitement,Immobility/stupor,Negativism,Waxy flexibility,Withdrawal | 95.1 | 93.0 | 97.1 |
| 5 | Excitement,Immobility/stupor,Posturing/catalepsy,Mannerisms,Withdrawal | 95.1 | 93.0 | 97.1 |
| 5 | Excitement,Immobility/stupor,Stereotypy,Verbigeration,Negativism | 95.1 | 93.0 | 97.1 |
| 5 | Excitement,Staring,Echopraxia/echolalia,Verbigeration,Negativism | 95.1 | 93.0 | 97.1 |
| 5 | Excitement,Staring,Echopraxia/echolalia,Verbigeration,Withdrawal | 95.1 | 92.8 | 96.9 |
| 5 | Excitement,Staring,Grimacing,Echopraxia/echolalia,Negativism | 95.1 | 93.0 | 97.1 |
| 5 | Excitement,Staring,Grimacing,Negativism,Waxy flexibility | 95.1 | 93.0 | 97.1 |
| 5 | Excitement,Staring,Posturing/catalepsy,Grimacing,Mannerisms | 95.1 | 93.0 | 96.9 |
| 5 | Immobility/stupor,Mannerisms,Rigidity,Negativism,Waxy flexibility | 95.1 | 93.0 | 96.9 |
| 5 | Immobility/stupor,Mutism,Staring,Mannerisms,Withdrawal | 95.1 | 93.0 | 97.1 |
| 5 | Immobility/stupor,Mutism,Staring,Negativism,Withdrawal | 95.1 | 92.8 | 97.1 |
| 5 | Immobility/stupor,Mutism,Staring,Verbigeration,Rigidity | 95.1 | 93.0 | 96.9 |
| 5 | Immobility/stupor,Staring,Grimacing,Rigidity,Negativism | 95.1 | 93.0 | 96.9 |
| 5 | Immobility/stupor,Staring,Grimacing,Stereotypy,Negativism | 95.1 | 93.0 | 96.9 |
| 5 | Immobility/stupor,Staring,Grimacing,Stereotypy,Rigidity | 95.1 | 93.0 | 97.1 |
| 5 | Immobility/stupor,Staring,Grimacing,Verbigeration,Negativism | 95.1 | 93.0 | 97.1 |
| 5 | Immobility/stupor,Staring,Mannerisms,Rigidity,Waxy flexibility | 95.1 | 93.0 | 97.1 |
| 5 | Immobility/stupor,Staring,Posturing/catalepsy,Rigidity,Negativism | 95.1 | 93.0 | 97.1 |
| 5 | Immobility/stupor,Staring,Posturing/catalepsy,Verbigeration,Negativism | 95.1 | 93.0 | 96.9 |
| 5 | Immobility/stupor,Staring,Stereotypy,Mannerisms,Withdrawal | 95.1 | 93.0 | 97.1 |
| 5 | Immobility/stupor,Staring,Stereotypy,Negativism,Waxy flexibility | 95.1 | 93.0 | 96.9 |
| 5 | Mutism,Staring,Grimacing,Echopraxia/echolalia,Verbigeration | 95.1 | 93.0 | 97.1 |
| 5 | Mutism,Staring,Grimacing,Mannerisms,Verbigeration | 95.1 | 93.0 | 97.1 |
| 5 | Mutism,Staring,Posturing/catalepsy,Grimacing,Verbigeration | 95.1 | 93.0 | 97.1 |
| 5 | Mutism,Staring,Stereotypy,Mannerisms,Verbigeration | 95.1 | 93.0 | 96.9 |
| 5 | Mutism,Staring,Stereotypy,Rigidity,Waxy flexibility | 95.1 | 92.8 | 96.9 |
| 5 | Mutism,Staring,Stereotypy,Waxy flexibility,Withdrawal | 95.1 | 93.0 | 96.9 |
| 5 | Mutism,Staring,Verbigeration,Rigidity,Waxy flexibility | 95.1 | 93.0 | 96.9 |
| 5 | Staring,Echopraxia/echolalia,Verbigeration,Negativism,Withdrawal | 95.1 | 93.0 | 97.1 |
| 5 | Staring,Grimacing,Stereotypy,Verbigeration,Rigidity | 95.1 | 93.0 | 97.1 |
| 5 | Staring,Posturing/catalepsy,Echopraxia/echolalia,Mannerisms,Withdrawal | 95.1 | 93.0 | 97.1 |
| 5 | Staring,Posturing/catalepsy,Echopraxia/echolalia,Rigidity,Negativism | 95.1 | 93.0 | 97.1 |
| 5 | Staring,Posturing/catalepsy,Grimacing,Rigidity,Withdrawal | 95.1 | 93.0 | 96.9 |
| 5 | Staring,Posturing/catalepsy,Verbigeration,Rigidity,Withdrawal | 95.1 | 93.0 | 97.1 |
| 5 | Staring,Stereotypy,Mannerisms,Negativism,Waxy flexibility | 95.1 | 93.0 | 97.1 |
| 5 | Staring,Stereotypy,Rigidity,Negativism,Waxy flexibility | 95.1 | 93.0 | 97.1 |
| 5 | Immobility/stupor,Mutism,Staring,Stereotypy,Negativism | 95.0 | 92.8 | 96.9 |
| 5 | Staring,Grimacing,Mannerisms,Waxy flexibility,Withdrawal | 95.0 | 92.8 | 97.1 |
| 5 | Excitement,Immobility/stupor,Mutism,Posturing/catalepsy,Stereotypy | 94.9 | 92.8 | 96.9 |
| 5 | Excitement,Staring,Grimacing,Echopraxia/echolalia,Waxy flexibility | 94.9 | 92.6 | 96.9 |
| 5 | Excitement,Staring,Posturing/catalepsy,Grimacing,Stereotypy | 94.9 | 92.8 | 96.9 |
| 5 | Excitement,Staring,Posturing/catalepsy,Grimacing,Verbigeration | 94.9 | 92.6 | 96.9 |
| 5 | Immobility/stupor,Mutism,Verbigeration,Rigidity,Negativism | 94.9 | 92.6 | 96.9 |
| 5 | Mutism,Staring,Posturing/catalepsy,Grimacing,Echopraxia/echolalia | 94.9 | 92.8 | 96.9 |
| 5 | Mutism,Staring,Posturing/catalepsy,Grimacing,Stereotypy | 94.9 | 92.8 | 96.6 |
| 5 | Mutism,Staring,Posturing/catalepsy,Waxy flexibility,Withdrawal | 94.9 | 92.6 | 96.9 |
| 5 | Mutism,Staring,Stereotypy,Verbigeration,Waxy flexibility | 94.9 | 92.6 | 96.9 |
| 5 | Staring,Echopraxia/echolalia,Stereotypy,Mannerisms,Withdrawal | 94.9 | 92.8 | 96.9 |
| 5 | Staring,Echopraxia/echolalia,Verbigeration,Rigidity,Negativism | 94.9 | 92.6 | 96.9 |
| 5 | Staring,Grimacing,Stereotypy,Rigidity,Negativism | 94.9 | 92.8 | 96.9 |
| 5 | Staring,Grimacing,Stereotypy,Verbigeration,Withdrawal | 94.9 | 92.6 | 96.9 |
| 5 | Staring,Posturing/catalepsy,Mannerisms,Verbigeration,Negativism | 94.9 | 92.8 | 96.9 |
| 5 | Staring,Posturing/catalepsy,Stereotypy,Verbigeration,Negativism | 94.9 | 92.8 | 96.9 |
| 5 | Excitement,Immobility/stupor,Echopraxia/echolalia,Stereotypy,Negativism | 94.8 | 92.6 | 96.9 |
| 5 | Excitement,Immobility/stupor,Echopraxia/echolalia,Waxy flexibility,Withdrawal | 94.8 | 92.6 | 96.9 |
| 5 | Excitement,Immobility/stupor,Mannerisms,Rigidity,Waxy flexibility | 94.8 | 92.6 | 96.9 |
| 5 | Excitement,Immobility/stupor,Mutism,Echopraxia/echolalia,Waxy flexibility | 94.8 | 92.6 | 96.6 |
| 5 | Excitement,Immobility/stupor,Mutism,Posturing/catalepsy,Mannerisms | 94.8 | 92.8 | 96.9 |
| 5 | Excitement,Immobility/stupor,Mutism,Posturing/catalepsy,Waxy flexibility | 94.8 | 92.6 | 96.9 |
| 5 | Excitement,Immobility/stupor,Mutism,Stereotypy,Withdrawal | 94.8 | 92.8 | 96.9 |
| 5 | Excitement,Immobility/stupor,Posturing/catalepsy,Grimacing,Echopraxia/echolalia | 94.8 | 92.6 | 96.9 |
| 5 | Excitement,Immobility/stupor,Stereotypy,Negativism,Waxy flexibility | 94.8 | 92.6 | 96.9 |
| 5 | Excitement,Staring,Stereotypy,Mannerisms,Negativism | 94.8 | 92.8 | 96.9 |
| 5 | Immobility/stupor,Echopraxia/echolalia,Rigidity,Negativism,Withdrawal | 94.8 | 92.8 | 96.9 |
| 5 | Immobility/stupor,Mutism,Staring,Rigidity,Negativism | 94.8 | 92.6 | 96.9 |
| 5 | Immobility/stupor,Mutism,Staring,Rigidity,Withdrawal | 94.8 | 92.8 | 96.9 |
| 5 | Immobility/stupor,Posturing/catalepsy,Mannerisms,Negativism,Withdrawal | 94.8 | 92.6 | 96.6 |
| 5 | Immobility/stupor,Posturing/catalepsy,Stereotypy,Rigidity,Negativism | 94.8 | 92.6 | 96.9 |
| 5 | Immobility/stupor,Staring,Echopraxia/echolalia,Mannerisms,Withdrawal | 94.8 | 92.6 | 96.9 |
| 5 | Immobility/stupor,Staring,Echopraxia/echolalia,Verbigeration,Negativism | 94.8 | 92.6 | 96.9 |
| 5 | Immobility/stupor,Staring,Grimacing,Mannerisms,Withdrawal | 94.8 | 92.8 | 96.9 |
| 5 | Immobility/stupor,Staring,Grimacing,Negativism,Withdrawal | 94.8 | 92.6 | 96.9 |
| 5 | Immobility/stupor,Staring,Grimacing,Rigidity,Withdrawal | 94.8 | 92.8 | 96.9 |
| 5 | Immobility/stupor,Staring,Negativism,Waxy flexibility,Withdrawal | 94.8 | 92.8 | 96.9 |
| 5 | Immobility/stupor,Staring,Posturing/catalepsy,Rigidity,Withdrawal | 94.8 | 92.8 | 96.9 |
| 5 | Mutism,Staring,Echopraxia/echolalia,Mannerisms,Waxy flexibility | 94.8 | 92.8 | 96.9 |
| 5 | Mutism,Staring,Grimacing,Mannerisms,Waxy flexibility | 94.8 | 92.6 | 96.9 |
| 5 | Mutism,Staring,Posturing/catalepsy,Negativism,Waxy flexibility | 94.8 | 92.8 | 96.9 |
| 5 | Mutism,Staring,Stereotypy,Mannerisms,Waxy flexibility | 94.8 | 92.6 | 96.9 |
| 5 | Staring,Echopraxia/echolalia,Mannerisms,Negativism,Waxy flexibility | 94.8 | 92.6 | 96.9 |
| 5 | Staring,Grimacing,Echopraxia/echolalia,Stereotypy,Withdrawal | 94.8 | 92.6 | 96.9 |
| 5 | Staring,Grimacing,Mannerisms,Verbigeration,Withdrawal | 94.8 | 92.6 | 96.9 |
| 5 | Staring,Posturing/catalepsy,Grimacing,Mannerisms,Negativism | 94.8 | 92.8 | 96.9 |
| 5 | Staring,Posturing/catalepsy,Grimacing,Mannerisms,Rigidity | 94.8 | 92.8 | 96.9 |
| 5 | Staring,Posturing/catalepsy,Mannerisms,Verbigeration,Withdrawal | 94.8 | 92.6 | 96.9 |
| 5 | Staring,Stereotypy,Mannerisms,Waxy flexibility,Withdrawal | 94.8 | 92.6 | 96.9 |
| 5 | Excitement,Immobility/stupor,Echopraxia/echolalia,Stereotypy,Withdrawal | 94.6 | 92.4 | 96.6 |
| 5 | Excitement,Immobility/stupor,Echopraxia/echolalia,Verbigeration,Negativism | 94.6 | 92.4 | 96.6 |
| 5 | Excitement,Immobility/stupor,Echopraxia/echolalia,Verbigeration,Withdrawal | 94.6 | 92.4 | 96.6 |
| 5 | Excitement,Immobility/stupor,Grimacing,Echopraxia/echolalia,Negativism | 94.6 | 92.4 | 96.6 |
| 5 | Excitement,Immobility/stupor,Grimacing,Negativism,Waxy flexibility | 94.6 | 92.4 | 96.6 |
| 5 | Excitement,Immobility/stupor,Grimacing,Negativism,Withdrawal | 94.6 | 92.4 | 96.6 |
| 5 | Excitement,Immobility/stupor,Mannerisms,Negativism,Withdrawal | 94.6 | 92.4 | 96.6 |
| 5 | Excitement,Immobility/stupor,Mutism,Grimacing,Negativism | 94.6 | 92.4 | 96.6 |
| 5 | Excitement,Immobility/stupor,Mutism,Verbigeration,Withdrawal | 94.6 | 92.4 | 96.6 |
| 5 | Excitement,Immobility/stupor,Posturing/catalepsy,Stereotypy,Withdrawal | 94.6 | 92.4 | 96.6 |
| 5 | Excitement,Immobility/stupor,Posturing/catalepsy,Waxy flexibility,Withdrawal | 94.6 | 92.4 | 96.6 |
| 5 | Excitement,Immobility/stupor,Stereotypy,Mannerisms,Rigidity | 94.6 | 92.4 | 96.6 |
| 5 | Excitement,Immobility/stupor,Verbigeration,Negativism,Waxy flexibility | 94.6 | 92.4 | 96.6 |
| 5 | Excitement,Staring,Grimacing,Stereotypy,Negativism | 94.6 | 92.4 | 96.6 |
| 5 | Excitement,Staring,Mannerisms,Verbigeration,Negativism | 94.6 | 92.4 | 96.6 |
| 5 | Excitement,Staring,Posturing/catalepsy,Mannerisms,Verbigeration | 94.6 | 92.4 | 96.6 |
| 5 | Immobility/stupor,Mutism,Staring,Echopraxia/echolalia,Rigidity | 94.6 | 92.4 | 96.6 |
| 5 | Immobility/stupor,Mutism,Staring,Posturing/catalepsy,Negativism | 94.6 | 92.4 | 96.6 |
| 5 | Immobility/stupor,Mutism,Staring,Stereotypy,Rigidity | 94.6 | 92.4 | 96.6 |
| 5 | Immobility/stupor,Mutism,Staring,Verbigeration,Negativism | 94.6 | 92.4 | 96.6 |
| 5 | Immobility/stupor,Mutism,Stereotypy,Rigidity,Negativism | 94.6 | 92.4 | 96.6 |
| 5 | Immobility/stupor,Posturing/catalepsy,Rigidity,Negativism,Withdrawal | 94.6 | 92.4 | 96.6 |
| 5 | Immobility/stupor,Staring,Echopraxia/echolalia,Stereotypy,Rigidity | 94.6 | 92.6 | 96.6 |
| 5 | Immobility/stupor,Staring,Echopraxia/echolalia,Verbigeration,Rigidity | 94.6 | 92.4 | 96.6 |
| 5 | Immobility/stupor,Staring,Grimacing,Echopraxia/echolalia,Negativism | 94.6 | 92.4 | 96.6 |
| 5 | Immobility/stupor,Staring,Posturing/catalepsy,Stereotypy,Rigidity | 94.6 | 92.4 | 96.6 |
| 5 | Immobility/stupor,Staring,Rigidity,Negativism,Waxy flexibility | 94.6 | 92.4 | 96.6 |
| 5 | Immobility/stupor,Stereotypy,Mannerisms,Negativism,Withdrawal | 94.6 | 92.4 | 96.6 |
| 5 | Mutism,Staring,Echopraxia/echolalia,Mannerisms,Verbigeration | 94.6 | 92.4 | 96.6 |
| 5 | Mutism,Staring,Echopraxia/echolalia,Rigidity,Waxy flexibility | 94.6 | 92.4 | 96.6 |
| 5 | Mutism,Staring,Echopraxia/echolalia,Stereotypy,Verbigeration | 94.6 | 92.4 | 96.6 |
| 5 | Mutism,Staring,Echopraxia/echolalia,Stereotypy,Waxy flexibility | 94.6 | 92.4 | 96.6 |
| 5 | Mutism,Staring,Echopraxia/echolalia,Verbigeration,Withdrawal | 94.6 | 92.4 | 96.6 |
| 5 | Mutism,Staring,Echopraxia/echolalia,Waxy flexibility,Withdrawal | 94.6 | 92.4 | 96.6 |
| 5 | Mutism,Staring,Grimacing,Verbigeration,Waxy flexibility | 94.6 | 92.4 | 96.6 |
| 5 | Mutism,Staring,Verbigeration,Waxy flexibility,Withdrawal | 94.6 | 92.4 | 96.6 |
| 5 | Staring,Echopraxia/echolalia,Verbigeration,Rigidity,Withdrawal | 94.6 | 92.4 | 96.6 |
| 5 | Staring,Grimacing,Echopraxia/echolalia,Mannerisms,Rigidity | 94.6 | 92.4 | 96.6 |
| 5 | Staring,Grimacing,Mannerisms,Rigidity,Waxy flexibility | 94.6 | 92.6 | 96.6 |
| 5 | Staring,Grimacing,Mannerisms,Verbigeration,Rigidity | 94.6 | 92.6 | 96.6 |
| 5 | Staring,Grimacing,Rigidity,Waxy flexibility,Withdrawal | 94.6 | 92.4 | 96.6 |
| 5 | Staring,Grimacing,Verbigeration,Rigidity,Negativism | 94.6 | 92.4 | 96.6 |
| 5 | Staring,Posturing/catalepsy,Echopraxia/echolalia,Stereotypy,Negativism | 94.6 | 92.4 | 96.6 |
| 5 | Staring,Posturing/catalepsy,Grimacing,Stereotypy,Withdrawal | 94.6 | 92.4 | 96.6 |
| 5 | Staring,Posturing/catalepsy,Mannerisms,Waxy flexibility,Withdrawal | 94.6 | 92.4 | 96.6 |
| 5 | Staring,Posturing/catalepsy,Verbigeration,Rigidity,Negativism | 94.6 | 92.4 | 96.6 |
| 5 | Staring,Stereotypy,Mannerisms,Verbigeration,Negativism | 94.6 | 92.4 | 96.6 |
| 5 | Staring,Verbigeration,Rigidity,Waxy flexibility,Withdrawal | 94.6 | 92.4 | 96.6 |
| 5 | Excitement,Immobility/stupor,Grimacing,Echopraxia/echolalia,Withdrawal | 94.4 | 92.2 | 96.4 |
| 5 | Excitement,Immobility/stupor,Grimacing,Verbigeration,Negativism | 94.4 | 92.2 | 96.4 |
| 5 | Excitement,Immobility/stupor,Mutism,Grimacing,Waxy flexibility | 94.4 | 92.2 | 96.4 |
| 5 | Excitement,Immobility/stupor,Mutism,Grimacing,Withdrawal | 94.4 | 92.2 | 96.4 |
| 5 | Excitement,Immobility/stupor,Mutism,Stereotypy,Waxy flexibility | 94.4 | 92.2 | 96.4 |
| 5 | Excitement,Immobility/stupor,Mutism,Verbigeration,Waxy flexibility | 94.4 | 92.2 | 96.4 |
| 5 | Excitement,Immobility/stupor,Posturing/catalepsy,Echopraxia/echolalia,Verbigeration | 94.4 | 92.2 | 96.4 |
| 5 | Excitement,Immobility/stupor,Posturing/catalepsy,Grimacing,Verbigeration | 94.4 | 92.2 | 96.4 |
| 5 | Excitement,Staring,Grimacing,Mannerisms,Waxy flexibility | 94.4 | 92.2 | 96.4 |
| 5 | Excitement,Staring,Grimacing,Stereotypy,Waxy flexibility | 94.4 | 92.2 | 96.4 |
| 5 | Excitement,Staring,Grimacing,Verbigeration,Negativism | 94.4 | 92.2 | 96.4 |
| 5 | Excitement,Staring,Posturing/catalepsy,Echopraxia/echolalia,Mannerisms | 94.4 | 92.2 | 96.4 |
| 5 | Excitement,Staring,Posturing/catalepsy,Echopraxia/echolalia,Stereotypy | 94.4 | 92.2 | 96.4 |
| 5 | Excitement,Staring,Posturing/catalepsy,Echopraxia/echolalia,Verbigeration | 94.4 | 92.2 | 96.4 |
| 5 | Excitement,Staring,Posturing/catalepsy,Echopraxia/echolalia,Waxy flexibility | 94.4 | 92.2 | 96.4 |
| 5 | Excitement,Staring,Posturing/catalepsy,Grimacing,Waxy flexibility | 94.4 | 92.2 | 96.4 |
| 5 | Excitement,Staring,Posturing/catalepsy,Verbigeration,Waxy flexibility | 94.4 | 92.2 | 96.4 |
| 5 | Immobility/stupor,Echopraxia/echolalia,Stereotypy,Negativism,Withdrawal | 94.4 | 92.2 | 96.4 |
| 5 | Immobility/stupor,Grimacing,Stereotypy,Rigidity,Negativism | 94.4 | 92.2 | 96.4 |
| 5 | Immobility/stupor,Grimacing,Verbigeration,Rigidity,Negativism | 94.4 | 92.2 | 96.4 |
| 5 | Immobility/stupor,Mutism,Mannerisms,Rigidity,Withdrawal | 94.4 | 92.2 | 96.4 |
| 5 | Immobility/stupor,Mutism,Staring,Echopraxia/echolalia,Mannerisms | 94.4 | 92.2 | 96.4 |
| 5 | Immobility/stupor,Mutism,Staring,Echopraxia/echolalia,Negativism | 94.4 | 92.2 | 96.4 |
| 5 | Immobility/stupor,Mutism,Staring,Posturing/catalepsy,Mannerisms | 94.4 | 92.2 | 96.4 |
| 5 | Immobility/stupor,Mutism,Staring,Posturing/catalepsy,Withdrawal | 94.4 | 92.2 | 96.4 |
| 5 | Immobility/stupor,Posturing/catalepsy,Stereotypy,Negativism,Withdrawal | 94.4 | 92.2 | 96.4 |
| 5 | Immobility/stupor,Posturing/catalepsy,Verbigeration,Rigidity,Negativism | 94.4 | 92.2 | 96.4 |
| 5 | Immobility/stupor,Staring,Echopraxia/echolalia,Stereotypy,Mannerisms | 94.4 | 92.2 | 96.4 |
| 5 | Immobility/stupor,Staring,Grimacing,Echopraxia/echolalia,Mannerisms | 94.4 | 92.2 | 96.4 |
| 5 | Immobility/stupor,Staring,Grimacing,Echopraxia/echolalia,Rigidity | 94.4 | 92.2 | 96.4 |
| 5 | Immobility/stupor,Staring,Grimacing,Stereotypy,Mannerisms | 94.4 | 92.2 | 96.4 |
| 5 | Immobility/stupor,Staring,Grimacing,Stereotypy,Verbigeration | 94.4 | 92.2 | 96.4 |
| 5 | Immobility/stupor,Staring,Mannerisms,Verbigeration,Withdrawal | 94.4 | 92.2 | 96.4 |
| 5 | Immobility/stupor,Staring,Mannerisms,Waxy flexibility,Withdrawal | 94.4 | 92.2 | 96.4 |
| 5 | Immobility/stupor,Staring,Posturing/catalepsy,Grimacing,Negativism | 94.4 | 92.2 | 96.4 |
| 5 | Immobility/stupor,Staring,Posturing/catalepsy,Mannerisms,Verbigeration | 94.4 | 92.2 | 96.4 |
| 5 | Immobility/stupor,Staring,Posturing/catalepsy,Verbigeration,Rigidity | 94.4 | 92.2 | 96.4 |
| 5 | Immobility/stupor,Staring,Rigidity,Waxy flexibility,Withdrawal | 94.4 | 92.2 | 96.4 |
| 5 | Immobility/stupor,Staring,Stereotypy,Mannerisms,Verbigeration | 94.4 | 92.2 | 96.4 |
| 5 | Immobility/stupor,Stereotypy,Rigidity,Negativism,Waxy flexibility | 94.4 | 92.2 | 96.4 |
| 5 | Immobility/stupor,Stereotypy,Verbigeration,Negativism,Withdrawal | 94.4 | 92.2 | 96.4 |
| 5 | Mutism,Staring,Grimacing,Negativism,Waxy flexibility | 94.4 | 92.2 | 96.4 |
| 5 | Mutism,Staring,Grimacing,Stereotypy,Waxy flexibility | 94.4 | 92.2 | 96.4 |
| 5 | Mutism,Staring,Grimacing,Waxy flexibility,Withdrawal | 94.4 | 92.2 | 96.4 |
| 5 | Mutism,Staring,Mannerisms,Verbigeration,Waxy flexibility | 94.4 | 92.2 | 96.4 |
| 5 | Mutism,Staring,Posturing/catalepsy,Grimacing,Rigidity | 94.4 | 92.2 | 96.4 |
| 5 | Mutism,Staring,Posturing/catalepsy,Mannerisms,Waxy flexibility | 94.4 | 92.2 | 96.4 |
| 5 | Staring,Echopraxia/echolalia,Mannerisms,Waxy flexibility,Withdrawal | 94.4 | 92.2 | 96.4 |
| 5 | Staring,Echopraxia/echolalia,Rigidity,Negativism,Waxy flexibility | 94.4 | 92.2 | 96.4 |
| 5 | Staring,Echopraxia/echolalia,Rigidity,Waxy flexibility,Withdrawal | 94.4 | 92.2 | 96.4 |
| 5 | Staring,Grimacing,Echopraxia/echolalia,Mannerisms,Negativism | 94.4 | 92.2 | 96.4 |
| 5 | Staring,Grimacing,Echopraxia/echolalia,Rigidity,Negativism | 94.4 | 92.2 | 96.4 |
| 5 | Staring,Grimacing,Negativism,Waxy flexibility,Withdrawal | 94.4 | 92.2 | 96.4 |
| 5 | Staring,Grimacing,Stereotypy,Mannerisms,Negativism | 94.4 | 92.2 | 96.4 |
| 5 | Staring,Posturing/catalepsy,Grimacing,Echopraxia/echolalia,Withdrawal | 94.4 | 92.2 | 96.4 |
| 5 | Staring,Posturing/catalepsy,Mannerisms,Negativism,Waxy flexibility | 94.4 | 92.2 | 96.4 |
| 5 | Staring,Posturing/catalepsy,Mannerisms,Verbigeration,Rigidity | 94.4 | 92.2 | 96.4 |
| 5 | Staring,Posturing/catalepsy,Stereotypy,Mannerisms,Rigidity | 94.4 | 92.2 | 96.4 |
| 5 | Staring,Stereotypy,Mannerisms,Verbigeration,Rigidity | 94.4 | 92.2 | 96.4 |
| 5 | Staring,Stereotypy,Mannerisms,Verbigeration,Withdrawal | 94.4 | 92.2 | 96.4 |
| 5 | Excitement,Immobility/stupor,Grimacing,Echopraxia/echolalia,Waxy flexibility | 94.2 | 91.9 | 96.2 |
| 5 | Excitement,Immobility/stupor,Grimacing,Stereotypy,Negativism | 94.2 | 91.9 | 96.2 |
| 5 | Excitement,Immobility/stupor,Mannerisms,Negativism,Waxy flexibility | 94.2 | 91.9 | 96.2 |
| 5 | Excitement,Immobility/stupor,Mutism,Echopraxia/echolalia,Stereotypy | 94.2 | 91.9 | 96.2 |
| 5 | Excitement,Immobility/stupor,Mutism,Echopraxia/echolalia,Verbigeration | 94.2 | 91.9 | 96.2 |
| 5 | Excitement,Immobility/stupor,Posturing/catalepsy,Grimacing,Mannerisms | 94.2 | 91.9 | 96.2 |
| 5 | Excitement,Immobility/stupor,Stereotypy,Verbigeration,Withdrawal | 94.2 | 91.9 | 96.2 |
| 5 | Excitement,Staring,Echopraxia/echolalia,Stereotypy,Waxy flexibility | 94.2 | 91.9 | 96.2 |
| 5 | Excitement,Staring,Grimacing,Echopraxia/echolalia,Stereotypy | 94.2 | 91.9 | 96.2 |
| 5 | Excitement,Staring,Grimacing,Mannerisms,Negativism | 94.2 | 91.9 | 96.2 |
| 5 | Excitement,Staring,Grimacing,Verbigeration,Waxy flexibility | 94.2 | 91.9 | 96.2 |
| 5 | Excitement,Staring,Posturing/catalepsy,Mannerisms,Waxy flexibility | 94.2 | 91.9 | 96.2 |
| 5 | Excitement,Staring,Posturing/catalepsy,Stereotypy,Mannerisms | 94.2 | 91.9 | 96.2 |
| 5 | Excitement,Staring,Posturing/catalepsy,Stereotypy,Verbigeration | 94.2 | 91.9 | 96.2 |
| 5 | Excitement,Staring,Posturing/catalepsy,Stereotypy,Waxy flexibility | 94.2 | 91.9 | 96.2 |
| 5 | Immobility/stupor,Echopraxia/echolalia,Stereotypy,Rigidity,Negativism | 94.2 | 91.9 | 96.2 |
| 5 | Immobility/stupor,Echopraxia/echolalia,Verbigeration,Rigidity,Negativism | 94.2 | 91.9 | 96.2 |
| 5 | Immobility/stupor,Grimacing,Mannerisms,Rigidity,Withdrawal | 94.2 | 91.9 | 96.2 |
| 5 | Immobility/stupor,Mutism,Staring,Mannerisms,Verbigeration | 94.2 | 91.9 | 96.4 |
| 5 | Immobility/stupor,Mutism,Staring,Stereotypy,Mannerisms | 94.2 | 91.9 | 96.2 |
| 5 | Immobility/stupor,Posturing/catalepsy,Verbigeration,Negativism,Withdrawal | 94.2 | 91.9 | 96.2 |
| 5 | Immobility/stupor,Staring,Echopraxia/echolalia,Negativism,Waxy flexibility | 94.2 | 91.9 | 96.2 |
| 5 | Immobility/stupor,Staring,Grimacing,Mannerisms,Verbigeration | 94.2 | 91.9 | 96.2 |
| 5 | Immobility/stupor,Staring,Grimacing,Stereotypy,Withdrawal | 94.2 | 91.9 | 96.2 |
| 5 | Immobility/stupor,Staring,Posturing/catalepsy,Echopraxia/echolalia,Mannerisms | 94.2 | 91.9 | 96.2 |
| 5 | Immobility/stupor,Staring,Posturing/catalepsy,Echopraxia/echolalia,Rigidity | 94.2 | 91.9 | 96.2 |
| 5 | Immobility/stupor,Staring,Posturing/catalepsy,Echopraxia/echolalia,Withdrawal | 94.2 | 91.9 | 96.2 |
| 5 | Immobility/stupor,Staring,Posturing/catalepsy,Grimacing,Mannerisms | 94.2 | 91.9 | 96.2 |
| 5 | Immobility/stupor,Staring,Posturing/catalepsy,Stereotypy,Mannerisms | 94.2 | 91.9 | 96.2 |
| 5 | Immobility/stupor,Staring,Posturing/catalepsy,Stereotypy,Verbigeration | 94.2 | 91.9 | 96.2 |
| 5 | Immobility/stupor,Staring,Posturing/catalepsy,Stereotypy,Withdrawal | 94.2 | 91.9 | 96.2 |
| 5 | Immobility/stupor,Staring,Stereotypy,Rigidity,Waxy flexibility | 94.2 | 91.9 | 96.2 |
| 5 | Immobility/stupor,Staring,Verbigeration,Negativism,Waxy flexibility | 94.2 | 91.9 | 96.2 |
| 5 | Mutism,Staring,Echopraxia/echolalia,Verbigeration,Waxy flexibility | 94.2 | 91.9 | 96.2 |
| 5 | Mutism,Staring,Grimacing,Echopraxia/echolalia,Waxy flexibility | 94.2 | 91.9 | 96.2 |
| 5 | Mutism,Staring,Grimacing,Rigidity,Waxy flexibility | 94.2 | 91.9 | 96.2 |
| 5 | Mutism,Staring,Posturing/catalepsy,Echopraxia/echolalia,Waxy flexibility | 94.2 | 91.9 | 96.2 |
| 5 | Mutism,Staring,Posturing/catalepsy,Stereotypy,Waxy flexibility | 94.2 | 91.9 | 96.2 |
| 5 | Mutism,Staring,Posturing/catalepsy,Verbigeration,Waxy flexibility | 94.2 | 91.9 | 96.2 |
| 5 | Staring,Grimacing,Echopraxia/echolalia,Stereotypy,Rigidity | 94.2 | 91.9 | 96.2 |
| 5 | Staring,Mannerisms,Verbigeration,Negativism,Waxy flexibility | 94.2 | 91.9 | 96.2 |
| 5 | Staring,Posturing/catalepsy,Echopraxia/echolalia,Mannerisms,Rigidity | 94.2 | 91.9 | 96.2 |
| 5 | Staring,Posturing/catalepsy,Grimacing,Stereotypy,Negativism | 94.2 | 91.9 | 96.2 |
| 5 | Staring,Posturing/catalepsy,Grimacing,Stereotypy,Rigidity | 94.2 | 91.9 | 96.2 |
| 5 | Staring,Posturing/catalepsy,Grimacing,Verbigeration,Withdrawal | 94.2 | 91.9 | 96.2 |
| 5 | Staring,Posturing/catalepsy,Rigidity,Waxy flexibility,Withdrawal | 94.2 | 91.9 | 96.2 |
| 5 | Staring,Posturing/catalepsy,Stereotypy,Verbigeration,Rigidity | 94.2 | 91.9 | 96.2 |
| 5 | Staring,Stereotypy,Verbigeration,Negativism,Waxy flexibility | 94.2 | 91.9 | 96.2 |
| 5 | Staring,Verbigeration,Rigidity,Negativism,Waxy flexibility | 94.2 | 91.9 | 96.2 |
| 5 | Staring,Posturing/catalepsy,Echopraxia/echolalia,Stereotypy,Withdrawal | 94.1 | 91.9 | 96.2 |
| 5 | Excitement,Immobility/stupor,Echopraxia/echolalia,Mannerisms,Withdrawal | 94.0 | 91.7 | 96.0 |
| 5 | Excitement,Immobility/stupor,Mutism,Grimacing,Echopraxia/echolalia | 94.0 | 91.7 | 96.0 |
| 5 | Excitement,Immobility/stupor,Mutism,Mannerisms,Negativism | 94.0 | 91.7 | 96.2 |
| 5 | Excitement,Immobility/stupor,Mutism,Stereotypy,Verbigeration | 94.0 | 91.7 | 96.0 |
| 5 | Excitement,Immobility/stupor,Posturing/catalepsy,Grimacing,Waxy flexibility | 94.0 | 91.7 | 96.0 |
| 5 | Immobility/stupor,Grimacing,Rigidity,Negativism,Withdrawal | 94.0 | 91.7 | 96.0 |
| 5 | Immobility/stupor,Mannerisms,Verbigeration,Negativism,Withdrawal | 94.0 | 91.7 | 96.0 |
| 5 | Immobility/stupor,Mutism,Posturing/catalepsy,Mannerisms,Negativism | 94.0 | 91.7 | 96.0 |
| 5 | Immobility/stupor,Mutism,Rigidity,Negativism,Withdrawal | 94.0 | 91.7 | 96.0 |
| 5 | Immobility/stupor,Mutism,Staring,Grimacing,Mannerisms | 94.0 | 91.7 | 96.0 |
| 5 | Immobility/stupor,Mutism,Staring,Grimacing,Verbigeration | 94.0 | 91.7 | 96.0 |
| 5 | Immobility/stupor,Mutism,Staring,Stereotypy,Verbigeration | 94.0 | 91.7 | 96.0 |
| 5 | Immobility/stupor,Posturing/catalepsy,Mannerisms,Rigidity,Withdrawal | 94.0 | 91.7 | 96.0 |
| 5 | Immobility/stupor,Posturing/catalepsy,Stereotypy,Verbigeration,Negativism | 94.0 | 91.7 | 96.2 |
| 5 | Immobility/stupor,Staring,Echopraxia/echolalia,Stereotypy,Withdrawal | 94.0 | 91.7 | 96.0 |
| 5 | Staring,Echopraxia/echolalia,Mannerisms,Verbigeration,Negativism | 94.0 | 91.7 | 96.2 |
| 5 | Staring,Grimacing,Echopraxia/echolalia,Stereotypy,Negativism | 94.0 | 91.7 | 96.0 |
| 5 | Staring,Grimacing,Stereotypy,Rigidity,Waxy flexibility | 94.0 | 91.7 | 96.0 |
| 5 | Staring,Posturing/catalepsy,Grimacing,Rigidity,Negativism | 94.0 | 91.7 | 96.0 |
| 5 | Staring,Posturing/catalepsy,Stereotypy,Verbigeration,Withdrawal | 94.0 | 91.7 | 96.2 |
| 5 | Excitement,Immobility/stupor,Mannerisms,Verbigeration,Negativism | 93.9 | 91.7 | 96.0 |
| 5 | Excitement,Immobility/stupor,Mutism,Grimacing,Verbigeration | 93.9 | 91.7 | 96.0 |
| 5 | Excitement,Immobility/stupor,Posturing/catalepsy,Echopraxia/echolalia,Waxy flexibility | 93.9 | 91.7 | 96.0 |
| 5 | Excitement,Immobility/stupor,Stereotypy,Waxy flexibility,Withdrawal | 93.9 | 91.7 | 96.0 |
| 5 | Excitement,Immobility/stupor,Verbigeration,Waxy flexibility,Withdrawal | 93.9 | 91.7 | 96.2 |
| 5 | Excitement,Staring,Stereotypy,Verbigeration,Waxy flexibility | 93.9 | 91.7 | 96.0 |
| 5 | Immobility/stupor,Echopraxia/echolalia,Mannerisms,Negativism,Withdrawal | 93.9 | 91.7 | 96.0 |
| 5 | Immobility/stupor,Echopraxia/echolalia,Mannerisms,Rigidity,Withdrawal | 93.9 | 91.7 | 96.0 |
| 5 | Immobility/stupor,Mutism,Staring,Posturing/catalepsy,Echopraxia/echolalia | 93.9 | 91.7 | 96.0 |
| 5 | Immobility/stupor,Mutism,Staring,Posturing/catalepsy,Verbigeration | 93.9 | 91.7 | 96.0 |
| 5 | Immobility/stupor,Posturing/catalepsy,Stereotypy,Mannerisms,Negativism | 93.9 | 91.7 | 96.0 |
| 5 | Immobility/stupor,Staring,Grimacing,Echopraxia/echolalia,Stereotypy | 93.9 | 91.7 | 96.0 |
| 5 | Immobility/stupor,Staring,Stereotypy,Verbigeration,Withdrawal | 93.9 | 91.7 | 96.0 |
| 5 | Immobility/stupor,Staring,Verbigeration,Rigidity,Waxy flexibility | 93.9 | 91.7 | 96.0 |
| 5 | Immobility/stupor,Stereotypy,Mannerisms,Rigidity,Withdrawal | 93.9 | 91.7 | 96.2 |
| 5 | Staring,Echopraxia/echolalia,Stereotypy,Mannerisms,Rigidity | 93.9 | 91.7 | 96.0 |
| 5 | Staring,Echopraxia/echolalia,Stereotypy,Negativism,Waxy flexibility | 93.9 | 91.7 | 96.0 |
| 5 | Staring,Grimacing,Mannerisms,Negativism,Waxy flexibility | 93.9 | 91.7 | 96.0 |
| 5 | Staring,Grimacing,Stereotypy,Verbigeration,Negativism | 93.9 | 91.7 | 96.0 |
| 5 | Staring,Grimacing,Stereotypy,Waxy flexibility,Withdrawal | 93.9 | 91.5 | 96.0 |
| 5 | Staring,Mannerisms,Verbigeration,Waxy flexibility,Withdrawal | 93.9 | 91.5 | 96.0 |
| 5 | Staring,Posturing/catalepsy,Echopraxia/echolalia,Verbigeration,Negativism | 93.9 | 91.7 | 96.2 |
| 5 | Staring,Posturing/catalepsy,Grimacing,Echopraxia/echolalia,Negativism | 93.9 | 91.5 | 96.0 |
| 5 | Staring,Posturing/catalepsy,Grimacing,Verbigeration,Negativism | 93.9 | 91.5 | 96.0 |
| 5 | Staring,Posturing/catalepsy,Grimacing,Verbigeration,Rigidity | 93.9 | 91.7 | 96.0 |
| 5 | Staring,Posturing/catalepsy,Stereotypy,Negativism,Waxy flexibility | 93.9 | 91.7 | 96.0 |
| 5 | Staring,Stereotypy,Mannerisms,Rigidity,Waxy flexibility | 93.9 | 91.5 | 96.0 |
| 5 | Excitement,Immobility/stupor,Echopraxia/echolalia,Mannerisms,Negativism | 93.7 | 91.5 | 96.0 |
| 5 | Excitement,Immobility/stupor,Grimacing,Waxy flexibility,Withdrawal | 93.7 | 91.3 | 96.0 |
| 5 | Excitement,Immobility/stupor,Mannerisms,Verbigeration,Withdrawal | 93.7 | 91.5 | 95.7 |
| 5 | Excitement,Immobility/stupor,Mannerisms,Waxy flexibility,Withdrawal | 93.7 | 91.5 | 96.0 |
| 5 | Excitement,Immobility/stupor,Mutism,Grimacing,Stereotypy | 93.7 | 91.5 | 96.0 |
| 5 | Excitement,Immobility/stupor,Mutism,Mannerisms,Withdrawal | 93.7 | 91.3 | 96.0 |
| 5 | Excitement,Immobility/stupor,Posturing/catalepsy,Echopraxia/echolalia,Mannerisms | 93.7 | 91.5 | 96.0 |
| 5 | Excitement,Immobility/stupor,Posturing/catalepsy,Grimacing,Stereotypy | 93.7 | 91.5 | 96.0 |
| 5 | Excitement,Immobility/stupor,Posturing/catalepsy,Mannerisms,Verbigeration | 93.7 | 91.5 | 96.0 |
| 5 | Excitement,Immobility/stupor,Posturing/catalepsy,Verbigeration,Waxy flexibility | 93.7 | 91.5 | 96.0 |
| 5 | Excitement,Immobility/stupor,Stereotypy,Mannerisms,Negativism | 93.7 | 91.5 | 96.0 |
| 5 | Excitement,Mutism,Posturing/catalepsy,Rigidity,Negativism | 93.7 | 91.3 | 96.0 |
| 5 | Excitement,Mutism,Posturing/catalepsy,Rigidity,Withdrawal | 93.7 | 91.3 | 96.0 |
| 5 | Excitement,Mutism,Rigidity,Negativism,Withdrawal | 93.7 | 91.5 | 96.0 |
| 5 | Excitement,Staring,Echopraxia/echolalia,Verbigeration,Waxy flexibility | 93.7 | 91.5 | 96.0 |
| 5 | Excitement,Staring,Grimacing,Echopraxia/echolalia,Mannerisms | 93.7 | 91.5 | 96.0 |
| 5 | Excitement,Staring,Mannerisms,Verbigeration,Waxy flexibility | 93.7 | 91.5 | 95.7 |
| 5 | Immobility/stupor,Echopraxia/echolalia,Stereotypy,Mannerisms,Negativism | 93.7 | 91.5 | 96.0 |
| 5 | Immobility/stupor,Echopraxia/echolalia,Stereotypy,Rigidity,Withdrawal | 93.7 | 91.5 | 96.0 |
| 5 | Immobility/stupor,Grimacing,Echopraxia/echolalia,Rigidity,Negativism | 93.7 | 91.5 | 95.7 |
| 5 | Immobility/stupor,Grimacing,Stereotypy,Rigidity,Withdrawal | 93.7 | 91.5 | 96.0 |
| 5 | Immobility/stupor,Mutism,Echopraxia/echolalia,Rigidity,Negativism | 93.7 | 91.3 | 95.7 |
| 5 | Immobility/stupor,Mutism,Grimacing,Mannerisms,Rigidity | 93.7 | 91.3 | 96.0 |
| 5 | Immobility/stupor,Mutism,Staring,Echopraxia/echolalia,Withdrawal | 93.7 | 91.5 | 96.0 |
| 5 | Immobility/stupor,Mutism,Staring,Mannerisms,Waxy flexibility | 93.7 | 91.3 | 95.7 |
| 5 | Immobility/stupor,Mutism,Staring,Posturing/catalepsy,Stereotypy | 93.7 | 91.3 | 96.0 |
| 5 | Immobility/stupor,Mutism,Staring,Stereotypy,Withdrawal | 93.7 | 91.5 | 96.0 |
| 5 | Immobility/stupor,Mutism,Staring,Verbigeration,Withdrawal | 93.7 | 91.5 | 96.0 |
| 5 | Immobility/stupor,Posturing/catalepsy,Echopraxia/echolalia,Mannerisms,Negativism | 93.7 | 91.5 | 96.0 |
| 5 | Immobility/stupor,Posturing/catalepsy,Echopraxia/echolalia,Negativism,Withdrawal | 93.7 | 91.5 | 96.0 |
| 5 | Immobility/stupor,Posturing/catalepsy,Echopraxia/echolalia,Rigidity,Negativism | 93.7 | 91.3 | 95.7 |
| 5 | Immobility/stupor,Posturing/catalepsy,Grimacing,Mannerisms,Negativism | 93.7 | 91.5 | 96.0 |
| 5 | Immobility/stupor,Posturing/catalepsy,Grimacing,Rigidity,Negativism | 93.7 | 91.5 | 96.0 |
| 5 | Immobility/stupor,Posturing/catalepsy,Mannerisms,Verbigeration,Negativism | 93.7 | 91.5 | 96.0 |
| 5 | Immobility/stupor,Staring,Echopraxia/echolalia,Mannerisms,Verbigeration | 93.7 | 91.3 | 96.0 |
| 5 | Immobility/stupor,Staring,Grimacing,Echopraxia/echolalia,Withdrawal | 93.7 | 91.5 | 96.0 |
| 5 | Immobility/stupor,Staring,Grimacing,Mannerisms,Waxy flexibility | 93.7 | 91.5 | 95.7 |
| 5 | Immobility/stupor,Staring,Grimacing,Verbigeration,Withdrawal | 93.7 | 91.5 | 96.0 |
| 5 | Immobility/stupor,Staring,Mannerisms,Verbigeration,Waxy flexibility | 93.7 | 91.5 | 96.0 |
| 5 | Immobility/stupor,Staring,Posturing/catalepsy,Echopraxia/echolalia,Stereotypy | 93.7 | 91.5 | 95.7 |
| 5 | Immobility/stupor,Staring,Posturing/catalepsy,Grimacing,Verbigeration | 93.7 | 91.3 | 96.0 |
| 5 | Immobility/stupor,Staring,Posturing/catalepsy,Grimacing,Withdrawal | 93.7 | 91.5 | 96.0 |
| 5 | Immobility/stupor,Staring,Posturing/catalepsy,Verbigeration,Withdrawal | 93.7 | 91.5 | 96.0 |
| 5 | Immobility/stupor,Staring,Stereotypy,Mannerisms,Waxy flexibility | 93.7 | 91.5 | 95.7 |
| 5 | Immobility/stupor,Staring,Stereotypy,Verbigeration,Waxy flexibility | 93.7 | 91.5 | 95.7 |
| 5 | Immobility/stupor,Stereotypy,Mannerisms,Negativism,Waxy flexibility | 93.7 | 91.5 | 95.7 |
| 5 | Immobility/stupor,Stereotypy,Mannerisms,Verbigeration,Negativism | 93.7 | 91.5 | 96.0 |
| 5 | Mutism,Staring,Posturing/catalepsy,Rigidity,Waxy flexibility | 93.7 | 91.5 | 96.0 |
| 5 | Staring,Echopraxia/echolalia,Stereotypy,Verbigeration,Negativism | 93.7 | 91.5 | 96.0 |
| 5 | Staring,Echopraxia/echolalia,Stereotypy,Waxy flexibility,Withdrawal | 93.7 | 91.5 | 95.7 |
| 5 | Staring,Grimacing,Echopraxia/echolalia,Verbigeration,Rigidity | 93.7 | 91.5 | 96.0 |
| 5 | Staring,Grimacing,Rigidity,Negativism,Waxy flexibility | 93.7 | 91.5 | 96.0 |
| 5 | Staring,Grimacing,Verbigeration,Rigidity,Waxy flexibility | 93.7 | 91.5 | 96.0 |
| 5 | Staring,Stereotypy,Verbigeration,Rigidity,Waxy flexibility | 93.7 | 91.5 | 96.0 |
| 5 | Staring,Stereotypy,Verbigeration,Waxy flexibility,Withdrawal | 93.7 | 91.5 | 96.0 |
| 5 | Excitement,Immobility/stupor,Echopraxia/echolalia,Verbigeration,Waxy flexibility | 93.5 | 91.3 | 95.7 |
| 5 | Excitement,Immobility/stupor,Grimacing,Mannerisms,Negativism | 93.5 | 91.0 | 95.7 |
| 5 | Excitement,Immobility/stupor,Grimacing,Stereotypy,Withdrawal | 93.5 | 91.3 | 95.7 |
| 5 | Excitement,Immobility/stupor,Grimacing,Verbigeration,Withdrawal | 93.5 | 91.0 | 95.7 |
| 5 | Excitement,Immobility/stupor,Mutism,Mannerisms,Waxy flexibility | 93.5 | 91.0 | 95.7 |
| 5 | Excitement,Immobility/stupor,Posturing/catalepsy,Echopraxia/echolalia,Stereotypy | 93.5 | 91.0 | 95.7 |
| 5 | Excitement,Immobility/stupor,Posturing/catalepsy,Stereotypy,Verbigeration | 93.5 | 91.3 | 95.7 |
| 5 | Excitement,Staring,Echopraxia/echolalia,Mannerisms,Waxy flexibility | 93.5 | 91.3 | 95.7 |
| 5 | Excitement,Staring,Grimacing,Stereotypy,Verbigeration | 93.5 | 91.0 | 95.7 |
| 5 | Excitement,Staring,Stereotypy,Mannerisms,Waxy flexibility | 93.5 | 91.0 | 95.7 |
| 5 | Immobility/stupor,Grimacing,Stereotypy,Negativism,Withdrawal | 93.5 | 91.3 | 95.7 |
| 5 | Immobility/stupor,Mannerisms,Negativism,Waxy flexibility,Withdrawal | 93.5 | 91.0 | 95.7 |
| 5 | Immobility/stupor,Mannerisms,Verbigeration,Rigidity,Withdrawal | 93.5 | 91.0 | 95.7 |
| 5 | Immobility/stupor,Mutism,Echopraxia/echolalia,Mannerisms,Rigidity | 93.5 | 91.3 | 95.7 |
| 5 | Immobility/stupor,Mutism,Mannerisms,Negativism,Withdrawal | 93.5 | 91.3 | 95.7 |
| 5 | Immobility/stupor,Mutism,Mannerisms,Verbigeration,Rigidity | 93.5 | 91.3 | 95.7 |
| 5 | Immobility/stupor,Mutism,Posturing/catalepsy,Negativism,Withdrawal | 93.5 | 91.0 | 95.7 |
| 5 | Immobility/stupor,Mutism,Staring,Echopraxia/echolalia,Stereotypy | 93.5 | 91.3 | 95.7 |
| 5 | Immobility/stupor,Mutism,Staring,Echopraxia/echolalia,Verbigeration | 93.5 | 91.0 | 95.5 |
| 5 | Immobility/stupor,Mutism,Staring,Grimacing,Negativism | 93.5 | 91.0 | 95.7 |
| 5 | Immobility/stupor,Mutism,Staring,Grimacing,Rigidity | 93.5 | 91.3 | 95.7 |
| 5 | Immobility/stupor,Mutism,Staring,Grimacing,Stereotypy | 93.5 | 91.0 | 95.7 |
| 5 | Immobility/stupor,Mutism,Staring,Grimacing,Withdrawal | 93.5 | 91.3 | 95.7 |
| 5 | Immobility/stupor,Mutism,Staring,Negativism,Waxy flexibility | 93.5 | 91.3 | 95.7 |
| 5 | Immobility/stupor,Mutism,Staring,Posturing/catalepsy,Rigidity | 93.5 | 91.3 | 95.7 |
| 5 | Immobility/stupor,Mutism,Stereotypy,Mannerisms,Negativism | 93.5 | 91.3 | 95.7 |
| 5 | Immobility/stupor,Mutism,Stereotypy,Rigidity,Withdrawal | 93.5 | 91.0 | 95.7 |
| 5 | Immobility/stupor,Mutism,Verbigeration,Rigidity,Withdrawal | 93.5 | 91.0 | 95.7 |
| 5 | Immobility/stupor,Posturing/catalepsy,Grimacing,Negativism,Withdrawal | 93.5 | 91.0 | 95.7 |
| 5 | Immobility/stupor,Rigidity,Negativism,Waxy flexibility,Withdrawal | 93.5 | 91.0 | 95.7 |
| 5 | Immobility/stupor,Staring,Echopraxia/echolalia,Mannerisms,Waxy flexibility | 93.5 | 91.3 | 95.7 |
| 5 | Immobility/stupor,Staring,Grimacing,Echopraxia/echolalia,Verbigeration | 93.5 | 91.3 | 95.7 |
| 5 | Immobility/stupor,Staring,Grimacing,Negativism,Waxy flexibility | 93.5 | 91.3 | 95.7 |
| 5 | Immobility/stupor,Staring,Posturing/catalepsy,Grimacing,Rigidity | 93.5 | 91.3 | 95.7 |
| 5 | Immobility/stupor,Staring,Posturing/catalepsy,Grimacing,Stereotypy | 93.5 | 91.3 | 95.7 |
| 5 | Immobility/stupor,Staring,Posturing/catalepsy,Negativism,Waxy flexibility | 93.5 | 91.0 | 95.7 |
| 5 | Immobility/stupor,Staring,Stereotypy,Waxy flexibility,Withdrawal | 93.5 | 91.0 | 95.7 |
| 5 | Immobility/stupor,Stereotypy,Verbigeration,Rigidity,Withdrawal | 93.5 | 91.0 | 95.7 |
| 5 | Staring,Grimacing,Echopraxia/echolalia,Verbigeration,Withdrawal | 93.5 | 91.3 | 95.7 |
| 5 | Staring,Grimacing,Echopraxia/echolalia,Waxy flexibility,Withdrawal | 93.5 | 91.0 | 95.7 |
| 5 | Staring,Grimacing,Mannerisms,Verbigeration,Negativism | 93.5 | 91.0 | 95.7 |
| 5 | Staring,Grimacing,Verbigeration,Waxy flexibility,Withdrawal | 93.5 | 91.3 | 95.7 |
| 5 | Staring,Mannerisms,Verbigeration,Rigidity,Waxy flexibility | 93.5 | 91.0 | 95.7 |
| 5 | Staring,Posturing/catalepsy,Echopraxia/echolalia,Stereotypy,Rigidity | 93.5 | 91.3 | 95.7 |
| 5 | Staring,Posturing/catalepsy,Grimacing,Echopraxia/echolalia,Rigidity | 93.5 | 91.3 | 95.7 |
| 5 | Staring,Posturing/catalepsy,Grimacing,Stereotypy,Mannerisms | 93.5 | 91.0 | 95.7 |
| 5 | Staring,Posturing/catalepsy,Stereotypy,Waxy flexibility,Withdrawal | 93.5 | 91.0 | 95.7 |
| 5 | Excitement,Immobility/stupor,Echopraxia/echolalia,Stereotypy,Waxy flexibility | 93.3 | 90.8 | 95.5 |
| 5 | Excitement,Immobility/stupor,Stereotypy,Mannerisms,Withdrawal | 93.3 | 90.8 | 95.5 |
| 5 | Excitement,Mutism,Posturing/catalepsy,Negativism,Withdrawal | 93.3 | 90.8 | 95.5 |
| 5 | Excitement,Staring,Grimacing,Echopraxia/echolalia,Verbigeration | 93.3 | 90.8 | 95.5 |
| 5 | Excitement,Staring,Grimacing,Stereotypy,Mannerisms | 93.3 | 90.8 | 95.5 |
| 5 | Immobility/stupor,Grimacing,Echopraxia/echolalia,Rigidity,Withdrawal | 93.3 | 90.8 | 95.5 |
| 5 | Immobility/stupor,Grimacing,Stereotypy,Mannerisms,Negativism | 93.3 | 90.8 | 95.5 |
| 5 | Immobility/stupor,Mutism,Echopraxia/echolalia,Mannerisms,Negativism | 93.3 | 90.8 | 95.5 |
| 5 | Immobility/stupor,Mutism,Grimacing,Verbigeration,Rigidity | 93.3 | 90.8 | 95.5 |
| 5 | Immobility/stupor,Mutism,Posturing/catalepsy,Mannerisms,Rigidity | 93.3 | 90.8 | 95.5 |
| 5 | Immobility/stupor,Mutism,Posturing/catalepsy,Rigidity,Negativism | 93.3 | 90.8 | 95.5 |
| 5 | Immobility/stupor,Mutism,Posturing/catalepsy,Stereotypy,Negativism | 93.3 | 90.8 | 95.5 |
| 5 | Immobility/stupor,Mutism,Posturing/catalepsy,Verbigeration,Negativism | 93.3 | 90.8 | 95.5 |
| 5 | Immobility/stupor,Mutism,Staring,Grimacing,Echopraxia/echolalia | 93.3 | 90.8 | 95.5 |
| 5 | Immobility/stupor,Mutism,Staring,Verbigeration,Waxy flexibility | 93.3 | 90.8 | 95.5 |
| 5 | Immobility/stupor,Mutism,Staring,Waxy flexibility,Withdrawal | 93.3 | 90.8 | 95.5 |
| 5 | Immobility/stupor,Mutism,Stereotypy,Mannerisms,Rigidity | 93.3 | 91.0 | 95.5 |
| 5 | Immobility/stupor,Mutism,Stereotypy,Negativism,Withdrawal | 93.3 | 90.8 | 95.5 |
| 5 | Immobility/stupor,Mutism,Stereotypy,Verbigeration,Rigidity | 93.3 | 90.8 | 95.5 |
| 5 | Immobility/stupor,Posturing/catalepsy,Echopraxia/echolalia,Stereotypy,Negativism | 93.3 | 90.8 | 95.5 |
| 5 | Immobility/stupor,Posturing/catalepsy,Grimacing,Stereotypy,Negativism | 93.3 | 90.8 | 95.5 |
| 5 | Immobility/stupor,Posturing/catalepsy,Grimacing,Verbigeration,Negativism | 93.3 | 91.0 | 95.5 |
| 5 | Immobility/stupor,Staring,Echopraxia/echolalia,Rigidity,Waxy flexibility | 93.3 | 90.8 | 95.5 |
| 5 | Immobility/stupor,Staring,Echopraxia/echolalia,Stereotypy,Verbigeration | 93.3 | 90.8 | 95.5 |
| 5 | Immobility/stupor,Staring,Grimacing,Rigidity,Waxy flexibility | 93.3 | 90.8 | 95.5 |
| 5 | Immobility/stupor,Staring,Grimacing,Verbigeration,Waxy flexibility | 93.3 | 90.8 | 95.5 |
| 5 | Immobility/stupor,Staring,Posturing/catalepsy,Echopraxia/echolalia,Verbigeration | 93.3 | 90.8 | 95.5 |
| 5 | Immobility/stupor,Staring,Posturing/catalepsy,Grimacing,Echopraxia/echolalia | 93.3 | 90.8 | 95.5 |
| 5 | Immobility/stupor,Stereotypy,Negativism,Waxy flexibility,Withdrawal | 93.3 | 90.8 | 95.5 |
| 5 | Immobility/stupor,Verbigeration,Rigidity,Negativism,Waxy flexibility | 93.3 | 90.8 | 95.5 |
| 5 | Staring,Echopraxia/echolalia,Mannerisms,Rigidity,Waxy flexibility | 93.3 | 90.8 | 95.5 |
| 5 | Staring,Echopraxia/echolalia,Mannerisms,Verbigeration,Rigidity | 93.3 | 90.8 | 95.5 |
| 5 | Staring,Echopraxia/echolalia,Mannerisms,Verbigeration,Withdrawal | 93.3 | 90.8 | 95.5 |
| 5 | Staring,Echopraxia/echolalia,Stereotypy,Verbigeration,Rigidity | 93.3 | 90.8 | 95.5 |
| 5 | Staring,Echopraxia/echolalia,Verbigeration,Negativism,Waxy flexibility | 93.3 | 90.8 | 95.5 |
| 5 | Staring,Grimacing,Echopraxia/echolalia,Stereotypy,Mannerisms | 93.3 | 90.8 | 95.5 |
| 5 | Staring,Grimacing,Echopraxia/echolalia,Verbigeration,Negativism | 93.3 | 90.8 | 95.5 |
| 5 | Staring,Grimacing,Stereotypy,Negativism,Waxy flexibility | 93.3 | 90.8 | 95.5 |
| 5 | Staring,Posturing/catalepsy,Echopraxia/echolalia,Negativism,Waxy flexibility | 93.3 | 90.8 | 95.5 |
| 5 | Staring,Posturing/catalepsy,Echopraxia/echolalia,Verbigeration,Rigidity | 93.3 | 90.8 | 95.5 |
| 5 | Staring,Posturing/catalepsy,Echopraxia/echolalia,Verbigeration,Withdrawal | 93.3 | 90.8 | 95.5 |
| 5 | Staring,Posturing/catalepsy,Echopraxia/echolalia,Waxy flexibility,Withdrawal | 93.3 | 90.8 | 95.5 |
| 5 | Staring,Posturing/catalepsy,Grimacing,Echopraxia/echolalia,Mannerisms | 93.3 | 90.8 | 95.5 |
| 5 | Staring,Posturing/catalepsy,Rigidity,Negativism,Waxy flexibility | 93.3 | 90.8 | 95.5 |
| 5 | Excitement,Immobility/stupor,Grimacing,Mannerisms,Withdrawal | 93.1 | 90.6 | 95.3 |
| 5 | Excitement,Immobility/stupor,Stereotypy,Verbigeration,Waxy flexibility | 93.1 | 90.6 | 95.3 |
| 5 | Excitement,Staring,Echopraxia/echolalia,Stereotypy,Mannerisms | 93.1 | 90.6 | 95.3 |
| 5 | Excitement,Staring,Grimacing,Mannerisms,Verbigeration | 93.1 | 90.6 | 95.3 |
| 5 | Immobility/stupor,Echopraxia/echolalia,Verbigeration,Negativism,Withdrawal | 93.1 | 90.6 | 95.3 |
| 5 | Immobility/stupor,Grimacing,Mannerisms,Negativism,Withdrawal | 93.1 | 90.6 | 95.3 |
| 5 | Immobility/stupor,Mannerisms,Rigidity,Waxy flexibility,Withdrawal | 93.1 | 90.6 | 95.3 |
| 5 | Immobility/stupor,Mutism,Echopraxia/echolalia,Rigidity,Withdrawal | 93.1 | 90.6 | 95.3 |
| 5 | Immobility/stupor,Mutism,Stereotypy,Verbigeration,Negativism | 93.1 | 90.6 | 95.3 |
| 5 | Immobility/stupor,Posturing/catalepsy,Grimacing,Mannerisms,Rigidity | 93.1 | 90.6 | 95.3 |
| 5 | Immobility/stupor,Staring,Echopraxia/echolalia,Waxy flexibility,Withdrawal | 93.1 | 90.6 | 95.3 |
| 5 | Immobility/stupor,Staring,Grimacing,Stereotypy,Waxy flexibility | 93.1 | 90.6 | 95.3 |
| 5 | Immobility/stupor,Staring,Verbigeration,Waxy flexibility,Withdrawal | 93.1 | 90.6 | 95.3 |
| 5 | Staring,Grimacing,Echopraxia/echolalia,Rigidity,Waxy flexibility | 93.1 | 90.6 | 95.3 |
| 5 | Staring,Grimacing,Stereotypy,Mannerisms,Waxy flexibility | 93.1 | 90.6 | 95.3 |
| 5 | Staring,Grimacing,Verbigeration,Negativism,Waxy flexibility | 93.1 | 90.6 | 95.3 |
| 5 | Staring,Posturing/catalepsy,Grimacing,Mannerisms,Verbigeration | 93.1 | 90.6 | 95.3 |
| 5 | Staring,Posturing/catalepsy,Grimacing,Waxy flexibility,Withdrawal | 93.1 | 90.6 | 95.3 |
| 5 | Staring,Posturing/catalepsy,Mannerisms,Rigidity,Waxy flexibility | 93.1 | 90.6 | 95.3 |
| 5 | Excitement,Immobility/stupor,Grimacing,Mannerisms,Waxy flexibility | 93.0 | 90.6 | 95.3 |
| 5 | Excitement,Immobility/stupor,Grimacing,Stereotypy,Waxy flexibility | 93.0 | 90.6 | 95.3 |
| 5 | Excitement,Immobility/stupor,Grimacing,Verbigeration,Waxy flexibility | 93.0 | 90.6 | 95.3 |
| 5 | Excitement,Immobility/stupor,Mutism,Echopraxia/echolalia,Mannerisms | 93.0 | 90.6 | 95.3 |
| 5 | Excitement,Immobility/stupor,Mutism,Mannerisms,Verbigeration | 93.0 | 90.6 | 95.3 |
| 5 | Excitement,Mutism,Rigidity,Waxy flexibility,Withdrawal | 93.0 | 90.6 | 95.3 |
| 5 | Immobility/stupor,Grimacing,Echopraxia/echolalia,Mannerisms,Rigidity | 93.0 | 90.6 | 95.3 |
| 5 | Immobility/stupor,Grimacing,Rigidity,Negativism,Waxy flexibility | 93.0 | 90.6 | 95.3 |
| 5 | Immobility/stupor,Grimacing,Stereotypy,Mannerisms,Rigidity | 93.0 | 90.6 | 95.3 |
| 5 | Immobility/stupor,Grimacing,Stereotypy,Verbigeration,Negativism | 93.0 | 90.6 | 95.3 |
| 5 | Immobility/stupor,Grimacing,Stereotypy,Verbigeration,Rigidity | 93.0 | 90.6 | 95.3 |
| 5 | Immobility/stupor,Grimacing,Verbigeration,Rigidity,Withdrawal | 93.0 | 90.6 | 95.3 |
| 5 | Immobility/stupor,Mutism,Mannerisms,Negativism,Waxy flexibility | 93.0 | 90.6 | 95.3 |
| 5 | Immobility/stupor,Mutism,Staring,Rigidity,Waxy flexibility | 93.0 | 90.6 | 95.3 |
| 5 | Immobility/stupor,Mutism,Staring,Stereotypy,Waxy flexibility | 93.0 | 90.6 | 95.3 |
| 5 | Immobility/stupor,Posturing/catalepsy,Stereotypy,Rigidity,Withdrawal | 93.0 | 90.6 | 95.3 |
| 5 | Immobility/stupor,Staring,Echopraxia/echolalia,Stereotypy,Waxy flexibility | 93.0 | 90.6 | 95.3 |
| 5 | Immobility/stupor,Staring,Echopraxia/echolalia,Verbigeration,Withdrawal | 93.0 | 90.6 | 95.3 |
| 5 | Mutism,Mannerisms,Rigidity,Negativism,Withdrawal | 93.0 | 90.6 | 95.3 |
| 5 | Staring,Echopraxia/echolalia,Stereotypy,Verbigeration,Withdrawal | 93.0 | 90.6 | 95.3 |
| 5 | Staring,Posturing/catalepsy,Grimacing,Stereotypy,Verbigeration | 93.0 | 90.6 | 95.3 |
| 5 | Staring,Posturing/catalepsy,Verbigeration,Waxy flexibility,Withdrawal | 93.0 | 90.6 | 95.3 |
| 5 | Excitement,Immobility/stupor,Echopraxia/echolalia,Stereotypy,Verbigeration | 92.8 | 90.4 | 95.1 |
| 5 | Excitement,Immobility/stupor,Grimacing,Echopraxia/echolalia,Stereotypy | 92.8 | 90.4 | 95.1 |
| 5 | Excitement,Immobility/stupor,Grimacing,Echopraxia/echolalia,Verbigeration | 92.8 | 90.4 | 95.1 |
| 5 | Excitement,Immobility/stupor,Mutism,Grimacing,Mannerisms | 92.8 | 90.4 | 95.1 |
| 5 | Excitement,Immobility/stupor,Mutism,Stereotypy,Mannerisms | 92.8 | 90.4 | 95.1 |
| 5 | Excitement,Immobility/stupor,Posturing/catalepsy,Mannerisms,Waxy flexibility | 92.8 | 90.4 | 95.1 |
| 5 | Excitement,Immobility/stupor,Posturing/catalepsy,Stereotypy,Waxy flexibility | 92.8 | 90.4 | 95.1 |
| 5 | Excitement,Mutism,Posturing/catalepsy,Echopraxia/echolalia,Rigidity | 92.8 | 90.4 | 95.1 |
| 5 | Excitement,Staring,Echopraxia/echolalia,Stereotypy,Verbigeration | 92.8 | 90.4 | 95.1 |
| 5 | Immobility/stupor,Echopraxia/echolalia,Rigidity,Negativism,Waxy flexibility | 92.8 | 90.4 | 95.1 |
| 5 | Immobility/stupor,Echopraxia/echolalia,Stereotypy,Negativism,Waxy flexibility | 92.8 | 90.4 | 95.1 |
| 5 | Immobility/stupor,Echopraxia/echolalia,Stereotypy,Verbigeration,Negativism | 92.8 | 90.4 | 95.1 |
| 5 | Immobility/stupor,Echopraxia/echolalia,Verbigeration,Rigidity,Withdrawal | 92.8 | 90.4 | 95.1 |
| 5 | Immobility/stupor,Grimacing,Echopraxia/echolalia,Stereotypy,Negativism | 92.8 | 90.4 | 95.1 |
| 5 | Immobility/stupor,Mutism,Grimacing,Rigidity,Negativism | 92.8 | 90.4 | 95.1 |
| 5 | Immobility/stupor,Mutism,Grimacing,Rigidity,Withdrawal | 92.8 | 90.4 | 95.1 |
| 5 | Immobility/stupor,Mutism,Grimacing,Stereotypy,Rigidity | 92.8 | 90.4 | 95.1 |
| 5 | Immobility/stupor,Mutism,Mannerisms,Verbigeration,Negativism | 92.8 | 90.4 | 95.1 |
| 5 | Immobility/stupor,Mutism,Posturing/catalepsy,Echopraxia/echolalia,Negativism | 92.8 | 90.4 | 95.1 |
| 5 | Immobility/stupor,Mutism,Rigidity,Negativism,Waxy flexibility | 92.8 | 90.4 | 95.1 |
| 5 | Immobility/stupor,Mutism,Staring,Echopraxia/echolalia,Waxy flexibility | 92.8 | 90.4 | 95.1 |
| 5 | Immobility/stupor,Mutism,Verbigeration,Negativism,Withdrawal | 92.8 | 90.4 | 95.1 |
| 5 | Immobility/stupor,Posturing/catalepsy,Echopraxia/echolalia,Verbigeration,Negativism | 92.8 | 90.4 | 95.1 |
| 5 | Immobility/stupor,Posturing/catalepsy,Grimacing,Rigidity,Withdrawal | 92.8 | 90.4 | 95.1 |
| 5 | Immobility/stupor,Posturing/catalepsy,Mannerisms,Negativism,Waxy flexibility | 92.8 | 90.4 | 95.1 |
| 5 | Immobility/stupor,Posturing/catalepsy,Stereotypy,Negativism,Waxy flexibility | 92.8 | 90.4 | 95.1 |
| 5 | Immobility/stupor,Staring,Posturing/catalepsy,Mannerisms,Waxy flexibility | 92.8 | 90.4 | 95.1 |
| 5 | Immobility/stupor,Staring,Posturing/catalepsy,Waxy flexibility,Withdrawal | 92.8 | 90.4 | 95.1 |
| 5 | Immobility/stupor,Stereotypy,Verbigeration,Negativism,Waxy flexibility | 92.8 | 90.4 | 95.1 |
| 5 | Mutism,Staring,Posturing/catalepsy,Grimacing,Waxy flexibility | 92.8 | 90.4 | 95.1 |
| 5 | Staring,Grimacing,Echopraxia/echolalia,Mannerisms,Waxy flexibility | 92.8 | 90.4 | 95.1 |
| 5 | Staring,Grimacing,Echopraxia/echolalia,Negativism,Waxy flexibility | 92.8 | 90.4 | 95.1 |
| 5 | Staring,Posturing/catalepsy,Grimacing,Echopraxia/echolalia,Stereotypy | 92.8 | 90.4 | 95.1 |
| 5 | Staring,Posturing/catalepsy,Verbigeration,Negativism,Waxy flexibility | 92.8 | 90.4 | 95.1 |
| 5 | Excitement,Immobility/stupor,Posturing/catalepsy,Stereotypy,Mannerisms | 92.6 | 90.1 | 94.8 |
| 5 | Excitement,Mutism,Posturing/catalepsy,Verbigeration,Rigidity | 92.6 | 89.9 | 94.8 |
| 5 | Excitement,Staring,Stereotypy,Mannerisms,Verbigeration | 92.6 | 90.1 | 95.1 |
| 5 | Immobility/stupor,Echopraxia/echolalia,Mannerisms,Negativism,Waxy flexibility | 92.6 | 90.1 | 94.8 |
| 5 | Immobility/stupor,Echopraxia/echolalia,Mannerisms,Verbigeration,Negativism | 92.6 | 90.1 | 94.8 |
| 5 | Immobility/stupor,Grimacing,Echopraxia/echolalia,Mannerisms,Negativism | 92.6 | 90.1 | 94.8 |
| 5 | Immobility/stupor,Grimacing,Mannerisms,Verbigeration,Rigidity | 92.6 | 90.1 | 94.8 |
| 5 | Immobility/stupor,Grimacing,Verbigeration,Negativism,Withdrawal | 92.6 | 90.1 | 94.8 |
| 5 | Immobility/stupor,Mutism,Grimacing,Mannerisms,Negativism | 92.6 | 90.1 | 94.8 |
| 5 | Immobility/stupor,Mutism,Mannerisms,Rigidity,Waxy flexibility | 92.6 | 90.1 | 94.8 |
| 5 | Immobility/stupor,Mutism,Posturing/catalepsy,Mannerisms,Withdrawal | 92.6 | 90.1 | 95.1 |
| 5 | Immobility/stupor,Mutism,Staring,Posturing/catalepsy,Grimacing | 92.6 | 90.1 | 94.8 |
| 5 | Immobility/stupor,Posturing/catalepsy,Echopraxia/echolalia,Rigidity,Withdrawal | 92.6 | 90.1 | 94.8 |
| 5 | Immobility/stupor,Posturing/catalepsy,Grimacing,Echopraxia/echolalia,Negativism | 92.6 | 90.1 | 95.1 |
| 5 | Immobility/stupor,Posturing/catalepsy,Rigidity,Negativism,Waxy flexibility | 92.6 | 90.1 | 94.8 |
| 5 | Immobility/stupor,Posturing/catalepsy,Verbigeration,Rigidity,Withdrawal | 92.6 | 90.1 | 94.8 |
| 5 | Immobility/stupor,Staring,Echopraxia/echolalia,Verbigeration,Waxy flexibility | 92.6 | 90.1 | 95.1 |
| 5 | Immobility/stupor,Staring,Grimacing,Echopraxia/echolalia,Waxy flexibility | 92.6 | 90.1 | 94.8 |
| 5 | Immobility/stupor,Staring,Grimacing,Waxy flexibility,Withdrawal | 92.6 | 90.1 | 95.1 |
| 5 | Immobility/stupor,Stereotypy,Rigidity,Waxy flexibility,Withdrawal | 92.6 | 90.1 | 94.8 |
| 5 | Mutism,Posturing/catalepsy,Mannerisms,Rigidity,Negativism | 92.6 | 90.1 | 94.8 |
| 5 | Mutism,Stereotypy,Rigidity,Negativism,Withdrawal | 92.6 | 90.1 | 94.8 |
| 5 | Staring,Echopraxia/echolalia,Stereotypy,Rigidity,Waxy flexibility | 92.6 | 90.1 | 94.8 |
| 5 | Staring,Echopraxia/echolalia,Verbigeration,Waxy flexibility,Withdrawal | 92.6 | 90.1 | 94.8 |
| 5 | Staring,Grimacing,Stereotypy,Mannerisms,Verbigeration | 92.6 | 90.1 | 94.8 |
| 5 | Staring,Grimacing,Stereotypy,Verbigeration,Waxy flexibility | 92.6 | 90.1 | 94.8 |
| 5 | Staring,Posturing/catalepsy,Echopraxia/echolalia,Stereotypy,Mannerisms | 92.6 | 90.1 | 94.8 |
| 5 | Staring,Posturing/catalepsy,Stereotypy,Mannerisms,Verbigeration | 92.6 | 90.1 | 94.8 |
| 5 | Excitement,Immobility/stupor,Echopraxia/echolalia,Mannerisms,Waxy flexibility | 92.4 | 89.9 | 94.8 |
| 5 | Excitement,Immobility/stupor,Grimacing,Echopraxia/echolalia,Mannerisms | 92.4 | 89.9 | 94.6 |
| 5 | Excitement,Immobility/stupor,Grimacing,Stereotypy,Verbigeration | 92.4 | 89.9 | 94.8 |
| 5 | Excitement,Immobility/stupor,Mannerisms,Verbigeration,Waxy flexibility | 92.4 | 89.9 | 94.8 |
| 5 | Excitement,Mutism,Echopraxia/echolalia,Rigidity,Negativism | 92.4 | 89.9 | 94.6 |
| 5 | Excitement,Mutism,Stereotypy,Rigidity,Negativism | 92.4 | 89.7 | 94.8 |
| 5 | Immobility/stupor,Echopraxia/echolalia,Negativism,Waxy flexibility,Withdrawal | 92.4 | 89.9 | 94.6 |
| 5 | Immobility/stupor,Grimacing,Echopraxia/echolalia,Stereotypy,Rigidity | 92.4 | 89.9 | 94.6 |
| 5 | Immobility/stupor,Grimacing,Echopraxia/echolalia,Verbigeration,Negativism | 92.4 | 89.9 | 94.8 |
| 5 | Immobility/stupor,Grimacing,Mannerisms,Rigidity,Waxy flexibility | 92.4 | 89.7 | 94.6 |
| 5 | Immobility/stupor,Grimacing,Mannerisms,Verbigeration,Negativism | 92.4 | 89.9 | 94.6 |
| 5 | Immobility/stupor,Grimacing,Stereotypy,Negativism,Waxy flexibility | 92.4 | 89.9 | 94.8 |
| 5 | Immobility/stupor,Mutism,Echopraxia/echolalia,Negativism,Withdrawal | 92.4 | 89.9 | 94.8 |
| 5 | Immobility/stupor,Mutism,Echopraxia/echolalia,Stereotypy,Negativism | 92.4 | 89.9 | 94.6 |
| 5 | Immobility/stupor,Mutism,Echopraxia/echolalia,Stereotypy,Rigidity | 92.4 | 89.9 | 94.6 |
| 5 | Immobility/stupor,Mutism,Echopraxia/echolalia,Verbigeration,Rigidity | 92.4 | 89.9 | 94.8 |
| 5 | Immobility/stupor,Mutism,Posturing/catalepsy,Grimacing,Negativism | 92.4 | 89.7 | 94.8 |
| 5 | Immobility/stupor,Mutism,Posturing/catalepsy,Stereotypy,Rigidity | 92.4 | 89.9 | 94.8 |
| 5 | Immobility/stupor,Mutism,Stereotypy,Negativism,Waxy flexibility | 92.4 | 89.9 | 94.8 |
| 5 | Immobility/stupor,Posturing/catalepsy,Grimacing,Mannerisms,Withdrawal | 92.4 | 89.7 | 94.6 |
| 5 | Immobility/stupor,Posturing/catalepsy,Grimacing,Stereotypy,Rigidity | 92.4 | 89.9 | 94.6 |
| 5 | Immobility/stupor,Posturing/catalepsy,Grimacing,Verbigeration,Rigidity | 92.4 | 89.7 | 94.8 |
| 5 | Immobility/stupor,Posturing/catalepsy,Negativism,Waxy flexibility,Withdrawal | 92.4 | 89.9 | 94.6 |
| 5 | Immobility/stupor,Staring,Posturing/catalepsy,Stereotypy,Waxy flexibility | 92.4 | 89.9 | 94.8 |
| 5 | Immobility/stupor,Stereotypy,Mannerisms,Verbigeration,Rigidity | 92.4 | 89.9 | 94.8 |
| 5 | Immobility/stupor,Verbigeration,Negativism,Waxy flexibility,Withdrawal | 92.4 | 89.9 | 94.8 |
| 5 | Mutism,Verbigeration,Rigidity,Negativism,Withdrawal | 92.4 | 89.9 | 94.6 |
| 5 | Staring,Grimacing,Echopraxia/echolalia,Stereotypy,Waxy flexibility | 92.4 | 89.9 | 94.8 |
| 5 | Staring,Grimacing,Mannerisms,Verbigeration,Waxy flexibility | 92.4 | 89.9 | 94.8 |
| 5 | Staring,Posturing/catalepsy,Grimacing,Negativism,Waxy flexibility | 92.4 | 89.9 | 94.8 |
| 5 | Staring,Posturing/catalepsy,Stereotypy,Rigidity,Waxy flexibility | 92.4 | 89.9 | 94.8 |
| 5 | Immobility/stupor,Mutism,Posturing/catalepsy,Verbigeration,Rigidity | 92.3 | 89.9 | 94.6 |
| 5 | Excitement,Immobility/stupor,Echopraxia/echolalia,Mannerisms,Verbigeration | 92.2 | 89.5 | 94.6 |
| 5 | Excitement,Mutism,Posturing/catalepsy,Grimacing,Rigidity | 92.2 | 89.5 | 94.6 |
| 5 | Excitement,Mutism,Posturing/catalepsy,Rigidity,Waxy flexibility | 92.2 | 89.7 | 94.6 |
| 5 | Excitement,Mutism,Verbigeration,Rigidity,Negativism | 92.2 | 89.7 | 94.6 |
| 5 | Excitement,Posturing/catalepsy,Rigidity,Negativism,Withdrawal | 92.2 | 89.5 | 94.6 |
| 5 | Excitement,Staring,Echopraxia/echolalia,Mannerisms,Verbigeration | 92.2 | 89.7 | 94.6 |
| 5 | Immobility/stupor,Grimacing,Echopraxia/echolalia,Negativism,Withdrawal | 92.2 | 89.7 | 94.4 |
| 5 | Immobility/stupor,Grimacing,Echopraxia/echolalia,Verbigeration,Rigidity | 92.2 | 89.7 | 94.6 |
| 5 | Immobility/stupor,Grimacing,Mannerisms,Negativism,Waxy flexibility | 92.2 | 89.7 | 94.6 |
| 5 | Immobility/stupor,Mannerisms,Verbigeration,Negativism,Waxy flexibility | 92.2 | 89.5 | 94.6 |
| 5 | Immobility/stupor,Mutism,Echopraxia/echolalia,Verbigeration,Negativism | 92.2 | 89.5 | 94.4 |
| 5 | Immobility/stupor,Mutism,Grimacing,Stereotypy,Negativism | 92.2 | 89.5 | 94.6 |
| 5 | Immobility/stupor,Mutism,Grimacing,Verbigeration,Negativism | 92.2 | 89.5 | 94.6 |
| 5 | Immobility/stupor,Mutism,Posturing/catalepsy,Grimacing,Withdrawal | 92.2 | 89.7 | 94.6 |
| 5 | Immobility/stupor,Posturing/catalepsy,Echopraxia/echolalia,Mannerisms,Rigidity | 92.2 | 89.7 | 94.6 |
| 5 | Immobility/stupor,Posturing/catalepsy,Mannerisms,Verbigeration,Rigidity | 92.2 | 89.7 | 94.6 |
| 5 | Immobility/stupor,Posturing/catalepsy,Stereotypy,Verbigeration,Rigidity | 92.2 | 89.7 | 94.6 |
| 5 | Immobility/stupor,Staring,Posturing/catalepsy,Verbigeration,Waxy flexibility | 92.2 | 89.7 | 94.6 |
| 5 | Staring,Echopraxia/echolalia,Stereotypy,Mannerisms,Waxy flexibility | 92.2 | 89.5 | 94.6 |
| 5 | Staring,Grimacing,Echopraxia/echolalia,Stereotypy,Verbigeration | 92.2 | 89.7 | 94.6 |
| 5 | Excitement,Immobility/stupor,Grimacing,Mannerisms,Verbigeration | 92.1 | 89.5 | 94.6 |
| 5 | Immobility/stupor,Echopraxia/echolalia,Stereotypy,Mannerisms,Rigidity | 92.1 | 89.5 | 94.6 |
| 5 | Immobility/stupor,Grimacing,Echopraxia/echolalia,Stereotypy,Withdrawal | 92.1 | 89.5 | 94.6 |
| 5 | Immobility/stupor,Mutism,Posturing/catalepsy,Rigidity,Withdrawal | 92.1 | 89.7 | 94.6 |
| 5 | Immobility/stupor,Posturing/catalepsy,Stereotypy,Mannerisms,Rigidity | 92.1 | 89.7 | 94.4 |
| 5 | Mutism,Posturing/catalepsy,Rigidity,Negativism,Withdrawal | 92.1 | 89.7 | 94.6 |
| 5 | Staring,Echopraxia/echolalia,Verbigeration,Rigidity,Waxy flexibility | 92.1 | 89.7 | 94.6 |
| 5 | Staring,Posturing/catalepsy,Grimacing,Echopraxia/echolalia,Verbigeration | 92.1 | 89.5 | 94.6 |
| 5 | Immobility/stupor,Mutism,Staring,Grimacing,Waxy flexibility | 92.0 | 89.5 | 94.4 |
| 5 | Immobility/stupor,Staring,Posturing/catalepsy,Rigidity,Waxy flexibility | 92.0 | 89.5 | 94.4 |
| 5 | Excitement,Immobility/stupor,Echopraxia/echolalia,Stereotypy,Mannerisms | 91.9 | 89.5 | 94.4 |
| 5 | Excitement,Immobility/stupor,Stereotypy,Mannerisms,Verbigeration | 91.9 | 89.2 | 94.4 |
| 5 | Excitement,Immobility/stupor,Stereotypy,Mannerisms,Waxy flexibility | 91.9 | 89.2 | 94.4 |
| 5 | Excitement,Mutism,Echopraxia/echolalia,Rigidity,Withdrawal | 91.9 | 89.5 | 94.4 |
| 5 | Excitement,Mutism,Grimacing,Rigidity,Negativism | 91.9 | 89.2 | 94.4 |
| 5 | Excitement,Mutism,Grimacing,Rigidity,Withdrawal | 91.9 | 89.2 | 94.4 |
| 5 | Excitement,Mutism,Posturing/catalepsy,Mannerisms,Rigidity | 91.9 | 89.2 | 94.4 |
| 5 | Excitement,Mutism,Rigidity,Negativism,Waxy flexibility | 91.9 | 89.5 | 94.4 |
| 5 | Excitement,Mutism,Stereotypy,Rigidity,Withdrawal | 91.9 | 89.2 | 94.4 |
| 5 | Immobility/stupor,Echopraxia/echolalia,Rigidity,Waxy flexibility,Withdrawal | 91.9 | 89.2 | 94.4 |
| 5 | Immobility/stupor,Echopraxia/echolalia,Stereotypy,Verbigeration,Rigidity | 91.9 | 89.2 | 94.4 |
| 5 | Immobility/stupor,Grimacing,Stereotypy,Rigidity,Waxy flexibility | 91.9 | 89.5 | 94.4 |
| 5 | Immobility/stupor,Mutism,Echopraxia/echolalia,Mannerisms,Withdrawal | 91.9 | 89.5 | 94.4 |
| 5 | Immobility/stupor,Mutism,Grimacing,Echopraxia/echolalia,Rigidity | 91.9 | 89.2 | 94.4 |
| 5 | Immobility/stupor,Mutism,Grimacing,Mannerisms,Withdrawal | 91.9 | 89.2 | 94.4 |
| 5 | Immobility/stupor,Mutism,Negativism,Waxy flexibility,Withdrawal | 91.9 | 89.2 | 94.4 |
| 5 | Immobility/stupor,Mutism,Posturing/catalepsy,Echopraxia/echolalia,Rigidity | 91.9 | 89.2 | 94.4 |
| 5 | Immobility/stupor,Mutism,Posturing/catalepsy,Echopraxia/echolalia,Withdrawal | 91.9 | 89.2 | 94.4 |
| 5 | Immobility/stupor,Posturing/catalepsy,Echopraxia/echolalia,Mannerisms,Withdrawal | 91.9 | 89.2 | 94.4 |
| 5 | Immobility/stupor,Posturing/catalepsy,Grimacing,Stereotypy,Withdrawal | 91.9 | 89.5 | 94.4 |
| 5 | Immobility/stupor,Posturing/catalepsy,Verbigeration,Negativism,Waxy flexibility | 91.9 | 89.2 | 94.4 |
| 5 | Immobility/stupor,Staring,Posturing/catalepsy,Echopraxia/echolalia,Waxy flexibility | 91.9 | 89.2 | 94.4 |
| 5 | Immobility/stupor,Verbigeration,Rigidity,Waxy flexibility,Withdrawal | 91.9 | 89.2 | 94.4 |
| 5 | Mutism,Posturing/catalepsy,Mannerisms,Negativism,Withdrawal | 91.9 | 89.2 | 94.4 |
| 5 | Mutism,Posturing/catalepsy,Verbigeration,Rigidity,Negativism | 91.9 | 89.5 | 94.4 |
| 5 | Staring,Grimacing,Echopraxia/echolalia,Mannerisms,Verbigeration | 91.9 | 89.2 | 94.4 |
| 5 | Staring,Posturing/catalepsy,Echopraxia/echolalia,Mannerisms,Verbigeration | 91.9 | 89.2 | 94.4 |
| 5 | Staring,Posturing/catalepsy,Grimacing,Mannerisms,Waxy flexibility | 91.9 | 89.2 | 94.4 |
| 5 | Staring,Posturing/catalepsy,Verbigeration,Rigidity,Waxy flexibility | 91.9 | 89.2 | 94.4 |
| 5 | Staring,Stereotypy,Mannerisms,Verbigeration,Waxy flexibility | 91.9 | 89.2 | 94.4 |
| 5 | Immobility/stupor,Mutism,Rigidity,Waxy flexibility,Withdrawal | 91.8 | 89.0 | 94.2 |
| 5 | Excitement,Mutism,Posturing/catalepsy,Echopraxia/echolalia,Negativism | 91.7 | 89.0 | 94.2 |
| 5 | Excitement,Mutism,Posturing/catalepsy,Stereotypy,Rigidity | 91.7 | 89.0 | 94.2 |
| 5 | Excitement,Mutism,Posturing/catalepsy,Waxy flexibility,Withdrawal | 91.7 | 89.0 | 94.2 |
| 5 | Excitement,Mutism,Verbigeration,Rigidity,Withdrawal | 91.7 | 89.0 | 94.2 |
| 5 | Immobility/stupor,Echopraxia/echolalia,Mannerisms,Verbigeration,Rigidity | 91.7 | 89.0 | 94.2 |
| 5 | Immobility/stupor,Echopraxia/echolalia,Verbigeration,Negativism,Waxy flexibility | 91.7 | 89.0 | 94.2 |
| 5 | Immobility/stupor,Grimacing,Echopraxia/echolalia,Negativism,Waxy flexibility | 91.7 | 89.0 | 94.2 |
| 5 | Immobility/stupor,Grimacing,Rigidity,Waxy flexibility,Withdrawal | 91.7 | 89.0 | 94.2 |
| 5 | Immobility/stupor,Mutism,Echopraxia/echolalia,Negativism,Waxy flexibility | 91.7 | 89.0 | 94.2 |
| 5 | Immobility/stupor,Mutism,Grimacing,Negativism,Withdrawal | 91.7 | 89.0 | 94.2 |
| 5 | Immobility/stupor,Mutism,Posturing/catalepsy,Grimacing,Mannerisms | 91.7 | 89.0 | 94.2 |
| 5 | Immobility/stupor,Mutism,Posturing/catalepsy,Grimacing,Verbigeration | 91.7 | 89.0 | 94.2 |
| 5 | Immobility/stupor,Mutism,Posturing/catalepsy,Verbigeration,Withdrawal | 91.7 | 89.2 | 94.2 |
| 5 | Immobility/stupor,Mutism,Staring,Posturing/catalepsy,Waxy flexibility | 91.7 | 89.0 | 94.2 |
| 5 | Immobility/stupor,Mutism,Verbigeration,Negativism,Waxy flexibility | 91.7 | 89.0 | 94.2 |
| 5 | Immobility/stupor,Mutism,Verbigeration,Rigidity,Waxy flexibility | 91.7 | 89.0 | 94.2 |
| 5 | Immobility/stupor,Posturing/catalepsy,Echopraxia/echolalia,Stereotypy,Rigidity | 91.7 | 89.0 | 94.2 |
| 5 | Immobility/stupor,Posturing/catalepsy,Grimacing,Echopraxia/echolalia,Rigidity | 91.7 | 89.0 | 94.2 |
| 5 | Immobility/stupor,Posturing/catalepsy,Grimacing,Echopraxia/echolalia,Withdrawal | 91.7 | 89.0 | 94.2 |
| 5 | Immobility/stupor,Posturing/catalepsy,Grimacing,Negativism,Waxy flexibility | 91.7 | 89.0 | 94.2 |
| 5 | Immobility/stupor,Posturing/catalepsy,Mannerisms,Verbigeration,Withdrawal | 91.7 | 89.0 | 94.2 |
| 5 | Immobility/stupor,Posturing/catalepsy,Stereotypy,Mannerisms,Withdrawal | 91.7 | 89.0 | 94.2 |
| 5 | Immobility/stupor,Stereotypy,Mannerisms,Rigidity,Waxy flexibility | 91.7 | 89.0 | 94.2 |
| 5 | Mutism,Echopraxia/echolalia,Rigidity,Negativism,Withdrawal | 91.7 | 89.0 | 94.2 |
| 5 | Mutism,Stereotypy,Mannerisms,Rigidity,Negativism | 91.7 | 89.0 | 94.2 |
| 5 | Mutism,Stereotypy,Verbigeration,Rigidity,Negativism | 91.7 | 89.0 | 94.2 |
| 5 | Posturing/catalepsy,Mannerisms,Rigidity,Negativism,Withdrawal | 91.7 | 89.0 | 94.2 |
| 5 | Staring,Grimacing,Echopraxia/echolalia,Verbigeration,Waxy flexibility | 91.7 | 89.0 | 94.2 |
| 5 | Staring,Posturing/catalepsy,Echopraxia/echolalia,Rigidity,Waxy flexibility | 91.7 | 89.0 | 94.2 |
| 5 | Staring,Posturing/catalepsy,Echopraxia/echolalia,Stereotypy,Verbigeration | 91.7 | 89.0 | 94.2 |
| 5 | Staring,Posturing/catalepsy,Grimacing,Rigidity,Waxy flexibility | 91.7 | 89.0 | 94.2 |
| 5 | Staring,Posturing/catalepsy,Stereotypy,Mannerisms,Waxy flexibility | 91.7 | 89.2 | 94.2 |
| 5 | Staring,Posturing/catalepsy,Stereotypy,Verbigeration,Waxy flexibility | 91.7 | 89.0 | 94.2 |
| 5 | Excitement,Immobility/stupor,Grimacing,Stereotypy,Mannerisms | 91.5 | 88.8 | 93.9 |
| 5 | Excitement,Mutism,Posturing/catalepsy,Echopraxia/echolalia,Withdrawal | 91.5 | 88.8 | 93.9 |
| 5 | Excitement,Mutism,Posturing/catalepsy,Verbigeration,Negativism | 91.5 | 88.8 | 93.9 |
| 5 | Immobility/stupor,Grimacing,Stereotypy,Mannerisms,Withdrawal | 91.5 | 88.8 | 93.9 |
| 5 | Immobility/stupor,Grimacing,Verbigeration,Negativism,Waxy flexibility | 91.5 | 88.8 | 93.9 |
| 5 | Immobility/stupor,Grimacing,Verbigeration,Rigidity,Waxy flexibility | 91.5 | 88.8 | 93.9 |
| 5 | Immobility/stupor,Mutism,Grimacing,Stereotypy,Withdrawal | 91.5 | 88.8 | 93.9 |
| 5 | Immobility/stupor,Mutism,Grimacing,Verbigeration,Withdrawal | 91.5 | 88.8 | 93.9 |
| 5 | Immobility/stupor,Mutism,Mannerisms,Verbigeration,Withdrawal | 91.5 | 88.8 | 93.9 |
| 5 | Immobility/stupor,Mutism,Posturing/catalepsy,Echopraxia/echolalia,Mannerisms | 91.5 | 88.8 | 93.9 |
| 5 | Immobility/stupor,Mutism,Posturing/catalepsy,Grimacing,Rigidity | 91.5 | 88.8 | 93.9 |
| 5 | Immobility/stupor,Mutism,Posturing/catalepsy,Negativism,Waxy flexibility | 91.5 | 88.8 | 93.9 |
| 5 | Immobility/stupor,Mutism,Posturing/catalepsy,Stereotypy,Withdrawal | 91.5 | 88.8 | 93.9 |
| 5 | Immobility/stupor,Mutism,Stereotypy,Mannerisms,Withdrawal | 91.5 | 88.8 | 93.9 |
| 5 | Immobility/stupor,Mutism,Stereotypy,Rigidity,Waxy flexibility | 91.5 | 88.8 | 93.9 |
| 5 | Immobility/stupor,Posturing/catalepsy,Echopraxia/echolalia,Negativism,Waxy flexibility | 91.5 | 88.8 | 93.9 |
| 5 | Immobility/stupor,Posturing/catalepsy,Echopraxia/echolalia,Stereotypy,Withdrawal | 91.5 | 88.8 | 93.9 |
| 5 | Immobility/stupor,Stereotypy,Verbigeration,Rigidity,Waxy flexibility | 91.5 | 88.8 | 93.9 |
| 5 | Mutism,Echopraxia/echolalia,Mannerisms,Rigidity,Negativism | 91.5 | 88.8 | 93.9 |
| 5 | Mutism,Posturing/catalepsy,Stereotypy,Rigidity,Negativism | 91.5 | 88.8 | 93.9 |
| 5 | Staring,Posturing/catalepsy,Grimacing,Stereotypy,Waxy flexibility | 91.5 | 88.8 | 93.9 |
| 5 | Staring,Posturing/catalepsy,Mannerisms,Verbigeration,Waxy flexibility | 91.5 | 88.8 | 93.9 |
| 5 | Mutism,Posturing/catalepsy,Mannerisms,Rigidity,Withdrawal | 91.4 | 88.8 | 93.9 |
| 5 | Excitement,Mutism,Posturing/catalepsy,Grimacing,Negativism | 91.3 | 88.6 | 93.7 |
| 5 | Excitement,Mutism,Posturing/catalepsy,Verbigeration,Withdrawal | 91.3 | 88.6 | 93.7 |
| 5 | Immobility/stupor,Grimacing,Echopraxia/echolalia,Mannerisms,Withdrawal | 91.3 | 88.6 | 93.7 |
| 5 | Immobility/stupor,Grimacing,Echopraxia/echolalia,Rigidity,Waxy flexibility | 91.3 | 88.6 | 93.7 |
| 5 | Immobility/stupor,Mutism,Echopraxia/echolalia,Stereotypy,Withdrawal | 91.3 | 88.6 | 93.7 |
| 5 | Immobility/stupor,Mutism,Grimacing,Echopraxia/echolalia,Negativism | 91.3 | 88.6 | 93.7 |
| 5 | Immobility/stupor,Mutism,Grimacing,Echopraxia/echolalia,Withdrawal | 91.3 | 88.6 | 93.7 |
| 5 | Immobility/stupor,Mutism,Posturing/catalepsy,Mannerisms,Verbigeration | 91.3 | 88.6 | 93.7 |
| 5 | Immobility/stupor,Mutism,Posturing/catalepsy,Stereotypy,Verbigeration | 91.3 | 88.6 | 93.7 |
| 5 | Immobility/stupor,Mutism,Stereotypy,Verbigeration,Withdrawal | 91.3 | 88.6 | 93.7 |
| 5 | Immobility/stupor,Posturing/catalepsy,Grimacing,Verbigeration,Withdrawal | 91.3 | 88.6 | 93.7 |
| 5 | Immobility/stupor,Posturing/catalepsy,Stereotypy,Verbigeration,Withdrawal | 91.3 | 88.6 | 93.7 |
| 5 | Immobility/stupor,Staring,Posturing/catalepsy,Grimacing,Waxy flexibility | 91.3 | 88.6 | 93.7 |
| 5 | Mutism,Posturing/catalepsy,Echopraxia/echolalia,Rigidity,Negativism | 91.3 | 88.6 | 93.7 |
| 5 | Mutism,Rigidity,Negativism,Waxy flexibility,Withdrawal | 91.3 | 88.6 | 93.7 |
| 5 | Posturing/catalepsy,Stereotypy,Rigidity,Negativism,Withdrawal | 91.3 | 88.6 | 93.7 |
| 5 | Staring,Posturing/catalepsy,Echopraxia/echolalia,Mannerisms,Waxy flexibility | 91.3 | 88.6 | 93.7 |
| 5 | Staring,Posturing/catalepsy,Echopraxia/echolalia,Stereotypy,Waxy flexibility | 91.3 | 88.6 | 93.7 |
| 5 | Staring,Posturing/catalepsy,Grimacing,Verbigeration,Waxy flexibility | 91.3 | 88.6 | 93.7 |
| 5 | Excitement,Mutism,Mannerisms,Rigidity,Withdrawal | 91.2 | 88.6 | 93.7 |
| 5 | Excitement,Mutism,Posturing/catalepsy,Grimacing,Withdrawal | 91.2 | 88.6 | 93.7 |
| 5 | Excitement,Mutism,Posturing/catalepsy,Stereotypy,Negativism | 91.2 | 88.6 | 93.7 |
| 5 | Immobility/stupor,Echopraxia/echolalia,Stereotypy,Mannerisms,Withdrawal | 91.2 | 88.6 | 93.7 |
| 5 | Immobility/stupor,Grimacing,Stereotypy,Verbigeration,Withdrawal | 91.2 | 88.6 | 93.7 |
| 5 | Immobility/stupor,Posturing/catalepsy,Echopraxia/echolalia,Verbigeration,Rigidity | 91.2 | 88.6 | 93.7 |
| 5 | Mutism,Grimacing,Rigidity,Negativism,Withdrawal | 91.2 | 88.6 | 93.7 |
| 5 | Immobility/stupor,Mutism,Mannerisms,Waxy flexibility,Withdrawal | 91.1 | 88.3 | 93.5 |
| 5 | Staring,Echopraxia/echolalia,Mannerisms,Verbigeration,Waxy flexibility | 91.1 | 88.3 | 93.5 |
| 5 | Excitement,Mutism,Mannerisms,Rigidity,Negativism | 91.0 | 88.3 | 93.5 |
| 5 | Excitement,Mutism,Posturing/catalepsy,Mannerisms,Withdrawal | 91.0 | 88.1 | 93.7 |
| 5 | Excitement,Mutism,Posturing/catalepsy,Stereotypy,Withdrawal | 91.0 | 88.3 | 93.5 |
| 5 | Excitement,Mutism,Stereotypy,Rigidity,Waxy flexibility | 91.0 | 88.3 | 93.5 |
| 5 | Immobility/stupor,Echopraxia/echolalia,Mannerisms,Rigidity,Waxy flexibility | 91.0 | 88.3 | 93.5 |
| 5 | Immobility/stupor,Echopraxia/echolalia,Stereotypy,Rigidity,Waxy flexibility | 91.0 | 88.3 | 93.5 |
| 5 | Immobility/stupor,Grimacing,Negativism,Waxy flexibility,Withdrawal | 91.0 | 88.3 | 93.5 |
| 5 | Immobility/stupor,Mannerisms,Verbigeration,Rigidity,Waxy flexibility | 91.0 | 88.3 | 93.5 |
| 5 | Immobility/stupor,Mutism,Echopraxia/echolalia,Rigidity,Waxy flexibility | 91.0 | 88.3 | 93.7 |
| 5 | Immobility/stupor,Mutism,Grimacing,Rigidity,Waxy flexibility | 91.0 | 88.3 | 93.5 |
| 5 | Immobility/stupor,Mutism,Grimacing,Stereotypy,Verbigeration | 91.0 | 88.3 | 93.5 |
| 5 | Immobility/stupor,Mutism,Posturing/catalepsy,Echopraxia/echolalia,Verbigeration | 91.0 | 88.3 | 93.7 |
| 5 | Immobility/stupor,Mutism,Posturing/catalepsy,Grimacing,Echopraxia/echolalia | 91.0 | 88.3 | 93.5 |
| 5 | Immobility/stupor,Mutism,Posturing/catalepsy,Grimacing,Stereotypy | 91.0 | 88.3 | 93.5 |
| 5 | Mutism,Grimacing,Mannerisms,Rigidity,Negativism | 91.0 | 88.3 | 93.7 |
| 5 | Mutism,Mannerisms,Rigidity,Negativism,Waxy flexibility | 91.0 | 88.3 | 93.5 |
| 5 | Mutism,Mannerisms,Verbigeration,Rigidity,Negativism | 91.0 | 88.3 | 93.7 |
| 5 | Mutism,Posturing/catalepsy,Stereotypy,Negativism,Withdrawal | 91.0 | 88.3 | 93.5 |
| 5 | Mutism,Posturing/catalepsy,Verbigeration,Negativism,Withdrawal | 91.0 | 88.3 | 93.5 |
| 5 | Staring,Echopraxia/echolalia,Stereotypy,Mannerisms,Verbigeration | 91.0 | 88.3 | 93.7 |
| 5 | Staring,Echopraxia/echolalia,Stereotypy,Verbigeration,Waxy flexibility | 91.0 | 88.3 | 93.5 |
| 5 | Staring,Posturing/catalepsy,Grimacing,Echopraxia/echolalia,Waxy flexibility | 91.0 | 88.3 | 93.5 |
| 5 | Excitement,Mutism,Posturing/catalepsy,Negativism,Waxy flexibility | 90.8 | 88.1 | 93.5 |
| 5 | Excitement,Mutism,Stereotypy,Negativism,Withdrawal | 90.8 | 88.1 | 93.5 |
| 5 | Immobility/stupor,Grimacing,Mannerisms,Verbigeration,Withdrawal | 90.8 | 88.1 | 93.5 |
| 5 | Immobility/stupor,Mutism,Echopraxia/echolalia,Verbigeration,Withdrawal | 90.8 | 87.9 | 93.5 |
| 5 | Immobility/stupor,Mutism,Echopraxia/echolalia,Waxy flexibility,Withdrawal | 90.8 | 88.1 | 93.5 |
| 5 | Immobility/stupor,Mutism,Grimacing,Echopraxia/echolalia,Mannerisms | 90.8 | 88.1 | 93.5 |
| 5 | Immobility/stupor,Mutism,Grimacing,Echopraxia/echolalia,Verbigeration | 90.8 | 87.9 | 93.3 |
| 5 | Immobility/stupor,Mutism,Grimacing,Negativism,Waxy flexibility | 90.8 | 88.1 | 93.5 |
| 5 | Immobility/stupor,Mutism,Posturing/catalepsy,Echopraxia/echolalia,Stereotypy | 90.8 | 87.9 | 93.5 |
| 5 | Immobility/stupor,Posturing/catalepsy,Echopraxia/echolalia,Verbigeration,Withdrawal | 90.8 | 88.1 | 93.5 |
| 5 | Immobility/stupor,Posturing/catalepsy,Grimacing,Echopraxia/echolalia,Mannerisms | 90.8 | 88.1 | 93.5 |
| 5 | Immobility/stupor,Posturing/catalepsy,Grimacing,Stereotypy,Verbigeration | 90.8 | 87.9 | 93.5 |
| 5 | Immobility/stupor,Posturing/catalepsy,Mannerisms,Rigidity,Waxy flexibility | 90.8 | 88.1 | 93.5 |
| 5 | Immobility/stupor,Posturing/catalepsy,Rigidity,Waxy flexibility,Withdrawal | 90.8 | 88.1 | 93.5 |
| 5 | Immobility/stupor,Stereotypy,Mannerisms,Verbigeration,Withdrawal | 90.8 | 88.1 | 93.3 |
| 5 | Mutism,Posturing/catalepsy,Echopraxia/echolalia,Negativism,Withdrawal | 90.8 | 88.1 | 93.5 |
| 5 | Mutism,Stereotypy,Rigidity,Negativism,Waxy flexibility | 90.8 | 88.1 | 93.5 |
| 5 | Posturing/catalepsy,Verbigeration,Rigidity,Negativism,Withdrawal | 90.8 | 88.1 | 93.5 |
| 5 | Excitement,Mutism,Echopraxia/echolalia,Rigidity,Waxy flexibility | 90.6 | 87.9 | 93.0 |
| 5 | Excitement,Mutism,Posturing/catalepsy,Mannerisms,Negativism | 90.6 | 87.7 | 93.3 |
| 5 | Excitement,Posturing/catalepsy,Echopraxia/echolalia,Rigidity,Withdrawal | 90.6 | 87.9 | 93.3 |
| 5 | Immobility/stupor,Echopraxia/echolalia,Stereotypy,Verbigeration,Withdrawal | 90.6 | 87.9 | 93.3 |
| 5 | Immobility/stupor,Grimacing,Echopraxia/echolalia,Verbigeration,Withdrawal | 90.6 | 87.9 | 93.3 |
| 5 | Immobility/stupor,Grimacing,Mannerisms,Waxy flexibility,Withdrawal | 90.6 | 87.7 | 93.0 |
| 5 | Immobility/stupor,Mutism,Echopraxia/echolalia,Stereotypy,Mannerisms | 90.6 | 87.9 | 93.3 |
| 5 | Immobility/stupor,Mutism,Grimacing,Echopraxia/echolalia,Stereotypy | 90.6 | 87.9 | 93.3 |
| 5 | Immobility/stupor,Mutism,Grimacing,Mannerisms,Verbigeration | 90.6 | 87.9 | 93.0 |
| 5 | Immobility/stupor,Mutism,Posturing/catalepsy,Stereotypy,Mannerisms | 90.6 | 87.7 | 93.3 |
| 5 | Immobility/stupor,Mutism,Stereotypy,Waxy flexibility,Withdrawal | 90.6 | 87.9 | 93.3 |
| 5 | Immobility/stupor,Mutism,Verbigeration,Waxy flexibility,Withdrawal | 90.6 | 87.7 | 93.0 |
| 5 | Immobility/stupor,Posturing/catalepsy,Grimacing,Echopraxia/echolalia,Stereotypy | 90.6 | 87.9 | 93.3 |
| 5 | Immobility/stupor,Posturing/catalepsy,Grimacing,Mannerisms,Verbigeration | 90.6 | 87.7 | 93.3 |
| 5 | Immobility/stupor,Posturing/catalepsy,Grimacing,Stereotypy,Mannerisms | 90.6 | 87.9 | 93.3 |
| 5 | Mutism,Echopraxia/echolalia,Stereotypy,Rigidity,Negativism | 90.6 | 87.9 | 93.3 |
| 5 | Mutism,Grimacing,Verbigeration,Rigidity,Negativism | 90.6 | 87.9 | 93.3 |
| 5 | Mutism,Mannerisms,Rigidity,Waxy flexibility,Withdrawal | 90.6 | 87.7 | 93.3 |
| 5 | Mutism,Posturing/catalepsy,Grimacing,Negativism,Withdrawal | 90.6 | 87.9 | 93.3 |
| 5 | Excitement,Mutism,Grimacing,Rigidity,Waxy flexibility | 90.4 | 87.4 | 93.0 |
| 5 | Excitement,Mutism,Negativism,Waxy flexibility,Withdrawal | 90.4 | 87.4 | 93.0 |
| 5 | Excitement,Posturing/catalepsy,Rigidity,Waxy flexibility,Withdrawal | 90.4 | 87.7 | 93.0 |
| 5 | Excitement,Posturing/catalepsy,Verbigeration,Rigidity,Withdrawal | 90.4 | 87.7 | 93.0 |
| 5 | Immobility/stupor,Echopraxia/echolalia,Stereotypy,Waxy flexibility,Withdrawal | 90.4 | 87.7 | 93.0 |
| 5 | Immobility/stupor,Mutism,Echopraxia/echolalia,Mannerisms,Verbigeration | 90.4 | 87.4 | 93.0 |
| 5 | Immobility/stupor,Mutism,Echopraxia/echolalia,Mannerisms,Waxy flexibility | 90.4 | 87.7 | 93.0 |
| 5 | Immobility/stupor,Mutism,Grimacing,Stereotypy,Mannerisms | 90.4 | 87.7 | 93.0 |
| 5 | Immobility/stupor,Mutism,Stereotypy,Mannerisms,Verbigeration | 90.4 | 87.4 | 93.0 |
| 5 | Immobility/stupor,Posturing/catalepsy,Stereotypy,Rigidity,Waxy flexibility | 90.4 | 87.4 | 93.0 |
| 5 | Mutism,Grimacing,Mannerisms,Rigidity,Withdrawal | 90.4 | 87.4 | 93.0 |
| 5 | Mutism,Grimacing,Stereotypy,Rigidity,Negativism | 90.4 | 87.4 | 93.0 |
| 5 | Mutism,Posturing/catalepsy,Grimacing,Rigidity,Negativism | 90.4 | 87.4 | 93.0 |
| 5 | Mutism,Posturing/catalepsy,Stereotypy,Rigidity,Withdrawal | 90.4 | 87.7 | 93.0 |
| 5 | Mutism,Posturing/catalepsy,Verbigeration,Rigidity,Withdrawal | 90.4 | 87.7 | 93.0 |
| 5 | Mutism,Stereotypy,Rigidity,Waxy flexibility,Withdrawal | 90.4 | 87.7 | 93.0 |
| 5 | Staring,Posturing/catalepsy,Echopraxia/echolalia,Verbigeration,Waxy flexibility | 90.4 | 87.4 | 93.0 |
| 5 | Excitement,Posturing/catalepsy,Stereotypy,Negativism,Withdrawal | 90.3 | 87.4 | 93.0 |
| 5 | Immobility/stupor,Echopraxia/echolalia,Mannerisms,Verbigeration,Withdrawal | 90.3 | 87.4 | 93.0 |
| 5 | Immobility/stupor,Grimacing,Echopraxia/echolalia,Stereotypy,Mannerisms | 90.3 | 87.4 | 93.0 |
| 5 | Immobility/stupor,Grimacing,Echopraxia/echolalia,Waxy flexibility,Withdrawal | 90.3 | 87.4 | 93.0 |
| 5 | Immobility/stupor,Grimacing,Stereotypy,Waxy flexibility,Withdrawal | 90.3 | 87.4 | 93.0 |
| 5 | Immobility/stupor,Posturing/catalepsy,Grimacing,Echopraxia/echolalia,Verbigeration | 90.3 | 87.4 | 93.0 |
| 5 | Immobility/stupor,Posturing/catalepsy,Grimacing,Rigidity,Waxy flexibility | 90.3 | 87.4 | 93.0 |
| 5 | Immobility/stupor,Posturing/catalepsy,Mannerisms,Waxy flexibility,Withdrawal | 90.3 | 87.4 | 93.0 |
| 5 | Mutism,Echopraxia/echolalia,Verbigeration,Rigidity,Negativism | 90.3 | 87.4 | 93.0 |
| 5 | Immobility/stupor,Grimacing,Echopraxia/echolalia,Stereotypy,Waxy flexibility | 90.2 | 87.4 | 92.8 |
| 5 | Excitement,Mutism,Echopraxia/echolalia,Negativism,Withdrawal | 90.1 | 87.2 | 92.8 |
| 5 | Excitement,Mutism,Echopraxia/echolalia,Stereotypy,Rigidity | 90.1 | 87.2 | 92.8 |
| 5 | Excitement,Mutism,Verbigeration,Rigidity,Waxy flexibility | 90.1 | 87.2 | 92.8 |
| 5 | Excitement,Posturing/catalepsy,Echopraxia/echolalia,Negativism,Withdrawal | 90.1 | 87.2 | 92.8 |
| 5 | Excitement,Posturing/catalepsy,Mannerisms,Rigidity,Withdrawal | 90.1 | 87.2 | 92.8 |
| 5 | Excitement,Posturing/catalepsy,Verbigeration,Negativism,Withdrawal | 90.1 | 87.2 | 92.8 |
| 5 | Immobility/stupor,Echopraxia/echolalia,Mannerisms,Waxy flexibility,Withdrawal | 90.1 | 87.2 | 92.8 |
| 5 | Immobility/stupor,Echopraxia/echolalia,Verbigeration,Rigidity,Waxy flexibility | 90.1 | 87.2 | 92.8 |
| 5 | Immobility/stupor,Mutism,Echopraxia/echolalia,Stereotypy,Verbigeration | 90.1 | 87.2 | 92.8 |
| 5 | Immobility/stupor,Mutism,Grimacing,Mannerisms,Waxy flexibility | 90.1 | 87.2 | 92.8 |
| 5 | Immobility/stupor,Mutism,Grimacing,Verbigeration,Waxy flexibility | 90.1 | 87.2 | 92.8 |
| 5 | Immobility/stupor,Mutism,Grimacing,Waxy flexibility,Withdrawal | 90.1 | 87.2 | 92.8 |
| 5 | Immobility/stupor,Mutism,Posturing/catalepsy,Waxy flexibility,Withdrawal | 90.1 | 87.4 | 92.8 |
| 5 | Immobility/stupor,Posturing/catalepsy,Stereotypy,Waxy flexibility,Withdrawal | 90.1 | 87.2 | 92.8 |
| 5 | Immobility/stupor,Stereotypy,Mannerisms,Waxy flexibility,Withdrawal | 90.1 | 87.2 | 92.8 |
| 5 | Mutism,Posturing/catalepsy,Echopraxia/echolalia,Mannerisms,Negativism | 90.1 | 87.2 | 92.8 |
| 5 | Mutism,Posturing/catalepsy,Echopraxia/echolalia,Rigidity,Withdrawal | 90.1 | 87.2 | 92.8 |
| 5 | Mutism,Posturing/catalepsy,Grimacing,Rigidity,Withdrawal | 90.1 | 87.2 | 92.8 |
| 5 | Mutism,Verbigeration,Rigidity,Negativism,Waxy flexibility | 90.1 | 87.2 | 92.8 |
| 5 | Posturing/catalepsy,Echopraxia/echolalia,Rigidity,Negativism,Withdrawal | 90.1 | 87.2 | 92.8 |
| 5 | Excitement,Mutism,Grimacing,Echopraxia/echolalia,Rigidity | 89.9 | 87.0 | 92.6 |
| 5 | Excitement,Mutism,Verbigeration,Negativism,Withdrawal | 89.9 | 87.0 | 92.6 |
| 5 | Excitement,Posturing/catalepsy,Grimacing,Rigidity,Withdrawal | 89.9 | 87.0 | 92.6 |
| 5 | Excitement,Posturing/catalepsy,Stereotypy,Rigidity,Withdrawal | 89.9 | 87.0 | 92.6 |
| 5 | Immobility/stupor,Grimacing,Echopraxia/echolalia,Stereotypy,Verbigeration | 89.9 | 87.0 | 92.6 |
| 5 | Immobility/stupor,Mutism,Echopraxia/echolalia,Stereotypy,Waxy flexibility | 89.9 | 87.0 | 92.6 |
| 5 | Immobility/stupor,Mutism,Grimacing,Echopraxia/echolalia,Waxy flexibility | 89.9 | 87.0 | 92.6 |
| 5 | Immobility/stupor,Mutism,Stereotypy,Verbigeration,Waxy flexibility | 89.9 | 87.0 | 92.6 |
| 5 | Immobility/stupor,Posturing/catalepsy,Echopraxia/echolalia,Stereotypy,Mannerisms | 89.9 | 87.0 | 92.6 |
| 5 | Immobility/stupor,Posturing/catalepsy,Echopraxia/echolalia,Stereotypy,Verbigeration | 89.9 | 87.0 | 92.6 |
| 5 | Immobility/stupor,Posturing/catalepsy,Grimacing,Waxy flexibility,Withdrawal | 89.9 | 87.2 | 92.6 |
| 5 | Immobility/stupor,Posturing/catalepsy,Stereotypy,Mannerisms,Verbigeration | 89.9 | 87.0 | 92.6 |
| 5 | Immobility/stupor,Posturing/catalepsy,Verbigeration,Waxy flexibility,Withdrawal | 89.9 | 87.0 | 92.6 |
| 5 | Immobility/stupor,Stereotypy,Verbigeration,Waxy flexibility,Withdrawal | 89.9 | 87.0 | 92.6 |
| 5 | Mutism,Posturing/catalepsy,Negativism,Waxy flexibility,Withdrawal | 89.9 | 87.0 | 92.6 |
| 5 | Mutism,Posturing/catalepsy,Rigidity,Negativism,Waxy flexibility | 89.9 | 87.0 | 92.6 |
| 5 | Mutism,Posturing/catalepsy,Stereotypy,Verbigeration,Negativism | 89.9 | 87.0 | 92.6 |
| 5 | Mutism,Stereotypy,Mannerisms,Rigidity,Withdrawal | 89.9 | 87.0 | 92.6 |
| 5 | Mutism,Verbigeration,Rigidity,Waxy flexibility,Withdrawal | 89.9 | 87.0 | 92.6 |
| 5 | Posturing/catalepsy,Grimacing,Rigidity,Negativism,Withdrawal | 89.9 | 87.0 | 92.6 |
| 5 | Excitement,Mutism,Grimacing,Verbigeration,Rigidity | 89.7 | 86.8 | 92.4 |
| 5 | Excitement,Mutism,Stereotypy,Verbigeration,Rigidity | 89.7 | 86.8 | 92.4 |
| 5 | Excitement,Mutism,Stereotypy,Waxy flexibility,Withdrawal | 89.7 | 86.8 | 92.4 |
| 5 | Excitement,Posturing/catalepsy,Grimacing,Negativism,Withdrawal | 89.7 | 86.8 | 92.4 |
| 5 | Excitement,Posturing/catalepsy,Mannerisms,Negativism,Withdrawal | 89.7 | 86.8 | 92.4 |
| 5 | Immobility/stupor,Grimacing,Echopraxia/echolalia,Mannerisms,Waxy flexibility | 89.7 | 86.8 | 92.4 |
| 5 | Immobility/stupor,Grimacing,Stereotypy,Mannerisms,Verbigeration | 89.7 | 86.8 | 92.4 |
| 5 | Immobility/stupor,Grimacing,Stereotypy,Mannerisms,Waxy flexibility | 89.7 | 86.8 | 92.4 |
| 5 | Immobility/stupor,Grimacing,Stereotypy,Verbigeration,Waxy flexibility | 89.7 | 86.8 | 92.4 |
| 5 | Immobility/stupor,Mannerisms,Verbigeration,Waxy flexibility,Withdrawal | 89.7 | 86.8 | 92.4 |
| 5 | Immobility/stupor,Mutism,Echopraxia/echolalia,Verbigeration,Waxy flexibility | 89.7 | 86.8 | 92.4 |
| 5 | Immobility/stupor,Mutism,Grimacing,Stereotypy,Waxy flexibility | 89.7 | 86.8 | 92.4 |
| 5 | Immobility/stupor,Mutism,Mannerisms,Verbigeration,Waxy flexibility | 89.7 | 86.8 | 92.4 |
| 5 | Immobility/stupor,Mutism,Posturing/catalepsy,Verbigeration,Waxy flexibility | 89.7 | 86.8 | 92.4 |
| 5 | Immobility/stupor,Mutism,Stereotypy,Mannerisms,Waxy flexibility | 89.7 | 86.8 | 92.4 |
| 5 | Immobility/stupor,Posturing/catalepsy,Verbigeration,Rigidity,Waxy flexibility | 89.7 | 86.8 | 92.4 |
| 5 | Mutism,Echopraxia/echolalia,Rigidity,Negativism,Waxy flexibility | 89.7 | 86.8 | 92.4 |
| 5 | Mutism,Grimacing,Echopraxia/echolalia,Rigidity,Negativism | 89.7 | 86.8 | 92.4 |
| 5 | Mutism,Grimacing,Rigidity,Waxy flexibility,Withdrawal | 89.7 | 86.8 | 92.4 |
| 5 | Mutism,Grimacing,Stereotypy,Rigidity,Withdrawal | 89.7 | 86.8 | 92.4 |
| 5 | Mutism,Grimacing,Verbigeration,Rigidity,Withdrawal | 89.7 | 86.8 | 92.4 |
| 5 | Mutism,Posturing/catalepsy,Echopraxia/echolalia,Mannerisms,Rigidity | 89.7 | 86.8 | 92.4 |
| 5 | Mutism,Posturing/catalepsy,Grimacing,Mannerisms,Negativism | 89.7 | 86.8 | 92.4 |
| 5 | Mutism,Posturing/catalepsy,Grimacing,Mannerisms,Rigidity | 89.7 | 86.8 | 92.4 |
| 5 | Mutism,Posturing/catalepsy,Mannerisms,Verbigeration,Rigidity | 89.7 | 86.8 | 92.4 |
| 5 | Mutism,Posturing/catalepsy,Stereotypy,Mannerisms,Negativism | 89.7 | 86.8 | 92.4 |
| 5 | Mutism,Stereotypy,Mannerisms,Negativism,Withdrawal | 89.7 | 86.8 | 92.4 |
| 5 | Mutism,Stereotypy,Verbigeration,Rigidity,Withdrawal | 89.7 | 86.8 | 92.6 |
| 5 | Posturing/catalepsy,Stereotypy,Mannerisms,Negativism,Withdrawal | 89.7 | 86.8 | 92.4 |
| 5 | Excitement,Mutism,Echopraxia/echolalia,Verbigeration,Rigidity | 89.5 | 86.5 | 92.2 |
| 5 | Excitement,Mutism,Grimacing,Negativism,Withdrawal | 89.5 | 86.5 | 92.4 |
| 5 | Immobility/stupor,Echopraxia/echolalia,Verbigeration,Waxy flexibility,Withdrawal | 89.5 | 86.5 | 92.2 |
| 5 | Immobility/stupor,Grimacing,Verbigeration,Waxy flexibility,Withdrawal | 89.5 | 86.5 | 92.2 |
| 5 | Immobility/stupor,Mutism,Posturing/catalepsy,Echopraxia/echolalia,Waxy flexibility | 89.5 | 86.5 | 92.2 |
| 5 | Immobility/stupor,Mutism,Posturing/catalepsy,Rigidity,Waxy flexibility | 89.5 | 86.5 | 92.2 |
| 5 | Immobility/stupor,Mutism,Posturing/catalepsy,Stereotypy,Waxy flexibility | 89.5 | 86.5 | 92.2 |
| 5 | Immobility/stupor,Posturing/catalepsy,Echopraxia/echolalia,Mannerisms,Verbigeration | 89.5 | 86.5 | 92.2 |
| 5 | Immobility/stupor,Posturing/catalepsy,Echopraxia/echolalia,Rigidity,Waxy flexibility | 89.5 | 86.5 | 92.2 |
| 5 | Immobility/stupor,Posturing/catalepsy,Echopraxia/echolalia,Waxy flexibility,Withdrawal | 89.5 | 86.5 | 92.2 |
| 5 | Immobility/stupor,Posturing/catalepsy,Grimacing,Verbigeration,Waxy flexibility | 89.5 | 86.5 | 92.2 |
| 5 | Mutism,Echopraxia/echolalia,Mannerisms,Rigidity,Withdrawal | 89.5 | 86.5 | 92.2 |
| 5 | Mutism,Echopraxia/echolalia,Stereotypy,Negativism,Withdrawal | 89.5 | 86.5 | 92.2 |
| 5 | Mutism,Echopraxia/echolalia,Stereotypy,Rigidity,Withdrawal | 89.5 | 86.5 | 92.2 |
| 5 | Mutism,Mannerisms,Negativism,Waxy flexibility,Withdrawal | 89.5 | 86.5 | 92.2 |
| 5 | Mutism,Mannerisms,Verbigeration,Rigidity,Withdrawal | 89.5 | 86.5 | 92.2 |
| 5 | Mutism,Posturing/catalepsy,Echopraxia/echolalia,Verbigeration,Negativism | 89.5 | 86.5 | 92.2 |
| 5 | Mutism,Posturing/catalepsy,Grimacing,Verbigeration,Rigidity | 89.5 | 86.5 | 92.2 |
| 5 | Mutism,Posturing/catalepsy,Mannerisms,Verbigeration,Negativism | 89.5 | 86.5 | 92.2 |
| 5 | Mutism,Posturing/catalepsy,Rigidity,Waxy flexibility,Withdrawal | 89.5 | 86.5 | 92.2 |
| 5 | Mutism,Stereotypy,Verbigeration,Negativism,Withdrawal | 89.5 | 86.5 | 92.4 |
| 5 | Posturing/catalepsy,Stereotypy,Verbigeration,Negativism,Withdrawal | 89.5 | 86.5 | 92.4 |
| 5 | Stereotypy,Mannerisms,Rigidity,Negativism,Withdrawal | 89.5 | 86.5 | 92.2 |
| 5 | Excitement,Mutism,Grimacing,Stereotypy,Rigidity | 89.4 | 86.5 | 92.2 |
| 5 | Immobility/stupor,Mutism,Posturing/catalepsy,Mannerisms,Waxy flexibility | 89.4 | 86.5 | 92.2 |
| 5 | Mutism,Posturing/catalepsy,Stereotypy,Verbigeration,Rigidity | 89.4 | 86.5 | 92.2 |
| 5 | Immobility/stupor,Posturing/catalepsy,Grimacing,Mannerisms,Waxy flexibility | 89.3 | 86.3 | 92.2 |
| 5 | Mutism,Posturing/catalepsy,Grimacing,Verbigeration,Negativism | 89.3 | 86.3 | 92.2 |
| 5 | Excitement,Posturing/catalepsy,Negativism,Waxy flexibility,Withdrawal | 89.2 | 86.3 | 91.9 |
| 5 | Excitement,Stereotypy,Rigidity,Negativism,Withdrawal | 89.2 | 86.3 | 91.9 |
| 5 | Immobility/stupor,Grimacing,Echopraxia/echolalia,Mannerisms,Verbigeration | 89.2 | 86.3 | 91.9 |
| 5 | Immobility/stupor,Grimacing,Echopraxia/echolalia,Verbigeration,Waxy flexibility | 89.2 | 86.3 | 91.9 |
| 5 | Immobility/stupor,Mutism,Posturing/catalepsy,Grimacing,Waxy flexibility | 89.2 | 86.1 | 92.2 |
| 5 | Immobility/stupor,Posturing/catalepsy,Grimacing,Stereotypy,Waxy flexibility | 89.2 | 86.3 | 91.9 |
| 5 | Mutism,Echopraxia/echolalia,Mannerisms,Negativism,Withdrawal | 89.2 | 86.3 | 91.9 |
| 5 | Mutism,Echopraxia/echolalia,Rigidity,Waxy flexibility,Withdrawal | 89.2 | 86.3 | 91.9 |
| 5 | Mutism,Grimacing,Rigidity,Negativism,Waxy flexibility | 89.2 | 86.3 | 91.9 |
| 5 | Mutism,Posturing/catalepsy,Echopraxia/echolalia,Stereotypy,Negativism | 89.2 | 86.3 | 91.9 |
| 5 | Mutism,Posturing/catalepsy,Grimacing,Mannerisms,Withdrawal | 89.2 | 86.3 | 91.9 |
| 5 | Mutism,Stereotypy,Negativism,Waxy flexibility,Withdrawal | 89.2 | 86.3 | 91.9 |
| 5 | Posturing/catalepsy,Rigidity,Negativism,Waxy flexibility,Withdrawal | 89.2 | 86.3 | 91.9 |
| 5 | Stereotypy,Verbigeration,Rigidity,Negativism,Withdrawal | 89.2 | 86.3 | 92.2 |
| 5 | Echopraxia/echolalia,Stereotypy,Rigidity,Negativism,Withdrawal | 89.0 | 86.1 | 91.7 |
| 5 | Excitement,Echopraxia/echolalia,Rigidity,Negativism,Withdrawal | 89.0 | 86.1 | 91.9 |
| 5 | Excitement,Mutism,Echopraxia/echolalia,Waxy flexibility,Withdrawal | 89.0 | 86.1 | 91.7 |
| 5 | Excitement,Mutism,Mannerisms,Rigidity,Waxy flexibility | 89.0 | 86.1 | 91.9 |
| 5 | Excitement,Mutism,Posturing/catalepsy,Echopraxia/echolalia,Waxy flexibility | 89.0 | 85.9 | 91.9 |
| 5 | Excitement,Mutism,Stereotypy,Negativism,Waxy flexibility | 89.0 | 86.1 | 91.9 |
| 5 | Immobility/stupor,Echopraxia/echolalia,Stereotypy,Mannerisms,Verbigeration | 89.0 | 86.1 | 91.7 |
| 5 | Immobility/stupor,Echopraxia/echolalia,Stereotypy,Mannerisms,Waxy flexibility | 89.0 | 86.1 | 91.7 |
| 5 | Immobility/stupor,Posturing/catalepsy,Grimacing,Echopraxia/echolalia,Waxy flexibility | 89.0 | 86.1 | 91.9 |
| 5 | Immobility/stupor,Posturing/catalepsy,Stereotypy,Verbigeration,Waxy flexibility | 89.0 | 86.1 | 91.9 |
| 5 | Mutism,Grimacing,Echopraxia/echolalia,Rigidity,Withdrawal | 89.0 | 86.1 | 91.9 |
| 5 | Mutism,Posturing/catalepsy,Echopraxia/echolalia,Verbigeration,Rigidity | 89.0 | 86.1 | 91.9 |
| 5 | Mutism,Posturing/catalepsy,Grimacing,Echopraxia/echolalia,Negativism | 89.0 | 86.1 | 91.9 |
| 5 | Mutism,Posturing/catalepsy,Grimacing,Stereotypy,Negativism | 89.0 | 86.1 | 91.7 |
| 5 | Mutism,Posturing/catalepsy,Stereotypy,Mannerisms,Rigidity | 89.0 | 86.1 | 91.9 |
| 5 | Posturing/catalepsy,Echopraxia/echolalia,Stereotypy,Negativism,Withdrawal | 89.0 | 86.1 | 91.7 |
| 5 | Posturing/catalepsy,Grimacing,Stereotypy,Negativism,Withdrawal | 89.0 | 86.1 | 91.9 |
| 5 | Posturing/catalepsy,Mannerisms,Verbigeration,Negativism,Withdrawal | 89.0 | 85.9 | 91.9 |
| 5 | Excitement,Mutism,Grimacing,Waxy flexibility,Withdrawal | 88.8 | 85.9 | 91.7 |
| 5 | Excitement,Mutism,Mannerisms,Negativism,Withdrawal | 88.8 | 85.9 | 91.7 |
| 5 | Excitement,Mutism,Posturing/catalepsy,Grimacing,Echopraxia/echolalia | 88.8 | 85.9 | 91.5 |
| 5 | Immobility/stupor,Echopraxia/echolalia,Stereotypy,Verbigeration,Waxy flexibility | 88.8 | 85.9 | 91.7 |
| 5 | Immobility/stupor,Grimacing,Mannerisms,Verbigeration,Waxy flexibility | 88.8 | 85.9 | 91.7 |
| 5 | Immobility/stupor,Posturing/catalepsy,Echopraxia/echolalia,Stereotypy,Waxy flexibility | 88.8 | 85.9 | 91.7 |
| 5 | Immobility/stupor,Stereotypy,Mannerisms,Verbigeration,Waxy flexibility | 88.8 | 85.9 | 91.7 |
| 5 | Mutism,Grimacing,Mannerisms,Negativism,Withdrawal | 88.8 | 85.9 | 91.7 |
| 5 | Mutism,Grimacing,Stereotypy,Negativism,Withdrawal | 88.8 | 85.9 | 91.7 |
| 5 | Mutism,Mannerisms,Verbigeration,Negativism,Withdrawal | 88.8 | 85.7 | 91.7 |
| 5 | Mutism,Posturing/catalepsy,Echopraxia/echolalia,Mannerisms,Withdrawal | 88.8 | 85.7 | 91.7 |
| 5 | Mutism,Posturing/catalepsy,Echopraxia/echolalia,Stereotypy,Rigidity | 88.8 | 85.9 | 91.7 |
| 5 | Mutism,Posturing/catalepsy,Grimacing,Stereotypy,Rigidity | 88.8 | 85.9 | 91.7 |
| 5 | Mutism,Posturing/catalepsy,Mannerisms,Negativism,Waxy flexibility | 88.8 | 85.9 | 91.7 |
| 5 | Mutism,Posturing/catalepsy,Mannerisms,Rigidity,Waxy flexibility | 88.8 | 85.7 | 91.7 |
| 5 | Mutism,Posturing/catalepsy,Mannerisms,Verbigeration,Withdrawal | 88.8 | 85.9 | 91.7 |
| 5 | Posturing/catalepsy,Echopraxia/echolalia,Mannerisms,Negativism,Withdrawal | 88.8 | 85.9 | 91.7 |
| 5 | Posturing/catalepsy,Grimacing,Mannerisms,Negativism,Withdrawal | 88.8 | 85.9 | 91.7 |
| 5 | Echopraxia/echolalia,Mannerisms,Rigidity,Negativism,Withdrawal | 88.6 | 85.7 | 91.5 |
| 5 | Excitement,Mutism,Echopraxia/echolalia,Mannerisms,Rigidity | 88.6 | 85.6 | 91.5 |
| 5 | Excitement,Mutism,Echopraxia/echolalia,Stereotypy,Negativism | 88.6 | 85.4 | 91.5 |
| 5 | Excitement,Mutism,Grimacing,Mannerisms,Rigidity | 88.6 | 85.4 | 91.5 |
| 5 | Excitement,Mutism,Posturing/catalepsy,Grimacing,Waxy flexibility | 88.6 | 85.7 | 91.5 |
| 5 | Excitement,Mutism,Posturing/catalepsy,Verbigeration,Waxy flexibility | 88.6 | 85.4 | 91.5 |
| 5 | Excitement,Mutism,Stereotypy,Mannerisms,Rigidity | 88.6 | 85.7 | 91.5 |
| 5 | Excitement,Mutism,Stereotypy,Verbigeration,Negativism | 88.6 | 85.4 | 91.5 |
| 5 | Grimacing,Stereotypy,Rigidity,Negativism,Withdrawal | 88.6 | 85.7 | 91.5 |
| 5 | Mutism,Echopraxia/echolalia,Verbigeration,Rigidity,Withdrawal | 88.6 | 85.4 | 91.5 |
| 5 | Mutism,Posturing/catalepsy,Grimacing,Echopraxia/echolalia,Rigidity | 88.6 | 85.7 | 91.5 |
| 5 | Mutism,Posturing/catalepsy,Grimacing,Stereotypy,Withdrawal | 88.6 | 85.7 | 91.5 |
| 5 | Mutism,Posturing/catalepsy,Stereotypy,Negativism,Waxy flexibility | 88.6 | 85.4 | 91.5 |
| 5 | Mutism,Posturing/catalepsy,Verbigeration,Rigidity,Waxy flexibility | 88.6 | 85.7 | 91.5 |
| 5 | Posturing/catalepsy,Stereotypy,Negativism,Waxy flexibility,Withdrawal | 88.6 | 85.7 | 91.5 |
| 5 | Stereotypy,Rigidity,Negativism,Waxy flexibility,Withdrawal | 88.6 | 85.4 | 91.5 |
| 5 | Excitement,Mutism,Posturing/catalepsy,Echopraxia/echolalia,Verbigeration | 88.5 | 85.4 | 91.5 |
| 5 | Excitement,Mutism,Verbigeration,Waxy flexibility,Withdrawal | 88.5 | 85.4 | 91.5 |
| 5 | Mutism,Posturing/catalepsy,Mannerisms,Waxy flexibility,Withdrawal | 88.5 | 85.4 | 91.5 |
| 5 | Mutism,Posturing/catalepsy,Stereotypy,Mannerisms,Withdrawal | 88.5 | 85.4 | 91.5 |
| 5 | Mutism,Posturing/catalepsy,Grimacing,Verbigeration,Withdrawal | 88.4 | 85.2 | 91.3 |
| 5 | Excitement,Mutism,Echopraxia/echolalia,Negativism,Waxy flexibility | 88.3 | 85.2 | 91.3 |
| 5 | Excitement,Mutism,Echopraxia/echolalia,Stereotypy,Withdrawal | 88.3 | 85.2 | 91.3 |
| 5 | Excitement,Mutism,Posturing/catalepsy,Echopraxia/echolalia,Stereotypy | 88.3 | 85.2 | 91.3 |
| 5 | Excitement,Mutism,Posturing/catalepsy,Grimacing,Verbigeration | 88.3 | 85.4 | 91.3 |
| 5 | Excitement,Mutism,Posturing/catalepsy,Stereotypy,Waxy flexibility | 88.3 | 85.2 | 91.3 |
| 5 | Excitement,Verbigeration,Rigidity,Negativism,Withdrawal | 88.3 | 85.2 | 91.3 |
| 5 | Immobility/stupor,Posturing/catalepsy,Stereotypy,Mannerisms,Waxy flexibility | 88.3 | 85.2 | 91.3 |
| 5 | Mutism,Echopraxia/echolalia,Negativism,Waxy flexibility,Withdrawal | 88.3 | 85.4 | 91.3 |
| 5 | Mutism,Posturing/catalepsy,Echopraxia/echolalia,Negativism,Waxy flexibility | 88.3 | 85.2 | 91.3 |
| 5 | Mutism,Posturing/catalepsy,Grimacing,Echopraxia/echolalia,Withdrawal | 88.3 | 85.4 | 91.3 |
| 5 | Mutism,Posturing/catalepsy,Verbigeration,Negativism,Waxy flexibility | 88.3 | 85.2 | 91.3 |
| 5 | Mutism,Verbigeration,Negativism,Waxy flexibility,Withdrawal | 88.3 | 85.2 | 91.3 |
| 5 | Excitement,Grimacing,Rigidity,Negativism,Withdrawal | 88.1 | 85.2 | 91.0 |
| 5 | Excitement,Mutism,Mannerisms,Verbigeration,Rigidity | 88.1 | 85.0 | 91.0 |
| 5 | Excitement,Mutism,Mannerisms,Waxy flexibility,Withdrawal | 88.1 | 85.0 | 91.0 |
| 5 | Excitement,Mutism,Posturing/catalepsy,Grimacing,Stereotypy | 88.1 | 85.0 | 91.0 |
| 5 | Excitement,Mutism,Posturing/catalepsy,Stereotypy,Verbigeration | 88.1 | 85.0 | 91.0 |
| 5 | Grimacing,Mannerisms,Rigidity,Negativism,Withdrawal | 88.1 | 85.0 | 91.0 |
| 5 | Immobility/stupor,Posturing/catalepsy,Echopraxia/echolalia,Mannerisms,Waxy flexibility | 88.1 | 85.2 | 91.0 |
| 5 | Immobility/stupor,Posturing/catalepsy,Mannerisms,Verbigeration,Waxy flexibility | 88.1 | 85.0 | 91.0 |
| 5 | Mannerisms,Verbigeration,Rigidity,Negativism,Withdrawal | 88.1 | 85.0 | 91.0 |
| 5 | Mutism,Echopraxia/echolalia,Verbigeration,Negativism,Withdrawal | 88.1 | 85.0 | 91.0 |
| 5 | Mutism,Grimacing,Verbigeration,Negativism,Withdrawal | 88.1 | 85.0 | 91.0 |
| 5 | Mutism,Posturing/catalepsy,Echopraxia/echolalia,Rigidity,Waxy flexibility | 88.1 | 85.0 | 91.0 |
| 5 | Mutism,Posturing/catalepsy,Echopraxia/echolalia,Stereotypy,Withdrawal | 88.1 | 85.0 | 91.0 |
| 5 | Mutism,Posturing/catalepsy,Stereotypy,Rigidity,Waxy flexibility | 88.1 | 85.0 | 91.0 |
| 5 | Mutism,Posturing/catalepsy,Stereotypy,Verbigeration,Withdrawal | 88.1 | 85.0 | 91.0 |
| 5 | Mutism,Posturing/catalepsy,Stereotypy,Waxy flexibility,Withdrawal | 88.1 | 85.0 | 91.0 |
| 5 | Mutism,Posturing/catalepsy,Verbigeration,Waxy flexibility,Withdrawal | 88.1 | 85.0 | 91.0 |
| 5 | Mutism,Stereotypy,Mannerisms,Negativism,Waxy flexibility | 88.1 | 85.0 | 91.0 |
| 5 | Mutism,Stereotypy,Verbigeration,Rigidity,Waxy flexibility | 88.1 | 85.0 | 91.0 |
| 5 | Posturing/catalepsy,Mannerisms,Negativism,Waxy flexibility,Withdrawal | 88.1 | 85.0 | 91.0 |
| 5 | Excitement,Mutism,Grimacing,Negativism,Waxy flexibility | 87.9 | 84.8 | 90.8 |
| 5 | Excitement,Mutism,Posturing/catalepsy,Echopraxia/echolalia,Mannerisms | 87.9 | 84.8 | 90.8 |
| 5 | Excitement,Mutism,Stereotypy,Verbigeration,Withdrawal | 87.9 | 84.8 | 90.8 |
| 5 | Excitement,Posturing/catalepsy,Grimacing,Echopraxia/echolalia,Withdrawal | 87.9 | 84.8 | 90.8 |
| 5 | Excitement,Rigidity,Negativism,Waxy flexibility,Withdrawal | 87.9 | 84.8 | 90.8 |
| 5 | Immobility/stupor,Posturing/catalepsy,Echopraxia/echolalia,Verbigeration,Waxy flexibility | 87.9 | 84.8 | 90.8 |
| 5 | Mannerisms,Rigidity,Negativism,Waxy flexibility,Withdrawal | 87.9 | 84.8 | 90.8 |
| 5 | Mutism,Echopraxia/echolalia,Stereotypy,Mannerisms,Negativism | 87.9 | 84.8 | 90.8 |
| 5 | Mutism,Grimacing,Mannerisms,Rigidity,Waxy flexibility | 87.9 | 84.8 | 90.8 |
| 5 | Mutism,Grimacing,Stereotypy,Rigidity,Waxy flexibility | 87.9 | 84.8 | 90.8 |
| 5 | Mutism,Grimacing,Stereotypy,Verbigeration,Rigidity | 87.9 | 84.8 | 90.8 |
| 5 | Mutism,Grimacing,Verbigeration,Rigidity,Waxy flexibility | 87.9 | 84.8 | 90.8 |
| 5 | Mutism,Posturing/catalepsy,Echopraxia/echolalia,Verbigeration,Withdrawal | 87.9 | 84.8 | 90.8 |
| 5 | Mutism,Posturing/catalepsy,Echopraxia/echolalia,Waxy flexibility,Withdrawal | 87.9 | 84.8 | 90.8 |
| 5 | Mutism,Posturing/catalepsy,Grimacing,Negativism,Waxy flexibility | 87.9 | 84.8 | 90.8 |
| 5 | Mutism,Posturing/catalepsy,Grimacing,Waxy flexibility,Withdrawal | 87.9 | 84.8 | 90.8 |
| 5 | Mutism,Stereotypy,Mannerisms,Rigidity,Waxy flexibility | 87.9 | 84.8 | 90.8 |
| 5 | Posturing/catalepsy,Echopraxia/echolalia,Verbigeration,Negativism,Withdrawal | 87.9 | 84.8 | 90.8 |
| 5 | Posturing/catalepsy,Grimacing,Echopraxia/echolalia,Negativism,Withdrawal | 87.9 | 84.8 | 90.8 |
| 5 | Posturing/catalepsy,Grimacing,Mannerisms,Rigidity,Withdrawal | 87.9 | 84.8 | 90.8 |
| 5 | Posturing/catalepsy,Grimacing,Verbigeration,Negativism,Withdrawal | 87.9 | 84.8 | 90.8 |
| 5 | Posturing/catalepsy,Mannerisms,Verbigeration,Rigidity,Withdrawal | 87.9 | 85.0 | 90.8 |
| 5 | Posturing/catalepsy,Stereotypy,Mannerisms,Rigidity,Withdrawal | 87.9 | 84.8 | 90.8 |
| 5 | Excitement,Mannerisms,Rigidity,Negativism,Withdrawal | 87.7 | 84.5 | 90.6 |
| 5 | Excitement,Mutism,Grimacing,Stereotypy,Negativism | 87.7 | 84.5 | 90.6 |
| 5 | Excitement,Mutism,Posturing/catalepsy,Grimacing,Mannerisms | 87.7 | 84.5 | 90.6 |
| 5 | Excitement,Mutism,Posturing/catalepsy,Mannerisms,Waxy flexibility | 87.7 | 84.5 | 90.6 |
| 5 | Excitement,Mutism,Verbigeration,Negativism,Waxy flexibility | 87.7 | 84.5 | 90.6 |
| 5 | Excitement,Stereotypy,Rigidity,Waxy flexibility,Withdrawal | 87.7 | 84.5 | 90.6 |
| 5 | Mutism,Grimacing,Echopraxia/echolalia,Negativism,Withdrawal | 87.7 | 84.5 | 90.6 |
| 5 | Mutism,Posturing/catalepsy,Grimacing,Rigidity,Waxy flexibility | 87.7 | 84.5 | 90.6 |
| 5 | Posturing/catalepsy,Echopraxia/echolalia,Mannerisms,Rigidity,Withdrawal | 87.7 | 84.5 | 90.8 |
| 5 | Immobility/stupor,Echopraxia/echolalia,Mannerisms,Verbigeration,Waxy flexibility | 87.6 | 84.5 | 90.6 |
| 5 | Excitement,Mutism,Posturing/catalepsy,Mannerisms,Verbigeration | 87.5 | 84.3 | 90.4 |
| 5 | Posturing/catalepsy,Mannerisms,Rigidity,Waxy flexibility,Withdrawal | 87.5 | 84.3 | 90.4 |
| 5 | Echopraxia/echolalia,Verbigeration,Rigidity,Negativism,Withdrawal | 87.4 | 84.3 | 90.4 |
| 5 | Excitement,Mutism,Echopraxia/echolalia,Verbigeration,Negativism | 87.4 | 84.3 | 90.4 |
| 5 | Excitement,Mutism,Grimacing,Echopraxia/echolalia,Negativism | 87.4 | 84.3 | 90.4 |
| 5 | Excitement,Mutism,Grimacing,Stereotypy,Withdrawal | 87.4 | 84.3 | 90.4 |
| 5 | Excitement,Posturing/catalepsy,Echopraxia/echolalia,Waxy flexibility,Withdrawal | 87.4 | 84.3 | 90.4 |
| 5 | Mutism,Echopraxia/echolalia,Stereotypy,Rigidity,Waxy flexibility | 87.4 | 84.3 | 90.4 |
| 5 | Mutism,Grimacing,Echopraxia/echolalia,Mannerisms,Rigidity | 87.4 | 84.3 | 90.4 |
| 5 | Mutism,Grimacing,Negativism,Waxy flexibility,Withdrawal | 87.4 | 84.3 | 90.4 |
| 5 | Mutism,Grimacing,Stereotypy,Mannerisms,Rigidity | 87.4 | 84.3 | 90.6 |
| 5 | Mutism,Stereotypy,Mannerisms,Verbigeration,Negativism | 87.4 | 84.3 | 90.4 |
| 5 | Posturing/catalepsy,Verbigeration,Negativism,Waxy flexibility,Withdrawal | 87.4 | 84.3 | 90.4 |
| 5 | Excitement,Mutism,Echopraxia/echolalia,Verbigeration,Withdrawal | 87.2 | 84.1 | 90.4 |
| 5 | Excitement,Mutism,Grimacing,Echopraxia/echolalia,Withdrawal | 87.2 | 84.1 | 90.1 |
| 5 | Excitement,Mutism,Posturing/catalepsy,Stereotypy,Mannerisms | 87.2 | 83.9 | 90.1 |
| 5 | Excitement,Mutism,Stereotypy,Mannerisms,Negativism | 87.2 | 84.1 | 90.1 |
| 5 | Excitement,Posturing/catalepsy,Echopraxia/echolalia,Verbigeration,Withdrawal | 87.2 | 84.1 | 90.1 |
| 5 | Grimacing,Echopraxia/echolalia,Rigidity,Negativism,Withdrawal | 87.2 | 84.1 | 90.1 |
| 5 | Grimacing,Verbigeration,Rigidity,Negativism,Withdrawal | 87.2 | 84.1 | 90.1 |
| 5 | Mutism,Echopraxia/echolalia,Mannerisms,Negativism,Waxy flexibility | 87.2 | 84.1 | 90.4 |
| 5 | Mutism,Echopraxia/echolalia,Mannerisms,Rigidity,Waxy flexibility | 87.2 | 84.1 | 90.1 |
| 5 | Mutism,Echopraxia/echolalia,Stereotypy,Mannerisms,Rigidity | 87.2 | 83.9 | 90.4 |
| 5 | Mutism,Echopraxia/echolalia,Stereotypy,Negativism,Waxy flexibility | 87.2 | 84.1 | 90.1 |
| 5 | Mutism,Grimacing,Echopraxia/echolalia,Rigidity,Waxy flexibility | 87.2 | 84.1 | 90.1 |
| 5 | Mutism,Grimacing,Mannerisms,Verbigeration,Rigidity | 87.2 | 84.3 | 90.1 |
| 5 | Mutism,Stereotypy,Verbigeration,Negativism,Waxy flexibility | 87.2 | 84.1 | 90.1 |
| 5 | Posturing/catalepsy,Echopraxia/echolalia,Stereotypy,Rigidity,Withdrawal | 87.2 | 84.1 | 90.4 |
| 5 | Posturing/catalepsy,Grimacing,Stereotypy,Rigidity,Withdrawal | 87.2 | 84.1 | 90.1 |
| 5 | Posturing/catalepsy,Stereotypy,Verbigeration,Rigidity,Withdrawal | 87.2 | 84.1 | 90.4 |
| 5 | Verbigeration,Rigidity,Negativism,Waxy flexibility,Withdrawal | 87.2 | 84.1 | 90.1 |
| 5 | Echopraxia/echolalia,Rigidity,Negativism,Waxy flexibility,Withdrawal | 87.0 | 83.9 | 90.1 |
| 5 | Excitement,Echopraxia/echolalia,Stereotypy,Rigidity,Withdrawal | 87.0 | 83.9 | 89.9 |
| 5 | Excitement,Mutism,Grimacing,Verbigeration,Negativism | 87.0 | 83.6 | 90.1 |
| 5 | Excitement,Mutism,Grimacing,Verbigeration,Withdrawal | 87.0 | 83.9 | 89.9 |
| 5 | Excitement,Mutism,Mannerisms,Negativism,Waxy flexibility | 87.0 | 83.9 | 90.1 |
| 5 | Excitement,Mutism,Stereotypy,Mannerisms,Withdrawal | 87.0 | 83.9 | 90.1 |
| 5 | Excitement,Posturing/catalepsy,Grimacing,Mannerisms,Withdrawal | 87.0 | 83.6 | 89.9 |
| 5 | Excitement,Posturing/catalepsy,Grimacing,Rigidity,Negativism | 87.0 | 83.6 | 89.9 |
| 5 | Excitement,Posturing/catalepsy,Grimacing,Stereotypy,Withdrawal | 87.0 | 83.9 | 90.1 |
| 5 | Excitement,Posturing/catalepsy,Grimacing,Verbigeration,Withdrawal | 87.0 | 83.9 | 90.1 |
| 5 | Excitement,Posturing/catalepsy,Grimacing,Waxy flexibility,Withdrawal | 87.0 | 83.9 | 90.1 |
| 5 | Excitement,Posturing/catalepsy,Stereotypy,Waxy flexibility,Withdrawal | 87.0 | 83.9 | 89.9 |
| 5 | Excitement,Posturing/catalepsy,Verbigeration,Waxy flexibility,Withdrawal | 87.0 | 83.9 | 90.1 |
| 5 | Mutism,Grimacing,Echopraxia/echolalia,Stereotypy,Rigidity | 87.0 | 83.9 | 89.9 |
| 5 | Mutism,Grimacing,Echopraxia/echolalia,Verbigeration,Rigidity | 87.0 | 83.9 | 89.9 |
| 5 | Mutism,Grimacing,Stereotypy,Mannerisms,Negativism | 87.0 | 83.9 | 90.1 |
| 5 | Mutism,Stereotypy,Mannerisms,Verbigeration,Rigidity | 87.0 | 83.9 | 89.9 |
| 5 | Posturing/catalepsy,Echopraxia/echolalia,Negativism,Waxy flexibility,Withdrawal | 87.0 | 83.9 | 90.1 |
| 5 | Posturing/catalepsy,Echopraxia/echolalia,Verbigeration,Rigidity,Withdrawal | 87.0 | 83.9 | 90.1 |
| 5 | Posturing/catalepsy,Grimacing,Echopraxia/echolalia,Rigidity,Withdrawal | 87.0 | 83.9 | 89.9 |
| 5 | Posturing/catalepsy,Grimacing,Verbigeration,Rigidity,Withdrawal | 87.0 | 83.9 | 89.9 |
| 5 | Excitement,Echopraxia/echolalia,Rigidity,Waxy flexibility,Withdrawal | 86.8 | 83.6 | 89.9 |
| 5 | Excitement,Posturing/catalepsy,Echopraxia/echolalia,Mannerisms,Withdrawal | 86.8 | 83.6 | 89.9 |
| 5 | Excitement,Posturing/catalepsy,Echopraxia/echolalia,Stereotypy,Withdrawal | 86.8 | 83.6 | 89.9 |
| 5 | Excitement,Posturing/catalepsy,Mannerisms,Verbigeration,Withdrawal | 86.8 | 83.6 | 89.9 |
| 5 | Excitement,Posturing/catalepsy,Stereotypy,Rigidity,Negativism | 86.8 | 83.6 | 89.9 |
| 5 | Excitement,Posturing/catalepsy,Verbigeration,Rigidity,Negativism | 86.8 | 83.6 | 89.9 |
| 5 | Mutism,Echopraxia/echolalia,Stereotypy,Verbigeration,Negativism | 86.8 | 83.6 | 89.9 |
| 5 | Mutism,Echopraxia/echolalia,Stereotypy,Verbigeration,Rigidity | 86.8 | 83.6 | 89.7 |
| 5 | Mutism,Echopraxia/echolalia,Stereotypy,Waxy flexibility,Withdrawal | 86.8 | 83.6 | 89.9 |
| 5 | Mutism,Stereotypy,Mannerisms,Waxy flexibility,Withdrawal | 86.8 | 83.6 | 89.9 |
| 5 | Posturing/catalepsy,Stereotypy,Rigidity,Waxy flexibility,Withdrawal | 86.8 | 83.6 | 89.9 |
| 5 | Posturing/catalepsy,Verbigeration,Rigidity,Waxy flexibility,Withdrawal | 86.8 | 83.6 | 89.9 |
| 5 | Excitement,Posturing/catalepsy,Echopraxia/echolalia,Rigidity,Negativism | 86.7 | 83.4 | 89.9 |
| 5 | Excitement,Posturing/catalepsy,Stereotypy,Verbigeration,Withdrawal | 86.7 | 83.6 | 89.9 |
| 5 | Mutism,Grimacing,Echopraxia/echolalia,Mannerisms,Negativism | 86.7 | 83.6 | 89.9 |
| 5 | Mutism,Grimacing,Stereotypy,Verbigeration,Negativism | 86.7 | 83.4 | 89.7 |
| 5 | Mutism,Mannerisms,Verbigeration,Rigidity,Waxy flexibility | 86.7 | 83.4 | 89.9 |
| 5 | Mutism,Echopraxia/echolalia,Verbigeration,Rigidity,Waxy flexibility | 86.6 | 83.4 | 89.7 |
| 5 | Mutism,Grimacing,Mannerisms,Negativism,Waxy flexibility | 86.6 | 83.4 | 89.7 |
| 5 | Mutism,Grimacing,Stereotypy,Negativism,Waxy flexibility | 86.6 | 83.4 | 89.7 |
| 5 | Excitement,Mutism,Echopraxia/echolalia,Mannerisms,Negativism | 86.5 | 83.2 | 89.7 |
| 5 | Excitement,Posturing/catalepsy,Mannerisms,Waxy flexibility,Withdrawal | 86.5 | 83.4 | 89.7 |
| 5 | Excitement,Verbigeration,Rigidity,Waxy flexibility,Withdrawal | 86.5 | 83.4 | 89.7 |
| 5 | Mutism,Grimacing,Echopraxia/echolalia,Stereotypy,Negativism | 86.5 | 83.4 | 89.7 |
| 5 | Mutism,Grimacing,Mannerisms,Waxy flexibility,Withdrawal | 86.5 | 83.2 | 89.7 |
| 5 | Posturing/catalepsy,Grimacing,Negativism,Waxy flexibility,Withdrawal | 86.5 | 83.4 | 89.7 |
| 5 | Excitement,Mutism,Echopraxia/echolalia,Mannerisms,Withdrawal | 86.3 | 83.2 | 89.5 |
| 5 | Excitement,Posturing/catalepsy,Stereotypy,Mannerisms,Withdrawal | 86.3 | 83.0 | 89.2 |
| 5 | Excitement,Stereotypy,Verbigeration,Rigidity,Withdrawal | 86.3 | 83.2 | 89.5 |
| 5 | Grimacing,Rigidity,Negativism,Waxy flexibility,Withdrawal | 86.3 | 83.0 | 89.5 |
| 5 | Mutism,Echopraxia/echolalia,Mannerisms,Verbigeration,Negativism | 86.3 | 83.0 | 89.5 |
| 5 | Mutism,Echopraxia/echolalia,Mannerisms,Verbigeration,Rigidity | 86.3 | 83.2 | 89.5 |
| 5 | Mutism,Grimacing,Stereotypy,Waxy flexibility,Withdrawal | 86.3 | 83.2 | 89.5 |
| 5 | Mutism,Stereotypy,Verbigeration,Waxy flexibility,Withdrawal | 86.3 | 83.0 | 89.5 |
| 5 | Posturing/catalepsy,Echopraxia/echolalia,Rigidity,Waxy flexibility,Withdrawal | 86.3 | 83.0 | 89.5 |
| 5 | Posturing/catalepsy,Stereotypy,Mannerisms,Rigidity,Negativism | 86.3 | 83.2 | 89.5 |
| 5 | Posturing/catalepsy,Stereotypy,Verbigeration,Rigidity,Negativism | 86.3 | 83.2 | 89.5 |
| 5 | Excitement,Grimacing,Stereotypy,Rigidity,Withdrawal | 86.1 | 82.7 | 89.2 |
| 5 | Excitement,Mutism,Echopraxia/echolalia,Stereotypy,Waxy flexibility | 86.1 | 82.7 | 89.2 |
| 5 | Excitement,Mutism,Grimacing,Mannerisms,Withdrawal | 86.1 | 82.7 | 89.2 |
| 5 | Excitement,Mutism,Mannerisms,Verbigeration,Withdrawal | 86.1 | 82.7 | 89.2 |
| 5 | Excitement,Posturing/catalepsy,Mannerisms,Rigidity,Negativism | 86.1 | 83.0 | 89.2 |
| 5 | Mutism,Echopraxia/echolalia,Mannerisms,Waxy flexibility,Withdrawal | 86.1 | 82.7 | 89.2 |
| 5 | Mutism,Echopraxia/echolalia,Verbigeration,Negativism,Waxy flexibility | 86.1 | 82.7 | 89.2 |
| 5 | Mutism,Grimacing,Echopraxia/echolalia,Negativism,Waxy flexibility | 86.1 | 82.7 | 89.2 |
| 5 | Mutism,Grimacing,Echopraxia/echolalia,Stereotypy,Withdrawal | 86.1 | 82.7 | 89.2 |
| 5 | Mutism,Grimacing,Echopraxia/echolalia,Verbigeration,Negativism | 86.1 | 82.7 | 89.2 |
| 5 | Mutism,Grimacing,Echopraxia/echolalia,Waxy flexibility,Withdrawal | 86.1 | 82.7 | 89.2 |
| 5 | Mutism,Posturing/catalepsy,Grimacing,Stereotypy,Verbigeration | 86.1 | 82.7 | 89.2 |
| 5 | Posturing/catalepsy,Echopraxia/echolalia,Mannerisms,Rigidity,Negativism | 86.1 | 82.7 | 89.2 |
| 5 | Posturing/catalepsy,Grimacing,Mannerisms,Rigidity,Negativism | 86.1 | 83.0 | 89.2 |
| 5 | Posturing/catalepsy,Grimacing,Rigidity,Waxy flexibility,Withdrawal | 86.1 | 82.7 | 89.2 |
| 5 | Echopraxia/echolalia,Stereotypy,Mannerisms,Negativism,Withdrawal | 85.9 | 82.5 | 89.0 |
| 5 | Excitement,Echopraxia/echolalia,Stereotypy,Negativism,Withdrawal | 85.9 | 82.5 | 89.0 |
| 5 | Excitement,Echopraxia/echolalia,Verbigeration,Rigidity,Withdrawal | 85.9 | 82.5 | 89.0 |
| 5 | Excitement,Grimacing,Echopraxia/echolalia,Rigidity,Withdrawal | 85.9 | 82.5 | 89.0 |
| 5 | Excitement,Mutism,Grimacing,Mannerisms,Negativism | 85.9 | 82.7 | 89.0 |
| 5 | Excitement,Mutism,Mannerisms,Verbigeration,Negativism | 85.9 | 82.5 | 89.0 |
| 5 | Excitement,Posturing/catalepsy,Rigidity,Negativism,Waxy flexibility | 85.9 | 82.5 | 89.0 |
| 5 | Excitement,Stereotypy,Mannerisms,Rigidity,Withdrawal | 85.9 | 82.7 | 89.0 |
| 5 | Excitement,Stereotypy,Negativism,Waxy flexibility,Withdrawal | 85.9 | 82.5 | 89.0 |
| 5 | Mutism,Echopraxia/echolalia,Stereotypy,Mannerisms,Withdrawal | 85.9 | 82.7 | 89.0 |
| 5 | Mutism,Grimacing,Verbigeration,Waxy flexibility,Withdrawal | 85.9 | 82.7 | 89.0 |
| 5 | Mutism,Mannerisms,Verbigeration,Negativism,Waxy flexibility | 85.9 | 82.5 | 89.0 |
| 5 | Mutism,Posturing/catalepsy,Grimacing,Echopraxia/echolalia,Mannerisms | 85.9 | 82.5 | 89.0 |
| 5 | Mutism,Posturing/catalepsy,Grimacing,Echopraxia/echolalia,Verbigeration | 85.9 | 82.5 | 89.0 |
| 5 | Posturing/catalepsy,Grimacing,Stereotypy,Mannerisms,Withdrawal | 85.9 | 82.5 | 89.2 |
| 5 | Posturing/catalepsy,Mannerisms,Verbigeration,Rigidity,Negativism | 85.9 | 82.7 | 89.0 |
| 5 | Stereotypy,Mannerisms,Negativism,Waxy flexibility,Withdrawal | 85.8 | 82.5 | 89.0 |
| 5 | Excitement,Grimacing,Rigidity,Waxy flexibility,Withdrawal | 85.7 | 82.3 | 88.8 |
| 5 | Mutism,Grimacing,Mannerisms,Verbigeration,Negativism | 85.7 | 82.3 | 88.8 |
| 5 | Mutism,Grimacing,Stereotypy,Verbigeration,Withdrawal | 85.7 | 82.3 | 88.8 |
| 5 | Mutism,Posturing/catalepsy,Grimacing,Verbigeration,Waxy flexibility | 85.7 | 82.3 | 88.8 |
| 5 | Posturing/catalepsy,Grimacing,Echopraxia/echolalia,Stereotypy,Withdrawal | 85.7 | 82.3 | 88.8 |
| 5 | Stereotypy,Mannerisms,Rigidity,Waxy flexibility,Withdrawal | 85.7 | 82.3 | 89.0 |
| 5 | Echopraxia/echolalia,Stereotypy,Negativism,Waxy flexibility,Withdrawal | 85.6 | 82.3 | 88.8 |
| 5 | Excitement,Mutism,Grimacing,Echopraxia/echolalia,Waxy flexibility | 85.6 | 82.3 | 88.8 |
| 5 | Excitement,Mutism,Grimacing,Stereotypy,Waxy flexibility | 85.6 | 82.3 | 88.8 |
| 5 | Mutism,Grimacing,Stereotypy,Mannerisms,Withdrawal | 85.6 | 82.3 | 88.8 |
| 5 | Mutism,Grimacing,Verbigeration,Negativism,Waxy flexibility | 85.6 | 82.3 | 88.8 |
| 5 | Mutism,Mannerisms,Verbigeration,Waxy flexibility,Withdrawal | 85.6 | 82.3 | 88.8 |
| 5 | Mutism,Posturing/catalepsy,Grimacing,Echopraxia/echolalia,Stereotypy | 85.6 | 82.3 | 88.8 |
| 5 | Mutism,Posturing/catalepsy,Grimacing,Mannerisms,Verbigeration | 85.6 | 82.3 | 88.8 |
| 5 | Posturing/catalepsy,Grimacing,Stereotypy,Rigidity,Negativism | 85.6 | 82.3 | 88.8 |
| 5 | Posturing/catalepsy,Grimacing,Verbigeration,Rigidity,Negativism | 85.6 | 82.3 | 88.8 |
| 5 | Excitement,Grimacing,Verbigeration,Rigidity,Withdrawal | 85.4 | 82.1 | 88.6 |
| 5 | Excitement,Stereotypy,Verbigeration,Negativism,Withdrawal | 85.4 | 82.1 | 88.6 |
| 5 | Mutism,Echopraxia/echolalia,Verbigeration,Waxy flexibility,Withdrawal | 85.4 | 82.1 | 88.6 |
| 5 | Mutism,Grimacing,Echopraxia/echolalia,Mannerisms,Withdrawal | 85.4 | 82.1 | 88.6 |
| 5 | Mutism,Posturing/catalepsy,Echopraxia/echolalia,Mannerisms,Verbigeration | 85.4 | 82.1 | 88.6 |
| 5 | Mutism,Posturing/catalepsy,Echopraxia/echolalia,Stereotypy,Mannerisms | 85.4 | 82.1 | 88.6 |
| 5 | Mutism,Posturing/catalepsy,Echopraxia/echolalia,Stereotypy,Verbigeration | 85.4 | 82.1 | 88.6 |
| 5 | Mutism,Posturing/catalepsy,Grimacing,Stereotypy,Mannerisms | 85.4 | 82.1 | 88.6 |
| 5 | Mutism,Posturing/catalepsy,Stereotypy,Verbigeration,Waxy flexibility | 85.4 | 82.1 | 88.6 |
| 5 | Posturing/catalepsy,Echopraxia/echolalia,Stereotypy,Rigidity,Negativism | 85.4 | 82.1 | 88.6 |
| 5 | Posturing/catalepsy,Grimacing,Echopraxia/echolalia,Mannerisms,Withdrawal | 85.4 | 82.1 | 88.6 |
| 5 | Echopraxia/echolalia,Stereotypy,Rigidity,Waxy flexibility,Withdrawal | 85.2 | 81.8 | 88.3 |
| 5 | Excitement,Echopraxia/echolalia,Mannerisms,Rigidity,Withdrawal | 85.2 | 81.8 | 88.3 |
| 5 | Excitement,Echopraxia/echolalia,Negativism,Waxy flexibility,Withdrawal | 85.2 | 81.8 | 88.3 |
| 5 | Excitement,Mannerisms,Rigidity,Waxy flexibility,Withdrawal | 85.2 | 81.8 | 88.6 |
| 5 | Excitement,Mannerisms,Verbigeration,Rigidity,Withdrawal | 85.2 | 81.8 | 88.3 |
| 5 | Excitement,Mutism,Stereotypy,Verbigeration,Waxy flexibility | 85.2 | 81.8 | 88.3 |
| 5 | Grimacing,Stereotypy,Mannerisms,Rigidity,Withdrawal | 85.2 | 81.8 | 88.3 |
| 5 | Grimacing,Stereotypy,Rigidity,Waxy flexibility,Withdrawal | 85.2 | 81.8 | 88.3 |
| 5 | Mutism,Echopraxia/echolalia,Stereotypy,Verbigeration,Withdrawal | 85.2 | 81.8 | 88.3 |
| 5 | Mutism,Grimacing,Mannerisms,Verbigeration,Withdrawal | 85.2 | 81.8 | 88.6 |
| 5 | Mutism,Posturing/catalepsy,Echopraxia/echolalia,Stereotypy,Waxy flexibility | 85.2 | 81.8 | 88.3 |
| 5 | Mutism,Posturing/catalepsy,Stereotypy,Mannerisms,Verbigeration | 85.2 | 81.8 | 88.3 |
| 5 | Mutism,Stereotypy,Mannerisms,Verbigeration,Withdrawal | 85.2 | 81.8 | 88.6 |
| 5 | Posturing/catalepsy,Grimacing,Mannerisms,Verbigeration,Withdrawal | 85.2 | 81.8 | 88.3 |
| 5 | Posturing/catalepsy,Grimacing,Stereotypy,Verbigeration,Withdrawal | 85.2 | 81.8 | 88.6 |
| 5 | Posturing/catalepsy,Mannerisms,Rigidity,Negativism,Waxy flexibility | 85.2 | 81.8 | 88.3 |
| 5 | Posturing/catalepsy,Stereotypy,Rigidity,Negativism,Waxy flexibility | 85.2 | 81.8 | 88.3 |
| 5 | Stereotypy,Mannerisms,Verbigeration,Negativism,Withdrawal | 85.2 | 81.8 | 88.3 |
| 5 | Stereotypy,Verbigeration,Negativism,Waxy flexibility,Withdrawal | 85.2 | 81.8 | 88.3 |
| 5 | Stereotypy,Verbigeration,Rigidity,Waxy flexibility,Withdrawal | 85.2 | 81.8 | 88.6 |
| 5 | Echopraxia/echolalia,Stereotypy,Mannerisms,Rigidity,Withdrawal | 85.0 | 81.6 | 88.1 |
| 5 | Grimacing,Echopraxia/echolalia,Stereotypy,Negativism,Withdrawal | 85.0 | 81.6 | 88.1 |
| 5 | Grimacing,Echopraxia/echolalia,Stereotypy,Rigidity,Withdrawal | 85.0 | 81.6 | 88.1 |
| 5 | Grimacing,Stereotypy,Mannerisms,Negativism,Withdrawal | 85.0 | 81.6 | 88.1 |
| 5 | Mutism,Grimacing,Echopraxia/echolalia,Verbigeration,Withdrawal | 85.0 | 81.6 | 88.3 |
| 5 | Mutism,Posturing/catalepsy,Echopraxia/echolalia,Verbigeration,Waxy flexibility | 85.0 | 81.4 | 88.3 |
| 5 | Mutism,Posturing/catalepsy,Grimacing,Mannerisms,Waxy flexibility | 85.0 | 81.6 | 88.3 |
| 5 | Mutism,Posturing/catalepsy,Grimacing,Stereotypy,Waxy flexibility | 85.0 | 81.6 | 88.3 |
| 5 | Posturing/catalepsy,Echopraxia/echolalia,Verbigeration,Rigidity,Negativism | 85.0 | 81.6 | 88.3 |
| 5 | Posturing/catalepsy,Grimacing,Echopraxia/echolalia,Rigidity,Negativism | 85.0 | 81.6 | 88.1 |
| 5 | Mutism,Posturing/catalepsy,Echopraxia/echolalia,Mannerisms,Waxy flexibility | 84.9 | 81.6 | 88.1 |
| 5 | Mutism,Posturing/catalepsy,Grimacing,Echopraxia/echolalia,Waxy flexibility | 84.9 | 81.6 | 88.1 |
| 5 | Echopraxia/echolalia,Stereotypy,Verbigeration,Negativism,Withdrawal | 84.8 | 81.4 | 88.1 |
| 5 | Excitement,Mutism,Grimacing,Verbigeration,Waxy flexibility | 84.8 | 81.4 | 88.1 |
| 5 | Excitement,Mutism,Stereotypy,Mannerisms,Waxy flexibility | 84.8 | 81.4 | 87.9 |
| 5 | Grimacing,Stereotypy,Verbigeration,Rigidity,Withdrawal | 84.8 | 81.4 | 87.9 |
| 5 | Mutism,Posturing/catalepsy,Mannerisms,Verbigeration,Waxy flexibility | 84.8 | 81.4 | 88.1 |
| 5 | Posturing/catalepsy,Echopraxia/echolalia,Stereotypy,Mannerisms,Withdrawal | 84.8 | 81.4 | 87.9 |
| 5 | Posturing/catalepsy,Grimacing,Stereotypy,Waxy flexibility,Withdrawal | 84.8 | 81.4 | 87.9 |
| 5 | Posturing/catalepsy,Stereotypy,Mannerisms,Verbigeration,Withdrawal | 84.8 | 81.4 | 88.1 |
| 5 | Posturing/catalepsy,Stereotypy,Mannerisms,Waxy flexibility,Withdrawal | 84.8 | 81.4 | 88.1 |
| 5 | Excitement,Grimacing,Stereotypy,Negativism,Withdrawal | 84.7 | 81.4 | 87.9 |
| 5 | Excitement,Mutism,Echopraxia/echolalia,Verbigeration,Waxy flexibility | 84.6 | 80.9 | 87.9 |
| 5 | Mutism,Posturing/catalepsy,Stereotypy,Mannerisms,Waxy flexibility | 84.6 | 81.2 | 87.9 |
| 5 | Grimacing,Stereotypy,Negativism,Waxy flexibility,Withdrawal | 84.5 | 81.2 | 87.9 |
| 5 | Grimacing,Stereotypy,Verbigeration,Negativism,Withdrawal | 84.5 | 81.2 | 87.9 |
| 5 | Mutism,Echopraxia/echolalia,Mannerisms,Verbigeration,Withdrawal | 84.5 | 80.9 | 87.9 |
| 5 | Posturing/catalepsy,Grimacing,Echopraxia/echolalia,Verbigeration,Withdrawal | 84.5 | 81.2 | 87.9 |
| 5 | Posturing/catalepsy,Grimacing,Mannerisms,Waxy flexibility,Withdrawal | 84.5 | 80.9 | 87.9 |
| 5 | Stereotypy,Mannerisms,Verbigeration,Rigidity,Withdrawal | 84.5 | 81.2 | 87.7 |
| 5 | Echopraxia/echolalia,Stereotypy,Verbigeration,Rigidity,Withdrawal | 84.3 | 80.9 | 87.7 |
| 5 | Excitement,Grimacing,Echopraxia/echolalia,Negativism,Withdrawal | 84.3 | 80.9 | 87.7 |
| 5 | Excitement,Grimacing,Mannerisms,Rigidity,Withdrawal | 84.3 | 80.9 | 87.7 |
| 5 | Excitement,Posturing/catalepsy,Grimacing,Echopraxia/echolalia,Negativism | 84.3 | 80.7 | 87.7 |
| 5 | Excitement,Stereotypy,Mannerisms,Negativism,Withdrawal | 84.3 | 80.9 | 87.7 |
| 5 | Posturing/catalepsy,Echopraxia/echolalia,Mannerisms,Verbigeration,Withdrawal | 84.3 | 80.9 | 87.7 |
| 5 | Posturing/catalepsy,Grimacing,Rigidity,Negativism,Waxy flexibility | 84.3 | 80.9 | 87.7 |
| 5 | Echopraxia/echolalia,Mannerisms,Negativism,Waxy flexibility,Withdrawal | 84.1 | 80.5 | 87.4 |
| 5 | Excitement,Mutism,Echopraxia/echolalia,Mannerisms,Waxy flexibility | 84.1 | 80.7 | 87.4 |
| 5 | Excitement,Mutism,Grimacing,Echopraxia/echolalia,Stereotypy | 84.1 | 80.5 | 87.4 |
| 5 | Excitement,Mutism,Grimacing,Mannerisms,Waxy flexibility | 84.1 | 80.5 | 87.4 |
| 5 | Excitement,Verbigeration,Negativism,Waxy flexibility,Withdrawal | 84.1 | 80.5 | 87.4 |
| 5 | Grimacing,Echopraxia/echolalia,Mannerisms,Rigidity,Withdrawal | 84.1 | 80.5 | 87.4 |
| 5 | Grimacing,Echopraxia/echolalia,Rigidity,Waxy flexibility,Withdrawal | 84.1 | 80.7 | 87.4 |
| 5 | Grimacing,Mannerisms,Rigidity,Waxy flexibility,Withdrawal | 84.1 | 80.5 | 87.4 |
| 5 | Grimacing,Mannerisms,Verbigeration,Rigidity,Withdrawal | 84.1 | 80.7 | 87.4 |
| 5 | Grimacing,Verbigeration,Rigidity,Waxy flexibility,Withdrawal | 84.1 | 80.5 | 87.4 |
| 5 | Mannerisms,Verbigeration,Rigidity,Waxy flexibility,Withdrawal | 84.1 | 80.5 | 87.4 |
| 5 | Posturing/catalepsy,Echopraxia/echolalia,Stereotypy,Waxy flexibility,Withdrawal | 84.1 | 80.5 | 87.2 |
| 5 | Posturing/catalepsy,Grimacing,Echopraxia/echolalia,Waxy flexibility,Withdrawal | 84.1 | 80.5 | 87.4 |
| 5 | Posturing/catalepsy,Mannerisms,Verbigeration,Waxy flexibility,Withdrawal | 84.1 | 80.7 | 87.2 |
| 5 | Posturing/catalepsy,Stereotypy,Verbigeration,Waxy flexibility,Withdrawal | 84.1 | 80.7 | 87.4 |
| 5 | Posturing/catalepsy,Verbigeration,Rigidity,Negativism,Waxy flexibility | 84.1 | 80.5 | 87.4 |
| 5 | Excitement,Echopraxia/echolalia,Verbigeration,Negativism,Withdrawal | 84.0 | 80.7 | 87.4 |
| 5 | Excitement,Grimacing,Negativism,Waxy flexibility,Withdrawal | 83.9 | 80.5 | 87.2 |
| 5 | Excitement,Posturing/catalepsy,Grimacing,Stereotypy,Negativism | 83.9 | 80.3 | 87.2 |
| 5 | Excitement,Posturing/catalepsy,Grimacing,Verbigeration,Negativism | 83.9 | 80.3 | 87.2 |
| 5 | Posturing/catalepsy,Echopraxia/echolalia,Mannerisms,Waxy flexibility,Withdrawal | 83.9 | 80.3 | 87.2 |
| 5 | Posturing/catalepsy,Echopraxia/echolalia,Rigidity,Negativism,Waxy flexibility | 83.8 | 80.3 | 87.2 |
| 5 | Posturing/catalepsy,Echopraxia/echolalia,Stereotypy,Verbigeration,Withdrawal | 83.8 | 80.3 | 87.2 |
| 5 | Echopraxia/echolalia,Mannerisms,Rigidity,Waxy flexibility,Withdrawal | 83.6 | 80.3 | 87.0 |
| 5 | Echopraxia/echolalia,Verbigeration,Rigidity,Waxy flexibility,Withdrawal | 83.6 | 80.0 | 87.0 |
| 5 | Excitement,Mannerisms,Negativism,Waxy flexibility,Withdrawal | 83.6 | 80.0 | 87.0 |
| 5 | Excitement,Mutism,Echopraxia/echolalia,Stereotypy,Verbigeration | 83.6 | 80.0 | 87.0 |
| 5 | Excitement,Mutism,Grimacing,Stereotypy,Verbigeration | 83.6 | 80.0 | 87.0 |
| 5 | Grimacing,Echopraxia/echolalia,Verbigeration,Rigidity,Withdrawal | 83.6 | 80.3 | 87.0 |
| 5 | Posturing/catalepsy,Grimacing,Verbigeration,Waxy flexibility,Withdrawal | 83.6 | 80.0 | 87.0 |
| 5 | Excitement,Echopraxia/echolalia,Mannerisms,Negativism,Withdrawal | 83.4 | 79.8 | 86.8 |
| 5 | Excitement,Grimacing,Verbigeration,Negativism,Withdrawal | 83.4 | 79.8 | 86.8 |
| 5 | Excitement,Posturing/catalepsy,Grimacing,Negativism,Waxy flexibility | 83.4 | 80.0 | 86.8 |
| 5 | Excitement,Posturing/catalepsy,Stereotypy,Verbigeration,Negativism | 83.4 | 79.8 | 86.8 |
| 5 | Grimacing,Mannerisms,Negativism,Waxy flexibility,Withdrawal | 83.4 | 79.8 | 86.8 |
| 5 | Mannerisms,Verbigeration,Negativism,Waxy flexibility,Withdrawal | 83.4 | 79.8 | 86.8 |
| 5 | Mutism,Grimacing,Echopraxia/echolalia,Stereotypy,Waxy flexibility | 83.4 | 79.8 | 86.8 |
| 5 | Echopraxia/echolalia,Mannerisms,Verbigeration,Rigidity,Withdrawal | 83.2 | 79.6 | 86.5 |
| 5 | Echopraxia/echolalia,Verbigeration,Negativism,Waxy flexibility,Withdrawal | 83.2 | 79.6 | 86.5 |
| 5 | Excitement,Mutism,Echopraxia/echolalia,Stereotypy,Mannerisms | 83.2 | 79.6 | 86.5 |
| 5 | Excitement,Mutism,Grimacing,Echopraxia/echolalia,Verbigeration | 83.2 | 79.6 | 86.5 |
| 5 | Excitement,Posturing/catalepsy,Echopraxia/echolalia,Stereotypy,Negativism | 83.2 | 79.6 | 86.5 |
| 5 | Excitement,Posturing/catalepsy,Grimacing,Mannerisms,Negativism | 83.2 | 79.6 | 86.5 |
| 5 | Grimacing,Echopraxia/echolalia,Mannerisms,Negativism,Withdrawal | 83.2 | 79.6 | 86.5 |
| 5 | Grimacing,Echopraxia/echolalia,Negativism,Waxy flexibility,Withdrawal | 83.2 | 79.6 | 86.5 |
| 5 | Mutism,Echopraxia/echolalia,Stereotypy,Mannerisms,Waxy flexibility | 83.2 | 79.6 | 86.5 |
| 5 | Posturing/catalepsy,Echopraxia/echolalia,Verbigeration,Waxy flexibility,Withdrawal | 83.2 | 79.6 | 86.5 |
| 5 | Echopraxia/echolalia,Mannerisms,Verbigeration,Negativism,Withdrawal | 83.0 | 79.4 | 86.3 |
| 5 | Excitement,Echopraxia/echolalia,Stereotypy,Waxy flexibility,Withdrawal | 83.0 | 79.6 | 86.3 |
| 5 | Excitement,Mannerisms,Verbigeration,Negativism,Withdrawal | 83.0 | 79.4 | 86.3 |
| 5 | Excitement,Posturing/catalepsy,Grimacing,Echopraxia/echolalia,Rigidity | 83.0 | 79.4 | 86.3 |
| 5 | Excitement,Posturing/catalepsy,Stereotypy,Negativism,Waxy flexibility | 83.0 | 79.4 | 86.3 |
| 5 | Mutism,Grimacing,Stereotypy,Verbigeration,Waxy flexibility | 83.0 | 79.4 | 86.3 |
| 5 | Posturing/catalepsy,Grimacing,Stereotypy,Mannerisms,Negativism | 83.0 | 79.6 | 86.3 |
| 5 | Posturing/catalepsy,Grimacing,Stereotypy,Verbigeration,Negativism | 83.0 | 79.4 | 86.3 |
| 5 | Excitement,Mutism,Mannerisms,Verbigeration,Waxy flexibility | 82.9 | 79.4 | 86.3 |
| 5 | Grimacing,Mannerisms,Verbigeration,Negativism,Withdrawal | 82.9 | 79.4 | 86.3 |
| 5 | Mutism,Grimacing,Echopraxia/echolalia,Verbigeration,Waxy flexibility | 82.8 | 79.1 | 86.1 |
| 5 | Mutism,Grimacing,Stereotypy,Mannerisms,Waxy flexibility | 82.8 | 79.1 | 86.1 |
| 5 | Excitement,Grimacing,Mannerisms,Negativism,Withdrawal | 82.7 | 79.1 | 86.1 |
| 5 | Excitement,Posturing/catalepsy,Echopraxia/echolalia,Negativism,Waxy flexibility | 82.7 | 79.1 | 86.3 |
| 5 | Grimacing,Echopraxia/echolalia,Verbigeration,Negativism,Withdrawal | 82.7 | 79.1 | 86.1 |
| 5 | Mutism,Grimacing,Echopraxia/echolalia,Mannerisms,Waxy flexibility | 82.7 | 79.1 | 86.1 |
| 5 | Excitement,Mutism,Grimacing,Stereotypy,Mannerisms | 82.5 | 78.9 | 85.9 |
| 5 | Excitement,Mutism,Stereotypy,Mannerisms,Verbigeration | 82.5 | 78.9 | 86.1 |
| 5 | Excitement,Posturing/catalepsy,Echopraxia/echolalia,Verbigeration,Negativism | 82.5 | 78.9 | 85.9 |
| 5 | Excitement,Posturing/catalepsy,Grimacing,Verbigeration,Rigidity | 82.5 | 78.9 | 85.9 |
| 5 | Posturing/catalepsy,Grimacing,Echopraxia/echolalia,Mannerisms,Negativism | 82.5 | 78.9 | 86.1 |
| 5 | Posturing/catalepsy,Grimacing,Echopraxia/echolalia,Stereotypy,Negativism | 82.5 | 78.9 | 85.9 |
| 5 | Excitement,Grimacing,Echopraxia/echolalia,Waxy flexibility,Withdrawal | 82.3 | 78.7 | 85.7 |
| 5 | Excitement,Mutism,Grimacing,Echopraxia/echolalia,Mannerisms | 82.3 | 78.7 | 85.9 |
| 5 | Excitement,Posturing/catalepsy,Stereotypy,Mannerisms,Negativism | 82.3 | 78.7 | 85.7 |
| 5 | Excitement,Posturing/catalepsy,Verbigeration,Negativism,Waxy flexibility | 82.3 | 78.7 | 85.9 |
| 5 | Grimacing,Verbigeration,Negativism,Waxy flexibility,Withdrawal | 82.3 | 78.7 | 85.7 |
| 5 | Mutism,Echopraxia/echolalia,Stereotypy,Verbigeration,Waxy flexibility | 82.3 | 78.7 | 85.9 |
| 5 | Mutism,Stereotypy,Mannerisms,Verbigeration,Waxy flexibility | 82.3 | 78.7 | 85.9 |
| 5 | Posturing/catalepsy,Grimacing,Stereotypy,Negativism,Waxy flexibility | 82.3 | 78.7 | 85.9 |
| 5 | Excitement,Posturing/catalepsy,Echopraxia/echolalia,Mannerisms,Negativism | 82.1 | 78.3 | 85.4 |
| 5 | Excitement,Stereotypy,Verbigeration,Waxy flexibility,Withdrawal | 82.1 | 78.5 | 85.4 |
| 5 | Posturing/catalepsy,Echopraxia/echolalia,Stereotypy,Mannerisms,Negativism | 82.1 | 78.5 | 85.7 |
| 5 | Posturing/catalepsy,Grimacing,Echopraxia/echolalia,Verbigeration,Negativism | 82.1 | 78.5 | 85.4 |
| 5 | Posturing/catalepsy,Grimacing,Mannerisms,Verbigeration,Negativism | 82.1 | 78.5 | 85.7 |
| 5 | Stereotypy,Mannerisms,Rigidity,Negativism,Waxy flexibility | 82.1 | 78.5 | 85.7 |
| 5 | Excitement,Posturing/catalepsy,Grimacing,Rigidity,Waxy flexibility | 82.0 | 78.5 | 85.4 |
| 5 | Excitement,Stereotypy,Rigidity,Negativism,Waxy flexibility | 82.0 | 78.3 | 85.7 |
| 5 | Mutism,Grimacing,Echopraxia/echolalia,Stereotypy,Mannerisms | 82.0 | 78.5 | 85.4 |
| 5 | Excitement,Grimacing,Stereotypy,Waxy flexibility,Withdrawal | 81.8 | 78.3 | 85.4 |
| 5 | Excitement,Mutism,Grimacing,Mannerisms,Verbigeration | 81.8 | 78.3 | 85.4 |
| 5 | Excitement,Posturing/catalepsy,Mannerisms,Verbigeration,Negativism | 81.8 | 78.3 | 85.4 |
| 5 | Mutism,Grimacing,Echopraxia/echolalia,Stereotypy,Verbigeration | 81.8 | 78.3 | 85.2 |
| 5 | Mutism,Grimacing,Mannerisms,Verbigeration,Waxy flexibility | 81.8 | 78.3 | 85.4 |
| 5 | Posturing/catalepsy,Stereotypy,Mannerisms,Negativism,Waxy flexibility | 81.8 | 78.3 | 85.2 |
| 5 | Posturing/catalepsy,Stereotypy,Mannerisms,Verbigeration,Negativism | 81.8 | 78.3 | 85.2 |
| 5 | Posturing/catalepsy,Stereotypy,Verbigeration,Negativism,Waxy flexibility | 81.8 | 78.3 | 85.4 |
| 5 | Excitement,Echopraxia/echolalia,Verbigeration,Waxy flexibility,Withdrawal | 81.6 | 78.0 | 85.2 |
| 5 | Excitement,Posturing/catalepsy,Echopraxia/echolalia,Rigidity,Waxy flexibility | 81.6 | 78.0 | 85.2 |
| 5 | Excitement,Posturing/catalepsy,Echopraxia/echolalia,Verbigeration,Rigidity | 81.6 | 78.0 | 85.2 |
| 5 | Excitement,Posturing/catalepsy,Grimacing,Stereotypy,Rigidity | 81.6 | 78.0 | 85.2 |
| 5 | Excitement,Posturing/catalepsy,Mannerisms,Negativism,Waxy flexibility | 81.6 | 78.0 | 85.0 |
| 5 | Grimacing,Echopraxia/echolalia,Stereotypy,Waxy flexibility,Withdrawal | 81.6 | 78.0 | 85.2 |
| 5 | Grimacing,Stereotypy,Rigidity,Negativism,Waxy flexibility | 81.6 | 78.0 | 85.2 |
| 5 | Posturing/catalepsy,Echopraxia/echolalia,Stereotypy,Negativism,Waxy flexibility | 81.6 | 78.0 | 85.0 |
| 5 | Posturing/catalepsy,Grimacing,Mannerisms,Negativism,Waxy flexibility | 81.6 | 77.8 | 85.2 |
| 5 | Posturing/catalepsy,Grimacing,Verbigeration,Negativism,Waxy flexibility | 81.6 | 78.0 | 85.0 |
| 5 | Excitement,Echopraxia/echolalia,Stereotypy,Rigidity,Negativism | 81.4 | 77.8 | 85.0 |
| 5 | Excitement,Grimacing,Stereotypy,Rigidity,Negativism | 81.4 | 77.8 | 85.0 |
| 5 | Excitement,Mutism,Echopraxia/echolalia,Mannerisms,Verbigeration | 81.4 | 77.8 | 85.0 |
| 5 | Excitement,Posturing/catalepsy,Grimacing,Mannerisms,Rigidity | 81.4 | 77.8 | 85.0 |
| 5 | Grimacing,Stereotypy,Mannerisms,Rigidity,Negativism | 81.4 | 77.8 | 85.0 |
| 5 | Mutism,Echopraxia/echolalia,Mannerisms,Verbigeration,Waxy flexibility | 81.4 | 77.8 | 85.0 |
| 5 | Mutism,Grimacing,Stereotypy,Mannerisms,Verbigeration | 81.4 | 77.6 | 85.0 |
| 5 | Posturing/catalepsy,Echopraxia/echolalia,Stereotypy,Verbigeration,Negativism | 81.4 | 77.6 | 85.0 |
| 5 | Posturing/catalepsy,Grimacing,Echopraxia/echolalia,Negativism,Waxy flexibility | 81.4 | 77.8 | 85.0 |
| 5 | Stereotypy,Verbigeration,Rigidity,Negativism,Waxy flexibility | 81.4 | 77.8 | 85.0 |
| 5 | Excitement,Posturing/catalepsy,Echopraxia/echolalia,Stereotypy,Rigidity | 81.2 | 77.4 | 84.8 |
| 5 | Excitement,Posturing/catalepsy,Verbigeration,Rigidity,Waxy flexibility | 81.2 | 77.6 | 84.8 |
| 5 | Excitement,Stereotypy,Mannerisms,Waxy flexibility,Withdrawal | 81.2 | 77.4 | 84.8 |
| 5 | Mutism,Grimacing,Echopraxia/echolalia,Mannerisms,Verbigeration | 81.2 | 77.6 | 84.8 |
| 5 | Echopraxia/echolalia,Stereotypy,Mannerisms,Rigidity,Negativism | 81.0 | 77.1 | 84.5 |
| 5 | Echopraxia/echolalia,Stereotypy,Rigidity,Negativism,Waxy flexibility | 81.0 | 77.4 | 84.5 |
| 5 | Excitement,Posturing/catalepsy,Stereotypy,Verbigeration,Rigidity | 81.0 | 77.4 | 84.5 |
| 5 | Grimacing,Stereotypy,Verbigeration,Rigidity,Negativism | 81.0 | 77.1 | 84.5 |
| 5 | Excitement,Grimacing,Echopraxia/echolalia,Rigidity,Negativism | 80.9 | 77.1 | 84.5 |
| 5 | Excitement,Grimacing,Rigidity,Negativism,Waxy flexibility | 80.9 | 77.4 | 84.5 |
| 5 | Mutism,Echopraxia/echolalia,Stereotypy,Mannerisms,Verbigeration | 80.9 | 77.4 | 84.5 |
| 5 | Excitement,Echopraxia/echolalia,Mannerisms,Waxy flexibility,Withdrawal | 80.7 | 76.9 | 84.3 |
| 5 | Excitement,Posturing/catalepsy,Echopraxia/echolalia,Mannerisms,Rigidity | 80.7 | 76.9 | 84.5 |
| 5 | Excitement,Posturing/catalepsy,Stereotypy,Rigidity,Waxy flexibility | 80.7 | 76.9 | 84.3 |
| 5 | Excitement,Stereotypy,Verbigeration,Rigidity,Negativism | 80.7 | 77.1 | 84.3 |
| 5 | Grimacing,Stereotypy,Mannerisms,Waxy flexibility,Withdrawal | 80.7 | 76.9 | 84.3 |
| 5 | Posturing/catalepsy,Echopraxia/echolalia,Mannerisms,Negativism,Waxy flexibility | 80.7 | 76.9 | 84.3 |
| 5 | Stereotypy,Mannerisms,Verbigeration,Rigidity,Negativism | 80.7 | 77.1 | 84.3 |
| 5 | Excitement,Grimacing,Echopraxia/echolalia,Stereotypy,Withdrawal | 80.5 | 76.7 | 84.1 |
| 5 | Excitement,Grimacing,Verbigeration,Waxy flexibility,Withdrawal | 80.5 | 76.9 | 84.1 |
| 5 | Excitement,Posturing/catalepsy,Mannerisms,Verbigeration,Rigidity | 80.5 | 76.7 | 84.1 |
| 5 | Grimacing,Echopraxia/echolalia,Stereotypy,Rigidity,Negativism | 80.5 | 76.7 | 84.1 |
| 5 | Posturing/catalepsy,Echopraxia/echolalia,Mannerisms,Verbigeration,Negativism | 80.5 | 76.7 | 84.1 |
| 5 | Echopraxia/echolalia,Stereotypy,Mannerisms,Waxy flexibility,Withdrawal | 80.3 | 76.5 | 83.9 |
| 5 | Excitement,Echopraxia/echolalia,Stereotypy,Verbigeration,Withdrawal | 80.3 | 76.5 | 83.9 |
| 5 | Grimacing,Mannerisms,Rigidity,Negativism,Waxy flexibility | 80.3 | 76.5 | 83.9 |
| 5 | Posturing/catalepsy,Grimacing,Stereotypy,Verbigeration,Rigidity | 80.3 | 76.7 | 83.9 |
| 5 | Posturing/catalepsy,Mannerisms,Verbigeration,Negativism,Waxy flexibility | 80.2 | 76.5 | 83.9 |
| 5 | Excitement,Mannerisms,Verbigeration,Waxy flexibility,Withdrawal | 80.1 | 76.2 | 83.6 |
| 5 | Grimacing,Echopraxia/echolalia,Mannerisms,Rigidity,Negativism | 80.1 | 76.2 | 83.6 |
| 5 | Posturing/catalepsy,Grimacing,Echopraxia/echolalia,Mannerisms,Rigidity | 80.1 | 76.2 | 83.6 |
| 5 | Posturing/catalepsy,Grimacing,Echopraxia/echolalia,Verbigeration,Rigidity | 80.1 | 76.2 | 83.6 |
| 5 | Excitement,Echopraxia/echolalia,Rigidity,Negativism,Waxy flexibility | 80.0 | 76.2 | 83.6 |
| 5 | Excitement,Stereotypy,Mannerisms,Rigidity,Negativism | 80.0 | 76.2 | 83.6 |
| 5 | Grimacing,Stereotypy,Verbigeration,Waxy flexibility,Withdrawal | 80.0 | 76.2 | 83.6 |
| 5 | Posturing/catalepsy,Echopraxia/echolalia,Verbigeration,Negativism,Waxy flexibility | 80.0 | 76.2 | 83.6 |
| 5 | Posturing/catalepsy,Grimacing,Mannerisms,Verbigeration,Rigidity | 79.9 | 76.2 | 83.4 |
| 5 | Echopraxia/echolalia,Stereotypy,Verbigeration,Rigidity,Negativism | 79.8 | 76.0 | 83.6 |
| 5 | Excitement,Grimacing,Echopraxia/echolalia,Verbigeration,Withdrawal | 79.8 | 76.0 | 83.4 |
| 5 | Excitement,Grimacing,Verbigeration,Rigidity,Negativism | 79.8 | 76.0 | 83.4 |
| 5 | Excitement,Posturing/catalepsy,Mannerisms,Rigidity,Waxy flexibility | 79.8 | 76.0 | 83.6 |
| 5 | Grimacing,Echopraxia/echolalia,Mannerisms,Waxy flexibility,Withdrawal | 79.8 | 76.0 | 83.6 |
| 5 | Grimacing,Echopraxia/echolalia,Rigidity,Negativism,Waxy flexibility | 79.8 | 76.0 | 83.6 |
| 5 | Grimacing,Echopraxia/echolalia,Stereotypy,Mannerisms,Withdrawal | 79.8 | 76.0 | 83.4 |
| 5 | Echopraxia/echolalia,Stereotypy,Verbigeration,Waxy flexibility,Withdrawal | 79.6 | 75.8 | 83.2 |
| 5 | Excitement,Echopraxia/echolalia,Stereotypy,Mannerisms,Withdrawal | 79.6 | 75.8 | 83.2 |
| 5 | Excitement,Grimacing,Mannerisms,Waxy flexibility,Withdrawal | 79.6 | 75.8 | 83.2 |
| 5 | Excitement,Grimacing,Stereotypy,Verbigeration,Withdrawal | 79.6 | 75.8 | 83.2 |
| 5 | Excitement,Posturing/catalepsy,Stereotypy,Mannerisms,Rigidity | 79.6 | 75.8 | 83.2 |
| 5 | Grimacing,Echopraxia/echolalia,Verbigeration,Waxy flexibility,Withdrawal | 79.6 | 75.8 | 83.2 |
| 5 | Grimacing,Verbigeration,Rigidity,Negativism,Waxy flexibility | 79.6 | 75.8 | 83.2 |
| 5 | Posturing/catalepsy,Grimacing,Echopraxia/echolalia,Stereotypy,Rigidity | 79.6 | 75.8 | 83.4 |
| 5 | Posturing/catalepsy,Grimacing,Stereotypy,Mannerisms,Rigidity | 79.6 | 75.8 | 83.2 |
| 5 | Posturing/catalepsy,Grimacing,Verbigeration,Rigidity,Waxy flexibility | 79.6 | 75.8 | 83.2 |
| 5 | Echopraxia/echolalia,Mannerisms,Rigidity,Negativism,Waxy flexibility | 79.4 | 75.6 | 83.0 |
| 5 | Excitement,Grimacing,Mannerisms,Rigidity,Negativism | 79.4 | 75.6 | 83.0 |
| 5 | Grimacing,Echopraxia/echolalia,Stereotypy,Verbigeration,Withdrawal | 79.4 | 75.6 | 83.2 |
| 5 | Stereotypy,Mannerisms,Verbigeration,Waxy flexibility,Withdrawal | 79.4 | 75.6 | 83.0 |
| 5 | Posturing/catalepsy,Grimacing,Echopraxia/echolalia,Rigidity,Waxy flexibility | 79.3 | 75.6 | 83.2 |
| 5 | Grimacing,Mannerisms,Verbigeration,Rigidity,Negativism | 79.2 | 75.3 | 83.0 |
| 5 | Posturing/catalepsy,Grimacing,Mannerisms,Rigidity,Waxy flexibility | 79.2 | 75.3 | 83.0 |
| 5 | Excitement,Verbigeration,Rigidity,Negativism,Waxy flexibility | 79.1 | 75.3 | 83.0 |
| 5 | Posturing/catalepsy,Grimacing,Stereotypy,Rigidity,Waxy flexibility | 79.1 | 75.3 | 82.7 |
| 5 | Excitement,Stereotypy,Mannerisms,Verbigeration,Withdrawal | 78.9 | 75.1 | 82.7 |
| 5 | Grimacing,Echopraxia/echolalia,Verbigeration,Rigidity,Negativism | 78.9 | 75.1 | 82.7 |
| 5 | Grimacing,Mannerisms,Verbigeration,Waxy flexibility,Withdrawal | 78.9 | 75.1 | 82.7 |
| 5 | Excitement,Grimacing,Echopraxia/echolalia,Mannerisms,Withdrawal | 78.7 | 74.9 | 82.3 |
| 5 | Excitement,Grimacing,Stereotypy,Mannerisms,Withdrawal | 78.7 | 74.9 | 82.5 |
| 5 | Excitement,Mannerisms,Rigidity,Negativism,Waxy flexibility | 78.7 | 74.9 | 82.3 |
| 5 | Grimacing,Stereotypy,Mannerisms,Verbigeration,Withdrawal | 78.7 | 74.9 | 82.5 |
| 5 | Mannerisms,Verbigeration,Rigidity,Negativism,Waxy flexibility | 78.7 | 74.9 | 82.3 |
| 5 | Posturing/catalepsy,Echopraxia/echolalia,Stereotypy,Verbigeration,Rigidity | 78.7 | 74.9 | 82.5 |
| 5 | Excitement,Echopraxia/echolalia,Mannerisms,Rigidity,Negativism | 78.5 | 74.7 | 82.3 |
| 5 | Excitement,Echopraxia/echolalia,Verbigeration,Rigidity,Negativism | 78.5 | 74.7 | 82.3 |
| 5 | Posturing/catalepsy,Stereotypy,Mannerisms,Verbigeration,Rigidity | 78.5 | 74.7 | 82.3 |
| 5 | Posturing/catalepsy,Stereotypy,Verbigeration,Rigidity,Waxy flexibility | 78.5 | 74.7 | 82.1 |
| 5 | Excitement,Echopraxia/echolalia,Mannerisms,Verbigeration,Withdrawal | 78.3 | 74.2 | 82.1 |
| 5 | Posturing/catalepsy,Echopraxia/echolalia,Stereotypy,Mannerisms,Rigidity | 78.3 | 74.4 | 82.1 |
| 5 | Excitement,Grimacing,Mannerisms,Verbigeration,Withdrawal | 78.2 | 74.4 | 82.1 |
| 5 | Excitement,Posturing/catalepsy,Grimacing,Echopraxia/echolalia,Waxy flexibility | 78.2 | 74.4 | 82.1 |
| 5 | Posturing/catalepsy,Echopraxia/echolalia,Mannerisms,Verbigeration,Rigidity | 78.2 | 74.4 | 81.8 |
| 5 | Posturing/catalepsy,Echopraxia/echolalia,Stereotypy,Rigidity,Waxy flexibility | 78.2 | 74.4 | 81.8 |
| 5 | Echopraxia/echolalia,Mannerisms,Verbigeration,Waxy flexibility,Withdrawal | 78.0 | 74.2 | 81.8 |
| 5 | Grimacing,Echopraxia/echolalia,Mannerisms,Verbigeration,Withdrawal | 78.0 | 74.2 | 81.6 |
| 5 | Echopraxia/echolalia,Mannerisms,Verbigeration,Rigidity,Negativism | 77.8 | 74.0 | 81.6 |
| 5 | Echopraxia/echolalia,Stereotypy,Mannerisms,Verbigeration,Withdrawal | 77.8 | 73.8 | 81.6 |
| 5 | Echopraxia/echolalia,Verbigeration,Rigidity,Negativism,Waxy flexibility | 77.8 | 73.8 | 81.6 |
| 5 | Posturing/catalepsy,Echopraxia/echolalia,Mannerisms,Rigidity,Waxy flexibility | 77.8 | 73.8 | 81.6 |
| 5 | Excitement,Mannerisms,Verbigeration,Rigidity,Negativism | 77.6 | 73.8 | 81.4 |
| 5 | Excitement,Posturing/catalepsy,Grimacing,Echopraxia/echolalia,Verbigeration | 77.6 | 73.5 | 81.4 |
| 5 | Posturing/catalepsy,Echopraxia/echolalia,Verbigeration,Rigidity,Waxy flexibility | 77.6 | 73.8 | 81.4 |
| 5 | Posturing/catalepsy,Mannerisms,Verbigeration,Rigidity,Waxy flexibility | 77.6 | 73.8 | 81.4 |
| 5 | Posturing/catalepsy,Stereotypy,Mannerisms,Rigidity,Waxy flexibility | 77.6 | 73.5 | 81.4 |
| 5 | Excitement,Posturing/catalepsy,Grimacing,Verbigeration,Waxy flexibility | 77.4 | 73.3 | 81.2 |
| 5 | Excitement,Posturing/catalepsy,Grimacing,Echopraxia/echolalia,Stereotypy | 77.1 | 73.1 | 80.9 |
| 5 | Excitement,Posturing/catalepsy,Grimacing,Stereotypy,Verbigeration | 77.1 | 73.1 | 80.9 |
| 5 | Excitement,Grimacing,Stereotypy,Rigidity,Waxy flexibility | 76.9 | 72.9 | 80.7 |
| 5 | Excitement,Posturing/catalepsy,Grimacing,Echopraxia/echolalia,Mannerisms | 76.9 | 72.9 | 80.7 |
| 5 | Excitement,Posturing/catalepsy,Grimacing,Stereotypy,Waxy flexibility | 76.9 | 72.9 | 80.7 |
| 5 | Excitement,Grimacing,Echopraxia/echolalia,Rigidity,Waxy flexibility | 76.5 | 72.4 | 80.3 |
| 5 | Excitement,Grimacing,Stereotypy,Negativism,Waxy flexibility | 76.5 | 72.4 | 80.3 |
| 5 | Grimacing,Stereotypy,Mannerisms,Negativism,Waxy flexibility | 76.4 | 72.4 | 80.3 |
| 5 | Excitement,Posturing/catalepsy,Grimacing,Mannerisms,Verbigeration | 76.2 | 72.2 | 80.0 |
| 5 | Excitement,Posturing/catalepsy,Grimacing,Mannerisms,Waxy flexibility | 76.0 | 72.0 | 80.0 |
| 5 | Echopraxia/echolalia,Stereotypy,Mannerisms,Negativism,Waxy flexibility | 75.8 | 72.0 | 79.6 |
| 5 | Excitement,Echopraxia/echolalia,Stereotypy,Negativism,Waxy flexibility | 75.8 | 71.7 | 79.8 |
| 5 | Grimacing,Echopraxia/echolalia,Stereotypy,Negativism,Waxy flexibility | 75.7 | 71.7 | 79.6 |
| 5 | Excitement,Grimacing,Echopraxia/echolalia,Negativism,Waxy flexibility | 75.6 | 71.5 | 79.6 |
| 5 | Excitement,Posturing/catalepsy,Grimacing,Stereotypy,Mannerisms | 75.6 | 71.5 | 79.4 |
| 5 | Excitement,Echopraxia/echolalia,Stereotypy,Rigidity,Waxy flexibility | 75.4 | 71.3 | 79.4 |
| 5 | Excitement,Grimacing,Echopraxia/echolalia,Stereotypy,Rigidity | 75.3 | 71.3 | 79.4 |
| 5 | Excitement,Grimacing,Verbigeration,Rigidity,Waxy flexibility | 75.3 | 71.3 | 79.4 |
| 5 | Excitement,Posturing/catalepsy,Stereotypy,Verbigeration,Waxy flexibility | 75.3 | 71.3 | 79.1 |
| 5 | Excitement,Stereotypy,Verbigeration,Negativism,Waxy flexibility | 75.3 | 71.3 | 79.4 |
| 5 | Grimacing,Stereotypy,Verbigeration,Negativism,Waxy flexibility | 75.3 | 71.3 | 79.1 |
| 5 | Posturing/catalepsy,Grimacing,Stereotypy,Verbigeration,Waxy flexibility | 75.3 | 71.3 | 79.1 |
| 5 | Excitement,Posturing/catalepsy,Echopraxia/echolalia,Stereotypy,Waxy flexibility | 75.1 | 71.1 | 79.1 |
| 5 | Excitement,Posturing/catalepsy,Echopraxia/echolalia,Verbigeration,Waxy flexibility | 75.1 | 71.1 | 79.1 |
| 5 | Grimacing,Echopraxia/echolalia,Stereotypy,Rigidity,Waxy flexibility | 75.1 | 71.1 | 79.1 |
| 5 | Grimacing,Stereotypy,Verbigeration,Rigidity,Waxy flexibility | 75.1 | 70.9 | 78.9 |
| 5 | Posturing/catalepsy,Grimacing,Echopraxia/echolalia,Stereotypy,Waxy flexibility | 75.1 | 71.1 | 79.1 |
| 5 | Grimacing,Stereotypy,Mannerisms,Rigidity,Waxy flexibility | 74.9 | 70.9 | 78.9 |
| 5 | Posturing/catalepsy,Grimacing,Echopraxia/echolalia,Stereotypy,Verbigeration | 74.9 | 70.9 | 78.7 |
| 5 | Excitement,Grimacing,Echopraxia/echolalia,Stereotypy,Negativism | 74.7 | 70.6 | 78.7 |
| 5 | Excitement,Posturing/catalepsy,Echopraxia/echolalia,Stereotypy,Verbigeration | 74.7 | 70.6 | 78.7 |
| 5 | Excitement,Stereotypy,Mannerisms,Negativism,Waxy flexibility | 74.7 | 70.6 | 78.7 |
| 5 | Posturing/catalepsy,Grimacing,Echopraxia/echolalia,Stereotypy,Mannerisms | 74.7 | 70.6 | 78.7 |
| 5 | Stereotypy,Mannerisms,Verbigeration,Negativism,Waxy flexibility | 74.7 | 70.6 | 78.7 |
| 5 | Grimacing,Echopraxia/echolalia,Stereotypy,Mannerisms,Negativism | 74.6 | 70.4 | 78.7 |
| 5 | Posturing/catalepsy,Grimacing,Stereotypy,Mannerisms,Verbigeration | 74.6 | 70.6 | 78.7 |
| 5 | Excitement,Stereotypy,Verbigeration,Rigidity,Waxy flexibility | 74.5 | 70.2 | 78.5 |
| 5 | Posturing/catalepsy,Grimacing,Echopraxia/echolalia,Verbigeration,Waxy flexibility | 74.5 | 70.4 | 78.5 |
| 5 | Excitement,Grimacing,Stereotypy,Verbigeration,Rigidity | 74.4 | 70.2 | 78.5 |
| 5 | Grimacing,Echopraxia/echolalia,Mannerisms,Negativism,Waxy flexibility | 74.4 | 70.2 | 78.5 |
| 5 | Posturing/catalepsy,Grimacing,Echopraxia/echolalia,Mannerisms,Verbigeration | 74.4 | 70.4 | 78.5 |
| 5 | Excitement,Grimacing,Echopraxia/echolalia,Verbigeration,Rigidity | 74.2 | 70.2 | 78.3 |
| 5 | Excitement,Grimacing,Verbigeration,Negativism,Waxy flexibility | 74.0 | 69.7 | 78.0 |
| 5 | Posturing/catalepsy,Grimacing,Mannerisms,Verbigeration,Waxy flexibility | 74.0 | 70.0 | 78.0 |
| 5 | Posturing/catalepsy,Grimacing,Stereotypy,Mannerisms,Waxy flexibility | 74.0 | 69.7 | 78.0 |
| 5 | Echopraxia/echolalia,Stereotypy,Verbigeration,Negativism,Waxy flexibility | 73.8 | 69.7 | 77.8 |
| 5 | Excitement,Grimacing,Mannerisms,Negativism,Waxy flexibility | 73.8 | 69.5 | 77.8 |
| 5 | Excitement,Grimacing,Stereotypy,Verbigeration,Negativism | 73.8 | 69.7 | 77.8 |
| 5 | Excitement,Posturing/catalepsy,Stereotypy,Mannerisms,Verbigeration | 73.8 | 69.7 | 77.8 |
| 5 | Grimacing,Echopraxia/echolalia,Verbigeration,Rigidity,Waxy flexibility | 73.8 | 69.7 | 77.8 |
| 5 | Excitement,Posturing/catalepsy,Echopraxia/echolalia,Mannerisms,Waxy flexibility | 73.7 | 69.7 | 77.8 |
| 5 | Posturing/catalepsy,Grimacing,Echopraxia/echolalia,Mannerisms,Waxy flexibility | 73.7 | 69.5 | 77.8 |
| 5 | Excitement,Grimacing,Stereotypy,Mannerisms,Rigidity | 73.6 | 69.5 | 77.6 |
| 5 | Excitement,Posturing/catalepsy,Echopraxia/echolalia,Stereotypy,Mannerisms | 73.6 | 69.3 | 77.6 |
| 5 | Excitement,Posturing/catalepsy,Echopraxia/echolalia,Mannerisms,Verbigeration | 73.5 | 69.3 | 77.6 |
| 5 | Excitement,Grimacing,Mannerisms,Rigidity,Waxy flexibility | 73.3 | 69.3 | 77.4 |
| 5 | Excitement,Posturing/catalepsy,Mannerisms,Verbigeration,Waxy flexibility | 73.3 | 69.3 | 77.4 |
| 5 | Grimacing,Echopraxia/echolalia,Mannerisms,Rigidity,Waxy flexibility | 73.3 | 69.3 | 77.4 |
| 5 | Grimacing,Echopraxia/echolalia,Stereotypy,Mannerisms,Rigidity | 73.3 | 69.3 | 77.4 |
| 5 | Grimacing,Echopraxia/echolalia,Stereotypy,Verbigeration,Negativism | 73.3 | 69.3 | 77.4 |
| 5 | Grimacing,Echopraxia/echolalia,Verbigeration,Negativism,Waxy flexibility | 73.3 | 69.1 | 77.4 |
| 5 | Grimacing,Stereotypy,Mannerisms,Verbigeration,Negativism | 73.3 | 69.1 | 77.4 |
| 5 | Excitement,Grimacing,Echopraxia/echolalia,Mannerisms,Rigidity | 73.1 | 68.8 | 77.1 |
| 5 | Excitement,Grimacing,Stereotypy,Mannerisms,Negativism | 73.1 | 69.1 | 77.1 |
| 5 | Excitement,Posturing/catalepsy,Stereotypy,Mannerisms,Waxy flexibility | 73.1 | 68.8 | 77.1 |
| 5 | Excitement,Stereotypy,Mannerisms,Rigidity,Waxy flexibility | 73.1 | 68.8 | 77.1 |
| 5 | Grimacing,Echopraxia/echolalia,Stereotypy,Verbigeration,Rigidity | 73.1 | 68.8 | 77.1 |
| 5 | Excitement,Echopraxia/echolalia,Stereotypy,Mannerisms,Negativism | 72.9 | 68.6 | 76.9 |
| 5 | Excitement,Echopraxia/echolalia,Stereotypy,Verbigeration,Negativism | 72.9 | 68.6 | 77.1 |
| 5 | Grimacing,Stereotypy,Mannerisms,Verbigeration,Rigidity | 72.9 | 68.6 | 76.9 |
| 5 | Excitement,Echopraxia/echolalia,Stereotypy,Verbigeration,Rigidity | 72.6 | 68.4 | 76.7 |
| 5 | Excitement,Echopraxia/echolalia,Verbigeration,Rigidity,Waxy flexibility | 72.6 | 68.6 | 76.7 |
| 5 | Grimacing,Mannerisms,Verbigeration,Negativism,Waxy flexibility | 72.6 | 68.4 | 76.7 |
| 5 | Grimacing,Mannerisms,Verbigeration,Rigidity,Waxy flexibility | 72.6 | 68.4 | 76.7 |
| 5 | Echopraxia/echolalia,Stereotypy,Mannerisms,Rigidity,Waxy flexibility | 72.4 | 68.2 | 76.5 |
| 5 | Excitement,Grimacing,Echopraxia/echolalia,Verbigeration,Negativism | 72.4 | 68.2 | 76.5 |
| 5 | Echopraxia/echolalia,Stereotypy,Mannerisms,Verbigeration,Negativism | 72.2 | 67.9 | 76.2 |
| 5 | Excitement,Echopraxia/echolalia,Mannerisms,Negativism,Waxy flexibility | 72.2 | 67.9 | 76.2 |
| 5 | Excitement,Echopraxia/echolalia,Stereotypy,Mannerisms,Rigidity | 72.2 | 67.9 | 76.2 |
| 5 | Excitement,Grimacing,Mannerisms,Verbigeration,Rigidity | 72.2 | 67.9 | 76.5 |
| 5 | Echopraxia/echolalia,Stereotypy,Verbigeration,Rigidity,Waxy flexibility | 72.0 | 67.9 | 76.0 |
| 5 | Excitement,Echopraxia/echolalia,Verbigeration,Negativism,Waxy flexibility | 72.0 | 67.7 | 76.0 |
| 5 | Excitement,Grimacing,Echopraxia/echolalia,Mannerisms,Negativism | 72.0 | 67.7 | 76.0 |
| 5 | Posturing/catalepsy,Echopraxia/echolalia,Stereotypy,Verbigeration,Waxy flexibility | 72.0 | 67.9 | 76.0 |
| 5 | Posturing/catalepsy,Stereotypy,Mannerisms,Verbigeration,Waxy flexibility | 71.8 | 67.5 | 75.8 |
| 5 | Excitement,Stereotypy,Mannerisms,Verbigeration,Negativism | 71.7 | 67.5 | 76.0 |
| 5 | Stereotypy,Mannerisms,Verbigeration,Rigidity,Waxy flexibility | 71.7 | 67.5 | 75.8 |
| 5 | Excitement,Echopraxia/echolalia,Mannerisms,Rigidity,Waxy flexibility | 71.5 | 67.3 | 75.8 |
| 5 | Grimacing,Echopraxia/echolalia,Mannerisms,Verbigeration,Rigidity | 71.5 | 67.3 | 75.6 |
| 5 | Posturing/catalepsy,Echopraxia/echolalia,Stereotypy,Mannerisms,Waxy flexibility | 71.5 | 67.3 | 75.8 |
| 5 | Posturing/catalepsy,Echopraxia/echolalia,Stereotypy,Mannerisms,Verbigeration | 71.3 | 67.0 | 75.6 |
| 5 | Excitement,Stereotypy,Mannerisms,Verbigeration,Rigidity | 71.1 | 66.8 | 75.3 |
| 5 | Grimacing,Echopraxia/echolalia,Mannerisms,Verbigeration,Negativism | 71.0 | 66.8 | 75.1 |
| 5 | Excitement,Mannerisms,Verbigeration,Negativism,Waxy flexibility | 70.8 | 66.6 | 75.1 |
| 5 | Echopraxia/echolalia,Mannerisms,Verbigeration,Negativism,Waxy flexibility | 70.7 | 66.4 | 74.9 |
| 5 | Excitement,Grimacing,Mannerisms,Verbigeration,Negativism | 70.6 | 66.4 | 74.7 |
| 5 | Excitement,Mannerisms,Verbigeration,Rigidity,Waxy flexibility | 70.4 | 66.1 | 74.7 |
| 5 | Posturing/catalepsy,Echopraxia/echolalia,Mannerisms,Verbigeration,Waxy flexibility | 70.4 | 66.1 | 74.7 |
| 5 | Echopraxia/echolalia,Stereotypy,Mannerisms,Verbigeration,Rigidity | 69.9 | 65.7 | 74.2 |
| 5 | Excitement,Echopraxia/echolalia,Mannerisms,Verbigeration,Rigidity | 69.5 | 65.2 | 73.5 |
| 5 | Echopraxia/echolalia,Mannerisms,Verbigeration,Rigidity,Waxy flexibility | 69.1 | 64.8 | 73.3 |
| 5 | Excitement,Echopraxia/echolalia,Mannerisms,Verbigeration,Negativism | 68.4 | 63.9 | 72.6 |
| 5 | Excitement,Grimacing,Echopraxia/echolalia,Stereotypy,Waxy flexibility | 67.9 | 63.7 | 72.2 |
| 5 | Excitement,Grimacing,Stereotypy,Verbigeration,Waxy flexibility | 67.1 | 62.6 | 71.3 |
| 5 | Excitement,Grimacing,Echopraxia/echolalia,Verbigeration,Waxy flexibility | 66.1 | 61.7 | 70.4 |
| 5 | Grimacing,Echopraxia/echolalia,Stereotypy,Mannerisms,Waxy flexibility | 65.9 | 61.4 | 70.2 |
| 5 | Grimacing,Echopraxia/echolalia,Stereotypy,Verbigeration,Waxy flexibility | 65.9 | 61.4 | 70.4 |
| 5 | Excitement,Grimacing,Stereotypy,Mannerisms,Waxy flexibility | 65.5 | 61.0 | 69.7 |
| 5 | Grimacing,Stereotypy,Mannerisms,Verbigeration,Waxy flexibility | 65.0 | 60.5 | 69.5 |
| 5 | Excitement,Grimacing,Echopraxia/echolalia,Mannerisms,Waxy flexibility | 64.4 | 59.9 | 68.8 |
| 5 | Excitement,Echopraxia/echolalia,Stereotypy,Verbigeration,Waxy flexibility | 64.1 | 59.6 | 68.6 |
| 5 | Excitement,Echopraxia/echolalia,Stereotypy,Mannerisms,Waxy flexibility | 63.2 | 58.7 | 67.7 |
| 5 | Excitement,Grimacing,Mannerisms,Verbigeration,Waxy flexibility | 63.2 | 58.7 | 67.5 |
| 5 | Grimacing,Echopraxia/echolalia,Mannerisms,Verbigeration,Waxy flexibility | 62.8 | 58.1 | 67.3 |
| 5 | Excitement,Stereotypy,Mannerisms,Verbigeration,Waxy flexibility | 62.3 | 57.8 | 66.8 |
| 5 | Excitement,Grimacing,Echopraxia/echolalia,Stereotypy,Verbigeration | 62.1 | 57.6 | 66.6 |
| 5 | Excitement,Grimacing,Echopraxia/echolalia,Stereotypy,Mannerisms | 61.2 | 56.7 | 65.7 |
| 5 | Echopraxia/echolalia,Stereotypy,Mannerisms,Verbigeration,Waxy flexibility | 60.8 | 56.3 | 65.2 |
| 5 | Grimacing,Echopraxia/echolalia,Stereotypy,Mannerisms,Verbigeration | 60.3 | 55.8 | 64.8 |
| 5 | Excitement,Grimacing,Stereotypy,Mannerisms,Verbigeration | 60.1 | 55.6 | 64.6 |
| 5 | Excitement,Echopraxia/echolalia,Mannerisms,Verbigeration,Waxy flexibility | 59.4 | 54.9 | 63.9 |
| 5 | Excitement,Grimacing,Echopraxia/echolalia,Mannerisms,Verbigeration | 58.9 | 54.3 | 63.7 |
| 5 | Excitement,Echopraxia/echolalia,Stereotypy,Mannerisms,Verbigeration | 57.0 | 52.5 | 61.7 |

Table S6: sensitivity (relative to the full BFCSI) of all six question combinations, and the 95% confidence interval derived from bootstrapping, sorted by decreasing sensitivity.

|  |  |  | 95% CI | |
| --- | --- | --- | --- | --- |
| # of Qs | Combination | Sensitivity | Lower | Upper |
| 6 | Excitement,Mutism,Staring,Posturing/catalepsy,Echopraxia/echolalia,Negativism | 99.6 | 98.9 | 100.0 |
| 6 | Excitement,Mutism,Staring,Posturing/catalepsy,Echopraxia/echolalia,Rigidity | 99.6 | 98.9 | 100.0 |
| 6 | Excitement,Mutism,Staring,Posturing/catalepsy,Negativism,Withdrawal | 99.6 | 98.9 | 100.0 |
| 6 | Excitement,Mutism,Staring,Echopraxia/echolalia,Rigidity,Negativism | 99.3 | 98.4 | 100.0 |
| 6 | Excitement,Mutism,Staring,Echopraxia/echolalia,Rigidity,Withdrawal | 99.3 | 98.4 | 100.0 |
| 6 | Excitement,Mutism,Staring,Posturing/catalepsy,Rigidity,Withdrawal | 99.3 | 98.4 | 100.0 |
| 6 | Excitement,Mutism,Staring,Posturing/catalepsy,Verbigeration,Negativism | 99.3 | 98.4 | 100.0 |
| 6 | Excitement,Immobility/stupor,Mutism,Staring,Posturing/catalepsy,Negativism | 99.1 | 98.2 | 99.8 |
| 6 | Excitement,Immobility/stupor,Staring,Echopraxia/echolalia,Rigidity,Withdrawal | 99.1 | 98.2 | 99.8 |
| 6 | Excitement,Mutism,Staring,Echopraxia/echolalia,Negativism,Waxy flexibility | 99.1 | 98.2 | 99.8 |
| 6 | Excitement,Mutism,Staring,Echopraxia/echolalia,Negativism,Withdrawal | 99.1 | 98.2 | 99.8 |
| 6 | Excitement,Mutism,Staring,Posturing/catalepsy,Echopraxia/echolalia,Withdrawal | 99.1 | 98.2 | 99.8 |
| 6 | Excitement,Mutism,Staring,Posturing/catalepsy,Mannerisms,Negativism | 99.1 | 98.2 | 99.8 |
| 6 | Excitement,Mutism,Staring,Posturing/catalepsy,Mannerisms,Rigidity | 99.1 | 98.2 | 99.8 |
| 6 | Excitement,Mutism,Staring,Posturing/catalepsy,Negativism,Waxy flexibility | 99.1 | 98.2 | 99.8 |
| 6 | Excitement,Mutism,Staring,Posturing/catalepsy,Rigidity,Negativism | 99.1 | 98.2 | 99.8 |
| 6 | Excitement,Mutism,Staring,Posturing/catalepsy,Stereotypy,Negativism | 99.1 | 98.2 | 99.8 |
| 6 | Excitement,Mutism,Staring,Posturing/catalepsy,Verbigeration,Rigidity | 99.1 | 98.2 | 99.8 |
| 6 | Excitement,Mutism,Staring,Rigidity,Negativism,Withdrawal | 99.1 | 98.2 | 99.8 |
| 6 | Excitement,Mutism,Staring,Rigidity,Waxy flexibility,Withdrawal | 99.1 | 98.2 | 99.8 |
| 6 | Excitement,Mutism,Staring,Stereotypy,Rigidity,Withdrawal | 99.1 | 98.2 | 99.8 |
| 6 | Excitement,Mutism,Staring,Verbigeration,Rigidity,Negativism | 99.1 | 98.2 | 99.8 |
| 6 | Excitement,Immobility/stupor,Mutism,Staring,Echopraxia/echolalia,Rigidity | 98.9 | 97.8 | 99.8 |
| 6 | Excitement,Immobility/stupor,Staring,Echopraxia/echolalia,Rigidity,Negativism | 98.9 | 97.8 | 99.8 |
| 6 | Excitement,Immobility/stupor,Staring,Mannerisms,Rigidity,Withdrawal | 98.9 | 97.8 | 99.8 |
| 6 | Excitement,Immobility/stupor,Staring,Posturing/catalepsy,Echopraxia/echolalia,Negativism | 98.9 | 97.8 | 99.8 |
| 6 | Excitement,Immobility/stupor,Staring,Posturing/catalepsy,Negativism,Withdrawal | 98.9 | 97.8 | 99.8 |
| 6 | Excitement,Immobility/stupor,Staring,Posturing/catalepsy,Rigidity,Negativism | 98.9 | 97.8 | 99.8 |
| 6 | Excitement,Immobility/stupor,Staring,Posturing/catalepsy,Rigidity,Withdrawal | 98.9 | 97.8 | 99.8 |
| 6 | Excitement,Immobility/stupor,Staring,Posturing/catalepsy,Verbigeration,Negativism | 98.9 | 97.8 | 99.8 |
| 6 | Excitement,Immobility/stupor,Staring,Rigidity,Negativism,Withdrawal | 98.9 | 97.8 | 99.8 |
| 6 | Excitement,Immobility/stupor,Staring,Rigidity,Waxy flexibility,Withdrawal | 98.9 | 97.8 | 99.8 |
| 6 | Excitement,Immobility/stupor,Staring,Verbigeration,Rigidity,Negativism | 98.9 | 97.8 | 99.8 |
| 6 | Excitement,Mutism,Staring,Echopraxia/echolalia,Rigidity,Waxy flexibility | 98.9 | 97.8 | 99.8 |
| 6 | Excitement,Mutism,Staring,Echopraxia/echolalia,Stereotypy,Negativism | 98.9 | 97.8 | 99.8 |
| 6 | Excitement,Mutism,Staring,Echopraxia/echolalia,Stereotypy,Rigidity | 98.9 | 97.8 | 99.8 |
| 6 | Excitement,Mutism,Staring,Grimacing,Echopraxia/echolalia,Rigidity | 98.9 | 97.8 | 99.8 |
| 6 | Excitement,Mutism,Staring,Grimacing,Rigidity,Withdrawal | 98.9 | 97.8 | 99.8 |
| 6 | Excitement,Mutism,Staring,Mannerisms,Rigidity,Withdrawal | 98.9 | 97.8 | 99.8 |
| 6 | Excitement,Mutism,Staring,Posturing/catalepsy,Grimacing,Negativism | 98.9 | 97.8 | 99.8 |
| 6 | Excitement,Mutism,Staring,Posturing/catalepsy,Stereotypy,Rigidity | 98.9 | 97.8 | 99.8 |
| 6 | Excitement,Mutism,Staring,Rigidity,Negativism,Waxy flexibility | 98.9 | 97.8 | 99.8 |
| 6 | Excitement,Mutism,Staring,Stereotypy,Negativism,Withdrawal | 98.9 | 97.8 | 99.8 |
| 6 | Excitement,Mutism,Staring,Stereotypy,Rigidity,Negativism | 98.9 | 97.8 | 99.8 |
| 6 | Excitement,Mutism,Staring,Stereotypy,Rigidity,Waxy flexibility | 98.9 | 97.8 | 99.8 |
| 6 | Excitement,Mutism,Staring,Verbigeration,Rigidity,Withdrawal | 98.9 | 97.8 | 99.8 |
| 6 | Excitement,Staring,Posturing/catalepsy,Mannerisms,Rigidity,Withdrawal | 98.9 | 97.8 | 99.8 |
| 6 | Mutism,Staring,Posturing/catalepsy,Mannerisms,Negativism,Withdrawal | 98.9 | 97.8 | 99.8 |
| 6 | Staring,Stereotypy,Mannerisms,Rigidity,Negativism,Withdrawal | 98.9 | 97.8 | 99.8 |
| 6 | Excitement,Immobility/stupor,Mutism,Staring,Posturing/catalepsy,Rigidity | 98.7 | 97.5 | 99.6 |
| 6 | Excitement,Immobility/stupor,Mutism,Staring,Rigidity,Negativism | 98.7 | 97.5 | 99.6 |
| 6 | Excitement,Immobility/stupor,Mutism,Staring,Rigidity,Waxy flexibility | 98.7 | 97.5 | 99.6 |
| 6 | Excitement,Immobility/stupor,Mutism,Staring,Verbigeration,Rigidity | 98.7 | 97.5 | 99.6 |
| 6 | Excitement,Immobility/stupor,Staring,Mannerisms,Rigidity,Negativism | 98.7 | 97.5 | 99.6 |
| 6 | Excitement,Immobility/stupor,Staring,Mannerisms,Verbigeration,Rigidity | 98.7 | 97.5 | 99.6 |
| 6 | Excitement,Immobility/stupor,Staring,Posturing/catalepsy,Echopraxia/echolalia,Rigidity | 98.7 | 97.5 | 99.6 |
| 6 | Excitement,Immobility/stupor,Staring,Posturing/catalepsy,Mannerisms,Negativism | 98.7 | 97.5 | 99.6 |
| 6 | Excitement,Immobility/stupor,Staring,Posturing/catalepsy,Mannerisms,Rigidity | 98.7 | 97.5 | 99.6 |
| 6 | Excitement,Immobility/stupor,Staring,Posturing/catalepsy,Verbigeration,Rigidity | 98.7 | 97.5 | 99.6 |
| 6 | Excitement,Immobility/stupor,Staring,Rigidity,Negativism,Waxy flexibility | 98.7 | 97.5 | 99.6 |
| 6 | Excitement,Immobility/stupor,Staring,Stereotypy,Rigidity,Negativism | 98.7 | 97.5 | 99.6 |
| 6 | Excitement,Immobility/stupor,Staring,Stereotypy,Rigidity,Withdrawal | 98.7 | 97.5 | 99.6 |
| 6 | Excitement,Immobility/stupor,Staring,Verbigeration,Rigidity,Waxy flexibility | 98.7 | 97.5 | 99.6 |
| 6 | Excitement,Immobility/stupor,Staring,Verbigeration,Rigidity,Withdrawal | 98.7 | 97.5 | 99.6 |
| 6 | Excitement,Mutism,Staring,Grimacing,Verbigeration,Rigidity | 98.7 | 97.5 | 99.6 |
| 6 | Excitement,Mutism,Staring,Negativism,Waxy flexibility,Withdrawal | 98.7 | 97.5 | 99.6 |
| 6 | Excitement,Mutism,Staring,Posturing/catalepsy,Echopraxia/echolalia,Waxy flexibility | 98.7 | 97.5 | 99.6 |
| 6 | Excitement,Mutism,Staring,Posturing/catalepsy,Grimacing,Withdrawal | 98.7 | 97.5 | 99.6 |
| 6 | Excitement,Mutism,Staring,Posturing/catalepsy,Mannerisms,Withdrawal | 98.7 | 97.5 | 99.6 |
| 6 | Excitement,Mutism,Staring,Posturing/catalepsy,Rigidity,Waxy flexibility | 98.7 | 97.5 | 99.6 |
| 6 | Excitement,Mutism,Staring,Posturing/catalepsy,Waxy flexibility,Withdrawal | 98.7 | 97.5 | 99.6 |
| 6 | Excitement,Mutism,Staring,Stereotypy,Negativism,Waxy flexibility | 98.7 | 97.5 | 99.6 |
| 6 | Excitement,Mutism,Staring,Verbigeration,Negativism,Waxy flexibility | 98.7 | 97.5 | 99.6 |
| 6 | Excitement,Mutism,Staring,Verbigeration,Negativism,Withdrawal | 98.7 | 97.5 | 99.6 |
| 6 | Excitement,Mutism,Staring,Verbigeration,Rigidity,Waxy flexibility | 98.7 | 97.5 | 99.6 |
| 6 | Excitement,Staring,Echopraxia/echolalia,Rigidity,Negativism,Withdrawal | 98.7 | 97.5 | 99.6 |
| 6 | Excitement,Staring,Mannerisms,Rigidity,Negativism,Withdrawal | 98.7 | 97.5 | 99.6 |
| 6 | Excitement,Staring,Posturing/catalepsy,Echopraxia/echolalia,Rigidity,Withdrawal | 98.7 | 97.5 | 99.6 |
| 6 | Excitement,Staring,Posturing/catalepsy,Rigidity,Negativism,Withdrawal | 98.7 | 97.5 | 99.6 |
| 6 | Mutism,Staring,Echopraxia/echolalia,Mannerisms,Rigidity,Negativism | 98.7 | 97.5 | 99.6 |
| 6 | Staring,Posturing/catalepsy,Mannerisms,Rigidity,Negativism,Withdrawal | 98.7 | 97.5 | 99.6 |
| 6 | Excitement,Immobility/stupor,Mutism,Staring,Rigidity,Withdrawal | 98.6 | 97.5 | 99.6 |
| 6 | Excitement,Immobility/stupor,Staring,Echopraxia/echolalia,Negativism,Withdrawal | 98.6 | 97.5 | 99.6 |
| 6 | Excitement,Mutism,Staring,Echopraxia/echolalia,Verbigeration,Negativism | 98.6 | 97.5 | 99.6 |
| 6 | Excitement,Mutism,Staring,Echopraxia/echolalia,Verbigeration,Rigidity | 98.6 | 97.5 | 99.6 |
| 6 | Excitement,Mutism,Staring,Mannerisms,Rigidity,Negativism | 98.6 | 97.5 | 99.6 |
| 6 | Excitement,Mutism,Staring,Posturing/catalepsy,Grimacing,Rigidity | 98.6 | 97.5 | 99.6 |
| 6 | Excitement,Mutism,Staring,Stereotypy,Verbigeration,Negativism | 98.6 | 97.5 | 99.6 |
| 6 | Excitement,Mutism,Staring,Stereotypy,Verbigeration,Rigidity | 98.6 | 97.5 | 99.6 |
| 6 | Excitement,Staring,Stereotypy,Rigidity,Negativism,Withdrawal | 98.6 | 97.5 | 99.6 |
| 6 | Immobility/stupor,Staring,Mannerisms,Rigidity,Negativism,Withdrawal | 98.6 | 97.5 | 99.6 |
| 6 | Mutism,Staring,Mannerisms,Rigidity,Negativism,Withdrawal | 98.6 | 97.5 | 99.6 |
| 6 | Staring,Echopraxia/echolalia,Mannerisms,Rigidity,Negativism,Withdrawal | 98.6 | 97.5 | 99.6 |
| 6 | Excitement,Immobility/stupor,Echopraxia/echolalia,Rigidity,Negativism,Withdrawal | 98.4 | 97.1 | 99.6 |
| 6 | Excitement,Immobility/stupor,Mutism,Staring,Echopraxia/echolalia,Negativism | 98.4 | 97.1 | 99.6 |
| 6 | Excitement,Immobility/stupor,Mutism,Staring,Mannerisms,Rigidity | 98.4 | 97.1 | 99.6 |
| 6 | Excitement,Immobility/stupor,Mutism,Staring,Negativism,Waxy flexibility | 98.4 | 97.1 | 99.6 |
| 6 | Excitement,Immobility/stupor,Mutism,Staring,Posturing/catalepsy,Echopraxia/echolalia | 98.4 | 97.1 | 99.6 |
| 6 | Excitement,Immobility/stupor,Mutism,Staring,Stereotypy,Rigidity | 98.4 | 97.1 | 99.6 |
| 6 | Excitement,Immobility/stupor,Mutism,Verbigeration,Rigidity,Negativism | 98.4 | 97.1 | 99.6 |
| 6 | Excitement,Immobility/stupor,Staring,Echopraxia/echolalia,Negativism,Waxy flexibility | 98.4 | 97.1 | 99.6 |
| 6 | Excitement,Immobility/stupor,Staring,Echopraxia/echolalia,Rigidity,Waxy flexibility | 98.4 | 97.1 | 99.6 |
| 6 | Excitement,Immobility/stupor,Staring,Echopraxia/echolalia,Verbigeration,Rigidity | 98.4 | 97.1 | 99.6 |
| 6 | Excitement,Immobility/stupor,Staring,Grimacing,Rigidity,Withdrawal | 98.4 | 97.1 | 99.6 |
| 6 | Excitement,Immobility/stupor,Staring,Mannerisms,Rigidity,Waxy flexibility | 98.4 | 97.1 | 99.6 |
| 6 | Excitement,Immobility/stupor,Staring,Posturing/catalepsy,Negativism,Waxy flexibility | 98.4 | 97.3 | 99.6 |
| 6 | Excitement,Immobility/stupor,Staring,Posturing/catalepsy,Stereotypy,Negativism | 98.4 | 97.3 | 99.6 |
| 6 | Excitement,Immobility/stupor,Staring,Stereotypy,Rigidity,Waxy flexibility | 98.4 | 97.1 | 99.6 |
| 6 | Excitement,Immobility/stupor,Staring,Stereotypy,Verbigeration,Rigidity | 98.4 | 97.1 | 99.6 |
| 6 | Excitement,Mutism,Staring,Echopraxia/echolalia,Mannerisms,Negativism | 98.4 | 97.3 | 99.6 |
| 6 | Excitement,Mutism,Staring,Echopraxia/echolalia,Mannerisms,Rigidity | 98.4 | 97.1 | 99.6 |
| 6 | Excitement,Mutism,Staring,Echopraxia/echolalia,Stereotypy,Withdrawal | 98.4 | 97.1 | 99.6 |
| 6 | Excitement,Mutism,Staring,Echopraxia/echolalia,Waxy flexibility,Withdrawal | 98.4 | 97.1 | 99.6 |
| 6 | Excitement,Mutism,Staring,Grimacing,Echopraxia/echolalia,Negativism | 98.4 | 97.1 | 99.6 |
| 6 | Excitement,Mutism,Staring,Grimacing,Rigidity,Negativism | 98.4 | 97.1 | 99.6 |
| 6 | Excitement,Mutism,Staring,Grimacing,Rigidity,Waxy flexibility | 98.4 | 97.3 | 99.6 |
| 6 | Excitement,Mutism,Staring,Grimacing,Stereotypy,Rigidity | 98.4 | 97.1 | 99.6 |
| 6 | Excitement,Mutism,Staring,Mannerisms,Negativism,Waxy flexibility | 98.4 | 97.1 | 99.6 |
| 6 | Excitement,Mutism,Staring,Mannerisms,Negativism,Withdrawal | 98.4 | 97.3 | 99.6 |
| 6 | Excitement,Mutism,Staring,Mannerisms,Rigidity,Waxy flexibility | 98.4 | 97.3 | 99.6 |
| 6 | Excitement,Mutism,Staring,Mannerisms,Verbigeration,Rigidity | 98.4 | 97.1 | 99.6 |
| 6 | Excitement,Mutism,Staring,Posturing/catalepsy,Echopraxia/echolalia,Stereotypy | 98.4 | 97.1 | 99.6 |
| 6 | Excitement,Mutism,Staring,Posturing/catalepsy,Echopraxia/echolalia,Verbigeration | 98.4 | 97.1 | 99.6 |
| 6 | Excitement,Mutism,Staring,Posturing/catalepsy,Grimacing,Echopraxia/echolalia | 98.4 | 97.1 | 99.6 |
| 6 | Excitement,Mutism,Staring,Posturing/catalepsy,Stereotypy,Withdrawal | 98.4 | 97.3 | 99.6 |
| 6 | Excitement,Mutism,Staring,Posturing/catalepsy,Verbigeration,Withdrawal | 98.4 | 97.3 | 99.6 |
| 6 | Excitement,Staring,Echopraxia/echolalia,Stereotypy,Rigidity,Withdrawal | 98.4 | 97.1 | 99.6 |
| 6 | Excitement,Staring,Mannerisms,Rigidity,Waxy flexibility,Withdrawal | 98.4 | 97.1 | 99.6 |
| 6 | Excitement,Staring,Posturing/catalepsy,Echopraxia/echolalia,Negativism,Withdrawal | 98.4 | 97.1 | 99.6 |
| 6 | Excitement,Staring,Posturing/catalepsy,Grimacing,Rigidity,Withdrawal | 98.4 | 97.3 | 99.6 |
| 6 | Excitement,Staring,Posturing/catalepsy,Mannerisms,Negativism,Withdrawal | 98.4 | 97.1 | 99.6 |
| 6 | Excitement,Staring,Rigidity,Negativism,Waxy flexibility,Withdrawal | 98.4 | 97.1 | 99.6 |
| 6 | Excitement,Staring,Stereotypy,Mannerisms,Rigidity,Withdrawal | 98.4 | 97.1 | 99.6 |
| 6 | Excitement,Staring,Stereotypy,Rigidity,Waxy flexibility,Withdrawal | 98.4 | 97.1 | 99.6 |
| 6 | Excitement,Staring,Verbigeration,Rigidity,Negativism,Withdrawal | 98.4 | 97.3 | 99.6 |
| 6 | Immobility/stupor,Staring,Echopraxia/echolalia,Mannerisms,Rigidity,Negativism | 98.4 | 97.3 | 99.6 |
| 6 | Immobility/stupor,Staring,Stereotypy,Mannerisms,Rigidity,Negativism | 98.4 | 97.1 | 99.6 |
| 6 | Mutism,Staring,Echopraxia/echolalia,Mannerisms,Negativism,Withdrawal | 98.4 | 97.1 | 99.6 |
| 6 | Mutism,Staring,Stereotypy,Mannerisms,Negativism,Withdrawal | 98.4 | 97.1 | 99.6 |
| 6 | Mutism,Staring,Stereotypy,Mannerisms,Rigidity,Negativism | 98.4 | 97.1 | 99.6 |
| 6 | Staring,Mannerisms,Rigidity,Negativism,Waxy flexibility,Withdrawal | 98.4 | 97.3 | 99.6 |
| 6 | Staring,Mannerisms,Verbigeration,Rigidity,Negativism,Withdrawal | 98.4 | 97.3 | 99.6 |
| 6 | Excitement,Immobility/stupor,Mutism,Echopraxia/echolalia,Rigidity,Negativism | 98.2 | 96.9 | 99.3 |
| 6 | Excitement,Immobility/stupor,Mutism,Echopraxia/echolalia,Rigidity,Withdrawal | 98.2 | 96.9 | 99.3 |
| 6 | Excitement,Immobility/stupor,Mutism,Posturing/catalepsy,Echopraxia/echolalia,Rigidity | 98.2 | 96.9 | 99.3 |
| 6 | Excitement,Immobility/stupor,Mutism,Posturing/catalepsy,Rigidity,Negativism | 98.2 | 96.9 | 99.3 |
| 6 | Excitement,Immobility/stupor,Mutism,Staring,Echopraxia/echolalia,Waxy flexibility | 98.2 | 96.9 | 99.3 |
| 6 | Excitement,Immobility/stupor,Mutism,Staring,Negativism,Withdrawal | 98.2 | 96.9 | 99.3 |
| 6 | Excitement,Immobility/stupor,Mutism,Staring,Posturing/catalepsy,Withdrawal | 98.2 | 96.9 | 99.3 |
| 6 | Excitement,Immobility/stupor,Mutism,Staring,Stereotypy,Negativism | 98.2 | 96.9 | 99.3 |
| 6 | Excitement,Immobility/stupor,Mutism,Staring,Verbigeration,Negativism | 98.2 | 96.9 | 99.3 |
| 6 | Excitement,Immobility/stupor,Posturing/catalepsy,Echopraxia/echolalia,Rigidity,Withdrawal | 98.2 | 96.9 | 99.3 |
| 6 | Excitement,Immobility/stupor,Posturing/catalepsy,Rigidity,Negativism,Withdrawal | 98.2 | 96.9 | 99.3 |
| 6 | Excitement,Immobility/stupor,Staring,Echopraxia/echolalia,Mannerisms,Rigidity | 98.2 | 96.9 | 99.3 |
| 6 | Excitement,Immobility/stupor,Staring,Echopraxia/echolalia,Stereotypy,Negativism | 98.2 | 96.9 | 99.3 |
| 6 | Excitement,Immobility/stupor,Staring,Echopraxia/echolalia,Stereotypy,Rigidity | 98.2 | 96.9 | 99.3 |
| 6 | Excitement,Immobility/stupor,Staring,Grimacing,Echopraxia/echolalia,Rigidity | 98.2 | 96.9 | 99.3 |
| 6 | Excitement,Immobility/stupor,Staring,Grimacing,Rigidity,Negativism | 98.2 | 96.9 | 99.3 |
| 6 | Excitement,Immobility/stupor,Staring,Grimacing,Verbigeration,Rigidity | 98.2 | 96.9 | 99.3 |
| 6 | Excitement,Immobility/stupor,Staring,Posturing/catalepsy,Echopraxia/echolalia,Withdrawal | 98.2 | 96.9 | 99.3 |
| 6 | Excitement,Immobility/stupor,Staring,Posturing/catalepsy,Grimacing,Negativism | 98.2 | 96.9 | 99.3 |
| 6 | Excitement,Immobility/stupor,Staring,Posturing/catalepsy,Grimacing,Rigidity | 98.2 | 96.9 | 99.3 |
| 6 | Excitement,Immobility/stupor,Staring,Posturing/catalepsy,Stereotypy,Rigidity | 98.2 | 96.9 | 99.3 |
| 6 | Excitement,Immobility/stupor,Staring,Stereotypy,Negativism,Withdrawal | 98.2 | 96.9 | 99.3 |
| 6 | Excitement,Immobility/stupor,Staring,Stereotypy,Verbigeration,Negativism | 98.2 | 96.9 | 99.3 |
| 6 | Excitement,Immobility/stupor,Staring,Verbigeration,Negativism,Waxy flexibility | 98.2 | 96.9 | 99.3 |
| 6 | Excitement,Immobility/stupor,Staring,Verbigeration,Negativism,Withdrawal | 98.2 | 96.9 | 99.3 |
| 6 | Excitement,Mutism,Staring,Echopraxia/echolalia,Stereotypy,Waxy flexibility | 98.2 | 96.9 | 99.3 |
| 6 | Excitement,Mutism,Staring,Grimacing,Echopraxia/echolalia,Waxy flexibility | 98.2 | 96.9 | 99.3 |
| 6 | Excitement,Mutism,Staring,Grimacing,Echopraxia/echolalia,Withdrawal | 98.2 | 96.9 | 99.3 |
| 6 | Excitement,Mutism,Staring,Grimacing,Mannerisms,Rigidity | 98.2 | 96.9 | 99.3 |
| 6 | Excitement,Mutism,Staring,Grimacing,Negativism,Waxy flexibility | 98.2 | 96.9 | 99.3 |
| 6 | Excitement,Mutism,Staring,Grimacing,Negativism,Withdrawal | 98.2 | 96.9 | 99.3 |
| 6 | Excitement,Mutism,Staring,Mannerisms,Verbigeration,Negativism | 98.2 | 96.9 | 99.3 |
| 6 | Excitement,Mutism,Staring,Posturing/catalepsy,Echopraxia/echolalia,Mannerisms | 98.2 | 96.9 | 99.3 |
| 6 | Excitement,Mutism,Staring,Posturing/catalepsy,Stereotypy,Waxy flexibility | 98.2 | 96.9 | 99.3 |
| 6 | Excitement,Mutism,Staring,Posturing/catalepsy,Verbigeration,Waxy flexibility | 98.2 | 96.9 | 99.3 |
| 6 | Excitement,Mutism,Staring,Stereotypy,Mannerisms,Rigidity | 98.2 | 96.9 | 99.3 |
| 6 | Excitement,Mutism,Staring,Stereotypy,Waxy flexibility,Withdrawal | 98.2 | 96.9 | 99.3 |
| 6 | Excitement,Staring,Echopraxia/echolalia,Mannerisms,Rigidity,Withdrawal | 98.2 | 96.9 | 99.3 |
| 6 | Excitement,Staring,Echopraxia/echolalia,Rigidity,Waxy flexibility,Withdrawal | 98.2 | 96.9 | 99.3 |
| 6 | Excitement,Staring,Grimacing,Echopraxia/echolalia,Rigidity,Withdrawal | 98.2 | 96.9 | 99.3 |
| 6 | Excitement,Staring,Grimacing,Mannerisms,Rigidity,Withdrawal | 98.2 | 96.9 | 99.3 |
| 6 | Excitement,Staring,Grimacing,Rigidity,Waxy flexibility,Withdrawal | 98.2 | 96.9 | 99.3 |
| 6 | Excitement,Staring,Grimacing,Stereotypy,Rigidity,Withdrawal | 98.2 | 96.9 | 99.3 |
| 6 | Excitement,Staring,Mannerisms,Verbigeration,Rigidity,Withdrawal | 98.2 | 96.9 | 99.3 |
| 6 | Excitement,Staring,Posturing/catalepsy,Rigidity,Waxy flexibility,Withdrawal | 98.2 | 96.9 | 99.3 |
| 6 | Excitement,Staring,Posturing/catalepsy,Stereotypy,Rigidity,Withdrawal | 98.2 | 96.9 | 99.3 |
| 6 | Excitement,Staring,Posturing/catalepsy,Verbigeration,Negativism,Withdrawal | 98.2 | 96.9 | 99.3 |
| 6 | Excitement,Staring,Posturing/catalepsy,Verbigeration,Rigidity,Withdrawal | 98.2 | 96.9 | 99.3 |
| 6 | Excitement,Staring,Stereotypy,Verbigeration,Rigidity,Withdrawal | 98.2 | 96.9 | 99.3 |
| 6 | Immobility/stupor,Staring,Mannerisms,Verbigeration,Rigidity,Negativism | 98.2 | 96.9 | 99.3 |
| 6 | Immobility/stupor,Staring,Posturing/catalepsy,Mannerisms,Negativism,Withdrawal | 98.2 | 96.9 | 99.3 |
| 6 | Immobility/stupor,Staring,Posturing/catalepsy,Mannerisms,Rigidity,Negativism | 98.2 | 96.9 | 99.3 |
| 6 | Mutism,Staring,Echopraxia/echolalia,Stereotypy,Mannerisms,Negativism | 98.2 | 96.9 | 99.3 |
| 6 | Mutism,Staring,Mannerisms,Verbigeration,Rigidity,Negativism | 98.2 | 96.9 | 99.3 |
| 6 | Mutism,Staring,Posturing/catalepsy,Echopraxia/echolalia,Mannerisms,Negativism | 98.2 | 96.9 | 99.3 |
| 6 | Mutism,Staring,Posturing/catalepsy,Mannerisms,Rigidity,Negativism | 98.2 | 96.9 | 99.3 |
| 6 | Mutism,Staring,Posturing/catalepsy,Mannerisms,Rigidity,Withdrawal | 98.2 | 96.9 | 99.3 |
| 6 | Mutism,Staring,Posturing/catalepsy,Stereotypy,Mannerisms,Negativism | 98.2 | 96.9 | 99.3 |
| 6 | Mutism,Staring,Stereotypy,Mannerisms,Rigidity,Withdrawal | 98.2 | 96.9 | 99.3 |
| 6 | Mutism,Staring,Stereotypy,Verbigeration,Rigidity,Negativism | 98.2 | 96.9 | 99.3 |
| 6 | Staring,Grimacing,Mannerisms,Rigidity,Negativism,Withdrawal | 98.2 | 96.9 | 99.3 |
| 6 | Staring,Stereotypy,Verbigeration,Rigidity,Negativism,Withdrawal | 98.2 | 96.9 | 99.3 |
| 6 | Excitement,Immobility/stupor,Echopraxia/echolalia,Stereotypy,Rigidity,Withdrawal | 98.0 | 96.6 | 99.1 |
| 6 | Excitement,Immobility/stupor,Echopraxia/echolalia,Verbigeration,Rigidity,Withdrawal | 98.0 | 96.6 | 99.1 |
| 6 | Excitement,Immobility/stupor,Grimacing,Echopraxia/echolalia,Rigidity,Withdrawal | 98.0 | 96.6 | 99.1 |
| 6 | Excitement,Immobility/stupor,Mutism,Posturing/catalepsy,Rigidity,Withdrawal | 98.0 | 96.6 | 99.1 |
| 6 | Excitement,Immobility/stupor,Mutism,Posturing/catalepsy,Verbigeration,Rigidity | 98.0 | 96.6 | 99.1 |
| 6 | Excitement,Immobility/stupor,Mutism,Rigidity,Negativism,Withdrawal | 98.0 | 96.6 | 99.1 |
| 6 | Excitement,Immobility/stupor,Mutism,Staring,Echopraxia/echolalia,Withdrawal | 98.0 | 96.6 | 99.1 |
| 6 | Excitement,Immobility/stupor,Mutism,Staring,Grimacing,Rigidity | 98.0 | 96.6 | 99.1 |
| 6 | Excitement,Immobility/stupor,Mutism,Staring,Posturing/catalepsy,Mannerisms | 98.0 | 96.6 | 99.1 |
| 6 | Excitement,Immobility/stupor,Mutism,Staring,Posturing/catalepsy,Verbigeration | 98.0 | 96.6 | 99.1 |
| 6 | Excitement,Immobility/stupor,Mutism,Staring,Posturing/catalepsy,Waxy flexibility | 98.0 | 96.6 | 99.1 |
| 6 | Excitement,Immobility/stupor,Mutism,Stereotypy,Rigidity,Negativism | 98.0 | 96.6 | 99.1 |
| 6 | Excitement,Immobility/stupor,Mutism,Verbigeration,Rigidity,Withdrawal | 98.0 | 96.6 | 99.1 |
| 6 | Excitement,Immobility/stupor,Posturing/catalepsy,Verbigeration,Rigidity,Negativism | 98.0 | 96.6 | 99.1 |
| 6 | Excitement,Immobility/stupor,Staring,Echopraxia/echolalia,Mannerisms,Negativism | 98.0 | 96.6 | 99.1 |
| 6 | Excitement,Immobility/stupor,Staring,Echopraxia/echolalia,Verbigeration,Negativism | 98.0 | 96.6 | 99.1 |
| 6 | Excitement,Immobility/stupor,Staring,Grimacing,Mannerisms,Rigidity | 98.0 | 96.6 | 99.1 |
| 6 | Excitement,Immobility/stupor,Staring,Grimacing,Rigidity,Waxy flexibility | 98.0 | 96.6 | 99.1 |
| 6 | Excitement,Immobility/stupor,Staring,Grimacing,Stereotypy,Rigidity | 98.0 | 96.6 | 99.1 |
| 6 | Excitement,Immobility/stupor,Staring,Mannerisms,Negativism,Waxy flexibility | 98.0 | 96.6 | 99.1 |
| 6 | Excitement,Immobility/stupor,Staring,Mannerisms,Negativism,Withdrawal | 98.0 | 96.6 | 99.1 |
| 6 | Excitement,Immobility/stupor,Staring,Mannerisms,Verbigeration,Negativism | 98.0 | 96.6 | 99.1 |
| 6 | Excitement,Immobility/stupor,Staring,Negativism,Waxy flexibility,Withdrawal | 98.0 | 96.6 | 99.1 |
| 6 | Excitement,Immobility/stupor,Staring,Posturing/catalepsy,Mannerisms,Withdrawal | 98.0 | 96.6 | 99.1 |
| 6 | Excitement,Immobility/stupor,Staring,Posturing/catalepsy,Rigidity,Waxy flexibility | 98.0 | 96.6 | 99.1 |
| 6 | Excitement,Immobility/stupor,Staring,Stereotypy,Mannerisms,Rigidity | 98.0 | 96.6 | 99.1 |
| 6 | Excitement,Immobility/stupor,Staring,Stereotypy,Negativism,Waxy flexibility | 98.0 | 96.6 | 99.1 |
| 6 | Excitement,Immobility/stupor,Stereotypy,Rigidity,Negativism,Withdrawal | 98.0 | 96.6 | 99.1 |
| 6 | Excitement,Immobility/stupor,Verbigeration,Rigidity,Negativism,Withdrawal | 98.0 | 96.6 | 99.1 |
| 6 | Excitement,Mutism,Staring,Grimacing,Stereotypy,Negativism | 98.0 | 96.6 | 99.1 |
| 6 | Excitement,Mutism,Staring,Grimacing,Verbigeration,Negativism | 98.0 | 96.6 | 99.1 |
| 6 | Excitement,Mutism,Staring,Grimacing,Waxy flexibility,Withdrawal | 98.0 | 96.6 | 99.1 |
| 6 | Excitement,Mutism,Staring,Posturing/catalepsy,Grimacing,Stereotypy | 98.0 | 96.6 | 99.1 |
| 6 | Excitement,Mutism,Staring,Posturing/catalepsy,Grimacing,Verbigeration | 98.0 | 96.6 | 99.1 |
| 6 | Excitement,Mutism,Staring,Posturing/catalepsy,Mannerisms,Verbigeration | 98.0 | 96.6 | 99.1 |
| 6 | Excitement,Mutism,Staring,Posturing/catalepsy,Mannerisms,Waxy flexibility | 98.0 | 96.6 | 99.1 |
| 6 | Excitement,Mutism,Staring,Posturing/catalepsy,Stereotypy,Verbigeration | 98.0 | 96.6 | 99.1 |
| 6 | Excitement,Mutism,Staring,Stereotypy,Mannerisms,Negativism | 98.0 | 96.6 | 99.1 |
| 6 | Excitement,Staring,Echopraxia/echolalia,Negativism,Waxy flexibility,Withdrawal | 98.0 | 96.6 | 99.1 |
| 6 | Excitement,Staring,Echopraxia/echolalia,Stereotypy,Negativism,Withdrawal | 98.0 | 96.6 | 99.1 |
| 6 | Excitement,Staring,Grimacing,Rigidity,Negativism,Withdrawal | 98.0 | 96.6 | 99.1 |
| 6 | Excitement,Staring,Grimacing,Verbigeration,Rigidity,Withdrawal | 98.0 | 96.6 | 99.1 |
| 6 | Excitement,Staring,Posturing/catalepsy,Grimacing,Negativism,Withdrawal | 98.0 | 96.6 | 99.1 |
| 6 | Excitement,Staring,Posturing/catalepsy,Negativism,Waxy flexibility,Withdrawal | 98.0 | 96.6 | 99.1 |
| 6 | Excitement,Staring,Posturing/catalepsy,Stereotypy,Negativism,Withdrawal | 98.0 | 96.6 | 99.1 |
| 6 | Excitement,Staring,Verbigeration,Rigidity,Waxy flexibility,Withdrawal | 98.0 | 96.6 | 99.1 |
| 6 | Immobility/stupor,Staring,Echopraxia/echolalia,Mannerisms,Negativism,Withdrawal | 98.0 | 96.6 | 99.1 |
| 6 | Immobility/stupor,Staring,Mannerisms,Rigidity,Negativism,Waxy flexibility | 98.0 | 96.6 | 99.1 |
| 6 | Immobility/stupor,Staring,Stereotypy,Mannerisms,Negativism,Withdrawal | 98.0 | 96.6 | 99.1 |
| 6 | Immobility/stupor,Staring,Stereotypy,Verbigeration,Rigidity,Negativism | 98.0 | 96.6 | 99.1 |
| 6 | Mutism,Staring,Echopraxia/echolalia,Rigidity,Negativism,Withdrawal | 98.0 | 96.6 | 99.1 |
| 6 | Mutism,Staring,Grimacing,Mannerisms,Negativism,Withdrawal | 98.0 | 96.6 | 99.1 |
| 6 | Mutism,Staring,Grimacing,Mannerisms,Rigidity,Withdrawal | 98.0 | 96.6 | 99.1 |
| 6 | Mutism,Staring,Mannerisms,Negativism,Waxy flexibility,Withdrawal | 98.0 | 96.6 | 99.1 |
| 6 | Mutism,Staring,Mannerisms,Rigidity,Negativism,Waxy flexibility | 98.0 | 96.6 | 99.1 |
| 6 | Mutism,Staring,Mannerisms,Verbigeration,Negativism,Withdrawal | 98.0 | 96.6 | 99.1 |
| 6 | Mutism,Staring,Posturing/catalepsy,Echopraxia/echolalia,Negativism,Withdrawal | 98.0 | 96.6 | 99.1 |
| 6 | Mutism,Staring,Posturing/catalepsy,Stereotypy,Negativism,Withdrawal | 98.0 | 96.6 | 99.1 |
| 6 | Mutism,Staring,Stereotypy,Rigidity,Negativism,Withdrawal | 98.0 | 96.6 | 99.1 |
| 6 | Mutism,Staring,Verbigeration,Rigidity,Negativism,Withdrawal | 98.0 | 96.6 | 99.1 |
| 6 | Staring,Echopraxia/echolalia,Stereotypy,Mannerisms,Negativism,Withdrawal | 98.0 | 96.6 | 99.1 |
| 6 | Staring,Grimacing,Stereotypy,Mannerisms,Rigidity,Withdrawal | 98.0 | 96.6 | 99.1 |
| 6 | Staring,Posturing/catalepsy,Echopraxia/echolalia,Mannerisms,Negativism,Withdrawal | 98.0 | 96.6 | 99.1 |
| 6 | Staring,Posturing/catalepsy,Grimacing,Mannerisms,Negativism,Withdrawal | 98.0 | 96.6 | 99.1 |
| 6 | Staring,Posturing/catalepsy,Stereotypy,Mannerisms,Negativism,Withdrawal | 98.0 | 96.6 | 99.1 |
| 6 | Excitement,Immobility/stupor,Echopraxia/echolalia,Mannerisms,Rigidity,Withdrawal | 97.8 | 96.4 | 99.1 |
| 6 | Excitement,Immobility/stupor,Echopraxia/echolalia,Rigidity,Waxy flexibility,Withdrawal | 97.8 | 96.2 | 99.1 |
| 6 | Excitement,Immobility/stupor,Grimacing,Echopraxia/echolalia,Rigidity,Negativism | 97.8 | 96.2 | 99.1 |
| 6 | Excitement,Immobility/stupor,Mannerisms,Rigidity,Negativism,Withdrawal | 97.8 | 96.2 | 99.1 |
| 6 | Excitement,Immobility/stupor,Mutism,Echopraxia/echolalia,Verbigeration,Rigidity | 97.8 | 96.2 | 99.1 |
| 6 | Excitement,Immobility/stupor,Mutism,Grimacing,Rigidity,Withdrawal | 97.8 | 96.2 | 99.1 |
| 6 | Excitement,Immobility/stupor,Mutism,Posturing/catalepsy,Grimacing,Rigidity | 97.8 | 96.4 | 99.1 |
| 6 | Excitement,Immobility/stupor,Mutism,Posturing/catalepsy,Negativism,Withdrawal | 97.8 | 96.2 | 99.1 |
| 6 | Excitement,Immobility/stupor,Mutism,Rigidity,Negativism,Waxy flexibility | 97.8 | 96.2 | 99.1 |
| 6 | Excitement,Immobility/stupor,Mutism,Rigidity,Waxy flexibility,Withdrawal | 97.8 | 96.2 | 99.1 |
| 6 | Excitement,Immobility/stupor,Mutism,Staring,Mannerisms,Negativism | 97.8 | 96.2 | 99.1 |
| 6 | Excitement,Immobility/stupor,Mutism,Staring,Posturing/catalepsy,Grimacing | 97.8 | 96.4 | 99.1 |
| 6 | Excitement,Immobility/stupor,Mutism,Staring,Posturing/catalepsy,Stereotypy | 97.8 | 96.2 | 99.1 |
| 6 | Excitement,Immobility/stupor,Mutism,Staring,Stereotypy,Waxy flexibility | 97.8 | 96.4 | 99.1 |
| 6 | Excitement,Immobility/stupor,Mutism,Staring,Verbigeration,Waxy flexibility | 97.8 | 96.2 | 99.1 |
| 6 | Excitement,Immobility/stupor,Mutism,Staring,Waxy flexibility,Withdrawal | 97.8 | 96.2 | 99.1 |
| 6 | Excitement,Immobility/stupor,Posturing/catalepsy,Echopraxia/echolalia,Negativism,Withdrawal | 97.8 | 96.2 | 99.1 |
| 6 | Excitement,Immobility/stupor,Posturing/catalepsy,Echopraxia/echolalia,Rigidity,Negativism | 97.8 | 96.2 | 99.1 |
| 6 | Excitement,Immobility/stupor,Posturing/catalepsy,Grimacing,Rigidity,Negativism | 97.8 | 96.2 | 99.1 |
| 6 | Excitement,Immobility/stupor,Posturing/catalepsy,Grimacing,Rigidity,Withdrawal | 97.8 | 96.4 | 99.1 |
| 6 | Excitement,Immobility/stupor,Posturing/catalepsy,Mannerisms,Rigidity,Withdrawal | 97.8 | 96.2 | 99.1 |
| 6 | Excitement,Immobility/stupor,Posturing/catalepsy,Verbigeration,Rigidity,Withdrawal | 97.8 | 96.2 | 99.1 |
| 6 | Excitement,Immobility/stupor,Staring,Echopraxia/echolalia,Waxy flexibility,Withdrawal | 97.8 | 96.4 | 99.1 |
| 6 | Excitement,Immobility/stupor,Staring,Grimacing,Echopraxia/echolalia,Negativism | 97.8 | 96.2 | 99.1 |
| 6 | Excitement,Immobility/stupor,Staring,Posturing/catalepsy,Mannerisms,Verbigeration | 97.8 | 96.2 | 99.1 |
| 6 | Excitement,Immobility/stupor,Stereotypy,Verbigeration,Rigidity,Negativism | 97.8 | 96.2 | 99.1 |
| 6 | Excitement,Mutism,Staring,Echopraxia/echolalia,Mannerisms,Withdrawal | 97.8 | 96.2 | 99.1 |
| 6 | Excitement,Mutism,Staring,Echopraxia/echolalia,Verbigeration,Waxy flexibility | 97.8 | 96.4 | 99.1 |
| 6 | Excitement,Mutism,Staring,Echopraxia/echolalia,Verbigeration,Withdrawal | 97.8 | 96.2 | 99.1 |
| 6 | Excitement,Mutism,Staring,Grimacing,Echopraxia/echolalia,Stereotypy | 97.8 | 96.4 | 99.1 |
| 6 | Excitement,Mutism,Staring,Grimacing,Stereotypy,Waxy flexibility | 97.8 | 96.2 | 99.1 |
| 6 | Excitement,Mutism,Staring,Grimacing,Stereotypy,Withdrawal | 97.8 | 96.2 | 99.1 |
| 6 | Excitement,Mutism,Staring,Posturing/catalepsy,Grimacing,Mannerisms | 97.8 | 96.2 | 99.1 |
| 6 | Excitement,Mutism,Staring,Posturing/catalepsy,Grimacing,Waxy flexibility | 97.8 | 96.4 | 99.1 |
| 6 | Excitement,Mutism,Staring,Posturing/catalepsy,Stereotypy,Mannerisms | 97.8 | 96.4 | 99.1 |
| 6 | Excitement,Mutism,Staring,Stereotypy,Verbigeration,Waxy flexibility | 97.8 | 96.2 | 99.1 |
| 6 | Excitement,Mutism,Staring,Stereotypy,Verbigeration,Withdrawal | 97.8 | 96.2 | 99.1 |
| 6 | Excitement,Mutism,Staring,Verbigeration,Waxy flexibility,Withdrawal | 97.8 | 96.4 | 99.1 |
| 6 | Excitement,Staring,Echopraxia/echolalia,Mannerisms,Negativism,Withdrawal | 97.8 | 96.2 | 99.1 |
| 6 | Excitement,Staring,Echopraxia/echolalia,Verbigeration,Rigidity,Withdrawal | 97.8 | 96.4 | 99.1 |
| 6 | Excitement,Staring,Mannerisms,Negativism,Waxy flexibility,Withdrawal | 97.8 | 96.4 | 99.1 |
| 6 | Excitement,Staring,Posturing/catalepsy,Mannerisms,Rigidity,Negativism | 97.8 | 96.2 | 99.1 |
| 6 | Excitement,Staring,Stereotypy,Verbigeration,Negativism,Withdrawal | 97.8 | 96.4 | 99.1 |
| 6 | Immobility/stupor,Mutism,Staring,Mannerisms,Rigidity,Negativism | 97.8 | 96.4 | 99.1 |
| 6 | Immobility/stupor,Staring,Grimacing,Mannerisms,Rigidity,Negativism | 97.8 | 96.2 | 99.1 |
| 6 | Immobility/stupor,Staring,Stereotypy,Rigidity,Negativism,Withdrawal | 97.8 | 96.2 | 99.1 |
| 6 | Immobility/stupor,Staring,Verbigeration,Rigidity,Negativism,Withdrawal | 97.8 | 96.2 | 98.9 |
| 6 | Mutism,Staring,Echopraxia/echolalia,Verbigeration,Rigidity,Negativism | 97.8 | 96.2 | 99.1 |
| 6 | Mutism,Staring,Grimacing,Echopraxia/echolalia,Mannerisms,Negativism | 97.8 | 96.2 | 99.1 |
| 6 | Mutism,Staring,Grimacing,Mannerisms,Rigidity,Negativism | 97.8 | 96.2 | 99.1 |
| 6 | Mutism,Staring,Grimacing,Stereotypy,Verbigeration,Rigidity | 97.8 | 96.4 | 99.1 |
| 6 | Mutism,Staring,Mannerisms,Verbigeration,Rigidity,Withdrawal | 97.8 | 96.4 | 99.1 |
| 6 | Mutism,Staring,Posturing/catalepsy,Echopraxia/echolalia,Mannerisms,Rigidity | 97.8 | 96.2 | 99.1 |
| 6 | Mutism,Staring,Posturing/catalepsy,Grimacing,Mannerisms,Negativism | 97.8 | 96.4 | 99.1 |
| 6 | Mutism,Staring,Posturing/catalepsy,Grimacing,Mannerisms,Withdrawal | 97.8 | 96.4 | 99.1 |
| 6 | Mutism,Staring,Posturing/catalepsy,Mannerisms,Verbigeration,Rigidity | 97.8 | 96.2 | 99.1 |
| 6 | Mutism,Staring,Posturing/catalepsy,Stereotypy,Verbigeration,Negativism | 97.8 | 96.2 | 99.1 |
| 6 | Mutism,Staring,Posturing/catalepsy,Verbigeration,Negativism,Withdrawal | 97.8 | 96.4 | 99.1 |
| 6 | Mutism,Staring,Stereotypy,Mannerisms,Verbigeration,Negativism | 97.8 | 96.4 | 99.1 |
| 6 | Mutism,Staring,Stereotypy,Verbigeration,Negativism,Withdrawal | 97.8 | 96.4 | 99.1 |
| 6 | Staring,Echopraxia/echolalia,Stereotypy,Rigidity,Negativism,Withdrawal | 97.8 | 96.2 | 99.1 |
| 6 | Staring,Stereotypy,Mannerisms,Verbigeration,Negativism,Withdrawal | 97.8 | 96.2 | 99.1 |
| 6 | Excitement,Immobility/stupor,Mutism,Grimacing,Rigidity,Negativism | 97.7 | 96.2 | 99.1 |
| 6 | Excitement,Immobility/stupor,Mutism,Stereotypy,Rigidity,Withdrawal | 97.7 | 96.2 | 99.1 |
| 6 | Excitement,Immobility/stupor,Posturing/catalepsy,Verbigeration,Negativism,Withdrawal | 97.7 | 96.2 | 99.1 |
| 6 | Excitement,Mutism,Staring,Mannerisms,Waxy flexibility,Withdrawal | 97.7 | 96.4 | 99.1 |
| 6 | Excitement,Staring,Posturing/catalepsy,Echopraxia/echolalia,Rigidity,Negativism | 97.7 | 96.4 | 99.1 |
| 6 | Immobility/stupor,Staring,Echopraxia/echolalia,Rigidity,Negativism,Withdrawal | 97.7 | 96.2 | 98.9 |
| 6 | Immobility/stupor,Staring,Echopraxia/echolalia,Stereotypy,Mannerisms,Negativism | 97.7 | 96.2 | 99.1 |
| 6 | Immobility/stupor,Staring,Posturing/catalepsy,Stereotypy,Mannerisms,Negativism | 97.7 | 96.2 | 99.1 |
| 6 | Mutism,Staring,Echopraxia/echolalia,Mannerisms,Negativism,Waxy flexibility | 97.7 | 96.2 | 99.1 |
| 6 | Mutism,Staring,Echopraxia/echolalia,Mannerisms,Rigidity,Withdrawal | 97.7 | 96.4 | 99.1 |
| 6 | Mutism,Staring,Posturing/catalepsy,Mannerisms,Verbigeration,Negativism | 97.7 | 96.2 | 99.1 |
| 6 | Mutism,Staring,Stereotypy,Mannerisms,Negativism,Waxy flexibility | 97.7 | 96.2 | 98.9 |
| 6 | Mutism,Staring,Stereotypy,Verbigeration,Rigidity,Withdrawal | 97.7 | 96.4 | 99.1 |
| 6 | Staring,Grimacing,Stereotypy,Mannerisms,Negativism,Withdrawal | 97.7 | 96.2 | 99.1 |
| 6 | Staring,Posturing/catalepsy,Mannerisms,Verbigeration,Negativism,Withdrawal | 97.7 | 96.2 | 99.1 |
| 6 | Excitement,Immobility/stupor,Echopraxia/echolalia,Stereotypy,Rigidity,Negativism | 97.5 | 96.0 | 98.9 |
| 6 | Excitement,Immobility/stupor,Echopraxia/echolalia,Verbigeration,Rigidity,Negativism | 97.5 | 96.0 | 98.9 |
| 6 | Excitement,Immobility/stupor,Grimacing,Rigidity,Negativism,Withdrawal | 97.5 | 96.0 | 98.9 |
| 6 | Excitement,Immobility/stupor,Mannerisms,Verbigeration,Rigidity,Withdrawal | 97.5 | 96.0 | 98.9 |
| 6 | Excitement,Immobility/stupor,Mutism,Echopraxia/echolalia,Rigidity,Waxy flexibility | 97.5 | 96.0 | 98.9 |
| 6 | Excitement,Immobility/stupor,Mutism,Echopraxia/echolalia,Stereotypy,Rigidity | 97.5 | 96.0 | 98.9 |
| 6 | Excitement,Immobility/stupor,Mutism,Grimacing,Echopraxia/echolalia,Rigidity | 97.5 | 96.0 | 98.9 |
| 6 | Excitement,Immobility/stupor,Mutism,Grimacing,Verbigeration,Rigidity | 97.5 | 96.0 | 98.9 |
| 6 | Excitement,Immobility/stupor,Mutism,Mannerisms,Rigidity,Negativism | 97.5 | 96.0 | 98.9 |
| 6 | Excitement,Immobility/stupor,Mutism,Mannerisms,Rigidity,Withdrawal | 97.5 | 96.0 | 98.9 |
| 6 | Excitement,Immobility/stupor,Mutism,Posturing/catalepsy,Mannerisms,Rigidity | 97.5 | 96.0 | 98.9 |
| 6 | Excitement,Immobility/stupor,Mutism,Posturing/catalepsy,Stereotypy,Rigidity | 97.5 | 96.0 | 98.9 |
| 6 | Excitement,Immobility/stupor,Mutism,Posturing/catalepsy,Verbigeration,Negativism | 97.5 | 96.0 | 98.9 |
| 6 | Excitement,Immobility/stupor,Mutism,Staring,Echopraxia/echolalia,Stereotypy | 97.5 | 96.0 | 98.9 |
| 6 | Excitement,Immobility/stupor,Mutism,Staring,Grimacing,Negativism | 97.5 | 96.0 | 98.9 |
| 6 | Excitement,Immobility/stupor,Mutism,Staring,Mannerisms,Waxy flexibility | 97.5 | 96.0 | 98.9 |
| 6 | Excitement,Immobility/stupor,Mutism,Stereotypy,Verbigeration,Rigidity | 97.5 | 96.0 | 98.9 |
| 6 | Excitement,Immobility/stupor,Mutism,Verbigeration,Rigidity,Waxy flexibility | 97.5 | 96.0 | 98.9 |
| 6 | Excitement,Immobility/stupor,Posturing/catalepsy,Mannerisms,Rigidity,Negativism | 97.5 | 96.0 | 98.9 |
| 6 | Excitement,Immobility/stupor,Posturing/catalepsy,Stereotypy,Rigidity,Negativism | 97.5 | 96.0 | 98.9 |
| 6 | Excitement,Immobility/stupor,Posturing/catalepsy,Stereotypy,Rigidity,Withdrawal | 97.5 | 96.0 | 98.9 |
| 6 | Excitement,Immobility/stupor,Staring,Echopraxia/echolalia,Mannerisms,Withdrawal | 97.5 | 96.0 | 98.9 |
| 6 | Excitement,Immobility/stupor,Staring,Echopraxia/echolalia,Stereotypy,Withdrawal | 97.5 | 96.0 | 98.9 |
| 6 | Excitement,Immobility/stupor,Staring,Grimacing,Negativism,Waxy flexibility | 97.5 | 96.0 | 98.9 |
| 6 | Excitement,Immobility/stupor,Staring,Grimacing,Negativism,Withdrawal | 97.5 | 96.0 | 98.9 |
| 6 | Excitement,Immobility/stupor,Staring,Grimacing,Verbigeration,Negativism | 97.5 | 96.0 | 98.9 |
| 6 | Excitement,Immobility/stupor,Staring,Posturing/catalepsy,Echopraxia/echolalia,Mannerisms | 97.5 | 96.0 | 98.9 |
| 6 | Excitement,Immobility/stupor,Staring,Posturing/catalepsy,Echopraxia/echolalia,Verbigeration | 97.5 | 96.0 | 98.9 |
| 6 | Excitement,Immobility/stupor,Staring,Posturing/catalepsy,Echopraxia/echolalia,Waxy flexibility | 97.5 | 96.0 | 98.9 |
| 6 | Excitement,Immobility/stupor,Staring,Posturing/catalepsy,Grimacing,Echopraxia/echolalia | 97.5 | 96.0 | 98.9 |
| 6 | Excitement,Immobility/stupor,Staring,Posturing/catalepsy,Grimacing,Withdrawal | 97.5 | 96.0 | 98.9 |
| 6 | Excitement,Immobility/stupor,Staring,Posturing/catalepsy,Verbigeration,Waxy flexibility | 97.5 | 96.0 | 98.9 |
| 6 | Excitement,Immobility/stupor,Staring,Posturing/catalepsy,Verbigeration,Withdrawal | 97.5 | 96.0 | 98.9 |
| 6 | Excitement,Immobility/stupor,Staring,Posturing/catalepsy,Waxy flexibility,Withdrawal | 97.5 | 96.0 | 98.9 |
| 6 | Excitement,Immobility/stupor,Staring,Stereotypy,Mannerisms,Negativism | 97.5 | 96.0 | 98.9 |
| 6 | Excitement,Immobility/stupor,Stereotypy,Verbigeration,Rigidity,Withdrawal | 97.5 | 96.0 | 98.9 |
| 6 | Excitement,Mutism,Staring,Echopraxia/echolalia,Mannerisms,Waxy flexibility | 97.5 | 96.0 | 98.9 |
| 6 | Excitement,Mutism,Staring,Echopraxia/echolalia,Stereotypy,Verbigeration | 97.5 | 96.0 | 98.9 |
| 6 | Excitement,Mutism,Staring,Grimacing,Mannerisms,Negativism | 97.5 | 96.0 | 98.9 |
| 6 | Excitement,Mutism,Staring,Grimacing,Verbigeration,Waxy flexibility | 97.5 | 96.0 | 98.9 |
| 6 | Excitement,Mutism,Staring,Grimacing,Verbigeration,Withdrawal | 97.5 | 96.0 | 98.9 |
| 6 | Excitement,Mutism,Staring,Stereotypy,Mannerisms,Withdrawal | 97.5 | 96.0 | 98.9 |
| 6 | Excitement,Staring,Echopraxia/echolalia,Rigidity,Negativism,Waxy flexibility | 97.5 | 96.0 | 98.9 |
| 6 | Excitement,Staring,Echopraxia/echolalia,Stereotypy,Rigidity,Negativism | 97.5 | 96.0 | 98.9 |
| 6 | Excitement,Staring,Grimacing,Echopraxia/echolalia,Negativism,Withdrawal | 97.5 | 96.0 | 98.9 |
| 6 | Excitement,Staring,Mannerisms,Rigidity,Negativism,Waxy flexibility | 97.5 | 96.0 | 98.9 |
| 6 | Excitement,Staring,Mannerisms,Verbigeration,Negativism,Withdrawal | 97.5 | 96.0 | 98.9 |
| 6 | Excitement,Staring,Posturing/catalepsy,Echopraxia/echolalia,Mannerisms,Withdrawal | 97.5 | 96.0 | 98.9 |
| 6 | Excitement,Staring,Posturing/catalepsy,Grimacing,Echopraxia/echolalia,Withdrawal | 97.5 | 96.0 | 98.9 |
| 6 | Excitement,Staring,Posturing/catalepsy,Grimacing,Mannerisms,Withdrawal | 97.5 | 96.0 | 98.9 |
| 6 | Excitement,Staring,Posturing/catalepsy,Stereotypy,Rigidity,Negativism | 97.5 | 96.0 | 98.9 |
| 6 | Excitement,Staring,Posturing/catalepsy,Verbigeration,Rigidity,Negativism | 97.5 | 96.0 | 98.9 |
| 6 | Excitement,Staring,Stereotypy,Mannerisms,Negativism,Withdrawal | 97.5 | 96.0 | 98.9 |
| 6 | Excitement,Staring,Stereotypy,Negativism,Waxy flexibility,Withdrawal | 97.5 | 96.0 | 98.9 |
| 6 | Excitement,Staring,Stereotypy,Rigidity,Negativism,Waxy flexibility | 97.5 | 96.0 | 98.9 |
| 6 | Excitement,Staring,Stereotypy,Verbigeration,Rigidity,Negativism | 97.5 | 96.0 | 98.9 |
| 6 | Excitement,Staring,Verbigeration,Negativism,Waxy flexibility,Withdrawal | 97.5 | 96.0 | 98.9 |
| 6 | Immobility/stupor,Echopraxia/echolalia,Mannerisms,Rigidity,Negativism,Withdrawal | 97.5 | 96.0 | 98.9 |
| 6 | Immobility/stupor,Mutism,Staring,Mannerisms,Negativism,Withdrawal | 97.5 | 96.0 | 98.9 |
| 6 | Immobility/stupor,Posturing/catalepsy,Mannerisms,Rigidity,Negativism,Withdrawal | 97.5 | 96.0 | 98.9 |
| 6 | Immobility/stupor,Staring,Mannerisms,Verbigeration,Negativism,Withdrawal | 97.5 | 96.0 | 98.9 |
| 6 | Immobility/stupor,Staring,Posturing/catalepsy,Echopraxia/echolalia,Mannerisms,Negativism | 97.5 | 96.0 | 98.9 |
| 6 | Immobility/stupor,Staring,Stereotypy,Mannerisms,Rigidity,Withdrawal | 97.5 | 96.0 | 98.9 |
| 6 | Immobility/stupor,Staring,Stereotypy,Mannerisms,Verbigeration,Negativism | 97.5 | 96.0 | 98.9 |
| 6 | Immobility/stupor,Stereotypy,Mannerisms,Rigidity,Negativism,Withdrawal | 97.5 | 96.0 | 98.9 |
| 6 | Mutism,Staring,Echopraxia/echolalia,Mannerisms,Verbigeration,Negativism | 97.5 | 96.0 | 98.9 |
| 6 | Mutism,Staring,Echopraxia/echolalia,Stereotypy,Negativism,Withdrawal | 97.5 | 96.0 | 98.9 |
| 6 | Mutism,Staring,Echopraxia/echolalia,Stereotypy,Rigidity,Negativism | 97.5 | 96.0 | 98.9 |
| 6 | Mutism,Staring,Echopraxia/echolalia,Stereotypy,Rigidity,Withdrawal | 97.5 | 96.0 | 98.9 |
| 6 | Mutism,Staring,Grimacing,Echopraxia/echolalia,Mannerisms,Rigidity | 97.5 | 96.0 | 98.9 |
| 6 | Mutism,Staring,Grimacing,Mannerisms,Verbigeration,Rigidity | 97.5 | 96.0 | 98.9 |
| 6 | Mutism,Staring,Grimacing,Stereotypy,Mannerisms,Negativism | 97.5 | 96.0 | 98.9 |
| 6 | Mutism,Staring,Grimacing,Stereotypy,Mannerisms,Rigidity | 97.5 | 96.0 | 98.9 |
| 6 | Mutism,Staring,Mannerisms,Rigidity,Waxy flexibility,Withdrawal | 97.5 | 96.0 | 98.9 |
| 6 | Mutism,Staring,Posturing/catalepsy,Echopraxia/echolalia,Mannerisms,Withdrawal | 97.5 | 96.0 | 98.9 |
| 6 | Mutism,Staring,Posturing/catalepsy,Echopraxia/echolalia,Rigidity,Negativism | 97.5 | 96.0 | 98.9 |
| 6 | Mutism,Staring,Posturing/catalepsy,Echopraxia/echolalia,Stereotypy,Negativism | 97.5 | 96.0 | 98.9 |
| 6 | Mutism,Staring,Posturing/catalepsy,Echopraxia/echolalia,Verbigeration,Negativism | 97.5 | 96.0 | 98.9 |
| 6 | Mutism,Staring,Posturing/catalepsy,Grimacing,Negativism,Withdrawal | 97.5 | 96.0 | 98.9 |
| 6 | Mutism,Staring,Posturing/catalepsy,Rigidity,Negativism,Withdrawal | 97.5 | 96.0 | 98.9 |
| 6 | Mutism,Staring,Posturing/catalepsy,Stereotypy,Mannerisms,Rigidity | 97.5 | 96.0 | 98.9 |
| 6 | Mutism,Staring,Posturing/catalepsy,Stereotypy,Mannerisms,Withdrawal | 97.5 | 96.0 | 98.9 |
| 6 | Mutism,Staring,Posturing/catalepsy,Stereotypy,Rigidity,Negativism | 97.5 | 96.0 | 98.9 |
| 6 | Mutism,Staring,Posturing/catalepsy,Stereotypy,Verbigeration,Rigidity | 97.5 | 96.0 | 98.9 |
| 6 | Mutism,Staring,Stereotypy,Mannerisms,Verbigeration,Rigidity | 97.5 | 96.0 | 98.9 |
| 6 | Staring,Echopraxia/echolalia,Mannerisms,Negativism,Waxy flexibility,Withdrawal | 97.5 | 96.0 | 98.9 |
| 6 | Staring,Grimacing,Echopraxia/echolalia,Mannerisms,Negativism,Withdrawal | 97.5 | 96.0 | 98.9 |
| 6 | Staring,Posturing/catalepsy,Echopraxia/echolalia,Rigidity,Negativism,Withdrawal | 97.5 | 96.0 | 98.9 |
| 6 | Staring,Posturing/catalepsy,Grimacing,Mannerisms,Rigidity,Withdrawal | 97.5 | 96.0 | 98.9 |
| 6 | Staring,Posturing/catalepsy,Mannerisms,Negativism,Waxy flexibility,Withdrawal | 97.5 | 96.0 | 98.9 |
| 6 | Staring,Posturing/catalepsy,Stereotypy,Mannerisms,Rigidity,Withdrawal | 97.5 | 96.0 | 98.9 |
| 6 | Staring,Posturing/catalepsy,Stereotypy,Rigidity,Negativism,Withdrawal | 97.5 | 96.0 | 98.9 |
| 6 | Staring,Stereotypy,Mannerisms,Negativism,Waxy flexibility,Withdrawal | 97.5 | 96.0 | 98.9 |
| 6 | Excitement,Immobility/stupor,Echopraxia/echolalia,Rigidity,Negativism,Waxy flexibility | 97.3 | 95.7 | 98.7 |
| 6 | Excitement,Immobility/stupor,Grimacing,Verbigeration,Rigidity,Negativism | 97.3 | 95.7 | 98.7 |
| 6 | Excitement,Immobility/stupor,Mannerisms,Verbigeration,Rigidity,Negativism | 97.3 | 95.7 | 98.7 |
| 6 | Excitement,Immobility/stupor,Mutism,Mannerisms,Verbigeration,Rigidity | 97.3 | 95.7 | 98.7 |
| 6 | Excitement,Immobility/stupor,Mutism,Posturing/catalepsy,Echopraxia/echolalia,Negativism | 97.3 | 95.7 | 98.7 |
| 6 | Excitement,Immobility/stupor,Mutism,Posturing/catalepsy,Grimacing,Negativism | 97.3 | 95.7 | 98.7 |
| 6 | Excitement,Immobility/stupor,Mutism,Staring,Echopraxia/echolalia,Verbigeration | 97.3 | 95.7 | 98.7 |
| 6 | Excitement,Immobility/stupor,Mutism,Staring,Grimacing,Echopraxia/echolalia | 97.3 | 95.7 | 98.7 |
| 6 | Excitement,Immobility/stupor,Mutism,Staring,Grimacing,Waxy flexibility | 97.3 | 95.7 | 98.7 |
| 6 | Excitement,Immobility/stupor,Mutism,Staring,Stereotypy,Withdrawal | 97.3 | 95.7 | 98.7 |
| 6 | Excitement,Immobility/stupor,Mutism,Stereotypy,Rigidity,Waxy flexibility | 97.3 | 95.7 | 98.7 |
| 6 | Excitement,Immobility/stupor,Posturing/catalepsy,Echopraxia/echolalia,Verbigeration,Rigidity | 97.3 | 95.7 | 98.7 |
| 6 | Excitement,Immobility/stupor,Posturing/catalepsy,Grimacing,Echopraxia/echolalia,Rigidity | 97.3 | 95.7 | 98.7 |
| 6 | Excitement,Immobility/stupor,Posturing/catalepsy,Grimacing,Negativism,Withdrawal | 97.3 | 95.7 | 98.7 |
| 6 | Excitement,Immobility/stupor,Posturing/catalepsy,Mannerisms,Negativism,Withdrawal | 97.3 | 95.7 | 98.7 |
| 6 | Excitement,Immobility/stupor,Posturing/catalepsy,Rigidity,Negativism,Waxy flexibility | 97.3 | 95.7 | 98.7 |
| 6 | Excitement,Immobility/stupor,Posturing/catalepsy,Rigidity,Waxy flexibility,Withdrawal | 97.3 | 95.7 | 98.7 |
| 6 | Excitement,Immobility/stupor,Posturing/catalepsy,Stereotypy,Negativism,Withdrawal | 97.3 | 95.7 | 98.7 |
| 6 | Excitement,Immobility/stupor,Rigidity,Negativism,Waxy flexibility,Withdrawal | 97.3 | 95.7 | 98.7 |
| 6 | Excitement,Immobility/stupor,Staring,Echopraxia/echolalia,Verbigeration,Waxy flexibility | 97.3 | 95.7 | 98.7 |
| 6 | Excitement,Immobility/stupor,Staring,Grimacing,Echopraxia/echolalia,Waxy flexibility | 97.3 | 95.7 | 98.7 |
| 6 | Excitement,Immobility/stupor,Staring,Grimacing,Echopraxia/echolalia,Withdrawal | 97.3 | 95.7 | 98.7 |
| 6 | Excitement,Immobility/stupor,Staring,Grimacing,Stereotypy,Negativism | 97.3 | 95.7 | 98.7 |
| 6 | Excitement,Immobility/stupor,Staring,Mannerisms,Verbigeration,Waxy flexibility | 97.3 | 95.7 | 98.7 |
| 6 | Excitement,Immobility/stupor,Staring,Mannerisms,Waxy flexibility,Withdrawal | 97.3 | 95.7 | 98.7 |
| 6 | Excitement,Immobility/stupor,Staring,Posturing/catalepsy,Grimacing,Mannerisms | 97.3 | 95.7 | 98.7 |
| 6 | Excitement,Immobility/stupor,Staring,Posturing/catalepsy,Grimacing,Verbigeration | 97.3 | 95.7 | 98.7 |
| 6 | Excitement,Immobility/stupor,Staring,Posturing/catalepsy,Mannerisms,Waxy flexibility | 97.3 | 95.7 | 98.7 |
| 6 | Excitement,Immobility/stupor,Staring,Posturing/catalepsy,Stereotypy,Withdrawal | 97.3 | 95.7 | 98.7 |
| 6 | Excitement,Immobility/stupor,Stereotypy,Rigidity,Negativism,Waxy flexibility | 97.3 | 95.7 | 98.7 |
| 6 | Excitement,Immobility/stupor,Stereotypy,Rigidity,Waxy flexibility,Withdrawal | 97.3 | 95.7 | 98.7 |
| 6 | Excitement,Mutism,Staring,Echopraxia/echolalia,Stereotypy,Mannerisms | 97.3 | 95.7 | 98.7 |
| 6 | Excitement,Mutism,Staring,Grimacing,Echopraxia/echolalia,Verbigeration | 97.3 | 95.7 | 98.7 |
| 6 | Excitement,Mutism,Staring,Grimacing,Mannerisms,Waxy flexibility | 97.3 | 95.7 | 98.7 |
| 6 | Excitement,Mutism,Staring,Grimacing,Mannerisms,Withdrawal | 97.3 | 95.7 | 98.7 |
| 6 | Excitement,Mutism,Staring,Grimacing,Stereotypy,Verbigeration | 97.3 | 95.7 | 98.7 |
| 6 | Excitement,Mutism,Staring,Mannerisms,Verbigeration,Withdrawal | 97.3 | 95.7 | 98.7 |
| 6 | Excitement,Mutism,Staring,Stereotypy,Mannerisms,Waxy flexibility | 97.3 | 95.7 | 98.7 |
| 6 | Excitement,Staring,Echopraxia/echolalia,Mannerisms,Rigidity,Negativism | 97.3 | 95.7 | 98.7 |
| 6 | Excitement,Staring,Echopraxia/echolalia,Verbigeration,Negativism,Withdrawal | 97.3 | 95.7 | 98.7 |
| 6 | Excitement,Staring,Grimacing,Stereotypy,Negativism,Withdrawal | 97.3 | 95.7 | 98.7 |
| 6 | Excitement,Staring,Posturing/catalepsy,Echopraxia/echolalia,Waxy flexibility,Withdrawal | 97.3 | 95.7 | 98.7 |
| 6 | Excitement,Staring,Posturing/catalepsy,Grimacing,Echopraxia/echolalia,Rigidity | 97.3 | 95.7 | 98.7 |
| 6 | Excitement,Staring,Posturing/catalepsy,Grimacing,Mannerisms,Rigidity | 97.3 | 95.7 | 98.7 |
| 6 | Excitement,Staring,Posturing/catalepsy,Mannerisms,Verbigeration,Withdrawal | 97.3 | 95.7 | 98.7 |
| 6 | Excitement,Staring,Posturing/catalepsy,Mannerisms,Waxy flexibility,Withdrawal | 97.3 | 95.7 | 98.7 |
| 6 | Excitement,Staring,Stereotypy,Mannerisms,Rigidity,Negativism | 97.3 | 95.7 | 98.7 |
| 6 | Excitement,Staring,Verbigeration,Rigidity,Negativism,Waxy flexibility | 97.3 | 95.7 | 98.7 |
| 6 | Immobility/stupor,Mannerisms,Verbigeration,Rigidity,Negativism,Withdrawal | 97.3 | 95.7 | 98.7 |
| 6 | Immobility/stupor,Mutism,Staring,Mannerisms,Rigidity,Withdrawal | 97.3 | 95.7 | 98.7 |
| 6 | Immobility/stupor,Mutism,Staring,Posturing/catalepsy,Mannerisms,Negativism | 97.3 | 95.7 | 98.7 |
| 6 | Immobility/stupor,Mutism,Staring,Stereotypy,Mannerisms,Negativism | 97.3 | 95.7 | 98.7 |
| 6 | Immobility/stupor,Staring,Grimacing,Mannerisms,Negativism,Withdrawal | 97.3 | 95.7 | 98.7 |
| 6 | Immobility/stupor,Staring,Grimacing,Mannerisms,Rigidity,Withdrawal | 97.3 | 95.7 | 98.7 |
| 6 | Immobility/stupor,Staring,Mannerisms,Negativism,Waxy flexibility,Withdrawal | 97.3 | 95.7 | 98.7 |
| 6 | Immobility/stupor,Staring,Posturing/catalepsy,Echopraxia/echolalia,Negativism,Withdrawal | 97.3 | 95.7 | 98.7 |
| 6 | Immobility/stupor,Staring,Posturing/catalepsy,Mannerisms,Rigidity,Withdrawal | 97.3 | 95.7 | 98.7 |
| 6 | Immobility/stupor,Staring,Posturing/catalepsy,Mannerisms,Verbigeration,Negativism | 97.3 | 95.7 | 98.7 |
| 6 | Immobility/stupor,Staring,Posturing/catalepsy,Rigidity,Negativism,Withdrawal | 97.3 | 95.7 | 98.7 |
| 6 | Immobility/stupor,Staring,Posturing/catalepsy,Stereotypy,Negativism,Withdrawal | 97.3 | 95.7 | 98.7 |
| 6 | Immobility/stupor,Staring,Posturing/catalepsy,Stereotypy,Rigidity,Negativism | 97.3 | 95.7 | 98.7 |
| 6 | Immobility/stupor,Staring,Posturing/catalepsy,Stereotypy,Verbigeration,Negativism | 97.3 | 95.7 | 98.7 |
| 6 | Immobility/stupor,Staring,Stereotypy,Mannerisms,Negativism,Waxy flexibility | 97.3 | 95.7 | 98.7 |
| 6 | Immobility/stupor,Staring,Stereotypy,Verbigeration,Negativism,Withdrawal | 97.3 | 95.7 | 98.7 |
| 6 | Immobility/stupor,Stereotypy,Verbigeration,Rigidity,Negativism,Withdrawal | 97.3 | 95.7 | 98.7 |
| 6 | Mutism,Staring,Echopraxia/echolalia,Stereotypy,Mannerisms,Rigidity | 97.3 | 95.7 | 98.7 |
| 6 | Mutism,Staring,Grimacing,Echopraxia/echolalia,Mannerisms,Withdrawal | 97.3 | 95.7 | 98.7 |
| 6 | Mutism,Staring,Grimacing,Stereotypy,Mannerisms,Withdrawal | 97.3 | 95.7 | 98.7 |
| 6 | Mutism,Staring,Grimacing,Stereotypy,Negativism,Withdrawal | 97.3 | 95.7 | 98.7 |
| 6 | Mutism,Staring,Grimacing,Stereotypy,Rigidity,Withdrawal | 97.3 | 95.7 | 98.7 |
| 6 | Mutism,Staring,Grimacing,Stereotypy,Verbigeration,Negativism | 97.3 | 95.7 | 98.7 |
| 6 | Mutism,Staring,Grimacing,Verbigeration,Rigidity,Negativism | 97.3 | 95.7 | 98.7 |
| 6 | Mutism,Staring,Grimacing,Verbigeration,Rigidity,Withdrawal | 97.3 | 95.7 | 98.7 |
| 6 | Mutism,Staring,Posturing/catalepsy,Echopraxia/echolalia,Rigidity,Withdrawal | 97.3 | 95.7 | 98.7 |
| 6 | Mutism,Staring,Posturing/catalepsy,Echopraxia/echolalia,Verbigeration,Rigidity | 97.3 | 95.7 | 98.7 |
| 6 | Mutism,Staring,Posturing/catalepsy,Grimacing,Mannerisms,Rigidity | 97.3 | 95.7 | 98.7 |
| 6 | Mutism,Staring,Posturing/catalepsy,Mannerisms,Negativism,Waxy flexibility | 97.3 | 95.7 | 98.7 |
| 6 | Mutism,Staring,Posturing/catalepsy,Mannerisms,Verbigeration,Withdrawal | 97.3 | 95.7 | 98.7 |
| 6 | Mutism,Staring,Posturing/catalepsy,Stereotypy,Rigidity,Withdrawal | 97.3 | 95.7 | 98.7 |
| 6 | Mutism,Staring,Posturing/catalepsy,Verbigeration,Rigidity,Negativism | 97.3 | 95.7 | 98.7 |
| 6 | Staring,Echopraxia/echolalia,Stereotypy,Mannerisms,Rigidity,Negativism | 97.3 | 95.7 | 98.7 |
| 6 | Staring,Echopraxia/echolalia,Stereotypy,Mannerisms,Rigidity,Withdrawal | 97.3 | 95.7 | 98.7 |
| 6 | Staring,Echopraxia/echolalia,Verbigeration,Rigidity,Negativism,Withdrawal | 97.3 | 95.7 | 98.7 |
| 6 | Staring,Grimacing,Echopraxia/echolalia,Mannerisms,Rigidity,Withdrawal | 97.3 | 95.7 | 98.7 |
| 6 | Staring,Grimacing,Mannerisms,Rigidity,Waxy flexibility,Withdrawal | 97.3 | 95.7 | 98.7 |
| 6 | Staring,Grimacing,Mannerisms,Verbigeration,Rigidity,Withdrawal | 97.3 | 95.7 | 98.7 |
| 6 | Staring,Grimacing,Stereotypy,Rigidity,Negativism,Withdrawal | 97.3 | 95.7 | 98.7 |
| 6 | Staring,Grimacing,Stereotypy,Verbigeration,Rigidity,Withdrawal | 97.3 | 95.7 | 98.7 |
| 6 | Staring,Posturing/catalepsy,Echopraxia/echolalia,Mannerisms,Rigidity,Negativism | 97.3 | 95.7 | 98.7 |
| 6 | Staring,Posturing/catalepsy,Stereotypy,Mannerisms,Rigidity,Negativism | 97.3 | 95.7 | 98.7 |
| 6 | Staring,Posturing/catalepsy,Verbigeration,Rigidity,Negativism,Withdrawal | 97.3 | 95.7 | 98.7 |
| 6 | Staring,Stereotypy,Mannerisms,Rigidity,Negativism,Waxy flexibility | 97.3 | 95.7 | 98.7 |
| 6 | Staring,Stereotypy,Mannerisms,Rigidity,Waxy flexibility,Withdrawal | 97.3 | 95.7 | 98.7 |
| 6 | Staring,Stereotypy,Mannerisms,Verbigeration,Rigidity,Negativism | 97.3 | 95.7 | 98.7 |
| 6 | Staring,Stereotypy,Mannerisms,Verbigeration,Rigidity,Withdrawal | 97.3 | 95.7 | 98.7 |
| 6 | Staring,Stereotypy,Rigidity,Negativism,Waxy flexibility,Withdrawal | 97.3 | 95.7 | 98.7 |
| 6 | Excitement,Immobility/stupor,Echopraxia/echolalia,Mannerisms,Rigidity,Negativism | 97.1 | 95.5 | 98.4 |
| 6 | Excitement,Immobility/stupor,Echopraxia/echolalia,Stereotypy,Negativism,Withdrawal | 97.1 | 95.5 | 98.7 |
| 6 | Excitement,Immobility/stupor,Grimacing,Mannerisms,Rigidity,Negativism | 97.1 | 95.5 | 98.4 |
| 6 | Excitement,Immobility/stupor,Grimacing,Mannerisms,Rigidity,Withdrawal | 97.1 | 95.5 | 98.4 |
| 6 | Excitement,Immobility/stupor,Grimacing,Rigidity,Negativism,Waxy flexibility | 97.1 | 95.3 | 98.4 |
| 6 | Excitement,Immobility/stupor,Grimacing,Stereotypy,Rigidity,Negativism | 97.1 | 95.5 | 98.4 |
| 6 | Excitement,Immobility/stupor,Grimacing,Stereotypy,Rigidity,Withdrawal | 97.1 | 95.3 | 98.7 |
| 6 | Excitement,Immobility/stupor,Grimacing,Verbigeration,Rigidity,Withdrawal | 97.1 | 95.5 | 98.4 |
| 6 | Excitement,Immobility/stupor,Mannerisms,Rigidity,Waxy flexibility,Withdrawal | 97.1 | 95.5 | 98.7 |
| 6 | Excitement,Immobility/stupor,Mutism,Echopraxia/echolalia,Mannerisms,Rigidity | 97.1 | 95.5 | 98.4 |
| 6 | Excitement,Immobility/stupor,Mutism,Grimacing,Rigidity,Waxy flexibility | 97.1 | 95.5 | 98.7 |
| 6 | Excitement,Immobility/stupor,Mutism,Grimacing,Stereotypy,Rigidity | 97.1 | 95.5 | 98.4 |
| 6 | Excitement,Immobility/stupor,Mutism,Posturing/catalepsy,Echopraxia/echolalia,Withdrawal | 97.1 | 95.5 | 98.4 |
| 6 | Excitement,Immobility/stupor,Mutism,Posturing/catalepsy,Rigidity,Waxy flexibility | 97.1 | 95.5 | 98.7 |
| 6 | Excitement,Immobility/stupor,Mutism,Posturing/catalepsy,Stereotypy,Negativism | 97.1 | 95.5 | 98.7 |
| 6 | Excitement,Immobility/stupor,Mutism,Staring,Echopraxia/echolalia,Mannerisms | 97.1 | 95.3 | 98.4 |
| 6 | Excitement,Immobility/stupor,Mutism,Staring,Grimacing,Withdrawal | 97.1 | 95.3 | 98.7 |
| 6 | Excitement,Immobility/stupor,Mutism,Staring,Mannerisms,Withdrawal | 97.1 | 95.5 | 98.4 |
| 6 | Excitement,Immobility/stupor,Mutism,Staring,Stereotypy,Verbigeration | 97.1 | 95.5 | 98.4 |
| 6 | Excitement,Immobility/stupor,Mutism,Staring,Verbigeration,Withdrawal | 97.1 | 95.5 | 98.7 |
| 6 | Excitement,Immobility/stupor,Posturing/catalepsy,Grimacing,Verbigeration,Rigidity | 97.1 | 95.5 | 98.7 |
| 6 | Excitement,Immobility/stupor,Staring,Echopraxia/echolalia,Mannerisms,Waxy flexibility | 97.1 | 95.5 | 98.4 |
| 6 | Excitement,Immobility/stupor,Staring,Echopraxia/echolalia,Stereotypy,Waxy flexibility | 97.1 | 95.5 | 98.4 |
| 6 | Excitement,Immobility/stupor,Staring,Echopraxia/echolalia,Verbigeration,Withdrawal | 97.1 | 95.5 | 98.7 |
| 6 | Excitement,Immobility/stupor,Staring,Grimacing,Mannerisms,Negativism | 97.1 | 95.5 | 98.4 |
| 6 | Excitement,Immobility/stupor,Staring,Mannerisms,Verbigeration,Withdrawal | 97.1 | 95.5 | 98.7 |
| 6 | Excitement,Immobility/stupor,Staring,Posturing/catalepsy,Echopraxia/echolalia,Stereotypy | 97.1 | 95.5 | 98.4 |
| 6 | Excitement,Immobility/stupor,Staring,Posturing/catalepsy,Stereotypy,Verbigeration | 97.1 | 95.5 | 98.4 |
| 6 | Excitement,Immobility/stupor,Staring,Stereotypy,Verbigeration,Waxy flexibility | 97.1 | 95.5 | 98.7 |
| 6 | Excitement,Immobility/stupor,Staring,Stereotypy,Waxy flexibility,Withdrawal | 97.1 | 95.5 | 98.7 |
| 6 | Excitement,Immobility/stupor,Staring,Verbigeration,Waxy flexibility,Withdrawal | 97.1 | 95.5 | 98.4 |
| 6 | Excitement,Immobility/stupor,Stereotypy,Mannerisms,Rigidity,Withdrawal | 97.1 | 95.3 | 98.4 |
| 6 | Excitement,Immobility/stupor,Stereotypy,Verbigeration,Negativism,Withdrawal | 97.1 | 95.5 | 98.4 |
| 6 | Excitement,Immobility/stupor,Verbigeration,Rigidity,Negativism,Waxy flexibility | 97.1 | 95.5 | 98.4 |
| 6 | Excitement,Immobility/stupor,Verbigeration,Rigidity,Waxy flexibility,Withdrawal | 97.1 | 95.5 | 98.4 |
| 6 | Excitement,Mutism,Staring,Grimacing,Echopraxia/echolalia,Mannerisms | 97.1 | 95.5 | 98.7 |
| 6 | Excitement,Mutism,Staring,Mannerisms,Verbigeration,Waxy flexibility | 97.1 | 95.5 | 98.4 |
| 6 | Excitement,Staring,Echopraxia/echolalia,Mannerisms,Waxy flexibility,Withdrawal | 97.1 | 95.3 | 98.7 |
| 6 | Excitement,Staring,Echopraxia/echolalia,Verbigeration,Rigidity,Negativism | 97.1 | 95.5 | 98.7 |
| 6 | Excitement,Staring,Grimacing,Echopraxia/echolalia,Rigidity,Waxy flexibility | 97.1 | 95.5 | 98.4 |
| 6 | Excitement,Staring,Grimacing,Echopraxia/echolalia,Waxy flexibility,Withdrawal | 97.1 | 95.5 | 98.4 |
| 6 | Excitement,Staring,Grimacing,Mannerisms,Negativism,Withdrawal | 97.1 | 95.5 | 98.7 |
| 6 | Excitement,Staring,Grimacing,Mannerisms,Rigidity,Waxy flexibility | 97.1 | 95.5 | 98.4 |
| 6 | Excitement,Staring,Grimacing,Negativism,Waxy flexibility,Withdrawal | 97.1 | 95.5 | 98.4 |
| 6 | Excitement,Staring,Grimacing,Stereotypy,Rigidity,Waxy flexibility | 97.1 | 95.5 | 98.7 |
| 6 | Excitement,Staring,Grimacing,Verbigeration,Negativism,Withdrawal | 97.1 | 95.5 | 98.4 |
| 6 | Excitement,Staring,Mannerisms,Verbigeration,Rigidity,Negativism | 97.1 | 95.5 | 98.4 |
| 6 | Excitement,Staring,Posturing/catalepsy,Echopraxia/echolalia,Mannerisms,Negativism | 97.1 | 95.5 | 98.4 |
| 6 | Excitement,Staring,Posturing/catalepsy,Echopraxia/echolalia,Negativism,Waxy flexibility | 97.1 | 95.5 | 98.4 |
| 6 | Excitement,Staring,Posturing/catalepsy,Grimacing,Rigidity,Negativism | 97.1 | 95.5 | 98.7 |
| 6 | Excitement,Staring,Posturing/catalepsy,Grimacing,Stereotypy,Rigidity | 97.1 | 95.5 | 98.4 |
| 6 | Excitement,Staring,Posturing/catalepsy,Grimacing,Verbigeration,Rigidity | 97.1 | 95.5 | 98.7 |
| 6 | Excitement,Staring,Posturing/catalepsy,Grimacing,Verbigeration,Withdrawal | 97.1 | 95.5 | 98.4 |
| 6 | Excitement,Staring,Posturing/catalepsy,Mannerisms,Verbigeration,Negativism | 97.1 | 95.3 | 98.4 |
| 6 | Excitement,Staring,Posturing/catalepsy,Mannerisms,Verbigeration,Rigidity | 97.1 | 95.5 | 98.4 |
| 6 | Excitement,Staring,Posturing/catalepsy,Rigidity,Negativism,Waxy flexibility | 97.1 | 95.5 | 98.4 |
| 6 | Immobility/stupor,Grimacing,Mannerisms,Rigidity,Negativism,Withdrawal | 97.1 | 95.5 | 98.4 |
| 6 | Immobility/stupor,Mutism,Echopraxia/echolalia,Mannerisms,Rigidity,Negativism | 97.1 | 95.3 | 98.7 |
| 6 | Immobility/stupor,Mutism,Mannerisms,Rigidity,Negativism,Withdrawal | 97.1 | 95.5 | 98.7 |
| 6 | Immobility/stupor,Mutism,Mannerisms,Verbigeration,Rigidity,Negativism | 97.1 | 95.5 | 98.4 |
| 6 | Immobility/stupor,Mutism,Staring,Echopraxia/echolalia,Mannerisms,Negativism | 97.1 | 95.5 | 98.4 |
| 6 | Immobility/stupor,Mutism,Staring,Mannerisms,Verbigeration,Rigidity | 97.1 | 95.5 | 98.4 |
| 6 | Immobility/stupor,Mutism,Staring,Posturing/catalepsy,Negativism,Withdrawal | 97.1 | 95.5 | 98.4 |
| 6 | Immobility/stupor,Mutism,Stereotypy,Mannerisms,Rigidity,Negativism | 97.1 | 95.5 | 98.7 |
| 6 | Immobility/stupor,Staring,Echopraxia/echolalia,Mannerisms,Negativism,Waxy flexibility | 97.1 | 95.3 | 98.4 |
| 6 | Immobility/stupor,Staring,Echopraxia/echolalia,Mannerisms,Rigidity,Withdrawal | 97.1 | 95.5 | 98.7 |
| 6 | Immobility/stupor,Staring,Echopraxia/echolalia,Stereotypy,Negativism,Withdrawal | 97.1 | 95.5 | 98.4 |
| 6 | Immobility/stupor,Staring,Echopraxia/echolalia,Stereotypy,Rigidity,Negativism | 97.1 | 95.3 | 98.4 |
| 6 | Immobility/stupor,Staring,Echopraxia/echolalia,Verbigeration,Rigidity,Negativism | 97.1 | 95.3 | 98.7 |
| 6 | Immobility/stupor,Staring,Grimacing,Echopraxia/echolalia,Mannerisms,Negativism | 97.1 | 95.5 | 98.4 |
| 6 | Immobility/stupor,Staring,Grimacing,Stereotypy,Mannerisms,Negativism | 97.1 | 95.5 | 98.7 |
| 6 | Immobility/stupor,Staring,Grimacing,Stereotypy,Mannerisms,Rigidity | 97.1 | 95.3 | 98.7 |
| 6 | Immobility/stupor,Staring,Grimacing,Stereotypy,Verbigeration,Rigidity | 97.1 | 95.5 | 98.4 |
| 6 | Immobility/stupor,Staring,Grimacing,Verbigeration,Rigidity,Negativism | 97.1 | 95.5 | 98.7 |
| 6 | Immobility/stupor,Staring,Mannerisms,Verbigeration,Rigidity,Withdrawal | 97.1 | 95.5 | 98.4 |
| 6 | Immobility/stupor,Staring,Posturing/catalepsy,Grimacing,Mannerisms,Negativism | 97.1 | 95.5 | 98.4 |
| 6 | Immobility/stupor,Staring,Posturing/catalepsy,Verbigeration,Negativism,Withdrawal | 97.1 | 95.5 | 98.4 |
| 6 | Immobility/stupor,Staring,Stereotypy,Mannerisms,Verbigeration,Rigidity | 97.1 | 95.5 | 98.7 |
| 6 | Immobility/stupor,Stereotypy,Mannerisms,Verbigeration,Rigidity,Negativism | 97.1 | 95.5 | 98.4 |
| 6 | Mutism,Staring,Echopraxia/echolalia,Mannerisms,Verbigeration,Rigidity | 97.1 | 95.5 | 98.4 |
| 6 | Mutism,Staring,Echopraxia/echolalia,Stereotypy,Mannerisms,Withdrawal | 97.1 | 95.5 | 98.4 |
| 6 | Mutism,Staring,Echopraxia/echolalia,Stereotypy,Verbigeration,Negativism | 97.1 | 95.5 | 98.4 |
| 6 | Mutism,Staring,Echopraxia/echolalia,Verbigeration,Negativism,Withdrawal | 97.1 | 95.5 | 98.4 |
| 6 | Mutism,Staring,Grimacing,Echopraxia/echolalia,Negativism,Withdrawal | 97.1 | 95.5 | 98.4 |
| 6 | Mutism,Staring,Grimacing,Echopraxia/echolalia,Rigidity,Negativism | 97.1 | 95.5 | 98.4 |
| 6 | Mutism,Staring,Grimacing,Echopraxia/echolalia,Rigidity,Withdrawal | 97.1 | 95.5 | 98.4 |
| 6 | Mutism,Staring,Grimacing,Echopraxia/echolalia,Stereotypy,Negativism | 97.1 | 95.5 | 98.7 |
| 6 | Mutism,Staring,Grimacing,Echopraxia/echolalia,Verbigeration,Negativism | 97.1 | 95.5 | 98.4 |
| 6 | Mutism,Staring,Grimacing,Echopraxia/echolalia,Verbigeration,Rigidity | 97.1 | 95.5 | 98.4 |
| 6 | Mutism,Staring,Grimacing,Mannerisms,Negativism,Waxy flexibility | 97.1 | 95.5 | 98.4 |
| 6 | Mutism,Staring,Grimacing,Mannerisms,Rigidity,Waxy flexibility | 97.1 | 95.5 | 98.7 |
| 6 | Mutism,Staring,Grimacing,Mannerisms,Verbigeration,Negativism | 97.1 | 95.5 | 98.4 |
| 6 | Mutism,Staring,Grimacing,Mannerisms,Waxy flexibility,Withdrawal | 97.1 | 95.5 | 98.4 |
| 6 | Mutism,Staring,Grimacing,Rigidity,Negativism,Withdrawal | 97.1 | 95.5 | 98.7 |
| 6 | Mutism,Staring,Grimacing,Stereotypy,Rigidity,Negativism | 97.1 | 95.5 | 98.7 |
| 6 | Mutism,Staring,Grimacing,Verbigeration,Negativism,Withdrawal | 97.1 | 95.5 | 98.4 |
| 6 | Mutism,Staring,Mannerisms,Verbigeration,Negativism,Waxy flexibility | 97.1 | 95.3 | 98.4 |
| 6 | Mutism,Staring,Posturing/catalepsy,Echopraxia/echolalia,Stereotypy,Rigidity | 97.1 | 95.5 | 98.7 |
| 6 | Mutism,Staring,Posturing/catalepsy,Grimacing,Echopraxia/echolalia,Negativism | 97.1 | 95.5 | 98.4 |
| 6 | Mutism,Staring,Posturing/catalepsy,Grimacing,Stereotypy,Negativism | 97.1 | 95.5 | 98.7 |
| 6 | Mutism,Staring,Posturing/catalepsy,Grimacing,Verbigeration,Negativism | 97.1 | 95.5 | 98.7 |
| 6 | Mutism,Staring,Posturing/catalepsy,Mannerisms,Waxy flexibility,Withdrawal | 97.1 | 95.5 | 98.4 |
| 6 | Mutism,Staring,Posturing/catalepsy,Negativism,Waxy flexibility,Withdrawal | 97.1 | 95.5 | 98.4 |
| 6 | Mutism,Staring,Posturing/catalepsy,Verbigeration,Rigidity,Withdrawal | 97.1 | 95.5 | 98.4 |
| 6 | Mutism,Staring,Rigidity,Negativism,Waxy flexibility,Withdrawal | 97.1 | 95.5 | 98.7 |
| 6 | Mutism,Staring,Stereotypy,Mannerisms,Rigidity,Waxy flexibility | 97.1 | 95.5 | 98.4 |
| 6 | Mutism,Staring,Stereotypy,Negativism,Waxy flexibility,Withdrawal | 97.1 | 95.5 | 98.4 |
| 6 | Mutism,Staring,Stereotypy,Rigidity,Negativism,Waxy flexibility | 97.1 | 95.5 | 98.4 |
| 6 | Mutism,Staring,Stereotypy,Rigidity,Waxy flexibility,Withdrawal | 97.1 | 95.3 | 98.7 |
| 6 | Mutism,Staring,Stereotypy,Verbigeration,Negativism,Waxy flexibility | 97.1 | 95.5 | 98.4 |
| 6 | Mutism,Staring,Stereotypy,Verbigeration,Rigidity,Waxy flexibility | 97.1 | 95.5 | 98.4 |
| 6 | Staring,Echopraxia/echolalia,Mannerisms,Rigidity,Negativism,Waxy flexibility | 97.1 | 95.5 | 98.4 |
| 6 | Staring,Echopraxia/echolalia,Rigidity,Negativism,Waxy flexibility,Withdrawal | 97.1 | 95.5 | 98.4 |
| 6 | Staring,Grimacing,Echopraxia/echolalia,Rigidity,Negativism,Withdrawal | 97.1 | 95.5 | 98.7 |
| 6 | Staring,Grimacing,Mannerisms,Negativism,Waxy flexibility,Withdrawal | 97.1 | 95.5 | 98.4 |
| 6 | Staring,Grimacing,Mannerisms,Verbigeration,Negativism,Withdrawal | 97.1 | 95.5 | 98.4 |
| 6 | Staring,Grimacing,Stereotypy,Verbigeration,Negativism,Withdrawal | 97.1 | 95.5 | 98.7 |
| 6 | Staring,Grimacing,Verbigeration,Rigidity,Negativism,Withdrawal | 97.1 | 95.5 | 98.4 |
| 6 | Staring,Mannerisms,Verbigeration,Negativism,Waxy flexibility,Withdrawal | 97.1 | 95.5 | 98.4 |
| 6 | Staring,Posturing/catalepsy,Echopraxia/echolalia,Mannerisms,Rigidity,Withdrawal | 97.1 | 95.3 | 98.4 |
| 6 | Staring,Posturing/catalepsy,Mannerisms,Verbigeration,Rigidity,Withdrawal | 97.1 | 95.5 | 98.7 |
| 6 | Staring,Posturing/catalepsy,Stereotypy,Verbigeration,Negativism,Withdrawal | 97.1 | 95.5 | 98.4 |
| 6 | Staring,Verbigeration,Rigidity,Negativism,Waxy flexibility,Withdrawal | 97.1 | 95.5 | 98.7 |
| 6 | Excitement,Immobility/stupor,Echopraxia/echolalia,Verbigeration,Negativism,Withdrawal | 96.9 | 95.1 | 98.4 |
| 6 | Excitement,Immobility/stupor,Grimacing,Rigidity,Waxy flexibility,Withdrawal | 96.9 | 95.1 | 98.4 |
| 6 | Excitement,Immobility/stupor,Mannerisms,Rigidity,Negativism,Waxy flexibility | 96.9 | 95.1 | 98.4 |
| 6 | Excitement,Immobility/stupor,Mutism,Posturing/catalepsy,Grimacing,Withdrawal | 96.9 | 95.1 | 98.4 |
| 6 | Excitement,Immobility/stupor,Mutism,Posturing/catalepsy,Mannerisms,Negativism | 96.9 | 95.1 | 98.4 |
| 6 | Excitement,Immobility/stupor,Mutism,Posturing/catalepsy,Negativism,Waxy flexibility | 96.9 | 95.1 | 98.4 |
| 6 | Excitement,Immobility/stupor,Mutism,Staring,Grimacing,Stereotypy | 96.9 | 95.1 | 98.4 |
| 6 | Excitement,Immobility/stupor,Mutism,Staring,Grimacing,Verbigeration | 96.9 | 95.3 | 98.4 |
| 6 | Excitement,Immobility/stupor,Mutism,Staring,Mannerisms,Verbigeration | 96.9 | 95.1 | 98.4 |
| 6 | Excitement,Immobility/stupor,Mutism,Stereotypy,Negativism,Withdrawal | 96.9 | 95.1 | 98.4 |
| 6 | Excitement,Immobility/stupor,Posturing/catalepsy,Grimacing,Echopraxia/echolalia,Negativism | 96.9 | 95.1 | 98.4 |
| 6 | Excitement,Immobility/stupor,Posturing/catalepsy,Grimacing,Mannerisms,Rigidity | 96.9 | 95.1 | 98.4 |
| 6 | Excitement,Immobility/stupor,Posturing/catalepsy,Mannerisms,Verbigeration,Rigidity | 96.9 | 95.1 | 98.4 |
| 6 | Excitement,Immobility/stupor,Posturing/catalepsy,Negativism,Waxy flexibility,Withdrawal | 96.9 | 95.1 | 98.4 |
| 6 | Excitement,Immobility/stupor,Staring,Grimacing,Verbigeration,Waxy flexibility | 96.9 | 95.1 | 98.4 |
| 6 | Excitement,Immobility/stupor,Staring,Grimacing,Waxy flexibility,Withdrawal | 96.9 | 95.1 | 98.4 |
| 6 | Excitement,Immobility/stupor,Staring,Posturing/catalepsy,Grimacing,Stereotypy | 96.9 | 95.1 | 98.4 |
| 6 | Excitement,Immobility/stupor,Staring,Posturing/catalepsy,Grimacing,Waxy flexibility | 96.9 | 95.3 | 98.4 |
| 6 | Excitement,Immobility/stupor,Staring,Posturing/catalepsy,Stereotypy,Mannerisms | 96.9 | 95.1 | 98.4 |
| 6 | Excitement,Immobility/stupor,Staring,Posturing/catalepsy,Stereotypy,Waxy flexibility | 96.9 | 95.1 | 98.4 |
| 6 | Excitement,Immobility/stupor,Staring,Stereotypy,Mannerisms,Withdrawal | 96.9 | 95.1 | 98.4 |
| 6 | Excitement,Immobility/stupor,Staring,Stereotypy,Verbigeration,Withdrawal | 96.9 | 95.1 | 98.4 |
| 6 | Excitement,Immobility/stupor,Stereotypy,Mannerisms,Rigidity,Negativism | 96.9 | 95.1 | 98.4 |
| 6 | Excitement,Mutism,Staring,Stereotypy,Mannerisms,Verbigeration | 96.9 | 95.1 | 98.4 |
| 6 | Excitement,Staring,Echopraxia/echolalia,Stereotypy,Waxy flexibility,Withdrawal | 96.9 | 95.1 | 98.4 |
| 6 | Excitement,Staring,Grimacing,Echopraxia/echolalia,Mannerisms,Withdrawal | 96.9 | 95.3 | 98.4 |
| 6 | Excitement,Staring,Grimacing,Echopraxia/echolalia,Rigidity,Negativism | 96.9 | 95.1 | 98.4 |
| 6 | Excitement,Staring,Grimacing,Echopraxia/echolalia,Stereotypy,Rigidity | 96.9 | 95.3 | 98.4 |
| 6 | Excitement,Staring,Grimacing,Echopraxia/echolalia,Stereotypy,Withdrawal | 96.9 | 95.1 | 98.4 |
| 6 | Excitement,Staring,Grimacing,Mannerisms,Waxy flexibility,Withdrawal | 96.9 | 95.1 | 98.4 |
| 6 | Excitement,Staring,Grimacing,Rigidity,Negativism,Waxy flexibility | 96.9 | 95.1 | 98.4 |
| 6 | Excitement,Staring,Grimacing,Stereotypy,Rigidity,Negativism | 96.9 | 95.1 | 98.4 |
| 6 | Excitement,Staring,Grimacing,Stereotypy,Verbigeration,Rigidity | 96.9 | 95.1 | 98.4 |
| 6 | Excitement,Staring,Grimacing,Verbigeration,Rigidity,Waxy flexibility | 96.9 | 95.1 | 98.4 |
| 6 | Excitement,Staring,Posturing/catalepsy,Echopraxia/echolalia,Mannerisms,Rigidity | 96.9 | 95.1 | 98.4 |
| 6 | Excitement,Staring,Posturing/catalepsy,Echopraxia/echolalia,Stereotypy,Negativism | 96.9 | 95.1 | 98.4 |
| 6 | Excitement,Staring,Posturing/catalepsy,Echopraxia/echolalia,Stereotypy,Withdrawal | 96.9 | 95.1 | 98.4 |
| 6 | Excitement,Staring,Posturing/catalepsy,Echopraxia/echolalia,Verbigeration,Negativism | 96.9 | 95.3 | 98.4 |
| 6 | Excitement,Staring,Posturing/catalepsy,Grimacing,Echopraxia/echolalia,Negativism | 96.9 | 95.1 | 98.4 |
| 6 | Excitement,Staring,Posturing/catalepsy,Grimacing,Stereotypy,Withdrawal | 96.9 | 95.1 | 98.4 |
| 6 | Excitement,Staring,Posturing/catalepsy,Grimacing,Waxy flexibility,Withdrawal | 96.9 | 95.1 | 98.4 |
| 6 | Excitement,Staring,Posturing/catalepsy,Stereotypy,Mannerisms,Withdrawal | 96.9 | 95.1 | 98.4 |
| 6 | Excitement,Staring,Posturing/catalepsy,Stereotypy,Verbigeration,Negativism | 96.9 | 95.1 | 98.4 |
| 6 | Excitement,Staring,Posturing/catalepsy,Verbigeration,Negativism,Waxy flexibility | 96.9 | 95.1 | 98.4 |
| 6 | Excitement,Staring,Posturing/catalepsy,Verbigeration,Waxy flexibility,Withdrawal | 96.9 | 95.1 | 98.4 |
| 6 | Immobility/stupor,Echopraxia/echolalia,Stereotypy,Rigidity,Negativism,Withdrawal | 96.9 | 95.3 | 98.4 |
| 6 | Immobility/stupor,Mannerisms,Rigidity,Negativism,Waxy flexibility,Withdrawal | 96.9 | 95.1 | 98.4 |
| 6 | Immobility/stupor,Mutism,Posturing/catalepsy,Mannerisms,Rigidity,Negativism | 96.9 | 95.1 | 98.4 |
| 6 | Immobility/stupor,Mutism,Staring,Echopraxia/echolalia,Mannerisms,Rigidity | 96.9 | 95.3 | 98.4 |
| 6 | Immobility/stupor,Mutism,Staring,Posturing/catalepsy,Mannerisms,Rigidity | 96.9 | 95.1 | 98.4 |
| 6 | Immobility/stupor,Mutism,Staring,Posturing/catalepsy,Mannerisms,Withdrawal | 96.9 | 95.1 | 98.4 |
| 6 | Immobility/stupor,Mutism,Staring,Rigidity,Negativism,Withdrawal | 96.9 | 95.1 | 98.4 |
| 6 | Immobility/stupor,Mutism,Staring,Stereotypy,Mannerisms,Rigidity | 96.9 | 95.1 | 98.4 |
| 6 | Immobility/stupor,Mutism,Staring,Stereotypy,Rigidity,Negativism | 96.9 | 95.1 | 98.4 |
| 6 | Immobility/stupor,Mutism,Staring,Stereotypy,Verbigeration,Rigidity | 96.9 | 95.1 | 98.4 |
| 6 | Immobility/stupor,Mutism,Staring,Verbigeration,Rigidity,Negativism | 96.9 | 95.1 | 98.4 |
| 6 | Immobility/stupor,Mutism,Stereotypy,Verbigeration,Rigidity,Negativism | 96.9 | 95.1 | 98.4 |
| 6 | Immobility/stupor,Posturing/catalepsy,Echopraxia/echolalia,Mannerisms,Rigidity,Negativism | 96.9 | 95.1 | 98.4 |
| 6 | Immobility/stupor,Posturing/catalepsy,Mannerisms,Verbigeration,Rigidity,Negativism | 96.9 | 95.1 | 98.4 |
| 6 | Immobility/stupor,Posturing/catalepsy,Stereotypy,Mannerisms,Rigidity,Negativism | 96.9 | 95.1 | 98.4 |
| 6 | Immobility/stupor,Staring,Grimacing,Mannerisms,Verbigeration,Rigidity | 96.9 | 95.1 | 98.4 |
| 6 | Immobility/stupor,Staring,Grimacing,Rigidity,Negativism,Withdrawal | 96.9 | 95.1 | 98.4 |
| 6 | Immobility/stupor,Staring,Grimacing,Stereotypy,Rigidity,Negativism | 96.9 | 95.1 | 98.4 |
| 6 | Immobility/stupor,Staring,Grimacing,Stereotypy,Verbigeration,Negativism | 96.9 | 95.1 | 98.4 |
| 6 | Immobility/stupor,Staring,Mannerisms,Rigidity,Waxy flexibility,Withdrawal | 96.9 | 95.1 | 98.4 |
| 6 | Immobility/stupor,Staring,Posturing/catalepsy,Echopraxia/echolalia,Rigidity,Negativism | 96.9 | 95.1 | 98.4 |
| 6 | Immobility/stupor,Staring,Posturing/catalepsy,Echopraxia/echolalia,Stereotypy,Negativism | 96.9 | 95.1 | 98.4 |
| 6 | Immobility/stupor,Staring,Posturing/catalepsy,Mannerisms,Verbigeration,Rigidity | 96.9 | 95.1 | 98.4 |
| 6 | Immobility/stupor,Staring,Rigidity,Negativism,Waxy flexibility,Withdrawal | 96.9 | 95.1 | 98.4 |
| 6 | Immobility/stupor,Staring,Stereotypy,Rigidity,Negativism,Waxy flexibility | 96.9 | 95.1 | 98.4 |
| 6 | Immobility/stupor,Staring,Stereotypy,Verbigeration,Rigidity,Withdrawal | 96.9 | 95.1 | 98.4 |
| 6 | Immobility/stupor,Stereotypy,Mannerisms,Rigidity,Negativism,Waxy flexibility | 96.9 | 95.1 | 98.4 |
| 6 | Mutism,Staring,Echopraxia/echolalia,Mannerisms,Rigidity,Waxy flexibility | 96.9 | 95.1 | 98.4 |
| 6 | Mutism,Staring,Echopraxia/echolalia,Negativism,Waxy flexibility,Withdrawal | 96.9 | 95.3 | 98.4 |
| 6 | Mutism,Staring,Echopraxia/echolalia,Rigidity,Negativism,Waxy flexibility | 96.9 | 95.1 | 98.4 |
| 6 | Mutism,Staring,Echopraxia/echolalia,Stereotypy,Negativism,Waxy flexibility | 96.9 | 95.1 | 98.4 |
| 6 | Mutism,Staring,Echopraxia/echolalia,Stereotypy,Verbigeration,Rigidity | 96.9 | 95.1 | 98.4 |
| 6 | Mutism,Staring,Echopraxia/echolalia,Verbigeration,Negativism,Waxy flexibility | 96.9 | 95.1 | 98.4 |
| 6 | Mutism,Staring,Echopraxia/echolalia,Verbigeration,Rigidity,Withdrawal | 96.9 | 95.1 | 98.4 |
| 6 | Mutism,Staring,Grimacing,Echopraxia/echolalia,Stereotypy,Mannerisms | 96.9 | 95.1 | 98.4 |
| 6 | Mutism,Staring,Grimacing,Echopraxia/echolalia,Stereotypy,Rigidity | 96.9 | 95.1 | 98.4 |
| 6 | Mutism,Staring,Grimacing,Echopraxia/echolalia,Stereotypy,Withdrawal | 96.9 | 95.1 | 98.4 |
| 6 | Mutism,Staring,Grimacing,Mannerisms,Verbigeration,Withdrawal | 96.9 | 95.1 | 98.4 |
| 6 | Mutism,Staring,Grimacing,Stereotypy,Verbigeration,Withdrawal | 96.9 | 95.1 | 98.4 |
| 6 | Mutism,Staring,Mannerisms,Verbigeration,Rigidity,Waxy flexibility | 96.9 | 95.1 | 98.4 |
| 6 | Mutism,Staring,Posturing/catalepsy,Grimacing,Echopraxia/echolalia,Mannerisms | 96.9 | 95.1 | 98.4 |
| 6 | Mutism,Staring,Posturing/catalepsy,Grimacing,Echopraxia/echolalia,Withdrawal | 96.9 | 95.1 | 98.4 |
| 6 | Mutism,Staring,Posturing/catalepsy,Grimacing,Stereotypy,Withdrawal | 96.9 | 95.1 | 98.4 |
| 6 | Mutism,Staring,Posturing/catalepsy,Grimacing,Verbigeration,Rigidity | 96.9 | 95.1 | 98.4 |
| 6 | Mutism,Staring,Posturing/catalepsy,Stereotypy,Negativism,Waxy flexibility | 96.9 | 95.1 | 98.4 |
| 6 | Mutism,Staring,Verbigeration,Negativism,Waxy flexibility,Withdrawal | 96.9 | 95.1 | 98.4 |
| 6 | Staring,Echopraxia/echolalia,Mannerisms,Verbigeration,Negativism,Withdrawal | 96.9 | 95.3 | 98.4 |
| 6 | Staring,Grimacing,Echopraxia/echolalia,Stereotypy,Mannerisms,Withdrawal | 96.9 | 95.1 | 98.4 |
| 6 | Staring,Grimacing,Echopraxia/echolalia,Stereotypy,Negativism,Withdrawal | 96.9 | 95.1 | 98.4 |
| 6 | Staring,Grimacing,Echopraxia/echolalia,Stereotypy,Rigidity,Withdrawal | 96.9 | 95.1 | 98.4 |
| 6 | Staring,Grimacing,Stereotypy,Mannerisms,Rigidity,Negativism | 96.9 | 95.1 | 98.4 |
| 6 | Staring,Posturing/catalepsy,Echopraxia/echolalia,Stereotypy,Negativism,Withdrawal | 96.9 | 95.1 | 98.4 |
| 6 | Staring,Posturing/catalepsy,Grimacing,Echopraxia/echolalia,Negativism,Withdrawal | 96.9 | 95.1 | 98.4 |
| 6 | Staring,Posturing/catalepsy,Grimacing,Rigidity,Negativism,Withdrawal | 96.9 | 95.3 | 98.4 |
| 6 | Staring,Posturing/catalepsy,Grimacing,Stereotypy,Mannerisms,Withdrawal | 96.9 | 95.1 | 98.4 |
| 6 | Staring,Posturing/catalepsy,Grimacing,Stereotypy,Negativism,Withdrawal | 96.9 | 95.1 | 98.4 |
| 6 | Staring,Posturing/catalepsy,Mannerisms,Verbigeration,Rigidity,Negativism | 96.9 | 95.1 | 98.4 |
| 6 | Excitement,Immobility/stupor,Grimacing,Echopraxia/echolalia,Rigidity,Waxy flexibility | 96.8 | 95.1 | 98.4 |
| 6 | Excitement,Immobility/stupor,Mutism,Grimacing,Mannerisms,Rigidity | 96.8 | 95.1 | 98.4 |
| 6 | Excitement,Immobility/stupor,Mutism,Mannerisms,Rigidity,Waxy flexibility | 96.8 | 95.1 | 98.4 |
| 6 | Excitement,Immobility/stupor,Posturing/catalepsy,Grimacing,Verbigeration,Negativism | 96.8 | 95.1 | 98.4 |
| 6 | Excitement,Immobility/stupor,Staring,Grimacing,Mannerisms,Waxy flexibility | 96.8 | 95.1 | 98.4 |
| 6 | Excitement,Staring,Posturing/catalepsy,Echopraxia/echolalia,Verbigeration,Rigidity | 96.8 | 95.1 | 98.4 |
| 6 | Excitement,Staring,Posturing/catalepsy,Echopraxia/echolalia,Verbigeration,Withdrawal | 96.8 | 95.1 | 98.4 |
| 6 | Excitement,Staring,Posturing/catalepsy,Mannerisms,Negativism,Waxy flexibility | 96.8 | 95.1 | 98.4 |
| 6 | Immobility/stupor,Grimacing,Echopraxia/echolalia,Mannerisms,Rigidity,Negativism | 96.8 | 95.1 | 98.4 |
| 6 | Immobility/stupor,Mutism,Staring,Posturing/catalepsy,Stereotypy,Negativism | 96.8 | 95.1 | 98.4 |
| 6 | Immobility/stupor,Posturing/catalepsy,Grimacing,Mannerisms,Rigidity,Negativism | 96.8 | 95.1 | 98.2 |
| 6 | Immobility/stupor,Staring,Echopraxia/echolalia,Mannerisms,Verbigeration,Negativism | 96.8 | 95.1 | 98.4 |
| 6 | Mutism,Staring,Posturing/catalepsy,Echopraxia/echolalia,Stereotypy,Mannerisms | 96.8 | 95.1 | 98.4 |
| 6 | Mutism,Staring,Posturing/catalepsy,Echopraxia/echolalia,Stereotypy,Withdrawal | 96.8 | 95.1 | 98.4 |
| 6 | Mutism,Staring,Stereotypy,Mannerisms,Waxy flexibility,Withdrawal | 96.8 | 95.1 | 98.4 |
| 6 | Mutism,Staring,Verbigeration,Rigidity,Negativism,Waxy flexibility | 96.8 | 95.1 | 98.4 |
| 6 | Excitement,Immobility/stupor,Posturing/catalepsy,Echopraxia/echolalia,Rigidity,Waxy flexibility | 96.7 | 94.8 | 98.2 |
| 6 | Excitement,Immobility/stupor,Posturing/catalepsy,Stereotypy,Verbigeration,Negativism | 96.7 | 94.8 | 98.2 |
| 6 | Excitement,Immobility/stupor,Staring,Grimacing,Stereotypy,Withdrawal | 96.7 | 94.8 | 98.2 |
| 6 | Excitement,Staring,Grimacing,Stereotypy,Mannerisms,Rigidity | 96.7 | 94.8 | 98.2 |
| 6 | Excitement,Staring,Stereotypy,Mannerisms,Waxy flexibility,Withdrawal | 96.7 | 94.8 | 98.2 |
| 6 | Mutism,Staring,Posturing/catalepsy,Echopraxia/echolalia,Mannerisms,Verbigeration | 96.7 | 94.8 | 98.2 |
| 6 | Mutism,Staring,Posturing/catalepsy,Grimacing,Rigidity,Withdrawal | 96.7 | 94.8 | 98.2 |
| 6 | Mutism,Staring,Posturing/catalepsy,Grimacing,Stereotypy,Rigidity | 96.7 | 94.8 | 98.2 |
| 6 | Staring,Posturing/catalepsy,Mannerisms,Rigidity,Waxy flexibility,Withdrawal | 96.7 | 94.8 | 98.2 |
| 6 | Excitement,Immobility/stupor,Echopraxia/echolalia,Negativism,Waxy flexibility,Withdrawal | 96.6 | 94.8 | 98.2 |
| 6 | Excitement,Immobility/stupor,Echopraxia/echolalia,Stereotypy,Verbigeration,Rigidity | 96.6 | 94.8 | 98.2 |
| 6 | Excitement,Immobility/stupor,Echopraxia/echolalia,Verbigeration,Rigidity,Waxy flexibility | 96.6 | 94.8 | 98.2 |
| 6 | Excitement,Immobility/stupor,Grimacing,Echopraxia/echolalia,Verbigeration,Rigidity | 96.6 | 94.8 | 98.2 |
| 6 | Excitement,Immobility/stupor,Mutism,Echopraxia/echolalia,Negativism,Withdrawal | 96.6 | 94.8 | 98.2 |
| 6 | Excitement,Immobility/stupor,Mutism,Posturing/catalepsy,Grimacing,Echopraxia/echolalia | 96.6 | 94.8 | 98.2 |
| 6 | Excitement,Immobility/stupor,Mutism,Posturing/catalepsy,Verbigeration,Withdrawal | 96.6 | 94.8 | 98.2 |
| 6 | Excitement,Immobility/stupor,Mutism,Staring,Stereotypy,Mannerisms | 96.6 | 94.8 | 98.2 |
| 6 | Excitement,Immobility/stupor,Mutism,Stereotypy,Mannerisms,Rigidity | 96.6 | 94.8 | 98.2 |
| 6 | Excitement,Immobility/stupor,Mutism,Stereotypy,Verbigeration,Negativism | 96.6 | 94.8 | 98.2 |
| 6 | Excitement,Immobility/stupor,Mutism,Verbigeration,Negativism,Withdrawal | 96.6 | 94.8 | 98.2 |
| 6 | Excitement,Immobility/stupor,Posturing/catalepsy,Echopraxia/echolalia,Mannerisms,Rigidity | 96.6 | 94.8 | 98.2 |
| 6 | Excitement,Immobility/stupor,Posturing/catalepsy,Echopraxia/echolalia,Stereotypy,Rigidity | 96.6 | 94.8 | 98.2 |
| 6 | Excitement,Immobility/stupor,Posturing/catalepsy,Echopraxia/echolalia,Verbigeration,Negativism | 96.6 | 94.8 | 98.2 |
| 6 | Excitement,Immobility/stupor,Posturing/catalepsy,Grimacing,Echopraxia/echolalia,Withdrawal | 96.6 | 94.8 | 98.2 |
| 6 | Excitement,Immobility/stupor,Posturing/catalepsy,Grimacing,Stereotypy,Rigidity | 96.6 | 94.8 | 98.2 |
| 6 | Excitement,Immobility/stupor,Posturing/catalepsy,Mannerisms,Verbigeration,Negativism | 96.6 | 94.8 | 98.2 |
| 6 | Excitement,Immobility/stupor,Posturing/catalepsy,Stereotypy,Verbigeration,Rigidity | 96.6 | 94.8 | 98.2 |
| 6 | Excitement,Immobility/stupor,Posturing/catalepsy,Verbigeration,Negativism,Waxy flexibility | 96.6 | 94.8 | 98.2 |
| 6 | Excitement,Immobility/stupor,Posturing/catalepsy,Verbigeration,Rigidity,Waxy flexibility | 96.6 | 94.8 | 98.2 |
| 6 | Excitement,Immobility/stupor,Staring,Echopraxia/echolalia,Mannerisms,Verbigeration | 96.6 | 94.8 | 98.2 |
| 6 | Excitement,Immobility/stupor,Staring,Echopraxia/echolalia,Stereotypy,Mannerisms | 96.6 | 94.8 | 98.2 |
| 6 | Excitement,Immobility/stupor,Staring,Echopraxia/echolalia,Stereotypy,Verbigeration | 96.6 | 94.8 | 98.2 |
| 6 | Excitement,Immobility/stupor,Staring,Grimacing,Echopraxia/echolalia,Mannerisms | 96.6 | 94.8 | 98.2 |
| 6 | Excitement,Immobility/stupor,Staring,Grimacing,Echopraxia/echolalia,Stereotypy | 96.6 | 94.8 | 98.2 |
| 6 | Excitement,Immobility/stupor,Staring,Grimacing,Mannerisms,Withdrawal | 96.6 | 94.8 | 98.2 |
| 6 | Excitement,Immobility/stupor,Staring,Grimacing,Stereotypy,Waxy flexibility | 96.6 | 94.8 | 98.2 |
| 6 | Excitement,Immobility/stupor,Staring,Grimacing,Verbigeration,Withdrawal | 96.6 | 94.8 | 98.2 |
| 6 | Excitement,Immobility/stupor,Staring,Stereotypy,Mannerisms,Verbigeration | 96.6 | 94.8 | 98.2 |
| 6 | Excitement,Immobility/stupor,Staring,Stereotypy,Mannerisms,Waxy flexibility | 96.6 | 94.8 | 98.2 |
| 6 | Excitement,Mutism,Staring,Echopraxia/echolalia,Mannerisms,Verbigeration | 96.6 | 94.8 | 98.2 |
| 6 | Excitement,Mutism,Staring,Grimacing,Mannerisms,Verbigeration | 96.6 | 94.8 | 98.2 |
| 6 | Excitement,Mutism,Staring,Grimacing,Stereotypy,Mannerisms | 96.6 | 94.8 | 98.2 |
| 6 | Excitement,Staring,Echopraxia/echolalia,Mannerisms,Negativism,Waxy flexibility | 96.6 | 94.8 | 98.2 |
| 6 | Excitement,Staring,Echopraxia/echolalia,Stereotypy,Mannerisms,Withdrawal | 96.6 | 94.8 | 98.2 |
| 6 | Excitement,Staring,Echopraxia/echolalia,Stereotypy,Negativism,Waxy flexibility | 96.6 | 94.8 | 98.2 |
| 6 | Excitement,Staring,Grimacing,Echopraxia/echolalia,Mannerisms,Rigidity | 96.6 | 94.8 | 98.2 |
| 6 | Excitement,Staring,Grimacing,Mannerisms,Rigidity,Negativism | 96.6 | 94.8 | 98.2 |
| 6 | Excitement,Staring,Grimacing,Mannerisms,Verbigeration,Rigidity | 96.6 | 94.8 | 98.2 |
| 6 | Excitement,Staring,Grimacing,Verbigeration,Rigidity,Negativism | 96.6 | 94.8 | 98.2 |
| 6 | Excitement,Staring,Mannerisms,Verbigeration,Waxy flexibility,Withdrawal | 96.6 | 94.8 | 98.2 |
| 6 | Excitement,Staring,Posturing/catalepsy,Echopraxia/echolalia,Stereotypy,Rigidity | 96.6 | 94.8 | 98.2 |
| 6 | Excitement,Staring,Posturing/catalepsy,Grimacing,Mannerisms,Negativism | 96.6 | 94.8 | 98.2 |
| 6 | Excitement,Staring,Posturing/catalepsy,Grimacing,Verbigeration,Negativism | 96.6 | 94.8 | 98.2 |
| 6 | Excitement,Staring,Posturing/catalepsy,Stereotypy,Mannerisms,Negativism | 96.6 | 94.8 | 98.2 |
| 6 | Excitement,Staring,Posturing/catalepsy,Stereotypy,Mannerisms,Rigidity | 96.6 | 94.8 | 98.2 |
| 6 | Excitement,Staring,Posturing/catalepsy,Stereotypy,Negativism,Waxy flexibility | 96.6 | 94.8 | 98.2 |
| 6 | Excitement,Staring,Posturing/catalepsy,Stereotypy,Verbigeration,Rigidity | 96.6 | 94.8 | 98.2 |
| 6 | Excitement,Staring,Posturing/catalepsy,Stereotypy,Waxy flexibility,Withdrawal | 96.6 | 94.8 | 98.2 |
| 6 | Excitement,Staring,Stereotypy,Verbigeration,Rigidity,Waxy flexibility | 96.6 | 94.8 | 98.2 |
| 6 | Immobility/stupor,Echopraxia/echolalia,Stereotypy,Mannerisms,Rigidity,Negativism | 96.6 | 94.8 | 98.2 |
| 6 | Immobility/stupor,Grimacing,Stereotypy,Mannerisms,Rigidity,Negativism | 96.6 | 94.8 | 98.2 |
| 6 | Immobility/stupor,Mutism,Grimacing,Mannerisms,Rigidity,Negativism | 96.6 | 94.8 | 98.2 |
| 6 | Immobility/stupor,Mutism,Mannerisms,Rigidity,Negativism,Waxy flexibility | 96.6 | 94.8 | 98.2 |
| 6 | Immobility/stupor,Mutism,Staring,Echopraxia/echolalia,Rigidity,Negativism | 96.6 | 94.8 | 98.2 |
| 6 | Immobility/stupor,Mutism,Staring,Grimacing,Mannerisms,Rigidity | 96.6 | 94.8 | 98.2 |
| 6 | Immobility/stupor,Mutism,Staring,Mannerisms,Negativism,Waxy flexibility | 96.6 | 94.8 | 98.2 |
| 6 | Immobility/stupor,Mutism,Staring,Mannerisms,Verbigeration,Negativism | 96.6 | 94.8 | 98.2 |
| 6 | Immobility/stupor,Mutism,Staring,Stereotypy,Negativism,Withdrawal | 96.6 | 94.8 | 98.2 |
| 6 | Immobility/stupor,Mutism,Staring,Stereotypy,Verbigeration,Negativism | 96.6 | 94.8 | 98.2 |
| 6 | Immobility/stupor,Posturing/catalepsy,Stereotypy,Rigidity,Negativism,Withdrawal | 96.6 | 94.8 | 98.2 |
| 6 | Immobility/stupor,Posturing/catalepsy,Stereotypy,Verbigeration,Rigidity,Negativism | 96.6 | 94.8 | 98.2 |
| 6 | Immobility/stupor,Staring,Echopraxia/echolalia,Stereotypy,Rigidity,Withdrawal | 96.6 | 94.8 | 98.2 |
| 6 | Immobility/stupor,Staring,Grimacing,Echopraxia/echolalia,Mannerisms,Rigidity | 96.6 | 94.8 | 98.2 |
| 6 | Immobility/stupor,Staring,Grimacing,Echopraxia/echolalia,Rigidity,Negativism | 96.6 | 94.8 | 98.2 |
| 6 | Immobility/stupor,Staring,Grimacing,Mannerisms,Verbigeration,Negativism | 96.6 | 94.8 | 98.2 |
| 6 | Immobility/stupor,Staring,Grimacing,Stereotypy,Negativism,Withdrawal | 96.6 | 94.8 | 98.2 |
| 6 | Immobility/stupor,Staring,Grimacing,Stereotypy,Rigidity,Withdrawal | 96.6 | 94.8 | 98.2 |
| 6 | Immobility/stupor,Staring,Mannerisms,Verbigeration,Negativism,Waxy flexibility | 96.6 | 94.8 | 98.2 |
| 6 | Immobility/stupor,Staring,Posturing/catalepsy,Grimacing,Mannerisms,Rigidity | 96.6 | 94.8 | 98.2 |
| 6 | Immobility/stupor,Staring,Posturing/catalepsy,Grimacing,Negativism,Withdrawal | 96.6 | 94.8 | 98.2 |
| 6 | Immobility/stupor,Staring,Posturing/catalepsy,Mannerisms,Negativism,Waxy flexibility | 96.6 | 94.8 | 98.2 |
| 6 | Immobility/stupor,Staring,Posturing/catalepsy,Stereotypy,Mannerisms,Rigidity | 96.6 | 94.8 | 98.2 |
| 6 | Immobility/stupor,Staring,Posturing/catalepsy,Stereotypy,Verbigeration,Rigidity | 96.6 | 94.8 | 98.2 |
| 6 | Immobility/stupor,Staring,Posturing/catalepsy,Verbigeration,Rigidity,Negativism | 96.6 | 94.8 | 98.2 |
| 6 | Immobility/stupor,Staring,Stereotypy,Verbigeration,Negativism,Waxy flexibility | 96.6 | 94.8 | 98.2 |
| 6 | Mutism,Staring,Echopraxia/echolalia,Mannerisms,Waxy flexibility,Withdrawal | 96.6 | 94.8 | 98.2 |
| 6 | Mutism,Staring,Grimacing,Echopraxia/echolalia,Mannerisms,Waxy flexibility | 96.6 | 94.8 | 98.2 |
| 6 | Mutism,Staring,Grimacing,Verbigeration,Rigidity,Waxy flexibility | 96.6 | 94.8 | 98.2 |
| 6 | Mutism,Staring,Posturing/catalepsy,Echopraxia/echolalia,Negativism,Waxy flexibility | 96.6 | 94.8 | 98.2 |
| 6 | Mutism,Staring,Posturing/catalepsy,Grimacing,Mannerisms,Verbigeration | 96.6 | 94.8 | 98.2 |
| 6 | Mutism,Staring,Posturing/catalepsy,Grimacing,Stereotypy,Mannerisms | 96.6 | 94.8 | 98.2 |
| 6 | Mutism,Staring,Posturing/catalepsy,Grimacing,Stereotypy,Verbigeration | 96.6 | 94.8 | 98.2 |
| 6 | Mutism,Staring,Posturing/catalepsy,Grimacing,Verbigeration,Withdrawal | 96.6 | 94.8 | 98.2 |
| 6 | Mutism,Staring,Posturing/catalepsy,Mannerisms,Rigidity,Waxy flexibility | 96.6 | 94.8 | 98.2 |
| 6 | Mutism,Staring,Posturing/catalepsy,Stereotypy,Mannerisms,Verbigeration | 96.6 | 94.8 | 98.2 |
| 6 | Mutism,Staring,Posturing/catalepsy,Stereotypy,Verbigeration,Withdrawal | 96.6 | 94.8 | 98.2 |
| 6 | Mutism,Staring,Stereotypy,Mannerisms,Verbigeration,Withdrawal | 96.6 | 94.8 | 98.2 |
| 6 | Mutism,Staring,Verbigeration,Rigidity,Waxy flexibility,Withdrawal | 96.6 | 94.8 | 98.2 |
| 6 | Staring,Echopraxia/echolalia,Mannerisms,Verbigeration,Rigidity,Negativism | 96.6 | 94.8 | 98.2 |
| 6 | Staring,Grimacing,Echopraxia/echolalia,Mannerisms,Rigidity,Negativism | 96.6 | 94.8 | 98.2 |
| 6 | Staring,Mannerisms,Verbigeration,Rigidity,Negativism,Waxy flexibility | 96.6 | 94.8 | 98.2 |
| 6 | Staring,Posturing/catalepsy,Grimacing,Echopraxia/echolalia,Mannerisms,Withdrawal | 96.6 | 94.8 | 98.2 |
| 6 | Staring,Posturing/catalepsy,Grimacing,Mannerisms,Rigidity,Negativism | 96.6 | 94.8 | 98.2 |
| 6 | Staring,Posturing/catalepsy,Grimacing,Stereotypy,Rigidity,Withdrawal | 96.6 | 94.8 | 98.2 |
| 6 | Staring,Posturing/catalepsy,Grimacing,Verbigeration,Negativism,Withdrawal | 96.6 | 94.8 | 98.2 |
| 6 | Staring,Posturing/catalepsy,Rigidity,Negativism,Waxy flexibility,Withdrawal | 96.6 | 94.8 | 98.2 |
| 6 | Staring,Posturing/catalepsy,Stereotypy,Verbigeration,Rigidity,Negativism | 96.6 | 94.8 | 98.2 |
| 6 | Staring,Stereotypy,Verbigeration,Negativism,Waxy flexibility,Withdrawal | 96.6 | 94.8 | 98.2 |
| 6 | Excitement,Immobility/stupor,Echopraxia/echolalia,Stereotypy,Rigidity,Waxy flexibility | 96.4 | 94.6 | 98.0 |
| 6 | Excitement,Immobility/stupor,Grimacing,Echopraxia/echolalia,Mannerisms,Rigidity | 96.4 | 94.6 | 98.0 |
| 6 | Excitement,Immobility/stupor,Grimacing,Echopraxia/echolalia,Negativism,Withdrawal | 96.4 | 94.6 | 98.0 |
| 6 | Excitement,Immobility/stupor,Grimacing,Echopraxia/echolalia,Stereotypy,Rigidity | 96.4 | 94.6 | 98.0 |
| 6 | Excitement,Immobility/stupor,Mutism,Echopraxia/echolalia,Negativism,Waxy flexibility | 96.4 | 94.6 | 98.0 |
| 6 | Excitement,Immobility/stupor,Mutism,Negativism,Waxy flexibility,Withdrawal | 96.4 | 94.6 | 98.0 |
| 6 | Excitement,Immobility/stupor,Mutism,Posturing/catalepsy,Echopraxia/echolalia,Verbigeration | 96.4 | 94.6 | 98.0 |
| 6 | Excitement,Immobility/stupor,Mutism,Posturing/catalepsy,Grimacing,Verbigeration | 96.4 | 94.6 | 98.0 |
| 6 | Excitement,Immobility/stupor,Mutism,Posturing/catalepsy,Mannerisms,Withdrawal | 96.4 | 94.6 | 98.0 |
| 6 | Excitement,Immobility/stupor,Mutism,Posturing/catalepsy,Stereotypy,Withdrawal | 96.4 | 94.6 | 98.0 |
| 6 | Excitement,Immobility/stupor,Mutism,Posturing/catalepsy,Waxy flexibility,Withdrawal | 96.4 | 94.6 | 98.0 |
| 6 | Excitement,Immobility/stupor,Mutism,Staring,Grimacing,Mannerisms | 96.4 | 94.6 | 98.0 |
| 6 | Excitement,Immobility/stupor,Mutism,Stereotypy,Negativism,Waxy flexibility | 96.4 | 94.6 | 98.0 |
| 6 | Excitement,Immobility/stupor,Mutism,Verbigeration,Negativism,Waxy flexibility | 96.4 | 94.6 | 98.0 |
| 6 | Excitement,Immobility/stupor,Posturing/catalepsy,Echopraxia/echolalia,Negativism,Waxy flexibility | 96.4 | 94.6 | 98.0 |
| 6 | Excitement,Immobility/stupor,Posturing/catalepsy,Echopraxia/echolalia,Verbigeration,Withdrawal | 96.4 | 94.6 | 98.0 |
| 6 | Excitement,Immobility/stupor,Posturing/catalepsy,Grimacing,Mannerisms,Negativism | 96.4 | 94.6 | 98.0 |
| 6 | Excitement,Immobility/stupor,Posturing/catalepsy,Grimacing,Negativism,Waxy flexibility | 96.4 | 94.6 | 98.0 |
| 6 | Excitement,Immobility/stupor,Posturing/catalepsy,Grimacing,Rigidity,Waxy flexibility | 96.4 | 94.6 | 98.0 |
| 6 | Excitement,Immobility/stupor,Staring,Grimacing,Echopraxia/echolalia,Verbigeration | 96.4 | 94.6 | 98.0 |
| 6 | Excitement,Immobility/stupor,Staring,Grimacing,Mannerisms,Verbigeration | 96.4 | 94.6 | 98.0 |
| 6 | Excitement,Immobility/stupor,Staring,Grimacing,Stereotypy,Verbigeration | 96.4 | 94.6 | 98.0 |
| 6 | Excitement,Immobility/stupor,Stereotypy,Verbigeration,Rigidity,Waxy flexibility | 96.4 | 94.6 | 98.0 |
| 6 | Excitement,Staring,Echopraxia/echolalia,Stereotypy,Rigidity,Waxy flexibility | 96.4 | 94.6 | 98.0 |
| 6 | Excitement,Staring,Echopraxia/echolalia,Verbigeration,Negativism,Waxy flexibility | 96.4 | 94.6 | 98.0 |
| 6 | Excitement,Staring,Echopraxia/echolalia,Verbigeration,Waxy flexibility,Withdrawal | 96.4 | 94.6 | 98.0 |
| 6 | Excitement,Staring,Grimacing,Echopraxia/echolalia,Verbigeration,Rigidity | 96.4 | 94.6 | 98.0 |
| 6 | Excitement,Staring,Grimacing,Mannerisms,Verbigeration,Withdrawal | 96.4 | 94.6 | 98.0 |
| 6 | Excitement,Staring,Grimacing,Stereotypy,Mannerisms,Withdrawal | 96.4 | 94.6 | 98.0 |
| 6 | Excitement,Staring,Grimacing,Stereotypy,Verbigeration,Withdrawal | 96.4 | 94.6 | 98.0 |
| 6 | Excitement,Staring,Grimacing,Stereotypy,Waxy flexibility,Withdrawal | 96.4 | 94.6 | 98.0 |
| 6 | Excitement,Staring,Grimacing,Verbigeration,Waxy flexibility,Withdrawal | 96.4 | 94.6 | 98.0 |
| 6 | Excitement,Staring,Mannerisms,Verbigeration,Negativism,Waxy flexibility | 96.4 | 94.6 | 98.0 |
| 6 | Excitement,Staring,Mannerisms,Verbigeration,Rigidity,Waxy flexibility | 96.4 | 94.6 | 98.0 |
| 6 | Excitement,Staring,Posturing/catalepsy,Echopraxia/echolalia,Rigidity,Waxy flexibility | 96.4 | 94.6 | 98.0 |
| 6 | Excitement,Staring,Posturing/catalepsy,Grimacing,Rigidity,Waxy flexibility | 96.4 | 94.6 | 98.0 |
| 6 | Excitement,Staring,Posturing/catalepsy,Grimacing,Stereotypy,Negativism | 96.4 | 94.6 | 98.0 |
| 6 | Excitement,Staring,Posturing/catalepsy,Mannerisms,Rigidity,Waxy flexibility | 96.4 | 94.6 | 98.0 |
| 6 | Excitement,Staring,Posturing/catalepsy,Stereotypy,Verbigeration,Withdrawal | 96.4 | 94.6 | 98.0 |
| 6 | Excitement,Staring,Posturing/catalepsy,Verbigeration,Rigidity,Waxy flexibility | 96.4 | 94.6 | 98.0 |
| 6 | Excitement,Staring,Stereotypy,Mannerisms,Rigidity,Waxy flexibility | 96.4 | 94.6 | 98.0 |
| 6 | Excitement,Staring,Stereotypy,Mannerisms,Verbigeration,Rigidity | 96.4 | 94.6 | 98.0 |
| 6 | Excitement,Staring,Stereotypy,Verbigeration,Negativism,Waxy flexibility | 96.4 | 94.6 | 98.0 |
| 6 | Excitement,Staring,Stereotypy,Verbigeration,Waxy flexibility,Withdrawal | 96.4 | 94.6 | 98.0 |
| 6 | Immobility/stupor,Echopraxia/echolalia,Mannerisms,Verbigeration,Rigidity,Negativism | 96.4 | 94.6 | 98.0 |
| 6 | Immobility/stupor,Echopraxia/echolalia,Stereotypy,Mannerisms,Negativism,Withdrawal | 96.4 | 94.6 | 98.0 |
| 6 | Immobility/stupor,Echopraxia/echolalia,Verbigeration,Rigidity,Negativism,Withdrawal | 96.4 | 94.6 | 98.0 |
| 6 | Immobility/stupor,Grimacing,Mannerisms,Rigidity,Negativism,Waxy flexibility | 96.4 | 94.6 | 98.0 |
| 6 | Immobility/stupor,Grimacing,Mannerisms,Verbigeration,Rigidity,Negativism | 96.4 | 94.6 | 98.0 |
| 6 | Immobility/stupor,Grimacing,Stereotypy,Rigidity,Negativism,Withdrawal | 96.4 | 94.6 | 98.0 |
| 6 | Immobility/stupor,Grimacing,Stereotypy,Verbigeration,Rigidity,Negativism | 96.4 | 94.6 | 98.0 |
| 6 | Immobility/stupor,Mutism,Staring,Echopraxia/echolalia,Negativism,Withdrawal | 96.4 | 94.6 | 98.0 |
| 6 | Immobility/stupor,Mutism,Staring,Echopraxia/echolalia,Rigidity,Withdrawal | 96.4 | 94.6 | 98.0 |
| 6 | Immobility/stupor,Mutism,Staring,Grimacing,Mannerisms,Negativism | 96.4 | 94.6 | 98.0 |
| 6 | Immobility/stupor,Mutism,Staring,Grimacing,Verbigeration,Rigidity | 96.4 | 94.6 | 98.0 |
| 6 | Immobility/stupor,Mutism,Staring,Mannerisms,Rigidity,Waxy flexibility | 96.4 | 94.6 | 98.0 |
| 6 | Immobility/stupor,Mutism,Staring,Posturing/catalepsy,Echopraxia/echolalia,Negativism | 96.4 | 94.6 | 98.0 |
| 6 | Immobility/stupor,Mutism,Staring,Posturing/catalepsy,Verbigeration,Negativism | 96.4 | 94.6 | 98.0 |
| 6 | Immobility/stupor,Mutism,Staring,Stereotypy,Rigidity,Withdrawal | 96.4 | 94.6 | 98.0 |
| 6 | Immobility/stupor,Mutism,Staring,Verbigeration,Negativism,Withdrawal | 96.4 | 94.6 | 98.0 |
| 6 | Immobility/stupor,Mutism,Staring,Verbigeration,Rigidity,Withdrawal | 96.4 | 94.6 | 98.0 |
| 6 | Immobility/stupor,Mutism,Stereotypy,Rigidity,Negativism,Withdrawal | 96.4 | 94.6 | 98.0 |
| 6 | Immobility/stupor,Mutism,Verbigeration,Rigidity,Negativism,Withdrawal | 96.4 | 94.6 | 98.0 |
| 6 | Immobility/stupor,Posturing/catalepsy,Mannerisms,Rigidity,Negativism,Waxy flexibility | 96.4 | 94.6 | 98.0 |
| 6 | Immobility/stupor,Posturing/catalepsy,Verbigeration,Rigidity,Negativism,Withdrawal | 96.4 | 94.6 | 98.0 |
| 6 | Immobility/stupor,Staring,Echopraxia/echolalia,Mannerisms,Verbigeration,Rigidity | 96.4 | 94.6 | 98.0 |
| 6 | Immobility/stupor,Staring,Echopraxia/echolalia,Stereotypy,Mannerisms,Rigidity | 96.4 | 94.6 | 98.0 |
| 6 | Immobility/stupor,Staring,Echopraxia/echolalia,Stereotypy,Verbigeration,Negativism | 96.4 | 94.6 | 98.0 |
| 6 | Immobility/stupor,Staring,Echopraxia/echolalia,Verbigeration,Negativism,Withdrawal | 96.4 | 94.6 | 98.0 |
| 6 | Immobility/stupor,Staring,Grimacing,Echopraxia/echolalia,Negativism,Withdrawal | 96.4 | 94.6 | 98.0 |
| 6 | Immobility/stupor,Staring,Grimacing,Echopraxia/echolalia,Stereotypy,Negativism | 96.4 | 94.6 | 98.0 |
| 6 | Immobility/stupor,Staring,Grimacing,Mannerisms,Negativism,Waxy flexibility | 96.4 | 94.6 | 98.0 |
| 6 | Immobility/stupor,Staring,Grimacing,Mannerisms,Rigidity,Waxy flexibility | 96.4 | 94.6 | 98.0 |
| 6 | Immobility/stupor,Staring,Grimacing,Stereotypy,Mannerisms,Withdrawal | 96.4 | 94.6 | 98.0 |
| 6 | Immobility/stupor,Staring,Grimacing,Verbigeration,Negativism,Withdrawal | 96.4 | 94.6 | 98.0 |
| 6 | Immobility/stupor,Staring,Grimacing,Verbigeration,Rigidity,Withdrawal | 96.4 | 94.6 | 98.0 |
| 6 | Immobility/stupor,Staring,Mannerisms,Verbigeration,Rigidity,Waxy flexibility | 96.4 | 94.6 | 98.0 |
| 6 | Immobility/stupor,Staring,Posturing/catalepsy,Echopraxia/echolalia,Mannerisms,Rigidity | 96.4 | 94.6 | 98.0 |
| 6 | Immobility/stupor,Staring,Posturing/catalepsy,Echopraxia/echolalia,Verbigeration,Negativism | 96.4 | 94.6 | 98.0 |
| 6 | Immobility/stupor,Staring,Posturing/catalepsy,Grimacing,Mannerisms,Withdrawal | 96.4 | 94.6 | 98.0 |
| 6 | Immobility/stupor,Staring,Posturing/catalepsy,Grimacing,Stereotypy,Negativism | 96.4 | 94.6 | 98.0 |
| 6 | Immobility/stupor,Staring,Posturing/catalepsy,Grimacing,Verbigeration,Negativism | 96.4 | 94.6 | 98.0 |
| 6 | Immobility/stupor,Staring,Posturing/catalepsy,Stereotypy,Mannerisms,Withdrawal | 96.4 | 94.6 | 98.0 |
| 6 | Immobility/stupor,Staring,Posturing/catalepsy,Stereotypy,Rigidity,Withdrawal | 96.4 | 94.6 | 98.0 |
| 6 | Immobility/stupor,Staring,Stereotypy,Mannerisms,Rigidity,Waxy flexibility | 96.4 | 94.6 | 98.0 |
| 6 | Immobility/stupor,Staring,Stereotypy,Negativism,Waxy flexibility,Withdrawal | 96.4 | 94.6 | 98.0 |
| 6 | Immobility/stupor,Staring,Stereotypy,Verbigeration,Rigidity,Waxy flexibility | 96.4 | 94.6 | 98.0 |
| 6 | Mutism,Staring,Echopraxia/echolalia,Rigidity,Waxy flexibility,Withdrawal | 96.4 | 94.6 | 98.0 |
| 6 | Mutism,Staring,Echopraxia/echolalia,Stereotypy,Mannerisms,Waxy flexibility | 96.4 | 94.6 | 98.0 |
| 6 | Mutism,Staring,Grimacing,Echopraxia/echolalia,Mannerisms,Verbigeration | 96.4 | 94.6 | 98.0 |
| 6 | Mutism,Staring,Grimacing,Echopraxia/echolalia,Stereotypy,Verbigeration | 96.4 | 94.6 | 98.0 |
| 6 | Mutism,Staring,Grimacing,Stereotypy,Mannerisms,Verbigeration | 96.4 | 94.6 | 98.0 |
| 6 | Mutism,Staring,Grimacing,Stereotypy,Mannerisms,Waxy flexibility | 96.4 | 94.6 | 98.0 |
| 6 | Mutism,Staring,Grimacing,Stereotypy,Negativism,Waxy flexibility | 96.4 | 94.6 | 98.0 |
| 6 | Mutism,Staring,Grimacing,Stereotypy,Rigidity,Waxy flexibility | 96.4 | 94.6 | 98.0 |
| 6 | Mutism,Staring,Grimacing,Stereotypy,Verbigeration,Waxy flexibility | 96.4 | 94.6 | 98.0 |
| 6 | Mutism,Staring,Grimacing,Verbigeration,Negativism,Waxy flexibility | 96.4 | 94.6 | 98.0 |
| 6 | Mutism,Staring,Posturing/catalepsy,Echopraxia/echolalia,Stereotypy,Verbigeration | 96.4 | 94.6 | 98.0 |
| 6 | Mutism,Staring,Posturing/catalepsy,Echopraxia/echolalia,Verbigeration,Withdrawal | 96.4 | 94.6 | 98.0 |
| 6 | Mutism,Staring,Posturing/catalepsy,Grimacing,Echopraxia/echolalia,Rigidity | 96.4 | 94.6 | 98.0 |
| 6 | Mutism,Staring,Posturing/catalepsy,Grimacing,Echopraxia/echolalia,Stereotypy | 96.4 | 94.6 | 98.0 |
| 6 | Mutism,Staring,Posturing/catalepsy,Grimacing,Echopraxia/echolalia,Verbigeration | 96.4 | 94.6 | 98.0 |
| 6 | Mutism,Staring,Posturing/catalepsy,Verbigeration,Negativism,Waxy flexibility | 96.4 | 94.6 | 98.0 |
| 6 | Staring,Echopraxia/echolalia,Mannerisms,Rigidity,Waxy flexibility,Withdrawal | 96.4 | 94.6 | 98.0 |
| 6 | Staring,Echopraxia/echolalia,Stereotypy,Negativism,Waxy flexibility,Withdrawal | 96.4 | 94.6 | 98.0 |
| 6 | Staring,Echopraxia/echolalia,Stereotypy,Verbigeration,Negativism,Withdrawal | 96.4 | 94.6 | 98.0 |
| 6 | Staring,Grimacing,Mannerisms,Rigidity,Negativism,Waxy flexibility | 96.4 | 94.6 | 98.0 |
| 6 | Staring,Grimacing,Rigidity,Negativism,Waxy flexibility,Withdrawal | 96.4 | 94.6 | 98.0 |
| 6 | Staring,Grimacing,Stereotypy,Mannerisms,Verbigeration,Withdrawal | 96.4 | 94.6 | 98.0 |
| 6 | Staring,Grimacing,Stereotypy,Mannerisms,Waxy flexibility,Withdrawal | 96.4 | 94.6 | 98.0 |
| 6 | Staring,Grimacing,Stereotypy,Rigidity,Waxy flexibility,Withdrawal | 96.4 | 94.6 | 98.0 |
| 6 | Staring,Grimacing,Stereotypy,Verbigeration,Rigidity,Negativism | 96.4 | 94.6 | 98.0 |
| 6 | Staring,Mannerisms,Verbigeration,Rigidity,Waxy flexibility,Withdrawal | 96.4 | 94.6 | 98.0 |
| 6 | Staring,Posturing/catalepsy,Echopraxia/echolalia,Negativism,Waxy flexibility,Withdrawal | 96.4 | 94.6 | 98.0 |
| 6 | Staring,Posturing/catalepsy,Echopraxia/echolalia,Stereotypy,Mannerisms,Negativism | 96.4 | 94.6 | 98.0 |
| 6 | Staring,Posturing/catalepsy,Echopraxia/echolalia,Stereotypy,Rigidity,Withdrawal | 96.4 | 94.6 | 98.0 |
| 6 | Staring,Posturing/catalepsy,Echopraxia/echolalia,Verbigeration,Negativism,Withdrawal | 96.4 | 94.6 | 98.0 |
| 6 | Staring,Posturing/catalepsy,Grimacing,Mannerisms,Verbigeration,Withdrawal | 96.4 | 94.6 | 98.0 |
| 6 | Staring,Posturing/catalepsy,Mannerisms,Rigidity,Negativism,Waxy flexibility | 96.4 | 94.6 | 98.0 |
| 6 | Staring,Posturing/catalepsy,Stereotypy,Negativism,Waxy flexibility,Withdrawal | 96.4 | 94.6 | 98.0 |
| 6 | Staring,Posturing/catalepsy,Stereotypy,Verbigeration,Rigidity,Withdrawal | 96.4 | 94.6 | 98.0 |
| 6 | Staring,Stereotypy,Verbigeration,Rigidity,Negativism,Waxy flexibility | 96.4 | 94.6 | 98.0 |
| 6 | Staring,Stereotypy,Verbigeration,Rigidity,Waxy flexibility,Withdrawal | 96.4 | 94.6 | 98.0 |
| 6 | Excitement,Immobility/stupor,Echopraxia/echolalia,Mannerisms,Negativism,Withdrawal | 96.2 | 94.4 | 97.8 |
| 6 | Excitement,Immobility/stupor,Echopraxia/echolalia,Mannerisms,Verbigeration,Rigidity | 96.2 | 94.4 | 97.8 |
| 6 | Excitement,Immobility/stupor,Grimacing,Echopraxia/echolalia,Negativism,Waxy flexibility | 96.2 | 94.4 | 97.8 |
| 6 | Excitement,Immobility/stupor,Grimacing,Mannerisms,Rigidity,Waxy flexibility | 96.2 | 94.4 | 97.8 |
| 6 | Excitement,Immobility/stupor,Grimacing,Mannerisms,Verbigeration,Rigidity | 96.2 | 94.4 | 97.8 |
| 6 | Excitement,Immobility/stupor,Grimacing,Stereotypy,Rigidity,Waxy flexibility | 96.2 | 94.4 | 98.0 |
| 6 | Excitement,Immobility/stupor,Grimacing,Stereotypy,Verbigeration,Rigidity | 96.2 | 94.4 | 98.0 |
| 6 | Excitement,Immobility/stupor,Grimacing,Verbigeration,Rigidity,Waxy flexibility | 96.2 | 94.4 | 98.0 |
| 6 | Excitement,Immobility/stupor,Mannerisms,Verbigeration,Negativism,Withdrawal | 96.2 | 94.4 | 97.8 |
| 6 | Excitement,Immobility/stupor,Mutism,Echopraxia/echolalia,Stereotypy,Negativism | 96.2 | 94.4 | 97.8 |
| 6 | Excitement,Immobility/stupor,Mutism,Echopraxia/echolalia,Stereotypy,Withdrawal | 96.2 | 94.4 | 97.8 |
| 6 | Excitement,Immobility/stupor,Mutism,Echopraxia/echolalia,Verbigeration,Negativism | 96.2 | 94.4 | 97.8 |
| 6 | Excitement,Immobility/stupor,Mutism,Echopraxia/echolalia,Waxy flexibility,Withdrawal | 96.2 | 94.4 | 97.8 |
| 6 | Excitement,Immobility/stupor,Mutism,Grimacing,Negativism,Waxy flexibility | 96.2 | 94.4 | 97.8 |
| 6 | Excitement,Immobility/stupor,Mutism,Posturing/catalepsy,Echopraxia/echolalia,Waxy flexibility | 96.2 | 94.4 | 97.8 |
| 6 | Excitement,Immobility/stupor,Posturing/catalepsy,Echopraxia/echolalia,Mannerisms,Negativism | 96.2 | 94.4 | 97.8 |
| 6 | Excitement,Immobility/stupor,Posturing/catalepsy,Echopraxia/echolalia,Mannerisms,Withdrawal | 96.2 | 94.4 | 98.0 |
| 6 | Excitement,Immobility/stupor,Posturing/catalepsy,Echopraxia/echolalia,Stereotypy,Negativism | 96.2 | 94.4 | 97.8 |
| 6 | Excitement,Immobility/stupor,Posturing/catalepsy,Grimacing,Stereotypy,Negativism | 96.2 | 94.4 | 97.8 |
| 6 | Excitement,Immobility/stupor,Posturing/catalepsy,Mannerisms,Verbigeration,Withdrawal | 96.2 | 94.4 | 97.8 |
| 6 | Excitement,Immobility/stupor,Stereotypy,Negativism,Waxy flexibility,Withdrawal | 96.2 | 94.4 | 97.8 |
| 6 | Excitement,Staring,Echopraxia/echolalia,Mannerisms,Rigidity,Waxy flexibility | 96.2 | 94.4 | 98.0 |
| 6 | Excitement,Staring,Echopraxia/echolalia,Stereotypy,Mannerisms,Negativism | 96.2 | 94.4 | 97.8 |
| 6 | Excitement,Staring,Echopraxia/echolalia,Stereotypy,Mannerisms,Rigidity | 96.2 | 94.4 | 97.8 |
| 6 | Excitement,Staring,Echopraxia/echolalia,Stereotypy,Verbigeration,Rigidity | 96.2 | 94.4 | 97.8 |
| 6 | Excitement,Staring,Echopraxia/echolalia,Verbigeration,Rigidity,Waxy flexibility | 96.2 | 94.2 | 97.8 |
| 6 | Excitement,Staring,Grimacing,Echopraxia/echolalia,Negativism,Waxy flexibility | 96.2 | 94.4 | 98.0 |
| 6 | Excitement,Staring,Grimacing,Echopraxia/echolalia,Verbigeration,Withdrawal | 96.2 | 94.4 | 97.8 |
| 6 | Excitement,Staring,Posturing/catalepsy,Grimacing,Negativism,Waxy flexibility | 96.2 | 94.4 | 98.0 |
| 6 | Excitement,Staring,Posturing/catalepsy,Stereotypy,Rigidity,Waxy flexibility | 96.2 | 94.4 | 98.0 |
| 6 | Excitement,Staring,Stereotypy,Mannerisms,Negativism,Waxy flexibility | 96.2 | 94.4 | 97.8 |
| 6 | Excitement,Staring,Stereotypy,Mannerisms,Verbigeration,Withdrawal | 96.2 | 94.4 | 97.8 |
| 6 | Immobility/stupor,Echopraxia/echolalia,Mannerisms,Rigidity,Negativism,Waxy flexibility | 96.2 | 94.4 | 97.8 |
| 6 | Immobility/stupor,Mannerisms,Verbigeration,Rigidity,Negativism,Waxy flexibility | 96.2 | 94.4 | 97.8 |
| 6 | Immobility/stupor,Mutism,Posturing/catalepsy,Mannerisms,Negativism,Withdrawal | 96.2 | 94.4 | 97.8 |
| 6 | Immobility/stupor,Mutism,Staring,Echopraxia/echolalia,Mannerisms,Withdrawal | 96.2 | 94.4 | 97.8 |
| 6 | Immobility/stupor,Mutism,Staring,Echopraxia/echolalia,Stereotypy,Negativism | 96.2 | 94.4 | 97.8 |
| 6 | Immobility/stupor,Mutism,Staring,Echopraxia/echolalia,Verbigeration,Rigidity | 96.2 | 94.4 | 98.0 |
| 6 | Immobility/stupor,Mutism,Staring,Grimacing,Mannerisms,Withdrawal | 96.2 | 94.4 | 97.8 |
| 6 | Immobility/stupor,Mutism,Staring,Posturing/catalepsy,Verbigeration,Rigidity | 96.2 | 94.4 | 97.8 |
| 6 | Immobility/stupor,Mutism,Staring,Stereotypy,Mannerisms,Withdrawal | 96.2 | 94.4 | 98.0 |
| 6 | Immobility/stupor,Posturing/catalepsy,Echopraxia/echolalia,Mannerisms,Negativism,Withdrawal | 96.2 | 94.4 | 97.8 |
| 6 | Immobility/stupor,Posturing/catalepsy,Mannerisms,Verbigeration,Negativism,Withdrawal | 96.2 | 94.4 | 97.8 |
| 6 | Immobility/stupor,Posturing/catalepsy,Stereotypy,Mannerisms,Negativism,Withdrawal | 96.2 | 94.4 | 97.8 |
| 6 | Immobility/stupor,Staring,Echopraxia/echolalia,Negativism,Waxy flexibility,Withdrawal | 96.2 | 94.4 | 97.8 |
| 6 | Immobility/stupor,Staring,Echopraxia/echolalia,Rigidity,Negativism,Waxy flexibility | 96.2 | 94.4 | 98.0 |
| 6 | Immobility/stupor,Staring,Echopraxia/echolalia,Stereotypy,Mannerisms,Withdrawal | 96.2 | 94.4 | 97.8 |
| 6 | Immobility/stupor,Staring,Echopraxia/echolalia,Stereotypy,Negativism,Waxy flexibility | 96.2 | 94.4 | 97.8 |
| 6 | Immobility/stupor,Staring,Grimacing,Echopraxia/echolalia,Mannerisms,Withdrawal | 96.2 | 94.4 | 97.8 |
| 6 | Immobility/stupor,Staring,Grimacing,Echopraxia/echolalia,Rigidity,Withdrawal | 96.2 | 94.4 | 97.8 |
| 6 | Immobility/stupor,Staring,Grimacing,Echopraxia/echolalia,Verbigeration,Negativism | 96.2 | 94.4 | 98.0 |
| 6 | Immobility/stupor,Staring,Posturing/catalepsy,Echopraxia/echolalia,Mannerisms,Withdrawal | 96.2 | 94.4 | 97.8 |
| 6 | Immobility/stupor,Staring,Posturing/catalepsy,Echopraxia/echolalia,Rigidity,Withdrawal | 96.2 | 94.4 | 98.0 |
| 6 | Immobility/stupor,Staring,Posturing/catalepsy,Grimacing,Echopraxia/echolalia,Negativism | 96.2 | 94.4 | 97.8 |
| 6 | Immobility/stupor,Staring,Posturing/catalepsy,Negativism,Waxy flexibility,Withdrawal | 96.2 | 94.4 | 97.8 |
| 6 | Immobility/stupor,Staring,Posturing/catalepsy,Stereotypy,Negativism,Waxy flexibility | 96.2 | 94.4 | 98.0 |
| 6 | Immobility/stupor,Staring,Stereotypy,Rigidity,Waxy flexibility,Withdrawal | 96.2 | 94.2 | 98.0 |
| 6 | Immobility/stupor,Staring,Verbigeration,Negativism,Waxy flexibility,Withdrawal | 96.2 | 94.4 | 97.8 |
| 6 | Immobility/stupor,Staring,Verbigeration,Rigidity,Negativism,Waxy flexibility | 96.2 | 94.4 | 97.8 |
| 6 | Immobility/stupor,Stereotypy,Mannerisms,Verbigeration,Negativism,Withdrawal | 96.2 | 94.4 | 98.0 |
| 6 | Immobility/stupor,Stereotypy,Rigidity,Negativism,Waxy flexibility,Withdrawal | 96.2 | 94.2 | 97.8 |
| 6 | Immobility/stupor,Stereotypy,Verbigeration,Rigidity,Negativism,Waxy flexibility | 96.2 | 94.4 | 97.8 |
| 6 | Mutism,Staring,Echopraxia/echolalia,Mannerisms,Verbigeration,Withdrawal | 96.2 | 94.4 | 97.8 |
| 6 | Mutism,Staring,Echopraxia/echolalia,Stereotypy,Rigidity,Waxy flexibility | 96.2 | 94.4 | 97.8 |
| 6 | Mutism,Staring,Echopraxia/echolalia,Stereotypy,Waxy flexibility,Withdrawal | 96.2 | 94.4 | 97.8 |
| 6 | Mutism,Staring,Echopraxia/echolalia,Verbigeration,Rigidity,Waxy flexibility | 96.2 | 94.4 | 98.0 |
| 6 | Mutism,Staring,Grimacing,Echopraxia/echolalia,Negativism,Waxy flexibility | 96.2 | 94.4 | 97.8 |
| 6 | Mutism,Staring,Grimacing,Echopraxia/echolalia,Stereotypy,Waxy flexibility | 96.2 | 94.2 | 97.8 |
| 6 | Mutism,Staring,Grimacing,Echopraxia/echolalia,Verbigeration,Waxy flexibility | 96.2 | 94.4 | 98.0 |
| 6 | Mutism,Staring,Grimacing,Echopraxia/echolalia,Verbigeration,Withdrawal | 96.2 | 94.4 | 97.8 |
| 6 | Mutism,Staring,Grimacing,Mannerisms,Verbigeration,Waxy flexibility | 96.2 | 94.4 | 97.8 |
| 6 | Mutism,Staring,Grimacing,Negativism,Waxy flexibility,Withdrawal | 96.2 | 94.4 | 97.8 |
| 6 | Mutism,Staring,Grimacing,Rigidity,Waxy flexibility,Withdrawal | 96.2 | 94.4 | 97.8 |
| 6 | Mutism,Staring,Grimacing,Stereotypy,Waxy flexibility,Withdrawal | 96.2 | 94.4 | 98.0 |
| 6 | Mutism,Staring,Mannerisms,Verbigeration,Waxy flexibility,Withdrawal | 96.2 | 94.4 | 97.8 |
| 6 | Mutism,Staring,Posturing/catalepsy,Echopraxia/echolalia,Mannerisms,Waxy flexibility | 96.2 | 94.4 | 97.8 |
| 6 | Mutism,Staring,Posturing/catalepsy,Echopraxia/echolalia,Waxy flexibility,Withdrawal | 96.2 | 94.4 | 98.0 |
| 6 | Mutism,Staring,Posturing/catalepsy,Grimacing,Rigidity,Negativism | 96.2 | 94.4 | 98.0 |
| 6 | Mutism,Staring,Posturing/catalepsy,Rigidity,Waxy flexibility,Withdrawal | 96.2 | 94.4 | 98.0 |
| 6 | Mutism,Staring,Posturing/catalepsy,Stereotypy,Verbigeration,Waxy flexibility | 96.2 | 94.4 | 97.8 |
| 6 | Mutism,Staring,Posturing/catalepsy,Stereotypy,Waxy flexibility,Withdrawal | 96.2 | 94.4 | 97.8 |
| 6 | Mutism,Staring,Posturing/catalepsy,Verbigeration,Rigidity,Waxy flexibility | 96.2 | 94.2 | 97.8 |
| 6 | Mutism,Staring,Stereotypy,Verbigeration,Waxy flexibility,Withdrawal | 96.2 | 94.4 | 97.8 |
| 6 | Staring,Echopraxia/echolalia,Mannerisms,Verbigeration,Rigidity,Withdrawal | 96.2 | 94.4 | 98.0 |
| 6 | Staring,Echopraxia/echolalia,Stereotypy,Mannerisms,Negativism,Waxy flexibility | 96.2 | 94.4 | 97.8 |
| 6 | Staring,Echopraxia/echolalia,Stereotypy,Verbigeration,Rigidity,Negativism | 96.2 | 94.4 | 97.8 |
| 6 | Staring,Echopraxia/echolalia,Stereotypy,Verbigeration,Rigidity,Withdrawal | 96.2 | 94.4 | 98.0 |
| 6 | Staring,Grimacing,Echopraxia/echolalia,Mannerisms,Waxy flexibility,Withdrawal | 96.2 | 94.4 | 97.8 |
| 6 | Staring,Grimacing,Mannerisms,Verbigeration,Rigidity,Negativism | 96.2 | 94.4 | 97.8 |
| 6 | Staring,Grimacing,Stereotypy,Mannerisms,Rigidity,Waxy flexibility | 96.2 | 94.4 | 97.8 |
| 6 | Staring,Grimacing,Stereotypy,Mannerisms,Verbigeration,Rigidity | 96.2 | 94.4 | 97.8 |
| 6 | Staring,Posturing/catalepsy,Echopraxia/echolalia,Stereotypy,Rigidity,Negativism | 96.2 | 94.4 | 97.8 |
| 6 | Staring,Posturing/catalepsy,Grimacing,Echopraxia/echolalia,Mannerisms,Negativism | 96.2 | 94.4 | 97.8 |
| 6 | Staring,Posturing/catalepsy,Grimacing,Echopraxia/echolalia,Rigidity,Withdrawal | 96.2 | 94.4 | 98.0 |
| 6 | Staring,Posturing/catalepsy,Grimacing,Stereotypy,Mannerisms,Negativism | 96.2 | 94.4 | 97.8 |
| 6 | Staring,Posturing/catalepsy,Grimacing,Stereotypy,Mannerisms,Rigidity | 96.2 | 94.4 | 97.8 |
| 6 | Staring,Posturing/catalepsy,Grimacing,Verbigeration,Rigidity,Withdrawal | 96.2 | 94.2 | 97.8 |
| 6 | Staring,Posturing/catalepsy,Stereotypy,Mannerisms,Verbigeration,Negativism | 96.2 | 94.4 | 97.8 |
| 6 | Staring,Posturing/catalepsy,Verbigeration,Negativism,Waxy flexibility,Withdrawal | 96.2 | 94.2 | 97.8 |
| 6 | Excitement,Immobility/stupor,Echopraxia/echolalia,Mannerisms,Rigidity,Waxy flexibility | 96.0 | 93.9 | 97.8 |
| 6 | Excitement,Immobility/stupor,Echopraxia/echolalia,Stereotypy,Mannerisms,Rigidity | 96.0 | 94.2 | 97.8 |
| 6 | Excitement,Immobility/stupor,Echopraxia/echolalia,Stereotypy,Negativism,Waxy flexibility | 96.0 | 93.9 | 97.8 |
| 6 | Excitement,Immobility/stupor,Grimacing,Stereotypy,Negativism,Withdrawal | 96.0 | 94.2 | 97.5 |
| 6 | Excitement,Immobility/stupor,Grimacing,Verbigeration,Negativism,Withdrawal | 96.0 | 93.9 | 97.8 |
| 6 | Excitement,Immobility/stupor,Mannerisms,Verbigeration,Rigidity,Waxy flexibility | 96.0 | 93.9 | 97.8 |
| 6 | Excitement,Immobility/stupor,Mutism,Echopraxia/echolalia,Verbigeration,Withdrawal | 96.0 | 94.2 | 97.8 |
| 6 | Excitement,Immobility/stupor,Mutism,Grimacing,Negativism,Withdrawal | 96.0 | 94.2 | 97.8 |
| 6 | Excitement,Immobility/stupor,Mutism,Grimacing,Verbigeration,Negativism | 96.0 | 94.2 | 97.8 |
| 6 | Excitement,Immobility/stupor,Mutism,Posturing/catalepsy,Echopraxia/echolalia,Stereotypy | 96.0 | 93.9 | 97.8 |
| 6 | Excitement,Immobility/stupor,Mutism,Posturing/catalepsy,Grimacing,Mannerisms | 96.0 | 94.2 | 97.8 |
| 6 | Excitement,Immobility/stupor,Mutism,Posturing/catalepsy,Grimacing,Stereotypy | 96.0 | 93.9 | 97.8 |
| 6 | Excitement,Immobility/stupor,Mutism,Posturing/catalepsy,Grimacing,Waxy flexibility | 96.0 | 94.2 | 97.8 |
| 6 | Excitement,Immobility/stupor,Mutism,Posturing/catalepsy,Verbigeration,Waxy flexibility | 96.0 | 93.9 | 97.8 |
| 6 | Excitement,Immobility/stupor,Mutism,Stereotypy,Verbigeration,Withdrawal | 96.0 | 93.9 | 97.8 |
| 6 | Excitement,Immobility/stupor,Mutism,Stereotypy,Waxy flexibility,Withdrawal | 96.0 | 94.2 | 97.8 |
| 6 | Excitement,Immobility/stupor,Posturing/catalepsy,Echopraxia/echolalia,Stereotypy,Withdrawal | 96.0 | 93.9 | 97.8 |
| 6 | Excitement,Immobility/stupor,Posturing/catalepsy,Echopraxia/echolalia,Waxy flexibility,Withdrawal | 96.0 | 93.9 | 97.8 |
| 6 | Excitement,Immobility/stupor,Posturing/catalepsy,Grimacing,Verbigeration,Withdrawal | 96.0 | 93.9 | 97.8 |
| 6 | Excitement,Immobility/stupor,Posturing/catalepsy,Mannerisms,Negativism,Waxy flexibility | 96.0 | 93.9 | 97.8 |
| 6 | Excitement,Immobility/stupor,Posturing/catalepsy,Mannerisms,Rigidity,Waxy flexibility | 96.0 | 94.2 | 97.8 |
| 6 | Excitement,Immobility/stupor,Posturing/catalepsy,Stereotypy,Mannerisms,Rigidity | 96.0 | 94.2 | 97.8 |
| 6 | Excitement,Immobility/stupor,Posturing/catalepsy,Stereotypy,Negativism,Waxy flexibility | 96.0 | 93.9 | 97.8 |
| 6 | Excitement,Immobility/stupor,Posturing/catalepsy,Stereotypy,Rigidity,Waxy flexibility | 96.0 | 93.9 | 97.8 |
| 6 | Excitement,Immobility/stupor,Staring,Grimacing,Stereotypy,Mannerisms | 96.0 | 93.9 | 97.8 |
| 6 | Excitement,Immobility/stupor,Stereotypy,Mannerisms,Negativism,Withdrawal | 96.0 | 93.9 | 97.8 |
| 6 | Excitement,Immobility/stupor,Stereotypy,Mannerisms,Verbigeration,Rigidity | 96.0 | 94.2 | 97.8 |
| 6 | Excitement,Immobility/stupor,Stereotypy,Verbigeration,Negativism,Waxy flexibility | 96.0 | 93.9 | 97.8 |
| 6 | Excitement,Immobility/stupor,Verbigeration,Negativism,Waxy flexibility,Withdrawal | 96.0 | 93.9 | 97.8 |
| 6 | Excitement,Staring,Echopraxia/echolalia,Stereotypy,Verbigeration,Negativism | 96.0 | 93.9 | 97.8 |
| 6 | Excitement,Staring,Echopraxia/echolalia,Stereotypy,Verbigeration,Withdrawal | 96.0 | 93.9 | 97.8 |
| 6 | Excitement,Staring,Grimacing,Echopraxia/echolalia,Stereotypy,Negativism | 96.0 | 93.9 | 97.8 |
| 6 | Immobility/stupor,Grimacing,Echopraxia/echolalia,Rigidity,Negativism,Withdrawal | 96.0 | 93.9 | 97.8 |
| 6 | Immobility/stupor,Grimacing,Verbigeration,Rigidity,Negativism,Withdrawal | 96.0 | 93.9 | 97.8 |
| 6 | Immobility/stupor,Mutism,Echopraxia/echolalia,Verbigeration,Rigidity,Negativism | 96.0 | 93.9 | 97.8 |
| 6 | Immobility/stupor,Mutism,Grimacing,Mannerisms,Rigidity,Withdrawal | 96.0 | 93.9 | 97.8 |
| 6 | Immobility/stupor,Mutism,Grimacing,Verbigeration,Rigidity,Negativism | 96.0 | 93.9 | 97.8 |
| 6 | Immobility/stupor,Mutism,Posturing/catalepsy,Stereotypy,Rigidity,Negativism | 96.0 | 93.9 | 97.8 |
| 6 | Immobility/stupor,Mutism,Posturing/catalepsy,Verbigeration,Rigidity,Negativism | 96.0 | 93.9 | 97.8 |
| 6 | Immobility/stupor,Mutism,Staring,Echopraxia/echolalia,Stereotypy,Rigidity | 96.0 | 93.9 | 97.8 |
| 6 | Immobility/stupor,Mutism,Staring,Echopraxia/echolalia,Verbigeration,Negativism | 96.0 | 93.9 | 97.8 |
| 6 | Immobility/stupor,Mutism,Staring,Grimacing,Stereotypy,Negativism | 96.0 | 94.2 | 97.8 |
| 6 | Immobility/stupor,Mutism,Staring,Grimacing,Stereotypy,Rigidity | 96.0 | 94.2 | 97.8 |
| 6 | Immobility/stupor,Mutism,Staring,Grimacing,Verbigeration,Negativism | 96.0 | 94.2 | 97.8 |
| 6 | Immobility/stupor,Mutism,Staring,Mannerisms,Waxy flexibility,Withdrawal | 96.0 | 93.9 | 97.8 |
| 6 | Immobility/stupor,Mutism,Staring,Posturing/catalepsy,Echopraxia/echolalia,Mannerisms | 96.0 | 93.9 | 97.8 |
| 6 | Immobility/stupor,Mutism,Staring,Posturing/catalepsy,Echopraxia/echolalia,Rigidity | 96.0 | 94.2 | 97.8 |
| 6 | Immobility/stupor,Mutism,Staring,Posturing/catalepsy,Echopraxia/echolalia,Withdrawal | 96.0 | 94.2 | 97.8 |
| 6 | Immobility/stupor,Mutism,Staring,Posturing/catalepsy,Mannerisms,Verbigeration | 96.0 | 94.2 | 97.8 |
| 6 | Immobility/stupor,Mutism,Staring,Posturing/catalepsy,Rigidity,Negativism | 96.0 | 93.9 | 97.8 |
| 6 | Immobility/stupor,Mutism,Staring,Posturing/catalepsy,Rigidity,Withdrawal | 96.0 | 94.2 | 97.8 |
| 6 | Immobility/stupor,Mutism,Staring,Posturing/catalepsy,Stereotypy,Rigidity | 96.0 | 93.9 | 97.8 |
| 6 | Immobility/stupor,Mutism,Staring,Stereotypy,Negativism,Waxy flexibility | 96.0 | 93.9 | 97.8 |
| 6 | Immobility/stupor,Posturing/catalepsy,Echopraxia/echolalia,Rigidity,Negativism,Withdrawal | 96.0 | 93.9 | 97.8 |
| 6 | Immobility/stupor,Posturing/catalepsy,Grimacing,Mannerisms,Negativism,Withdrawal | 96.0 | 93.9 | 97.8 |
| 6 | Immobility/stupor,Posturing/catalepsy,Grimacing,Rigidity,Negativism,Withdrawal | 96.0 | 93.9 | 97.8 |
| 6 | Immobility/stupor,Posturing/catalepsy,Grimacing,Verbigeration,Rigidity,Negativism | 96.0 | 93.9 | 97.8 |
| 6 | Immobility/stupor,Posturing/catalepsy,Stereotypy,Verbigeration,Negativism,Withdrawal | 96.0 | 94.2 | 97.8 |
| 6 | Immobility/stupor,Staring,Echopraxia/echolalia,Mannerisms,Rigidity,Waxy flexibility | 96.0 | 93.9 | 97.8 |
| 6 | Immobility/stupor,Staring,Echopraxia/echolalia,Stereotypy,Verbigeration,Rigidity | 96.0 | 93.9 | 97.8 |
| 6 | Immobility/stupor,Staring,Echopraxia/echolalia,Verbigeration,Rigidity,Withdrawal | 96.0 | 93.9 | 97.8 |
| 6 | Immobility/stupor,Staring,Grimacing,Echopraxia/echolalia,Stereotypy,Mannerisms | 96.0 | 93.9 | 97.8 |
| 6 | Immobility/stupor,Staring,Grimacing,Echopraxia/echolalia,Stereotypy,Rigidity | 96.0 | 94.2 | 97.8 |
| 6 | Immobility/stupor,Staring,Grimacing,Echopraxia/echolalia,Verbigeration,Rigidity | 96.0 | 93.9 | 97.8 |
| 6 | Immobility/stupor,Staring,Posturing/catalepsy,Grimacing,Rigidity,Negativism | 96.0 | 93.9 | 97.8 |
| 6 | Immobility/stupor,Staring,Posturing/catalepsy,Grimacing,Stereotypy,Rigidity | 96.0 | 94.2 | 97.8 |
| 6 | Immobility/stupor,Staring,Posturing/catalepsy,Grimacing,Verbigeration,Rigidity | 96.0 | 94.2 | 97.8 |
| 6 | Immobility/stupor,Staring,Posturing/catalepsy,Mannerisms,Verbigeration,Withdrawal | 96.0 | 93.9 | 97.8 |
| 6 | Mutism,Staring,Echopraxia/echolalia,Stereotypy,Mannerisms,Verbigeration | 96.0 | 94.2 | 97.8 |
| 6 | Mutism,Staring,Echopraxia/echolalia,Stereotypy,Verbigeration,Withdrawal | 96.0 | 93.9 | 97.8 |
| 6 | Mutism,Staring,Grimacing,Echopraxia/echolalia,Rigidity,Waxy flexibility | 96.0 | 94.2 | 97.8 |
| 6 | Mutism,Staring,Grimacing,Echopraxia/echolalia,Waxy flexibility,Withdrawal | 96.0 | 93.9 | 97.8 |
| 6 | Mutism,Staring,Grimacing,Rigidity,Negativism,Waxy flexibility | 96.0 | 94.2 | 97.8 |
| 6 | Mutism,Staring,Grimacing,Verbigeration,Waxy flexibility,Withdrawal | 96.0 | 93.9 | 97.8 |
| 6 | Mutism,Staring,Posturing/catalepsy,Echopraxia/echolalia,Rigidity,Waxy flexibility | 96.0 | 93.9 | 97.8 |
| 6 | Mutism,Staring,Posturing/catalepsy,Echopraxia/echolalia,Stereotypy,Waxy flexibility | 96.0 | 93.9 | 97.8 |
| 6 | Mutism,Staring,Posturing/catalepsy,Mannerisms,Verbigeration,Waxy flexibility | 96.0 | 93.9 | 97.8 |
| 6 | Mutism,Staring,Posturing/catalepsy,Rigidity,Negativism,Waxy flexibility | 96.0 | 93.9 | 97.8 |
| 6 | Mutism,Staring,Posturing/catalepsy,Stereotypy,Mannerisms,Waxy flexibility | 96.0 | 93.9 | 97.8 |
| 6 | Mutism,Staring,Posturing/catalepsy,Stereotypy,Rigidity,Waxy flexibility | 96.0 | 93.9 | 97.8 |
| 6 | Mutism,Staring,Posturing/catalepsy,Verbigeration,Waxy flexibility,Withdrawal | 96.0 | 93.9 | 97.8 |
| 6 | Mutism,Staring,Stereotypy,Mannerisms,Verbigeration,Waxy flexibility | 96.0 | 93.9 | 97.8 |
| 6 | Staring,Echopraxia/echolalia,Stereotypy,Rigidity,Waxy flexibility,Withdrawal | 96.0 | 94.2 | 97.8 |
| 6 | Staring,Echopraxia/echolalia,Verbigeration,Negativism,Waxy flexibility,Withdrawal | 96.0 | 94.2 | 97.8 |
| 6 | Staring,Grimacing,Echopraxia/echolalia,Stereotypy,Mannerisms,Negativism | 96.0 | 93.9 | 97.8 |
| 6 | Staring,Grimacing,Echopraxia/echolalia,Stereotypy,Mannerisms,Rigidity | 96.0 | 93.9 | 97.8 |
| 6 | Staring,Grimacing,Echopraxia/echolalia,Verbigeration,Negativism,Withdrawal | 96.0 | 93.9 | 97.8 |
| 6 | Staring,Grimacing,Echopraxia/echolalia,Verbigeration,Rigidity,Withdrawal | 96.0 | 93.9 | 97.8 |
| 6 | Staring,Grimacing,Stereotypy,Negativism,Waxy flexibility,Withdrawal | 96.0 | 94.2 | 97.8 |
| 6 | Staring,Grimacing,Verbigeration,Rigidity,Waxy flexibility,Withdrawal | 96.0 | 93.9 | 97.8 |
| 6 | Staring,Posturing/catalepsy,Echopraxia/echolalia,Stereotypy,Mannerisms,Withdrawal | 96.0 | 94.2 | 97.8 |
| 6 | Staring,Posturing/catalepsy,Echopraxia/echolalia,Verbigeration,Rigidity,Negativism | 96.0 | 93.9 | 97.8 |
| 6 | Staring,Posturing/catalepsy,Grimacing,Mannerisms,Waxy flexibility,Withdrawal | 96.0 | 93.9 | 97.8 |
| 6 | Excitement,Immobility/stupor,Posturing/catalepsy,Grimacing,Mannerisms,Withdrawal | 95.9 | 93.9 | 97.8 |
| 6 | Excitement,Staring,Posturing/catalepsy,Grimacing,Echopraxia/echolalia,Mannerisms | 95.9 | 93.9 | 97.8 |
| 6 | Immobility/stupor,Echopraxia/echolalia,Stereotypy,Verbigeration,Rigidity,Negativism | 95.9 | 94.2 | 97.8 |
| 6 | Immobility/stupor,Staring,Posturing/catalepsy,Verbigeration,Rigidity,Withdrawal | 95.9 | 93.9 | 97.8 |
| 6 | Staring,Posturing/catalepsy,Stereotypy,Mannerisms,Negativism,Waxy flexibility | 95.9 | 93.9 | 97.8 |
| 6 | Excitement,Immobility/stupor,Grimacing,Echopraxia/echolalia,Waxy flexibility,Withdrawal | 95.8 | 93.7 | 97.5 |
| 6 | Excitement,Immobility/stupor,Mutism,Grimacing,Echopraxia/echolalia,Negativism | 95.8 | 93.7 | 97.5 |
| 6 | Excitement,Immobility/stupor,Mutism,Grimacing,Echopraxia/echolalia,Waxy flexibility | 95.8 | 93.7 | 97.5 |
| 6 | Excitement,Immobility/stupor,Mutism,Grimacing,Stereotypy,Negativism | 95.8 | 93.7 | 97.5 |
| 6 | Excitement,Immobility/stupor,Mutism,Posturing/catalepsy,Echopraxia/echolalia,Mannerisms | 95.8 | 93.7 | 97.5 |
| 6 | Excitement,Staring,Echopraxia/echolalia,Mannerisms,Verbigeration,Withdrawal | 95.8 | 93.7 | 97.5 |
| 6 | Excitement,Staring,Grimacing,Verbigeration,Negativism,Waxy flexibility | 95.8 | 93.7 | 97.5 |
| 6 | Excitement,Staring,Stereotypy,Mannerisms,Verbigeration,Negativism | 95.8 | 93.7 | 97.5 |
| 6 | Immobility/stupor,Posturing/catalepsy,Echopraxia/echolalia,Stereotypy,Negativism,Withdrawal | 95.8 | 93.7 | 97.5 |
| 6 | Immobility/stupor,Staring,Posturing/catalepsy,Grimacing,Echopraxia/echolalia,Mannerisms | 95.8 | 93.7 | 97.5 |
| 6 | Immobility/stupor,Staring,Verbigeration,Rigidity,Waxy flexibility,Withdrawal | 95.8 | 93.7 | 97.5 |
| 6 | Staring,Echopraxia/echolalia,Stereotypy,Rigidity,Negativism,Waxy flexibility | 95.8 | 93.7 | 97.5 |
| 6 | Staring,Grimacing,Verbigeration,Negativism,Waxy flexibility,Withdrawal | 95.8 | 93.7 | 97.5 |
| 6 | Staring,Posturing/catalepsy,Grimacing,Mannerisms,Verbigeration,Rigidity | 95.8 | 93.9 | 97.5 |
| 6 | Staring,Stereotypy,Mannerisms,Verbigeration,Negativism,Waxy flexibility | 95.8 | 93.7 | 97.5 |
| 6 | Excitement,Immobility/stupor,Echopraxia/echolalia,Stereotypy,Verbigeration,Negativism | 95.7 | 93.7 | 97.5 |
| 6 | Excitement,Immobility/stupor,Echopraxia/echolalia,Verbigeration,Negativism,Waxy flexibility | 95.7 | 93.7 | 97.5 |
| 6 | Excitement,Immobility/stupor,Mannerisms,Negativism,Waxy flexibility,Withdrawal | 95.7 | 93.7 | 97.5 |
| 6 | Excitement,Immobility/stupor,Mutism,Grimacing,Echopraxia/echolalia,Withdrawal | 95.7 | 93.7 | 97.5 |
| 6 | Excitement,Immobility/stupor,Mutism,Grimacing,Waxy flexibility,Withdrawal | 95.7 | 93.7 | 97.5 |
| 6 | Excitement,Immobility/stupor,Mutism,Mannerisms,Negativism,Waxy flexibility | 95.7 | 93.7 | 97.5 |
| 6 | Excitement,Immobility/stupor,Mutism,Posturing/catalepsy,Mannerisms,Verbigeration | 95.7 | 93.7 | 97.5 |
| 6 | Excitement,Immobility/stupor,Mutism,Posturing/catalepsy,Stereotypy,Verbigeration | 95.7 | 93.7 | 97.5 |
| 6 | Excitement,Immobility/stupor,Mutism,Verbigeration,Waxy flexibility,Withdrawal | 95.7 | 93.7 | 97.5 |
| 6 | Excitement,Immobility/stupor,Posturing/catalepsy,Stereotypy,Mannerisms,Negativism | 95.7 | 93.7 | 97.5 |
| 6 | Excitement,Immobility/stupor,Posturing/catalepsy,Stereotypy,Verbigeration,Withdrawal | 95.7 | 93.7 | 97.5 |
| 6 | Excitement,Immobility/stupor,Posturing/catalepsy,Verbigeration,Waxy flexibility,Withdrawal | 95.7 | 93.7 | 97.5 |
| 6 | Excitement,Staring,Echopraxia/echolalia,Mannerisms,Verbigeration,Rigidity | 95.7 | 93.7 | 97.5 |
| 6 | Excitement,Staring,Grimacing,Mannerisms,Negativism,Waxy flexibility | 95.7 | 93.7 | 97.5 |
| 6 | Excitement,Staring,Grimacing,Stereotypy,Negativism,Waxy flexibility | 95.7 | 93.7 | 97.5 |
| 6 | Excitement,Staring,Posturing/catalepsy,Grimacing,Echopraxia/echolalia,Stereotypy | 95.7 | 93.7 | 97.5 |
| 6 | Excitement,Staring,Posturing/catalepsy,Grimacing,Echopraxia/echolalia,Waxy flexibility | 95.7 | 93.7 | 97.5 |
| 6 | Excitement,Staring,Posturing/catalepsy,Grimacing,Mannerisms,Verbigeration | 95.7 | 93.7 | 97.5 |
| 6 | Immobility/stupor,Grimacing,Echopraxia/echolalia,Mannerisms,Rigidity,Withdrawal | 95.7 | 93.7 | 97.5 |
| 6 | Immobility/stupor,Grimacing,Echopraxia/echolalia,Verbigeration,Rigidity,Negativism | 95.7 | 93.7 | 97.5 |
| 6 | Immobility/stupor,Mutism,Echopraxia/echolalia,Rigidity,Negativism,Withdrawal | 95.7 | 93.7 | 97.5 |
| 6 | Immobility/stupor,Mutism,Staring,Echopraxia/echolalia,Stereotypy,Mannerisms | 95.7 | 93.7 | 97.5 |
| 6 | Immobility/stupor,Mutism,Staring,Grimacing,Echopraxia/echolalia,Mannerisms | 95.7 | 93.7 | 97.5 |
| 6 | Immobility/stupor,Mutism,Staring,Grimacing,Negativism,Withdrawal | 95.7 | 93.7 | 97.5 |
| 6 | Immobility/stupor,Mutism,Staring,Grimacing,Rigidity,Withdrawal | 95.7 | 93.7 | 97.5 |
| 6 | Immobility/stupor,Mutism,Staring,Grimacing,Stereotypy,Verbigeration | 95.7 | 93.7 | 97.5 |
| 6 | Immobility/stupor,Mutism,Staring,Mannerisms,Verbigeration,Withdrawal | 95.7 | 93.7 | 97.5 |
| 6 | Immobility/stupor,Mutism,Staring,Negativism,Waxy flexibility,Withdrawal | 95.7 | 93.7 | 97.5 |
| 6 | Immobility/stupor,Mutism,Staring,Posturing/catalepsy,Grimacing,Mannerisms | 95.7 | 93.7 | 97.5 |
| 6 | Immobility/stupor,Mutism,Staring,Posturing/catalepsy,Stereotypy,Mannerisms | 95.7 | 93.7 | 97.5 |
| 6 | Immobility/stupor,Mutism,Staring,Posturing/catalepsy,Stereotypy,Verbigeration | 95.7 | 93.7 | 97.5 |
| 6 | Immobility/stupor,Mutism,Staring,Posturing/catalepsy,Stereotypy,Withdrawal | 95.7 | 93.7 | 97.5 |
| 6 | Immobility/stupor,Mutism,Staring,Verbigeration,Rigidity,Waxy flexibility | 95.7 | 93.7 | 97.5 |
| 6 | Immobility/stupor,Posturing/catalepsy,Echopraxia/echolalia,Verbigeration,Rigidity,Negativism | 95.7 | 93.7 | 97.5 |
| 6 | Immobility/stupor,Posturing/catalepsy,Grimacing,Stereotypy,Rigidity,Negativism | 95.7 | 93.7 | 97.5 |
| 6 | Immobility/stupor,Staring,Echopraxia/echolalia,Verbigeration,Negativism,Waxy flexibility | 95.7 | 93.7 | 97.5 |
| 6 | Immobility/stupor,Staring,Grimacing,Echopraxia/echolalia,Stereotypy,Withdrawal | 95.7 | 93.7 | 97.5 |
| 6 | Immobility/stupor,Staring,Grimacing,Mannerisms,Verbigeration,Withdrawal | 95.7 | 93.7 | 97.5 |
| 6 | Immobility/stupor,Staring,Grimacing,Mannerisms,Waxy flexibility,Withdrawal | 95.7 | 93.7 | 97.5 |
| 6 | Immobility/stupor,Staring,Grimacing,Rigidity,Negativism,Waxy flexibility | 95.7 | 93.7 | 97.5 |
| 6 | Immobility/stupor,Staring,Grimacing,Stereotypy,Mannerisms,Verbigeration | 95.7 | 93.7 | 97.5 |
| 6 | Immobility/stupor,Staring,Grimacing,Stereotypy,Negativism,Waxy flexibility | 95.7 | 93.7 | 97.5 |
| 6 | Immobility/stupor,Staring,Grimacing,Stereotypy,Rigidity,Waxy flexibility | 95.7 | 93.7 | 97.5 |
| 6 | Immobility/stupor,Staring,Grimacing,Stereotypy,Verbigeration,Withdrawal | 95.7 | 93.7 | 97.5 |
| 6 | Immobility/stupor,Staring,Grimacing,Verbigeration,Negativism,Waxy flexibility | 95.7 | 93.7 | 97.5 |
| 6 | Immobility/stupor,Staring,Grimacing,Verbigeration,Rigidity,Waxy flexibility | 95.7 | 93.7 | 97.5 |
| 6 | Immobility/stupor,Staring,Posturing/catalepsy,Echopraxia/echolalia,Stereotypy,Rigidity | 95.7 | 93.7 | 97.5 |
| 6 | Immobility/stupor,Staring,Posturing/catalepsy,Echopraxia/echolalia,Verbigeration,Rigidity | 95.7 | 93.7 | 97.5 |
| 6 | Immobility/stupor,Staring,Posturing/catalepsy,Grimacing,Mannerisms,Verbigeration | 95.7 | 93.7 | 97.5 |
| 6 | Immobility/stupor,Staring,Posturing/catalepsy,Grimacing,Rigidity,Withdrawal | 95.7 | 93.7 | 97.5 |
| 6 | Immobility/stupor,Staring,Posturing/catalepsy,Grimacing,Stereotypy,Mannerisms | 95.7 | 93.7 | 97.5 |
| 6 | Immobility/stupor,Staring,Posturing/catalepsy,Stereotypy,Mannerisms,Verbigeration | 95.7 | 93.7 | 97.5 |
| 6 | Immobility/stupor,Staring,Stereotypy,Mannerisms,Verbigeration,Withdrawal | 95.7 | 93.7 | 97.5 |
| 6 | Immobility/stupor,Staring,Stereotypy,Mannerisms,Waxy flexibility,Withdrawal | 95.7 | 93.7 | 97.5 |
| 6 | Immobility/stupor,Verbigeration,Rigidity,Negativism,Waxy flexibility,Withdrawal | 95.7 | 93.7 | 97.5 |
| 6 | Mutism,Staring,Echopraxia/echolalia,Mannerisms,Verbigeration,Waxy flexibility | 95.7 | 93.7 | 97.5 |
| 6 | Mutism,Staring,Echopraxia/echolalia,Stereotypy,Verbigeration,Waxy flexibility | 95.7 | 93.7 | 97.5 |
| 6 | Mutism,Staring,Posturing/catalepsy,Echopraxia/echolalia,Verbigeration,Waxy flexibility | 95.7 | 93.7 | 97.5 |
| 6 | Mutism,Staring,Posturing/catalepsy,Grimacing,Mannerisms,Waxy flexibility | 95.7 | 93.7 | 97.5 |
| 6 | Mutism,Staring,Posturing/catalepsy,Grimacing,Negativism,Waxy flexibility | 95.7 | 93.7 | 97.5 |
| 6 | Mutism,Staring,Posturing/catalepsy,Grimacing,Waxy flexibility,Withdrawal | 95.7 | 93.7 | 97.5 |
| 6 | Staring,Echopraxia/echolalia,Stereotypy,Mannerisms,Waxy flexibility,Withdrawal | 95.7 | 93.7 | 97.5 |
| 6 | Staring,Grimacing,Echopraxia/echolalia,Negativism,Waxy flexibility,Withdrawal | 95.7 | 93.7 | 97.5 |
| 6 | Staring,Grimacing,Echopraxia/echolalia,Rigidity,Waxy flexibility,Withdrawal | 95.7 | 93.7 | 97.5 |
| 6 | Staring,Grimacing,Echopraxia/echolalia,Stereotypy,Rigidity,Negativism | 95.7 | 93.7 | 97.5 |
| 6 | Staring,Grimacing,Mannerisms,Verbigeration,Waxy flexibility,Withdrawal | 95.7 | 93.7 | 97.5 |
| 6 | Staring,Grimacing,Stereotypy,Verbigeration,Rigidity,Waxy flexibility | 95.7 | 93.7 | 97.5 |
| 6 | Staring,Posturing/catalepsy,Echopraxia/echolalia,Mannerisms,Negativism,Waxy flexibility | 95.7 | 93.7 | 97.5 |
| 6 | Staring,Posturing/catalepsy,Echopraxia/echolalia,Mannerisms,Verbigeration,Negativism | 95.7 | 93.7 | 97.5 |
| 6 | Staring,Posturing/catalepsy,Echopraxia/echolalia,Verbigeration,Rigidity,Withdrawal | 95.7 | 93.7 | 97.5 |
| 6 | Staring,Posturing/catalepsy,Grimacing,Echopraxia/echolalia,Mannerisms,Rigidity | 95.7 | 93.7 | 97.5 |
| 6 | Staring,Posturing/catalepsy,Grimacing,Echopraxia/echolalia,Stereotypy,Withdrawal | 95.7 | 93.7 | 97.5 |
| 6 | Staring,Posturing/catalepsy,Grimacing,Mannerisms,Verbigeration,Negativism | 95.7 | 93.7 | 97.5 |
| 6 | Staring,Posturing/catalepsy,Grimacing,Negativism,Waxy flexibility,Withdrawal | 95.7 | 93.7 | 97.5 |
| 6 | Staring,Posturing/catalepsy,Grimacing,Stereotypy,Rigidity,Negativism | 95.7 | 93.7 | 97.5 |
| 6 | Staring,Posturing/catalepsy,Grimacing,Stereotypy,Verbigeration,Rigidity | 95.7 | 93.7 | 97.5 |
| 6 | Staring,Posturing/catalepsy,Grimacing,Stereotypy,Verbigeration,Withdrawal | 95.7 | 93.7 | 97.5 |
| 6 | Staring,Posturing/catalepsy,Stereotypy,Mannerisms,Verbigeration,Withdrawal | 95.7 | 93.7 | 97.5 |
| 6 | Staring,Posturing/catalepsy,Stereotypy,Mannerisms,Waxy flexibility,Withdrawal | 95.7 | 93.7 | 97.5 |
| 6 | Staring,Posturing/catalepsy,Stereotypy,Rigidity,Waxy flexibility,Withdrawal | 95.7 | 93.7 | 97.5 |
| 6 | Excitement,Immobility/stupor,Echopraxia/echolalia,Stereotypy,Verbigeration,Withdrawal | 95.5 | 93.5 | 97.3 |
| 6 | Excitement,Immobility/stupor,Echopraxia/echolalia,Stereotypy,Waxy flexibility,Withdrawal | 95.5 | 93.5 | 97.3 |
| 6 | Excitement,Immobility/stupor,Echopraxia/echolalia,Verbigeration,Waxy flexibility,Withdrawal | 95.5 | 93.5 | 97.3 |
| 6 | Excitement,Immobility/stupor,Grimacing,Echopraxia/echolalia,Stereotypy,Negativism | 95.5 | 93.5 | 97.3 |
| 6 | Excitement,Immobility/stupor,Grimacing,Echopraxia/echolalia,Verbigeration,Negativism | 95.5 | 93.5 | 97.3 |
| 6 | Excitement,Immobility/stupor,Grimacing,Negativism,Waxy flexibility,Withdrawal | 95.5 | 93.5 | 97.3 |
| 6 | Excitement,Immobility/stupor,Grimacing,Stereotypy,Mannerisms,Rigidity | 95.5 | 93.5 | 97.3 |
| 6 | Excitement,Immobility/stupor,Grimacing,Stereotypy,Negativism,Waxy flexibility | 95.5 | 93.5 | 97.3 |
| 6 | Excitement,Immobility/stupor,Grimacing,Stereotypy,Verbigeration,Negativism | 95.5 | 93.5 | 97.3 |
| 6 | Excitement,Immobility/stupor,Grimacing,Verbigeration,Negativism,Waxy flexibility | 95.5 | 93.5 | 97.3 |
| 6 | Excitement,Immobility/stupor,Mutism,Echopraxia/echolalia,Stereotypy,Waxy flexibility | 95.5 | 93.5 | 97.3 |
| 6 | Excitement,Immobility/stupor,Mutism,Echopraxia/echolalia,Verbigeration,Waxy flexibility | 95.5 | 93.5 | 97.3 |
| 6 | Excitement,Immobility/stupor,Mutism,Grimacing,Stereotypy,Withdrawal | 95.5 | 93.5 | 97.3 |
| 6 | Excitement,Immobility/stupor,Mutism,Grimacing,Verbigeration,Withdrawal | 95.5 | 93.5 | 97.3 |
| 6 | Excitement,Immobility/stupor,Mutism,Mannerisms,Negativism,Withdrawal | 95.5 | 93.5 | 97.3 |
| 6 | Excitement,Immobility/stupor,Mutism,Mannerisms,Verbigeration,Negativism | 95.5 | 93.5 | 97.3 |
| 6 | Excitement,Immobility/stupor,Mutism,Posturing/catalepsy,Stereotypy,Waxy flexibility | 95.5 | 93.5 | 97.3 |
| 6 | Excitement,Immobility/stupor,Posturing/catalepsy,Grimacing,Echopraxia/echolalia,Waxy flexibility | 95.5 | 93.5 | 97.3 |
| 6 | Excitement,Immobility/stupor,Posturing/catalepsy,Grimacing,Stereotypy,Withdrawal | 95.5 | 93.5 | 97.3 |
| 6 | Excitement,Immobility/stupor,Posturing/catalepsy,Grimacing,Waxy flexibility,Withdrawal | 95.5 | 93.5 | 97.3 |
| 6 | Excitement,Immobility/stupor,Stereotypy,Mannerisms,Rigidity,Waxy flexibility | 95.5 | 93.5 | 97.3 |
| 6 | Excitement,Mutism,Posturing/catalepsy,Rigidity,Negativism,Withdrawal | 95.5 | 93.5 | 97.3 |
| 6 | Excitement,Staring,Echopraxia/echolalia,Mannerisms,Verbigeration,Negativism | 95.5 | 93.5 | 97.3 |
| 6 | Excitement,Staring,Grimacing,Echopraxia/echolalia,Mannerisms,Negativism | 95.5 | 93.5 | 97.3 |
| 6 | Excitement,Staring,Grimacing,Echopraxia/echolalia,Mannerisms,Waxy flexibility | 95.5 | 93.5 | 97.3 |
| 6 | Excitement,Staring,Grimacing,Echopraxia/echolalia,Stereotypy,Waxy flexibility | 95.5 | 93.5 | 97.3 |
| 6 | Excitement,Staring,Grimacing,Stereotypy,Verbigeration,Negativism | 95.5 | 93.5 | 97.3 |
| 6 | Excitement,Staring,Posturing/catalepsy,Grimacing,Echopraxia/echolalia,Verbigeration | 95.5 | 93.5 | 97.3 |
| 6 | Excitement,Staring,Posturing/catalepsy,Grimacing,Mannerisms,Waxy flexibility | 95.5 | 93.5 | 97.3 |
| 6 | Excitement,Staring,Posturing/catalepsy,Grimacing,Stereotypy,Mannerisms | 95.5 | 93.5 | 97.3 |
| 6 | Excitement,Staring,Posturing/catalepsy,Grimacing,Stereotypy,Verbigeration | 95.5 | 93.5 | 97.3 |
| 6 | Immobility/stupor,Echopraxia/echolalia,Rigidity,Negativism,Waxy flexibility,Withdrawal | 95.5 | 93.5 | 97.3 |
| 6 | Immobility/stupor,Echopraxia/echolalia,Stereotypy,Verbigeration,Negativism,Withdrawal | 95.5 | 93.5 | 97.3 |
| 6 | Immobility/stupor,Grimacing,Echopraxia/echolalia,Stereotypy,Negativism,Withdrawal | 95.5 | 93.5 | 97.3 |
| 6 | Immobility/stupor,Grimacing,Echopraxia/echolalia,Stereotypy,Rigidity,Negativism | 95.5 | 93.5 | 97.3 |
| 6 | Immobility/stupor,Grimacing,Echopraxia/echolalia,Stereotypy,Rigidity,Withdrawal | 95.5 | 93.5 | 97.3 |
| 6 | Immobility/stupor,Grimacing,Stereotypy,Mannerisms,Negativism,Withdrawal | 95.5 | 93.5 | 97.3 |
| 6 | Immobility/stupor,Grimacing,Stereotypy,Mannerisms,Rigidity,Withdrawal | 95.5 | 93.5 | 97.3 |
| 6 | Immobility/stupor,Grimacing,Stereotypy,Rigidity,Negativism,Waxy flexibility | 95.5 | 93.5 | 97.3 |
| 6 | Immobility/stupor,Mutism,Echopraxia/echolalia,Mannerisms,Negativism,Withdrawal | 95.5 | 93.5 | 97.3 |
| 6 | Immobility/stupor,Mutism,Echopraxia/echolalia,Mannerisms,Rigidity,Withdrawal | 95.5 | 93.5 | 97.3 |
| 6 | Immobility/stupor,Mutism,Echopraxia/echolalia,Stereotypy,Rigidity,Negativism | 95.5 | 93.5 | 97.3 |
| 6 | Immobility/stupor,Mutism,Grimacing,Stereotypy,Rigidity,Negativism | 95.5 | 93.5 | 97.3 |
| 6 | Immobility/stupor,Mutism,Mannerisms,Verbigeration,Rigidity,Withdrawal | 95.5 | 93.5 | 97.3 |
| 6 | Immobility/stupor,Mutism,Posturing/catalepsy,Echopraxia/echolalia,Mannerisms,Negativism | 95.5 | 93.5 | 97.3 |
| 6 | Immobility/stupor,Mutism,Posturing/catalepsy,Grimacing,Mannerisms,Negativism | 95.5 | 93.5 | 97.3 |
| 6 | Immobility/stupor,Mutism,Posturing/catalepsy,Mannerisms,Rigidity,Withdrawal | 95.5 | 93.5 | 97.3 |
| 6 | Immobility/stupor,Mutism,Staring,Echopraxia/echolalia,Mannerisms,Waxy flexibility | 95.5 | 93.5 | 97.3 |
| 6 | Immobility/stupor,Mutism,Staring,Grimacing,Echopraxia/echolalia,Negativism | 95.5 | 93.5 | 97.3 |
| 6 | Immobility/stupor,Mutism,Staring,Grimacing,Echopraxia/echolalia,Rigidity | 95.5 | 93.5 | 97.3 |
| 6 | Immobility/stupor,Mutism,Staring,Grimacing,Mannerisms,Verbigeration | 95.5 | 93.5 | 97.3 |
| 6 | Immobility/stupor,Mutism,Staring,Grimacing,Rigidity,Negativism | 95.5 | 93.5 | 97.3 |
| 6 | Immobility/stupor,Mutism,Staring,Grimacing,Stereotypy,Mannerisms | 95.5 | 93.5 | 97.3 |
| 6 | Immobility/stupor,Mutism,Staring,Grimacing,Stereotypy,Withdrawal | 95.5 | 93.5 | 97.3 |
| 6 | Immobility/stupor,Mutism,Staring,Posturing/catalepsy,Echopraxia/echolalia,Stereotypy | 95.5 | 93.5 | 97.3 |
| 6 | Immobility/stupor,Mutism,Staring,Posturing/catalepsy,Echopraxia/echolalia,Verbigeration | 95.5 | 93.5 | 97.3 |
| 6 | Immobility/stupor,Mutism,Staring,Posturing/catalepsy,Grimacing,Negativism | 95.5 | 93.5 | 97.3 |
| 6 | Immobility/stupor,Mutism,Staring,Posturing/catalepsy,Grimacing,Verbigeration | 95.5 | 93.5 | 97.3 |
| 6 | Immobility/stupor,Mutism,Staring,Posturing/catalepsy,Grimacing,Withdrawal | 95.5 | 93.5 | 97.3 |
| 6 | Immobility/stupor,Mutism,Staring,Posturing/catalepsy,Verbigeration,Withdrawal | 95.5 | 93.5 | 97.3 |
| 6 | Immobility/stupor,Mutism,Staring,Rigidity,Negativism,Waxy flexibility | 95.5 | 93.5 | 97.3 |
| 6 | Immobility/stupor,Mutism,Staring,Rigidity,Waxy flexibility,Withdrawal | 95.5 | 93.5 | 97.3 |
| 6 | Immobility/stupor,Mutism,Staring,Stereotypy,Mannerisms,Verbigeration | 95.5 | 93.5 | 97.3 |
| 6 | Immobility/stupor,Mutism,Staring,Stereotypy,Rigidity,Waxy flexibility | 95.5 | 93.5 | 97.3 |
| 6 | Immobility/stupor,Mutism,Staring,Verbigeration,Negativism,Waxy flexibility | 95.5 | 93.5 | 97.3 |
| 6 | Immobility/stupor,Mutism,Stereotypy,Mannerisms,Negativism,Withdrawal | 95.5 | 93.5 | 97.3 |
| 6 | Immobility/stupor,Mutism,Stereotypy,Mannerisms,Rigidity,Withdrawal | 95.5 | 93.5 | 97.3 |
| 6 | Immobility/stupor,Mutism,Stereotypy,Rigidity,Negativism,Waxy flexibility | 95.5 | 93.5 | 97.3 |
| 6 | Immobility/stupor,Mutism,Verbigeration,Rigidity,Negativism,Waxy flexibility | 95.5 | 93.5 | 97.3 |
| 6 | Immobility/stupor,Posturing/catalepsy,Echopraxia/echolalia,Stereotypy,Rigidity,Negativism | 95.5 | 93.5 | 97.3 |
| 6 | Immobility/stupor,Posturing/catalepsy,Grimacing,Mannerisms,Rigidity,Withdrawal | 95.5 | 93.5 | 97.3 |
| 6 | Immobility/stupor,Posturing/catalepsy,Grimacing,Stereotypy,Negativism,Withdrawal | 95.5 | 93.5 | 97.3 |
| 6 | Immobility/stupor,Posturing/catalepsy,Stereotypy,Rigidity,Negativism,Waxy flexibility | 95.5 | 93.5 | 97.3 |
| 6 | Immobility/stupor,Staring,Echopraxia/echolalia,Mannerisms,Waxy flexibility,Withdrawal | 95.5 | 93.5 | 97.3 |
| 6 | Immobility/stupor,Staring,Echopraxia/echolalia,Rigidity,Waxy flexibility,Withdrawal | 95.5 | 93.5 | 97.3 |
| 6 | Immobility/stupor,Staring,Grimacing,Echopraxia/echolalia,Mannerisms,Waxy flexibility | 95.5 | 93.5 | 97.3 |
| 6 | Immobility/stupor,Staring,Grimacing,Stereotypy,Mannerisms,Waxy flexibility | 95.5 | 93.5 | 97.3 |
| 6 | Immobility/stupor,Staring,Posturing/catalepsy,Echopraxia/echolalia,Negativism,Waxy flexibility | 95.5 | 93.5 | 97.3 |
| 6 | Immobility/stupor,Staring,Posturing/catalepsy,Echopraxia/echolalia,Stereotypy,Mannerisms | 95.5 | 93.5 | 97.3 |
| 6 | Immobility/stupor,Staring,Posturing/catalepsy,Echopraxia/echolalia,Stereotypy,Withdrawal | 95.5 | 93.5 | 97.3 |
| 6 | Immobility/stupor,Staring,Posturing/catalepsy,Grimacing,Stereotypy,Verbigeration | 95.5 | 93.5 | 97.3 |
| 6 | Immobility/stupor,Staring,Posturing/catalepsy,Grimacing,Stereotypy,Withdrawal | 95.5 | 93.5 | 97.3 |
| 6 | Immobility/stupor,Staring,Posturing/catalepsy,Mannerisms,Rigidity,Waxy flexibility | 95.5 | 93.5 | 97.3 |
| 6 | Immobility/stupor,Staring,Posturing/catalepsy,Mannerisms,Waxy flexibility,Withdrawal | 95.5 | 93.5 | 97.3 |
| 6 | Immobility/stupor,Stereotypy,Mannerisms,Negativism,Waxy flexibility,Withdrawal | 95.5 | 93.5 | 97.3 |
| 6 | Mutism,Staring,Echopraxia/echolalia,Verbigeration,Waxy flexibility,Withdrawal | 95.5 | 93.5 | 97.3 |
| 6 | Mutism,Staring,Posturing/catalepsy,Grimacing,Verbigeration,Waxy flexibility | 95.5 | 93.5 | 97.3 |
| 6 | Staring,Echopraxia/echolalia,Stereotypy,Mannerisms,Verbigeration,Negativism | 95.5 | 93.5 | 97.3 |
| 6 | Staring,Echopraxia/echolalia,Verbigeration,Rigidity,Negativism,Waxy flexibility | 95.5 | 93.5 | 97.3 |
| 6 | Staring,Grimacing,Echopraxia/echolalia,Mannerisms,Negativism,Waxy flexibility | 95.5 | 93.5 | 97.3 |
| 6 | Staring,Grimacing,Echopraxia/echolalia,Mannerisms,Rigidity,Waxy flexibility | 95.5 | 93.5 | 97.3 |
| 6 | Staring,Grimacing,Echopraxia/echolalia,Mannerisms,Verbigeration,Withdrawal | 95.5 | 93.5 | 97.3 |
| 6 | Staring,Grimacing,Echopraxia/echolalia,Verbigeration,Rigidity,Negativism | 95.5 | 93.5 | 97.3 |
| 6 | Staring,Grimacing,Mannerisms,Verbigeration,Rigidity,Waxy flexibility | 95.5 | 93.5 | 97.3 |
| 6 | Staring,Grimacing,Stereotypy,Mannerisms,Negativism,Waxy flexibility | 95.5 | 93.5 | 97.3 |
| 6 | Staring,Grimacing,Stereotypy,Rigidity,Negativism,Waxy flexibility | 95.5 | 93.5 | 97.3 |
| 6 | Staring,Posturing/catalepsy,Echopraxia/echolalia,Mannerisms,Waxy flexibility,Withdrawal | 95.5 | 93.5 | 97.3 |
| 6 | Staring,Posturing/catalepsy,Grimacing,Echopraxia/echolalia,Rigidity,Negativism | 95.5 | 93.5 | 97.3 |
| 6 | Staring,Posturing/catalepsy,Grimacing,Stereotypy,Verbigeration,Negativism | 95.5 | 93.5 | 97.3 |
| 6 | Staring,Posturing/catalepsy,Grimacing,Verbigeration,Rigidity,Negativism | 95.5 | 93.5 | 97.3 |
| 6 | Staring,Posturing/catalepsy,Stereotypy,Mannerisms,Verbigeration,Rigidity | 95.5 | 93.5 | 97.3 |
| 6 | Staring,Posturing/catalepsy,Stereotypy,Rigidity,Negativism,Waxy flexibility | 95.5 | 93.5 | 97.3 |
| 6 | Excitement,Immobility/stupor,Echopraxia/echolalia,Mannerisms,Negativism,Waxy flexibility | 95.3 | 93.3 | 97.1 |
| 6 | Excitement,Immobility/stupor,Echopraxia/echolalia,Mannerisms,Waxy flexibility,Withdrawal | 95.3 | 93.3 | 97.1 |
| 6 | Excitement,Immobility/stupor,Grimacing,Echopraxia/echolalia,Stereotypy,Withdrawal | 95.3 | 93.3 | 97.1 |
| 6 | Excitement,Immobility/stupor,Grimacing,Mannerisms,Negativism,Waxy flexibility | 95.3 | 93.3 | 97.1 |
| 6 | Excitement,Immobility/stupor,Grimacing,Mannerisms,Negativism,Withdrawal | 95.3 | 93.3 | 97.1 |
| 6 | Excitement,Immobility/stupor,Mannerisms,Verbigeration,Negativism,Waxy flexibility | 95.3 | 93.3 | 97.1 |
| 6 | Excitement,Immobility/stupor,Mutism,Grimacing,Stereotypy,Waxy flexibility | 95.3 | 93.3 | 97.1 |
| 6 | Excitement,Immobility/stupor,Mutism,Grimacing,Verbigeration,Waxy flexibility | 95.3 | 93.3 | 97.1 |
| 6 | Excitement,Immobility/stupor,Mutism,Mannerisms,Waxy flexibility,Withdrawal | 95.3 | 93.3 | 97.1 |
| 6 | Excitement,Immobility/stupor,Mutism,Posturing/catalepsy,Mannerisms,Waxy flexibility | 95.3 | 93.3 | 97.1 |
| 6 | Excitement,Immobility/stupor,Mutism,Stereotypy,Mannerisms,Negativism | 95.3 | 93.3 | 97.1 |
| 6 | Excitement,Immobility/stupor,Mutism,Stereotypy,Verbigeration,Waxy flexibility | 95.3 | 93.3 | 97.1 |
| 6 | Excitement,Immobility/stupor,Posturing/catalepsy,Grimacing,Echopraxia/echolalia,Mannerisms | 95.3 | 93.3 | 97.1 |
| 6 | Excitement,Immobility/stupor,Posturing/catalepsy,Grimacing,Echopraxia/echolalia,Verbigeration | 95.3 | 93.3 | 97.1 |
| 6 | Excitement,Immobility/stupor,Posturing/catalepsy,Mannerisms,Waxy flexibility,Withdrawal | 95.3 | 93.3 | 97.1 |
| 6 | Excitement,Immobility/stupor,Posturing/catalepsy,Stereotypy,Mannerisms,Withdrawal | 95.3 | 93.3 | 97.1 |
| 6 | Excitement,Immobility/stupor,Stereotypy,Mannerisms,Verbigeration,Negativism | 95.3 | 93.3 | 97.1 |
| 6 | Excitement,Staring,Grimacing,Echopraxia/echolalia,Verbigeration,Negativism | 95.3 | 93.3 | 97.1 |
| 6 | Excitement,Staring,Posturing/catalepsy,Grimacing,Stereotypy,Waxy flexibility | 95.3 | 93.3 | 97.1 |
| 6 | Excitement,Staring,Posturing/catalepsy,Grimacing,Verbigeration,Waxy flexibility | 95.3 | 93.3 | 97.1 |
| 6 | Excitement,Staring,Posturing/catalepsy,Mannerisms,Verbigeration,Waxy flexibility | 95.3 | 93.3 | 97.1 |
| 6 | Immobility/stupor,Echopraxia/echolalia,Mannerisms,Verbigeration,Negativism,Withdrawal | 95.3 | 93.3 | 97.1 |
| 6 | Immobility/stupor,Echopraxia/echolalia,Stereotypy,Mannerisms,Negativism,Waxy flexibility | 95.3 | 93.3 | 97.1 |
| 6 | Immobility/stupor,Echopraxia/echolalia,Stereotypy,Mannerisms,Rigidity,Withdrawal | 95.3 | 93.3 | 97.1 |
| 6 | Immobility/stupor,Echopraxia/echolalia,Stereotypy,Rigidity,Negativism,Waxy flexibility | 95.3 | 93.3 | 97.1 |
| 6 | Immobility/stupor,Grimacing,Stereotypy,Verbigeration,Negativism,Withdrawal | 95.3 | 93.3 | 97.1 |
| 6 | Immobility/stupor,Grimacing,Stereotypy,Verbigeration,Rigidity,Withdrawal | 95.3 | 93.3 | 97.1 |
| 6 | Immobility/stupor,Grimacing,Verbigeration,Rigidity,Negativism,Waxy flexibility | 95.3 | 93.3 | 97.1 |
| 6 | Immobility/stupor,Mutism,Grimacing,Stereotypy,Rigidity,Withdrawal | 95.3 | 93.3 | 97.1 |
| 6 | Immobility/stupor,Mutism,Grimacing,Stereotypy,Verbigeration,Rigidity | 95.3 | 93.3 | 97.1 |
| 6 | Immobility/stupor,Mutism,Grimacing,Verbigeration,Rigidity,Withdrawal | 95.3 | 93.3 | 97.1 |
| 6 | Immobility/stupor,Mutism,Mannerisms,Negativism,Waxy flexibility,Withdrawal | 95.3 | 93.3 | 97.1 |
| 6 | Immobility/stupor,Mutism,Mannerisms,Verbigeration,Negativism,Withdrawal | 95.3 | 93.3 | 97.1 |
| 6 | Immobility/stupor,Mutism,Posturing/catalepsy,Echopraxia/echolalia,Negativism,Withdrawal | 95.3 | 93.3 | 97.1 |
| 6 | Immobility/stupor,Mutism,Posturing/catalepsy,Echopraxia/echolalia,Rigidity,Negativism | 95.3 | 93.3 | 97.1 |
| 6 | Immobility/stupor,Mutism,Posturing/catalepsy,Mannerisms,Verbigeration,Negativism | 95.3 | 93.3 | 97.1 |
| 6 | Immobility/stupor,Mutism,Posturing/catalepsy,Rigidity,Negativism,Withdrawal | 95.3 | 93.3 | 97.3 |
| 6 | Immobility/stupor,Mutism,Posturing/catalepsy,Stereotypy,Mannerisms,Negativism | 95.3 | 93.3 | 97.1 |
| 6 | Immobility/stupor,Mutism,Posturing/catalepsy,Stereotypy,Negativism,Withdrawal | 95.3 | 93.3 | 97.1 |
| 6 | Immobility/stupor,Mutism,Posturing/catalepsy,Stereotypy,Verbigeration,Negativism | 95.3 | 93.3 | 97.1 |
| 6 | Immobility/stupor,Mutism,Posturing/catalepsy,Verbigeration,Negativism,Withdrawal | 95.3 | 93.3 | 97.1 |
| 6 | Immobility/stupor,Mutism,Staring,Echopraxia/echolalia,Mannerisms,Verbigeration | 95.3 | 93.3 | 97.1 |
| 6 | Immobility/stupor,Mutism,Staring,Echopraxia/echolalia,Negativism,Waxy flexibility | 95.3 | 93.3 | 97.1 |
| 6 | Immobility/stupor,Mutism,Staring,Echopraxia/echolalia,Stereotypy,Withdrawal | 95.3 | 93.3 | 97.1 |
| 6 | Immobility/stupor,Mutism,Staring,Grimacing,Echopraxia/echolalia,Stereotypy | 95.3 | 93.3 | 97.1 |
| 6 | Immobility/stupor,Mutism,Staring,Grimacing,Echopraxia/echolalia,Verbigeration | 95.3 | 93.3 | 97.1 |
| 6 | Immobility/stupor,Mutism,Staring,Grimacing,Echopraxia/echolalia,Withdrawal | 95.3 | 93.3 | 97.1 |
| 6 | Immobility/stupor,Mutism,Staring,Grimacing,Mannerisms,Waxy flexibility | 95.3 | 93.3 | 97.1 |
| 6 | Immobility/stupor,Mutism,Staring,Grimacing,Verbigeration,Withdrawal | 95.3 | 93.3 | 97.1 |
| 6 | Immobility/stupor,Mutism,Staring,Mannerisms,Verbigeration,Waxy flexibility | 95.3 | 93.3 | 97.1 |
| 6 | Immobility/stupor,Mutism,Staring,Stereotypy,Mannerisms,Waxy flexibility | 95.3 | 93.3 | 97.1 |
| 6 | Immobility/stupor,Mutism,Staring,Stereotypy,Verbigeration,Waxy flexibility | 95.3 | 93.3 | 97.1 |
| 6 | Immobility/stupor,Mutism,Staring,Stereotypy,Verbigeration,Withdrawal | 95.3 | 93.3 | 97.1 |
| 6 | Immobility/stupor,Mutism,Stereotypy,Verbigeration,Negativism,Withdrawal | 95.3 | 93.3 | 97.1 |
| 6 | Immobility/stupor,Mutism,Stereotypy,Verbigeration,Rigidity,Withdrawal | 95.3 | 93.3 | 97.1 |
| 6 | Immobility/stupor,Posturing/catalepsy,Echopraxia/echolalia,Stereotypy,Mannerisms,Negativism | 95.3 | 93.3 | 97.1 |
| 6 | Immobility/stupor,Posturing/catalepsy,Grimacing,Echopraxia/echolalia,Mannerisms,Negativism | 95.3 | 93.3 | 97.1 |
| 6 | Immobility/stupor,Posturing/catalepsy,Grimacing,Echopraxia/echolalia,Rigidity,Negativism | 95.3 | 93.3 | 97.1 |
| 6 | Immobility/stupor,Posturing/catalepsy,Grimacing,Stereotypy,Mannerisms,Negativism | 95.3 | 93.3 | 97.1 |
| 6 | Immobility/stupor,Posturing/catalepsy,Mannerisms,Negativism,Waxy flexibility,Withdrawal | 95.3 | 93.3 | 97.1 |
| 6 | Immobility/stupor,Posturing/catalepsy,Stereotypy,Mannerisms,Verbigeration,Negativism | 95.3 | 93.3 | 97.1 |
| 6 | Immobility/stupor,Staring,Echopraxia/echolalia,Stereotypy,Mannerisms,Waxy flexibility | 95.3 | 93.3 | 97.1 |
| 6 | Immobility/stupor,Staring,Grimacing,Echopraxia/echolalia,Mannerisms,Verbigeration | 95.3 | 93.3 | 97.1 |
| 6 | Immobility/stupor,Staring,Grimacing,Echopraxia/echolalia,Negativism,Waxy flexibility | 95.3 | 93.3 | 97.1 |
| 6 | Immobility/stupor,Staring,Grimacing,Mannerisms,Verbigeration,Waxy flexibility | 95.3 | 93.3 | 97.1 |
| 6 | Immobility/stupor,Staring,Grimacing,Negativism,Waxy flexibility,Withdrawal | 95.3 | 93.3 | 97.1 |
| 6 | Immobility/stupor,Staring,Grimacing,Rigidity,Waxy flexibility,Withdrawal | 95.3 | 93.3 | 97.1 |
| 6 | Immobility/stupor,Staring,Grimacing,Stereotypy,Verbigeration,Waxy flexibility | 95.3 | 93.3 | 97.1 |
| 6 | Immobility/stupor,Staring,Posturing/catalepsy,Echopraxia/echolalia,Mannerisms,Verbigeration | 95.3 | 93.3 | 97.1 |
| 6 | Immobility/stupor,Staring,Posturing/catalepsy,Grimacing,Echopraxia/echolalia,Rigidity | 95.3 | 93.3 | 97.1 |
| 6 | Immobility/stupor,Staring,Posturing/catalepsy,Grimacing,Echopraxia/echolalia,Withdrawal | 95.3 | 93.3 | 97.1 |
| 6 | Immobility/stupor,Staring,Posturing/catalepsy,Rigidity,Negativism,Waxy flexibility | 95.3 | 93.3 | 97.1 |
| 6 | Immobility/stupor,Staring,Posturing/catalepsy,Stereotypy,Verbigeration,Withdrawal | 95.3 | 93.3 | 97.1 |
| 6 | Immobility/stupor,Staring,Posturing/catalepsy,Verbigeration,Negativism,Waxy flexibility | 95.3 | 93.3 | 97.1 |
| 6 | Immobility/stupor,Staring,Stereotypy,Mannerisms,Verbigeration,Waxy flexibility | 95.3 | 93.3 | 97.1 |
| 6 | Mutism,Staring,Posturing/catalepsy,Grimacing,Echopraxia/echolalia,Waxy flexibility | 95.3 | 93.3 | 97.1 |
| 6 | Mutism,Staring,Posturing/catalepsy,Grimacing,Stereotypy,Waxy flexibility | 95.3 | 93.3 | 97.1 |
| 6 | Staring,Echopraxia/echolalia,Mannerisms,Verbigeration,Negativism,Waxy flexibility | 95.3 | 93.3 | 97.1 |
| 6 | Staring,Grimacing,Echopraxia/echolalia,Mannerisms,Verbigeration,Rigidity | 95.3 | 93.3 | 97.1 |
| 6 | Staring,Grimacing,Echopraxia/echolalia,Stereotypy,Verbigeration,Rigidity | 95.3 | 93.3 | 97.1 |
| 6 | Staring,Grimacing,Echopraxia/echolalia,Stereotypy,Verbigeration,Withdrawal | 95.3 | 93.3 | 97.1 |
| 6 | Staring,Grimacing,Echopraxia/echolalia,Stereotypy,Waxy flexibility,Withdrawal | 95.3 | 93.3 | 97.1 |
| 6 | Staring,Grimacing,Stereotypy,Mannerisms,Verbigeration,Negativism | 95.3 | 93.3 | 97.1 |
| 6 | Staring,Grimacing,Stereotypy,Verbigeration,Waxy flexibility,Withdrawal | 95.3 | 93.3 | 97.1 |
| 6 | Staring,Grimacing,Verbigeration,Rigidity,Negativism,Waxy flexibility | 95.3 | 93.3 | 97.1 |
| 6 | Staring,Posturing/catalepsy,Echopraxia/echolalia,Mannerisms,Verbigeration,Withdrawal | 95.3 | 93.3 | 97.1 |
| 6 | Staring,Posturing/catalepsy,Echopraxia/echolalia,Rigidity,Waxy flexibility,Withdrawal | 95.3 | 93.3 | 97.1 |
| 6 | Staring,Posturing/catalepsy,Echopraxia/echolalia,Stereotypy,Verbigeration,Negativism | 95.3 | 93.3 | 97.1 |
| 6 | Staring,Posturing/catalepsy,Grimacing,Echopraxia/echolalia,Stereotypy,Negativism | 95.3 | 93.3 | 97.1 |
| 6 | Staring,Posturing/catalepsy,Mannerisms,Verbigeration,Negativism,Waxy flexibility | 95.3 | 93.3 | 97.1 |
| 6 | Staring,Posturing/catalepsy,Mannerisms,Verbigeration,Waxy flexibility,Withdrawal | 95.3 | 93.3 | 97.1 |
| 6 | Staring,Posturing/catalepsy,Stereotypy,Verbigeration,Negativism,Waxy flexibility | 95.3 | 93.3 | 97.1 |
| 6 | Staring,Posturing/catalepsy,Verbigeration,Rigidity,Waxy flexibility,Withdrawal | 95.3 | 93.3 | 97.1 |
| 6 | Staring,Stereotypy,Mannerisms,Verbigeration,Waxy flexibility,Withdrawal | 95.3 | 93.3 | 97.1 |
| 6 | Excitement,Immobility/stupor,Echopraxia/echolalia,Mannerisms,Verbigeration,Withdrawal | 95.1 | 93.0 | 97.1 |
| 6 | Excitement,Immobility/stupor,Echopraxia/echolalia,Stereotypy,Mannerisms,Negativism | 95.1 | 93.0 | 97.1 |
| 6 | Excitement,Immobility/stupor,Echopraxia/echolalia,Stereotypy,Mannerisms,Withdrawal | 95.1 | 93.0 | 97.1 |
| 6 | Excitement,Immobility/stupor,Grimacing,Echopraxia/echolalia,Verbigeration,Withdrawal | 95.1 | 93.0 | 97.1 |
| 6 | Excitement,Immobility/stupor,Grimacing,Mannerisms,Verbigeration,Negativism | 95.1 | 93.0 | 97.1 |
| 6 | Excitement,Immobility/stupor,Mutism,Echopraxia/echolalia,Mannerisms,Negativism | 95.1 | 93.0 | 96.9 |
| 6 | Excitement,Immobility/stupor,Mutism,Echopraxia/echolalia,Mannerisms,Withdrawal | 95.1 | 93.0 | 96.9 |
| 6 | Excitement,Immobility/stupor,Mutism,Echopraxia/echolalia,Stereotypy,Verbigeration | 95.1 | 93.0 | 96.9 |
| 6 | Excitement,Immobility/stupor,Mutism,Mannerisms,Verbigeration,Withdrawal | 95.1 | 93.0 | 96.9 |
| 6 | Excitement,Immobility/stupor,Mutism,Posturing/catalepsy,Stereotypy,Mannerisms | 95.1 | 93.0 | 97.1 |
| 6 | Excitement,Immobility/stupor,Mutism,Stereotypy,Mannerisms,Withdrawal | 95.1 | 93.0 | 97.1 |
| 6 | Excitement,Immobility/stupor,Posturing/catalepsy,Echopraxia/echolalia,Verbigeration,Waxy flexibility | 95.1 | 93.0 | 97.1 |
| 6 | Excitement,Immobility/stupor,Posturing/catalepsy,Grimacing,Mannerisms,Verbigeration | 95.1 | 93.0 | 97.1 |
| 6 | Excitement,Immobility/stupor,Posturing/catalepsy,Grimacing,Verbigeration,Waxy flexibility | 95.1 | 93.0 | 97.1 |
| 6 | Excitement,Immobility/stupor,Posturing/catalepsy,Stereotypy,Waxy flexibility,Withdrawal | 95.1 | 93.0 | 96.9 |
| 6 | Excitement,Immobility/stupor,Stereotypy,Mannerisms,Negativism,Waxy flexibility | 95.1 | 93.0 | 97.1 |
| 6 | Excitement,Immobility/stupor,Stereotypy,Verbigeration,Waxy flexibility,Withdrawal | 95.1 | 93.0 | 97.1 |
| 6 | Excitement,Mutism,Posturing/catalepsy,Verbigeration,Rigidity,Negativism | 95.1 | 93.0 | 96.9 |
| 6 | Excitement,Staring,Grimacing,Echopraxia/echolalia,Verbigeration,Waxy flexibility | 95.1 | 93.0 | 96.9 |
| 6 | Excitement,Staring,Grimacing,Mannerisms,Verbigeration,Negativism | 95.1 | 93.0 | 96.9 |
| 6 | Excitement,Staring,Grimacing,Mannerisms,Verbigeration,Waxy flexibility | 95.1 | 93.0 | 96.9 |
| 6 | Excitement,Staring,Grimacing,Stereotypy,Mannerisms,Negativism | 95.1 | 93.0 | 97.1 |
| 6 | Excitement,Staring,Grimacing,Stereotypy,Mannerisms,Waxy flexibility | 95.1 | 93.0 | 97.1 |
| 6 | Excitement,Staring,Grimacing,Stereotypy,Verbigeration,Waxy flexibility | 95.1 | 93.0 | 97.1 |
| 6 | Excitement,Staring,Posturing/catalepsy,Echopraxia/echolalia,Mannerisms,Verbigeration | 95.1 | 93.0 | 96.9 |
| 6 | Excitement,Staring,Posturing/catalepsy,Echopraxia/echolalia,Mannerisms,Waxy flexibility | 95.1 | 93.0 | 96.9 |
| 6 | Excitement,Staring,Posturing/catalepsy,Echopraxia/echolalia,Stereotypy,Mannerisms | 95.1 | 93.0 | 97.1 |
| 6 | Excitement,Staring,Posturing/catalepsy,Echopraxia/echolalia,Stereotypy,Waxy flexibility | 95.1 | 93.0 | 96.9 |
| 6 | Excitement,Staring,Posturing/catalepsy,Echopraxia/echolalia,Verbigeration,Waxy flexibility | 95.1 | 92.8 | 96.9 |
| 6 | Excitement,Staring,Posturing/catalepsy,Stereotypy,Mannerisms,Verbigeration | 95.1 | 93.0 | 96.9 |
| 6 | Excitement,Staring,Posturing/catalepsy,Stereotypy,Verbigeration,Waxy flexibility | 95.1 | 93.0 | 96.9 |
| 6 | Immobility/stupor,Echopraxia/echolalia,Mannerisms,Negativism,Waxy flexibility,Withdrawal | 95.1 | 93.0 | 96.9 |
| 6 | Immobility/stupor,Echopraxia/echolalia,Stereotypy,Negativism,Waxy flexibility,Withdrawal | 95.1 | 93.0 | 96.9 |
| 6 | Immobility/stupor,Echopraxia/echolalia,Verbigeration,Rigidity,Negativism,Waxy flexibility | 95.1 | 93.0 | 96.9 |
| 6 | Immobility/stupor,Grimacing,Echopraxia/echolalia,Rigidity,Negativism,Waxy flexibility | 95.1 | 93.0 | 97.1 |
| 6 | Immobility/stupor,Grimacing,Echopraxia/echolalia,Stereotypy,Mannerisms,Negativism | 95.1 | 93.0 | 97.1 |
| 6 | Immobility/stupor,Grimacing,Mannerisms,Verbigeration,Rigidity,Withdrawal | 95.1 | 93.0 | 96.9 |
| 6 | Immobility/stupor,Mutism,Echopraxia/echolalia,Stereotypy,Mannerisms,Negativism | 95.1 | 93.0 | 97.1 |
| 6 | Immobility/stupor,Mutism,Echopraxia/echolalia,Stereotypy,Negativism,Withdrawal | 95.1 | 93.0 | 96.9 |
| 6 | Immobility/stupor,Mutism,Echopraxia/echolalia,Stereotypy,Rigidity,Withdrawal | 95.1 | 93.0 | 97.1 |
| 6 | Immobility/stupor,Mutism,Grimacing,Echopraxia/echolalia,Mannerisms,Rigidity | 95.1 | 93.0 | 97.1 |
| 6 | Immobility/stupor,Mutism,Grimacing,Mannerisms,Verbigeration,Rigidity | 95.1 | 93.0 | 96.9 |
| 6 | Immobility/stupor,Mutism,Grimacing,Rigidity,Negativism,Withdrawal | 95.1 | 93.0 | 97.1 |
| 6 | Immobility/stupor,Mutism,Grimacing,Stereotypy,Mannerisms,Rigidity | 95.1 | 93.0 | 97.1 |
| 6 | Immobility/stupor,Mutism,Mannerisms,Rigidity,Waxy flexibility,Withdrawal | 95.1 | 93.0 | 96.9 |
| 6 | Immobility/stupor,Mutism,Posturing/catalepsy,Grimacing,Mannerisms,Rigidity | 95.1 | 92.8 | 96.9 |
| 6 | Immobility/stupor,Mutism,Posturing/catalepsy,Grimacing,Negativism,Withdrawal | 95.1 | 93.0 | 96.9 |
| 6 | Immobility/stupor,Mutism,Staring,Grimacing,Verbigeration,Waxy flexibility | 95.1 | 93.0 | 96.9 |
| 6 | Immobility/stupor,Mutism,Staring,Posturing/catalepsy,Grimacing,Echopraxia/echolalia | 95.1 | 93.0 | 97.1 |
| 6 | Immobility/stupor,Mutism,Staring,Posturing/catalepsy,Grimacing,Stereotypy | 95.1 | 93.0 | 97.1 |
| 6 | Immobility/stupor,Mutism,Staring,Posturing/catalepsy,Negativism,Waxy flexibility | 95.1 | 93.0 | 96.9 |
| 6 | Immobility/stupor,Mutism,Staring,Stereotypy,Waxy flexibility,Withdrawal | 95.1 | 93.0 | 96.9 |
| 6 | Immobility/stupor,Mutism,Stereotypy,Mannerisms,Negativism,Waxy flexibility | 95.1 | 93.0 | 96.9 |
| 6 | Immobility/stupor,Mutism,Stereotypy,Mannerisms,Verbigeration,Negativism | 95.1 | 93.0 | 97.1 |
| 6 | Immobility/stupor,Posturing/catalepsy,Echopraxia/echolalia,Mannerisms,Rigidity,Withdrawal | 95.1 | 92.8 | 97.1 |
| 6 | Immobility/stupor,Posturing/catalepsy,Grimacing,Echopraxia/echolalia,Negativism,Withdrawal | 95.1 | 93.0 | 97.1 |
| 6 | Immobility/stupor,Posturing/catalepsy,Grimacing,Mannerisms,Verbigeration,Negativism | 95.1 | 93.0 | 96.9 |
| 6 | Immobility/stupor,Posturing/catalepsy,Grimacing,Verbigeration,Negativism,Withdrawal | 95.1 | 92.8 | 96.9 |
| 6 | Immobility/stupor,Posturing/catalepsy,Rigidity,Negativism,Waxy flexibility,Withdrawal | 95.1 | 93.0 | 97.1 |
| 6 | Immobility/stupor,Posturing/catalepsy,Stereotypy,Mannerisms,Rigidity,Withdrawal | 95.1 | 93.0 | 97.1 |
| 6 | Immobility/stupor,Posturing/catalepsy,Verbigeration,Rigidity,Negativism,Waxy flexibility | 95.1 | 93.0 | 96.9 |
| 6 | Immobility/stupor,Staring,Echopraxia/echolalia,Mannerisms,Verbigeration,Withdrawal | 95.1 | 93.0 | 96.9 |
| 6 | Immobility/stupor,Staring,Echopraxia/echolalia,Stereotypy,Mannerisms,Verbigeration | 95.1 | 93.0 | 97.1 |
| 6 | Immobility/stupor,Staring,Echopraxia/echolalia,Stereotypy,Rigidity,Waxy flexibility | 95.1 | 93.0 | 97.1 |
| 6 | Immobility/stupor,Staring,Echopraxia/echolalia,Verbigeration,Rigidity,Waxy flexibility | 95.1 | 93.0 | 96.9 |
| 6 | Immobility/stupor,Staring,Grimacing,Echopraxia/echolalia,Stereotypy,Verbigeration | 95.1 | 93.0 | 96.9 |
| 6 | Immobility/stupor,Staring,Mannerisms,Verbigeration,Waxy flexibility,Withdrawal | 95.1 | 93.0 | 97.1 |
| 6 | Immobility/stupor,Staring,Posturing/catalepsy,Grimacing,Echopraxia/echolalia,Stereotypy | 95.1 | 93.0 | 97.1 |
| 6 | Immobility/stupor,Staring,Posturing/catalepsy,Grimacing,Verbigeration,Withdrawal | 95.1 | 93.0 | 97.1 |
| 6 | Immobility/stupor,Staring,Posturing/catalepsy,Rigidity,Waxy flexibility,Withdrawal | 95.1 | 93.0 | 96.9 |
| 6 | Staring,Echopraxia/echolalia,Verbigeration,Rigidity,Waxy flexibility,Withdrawal | 95.1 | 93.0 | 96.9 |
| 6 | Staring,Grimacing,Echopraxia/echolalia,Rigidity,Negativism,Waxy flexibility | 95.1 | 92.8 | 96.9 |
| 6 | Staring,Posturing/catalepsy,Echopraxia/echolalia,Mannerisms,Verbigeration,Rigidity | 95.1 | 93.0 | 96.9 |
| 6 | Staring,Posturing/catalepsy,Echopraxia/echolalia,Rigidity,Negativism,Waxy flexibility | 95.1 | 93.0 | 96.9 |
| 6 | Staring,Posturing/catalepsy,Echopraxia/echolalia,Stereotypy,Mannerisms,Rigidity | 95.1 | 93.0 | 96.9 |
| 6 | Staring,Posturing/catalepsy,Echopraxia/echolalia,Stereotypy,Negativism,Waxy flexibility | 95.1 | 93.0 | 97.1 |
| 6 | Staring,Posturing/catalepsy,Grimacing,Echopraxia/echolalia,Stereotypy,Rigidity | 95.1 | 93.0 | 96.9 |
| 6 | Staring,Posturing/catalepsy,Grimacing,Echopraxia/echolalia,Verbigeration,Negativism | 95.1 | 93.0 | 96.9 |
| 6 | Staring,Posturing/catalepsy,Grimacing,Mannerisms,Negativism,Waxy flexibility | 95.1 | 93.0 | 97.1 |
| 6 | Staring,Posturing/catalepsy,Grimacing,Rigidity,Waxy flexibility,Withdrawal | 95.1 | 93.0 | 96.9 |
| 6 | Staring,Stereotypy,Mannerisms,Verbigeration,Rigidity,Waxy flexibility | 95.1 | 93.0 | 97.1 |
| 6 | Immobility/stupor,Mutism,Grimacing,Mannerisms,Negativism,Withdrawal | 95.0 | 92.8 | 96.9 |
| 6 | Immobility/stupor,Mutism,Staring,Echopraxia/echolalia,Rigidity,Waxy flexibility | 95.0 | 92.8 | 96.9 |
| 6 | Immobility/stupor,Posturing/catalepsy,Echopraxia/echolalia,Verbigeration,Negativism,Withdrawal | 95.0 | 93.0 | 97.1 |
| 6 | Immobility/stupor,Posturing/catalepsy,Grimacing,Stereotypy,Verbigeration,Negativism | 95.0 | 93.0 | 96.9 |
| 6 | Excitement,Immobility/stupor,Grimacing,Echopraxia/echolalia,Mannerisms,Withdrawal | 94.9 | 92.8 | 96.9 |
| 6 | Excitement,Immobility/stupor,Mutism,Grimacing,Mannerisms,Negativism | 94.9 | 92.8 | 96.6 |
| 6 | Excitement,Mutism,Posturing/catalepsy,Echopraxia/echolalia,Rigidity,Negativism | 94.9 | 92.8 | 96.9 |
| 6 | Immobility/stupor,Grimacing,Rigidity,Negativism,Waxy flexibility,Withdrawal | 94.9 | 92.6 | 96.9 |
| 6 | Immobility/stupor,Mutism,Posturing/catalepsy,Grimacing,Mannerisms,Withdrawal | 94.9 | 92.8 | 96.9 |
| 6 | Immobility/stupor,Mutism,Posturing/catalepsy,Mannerisms,Verbigeration,Rigidity | 94.9 | 92.6 | 96.9 |
| 6 | Immobility/stupor,Mutism,Staring,Echopraxia/echolalia,Stereotypy,Verbigeration | 94.9 | 92.6 | 96.9 |
| 6 | Immobility/stupor,Mutism,Staring,Echopraxia/echolalia,Verbigeration,Withdrawal | 94.9 | 92.8 | 96.9 |
| 6 | Immobility/stupor,Posturing/catalepsy,Stereotypy,Mannerisms,Negativism,Waxy flexibility | 94.9 | 92.8 | 96.9 |
| 6 | Immobility/stupor,Staring,Posturing/catalepsy,Stereotypy,Rigidity,Waxy flexibility | 94.9 | 92.8 | 96.9 |
| 6 | Immobility/stupor,Staring,Stereotypy,Verbigeration,Waxy flexibility,Withdrawal | 94.9 | 92.6 | 96.9 |
| 6 | Immobility/stupor,Stereotypy,Mannerisms,Verbigeration,Rigidity,Withdrawal | 94.9 | 92.6 | 96.9 |
| 6 | Staring,Grimacing,Echopraxia/echolalia,Stereotypy,Rigidity,Waxy flexibility | 94.9 | 92.8 | 96.9 |
| 6 | Staring,Posturing/catalepsy,Grimacing,Stereotypy,Waxy flexibility,Withdrawal | 94.9 | 92.8 | 96.9 |
| 6 | Excitement,Immobility/stupor,Echopraxia/echolalia,Mannerisms,Verbigeration,Negativism | 94.8 | 92.6 | 96.9 |
| 6 | Excitement,Immobility/stupor,Grimacing,Echopraxia/echolalia,Mannerisms,Negativism | 94.8 | 92.8 | 96.9 |
| 6 | Excitement,Immobility/stupor,Mannerisms,Verbigeration,Waxy flexibility,Withdrawal | 94.8 | 92.6 | 96.9 |
| 6 | Excitement,Immobility/stupor,Mutism,Echopraxia/echolalia,Mannerisms,Waxy flexibility | 94.8 | 92.6 | 96.9 |
| 6 | Excitement,Immobility/stupor,Mutism,Grimacing,Echopraxia/echolalia,Stereotypy | 94.8 | 92.6 | 96.9 |
| 6 | Excitement,Immobility/stupor,Mutism,Grimacing,Echopraxia/echolalia,Verbigeration | 94.8 | 92.6 | 96.9 |
| 6 | Excitement,Immobility/stupor,Mutism,Grimacing,Mannerisms,Waxy flexibility | 94.8 | 92.8 | 96.9 |
| 6 | Excitement,Immobility/stupor,Mutism,Grimacing,Stereotypy,Verbigeration | 94.8 | 92.6 | 96.9 |
| 6 | Excitement,Immobility/stupor,Posturing/catalepsy,Echopraxia/echolalia,Mannerisms,Verbigeration | 94.8 | 92.8 | 96.9 |
| 6 | Excitement,Immobility/stupor,Posturing/catalepsy,Grimacing,Echopraxia/echolalia,Stereotypy | 94.8 | 92.6 | 96.9 |
| 6 | Excitement,Immobility/stupor,Posturing/catalepsy,Grimacing,Mannerisms,Waxy flexibility | 94.8 | 92.6 | 96.9 |
| 6 | Excitement,Immobility/stupor,Stereotypy,Mannerisms,Verbigeration,Withdrawal | 94.8 | 92.6 | 96.9 |
| 6 | Excitement,Mutism,Echopraxia/echolalia,Rigidity,Negativism,Withdrawal | 94.8 | 92.6 | 96.9 |
| 6 | Excitement,Mutism,Posturing/catalepsy,Rigidity,Waxy flexibility,Withdrawal | 94.8 | 92.6 | 96.9 |
| 6 | Excitement,Mutism,Stereotypy,Rigidity,Negativism,Withdrawal | 94.8 | 92.6 | 96.9 |
| 6 | Excitement,Mutism,Verbigeration,Rigidity,Negativism,Withdrawal | 94.8 | 92.6 | 96.9 |
| 6 | Excitement,Staring,Echopraxia/echolalia,Stereotypy,Mannerisms,Waxy flexibility | 94.8 | 92.8 | 96.9 |
| 6 | Excitement,Staring,Grimacing,Echopraxia/echolalia,Stereotypy,Mannerisms | 94.8 | 92.6 | 96.9 |
| 6 | Excitement,Staring,Posturing/catalepsy,Echopraxia/echolalia,Stereotypy,Verbigeration | 94.8 | 92.6 | 96.9 |
| 6 | Excitement,Staring,Posturing/catalepsy,Stereotypy,Mannerisms,Waxy flexibility | 94.8 | 92.6 | 96.9 |
| 6 | Immobility/stupor,Echopraxia/echolalia,Stereotypy,Mannerisms,Verbigeration,Negativism | 94.8 | 92.6 | 96.9 |
| 6 | Immobility/stupor,Grimacing,Echopraxia/echolalia,Mannerisms,Negativism,Withdrawal | 94.8 | 92.8 | 96.9 |
| 6 | Immobility/stupor,Grimacing,Mannerisms,Rigidity,Waxy flexibility,Withdrawal | 94.8 | 92.8 | 96.9 |
| 6 | Immobility/stupor,Grimacing,Mannerisms,Verbigeration,Negativism,Withdrawal | 94.8 | 92.6 | 96.9 |
| 6 | Immobility/stupor,Grimacing,Stereotypy,Mannerisms,Negativism,Waxy flexibility | 94.8 | 92.6 | 96.9 |
| 6 | Immobility/stupor,Mutism,Echopraxia/echolalia,Mannerisms,Negativism,Waxy flexibility | 94.8 | 92.6 | 96.9 |
| 6 | Immobility/stupor,Mutism,Echopraxia/echolalia,Rigidity,Negativism,Waxy flexibility | 94.8 | 92.6 | 96.9 |
| 6 | Immobility/stupor,Mutism,Grimacing,Echopraxia/echolalia,Rigidity,Negativism | 94.8 | 92.8 | 96.9 |
| 6 | Immobility/stupor,Mutism,Grimacing,Echopraxia/echolalia,Rigidity,Withdrawal | 94.8 | 92.6 | 96.9 |
| 6 | Immobility/stupor,Mutism,Posturing/catalepsy,Echopraxia/echolalia,Mannerisms,Rigidity | 94.8 | 92.6 | 96.9 |
| 6 | Immobility/stupor,Mutism,Posturing/catalepsy,Echopraxia/echolalia,Verbigeration,Negativism | 94.8 | 92.6 | 96.9 |
| 6 | Immobility/stupor,Mutism,Posturing/catalepsy,Grimacing,Verbigeration,Negativism | 94.8 | 92.6 | 96.9 |
| 6 | Immobility/stupor,Mutism,Posturing/catalepsy,Stereotypy,Verbigeration,Rigidity | 94.8 | 92.6 | 96.9 |
| 6 | Immobility/stupor,Mutism,Rigidity,Negativism,Waxy flexibility,Withdrawal | 94.8 | 92.6 | 96.9 |
| 6 | Immobility/stupor,Mutism,Staring,Echopraxia/echolalia,Stereotypy,Waxy flexibility | 94.8 | 92.6 | 96.9 |
| 6 | Immobility/stupor,Mutism,Staring,Echopraxia/echolalia,Verbigeration,Waxy flexibility | 94.8 | 92.6 | 96.9 |
| 6 | Immobility/stupor,Mutism,Staring,Echopraxia/echolalia,Waxy flexibility,Withdrawal | 94.8 | 92.6 | 96.9 |
| 6 | Immobility/stupor,Mutism,Staring,Posturing/catalepsy,Mannerisms,Waxy flexibility | 94.8 | 92.6 | 96.9 |
| 6 | Immobility/stupor,Mutism,Staring,Posturing/catalepsy,Waxy flexibility,Withdrawal | 94.8 | 92.6 | 96.9 |
| 6 | Immobility/stupor,Mutism,Staring,Verbigeration,Waxy flexibility,Withdrawal | 94.8 | 92.6 | 96.9 |
| 6 | Immobility/stupor,Mutism,Stereotypy,Mannerisms,Verbigeration,Rigidity | 94.8 | 92.8 | 96.9 |
| 6 | Immobility/stupor,Posturing/catalepsy,Echopraxia/echolalia,Mannerisms,Verbigeration,Negativism | 94.8 | 92.6 | 96.9 |
| 6 | Immobility/stupor,Posturing/catalepsy,Grimacing,Rigidity,Negativism,Waxy flexibility | 94.8 | 92.6 | 96.9 |
| 6 | Immobility/stupor,Posturing/catalepsy,Grimacing,Stereotypy,Rigidity,Withdrawal | 94.8 | 92.6 | 96.9 |
| 6 | Immobility/stupor,Posturing/catalepsy,Mannerisms,Verbigeration,Rigidity,Withdrawal | 94.8 | 92.8 | 96.9 |
| 6 | Immobility/stupor,Posturing/catalepsy,Stereotypy,Negativism,Waxy flexibility,Withdrawal | 94.8 | 92.6 | 96.9 |
| 6 | Immobility/stupor,Staring,Echopraxia/echolalia,Mannerisms,Verbigeration,Waxy flexibility | 94.8 | 92.6 | 96.9 |
| 6 | Immobility/stupor,Staring,Echopraxia/echolalia,Stereotypy,Waxy flexibility,Withdrawal | 94.8 | 92.6 | 96.9 |
| 6 | Immobility/stupor,Staring,Grimacing,Echopraxia/echolalia,Rigidity,Waxy flexibility | 94.8 | 92.8 | 96.9 |
| 6 | Immobility/stupor,Staring,Grimacing,Echopraxia/echolalia,Stereotypy,Waxy flexibility | 94.8 | 92.6 | 96.6 |
| 6 | Immobility/stupor,Staring,Grimacing,Stereotypy,Waxy flexibility,Withdrawal | 94.8 | 92.6 | 96.9 |
| 6 | Immobility/stupor,Staring,Posturing/catalepsy,Echopraxia/echolalia,Stereotypy,Verbigeration | 94.8 | 92.6 | 96.9 |
| 6 | Immobility/stupor,Staring,Posturing/catalepsy,Grimacing,Echopraxia/echolalia,Verbigeration | 94.8 | 92.6 | 96.9 |
| 6 | Immobility/stupor,Staring,Posturing/catalepsy,Grimacing,Negativism,Waxy flexibility | 94.8 | 92.8 | 96.9 |
| 6 | Immobility/stupor,Staring,Posturing/catalepsy,Mannerisms,Verbigeration,Waxy flexibility | 94.8 | 92.8 | 96.9 |
| 6 | Immobility/stupor,Staring,Posturing/catalepsy,Stereotypy,Verbigeration,Waxy flexibility | 94.8 | 92.6 | 96.9 |
| 6 | Immobility/stupor,Stereotypy,Mannerisms,Verbigeration,Negativism,Waxy flexibility | 94.8 | 92.6 | 96.9 |
| 6 | Immobility/stupor,Stereotypy,Verbigeration,Negativism,Waxy flexibility,Withdrawal | 94.8 | 92.6 | 96.9 |
| 6 | Mutism,Posturing/catalepsy,Mannerisms,Rigidity,Negativism,Withdrawal | 94.8 | 92.6 | 96.9 |
| 6 | Staring,Echopraxia/echolalia,Stereotypy,Mannerisms,Verbigeration,Withdrawal | 94.8 | 92.6 | 96.9 |
| 6 | Staring,Echopraxia/echolalia,Stereotypy,Verbigeration,Negativism,Waxy flexibility | 94.8 | 92.6 | 96.9 |
| 6 | Staring,Grimacing,Echopraxia/echolalia,Mannerisms,Verbigeration,Negativism | 94.8 | 92.6 | 96.9 |
| 6 | Staring,Grimacing,Echopraxia/echolalia,Stereotypy,Verbigeration,Negativism | 94.8 | 92.6 | 96.9 |
| 6 | Staring,Grimacing,Mannerisms,Verbigeration,Negativism,Waxy flexibility | 94.8 | 92.8 | 96.9 |
| 6 | Staring,Grimacing,Stereotypy,Verbigeration,Negativism,Waxy flexibility | 94.8 | 92.8 | 96.9 |
| 6 | Staring,Posturing/catalepsy,Grimacing,Echopraxia/echolalia,Stereotypy,Mannerisms | 94.8 | 92.6 | 96.9 |
| 6 | Staring,Posturing/catalepsy,Grimacing,Echopraxia/echolalia,Verbigeration,Rigidity | 94.8 | 92.6 | 96.9 |
| 6 | Staring,Posturing/catalepsy,Grimacing,Echopraxia/echolalia,Verbigeration,Withdrawal | 94.8 | 92.6 | 96.9 |
| 6 | Staring,Posturing/catalepsy,Grimacing,Mannerisms,Rigidity,Waxy flexibility | 94.8 | 92.6 | 96.9 |
| 6 | Excitement,Immobility/stupor,Grimacing,Echopraxia/echolalia,Mannerisms,Waxy flexibility | 94.6 | 92.4 | 96.6 |
| 6 | Excitement,Immobility/stupor,Grimacing,Echopraxia/echolalia,Stereotypy,Waxy flexibility | 94.6 | 92.4 | 96.6 |
| 6 | Excitement,Immobility/stupor,Grimacing,Echopraxia/echolalia,Verbigeration,Waxy flexibility | 94.6 | 92.4 | 96.6 |
| 6 | Excitement,Immobility/stupor,Grimacing,Mannerisms,Waxy flexibility,Withdrawal | 94.6 | 92.4 | 96.6 |
| 6 | Excitement,Immobility/stupor,Grimacing,Stereotypy,Verbigeration,Withdrawal | 94.6 | 92.4 | 96.6 |
| 6 | Excitement,Immobility/stupor,Grimacing,Stereotypy,Waxy flexibility,Withdrawal | 94.6 | 92.4 | 96.6 |
| 6 | Excitement,Immobility/stupor,Mutism,Grimacing,Mannerisms,Withdrawal | 94.6 | 92.4 | 96.6 |
| 6 | Excitement,Immobility/stupor,Mutism,Mannerisms,Verbigeration,Waxy flexibility | 94.6 | 92.4 | 96.6 |
| 6 | Excitement,Immobility/stupor,Posturing/catalepsy,Echopraxia/echolalia,Stereotypy,Verbigeration | 94.6 | 92.4 | 96.6 |
| 6 | Excitement,Immobility/stupor,Posturing/catalepsy,Grimacing,Stereotypy,Verbigeration | 94.6 | 92.4 | 96.6 |
| 6 | Excitement,Mutism,Grimacing,Rigidity,Negativism,Withdrawal | 94.6 | 92.4 | 96.6 |
| 6 | Excitement,Mutism,Posturing/catalepsy,Echopraxia/echolalia,Rigidity,Withdrawal | 94.6 | 92.4 | 96.6 |
| 6 | Excitement,Mutism,Posturing/catalepsy,Verbigeration,Rigidity,Withdrawal | 94.6 | 92.4 | 96.6 |
| 6 | Excitement,Mutism,Rigidity,Negativism,Waxy flexibility,Withdrawal | 94.6 | 92.4 | 96.6 |
| 6 | Excitement,Staring,Echopraxia/echolalia,Stereotypy,Verbigeration,Waxy flexibility | 94.6 | 92.4 | 96.6 |
| 6 | Excitement,Staring,Stereotypy,Mannerisms,Verbigeration,Waxy flexibility | 94.6 | 92.4 | 96.6 |
| 6 | Immobility/stupor,Echopraxia/echolalia,Mannerisms,Verbigeration,Rigidity,Withdrawal | 94.6 | 92.6 | 96.6 |
| 6 | Immobility/stupor,Echopraxia/echolalia,Stereotypy,Verbigeration,Rigidity,Withdrawal | 94.6 | 92.4 | 96.6 |
| 6 | Immobility/stupor,Grimacing,Echopraxia/echolalia,Mannerisms,Negativism,Waxy flexibility | 94.6 | 92.4 | 96.6 |
| 6 | Immobility/stupor,Grimacing,Echopraxia/echolalia,Verbigeration,Rigidity,Withdrawal | 94.6 | 92.6 | 96.6 |
| 6 | Immobility/stupor,Grimacing,Stereotypy,Mannerisms,Verbigeration,Negativism | 94.6 | 92.4 | 96.6 |
| 6 | Immobility/stupor,Mannerisms,Verbigeration,Negativism,Waxy flexibility,Withdrawal | 94.6 | 92.4 | 96.6 |
| 6 | Immobility/stupor,Mutism,Echopraxia/echolalia,Mannerisms,Verbigeration,Negativism | 94.6 | 92.4 | 96.6 |
| 6 | Immobility/stupor,Mutism,Echopraxia/echolalia,Mannerisms,Verbigeration,Rigidity | 94.6 | 92.4 | 96.6 |
| 6 | Immobility/stupor,Mutism,Echopraxia/echolalia,Stereotypy,Mannerisms,Rigidity | 94.6 | 92.4 | 96.6 |
| 6 | Immobility/stupor,Mutism,Echopraxia/echolalia,Verbigeration,Rigidity,Withdrawal | 94.6 | 92.4 | 96.6 |
| 6 | Immobility/stupor,Mutism,Grimacing,Echopraxia/echolalia,Mannerisms,Negativism | 94.6 | 92.4 | 96.6 |
| 6 | Immobility/stupor,Mutism,Grimacing,Mannerisms,Rigidity,Waxy flexibility | 94.6 | 92.4 | 96.6 |
| 6 | Immobility/stupor,Mutism,Grimacing,Stereotypy,Mannerisms,Negativism | 94.6 | 92.4 | 96.6 |
| 6 | Immobility/stupor,Mutism,Grimacing,Stereotypy,Negativism,Withdrawal | 94.6 | 92.4 | 96.6 |
| 6 | Immobility/stupor,Mutism,Posturing/catalepsy,Echopraxia/echolalia,Stereotypy,Negativism | 94.6 | 92.4 | 96.6 |
| 6 | Immobility/stupor,Mutism,Posturing/catalepsy,Grimacing,Rigidity,Negativism | 94.6 | 92.4 | 96.6 |
| 6 | Immobility/stupor,Mutism,Posturing/catalepsy,Grimacing,Stereotypy,Negativism | 94.6 | 92.4 | 96.6 |
| 6 | Immobility/stupor,Mutism,Posturing/catalepsy,Grimacing,Verbigeration,Rigidity | 94.6 | 92.4 | 96.6 |
| 6 | Immobility/stupor,Mutism,Posturing/catalepsy,Mannerisms,Negativism,Waxy flexibility | 94.6 | 92.4 | 96.6 |
| 6 | Immobility/stupor,Mutism,Posturing/catalepsy,Stereotypy,Mannerisms,Rigidity | 94.6 | 92.4 | 96.6 |
| 6 | Immobility/stupor,Mutism,Posturing/catalepsy,Stereotypy,Rigidity,Withdrawal | 94.6 | 92.4 | 96.6 |
| 6 | Immobility/stupor,Mutism,Staring,Grimacing,Negativism,Waxy flexibility | 94.6 | 92.4 | 96.6 |
| 6 | Immobility/stupor,Mutism,Staring,Grimacing,Stereotypy,Waxy flexibility | 94.6 | 92.4 | 96.6 |
| 6 | Immobility/stupor,Mutism,Staring,Posturing/catalepsy,Grimacing,Rigidity | 94.6 | 92.4 | 96.6 |
| 6 | Immobility/stupor,Mutism,Staring,Posturing/catalepsy,Verbigeration,Waxy flexibility | 94.6 | 92.4 | 96.6 |
| 6 | Immobility/stupor,Posturing/catalepsy,Echopraxia/echolalia,Rigidity,Negativism,Waxy flexibility | 94.6 | 92.4 | 96.6 |
| 6 | Immobility/stupor,Posturing/catalepsy,Echopraxia/echolalia,Stereotypy,Rigidity,Withdrawal | 94.6 | 92.4 | 96.6 |
| 6 | Immobility/stupor,Posturing/catalepsy,Echopraxia/echolalia,Stereotypy,Verbigeration,Negativism | 94.6 | 92.4 | 96.6 |
| 6 | Immobility/stupor,Posturing/catalepsy,Grimacing,Echopraxia/echolalia,Rigidity,Withdrawal | 94.6 | 92.4 | 96.6 |
| 6 | Immobility/stupor,Posturing/catalepsy,Grimacing,Echopraxia/echolalia,Stereotypy,Negativism | 94.6 | 92.4 | 96.6 |
| 6 | Immobility/stupor,Posturing/catalepsy,Grimacing,Echopraxia/echolalia,Verbigeration,Negativism | 94.6 | 92.4 | 96.6 |
| 6 | Immobility/stupor,Posturing/catalepsy,Stereotypy,Verbigeration,Negativism,Waxy flexibility | 94.6 | 92.4 | 96.6 |
| 6 | Immobility/stupor,Posturing/catalepsy,Verbigeration,Negativism,Waxy flexibility,Withdrawal | 94.6 | 92.4 | 96.6 |
| 6 | Immobility/stupor,Staring,Echopraxia/echolalia,Stereotypy,Verbigeration,Withdrawal | 94.6 | 92.4 | 96.6 |
| 6 | Immobility/stupor,Staring,Grimacing,Echopraxia/echolalia,Verbigeration,Waxy flexibility | 94.6 | 92.4 | 96.6 |
| 6 | Immobility/stupor,Staring,Grimacing,Echopraxia/echolalia,Verbigeration,Withdrawal | 94.6 | 92.4 | 96.6 |
| 6 | Immobility/stupor,Staring,Posturing/catalepsy,Echopraxia/echolalia,Mannerisms,Waxy flexibility | 94.6 | 92.4 | 96.6 |
| 6 | Immobility/stupor,Staring,Posturing/catalepsy,Echopraxia/echolalia,Verbigeration,Withdrawal | 94.6 | 92.4 | 96.6 |
| 6 | Immobility/stupor,Staring,Posturing/catalepsy,Grimacing,Mannerisms,Waxy flexibility | 94.6 | 92.4 | 96.6 |
| 6 | Immobility/stupor,Staring,Posturing/catalepsy,Stereotypy,Mannerisms,Waxy flexibility | 94.6 | 92.4 | 96.6 |
| 6 | Immobility/stupor,Staring,Posturing/catalepsy,Stereotypy,Waxy flexibility,Withdrawal | 94.6 | 92.4 | 96.6 |
| 6 | Immobility/stupor,Staring,Posturing/catalepsy,Verbigeration,Rigidity,Waxy flexibility | 94.6 | 92.4 | 96.6 |
| 6 | Immobility/stupor,Stereotypy,Mannerisms,Rigidity,Waxy flexibility,Withdrawal | 94.6 | 92.4 | 96.6 |
| 6 | Mutism,Staring,Posturing/catalepsy,Grimacing,Rigidity,Waxy flexibility | 94.6 | 92.4 | 96.6 |
| 6 | Staring,Echopraxia/echolalia,Mannerisms,Verbigeration,Waxy flexibility,Withdrawal | 94.6 | 92.4 | 96.6 |
| 6 | Staring,Echopraxia/echolalia,Stereotypy,Mannerisms,Rigidity,Waxy flexibility | 94.6 | 92.6 | 96.6 |
| 6 | Staring,Echopraxia/echolalia,Stereotypy,Mannerisms,Verbigeration,Rigidity | 94.6 | 92.4 | 96.6 |
| 6 | Staring,Grimacing,Echopraxia/echolalia,Stereotypy,Mannerisms,Waxy flexibility | 94.6 | 92.4 | 96.6 |
| 6 | Staring,Grimacing,Echopraxia/echolalia,Stereotypy,Negativism,Waxy flexibility | 94.6 | 92.4 | 96.6 |
| 6 | Staring,Grimacing,Echopraxia/echolalia,Verbigeration,Rigidity,Waxy flexibility | 94.6 | 92.4 | 96.6 |
| 6 | Staring,Posturing/catalepsy,Echopraxia/echolalia,Stereotypy,Verbigeration,Rigidity | 94.6 | 92.4 | 96.6 |
| 6 | Staring,Posturing/catalepsy,Echopraxia/echolalia,Stereotypy,Waxy flexibility,Withdrawal | 94.6 | 92.4 | 96.6 |
| 6 | Staring,Posturing/catalepsy,Grimacing,Echopraxia/echolalia,Waxy flexibility,Withdrawal | 94.6 | 92.4 | 96.6 |
| 6 | Staring,Posturing/catalepsy,Grimacing,Stereotypy,Mannerisms,Verbigeration | 94.6 | 92.4 | 96.6 |
| 6 | Staring,Posturing/catalepsy,Stereotypy,Verbigeration,Waxy flexibility,Withdrawal | 94.6 | 92.4 | 96.6 |
| 6 | Staring,Posturing/catalepsy,Verbigeration,Rigidity,Negativism,Waxy flexibility | 94.6 | 92.4 | 96.6 |
| 6 | Excitement,Immobility/stupor,Echopraxia/echolalia,Stereotypy,Verbigeration,Waxy flexibility | 94.4 | 92.2 | 96.4 |
| 6 | Excitement,Immobility/stupor,Grimacing,Mannerisms,Verbigeration,Withdrawal | 94.4 | 92.2 | 96.4 |
| 6 | Excitement,Immobility/stupor,Grimacing,Stereotypy,Mannerisms,Negativism | 94.4 | 92.2 | 96.4 |
| 6 | Excitement,Immobility/stupor,Grimacing,Verbigeration,Waxy flexibility,Withdrawal | 94.4 | 92.2 | 96.4 |
| 6 | Excitement,Immobility/stupor,Mutism,Stereotypy,Mannerisms,Waxy flexibility | 94.4 | 92.2 | 96.4 |
| 6 | Excitement,Immobility/stupor,Posturing/catalepsy,Echopraxia/echolalia,Mannerisms,Waxy flexibility | 94.4 | 92.2 | 96.4 |
| 6 | Excitement,Immobility/stupor,Posturing/catalepsy,Grimacing,Stereotypy,Waxy flexibility | 94.4 | 92.2 | 96.4 |
| 6 | Excitement,Immobility/stupor,Posturing/catalepsy,Mannerisms,Verbigeration,Waxy flexibility | 94.4 | 92.2 | 96.4 |
| 6 | Excitement,Immobility/stupor,Posturing/catalepsy,Stereotypy,Verbigeration,Waxy flexibility | 94.4 | 92.2 | 96.4 |
| 6 | Excitement,Immobility/stupor,Stereotypy,Mannerisms,Waxy flexibility,Withdrawal | 94.4 | 92.2 | 96.4 |
| 6 | Excitement,Mutism,Posturing/catalepsy,Grimacing,Rigidity,Negativism | 94.4 | 92.2 | 96.4 |
| 6 | Excitement,Mutism,Posturing/catalepsy,Grimacing,Rigidity,Withdrawal | 94.4 | 92.2 | 96.4 |
| 6 | Excitement,Mutism,Posturing/catalepsy,Mannerisms,Rigidity,Withdrawal | 94.4 | 92.2 | 96.4 |
| 6 | Excitement,Mutism,Stereotypy,Rigidity,Waxy flexibility,Withdrawal | 94.4 | 92.2 | 96.4 |
| 6 | Excitement,Staring,Echopraxia/echolalia,Mannerisms,Verbigeration,Waxy flexibility | 94.4 | 92.2 | 96.4 |
| 6 | Excitement,Staring,Grimacing,Echopraxia/echolalia,Stereotypy,Verbigeration | 94.4 | 92.2 | 96.4 |
| 6 | Immobility/stupor,Echopraxia/echolalia,Mannerisms,Rigidity,Waxy flexibility,Withdrawal | 94.4 | 92.2 | 96.4 |
| 6 | Immobility/stupor,Grimacing,Echopraxia/echolalia,Stereotypy,Mannerisms,Rigidity | 94.4 | 92.2 | 96.4 |
| 6 | Immobility/stupor,Grimacing,Echopraxia/echolalia,Stereotypy,Negativism,Waxy flexibility | 94.4 | 92.2 | 96.4 |
| 6 | Immobility/stupor,Grimacing,Echopraxia/echolalia,Stereotypy,Verbigeration,Negativism | 94.4 | 92.2 | 96.4 |
| 6 | Immobility/stupor,Grimacing,Echopraxia/echolalia,Verbigeration,Negativism,Withdrawal | 94.4 | 92.2 | 96.4 |
| 6 | Immobility/stupor,Grimacing,Mannerisms,Negativism,Waxy flexibility,Withdrawal | 94.4 | 92.2 | 96.4 |
| 6 | Immobility/stupor,Grimacing,Stereotypy,Rigidity,Waxy flexibility,Withdrawal | 94.4 | 92.2 | 96.4 |
| 6 | Immobility/stupor,Mutism,Echopraxia/echolalia,Verbigeration,Negativism,Withdrawal | 94.4 | 92.2 | 96.4 |
| 6 | Immobility/stupor,Mutism,Grimacing,Echopraxia/echolalia,Verbigeration,Rigidity | 94.4 | 92.2 | 96.4 |
| 6 | Immobility/stupor,Mutism,Grimacing,Mannerisms,Negativism,Waxy flexibility | 94.4 | 92.2 | 96.4 |
| 6 | Immobility/stupor,Mutism,Grimacing,Rigidity,Negativism,Waxy flexibility | 94.4 | 92.2 | 96.4 |
| 6 | Immobility/stupor,Mutism,Grimacing,Stereotypy,Verbigeration,Negativism | 94.4 | 92.2 | 96.4 |
| 6 | Immobility/stupor,Mutism,Grimacing,Verbigeration,Negativism,Withdrawal | 94.4 | 92.2 | 96.4 |
| 6 | Immobility/stupor,Mutism,Posturing/catalepsy,Echopraxia/echolalia,Mannerisms,Withdrawal | 94.4 | 92.2 | 96.4 |
| 6 | Immobility/stupor,Mutism,Posturing/catalepsy,Echopraxia/echolalia,Rigidity,Withdrawal | 94.4 | 92.2 | 96.4 |
| 6 | Immobility/stupor,Mutism,Posturing/catalepsy,Grimacing,Echopraxia/echolalia,Negativism | 94.4 | 92.2 | 96.4 |
| 6 | Immobility/stupor,Mutism,Posturing/catalepsy,Grimacing,Rigidity,Withdrawal | 94.4 | 92.2 | 96.4 |
| 6 | Immobility/stupor,Mutism,Posturing/catalepsy,Grimacing,Stereotypy,Rigidity | 94.4 | 92.2 | 96.4 |
| 6 | Immobility/stupor,Mutism,Posturing/catalepsy,Verbigeration,Rigidity,Withdrawal | 94.4 | 92.2 | 96.4 |
| 6 | Immobility/stupor,Mutism,Staring,Grimacing,Echopraxia/echolalia,Waxy flexibility | 94.4 | 92.2 | 96.4 |
| 6 | Immobility/stupor,Mutism,Staring,Grimacing,Rigidity,Waxy flexibility | 94.4 | 92.2 | 96.4 |
| 6 | Immobility/stupor,Mutism,Staring,Grimacing,Waxy flexibility,Withdrawal | 94.4 | 92.2 | 96.4 |
| 6 | Immobility/stupor,Mutism,Staring,Posturing/catalepsy,Echopraxia/echolalia,Waxy flexibility | 94.4 | 92.2 | 96.4 |
| 6 | Immobility/stupor,Mutism,Staring,Posturing/catalepsy,Stereotypy,Waxy flexibility | 94.4 | 92.2 | 96.4 |
| 6 | Immobility/stupor,Mutism,Stereotypy,Negativism,Waxy flexibility,Withdrawal | 94.4 | 92.2 | 96.4 |
| 6 | Immobility/stupor,Mutism,Stereotypy,Rigidity,Waxy flexibility,Withdrawal | 94.4 | 92.2 | 96.4 |
| 6 | Immobility/stupor,Mutism,Verbigeration,Rigidity,Waxy flexibility,Withdrawal | 94.4 | 92.2 | 96.4 |
| 6 | Immobility/stupor,Posturing/catalepsy,Echopraxia/echolalia,Mannerisms,Negativism,Waxy flexibility | 94.4 | 92.2 | 96.4 |
| 6 | Immobility/stupor,Posturing/catalepsy,Grimacing,Echopraxia/echolalia,Mannerisms,Rigidity | 94.4 | 92.2 | 96.4 |
| 6 | Immobility/stupor,Posturing/catalepsy,Grimacing,Mannerisms,Negativism,Waxy flexibility | 94.4 | 92.2 | 96.4 |
| 6 | Immobility/stupor,Posturing/catalepsy,Grimacing,Stereotypy,Mannerisms,Rigidity | 94.4 | 92.2 | 96.4 |
| 6 | Immobility/stupor,Posturing/catalepsy,Grimacing,Stereotypy,Verbigeration,Rigidity | 94.4 | 92.2 | 96.4 |
| 6 | Immobility/stupor,Posturing/catalepsy,Grimacing,Verbigeration,Rigidity,Withdrawal | 94.4 | 92.2 | 96.4 |
| 6 | Immobility/stupor,Posturing/catalepsy,Mannerisms,Verbigeration,Negativism,Waxy flexibility | 94.4 | 92.2 | 96.4 |
| 6 | Immobility/stupor,Posturing/catalepsy,Stereotypy,Verbigeration,Rigidity,Withdrawal | 94.4 | 92.2 | 96.4 |
| 6 | Immobility/stupor,Staring,Echopraxia/echolalia,Stereotypy,Verbigeration,Waxy flexibility | 94.4 | 92.2 | 96.4 |
| 6 | Immobility/stupor,Staring,Grimacing,Echopraxia/echolalia,Waxy flexibility,Withdrawal | 94.4 | 92.2 | 96.4 |
| 6 | Immobility/stupor,Staring,Grimacing,Verbigeration,Waxy flexibility,Withdrawal | 94.4 | 92.2 | 96.4 |
| 6 | Immobility/stupor,Staring,Posturing/catalepsy,Echopraxia/echolalia,Waxy flexibility,Withdrawal | 94.4 | 92.2 | 96.4 |
| 6 | Mutism,Stereotypy,Mannerisms,Rigidity,Negativism,Withdrawal | 94.4 | 92.2 | 96.4 |
| 6 | Mutism,Stereotypy,Verbigeration,Rigidity,Negativism,Withdrawal | 94.4 | 92.2 | 96.4 |
| 6 | Staring,Grimacing,Echopraxia/echolalia,Verbigeration,Negativism,Waxy flexibility | 94.4 | 92.2 | 96.4 |
| 6 | Staring,Grimacing,Echopraxia/echolalia,Verbigeration,Waxy flexibility,Withdrawal | 94.4 | 92.2 | 96.4 |
| 6 | Staring,Posturing/catalepsy,Echopraxia/echolalia,Stereotypy,Verbigeration,Withdrawal | 94.4 | 92.2 | 96.4 |
| 6 | Staring,Posturing/catalepsy,Echopraxia/echolalia,Verbigeration,Negativism,Waxy flexibility | 94.4 | 92.2 | 96.4 |
| 6 | Staring,Posturing/catalepsy,Grimacing,Stereotypy,Negativism,Waxy flexibility | 94.4 | 92.2 | 96.4 |
| 6 | Staring,Posturing/catalepsy,Grimacing,Verbigeration,Waxy flexibility,Withdrawal | 94.4 | 92.2 | 96.4 |
| 6 | Staring,Posturing/catalepsy,Mannerisms,Verbigeration,Rigidity,Waxy flexibility | 94.4 | 92.2 | 96.4 |
| 6 | Staring,Posturing/catalepsy,Stereotypy,Mannerisms,Rigidity,Waxy flexibility | 94.4 | 92.2 | 96.4 |
| 6 | Staring,Posturing/catalepsy,Stereotypy,Verbigeration,Rigidity,Waxy flexibility | 94.4 | 92.2 | 96.4 |
| 6 | Excitement,Immobility/stupor,Mutism,Echopraxia/echolalia,Mannerisms,Verbigeration | 94.2 | 91.9 | 96.2 |
| 6 | Excitement,Immobility/stupor,Mutism,Echopraxia/echolalia,Stereotypy,Mannerisms | 94.2 | 91.9 | 96.2 |
| 6 | Excitement,Immobility/stupor,Mutism,Grimacing,Mannerisms,Verbigeration | 94.2 | 91.9 | 96.2 |
| 6 | Excitement,Immobility/stupor,Mutism,Stereotypy,Mannerisms,Verbigeration | 94.2 | 91.9 | 96.2 |
| 6 | Excitement,Immobility/stupor,Posturing/catalepsy,Grimacing,Stereotypy,Mannerisms | 94.2 | 91.9 | 96.2 |
| 6 | Excitement,Immobility/stupor,Posturing/catalepsy,Stereotypy,Mannerisms,Verbigeration | 94.2 | 91.9 | 96.2 |
| 6 | Excitement,Mutism,Mannerisms,Rigidity,Negativism,Withdrawal | 94.2 | 91.9 | 96.2 |
| 6 | Excitement,Mutism,Posturing/catalepsy,Mannerisms,Rigidity,Negativism | 94.2 | 91.9 | 96.2 |
| 6 | Excitement,Mutism,Posturing/catalepsy,Stereotypy,Rigidity,Negativism | 94.2 | 91.9 | 96.2 |
| 6 | Excitement,Mutism,Posturing/catalepsy,Stereotypy,Rigidity,Withdrawal | 94.2 | 91.9 | 96.2 |
| 6 | Excitement,Mutism,Posturing/catalepsy,Verbigeration,Negativism,Withdrawal | 94.2 | 91.9 | 96.2 |
| 6 | Excitement,Staring,Grimacing,Stereotypy,Mannerisms,Verbigeration | 94.2 | 91.9 | 96.2 |
| 6 | Immobility/stupor,Echopraxia/echolalia,Stereotypy,Rigidity,Waxy flexibility,Withdrawal | 94.2 | 91.9 | 96.2 |
| 6 | Immobility/stupor,Echopraxia/echolalia,Stereotypy,Verbigeration,Negativism,Waxy flexibility | 94.2 | 91.9 | 96.2 |
| 6 | Immobility/stupor,Echopraxia/echolalia,Verbigeration,Negativism,Waxy flexibility,Withdrawal | 94.2 | 91.9 | 96.2 |
| 6 | Immobility/stupor,Grimacing,Echopraxia/echolalia,Mannerisms,Verbigeration,Negativism | 94.2 | 91.7 | 96.2 |
| 6 | Immobility/stupor,Grimacing,Echopraxia/echolalia,Rigidity,Waxy flexibility,Withdrawal | 94.2 | 91.9 | 96.2 |
| 6 | Immobility/stupor,Grimacing,Stereotypy,Mannerisms,Rigidity,Waxy flexibility | 94.2 | 91.9 | 96.2 |
| 6 | Immobility/stupor,Grimacing,Stereotypy,Mannerisms,Verbigeration,Rigidity | 94.2 | 91.9 | 96.2 |
| 6 | Immobility/stupor,Grimacing,Stereotypy,Negativism,Waxy flexibility,Withdrawal | 94.2 | 91.9 | 96.2 |
| 6 | Immobility/stupor,Grimacing,Stereotypy,Verbigeration,Negativism,Waxy flexibility | 94.2 | 91.9 | 96.2 |
| 6 | Immobility/stupor,Mutism,Echopraxia/echolalia,Mannerisms,Rigidity,Waxy flexibility | 94.2 | 91.9 | 96.2 |
| 6 | Immobility/stupor,Mutism,Echopraxia/echolalia,Negativism,Waxy flexibility,Withdrawal | 94.2 | 91.9 | 96.2 |
| 6 | Immobility/stupor,Mutism,Echopraxia/echolalia,Stereotypy,Verbigeration,Negativism | 94.2 | 91.9 | 96.2 |
| 6 | Immobility/stupor,Mutism,Echopraxia/echolalia,Stereotypy,Verbigeration,Rigidity | 94.2 | 91.9 | 96.2 |
| 6 | Immobility/stupor,Mutism,Grimacing,Echopraxia/echolalia,Stereotypy,Rigidity | 94.2 | 91.9 | 96.2 |
| 6 | Immobility/stupor,Mutism,Grimacing,Echopraxia/echolalia,Verbigeration,Negativism | 94.2 | 91.9 | 96.2 |
| 6 | Immobility/stupor,Mutism,Grimacing,Mannerisms,Verbigeration,Negativism | 94.2 | 91.9 | 96.2 |
| 6 | Immobility/stupor,Mutism,Grimacing,Verbigeration,Rigidity,Waxy flexibility | 94.2 | 91.9 | 96.2 |
| 6 | Immobility/stupor,Mutism,Mannerisms,Verbigeration,Negativism,Waxy flexibility | 94.2 | 91.9 | 96.2 |
| 6 | Immobility/stupor,Mutism,Mannerisms,Verbigeration,Rigidity,Waxy flexibility | 94.2 | 91.9 | 96.2 |
| 6 | Immobility/stupor,Mutism,Posturing/catalepsy,Echopraxia/echolalia,Stereotypy,Rigidity | 94.2 | 91.9 | 96.2 |
| 6 | Immobility/stupor,Mutism,Posturing/catalepsy,Echopraxia/echolalia,Verbigeration,Rigidity | 94.2 | 91.9 | 96.2 |
| 6 | Immobility/stupor,Mutism,Posturing/catalepsy,Grimacing,Echopraxia/echolalia,Withdrawal | 94.2 | 91.9 | 96.2 |
| 6 | Immobility/stupor,Mutism,Posturing/catalepsy,Negativism,Waxy flexibility,Withdrawal | 94.2 | 91.9 | 96.2 |
| 6 | Immobility/stupor,Mutism,Posturing/catalepsy,Rigidity,Negativism,Waxy flexibility | 94.2 | 91.9 | 96.2 |
| 6 | Immobility/stupor,Mutism,Posturing/catalepsy,Stereotypy,Negativism,Waxy flexibility | 94.2 | 91.9 | 96.2 |
| 6 | Immobility/stupor,Mutism,Stereotypy,Mannerisms,Rigidity,Waxy flexibility | 94.2 | 91.9 | 96.2 |
| 6 | Immobility/stupor,Mutism,Stereotypy,Verbigeration,Negativism,Waxy flexibility | 94.2 | 91.9 | 96.2 |
| 6 | Immobility/stupor,Mutism,Stereotypy,Verbigeration,Rigidity,Waxy flexibility | 94.2 | 91.9 | 96.2 |
| 6 | Immobility/stupor,Mutism,Verbigeration,Negativism,Waxy flexibility,Withdrawal | 94.2 | 91.9 | 96.2 |
| 6 | Immobility/stupor,Posturing/catalepsy,Echopraxia/echolalia,Negativism,Waxy flexibility,Withdrawal | 94.2 | 91.9 | 96.2 |
| 6 | Immobility/stupor,Posturing/catalepsy,Echopraxia/echolalia,Stereotypy,Negativism,Waxy flexibility | 94.2 | 91.9 | 96.2 |
| 6 | Immobility/stupor,Posturing/catalepsy,Grimacing,Echopraxia/echolalia,Mannerisms,Withdrawal | 94.2 | 91.9 | 96.2 |
| 6 | Immobility/stupor,Posturing/catalepsy,Grimacing,Mannerisms,Verbigeration,Rigidity | 94.2 | 91.9 | 96.2 |
| 6 | Immobility/stupor,Posturing/catalepsy,Grimacing,Stereotypy,Negativism,Waxy flexibility | 94.2 | 91.9 | 96.2 |
| 6 | Immobility/stupor,Posturing/catalepsy,Grimacing,Verbigeration,Negativism,Waxy flexibility | 94.2 | 91.9 | 96.2 |
| 6 | Immobility/stupor,Posturing/catalepsy,Mannerisms,Rigidity,Waxy flexibility,Withdrawal | 94.2 | 91.9 | 96.2 |
| 6 | Immobility/stupor,Staring,Posturing/catalepsy,Echopraxia/echolalia,Rigidity,Waxy flexibility | 94.2 | 91.9 | 96.2 |
| 6 | Immobility/stupor,Staring,Posturing/catalepsy,Echopraxia/echolalia,Stereotypy,Waxy flexibility | 94.2 | 91.9 | 96.2 |
| 6 | Immobility/stupor,Staring,Posturing/catalepsy,Grimacing,Verbigeration,Waxy flexibility | 94.2 | 91.9 | 96.2 |
| 6 | Immobility/stupor,Staring,Posturing/catalepsy,Verbigeration,Waxy flexibility,Withdrawal | 94.2 | 91.9 | 96.2 |
| 6 | Immobility/stupor,Stereotypy,Verbigeration,Rigidity,Waxy flexibility,Withdrawal | 94.2 | 91.9 | 96.2 |
| 6 | Mutism,Grimacing,Mannerisms,Rigidity,Negativism,Withdrawal | 94.2 | 91.9 | 96.2 |
| 6 | Mutism,Mannerisms,Verbigeration,Rigidity,Negativism,Withdrawal | 94.2 | 91.9 | 96.2 |
| 6 | Staring,Echopraxia/echolalia,Mannerisms,Verbigeration,Rigidity,Waxy flexibility | 94.2 | 91.9 | 96.2 |
| 6 | Staring,Echopraxia/echolalia,Stereotypy,Verbigeration,Waxy flexibility,Withdrawal | 94.2 | 91.9 | 96.2 |
| 6 | Staring,Grimacing,Stereotypy,Mannerisms,Verbigeration,Waxy flexibility | 94.2 | 91.9 | 96.2 |
| 6 | Staring,Posturing/catalepsy,Echopraxia/echolalia,Mannerisms,Rigidity,Waxy flexibility | 94.2 | 91.9 | 96.2 |
| 6 | Staring,Posturing/catalepsy,Grimacing,Echopraxia/echolalia,Mannerisms,Verbigeration | 94.2 | 91.9 | 96.2 |
| 6 | Staring,Posturing/catalepsy,Grimacing,Echopraxia/echolalia,Negativism,Waxy flexibility | 94.2 | 91.9 | 96.4 |
| 6 | Staring,Posturing/catalepsy,Grimacing,Rigidity,Negativism,Waxy flexibility | 94.2 | 91.9 | 96.2 |
| 6 | Staring,Posturing/catalepsy,Grimacing,Stereotypy,Rigidity,Waxy flexibility | 94.2 | 91.9 | 96.2 |
| 6 | Staring,Posturing/catalepsy,Grimacing,Verbigeration,Negativism,Waxy flexibility | 94.2 | 91.9 | 96.2 |
| 6 | Excitement,Immobility/stupor,Posturing/catalepsy,Echopraxia/echolalia,Stereotypy,Waxy flexibility | 94.1 | 91.9 | 96.2 |
| 6 | Mutism,Echopraxia/echolalia,Mannerisms,Rigidity,Negativism,Withdrawal | 94.1 | 91.9 | 96.2 |
| 6 | Mutism,Mannerisms,Rigidity,Negativism,Waxy flexibility,Withdrawal | 94.1 | 91.9 | 96.2 |
| 6 | Excitement,Immobility/stupor,Echopraxia/echolalia,Mannerisms,Verbigeration,Waxy flexibility | 94.0 | 91.7 | 96.0 |
| 6 | Excitement,Immobility/stupor,Grimacing,Mannerisms,Verbigeration,Waxy flexibility | 94.0 | 91.7 | 96.0 |
| 6 | Excitement,Immobility/stupor,Grimacing,Stereotypy,Verbigeration,Waxy flexibility | 94.0 | 91.7 | 96.2 |
| 6 | Excitement,Immobility/stupor,Mutism,Grimacing,Echopraxia/echolalia,Mannerisms | 94.0 | 91.7 | 96.0 |
| 6 | Excitement,Mutism,Verbigeration,Rigidity,Waxy flexibility,Withdrawal | 94.0 | 91.7 | 96.2 |
| 6 | Immobility/stupor,Echopraxia/echolalia,Mannerisms,Verbigeration,Negativism,Waxy flexibility | 94.0 | 91.7 | 96.2 |
| 6 | Immobility/stupor,Grimacing,Echopraxia/echolalia,Mannerisms,Rigidity,Waxy flexibility | 94.0 | 91.7 | 96.2 |
| 6 | Immobility/stupor,Grimacing,Echopraxia/echolalia,Mannerisms,Verbigeration,Rigidity | 94.0 | 91.7 | 96.2 |
| 6 | Immobility/stupor,Grimacing,Echopraxia/echolalia,Verbigeration,Negativism,Waxy flexibility | 94.0 | 91.7 | 96.0 |
| 6 | Immobility/stupor,Mutism,Echopraxia/echolalia,Rigidity,Waxy flexibility,Withdrawal | 94.0 | 91.7 | 96.0 |
| 6 | Immobility/stupor,Mutism,Grimacing,Rigidity,Waxy flexibility,Withdrawal | 94.0 | 91.7 | 96.0 |
| 6 | Immobility/stupor,Mutism,Posturing/catalepsy,Grimacing,Stereotypy,Withdrawal | 94.0 | 91.7 | 96.0 |
| 6 | Immobility/stupor,Mutism,Posturing/catalepsy,Mannerisms,Verbigeration,Withdrawal | 94.0 | 91.7 | 96.0 |
| 6 | Immobility/stupor,Posturing/catalepsy,Echopraxia/echolalia,Verbigeration,Rigidity,Withdrawal | 94.0 | 91.7 | 96.2 |
| 6 | Immobility/stupor,Staring,Posturing/catalepsy,Grimacing,Stereotypy,Waxy flexibility | 94.0 | 91.7 | 96.2 |
| 6 | Immobility/stupor,Staring,Posturing/catalepsy,Grimacing,Waxy flexibility,Withdrawal | 94.0 | 91.7 | 96.0 |
| 6 | Mutism,Posturing/catalepsy,Mannerisms,Verbigeration,Rigidity,Negativism | 94.0 | 91.7 | 96.0 |
| 6 | Staring,Posturing/catalepsy,Grimacing,Stereotypy,Mannerisms,Waxy flexibility | 94.0 | 91.7 | 96.0 |
| 6 | Staring,Posturing/catalepsy,Grimacing,Verbigeration,Rigidity,Waxy flexibility | 94.0 | 91.7 | 96.2 |
| 6 | Excitement,Immobility/stupor,Grimacing,Stereotypy,Mannerisms,Withdrawal | 93.9 | 91.7 | 96.2 |
| 6 | Excitement,Immobility/stupor,Posturing/catalepsy,Echopraxia/echolalia,Stereotypy,Mannerisms | 93.9 | 91.7 | 96.0 |
| 6 | Excitement,Mutism,Grimacing,Rigidity,Waxy flexibility,Withdrawal | 93.9 | 91.7 | 96.2 |
| 6 | Excitement,Mutism,Posturing/catalepsy,Echopraxia/echolalia,Negativism,Withdrawal | 93.9 | 91.7 | 96.0 |
| 6 | Excitement,Mutism,Posturing/catalepsy,Rigidity,Negativism,Waxy flexibility | 93.9 | 91.7 | 96.0 |
| 6 | Excitement,Mutism,Posturing/catalepsy,Stereotypy,Negativism,Withdrawal | 93.9 | 91.7 | 96.2 |
| 6 | Excitement,Mutism,Stereotypy,Verbigeration,Rigidity,Negativism | 93.9 | 91.7 | 96.0 |
| 6 | Excitement,Staring,Grimacing,Echopraxia/echolalia,Mannerisms,Verbigeration | 93.9 | 91.7 | 96.0 |
| 6 | Immobility/stupor,Grimacing,Echopraxia/echolalia,Stereotypy,Mannerisms,Withdrawal | 93.9 | 91.7 | 96.0 |
| 6 | Immobility/stupor,Grimacing,Echopraxia/echolalia,Stereotypy,Verbigeration,Rigidity | 93.9 | 91.7 | 96.0 |
| 6 | Immobility/stupor,Grimacing,Stereotypy,Verbigeration,Rigidity,Waxy flexibility | 93.9 | 91.7 | 96.0 |
| 6 | Immobility/stupor,Mannerisms,Verbigeration,Rigidity,Waxy flexibility,Withdrawal | 93.9 | 91.7 | 96.0 |
| 6 | Immobility/stupor,Mutism,Echopraxia/echolalia,Stereotypy,Negativism,Waxy flexibility | 93.9 | 91.7 | 96.2 |
| 6 | Immobility/stupor,Mutism,Echopraxia/echolalia,Verbigeration,Negativism,Waxy flexibility | 93.9 | 91.7 | 96.0 |
| 6 | Immobility/stupor,Mutism,Grimacing,Echopraxia/echolalia,Mannerisms,Withdrawal | 93.9 | 91.7 | 96.0 |
| 6 | Immobility/stupor,Mutism,Grimacing,Echopraxia/echolalia,Negativism,Withdrawal | 93.9 | 91.7 | 96.0 |
| 6 | Immobility/stupor,Mutism,Grimacing,Echopraxia/echolalia,Stereotypy,Negativism | 93.9 | 91.7 | 96.0 |
| 6 | Immobility/stupor,Mutism,Grimacing,Echopraxia/echolalia,Stereotypy,Withdrawal | 93.9 | 91.7 | 96.0 |
| 6 | Immobility/stupor,Mutism,Grimacing,Stereotypy,Rigidity,Waxy flexibility | 93.9 | 91.7 | 96.0 |
| 6 | Immobility/stupor,Mutism,Posturing/catalepsy,Verbigeration,Negativism,Waxy flexibility | 93.9 | 91.7 | 96.2 |
| 6 | Immobility/stupor,Mutism,Staring,Posturing/catalepsy,Rigidity,Waxy flexibility | 93.9 | 91.7 | 96.0 |
| 6 | Immobility/stupor,Posturing/catalepsy,Grimacing,Echopraxia/echolalia,Stereotypy,Withdrawal | 93.9 | 91.7 | 96.2 |
| 6 | Immobility/stupor,Posturing/catalepsy,Grimacing,Negativism,Waxy flexibility,Withdrawal | 93.9 | 91.7 | 96.2 |
| 6 | Immobility/stupor,Posturing/catalepsy,Grimacing,Stereotypy,Mannerisms,Withdrawal | 93.9 | 91.5 | 96.0 |
| 6 | Immobility/stupor,Staring,Echopraxia/echolalia,Verbigeration,Waxy flexibility,Withdrawal | 93.9 | 91.7 | 96.0 |
| 6 | Mutism,Posturing/catalepsy,Echopraxia/echolalia,Mannerisms,Rigidity,Negativism | 93.9 | 91.7 | 96.0 |
| 6 | Mutism,Posturing/catalepsy,Stereotypy,Rigidity,Negativism,Withdrawal | 93.9 | 91.7 | 96.0 |
| 6 | Mutism,Posturing/catalepsy,Verbigeration,Rigidity,Negativism,Withdrawal | 93.9 | 91.7 | 96.0 |
| 6 | Staring,Echopraxia/echolalia,Stereotypy,Verbigeration,Rigidity,Waxy flexibility | 93.9 | 91.7 | 96.2 |
| 6 | Immobility/stupor,Mutism,Posturing/catalepsy,Grimacing,Echopraxia/echolalia,Rigidity | 93.8 | 91.3 | 96.0 |
| 6 | Excitement,Immobility/stupor,Echopraxia/echolalia,Stereotypy,Mannerisms,Waxy flexibility | 93.7 | 91.5 | 96.0 |
| 6 | Excitement,Immobility/stupor,Mutism,Grimacing,Stereotypy,Mannerisms | 93.7 | 91.3 | 95.7 |
| 6 | Excitement,Mutism,Echopraxia/echolalia,Rigidity,Waxy flexibility,Withdrawal | 93.7 | 91.5 | 96.0 |
| 6 | Excitement,Mutism,Echopraxia/echolalia,Stereotypy,Rigidity,Negativism | 93.7 | 91.5 | 96.0 |
| 6 | Excitement,Mutism,Posturing/catalepsy,Echopraxia/echolalia,Rigidity,Waxy flexibility | 93.7 | 91.5 | 96.0 |
| 6 | Excitement,Mutism,Posturing/catalepsy,Echopraxia/echolalia,Verbigeration,Rigidity | 93.7 | 91.5 | 96.0 |
| 6 | Excitement,Mutism,Posturing/catalepsy,Grimacing,Negativism,Withdrawal | 93.7 | 91.5 | 96.0 |
| 6 | Excitement,Mutism,Posturing/catalepsy,Mannerisms,Negativism,Withdrawal | 93.7 | 91.5 | 96.0 |
| 6 | Excitement,Mutism,Posturing/catalepsy,Verbigeration,Rigidity,Waxy flexibility | 93.7 | 91.5 | 96.0 |
| 6 | Excitement,Mutism,Stereotypy,Rigidity,Negativism,Waxy flexibility | 93.7 | 91.3 | 96.0 |
| 6 | Immobility/stupor,Grimacing,Echopraxia/echolalia,Negativism,Waxy flexibility,Withdrawal | 93.7 | 91.3 | 96.0 |
| 6 | Immobility/stupor,Grimacing,Mannerisms,Verbigeration,Negativism,Waxy flexibility | 93.7 | 91.5 | 95.7 |
| 6 | Immobility/stupor,Grimacing,Verbigeration,Rigidity,Waxy flexibility,Withdrawal | 93.7 | 91.5 | 96.0 |
| 6 | Immobility/stupor,Mutism,Grimacing,Stereotypy,Negativism,Waxy flexibility | 93.7 | 91.5 | 95.7 |
| 6 | Immobility/stupor,Mutism,Posturing/catalepsy,Echopraxia/echolalia,Negativism,Waxy flexibility | 93.7 | 91.5 | 96.0 |
| 6 | Immobility/stupor,Mutism,Posturing/catalepsy,Grimacing,Echopraxia/echolalia,Mannerisms | 93.7 | 91.5 | 95.7 |
| 6 | Immobility/stupor,Mutism,Posturing/catalepsy,Grimacing,Verbigeration,Withdrawal | 93.7 | 91.3 | 96.0 |
| 6 | Immobility/stupor,Mutism,Posturing/catalepsy,Stereotypy,Mannerisms,Withdrawal | 93.7 | 91.5 | 96.0 |
| 6 | Immobility/stupor,Posturing/catalepsy,Echopraxia/echolalia,Verbigeration,Negativism,Waxy flexibility | 93.7 | 91.3 | 96.0 |
| 6 | Immobility/stupor,Posturing/catalepsy,Grimacing,Echopraxia/echolalia,Negativism,Waxy flexibility | 93.7 | 91.5 | 96.0 |
| 6 | Immobility/stupor,Posturing/catalepsy,Grimacing,Echopraxia/echolalia,Stereotypy,Rigidity | 93.7 | 91.3 | 95.7 |
| 6 | Immobility/stupor,Posturing/catalepsy,Grimacing,Echopraxia/echolalia,Verbigeration,Rigidity | 93.7 | 91.5 | 96.0 |
| 6 | Immobility/stupor,Posturing/catalepsy,Stereotypy,Mannerisms,Verbigeration,Rigidity | 93.7 | 91.3 | 95.7 |
| 6 | Immobility/stupor,Staring,Posturing/catalepsy,Echopraxia/echolalia,Verbigeration,Waxy flexibility | 93.7 | 91.5 | 96.0 |
| 6 | Immobility/stupor,Staring,Posturing/catalepsy,Grimacing,Echopraxia/echolalia,Waxy flexibility | 93.7 | 91.5 | 96.0 |
| 6 | Immobility/stupor,Staring,Posturing/catalepsy,Grimacing,Rigidity,Waxy flexibility | 93.7 | 91.3 | 95.7 |
| 6 | Mutism,Echopraxia/echolalia,Stereotypy,Rigidity,Negativism,Withdrawal | 93.7 | 91.3 | 96.0 |
| 6 | Mutism,Posturing/catalepsy,Stereotypy,Verbigeration,Rigidity,Negativism | 93.7 | 91.3 | 96.0 |
| 6 | Mutism,Stereotypy,Rigidity,Negativism,Waxy flexibility,Withdrawal | 93.7 | 91.5 | 96.0 |
| 6 | Staring,Grimacing,Echopraxia/echolalia,Mannerisms,Verbigeration,Waxy flexibility | 93.7 | 91.5 | 96.0 |
| 6 | Staring,Grimacing,Echopraxia/echolalia,Stereotypy,Mannerisms,Verbigeration | 93.7 | 91.5 | 96.0 |
| 6 | Staring,Posturing/catalepsy,Echopraxia/echolalia,Verbigeration,Waxy flexibility,Withdrawal | 93.7 | 91.5 | 96.0 |
| 6 | Staring,Posturing/catalepsy,Grimacing,Echopraxia/echolalia,Mannerisms,Waxy flexibility | 93.7 | 91.5 | 95.7 |
| 6 | Staring,Posturing/catalepsy,Grimacing,Echopraxia/echolalia,Stereotypy,Verbigeration | 93.7 | 91.5 | 95.7 |
| 6 | Excitement,Immobility/stupor,Grimacing,Echopraxia/echolalia,Stereotypy,Verbigeration | 93.5 | 91.3 | 95.7 |
| 6 | Excitement,Immobility/stupor,Grimacing,Stereotypy,Mannerisms,Waxy flexibility | 93.5 | 91.3 | 95.7 |
| 6 | Excitement,Immobility/stupor,Stereotypy,Mannerisms,Verbigeration,Waxy flexibility | 93.5 | 91.0 | 95.7 |
| 6 | Excitement,Mutism,Grimacing,Echopraxia/echolalia,Rigidity,Negativism | 93.5 | 91.0 | 95.7 |
| 6 | Excitement,Mutism,Mannerisms,Rigidity,Waxy flexibility,Withdrawal | 93.5 | 91.3 | 95.7 |
| 6 | Excitement,Mutism,Posturing/catalepsy,Grimacing,Echopraxia/echolalia,Rigidity | 93.5 | 91.0 | 95.7 |
| 6 | Excitement,Mutism,Posturing/catalepsy,Grimacing,Verbigeration,Rigidity | 93.5 | 91.3 | 95.7 |
| 6 | Excitement,Mutism,Posturing/catalepsy,Negativism,Waxy flexibility,Withdrawal | 93.5 | 91.0 | 95.7 |
| 6 | Excitement,Posturing/catalepsy,Verbigeration,Rigidity,Negativism,Withdrawal | 93.5 | 91.0 | 95.7 |
| 6 | Excitement,Staring,Echopraxia/echolalia,Stereotypy,Mannerisms,Verbigeration | 93.5 | 91.3 | 95.7 |
| 6 | Immobility/stupor,Echopraxia/echolalia,Stereotypy,Mannerisms,Verbigeration,Rigidity | 93.5 | 91.0 | 95.5 |
| 6 | Immobility/stupor,Echopraxia/echolalia,Verbigeration,Rigidity,Waxy flexibility,Withdrawal | 93.5 | 91.0 | 95.7 |
| 6 | Immobility/stupor,Grimacing,Echopraxia/echolalia,Stereotypy,Rigidity,Waxy flexibility | 93.5 | 91.0 | 95.7 |
| 6 | Immobility/stupor,Grimacing,Mannerisms,Verbigeration,Rigidity,Waxy flexibility | 93.5 | 91.3 | 95.7 |
| 6 | Immobility/stupor,Mutism,Echopraxia/echolalia,Stereotypy,Mannerisms,Withdrawal | 93.5 | 91.0 | 95.7 |
| 6 | Immobility/stupor,Mutism,Grimacing,Echopraxia/echolalia,Negativism,Waxy flexibility | 93.5 | 91.0 | 95.7 |
| 6 | Immobility/stupor,Mutism,Grimacing,Mannerisms,Verbigeration,Withdrawal | 93.5 | 91.3 | 95.7 |
| 6 | Immobility/stupor,Mutism,Grimacing,Mannerisms,Waxy flexibility,Withdrawal | 93.5 | 91.0 | 95.7 |
| 6 | Immobility/stupor,Mutism,Grimacing,Stereotypy,Mannerisms,Withdrawal | 93.5 | 91.0 | 95.7 |
| 6 | Immobility/stupor,Mutism,Grimacing,Stereotypy,Verbigeration,Withdrawal | 93.5 | 91.0 | 95.5 |
| 6 | Immobility/stupor,Mutism,Grimacing,Verbigeration,Negativism,Waxy flexibility | 93.5 | 91.0 | 95.7 |
| 6 | Immobility/stupor,Mutism,Posturing/catalepsy,Echopraxia/echolalia,Stereotypy,Withdrawal | 93.5 | 91.3 | 95.7 |
| 6 | Immobility/stupor,Mutism,Posturing/catalepsy,Grimacing,Echopraxia/echolalia,Verbigeration | 93.5 | 91.0 | 95.7 |
| 6 | Immobility/stupor,Mutism,Posturing/catalepsy,Grimacing,Mannerisms,Verbigeration | 93.5 | 91.0 | 95.7 |
| 6 | Immobility/stupor,Mutism,Posturing/catalepsy,Grimacing,Negativism,Waxy flexibility | 93.5 | 91.3 | 95.7 |
| 6 | Immobility/stupor,Mutism,Posturing/catalepsy,Grimacing,Stereotypy,Verbigeration | 93.5 | 91.3 | 95.7 |
| 6 | Immobility/stupor,Mutism,Posturing/catalepsy,Mannerisms,Rigidity,Waxy flexibility | 93.5 | 91.0 | 95.7 |
| 6 | Immobility/stupor,Posturing/catalepsy,Echopraxia/echolalia,Mannerisms,Verbigeration,Rigidity | 93.5 | 91.0 | 95.7 |
| 6 | Immobility/stupor,Posturing/catalepsy,Echopraxia/echolalia,Stereotypy,Mannerisms,Rigidity | 93.5 | 91.0 | 95.7 |
| 6 | Immobility/stupor,Posturing/catalepsy,Echopraxia/echolalia,Stereotypy,Mannerisms,Withdrawal | 93.5 | 91.0 | 95.7 |
| 6 | Immobility/stupor,Posturing/catalepsy,Echopraxia/echolalia,Stereotypy,Verbigeration,Rigidity | 93.5 | 91.0 | 95.7 |
| 6 | Immobility/stupor,Posturing/catalepsy,Grimacing,Mannerisms,Rigidity,Waxy flexibility | 93.5 | 91.0 | 95.7 |
| 6 | Immobility/stupor,Posturing/catalepsy,Grimacing,Mannerisms,Verbigeration,Withdrawal | 93.5 | 91.0 | 95.7 |
| 6 | Immobility/stupor,Posturing/catalepsy,Stereotypy,Rigidity,Waxy flexibility,Withdrawal | 93.5 | 91.0 | 95.7 |
| 6 | Mutism,Grimacing,Stereotypy,Rigidity,Negativism,Withdrawal | 93.5 | 91.0 | 95.7 |
| 6 | Mutism,Grimacing,Verbigeration,Rigidity,Negativism,Withdrawal | 93.5 | 91.0 | 95.7 |
| 6 | Mutism,Posturing/catalepsy,Echopraxia/echolalia,Rigidity,Negativism,Withdrawal | 93.5 | 91.0 | 95.7 |
| 6 | Mutism,Posturing/catalepsy,Grimacing,Mannerisms,Rigidity,Negativism | 93.5 | 91.0 | 95.7 |
| 6 | Mutism,Posturing/catalepsy,Stereotypy,Mannerisms,Rigidity,Negativism | 93.5 | 91.0 | 95.7 |
| 6 | Mutism,Verbigeration,Rigidity,Negativism,Waxy flexibility,Withdrawal | 93.5 | 91.3 | 95.7 |
| 6 | Staring,Grimacing,Echopraxia/echolalia,Stereotypy,Verbigeration,Waxy flexibility | 93.5 | 91.0 | 95.7 |
| 6 | Staring,Posturing/catalepsy,Echopraxia/echolalia,Stereotypy,Rigidity,Waxy flexibility | 93.5 | 91.0 | 95.7 |
| 6 | Staring,Posturing/catalepsy,Grimacing,Echopraxia/echolalia,Rigidity,Waxy flexibility | 93.5 | 91.0 | 95.7 |
| 6 | Staring,Posturing/catalepsy,Grimacing,Mannerisms,Verbigeration,Waxy flexibility | 93.5 | 91.0 | 95.7 |
| 6 | Staring,Posturing/catalepsy,Grimacing,Stereotypy,Verbigeration,Waxy flexibility | 93.5 | 91.3 | 95.7 |
| 6 | Excitement,Immobility/stupor,Echopraxia/echolalia,Stereotypy,Mannerisms,Verbigeration | 93.3 | 90.8 | 95.5 |
| 6 | Excitement,Immobility/stupor,Grimacing,Echopraxia/echolalia,Mannerisms,Verbigeration | 93.3 | 90.8 | 95.5 |
| 6 | Excitement,Immobility/stupor,Grimacing,Echopraxia/echolalia,Stereotypy,Mannerisms | 93.3 | 90.8 | 95.5 |
| 6 | Excitement,Immobility/stupor,Posturing/catalepsy,Stereotypy,Mannerisms,Waxy flexibility | 93.3 | 90.8 | 95.5 |
| 6 | Excitement,Mutism,Echopraxia/echolalia,Rigidity,Negativism,Waxy flexibility | 93.3 | 90.8 | 95.5 |
| 6 | Excitement,Mutism,Echopraxia/echolalia,Verbigeration,Rigidity,Negativism | 93.3 | 90.8 | 95.5 |
| 6 | Excitement,Mutism,Grimacing,Verbigeration,Rigidity,Negativism | 93.3 | 91.0 | 95.5 |
| 6 | Excitement,Mutism,Posturing/catalepsy,Mannerisms,Verbigeration,Rigidity | 93.3 | 90.8 | 95.5 |
| 6 | Excitement,Mutism,Verbigeration,Rigidity,Negativism,Waxy flexibility | 93.3 | 90.8 | 95.5 |
| 6 | Excitement,Posturing/catalepsy,Echopraxia/echolalia,Rigidity,Negativism,Withdrawal | 93.3 | 90.8 | 95.5 |
| 6 | Excitement,Posturing/catalepsy,Stereotypy,Rigidity,Negativism,Withdrawal | 93.3 | 90.8 | 95.5 |
| 6 | Immobility/stupor,Echopraxia/echolalia,Stereotypy,Mannerisms,Rigidity,Waxy flexibility | 93.3 | 90.8 | 95.5 |
| 6 | Immobility/stupor,Grimacing,Echopraxia/echolalia,Stereotypy,Waxy flexibility,Withdrawal | 93.3 | 90.8 | 95.5 |
| 6 | Immobility/stupor,Grimacing,Echopraxia/echolalia,Verbigeration,Rigidity,Waxy flexibility | 93.3 | 90.8 | 95.5 |
| 6 | Immobility/stupor,Grimacing,Verbigeration,Negativism,Waxy flexibility,Withdrawal | 93.3 | 90.8 | 95.5 |
| 6 | Immobility/stupor,Mutism,Echopraxia/echolalia,Mannerisms,Waxy flexibility,Withdrawal | 93.3 | 90.8 | 95.5 |
| 6 | Immobility/stupor,Mutism,Echopraxia/echolalia,Stereotypy,Rigidity,Waxy flexibility | 93.3 | 90.8 | 95.5 |
| 6 | Immobility/stupor,Mutism,Echopraxia/echolalia,Verbigeration,Rigidity,Waxy flexibility | 93.3 | 90.8 | 95.5 |
| 6 | Immobility/stupor,Mutism,Grimacing,Echopraxia/echolalia,Rigidity,Waxy flexibility | 93.3 | 90.8 | 95.5 |
| 6 | Immobility/stupor,Mutism,Grimacing,Echopraxia/echolalia,Verbigeration,Withdrawal | 93.3 | 90.8 | 95.5 |
| 6 | Immobility/stupor,Mutism,Grimacing,Negativism,Waxy flexibility,Withdrawal | 93.3 | 90.8 | 95.5 |
| 6 | Immobility/stupor,Mutism,Posturing/catalepsy,Echopraxia/echolalia,Verbigeration,Withdrawal | 93.3 | 90.8 | 95.5 |
| 6 | Immobility/stupor,Mutism,Posturing/catalepsy,Mannerisms,Waxy flexibility,Withdrawal | 93.3 | 90.8 | 95.5 |
| 6 | Immobility/stupor,Mutism,Posturing/catalepsy,Stereotypy,Verbigeration,Withdrawal | 93.3 | 90.8 | 95.5 |
| 6 | Immobility/stupor,Mutism,Staring,Posturing/catalepsy,Grimacing,Waxy flexibility | 93.3 | 90.8 | 95.5 |
| 6 | Immobility/stupor,Posturing/catalepsy,Grimacing,Rigidity,Waxy flexibility,Withdrawal | 93.3 | 90.8 | 95.5 |
| 6 | Immobility/stupor,Posturing/catalepsy,Grimacing,Stereotypy,Verbigeration,Withdrawal | 93.3 | 90.8 | 95.5 |
| 6 | Immobility/stupor,Posturing/catalepsy,Verbigeration,Rigidity,Waxy flexibility,Withdrawal | 93.3 | 90.8 | 95.5 |
| 6 | Immobility/stupor,Stereotypy,Mannerisms,Verbigeration,Rigidity,Waxy flexibility | 93.3 | 90.8 | 95.5 |
| 6 | Mutism,Echopraxia/echolalia,Verbigeration,Rigidity,Negativism,Withdrawal | 93.3 | 90.8 | 95.5 |
| 6 | Mutism,Posturing/catalepsy,Echopraxia/echolalia,Verbigeration,Rigidity,Negativism | 93.3 | 90.8 | 95.5 |
| 6 | Mutism,Posturing/catalepsy,Grimacing,Rigidity,Negativism,Withdrawal | 93.3 | 90.8 | 95.5 |
| 6 | Mutism,Stereotypy,Mannerisms,Rigidity,Negativism,Waxy flexibility | 93.3 | 90.8 | 95.5 |
| 6 | Posturing/catalepsy,Stereotypy,Mannerisms,Rigidity,Negativism,Withdrawal | 93.3 | 90.8 | 95.5 |
| 6 | Staring,Posturing/catalepsy,Echopraxia/echolalia,Stereotypy,Mannerisms,Verbigeration | 93.3 | 90.8 | 95.5 |
| 6 | Staring,Posturing/catalepsy,Echopraxia/echolalia,Stereotypy,Mannerisms,Waxy flexibility | 93.3 | 90.8 | 95.5 |
| 6 | Staring,Posturing/catalepsy,Echopraxia/echolalia,Verbigeration,Rigidity,Waxy flexibility | 93.3 | 90.8 | 95.5 |
| 6 | Staring,Posturing/catalepsy,Grimacing,Echopraxia/echolalia,Stereotypy,Waxy flexibility | 93.3 | 90.8 | 95.5 |
| 6 | Staring,Posturing/catalepsy,Stereotypy,Mannerisms,Verbigeration,Waxy flexibility | 93.3 | 90.8 | 95.5 |
| 6 | Mutism,Stereotypy,Mannerisms,Verbigeration,Rigidity,Negativism | 93.2 | 90.8 | 95.5 |
| 6 | Excitement,Mutism,Echopraxia/echolalia,Stereotypy,Rigidity,Withdrawal | 93.1 | 90.6 | 95.3 |
| 6 | Excitement,Mutism,Grimacing,Rigidity,Negativism,Waxy flexibility | 93.1 | 90.6 | 95.3 |
| 6 | Excitement,Mutism,Grimacing,Stereotypy,Rigidity,Negativism | 93.1 | 90.6 | 95.3 |
| 6 | Excitement,Mutism,Posturing/catalepsy,Grimacing,Rigidity,Waxy flexibility | 93.1 | 90.6 | 95.3 |
| 6 | Excitement,Mutism,Stereotypy,Verbigeration,Rigidity,Withdrawal | 93.1 | 90.6 | 95.3 |
| 6 | Excitement,Posturing/catalepsy,Mannerisms,Rigidity,Negativism,Withdrawal | 93.1 | 90.6 | 95.3 |
| 6 | Immobility/stupor,Mutism,Echopraxia/echolalia,Mannerisms,Verbigeration,Withdrawal | 93.1 | 90.6 | 95.3 |
| 6 | Immobility/stupor,Mutism,Grimacing,Echopraxia/echolalia,Waxy flexibility,Withdrawal | 93.1 | 90.6 | 95.3 |
| 6 | Immobility/stupor,Mutism,Posturing/catalepsy,Echopraxia/echolalia,Mannerisms,Verbigeration | 93.1 | 90.6 | 95.3 |
| 6 | Immobility/stupor,Mutism,Posturing/catalepsy,Grimacing,Stereotypy,Mannerisms | 93.1 | 90.6 | 95.3 |
| 6 | Immobility/stupor,Posturing/catalepsy,Echopraxia/echolalia,Rigidity,Waxy flexibility,Withdrawal | 93.1 | 90.6 | 95.3 |
| 6 | Mutism,Posturing/catalepsy,Echopraxia/echolalia,Mannerisms,Negativism,Withdrawal | 93.1 | 90.6 | 95.3 |
| 6 | Mutism,Posturing/catalepsy,Grimacing,Mannerisms,Negativism,Withdrawal | 93.1 | 90.6 | 95.3 |
| 6 | Mutism,Posturing/catalepsy,Mannerisms,Rigidity,Negativism,Waxy flexibility | 93.1 | 90.6 | 95.3 |
| 6 | Mutism,Posturing/catalepsy,Mannerisms,Verbigeration,Negativism,Withdrawal | 93.1 | 90.6 | 95.3 |
| 6 | Excitement,Mutism,Posturing/catalepsy,Echopraxia/echolalia,Mannerisms,Rigidity | 93.0 | 90.6 | 95.3 |
| 6 | Excitement,Mutism,Posturing/catalepsy,Echopraxia/echolalia,Stereotypy,Rigidity | 93.0 | 90.6 | 95.3 |
| 6 | Excitement,Mutism,Posturing/catalepsy,Stereotypy,Verbigeration,Rigidity | 93.0 | 90.6 | 95.3 |
| 6 | Immobility/stupor,Grimacing,Echopraxia/echolalia,Mannerisms,Waxy flexibility,Withdrawal | 93.0 | 90.6 | 95.3 |
| 6 | Immobility/stupor,Grimacing,Echopraxia/echolalia,Stereotypy,Verbigeration,Withdrawal | 93.0 | 90.6 | 95.3 |
| 6 | Immobility/stupor,Mutism,Posturing/catalepsy,Grimacing,Echopraxia/echolalia,Stereotypy | 93.0 | 90.6 | 95.3 |
| 6 | Immobility/stupor,Mutism,Posturing/catalepsy,Stereotypy,Rigidity,Waxy flexibility | 93.0 | 90.6 | 95.3 |
| 6 | Immobility/stupor,Mutism,Posturing/catalepsy,Verbigeration,Rigidity,Waxy flexibility | 93.0 | 90.6 | 95.3 |
| 6 | Immobility/stupor,Mutism,Stereotypy,Mannerisms,Verbigeration,Withdrawal | 93.0 | 90.6 | 95.3 |
| 6 | Immobility/stupor,Posturing/catalepsy,Echopraxia/echolalia,Mannerisms,Verbigeration,Withdrawal | 93.0 | 90.6 | 95.3 |
| 6 | Immobility/stupor,Posturing/catalepsy,Grimacing,Stereotypy,Rigidity,Waxy flexibility | 93.0 | 90.6 | 95.3 |
| 6 | Immobility/stupor,Posturing/catalepsy,Grimacing,Verbigeration,Rigidity,Waxy flexibility | 93.0 | 90.6 | 95.3 |
| 6 | Immobility/stupor,Posturing/catalepsy,Stereotypy,Mannerisms,Verbigeration,Withdrawal | 93.0 | 90.6 | 95.3 |
| 6 | Mutism,Echopraxia/echolalia,Stereotypy,Mannerisms,Rigidity,Negativism | 93.0 | 90.6 | 95.3 |
| 6 | Mutism,Posturing/catalepsy,Grimacing,Verbigeration,Rigidity,Negativism | 93.0 | 90.6 | 95.3 |
| 6 | Mutism,Stereotypy,Verbigeration,Rigidity,Negativism,Waxy flexibility | 93.0 | 90.6 | 95.3 |
| 6 | Posturing/catalepsy,Mannerisms,Verbigeration,Rigidity,Negativism,Withdrawal | 93.0 | 90.6 | 95.3 |
| 6 | Immobility/stupor,Posturing/catalepsy,Grimacing,Mannerisms,Waxy flexibility,Withdrawal | 92.9 | 90.4 | 95.1 |
| 6 | Excitement,Immobility/stupor,Grimacing,Stereotypy,Mannerisms,Verbigeration | 92.8 | 90.4 | 95.1 |
| 6 | Excitement,Mutism,Grimacing,Echopraxia/echolalia,Rigidity,Withdrawal | 92.8 | 90.4 | 95.1 |
| 6 | Excitement,Mutism,Grimacing,Verbigeration,Rigidity,Withdrawal | 92.8 | 90.4 | 95.1 |
| 6 | Excitement,Mutism,Posturing/catalepsy,Grimacing,Mannerisms,Rigidity | 92.8 | 90.4 | 95.1 |
| 6 | Excitement,Mutism,Posturing/catalepsy,Mannerisms,Rigidity,Waxy flexibility | 92.8 | 90.4 | 95.1 |
| 6 | Excitement,Mutism,Posturing/catalepsy,Stereotypy,Rigidity,Waxy flexibility | 92.8 | 90.4 | 95.1 |
| 6 | Excitement,Posturing/catalepsy,Grimacing,Rigidity,Negativism,Withdrawal | 92.8 | 90.4 | 95.1 |
| 6 | Immobility/stupor,Echopraxia/echolalia,Stereotypy,Verbigeration,Rigidity,Waxy flexibility | 92.8 | 90.4 | 95.1 |
| 6 | Immobility/stupor,Grimacing,Stereotypy,Mannerisms,Verbigeration,Withdrawal | 92.8 | 90.4 | 95.1 |
| 6 | Immobility/stupor,Grimacing,Stereotypy,Mannerisms,Waxy flexibility,Withdrawal | 92.8 | 90.4 | 95.1 |
| 6 | Immobility/stupor,Mutism,Echopraxia/echolalia,Stereotypy,Verbigeration,Withdrawal | 92.8 | 90.4 | 95.1 |
| 6 | Immobility/stupor,Mutism,Echopraxia/echolalia,Stereotypy,Waxy flexibility,Withdrawal | 92.8 | 90.4 | 95.1 |
| 6 | Immobility/stupor,Mutism,Grimacing,Echopraxia/echolalia,Mannerisms,Waxy flexibility | 92.8 | 90.4 | 95.1 |
| 6 | Immobility/stupor,Mutism,Posturing/catalepsy,Echopraxia/echolalia,Stereotypy,Mannerisms | 92.8 | 90.4 | 95.1 |
| 6 | Immobility/stupor,Mutism,Posturing/catalepsy,Echopraxia/echolalia,Stereotypy,Verbigeration | 92.8 | 90.4 | 95.1 |
| 6 | Immobility/stupor,Mutism,Posturing/catalepsy,Echopraxia/echolalia,Waxy flexibility,Withdrawal | 92.8 | 90.4 | 95.1 |
| 6 | Immobility/stupor,Mutism,Posturing/catalepsy,Grimacing,Waxy flexibility,Withdrawal | 92.8 | 90.4 | 95.1 |
| 6 | Immobility/stupor,Mutism,Posturing/catalepsy,Rigidity,Waxy flexibility,Withdrawal | 92.8 | 90.4 | 95.1 |
| 6 | Immobility/stupor,Mutism,Stereotypy,Mannerisms,Waxy flexibility,Withdrawal | 92.8 | 90.4 | 95.1 |
| 6 | Immobility/stupor,Posturing/catalepsy,Grimacing,Echopraxia/echolalia,Verbigeration,Withdrawal | 92.8 | 90.4 | 95.1 |
| 6 | Immobility/stupor,Posturing/catalepsy,Stereotypy,Verbigeration,Rigidity,Waxy flexibility | 92.8 | 90.4 | 95.1 |
| 6 | Mutism,Echopraxia/echolalia,Rigidity,Negativism,Waxy flexibility,Withdrawal | 92.8 | 90.4 | 95.1 |
| 6 | Mutism,Grimacing,Echopraxia/echolalia,Mannerisms,Rigidity,Negativism | 92.8 | 90.4 | 95.1 |
| 6 | Mutism,Grimacing,Echopraxia/echolalia,Rigidity,Negativism,Withdrawal | 92.8 | 90.4 | 95.1 |
| 6 | Mutism,Grimacing,Stereotypy,Verbigeration,Rigidity,Negativism | 92.8 | 90.4 | 95.1 |
| 6 | Mutism,Posturing/catalepsy,Grimacing,Mannerisms,Rigidity,Withdrawal | 92.8 | 90.4 | 95.1 |
| 6 | Mutism,Posturing/catalepsy,Rigidity,Negativism,Waxy flexibility,Withdrawal | 92.8 | 90.4 | 95.1 |
| 6 | Mutism,Posturing/catalepsy,Stereotypy,Mannerisms,Negativism,Withdrawal | 92.8 | 90.4 | 95.1 |
| 6 | Posturing/catalepsy,Echopraxia/echolalia,Mannerisms,Rigidity,Negativism,Withdrawal | 92.8 | 90.4 | 95.1 |
| 6 | Posturing/catalepsy,Stereotypy,Verbigeration,Rigidity,Negativism,Withdrawal | 92.8 | 90.4 | 95.1 |
| 6 | Staring,Echopraxia/echolalia,Stereotypy,Mannerisms,Verbigeration,Waxy flexibility | 92.8 | 90.4 | 95.1 |
| 6 | Excitement,Mutism,Echopraxia/echolalia,Mannerisms,Rigidity,Negativism | 92.6 | 90.1 | 94.8 |
| 6 | Excitement,Mutism,Echopraxia/echolalia,Verbigeration,Rigidity,Withdrawal | 92.6 | 89.9 | 94.8 |
| 6 | Excitement,Mutism,Grimacing,Stereotypy,Rigidity,Withdrawal | 92.6 | 90.1 | 94.8 |
| 6 | Excitement,Mutism,Mannerisms,Verbigeration,Rigidity,Negativism | 92.6 | 90.1 | 94.8 |
| 6 | Excitement,Mutism,Posturing/catalepsy,Echopraxia/echolalia,Verbigeration,Negativism | 92.6 | 90.1 | 95.1 |
| 6 | Excitement,Mutism,Posturing/catalepsy,Echopraxia/echolalia,Waxy flexibility,Withdrawal | 92.6 | 90.1 | 94.8 |
| 6 | Excitement,Mutism,Posturing/catalepsy,Grimacing,Echopraxia/echolalia,Negativism | 92.6 | 90.1 | 95.1 |
| 6 | Excitement,Mutism,Posturing/catalepsy,Grimacing,Stereotypy,Rigidity | 92.6 | 90.1 | 94.8 |
| 6 | Excitement,Mutism,Posturing/catalepsy,Stereotypy,Verbigeration,Negativism | 92.6 | 90.1 | 94.8 |
| 6 | Excitement,Mutism,Posturing/catalepsy,Verbigeration,Waxy flexibility,Withdrawal | 92.6 | 90.1 | 94.8 |
| 6 | Excitement,Mutism,Stereotypy,Mannerisms,Rigidity,Negativism | 92.6 | 90.1 | 94.8 |
| 6 | Excitement,Posturing/catalepsy,Rigidity,Negativism,Waxy flexibility,Withdrawal | 92.6 | 90.1 | 94.8 |
| 6 | Immobility/stupor,Echopraxia/echolalia,Mannerisms,Verbigeration,Rigidity,Waxy flexibility | 92.6 | 90.1 | 94.8 |
| 6 | Immobility/stupor,Grimacing,Echopraxia/echolalia,Mannerisms,Verbigeration,Withdrawal | 92.6 | 90.1 | 94.8 |
| 6 | Immobility/stupor,Mutism,Grimacing,Echopraxia/echolalia,Mannerisms,Verbigeration | 92.6 | 90.1 | 94.8 |
| 6 | Immobility/stupor,Mutism,Grimacing,Echopraxia/echolalia,Stereotypy,Mannerisms | 92.6 | 90.1 | 95.1 |
| 6 | Immobility/stupor,Mutism,Grimacing,Echopraxia/echolalia,Stereotypy,Verbigeration | 92.6 | 89.9 | 95.1 |
| 6 | Immobility/stupor,Mutism,Grimacing,Echopraxia/echolalia,Verbigeration,Waxy flexibility | 92.6 | 89.9 | 95.1 |
| 6 | Immobility/stupor,Mutism,Grimacing,Stereotypy,Waxy flexibility,Withdrawal | 92.6 | 90.1 | 94.8 |
| 6 | Immobility/stupor,Mutism,Grimacing,Verbigeration,Waxy flexibility,Withdrawal | 92.6 | 90.1 | 94.8 |
| 6 | Immobility/stupor,Mutism,Mannerisms,Verbigeration,Waxy flexibility,Withdrawal | 92.6 | 90.1 | 94.8 |
| 6 | Immobility/stupor,Mutism,Posturing/catalepsy,Echopraxia/echolalia,Rigidity,Waxy flexibility | 92.6 | 90.1 | 94.8 |
| 6 | Immobility/stupor,Mutism,Posturing/catalepsy,Grimacing,Verbigeration,Waxy flexibility | 92.6 | 90.1 | 94.8 |
| 6 | Immobility/stupor,Mutism,Posturing/catalepsy,Stereotypy,Mannerisms,Verbigeration | 92.6 | 90.1 | 94.8 |
| 6 | Immobility/stupor,Mutism,Posturing/catalepsy,Stereotypy,Waxy flexibility,Withdrawal | 92.6 | 90.1 | 94.8 |
| 6 | Immobility/stupor,Mutism,Posturing/catalepsy,Verbigeration,Waxy flexibility,Withdrawal | 92.6 | 90.1 | 95.1 |
| 6 | Immobility/stupor,Posturing/catalepsy,Echopraxia/echolalia,Mannerisms,Rigidity,Waxy flexibility | 92.6 | 90.1 | 95.1 |
| 6 | Immobility/stupor,Posturing/catalepsy,Echopraxia/echolalia,Stereotypy,Verbigeration,Withdrawal | 92.6 | 90.1 | 94.8 |
| 6 | Immobility/stupor,Posturing/catalepsy,Grimacing,Echopraxia/echolalia,Rigidity,Waxy flexibility | 92.6 | 90.1 | 94.8 |
| 6 | Immobility/stupor,Posturing/catalepsy,Grimacing,Echopraxia/echolalia,Stereotypy,Mannerisms | 92.6 | 90.1 | 94.8 |
| 6 | Immobility/stupor,Posturing/catalepsy,Mannerisms,Verbigeration,Rigidity,Waxy flexibility | 92.6 | 90.1 | 95.1 |
| 6 | Immobility/stupor,Posturing/catalepsy,Stereotypy,Mannerisms,Rigidity,Waxy flexibility | 92.6 | 90.1 | 94.8 |
| 6 | Mutism,Echopraxia/echolalia,Mannerisms,Rigidity,Negativism,Waxy flexibility | 92.6 | 90.1 | 95.1 |
| 6 | Mutism,Grimacing,Mannerisms,Rigidity,Waxy flexibility,Withdrawal | 92.6 | 90.1 | 94.8 |
| 6 | Mutism,Grimacing,Rigidity,Negativism,Waxy flexibility,Withdrawal | 92.6 | 90.1 | 94.8 |
| 6 | Mutism,Grimacing,Stereotypy,Mannerisms,Rigidity,Negativism | 92.6 | 90.1 | 95.1 |
| 6 | Mutism,Posturing/catalepsy,Echopraxia/echolalia,Stereotypy,Rigidity,Negativism | 92.6 | 90.1 | 94.8 |
| 6 | Mutism,Posturing/catalepsy,Mannerisms,Negativism,Waxy flexibility,Withdrawal | 92.6 | 90.1 | 95.1 |
| 6 | Mutism,Posturing/catalepsy,Mannerisms,Rigidity,Waxy flexibility,Withdrawal | 92.6 | 90.1 | 94.8 |
| 6 | Mutism,Posturing/catalepsy,Mannerisms,Verbigeration,Rigidity,Withdrawal | 92.6 | 90.1 | 94.8 |
| 6 | Mutism,Posturing/catalepsy,Stereotypy,Verbigeration,Negativism,Withdrawal | 92.6 | 90.1 | 94.8 |
| 6 | Mutism,Posturing/catalepsy,Verbigeration,Rigidity,Negativism,Waxy flexibility | 92.6 | 90.1 | 94.8 |
| 6 | Posturing/catalepsy,Grimacing,Mannerisms,Rigidity,Negativism,Withdrawal | 92.6 | 90.1 | 95.1 |
| 6 | Staring,Posturing/catalepsy,Echopraxia/echolalia,Mannerisms,Verbigeration,Waxy flexibility | 92.6 | 90.1 | 94.8 |
| 6 | Staring,Posturing/catalepsy,Grimacing,Echopraxia/echolalia,Verbigeration,Waxy flexibility | 92.6 | 90.1 | 94.8 |
| 6 | Excitement,Mutism,Echopraxia/echolalia,Stereotypy,Rigidity,Waxy flexibility | 92.4 | 89.9 | 94.8 |
| 6 | Excitement,Mutism,Grimacing,Mannerisms,Rigidity,Negativism | 92.4 | 89.9 | 94.8 |
| 6 | Excitement,Mutism,Grimacing,Mannerisms,Rigidity,Withdrawal | 92.4 | 89.7 | 94.8 |
| 6 | Excitement,Mutism,Mannerisms,Rigidity,Negativism,Waxy flexibility | 92.4 | 89.9 | 94.6 |
| 6 | Excitement,Mutism,Mannerisms,Verbigeration,Rigidity,Withdrawal | 92.4 | 89.7 | 94.6 |
| 6 | Excitement,Mutism,Posturing/catalepsy,Grimacing,Verbigeration,Negativism | 92.4 | 89.9 | 94.8 |
| 6 | Excitement,Mutism,Posturing/catalepsy,Grimacing,Waxy flexibility,Withdrawal | 92.4 | 89.9 | 94.8 |
| 6 | Excitement,Mutism,Posturing/catalepsy,Stereotypy,Waxy flexibility,Withdrawal | 92.4 | 89.9 | 94.8 |
| 6 | Excitement,Mutism,Stereotypy,Mannerisms,Rigidity,Withdrawal | 92.4 | 89.7 | 94.6 |
| 6 | Excitement,Mutism,Stereotypy,Verbigeration,Rigidity,Waxy flexibility | 92.4 | 89.9 | 94.8 |
| 6 | Immobility/stupor,Echopraxia/echolalia,Stereotypy,Mannerisms,Verbigeration,Withdrawal | 92.4 | 89.9 | 94.6 |
| 6 | Immobility/stupor,Echopraxia/echolalia,Stereotypy,Mannerisms,Waxy flexibility,Withdrawal | 92.4 | 89.9 | 94.6 |
| 6 | Immobility/stupor,Grimacing,Echopraxia/echolalia,Stereotypy,Mannerisms,Waxy flexibility | 92.4 | 89.9 | 94.8 |
| 6 | Immobility/stupor,Mutism,Echopraxia/echolalia,Verbigeration,Waxy flexibility,Withdrawal | 92.4 | 89.9 | 94.6 |
| 6 | Immobility/stupor,Mutism,Grimacing,Echopraxia/echolalia,Stereotypy,Waxy flexibility | 92.4 | 89.7 | 94.8 |
| 6 | Immobility/stupor,Mutism,Grimacing,Stereotypy,Mannerisms,Verbigeration | 92.4 | 89.9 | 94.8 |
| 6 | Immobility/stupor,Mutism,Grimacing,Stereotypy,Verbigeration,Waxy flexibility | 92.4 | 89.9 | 94.8 |
| 6 | Immobility/stupor,Mutism,Posturing/catalepsy,Grimacing,Mannerisms,Waxy flexibility | 92.4 | 89.9 | 94.6 |
| 6 | Immobility/stupor,Mutism,Posturing/catalepsy,Grimacing,Rigidity,Waxy flexibility | 92.4 | 89.9 | 94.6 |
| 6 | Immobility/stupor,Mutism,Stereotypy,Verbigeration,Waxy flexibility,Withdrawal | 92.4 | 89.9 | 94.6 |
| 6 | Immobility/stupor,Posturing/catalepsy,Echopraxia/echolalia,Mannerisms,Waxy flexibility,Withdrawal | 92.4 | 89.9 | 94.6 |
| 6 | Immobility/stupor,Posturing/catalepsy,Echopraxia/echolalia,Stereotypy,Rigidity,Waxy flexibility | 92.4 | 89.9 | 94.6 |
| 6 | Immobility/stupor,Posturing/catalepsy,Grimacing,Echopraxia/echolalia,Mannerisms,Verbigeration | 92.4 | 89.9 | 94.8 |
| 6 | Immobility/stupor,Posturing/catalepsy,Grimacing,Echopraxia/echolalia,Waxy flexibility,Withdrawal | 92.4 | 89.9 | 94.6 |
| 6 | Immobility/stupor,Posturing/catalepsy,Grimacing,Stereotypy,Waxy flexibility,Withdrawal | 92.4 | 89.9 | 94.8 |
| 6 | Immobility/stupor,Posturing/catalepsy,Stereotypy,Mannerisms,Waxy flexibility,Withdrawal | 92.4 | 89.9 | 94.8 |
| 6 | Mutism,Echopraxia/echolalia,Mannerisms,Verbigeration,Rigidity,Negativism | 92.4 | 89.7 | 94.8 |
| 6 | Mutism,Echopraxia/echolalia,Stereotypy,Verbigeration,Rigidity,Negativism | 92.4 | 89.9 | 94.8 |
| 6 | Mutism,Grimacing,Mannerisms,Rigidity,Negativism,Waxy flexibility | 92.4 | 89.9 | 94.8 |
| 6 | Mutism,Grimacing,Mannerisms,Verbigeration,Rigidity,Negativism | 92.4 | 89.9 | 94.6 |
| 6 | Mutism,Mannerisms,Verbigeration,Rigidity,Negativism,Waxy flexibility | 92.4 | 89.9 | 94.8 |
| 6 | Mutism,Posturing/catalepsy,Echopraxia/echolalia,Mannerisms,Rigidity,Withdrawal | 92.4 | 89.9 | 94.8 |
| 6 | Mutism,Posturing/catalepsy,Grimacing,Echopraxia/echolalia,Rigidity,Negativism | 92.4 | 89.9 | 94.8 |
| 6 | Mutism,Posturing/catalepsy,Grimacing,Stereotypy,Rigidity,Negativism | 92.4 | 89.7 | 94.8 |
| 6 | Mutism,Posturing/catalepsy,Stereotypy,Mannerisms,Rigidity,Withdrawal | 92.4 | 89.9 | 94.8 |
| 6 | Mutism,Stereotypy,Mannerisms,Rigidity,Waxy flexibility,Withdrawal | 92.4 | 89.9 | 94.8 |
| 6 | Posturing/catalepsy,Mannerisms,Rigidity,Negativism,Waxy flexibility,Withdrawal | 92.4 | 89.7 | 94.8 |
| 6 | Staring,Posturing/catalepsy,Echopraxia/echolalia,Stereotypy,Verbigeration,Waxy flexibility | 92.4 | 89.9 | 94.8 |
| 6 | Excitement,Mutism,Echopraxia/echolalia,Mannerisms,Rigidity,Withdrawal | 92.2 | 89.5 | 94.6 |
| 6 | Excitement,Mutism,Echopraxia/echolalia,Stereotypy,Negativism,Withdrawal | 92.2 | 89.7 | 94.6 |
| 6 | Excitement,Mutism,Grimacing,Echopraxia/echolalia,Rigidity,Waxy flexibility | 92.2 | 89.7 | 94.6 |
| 6 | Excitement,Mutism,Posturing/catalepsy,Echopraxia/echolalia,Negativism,Waxy flexibility | 92.2 | 89.7 | 94.6 |
| 6 | Excitement,Mutism,Posturing/catalepsy,Echopraxia/echolalia,Stereotypy,Negativism | 92.2 | 89.7 | 94.4 |
| 6 | Excitement,Mutism,Posturing/catalepsy,Verbigeration,Negativism,Waxy flexibility | 92.2 | 89.7 | 94.6 |
| 6 | Excitement,Mutism,Stereotypy,Negativism,Waxy flexibility,Withdrawal | 92.2 | 89.7 | 94.6 |
| 6 | Excitement,Mutism,Stereotypy,Verbigeration,Negativism,Withdrawal | 92.2 | 89.5 | 94.6 |
| 6 | Immobility/stupor,Grimacing,Stereotypy,Verbigeration,Waxy flexibility,Withdrawal | 92.2 | 89.7 | 94.6 |
| 6 | Immobility/stupor,Mutism,Posturing/catalepsy,Stereotypy,Verbigeration,Waxy flexibility | 92.2 | 89.7 | 94.6 |
| 6 | Immobility/stupor,Posturing/catalepsy,Echopraxia/echolalia,Stereotypy,Waxy flexibility,Withdrawal | 92.2 | 89.7 | 94.6 |
| 6 | Immobility/stupor,Posturing/catalepsy,Grimacing,Stereotypy,Mannerisms,Verbigeration | 92.2 | 89.5 | 94.6 |
| 6 | Immobility/stupor,Posturing/catalepsy,Mannerisms,Verbigeration,Waxy flexibility,Withdrawal | 92.2 | 89.7 | 94.6 |
| 6 | Mutism,Posturing/catalepsy,Echopraxia/echolalia,Rigidity,Negativism,Waxy flexibility | 92.2 | 89.7 | 94.6 |
| 6 | Mutism,Posturing/catalepsy,Grimacing,Stereotypy,Negativism,Withdrawal | 92.2 | 89.5 | 94.6 |
| 6 | Mutism,Posturing/catalepsy,Stereotypy,Rigidity,Negativism,Waxy flexibility | 92.2 | 89.5 | 94.6 |
| 6 | Mutism,Stereotypy,Verbigeration,Rigidity,Waxy flexibility,Withdrawal | 92.2 | 89.7 | 94.6 |
| 6 | Posturing/catalepsy,Grimacing,Stereotypy,Rigidity,Negativism,Withdrawal | 92.2 | 89.7 | 94.6 |
| 6 | Excitement,Mutism,Posturing/catalepsy,Echopraxia/echolalia,Verbigeration,Withdrawal | 92.1 | 89.5 | 94.6 |
| 6 | Excitement,Mutism,Posturing/catalepsy,Grimacing,Echopraxia/echolalia,Withdrawal | 92.1 | 89.5 | 94.6 |
| 6 | Excitement,Mutism,Posturing/catalepsy,Mannerisms,Waxy flexibility,Withdrawal | 92.1 | 89.7 | 94.6 |
| 6 | Excitement,Mutism,Posturing/catalepsy,Stereotypy,Mannerisms,Rigidity | 92.1 | 89.5 | 94.4 |
| 6 | Immobility/stupor,Mutism,Echopraxia/echolalia,Stereotypy,Mannerisms,Waxy flexibility | 92.1 | 89.7 | 94.6 |
| 6 | Immobility/stupor,Mutism,Grimacing,Mannerisms,Verbigeration,Waxy flexibility | 92.1 | 89.5 | 94.6 |
| 6 | Immobility/stupor,Mutism,Grimacing,Stereotypy,Mannerisms,Waxy flexibility | 92.1 | 89.5 | 94.6 |
| 6 | Immobility/stupor,Mutism,Posturing/catalepsy,Echopraxia/echolalia,Mannerisms,Waxy flexibility | 92.1 | 89.5 | 94.6 |
| 6 | Immobility/stupor,Mutism,Posturing/catalepsy,Grimacing,Echopraxia/echolalia,Waxy flexibility | 92.1 | 89.5 | 94.6 |
| 6 | Immobility/stupor,Posturing/catalepsy,Grimacing,Echopraxia/echolalia,Stereotypy,Verbigeration | 92.1 | 89.5 | 94.4 |
| 6 | Mutism,Posturing/catalepsy,Echopraxia/echolalia,Stereotypy,Negativism,Withdrawal | 92.1 | 89.7 | 94.6 |
| 6 | Posturing/catalepsy,Echopraxia/echolalia,Stereotypy,Rigidity,Negativism,Withdrawal | 92.1 | 89.7 | 94.6 |
| 6 | Immobility/stupor,Mutism,Posturing/catalepsy,Echopraxia/echolalia,Stereotypy,Waxy flexibility | 92.0 | 89.2 | 94.4 |
| 6 | Mutism,Posturing/catalepsy,Grimacing,Verbigeration,Negativism,Withdrawal | 92.0 | 89.2 | 94.4 |
| 6 | Excitement,Mutism,Grimacing,Stereotypy,Rigidity,Waxy flexibility | 91.9 | 89.5 | 94.4 |
| 6 | Excitement,Mutism,Grimacing,Verbigeration,Rigidity,Waxy flexibility | 91.9 | 89.2 | 94.4 |
| 6 | Excitement,Mutism,Posturing/catalepsy,Echopraxia/echolalia,Mannerisms,Negativism | 91.9 | 89.2 | 94.4 |
| 6 | Excitement,Mutism,Posturing/catalepsy,Echopraxia/echolalia,Stereotypy,Withdrawal | 91.9 | 89.2 | 94.4 |
| 6 | Excitement,Mutism,Posturing/catalepsy,Grimacing,Negativism,Waxy flexibility | 91.9 | 89.2 | 94.4 |
| 6 | Excitement,Mutism,Posturing/catalepsy,Grimacing,Stereotypy,Negativism | 91.9 | 89.2 | 94.4 |
| 6 | Excitement,Mutism,Posturing/catalepsy,Grimacing,Verbigeration,Withdrawal | 91.9 | 89.5 | 94.4 |
| 6 | Excitement,Mutism,Posturing/catalepsy,Mannerisms,Verbigeration,Negativism | 91.9 | 89.2 | 94.4 |
| 6 | Excitement,Mutism,Posturing/catalepsy,Mannerisms,Verbigeration,Withdrawal | 91.9 | 89.2 | 94.4 |
| 6 | Excitement,Mutism,Posturing/catalepsy,Stereotypy,Verbigeration,Withdrawal | 91.9 | 89.2 | 94.4 |
| 6 | Excitement,Posturing/catalepsy,Echopraxia/echolalia,Rigidity,Waxy flexibility,Withdrawal | 91.9 | 89.2 | 94.4 |
| 6 | Excitement,Posturing/catalepsy,Echopraxia/echolalia,Verbigeration,Rigidity,Withdrawal | 91.9 | 89.2 | 94.4 |
| 6 | Excitement,Posturing/catalepsy,Verbigeration,Rigidity,Waxy flexibility,Withdrawal | 91.9 | 89.5 | 94.4 |
| 6 | Immobility/stupor,Grimacing,Echopraxia/echolalia,Verbigeration,Waxy flexibility,Withdrawal | 91.9 | 89.2 | 94.4 |
| 6 | Immobility/stupor,Grimacing,Mannerisms,Verbigeration,Waxy flexibility,Withdrawal | 91.9 | 89.5 | 94.4 |
| 6 | Immobility/stupor,Mutism,Echopraxia/echolalia,Mannerisms,Verbigeration,Waxy flexibility | 91.9 | 89.2 | 94.4 |
| 6 | Immobility/stupor,Mutism,Echopraxia/echolalia,Stereotypy,Mannerisms,Verbigeration | 91.9 | 89.5 | 94.4 |
| 6 | Immobility/stupor,Mutism,Posturing/catalepsy,Echopraxia/echolalia,Verbigeration,Waxy flexibility | 91.9 | 89.2 | 94.4 |
| 6 | Immobility/stupor,Mutism,Posturing/catalepsy,Grimacing,Stereotypy,Waxy flexibility | 91.9 | 89.2 | 94.4 |
| 6 | Immobility/stupor,Mutism,Posturing/catalepsy,Mannerisms,Verbigeration,Waxy flexibility | 91.9 | 89.2 | 94.4 |
| 6 | Immobility/stupor,Posturing/catalepsy,Echopraxia/echolalia,Verbigeration,Rigidity,Waxy flexibility | 91.9 | 89.2 | 94.4 |
| 6 | Immobility/stupor,Posturing/catalepsy,Grimacing,Verbigeration,Waxy flexibility,Withdrawal | 91.9 | 89.2 | 94.4 |
| 6 | Immobility/stupor,Posturing/catalepsy,Stereotypy,Verbigeration,Waxy flexibility,Withdrawal | 91.9 | 89.2 | 94.4 |
| 6 | Mutism,Echopraxia/echolalia,Stereotypy,Rigidity,Negativism,Waxy flexibility | 91.9 | 89.2 | 94.4 |
| 6 | Mutism,Grimacing,Echopraxia/echolalia,Verbigeration,Rigidity,Negativism | 91.9 | 89.2 | 94.4 |
| 6 | Mutism,Grimacing,Stereotypy,Rigidity,Negativism,Waxy flexibility | 91.9 | 89.2 | 94.4 |
| 6 | Mutism,Grimacing,Stereotypy,Rigidity,Waxy flexibility,Withdrawal | 91.9 | 89.2 | 94.4 |
| 6 | Mutism,Grimacing,Verbigeration,Rigidity,Negativism,Waxy flexibility | 91.9 | 89.2 | 94.4 |
| 6 | Mutism,Grimacing,Verbigeration,Rigidity,Waxy flexibility,Withdrawal | 91.9 | 89.5 | 94.4 |
| 6 | Mutism,Posturing/catalepsy,Echopraxia/echolalia,Verbigeration,Negativism,Withdrawal | 91.9 | 89.2 | 94.4 |
| 6 | Mutism,Posturing/catalepsy,Grimacing,Echopraxia/echolalia,Negativism,Withdrawal | 91.9 | 89.2 | 94.4 |
| 6 | Mutism,Posturing/catalepsy,Grimacing,Stereotypy,Rigidity,Withdrawal | 91.9 | 89.5 | 94.4 |
| 6 | Mutism,Posturing/catalepsy,Grimacing,Verbigeration,Rigidity,Withdrawal | 91.9 | 89.2 | 94.4 |
| 6 | Mutism,Posturing/catalepsy,Stereotypy,Verbigeration,Rigidity,Withdrawal | 91.9 | 89.5 | 94.4 |
| 6 | Mutism,Posturing/catalepsy,Verbigeration,Rigidity,Waxy flexibility,Withdrawal | 91.9 | 89.2 | 94.4 |
| 6 | Posturing/catalepsy,Echopraxia/echolalia,Verbigeration,Rigidity,Negativism,Withdrawal | 91.9 | 89.2 | 94.4 |
| 6 | Posturing/catalepsy,Stereotypy,Rigidity,Negativism,Waxy flexibility,Withdrawal | 91.9 | 89.2 | 94.4 |
| 6 | Excitement,Mutism,Echopraxia/echolalia,Verbigeration,Rigidity,Waxy flexibility | 91.7 | 89.0 | 94.2 |
| 6 | Excitement,Mutism,Posturing/catalepsy,Echopraxia/echolalia,Mannerisms,Withdrawal | 91.7 | 89.2 | 94.2 |
| 6 | Excitement,Mutism,Posturing/catalepsy,Grimacing,Mannerisms,Negativism | 91.7 | 89.0 | 94.2 |
| 6 | Excitement,Mutism,Posturing/catalepsy,Grimacing,Mannerisms,Withdrawal | 91.7 | 89.0 | 94.2 |
| 6 | Excitement,Mutism,Posturing/catalepsy,Grimacing,Stereotypy,Withdrawal | 91.7 | 89.0 | 94.2 |
| 6 | Excitement,Mutism,Posturing/catalepsy,Stereotypy,Negativism,Waxy flexibility | 91.7 | 89.0 | 94.2 |
| 6 | Excitement,Posturing/catalepsy,Mannerisms,Verbigeration,Rigidity,Withdrawal | 91.7 | 89.0 | 94.2 |
| 6 | Excitement,Posturing/catalepsy,Stereotypy,Verbigeration,Negativism,Withdrawal | 91.7 | 89.0 | 94.2 |
| 6 | Immobility/stupor,Echopraxia/echolalia,Stereotypy,Verbigeration,Waxy flexibility,Withdrawal | 91.7 | 89.0 | 94.2 |
| 6 | Immobility/stupor,Grimacing,Echopraxia/echolalia,Stereotypy,Verbigeration,Waxy flexibility | 91.7 | 89.2 | 94.2 |
| 6 | Immobility/stupor,Mutism,Echopraxia/echolalia,Stereotypy,Verbigeration,Waxy flexibility | 91.7 | 89.0 | 94.2 |
| 6 | Immobility/stupor,Mutism,Stereotypy,Mannerisms,Verbigeration,Waxy flexibility | 91.7 | 89.0 | 94.2 |
| 6 | Immobility/stupor,Posturing/catalepsy,Grimacing,Echopraxia/echolalia,Mannerisms,Waxy flexibility | 91.7 | 89.0 | 94.2 |
| 6 | Immobility/stupor,Posturing/catalepsy,Grimacing,Echopraxia/echolalia,Stereotypy,Waxy flexibility | 91.7 | 89.0 | 94.2 |
| 6 | Immobility/stupor,Posturing/catalepsy,Grimacing,Stereotypy,Verbigeration,Waxy flexibility | 91.7 | 89.0 | 94.2 |
| 6 | Immobility/stupor,Stereotypy,Mannerisms,Verbigeration,Waxy flexibility,Withdrawal | 91.7 | 89.0 | 94.2 |
| 6 | Mutism,Echopraxia/echolalia,Verbigeration,Rigidity,Negativism,Waxy flexibility | 91.7 | 89.0 | 94.2 |
| 6 | Mutism,Grimacing,Echopraxia/echolalia,Stereotypy,Rigidity,Negativism | 91.7 | 89.0 | 94.2 |
| 6 | Mutism,Mannerisms,Verbigeration,Rigidity,Waxy flexibility,Withdrawal | 91.7 | 89.0 | 94.2 |
| 6 | Mutism,Posturing/catalepsy,Echopraxia/echolalia,Stereotypy,Rigidity,Withdrawal | 91.7 | 89.0 | 94.2 |
| 6 | Mutism,Posturing/catalepsy,Grimacing,Echopraxia/echolalia,Mannerisms,Negativism | 91.7 | 89.0 | 94.2 |
| 6 | Mutism,Posturing/catalepsy,Grimacing,Echopraxia/echolalia,Rigidity,Withdrawal | 91.7 | 89.0 | 94.2 |
| 6 | Mutism,Posturing/catalepsy,Stereotypy,Negativism,Waxy flexibility,Withdrawal | 91.7 | 89.0 | 94.2 |
| 6 | Mutism,Posturing/catalepsy,Stereotypy,Rigidity,Waxy flexibility,Withdrawal | 91.7 | 89.0 | 94.2 |
| 6 | Mutism,Posturing/catalepsy,Verbigeration,Negativism,Waxy flexibility,Withdrawal | 91.7 | 89.0 | 94.2 |
| 6 | Posturing/catalepsy,Grimacing,Verbigeration,Rigidity,Negativism,Withdrawal | 91.7 | 89.0 | 93.9 |
| 6 | Posturing/catalepsy,Verbigeration,Rigidity,Negativism,Waxy flexibility,Withdrawal | 91.7 | 89.0 | 94.2 |
| 6 | Excitement,Mutism,Echopraxia/echolalia,Negativism,Waxy flexibility,Withdrawal | 91.5 | 88.8 | 93.9 |
| 6 | Excitement,Mutism,Posturing/catalepsy,Stereotypy,Mannerisms,Negativism | 91.5 | 88.8 | 93.9 |
| 6 | Excitement,Mutism,Posturing/catalepsy,Stereotypy,Mannerisms,Withdrawal | 91.5 | 88.8 | 93.9 |
| 6 | Excitement,Posturing/catalepsy,Echopraxia/echolalia,Mannerisms,Rigidity,Withdrawal | 91.5 | 88.8 | 93.9 |
| 6 | Excitement,Posturing/catalepsy,Grimacing,Echopraxia/echolalia,Rigidity,Withdrawal | 91.5 | 88.8 | 93.9 |
| 6 | Excitement,Posturing/catalepsy,Mannerisms,Rigidity,Waxy flexibility,Withdrawal | 91.5 | 88.8 | 93.9 |
| 6 | Excitement,Posturing/catalepsy,Stereotypy,Rigidity,Waxy flexibility,Withdrawal | 91.5 | 88.8 | 93.9 |
| 6 | Immobility/stupor,Echopraxia/echolalia,Mannerisms,Verbigeration,Waxy flexibility,Withdrawal | 91.5 | 88.8 | 93.9 |
| 6 | Immobility/stupor,Grimacing,Echopraxia/echolalia,Stereotypy,Mannerisms,Verbigeration | 91.5 | 89.0 | 93.9 |
| 6 | Immobility/stupor,Grimacing,Stereotypy,Mannerisms,Verbigeration,Waxy flexibility | 91.5 | 88.8 | 93.9 |
| 6 | Immobility/stupor,Mutism,Posturing/catalepsy,Stereotypy,Mannerisms,Waxy flexibility | 91.5 | 88.8 | 93.9 |
| 6 | Immobility/stupor,Posturing/catalepsy,Echopraxia/echolalia,Stereotypy,Mannerisms,Verbigeration | 91.5 | 88.8 | 93.9 |
| 6 | Immobility/stupor,Posturing/catalepsy,Echopraxia/echolalia,Verbigeration,Waxy flexibility,Withdrawal | 91.5 | 88.8 | 93.9 |
| 6 | Immobility/stupor,Posturing/catalepsy,Grimacing,Echopraxia/echolalia,Verbigeration,Waxy flexibility | 91.5 | 88.8 | 93.9 |
| 6 | Immobility/stupor,Posturing/catalepsy,Grimacing,Mannerisms,Verbigeration,Waxy flexibility | 91.5 | 88.8 | 93.9 |
| 6 | Immobility/stupor,Posturing/catalepsy,Grimacing,Stereotypy,Mannerisms,Waxy flexibility | 91.5 | 88.8 | 93.9 |
| 6 | Mutism,Echopraxia/echolalia,Mannerisms,Rigidity,Waxy flexibility,Withdrawal | 91.5 | 88.8 | 93.9 |
| 6 | Mutism,Echopraxia/echolalia,Stereotypy,Mannerisms,Negativism,Withdrawal | 91.5 | 88.8 | 93.9 |
| 6 | Mutism,Echopraxia/echolalia,Stereotypy,Rigidity,Waxy flexibility,Withdrawal | 91.5 | 88.8 | 93.9 |
| 6 | Mutism,Grimacing,Echopraxia/echolalia,Mannerisms,Rigidity,Withdrawal | 91.5 | 88.8 | 93.9 |
| 6 | Mutism,Grimacing,Echopraxia/echolalia,Rigidity,Negativism,Waxy flexibility | 91.5 | 88.8 | 93.9 |
| 6 | Mutism,Grimacing,Echopraxia/echolalia,Rigidity,Waxy flexibility,Withdrawal | 91.5 | 88.8 | 93.9 |
| 6 | Mutism,Grimacing,Mannerisms,Verbigeration,Rigidity,Withdrawal | 91.5 | 88.8 | 93.9 |
| 6 | Mutism,Grimacing,Stereotypy,Mannerisms,Rigidity,Withdrawal | 91.5 | 88.8 | 93.9 |
| 6 | Mutism,Grimacing,Stereotypy,Verbigeration,Rigidity,Withdrawal | 91.5 | 88.8 | 93.9 |
| 6 | Mutism,Posturing/catalepsy,Echopraxia/echolalia,Negativism,Waxy flexibility,Withdrawal | 91.5 | 88.8 | 93.9 |
| 6 | Mutism,Posturing/catalepsy,Echopraxia/echolalia,Rigidity,Waxy flexibility,Withdrawal | 91.5 | 88.8 | 93.9 |
| 6 | Mutism,Posturing/catalepsy,Echopraxia/echolalia,Verbigeration,Rigidity,Withdrawal | 91.5 | 88.8 | 93.9 |
| 6 | Mutism,Posturing/catalepsy,Grimacing,Rigidity,Negativism,Waxy flexibility | 91.5 | 88.8 | 93.9 |
| 6 | Mutism,Posturing/catalepsy,Grimacing,Rigidity,Waxy flexibility,Withdrawal | 91.5 | 88.8 | 93.9 |
| 6 | Mutism,Stereotypy,Mannerisms,Negativism,Waxy flexibility,Withdrawal | 91.5 | 88.8 | 93.9 |
| 6 | Posturing/catalepsy,Grimacing,Echopraxia/echolalia,Rigidity,Negativism,Withdrawal | 91.5 | 88.8 | 93.9 |
| 6 | Excitement,Mutism,Grimacing,Stereotypy,Negativism,Withdrawal | 91.3 | 88.6 | 93.7 |
| 6 | Excitement,Mutism,Posturing/catalepsy,Mannerisms,Negativism,Waxy flexibility | 91.3 | 88.6 | 93.7 |
| 6 | Excitement,Mutism,Stereotypy,Mannerisms,Rigidity,Waxy flexibility | 91.3 | 88.6 | 93.7 |
| 6 | Excitement,Mutism,Verbigeration,Negativism,Waxy flexibility,Withdrawal | 91.3 | 88.6 | 93.7 |
| 6 | Excitement,Posturing/catalepsy,Echopraxia/echolalia,Stereotypy,Negativism,Withdrawal | 91.3 | 88.6 | 93.7 |
| 6 | Excitement,Posturing/catalepsy,Echopraxia/echolalia,Verbigeration,Negativism,Withdrawal | 91.3 | 88.6 | 93.7 |
| 6 | Excitement,Posturing/catalepsy,Grimacing,Rigidity,Waxy flexibility,Withdrawal | 91.3 | 88.6 | 93.7 |
| 6 | Excitement,Posturing/catalepsy,Grimacing,Verbigeration,Rigidity,Withdrawal | 91.3 | 88.6 | 93.7 |
| 6 | Excitement,Posturing/catalepsy,Mannerisms,Verbigeration,Negativism,Withdrawal | 91.3 | 88.6 | 93.7 |
| 6 | Excitement,Posturing/catalepsy,Stereotypy,Verbigeration,Rigidity,Withdrawal | 91.3 | 88.6 | 93.7 |
| 6 | Mutism,Posturing/catalepsy,Echopraxia/echolalia,Mannerisms,Verbigeration,Negativism | 91.3 | 88.6 | 93.7 |
| 6 | Mutism,Posturing/catalepsy,Echopraxia/echolalia,Stereotypy,Mannerisms,Negativism | 91.3 | 88.6 | 93.9 |
| 6 | Mutism,Posturing/catalepsy,Grimacing,Echopraxia/echolalia,Mannerisms,Rigidity | 91.3 | 88.6 | 93.7 |
| 6 | Mutism,Posturing/catalepsy,Grimacing,Mannerisms,Verbigeration,Rigidity | 91.3 | 88.6 | 93.7 |
| 6 | Mutism,Posturing/catalepsy,Stereotypy,Mannerisms,Verbigeration,Negativism | 91.3 | 88.6 | 93.7 |
| 6 | Mutism,Stereotypy,Mannerisms,Verbigeration,Negativism,Withdrawal | 91.3 | 88.6 | 93.7 |
| 6 | Posturing/catalepsy,Stereotypy,Mannerisms,Verbigeration,Negativism,Withdrawal | 91.3 | 88.6 | 93.7 |
| 6 | Echopraxia/echolalia,Stereotypy,Mannerisms,Rigidity,Negativism,Withdrawal | 91.2 | 88.6 | 93.7 |
| 6 | Excitement,Mutism,Echopraxia/echolalia,Verbigeration,Negativism,Withdrawal | 91.2 | 88.6 | 93.9 |
| 6 | Excitement,Posturing/catalepsy,Echopraxia/echolalia,Stereotypy,Rigidity,Withdrawal | 91.2 | 88.6 | 93.7 |
| 6 | Excitement,Posturing/catalepsy,Grimacing,Echopraxia/echolalia,Negativism,Withdrawal | 91.2 | 88.6 | 93.7 |
| 6 | Immobility/stupor,Grimacing,Echopraxia/echolalia,Mannerisms,Verbigeration,Waxy flexibility | 91.2 | 88.6 | 93.7 |
| 6 | Mutism,Posturing/catalepsy,Grimacing,Negativism,Waxy flexibility,Withdrawal | 91.2 | 88.6 | 93.7 |
| 6 | Mutism,Posturing/catalepsy,Grimacing,Stereotypy,Verbigeration,Rigidity | 91.2 | 88.6 | 93.7 |
| 6 | Mutism,Grimacing,Echopraxia/echolalia,Stereotypy,Rigidity,Withdrawal | 91.1 | 88.3 | 93.5 |
| 6 | Mutism,Posturing/catalepsy,Grimacing,Mannerisms,Verbigeration,Negativism | 91.1 | 88.3 | 93.5 |
| 6 | Mutism,Stereotypy,Mannerisms,Verbigeration,Rigidity,Withdrawal | 91.1 | 88.3 | 93.5 |
| 6 | Posturing/catalepsy,Echopraxia/echolalia,Stereotypy,Mannerisms,Negativism,Withdrawal | 91.1 | 88.3 | 93.7 |
| 6 | Excitement,Echopraxia/echolalia,Stereotypy,Rigidity,Negativism,Withdrawal | 91.0 | 88.3 | 93.5 |
| 6 | Excitement,Mutism,Echopraxia/echolalia,Stereotypy,Verbigeration,Rigidity | 91.0 | 88.3 | 93.5 |
| 6 | Excitement,Mutism,Grimacing,Mannerisms,Rigidity,Waxy flexibility | 91.0 | 88.3 | 93.5 |
| 6 | Excitement,Mutism,Grimacing,Negativism,Waxy flexibility,Withdrawal | 91.0 | 88.3 | 93.5 |
| 6 | Excitement,Mutism,Stereotypy,Mannerisms,Negativism,Withdrawal | 91.0 | 88.3 | 93.5 |
| 6 | Excitement,Posturing/catalepsy,Grimacing,Mannerisms,Rigidity,Withdrawal | 91.0 | 88.1 | 93.5 |
| 6 | Excitement,Posturing/catalepsy,Grimacing,Stereotypy,Negativism,Withdrawal | 91.0 | 88.3 | 93.5 |
| 6 | Excitement,Posturing/catalepsy,Grimacing,Verbigeration,Negativism,Withdrawal | 91.0 | 88.3 | 93.7 |
| 6 | Excitement,Posturing/catalepsy,Stereotypy,Mannerisms,Negativism,Withdrawal | 91.0 | 88.3 | 93.5 |
| 6 | Excitement,Posturing/catalepsy,Stereotypy,Mannerisms,Rigidity,Withdrawal | 91.0 | 88.3 | 93.5 |
| 6 | Excitement,Posturing/catalepsy,Stereotypy,Negativism,Waxy flexibility,Withdrawal | 91.0 | 88.3 | 93.7 |
| 6 | Mutism,Echopraxia/echolalia,Stereotypy,Mannerisms,Rigidity,Withdrawal | 91.0 | 88.1 | 93.7 |
| 6 | Mutism,Posturing/catalepsy,Echopraxia/echolalia,Mannerisms,Verbigeration,Rigidity | 91.0 | 88.3 | 93.5 |
| 6 | Mutism,Posturing/catalepsy,Grimacing,Echopraxia/echolalia,Verbigeration,Negativism | 91.0 | 88.3 | 93.5 |
| 6 | Mutism,Posturing/catalepsy,Grimacing,Stereotypy,Mannerisms,Negativism | 91.0 | 88.1 | 93.7 |
| 6 | Mutism,Posturing/catalepsy,Grimacing,Stereotypy,Verbigeration,Negativism | 91.0 | 88.3 | 93.5 |
| 6 | Posturing/catalepsy,Echopraxia/echolalia,Rigidity,Negativism,Waxy flexibility,Withdrawal | 91.0 | 88.3 | 93.5 |
| 6 | Posturing/catalepsy,Grimacing,Stereotypy,Mannerisms,Negativism,Withdrawal | 91.0 | 88.3 | 93.7 |
| 6 | Stereotypy,Mannerisms,Rigidity,Negativism,Waxy flexibility,Withdrawal | 91.0 | 88.3 | 93.5 |
| 6 | Stereotypy,Mannerisms,Verbigeration,Rigidity,Negativism,Withdrawal | 91.0 | 88.3 | 93.7 |
| 6 | Excitement,Mutism,Echopraxia/echolalia,Mannerisms,Rigidity,Waxy flexibility | 90.8 | 88.1 | 93.3 |
| 6 | Excitement,Mutism,Echopraxia/echolalia,Stereotypy,Waxy flexibility,Withdrawal | 90.8 | 88.1 | 93.3 |
| 6 | Excitement,Mutism,Grimacing,Echopraxia/echolalia,Negativism,Withdrawal | 90.8 | 88.1 | 93.5 |
| 6 | Excitement,Mutism,Grimacing,Echopraxia/echolalia,Stereotypy,Rigidity | 90.8 | 88.1 | 93.3 |
| 6 | Excitement,Mutism,Grimacing,Echopraxia/echolalia,Verbigeration,Rigidity | 90.8 | 87.9 | 93.5 |
| 6 | Excitement,Mutism,Grimacing,Stereotypy,Verbigeration,Rigidity | 90.8 | 88.1 | 93.3 |
| 6 | Excitement,Mutism,Grimacing,Verbigeration,Negativism,Withdrawal | 90.8 | 88.1 | 93.5 |
| 6 | Excitement,Mutism,Mannerisms,Negativism,Waxy flexibility,Withdrawal | 90.8 | 88.1 | 93.5 |
| 6 | Excitement,Posturing/catalepsy,Echopraxia/echolalia,Mannerisms,Negativism,Withdrawal | 90.8 | 88.1 | 93.5 |
| 6 | Excitement,Posturing/catalepsy,Echopraxia/echolalia,Negativism,Waxy flexibility,Withdrawal | 90.8 | 87.9 | 93.5 |
| 6 | Excitement,Posturing/catalepsy,Grimacing,Mannerisms,Negativism,Withdrawal | 90.8 | 88.1 | 93.5 |
| 6 | Excitement,Posturing/catalepsy,Grimacing,Stereotypy,Rigidity,Withdrawal | 90.8 | 88.1 | 93.5 |
| 6 | Excitement,Posturing/catalepsy,Verbigeration,Negativism,Waxy flexibility,Withdrawal | 90.8 | 88.1 | 93.5 |
| 6 | Excitement,Stereotypy,Verbigeration,Rigidity,Negativism,Withdrawal | 90.8 | 88.1 | 93.5 |
| 6 | Grimacing,Stereotypy,Mannerisms,Rigidity,Negativism,Withdrawal | 90.8 | 88.1 | 93.3 |
| 6 | Immobility/stupor,Echopraxia/echolalia,Stereotypy,Mannerisms,Verbigeration,Waxy flexibility | 90.8 | 88.1 | 93.5 |
| 6 | Immobility/stupor,Posturing/catalepsy,Echopraxia/echolalia,Stereotypy,Mannerisms,Waxy flexibility | 90.8 | 87.9 | 93.3 |
| 6 | Immobility/stupor,Posturing/catalepsy,Echopraxia/echolalia,Stereotypy,Verbigeration,Waxy flexibility | 90.8 | 88.1 | 93.3 |
| 6 | Immobility/stupor,Posturing/catalepsy,Stereotypy,Mannerisms,Verbigeration,Waxy flexibility | 90.8 | 88.1 | 93.3 |
| 6 | Mutism,Echopraxia/echolalia,Mannerisms,Negativism,Waxy flexibility,Withdrawal | 90.8 | 88.1 | 93.3 |
| 6 | Mutism,Echopraxia/echolalia,Stereotypy,Negativism,Waxy flexibility,Withdrawal | 90.8 | 88.1 | 93.5 |
| 6 | Mutism,Echopraxia/echolalia,Stereotypy,Verbigeration,Negativism,Withdrawal | 90.8 | 88.1 | 93.5 |
| 6 | Mutism,Echopraxia/echolalia,Verbigeration,Rigidity,Waxy flexibility,Withdrawal | 90.8 | 88.1 | 93.5 |
| 6 | Mutism,Grimacing,Stereotypy,Mannerisms,Negativism,Withdrawal | 90.8 | 88.1 | 93.3 |
| 6 | Mutism,Posturing/catalepsy,Echopraxia/echolalia,Mannerisms,Negativism,Waxy flexibility | 90.8 | 88.1 | 93.3 |
| 6 | Mutism,Posturing/catalepsy,Echopraxia/echolalia,Mannerisms,Rigidity,Waxy flexibility | 90.8 | 88.1 | 93.5 |
| 6 | Mutism,Posturing/catalepsy,Echopraxia/echolalia,Stereotypy,Verbigeration,Negativism | 90.8 | 88.1 | 93.5 |
| 6 | Mutism,Posturing/catalepsy,Grimacing,Echopraxia/echolalia,Verbigeration,Rigidity | 90.8 | 88.1 | 93.5 |
| 6 | Mutism,Posturing/catalepsy,Grimacing,Mannerisms,Rigidity,Waxy flexibility | 90.8 | 87.9 | 93.5 |
| 6 | Mutism,Posturing/catalepsy,Grimacing,Stereotypy,Mannerisms,Rigidity | 90.8 | 88.1 | 93.5 |
| 6 | Mutism,Posturing/catalepsy,Grimacing,Verbigeration,Rigidity,Waxy flexibility | 90.8 | 88.1 | 93.3 |
| 6 | Mutism,Posturing/catalepsy,Mannerisms,Verbigeration,Rigidity,Waxy flexibility | 90.8 | 88.1 | 93.3 |
| 6 | Mutism,Posturing/catalepsy,Stereotypy,Mannerisms,Verbigeration,Rigidity | 90.8 | 88.1 | 93.5 |
| 6 | Mutism,Posturing/catalepsy,Stereotypy,Verbigeration,Rigidity,Waxy flexibility | 90.8 | 88.1 | 93.5 |
| 6 | Mutism,Stereotypy,Verbigeration,Negativism,Waxy flexibility,Withdrawal | 90.8 | 88.1 | 93.5 |
| 6 | Posturing/catalepsy,Grimacing,Rigidity,Negativism,Waxy flexibility,Withdrawal | 90.8 | 87.9 | 93.3 |
| 6 | Posturing/catalepsy,Stereotypy,Mannerisms,Negativism,Waxy flexibility,Withdrawal | 90.8 | 87.9 | 93.5 |
| 6 | Excitement,Mutism,Mannerisms,Verbigeration,Rigidity,Waxy flexibility | 90.6 | 87.9 | 93.3 |
| 6 | Excitement,Mutism,Stereotypy,Verbigeration,Waxy flexibility,Withdrawal | 90.6 | 87.7 | 93.3 |
| 6 | Excitement,Stereotypy,Rigidity,Negativism,Waxy flexibility,Withdrawal | 90.6 | 87.7 | 93.3 |
| 6 | Mutism,Echopraxia/echolalia,Stereotypy,Verbigeration,Rigidity,Withdrawal | 90.6 | 87.9 | 93.3 |
| 6 | Mutism,Grimacing,Echopraxia/echolalia,Stereotypy,Negativism,Withdrawal | 90.6 | 87.9 | 93.3 |
| 6 | Mutism,Grimacing,Echopraxia/echolalia,Verbigeration,Rigidity,Withdrawal | 90.6 | 87.9 | 93.0 |
| 6 | Mutism,Grimacing,Mannerisms,Negativism,Waxy flexibility,Withdrawal | 90.6 | 87.7 | 93.3 |
| 6 | Mutism,Grimacing,Stereotypy,Verbigeration,Negativism,Withdrawal | 90.6 | 87.7 | 93.3 |
| 6 | Mutism,Posturing/catalepsy,Echopraxia/echolalia,Stereotypy,Mannerisms,Rigidity | 90.6 | 87.9 | 93.3 |
| 6 | Mutism,Posturing/catalepsy,Echopraxia/echolalia,Stereotypy,Verbigeration,Rigidity | 90.6 | 87.9 | 93.3 |
| 6 | Mutism,Posturing/catalepsy,Grimacing,Echopraxia/echolalia,Mannerisms,Withdrawal | 90.6 | 87.7 | 93.3 |
| 6 | Mutism,Posturing/catalepsy,Grimacing,Echopraxia/echolalia,Stereotypy,Negativism | 90.6 | 87.9 | 93.3 |
| 6 | Mutism,Posturing/catalepsy,Grimacing,Mannerisms,Negativism,Waxy flexibility | 90.6 | 87.9 | 93.3 |
| 6 | Mutism,Posturing/catalepsy,Grimacing,Mannerisms,Waxy flexibility,Withdrawal | 90.6 | 87.9 | 93.3 |
| 6 | Mutism,Posturing/catalepsy,Stereotypy,Mannerisms,Negativism,Waxy flexibility | 90.6 | 87.9 | 93.0 |
| 6 | Mutism,Posturing/catalepsy,Stereotypy,Verbigeration,Negativism,Waxy flexibility | 90.6 | 87.9 | 93.3 |
| 6 | Posturing/catalepsy,Grimacing,Stereotypy,Verbigeration,Negativism,Withdrawal | 90.6 | 87.7 | 93.3 |
| 6 | Stereotypy,Verbigeration,Rigidity,Negativism,Waxy flexibility,Withdrawal | 90.6 | 87.7 | 93.3 |
| 6 | Echopraxia/echolalia,Stereotypy,Verbigeration,Rigidity,Negativism,Withdrawal | 90.4 | 87.7 | 93.0 |
| 6 | Excitement,Grimacing,Echopraxia/echolalia,Rigidity,Negativism,Withdrawal | 90.4 | 87.7 | 93.0 |
| 6 | Excitement,Mutism,Echopraxia/echolalia,Mannerisms,Negativism,Withdrawal | 90.4 | 87.4 | 93.0 |
| 6 | Excitement,Mutism,Echopraxia/echolalia,Stereotypy,Mannerisms,Rigidity | 90.4 | 87.4 | 93.0 |
| 6 | Excitement,Mutism,Echopraxia/echolalia,Stereotypy,Negativism,Waxy flexibility | 90.4 | 87.7 | 93.0 |
| 6 | Excitement,Mutism,Mannerisms,Verbigeration,Negativism,Withdrawal | 90.4 | 87.4 | 93.0 |
| 6 | Excitement,Mutism,Stereotypy,Verbigeration,Negativism,Waxy flexibility | 90.4 | 87.7 | 93.0 |
| 6 | Excitement,Posturing/catalepsy,Mannerisms,Negativism,Waxy flexibility,Withdrawal | 90.4 | 87.4 | 93.0 |
| 6 | Grimacing,Stereotypy,Verbigeration,Rigidity,Negativism,Withdrawal | 90.4 | 87.7 | 93.0 |
| 6 | Mutism,Echopraxia/echolalia,Mannerisms,Verbigeration,Rigidity,Withdrawal | 90.4 | 87.4 | 93.0 |
| 6 | Mutism,Grimacing,Echopraxia/echolalia,Mannerisms,Negativism,Withdrawal | 90.4 | 87.4 | 93.0 |
| 6 | Mutism,Grimacing,Stereotypy,Verbigeration,Rigidity,Waxy flexibility | 90.4 | 87.7 | 93.0 |
| 6 | Mutism,Mannerisms,Verbigeration,Negativism,Waxy flexibility,Withdrawal | 90.4 | 87.4 | 93.0 |
| 6 | Mutism,Posturing/catalepsy,Echopraxia/echolalia,Verbigeration,Negativism,Waxy flexibility | 90.4 | 87.4 | 92.8 |
| 6 | Mutism,Posturing/catalepsy,Grimacing,Mannerisms,Verbigeration,Withdrawal | 90.4 | 87.4 | 93.0 |
| 6 | Mutism,Posturing/catalepsy,Grimacing,Stereotypy,Mannerisms,Withdrawal | 90.4 | 87.4 | 93.0 |
| 6 | Mutism,Posturing/catalepsy,Grimacing,Verbigeration,Negativism,Waxy flexibility | 90.4 | 87.4 | 93.0 |
| 6 | Mutism,Posturing/catalepsy,Mannerisms,Verbigeration,Negativism,Waxy flexibility | 90.4 | 87.4 | 93.0 |
| 6 | Posturing/catalepsy,Grimacing,Echopraxia/echolalia,Mannerisms,Negativism,Withdrawal | 90.4 | 87.4 | 93.0 |
| 6 | Posturing/catalepsy,Grimacing,Echopraxia/echolalia,Stereotypy,Negativism,Withdrawal | 90.4 | 87.4 | 93.0 |
| 6 | Posturing/catalepsy,Stereotypy,Verbigeration,Negativism,Waxy flexibility,Withdrawal | 90.4 | 87.4 | 93.0 |
| 6 | Excitement,Grimacing,Stereotypy,Rigidity,Negativism,Withdrawal | 90.3 | 87.4 | 93.0 |
| 6 | Immobility/stupor,Posturing/catalepsy,Echopraxia/echolalia,Mannerisms,Verbigeration,Waxy flexibility | 90.3 | 87.4 | 93.0 |
| 6 | Mutism,Echopraxia/echolalia,Mannerisms,Verbigeration,Negativism,Withdrawal | 90.3 | 87.4 | 93.0 |
| 6 | Mutism,Posturing/catalepsy,Echopraxia/echolalia,Verbigeration,Rigidity,Waxy flexibility | 90.3 | 87.7 | 93.0 |
| 6 | Mutism,Posturing/catalepsy,Grimacing,Echopraxia/echolalia,Stereotypy,Rigidity | 90.3 | 87.4 | 93.0 |
| 6 | Posturing/catalepsy,Grimacing,Mannerisms,Verbigeration,Negativism,Withdrawal | 90.3 | 87.7 | 93.0 |
| 6 | Excitement,Echopraxia/echolalia,Rigidity,Negativism,Waxy flexibility,Withdrawal | 90.2 | 87.2 | 92.8 |
| 6 | Grimacing,Echopraxia/echolalia,Stereotypy,Rigidity,Negativism,Withdrawal | 90.2 | 87.2 | 92.8 |
| 6 | Mutism,Posturing/catalepsy,Stereotypy,Mannerisms,Rigidity,Waxy flexibility | 90.2 | 87.2 | 92.8 |
| 6 | Echopraxia/echolalia,Stereotypy,Rigidity,Negativism,Waxy flexibility,Withdrawal | 90.1 | 87.2 | 92.8 |
| 6 | Excitement,Echopraxia/echolalia,Verbigeration,Rigidity,Negativism,Withdrawal | 90.1 | 87.2 | 92.8 |
| 6 | Excitement,Mutism,Grimacing,Echopraxia/echolalia,Mannerisms,Rigidity | 90.1 | 87.2 | 92.8 |
| 6 | Excitement,Mutism,Grimacing,Echopraxia/echolalia,Waxy flexibility,Withdrawal | 90.1 | 87.4 | 92.8 |
| 6 | Excitement,Mutism,Grimacing,Mannerisms,Verbigeration,Rigidity | 90.1 | 87.2 | 92.8 |
| 6 | Excitement,Mutism,Grimacing,Stereotypy,Waxy flexibility,Withdrawal | 90.1 | 87.2 | 92.8 |
| 6 | Excitement,Mutism,Posturing/catalepsy,Grimacing,Echopraxia/echolalia,Waxy flexibility | 90.1 | 87.2 | 92.8 |
| 6 | Excitement,Mutism,Stereotypy,Mannerisms,Verbigeration,Rigidity | 90.1 | 87.2 | 92.8 |
| 6 | Excitement,Posturing/catalepsy,Grimacing,Negativism,Waxy flexibility,Withdrawal | 90.1 | 87.2 | 92.8 |
| 6 | Excitement,Stereotypy,Mannerisms,Rigidity,Negativism,Withdrawal | 90.1 | 87.2 | 92.8 |
| 6 | Grimacing,Echopraxia/echolalia,Mannerisms,Rigidity,Negativism,Withdrawal | 90.1 | 87.2 | 92.8 |
| 6 | Mutism,Grimacing,Mannerisms,Verbigeration,Negativism,Withdrawal | 90.1 | 87.2 | 92.8 |
| 6 | Mutism,Grimacing,Stereotypy,Mannerisms,Rigidity,Waxy flexibility | 90.1 | 87.2 | 92.8 |
| 6 | Mutism,Grimacing,Stereotypy,Negativism,Waxy flexibility,Withdrawal | 90.1 | 87.2 | 92.8 |
| 6 | Mutism,Posturing/catalepsy,Echopraxia/echolalia,Mannerisms,Waxy flexibility,Withdrawal | 90.1 | 87.2 | 92.8 |
| 6 | Mutism,Posturing/catalepsy,Echopraxia/echolalia,Stereotypy,Negativism,Waxy flexibility | 90.1 | 87.2 | 92.8 |
| 6 | Mutism,Posturing/catalepsy,Echopraxia/echolalia,Stereotypy,Rigidity,Waxy flexibility | 90.1 | 87.4 | 92.8 |
| 6 | Mutism,Posturing/catalepsy,Grimacing,Echopraxia/echolalia,Negativism,Waxy flexibility | 90.1 | 87.2 | 92.8 |
| 6 | Mutism,Posturing/catalepsy,Grimacing,Echopraxia/echolalia,Rigidity,Waxy flexibility | 90.1 | 87.2 | 92.8 |
| 6 | Mutism,Posturing/catalepsy,Grimacing,Echopraxia/echolalia,Stereotypy,Withdrawal | 90.1 | 87.2 | 92.8 |
| 6 | Mutism,Posturing/catalepsy,Grimacing,Stereotypy,Rigidity,Waxy flexibility | 90.1 | 87.2 | 92.8 |
| 6 | Mutism,Posturing/catalepsy,Mannerisms,Verbigeration,Waxy flexibility,Withdrawal | 90.1 | 87.2 | 92.8 |
| 6 | Mutism,Posturing/catalepsy,Stereotypy,Mannerisms,Waxy flexibility,Withdrawal | 90.1 | 87.2 | 92.8 |
| 6 | Posturing/catalepsy,Echopraxia/echolalia,Mannerisms,Verbigeration,Negativism,Withdrawal | 90.1 | 87.2 | 92.8 |
| 6 | Posturing/catalepsy,Echopraxia/echolalia,Stereotypy,Verbigeration,Negativism,Withdrawal | 90.1 | 87.2 | 92.8 |
| 6 | Echopraxia/echolalia,Mannerisms,Rigidity,Negativism,Waxy flexibility,Withdrawal | 89.9 | 87.0 | 92.6 |
| 6 | Excitement,Echopraxia/echolalia,Mannerisms,Rigidity,Negativism,Withdrawal | 89.9 | 87.0 | 92.6 |
| 6 | Excitement,Mutism,Grimacing,Mannerisms,Negativism,Withdrawal | 89.9 | 87.0 | 92.6 |
| 6 | Excitement,Mutism,Posturing/catalepsy,Echopraxia/echolalia,Verbigeration,Waxy flexibility | 89.9 | 87.0 | 92.6 |
| 6 | Excitement,Mutism,Posturing/catalepsy,Grimacing,Verbigeration,Waxy flexibility | 89.9 | 87.0 | 92.6 |
| 6 | Excitement,Mutism,Stereotypy,Mannerisms,Waxy flexibility,Withdrawal | 89.9 | 87.0 | 92.6 |
| 6 | Grimacing,Stereotypy,Rigidity,Negativism,Waxy flexibility,Withdrawal | 89.9 | 87.0 | 92.6 |
| 6 | Mutism,Echopraxia/echolalia,Stereotypy,Mannerisms,Negativism,Waxy flexibility | 89.9 | 87.0 | 92.6 |
| 6 | Mutism,Echopraxia/echolalia,Verbigeration,Negativism,Waxy flexibility,Withdrawal | 89.9 | 87.0 | 92.6 |
| 6 | Mutism,Grimacing,Echopraxia/echolalia,Mannerisms,Rigidity,Waxy flexibility | 89.9 | 87.0 | 92.6 |
| 6 | Mutism,Posturing/catalepsy,Echopraxia/echolalia,Mannerisms,Verbigeration,Withdrawal | 89.9 | 87.0 | 92.6 |
| 6 | Mutism,Posturing/catalepsy,Echopraxia/echolalia,Stereotypy,Mannerisms,Withdrawal | 89.9 | 87.0 | 92.6 |
| 6 | Mutism,Posturing/catalepsy,Grimacing,Echopraxia/echolalia,Waxy flexibility,Withdrawal | 89.9 | 87.0 | 92.6 |
| 6 | Mutism,Posturing/catalepsy,Grimacing,Stereotypy,Negativism,Waxy flexibility | 89.9 | 87.0 | 92.6 |
| 6 | Mutism,Posturing/catalepsy,Grimacing,Stereotypy,Verbigeration,Withdrawal | 89.9 | 87.0 | 92.6 |
| 6 | Mutism,Posturing/catalepsy,Grimacing,Stereotypy,Waxy flexibility,Withdrawal | 89.9 | 87.0 | 92.6 |
| 6 | Mutism,Posturing/catalepsy,Grimacing,Verbigeration,Waxy flexibility,Withdrawal | 89.9 | 87.0 | 92.6 |
| 6 | Posturing/catalepsy,Echopraxia/echolalia,Stereotypy,Negativism,Waxy flexibility,Withdrawal | 89.9 | 87.0 | 92.6 |
| 6 | Posturing/catalepsy,Mannerisms,Verbigeration,Negativism,Waxy flexibility,Withdrawal | 89.9 | 87.0 | 92.6 |
| 6 | Echopraxia/echolalia,Mannerisms,Verbigeration,Rigidity,Negativism,Withdrawal | 89.7 | 86.8 | 92.4 |
| 6 | Excitement,Grimacing,Verbigeration,Rigidity,Negativism,Withdrawal | 89.7 | 86.8 | 92.4 |
| 6 | Excitement,Mannerisms,Verbigeration,Rigidity,Negativism,Withdrawal | 89.7 | 86.8 | 92.4 |
| 6 | Excitement,Mutism,Echopraxia/echolalia,Mannerisms,Verbigeration,Rigidity | 89.7 | 86.8 | 92.4 |
| 6 | Excitement,Mutism,Echopraxia/echolalia,Stereotypy,Verbigeration,Negativism | 89.7 | 86.8 | 92.4 |
| 6 | Excitement,Mutism,Echopraxia/echolalia,Verbigeration,Waxy flexibility,Withdrawal | 89.7 | 86.8 | 92.4 |
| 6 | Excitement,Mutism,Grimacing,Echopraxia/echolalia,Negativism,Waxy flexibility | 89.7 | 86.8 | 92.4 |
| 6 | Excitement,Mutism,Grimacing,Stereotypy,Mannerisms,Rigidity | 89.7 | 86.8 | 92.4 |
| 6 | Excitement,Mutism,Grimacing,Stereotypy,Negativism,Waxy flexibility | 89.7 | 86.8 | 92.4 |
| 6 | Excitement,Mutism,Posturing/catalepsy,Echopraxia/echolalia,Stereotypy,Waxy flexibility | 89.7 | 86.8 | 92.4 |
| 6 | Excitement,Mutism,Posturing/catalepsy,Stereotypy,Verbigeration,Waxy flexibility | 89.7 | 86.8 | 92.4 |
| 6 | Excitement,Verbigeration,Rigidity,Negativism,Waxy flexibility,Withdrawal | 89.7 | 86.8 | 92.4 |
| 6 | Grimacing,Mannerisms,Rigidity,Negativism,Waxy flexibility,Withdrawal | 89.7 | 86.8 | 92.4 |
| 6 | Grimacing,Mannerisms,Verbigeration,Rigidity,Negativism,Withdrawal | 89.7 | 86.8 | 92.4 |
| 6 | Mannerisms,Verbigeration,Rigidity,Negativism,Waxy flexibility,Withdrawal | 89.7 | 86.8 | 92.4 |
| 6 | Mutism,Echopraxia/echolalia,Stereotypy,Mannerisms,Rigidity,Waxy flexibility | 89.7 | 86.8 | 92.4 |
| 6 | Mutism,Grimacing,Echopraxia/echolalia,Negativism,Waxy flexibility,Withdrawal | 89.7 | 86.8 | 92.4 |
| 6 | Mutism,Grimacing,Echopraxia/echolalia,Stereotypy,Rigidity,Waxy flexibility | 89.7 | 86.8 | 92.4 |
| 6 | Mutism,Grimacing,Echopraxia/echolalia,Verbigeration,Negativism,Withdrawal | 89.7 | 86.8 | 92.4 |
| 6 | Mutism,Grimacing,Echopraxia/echolalia,Verbigeration,Rigidity,Waxy flexibility | 89.7 | 86.8 | 92.4 |
| 6 | Mutism,Grimacing,Mannerisms,Verbigeration,Rigidity,Waxy flexibility | 89.7 | 86.8 | 92.4 |
| 6 | Mutism,Posturing/catalepsy,Echopraxia/echolalia,Stereotypy,Waxy flexibility,Withdrawal | 89.7 | 86.8 | 92.4 |
| 6 | Mutism,Posturing/catalepsy,Grimacing,Echopraxia/echolalia,Verbigeration,Withdrawal | 89.7 | 86.8 | 92.4 |
| 6 | Mutism,Posturing/catalepsy,Stereotypy,Mannerisms,Verbigeration,Withdrawal | 89.7 | 86.8 | 92.4 |
| 6 | Mutism,Posturing/catalepsy,Stereotypy,Verbigeration,Waxy flexibility,Withdrawal | 89.7 | 86.8 | 92.4 |
| 6 | Mutism,Stereotypy,Mannerisms,Verbigeration,Rigidity,Waxy flexibility | 89.7 | 86.8 | 92.4 |
| 6 | Posturing/catalepsy,Echopraxia/echolalia,Mannerisms,Negativism,Waxy flexibility,Withdrawal | 89.7 | 86.8 | 92.4 |
| 6 | Posturing/catalepsy,Grimacing,Echopraxia/echolalia,Mannerisms,Rigidity,Withdrawal | 89.7 | 86.8 | 92.4 |
| 6 | Posturing/catalepsy,Grimacing,Mannerisms,Verbigeration,Rigidity,Withdrawal | 89.7 | 86.8 | 92.4 |
| 6 | Posturing/catalepsy,Grimacing,Stereotypy,Mannerisms,Rigidity,Withdrawal | 89.7 | 86.8 | 92.4 |
| 6 | Posturing/catalepsy,Grimacing,Stereotypy,Negativism,Waxy flexibility,Withdrawal | 89.7 | 86.8 | 92.4 |
| 6 | Excitement,Echopraxia/echolalia,Stereotypy,Rigidity,Waxy flexibility,Withdrawal | 89.5 | 86.5 | 92.4 |
| 6 | Excitement,Grimacing,Mannerisms,Rigidity,Negativism,Withdrawal | 89.5 | 86.5 | 92.2 |
| 6 | Excitement,Grimacing,Rigidity,Negativism,Waxy flexibility,Withdrawal | 89.5 | 86.5 | 92.2 |
| 6 | Excitement,Mutism,Grimacing,Verbigeration,Waxy flexibility,Withdrawal | 89.5 | 86.5 | 92.2 |
| 6 | Excitement,Mutism,Posturing/catalepsy,Grimacing,Echopraxia/echolalia,Verbigeration | 89.5 | 86.5 | 92.4 |
| 6 | Excitement,Mutism,Posturing/catalepsy,Grimacing,Stereotypy,Waxy flexibility | 89.5 | 86.5 | 92.2 |
| 6 | Excitement,Mutism,Stereotypy,Mannerisms,Negativism,Waxy flexibility | 89.5 | 86.5 | 92.2 |
| 6 | Excitement,Posturing/catalepsy,Grimacing,Echopraxia/echolalia,Waxy flexibility,Withdrawal | 89.5 | 86.5 | 92.2 |
| 6 | Mutism,Grimacing,Verbigeration,Negativism,Waxy flexibility,Withdrawal | 89.5 | 86.5 | 92.2 |
| 6 | Mutism,Posturing/catalepsy,Echopraxia/echolalia,Verbigeration,Waxy flexibility,Withdrawal | 89.5 | 86.5 | 92.2 |
| 6 | Mutism,Stereotypy,Mannerisms,Verbigeration,Negativism,Waxy flexibility | 89.5 | 86.5 | 92.2 |
| 6 | Posturing/catalepsy,Echopraxia/echolalia,Mannerisms,Verbigeration,Rigidity,Withdrawal | 89.5 | 86.5 | 92.2 |
| 6 | Posturing/catalepsy,Grimacing,Mannerisms,Negativism,Waxy flexibility,Withdrawal | 89.5 | 86.5 | 92.2 |
| 6 | Posturing/catalepsy,Grimacing,Mannerisms,Rigidity,Waxy flexibility,Withdrawal | 89.5 | 86.5 | 92.2 |
| 6 | Posturing/catalepsy,Mannerisms,Verbigeration,Rigidity,Waxy flexibility,Withdrawal | 89.5 | 86.5 | 92.2 |
| 6 | Posturing/catalepsy,Stereotypy,Mannerisms,Rigidity,Waxy flexibility,Withdrawal | 89.5 | 86.5 | 92.2 |
| 6 | Posturing/catalepsy,Stereotypy,Mannerisms,Verbigeration,Rigidity,Withdrawal | 89.5 | 86.5 | 92.2 |
| 6 | Posturing/catalepsy,Grimacing,Echopraxia/echolalia,Verbigeration,Negativism,Withdrawal | 89.4 | 86.5 | 92.2 |
| 6 | Excitement,Mannerisms,Rigidity,Negativism,Waxy flexibility,Withdrawal | 89.3 | 86.3 | 91.9 |
| 6 | Excitement,Mutism,Echopraxia/echolalia,Mannerisms,Waxy flexibility,Withdrawal | 89.3 | 86.3 | 92.2 |
| 6 | Excitement,Mutism,Echopraxia/echolalia,Verbigeration,Negativism,Waxy flexibility | 89.3 | 86.3 | 92.2 |
| 6 | Excitement,Mutism,Grimacing,Echopraxia/echolalia,Stereotypy,Negativism | 89.3 | 86.3 | 92.2 |
| 6 | Excitement,Mutism,Posturing/catalepsy,Echopraxia/echolalia,Mannerisms,Waxy flexibility | 89.3 | 86.3 | 92.2 |
| 6 | Excitement,Mutism,Posturing/catalepsy,Grimacing,Stereotypy,Verbigeration | 89.3 | 86.3 | 91.9 |
| 6 | Posturing/catalepsy,Echopraxia/echolalia,Mannerisms,Rigidity,Waxy flexibility,Withdrawal | 89.3 | 86.3 | 91.9 |
| 6 | Excitement,Mutism,Echopraxia/echolalia,Stereotypy,Verbigeration,Withdrawal | 89.2 | 86.3 | 91.9 |
| 6 | Excitement,Mutism,Grimacing,Mannerisms,Waxy flexibility,Withdrawal | 89.2 | 86.3 | 91.9 |
| 6 | Excitement,Mutism,Grimacing,Stereotypy,Verbigeration,Negativism | 89.2 | 86.3 | 91.9 |
| 6 | Excitement,Mutism,Posturing/catalepsy,Echopraxia/echolalia,Stereotypy,Verbigeration | 89.2 | 86.3 | 91.9 |
| 6 | Excitement,Mutism,Posturing/catalepsy,Grimacing,Echopraxia/echolalia,Stereotypy | 89.2 | 86.3 | 91.9 |
| 6 | Excitement,Mutism,Posturing/catalepsy,Grimacing,Mannerisms,Waxy flexibility | 89.2 | 86.3 | 91.9 |
| 6 | Excitement,Stereotypy,Verbigeration,Rigidity,Waxy flexibility,Withdrawal | 89.2 | 86.3 | 91.9 |
| 6 | Mutism,Echopraxia/echolalia,Stereotypy,Verbigeration,Rigidity,Waxy flexibility | 89.2 | 86.3 | 91.9 |
| 6 | Mutism,Grimacing,Echopraxia/echolalia,Mannerisms,Negativism,Waxy flexibility | 89.2 | 86.3 | 91.9 |
| 6 | Mutism,Grimacing,Stereotypy,Mannerisms,Negativism,Waxy flexibility | 89.2 | 86.3 | 91.9 |
| 6 | Mutism,Posturing/catalepsy,Echopraxia/echolalia,Stereotypy,Verbigeration,Withdrawal | 89.2 | 86.3 | 92.2 |
| 6 | Posturing/catalepsy,Echopraxia/echolalia,Stereotypy,Mannerisms,Rigidity,Withdrawal | 89.2 | 86.3 | 91.9 |
| 6 | Posturing/catalepsy,Grimacing,Echopraxia/echolalia,Stereotypy,Rigidity,Withdrawal | 89.2 | 86.3 | 91.9 |
| 6 | Posturing/catalepsy,Grimacing,Stereotypy,Verbigeration,Rigidity,Withdrawal | 89.2 | 86.3 | 91.9 |
| 6 | Echopraxia/echolalia,Verbigeration,Rigidity,Negativism,Waxy flexibility,Withdrawal | 89.0 | 86.1 | 91.9 |
| 6 | Excitement,Mutism,Echopraxia/echolalia,Stereotypy,Mannerisms,Negativism | 89.0 | 86.1 | 91.9 |
| 6 | Excitement,Mutism,Grimacing,Verbigeration,Negativism,Waxy flexibility | 89.0 | 86.1 | 91.7 |
| 6 | Excitement,Mutism,Mannerisms,Verbigeration,Waxy flexibility,Withdrawal | 89.0 | 86.1 | 91.9 |
| 6 | Excitement,Mutism,Posturing/catalepsy,Grimacing,Echopraxia/echolalia,Mannerisms | 89.0 | 86.1 | 91.9 |
| 6 | Excitement,Mutism,Posturing/catalepsy,Mannerisms,Verbigeration,Waxy flexibility | 89.0 | 86.1 | 91.7 |
| 6 | Excitement,Mutism,Stereotypy,Mannerisms,Verbigeration,Negativism | 89.0 | 86.1 | 91.7 |
| 6 | Excitement,Posturing/catalepsy,Echopraxia/echolalia,Verbigeration,Waxy flexibility,Withdrawal | 89.0 | 86.1 | 91.7 |
| 6 | Excitement,Posturing/catalepsy,Grimacing,Echopraxia/echolalia,Mannerisms,Withdrawal | 89.0 | 86.1 | 91.7 |
| 6 | Excitement,Posturing/catalepsy,Grimacing,Echopraxia/echolalia,Verbigeration,Withdrawal | 89.0 | 86.1 | 91.7 |
| 6 | Grimacing,Echopraxia/echolalia,Rigidity,Negativism,Waxy flexibility,Withdrawal | 89.0 | 86.1 | 91.7 |
| 6 | Grimacing,Echopraxia/echolalia,Verbigeration,Rigidity,Negativism,Withdrawal | 89.0 | 85.9 | 91.9 |
| 6 | Mutism,Echopraxia/echolalia,Stereotypy,Mannerisms,Verbigeration,Negativism | 89.0 | 86.1 | 91.9 |
| 6 | Mutism,Grimacing,Echopraxia/echolalia,Stereotypy,Mannerisms,Negativism | 89.0 | 86.1 | 91.9 |
| 6 | Mutism,Grimacing,Echopraxia/echolalia,Stereotypy,Mannerisms,Rigidity | 89.0 | 86.1 | 91.9 |
| 6 | Mutism,Grimacing,Stereotypy,Mannerisms,Verbigeration,Rigidity | 89.0 | 86.1 | 91.7 |
| 6 | Posturing/catalepsy,Echopraxia/echolalia,Verbigeration,Negativism,Waxy flexibility,Withdrawal | 89.0 | 86.1 | 91.9 |
| 6 | Posturing/catalepsy,Grimacing,Echopraxia/echolalia,Verbigeration,Rigidity,Withdrawal | 89.0 | 86.1 | 91.9 |
| 6 | Posturing/catalepsy,Stereotypy,Verbigeration,Rigidity,Waxy flexibility,Withdrawal | 89.0 | 86.1 | 91.9 |
| 6 | Excitement,Grimacing,Stereotypy,Rigidity,Waxy flexibility,Withdrawal | 88.8 | 85.9 | 91.7 |
| 6 | Excitement,Mutism,Echopraxia/echolalia,Mannerisms,Negativism,Waxy flexibility | 88.8 | 85.7 | 91.7 |
| 6 | Excitement,Mutism,Grimacing,Echopraxia/echolalia,Stereotypy,Withdrawal | 88.8 | 85.9 | 91.7 |
| 6 | Excitement,Mutism,Posturing/catalepsy,Echopraxia/echolalia,Mannerisms,Verbigeration | 88.8 | 85.9 | 91.7 |
| 6 | Excitement,Mutism,Posturing/catalepsy,Grimacing,Mannerisms,Verbigeration | 88.8 | 85.9 | 91.7 |
| 6 | Excitement,Posturing/catalepsy,Stereotypy,Verbigeration,Waxy flexibility,Withdrawal | 88.8 | 85.9 | 91.7 |
| 6 | Excitement,Stereotypy,Mannerisms,Rigidity,Waxy flexibility,Withdrawal | 88.8 | 85.7 | 91.7 |
| 6 | Grimacing,Verbigeration,Rigidity,Negativism,Waxy flexibility,Withdrawal | 88.8 | 85.9 | 91.5 |
| 6 | Mutism,Echopraxia/echolalia,Mannerisms,Verbigeration,Rigidity,Waxy flexibility | 88.8 | 85.9 | 91.5 |
| 6 | Mutism,Echopraxia/echolalia,Stereotypy,Verbigeration,Negativism,Waxy flexibility | 88.8 | 85.7 | 91.7 |
| 6 | Mutism,Grimacing,Echopraxia/echolalia,Mannerisms,Verbigeration,Rigidity | 88.8 | 85.7 | 91.7 |
| 6 | Mutism,Grimacing,Echopraxia/echolalia,Stereotypy,Negativism,Waxy flexibility | 88.8 | 85.9 | 91.7 |
| 6 | Mutism,Grimacing,Echopraxia/echolalia,Stereotypy,Verbigeration,Rigidity | 88.8 | 85.9 | 91.7 |
| 6 | Mutism,Grimacing,Echopraxia/echolalia,Stereotypy,Waxy flexibility,Withdrawal | 88.8 | 85.7 | 91.5 |
| 6 | Posturing/catalepsy,Echopraxia/echolalia,Stereotypy,Rigidity,Waxy flexibility,Withdrawal | 88.8 | 85.7 | 91.7 |
| 6 | Posturing/catalepsy,Echopraxia/echolalia,Stereotypy,Verbigeration,Rigidity,Withdrawal | 88.8 | 85.9 | 91.7 |
| 6 | Posturing/catalepsy,Echopraxia/echolalia,Verbigeration,Rigidity,Waxy flexibility,Withdrawal | 88.8 | 85.9 | 91.7 |
| 6 | Posturing/catalepsy,Grimacing,Echopraxia/echolalia,Negativism,Waxy flexibility,Withdrawal | 88.8 | 85.9 | 91.7 |
| 6 | Posturing/catalepsy,Grimacing,Echopraxia/echolalia,Rigidity,Waxy flexibility,Withdrawal | 88.8 | 85.7 | 91.7 |
| 6 | Posturing/catalepsy,Grimacing,Stereotypy,Rigidity,Waxy flexibility,Withdrawal | 88.8 | 85.9 | 91.7 |
| 6 | Posturing/catalepsy,Grimacing,Verbigeration,Negativism,Waxy flexibility,Withdrawal | 88.8 | 85.9 | 91.7 |
| 6 | Posturing/catalepsy,Grimacing,Verbigeration,Rigidity,Waxy flexibility,Withdrawal | 88.8 | 85.9 | 91.5 |
| 6 | Excitement,Echopraxia/echolalia,Verbigeration,Rigidity,Waxy flexibility,Withdrawal | 88.6 | 85.4 | 91.5 |
| 6 | Excitement,Grimacing,Echopraxia/echolalia,Rigidity,Waxy flexibility,Withdrawal | 88.6 | 85.7 | 91.5 |
| 6 | Excitement,Mutism,Echopraxia/echolalia,Stereotypy,Mannerisms,Withdrawal | 88.6 | 85.7 | 91.5 |
| 6 | Excitement,Mutism,Grimacing,Echopraxia/echolalia,Verbigeration,Negativism | 88.6 | 85.4 | 91.5 |
| 6 | Excitement,Mutism,Grimacing,Mannerisms,Negativism,Waxy flexibility | 88.6 | 85.7 | 91.5 |
| 6 | Excitement,Mutism,Grimacing,Stereotypy,Verbigeration,Withdrawal | 88.6 | 85.7 | 91.5 |
| 6 | Excitement,Mutism,Posturing/catalepsy,Stereotypy,Mannerisms,Verbigeration | 88.6 | 85.7 | 91.5 |
| 6 | Excitement,Mutism,Posturing/catalepsy,Stereotypy,Mannerisms,Waxy flexibility | 88.6 | 85.7 | 91.5 |
| 6 | Excitement,Posturing/catalepsy,Echopraxia/echolalia,Stereotypy,Waxy flexibility,Withdrawal | 88.6 | 85.6 | 91.5 |
| 6 | Excitement,Posturing/catalepsy,Grimacing,Echopraxia/echolalia,Stereotypy,Withdrawal | 88.6 | 85.7 | 91.5 |
| 6 | Excitement,Posturing/catalepsy,Grimacing,Mannerisms,Verbigeration,Withdrawal | 88.6 | 85.4 | 91.5 |
| 6 | Excitement,Posturing/catalepsy,Grimacing,Mannerisms,Waxy flexibility,Withdrawal | 88.6 | 85.7 | 91.5 |
| 6 | Excitement,Posturing/catalepsy,Grimacing,Stereotypy,Waxy flexibility,Withdrawal | 88.6 | 85.7 | 91.5 |
| 6 | Excitement,Posturing/catalepsy,Grimacing,Verbigeration,Rigidity,Negativism | 88.6 | 85.4 | 91.5 |
| 6 | Excitement,Posturing/catalepsy,Grimacing,Verbigeration,Waxy flexibility,Withdrawal | 88.6 | 85.7 | 91.5 |
| 6 | Excitement,Posturing/catalepsy,Mannerisms,Verbigeration,Waxy flexibility,Withdrawal | 88.6 | 85.4 | 91.5 |
| 6 | Mutism,Echopraxia/echolalia,Stereotypy,Mannerisms,Waxy flexibility,Withdrawal | 88.6 | 85.4 | 91.5 |
| 6 | Mutism,Grimacing,Echopraxia/echolalia,Mannerisms,Waxy flexibility,Withdrawal | 88.6 | 85.7 | 91.5 |
| 6 | Mutism,Grimacing,Stereotypy,Mannerisms,Verbigeration,Negativism | 88.6 | 85.7 | 91.5 |
| 6 | Mutism,Grimacing,Stereotypy,Mannerisms,Waxy flexibility,Withdrawal | 88.6 | 85.7 | 91.5 |
| 6 | Mutism,Grimacing,Stereotypy,Verbigeration,Negativism,Waxy flexibility | 88.6 | 85.7 | 91.5 |
| 6 | Excitement,Mutism,Posturing/catalepsy,Echopraxia/echolalia,Stereotypy,Mannerisms | 88.5 | 85.7 | 91.5 |
| 6 | Excitement,Posturing/catalepsy,Echopraxia/echolalia,Mannerisms,Waxy flexibility,Withdrawal | 88.5 | 85.4 | 91.5 |
| 6 | Excitement,Posturing/catalepsy,Grimacing,Echopraxia/echolalia,Rigidity,Negativism | 88.5 | 85.4 | 91.5 |
| 6 | Excitement,Posturing/catalepsy,Grimacing,Stereotypy,Verbigeration,Withdrawal | 88.4 | 85.4 | 91.3 |
| 6 | Mutism,Grimacing,Echopraxia/echolalia,Stereotypy,Verbigeration,Negativism | 88.4 | 85.4 | 91.3 |
| 6 | Excitement,Mutism,Mannerisms,Verbigeration,Negativism,Waxy flexibility | 88.3 | 85.2 | 91.3 |
| 6 | Excitement,Mutism,Posturing/catalepsy,Grimacing,Stereotypy,Mannerisms | 88.3 | 85.2 | 91.3 |
| 6 | Excitement,Mutism,Stereotypy,Mannerisms,Verbigeration,Withdrawal | 88.3 | 85.2 | 91.3 |
| 6 | Excitement,Posturing/catalepsy,Echopraxia/echolalia,Mannerisms,Verbigeration,Withdrawal | 88.3 | 85.2 | 91.3 |
| 6 | Excitement,Posturing/catalepsy,Stereotypy,Verbigeration,Rigidity,Negativism | 88.3 | 85.2 | 91.3 |
| 6 | Mutism,Echopraxia/echolalia,Mannerisms,Verbigeration,Negativism,Waxy flexibility | 88.3 | 85.2 | 91.3 |
| 6 | Mutism,Echopraxia/echolalia,Stereotypy,Mannerisms,Verbigeration,Rigidity | 88.3 | 85.2 | 91.3 |
| 6 | Mutism,Grimacing,Echopraxia/echolalia,Verbigeration,Negativism,Waxy flexibility | 88.3 | 85.2 | 91.3 |
| 6 | Echopraxia/echolalia,Stereotypy,Mannerisms,Negativism,Waxy flexibility,Withdrawal | 88.1 | 85.0 | 91.0 |
| 6 | Excitement,Echopraxia/echolalia,Stereotypy,Mannerisms,Rigidity,Withdrawal | 88.1 | 85.0 | 91.0 |
| 6 | Excitement,Echopraxia/echolalia,Stereotypy,Verbigeration,Rigidity,Withdrawal | 88.1 | 85.0 | 91.0 |
| 6 | Excitement,Grimacing,Verbigeration,Rigidity,Waxy flexibility,Withdrawal | 88.1 | 85.0 | 91.0 |
| 6 | Excitement,Mannerisms,Verbigeration,Rigidity,Waxy flexibility,Withdrawal | 88.1 | 85.0 | 91.0 |
| 6 | Excitement,Mutism,Grimacing,Echopraxia/echolalia,Verbigeration,Withdrawal | 88.1 | 85.0 | 91.0 |
| 6 | Excitement,Mutism,Grimacing,Stereotypy,Mannerisms,Negativism | 88.1 | 85.0 | 91.0 |
| 6 | Excitement,Posturing/catalepsy,Echopraxia/echolalia,Stereotypy,Rigidity,Negativism | 88.1 | 85.0 | 91.0 |
| 6 | Excitement,Posturing/catalepsy,Echopraxia/echolalia,Stereotypy,Verbigeration,Withdrawal | 88.1 | 85.0 | 91.0 |
| 6 | Excitement,Posturing/catalepsy,Echopraxia/echolalia,Verbigeration,Rigidity,Negativism | 88.1 | 85.0 | 91.0 |
| 6 | Excitement,Posturing/catalepsy,Grimacing,Stereotypy,Mannerisms,Withdrawal | 88.1 | 85.0 | 91.0 |
| 6 | Excitement,Posturing/catalepsy,Grimacing,Stereotypy,Rigidity,Negativism | 88.1 | 85.0 | 91.0 |
| 6 | Excitement,Posturing/catalepsy,Stereotypy,Mannerisms,Verbigeration,Withdrawal | 88.1 | 85.0 | 91.0 |
| 6 | Excitement,Posturing/catalepsy,Stereotypy,Mannerisms,Waxy flexibility,Withdrawal | 88.1 | 85.2 | 91.0 |
| 6 | Grimacing,Stereotypy,Mannerisms,Rigidity,Waxy flexibility,Withdrawal | 88.1 | 85.0 | 91.0 |
| 6 | Mutism,Grimacing,Echopraxia/echolalia,Mannerisms,Verbigeration,Negativism | 88.1 | 85.0 | 91.0 |
| 6 | Mutism,Grimacing,Stereotypy,Verbigeration,Waxy flexibility,Withdrawal | 88.1 | 85.0 | 91.0 |
| 6 | Excitement,Echopraxia/echolalia,Mannerisms,Rigidity,Waxy flexibility,Withdrawal | 87.9 | 84.8 | 90.8 |
| 6 | Excitement,Echopraxia/echolalia,Stereotypy,Negativism,Waxy flexibility,Withdrawal | 87.9 | 84.8 | 90.8 |
| 6 | Excitement,Grimacing,Echopraxia/echolalia,Stereotypy,Rigidity,Withdrawal | 87.9 | 84.8 | 90.8 |
| 6 | Excitement,Mutism,Echopraxia/echolalia,Mannerisms,Verbigeration,Negativism | 87.9 | 85.0 | 90.8 |
| 6 | Excitement,Mutism,Grimacing,Echopraxia/echolalia,Mannerisms,Negativism | 87.9 | 84.8 | 90.8 |
| 6 | Excitement,Posturing/catalepsy,Echopraxia/echolalia,Stereotypy,Mannerisms,Withdrawal | 87.9 | 84.8 | 90.8 |
| 6 | Excitement,Posturing/catalepsy,Grimacing,Mannerisms,Rigidity,Negativism | 87.9 | 84.8 | 90.8 |
| 6 | Excitement,Posturing/catalepsy,Grimacing,Rigidity,Negativism,Waxy flexibility | 87.9 | 84.8 | 90.8 |
| 6 | Excitement,Posturing/catalepsy,Mannerisms,Verbigeration,Rigidity,Negativism | 87.9 | 84.8 | 90.8 |
| 6 | Mutism,Echopraxia/echolalia,Stereotypy,Verbigeration,Waxy flexibility,Withdrawal | 87.9 | 84.8 | 90.8 |
| 6 | Mutism,Grimacing,Echopraxia/echolalia,Verbigeration,Waxy flexibility,Withdrawal | 87.9 | 84.8 | 90.8 |
| 6 | Mutism,Grimacing,Mannerisms,Verbigeration,Negativism,Waxy flexibility | 87.9 | 84.8 | 90.8 |
| 6 | Mutism,Grimacing,Mannerisms,Verbigeration,Waxy flexibility,Withdrawal | 87.9 | 84.8 | 90.8 |
| 6 | Mutism,Stereotypy,Mannerisms,Verbigeration,Waxy flexibility,Withdrawal | 87.9 | 84.8 | 90.8 |
| 6 | Posturing/catalepsy,Grimacing,Echopraxia/echolalia,Mannerisms,Rigidity,Negativism | 87.9 | 84.8 | 90.8 |
| 6 | Posturing/catalepsy,Grimacing,Echopraxia/echolalia,Stereotypy,Mannerisms,Withdrawal | 87.9 | 84.8 | 90.8 |
| 6 | Posturing/catalepsy,Grimacing,Stereotypy,Mannerisms,Rigidity,Negativism | 87.9 | 84.8 | 90.8 |
| 6 | Posturing/catalepsy,Grimacing,Stereotypy,Verbigeration,Rigidity,Negativism | 87.9 | 84.8 | 90.8 |
| 6 | Posturing/catalepsy,Stereotypy,Mannerisms,Verbigeration,Rigidity,Negativism | 87.9 | 84.8 | 90.8 |
| 6 | Echopraxia/echolalia,Stereotypy,Mannerisms,Rigidity,Waxy flexibility,Withdrawal | 87.7 | 84.5 | 90.6 |
| 6 | Excitement,Mutism,Grimacing,Mannerisms,Verbigeration,Negativism | 87.7 | 84.5 | 90.6 |
| 6 | Excitement,Mutism,Grimacing,Stereotypy,Mannerisms,Withdrawal | 87.7 | 84.5 | 90.6 |
| 6 | Excitement,Posturing/catalepsy,Echopraxia/echolalia,Mannerisms,Rigidity,Negativism | 87.7 | 84.5 | 90.6 |
| 6 | Excitement,Posturing/catalepsy,Echopraxia/echolalia,Rigidity,Negativism,Waxy flexibility | 87.7 | 84.5 | 90.6 |
| 6 | Excitement,Posturing/catalepsy,Stereotypy,Rigidity,Negativism,Waxy flexibility | 87.7 | 84.5 | 90.8 |
| 6 | Excitement,Posturing/catalepsy,Verbigeration,Rigidity,Negativism,Waxy flexibility | 87.7 | 84.5 | 90.6 |
| 6 | Excitement,Stereotypy,Mannerisms,Verbigeration,Rigidity,Withdrawal | 87.7 | 84.5 | 90.6 |
| 6 | Grimacing,Echopraxia/echolalia,Stereotypy,Rigidity,Waxy flexibility,Withdrawal | 87.7 | 84.5 | 90.6 |
| 6 | Mutism,Grimacing,Echopraxia/echolalia,Stereotypy,Mannerisms,Withdrawal | 87.7 | 84.5 | 90.6 |
| 6 | Mutism,Posturing/catalepsy,Grimacing,Echopraxia/echolalia,Mannerisms,Verbigeration | 87.7 | 84.5 | 90.6 |
| 6 | Mutism,Posturing/catalepsy,Grimacing,Echopraxia/echolalia,Verbigeration,Waxy flexibility | 87.7 | 84.5 | 90.8 |
| 6 | Mutism,Posturing/catalepsy,Grimacing,Stereotypy,Verbigeration,Waxy flexibility | 87.7 | 84.5 | 90.8 |
| 6 | Posturing/catalepsy,Echopraxia/echolalia,Stereotypy,Mannerisms,Rigidity,Negativism | 87.7 | 84.5 | 90.6 |
| 6 | Posturing/catalepsy,Grimacing,Mannerisms,Verbigeration,Rigidity,Negativism | 87.7 | 84.5 | 90.6 |
| 6 | Posturing/catalepsy,Grimacing,Stereotypy,Mannerisms,Waxy flexibility,Withdrawal | 87.7 | 84.8 | 90.6 |
| 6 | Excitement,Mutism,Echopraxia/echolalia,Mannerisms,Verbigeration,Withdrawal | 87.5 | 84.3 | 90.4 |
| 6 | Excitement,Mutism,Grimacing,Echopraxia/echolalia,Stereotypy,Waxy flexibility | 87.5 | 84.3 | 90.4 |
| 6 | Excitement,Posturing/catalepsy,Stereotypy,Mannerisms,Rigidity,Negativism | 87.5 | 84.3 | 90.4 |
| 6 | Excitement,Stereotypy,Verbigeration,Negativism,Waxy flexibility,Withdrawal | 87.5 | 84.3 | 90.4 |
| 6 | Mutism,Posturing/catalepsy,Grimacing,Echopraxia/echolalia,Stereotypy,Mannerisms | 87.5 | 84.3 | 90.4 |
| 6 | Mutism,Posturing/catalepsy,Grimacing,Echopraxia/echolalia,Stereotypy,Waxy flexibility | 87.5 | 84.3 | 90.4 |
| 6 | Posturing/catalepsy,Grimacing,Echopraxia/echolalia,Mannerisms,Verbigeration,Withdrawal | 87.5 | 84.3 | 90.4 |
| 6 | Excitement,Grimacing,Echopraxia/echolalia,Verbigeration,Rigidity,Withdrawal | 87.4 | 84.3 | 90.4 |
| 6 | Excitement,Grimacing,Stereotypy,Verbigeration,Rigidity,Withdrawal | 87.4 | 84.3 | 90.4 |
| 6 | Excitement,Mutism,Grimacing,Echopraxia/echolalia,Mannerisms,Withdrawal | 87.4 | 84.3 | 90.4 |
| 6 | Excitement,Mutism,Grimacing,Mannerisms,Verbigeration,Withdrawal | 87.4 | 84.3 | 90.4 |
| 6 | Grimacing,Stereotypy,Verbigeration,Rigidity,Waxy flexibility,Withdrawal | 87.4 | 84.3 | 90.6 |
| 6 | Mutism,Posturing/catalepsy,Grimacing,Echopraxia/echolalia,Mannerisms,Waxy flexibility | 87.4 | 84.3 | 90.4 |
| 6 | Mutism,Posturing/catalepsy,Grimacing,Echopraxia/echolalia,Stereotypy,Verbigeration | 87.4 | 84.3 | 90.4 |
| 6 | Mutism,Posturing/catalepsy,Grimacing,Mannerisms,Verbigeration,Waxy flexibility | 87.4 | 84.3 | 90.4 |
| 6 | Posturing/catalepsy,Echopraxia/echolalia,Mannerisms,Verbigeration,Rigidity,Negativism | 87.4 | 84.3 | 90.4 |
| 6 | Posturing/catalepsy,Grimacing,Echopraxia/echolalia,Stereotypy,Waxy flexibility,Withdrawal | 87.4 | 84.3 | 90.4 |
| 6 | Posturing/catalepsy,Grimacing,Stereotypy,Mannerisms,Verbigeration,Withdrawal | 87.4 | 84.3 | 90.4 |
| 6 | Posturing/catalepsy,Stereotypy,Mannerisms,Rigidity,Negativism,Waxy flexibility | 87.4 | 84.3 | 90.4 |
| 6 | Posturing/catalepsy,Stereotypy,Verbigeration,Rigidity,Negativism,Waxy flexibility | 87.4 | 84.3 | 90.4 |
| 6 | Stereotypy,Mannerisms,Verbigeration,Negativism,Waxy flexibility,Withdrawal | 87.4 | 84.3 | 90.4 |
| 6 | Stereotypy,Mannerisms,Verbigeration,Rigidity,Waxy flexibility,Withdrawal | 87.4 | 84.3 | 90.4 |
| 6 | Excitement,Grimacing,Mannerisms,Rigidity,Waxy flexibility,Withdrawal | 87.2 | 84.1 | 90.1 |
| 6 | Excitement,Grimacing,Stereotypy,Mannerisms,Rigidity,Withdrawal | 87.2 | 84.1 | 90.4 |
| 6 | Grimacing,Echopraxia/echolalia,Stereotypy,Mannerisms,Negativism,Withdrawal | 87.2 | 84.1 | 90.4 |
| 6 | Grimacing,Echopraxia/echolalia,Stereotypy,Mannerisms,Rigidity,Withdrawal | 87.2 | 84.1 | 90.4 |
| 6 | Grimacing,Stereotypy,Mannerisms,Negativism,Waxy flexibility,Withdrawal | 87.2 | 84.1 | 90.4 |
| 6 | Mutism,Echopraxia/echolalia,Mannerisms,Verbigeration,Waxy flexibility,Withdrawal | 87.2 | 84.1 | 90.1 |
| 6 | Mutism,Grimacing,Echopraxia/echolalia,Stereotypy,Verbigeration,Withdrawal | 87.2 | 84.1 | 90.4 |
| 6 | Mutism,Posturing/catalepsy,Grimacing,Stereotypy,Mannerisms,Verbigeration | 87.2 | 84.1 | 90.1 |
| 6 | Posturing/catalepsy,Echopraxia/echolalia,Mannerisms,Rigidity,Negativism,Waxy flexibility | 87.2 | 84.1 | 90.1 |
| 6 | Posturing/catalepsy,Echopraxia/echolalia,Stereotypy,Verbigeration,Rigidity,Negativism | 87.2 | 84.1 | 90.1 |
| 6 | Posturing/catalepsy,Grimacing,Echopraxia/echolalia,Mannerisms,Waxy flexibility,Withdrawal | 87.2 | 84.1 | 90.1 |
| 6 | Posturing/catalepsy,Grimacing,Echopraxia/echolalia,Verbigeration,Rigidity,Negativism | 87.2 | 84.1 | 90.4 |
| 6 | Posturing/catalepsy,Grimacing,Mannerisms,Rigidity,Negativism,Waxy flexibility | 87.2 | 84.1 | 90.4 |
| 6 | Echopraxia/echolalia,Stereotypy,Mannerisms,Verbigeration,Negativism,Withdrawal | 87.0 | 83.9 | 90.1 |
| 6 | Echopraxia/echolalia,Stereotypy,Verbigeration,Negativism,Waxy flexibility,Withdrawal | 87.0 | 83.9 | 89.9 |
| 6 | Echopraxia/echolalia,Stereotypy,Verbigeration,Rigidity,Waxy flexibility,Withdrawal | 87.0 | 83.9 | 90.1 |
| 6 | Excitement,Echopraxia/echolalia,Mannerisms,Verbigeration,Rigidity,Withdrawal | 87.0 | 83.9 | 89.9 |
| 6 | Excitement,Echopraxia/echolalia,Stereotypy,Verbigeration,Negativism,Withdrawal | 87.0 | 83.9 | 90.1 |
| 6 | Excitement,Grimacing,Echopraxia/echolalia,Mannerisms,Rigidity,Withdrawal | 87.0 | 83.9 | 90.1 |
| 6 | Excitement,Grimacing,Mannerisms,Verbigeration,Rigidity,Withdrawal | 87.0 | 83.9 | 89.9 |
| 6 | Excitement,Mutism,Echopraxia/echolalia,Stereotypy,Verbigeration,Waxy flexibility | 87.0 | 83.9 | 89.9 |
| 6 | Excitement,Posturing/catalepsy,Mannerisms,Rigidity,Negativism,Waxy flexibility | 87.0 | 83.9 | 89.9 |
| 6 | Grimacing,Echopraxia/echolalia,Mannerisms,Rigidity,Waxy flexibility,Withdrawal | 87.0 | 83.9 | 90.1 |
| 6 | Grimacing,Echopraxia/echolalia,Stereotypy,Negativism,Waxy flexibility,Withdrawal | 87.0 | 83.9 | 90.1 |
| 6 | Grimacing,Mannerisms,Verbigeration,Rigidity,Waxy flexibility,Withdrawal | 87.0 | 83.9 | 90.1 |
| 6 | Mutism,Grimacing,Stereotypy,Mannerisms,Verbigeration,Withdrawal | 87.0 | 83.9 | 90.1 |
| 6 | Mutism,Posturing/catalepsy,Echopraxia/echolalia,Mannerisms,Verbigeration,Waxy flexibility | 87.0 | 83.9 | 89.9 |
| 6 | Mutism,Posturing/catalepsy,Echopraxia/echolalia,Stereotypy,Mannerisms,Waxy flexibility | 87.0 | 83.9 | 90.1 |
| 6 | Mutism,Posturing/catalepsy,Echopraxia/echolalia,Stereotypy,Verbigeration,Waxy flexibility | 87.0 | 83.9 | 89.9 |
| 6 | Mutism,Posturing/catalepsy,Grimacing,Stereotypy,Mannerisms,Waxy flexibility | 87.0 | 83.9 | 90.1 |
| 6 | Posturing/catalepsy,Grimacing,Echopraxia/echolalia,Stereotypy,Rigidity,Negativism | 87.0 | 83.9 | 90.1 |
| 6 | Posturing/catalepsy,Grimacing,Echopraxia/echolalia,Stereotypy,Verbigeration,Withdrawal | 87.0 | 83.9 | 89.9 |
| 6 | Posturing/catalepsy,Grimacing,Mannerisms,Verbigeration,Waxy flexibility,Withdrawal | 87.0 | 83.9 | 90.1 |
| 6 | Posturing/catalepsy,Grimacing,Stereotypy,Rigidity,Negativism,Waxy flexibility | 87.0 | 83.9 | 90.1 |
| 6 | Posturing/catalepsy,Grimacing,Stereotypy,Verbigeration,Waxy flexibility,Withdrawal | 87.0 | 83.6 | 90.1 |
| 6 | Posturing/catalepsy,Grimacing,Verbigeration,Rigidity,Negativism,Waxy flexibility | 87.0 | 83.9 | 89.9 |
| 6 | Posturing/catalepsy,Mannerisms,Verbigeration,Rigidity,Negativism,Waxy flexibility | 87.0 | 83.9 | 90.1 |
| 6 | Excitement,Grimacing,Echopraxia/echolalia,Negativism,Waxy flexibility,Withdrawal | 86.8 | 83.4 | 89.9 |
| 6 | Excitement,Grimacing,Echopraxia/echolalia,Stereotypy,Negativism,Withdrawal | 86.8 | 83.6 | 89.9 |
| 6 | Excitement,Grimacing,Stereotypy,Negativism,Waxy flexibility,Withdrawal | 86.8 | 83.6 | 89.9 |
| 6 | Excitement,Mutism,Echopraxia/echolalia,Stereotypy,Mannerisms,Waxy flexibility | 86.8 | 83.6 | 89.9 |
| 6 | Excitement,Mutism,Grimacing,Stereotypy,Verbigeration,Waxy flexibility | 86.8 | 83.6 | 89.7 |
| 6 | Excitement,Stereotypy,Mannerisms,Negativism,Waxy flexibility,Withdrawal | 86.8 | 83.4 | 89.9 |
| 6 | Grimacing,Echopraxia/echolalia,Verbigeration,Rigidity,Waxy flexibility,Withdrawal | 86.8 | 83.6 | 89.9 |
| 6 | Grimacing,Stereotypy,Mannerisms,Verbigeration,Rigidity,Withdrawal | 86.8 | 83.6 | 89.7 |
| 6 | Mutism,Echopraxia/echolalia,Stereotypy,Mannerisms,Verbigeration,Withdrawal | 86.8 | 83.6 | 89.9 |
| 6 | Mutism,Grimacing,Echopraxia/echolalia,Mannerisms,Verbigeration,Withdrawal | 86.8 | 83.6 | 89.9 |
| 6 | Posturing/catalepsy,Echopraxia/echolalia,Stereotypy,Mannerisms,Waxy flexibility,Withdrawal | 86.8 | 83.6 | 89.9 |
| 6 | Posturing/catalepsy,Echopraxia/echolalia,Stereotypy,Rigidity,Negativism,Waxy flexibility | 86.8 | 83.6 | 89.9 |
| 6 | Excitement,Echopraxia/echolalia,Stereotypy,Mannerisms,Negativism,Withdrawal | 86.7 | 83.6 | 89.7 |
| 6 | Mutism,Posturing/catalepsy,Echopraxia/echolalia,Stereotypy,Mannerisms,Verbigeration | 86.7 | 83.4 | 89.7 |
| 6 | Mutism,Posturing/catalepsy,Stereotypy,Mannerisms,Verbigeration,Waxy flexibility | 86.7 | 83.6 | 89.9 |
| 6 | Posturing/catalepsy,Stereotypy,Mannerisms,Verbigeration,Waxy flexibility,Withdrawal | 86.7 | 83.6 | 89.9 |
| 6 | Grimacing,Echopraxia/echolalia,Stereotypy,Verbigeration,Rigidity,Withdrawal | 86.6 | 83.4 | 89.7 |
| 6 | Excitement,Mutism,Grimacing,Echopraxia/echolalia,Verbigeration,Waxy flexibility | 86.5 | 83.4 | 89.7 |
| 6 | Grimacing,Stereotypy,Mannerisms,Verbigeration,Negativism,Withdrawal | 86.5 | 83.4 | 89.7 |
| 6 | Posturing/catalepsy,Grimacing,Echopraxia/echolalia,Rigidity,Negativism,Waxy flexibility | 86.5 | 83.2 | 89.7 |
| 6 | Posturing/catalepsy,Grimacing,Echopraxia/echolalia,Verbigeration,Waxy flexibility,Withdrawal | 86.5 | 83.4 | 89.5 |
| 6 | Grimacing,Stereotypy,Verbigeration,Negativism,Waxy flexibility,Withdrawal | 86.4 | 83.2 | 89.5 |
| 6 | Echopraxia/echolalia,Stereotypy,Mannerisms,Verbigeration,Rigidity,Withdrawal | 86.3 | 83.0 | 89.5 |
| 6 | Excitement,Echopraxia/echolalia,Verbigeration,Negativism,Waxy flexibility,Withdrawal | 86.3 | 83.2 | 89.5 |
| 6 | Excitement,Grimacing,Stereotypy,Verbigeration,Negativism,Withdrawal | 86.3 | 83.0 | 89.5 |
| 6 | Excitement,Stereotypy,Mannerisms,Verbigeration,Negativism,Withdrawal | 86.3 | 83.2 | 89.5 |
| 6 | Grimacing,Echopraxia/echolalia,Stereotypy,Verbigeration,Negativism,Withdrawal | 86.3 | 83.2 | 89.5 |
| 6 | Posturing/catalepsy,Echopraxia/echolalia,Mannerisms,Verbigeration,Waxy flexibility,Withdrawal | 86.3 | 83.0 | 89.5 |
| 6 | Posturing/catalepsy,Echopraxia/echolalia,Stereotypy,Mannerisms,Verbigeration,Withdrawal | 86.3 | 83.0 | 89.5 |
| 6 | Posturing/catalepsy,Echopraxia/echolalia,Verbigeration,Rigidity,Negativism,Waxy flexibility | 86.3 | 83.0 | 89.5 |
| 6 | Echopraxia/echolalia,Mannerisms,Verbigeration,Rigidity,Waxy flexibility,Withdrawal | 86.1 | 82.7 | 89.2 |
| 6 | Excitement,Echopraxia/echolalia,Mannerisms,Negativism,Waxy flexibility,Withdrawal | 86.1 | 82.7 | 89.2 |
| 6 | Excitement,Mutism,Grimacing,Echopraxia/echolalia,Mannerisms,Waxy flexibility | 86.1 | 82.7 | 89.2 |
| 6 | Excitement,Mutism,Grimacing,Stereotypy,Mannerisms,Waxy flexibility | 86.1 | 82.7 | 89.2 |
| 6 | Excitement,Mutism,Stereotypy,Mannerisms,Verbigeration,Waxy flexibility | 86.1 | 82.7 | 89.2 |
| 6 | Grimacing,Echopraxia/echolalia,Mannerisms,Negativism,Waxy flexibility,Withdrawal | 86.1 | 82.7 | 89.2 |
| 6 | Grimacing,Echopraxia/echolalia,Mannerisms,Verbigeration,Rigidity,Withdrawal | 86.1 | 82.7 | 89.2 |
| 6 | Posturing/catalepsy,Echopraxia/echolalia,Stereotypy,Verbigeration,Waxy flexibility,Withdrawal | 85.9 | 82.7 | 89.0 |
| 6 | Excitement,Mannerisms,Verbigeration,Negativism,Waxy flexibility,Withdrawal | 85.7 | 82.3 | 88.8 |
| 6 | Excitement,Grimacing,Echopraxia/echolalia,Verbigeration,Negativism,Withdrawal | 85.6 | 82.3 | 88.8 |
| 6 | Excitement,Grimacing,Stereotypy,Mannerisms,Negativism,Withdrawal | 85.6 | 82.3 | 88.8 |
| 6 | Mutism,Grimacing,Echopraxia/echolalia,Stereotypy,Mannerisms,Waxy flexibility | 85.6 | 82.3 | 88.8 |
| 6 | Echopraxia/echolalia,Mannerisms,Verbigeration,Negativism,Waxy flexibility,Withdrawal | 85.4 | 82.1 | 88.6 |
| 6 | Excitement,Grimacing,Mannerisms,Negativism,Waxy flexibility,Withdrawal | 85.4 | 82.1 | 88.6 |
| 6 | Excitement,Grimacing,Verbigeration,Negativism,Waxy flexibility,Withdrawal | 85.4 | 82.1 | 88.6 |
| 6 | Excitement,Mutism,Grimacing,Mannerisms,Verbigeration,Waxy flexibility | 85.4 | 82.1 | 88.8 |
| 6 | Excitement,Posturing/catalepsy,Grimacing,Echopraxia/echolalia,Negativism,Waxy flexibility | 85.4 | 82.1 | 88.6 |
| 6 | Excitement,Posturing/catalepsy,Grimacing,Echopraxia/echolalia,Verbigeration,Negativism | 85.4 | 82.1 | 88.6 |
| 6 | Excitement,Posturing/catalepsy,Grimacing,Stereotypy,Verbigeration,Negativism | 85.4 | 82.1 | 88.6 |
| 6 | Excitement,Grimacing,Echopraxia/echolalia,Mannerisms,Negativism,Withdrawal | 85.2 | 81.8 | 88.3 |
| 6 | Excitement,Mutism,Echopraxia/echolalia,Mannerisms,Verbigeration,Waxy flexibility | 85.2 | 81.8 | 88.3 |
| 6 | Excitement,Posturing/catalepsy,Grimacing,Echopraxia/echolalia,Stereotypy,Negativism | 85.2 | 81.8 | 88.3 |
| 6 | Excitement,Posturing/catalepsy,Grimacing,Verbigeration,Negativism,Waxy flexibility | 85.2 | 81.8 | 88.3 |
| 6 | Grimacing,Echopraxia/echolalia,Verbigeration,Negativism,Waxy flexibility,Withdrawal | 85.2 | 81.8 | 88.3 |
| 6 | Grimacing,Mannerisms,Verbigeration,Negativism,Waxy flexibility,Withdrawal | 85.2 | 81.8 | 88.3 |
| 6 | Mutism,Grimacing,Echopraxia/echolalia,Stereotypy,Verbigeration,Waxy flexibility | 85.2 | 81.8 | 88.3 |
| 6 | Excitement,Echopraxia/echolalia,Mannerisms,Verbigeration,Negativism,Withdrawal | 85.0 | 81.6 | 88.3 |
| 6 | Excitement,Grimacing,Mannerisms,Verbigeration,Negativism,Withdrawal | 85.0 | 81.6 | 88.1 |
| 6 | Excitement,Mutism,Grimacing,Echopraxia/echolalia,Stereotypy,Verbigeration | 85.0 | 81.6 | 88.3 |
| 6 | Excitement,Posturing/catalepsy,Grimacing,Echopraxia/echolalia,Mannerisms,Negativism | 85.0 | 81.6 | 88.3 |
| 6 | Excitement,Posturing/catalepsy,Grimacing,Echopraxia/echolalia,Rigidity,Waxy flexibility | 85.0 | 81.6 | 88.1 |
| 6 | Excitement,Posturing/catalepsy,Grimacing,Mannerisms,Verbigeration,Negativism | 85.0 | 81.6 | 88.1 |
| 6 | Excitement,Posturing/catalepsy,Grimacing,Stereotypy,Negativism,Waxy flexibility | 85.0 | 81.6 | 88.3 |
| 6 | Grimacing,Echopraxia/echolalia,Mannerisms,Verbigeration,Negativism,Withdrawal | 84.9 | 81.6 | 88.1 |
| 6 | Posturing/catalepsy,Grimacing,Echopraxia/echolalia,Stereotypy,Mannerisms,Negativism | 84.8 | 81.4 | 88.1 |
| 6 | Excitement,Grimacing,Echopraxia/echolalia,Stereotypy,Waxy flexibility,Withdrawal | 84.7 | 81.4 | 87.9 |
| 6 | Excitement,Posturing/catalepsy,Stereotypy,Verbigeration,Negativism,Waxy flexibility | 84.7 | 81.4 | 87.9 |
| 6 | Mutism,Grimacing,Echopraxia/echolalia,Mannerisms,Verbigeration,Waxy flexibility | 84.7 | 81.4 | 87.9 |
| 6 | Mutism,Grimacing,Stereotypy,Mannerisms,Verbigeration,Waxy flexibility | 84.7 | 81.4 | 88.1 |
| 6 | Excitement,Posturing/catalepsy,Grimacing,Verbigeration,Rigidity,Waxy flexibility | 84.6 | 81.2 | 87.9 |
| 6 | Posturing/catalepsy,Grimacing,Stereotypy,Mannerisms,Negativism,Waxy flexibility | 84.6 | 80.9 | 87.9 |
| 6 | Posturing/catalepsy,Grimacing,Stereotypy,Verbigeration,Negativism,Waxy flexibility | 84.6 | 81.2 | 87.9 |
| 6 | Excitement,Echopraxia/echolalia,Stereotypy,Verbigeration,Waxy flexibility,Withdrawal | 84.5 | 81.2 | 87.7 |
| 6 | Excitement,Mutism,Grimacing,Echopraxia/echolalia,Stereotypy,Mannerisms | 84.5 | 81.2 | 87.7 |
| 6 | Excitement,Posturing/catalepsy,Echopraxia/echolalia,Stereotypy,Negativism,Waxy flexibility | 84.5 | 80.9 | 87.9 |
| 6 | Excitement,Posturing/catalepsy,Grimacing,Echopraxia/echolalia,Verbigeration,Rigidity | 84.5 | 81.2 | 87.7 |
| 6 | Excitement,Posturing/catalepsy,Grimacing,Mannerisms,Negativism,Waxy flexibility | 84.5 | 81.2 | 87.9 |
| 6 | Excitement,Posturing/catalepsy,Grimacing,Stereotypy,Mannerisms,Negativism | 84.5 | 81.2 | 87.7 |
| 6 | Mutism,Echopraxia/echolalia,Stereotypy,Mannerisms,Verbigeration,Waxy flexibility | 84.5 | 80.9 | 87.7 |
| 6 | Posturing/catalepsy,Grimacing,Stereotypy,Mannerisms,Verbigeration,Negativism | 84.5 | 80.9 | 87.9 |
| 6 | Excitement,Echopraxia/echolalia,Stereotypy,Mannerisms,Waxy flexibility,Withdrawal | 84.3 | 80.7 | 87.7 |
| 6 | Excitement,Grimacing,Stereotypy,Rigidity,Negativism,Waxy flexibility | 84.3 | 80.9 | 87.7 |
| 6 | Excitement,Mutism,Echopraxia/echolalia,Stereotypy,Mannerisms,Verbigeration | 84.3 | 80.9 | 87.7 |
| 6 | Excitement,Posturing/catalepsy,Echopraxia/echolalia,Stereotypy,Verbigeration,Negativism | 84.3 | 80.9 | 87.4 |
| 6 | Grimacing,Echopraxia/echolalia,Stereotypy,Mannerisms,Waxy flexibility,Withdrawal | 84.3 | 80.9 | 87.7 |
| 6 | Grimacing,Stereotypy,Mannerisms,Rigidity,Negativism,Waxy flexibility | 84.3 | 80.7 | 87.7 |
| 6 | Posturing/catalepsy,Grimacing,Echopraxia/echolalia,Stereotypy,Negativism,Waxy flexibility | 84.3 | 80.9 | 87.7 |
| 6 | Echopraxia/echolalia,Stereotypy,Mannerisms,Rigidity,Negativism,Waxy flexibility | 84.1 | 80.5 | 87.4 |
| 6 | Excitement,Echopraxia/echolalia,Stereotypy,Rigidity,Negativism,Waxy flexibility | 84.1 | 80.7 | 87.4 |
| 6 | Excitement,Grimacing,Echopraxia/echolalia,Verbigeration,Waxy flexibility,Withdrawal | 84.1 | 80.7 | 87.4 |
| 6 | Excitement,Mutism,Grimacing,Stereotypy,Mannerisms,Verbigeration | 84.1 | 80.7 | 87.4 |
| 6 | Excitement,Posturing/catalepsy,Echopraxia/echolalia,Verbigeration,Negativism,Waxy flexibility | 84.1 | 80.7 | 87.2 |
| 6 | Posturing/catalepsy,Grimacing,Echopraxia/echolalia,Mannerisms,Negativism,Waxy flexibility | 84.1 | 80.5 | 87.4 |
| 6 | Posturing/catalepsy,Grimacing,Echopraxia/echolalia,Mannerisms,Verbigeration,Negativism | 84.1 | 80.7 | 87.4 |
| 6 | Posturing/catalepsy,Grimacing,Echopraxia/echolalia,Stereotypy,Verbigeration,Negativism | 84.1 | 80.7 | 87.4 |
| 6 | Excitement,Posturing/catalepsy,Stereotypy,Mannerisms,Verbigeration,Negativism | 84.0 | 80.5 | 87.4 |
| 6 | Excitement,Posturing/catalepsy,Echopraxia/echolalia,Stereotypy,Mannerisms,Negativism | 83.9 | 80.3 | 87.2 |
| 6 | Excitement,Posturing/catalepsy,Grimacing,Echopraxia/echolalia,Mannerisms,Rigidity | 83.9 | 80.5 | 87.2 |
| 6 | Excitement,Stereotypy,Verbigeration,Rigidity,Negativism,Waxy flexibility | 83.9 | 80.5 | 87.2 |
| 6 | Posturing/catalepsy,Echopraxia/echolalia,Stereotypy,Mannerisms,Negativism,Waxy flexibility | 83.9 | 80.3 | 87.2 |
| 6 | Stereotypy,Mannerisms,Verbigeration,Rigidity,Negativism,Waxy flexibility | 83.9 | 80.3 | 87.2 |
| 6 | Grimacing,Stereotypy,Verbigeration,Rigidity,Negativism,Waxy flexibility | 83.8 | 80.3 | 87.2 |
| 6 | Posturing/catalepsy,Grimacing,Echopraxia/echolalia,Verbigeration,Negativism,Waxy flexibility | 83.8 | 80.3 | 87.2 |
| 6 | Posturing/catalepsy,Grimacing,Mannerisms,Verbigeration,Negativism,Waxy flexibility | 83.8 | 80.5 | 87.2 |
| 6 | Excitement,Mutism,Grimacing,Echopraxia/echolalia,Mannerisms,Verbigeration | 83.7 | 80.3 | 87.0 |
| 6 | Excitement,Posturing/catalepsy,Echopraxia/echolalia,Mannerisms,Negativism,Waxy flexibility | 83.7 | 80.3 | 87.0 |
| 6 | Excitement,Posturing/catalepsy,Echopraxia/echolalia,Verbigeration,Rigidity,Waxy flexibility | 83.7 | 80.3 | 87.0 |
| 6 | Excitement,Posturing/catalepsy,Grimacing,Echopraxia/echolalia,Stereotypy,Rigidity | 83.7 | 80.3 | 87.0 |
| 6 | Excitement,Posturing/catalepsy,Mannerisms,Verbigeration,Negativism,Waxy flexibility | 83.7 | 80.3 | 87.0 |
| 6 | Posturing/catalepsy,Stereotypy,Mannerisms,Verbigeration,Negativism,Waxy flexibility | 83.7 | 80.3 | 87.0 |
| 6 | Excitement,Grimacing,Echopraxia/echolalia,Mannerisms,Waxy flexibility,Withdrawal | 83.6 | 80.0 | 87.0 |
| 6 | Excitement,Grimacing,Echopraxia/echolalia,Rigidity,Negativism,Waxy flexibility | 83.6 | 80.0 | 87.0 |
| 6 | Excitement,Posturing/catalepsy,Grimacing,Mannerisms,Verbigeration,Rigidity | 83.6 | 80.0 | 87.0 |
| 6 | Excitement,Posturing/catalepsy,Grimacing,Stereotypy,Rigidity,Waxy flexibility | 83.6 | 80.0 | 87.0 |
| 6 | Excitement,Posturing/catalepsy,Grimacing,Stereotypy,Verbigeration,Rigidity | 83.6 | 80.0 | 87.0 |
| 6 | Excitement,Posturing/catalepsy,Stereotypy,Mannerisms,Negativism,Waxy flexibility | 83.6 | 80.0 | 87.0 |
| 6 | Excitement,Grimacing,Echopraxia/echolalia,Stereotypy,Rigidity,Negativism | 83.4 | 79.8 | 86.8 |
| 6 | Excitement,Grimacing,Stereotypy,Verbigeration,Waxy flexibility,Withdrawal | 83.4 | 79.8 | 86.8 |
| 6 | Excitement,Posturing/catalepsy,Echopraxia/echolalia,Mannerisms,Verbigeration,Negativism | 83.4 | 80.0 | 86.8 |
| 6 | Excitement,Posturing/catalepsy,Grimacing,Mannerisms,Rigidity,Waxy flexibility | 83.4 | 79.8 | 86.8 |
| 6 | Excitement,Stereotypy,Mannerisms,Verbigeration,Waxy flexibility,Withdrawal | 83.4 | 80.0 | 86.8 |
| 6 | Grimacing,Echopraxia/echolalia,Stereotypy,Rigidity,Negativism,Waxy flexibility | 83.4 | 80.0 | 86.8 |
| 6 | Grimacing,Echopraxia/echolalia,Stereotypy,Verbigeration,Waxy flexibility,Withdrawal | 83.4 | 79.8 | 86.8 |
| 6 | Mutism,Grimacing,Echopraxia/echolalia,Stereotypy,Mannerisms,Verbigeration | 83.4 | 80.0 | 86.8 |
| 6 | Excitement,Grimacing,Stereotypy,Mannerisms,Waxy flexibility,Withdrawal | 83.2 | 79.6 | 86.5 |
| 6 | Excitement,Posturing/catalepsy,Stereotypy,Verbigeration,Rigidity,Waxy flexibility | 83.2 | 79.6 | 86.5 |
| 6 | Excitement,Stereotypy,Mannerisms,Rigidity,Negativism,Waxy flexibility | 83.2 | 79.6 | 86.5 |
| 6 | Grimacing,Echopraxia/echolalia,Stereotypy,Mannerisms,Rigidity,Negativism | 83.2 | 79.6 | 86.5 |
| 6 | Posturing/catalepsy,Echopraxia/echolalia,Stereotypy,Mannerisms,Verbigeration,Negativism | 83.2 | 79.6 | 86.5 |
| 6 | Posturing/catalepsy,Echopraxia/echolalia,Stereotypy,Verbigeration,Negativism,Waxy flexibility | 83.2 | 79.6 | 86.5 |
| 6 | Excitement,Posturing/catalepsy,Echopraxia/echolalia,Stereotypy,Rigidity,Waxy flexibility | 83.1 | 79.6 | 86.5 |
| 6 | Excitement,Echopraxia/echolalia,Mannerisms,Verbigeration,Waxy flexibility,Withdrawal | 83.0 | 79.4 | 86.3 |
| 6 | Excitement,Posturing/catalepsy,Echopraxia/echolalia,Stereotypy,Verbigeration,Rigidity | 83.0 | 79.4 | 86.3 |
| 6 | Grimacing,Echopraxia/echolalia,Mannerisms,Rigidity,Negativism,Waxy flexibility | 83.0 | 79.4 | 86.3 |
| 6 | Grimacing,Stereotypy,Mannerisms,Verbigeration,Rigidity,Negativism | 83.0 | 79.4 | 86.3 |
| 6 | Excitement,Grimacing,Stereotypy,Verbigeration,Rigidity,Negativism | 82.9 | 79.4 | 86.3 |
| 6 | Echopraxia/echolalia,Stereotypy,Verbigeration,Rigidity,Negativism,Waxy flexibility | 82.8 | 79.1 | 86.1 |
| 6 | Excitement,Grimacing,Verbigeration,Rigidity,Negativism,Waxy flexibility | 82.7 | 79.1 | 86.1 |
| 6 | Excitement,Posturing/catalepsy,Echopraxia/echolalia,Mannerisms,Rigidity,Waxy flexibility | 82.7 | 79.1 | 86.3 |
| 6 | Excitement,Posturing/catalepsy,Echopraxia/echolalia,Mannerisms,Verbigeration,Rigidity | 82.7 | 79.1 | 86.1 |
| 6 | Grimacing,Stereotypy,Mannerisms,Verbigeration,Waxy flexibility,Withdrawal | 82.7 | 79.1 | 86.1 |
| 6 | Excitement,Echopraxia/echolalia,Stereotypy,Mannerisms,Rigidity,Negativism | 82.5 | 78.9 | 86.1 |
| 6 | Excitement,Echopraxia/echolalia,Stereotypy,Verbigeration,Rigidity,Negativism | 82.5 | 78.9 | 86.1 |
| 6 | Excitement,Grimacing,Mannerisms,Verbigeration,Waxy flexibility,Withdrawal | 82.5 | 78.9 | 86.1 |
| 6 | Excitement,Grimacing,Stereotypy,Mannerisms,Rigidity,Negativism | 82.5 | 78.9 | 85.9 |
| 6 | Excitement,Posturing/catalepsy,Grimacing,Stereotypy,Mannerisms,Rigidity | 82.5 | 78.9 | 85.9 |
| 6 | Excitement,Posturing/catalepsy,Mannerisms,Verbigeration,Rigidity,Waxy flexibility | 82.5 | 78.9 | 85.9 |
| 6 | Grimacing,Echopraxia/echolalia,Mannerisms,Verbigeration,Waxy flexibility,Withdrawal | 82.5 | 78.9 | 85.9 |
| 6 | Posturing/catalepsy,Echopraxia/echolalia,Mannerisms,Verbigeration,Negativism,Waxy flexibility | 82.5 | 78.9 | 85.9 |
| 6 | Posturing/catalepsy,Grimacing,Echopraxia/echolalia,Verbigeration,Rigidity,Waxy flexibility | 82.5 | 78.9 | 86.1 |
| 6 | Posturing/catalepsy,Grimacing,Stereotypy,Verbigeration,Rigidity,Waxy flexibility | 82.5 | 78.9 | 85.9 |
| 6 | Echopraxia/echolalia,Stereotypy,Mannerisms,Verbigeration,Rigidity,Negativism | 82.3 | 78.7 | 85.9 |
| 6 | Echopraxia/echolalia,Stereotypy,Mannerisms,Verbigeration,Waxy flexibility,Withdrawal | 82.3 | 78.7 | 85.9 |
| 6 | Excitement,Grimacing,Mannerisms,Rigidity,Negativism,Waxy flexibility | 82.3 | 78.7 | 85.9 |
| 6 | Grimacing,Echopraxia/echolalia,Stereotypy,Verbigeration,Rigidity,Negativism | 82.3 | 78.7 | 85.7 |
| 6 | Posturing/catalepsy,Grimacing,Echopraxia/echolalia,Mannerisms,Rigidity,Waxy flexibility | 82.3 | 78.7 | 85.9 |
| 6 | Posturing/catalepsy,Grimacing,Echopraxia/echolalia,Mannerisms,Verbigeration,Rigidity | 82.3 | 78.7 | 85.9 |
| 6 | Excitement,Grimacing,Echopraxia/echolalia,Verbigeration,Rigidity,Negativism | 82.1 | 78.5 | 85.7 |
| 6 | Excitement,Posturing/catalepsy,Echopraxia/echolalia,Stereotypy,Mannerisms,Rigidity | 82.1 | 78.5 | 85.7 |
| 6 | Excitement,Stereotypy,Mannerisms,Verbigeration,Rigidity,Negativism | 82.1 | 78.5 | 85.4 |
| 6 | Grimacing,Echopraxia/echolalia,Verbigeration,Rigidity,Negativism,Waxy flexibility | 82.1 | 78.5 | 85.4 |
| 6 | Posturing/catalepsy,Grimacing,Echopraxia/echolalia,Stereotypy,Rigidity,Waxy flexibility | 82.1 | 78.5 | 85.4 |
| 6 | Posturing/catalepsy,Grimacing,Echopraxia/echolalia,Stereotypy,Verbigeration,Rigidity | 82.1 | 78.5 | 85.7 |
| 6 | Posturing/catalepsy,Grimacing,Mannerisms,Verbigeration,Rigidity,Waxy flexibility | 82.1 | 78.5 | 85.7 |
| 6 | Posturing/catalepsy,Grimacing,Stereotypy,Mannerisms,Verbigeration,Rigidity | 82.1 | 78.5 | 85.4 |
| 6 | Excitement,Grimacing,Echopraxia/echolalia,Stereotypy,Verbigeration,Withdrawal | 82.0 | 78.3 | 85.7 |
| 6 | Excitement,Posturing/catalepsy,Stereotypy,Mannerisms,Verbigeration,Rigidity | 82.0 | 78.3 | 85.7 |
| 6 | Grimacing,Mannerisms,Verbigeration,Rigidity,Negativism,Waxy flexibility | 82.0 | 78.5 | 85.4 |
| 6 | Excitement,Grimacing,Echopraxia/echolalia,Mannerisms,Rigidity,Negativism | 81.8 | 78.0 | 85.4 |
| 6 | Excitement,Grimacing,Echopraxia/echolalia,Stereotypy,Mannerisms,Withdrawal | 81.8 | 78.3 | 85.4 |
| 6 | Posturing/catalepsy,Grimacing,Echopraxia/echolalia,Stereotypy,Mannerisms,Rigidity | 81.8 | 78.3 | 85.4 |
| 6 | Posturing/catalepsy,Grimacing,Stereotypy,Mannerisms,Rigidity,Waxy flexibility | 81.8 | 78.3 | 85.4 |
| 6 | Excitement,Echopraxia/echolalia,Stereotypy,Mannerisms,Verbigeration,Withdrawal | 81.6 | 78.0 | 85.2 |
| 6 | Excitement,Posturing/catalepsy,Stereotypy,Mannerisms,Rigidity,Waxy flexibility | 81.6 | 78.0 | 85.2 |
| 6 | Grimacing,Echopraxia/echolalia,Stereotypy,Mannerisms,Verbigeration,Withdrawal | 81.6 | 78.0 | 85.2 |
| 6 | Excitement,Echopraxia/echolalia,Mannerisms,Rigidity,Negativism,Waxy flexibility | 81.4 | 77.6 | 85.0 |
| 6 | Excitement,Echopraxia/echolalia,Verbigeration,Rigidity,Negativism,Waxy flexibility | 81.4 | 77.6 | 85.0 |
| 6 | Excitement,Grimacing,Mannerisms,Verbigeration,Rigidity,Negativism | 81.4 | 77.6 | 85.0 |
| 6 | Excitement,Grimacing,Echopraxia/echolalia,Mannerisms,Verbigeration,Withdrawal | 81.2 | 77.6 | 84.8 |
| 6 | Grimacing,Echopraxia/echolalia,Mannerisms,Verbigeration,Rigidity,Negativism | 81.2 | 77.6 | 84.8 |
| 6 | Echopraxia/echolalia,Mannerisms,Verbigeration,Rigidity,Negativism,Waxy flexibility | 80.9 | 77.1 | 84.5 |
| 6 | Excitement,Grimacing,Stereotypy,Mannerisms,Verbigeration,Withdrawal | 80.9 | 77.1 | 84.5 |
| 6 | Excitement,Mannerisms,Verbigeration,Rigidity,Negativism,Waxy flexibility | 80.9 | 77.4 | 84.5 |
| 6 | Posturing/catalepsy,Echopraxia/echolalia,Stereotypy,Verbigeration,Rigidity,Waxy flexibility | 80.9 | 77.1 | 84.5 |
| 6 | Posturing/catalepsy,Stereotypy,Mannerisms,Verbigeration,Rigidity,Waxy flexibility | 80.7 | 76.9 | 84.3 |
| 6 | Posturing/catalepsy,Echopraxia/echolalia,Mannerisms,Verbigeration,Rigidity,Waxy flexibility | 80.5 | 76.7 | 84.1 |
| 6 | Posturing/catalepsy,Echopraxia/echolalia,Stereotypy,Mannerisms,Rigidity,Waxy flexibility | 80.5 | 76.7 | 84.1 |
| 6 | Posturing/catalepsy,Echopraxia/echolalia,Stereotypy,Mannerisms,Verbigeration,Rigidity | 80.5 | 76.7 | 84.1 |
| 6 | Excitement,Posturing/catalepsy,Grimacing,Echopraxia/echolalia,Verbigeration,Waxy flexibility | 80.1 | 76.2 | 83.6 |
| 6 | Excitement,Echopraxia/echolalia,Mannerisms,Verbigeration,Rigidity,Negativism | 79.8 | 76.0 | 83.4 |
| 6 | Excitement,Grimacing,Echopraxia/echolalia,Stereotypy,Rigidity,Waxy flexibility | 79.6 | 75.8 | 83.2 |
| 6 | Excitement,Posturing/catalepsy,Grimacing,Echopraxia/echolalia,Stereotypy,Waxy flexibility | 79.6 | 75.8 | 83.2 |
| 6 | Excitement,Posturing/catalepsy,Grimacing,Stereotypy,Verbigeration,Waxy flexibility | 79.6 | 75.8 | 83.2 |
| 6 | Excitement,Posturing/catalepsy,Grimacing,Echopraxia/echolalia,Mannerisms,Waxy flexibility | 79.4 | 75.6 | 83.2 |
| 6 | Grimacing,Echopraxia/echolalia,Stereotypy,Mannerisms,Negativism,Waxy flexibility | 79.1 | 75.3 | 83.0 |
| 6 | Excitement,Grimacing,Stereotypy,Verbigeration,Rigidity,Waxy flexibility | 78.9 | 75.1 | 82.5 |
| 6 | Excitement,Posturing/catalepsy,Grimacing,Mannerisms,Verbigeration,Waxy flexibility | 78.9 | 75.1 | 82.7 |
| 6 | Excitement,Grimacing,Echopraxia/echolalia,Stereotypy,Negativism,Waxy flexibility | 78.7 | 74.9 | 82.5 |
| 6 | Excitement,Grimacing,Echopraxia/echolalia,Verbigeration,Rigidity,Waxy flexibility | 78.7 | 74.9 | 82.5 |
| 6 | Excitement,Posturing/catalepsy,Grimacing,Echopraxia/echolalia,Mannerisms,Verbigeration | 78.7 | 74.7 | 82.5 |
| 6 | Excitement,Posturing/catalepsy,Grimacing,Echopraxia/echolalia,Stereotypy,Verbigeration | 78.7 | 74.7 | 82.3 |
| 6 | Excitement,Posturing/catalepsy,Grimacing,Echopraxia/echolalia,Stereotypy,Mannerisms | 78.3 | 74.4 | 82.1 |
| 6 | Excitement,Posturing/catalepsy,Grimacing,Stereotypy,Mannerisms,Verbigeration | 78.3 | 74.2 | 82.3 |
| 6 | Excitement,Grimacing,Stereotypy,Verbigeration,Negativism,Waxy flexibility | 78.2 | 74.2 | 82.1 |
| 6 | Excitement,Grimacing,Stereotypy,Mannerisms,Rigidity,Waxy flexibility | 78.0 | 74.0 | 81.8 |
| 6 | Excitement,Posturing/catalepsy,Grimacing,Stereotypy,Mannerisms,Waxy flexibility | 78.0 | 74.2 | 81.8 |
| 6 | Grimacing,Stereotypy,Mannerisms,Verbigeration,Negativism,Waxy flexibility | 78.0 | 74.2 | 81.8 |
| 6 | Excitement,Grimacing,Stereotypy,Mannerisms,Negativism,Waxy flexibility | 77.8 | 74.0 | 81.6 |
| 6 | Grimacing,Echopraxia/echolalia,Stereotypy,Mannerisms,Rigidity,Waxy flexibility | 77.8 | 74.0 | 81.6 |
| 6 | Posturing/catalepsy,Grimacing,Echopraxia/echolalia,Stereotypy,Verbigeration,Waxy flexibility | 77.8 | 74.0 | 81.6 |
| 6 | Excitement,Grimacing,Echopraxia/echolalia,Mannerisms,Rigidity,Waxy flexibility | 77.6 | 73.5 | 81.4 |
| 6 | Grimacing,Echopraxia/echolalia,Stereotypy,Verbigeration,Negativism,Waxy flexibility | 77.6 | 73.5 | 81.4 |
| 6 | Grimacing,Echopraxia/echolalia,Stereotypy,Verbigeration,Rigidity,Waxy flexibility | 77.6 | 73.8 | 81.4 |
| 6 | Posturing/catalepsy,Grimacing,Echopraxia/echolalia,Stereotypy,Mannerisms,Waxy flexibility | 77.6 | 73.8 | 81.4 |
| 6 | Posturing/catalepsy,Grimacing,Stereotypy,Mannerisms,Verbigeration,Waxy flexibility | 77.6 | 73.8 | 81.4 |
| 6 | Excitement,Echopraxia/echolalia,Stereotypy,Mannerisms,Negativism,Waxy flexibility | 77.4 | 73.5 | 81.2 |
| 6 | Excitement,Posturing/catalepsy,Echopraxia/echolalia,Stereotypy,Verbigeration,Waxy flexibility | 77.4 | 73.5 | 81.2 |
| 6 | Grimacing,Stereotypy,Mannerisms,Verbigeration,Rigidity,Waxy flexibility | 77.4 | 73.5 | 81.2 |
| 6 | Posturing/catalepsy,Grimacing,Echopraxia/echolalia,Mannerisms,Verbigeration,Waxy flexibility | 77.4 | 73.5 | 81.2 |
| 6 | Excitement,Echopraxia/echolalia,Stereotypy,Verbigeration,Negativism,Waxy flexibility | 77.2 | 73.1 | 80.9 |
| 6 | Excitement,Echopraxia/echolalia,Stereotypy,Verbigeration,Rigidity,Waxy flexibility | 77.1 | 73.3 | 80.9 |
| 6 | Echopraxia/echolalia,Stereotypy,Mannerisms,Verbigeration,Negativism,Waxy flexibility | 76.9 | 72.9 | 80.7 |
| 6 | Excitement,Grimacing,Echopraxia/echolalia,Mannerisms,Negativism,Waxy flexibility | 76.9 | 72.9 | 80.7 |
| 6 | Excitement,Grimacing,Echopraxia/echolalia,Verbigeration,Negativism,Waxy flexibility | 76.9 | 72.9 | 80.7 |
| 6 | Excitement,Grimacing,Mannerisms,Verbigeration,Rigidity,Waxy flexibility | 76.9 | 73.1 | 80.7 |
| 6 | Posturing/catalepsy,Grimacing,Echopraxia/echolalia,Stereotypy,Mannerisms,Verbigeration | 76.9 | 73.1 | 80.7 |
| 6 | Excitement,Echopraxia/echolalia,Stereotypy,Mannerisms,Rigidity,Waxy flexibility | 76.7 | 72.6 | 80.5 |
| 6 | Excitement,Grimacing,Echopraxia/echolalia,Stereotypy,Verbigeration,Rigidity | 76.7 | 72.6 | 80.5 |
| 6 | Excitement,Stereotypy,Mannerisms,Verbigeration,Negativism,Waxy flexibility | 76.7 | 72.6 | 80.5 |
| 6 | Excitement,Grimacing,Echopraxia/echolalia,Stereotypy,Mannerisms,Rigidity | 76.5 | 72.4 | 80.3 |
| 6 | Excitement,Posturing/catalepsy,Echopraxia/echolalia,Mannerisms,Verbigeration,Waxy flexibility | 76.4 | 72.4 | 80.3 |
| 6 | Excitement,Posturing/catalepsy,Stereotypy,Mannerisms,Verbigeration,Waxy flexibility | 76.4 | 72.4 | 80.3 |
| 6 | Excitement,Posturing/catalepsy,Echopraxia/echolalia,Stereotypy,Mannerisms,Waxy flexibility | 76.2 | 72.2 | 80.0 |
| 6 | Grimacing,Echopraxia/echolalia,Mannerisms,Verbigeration,Rigidity,Waxy flexibility | 76.2 | 72.4 | 80.0 |
| 6 | Excitement,Grimacing,Echopraxia/echolalia,Stereotypy,Mannerisms,Negativism | 76.0 | 72.0 | 79.8 |
| 6 | Excitement,Grimacing,Echopraxia/echolalia,Stereotypy,Verbigeration,Negativism | 76.0 | 72.0 | 79.8 |
| 6 | Grimacing,Echopraxia/echolalia,Mannerisms,Verbigeration,Negativism,Waxy flexibility | 76.0 | 72.0 | 80.0 |
| 6 | Excitement,Grimacing,Mannerisms,Verbigeration,Negativism,Waxy flexibility | 75.8 | 71.7 | 79.6 |
| 6 | Excitement,Posturing/catalepsy,Echopraxia/echolalia,Stereotypy,Mannerisms,Verbigeration | 75.8 | 72.0 | 79.6 |
| 6 | Excitement,Stereotypy,Mannerisms,Verbigeration,Rigidity,Waxy flexibility | 75.8 | 71.7 | 79.8 |
| 6 | Grimacing,Echopraxia/echolalia,Stereotypy,Mannerisms,Verbigeration,Negativism | 75.8 | 71.7 | 79.6 |
| 6 | Excitement,Grimacing,Stereotypy,Mannerisms,Verbigeration,Rigidity | 75.7 | 71.7 | 79.6 |
| 6 | Excitement,Grimacing,Echopraxia/echolalia,Mannerisms,Verbigeration,Rigidity | 75.4 | 71.3 | 79.1 |
| 6 | Excitement,Grimacing,Stereotypy,Mannerisms,Verbigeration,Negativism | 75.1 | 70.9 | 78.9 |
| 6 | Grimacing,Echopraxia/echolalia,Stereotypy,Mannerisms,Verbigeration,Rigidity | 75.1 | 71.1 | 78.9 |
| 6 | Echopraxia/echolalia,Stereotypy,Mannerisms,Verbigeration,Rigidity,Waxy flexibility | 74.7 | 70.6 | 78.5 |
| 6 | Posturing/catalepsy,Echopraxia/echolalia,Stereotypy,Mannerisms,Verbigeration,Waxy flexibility | 74.5 | 70.4 | 78.5 |
| 6 | Excitement,Echopraxia/echolalia,Mannerisms,Verbigeration,Rigidity,Waxy flexibility | 74.2 | 70.2 | 78.3 |
| 6 | Excitement,Echopraxia/echolalia,Stereotypy,Mannerisms,Verbigeration,Negativism | 74.2 | 70.2 | 78.3 |
| 6 | Excitement,Echopraxia/echolalia,Stereotypy,Mannerisms,Verbigeration,Rigidity | 74.0 | 70.0 | 78.0 |
| 6 | Excitement,Echopraxia/echolalia,Mannerisms,Verbigeration,Negativism,Waxy flexibility | 73.6 | 69.3 | 77.6 |
| 6 | Excitement,Grimacing,Echopraxia/echolalia,Mannerisms,Verbigeration,Negativism | 73.5 | 69.3 | 77.6 |
| 6 | Excitement,Grimacing,Echopraxia/echolalia,Stereotypy,Verbigeration,Waxy flexibility | 70.4 | 66.1 | 74.7 |
| 6 | Excitement,Grimacing,Echopraxia/echolalia,Stereotypy,Mannerisms,Waxy flexibility | 69.7 | 65.5 | 74.0 |
| 6 | Excitement,Grimacing,Stereotypy,Mannerisms,Verbigeration,Waxy flexibility | 68.8 | 64.3 | 72.9 |
| 6 | Grimacing,Echopraxia/echolalia,Stereotypy,Mannerisms,Verbigeration,Waxy flexibility | 68.8 | 64.6 | 73.1 |
| 6 | Excitement,Grimacing,Echopraxia/echolalia,Mannerisms,Verbigeration,Waxy flexibility | 67.7 | 63.5 | 72.0 |
| 6 | Excitement,Echopraxia/echolalia,Stereotypy,Mannerisms,Verbigeration,Waxy flexibility | 66.1 | 61.7 | 70.4 |
| 6 | Excitement,Grimacing,Echopraxia/echolalia,Stereotypy,Mannerisms,Verbigeration | 63.9 | 59.4 | 68.4 |

Table S7: sensitivity (relative to the full BFCSI) of all seven question combinations, and the 95% confidence interval derived from bootstrapping, sorted by decreasing sensitivity.

|  |  |  | 95% CI | |
| --- | --- | --- | --- | --- |
| # of Qs | Combination | Sensitivity | Lower | Upper |
| 7 | Excitement,Mutism,Staring,Posturing/catalepsy,Echopraxia/echolalia,Negativism,Withdrawal | 100.0 | 100.0 | 100.0 |
| 7 | Excitement,Mutism,Staring,Posturing/catalepsy,Echopraxia/echolalia,Rigidity,Withdrawal | 100.0 | 100.0 | 100.0 |
| 7 | Excitement,Mutism,Staring,Echopraxia/echolalia,Rigidity,Negativism,Withdrawal | 99.8 | 99.3 | 100.0 |
| 7 | Excitement,Mutism,Staring,Echopraxia/echolalia,Stereotypy,Rigidity,Withdrawal | 99.8 | 99.3 | 100.0 |
| 7 | Excitement,Mutism,Staring,Posturing/catalepsy,Echopraxia/echolalia,Negativism,Waxy flexibility | 99.8 | 99.3 | 100.0 |
| 7 | Excitement,Mutism,Staring,Posturing/catalepsy,Echopraxia/echolalia,Rigidity,Negativism | 99.8 | 99.3 | 100.0 |
| 7 | Excitement,Mutism,Staring,Posturing/catalepsy,Echopraxia/echolalia,Verbigeration,Negativism | 99.8 | 99.3 | 100.0 |
| 7 | Excitement,Mutism,Staring,Posturing/catalepsy,Echopraxia/echolalia,Verbigeration,Rigidity | 99.8 | 99.3 | 100.0 |
| 7 | Excitement,Mutism,Staring,Posturing/catalepsy,Mannerisms,Negativism,Withdrawal | 99.8 | 99.3 | 100.0 |
| 7 | Excitement,Mutism,Staring,Posturing/catalepsy,Mannerisms,Rigidity,Withdrawal | 99.8 | 99.3 | 100.0 |
| 7 | Excitement,Mutism,Staring,Posturing/catalepsy,Stereotypy,Negativism,Withdrawal | 99.8 | 99.3 | 100.0 |
| 7 | Excitement,Mutism,Staring,Posturing/catalepsy,Verbigeration,Negativism,Withdrawal | 99.8 | 99.3 | 100.0 |
| 7 | Excitement,Mutism,Staring,Stereotypy,Rigidity,Waxy flexibility,Withdrawal | 99.8 | 99.3 | 100.0 |
| 7 | Excitement,Immobility/stupor,Mutism,Staring,Echopraxia/echolalia,Rigidity,Withdrawal | 99.6 | 98.9 | 100.0 |
| 7 | Excitement,Immobility/stupor,Mutism,Staring,Posturing/catalepsy,Echopraxia/echolalia,Negativism | 99.6 | 98.9 | 100.0 |
| 7 | Excitement,Immobility/stupor,Staring,Echopraxia/echolalia,Rigidity,Negativism,Withdrawal | 99.6 | 98.9 | 100.0 |
| 7 | Excitement,Immobility/stupor,Staring,Posturing/catalepsy,Echopraxia/echolalia,Negativism,Withdrawal | 99.6 | 98.9 | 100.0 |
| 7 | Excitement,Immobility/stupor,Staring,Posturing/catalepsy,Mannerisms,Rigidity,Withdrawal | 99.6 | 98.9 | 100.0 |
| 7 | Excitement,Mutism,Staring,Echopraxia/echolalia,Rigidity,Waxy flexibility,Withdrawal | 99.6 | 98.9 | 100.0 |
| 7 | Excitement,Mutism,Staring,Echopraxia/echolalia,Stereotypy,Negativism,Withdrawal | 99.6 | 98.9 | 100.0 |
| 7 | Excitement,Mutism,Staring,Echopraxia/echolalia,Stereotypy,Rigidity,Negativism | 99.6 | 98.9 | 100.0 |
| 7 | Excitement,Mutism,Staring,Echopraxia/echolalia,Verbigeration,Rigidity,Negativism | 99.6 | 98.9 | 100.0 |
| 7 | Excitement,Mutism,Staring,Grimacing,Echopraxia/echolalia,Rigidity,Withdrawal | 99.6 | 98.9 | 100.0 |
| 7 | Excitement,Mutism,Staring,Posturing/catalepsy,Echopraxia/echolalia,Mannerisms,Rigidity | 99.6 | 98.9 | 100.0 |
| 7 | Excitement,Mutism,Staring,Posturing/catalepsy,Echopraxia/echolalia,Rigidity,Waxy flexibility | 99.6 | 98.9 | 100.0 |
| 7 | Excitement,Mutism,Staring,Posturing/catalepsy,Echopraxia/echolalia,Stereotypy,Rigidity | 99.6 | 98.9 | 100.0 |
| 7 | Excitement,Mutism,Staring,Posturing/catalepsy,Grimacing,Echopraxia/echolalia,Negativism | 99.6 | 98.9 | 100.0 |
| 7 | Excitement,Mutism,Staring,Posturing/catalepsy,Grimacing,Negativism,Withdrawal | 99.6 | 98.9 | 100.0 |
| 7 | Excitement,Mutism,Staring,Posturing/catalepsy,Grimacing,Rigidity,Withdrawal | 99.6 | 98.9 | 100.0 |
| 7 | Excitement,Mutism,Staring,Posturing/catalepsy,Mannerisms,Verbigeration,Negativism | 99.6 | 98.9 | 100.0 |
| 7 | Excitement,Mutism,Staring,Posturing/catalepsy,Negativism,Waxy flexibility,Withdrawal | 99.6 | 98.9 | 100.0 |
| 7 | Excitement,Mutism,Staring,Posturing/catalepsy,Rigidity,Negativism,Withdrawal | 99.6 | 98.9 | 100.0 |
| 7 | Excitement,Mutism,Staring,Posturing/catalepsy,Stereotypy,Rigidity,Withdrawal | 99.6 | 98.9 | 100.0 |
| 7 | Excitement,Mutism,Staring,Posturing/catalepsy,Stereotypy,Verbigeration,Negativism | 99.6 | 98.9 | 100.0 |
| 7 | Excitement,Mutism,Staring,Posturing/catalepsy,Verbigeration,Negativism,Waxy flexibility | 99.6 | 98.9 | 100.0 |
| 7 | Excitement,Mutism,Staring,Posturing/catalepsy,Verbigeration,Rigidity,Negativism | 99.6 | 98.9 | 100.0 |
| 7 | Excitement,Mutism,Staring,Posturing/catalepsy,Verbigeration,Rigidity,Withdrawal | 99.6 | 98.9 | 100.0 |
| 7 | Excitement,Mutism,Staring,Verbigeration,Rigidity,Negativism,Withdrawal | 99.6 | 98.9 | 100.0 |
| 7 | Excitement,Immobility/stupor,Mutism,Staring,Posturing/catalepsy,Echopraxia/echolalia,Rigidity | 99.5 | 98.9 | 100.0 |
| 7 | Excitement,Immobility/stupor,Mutism,Staring,Posturing/catalepsy,Negativism,Withdrawal | 99.5 | 98.9 | 100.0 |
| 7 | Excitement,Immobility/stupor,Mutism,Staring,Posturing/catalepsy,Verbigeration,Negativism | 99.5 | 98.9 | 100.0 |
| 7 | Excitement,Immobility/stupor,Staring,Posturing/catalepsy,Echopraxia/echolalia,Rigidity,Withdrawal | 99.5 | 98.9 | 100.0 |
| 7 | Excitement,Mutism,Staring,Echopraxia/echolalia,Rigidity,Negativism,Waxy flexibility | 99.5 | 98.9 | 100.0 |
| 7 | Excitement,Mutism,Staring,Posturing/catalepsy,Echopraxia/echolalia,Mannerisms,Negativism | 99.5 | 98.9 | 100.0 |
| 7 | Excitement,Mutism,Staring,Posturing/catalepsy,Echopraxia/echolalia,Stereotypy,Negativism | 99.5 | 98.9 | 100.0 |
| 7 | Excitement,Mutism,Staring,Posturing/catalepsy,Grimacing,Echopraxia/echolalia,Rigidity | 99.5 | 98.9 | 100.0 |
| 7 | Excitement,Mutism,Staring,Posturing/catalepsy,Mannerisms,Verbigeration,Rigidity | 99.5 | 98.9 | 100.0 |
| 7 | Excitement,Mutism,Staring,Posturing/catalepsy,Rigidity,Waxy flexibility,Withdrawal | 99.5 | 98.9 | 100.0 |
| 7 | Excitement,Mutism,Staring,Stereotypy,Rigidity,Negativism,Withdrawal | 99.5 | 98.9 | 100.0 |
| 7 | Excitement,Mutism,Staring,Stereotypy,Verbigeration,Rigidity,Negativism | 99.5 | 98.9 | 100.0 |
| 7 | Excitement,Mutism,Staring,Stereotypy,Verbigeration,Rigidity,Withdrawal | 99.5 | 98.9 | 100.0 |
| 7 | Excitement,Immobility/stupor,Mutism,Staring,Echopraxia/echolalia,Rigidity,Negativism | 99.3 | 98.4 | 100.0 |
| 7 | Excitement,Immobility/stupor,Mutism,Staring,Echopraxia/echolalia,Rigidity,Waxy flexibility | 99.3 | 98.4 | 100.0 |
| 7 | Excitement,Immobility/stupor,Mutism,Staring,Posturing/catalepsy,Mannerisms,Negativism | 99.3 | 98.4 | 100.0 |
| 7 | Excitement,Immobility/stupor,Mutism,Staring,Posturing/catalepsy,Mannerisms,Rigidity | 99.3 | 98.4 | 100.0 |
| 7 | Excitement,Immobility/stupor,Mutism,Staring,Posturing/catalepsy,Negativism,Waxy flexibility | 99.3 | 98.4 | 100.0 |
| 7 | Excitement,Immobility/stupor,Mutism,Staring,Posturing/catalepsy,Rigidity,Negativism | 99.3 | 98.4 | 100.0 |
| 7 | Excitement,Immobility/stupor,Mutism,Staring,Posturing/catalepsy,Rigidity,Withdrawal | 99.3 | 98.4 | 100.0 |
| 7 | Excitement,Immobility/stupor,Mutism,Staring,Posturing/catalepsy,Stereotypy,Negativism | 99.3 | 98.4 | 100.0 |
| 7 | Excitement,Immobility/stupor,Mutism,Staring,Posturing/catalepsy,Verbigeration,Rigidity | 99.3 | 98.4 | 100.0 |
| 7 | Excitement,Immobility/stupor,Mutism,Staring,Rigidity,Waxy flexibility,Withdrawal | 99.3 | 98.4 | 100.0 |
| 7 | Excitement,Immobility/stupor,Mutism,Staring,Stereotypy,Rigidity,Waxy flexibility | 99.3 | 98.4 | 100.0 |
| 7 | Excitement,Immobility/stupor,Mutism,Staring,Verbigeration,Rigidity,Negativism | 99.3 | 98.4 | 100.0 |
| 7 | Excitement,Immobility/stupor,Mutism,Staring,Verbigeration,Rigidity,Waxy flexibility | 99.3 | 98.4 | 100.0 |
| 7 | Excitement,Immobility/stupor,Staring,Echopraxia/echolalia,Mannerisms,Rigidity,Withdrawal | 99.3 | 98.4 | 100.0 |
| 7 | Excitement,Immobility/stupor,Staring,Echopraxia/echolalia,Rigidity,Waxy flexibility,Withdrawal | 99.3 | 98.4 | 100.0 |
| 7 | Excitement,Immobility/stupor,Staring,Echopraxia/echolalia,Stereotypy,Rigidity,Withdrawal | 99.3 | 98.4 | 100.0 |
| 7 | Excitement,Immobility/stupor,Staring,Mannerisms,Rigidity,Negativism,Withdrawal | 99.3 | 98.4 | 100.0 |
| 7 | Excitement,Immobility/stupor,Staring,Mannerisms,Rigidity,Waxy flexibility,Withdrawal | 99.3 | 98.4 | 100.0 |
| 7 | Excitement,Immobility/stupor,Staring,Mannerisms,Verbigeration,Rigidity,Negativism | 99.3 | 98.4 | 100.0 |
| 7 | Excitement,Immobility/stupor,Staring,Mannerisms,Verbigeration,Rigidity,Withdrawal | 99.3 | 98.4 | 100.0 |
| 7 | Excitement,Immobility/stupor,Staring,Posturing/catalepsy,Echopraxia/echolalia,Rigidity,Negativism | 99.3 | 98.4 | 100.0 |
| 7 | Excitement,Immobility/stupor,Staring,Posturing/catalepsy,Mannerisms,Negativism,Withdrawal | 99.3 | 98.4 | 100.0 |
| 7 | Excitement,Immobility/stupor,Staring,Posturing/catalepsy,Mannerisms,Rigidity,Negativism | 99.3 | 98.4 | 100.0 |
| 7 | Excitement,Immobility/stupor,Staring,Posturing/catalepsy,Mannerisms,Verbigeration,Negativism | 99.3 | 98.4 | 100.0 |
| 7 | Excitement,Immobility/stupor,Staring,Posturing/catalepsy,Mannerisms,Verbigeration,Rigidity | 99.3 | 98.4 | 100.0 |
| 7 | Excitement,Immobility/stupor,Staring,Posturing/catalepsy,Rigidity,Negativism,Withdrawal | 99.3 | 98.4 | 100.0 |
| 7 | Excitement,Immobility/stupor,Staring,Posturing/catalepsy,Verbigeration,Negativism,Withdrawal | 99.3 | 98.4 | 100.0 |
| 7 | Excitement,Immobility/stupor,Staring,Posturing/catalepsy,Verbigeration,Rigidity,Negativism | 99.3 | 98.4 | 100.0 |
| 7 | Excitement,Immobility/stupor,Staring,Stereotypy,Rigidity,Negativism,Withdrawal | 99.3 | 98.4 | 100.0 |
| 7 | Excitement,Immobility/stupor,Staring,Stereotypy,Rigidity,Waxy flexibility,Withdrawal | 99.3 | 98.4 | 100.0 |
| 7 | Excitement,Immobility/stupor,Staring,Stereotypy,Verbigeration,Rigidity,Negativism | 99.3 | 98.4 | 100.0 |
| 7 | Excitement,Immobility/stupor,Staring,Verbigeration,Rigidity,Negativism,Withdrawal | 99.3 | 98.4 | 100.0 |
| 7 | Excitement,Mutism,Staring,Echopraxia/echolalia,Mannerisms,Rigidity,Negativism | 99.3 | 98.4 | 100.0 |
| 7 | Excitement,Mutism,Staring,Echopraxia/echolalia,Mannerisms,Rigidity,Withdrawal | 99.3 | 98.4 | 100.0 |
| 7 | Excitement,Mutism,Staring,Echopraxia/echolalia,Negativism,Waxy flexibility,Withdrawal | 99.3 | 98.4 | 100.0 |
| 7 | Excitement,Mutism,Staring,Echopraxia/echolalia,Stereotypy,Negativism,Waxy flexibility | 99.3 | 98.4 | 100.0 |
| 7 | Excitement,Mutism,Staring,Echopraxia/echolalia,Stereotypy,Rigidity,Waxy flexibility | 99.3 | 98.4 | 100.0 |
| 7 | Excitement,Mutism,Staring,Echopraxia/echolalia,Verbigeration,Negativism,Waxy flexibility | 99.3 | 98.4 | 100.0 |
| 7 | Excitement,Mutism,Staring,Echopraxia/echolalia,Verbigeration,Rigidity,Withdrawal | 99.3 | 98.4 | 100.0 |
| 7 | Excitement,Mutism,Staring,Grimacing,Echopraxia/echolalia,Rigidity,Negativism | 99.3 | 98.4 | 100.0 |
| 7 | Excitement,Mutism,Staring,Grimacing,Echopraxia/echolalia,Rigidity,Waxy flexibility | 99.3 | 98.4 | 100.0 |
| 7 | Excitement,Mutism,Staring,Grimacing,Rigidity,Waxy flexibility,Withdrawal | 99.3 | 98.4 | 100.0 |
| 7 | Excitement,Mutism,Staring,Grimacing,Stereotypy,Rigidity,Withdrawal | 99.3 | 98.4 | 100.0 |
| 7 | Excitement,Mutism,Staring,Grimacing,Verbigeration,Rigidity,Withdrawal | 99.3 | 98.4 | 100.0 |
| 7 | Excitement,Mutism,Staring,Mannerisms,Rigidity,Negativism,Withdrawal | 99.3 | 98.4 | 100.0 |
| 7 | Excitement,Mutism,Staring,Mannerisms,Rigidity,Waxy flexibility,Withdrawal | 99.3 | 98.4 | 100.0 |
| 7 | Excitement,Mutism,Staring,Mannerisms,Verbigeration,Rigidity,Negativism | 99.3 | 98.4 | 100.0 |
| 7 | Excitement,Mutism,Staring,Mannerisms,Verbigeration,Rigidity,Withdrawal | 99.3 | 98.4 | 100.0 |
| 7 | Excitement,Mutism,Staring,Posturing/catalepsy,Echopraxia/echolalia,Waxy flexibility,Withdrawal | 99.3 | 98.4 | 100.0 |
| 7 | Excitement,Mutism,Staring,Posturing/catalepsy,Grimacing,Echopraxia/echolalia,Withdrawal | 99.3 | 98.4 | 100.0 |
| 7 | Excitement,Mutism,Staring,Posturing/catalepsy,Grimacing,Verbigeration,Negativism | 99.3 | 98.4 | 100.0 |
| 7 | Excitement,Mutism,Staring,Posturing/catalepsy,Grimacing,Verbigeration,Rigidity | 99.3 | 98.4 | 100.0 |
| 7 | Excitement,Mutism,Staring,Posturing/catalepsy,Mannerisms,Negativism,Waxy flexibility | 99.3 | 98.4 | 100.0 |
| 7 | Excitement,Mutism,Staring,Posturing/catalepsy,Mannerisms,Rigidity,Negativism | 99.3 | 98.4 | 100.0 |
| 7 | Excitement,Mutism,Staring,Posturing/catalepsy,Stereotypy,Negativism,Waxy flexibility | 99.3 | 98.4 | 100.0 |
| 7 | Excitement,Mutism,Staring,Posturing/catalepsy,Stereotypy,Rigidity,Negativism | 99.3 | 98.4 | 100.0 |
| 7 | Excitement,Mutism,Staring,Posturing/catalepsy,Stereotypy,Verbigeration,Rigidity | 99.3 | 98.4 | 100.0 |
| 7 | Excitement,Mutism,Staring,Posturing/catalepsy,Verbigeration,Rigidity,Waxy flexibility | 99.3 | 98.4 | 100.0 |
| 7 | Excitement,Mutism,Staring,Rigidity,Negativism,Waxy flexibility,Withdrawal | 99.3 | 98.4 | 100.0 |
| 7 | Excitement,Mutism,Staring,Stereotypy,Mannerisms,Rigidity,Withdrawal | 99.3 | 98.4 | 100.0 |
| 7 | Excitement,Mutism,Staring,Stereotypy,Rigidity,Negativism,Waxy flexibility | 99.3 | 98.4 | 100.0 |
| 7 | Excitement,Mutism,Staring,Stereotypy,Verbigeration,Negativism,Withdrawal | 99.3 | 98.4 | 100.0 |
| 7 | Excitement,Mutism,Staring,Stereotypy,Verbigeration,Rigidity,Waxy flexibility | 99.3 | 98.4 | 100.0 |
| 7 | Excitement,Mutism,Staring,Verbigeration,Rigidity,Negativism,Waxy flexibility | 99.3 | 98.4 | 100.0 |
| 7 | Excitement,Mutism,Staring,Verbigeration,Rigidity,Waxy flexibility,Withdrawal | 99.3 | 98.4 | 100.0 |
| 7 | Excitement,Staring,Posturing/catalepsy,Mannerisms,Rigidity,Negativism,Withdrawal | 99.3 | 98.4 | 100.0 |
| 7 | Immobility/stupor,Staring,Echopraxia/echolalia,Mannerisms,Rigidity,Negativism,Withdrawal | 99.3 | 98.4 | 100.0 |
| 7 | Immobility/stupor,Staring,Stereotypy,Mannerisms,Rigidity,Negativism,Withdrawal | 99.3 | 98.4 | 100.0 |
| 7 | Mutism,Staring,Echopraxia/echolalia,Mannerisms,Rigidity,Negativism,Withdrawal | 99.3 | 98.4 | 100.0 |
| 7 | Mutism,Staring,Posturing/catalepsy,Echopraxia/echolalia,Mannerisms,Negativism,Withdrawal | 99.3 | 98.4 | 100.0 |
| 7 | Mutism,Staring,Posturing/catalepsy,Grimacing,Mannerisms,Negativism,Withdrawal | 99.3 | 98.4 | 100.0 |
| 7 | Mutism,Staring,Posturing/catalepsy,Stereotypy,Mannerisms,Negativism,Withdrawal | 99.3 | 98.4 | 100.0 |
| 7 | Mutism,Staring,Stereotypy,Mannerisms,Rigidity,Negativism,Withdrawal | 99.3 | 98.4 | 100.0 |
| 7 | Staring,Echopraxia/echolalia,Stereotypy,Mannerisms,Rigidity,Negativism,Withdrawal | 99.3 | 98.4 | 100.0 |
| 7 | Staring,Stereotypy,Mannerisms,Verbigeration,Rigidity,Negativism,Withdrawal | 99.3 | 98.4 | 100.0 |
| 7 | Excitement,Immobility/stupor,Mutism,Posturing/catalepsy,Verbigeration,Rigidity,Negativism | 99.1 | 98.2 | 99.8 |
| 7 | Excitement,Immobility/stupor,Mutism,Staring,Echopraxia/echolalia,Negativism,Waxy flexibility | 99.1 | 98.2 | 99.8 |
| 7 | Excitement,Immobility/stupor,Mutism,Staring,Echopraxia/echolalia,Negativism,Withdrawal | 99.1 | 98.2 | 99.8 |
| 7 | Excitement,Immobility/stupor,Mutism,Staring,Echopraxia/echolalia,Stereotypy,Rigidity | 99.1 | 98.2 | 99.8 |
| 7 | Excitement,Immobility/stupor,Mutism,Staring,Echopraxia/echolalia,Verbigeration,Rigidity | 99.1 | 98.2 | 99.8 |
| 7 | Excitement,Immobility/stupor,Mutism,Staring,Mannerisms,Rigidity,Waxy flexibility | 99.1 | 98.2 | 99.8 |
| 7 | Excitement,Immobility/stupor,Mutism,Staring,Mannerisms,Rigidity,Withdrawal | 99.1 | 98.2 | 99.8 |
| 7 | Excitement,Immobility/stupor,Mutism,Staring,Mannerisms,Verbigeration,Rigidity | 99.1 | 98.2 | 99.8 |
| 7 | Excitement,Immobility/stupor,Mutism,Staring,Posturing/catalepsy,Echopraxia/echolalia,Withdrawal | 99.1 | 98.2 | 99.8 |
| 7 | Excitement,Immobility/stupor,Mutism,Staring,Posturing/catalepsy,Grimacing,Negativism | 99.1 | 98.2 | 99.8 |
| 7 | Excitement,Immobility/stupor,Mutism,Staring,Posturing/catalepsy,Stereotypy,Rigidity | 99.1 | 98.2 | 99.8 |
| 7 | Excitement,Immobility/stupor,Mutism,Staring,Rigidity,Negativism,Waxy flexibility | 99.1 | 98.2 | 99.8 |
| 7 | Excitement,Immobility/stupor,Mutism,Staring,Rigidity,Negativism,Withdrawal | 99.1 | 98.2 | 99.8 |
| 7 | Excitement,Immobility/stupor,Mutism,Staring,Stereotypy,Rigidity,Negativism | 99.1 | 98.2 | 99.8 |
| 7 | Excitement,Immobility/stupor,Mutism,Staring,Stereotypy,Rigidity,Withdrawal | 99.1 | 98.2 | 99.8 |
| 7 | Excitement,Immobility/stupor,Mutism,Staring,Stereotypy,Verbigeration,Rigidity | 99.1 | 98.2 | 99.8 |
| 7 | Excitement,Immobility/stupor,Mutism,Staring,Verbigeration,Rigidity,Withdrawal | 99.1 | 98.2 | 99.8 |
| 7 | Excitement,Immobility/stupor,Mutism,Stereotypy,Verbigeration,Rigidity,Negativism | 99.1 | 98.2 | 99.8 |
| 7 | Excitement,Immobility/stupor,Staring,Echopraxia/echolalia,Mannerisms,Rigidity,Negativism | 99.1 | 98.2 | 99.8 |
| 7 | Excitement,Immobility/stupor,Staring,Echopraxia/echolalia,Rigidity,Negativism,Waxy flexibility | 99.1 | 98.2 | 99.8 |
| 7 | Excitement,Immobility/stupor,Staring,Echopraxia/echolalia,Stereotypy,Negativism,Withdrawal | 99.1 | 98.2 | 99.8 |
| 7 | Excitement,Immobility/stupor,Staring,Echopraxia/echolalia,Stereotypy,Rigidity,Negativism | 99.1 | 98.2 | 99.8 |
| 7 | Excitement,Immobility/stupor,Staring,Echopraxia/echolalia,Verbigeration,Rigidity,Negativism | 99.1 | 98.2 | 99.8 |
| 7 | Excitement,Immobility/stupor,Staring,Echopraxia/echolalia,Verbigeration,Rigidity,Withdrawal | 99.1 | 98.2 | 99.8 |
| 7 | Excitement,Immobility/stupor,Staring,Grimacing,Echopraxia/echolalia,Rigidity,Withdrawal | 99.1 | 98.2 | 99.8 |
| 7 | Excitement,Immobility/stupor,Staring,Mannerisms,Rigidity,Negativism,Waxy flexibility | 99.1 | 98.2 | 99.8 |
| 7 | Excitement,Immobility/stupor,Staring,Mannerisms,Verbigeration,Rigidity,Waxy flexibility | 99.1 | 98.2 | 99.8 |
| 7 | Excitement,Immobility/stupor,Staring,Posturing/catalepsy,Echopraxia/echolalia,Mannerisms,Negativism | 99.1 | 98.2 | 99.8 |
| 7 | Excitement,Immobility/stupor,Staring,Posturing/catalepsy,Echopraxia/echolalia,Negativism,Waxy flexibility | 99.1 | 98.2 | 99.8 |
| 7 | Excitement,Immobility/stupor,Staring,Posturing/catalepsy,Echopraxia/echolalia,Verbigeration,Negativism | 99.1 | 98.2 | 99.8 |
| 7 | Excitement,Immobility/stupor,Staring,Posturing/catalepsy,Echopraxia/echolalia,Verbigeration,Rigidity | 99.1 | 98.2 | 99.8 |
| 7 | Excitement,Immobility/stupor,Staring,Posturing/catalepsy,Grimacing,Rigidity,Withdrawal | 99.1 | 98.2 | 99.8 |
| 7 | Excitement,Immobility/stupor,Staring,Posturing/catalepsy,Rigidity,Waxy flexibility,Withdrawal | 99.1 | 98.2 | 99.8 |
| 7 | Excitement,Immobility/stupor,Staring,Posturing/catalepsy,Stereotypy,Negativism,Withdrawal | 99.1 | 98.2 | 99.8 |
| 7 | Excitement,Immobility/stupor,Staring,Posturing/catalepsy,Stereotypy,Rigidity,Negativism | 99.1 | 98.2 | 99.8 |
| 7 | Excitement,Immobility/stupor,Staring,Posturing/catalepsy,Stereotypy,Rigidity,Withdrawal | 99.1 | 98.2 | 99.8 |
| 7 | Excitement,Immobility/stupor,Staring,Posturing/catalepsy,Stereotypy,Verbigeration,Negativism | 99.1 | 98.2 | 99.8 |
| 7 | Excitement,Immobility/stupor,Staring,Posturing/catalepsy,Verbigeration,Negativism,Waxy flexibility | 99.1 | 98.2 | 99.8 |
| 7 | Excitement,Immobility/stupor,Staring,Posturing/catalepsy,Verbigeration,Rigidity,Withdrawal | 99.1 | 98.2 | 99.8 |
| 7 | Excitement,Immobility/stupor,Staring,Rigidity,Negativism,Waxy flexibility,Withdrawal | 99.1 | 98.2 | 99.8 |
| 7 | Excitement,Immobility/stupor,Staring,Stereotypy,Mannerisms,Rigidity,Withdrawal | 99.1 | 98.2 | 99.8 |
| 7 | Excitement,Immobility/stupor,Staring,Stereotypy,Rigidity,Negativism,Waxy flexibility | 99.1 | 98.2 | 99.8 |
| 7 | Excitement,Immobility/stupor,Staring,Stereotypy,Verbigeration,Rigidity,Waxy flexibility | 99.1 | 98.2 | 99.8 |
| 7 | Excitement,Immobility/stupor,Staring,Stereotypy,Verbigeration,Rigidity,Withdrawal | 99.1 | 98.2 | 99.8 |
| 7 | Excitement,Immobility/stupor,Staring,Verbigeration,Rigidity,Negativism,Waxy flexibility | 99.1 | 98.2 | 99.8 |
| 7 | Excitement,Immobility/stupor,Staring,Verbigeration,Rigidity,Waxy flexibility,Withdrawal | 99.1 | 98.2 | 99.8 |
| 7 | Excitement,Mutism,Staring,Echopraxia/echolalia,Mannerisms,Negativism,Waxy flexibility | 99.1 | 98.2 | 99.8 |
| 7 | Excitement,Mutism,Staring,Echopraxia/echolalia,Mannerisms,Negativism,Withdrawal | 99.1 | 98.2 | 99.8 |
| 7 | Excitement,Mutism,Staring,Echopraxia/echolalia,Stereotypy,Verbigeration,Negativism | 99.1 | 98.2 | 99.8 |
| 7 | Excitement,Mutism,Staring,Echopraxia/echolalia,Stereotypy,Verbigeration,Rigidity | 99.1 | 98.2 | 99.8 |
| 7 | Excitement,Mutism,Staring,Echopraxia/echolalia,Verbigeration,Negativism,Withdrawal | 99.1 | 98.2 | 99.8 |
| 7 | Excitement,Mutism,Staring,Echopraxia/echolalia,Verbigeration,Rigidity,Waxy flexibility | 99.1 | 98.2 | 99.8 |
| 7 | Excitement,Mutism,Staring,Grimacing,Echopraxia/echolalia,Negativism,Waxy flexibility | 99.1 | 98.2 | 99.8 |
| 7 | Excitement,Mutism,Staring,Grimacing,Echopraxia/echolalia,Negativism,Withdrawal | 99.1 | 98.2 | 99.8 |
| 7 | Excitement,Mutism,Staring,Grimacing,Echopraxia/echolalia,Stereotypy,Rigidity | 99.1 | 98.2 | 99.8 |
| 7 | Excitement,Mutism,Staring,Grimacing,Echopraxia/echolalia,Verbigeration,Rigidity | 99.1 | 98.2 | 99.8 |
| 7 | Excitement,Mutism,Staring,Grimacing,Mannerisms,Rigidity,Withdrawal | 99.1 | 98.2 | 99.8 |
| 7 | Excitement,Mutism,Staring,Grimacing,Rigidity,Negativism,Withdrawal | 99.1 | 98.2 | 99.8 |
| 7 | Excitement,Mutism,Staring,Grimacing,Stereotypy,Rigidity,Waxy flexibility | 99.1 | 98.2 | 99.8 |
| 7 | Excitement,Mutism,Staring,Grimacing,Stereotypy,Verbigeration,Rigidity | 99.1 | 98.2 | 99.8 |
| 7 | Excitement,Mutism,Staring,Grimacing,Verbigeration,Rigidity,Negativism | 99.1 | 98.2 | 99.8 |
| 7 | Excitement,Mutism,Staring,Grimacing,Verbigeration,Rigidity,Waxy flexibility | 99.1 | 98.2 | 99.8 |
| 7 | Excitement,Mutism,Staring,Mannerisms,Rigidity,Negativism,Waxy flexibility | 99.1 | 98.2 | 99.8 |
| 7 | Excitement,Mutism,Staring,Posturing/catalepsy,Echopraxia/echolalia,Mannerisms,Withdrawal | 99.1 | 98.2 | 99.8 |
| 7 | Excitement,Mutism,Staring,Posturing/catalepsy,Echopraxia/echolalia,Stereotypy,Withdrawal | 99.1 | 98.2 | 99.8 |
| 7 | Excitement,Mutism,Staring,Posturing/catalepsy,Echopraxia/echolalia,Verbigeration,Withdrawal | 99.1 | 98.2 | 99.8 |
| 7 | Excitement,Mutism,Staring,Posturing/catalepsy,Grimacing,Mannerisms,Negativism | 99.1 | 98.2 | 99.8 |
| 7 | Excitement,Mutism,Staring,Posturing/catalepsy,Grimacing,Mannerisms,Rigidity | 99.1 | 98.2 | 99.8 |
| 7 | Excitement,Mutism,Staring,Posturing/catalepsy,Grimacing,Negativism,Waxy flexibility | 99.1 | 98.2 | 99.8 |
| 7 | Excitement,Mutism,Staring,Posturing/catalepsy,Grimacing,Rigidity,Negativism | 99.1 | 98.2 | 99.8 |
| 7 | Excitement,Mutism,Staring,Posturing/catalepsy,Grimacing,Stereotypy,Negativism | 99.1 | 98.2 | 99.8 |
| 7 | Excitement,Mutism,Staring,Posturing/catalepsy,Grimacing,Stereotypy,Rigidity | 99.1 | 98.2 | 99.8 |
| 7 | Excitement,Mutism,Staring,Posturing/catalepsy,Mannerisms,Rigidity,Waxy flexibility | 99.1 | 98.2 | 99.8 |
| 7 | Excitement,Mutism,Staring,Posturing/catalepsy,Rigidity,Negativism,Waxy flexibility | 99.1 | 98.2 | 99.8 |
| 7 | Excitement,Mutism,Staring,Posturing/catalepsy,Stereotypy,Mannerisms,Negativism | 99.1 | 98.2 | 99.8 |
| 7 | Excitement,Mutism,Staring,Posturing/catalepsy,Stereotypy,Mannerisms,Rigidity | 99.1 | 98.2 | 99.8 |
| 7 | Excitement,Mutism,Staring,Posturing/catalepsy,Stereotypy,Rigidity,Waxy flexibility | 99.1 | 98.2 | 99.8 |
| 7 | Excitement,Mutism,Staring,Stereotypy,Negativism,Waxy flexibility,Withdrawal | 99.1 | 98.2 | 99.8 |
| 7 | Excitement,Mutism,Staring,Stereotypy,Verbigeration,Negativism,Waxy flexibility | 99.1 | 98.2 | 99.8 |
| 7 | Excitement,Staring,Echopraxia/echolalia,Mannerisms,Rigidity,Negativism,Withdrawal | 99.1 | 98.2 | 99.8 |
| 7 | Excitement,Staring,Echopraxia/echolalia,Stereotypy,Rigidity,Negativism,Withdrawal | 99.1 | 98.2 | 99.8 |
| 7 | Excitement,Staring,Mannerisms,Rigidity,Negativism,Waxy flexibility,Withdrawal | 99.1 | 98.2 | 99.8 |
| 7 | Excitement,Staring,Mannerisms,Verbigeration,Rigidity,Negativism,Withdrawal | 99.1 | 98.2 | 99.8 |
| 7 | Excitement,Staring,Posturing/catalepsy,Echopraxia/echolalia,Mannerisms,Rigidity,Withdrawal | 99.1 | 98.2 | 99.8 |
| 7 | Excitement,Staring,Posturing/catalepsy,Echopraxia/echolalia,Rigidity,Negativism,Withdrawal | 99.1 | 98.2 | 99.8 |
| 7 | Excitement,Staring,Posturing/catalepsy,Grimacing,Mannerisms,Rigidity,Withdrawal | 99.1 | 98.2 | 99.8 |
| 7 | Excitement,Staring,Posturing/catalepsy,Mannerisms,Verbigeration,Rigidity,Withdrawal | 99.1 | 98.2 | 99.8 |
| 7 | Excitement,Staring,Stereotypy,Mannerisms,Rigidity,Negativism,Withdrawal | 99.1 | 98.2 | 99.8 |
| 7 | Excitement,Staring,Stereotypy,Verbigeration,Rigidity,Negativism,Withdrawal | 99.1 | 98.2 | 99.8 |
| 7 | Immobility/stupor,Staring,Mannerisms,Verbigeration,Rigidity,Negativism,Withdrawal | 99.1 | 98.2 | 99.8 |
| 7 | Immobility/stupor,Staring,Posturing/catalepsy,Mannerisms,Rigidity,Negativism,Withdrawal | 99.1 | 98.2 | 99.8 |
| 7 | Immobility/stupor,Staring,Stereotypy,Mannerisms,Verbigeration,Rigidity,Negativism | 99.1 | 98.2 | 99.8 |
| 7 | Mutism,Staring,Echopraxia/echolalia,Stereotypy,Mannerisms,Negativism,Withdrawal | 99.1 | 98.2 | 99.8 |
| 7 | Mutism,Staring,Echopraxia/echolalia,Stereotypy,Mannerisms,Rigidity,Negativism | 99.1 | 98.2 | 99.8 |
| 7 | Mutism,Staring,Mannerisms,Verbigeration,Rigidity,Negativism,Withdrawal | 99.1 | 98.2 | 99.8 |
| 7 | Mutism,Staring,Posturing/catalepsy,Echopraxia/echolalia,Mannerisms,Rigidity,Negativism | 99.1 | 98.2 | 99.8 |
| 7 | Mutism,Staring,Posturing/catalepsy,Mannerisms,Rigidity,Negativism,Withdrawal | 99.1 | 98.2 | 99.8 |
| 7 | Mutism,Staring,Posturing/catalepsy,Mannerisms,Verbigeration,Negativism,Withdrawal | 99.1 | 98.2 | 99.8 |
| 7 | Mutism,Staring,Stereotypy,Mannerisms,Verbigeration,Rigidity,Negativism | 99.1 | 98.2 | 99.8 |
| 7 | Mutism,Staring,Stereotypy,Verbigeration,Rigidity,Negativism,Withdrawal | 99.1 | 98.2 | 99.8 |
| 7 | Staring,Grimacing,Stereotypy,Mannerisms,Rigidity,Negativism,Withdrawal | 99.1 | 98.2 | 99.8 |
| 7 | Staring,Posturing/catalepsy,Echopraxia/echolalia,Mannerisms,Rigidity,Negativism,Withdrawal | 99.1 | 98.2 | 99.8 |
| 7 | Staring,Posturing/catalepsy,Stereotypy,Mannerisms,Rigidity,Negativism,Withdrawal | 99.1 | 98.2 | 99.8 |
| 7 | Staring,Stereotypy,Mannerisms,Rigidity,Negativism,Waxy flexibility,Withdrawal | 99.1 | 98.2 | 99.8 |
| 7 | Excitement,Immobility/stupor,Echopraxia/echolalia,Stereotypy,Rigidity,Negativism,Withdrawal | 98.9 | 97.8 | 99.8 |
| 7 | Excitement,Immobility/stupor,Echopraxia/echolalia,Verbigeration,Rigidity,Negativism,Withdrawal | 98.9 | 97.8 | 99.8 |
| 7 | Excitement,Immobility/stupor,Grimacing,Echopraxia/echolalia,Rigidity,Negativism,Withdrawal | 98.9 | 97.8 | 99.8 |
| 7 | Excitement,Immobility/stupor,Mutism,Echopraxia/echolalia,Rigidity,Negativism,Withdrawal | 98.9 | 97.8 | 99.8 |
| 7 | Excitement,Immobility/stupor,Mutism,Echopraxia/echolalia,Verbigeration,Rigidity,Negativism | 98.9 | 97.8 | 99.8 |
| 7 | Excitement,Immobility/stupor,Mutism,Posturing/catalepsy,Echopraxia/echolalia,Rigidity,Negativism | 98.9 | 97.8 | 99.8 |
| 7 | Excitement,Immobility/stupor,Mutism,Posturing/catalepsy,Echopraxia/echolalia,Rigidity,Withdrawal | 98.9 | 97.8 | 99.8 |
| 7 | Excitement,Immobility/stupor,Mutism,Posturing/catalepsy,Echopraxia/echolalia,Verbigeration,Rigidity | 98.9 | 97.8 | 99.8 |
| 7 | Excitement,Immobility/stupor,Mutism,Staring,Echopraxia/echolalia,Mannerisms,Rigidity | 98.9 | 97.8 | 99.8 |
| 7 | Excitement,Immobility/stupor,Mutism,Staring,Echopraxia/echolalia,Stereotypy,Negativism | 98.9 | 97.8 | 99.8 |
| 7 | Excitement,Immobility/stupor,Mutism,Staring,Grimacing,Echopraxia/echolalia,Rigidity | 98.9 | 97.8 | 99.8 |
| 7 | Excitement,Immobility/stupor,Mutism,Staring,Grimacing,Rigidity,Withdrawal | 98.9 | 97.8 | 99.8 |
| 7 | Excitement,Immobility/stupor,Mutism,Staring,Grimacing,Verbigeration,Rigidity | 98.9 | 97.8 | 99.8 |
| 7 | Excitement,Immobility/stupor,Mutism,Staring,Mannerisms,Rigidity,Negativism | 98.9 | 97.8 | 99.8 |
| 7 | Excitement,Immobility/stupor,Mutism,Staring,Posturing/catalepsy,Echopraxia/echolalia,Waxy flexibility | 98.9 | 97.8 | 99.8 |
| 7 | Excitement,Immobility/stupor,Mutism,Staring,Posturing/catalepsy,Grimacing,Rigidity | 98.9 | 97.8 | 99.8 |
| 7 | Excitement,Immobility/stupor,Mutism,Staring,Posturing/catalepsy,Rigidity,Waxy flexibility | 98.9 | 97.8 | 99.8 |
| 7 | Excitement,Immobility/stupor,Mutism,Staring,Stereotypy,Negativism,Waxy flexibility | 98.9 | 97.8 | 99.8 |
| 7 | Excitement,Immobility/stupor,Mutism,Staring,Stereotypy,Negativism,Withdrawal | 98.9 | 97.8 | 99.8 |
| 7 | Excitement,Immobility/stupor,Mutism,Staring,Stereotypy,Verbigeration,Negativism | 98.9 | 97.8 | 99.8 |
| 7 | Excitement,Immobility/stupor,Mutism,Staring,Verbigeration,Negativism,Waxy flexibility | 98.9 | 97.8 | 99.8 |
| 7 | Excitement,Immobility/stupor,Mutism,Verbigeration,Rigidity,Negativism,Withdrawal | 98.9 | 97.8 | 99.8 |
| 7 | Excitement,Immobility/stupor,Posturing/catalepsy,Echopraxia/echolalia,Rigidity,Negativism,Withdrawal | 98.9 | 97.8 | 99.8 |
| 7 | Excitement,Immobility/stupor,Posturing/catalepsy,Verbigeration,Rigidity,Negativism,Withdrawal | 98.9 | 97.8 | 99.8 |
| 7 | Excitement,Immobility/stupor,Staring,Echopraxia/echolalia,Mannerisms,Negativism,Withdrawal | 98.9 | 97.8 | 99.8 |
| 7 | Excitement,Immobility/stupor,Staring,Echopraxia/echolalia,Negativism,Waxy flexibility,Withdrawal | 98.9 | 97.8 | 99.8 |
| 7 | Excitement,Immobility/stupor,Staring,Echopraxia/echolalia,Verbigeration,Rigidity,Waxy flexibility | 98.9 | 97.8 | 99.8 |
| 7 | Excitement,Immobility/stupor,Staring,Grimacing,Echopraxia/echolalia,Rigidity,Negativism | 98.9 | 97.8 | 99.8 |
| 7 | Excitement,Immobility/stupor,Staring,Grimacing,Mannerisms,Rigidity,Withdrawal | 98.9 | 97.8 | 99.8 |
| 7 | Excitement,Immobility/stupor,Staring,Grimacing,Rigidity,Negativism,Withdrawal | 98.9 | 97.8 | 99.8 |
| 7 | Excitement,Immobility/stupor,Staring,Grimacing,Rigidity,Waxy flexibility,Withdrawal | 98.9 | 97.8 | 99.8 |
| 7 | Excitement,Immobility/stupor,Staring,Grimacing,Stereotypy,Rigidity,Withdrawal | 98.9 | 97.8 | 99.8 |
| 7 | Excitement,Immobility/stupor,Staring,Grimacing,Verbigeration,Rigidity,Negativism | 98.9 | 97.8 | 99.8 |
| 7 | Excitement,Immobility/stupor,Staring,Grimacing,Verbigeration,Rigidity,Withdrawal | 98.9 | 97.8 | 99.8 |
| 7 | Excitement,Immobility/stupor,Staring,Posturing/catalepsy,Echopraxia/echolalia,Mannerisms,Rigidity | 98.9 | 97.8 | 99.8 |
| 7 | Excitement,Immobility/stupor,Staring,Posturing/catalepsy,Echopraxia/echolalia,Stereotypy,Negativism | 98.9 | 97.8 | 99.8 |
| 7 | Excitement,Immobility/stupor,Staring,Posturing/catalepsy,Grimacing,Echopraxia/echolalia,Negativism | 98.9 | 97.8 | 99.8 |
| 7 | Excitement,Immobility/stupor,Staring,Posturing/catalepsy,Grimacing,Echopraxia/echolalia,Rigidity | 98.9 | 97.8 | 99.8 |
| 7 | Excitement,Immobility/stupor,Staring,Posturing/catalepsy,Grimacing,Mannerisms,Rigidity | 98.9 | 97.8 | 99.8 |
| 7 | Excitement,Immobility/stupor,Staring,Posturing/catalepsy,Grimacing,Negativism,Withdrawal | 98.9 | 97.8 | 99.8 |
| 7 | Excitement,Immobility/stupor,Staring,Posturing/catalepsy,Grimacing,Rigidity,Negativism | 98.9 | 97.8 | 99.8 |
| 7 | Excitement,Immobility/stupor,Staring,Posturing/catalepsy,Grimacing,Verbigeration,Negativism | 98.9 | 97.8 | 99.8 |
| 7 | Excitement,Immobility/stupor,Staring,Posturing/catalepsy,Grimacing,Verbigeration,Rigidity | 98.9 | 97.8 | 99.8 |
| 7 | Excitement,Immobility/stupor,Staring,Posturing/catalepsy,Mannerisms,Negativism,Waxy flexibility | 98.9 | 97.8 | 99.8 |
| 7 | Excitement,Immobility/stupor,Staring,Posturing/catalepsy,Negativism,Waxy flexibility,Withdrawal | 98.9 | 97.8 | 99.8 |
| 7 | Excitement,Immobility/stupor,Staring,Posturing/catalepsy,Rigidity,Negativism,Waxy flexibility | 98.9 | 97.8 | 99.8 |
| 7 | Excitement,Immobility/stupor,Staring,Posturing/catalepsy,Stereotypy,Verbigeration,Rigidity | 98.9 | 97.8 | 99.8 |
| 7 | Excitement,Immobility/stupor,Staring,Posturing/catalepsy,Verbigeration,Rigidity,Waxy flexibility | 98.9 | 97.8 | 99.8 |
| 7 | Excitement,Immobility/stupor,Staring,Stereotypy,Mannerisms,Rigidity,Negativism | 98.9 | 97.8 | 99.8 |
| 7 | Excitement,Immobility/stupor,Staring,Stereotypy,Mannerisms,Verbigeration,Rigidity | 98.9 | 97.8 | 99.8 |
| 7 | Excitement,Immobility/stupor,Staring,Stereotypy,Verbigeration,Negativism,Withdrawal | 98.9 | 97.8 | 99.8 |
| 7 | Excitement,Immobility/stupor,Stereotypy,Verbigeration,Rigidity,Negativism,Withdrawal | 98.9 | 97.8 | 99.8 |
| 7 | Excitement,Mutism,Staring,Echopraxia/echolalia,Mannerisms,Rigidity,Waxy flexibility | 98.9 | 97.8 | 99.8 |
| 7 | Excitement,Mutism,Staring,Echopraxia/echolalia,Stereotypy,Mannerisms,Negativism | 98.9 | 97.8 | 99.8 |
| 7 | Excitement,Mutism,Staring,Echopraxia/echolalia,Stereotypy,Mannerisms,Rigidity | 98.9 | 97.8 | 99.8 |
| 7 | Excitement,Mutism,Staring,Echopraxia/echolalia,Stereotypy,Waxy flexibility,Withdrawal | 98.9 | 97.8 | 99.8 |
| 7 | Excitement,Mutism,Staring,Grimacing,Echopraxia/echolalia,Mannerisms,Rigidity | 98.9 | 97.8 | 99.8 |
| 7 | Excitement,Mutism,Staring,Grimacing,Echopraxia/echolalia,Stereotypy,Negativism | 98.9 | 97.8 | 99.8 |
| 7 | Excitement,Mutism,Staring,Grimacing,Echopraxia/echolalia,Waxy flexibility,Withdrawal | 98.9 | 97.8 | 99.8 |
| 7 | Excitement,Mutism,Staring,Grimacing,Mannerisms,Rigidity,Waxy flexibility | 98.9 | 97.8 | 99.8 |
| 7 | Excitement,Mutism,Staring,Grimacing,Mannerisms,Verbigeration,Rigidity | 98.9 | 97.8 | 99.8 |
| 7 | Excitement,Mutism,Staring,Grimacing,Rigidity,Negativism,Waxy flexibility | 98.9 | 97.8 | 99.8 |
| 7 | Excitement,Mutism,Staring,Grimacing,Stereotypy,Negativism,Withdrawal | 98.9 | 97.8 | 99.8 |
| 7 | Excitement,Mutism,Staring,Grimacing,Stereotypy,Rigidity,Negativism | 98.9 | 97.8 | 99.8 |
| 7 | Excitement,Mutism,Staring,Mannerisms,Negativism,Waxy flexibility,Withdrawal | 98.9 | 97.8 | 99.8 |
| 7 | Excitement,Mutism,Staring,Mannerisms,Verbigeration,Negativism,Waxy flexibility | 98.9 | 97.8 | 99.8 |
| 7 | Excitement,Mutism,Staring,Mannerisms,Verbigeration,Negativism,Withdrawal | 98.9 | 97.8 | 99.8 |
| 7 | Excitement,Mutism,Staring,Mannerisms,Verbigeration,Rigidity,Waxy flexibility | 98.9 | 97.8 | 99.8 |
| 7 | Excitement,Mutism,Staring,Posturing/catalepsy,Echopraxia/echolalia,Stereotypy,Waxy flexibility | 98.9 | 97.8 | 99.8 |
| 7 | Excitement,Mutism,Staring,Posturing/catalepsy,Echopraxia/echolalia,Verbigeration,Waxy flexibility | 98.9 | 97.8 | 99.8 |
| 7 | Excitement,Mutism,Staring,Posturing/catalepsy,Grimacing,Echopraxia/echolalia,Waxy flexibility | 98.9 | 97.8 | 99.8 |
| 7 | Excitement,Mutism,Staring,Posturing/catalepsy,Grimacing,Mannerisms,Withdrawal | 98.9 | 97.8 | 99.8 |
| 7 | Excitement,Mutism,Staring,Posturing/catalepsy,Grimacing,Stereotypy,Withdrawal | 98.9 | 97.8 | 99.8 |
| 7 | Excitement,Mutism,Staring,Posturing/catalepsy,Grimacing,Verbigeration,Withdrawal | 98.9 | 97.8 | 99.8 |
| 7 | Excitement,Mutism,Staring,Posturing/catalepsy,Grimacing,Waxy flexibility,Withdrawal | 98.9 | 97.8 | 99.8 |
| 7 | Excitement,Mutism,Staring,Posturing/catalepsy,Mannerisms,Verbigeration,Withdrawal | 98.9 | 97.8 | 99.8 |
| 7 | Excitement,Mutism,Staring,Posturing/catalepsy,Mannerisms,Waxy flexibility,Withdrawal | 98.9 | 97.8 | 99.8 |
| 7 | Excitement,Mutism,Staring,Posturing/catalepsy,Stereotypy,Waxy flexibility,Withdrawal | 98.9 | 97.8 | 99.8 |
| 7 | Excitement,Mutism,Staring,Posturing/catalepsy,Verbigeration,Waxy flexibility,Withdrawal | 98.9 | 97.8 | 99.8 |
| 7 | Excitement,Mutism,Staring,Stereotypy,Mannerisms,Negativism,Withdrawal | 98.9 | 97.8 | 99.8 |
| 7 | Excitement,Mutism,Staring,Stereotypy,Mannerisms,Rigidity,Negativism | 98.9 | 97.8 | 99.8 |
| 7 | Excitement,Mutism,Staring,Stereotypy,Mannerisms,Rigidity,Waxy flexibility | 98.9 | 97.8 | 99.8 |
| 7 | Excitement,Mutism,Staring,Stereotypy,Mannerisms,Verbigeration,Rigidity | 98.9 | 97.8 | 99.8 |
| 7 | Excitement,Mutism,Staring,Verbigeration,Negativism,Waxy flexibility,Withdrawal | 98.9 | 97.8 | 99.8 |
| 7 | Excitement,Staring,Echopraxia/echolalia,Rigidity,Negativism,Waxy flexibility,Withdrawal | 98.9 | 97.8 | 99.8 |
| 7 | Excitement,Staring,Echopraxia/echolalia,Stereotypy,Mannerisms,Rigidity,Withdrawal | 98.9 | 97.8 | 99.8 |
| 7 | Excitement,Staring,Grimacing,Mannerisms,Rigidity,Waxy flexibility,Withdrawal | 98.9 | 97.8 | 99.8 |
| 7 | Excitement,Staring,Posturing/catalepsy,Echopraxia/echolalia,Mannerisms,Negativism,Withdrawal | 98.9 | 97.8 | 99.8 |
| 7 | Excitement,Staring,Posturing/catalepsy,Grimacing,Echopraxia/echolalia,Rigidity,Withdrawal | 98.9 | 97.8 | 99.8 |
| 7 | Excitement,Staring,Posturing/catalepsy,Mannerisms,Rigidity,Waxy flexibility,Withdrawal | 98.9 | 97.8 | 99.8 |
| 7 | Excitement,Staring,Posturing/catalepsy,Mannerisms,Verbigeration,Negativism,Withdrawal | 98.9 | 97.8 | 99.8 |
| 7 | Excitement,Staring,Posturing/catalepsy,Stereotypy,Mannerisms,Rigidity,Withdrawal | 98.9 | 97.8 | 99.8 |
| 7 | Excitement,Staring,Posturing/catalepsy,Stereotypy,Rigidity,Negativism,Withdrawal | 98.9 | 97.8 | 99.8 |
| 7 | Excitement,Staring,Posturing/catalepsy,Verbigeration,Rigidity,Negativism,Withdrawal | 98.9 | 97.8 | 99.8 |
| 7 | Excitement,Staring,Stereotypy,Mannerisms,Rigidity,Waxy flexibility,Withdrawal | 98.9 | 97.8 | 99.8 |
| 7 | Excitement,Staring,Stereotypy,Mannerisms,Verbigeration,Rigidity,Withdrawal | 98.9 | 97.8 | 99.8 |
| 7 | Excitement,Staring,Stereotypy,Rigidity,Negativism,Waxy flexibility,Withdrawal | 98.9 | 97.8 | 99.8 |
| 7 | Immobility/stupor,Mutism,Staring,Posturing/catalepsy,Mannerisms,Negativism,Withdrawal | 98.9 | 97.8 | 99.8 |
| 7 | Immobility/stupor,Staring,Echopraxia/echolalia,Stereotypy,Mannerisms,Negativism,Withdrawal | 98.9 | 97.8 | 99.8 |
| 7 | Immobility/stupor,Staring,Echopraxia/echolalia,Stereotypy,Mannerisms,Rigidity,Negativism | 98.9 | 97.8 | 99.8 |
| 7 | Immobility/stupor,Staring,Grimacing,Mannerisms,Rigidity,Negativism,Withdrawal | 98.9 | 97.8 | 99.8 |
| 7 | Immobility/stupor,Staring,Mannerisms,Rigidity,Negativism,Waxy flexibility,Withdrawal | 98.9 | 97.8 | 99.8 |
| 7 | Immobility/stupor,Staring,Posturing/catalepsy,Echopraxia/echolalia,Mannerisms,Negativism,Withdrawal | 98.9 | 97.8 | 99.8 |
| 7 | Immobility/stupor,Staring,Posturing/catalepsy,Echopraxia/echolalia,Mannerisms,Rigidity,Negativism | 98.9 | 97.8 | 99.8 |
| 7 | Immobility/stupor,Staring,Posturing/catalepsy,Stereotypy,Mannerisms,Negativism,Withdrawal | 98.9 | 97.8 | 99.8 |
| 7 | Immobility/stupor,Staring,Posturing/catalepsy,Stereotypy,Mannerisms,Rigidity,Negativism | 98.9 | 97.8 | 99.8 |
| 7 | Immobility/stupor,Staring,Stereotypy,Mannerisms,Rigidity,Negativism,Waxy flexibility | 98.9 | 97.8 | 99.8 |
| 7 | Immobility/stupor,Staring,Stereotypy,Verbigeration,Rigidity,Negativism,Withdrawal | 98.9 | 97.8 | 99.8 |
| 7 | Mutism,Staring,Echopraxia/echolalia,Mannerisms,Rigidity,Negativism,Waxy flexibility | 98.9 | 97.8 | 99.8 |
| 7 | Mutism,Staring,Echopraxia/echolalia,Mannerisms,Verbigeration,Rigidity,Negativism | 98.9 | 97.8 | 99.8 |
| 7 | Mutism,Staring,Grimacing,Echopraxia/echolalia,Mannerisms,Negativism,Withdrawal | 98.9 | 97.8 | 99.8 |
| 7 | Mutism,Staring,Grimacing,Echopraxia/echolalia,Mannerisms,Rigidity,Negativism | 98.9 | 97.8 | 99.8 |
| 7 | Mutism,Staring,Grimacing,Mannerisms,Rigidity,Negativism,Withdrawal | 98.9 | 97.8 | 99.8 |
| 7 | Mutism,Staring,Grimacing,Stereotypy,Mannerisms,Negativism,Withdrawal | 98.9 | 97.8 | 99.8 |
| 7 | Mutism,Staring,Grimacing,Stereotypy,Mannerisms,Rigidity,Withdrawal | 98.9 | 97.8 | 99.8 |
| 7 | Mutism,Staring,Mannerisms,Rigidity,Negativism,Waxy flexibility,Withdrawal | 98.9 | 97.8 | 99.8 |
| 7 | Mutism,Staring,Posturing/catalepsy,Echopraxia/echolalia,Stereotypy,Mannerisms,Negativism | 98.9 | 97.8 | 99.8 |
| 7 | Mutism,Staring,Posturing/catalepsy,Grimacing,Echopraxia/echolalia,Mannerisms,Negativism | 98.9 | 97.8 | 99.8 |
| 7 | Mutism,Staring,Posturing/catalepsy,Mannerisms,Negativism,Waxy flexibility,Withdrawal | 98.9 | 97.8 | 99.8 |
| 7 | Mutism,Staring,Posturing/catalepsy,Stereotypy,Mannerisms,Rigidity,Negativism | 98.9 | 97.8 | 99.8 |
| 7 | Mutism,Staring,Posturing/catalepsy,Stereotypy,Mannerisms,Rigidity,Withdrawal | 98.9 | 97.8 | 99.8 |
| 7 | Mutism,Staring,Stereotypy,Mannerisms,Rigidity,Negativism,Waxy flexibility | 98.9 | 97.8 | 99.8 |
| 7 | Mutism,Staring,Stereotypy,Mannerisms,Verbigeration,Negativism,Withdrawal | 98.9 | 97.8 | 99.8 |
| 7 | Staring,Echopraxia/echolalia,Mannerisms,Rigidity,Negativism,Waxy flexibility,Withdrawal | 98.9 | 97.8 | 99.8 |
| 7 | Staring,Grimacing,Echopraxia/echolalia,Mannerisms,Rigidity,Negativism,Withdrawal | 98.9 | 97.8 | 99.8 |
| 7 | Staring,Posturing/catalepsy,Grimacing,Mannerisms,Rigidity,Negativism,Withdrawal | 98.9 | 97.8 | 99.8 |
| 7 | Staring,Posturing/catalepsy,Mannerisms,Verbigeration,Rigidity,Negativism,Withdrawal | 98.9 | 97.8 | 99.8 |
| 7 | Excitement,Immobility/stupor,Mutism,Echopraxia/echolalia,Stereotypy,Rigidity,Negativism | 98.7 | 97.5 | 99.6 |
| 7 | Excitement,Immobility/stupor,Mutism,Echopraxia/echolalia,Stereotypy,Rigidity,Withdrawal | 98.7 | 97.5 | 99.6 |
| 7 | Excitement,Immobility/stupor,Mutism,Echopraxia/echolalia,Verbigeration,Rigidity,Withdrawal | 98.7 | 97.5 | 99.6 |
| 7 | Excitement,Immobility/stupor,Mutism,Grimacing,Echopraxia/echolalia,Rigidity,Negativism | 98.7 | 97.5 | 99.6 |
| 7 | Excitement,Immobility/stupor,Mutism,Grimacing,Verbigeration,Rigidity,Negativism | 98.7 | 97.5 | 99.6 |
| 7 | Excitement,Immobility/stupor,Mutism,Mannerisms,Verbigeration,Rigidity,Negativism | 98.7 | 97.5 | 99.6 |
| 7 | Excitement,Immobility/stupor,Mutism,Posturing/catalepsy,Grimacing,Echopraxia/echolalia,Rigidity | 98.7 | 97.5 | 99.6 |
| 7 | Excitement,Immobility/stupor,Mutism,Posturing/catalepsy,Grimacing,Rigidity,Negativism | 98.7 | 97.5 | 99.6 |
| 7 | Excitement,Immobility/stupor,Mutism,Posturing/catalepsy,Grimacing,Rigidity,Withdrawal | 98.7 | 97.5 | 99.6 |
| 7 | Excitement,Immobility/stupor,Mutism,Posturing/catalepsy,Grimacing,Verbigeration,Rigidity | 98.7 | 97.5 | 99.6 |
| 7 | Excitement,Immobility/stupor,Mutism,Posturing/catalepsy,Rigidity,Negativism,Withdrawal | 98.7 | 97.5 | 99.6 |
| 7 | Excitement,Immobility/stupor,Mutism,Posturing/catalepsy,Stereotypy,Rigidity,Negativism | 98.7 | 97.5 | 99.6 |
| 7 | Excitement,Immobility/stupor,Mutism,Posturing/catalepsy,Verbigeration,Rigidity,Withdrawal | 98.7 | 97.5 | 99.6 |
| 7 | Excitement,Immobility/stupor,Mutism,Staring,Echopraxia/echolalia,Verbigeration,Negativism | 98.7 | 97.5 | 99.6 |
| 7 | Excitement,Immobility/stupor,Mutism,Staring,Echopraxia/echolalia,Waxy flexibility,Withdrawal | 98.7 | 97.5 | 99.6 |
| 7 | Excitement,Immobility/stupor,Mutism,Staring,Grimacing,Rigidity,Negativism | 98.7 | 97.5 | 99.6 |
| 7 | Excitement,Immobility/stupor,Mutism,Staring,Grimacing,Rigidity,Waxy flexibility | 98.7 | 97.5 | 99.6 |
| 7 | Excitement,Immobility/stupor,Mutism,Staring,Grimacing,Stereotypy,Rigidity | 98.7 | 97.5 | 99.6 |
| 7 | Excitement,Immobility/stupor,Mutism,Staring,Mannerisms,Negativism,Waxy flexibility | 98.7 | 97.5 | 99.6 |
| 7 | Excitement,Immobility/stupor,Mutism,Staring,Negativism,Waxy flexibility,Withdrawal | 98.7 | 97.5 | 99.6 |
| 7 | Excitement,Immobility/stupor,Mutism,Staring,Posturing/catalepsy,Echopraxia/echolalia,Verbigeration | 98.7 | 97.5 | 99.6 |
| 7 | Excitement,Immobility/stupor,Mutism,Staring,Posturing/catalepsy,Grimacing,Echopraxia/echolalia | 98.7 | 97.5 | 99.6 |
| 7 | Excitement,Immobility/stupor,Mutism,Staring,Posturing/catalepsy,Grimacing,Withdrawal | 98.7 | 97.5 | 99.6 |
| 7 | Excitement,Immobility/stupor,Mutism,Staring,Posturing/catalepsy,Mannerisms,Withdrawal | 98.7 | 97.5 | 99.6 |
| 7 | Excitement,Immobility/stupor,Mutism,Staring,Posturing/catalepsy,Verbigeration,Waxy flexibility | 98.7 | 97.5 | 99.6 |
| 7 | Excitement,Immobility/stupor,Mutism,Staring,Posturing/catalepsy,Waxy flexibility,Withdrawal | 98.7 | 97.5 | 99.6 |
| 7 | Excitement,Immobility/stupor,Mutism,Staring,Stereotypy,Mannerisms,Rigidity | 98.7 | 97.5 | 99.6 |
| 7 | Excitement,Immobility/stupor,Mutism,Staring,Verbigeration,Negativism,Withdrawal | 98.7 | 97.5 | 99.6 |
| 7 | Excitement,Immobility/stupor,Mutism,Stereotypy,Rigidity,Negativism,Withdrawal | 98.7 | 97.5 | 99.6 |
| 7 | Excitement,Immobility/stupor,Mutism,Stereotypy,Verbigeration,Rigidity,Withdrawal | 98.7 | 97.5 | 99.6 |
| 7 | Excitement,Immobility/stupor,Mutism,Verbigeration,Rigidity,Negativism,Waxy flexibility | 98.7 | 97.5 | 99.6 |
| 7 | Excitement,Immobility/stupor,Posturing/catalepsy,Echopraxia/echolalia,Verbigeration,Rigidity,Withdrawal | 98.7 | 97.5 | 99.6 |
| 7 | Excitement,Immobility/stupor,Posturing/catalepsy,Grimacing,Echopraxia/echolalia,Rigidity,Withdrawal | 98.7 | 97.5 | 99.6 |
| 7 | Excitement,Immobility/stupor,Posturing/catalepsy,Grimacing,Rigidity,Negativism,Withdrawal | 98.7 | 97.5 | 99.6 |
| 7 | Excitement,Immobility/stupor,Posturing/catalepsy,Mannerisms,Rigidity,Negativism,Withdrawal | 98.7 | 97.5 | 99.6 |
| 7 | Excitement,Immobility/stupor,Posturing/catalepsy,Stereotypy,Rigidity,Negativism,Withdrawal | 98.7 | 97.5 | 99.6 |
| 7 | Excitement,Immobility/stupor,Staring,Echopraxia/echolalia,Mannerisms,Negativism,Waxy flexibility | 98.7 | 97.5 | 99.6 |
| 7 | Excitement,Immobility/stupor,Staring,Echopraxia/echolalia,Mannerisms,Verbigeration,Rigidity | 98.7 | 97.5 | 99.6 |
| 7 | Excitement,Immobility/stupor,Staring,Echopraxia/echolalia,Stereotypy,Rigidity,Waxy flexibility | 98.7 | 97.5 | 99.6 |
| 7 | Excitement,Immobility/stupor,Staring,Echopraxia/echolalia,Stereotypy,Verbigeration,Rigidity | 98.7 | 97.5 | 99.6 |
| 7 | Excitement,Immobility/stupor,Staring,Echopraxia/echolalia,Verbigeration,Negativism,Waxy flexibility | 98.7 | 97.5 | 99.6 |
| 7 | Excitement,Immobility/stupor,Staring,Echopraxia/echolalia,Verbigeration,Negativism,Withdrawal | 98.7 | 97.5 | 99.6 |
| 7 | Excitement,Immobility/stupor,Staring,Grimacing,Echopraxia/echolalia,Negativism,Withdrawal | 98.7 | 97.5 | 99.6 |
| 7 | Excitement,Immobility/stupor,Staring,Grimacing,Echopraxia/echolalia,Rigidity,Waxy flexibility | 98.7 | 97.5 | 99.6 |
| 7 | Excitement,Immobility/stupor,Staring,Grimacing,Mannerisms,Rigidity,Negativism | 98.7 | 97.5 | 99.6 |
| 7 | Excitement,Immobility/stupor,Staring,Grimacing,Mannerisms,Rigidity,Waxy flexibility | 98.7 | 97.5 | 99.6 |
| 7 | Excitement,Immobility/stupor,Staring,Grimacing,Mannerisms,Verbigeration,Rigidity | 98.7 | 97.5 | 99.6 |
| 7 | Excitement,Immobility/stupor,Staring,Grimacing,Rigidity,Negativism,Waxy flexibility | 98.7 | 97.5 | 99.6 |
| 7 | Excitement,Immobility/stupor,Staring,Grimacing,Stereotypy,Rigidity,Negativism | 98.7 | 97.5 | 99.6 |
| 7 | Excitement,Immobility/stupor,Staring,Grimacing,Stereotypy,Rigidity,Waxy flexibility | 98.7 | 97.5 | 99.6 |
| 7 | Excitement,Immobility/stupor,Staring,Grimacing,Stereotypy,Verbigeration,Rigidity | 98.7 | 97.5 | 99.6 |
| 7 | Excitement,Immobility/stupor,Staring,Grimacing,Verbigeration,Rigidity,Waxy flexibility | 98.7 | 97.5 | 99.6 |
| 7 | Excitement,Immobility/stupor,Staring,Mannerisms,Verbigeration,Negativism,Waxy flexibility | 98.7 | 97.5 | 99.6 |
| 7 | Excitement,Immobility/stupor,Staring,Mannerisms,Verbigeration,Negativism,Withdrawal | 98.7 | 97.5 | 99.6 |
| 7 | Excitement,Immobility/stupor,Staring,Posturing/catalepsy,Echopraxia/echolalia,Mannerisms,Withdrawal | 98.7 | 97.5 | 99.6 |
| 7 | Excitement,Immobility/stupor,Staring,Posturing/catalepsy,Grimacing,Mannerisms,Negativism | 98.7 | 97.5 | 99.6 |
| 7 | Excitement,Immobility/stupor,Staring,Posturing/catalepsy,Grimacing,Stereotypy,Rigidity | 98.7 | 97.5 | 99.6 |
| 7 | Excitement,Immobility/stupor,Staring,Posturing/catalepsy,Mannerisms,Rigidity,Waxy flexibility | 98.7 | 97.5 | 99.6 |
| 7 | Excitement,Immobility/stupor,Staring,Posturing/catalepsy,Stereotypy,Mannerisms,Negativism | 98.7 | 97.5 | 99.6 |
| 7 | Excitement,Immobility/stupor,Staring,Posturing/catalepsy,Stereotypy,Mannerisms,Rigidity | 98.7 | 97.5 | 99.6 |
| 7 | Excitement,Immobility/stupor,Staring,Posturing/catalepsy,Stereotypy,Negativism,Waxy flexibility | 98.7 | 97.5 | 99.6 |
| 7 | Excitement,Immobility/stupor,Staring,Stereotypy,Mannerisms,Rigidity,Waxy flexibility | 98.7 | 97.5 | 99.6 |
| 7 | Excitement,Immobility/stupor,Staring,Stereotypy,Verbigeration,Negativism,Waxy flexibility | 98.7 | 97.5 | 99.6 |
| 7 | Excitement,Mutism,Staring,Echopraxia/echolalia,Mannerisms,Verbigeration,Negativism | 98.7 | 97.5 | 99.6 |
| 7 | Excitement,Mutism,Staring,Echopraxia/echolalia,Mannerisms,Verbigeration,Rigidity | 98.7 | 97.5 | 99.6 |
| 7 | Excitement,Mutism,Staring,Grimacing,Echopraxia/echolalia,Stereotypy,Waxy flexibility | 98.7 | 97.5 | 99.6 |
| 7 | Excitement,Mutism,Staring,Grimacing,Echopraxia/echolalia,Stereotypy,Withdrawal | 98.7 | 97.5 | 99.6 |
| 7 | Excitement,Mutism,Staring,Grimacing,Echopraxia/echolalia,Verbigeration,Negativism | 98.7 | 97.5 | 99.6 |
| 7 | Excitement,Mutism,Staring,Grimacing,Mannerisms,Rigidity,Negativism | 98.7 | 97.5 | 99.6 |
| 7 | Excitement,Mutism,Staring,Grimacing,Negativism,Waxy flexibility,Withdrawal | 98.7 | 97.5 | 99.6 |
| 7 | Excitement,Mutism,Staring,Grimacing,Verbigeration,Negativism,Waxy flexibility | 98.7 | 97.5 | 99.6 |
| 7 | Excitement,Mutism,Staring,Grimacing,Verbigeration,Negativism,Withdrawal | 98.7 | 97.5 | 99.6 |
| 7 | Excitement,Mutism,Staring,Posturing/catalepsy,Echopraxia/echolalia,Mannerisms,Waxy flexibility | 98.7 | 97.5 | 99.6 |
| 7 | Excitement,Mutism,Staring,Posturing/catalepsy,Echopraxia/echolalia,Stereotypy,Verbigeration | 98.7 | 97.5 | 99.6 |
| 7 | Excitement,Mutism,Staring,Posturing/catalepsy,Grimacing,Echopraxia/echolalia,Stereotypy | 98.7 | 97.5 | 99.6 |
| 7 | Excitement,Mutism,Staring,Posturing/catalepsy,Grimacing,Echopraxia/echolalia,Verbigeration | 98.7 | 97.5 | 99.6 |
| 7 | Excitement,Mutism,Staring,Posturing/catalepsy,Grimacing,Rigidity,Waxy flexibility | 98.7 | 97.5 | 99.6 |
| 7 | Excitement,Mutism,Staring,Posturing/catalepsy,Stereotypy,Mannerisms,Withdrawal | 98.7 | 97.5 | 99.6 |
| 7 | Excitement,Mutism,Staring,Posturing/catalepsy,Stereotypy,Verbigeration,Waxy flexibility | 98.7 | 97.5 | 99.6 |
| 7 | Excitement,Mutism,Staring,Posturing/catalepsy,Stereotypy,Verbigeration,Withdrawal | 98.7 | 97.5 | 99.6 |
| 7 | Excitement,Mutism,Staring,Stereotypy,Mannerisms,Negativism,Waxy flexibility | 98.7 | 97.5 | 99.6 |
| 7 | Excitement,Mutism,Staring,Stereotypy,Mannerisms,Verbigeration,Negativism | 98.7 | 97.5 | 99.6 |
| 7 | Excitement,Staring,Echopraxia/echolalia,Mannerisms,Rigidity,Waxy flexibility,Withdrawal | 98.7 | 97.5 | 99.6 |
| 7 | Excitement,Staring,Echopraxia/echolalia,Stereotypy,Rigidity,Waxy flexibility,Withdrawal | 98.7 | 97.5 | 99.6 |
| 7 | Excitement,Staring,Echopraxia/echolalia,Verbigeration,Rigidity,Negativism,Withdrawal | 98.7 | 97.5 | 99.6 |
| 7 | Excitement,Staring,Grimacing,Echopraxia/echolalia,Mannerisms,Rigidity,Withdrawal | 98.7 | 97.5 | 99.6 |
| 7 | Excitement,Staring,Grimacing,Echopraxia/echolalia,Rigidity,Negativism,Withdrawal | 98.7 | 97.5 | 99.6 |
| 7 | Excitement,Staring,Grimacing,Echopraxia/echolalia,Rigidity,Waxy flexibility,Withdrawal | 98.7 | 97.5 | 99.6 |
| 7 | Excitement,Staring,Grimacing,Echopraxia/echolalia,Stereotypy,Rigidity,Withdrawal | 98.7 | 97.5 | 99.6 |
| 7 | Excitement,Staring,Grimacing,Mannerisms,Rigidity,Negativism,Withdrawal | 98.7 | 97.5 | 99.6 |
| 7 | Excitement,Staring,Grimacing,Mannerisms,Verbigeration,Rigidity,Withdrawal | 98.7 | 97.5 | 99.6 |
| 7 | Excitement,Staring,Grimacing,Stereotypy,Mannerisms,Rigidity,Withdrawal | 98.7 | 97.5 | 99.6 |
| 7 | Excitement,Staring,Grimacing,Stereotypy,Rigidity,Negativism,Withdrawal | 98.7 | 97.5 | 99.6 |
| 7 | Excitement,Staring,Grimacing,Stereotypy,Rigidity,Waxy flexibility,Withdrawal | 98.7 | 97.5 | 99.6 |
| 7 | Excitement,Staring,Grimacing,Stereotypy,Verbigeration,Rigidity,Withdrawal | 98.7 | 97.5 | 99.6 |
| 7 | Excitement,Staring,Mannerisms,Verbigeration,Rigidity,Waxy flexibility,Withdrawal | 98.7 | 97.5 | 99.6 |
| 7 | Excitement,Staring,Posturing/catalepsy,Echopraxia/echolalia,Negativism,Waxy flexibility,Withdrawal | 98.7 | 97.5 | 99.6 |
| 7 | Excitement,Staring,Posturing/catalepsy,Grimacing,Echopraxia/echolalia,Negativism,Withdrawal | 98.7 | 97.5 | 99.6 |
| 7 | Excitement,Staring,Posturing/catalepsy,Grimacing,Mannerisms,Negativism,Withdrawal | 98.7 | 97.5 | 99.6 |
| 7 | Excitement,Staring,Posturing/catalepsy,Grimacing,Rigidity,Negativism,Withdrawal | 98.7 | 97.5 | 99.6 |
| 7 | Excitement,Staring,Posturing/catalepsy,Grimacing,Stereotypy,Rigidity,Withdrawal | 98.7 | 97.5 | 99.6 |
| 7 | Excitement,Staring,Posturing/catalepsy,Grimacing,Verbigeration,Rigidity,Withdrawal | 98.7 | 97.5 | 99.6 |
| 7 | Excitement,Staring,Posturing/catalepsy,Mannerisms,Negativism,Waxy flexibility,Withdrawal | 98.7 | 97.5 | 99.6 |
| 7 | Excitement,Staring,Posturing/catalepsy,Rigidity,Negativism,Waxy flexibility,Withdrawal | 98.7 | 97.5 | 99.6 |
| 7 | Excitement,Staring,Verbigeration,Rigidity,Negativism,Waxy flexibility,Withdrawal | 98.7 | 97.5 | 99.6 |
| 7 | Immobility/stupor,Mutism,Staring,Mannerisms,Rigidity,Negativism,Withdrawal | 98.7 | 97.5 | 99.6 |
| 7 | Immobility/stupor,Mutism,Staring,Stereotypy,Mannerisms,Rigidity,Negativism | 98.7 | 97.5 | 99.6 |
| 7 | Immobility/stupor,Staring,Echopraxia/echolalia,Mannerisms,Rigidity,Negativism,Waxy flexibility | 98.7 | 97.5 | 99.6 |
| 7 | Immobility/stupor,Staring,Echopraxia/echolalia,Mannerisms,Verbigeration,Rigidity,Negativism | 98.7 | 97.5 | 99.6 |
| 7 | Immobility/stupor,Staring,Grimacing,Echopraxia/echolalia,Mannerisms,Rigidity,Negativism | 98.7 | 97.5 | 99.6 |
| 7 | Immobility/stupor,Staring,Grimacing,Stereotypy,Mannerisms,Rigidity,Negativism | 98.7 | 97.5 | 99.6 |
| 7 | Immobility/stupor,Staring,Posturing/catalepsy,Grimacing,Mannerisms,Negativism,Withdrawal | 98.7 | 97.5 | 99.6 |
| 7 | Immobility/stupor,Staring,Posturing/catalepsy,Mannerisms,Verbigeration,Negativism,Withdrawal | 98.7 | 97.5 | 99.6 |
| 7 | Immobility/stupor,Staring,Posturing/catalepsy,Mannerisms,Verbigeration,Rigidity,Negativism | 98.7 | 97.5 | 99.6 |
| 7 | Immobility/stupor,Staring,Stereotypy,Mannerisms,Verbigeration,Negativism,Withdrawal | 98.7 | 97.5 | 99.6 |
| 7 | Mutism,Staring,Echopraxia/echolalia,Mannerisms,Negativism,Waxy flexibility,Withdrawal | 98.7 | 97.5 | 99.6 |
| 7 | Mutism,Staring,Echopraxia/echolalia,Stereotypy,Mannerisms,Negativism,Waxy flexibility | 98.7 | 97.5 | 99.6 |
| 7 | Mutism,Staring,Echopraxia/echolalia,Stereotypy,Mannerisms,Rigidity,Withdrawal | 98.7 | 97.5 | 99.6 |
| 7 | Mutism,Staring,Echopraxia/echolalia,Stereotypy,Rigidity,Negativism,Withdrawal | 98.7 | 97.5 | 99.6 |
| 7 | Mutism,Staring,Grimacing,Echopraxia/echolalia,Mannerisms,Rigidity,Withdrawal | 98.7 | 97.5 | 99.6 |
| 7 | Mutism,Staring,Grimacing,Echopraxia/echolalia,Stereotypy,Mannerisms,Negativism | 98.7 | 97.5 | 99.6 |
| 7 | Mutism,Staring,Grimacing,Mannerisms,Verbigeration,Rigidity,Withdrawal | 98.7 | 97.5 | 99.6 |
| 7 | Mutism,Staring,Grimacing,Stereotypy,Mannerisms,Rigidity,Negativism | 98.7 | 97.5 | 99.6 |
| 7 | Mutism,Staring,Grimacing,Stereotypy,Verbigeration,Rigidity,Negativism | 98.7 | 97.5 | 99.6 |
| 7 | Mutism,Staring,Posturing/catalepsy,Echopraxia/echolalia,Mannerisms,Rigidity,Withdrawal | 98.7 | 97.5 | 99.6 |
| 7 | Mutism,Staring,Posturing/catalepsy,Echopraxia/echolalia,Mannerisms,Verbigeration,Negativism | 98.7 | 97.5 | 99.6 |
| 7 | Mutism,Staring,Posturing/catalepsy,Grimacing,Stereotypy,Mannerisms,Negativism | 98.7 | 97.5 | 99.6 |
| 7 | Mutism,Staring,Posturing/catalepsy,Stereotypy,Mannerisms,Verbigeration,Negativism | 98.7 | 97.5 | 99.6 |
| 7 | Mutism,Staring,Posturing/catalepsy,Stereotypy,Verbigeration,Negativism,Withdrawal | 98.7 | 97.5 | 99.6 |
| 7 | Mutism,Staring,Stereotypy,Mannerisms,Negativism,Waxy flexibility,Withdrawal | 98.7 | 97.5 | 99.6 |
| 7 | Mutism,Staring,Stereotypy,Mannerisms,Rigidity,Waxy flexibility,Withdrawal | 98.7 | 97.5 | 99.6 |
| 7 | Mutism,Staring,Stereotypy,Mannerisms,Verbigeration,Rigidity,Withdrawal | 98.7 | 97.5 | 99.6 |
| 7 | Staring,Echopraxia/echolalia,Mannerisms,Verbigeration,Rigidity,Negativism,Withdrawal | 98.7 | 97.5 | 99.6 |
| 7 | Staring,Grimacing,Echopraxia/echolalia,Stereotypy,Mannerisms,Negativism,Withdrawal | 98.7 | 97.5 | 99.6 |
| 7 | Staring,Grimacing,Mannerisms,Rigidity,Negativism,Waxy flexibility,Withdrawal | 98.7 | 97.5 | 99.6 |
| 7 | Staring,Grimacing,Mannerisms,Verbigeration,Rigidity,Negativism,Withdrawal | 98.7 | 97.5 | 99.6 |
| 7 | Staring,Mannerisms,Verbigeration,Rigidity,Negativism,Waxy flexibility,Withdrawal | 98.7 | 97.5 | 99.6 |
| 7 | Staring,Posturing/catalepsy,Grimacing,Echopraxia/echolalia,Mannerisms,Negativism,Withdrawal | 98.7 | 97.5 | 99.6 |
| 7 | Staring,Posturing/catalepsy,Grimacing,Stereotypy,Mannerisms,Negativism,Withdrawal | 98.7 | 97.5 | 99.6 |
| 7 | Staring,Posturing/catalepsy,Mannerisms,Rigidity,Negativism,Waxy flexibility,Withdrawal | 98.7 | 97.5 | 99.6 |
| 7 | Excitement,Immobility/stupor,Echopraxia/echolalia,Mannerisms,Rigidity,Negativism,Withdrawal | 98.6 | 97.5 | 99.6 |
| 7 | Excitement,Immobility/stupor,Mannerisms,Verbigeration,Rigidity,Negativism,Withdrawal | 98.6 | 97.5 | 99.6 |
| 7 | Excitement,Immobility/stupor,Mutism,Grimacing,Echopraxia/echolalia,Rigidity,Withdrawal | 98.6 | 97.5 | 99.6 |
| 7 | Excitement,Immobility/stupor,Staring,Echopraxia/echolalia,Mannerisms,Rigidity,Waxy flexibility | 98.6 | 97.5 | 99.6 |
| 7 | Excitement,Immobility/stupor,Staring,Echopraxia/echolalia,Stereotypy,Negativism,Waxy flexibility | 98.6 | 97.5 | 99.6 |
| 7 | Excitement,Immobility/stupor,Staring,Posturing/catalepsy,Echopraxia/echolalia,Rigidity,Waxy flexibility | 98.6 | 97.5 | 99.6 |
| 7 | Excitement,Immobility/stupor,Staring,Posturing/catalepsy,Echopraxia/echolalia,Stereotypy,Rigidity | 98.6 | 97.5 | 99.6 |
| 7 | Excitement,Mutism,Staring,Grimacing,Stereotypy,Negativism,Waxy flexibility | 98.6 | 97.5 | 99.6 |
| 7 | Excitement,Mutism,Staring,Grimacing,Stereotypy,Verbigeration,Negativism | 98.6 | 97.5 | 99.6 |
| 7 | Excitement,Staring,Posturing/catalepsy,Echopraxia/echolalia,Rigidity,Waxy flexibility,Withdrawal | 98.6 | 97.5 | 99.6 |
| 7 | Excitement,Staring,Posturing/catalepsy,Echopraxia/echolalia,Stereotypy,Rigidity,Withdrawal | 98.6 | 97.5 | 99.6 |
| 7 | Excitement,Staring,Posturing/catalepsy,Echopraxia/echolalia,Verbigeration,Rigidity,Withdrawal | 98.6 | 97.5 | 99.6 |
| 7 | Excitement,Staring,Stereotypy,Verbigeration,Rigidity,Waxy flexibility,Withdrawal | 98.6 | 97.5 | 99.6 |
| 7 | Immobility/stupor,Mutism,Staring,Echopraxia/echolalia,Mannerisms,Rigidity,Negativism | 98.6 | 97.5 | 99.6 |
| 7 | Mutism,Staring,Grimacing,Stereotypy,Verbigeration,Rigidity,Withdrawal | 98.6 | 97.5 | 99.6 |
| 7 | Mutism,Staring,Posturing/catalepsy,Grimacing,Mannerisms,Rigidity,Withdrawal | 98.6 | 97.5 | 99.6 |
| 7 | Mutism,Staring,Posturing/catalepsy,Mannerisms,Verbigeration,Rigidity,Negativism | 98.6 | 97.5 | 99.6 |
| 7 | Mutism,Staring,Posturing/catalepsy,Mannerisms,Verbigeration,Rigidity,Withdrawal | 98.6 | 97.5 | 99.6 |
| 7 | Mutism,Staring,Posturing/catalepsy,Stereotypy,Verbigeration,Rigidity,Negativism | 98.6 | 97.5 | 99.6 |
| 7 | Excitement,Immobility/stupor,Echopraxia/echolalia,Rigidity,Negativism,Waxy flexibility,Withdrawal | 98.4 | 97.3 | 99.6 |
| 7 | Excitement,Immobility/stupor,Echopraxia/echolalia,Stereotypy,Verbigeration,Rigidity,Withdrawal | 98.4 | 97.1 | 99.6 |
| 7 | Excitement,Immobility/stupor,Mutism,Echopraxia/echolalia,Rigidity,Negativism,Waxy flexibility | 98.4 | 97.1 | 99.6 |
| 7 | Excitement,Immobility/stupor,Mutism,Echopraxia/echolalia,Rigidity,Waxy flexibility,Withdrawal | 98.4 | 97.1 | 99.6 |
| 7 | Excitement,Immobility/stupor,Mutism,Grimacing,Rigidity,Negativism,Withdrawal | 98.4 | 97.1 | 99.6 |
| 7 | Excitement,Immobility/stupor,Mutism,Grimacing,Verbigeration,Rigidity,Withdrawal | 98.4 | 97.3 | 99.6 |
| 7 | Excitement,Immobility/stupor,Mutism,Mannerisms,Verbigeration,Rigidity,Withdrawal | 98.4 | 97.3 | 99.6 |
| 7 | Excitement,Immobility/stupor,Mutism,Posturing/catalepsy,Echopraxia/echolalia,Negativism,Withdrawal | 98.4 | 97.1 | 99.6 |
| 7 | Excitement,Immobility/stupor,Mutism,Posturing/catalepsy,Echopraxia/echolalia,Stereotypy,Rigidity | 98.4 | 97.1 | 99.6 |
| 7 | Excitement,Immobility/stupor,Mutism,Posturing/catalepsy,Mannerisms,Rigidity,Negativism | 98.4 | 97.1 | 99.6 |
| 7 | Excitement,Immobility/stupor,Mutism,Posturing/catalepsy,Mannerisms,Rigidity,Withdrawal | 98.4 | 97.1 | 99.6 |
| 7 | Excitement,Immobility/stupor,Mutism,Posturing/catalepsy,Mannerisms,Verbigeration,Rigidity | 98.4 | 97.1 | 99.6 |
| 7 | Excitement,Immobility/stupor,Mutism,Posturing/catalepsy,Stereotypy,Rigidity,Withdrawal | 98.4 | 97.3 | 99.6 |
| 7 | Excitement,Immobility/stupor,Mutism,Posturing/catalepsy,Stereotypy,Verbigeration,Rigidity | 98.4 | 97.1 | 99.6 |
| 7 | Excitement,Immobility/stupor,Mutism,Posturing/catalepsy,Verbigeration,Negativism,Withdrawal | 98.4 | 97.1 | 99.6 |
| 7 | Excitement,Immobility/stupor,Mutism,Staring,Echopraxia/echolalia,Mannerisms,Negativism | 98.4 | 97.1 | 99.6 |
| 7 | Excitement,Immobility/stupor,Mutism,Staring,Echopraxia/echolalia,Stereotypy,Waxy flexibility | 98.4 | 97.3 | 99.6 |
| 7 | Excitement,Immobility/stupor,Mutism,Staring,Echopraxia/echolalia,Stereotypy,Withdrawal | 98.4 | 97.1 | 99.6 |
| 7 | Excitement,Immobility/stupor,Mutism,Staring,Echopraxia/echolalia,Verbigeration,Waxy flexibility | 98.4 | 97.3 | 99.6 |
| 7 | Excitement,Immobility/stupor,Mutism,Staring,Grimacing,Echopraxia/echolalia,Negativism | 98.4 | 97.3 | 99.6 |
| 7 | Excitement,Immobility/stupor,Mutism,Staring,Grimacing,Echopraxia/echolalia,Waxy flexibility | 98.4 | 97.3 | 99.6 |
| 7 | Excitement,Immobility/stupor,Mutism,Staring,Grimacing,Mannerisms,Rigidity | 98.4 | 97.3 | 99.6 |
| 7 | Excitement,Immobility/stupor,Mutism,Staring,Grimacing,Negativism,Waxy flexibility | 98.4 | 97.3 | 99.6 |
| 7 | Excitement,Immobility/stupor,Mutism,Staring,Mannerisms,Negativism,Withdrawal | 98.4 | 97.1 | 99.6 |
| 7 | Excitement,Immobility/stupor,Mutism,Staring,Mannerisms,Verbigeration,Negativism | 98.4 | 97.3 | 99.6 |
| 7 | Excitement,Immobility/stupor,Mutism,Staring,Posturing/catalepsy,Echopraxia/echolalia,Mannerisms | 98.4 | 97.1 | 99.6 |
| 7 | Excitement,Immobility/stupor,Mutism,Staring,Posturing/catalepsy,Echopraxia/echolalia,Stereotypy | 98.4 | 97.1 | 99.6 |
| 7 | Excitement,Immobility/stupor,Mutism,Staring,Posturing/catalepsy,Grimacing,Verbigeration | 98.4 | 97.3 | 99.6 |
| 7 | Excitement,Immobility/stupor,Mutism,Staring,Posturing/catalepsy,Mannerisms,Verbigeration | 98.4 | 97.1 | 99.6 |
| 7 | Excitement,Immobility/stupor,Mutism,Staring,Posturing/catalepsy,Mannerisms,Waxy flexibility | 98.4 | 97.1 | 99.6 |
| 7 | Excitement,Immobility/stupor,Mutism,Staring,Posturing/catalepsy,Stereotypy,Waxy flexibility | 98.4 | 97.1 | 99.6 |
| 7 | Excitement,Immobility/stupor,Mutism,Staring,Posturing/catalepsy,Stereotypy,Withdrawal | 98.4 | 97.3 | 99.6 |
| 7 | Excitement,Immobility/stupor,Mutism,Staring,Posturing/catalepsy,Verbigeration,Withdrawal | 98.4 | 97.1 | 99.6 |
| 7 | Excitement,Immobility/stupor,Mutism,Stereotypy,Rigidity,Negativism,Waxy flexibility | 98.4 | 97.1 | 99.6 |
| 7 | Excitement,Immobility/stupor,Mutism,Stereotypy,Rigidity,Waxy flexibility,Withdrawal | 98.4 | 97.1 | 99.6 |
| 7 | Excitement,Immobility/stupor,Mutism,Verbigeration,Rigidity,Waxy flexibility,Withdrawal | 98.4 | 97.1 | 99.6 |
| 7 | Excitement,Immobility/stupor,Posturing/catalepsy,Echopraxia/echolalia,Mannerisms,Rigidity,Withdrawal | 98.4 | 97.1 | 99.6 |
| 7 | Excitement,Immobility/stupor,Posturing/catalepsy,Echopraxia/echolalia,Stereotypy,Rigidity,Withdrawal | 98.4 | 97.1 | 99.6 |
| 7 | Excitement,Immobility/stupor,Posturing/catalepsy,Echopraxia/echolalia,Verbigeration,Rigidity,Negativism | 98.4 | 97.1 | 99.6 |
| 7 | Excitement,Immobility/stupor,Posturing/catalepsy,Grimacing,Echopraxia/echolalia,Rigidity,Negativism | 98.4 | 97.3 | 99.6 |
| 7 | Excitement,Immobility/stupor,Posturing/catalepsy,Grimacing,Verbigeration,Rigidity,Negativism | 98.4 | 97.3 | 99.6 |
| 7 | Excitement,Immobility/stupor,Posturing/catalepsy,Mannerisms,Verbigeration,Rigidity,Negativism | 98.4 | 97.3 | 99.6 |
| 7 | Excitement,Immobility/stupor,Posturing/catalepsy,Mannerisms,Verbigeration,Rigidity,Withdrawal | 98.4 | 97.1 | 99.6 |
| 7 | Excitement,Immobility/stupor,Posturing/catalepsy,Stereotypy,Verbigeration,Rigidity,Negativism | 98.4 | 97.1 | 99.6 |
| 7 | Excitement,Immobility/stupor,Staring,Echopraxia/echolalia,Stereotypy,Mannerisms,Negativism | 98.4 | 97.3 | 99.6 |
| 7 | Excitement,Immobility/stupor,Staring,Echopraxia/echolalia,Stereotypy,Mannerisms,Rigidity | 98.4 | 97.1 | 99.6 |
| 7 | Excitement,Immobility/stupor,Staring,Echopraxia/echolalia,Stereotypy,Verbigeration,Negativism | 98.4 | 97.1 | 99.6 |
| 7 | Excitement,Immobility/stupor,Staring,Grimacing,Echopraxia/echolalia,Mannerisms,Rigidity | 98.4 | 97.3 | 99.6 |
| 7 | Excitement,Immobility/stupor,Staring,Grimacing,Echopraxia/echolalia,Negativism,Waxy flexibility | 98.4 | 97.1 | 99.6 |
| 7 | Excitement,Immobility/stupor,Staring,Grimacing,Echopraxia/echolalia,Stereotypy,Rigidity | 98.4 | 97.1 | 99.6 |
| 7 | Excitement,Immobility/stupor,Staring,Grimacing,Echopraxia/echolalia,Verbigeration,Rigidity | 98.4 | 97.1 | 99.6 |
| 7 | Excitement,Immobility/stupor,Staring,Mannerisms,Negativism,Waxy flexibility,Withdrawal | 98.4 | 97.1 | 99.6 |
| 7 | Excitement,Immobility/stupor,Staring,Posturing/catalepsy,Echopraxia/echolalia,Waxy flexibility,Withdrawal | 98.4 | 97.1 | 99.3 |
| 7 | Excitement,Immobility/stupor,Staring,Posturing/catalepsy,Grimacing,Echopraxia/echolalia,Withdrawal | 98.4 | 97.3 | 99.6 |
| 7 | Excitement,Immobility/stupor,Staring,Posturing/catalepsy,Grimacing,Negativism,Waxy flexibility | 98.4 | 97.1 | 99.6 |
| 7 | Excitement,Immobility/stupor,Staring,Posturing/catalepsy,Grimacing,Stereotypy,Negativism | 98.4 | 97.1 | 99.6 |
| 7 | Excitement,Immobility/stupor,Staring,Posturing/catalepsy,Mannerisms,Verbigeration,Withdrawal | 98.4 | 97.1 | 99.6 |
| 7 | Excitement,Immobility/stupor,Staring,Posturing/catalepsy,Stereotypy,Rigidity,Waxy flexibility | 98.4 | 97.1 | 99.6 |
| 7 | Excitement,Immobility/stupor,Staring,Stereotypy,Mannerisms,Negativism,Withdrawal | 98.4 | 97.1 | 99.6 |
| 7 | Excitement,Immobility/stupor,Staring,Stereotypy,Mannerisms,Verbigeration,Negativism | 98.4 | 97.3 | 99.6 |
| 7 | Excitement,Immobility/stupor,Staring,Stereotypy,Negativism,Waxy flexibility,Withdrawal | 98.4 | 97.1 | 99.6 |
| 7 | Excitement,Immobility/stupor,Staring,Verbigeration,Negativism,Waxy flexibility,Withdrawal | 98.4 | 97.3 | 99.6 |
| 7 | Excitement,Mutism,Staring,Echopraxia/echolalia,Mannerisms,Waxy flexibility,Withdrawal | 98.4 | 97.1 | 99.6 |
| 7 | Excitement,Mutism,Staring,Echopraxia/echolalia,Stereotypy,Mannerisms,Withdrawal | 98.4 | 97.1 | 99.6 |
| 7 | Excitement,Mutism,Staring,Echopraxia/echolalia,Stereotypy,Verbigeration,Waxy flexibility | 98.4 | 97.1 | 99.6 |
| 7 | Excitement,Mutism,Staring,Echopraxia/echolalia,Stereotypy,Verbigeration,Withdrawal | 98.4 | 97.1 | 99.6 |
| 7 | Excitement,Mutism,Staring,Echopraxia/echolalia,Verbigeration,Waxy flexibility,Withdrawal | 98.4 | 97.1 | 99.6 |
| 7 | Excitement,Mutism,Staring,Grimacing,Echopraxia/echolalia,Mannerisms,Negativism | 98.4 | 97.3 | 99.6 |
| 7 | Excitement,Mutism,Staring,Grimacing,Echopraxia/echolalia,Verbigeration,Waxy flexibility | 98.4 | 97.3 | 99.6 |
| 7 | Excitement,Mutism,Staring,Grimacing,Mannerisms,Negativism,Waxy flexibility | 98.4 | 97.1 | 99.6 |
| 7 | Excitement,Mutism,Staring,Grimacing,Mannerisms,Negativism,Withdrawal | 98.4 | 97.1 | 99.6 |
| 7 | Excitement,Mutism,Staring,Grimacing,Stereotypy,Mannerisms,Rigidity | 98.4 | 97.1 | 99.6 |
| 7 | Excitement,Mutism,Staring,Grimacing,Stereotypy,Waxy flexibility,Withdrawal | 98.4 | 97.3 | 99.6 |
| 7 | Excitement,Mutism,Staring,Posturing/catalepsy,Echopraxia/echolalia,Mannerisms,Verbigeration | 98.4 | 97.1 | 99.6 |
| 7 | Excitement,Mutism,Staring,Posturing/catalepsy,Echopraxia/echolalia,Stereotypy,Mannerisms | 98.4 | 97.1 | 99.6 |
| 7 | Excitement,Mutism,Staring,Posturing/catalepsy,Grimacing,Echopraxia/echolalia,Mannerisms | 98.4 | 97.1 | 99.6 |
| 7 | Excitement,Mutism,Staring,Posturing/catalepsy,Grimacing,Stereotypy,Verbigeration | 98.4 | 97.1 | 99.6 |
| 7 | Excitement,Mutism,Staring,Posturing/catalepsy,Grimacing,Stereotypy,Waxy flexibility | 98.4 | 97.1 | 99.6 |
| 7 | Excitement,Mutism,Staring,Posturing/catalepsy,Grimacing,Verbigeration,Waxy flexibility | 98.4 | 97.1 | 99.6 |
| 7 | Excitement,Mutism,Staring,Posturing/catalepsy,Mannerisms,Verbigeration,Waxy flexibility | 98.4 | 97.1 | 99.6 |
| 7 | Excitement,Mutism,Staring,Stereotypy,Verbigeration,Waxy flexibility,Withdrawal | 98.4 | 97.3 | 99.6 |
| 7 | Excitement,Staring,Echopraxia/echolalia,Mannerisms,Negativism,Waxy flexibility,Withdrawal | 98.4 | 97.1 | 99.6 |
| 7 | Excitement,Staring,Echopraxia/echolalia,Stereotypy,Mannerisms,Negativism,Withdrawal | 98.4 | 97.1 | 99.6 |
| 7 | Excitement,Staring,Echopraxia/echolalia,Stereotypy,Verbigeration,Rigidity,Withdrawal | 98.4 | 97.1 | 99.6 |
| 7 | Excitement,Staring,Grimacing,Rigidity,Negativism,Waxy flexibility,Withdrawal | 98.4 | 97.1 | 99.6 |
| 7 | Excitement,Staring,Grimacing,Verbigeration,Rigidity,Negativism,Withdrawal | 98.4 | 97.3 | 99.3 |
| 7 | Excitement,Staring,Grimacing,Verbigeration,Rigidity,Waxy flexibility,Withdrawal | 98.4 | 97.3 | 99.6 |
| 7 | Excitement,Staring,Posturing/catalepsy,Echopraxia/echolalia,Stereotypy,Negativism,Withdrawal | 98.4 | 97.1 | 99.6 |
| 7 | Excitement,Staring,Posturing/catalepsy,Echopraxia/echolalia,Verbigeration,Negativism,Withdrawal | 98.4 | 97.3 | 99.6 |
| 7 | Excitement,Staring,Posturing/catalepsy,Grimacing,Rigidity,Waxy flexibility,Withdrawal | 98.4 | 97.3 | 99.6 |
| 7 | Excitement,Staring,Posturing/catalepsy,Grimacing,Verbigeration,Negativism,Withdrawal | 98.4 | 97.1 | 99.6 |
| 7 | Excitement,Staring,Posturing/catalepsy,Stereotypy,Mannerisms,Negativism,Withdrawal | 98.4 | 97.1 | 99.6 |
| 7 | Excitement,Staring,Posturing/catalepsy,Stereotypy,Rigidity,Waxy flexibility,Withdrawal | 98.4 | 97.1 | 99.6 |
| 7 | Excitement,Staring,Posturing/catalepsy,Stereotypy,Verbigeration,Negativism,Withdrawal | 98.4 | 97.3 | 99.6 |
| 7 | Excitement,Staring,Posturing/catalepsy,Stereotypy,Verbigeration,Rigidity,Withdrawal | 98.4 | 97.1 | 99.6 |
| 7 | Excitement,Staring,Posturing/catalepsy,Verbigeration,Negativism,Waxy flexibility,Withdrawal | 98.4 | 97.3 | 99.6 |
| 7 | Excitement,Staring,Posturing/catalepsy,Verbigeration,Rigidity,Waxy flexibility,Withdrawal | 98.4 | 97.1 | 99.6 |
| 7 | Immobility/stupor,Echopraxia/echolalia,Stereotypy,Mannerisms,Rigidity,Negativism,Withdrawal | 98.4 | 97.1 | 99.6 |
| 7 | Immobility/stupor,Mutism,Staring,Echopraxia/echolalia,Mannerisms,Negativism,Withdrawal | 98.4 | 97.3 | 99.6 |
| 7 | Immobility/stupor,Mutism,Staring,Mannerisms,Verbigeration,Rigidity,Negativism | 98.4 | 97.1 | 99.6 |
| 7 | Immobility/stupor,Mutism,Staring,Posturing/catalepsy,Mannerisms,Rigidity,Negativism | 98.4 | 97.3 | 99.6 |
| 7 | Immobility/stupor,Mutism,Staring,Posturing/catalepsy,Stereotypy,Mannerisms,Negativism | 98.4 | 97.1 | 99.6 |
| 7 | Immobility/stupor,Mutism,Staring,Stereotypy,Mannerisms,Negativism,Withdrawal | 98.4 | 97.3 | 99.6 |
| 7 | Immobility/stupor,Mutism,Staring,Stereotypy,Verbigeration,Rigidity,Negativism | 98.4 | 97.1 | 99.6 |
| 7 | Immobility/stupor,Staring,Echopraxia/echolalia,Stereotypy,Rigidity,Negativism,Withdrawal | 98.4 | 97.1 | 99.6 |
| 7 | Immobility/stupor,Staring,Grimacing,Echopraxia/echolalia,Mannerisms,Negativism,Withdrawal | 98.4 | 97.1 | 99.6 |
| 7 | Immobility/stupor,Staring,Grimacing,Mannerisms,Verbigeration,Rigidity,Negativism | 98.4 | 97.1 | 99.6 |
| 7 | Immobility/stupor,Staring,Grimacing,Stereotypy,Mannerisms,Negativism,Withdrawal | 98.4 | 97.1 | 99.6 |
| 7 | Immobility/stupor,Staring,Grimacing,Stereotypy,Mannerisms,Rigidity,Withdrawal | 98.4 | 97.1 | 99.6 |
| 7 | Immobility/stupor,Staring,Grimacing,Stereotypy,Verbigeration,Rigidity,Negativism | 98.4 | 97.3 | 99.6 |
| 7 | Immobility/stupor,Staring,Mannerisms,Verbigeration,Rigidity,Negativism,Waxy flexibility | 98.4 | 97.1 | 99.6 |
| 7 | Immobility/stupor,Staring,Posturing/catalepsy,Echopraxia/echolalia,Stereotypy,Mannerisms,Negativism | 98.4 | 97.3 | 99.6 |
| 7 | Immobility/stupor,Staring,Posturing/catalepsy,Grimacing,Mannerisms,Rigidity,Negativism | 98.4 | 97.1 | 99.6 |
| 7 | Immobility/stupor,Staring,Posturing/catalepsy,Stereotypy,Mannerisms,Verbigeration,Negativism | 98.4 | 97.1 | 99.6 |
| 7 | Immobility/stupor,Staring,Posturing/catalepsy,Stereotypy,Verbigeration,Rigidity,Negativism | 98.4 | 97.1 | 99.6 |
| 7 | Immobility/stupor,Stereotypy,Mannerisms,Verbigeration,Rigidity,Negativism,Withdrawal | 98.4 | 97.1 | 99.6 |
| 7 | Mutism,Staring,Echopraxia/echolalia,Mannerisms,Verbigeration,Negativism,Withdrawal | 98.4 | 97.1 | 99.6 |
| 7 | Mutism,Staring,Echopraxia/echolalia,Stereotypy,Mannerisms,Verbigeration,Negativism | 98.4 | 97.3 | 99.6 |
| 7 | Mutism,Staring,Echopraxia/echolalia,Stereotypy,Verbigeration,Rigidity,Negativism | 98.4 | 97.1 | 99.6 |
| 7 | Mutism,Staring,Echopraxia/echolalia,Verbigeration,Rigidity,Negativism,Withdrawal | 98.4 | 97.3 | 99.6 |
| 7 | Mutism,Staring,Grimacing,Echopraxia/echolalia,Mannerisms,Negativism,Waxy flexibility | 98.4 | 97.1 | 99.6 |
| 7 | Mutism,Staring,Grimacing,Mannerisms,Negativism,Waxy flexibility,Withdrawal | 98.4 | 97.1 | 99.6 |
| 7 | Mutism,Staring,Grimacing,Mannerisms,Rigidity,Waxy flexibility,Withdrawal | 98.4 | 97.1 | 99.6 |
| 7 | Mutism,Staring,Grimacing,Mannerisms,Verbigeration,Negativism,Withdrawal | 98.4 | 97.1 | 99.6 |
| 7 | Mutism,Staring,Grimacing,Mannerisms,Verbigeration,Rigidity,Negativism | 98.4 | 97.1 | 99.6 |
| 7 | Mutism,Staring,Grimacing,Stereotypy,Mannerisms,Verbigeration,Rigidity | 98.4 | 97.3 | 99.6 |
| 7 | Mutism,Staring,Grimacing,Stereotypy,Verbigeration,Negativism,Withdrawal | 98.4 | 97.1 | 99.6 |
| 7 | Mutism,Staring,Mannerisms,Verbigeration,Rigidity,Negativism,Waxy flexibility | 98.4 | 97.3 | 99.6 |
| 7 | Mutism,Staring,Posturing/catalepsy,Echopraxia/echolalia,Mannerisms,Negativism,Waxy flexibility | 98.4 | 97.1 | 99.6 |
| 7 | Mutism,Staring,Posturing/catalepsy,Echopraxia/echolalia,Mannerisms,Verbigeration,Rigidity | 98.4 | 97.1 | 99.6 |
| 7 | Mutism,Staring,Posturing/catalepsy,Echopraxia/echolalia,Rigidity,Negativism,Withdrawal | 98.4 | 97.3 | 99.6 |
| 7 | Mutism,Staring,Posturing/catalepsy,Echopraxia/echolalia,Stereotypy,Negativism,Withdrawal | 98.4 | 97.1 | 99.6 |
| 7 | Mutism,Staring,Posturing/catalepsy,Echopraxia/echolalia,Verbigeration,Rigidity,Negativism | 98.4 | 97.1 | 99.6 |
| 7 | Mutism,Staring,Posturing/catalepsy,Grimacing,Echopraxia/echolalia,Mannerisms,Withdrawal | 98.4 | 97.1 | 99.6 |
| 7 | Mutism,Staring,Posturing/catalepsy,Grimacing,Echopraxia/echolalia,Negativism,Withdrawal | 98.4 | 97.1 | 99.6 |
| 7 | Mutism,Staring,Posturing/catalepsy,Grimacing,Mannerisms,Rigidity,Negativism | 98.4 | 97.1 | 99.6 |
| 7 | Mutism,Staring,Posturing/catalepsy,Grimacing,Mannerisms,Verbigeration,Negativism | 98.4 | 97.1 | 99.6 |
| 7 | Mutism,Staring,Posturing/catalepsy,Grimacing,Stereotypy,Mannerisms,Withdrawal | 98.4 | 97.3 | 99.6 |
| 7 | Mutism,Staring,Posturing/catalepsy,Grimacing,Stereotypy,Negativism,Withdrawal | 98.4 | 97.1 | 99.6 |
| 7 | Mutism,Staring,Posturing/catalepsy,Stereotypy,Mannerisms,Negativism,Waxy flexibility | 98.4 | 97.3 | 99.6 |
| 7 | Mutism,Staring,Posturing/catalepsy,Stereotypy,Mannerisms,Verbigeration,Rigidity | 98.4 | 97.3 | 99.6 |
| 7 | Mutism,Staring,Posturing/catalepsy,Stereotypy,Rigidity,Negativism,Withdrawal | 98.4 | 97.3 | 99.6 |
| 7 | Mutism,Staring,Stereotypy,Verbigeration,Rigidity,Negativism,Waxy flexibility | 98.4 | 97.1 | 99.6 |
| 7 | Staring,Grimacing,Echopraxia/echolalia,Stereotypy,Mannerisms,Rigidity,Withdrawal | 98.4 | 97.1 | 99.6 |
| 7 | Staring,Grimacing,Stereotypy,Mannerisms,Rigidity,Waxy flexibility,Withdrawal | 98.4 | 97.3 | 99.6 |
| 7 | Staring,Grimacing,Stereotypy,Mannerisms,Verbigeration,Negativism,Withdrawal | 98.4 | 97.1 | 99.6 |
| 7 | Staring,Grimacing,Stereotypy,Mannerisms,Verbigeration,Rigidity,Withdrawal | 98.4 | 97.1 | 99.6 |
| 7 | Staring,Grimacing,Stereotypy,Verbigeration,Rigidity,Negativism,Withdrawal | 98.4 | 97.1 | 99.6 |
| 7 | Staring,Posturing/catalepsy,Echopraxia/echolalia,Stereotypy,Mannerisms,Negativism,Withdrawal | 98.4 | 97.3 | 99.6 |
| 7 | Staring,Posturing/catalepsy,Grimacing,Mannerisms,Verbigeration,Negativism,Withdrawal | 98.4 | 97.3 | 99.6 |
| 7 | Staring,Posturing/catalepsy,Grimacing,Stereotypy,Mannerisms,Rigidity,Withdrawal | 98.4 | 97.3 | 99.6 |
| 7 | Staring,Posturing/catalepsy,Stereotypy,Mannerisms,Verbigeration,Negativism,Withdrawal | 98.4 | 97.1 | 99.6 |
| 7 | Excitement,Immobility/stupor,Echopraxia/echolalia,Mannerisms,Verbigeration,Rigidity,Withdrawal | 98.2 | 96.9 | 99.3 |
| 7 | Excitement,Immobility/stupor,Echopraxia/echolalia,Stereotypy,Mannerisms,Rigidity,Withdrawal | 98.2 | 96.9 | 99.3 |
| 7 | Excitement,Immobility/stupor,Echopraxia/echolalia,Stereotypy,Rigidity,Waxy flexibility,Withdrawal | 98.2 | 96.9 | 99.3 |
| 7 | Excitement,Immobility/stupor,Echopraxia/echolalia,Stereotypy,Verbigeration,Rigidity,Negativism | 98.2 | 96.9 | 99.3 |
| 7 | Excitement,Immobility/stupor,Echopraxia/echolalia,Verbigeration,Rigidity,Waxy flexibility,Withdrawal | 98.2 | 96.9 | 99.3 |
| 7 | Excitement,Immobility/stupor,Grimacing,Echopraxia/echolalia,Mannerisms,Rigidity,Withdrawal | 98.2 | 96.9 | 99.3 |
| 7 | Excitement,Immobility/stupor,Grimacing,Echopraxia/echolalia,Rigidity,Negativism,Waxy flexibility | 98.2 | 96.9 | 99.3 |
| 7 | Excitement,Immobility/stupor,Grimacing,Echopraxia/echolalia,Rigidity,Waxy flexibility,Withdrawal | 98.2 | 96.9 | 99.3 |
| 7 | Excitement,Immobility/stupor,Grimacing,Echopraxia/echolalia,Stereotypy,Rigidity,Withdrawal | 98.2 | 96.9 | 99.3 |
| 7 | Excitement,Immobility/stupor,Grimacing,Echopraxia/echolalia,Verbigeration,Rigidity,Negativism | 98.2 | 96.9 | 99.3 |
| 7 | Excitement,Immobility/stupor,Grimacing,Echopraxia/echolalia,Verbigeration,Rigidity,Withdrawal | 98.2 | 96.9 | 99.3 |
| 7 | Excitement,Immobility/stupor,Grimacing,Mannerisms,Rigidity,Negativism,Withdrawal | 98.2 | 96.9 | 99.3 |
| 7 | Excitement,Immobility/stupor,Grimacing,Stereotypy,Rigidity,Negativism,Withdrawal | 98.2 | 96.9 | 99.3 |
| 7 | Excitement,Immobility/stupor,Grimacing,Verbigeration,Rigidity,Negativism,Withdrawal | 98.2 | 96.9 | 99.3 |
| 7 | Excitement,Immobility/stupor,Mutism,Echopraxia/echolalia,Mannerisms,Rigidity,Negativism | 98.2 | 96.9 | 99.3 |
| 7 | Excitement,Immobility/stupor,Mutism,Echopraxia/echolalia,Mannerisms,Rigidity,Withdrawal | 98.2 | 96.9 | 99.3 |
| 7 | Excitement,Immobility/stupor,Mutism,Echopraxia/echolalia,Stereotypy,Verbigeration,Rigidity | 98.2 | 96.9 | 99.3 |
| 7 | Excitement,Immobility/stupor,Mutism,Echopraxia/echolalia,Verbigeration,Rigidity,Waxy flexibility | 98.2 | 96.9 | 99.3 |
| 7 | Excitement,Immobility/stupor,Mutism,Grimacing,Rigidity,Negativism,Waxy flexibility | 98.2 | 96.9 | 99.3 |
| 7 | Excitement,Immobility/stupor,Mutism,Grimacing,Rigidity,Waxy flexibility,Withdrawal | 98.2 | 96.9 | 99.3 |
| 7 | Excitement,Immobility/stupor,Mutism,Grimacing,Stereotypy,Rigidity,Negativism | 98.2 | 96.9 | 99.3 |
| 7 | Excitement,Immobility/stupor,Mutism,Grimacing,Stereotypy,Rigidity,Withdrawal | 98.2 | 96.9 | 99.3 |
| 7 | Excitement,Immobility/stupor,Mutism,Mannerisms,Rigidity,Negativism,Withdrawal | 98.2 | 96.9 | 99.3 |
| 7 | Excitement,Immobility/stupor,Mutism,Posturing/catalepsy,Echopraxia/echolalia,Mannerisms,Rigidity | 98.2 | 96.9 | 99.3 |
| 7 | Excitement,Immobility/stupor,Mutism,Posturing/catalepsy,Echopraxia/echolalia,Rigidity,Waxy flexibility | 98.2 | 96.9 | 99.3 |
| 7 | Excitement,Immobility/stupor,Mutism,Posturing/catalepsy,Grimacing,Mannerisms,Rigidity | 98.2 | 96.9 | 99.3 |
| 7 | Excitement,Immobility/stupor,Mutism,Posturing/catalepsy,Grimacing,Negativism,Withdrawal | 98.2 | 96.9 | 99.3 |
| 7 | Excitement,Immobility/stupor,Mutism,Posturing/catalepsy,Grimacing,Stereotypy,Rigidity | 98.2 | 96.9 | 99.3 |
| 7 | Excitement,Immobility/stupor,Mutism,Posturing/catalepsy,Rigidity,Negativism,Waxy flexibility | 98.2 | 96.9 | 99.3 |
| 7 | Excitement,Immobility/stupor,Mutism,Posturing/catalepsy,Rigidity,Waxy flexibility,Withdrawal | 98.2 | 96.9 | 99.3 |
| 7 | Excitement,Immobility/stupor,Mutism,Posturing/catalepsy,Stereotypy,Negativism,Withdrawal | 98.2 | 96.9 | 99.3 |
| 7 | Excitement,Immobility/stupor,Mutism,Posturing/catalepsy,Verbigeration,Rigidity,Waxy flexibility | 98.2 | 96.9 | 99.3 |
| 7 | Excitement,Immobility/stupor,Mutism,Rigidity,Negativism,Waxy flexibility,Withdrawal | 98.2 | 96.9 | 99.3 |
| 7 | Excitement,Immobility/stupor,Mutism,Staring,Echopraxia/echolalia,Mannerisms,Waxy flexibility | 98.2 | 96.9 | 99.3 |
| 7 | Excitement,Immobility/stupor,Mutism,Staring,Grimacing,Echopraxia/echolalia,Withdrawal | 98.2 | 96.9 | 99.3 |
| 7 | Excitement,Immobility/stupor,Mutism,Staring,Grimacing,Negativism,Withdrawal | 98.2 | 96.9 | 99.3 |
| 7 | Excitement,Immobility/stupor,Mutism,Staring,Grimacing,Stereotypy,Negativism | 98.2 | 96.9 | 99.3 |
| 7 | Excitement,Immobility/stupor,Mutism,Staring,Grimacing,Verbigeration,Negativism | 98.2 | 96.9 | 99.3 |
| 7 | Excitement,Immobility/stupor,Mutism,Staring,Posturing/catalepsy,Grimacing,Mannerisms | 98.2 | 96.9 | 99.3 |
| 7 | Excitement,Immobility/stupor,Mutism,Staring,Posturing/catalepsy,Grimacing,Stereotypy | 98.2 | 96.9 | 99.3 |
| 7 | Excitement,Immobility/stupor,Mutism,Staring,Posturing/catalepsy,Grimacing,Waxy flexibility | 98.2 | 96.9 | 99.3 |
| 7 | Excitement,Immobility/stupor,Mutism,Staring,Posturing/catalepsy,Stereotypy,Verbigeration | 98.2 | 96.9 | 99.3 |
| 7 | Excitement,Immobility/stupor,Mutism,Staring,Stereotypy,Mannerisms,Negativism | 98.2 | 96.9 | 99.3 |
| 7 | Excitement,Immobility/stupor,Mutism,Staring,Stereotypy,Verbigeration,Waxy flexibility | 98.2 | 96.9 | 99.3 |
| 7 | Excitement,Immobility/stupor,Mutism,Staring,Stereotypy,Waxy flexibility,Withdrawal | 98.2 | 96.9 | 99.3 |
| 7 | Excitement,Immobility/stupor,Mutism,Stereotypy,Verbigeration,Rigidity,Waxy flexibility | 98.2 | 96.9 | 99.3 |
| 7 | Excitement,Immobility/stupor,Posturing/catalepsy,Echopraxia/echolalia,Rigidity,Waxy flexibility,Withdrawal | 98.2 | 96.9 | 99.3 |
| 7 | Excitement,Immobility/stupor,Posturing/catalepsy,Echopraxia/echolalia,Verbigeration,Negativism,Withdrawal | 98.2 | 96.9 | 99.3 |
| 7 | Excitement,Immobility/stupor,Posturing/catalepsy,Grimacing,Echopraxia/echolalia,Negativism,Withdrawal | 98.2 | 96.9 | 99.3 |
| 7 | Excitement,Immobility/stupor,Posturing/catalepsy,Grimacing,Mannerisms,Rigidity,Negativism | 98.2 | 96.9 | 99.3 |
| 7 | Excitement,Immobility/stupor,Posturing/catalepsy,Grimacing,Mannerisms,Rigidity,Withdrawal | 98.2 | 96.9 | 99.3 |
| 7 | Excitement,Immobility/stupor,Posturing/catalepsy,Grimacing,Verbigeration,Rigidity,Withdrawal | 98.2 | 96.9 | 99.3 |
| 7 | Excitement,Immobility/stupor,Posturing/catalepsy,Mannerisms,Verbigeration,Negativism,Withdrawal | 98.2 | 96.9 | 99.3 |
| 7 | Excitement,Immobility/stupor,Posturing/catalepsy,Rigidity,Negativism,Waxy flexibility,Withdrawal | 98.2 | 96.9 | 99.3 |
| 7 | Excitement,Immobility/stupor,Posturing/catalepsy,Stereotypy,Verbigeration,Negativism,Withdrawal | 98.2 | 96.9 | 99.3 |
| 7 | Excitement,Immobility/stupor,Posturing/catalepsy,Stereotypy,Verbigeration,Rigidity,Withdrawal | 98.2 | 96.9 | 99.3 |
| 7 | Excitement,Immobility/stupor,Posturing/catalepsy,Verbigeration,Rigidity,Negativism,Waxy flexibility | 98.2 | 96.9 | 99.3 |
| 7 | Excitement,Immobility/stupor,Staring,Echopraxia/echolalia,Mannerisms,Verbigeration,Negativism | 98.2 | 96.9 | 99.3 |
| 7 | Excitement,Immobility/stupor,Staring,Echopraxia/echolalia,Mannerisms,Waxy flexibility,Withdrawal | 98.2 | 96.9 | 99.3 |
| 7 | Excitement,Immobility/stupor,Staring,Grimacing,Echopraxia/echolalia,Stereotypy,Negativism | 98.2 | 96.9 | 99.3 |
| 7 | Excitement,Immobility/stupor,Staring,Grimacing,Stereotypy,Mannerisms,Rigidity | 98.2 | 96.9 | 99.3 |
| 7 | Excitement,Immobility/stupor,Staring,Grimacing,Stereotypy,Negativism,Withdrawal | 98.2 | 96.9 | 99.3 |
| 7 | Excitement,Immobility/stupor,Staring,Grimacing,Stereotypy,Verbigeration,Negativism | 98.2 | 96.9 | 99.3 |
| 7 | Excitement,Immobility/stupor,Staring,Grimacing,Verbigeration,Negativism,Waxy flexibility | 98.2 | 96.9 | 99.3 |
| 7 | Excitement,Immobility/stupor,Staring,Grimacing,Verbigeration,Negativism,Withdrawal | 98.2 | 96.9 | 99.3 |
| 7 | Excitement,Immobility/stupor,Staring,Posturing/catalepsy,Echopraxia/echolalia,Stereotypy,Withdrawal | 98.2 | 96.9 | 99.3 |
| 7 | Excitement,Immobility/stupor,Staring,Posturing/catalepsy,Echopraxia/echolalia,Verbigeration,Withdrawal | 98.2 | 96.9 | 99.3 |
| 7 | Excitement,Immobility/stupor,Staring,Posturing/catalepsy,Grimacing,Mannerisms,Withdrawal | 98.2 | 96.9 | 99.3 |
| 7 | Excitement,Immobility/stupor,Staring,Posturing/catalepsy,Grimacing,Rigidity,Waxy flexibility | 98.2 | 96.9 | 99.3 |
| 7 | Excitement,Immobility/stupor,Staring,Posturing/catalepsy,Mannerisms,Verbigeration,Waxy flexibility | 98.2 | 96.9 | 99.3 |
| 7 | Excitement,Immobility/stupor,Staring,Posturing/catalepsy,Mannerisms,Waxy flexibility,Withdrawal | 98.2 | 96.9 | 99.3 |
| 7 | Excitement,Immobility/stupor,Staring,Stereotypy,Mannerisms,Negativism,Waxy flexibility | 98.2 | 96.9 | 99.3 |
| 7 | Excitement,Immobility/stupor,Stereotypy,Mannerisms,Rigidity,Negativism,Withdrawal | 98.2 | 96.9 | 99.3 |
| 7 | Excitement,Immobility/stupor,Stereotypy,Rigidity,Negativism,Waxy flexibility,Withdrawal | 98.2 | 96.9 | 99.3 |
| 7 | Excitement,Immobility/stupor,Stereotypy,Verbigeration,Rigidity,Negativism,Waxy flexibility | 98.2 | 96.9 | 99.3 |
| 7 | Excitement,Mutism,Staring,Echopraxia/echolalia,Stereotypy,Mannerisms,Waxy flexibility | 98.2 | 96.9 | 99.3 |
| 7 | Excitement,Mutism,Staring,Grimacing,Echopraxia/echolalia,Mannerisms,Waxy flexibility | 98.2 | 96.9 | 99.3 |
| 7 | Excitement,Mutism,Staring,Grimacing,Echopraxia/echolalia,Mannerisms,Withdrawal | 98.2 | 96.9 | 99.3 |
| 7 | Excitement,Mutism,Staring,Grimacing,Echopraxia/echolalia,Verbigeration,Withdrawal | 98.2 | 96.9 | 99.3 |
| 7 | Excitement,Mutism,Staring,Grimacing,Mannerisms,Verbigeration,Negativism | 98.2 | 96.9 | 99.3 |
| 7 | Excitement,Mutism,Staring,Grimacing,Mannerisms,Waxy flexibility,Withdrawal | 98.2 | 96.9 | 99.3 |
| 7 | Excitement,Mutism,Staring,Grimacing,Stereotypy,Verbigeration,Waxy flexibility | 98.2 | 96.9 | 99.3 |
| 7 | Excitement,Mutism,Staring,Grimacing,Stereotypy,Verbigeration,Withdrawal | 98.2 | 96.9 | 99.3 |
| 7 | Excitement,Mutism,Staring,Grimacing,Verbigeration,Waxy flexibility,Withdrawal | 98.2 | 96.9 | 99.3 |
| 7 | Excitement,Mutism,Staring,Posturing/catalepsy,Grimacing,Mannerisms,Verbigeration | 98.2 | 96.9 | 99.3 |
| 7 | Excitement,Mutism,Staring,Posturing/catalepsy,Grimacing,Mannerisms,Waxy flexibility | 98.2 | 96.9 | 99.3 |
| 7 | Excitement,Mutism,Staring,Posturing/catalepsy,Stereotypy,Mannerisms,Verbigeration | 98.2 | 96.9 | 99.3 |
| 7 | Excitement,Mutism,Staring,Posturing/catalepsy,Stereotypy,Mannerisms,Waxy flexibility | 98.2 | 96.9 | 99.3 |
| 7 | Excitement,Mutism,Staring,Stereotypy,Mannerisms,Waxy flexibility,Withdrawal | 98.2 | 96.9 | 99.3 |
| 7 | Excitement,Staring,Echopraxia/echolalia,Mannerisms,Verbigeration,Rigidity,Withdrawal | 98.2 | 96.9 | 99.3 |
| 7 | Excitement,Staring,Echopraxia/echolalia,Stereotypy,Negativism,Waxy flexibility,Withdrawal | 98.2 | 96.9 | 99.3 |
| 7 | Excitement,Staring,Echopraxia/echolalia,Verbigeration,Rigidity,Waxy flexibility,Withdrawal | 98.2 | 96.9 | 99.3 |
| 7 | Excitement,Staring,Grimacing,Echopraxia/echolalia,Stereotypy,Negativism,Withdrawal | 98.2 | 96.9 | 99.3 |
| 7 | Excitement,Staring,Grimacing,Echopraxia/echolalia,Verbigeration,Rigidity,Withdrawal | 98.2 | 96.9 | 99.3 |
| 7 | Excitement,Staring,Mannerisms,Verbigeration,Negativism,Waxy flexibility,Withdrawal | 98.2 | 96.9 | 99.3 |
| 7 | Excitement,Staring,Posturing/catalepsy,Echopraxia/echolalia,Mannerisms,Rigidity,Negativism | 98.2 | 96.9 | 99.3 |
| 7 | Excitement,Staring,Posturing/catalepsy,Grimacing,Echopraxia/echolalia,Mannerisms,Withdrawal | 98.2 | 96.9 | 99.3 |
| 7 | Excitement,Staring,Posturing/catalepsy,Grimacing,Stereotypy,Negativism,Withdrawal | 98.2 | 96.9 | 99.3 |
| 7 | Excitement,Staring,Posturing/catalepsy,Mannerisms,Verbigeration,Rigidity,Negativism | 98.2 | 96.9 | 99.3 |
| 7 | Excitement,Staring,Posturing/catalepsy,Stereotypy,Negativism,Waxy flexibility,Withdrawal | 98.2 | 96.9 | 99.3 |
| 7 | Excitement,Staring,Stereotypy,Mannerisms,Verbigeration,Negativism,Withdrawal | 98.2 | 96.9 | 99.3 |
| 7 | Immobility/stupor,Grimacing,Echopraxia/echolalia,Mannerisms,Rigidity,Negativism,Withdrawal | 98.2 | 96.9 | 99.3 |
| 7 | Immobility/stupor,Mutism,Staring,Echopraxia/echolalia,Stereotypy,Mannerisms,Negativism | 98.2 | 96.9 | 99.3 |
| 7 | Immobility/stupor,Mutism,Staring,Mannerisms,Rigidity,Negativism,Waxy flexibility | 98.2 | 96.9 | 99.3 |
| 7 | Immobility/stupor,Mutism,Staring,Posturing/catalepsy,Echopraxia/echolalia,Mannerisms,Negativism | 98.2 | 96.9 | 99.3 |
| 7 | Immobility/stupor,Mutism,Staring,Posturing/catalepsy,Mannerisms,Rigidity,Withdrawal | 98.2 | 96.9 | 99.3 |
| 7 | Immobility/stupor,Mutism,Staring,Stereotypy,Mannerisms,Rigidity,Withdrawal | 98.2 | 96.9 | 99.3 |
| 7 | Immobility/stupor,Mutism,Stereotypy,Mannerisms,Verbigeration,Rigidity,Negativism | 98.2 | 96.9 | 99.3 |
| 7 | Immobility/stupor,Posturing/catalepsy,Echopraxia/echolalia,Mannerisms,Rigidity,Negativism,Withdrawal | 98.2 | 96.9 | 99.3 |
| 7 | Immobility/stupor,Posturing/catalepsy,Grimacing,Mannerisms,Rigidity,Negativism,Withdrawal | 98.2 | 96.9 | 99.3 |
| 7 | Immobility/stupor,Posturing/catalepsy,Mannerisms,Verbigeration,Rigidity,Negativism,Withdrawal | 98.2 | 96.9 | 99.3 |
| 7 | Immobility/stupor,Posturing/catalepsy,Stereotypy,Mannerisms,Rigidity,Negativism,Withdrawal | 98.2 | 96.9 | 99.3 |
| 7 | Immobility/stupor,Staring,Echopraxia/echolalia,Mannerisms,Negativism,Waxy flexibility,Withdrawal | 98.2 | 96.9 | 99.3 |
| 7 | Immobility/stupor,Staring,Echopraxia/echolalia,Stereotypy,Mannerisms,Negativism,Waxy flexibility | 98.2 | 96.9 | 99.3 |
| 7 | Immobility/stupor,Staring,Echopraxia/echolalia,Verbigeration,Rigidity,Negativism,Withdrawal | 98.2 | 96.9 | 99.3 |
| 7 | Immobility/stupor,Staring,Grimacing,Echopraxia/echolalia,Stereotypy,Mannerisms,Negativism | 98.2 | 96.9 | 99.3 |
| 7 | Immobility/stupor,Staring,Grimacing,Mannerisms,Rigidity,Negativism,Waxy flexibility | 98.2 | 96.9 | 99.3 |
| 7 | Immobility/stupor,Staring,Posturing/catalepsy,Echopraxia/echolalia,Rigidity,Negativism,Withdrawal | 98.2 | 96.9 | 99.3 |
| 7 | Immobility/stupor,Staring,Posturing/catalepsy,Grimacing,Echopraxia/echolalia,Mannerisms,Negativism | 98.2 | 96.9 | 99.3 |
| 7 | Immobility/stupor,Staring,Posturing/catalepsy,Grimacing,Stereotypy,Mannerisms,Negativism | 98.2 | 96.9 | 99.3 |
| 7 | Immobility/stupor,Staring,Posturing/catalepsy,Mannerisms,Negativism,Waxy flexibility,Withdrawal | 98.2 | 96.9 | 99.3 |
| 7 | Immobility/stupor,Staring,Posturing/catalepsy,Mannerisms,Rigidity,Negativism,Waxy flexibility | 98.2 | 96.9 | 99.3 |
| 7 | Immobility/stupor,Staring,Posturing/catalepsy,Stereotypy,Mannerisms,Rigidity,Withdrawal | 98.2 | 96.9 | 99.3 |
| 7 | Immobility/stupor,Staring,Posturing/catalepsy,Stereotypy,Rigidity,Negativism,Withdrawal | 98.2 | 96.9 | 99.3 |
| 7 | Immobility/stupor,Staring,Posturing/catalepsy,Stereotypy,Verbigeration,Negativism,Withdrawal | 98.2 | 96.9 | 99.3 |
| 7 | Immobility/stupor,Staring,Stereotypy,Mannerisms,Negativism,Waxy flexibility,Withdrawal | 98.2 | 96.9 | 99.3 |
| 7 | Immobility/stupor,Staring,Stereotypy,Verbigeration,Rigidity,Negativism,Waxy flexibility | 98.2 | 96.9 | 99.3 |
| 7 | Mutism,Staring,Echopraxia/echolalia,Mannerisms,Verbigeration,Negativism,Waxy flexibility | 98.2 | 96.9 | 99.3 |
| 7 | Mutism,Staring,Grimacing,Echopraxia/echolalia,Mannerisms,Verbigeration,Negativism | 98.2 | 96.9 | 99.3 |
| 7 | Mutism,Staring,Grimacing,Echopraxia/echolalia,Mannerisms,Verbigeration,Rigidity | 98.2 | 96.9 | 99.3 |
| 7 | Mutism,Staring,Grimacing,Echopraxia/echolalia,Rigidity,Negativism,Withdrawal | 98.2 | 96.9 | 99.3 |
| 7 | Mutism,Staring,Grimacing,Echopraxia/echolalia,Stereotypy,Mannerisms,Rigidity | 98.2 | 96.9 | 99.3 |
| 7 | Mutism,Staring,Grimacing,Echopraxia/echolalia,Stereotypy,Mannerisms,Withdrawal | 98.2 | 96.9 | 99.3 |
| 7 | Mutism,Staring,Grimacing,Echopraxia/echolalia,Stereotypy,Negativism,Withdrawal | 98.2 | 96.9 | 99.3 |
| 7 | Mutism,Staring,Grimacing,Echopraxia/echolalia,Stereotypy,Rigidity,Withdrawal | 98.2 | 96.9 | 99.3 |
| 7 | Mutism,Staring,Grimacing,Echopraxia/echolalia,Verbigeration,Rigidity,Negativism | 98.2 | 96.9 | 99.3 |
| 7 | Mutism,Staring,Grimacing,Mannerisms,Rigidity,Negativism,Waxy flexibility | 98.2 | 96.9 | 99.3 |
| 7 | Mutism,Staring,Grimacing,Stereotypy,Mannerisms,Negativism,Waxy flexibility | 98.2 | 96.9 | 99.3 |
| 7 | Mutism,Staring,Grimacing,Stereotypy,Mannerisms,Rigidity,Waxy flexibility | 98.2 | 96.9 | 99.3 |
| 7 | Mutism,Staring,Grimacing,Stereotypy,Mannerisms,Verbigeration,Negativism | 98.2 | 96.9 | 99.3 |
| 7 | Mutism,Staring,Grimacing,Stereotypy,Rigidity,Negativism,Withdrawal | 98.2 | 96.9 | 99.3 |
| 7 | Mutism,Staring,Grimacing,Stereotypy,Verbigeration,Rigidity,Waxy flexibility | 98.2 | 96.9 | 99.3 |
| 7 | Mutism,Staring,Grimacing,Verbigeration,Rigidity,Negativism,Withdrawal | 98.2 | 96.9 | 99.3 |
| 7 | Mutism,Staring,Mannerisms,Verbigeration,Negativism,Waxy flexibility,Withdrawal | 98.2 | 96.9 | 99.3 |
| 7 | Mutism,Staring,Posturing/catalepsy,Echopraxia/echolalia,Stereotypy,Mannerisms,Rigidity | 98.2 | 96.9 | 99.3 |
| 7 | Mutism,Staring,Posturing/catalepsy,Echopraxia/echolalia,Stereotypy,Rigidity,Negativism | 98.2 | 96.9 | 99.3 |
| 7 | Mutism,Staring,Posturing/catalepsy,Echopraxia/echolalia,Stereotypy,Rigidity,Withdrawal | 98.2 | 96.9 | 99.3 |
| 7 | Mutism,Staring,Posturing/catalepsy,Echopraxia/echolalia,Stereotypy,Verbigeration,Negativism | 98.2 | 96.9 | 99.3 |
| 7 | Mutism,Staring,Posturing/catalepsy,Echopraxia/echolalia,Verbigeration,Negativism,Withdrawal | 98.2 | 96.9 | 99.3 |
| 7 | Mutism,Staring,Posturing/catalepsy,Grimacing,Echopraxia/echolalia,Mannerisms,Rigidity | 98.2 | 96.9 | 99.3 |
| 7 | Mutism,Staring,Posturing/catalepsy,Grimacing,Echopraxia/echolalia,Verbigeration,Negativism | 98.2 | 96.9 | 99.3 |
| 7 | Mutism,Staring,Posturing/catalepsy,Grimacing,Mannerisms,Verbigeration,Rigidity | 98.2 | 96.9 | 99.3 |
| 7 | Mutism,Staring,Posturing/catalepsy,Grimacing,Mannerisms,Verbigeration,Withdrawal | 98.2 | 96.9 | 99.3 |
| 7 | Mutism,Staring,Posturing/catalepsy,Grimacing,Stereotypy,Mannerisms,Rigidity | 98.2 | 96.9 | 99.3 |
| 7 | Mutism,Staring,Posturing/catalepsy,Grimacing,Stereotypy,Verbigeration,Negativism | 98.2 | 96.9 | 99.3 |
| 7 | Mutism,Staring,Posturing/catalepsy,Grimacing,Stereotypy,Verbigeration,Rigidity | 98.2 | 96.9 | 99.3 |
| 7 | Mutism,Staring,Posturing/catalepsy,Grimacing,Verbigeration,Negativism,Withdrawal | 98.2 | 96.9 | 99.3 |
| 7 | Mutism,Staring,Posturing/catalepsy,Mannerisms,Rigidity,Negativism,Waxy flexibility | 98.2 | 96.9 | 99.3 |
| 7 | Mutism,Staring,Posturing/catalepsy,Mannerisms,Rigidity,Waxy flexibility,Withdrawal | 98.2 | 96.9 | 99.3 |
| 7 | Mutism,Staring,Posturing/catalepsy,Stereotypy,Verbigeration,Rigidity,Withdrawal | 98.2 | 96.9 | 99.3 |
| 7 | Mutism,Staring,Posturing/catalepsy,Verbigeration,Rigidity,Negativism,Withdrawal | 98.2 | 96.9 | 99.3 |
| 7 | Mutism,Staring,Stereotypy,Mannerisms,Verbigeration,Negativism,Waxy flexibility | 98.2 | 96.9 | 99.3 |
| 7 | Mutism,Staring,Stereotypy,Rigidity,Negativism,Waxy flexibility,Withdrawal | 98.2 | 96.9 | 99.3 |
| 7 | Mutism,Staring,Stereotypy,Verbigeration,Rigidity,Waxy flexibility,Withdrawal | 98.2 | 96.9 | 99.3 |
| 7 | Staring,Echopraxia/echolalia,Stereotypy,Mannerisms,Negativism,Waxy flexibility,Withdrawal | 98.2 | 96.9 | 99.3 |
| 7 | Staring,Echopraxia/echolalia,Stereotypy,Verbigeration,Rigidity,Negativism,Withdrawal | 98.2 | 96.9 | 99.3 |
| 7 | Staring,Posturing/catalepsy,Echopraxia/echolalia,Mannerisms,Negativism,Waxy flexibility,Withdrawal | 98.2 | 96.9 | 99.3 |
| 7 | Staring,Posturing/catalepsy,Stereotypy,Mannerisms,Negativism,Waxy flexibility,Withdrawal | 98.2 | 96.9 | 99.3 |
| 7 | Staring,Posturing/catalepsy,Stereotypy,Verbigeration,Rigidity,Negativism,Withdrawal | 98.2 | 96.9 | 99.3 |
| 7 | Staring,Stereotypy,Verbigeration,Rigidity,Negativism,Waxy flexibility,Withdrawal | 98.2 | 96.9 | 99.3 |
| 7 | Excitement,Immobility/stupor,Echopraxia/echolalia,Mannerisms,Rigidity,Waxy flexibility,Withdrawal | 98.0 | 96.6 | 99.1 |
| 7 | Excitement,Immobility/stupor,Echopraxia/echolalia,Stereotypy,Rigidity,Negativism,Waxy flexibility | 98.0 | 96.6 | 99.1 |
| 7 | Excitement,Immobility/stupor,Echopraxia/echolalia,Verbigeration,Rigidity,Negativism,Waxy flexibility | 98.0 | 96.6 | 99.1 |
| 7 | Excitement,Immobility/stupor,Grimacing,Echopraxia/echolalia,Mannerisms,Rigidity,Negativism | 98.0 | 96.6 | 99.1 |
| 7 | Excitement,Immobility/stupor,Grimacing,Echopraxia/echolalia,Stereotypy,Rigidity,Negativism | 98.0 | 96.6 | 99.1 |
| 7 | Excitement,Immobility/stupor,Grimacing,Mannerisms,Verbigeration,Rigidity,Negativism | 98.0 | 96.6 | 99.1 |
| 7 | Excitement,Immobility/stupor,Grimacing,Stereotypy,Verbigeration,Rigidity,Negativism | 98.0 | 96.6 | 99.1 |
| 7 | Excitement,Immobility/stupor,Mannerisms,Rigidity,Negativism,Waxy flexibility,Withdrawal | 98.0 | 96.6 | 99.1 |
| 7 | Excitement,Immobility/stupor,Mutism,Echopraxia/echolalia,Stereotypy,Rigidity,Waxy flexibility | 98.0 | 96.6 | 99.1 |
| 7 | Excitement,Immobility/stupor,Mutism,Grimacing,Echopraxia/echolalia,Rigidity,Waxy flexibility | 98.0 | 96.6 | 99.1 |
| 7 | Excitement,Immobility/stupor,Mutism,Grimacing,Echopraxia/echolalia,Verbigeration,Rigidity | 98.0 | 96.6 | 99.1 |
| 7 | Excitement,Immobility/stupor,Mutism,Grimacing,Mannerisms,Rigidity,Negativism | 98.0 | 96.6 | 99.1 |
| 7 | Excitement,Immobility/stupor,Mutism,Grimacing,Mannerisms,Rigidity,Withdrawal | 98.0 | 96.6 | 99.1 |
| 7 | Excitement,Immobility/stupor,Mutism,Grimacing,Stereotypy,Verbigeration,Rigidity | 98.0 | 96.6 | 99.1 |
| 7 | Excitement,Immobility/stupor,Mutism,Grimacing,Verbigeration,Rigidity,Waxy flexibility | 98.0 | 96.6 | 99.1 |
| 7 | Excitement,Immobility/stupor,Mutism,Mannerisms,Rigidity,Negativism,Waxy flexibility | 98.0 | 96.6 | 99.1 |
| 7 | Excitement,Immobility/stupor,Mutism,Mannerisms,Rigidity,Waxy flexibility,Withdrawal | 98.0 | 96.6 | 99.1 |
| 7 | Excitement,Immobility/stupor,Mutism,Posturing/catalepsy,Echopraxia/echolalia,Verbigeration,Negativism | 98.0 | 96.6 | 99.1 |
| 7 | Excitement,Immobility/stupor,Mutism,Posturing/catalepsy,Grimacing,Echopraxia/echolalia,Negativism | 98.0 | 96.6 | 99.1 |
| 7 | Excitement,Immobility/stupor,Mutism,Posturing/catalepsy,Grimacing,Verbigeration,Negativism | 98.0 | 96.6 | 99.1 |
| 7 | Excitement,Immobility/stupor,Mutism,Posturing/catalepsy,Mannerisms,Negativism,Withdrawal | 98.0 | 96.6 | 99.1 |
| 7 | Excitement,Immobility/stupor,Mutism,Posturing/catalepsy,Stereotypy,Verbigeration,Negativism | 98.0 | 96.6 | 99.1 |
| 7 | Excitement,Immobility/stupor,Mutism,Staring,Echopraxia/echolalia,Mannerisms,Withdrawal | 98.0 | 96.6 | 99.1 |
| 7 | Excitement,Immobility/stupor,Mutism,Staring,Echopraxia/echolalia,Verbigeration,Withdrawal | 98.0 | 96.6 | 99.1 |
| 7 | Excitement,Immobility/stupor,Mutism,Staring,Grimacing,Stereotypy,Waxy flexibility | 98.0 | 96.6 | 99.1 |
| 7 | Excitement,Immobility/stupor,Mutism,Staring,Grimacing,Verbigeration,Waxy flexibility | 98.0 | 96.6 | 99.1 |
| 7 | Excitement,Immobility/stupor,Mutism,Staring,Grimacing,Waxy flexibility,Withdrawal | 98.0 | 96.6 | 99.1 |
| 7 | Excitement,Immobility/stupor,Mutism,Staring,Mannerisms,Verbigeration,Waxy flexibility | 98.0 | 96.6 | 99.1 |
| 7 | Excitement,Immobility/stupor,Mutism,Staring,Mannerisms,Waxy flexibility,Withdrawal | 98.0 | 96.6 | 99.1 |
| 7 | Excitement,Immobility/stupor,Mutism,Staring,Posturing/catalepsy,Stereotypy,Mannerisms | 98.0 | 96.6 | 99.1 |
| 7 | Excitement,Immobility/stupor,Mutism,Staring,Verbigeration,Waxy flexibility,Withdrawal | 98.0 | 96.6 | 99.1 |
| 7 | Excitement,Immobility/stupor,Mutism,Stereotypy,Mannerisms,Rigidity,Negativism | 98.0 | 96.6 | 99.1 |
| 7 | Excitement,Immobility/stupor,Mutism,Stereotypy,Mannerisms,Rigidity,Withdrawal | 98.0 | 96.6 | 99.1 |
| 7 | Excitement,Immobility/stupor,Mutism,Stereotypy,Verbigeration,Negativism,Withdrawal | 98.0 | 96.6 | 99.1 |
| 7 | Excitement,Immobility/stupor,Posturing/catalepsy,Echopraxia/echolalia,Mannerisms,Negativism,Withdrawal | 98.0 | 96.6 | 99.1 |
| 7 | Excitement,Immobility/stupor,Posturing/catalepsy,Echopraxia/echolalia,Mannerisms,Rigidity,Negativism | 98.0 | 96.6 | 99.1 |
| 7 | Excitement,Immobility/stupor,Posturing/catalepsy,Echopraxia/echolalia,Rigidity,Negativism,Waxy flexibility | 98.0 | 96.6 | 99.1 |
| 7 | Excitement,Immobility/stupor,Posturing/catalepsy,Echopraxia/echolalia,Stereotypy,Negativism,Withdrawal | 98.0 | 96.6 | 99.1 |
| 7 | Excitement,Immobility/stupor,Posturing/catalepsy,Echopraxia/echolalia,Stereotypy,Rigidity,Negativism | 98.0 | 96.6 | 99.1 |
| 7 | Excitement,Immobility/stupor,Posturing/catalepsy,Grimacing,Rigidity,Negativism,Waxy flexibility | 98.0 | 96.6 | 99.1 |
| 7 | Excitement,Immobility/stupor,Posturing/catalepsy,Grimacing,Stereotypy,Rigidity,Negativism | 98.0 | 96.6 | 99.1 |
| 7 | Excitement,Immobility/stupor,Posturing/catalepsy,Grimacing,Stereotypy,Rigidity,Withdrawal | 98.0 | 96.6 | 99.1 |
| 7 | Excitement,Immobility/stupor,Posturing/catalepsy,Grimacing,Verbigeration,Negativism,Withdrawal | 98.0 | 96.6 | 99.1 |
| 7 | Excitement,Immobility/stupor,Posturing/catalepsy,Stereotypy,Mannerisms,Rigidity,Withdrawal | 98.0 | 96.6 | 99.1 |
| 7 | Excitement,Immobility/stupor,Posturing/catalepsy,Verbigeration,Rigidity,Waxy flexibility,Withdrawal | 98.0 | 96.6 | 99.1 |
| 7 | Excitement,Immobility/stupor,Staring,Echopraxia/echolalia,Stereotypy,Mannerisms,Withdrawal | 98.0 | 96.6 | 99.1 |
| 7 | Excitement,Immobility/stupor,Staring,Echopraxia/echolalia,Stereotypy,Waxy flexibility,Withdrawal | 98.0 | 96.6 | 99.1 |
| 7 | Excitement,Immobility/stupor,Staring,Grimacing,Echopraxia/echolalia,Mannerisms,Negativism | 98.0 | 96.6 | 99.1 |
| 7 | Excitement,Immobility/stupor,Staring,Grimacing,Echopraxia/echolalia,Verbigeration,Negativism | 98.0 | 96.6 | 99.1 |
| 7 | Excitement,Immobility/stupor,Staring,Grimacing,Echopraxia/echolalia,Waxy flexibility,Withdrawal | 98.0 | 96.6 | 99.1 |
| 7 | Excitement,Immobility/stupor,Staring,Grimacing,Mannerisms,Negativism,Waxy flexibility | 98.0 | 96.6 | 99.1 |
| 7 | Excitement,Immobility/stupor,Staring,Grimacing,Mannerisms,Negativism,Withdrawal | 98.0 | 96.6 | 99.1 |
| 7 | Excitement,Immobility/stupor,Staring,Grimacing,Mannerisms,Verbigeration,Negativism | 98.0 | 96.6 | 99.1 |
| 7 | Excitement,Immobility/stupor,Staring,Grimacing,Negativism,Waxy flexibility,Withdrawal | 98.0 | 96.6 | 99.1 |
| 7 | Excitement,Immobility/stupor,Staring,Grimacing,Stereotypy,Negativism,Waxy flexibility | 98.0 | 96.6 | 99.1 |
| 7 | Excitement,Immobility/stupor,Staring,Posturing/catalepsy,Echopraxia/echolalia,Mannerisms,Verbigeration | 98.0 | 96.6 | 99.1 |
| 7 | Excitement,Immobility/stupor,Staring,Posturing/catalepsy,Echopraxia/echolalia,Mannerisms,Waxy flexibility | 98.0 | 96.6 | 99.1 |
| 7 | Excitement,Immobility/stupor,Staring,Posturing/catalepsy,Echopraxia/echolalia,Verbigeration,Waxy flexibility | 98.0 | 96.6 | 99.1 |
| 7 | Excitement,Immobility/stupor,Staring,Posturing/catalepsy,Grimacing,Echopraxia/echolalia,Mannerisms | 98.0 | 96.6 | 99.1 |
| 7 | Excitement,Immobility/stupor,Staring,Posturing/catalepsy,Grimacing,Echopraxia/echolalia,Waxy flexibility | 98.0 | 96.6 | 99.1 |
| 7 | Excitement,Immobility/stupor,Staring,Posturing/catalepsy,Grimacing,Mannerisms,Verbigeration | 98.0 | 96.6 | 99.1 |
| 7 | Excitement,Immobility/stupor,Staring,Posturing/catalepsy,Grimacing,Verbigeration,Withdrawal | 98.0 | 96.6 | 99.1 |
| 7 | Excitement,Immobility/stupor,Staring,Posturing/catalepsy,Stereotypy,Mannerisms,Withdrawal | 98.0 | 96.6 | 99.1 |
| 7 | Excitement,Immobility/stupor,Staring,Posturing/catalepsy,Verbigeration,Waxy flexibility,Withdrawal | 98.0 | 96.6 | 99.1 |
| 7 | Excitement,Immobility/stupor,Stereotypy,Mannerisms,Verbigeration,Rigidity,Negativism | 98.0 | 96.6 | 99.1 |
| 7 | Excitement,Immobility/stupor,Stereotypy,Mannerisms,Verbigeration,Rigidity,Withdrawal | 98.0 | 96.6 | 99.1 |
| 7 | Excitement,Immobility/stupor,Stereotypy,Verbigeration,Rigidity,Waxy flexibility,Withdrawal | 98.0 | 96.6 | 99.1 |
| 7 | Excitement,Immobility/stupor,Verbigeration,Rigidity,Negativism,Waxy flexibility,Withdrawal | 98.0 | 96.6 | 99.1 |
| 7 | Excitement,Mutism,Staring,Grimacing,Echopraxia/echolalia,Stereotypy,Verbigeration | 98.0 | 96.6 | 99.1 |
| 7 | Excitement,Mutism,Staring,Grimacing,Stereotypy,Mannerisms,Negativism | 98.0 | 96.6 | 99.1 |
| 7 | Excitement,Mutism,Staring,Mannerisms,Verbigeration,Waxy flexibility,Withdrawal | 98.0 | 96.6 | 99.1 |
| 7 | Excitement,Mutism,Staring,Posturing/catalepsy,Grimacing,Stereotypy,Mannerisms | 98.0 | 96.6 | 99.1 |
| 7 | Excitement,Mutism,Staring,Stereotypy,Mannerisms,Verbigeration,Withdrawal | 98.0 | 96.6 | 99.1 |
| 7 | Excitement,Staring,Echopraxia/echolalia,Mannerisms,Rigidity,Negativism,Waxy flexibility | 98.0 | 96.6 | 99.1 |
| 7 | Excitement,Staring,Echopraxia/echolalia,Stereotypy,Mannerisms,Rigidity,Negativism | 98.0 | 96.6 | 99.1 |
| 7 | Excitement,Staring,Echopraxia/echolalia,Stereotypy,Rigidity,Negativism,Waxy flexibility | 98.0 | 96.6 | 99.1 |
| 7 | Excitement,Staring,Echopraxia/echolalia,Stereotypy,Verbigeration,Negativism,Withdrawal | 98.0 | 96.6 | 99.1 |
| 7 | Excitement,Staring,Echopraxia/echolalia,Verbigeration,Negativism,Waxy flexibility,Withdrawal | 98.0 | 96.6 | 99.1 |
| 7 | Excitement,Staring,Grimacing,Echopraxia/echolalia,Mannerisms,Negativism,Withdrawal | 98.0 | 96.6 | 99.1 |
| 7 | Excitement,Staring,Grimacing,Echopraxia/echolalia,Negativism,Waxy flexibility,Withdrawal | 98.0 | 96.6 | 99.1 |
| 7 | Excitement,Staring,Grimacing,Stereotypy,Verbigeration,Negativism,Withdrawal | 98.0 | 96.6 | 99.1 |
| 7 | Excitement,Staring,Mannerisms,Verbigeration,Rigidity,Negativism,Waxy flexibility | 98.0 | 96.6 | 99.1 |
| 7 | Excitement,Staring,Posturing/catalepsy,Echopraxia/echolalia,Mannerisms,Waxy flexibility,Withdrawal | 98.0 | 96.6 | 99.1 |
| 7 | Excitement,Staring,Posturing/catalepsy,Echopraxia/echolalia,Stereotypy,Rigidity,Negativism | 98.0 | 96.6 | 99.1 |
| 7 | Excitement,Staring,Posturing/catalepsy,Echopraxia/echolalia,Verbigeration,Rigidity,Negativism | 98.0 | 96.6 | 99.1 |
| 7 | Excitement,Staring,Posturing/catalepsy,Grimacing,Mannerisms,Verbigeration,Withdrawal | 98.0 | 96.6 | 99.1 |
| 7 | Excitement,Staring,Posturing/catalepsy,Grimacing,Negativism,Waxy flexibility,Withdrawal | 98.0 | 96.6 | 99.1 |
| 7 | Excitement,Staring,Posturing/catalepsy,Stereotypy,Mannerisms,Rigidity,Negativism | 98.0 | 96.6 | 99.1 |
| 7 | Excitement,Staring,Posturing/catalepsy,Stereotypy,Verbigeration,Rigidity,Negativism | 98.0 | 96.6 | 99.1 |
| 7 | Excitement,Staring,Stereotypy,Mannerisms,Negativism,Waxy flexibility,Withdrawal | 98.0 | 96.6 | 99.1 |
| 7 | Excitement,Staring,Stereotypy,Mannerisms,Rigidity,Negativism,Waxy flexibility | 98.0 | 96.6 | 99.1 |
| 7 | Excitement,Staring,Stereotypy,Mannerisms,Verbigeration,Rigidity,Negativism | 98.0 | 96.6 | 99.1 |
| 7 | Excitement,Staring,Stereotypy,Verbigeration,Negativism,Waxy flexibility,Withdrawal | 98.0 | 96.6 | 99.1 |
| 7 | Excitement,Staring,Stereotypy,Verbigeration,Rigidity,Negativism,Waxy flexibility | 98.0 | 96.6 | 99.1 |
| 7 | Immobility/stupor,Echopraxia/echolalia,Mannerisms,Verbigeration,Rigidity,Negativism,Withdrawal | 98.0 | 96.6 | 99.1 |
| 7 | Immobility/stupor,Grimacing,Stereotypy,Mannerisms,Rigidity,Negativism,Withdrawal | 98.0 | 96.6 | 99.1 |
| 7 | Immobility/stupor,Mutism,Echopraxia/echolalia,Mannerisms,Rigidity,Negativism,Withdrawal | 98.0 | 96.6 | 99.1 |
| 7 | Immobility/stupor,Mutism,Mannerisms,Verbigeration,Rigidity,Negativism,Withdrawal | 98.0 | 96.6 | 99.1 |
| 7 | Immobility/stupor,Mutism,Staring,Echopraxia/echolalia,Mannerisms,Rigidity,Withdrawal | 98.0 | 96.6 | 99.1 |
| 7 | Immobility/stupor,Mutism,Staring,Echopraxia/echolalia,Rigidity,Negativism,Withdrawal | 98.0 | 96.6 | 99.1 |
| 7 | Immobility/stupor,Mutism,Staring,Grimacing,Mannerisms,Negativism,Withdrawal | 98.0 | 96.6 | 99.1 |
| 7 | Immobility/stupor,Mutism,Staring,Grimacing,Mannerisms,Rigidity,Negativism | 98.0 | 96.6 | 99.1 |
| 7 | Immobility/stupor,Mutism,Staring,Grimacing,Mannerisms,Rigidity,Withdrawal | 98.0 | 96.6 | 99.1 |
| 7 | Immobility/stupor,Mutism,Staring,Grimacing,Stereotypy,Verbigeration,Rigidity | 98.0 | 96.6 | 99.1 |
| 7 | Immobility/stupor,Mutism,Staring,Mannerisms,Negativism,Waxy flexibility,Withdrawal | 98.0 | 96.6 | 99.1 |
| 7 | Immobility/stupor,Mutism,Staring,Mannerisms,Verbigeration,Negativism,Withdrawal | 98.0 | 96.6 | 99.1 |
| 7 | Immobility/stupor,Mutism,Staring,Mannerisms,Verbigeration,Rigidity,Withdrawal | 98.0 | 96.6 | 99.1 |
| 7 | Immobility/stupor,Mutism,Staring,Posturing/catalepsy,Echopraxia/echolalia,Negativism,Withdrawal | 98.0 | 96.6 | 99.1 |
| 7 | Immobility/stupor,Mutism,Staring,Posturing/catalepsy,Grimacing,Mannerisms,Negativism | 98.0 | 96.6 | 99.1 |
| 7 | Immobility/stupor,Mutism,Staring,Posturing/catalepsy,Mannerisms,Verbigeration,Negativism | 98.0 | 96.6 | 99.1 |
| 7 | Immobility/stupor,Mutism,Staring,Posturing/catalepsy,Mannerisms,Verbigeration,Rigidity | 98.0 | 96.6 | 99.1 |
| 7 | Immobility/stupor,Mutism,Staring,Posturing/catalepsy,Stereotypy,Negativism,Withdrawal | 98.0 | 96.6 | 99.1 |
| 7 | Immobility/stupor,Mutism,Staring,Posturing/catalepsy,Stereotypy,Verbigeration,Negativism | 98.0 | 96.6 | 99.1 |
| 7 | Immobility/stupor,Mutism,Staring,Stereotypy,Mannerisms,Negativism,Waxy flexibility | 98.0 | 96.6 | 99.1 |
| 7 | Immobility/stupor,Mutism,Staring,Stereotypy,Mannerisms,Verbigeration,Negativism | 98.0 | 96.4 | 99.1 |
| 7 | Immobility/stupor,Mutism,Staring,Stereotypy,Mannerisms,Verbigeration,Rigidity | 98.0 | 96.6 | 99.1 |
| 7 | Immobility/stupor,Mutism,Staring,Stereotypy,Rigidity,Negativism,Withdrawal | 98.0 | 96.6 | 99.1 |
| 7 | Immobility/stupor,Mutism,Staring,Verbigeration,Rigidity,Negativism,Withdrawal | 98.0 | 96.6 | 99.1 |
| 7 | Immobility/stupor,Mutism,Stereotypy,Mannerisms,Rigidity,Negativism,Withdrawal | 98.0 | 96.6 | 99.1 |
| 7 | Immobility/stupor,Mutism,Stereotypy,Verbigeration,Rigidity,Negativism,Withdrawal | 98.0 | 96.6 | 99.1 |
| 7 | Immobility/stupor,Staring,Echopraxia/echolalia,Mannerisms,Verbigeration,Negativism,Withdrawal | 98.0 | 96.6 | 99.1 |
| 7 | Immobility/stupor,Staring,Echopraxia/echolalia,Stereotypy,Mannerisms,Rigidity,Withdrawal | 98.0 | 96.6 | 99.1 |
| 7 | Immobility/stupor,Staring,Echopraxia/echolalia,Stereotypy,Mannerisms,Verbigeration,Negativism | 98.0 | 96.6 | 99.1 |
| 7 | Immobility/stupor,Staring,Echopraxia/echolalia,Stereotypy,Verbigeration,Rigidity,Negativism | 98.0 | 96.6 | 99.1 |
| 7 | Immobility/stupor,Staring,Grimacing,Echopraxia/echolalia,Mannerisms,Rigidity,Withdrawal | 98.0 | 96.6 | 99.1 |
| 7 | Immobility/stupor,Staring,Grimacing,Echopraxia/echolalia,Rigidity,Negativism,Withdrawal | 98.0 | 96.6 | 99.1 |
| 7 | Immobility/stupor,Staring,Grimacing,Mannerisms,Verbigeration,Negativism,Withdrawal | 98.0 | 96.6 | 99.1 |
| 7 | Immobility/stupor,Staring,Grimacing,Mannerisms,Verbigeration,Rigidity,Withdrawal | 98.0 | 96.6 | 99.1 |
| 7 | Immobility/stupor,Staring,Grimacing,Stereotypy,Mannerisms,Verbigeration,Negativism | 98.0 | 96.6 | 99.1 |
| 7 | Immobility/stupor,Staring,Grimacing,Stereotypy,Mannerisms,Verbigeration,Rigidity | 98.0 | 96.6 | 99.1 |
| 7 | Immobility/stupor,Staring,Grimacing,Stereotypy,Rigidity,Negativism,Withdrawal | 98.0 | 96.6 | 99.1 |
| 7 | Immobility/stupor,Staring,Grimacing,Stereotypy,Verbigeration,Negativism,Withdrawal | 98.0 | 96.6 | 99.1 |
| 7 | Immobility/stupor,Staring,Grimacing,Stereotypy,Verbigeration,Rigidity,Withdrawal | 98.0 | 96.6 | 99.1 |
| 7 | Immobility/stupor,Staring,Grimacing,Verbigeration,Rigidity,Negativism,Withdrawal | 98.0 | 96.6 | 99.1 |
| 7 | Immobility/stupor,Staring,Posturing/catalepsy,Echopraxia/echolalia,Mannerisms,Verbigeration,Negativism | 98.0 | 96.6 | 99.1 |
| 7 | Immobility/stupor,Staring,Posturing/catalepsy,Echopraxia/echolalia,Stereotypy,Negativism,Withdrawal | 98.0 | 96.6 | 99.1 |
| 7 | Immobility/stupor,Staring,Posturing/catalepsy,Grimacing,Mannerisms,Rigidity,Withdrawal | 98.0 | 96.6 | 99.1 |
| 7 | Immobility/stupor,Staring,Posturing/catalepsy,Grimacing,Mannerisms,Verbigeration,Negativism | 98.0 | 96.6 | 99.1 |
| 7 | Immobility/stupor,Staring,Posturing/catalepsy,Stereotypy,Mannerisms,Negativism,Waxy flexibility | 98.0 | 96.6 | 99.1 |
| 7 | Immobility/stupor,Staring,Posturing/catalepsy,Verbigeration,Rigidity,Negativism,Withdrawal | 98.0 | 96.6 | 99.1 |
| 7 | Immobility/stupor,Staring,Stereotypy,Mannerisms,Rigidity,Waxy flexibility,Withdrawal | 98.0 | 96.6 | 99.1 |
| 7 | Immobility/stupor,Staring,Stereotypy,Mannerisms,Verbigeration,Negativism,Waxy flexibility | 98.0 | 96.6 | 99.1 |
| 7 | Immobility/stupor,Staring,Stereotypy,Mannerisms,Verbigeration,Rigidity,Withdrawal | 98.0 | 96.6 | 99.1 |
| 7 | Immobility/stupor,Staring,Stereotypy,Rigidity,Negativism,Waxy flexibility,Withdrawal | 98.0 | 96.6 | 99.1 |
| 7 | Immobility/stupor,Stereotypy,Mannerisms,Rigidity,Negativism,Waxy flexibility,Withdrawal | 98.0 | 96.6 | 99.1 |
| 7 | Mutism,Staring,Echopraxia/echolalia,Mannerisms,Rigidity,Waxy flexibility,Withdrawal | 98.0 | 96.6 | 99.1 |
| 7 | Mutism,Staring,Echopraxia/echolalia,Mannerisms,Verbigeration,Rigidity,Withdrawal | 98.0 | 96.6 | 99.1 |
| 7 | Mutism,Staring,Echopraxia/echolalia,Rigidity,Negativism,Waxy flexibility,Withdrawal | 98.0 | 96.6 | 99.1 |
| 7 | Mutism,Staring,Echopraxia/echolalia,Stereotypy,Verbigeration,Negativism,Withdrawal | 98.0 | 96.6 | 99.1 |
| 7 | Mutism,Staring,Echopraxia/echolalia,Stereotypy,Verbigeration,Rigidity,Withdrawal | 98.0 | 96.6 | 99.1 |
| 7 | Mutism,Staring,Echopraxia/echolalia,Verbigeration,Rigidity,Negativism,Waxy flexibility | 98.0 | 96.6 | 99.1 |
| 7 | Mutism,Staring,Grimacing,Echopraxia/echolalia,Mannerisms,Rigidity,Waxy flexibility | 98.0 | 96.6 | 99.1 |
| 7 | Mutism,Staring,Grimacing,Echopraxia/echolalia,Mannerisms,Waxy flexibility,Withdrawal | 98.0 | 96.6 | 99.1 |
| 7 | Mutism,Staring,Grimacing,Echopraxia/echolalia,Stereotypy,Rigidity,Negativism | 98.0 | 96.6 | 99.1 |
| 7 | Mutism,Staring,Grimacing,Echopraxia/echolalia,Stereotypy,Verbigeration,Negativism | 98.0 | 96.6 | 99.1 |
| 7 | Mutism,Staring,Grimacing,Echopraxia/echolalia,Stereotypy,Verbigeration,Rigidity | 98.0 | 96.6 | 99.1 |
| 7 | Mutism,Staring,Grimacing,Mannerisms,Verbigeration,Rigidity,Waxy flexibility | 98.0 | 96.6 | 99.1 |
| 7 | Mutism,Staring,Grimacing,Stereotypy,Mannerisms,Waxy flexibility,Withdrawal | 98.0 | 96.6 | 99.1 |
| 7 | Mutism,Staring,Mannerisms,Verbigeration,Rigidity,Waxy flexibility,Withdrawal | 98.0 | 96.6 | 99.1 |
| 7 | Mutism,Staring,Posturing/catalepsy,Echopraxia/echolalia,Negativism,Waxy flexibility,Withdrawal | 98.0 | 96.6 | 99.1 |
| 7 | Mutism,Staring,Posturing/catalepsy,Echopraxia/echolalia,Stereotypy,Mannerisms,Withdrawal | 98.0 | 96.6 | 99.1 |
| 7 | Mutism,Staring,Posturing/catalepsy,Echopraxia/echolalia,Stereotypy,Verbigeration,Rigidity | 98.0 | 96.6 | 99.1 |
| 7 | Mutism,Staring,Posturing/catalepsy,Grimacing,Echopraxia/echolalia,Stereotypy,Negativism | 98.0 | 96.6 | 99.1 |
| 7 | Mutism,Staring,Posturing/catalepsy,Grimacing,Mannerisms,Negativism,Waxy flexibility | 98.0 | 96.6 | 99.1 |
| 7 | Mutism,Staring,Posturing/catalepsy,Grimacing,Mannerisms,Waxy flexibility,Withdrawal | 98.0 | 96.6 | 99.1 |
| 7 | Mutism,Staring,Posturing/catalepsy,Grimacing,Stereotypy,Rigidity,Withdrawal | 98.0 | 96.6 | 99.1 |
| 7 | Mutism,Staring,Posturing/catalepsy,Mannerisms,Verbigeration,Negativism,Waxy flexibility | 98.0 | 96.6 | 99.1 |
| 7 | Mutism,Staring,Posturing/catalepsy,Stereotypy,Negativism,Waxy flexibility,Withdrawal | 98.0 | 96.6 | 99.1 |
| 7 | Mutism,Staring,Posturing/catalepsy,Stereotypy,Verbigeration,Negativism,Waxy flexibility | 98.0 | 96.6 | 99.1 |
| 7 | Mutism,Staring,Stereotypy,Mannerisms,Verbigeration,Rigidity,Waxy flexibility | 98.0 | 96.6 | 99.1 |
| 7 | Mutism,Staring,Stereotypy,Verbigeration,Negativism,Waxy flexibility,Withdrawal | 98.0 | 96.6 | 99.1 |
| 7 | Mutism,Staring,Verbigeration,Rigidity,Negativism,Waxy flexibility,Withdrawal | 98.0 | 96.6 | 99.1 |
| 7 | Staring,Echopraxia/echolalia,Stereotypy,Mannerisms,Verbigeration,Negativism,Withdrawal | 98.0 | 96.6 | 99.1 |
| 7 | Staring,Grimacing,Echopraxia/echolalia,Mannerisms,Negativism,Waxy flexibility,Withdrawal | 98.0 | 96.6 | 99.1 |
| 7 | Staring,Grimacing,Echopraxia/echolalia,Stereotypy,Rigidity,Negativism,Withdrawal | 98.0 | 96.6 | 99.1 |
| 7 | Staring,Grimacing,Stereotypy,Mannerisms,Negativism,Waxy flexibility,Withdrawal | 98.0 | 96.6 | 99.1 |
| 7 | Staring,Posturing/catalepsy,Echopraxia/echolalia,Mannerisms,Verbigeration,Negativism,Withdrawal | 98.0 | 96.6 | 99.1 |
| 7 | Staring,Posturing/catalepsy,Echopraxia/echolalia,Stereotypy,Rigidity,Negativism,Withdrawal | 98.0 | 96.6 | 99.1 |
| 7 | Staring,Posturing/catalepsy,Grimacing,Echopraxia/echolalia,Mannerisms,Rigidity,Withdrawal | 98.0 | 96.6 | 99.1 |
| 7 | Staring,Posturing/catalepsy,Grimacing,Mannerisms,Negativism,Waxy flexibility,Withdrawal | 98.0 | 96.6 | 99.1 |
| 7 | Staring,Posturing/catalepsy,Grimacing,Mannerisms,Verbigeration,Rigidity,Withdrawal | 98.0 | 96.6 | 99.1 |
| 7 | Staring,Posturing/catalepsy,Mannerisms,Verbigeration,Negativism,Waxy flexibility,Withdrawal | 98.0 | 96.6 | 99.1 |
| 7 | Staring,Stereotypy,Mannerisms,Verbigeration,Negativism,Waxy flexibility,Withdrawal | 98.0 | 96.6 | 99.1 |
| 7 | Excitement,Immobility/stupor,Echopraxia/echolalia,Mannerisms,Verbigeration,Rigidity,Negativism | 97.8 | 96.4 | 99.1 |
| 7 | Excitement,Immobility/stupor,Echopraxia/echolalia,Stereotypy,Mannerisms,Rigidity,Negativism | 97.8 | 96.4 | 99.1 |
| 7 | Excitement,Immobility/stupor,Echopraxia/echolalia,Stereotypy,Verbigeration,Negativism,Withdrawal | 97.8 | 96.2 | 99.1 |
| 7 | Excitement,Immobility/stupor,Grimacing,Mannerisms,Rigidity,Negativism,Waxy flexibility | 97.8 | 96.4 | 99.1 |
| 7 | Excitement,Immobility/stupor,Grimacing,Mannerisms,Verbigeration,Rigidity,Withdrawal | 97.8 | 96.2 | 99.1 |
| 7 | Excitement,Immobility/stupor,Grimacing,Rigidity,Negativism,Waxy flexibility,Withdrawal | 97.8 | 96.2 | 99.1 |
| 7 | Excitement,Immobility/stupor,Grimacing,Stereotypy,Rigidity,Negativism,Waxy flexibility | 97.8 | 96.2 | 99.1 |
| 7 | Excitement,Immobility/stupor,Grimacing,Verbigeration,Rigidity,Negativism,Waxy flexibility | 97.8 | 96.2 | 99.1 |
| 7 | Excitement,Immobility/stupor,Mannerisms,Verbigeration,Rigidity,Negativism,Waxy flexibility | 97.8 | 96.2 | 99.1 |
| 7 | Excitement,Immobility/stupor,Mannerisms,Verbigeration,Rigidity,Waxy flexibility,Withdrawal | 97.8 | 96.2 | 99.1 |
| 7 | Excitement,Immobility/stupor,Mutism,Echopraxia/echolalia,Mannerisms,Verbigeration,Rigidity | 97.8 | 96.2 | 99.1 |
| 7 | Excitement,Immobility/stupor,Mutism,Echopraxia/echolalia,Stereotypy,Negativism,Withdrawal | 97.8 | 96.4 | 99.1 |
| 7 | Excitement,Immobility/stupor,Mutism,Grimacing,Echopraxia/echolalia,Stereotypy,Rigidity | 97.8 | 96.4 | 99.1 |
| 7 | Excitement,Immobility/stupor,Mutism,Grimacing,Stereotypy,Rigidity,Waxy flexibility | 97.8 | 96.2 | 99.1 |
| 7 | Excitement,Immobility/stupor,Mutism,Mannerisms,Verbigeration,Rigidity,Waxy flexibility | 97.8 | 96.2 | 99.1 |
| 7 | Excitement,Immobility/stupor,Mutism,Posturing/catalepsy,Grimacing,Rigidity,Waxy flexibility | 97.8 | 96.2 | 99.1 |
| 7 | Excitement,Immobility/stupor,Mutism,Posturing/catalepsy,Mannerisms,Verbigeration,Negativism | 97.8 | 96.4 | 99.1 |
| 7 | Excitement,Immobility/stupor,Mutism,Posturing/catalepsy,Negativism,Waxy flexibility,Withdrawal | 97.8 | 96.4 | 99.1 |
| 7 | Excitement,Immobility/stupor,Mutism,Posturing/catalepsy,Stereotypy,Mannerisms,Rigidity | 97.8 | 96.4 | 99.1 |
| 7 | Excitement,Immobility/stupor,Mutism,Posturing/catalepsy,Stereotypy,Rigidity,Waxy flexibility | 97.8 | 96.2 | 99.1 |
| 7 | Excitement,Immobility/stupor,Mutism,Posturing/catalepsy,Verbigeration,Negativism,Waxy flexibility | 97.8 | 96.2 | 99.1 |
| 7 | Excitement,Immobility/stupor,Mutism,Staring,Echopraxia/echolalia,Stereotypy,Verbigeration | 97.8 | 96.4 | 99.1 |
| 7 | Excitement,Immobility/stupor,Mutism,Staring,Grimacing,Echopraxia/echolalia,Stereotypy | 97.8 | 96.2 | 99.1 |
| 7 | Excitement,Immobility/stupor,Mutism,Staring,Grimacing,Mannerisms,Negativism | 97.8 | 96.4 | 99.1 |
| 7 | Excitement,Immobility/stupor,Mutism,Staring,Grimacing,Mannerisms,Waxy flexibility | 97.8 | 96.4 | 99.1 |
| 7 | Excitement,Immobility/stupor,Mutism,Staring,Grimacing,Stereotypy,Withdrawal | 97.8 | 96.4 | 98.9 |
| 7 | Excitement,Immobility/stupor,Mutism,Staring,Stereotypy,Mannerisms,Waxy flexibility | 97.8 | 96.4 | 99.1 |
| 7 | Excitement,Immobility/stupor,Mutism,Staring,Stereotypy,Verbigeration,Withdrawal | 97.8 | 96.4 | 99.1 |
| 7 | Excitement,Immobility/stupor,Mutism,Stereotypy,Mannerisms,Verbigeration,Rigidity | 97.8 | 96.2 | 99.1 |
| 7 | Excitement,Immobility/stupor,Posturing/catalepsy,Echopraxia/echolalia,Negativism,Waxy flexibility,Withdrawal | 97.8 | 96.2 | 99.1 |
| 7 | Excitement,Immobility/stupor,Posturing/catalepsy,Grimacing,Echopraxia/echolalia,Verbigeration,Rigidity | 97.8 | 96.4 | 99.1 |
| 7 | Excitement,Immobility/stupor,Posturing/catalepsy,Grimacing,Mannerisms,Negativism,Withdrawal | 97.8 | 96.2 | 99.1 |
| 7 | Excitement,Immobility/stupor,Posturing/catalepsy,Grimacing,Rigidity,Waxy flexibility,Withdrawal | 97.8 | 96.4 | 99.1 |
| 7 | Excitement,Immobility/stupor,Posturing/catalepsy,Mannerisms,Rigidity,Negativism,Waxy flexibility | 97.8 | 96.2 | 99.1 |
| 7 | Excitement,Immobility/stupor,Posturing/catalepsy,Mannerisms,Rigidity,Waxy flexibility,Withdrawal | 97.8 | 96.4 | 99.1 |
| 7 | Excitement,Immobility/stupor,Posturing/catalepsy,Stereotypy,Mannerisms,Rigidity,Negativism | 97.8 | 96.2 | 99.1 |
| 7 | Excitement,Immobility/stupor,Posturing/catalepsy,Stereotypy,Rigidity,Negativism,Waxy flexibility | 97.8 | 96.2 | 99.1 |
| 7 | Excitement,Immobility/stupor,Posturing/catalepsy,Stereotypy,Rigidity,Waxy flexibility,Withdrawal | 97.8 | 96.2 | 99.1 |
| 7 | Excitement,Immobility/stupor,Posturing/catalepsy,Verbigeration,Negativism,Waxy flexibility,Withdrawal | 97.8 | 96.4 | 99.1 |
| 7 | Excitement,Immobility/stupor,Staring,Echopraxia/echolalia,Mannerisms,Verbigeration,Waxy flexibility | 97.8 | 96.4 | 99.1 |
| 7 | Excitement,Immobility/stupor,Staring,Echopraxia/echolalia,Verbigeration,Waxy flexibility,Withdrawal | 97.8 | 96.4 | 99.1 |
| 7 | Excitement,Immobility/stupor,Staring,Grimacing,Echopraxia/echolalia,Mannerisms,Waxy flexibility | 97.8 | 96.2 | 99.1 |
| 7 | Excitement,Immobility/stupor,Staring,Grimacing,Echopraxia/echolalia,Mannerisms,Withdrawal | 97.8 | 96.2 | 99.1 |
| 7 | Excitement,Immobility/stupor,Staring,Grimacing,Echopraxia/echolalia,Stereotypy,Withdrawal | 97.8 | 96.2 | 99.1 |
| 7 | Excitement,Immobility/stupor,Staring,Mannerisms,Verbigeration,Waxy flexibility,Withdrawal | 97.8 | 96.2 | 99.1 |
| 7 | Excitement,Immobility/stupor,Staring,Posturing/catalepsy,Grimacing,Echopraxia/echolalia,Verbigeration | 97.8 | 96.2 | 99.1 |
| 7 | Excitement,Immobility/stupor,Staring,Posturing/catalepsy,Grimacing,Mannerisms,Waxy flexibility | 97.8 | 96.4 | 99.1 |
| 7 | Excitement,Immobility/stupor,Staring,Posturing/catalepsy,Grimacing,Stereotypy,Withdrawal | 97.8 | 96.2 | 99.1 |
| 7 | Excitement,Immobility/stupor,Staring,Posturing/catalepsy,Grimacing,Waxy flexibility,Withdrawal | 97.8 | 96.2 | 99.1 |
| 7 | Excitement,Immobility/stupor,Staring,Posturing/catalepsy,Stereotypy,Mannerisms,Verbigeration | 97.8 | 96.2 | 99.1 |
| 7 | Excitement,Immobility/stupor,Staring,Posturing/catalepsy,Stereotypy,Verbigeration,Withdrawal | 97.8 | 96.2 | 99.1 |
| 7 | Excitement,Mutism,Staring,Echopraxia/echolalia,Mannerisms,Verbigeration,Waxy flexibility | 97.8 | 96.4 | 99.1 |
| 7 | Excitement,Mutism,Staring,Echopraxia/echolalia,Mannerisms,Verbigeration,Withdrawal | 97.8 | 96.2 | 99.1 |
| 7 | Excitement,Mutism,Staring,Grimacing,Echopraxia/echolalia,Stereotypy,Mannerisms | 97.8 | 96.4 | 99.1 |
| 7 | Excitement,Mutism,Staring,Grimacing,Mannerisms,Verbigeration,Waxy flexibility | 97.8 | 96.4 | 99.1 |
| 7 | Excitement,Mutism,Staring,Grimacing,Mannerisms,Verbigeration,Withdrawal | 97.8 | 96.4 | 99.1 |
| 7 | Excitement,Mutism,Staring,Grimacing,Stereotypy,Mannerisms,Waxy flexibility | 97.8 | 96.4 | 99.1 |
| 7 | Excitement,Mutism,Staring,Grimacing,Stereotypy,Mannerisms,Withdrawal | 97.8 | 96.2 | 99.1 |
| 7 | Excitement,Mutism,Staring,Stereotypy,Mannerisms,Verbigeration,Waxy flexibility | 97.8 | 96.4 | 99.1 |
| 7 | Excitement,Staring,Echopraxia/echolalia,Mannerisms,Verbigeration,Negativism,Withdrawal | 97.8 | 96.2 | 99.1 |
| 7 | Excitement,Staring,Echopraxia/echolalia,Stereotypy,Verbigeration,Rigidity,Negativism | 97.8 | 96.2 | 99.1 |
| 7 | Excitement,Staring,Echopraxia/echolalia,Verbigeration,Rigidity,Negativism,Waxy flexibility | 97.8 | 96.4 | 99.1 |
| 7 | Excitement,Staring,Grimacing,Echopraxia/echolalia,Mannerisms,Waxy flexibility,Withdrawal | 97.8 | 96.2 | 99.1 |
| 7 | Excitement,Staring,Grimacing,Mannerisms,Negativism,Waxy flexibility,Withdrawal | 97.8 | 96.4 | 99.1 |
| 7 | Excitement,Staring,Grimacing,Mannerisms,Verbigeration,Negativism,Withdrawal | 97.8 | 96.4 | 99.1 |
| 7 | Excitement,Staring,Grimacing,Stereotypy,Mannerisms,Negativism,Withdrawal | 97.8 | 96.2 | 99.1 |
| 7 | Excitement,Staring,Posturing/catalepsy,Echopraxia/echolalia,Rigidity,Negativism,Waxy flexibility | 97.8 | 96.2 | 99.1 |
| 7 | Excitement,Staring,Posturing/catalepsy,Grimacing,Echopraxia/echolalia,Mannerisms,Rigidity | 97.8 | 96.2 | 99.1 |
| 7 | Excitement,Staring,Posturing/catalepsy,Grimacing,Echopraxia/echolalia,Rigidity,Negativism | 97.8 | 96.4 | 99.1 |
| 7 | Excitement,Staring,Posturing/catalepsy,Grimacing,Echopraxia/echolalia,Waxy flexibility,Withdrawal | 97.8 | 96.2 | 99.1 |
| 7 | Excitement,Staring,Posturing/catalepsy,Grimacing,Mannerisms,Rigidity,Negativism | 97.8 | 96.4 | 99.1 |
| 7 | Excitement,Staring,Posturing/catalepsy,Grimacing,Mannerisms,Verbigeration,Rigidity | 97.8 | 96.2 | 99.1 |
| 7 | Excitement,Staring,Posturing/catalepsy,Grimacing,Mannerisms,Waxy flexibility,Withdrawal | 97.8 | 96.4 | 99.1 |
| 7 | Excitement,Staring,Posturing/catalepsy,Mannerisms,Rigidity,Negativism,Waxy flexibility | 97.8 | 96.2 | 99.1 |
| 7 | Excitement,Staring,Posturing/catalepsy,Mannerisms,Verbigeration,Waxy flexibility,Withdrawal | 97.8 | 96.4 | 99.1 |
| 7 | Immobility/stupor,Echopraxia/echolalia,Mannerisms,Rigidity,Negativism,Waxy flexibility,Withdrawal | 97.8 | 96.2 | 99.1 |
| 7 | Immobility/stupor,Echopraxia/echolalia,Stereotypy,Verbigeration,Rigidity,Negativism,Withdrawal | 97.8 | 96.2 | 99.1 |
| 7 | Immobility/stupor,Grimacing,Mannerisms,Verbigeration,Rigidity,Negativism,Withdrawal | 97.8 | 96.4 | 99.1 |
| 7 | Immobility/stupor,Grimacing,Stereotypy,Verbigeration,Rigidity,Negativism,Withdrawal | 97.8 | 96.2 | 99.1 |
| 7 | Immobility/stupor,Mutism,Echopraxia/echolalia,Mannerisms,Verbigeration,Rigidity,Negativism | 97.8 | 96.2 | 99.1 |
| 7 | Immobility/stupor,Mutism,Echopraxia/echolalia,Stereotypy,Mannerisms,Rigidity,Negativism | 97.8 | 96.2 | 99.1 |
| 7 | Immobility/stupor,Mutism,Grimacing,Echopraxia/echolalia,Mannerisms,Rigidity,Negativism | 97.8 | 96.2 | 99.1 |
| 7 | Immobility/stupor,Mutism,Grimacing,Mannerisms,Rigidity,Negativism,Withdrawal | 97.8 | 96.4 | 99.1 |
| 7 | Immobility/stupor,Mutism,Posturing/catalepsy,Echopraxia/echolalia,Mannerisms,Rigidity,Negativism | 97.8 | 96.4 | 99.1 |
| 7 | Immobility/stupor,Mutism,Posturing/catalepsy,Mannerisms,Rigidity,Negativism,Withdrawal | 97.8 | 96.2 | 99.1 |
| 7 | Immobility/stupor,Mutism,Posturing/catalepsy,Mannerisms,Verbigeration,Rigidity,Negativism | 97.8 | 96.2 | 99.1 |
| 7 | Immobility/stupor,Mutism,Posturing/catalepsy,Stereotypy,Mannerisms,Rigidity,Negativism | 97.8 | 96.2 | 99.1 |
| 7 | Immobility/stupor,Mutism,Staring,Echopraxia/echolalia,Mannerisms,Negativism,Waxy flexibility | 97.8 | 96.2 | 99.1 |
| 7 | Immobility/stupor,Mutism,Staring,Grimacing,Mannerisms,Verbigeration,Rigidity | 97.8 | 96.2 | 99.1 |
| 7 | Immobility/stupor,Mutism,Staring,Grimacing,Stereotypy,Mannerisms,Negativism | 97.8 | 96.4 | 99.1 |
| 7 | Immobility/stupor,Mutism,Staring,Grimacing,Stereotypy,Mannerisms,Rigidity | 97.8 | 96.4 | 99.1 |
| 7 | Immobility/stupor,Mutism,Staring,Mannerisms,Rigidity,Waxy flexibility,Withdrawal | 97.8 | 96.4 | 99.1 |
| 7 | Immobility/stupor,Mutism,Staring,Posturing/catalepsy,Echopraxia/echolalia,Mannerisms,Rigidity | 97.8 | 96.2 | 99.1 |
| 7 | Immobility/stupor,Mutism,Staring,Posturing/catalepsy,Grimacing,Mannerisms,Withdrawal | 97.8 | 96.2 | 99.1 |
| 7 | Immobility/stupor,Mutism,Staring,Posturing/catalepsy,Stereotypy,Mannerisms,Rigidity | 97.8 | 96.2 | 99.1 |
| 7 | Immobility/stupor,Mutism,Staring,Posturing/catalepsy,Stereotypy,Rigidity,Negativism | 97.8 | 96.2 | 99.1 |
| 7 | Immobility/stupor,Mutism,Staring,Posturing/catalepsy,Stereotypy,Verbigeration,Rigidity | 97.8 | 96.2 | 99.1 |
| 7 | Immobility/stupor,Mutism,Staring,Stereotypy,Verbigeration,Negativism,Withdrawal | 97.8 | 96.4 | 99.1 |
| 7 | Immobility/stupor,Mutism,Staring,Stereotypy,Verbigeration,Rigidity,Withdrawal | 97.8 | 96.2 | 99.1 |
| 7 | Immobility/stupor,Posturing/catalepsy,Grimacing,Echopraxia/echolalia,Mannerisms,Rigidity,Negativism | 97.8 | 96.2 | 99.1 |
| 7 | Immobility/stupor,Posturing/catalepsy,Mannerisms,Rigidity,Negativism,Waxy flexibility,Withdrawal | 97.8 | 96.2 | 99.1 |
| 7 | Immobility/stupor,Posturing/catalepsy,Stereotypy,Verbigeration,Rigidity,Negativism,Withdrawal | 97.8 | 96.4 | 99.1 |
| 7 | Immobility/stupor,Staring,Echopraxia/echolalia,Rigidity,Negativism,Waxy flexibility,Withdrawal | 97.8 | 96.2 | 99.1 |
| 7 | Immobility/stupor,Staring,Grimacing,Echopraxia/echolalia,Stereotypy,Negativism,Withdrawal | 97.8 | 96.2 | 99.1 |
| 7 | Immobility/stupor,Staring,Grimacing,Echopraxia/echolalia,Verbigeration,Rigidity,Negativism | 97.8 | 96.2 | 99.1 |
| 7 | Immobility/stupor,Staring,Grimacing,Mannerisms,Negativism,Waxy flexibility,Withdrawal | 97.8 | 96.4 | 99.1 |
| 7 | Immobility/stupor,Staring,Grimacing,Mannerisms,Rigidity,Waxy flexibility,Withdrawal | 97.8 | 96.2 | 98.9 |
| 7 | Immobility/stupor,Staring,Grimacing,Stereotypy,Mannerisms,Negativism,Waxy flexibility | 97.8 | 96.2 | 99.1 |
| 7 | Immobility/stupor,Staring,Grimacing,Stereotypy,Mannerisms,Rigidity,Waxy flexibility | 97.8 | 96.2 | 98.9 |
| 7 | Immobility/stupor,Staring,Mannerisms,Verbigeration,Negativism,Waxy flexibility,Withdrawal | 97.8 | 96.2 | 99.1 |
| 7 | Immobility/stupor,Staring,Posturing/catalepsy,Echopraxia/echolalia,Mannerisms,Negativism,Waxy flexibility | 97.8 | 96.4 | 99.1 |
| 7 | Immobility/stupor,Staring,Posturing/catalepsy,Echopraxia/echolalia,Mannerisms,Rigidity,Withdrawal | 97.8 | 96.2 | 99.1 |
| 7 | Immobility/stupor,Staring,Posturing/catalepsy,Echopraxia/echolalia,Stereotypy,Rigidity,Negativism | 97.8 | 96.2 | 99.1 |
| 7 | Immobility/stupor,Staring,Posturing/catalepsy,Grimacing,Stereotypy,Mannerisms,Rigidity | 97.8 | 96.2 | 99.1 |
| 7 | Immobility/stupor,Staring,Posturing/catalepsy,Grimacing,Stereotypy,Negativism,Withdrawal | 97.8 | 96.2 | 99.1 |
| 7 | Immobility/stupor,Staring,Posturing/catalepsy,Grimacing,Stereotypy,Verbigeration,Negativism | 97.8 | 96.2 | 99.1 |
| 7 | Immobility/stupor,Staring,Posturing/catalepsy,Mannerisms,Verbigeration,Rigidity,Withdrawal | 97.8 | 96.4 | 99.1 |
| 7 | Immobility/stupor,Staring,Posturing/catalepsy,Stereotypy,Mannerisms,Verbigeration,Rigidity | 97.8 | 96.2 | 99.1 |
| 7 | Immobility/stupor,Staring,Verbigeration,Rigidity,Negativism,Waxy flexibility,Withdrawal | 97.8 | 96.2 | 99.1 |
| 7 | Mutism,Staring,Echopraxia/echolalia,Stereotypy,Mannerisms,Rigidity,Waxy flexibility | 97.8 | 96.4 | 99.1 |
| 7 | Mutism,Staring,Echopraxia/echolalia,Stereotypy,Mannerisms,Verbigeration,Rigidity | 97.8 | 96.2 | 99.1 |
| 7 | Mutism,Staring,Echopraxia/echolalia,Stereotypy,Negativism,Waxy flexibility,Withdrawal | 97.8 | 96.2 | 99.1 |
| 7 | Mutism,Staring,Echopraxia/echolalia,Stereotypy,Rigidity,Negativism,Waxy flexibility | 97.8 | 96.2 | 99.1 |
| 7 | Mutism,Staring,Grimacing,Echopraxia/echolalia,Stereotypy,Mannerisms,Waxy flexibility | 97.8 | 96.4 | 99.1 |
| 7 | Mutism,Staring,Grimacing,Echopraxia/echolalia,Verbigeration,Negativism,Waxy flexibility | 97.8 | 96.2 | 99.1 |
| 7 | Mutism,Staring,Grimacing,Echopraxia/echolalia,Verbigeration,Negativism,Withdrawal | 97.8 | 96.4 | 99.1 |
| 7 | Mutism,Staring,Grimacing,Echopraxia/echolalia,Verbigeration,Rigidity,Withdrawal | 97.8 | 96.2 | 99.1 |
| 7 | Mutism,Staring,Grimacing,Mannerisms,Verbigeration,Negativism,Waxy flexibility | 97.8 | 96.2 | 99.1 |
| 7 | Mutism,Staring,Grimacing,Stereotypy,Mannerisms,Verbigeration,Withdrawal | 97.8 | 96.4 | 99.1 |
| 7 | Mutism,Staring,Grimacing,Stereotypy,Rigidity,Waxy flexibility,Withdrawal | 97.8 | 96.2 | 99.1 |
| 7 | Mutism,Staring,Grimacing,Stereotypy,Verbigeration,Negativism,Waxy flexibility | 97.8 | 96.2 | 99.1 |
| 7 | Mutism,Staring,Posturing/catalepsy,Echopraxia/echolalia,Mannerisms,Rigidity,Waxy flexibility | 97.8 | 96.2 | 99.1 |
| 7 | Mutism,Staring,Posturing/catalepsy,Echopraxia/echolalia,Mannerisms,Verbigeration,Withdrawal | 97.8 | 96.2 | 99.1 |
| 7 | Mutism,Staring,Posturing/catalepsy,Echopraxia/echolalia,Mannerisms,Waxy flexibility,Withdrawal | 97.8 | 96.4 | 99.1 |
| 7 | Mutism,Staring,Posturing/catalepsy,Echopraxia/echolalia,Stereotypy,Negativism,Waxy flexibility | 97.8 | 96.2 | 99.1 |
| 7 | Mutism,Staring,Posturing/catalepsy,Echopraxia/echolalia,Verbigeration,Negativism,Waxy flexibility | 97.8 | 96.2 | 99.1 |
| 7 | Mutism,Staring,Posturing/catalepsy,Echopraxia/echolalia,Verbigeration,Rigidity,Withdrawal | 97.8 | 96.2 | 99.1 |
| 7 | Mutism,Staring,Posturing/catalepsy,Grimacing,Echopraxia/echolalia,Mannerisms,Verbigeration | 97.8 | 96.2 | 99.1 |
| 7 | Mutism,Staring,Posturing/catalepsy,Grimacing,Echopraxia/echolalia,Rigidity,Negativism | 97.8 | 96.2 | 99.1 |
| 7 | Mutism,Staring,Posturing/catalepsy,Grimacing,Echopraxia/echolalia,Rigidity,Withdrawal | 97.8 | 96.2 | 99.1 |
| 7 | Mutism,Staring,Posturing/catalepsy,Grimacing,Echopraxia/echolalia,Stereotypy,Mannerisms | 97.8 | 96.2 | 99.1 |
| 7 | Mutism,Staring,Posturing/catalepsy,Grimacing,Echopraxia/echolalia,Stereotypy,Withdrawal | 97.8 | 96.2 | 99.1 |
| 7 | Mutism,Staring,Posturing/catalepsy,Grimacing,Echopraxia/echolalia,Verbigeration,Rigidity | 97.8 | 96.2 | 99.1 |
| 7 | Mutism,Staring,Posturing/catalepsy,Grimacing,Rigidity,Negativism,Withdrawal | 97.8 | 96.4 | 99.1 |
| 7 | Mutism,Staring,Posturing/catalepsy,Grimacing,Stereotypy,Rigidity,Negativism | 97.8 | 96.2 | 99.1 |
| 7 | Mutism,Staring,Posturing/catalepsy,Grimacing,Stereotypy,Verbigeration,Withdrawal | 97.8 | 96.2 | 99.1 |
| 7 | Mutism,Staring,Posturing/catalepsy,Grimacing,Verbigeration,Rigidity,Withdrawal | 97.8 | 96.2 | 99.1 |
| 7 | Mutism,Staring,Posturing/catalepsy,Mannerisms,Verbigeration,Rigidity,Waxy flexibility | 97.8 | 96.2 | 99.1 |
| 7 | Mutism,Staring,Posturing/catalepsy,Stereotypy,Mannerisms,Waxy flexibility,Withdrawal | 97.8 | 96.2 | 99.1 |
| 7 | Mutism,Staring,Posturing/catalepsy,Stereotypy,Verbigeration,Rigidity,Waxy flexibility | 97.8 | 96.2 | 99.1 |
| 7 | Mutism,Staring,Posturing/catalepsy,Verbigeration,Negativism,Waxy flexibility,Withdrawal | 97.8 | 96.2 | 99.1 |
| 7 | Staring,Echopraxia/echolalia,Stereotypy,Rigidity,Negativism,Waxy flexibility,Withdrawal | 97.8 | 96.2 | 99.1 |
| 7 | Staring,Grimacing,Echopraxia/echolalia,Mannerisms,Rigidity,Waxy flexibility,Withdrawal | 97.8 | 96.2 | 99.1 |
| 7 | Staring,Grimacing,Mannerisms,Verbigeration,Rigidity,Waxy flexibility,Withdrawal | 97.8 | 96.2 | 99.1 |
| 7 | Staring,Posturing/catalepsy,Echopraxia/echolalia,Stereotypy,Mannerisms,Rigidity,Negativism | 97.8 | 96.4 | 99.1 |
| 7 | Staring,Posturing/catalepsy,Echopraxia/echolalia,Stereotypy,Mannerisms,Rigidity,Withdrawal | 97.8 | 96.2 | 99.1 |
| 7 | Staring,Posturing/catalepsy,Echopraxia/echolalia,Verbigeration,Rigidity,Negativism,Withdrawal | 97.8 | 96.2 | 99.1 |
| 7 | Staring,Posturing/catalepsy,Grimacing,Echopraxia/echolalia,Rigidity,Negativism,Withdrawal | 97.8 | 96.4 | 99.1 |
| 7 | Staring,Posturing/catalepsy,Grimacing,Stereotypy,Rigidity,Negativism,Withdrawal | 97.8 | 96.2 | 99.1 |
| 7 | Staring,Posturing/catalepsy,Grimacing,Stereotypy,Verbigeration,Negativism,Withdrawal | 97.8 | 96.4 | 99.1 |
| 7 | Staring,Posturing/catalepsy,Stereotypy,Mannerisms,Verbigeration,Rigidity,Negativism | 97.8 | 96.4 | 99.1 |
| 7 | Staring,Posturing/catalepsy,Stereotypy,Mannerisms,Verbigeration,Rigidity,Withdrawal | 97.8 | 96.2 | 99.1 |
| 7 | Staring,Stereotypy,Mannerisms,Verbigeration,Rigidity,Negativism,Waxy flexibility | 97.8 | 96.4 | 99.1 |
| 7 | Excitement,Immobility/stupor,Grimacing,Stereotypy,Verbigeration,Rigidity,Withdrawal | 97.7 | 96.2 | 99.1 |
| 7 | Excitement,Immobility/stupor,Mutism,Grimacing,Mannerisms,Verbigeration,Rigidity | 97.7 | 96.2 | 99.1 |
| 7 | Excitement,Immobility/stupor,Mutism,Posturing/catalepsy,Grimacing,Echopraxia/echolalia,Withdrawal | 97.7 | 96.2 | 98.9 |
| 7 | Excitement,Immobility/stupor,Staring,Posturing/catalepsy,Grimacing,Verbigeration,Waxy flexibility | 97.7 | 96.2 | 99.1 |
| 7 | Excitement,Immobility/stupor,Staring,Posturing/catalepsy,Stereotypy,Verbigeration,Waxy flexibility | 97.7 | 96.4 | 99.1 |
| 7 | Excitement,Immobility/stupor,Staring,Posturing/catalepsy,Stereotypy,Waxy flexibility,Withdrawal | 97.7 | 96.2 | 98.9 |
| 7 | Immobility/stupor,Mutism,Posturing/catalepsy,Stereotypy,Verbigeration,Rigidity,Negativism | 97.7 | 96.2 | 99.1 |
| 7 | Immobility/stupor,Mutism,Staring,Echopraxia/echolalia,Verbigeration,Rigidity,Negativism | 97.7 | 96.2 | 99.1 |
| 7 | Immobility/stupor,Mutism,Staring,Grimacing,Echopraxia/echolalia,Mannerisms,Negativism | 97.7 | 96.2 | 99.1 |
| 7 | Immobility/stupor,Mutism,Staring,Posturing/catalepsy,Verbigeration,Negativism,Withdrawal | 97.7 | 96.4 | 99.1 |
| 7 | Immobility/stupor,Mutism,Stereotypy,Mannerisms,Rigidity,Negativism,Waxy flexibility | 97.7 | 96.4 | 99.1 |
| 7 | Immobility/stupor,Posturing/catalepsy,Stereotypy,Mannerisms,Verbigeration,Rigidity,Negativism | 97.7 | 96.2 | 99.1 |
| 7 | Immobility/stupor,Staring,Grimacing,Echopraxia/echolalia,Mannerisms,Negativism,Waxy flexibility | 97.7 | 96.2 | 99.1 |
| 7 | Immobility/stupor,Staring,Posturing/catalepsy,Echopraxia/echolalia,Verbigeration,Rigidity,Negativism | 97.7 | 96.2 | 99.1 |
| 7 | Immobility/stupor,Staring,Posturing/catalepsy,Grimacing,Echopraxia/echolalia,Negativism,Withdrawal | 97.7 | 96.2 | 99.1 |
| 7 | Immobility/stupor,Stereotypy,Mannerisms,Verbigeration,Rigidity,Negativism,Waxy flexibility | 97.7 | 96.2 | 99.1 |
| 7 | Mutism,Staring,Echopraxia/echolalia,Stereotypy,Rigidity,Waxy flexibility,Withdrawal | 97.7 | 96.4 | 98.9 |
| 7 | Mutism,Staring,Echopraxia/echolalia,Stereotypy,Verbigeration,Negativism,Waxy flexibility | 97.7 | 96.2 | 99.1 |
| 7 | Mutism,Staring,Posturing/catalepsy,Grimacing,Verbigeration,Rigidity,Negativism | 97.7 | 96.2 | 99.1 |
| 7 | Mutism,Staring,Posturing/catalepsy,Stereotypy,Mannerisms,Verbigeration,Withdrawal | 97.7 | 96.2 | 99.1 |
| 7 | Staring,Echopraxia/echolalia,Stereotypy,Mannerisms,Rigidity,Negativism,Waxy flexibility | 97.7 | 96.2 | 99.1 |
| 7 | Excitement,Immobility/stupor,Mutism,Grimacing,Echopraxia/echolalia,Mannerisms,Rigidity | 97.6 | 96.0 | 98.9 |
| 7 | Immobility/stupor,Mutism,Staring,Stereotypy,Mannerisms,Rigidity,Waxy flexibility | 97.6 | 96.0 | 98.9 |
| 7 | Staring,Posturing/catalepsy,Grimacing,Echopraxia/echolalia,Mannerisms,Rigidity,Negativism | 97.6 | 96.0 | 98.9 |
| 7 | Excitement,Immobility/stupor,Echopraxia/echolalia,Mannerisms,Rigidity,Negativism,Waxy flexibility | 97.5 | 96.0 | 98.9 |
| 7 | Excitement,Immobility/stupor,Grimacing,Mannerisms,Rigidity,Waxy flexibility,Withdrawal | 97.5 | 96.0 | 98.9 |
| 7 | Excitement,Immobility/stupor,Grimacing,Stereotypy,Rigidity,Waxy flexibility,Withdrawal | 97.5 | 96.0 | 98.9 |
| 7 | Excitement,Immobility/stupor,Mutism,Echopraxia/echolalia,Mannerisms,Rigidity,Waxy flexibility | 97.5 | 96.0 | 98.9 |
| 7 | Excitement,Immobility/stupor,Mutism,Echopraxia/echolalia,Stereotypy,Mannerisms,Rigidity | 97.5 | 96.0 | 98.9 |
| 7 | Excitement,Immobility/stupor,Mutism,Echopraxia/echolalia,Verbigeration,Negativism,Withdrawal | 97.5 | 96.0 | 98.9 |
| 7 | Excitement,Immobility/stupor,Mutism,Grimacing,Mannerisms,Rigidity,Waxy flexibility | 97.5 | 96.0 | 98.9 |
| 7 | Excitement,Immobility/stupor,Mutism,Posturing/catalepsy,Echopraxia/echolalia,Negativism,Waxy flexibility | 97.5 | 96.0 | 98.9 |
| 7 | Excitement,Immobility/stupor,Mutism,Posturing/catalepsy,Echopraxia/echolalia,Stereotypy,Negativism | 97.5 | 96.0 | 98.9 |
| 7 | Excitement,Immobility/stupor,Mutism,Posturing/catalepsy,Echopraxia/echolalia,Verbigeration,Withdrawal | 97.5 | 96.0 | 98.9 |
| 7 | Excitement,Immobility/stupor,Mutism,Posturing/catalepsy,Grimacing,Mannerisms,Negativism | 97.5 | 96.0 | 98.9 |
| 7 | Excitement,Immobility/stupor,Mutism,Posturing/catalepsy,Grimacing,Negativism,Waxy flexibility | 97.5 | 96.0 | 98.9 |
| 7 | Excitement,Immobility/stupor,Mutism,Posturing/catalepsy,Grimacing,Stereotypy,Negativism | 97.5 | 96.0 | 98.9 |
| 7 | Excitement,Immobility/stupor,Mutism,Posturing/catalepsy,Mannerisms,Rigidity,Waxy flexibility | 97.5 | 96.0 | 98.9 |
| 7 | Excitement,Immobility/stupor,Mutism,Staring,Echopraxia/echolalia,Stereotypy,Mannerisms | 97.5 | 96.0 | 98.9 |
| 7 | Excitement,Immobility/stupor,Mutism,Staring,Grimacing,Echopraxia/echolalia,Verbigeration | 97.5 | 96.0 | 98.9 |
| 7 | Excitement,Immobility/stupor,Mutism,Staring,Grimacing,Stereotypy,Verbigeration | 97.5 | 96.0 | 98.9 |
| 7 | Excitement,Immobility/stupor,Mutism,Staring,Grimacing,Verbigeration,Withdrawal | 97.5 | 96.0 | 98.9 |
| 7 | Excitement,Immobility/stupor,Mutism,Staring,Mannerisms,Verbigeration,Withdrawal | 97.5 | 96.0 | 98.9 |
| 7 | Excitement,Immobility/stupor,Mutism,Staring,Stereotypy,Mannerisms,Withdrawal | 97.5 | 96.0 | 98.9 |
| 7 | Excitement,Immobility/stupor,Posturing/catalepsy,Echopraxia/echolalia,Mannerisms,Verbigeration,Rigidity | 97.5 | 96.0 | 98.9 |
| 7 | Excitement,Immobility/stupor,Posturing/catalepsy,Echopraxia/echolalia,Stereotypy,Verbigeration,Rigidity | 97.5 | 96.0 | 98.9 |
| 7 | Excitement,Immobility/stupor,Posturing/catalepsy,Echopraxia/echolalia,Verbigeration,Rigidity,Waxy flexibility | 97.5 | 96.0 | 98.9 |
| 7 | Excitement,Immobility/stupor,Posturing/catalepsy,Grimacing,Echopraxia/echolalia,Mannerisms,Rigidity | 97.5 | 96.0 | 98.9 |
| 7 | Excitement,Immobility/stupor,Posturing/catalepsy,Grimacing,Echopraxia/echolalia,Rigidity,Waxy flexibility | 97.5 | 96.0 | 98.9 |
| 7 | Excitement,Immobility/stupor,Posturing/catalepsy,Grimacing,Mannerisms,Verbigeration,Rigidity | 97.5 | 96.0 | 98.9 |
| 7 | Excitement,Immobility/stupor,Posturing/catalepsy,Grimacing,Stereotypy,Negativism,Withdrawal | 97.5 | 96.0 | 98.9 |
| 7 | Excitement,Immobility/stupor,Posturing/catalepsy,Stereotypy,Mannerisms,Negativism,Withdrawal | 97.5 | 96.0 | 98.9 |
| 7 | Excitement,Immobility/stupor,Staring,Echopraxia/echolalia,Mannerisms,Verbigeration,Withdrawal | 97.5 | 96.0 | 98.9 |
| 7 | Excitement,Immobility/stupor,Staring,Echopraxia/echolalia,Stereotypy,Mannerisms,Waxy flexibility | 97.5 | 96.0 | 98.9 |
| 7 | Excitement,Immobility/stupor,Staring,Echopraxia/echolalia,Stereotypy,Verbigeration,Waxy flexibility | 97.5 | 96.0 | 98.9 |
| 7 | Excitement,Immobility/stupor,Staring,Echopraxia/echolalia,Stereotypy,Verbigeration,Withdrawal | 97.5 | 96.0 | 98.9 |
| 7 | Excitement,Immobility/stupor,Staring,Grimacing,Echopraxia/echolalia,Stereotypy,Waxy flexibility | 97.5 | 96.0 | 98.9 |
| 7 | Excitement,Immobility/stupor,Staring,Grimacing,Echopraxia/echolalia,Verbigeration,Waxy flexibility | 97.5 | 96.0 | 98.9 |
| 7 | Excitement,Immobility/stupor,Staring,Grimacing,Mannerisms,Verbigeration,Waxy flexibility | 97.5 | 96.0 | 98.9 |
| 7 | Excitement,Immobility/stupor,Staring,Grimacing,Mannerisms,Waxy flexibility,Withdrawal | 97.5 | 96.0 | 98.9 |
| 7 | Excitement,Immobility/stupor,Staring,Grimacing,Stereotypy,Mannerisms,Negativism | 97.5 | 96.0 | 98.9 |
| 7 | Excitement,Immobility/stupor,Staring,Posturing/catalepsy,Echopraxia/echolalia,Stereotypy,Mannerisms | 97.5 | 96.0 | 98.9 |
| 7 | Excitement,Immobility/stupor,Staring,Posturing/catalepsy,Echopraxia/echolalia,Stereotypy,Verbigeration | 97.5 | 96.0 | 98.9 |
| 7 | Excitement,Immobility/stupor,Staring,Posturing/catalepsy,Echopraxia/echolalia,Stereotypy,Waxy flexibility | 97.5 | 96.0 | 98.9 |
| 7 | Excitement,Immobility/stupor,Staring,Posturing/catalepsy,Grimacing,Echopraxia/echolalia,Stereotypy | 97.5 | 96.0 | 98.9 |
| 7 | Excitement,Immobility/stupor,Staring,Posturing/catalepsy,Grimacing,Stereotypy,Verbigeration | 97.5 | 96.0 | 98.9 |
| 7 | Excitement,Immobility/stupor,Staring,Stereotypy,Mannerisms,Verbigeration,Waxy flexibility | 97.5 | 96.0 | 98.9 |
| 7 | Excitement,Immobility/stupor,Staring,Stereotypy,Mannerisms,Verbigeration,Withdrawal | 97.5 | 96.0 | 98.9 |
| 7 | Excitement,Immobility/stupor,Staring,Stereotypy,Mannerisms,Waxy flexibility,Withdrawal | 97.5 | 96.0 | 98.9 |
| 7 | Excitement,Immobility/stupor,Staring,Stereotypy,Verbigeration,Waxy flexibility,Withdrawal | 97.5 | 96.0 | 98.9 |
| 7 | Excitement,Immobility/stupor,Stereotypy,Mannerisms,Rigidity,Negativism,Waxy flexibility | 97.5 | 96.0 | 98.9 |
| 7 | Excitement,Immobility/stupor,Stereotypy,Mannerisms,Rigidity,Waxy flexibility,Withdrawal | 97.5 | 96.0 | 98.9 |
| 7 | Excitement,Mutism,Staring,Echopraxia/echolalia,Stereotypy,Mannerisms,Verbigeration | 97.5 | 96.0 | 98.9 |
| 7 | Excitement,Staring,Echopraxia/echolalia,Mannerisms,Verbigeration,Rigidity,Negativism | 97.5 | 96.0 | 98.9 |
| 7 | Excitement,Staring,Echopraxia/echolalia,Stereotypy,Mannerisms,Waxy flexibility,Withdrawal | 97.5 | 96.0 | 98.9 |
| 7 | Excitement,Staring,Grimacing,Echopraxia/echolalia,Mannerisms,Rigidity,Waxy flexibility | 97.5 | 96.0 | 98.9 |
| 7 | Excitement,Staring,Grimacing,Echopraxia/echolalia,Rigidity,Negativism,Waxy flexibility | 97.5 | 96.0 | 98.9 |
| 7 | Excitement,Staring,Grimacing,Echopraxia/echolalia,Stereotypy,Mannerisms,Withdrawal | 97.5 | 96.0 | 98.9 |
| 7 | Excitement,Staring,Grimacing,Echopraxia/echolalia,Stereotypy,Rigidity,Negativism | 97.5 | 96.0 | 98.9 |
| 7 | Excitement,Staring,Grimacing,Echopraxia/echolalia,Stereotypy,Rigidity,Waxy flexibility | 97.5 | 96.0 | 98.9 |
| 7 | Excitement,Staring,Grimacing,Echopraxia/echolalia,Verbigeration,Negativism,Withdrawal | 97.5 | 96.0 | 98.9 |
| 7 | Excitement,Staring,Grimacing,Mannerisms,Rigidity,Negativism,Waxy flexibility | 97.5 | 96.0 | 98.9 |
| 7 | Excitement,Staring,Grimacing,Mannerisms,Verbigeration,Rigidity,Waxy flexibility | 97.5 | 96.0 | 98.9 |
| 7 | Excitement,Staring,Grimacing,Stereotypy,Mannerisms,Rigidity,Waxy flexibility | 97.5 | 96.0 | 98.9 |
| 7 | Excitement,Staring,Grimacing,Stereotypy,Negativism,Waxy flexibility,Withdrawal | 97.5 | 96.0 | 98.9 |
| 7 | Excitement,Staring,Grimacing,Stereotypy,Rigidity,Negativism,Waxy flexibility | 97.5 | 96.0 | 98.9 |
| 7 | Excitement,Staring,Grimacing,Stereotypy,Verbigeration,Rigidity,Negativism | 97.5 | 96.0 | 98.9 |
| 7 | Excitement,Staring,Grimacing,Stereotypy,Verbigeration,Rigidity,Waxy flexibility | 97.5 | 96.0 | 98.9 |
| 7 | Excitement,Staring,Grimacing,Verbigeration,Negativism,Waxy flexibility,Withdrawal | 97.5 | 96.0 | 98.9 |
| 7 | Excitement,Staring,Posturing/catalepsy,Echopraxia/echolalia,Mannerisms,Negativism,Waxy flexibility | 97.5 | 96.0 | 98.9 |
| 7 | Excitement,Staring,Posturing/catalepsy,Echopraxia/echolalia,Mannerisms,Verbigeration,Withdrawal | 97.5 | 96.0 | 98.9 |
| 7 | Excitement,Staring,Posturing/catalepsy,Echopraxia/echolalia,Stereotypy,Mannerisms,Withdrawal | 97.5 | 96.0 | 98.9 |
| 7 | Excitement,Staring,Posturing/catalepsy,Grimacing,Echopraxia/echolalia,Stereotypy,Rigidity | 97.5 | 96.0 | 98.9 |
| 7 | Excitement,Staring,Posturing/catalepsy,Grimacing,Echopraxia/echolalia,Stereotypy,Withdrawal | 97.5 | 96.0 | 98.9 |
| 7 | Excitement,Staring,Posturing/catalepsy,Grimacing,Echopraxia/echolalia,Verbigeration,Rigidity | 97.5 | 96.0 | 98.9 |
| 7 | Excitement,Staring,Posturing/catalepsy,Grimacing,Echopraxia/echolalia,Verbigeration,Withdrawal | 97.5 | 96.0 | 98.9 |
| 7 | Excitement,Staring,Posturing/catalepsy,Grimacing,Stereotypy,Mannerisms,Rigidity | 97.5 | 96.0 | 98.9 |
| 7 | Excitement,Staring,Posturing/catalepsy,Grimacing,Stereotypy,Mannerisms,Withdrawal | 97.5 | 96.0 | 98.9 |
| 7 | Excitement,Staring,Posturing/catalepsy,Grimacing,Stereotypy,Rigidity,Negativism | 97.5 | 96.0 | 98.9 |
| 7 | Excitement,Staring,Posturing/catalepsy,Grimacing,Stereotypy,Verbigeration,Rigidity | 97.5 | 96.0 | 98.9 |
| 7 | Excitement,Staring,Posturing/catalepsy,Grimacing,Verbigeration,Rigidity,Negativism | 97.5 | 96.0 | 98.9 |
| 7 | Excitement,Staring,Posturing/catalepsy,Mannerisms,Verbigeration,Negativism,Waxy flexibility | 97.5 | 96.0 | 98.9 |
| 7 | Excitement,Staring,Posturing/catalepsy,Stereotypy,Rigidity,Negativism,Waxy flexibility | 97.5 | 96.0 | 98.9 |
| 7 | Excitement,Staring,Posturing/catalepsy,Verbigeration,Rigidity,Negativism,Waxy flexibility | 97.5 | 96.0 | 98.9 |
| 7 | Immobility/stupor,Echopraxia/echolalia,Stereotypy,Mannerisms,Rigidity,Negativism,Waxy flexibility | 97.5 | 96.0 | 98.9 |
| 7 | Immobility/stupor,Echopraxia/echolalia,Stereotypy,Mannerisms,Verbigeration,Rigidity,Negativism | 97.5 | 96.0 | 98.9 |
| 7 | Immobility/stupor,Grimacing,Echopraxia/echolalia,Mannerisms,Rigidity,Negativism,Waxy flexibility | 97.5 | 96.0 | 98.9 |
| 7 | Immobility/stupor,Grimacing,Echopraxia/echolalia,Stereotypy,Mannerisms,Rigidity,Negativism | 97.5 | 96.0 | 98.9 |
| 7 | Immobility/stupor,Grimacing,Echopraxia/echolalia,Stereotypy,Rigidity,Negativism,Withdrawal | 97.5 | 96.0 | 98.9 |
| 7 | Immobility/stupor,Grimacing,Mannerisms,Rigidity,Negativism,Waxy flexibility,Withdrawal | 97.5 | 96.0 | 98.9 |
| 7 | Immobility/stupor,Grimacing,Stereotypy,Mannerisms,Rigidity,Negativism,Waxy flexibility | 97.5 | 96.0 | 98.9 |
| 7 | Immobility/stupor,Grimacing,Stereotypy,Mannerisms,Verbigeration,Rigidity,Negativism | 97.5 | 96.0 | 98.9 |
| 7 | Immobility/stupor,Mannerisms,Verbigeration,Rigidity,Negativism,Waxy flexibility,Withdrawal | 97.5 | 96.0 | 98.9 |
| 7 | Immobility/stupor,Mutism,Echopraxia/echolalia,Mannerisms,Rigidity,Negativism,Waxy flexibility | 97.5 | 96.0 | 98.9 |
| 7 | Immobility/stupor,Mutism,Grimacing,Mannerisms,Verbigeration,Rigidity,Negativism | 97.5 | 96.0 | 98.9 |
| 7 | Immobility/stupor,Mutism,Grimacing,Stereotypy,Mannerisms,Rigidity,Negativism | 97.5 | 96.0 | 98.9 |
| 7 | Immobility/stupor,Mutism,Grimacing,Stereotypy,Verbigeration,Rigidity,Negativism | 97.5 | 96.0 | 98.9 |
| 7 | Immobility/stupor,Mutism,Mannerisms,Rigidity,Negativism,Waxy flexibility,Withdrawal | 97.5 | 96.0 | 98.9 |
| 7 | Immobility/stupor,Mutism,Mannerisms,Verbigeration,Rigidity,Negativism,Waxy flexibility | 97.5 | 96.0 | 98.9 |
| 7 | Immobility/stupor,Mutism,Posturing/catalepsy,Grimacing,Mannerisms,Rigidity,Negativism | 97.5 | 96.0 | 98.9 |
| 7 | Immobility/stupor,Mutism,Staring,Echopraxia/echolalia,Mannerisms,Verbigeration,Negativism | 97.5 | 96.0 | 98.9 |
| 7 | Immobility/stupor,Mutism,Staring,Echopraxia/echolalia,Mannerisms,Verbigeration,Rigidity | 97.5 | 96.0 | 98.9 |
| 7 | Immobility/stupor,Mutism,Staring,Echopraxia/echolalia,Stereotypy,Mannerisms,Rigidity | 97.5 | 96.0 | 98.9 |
| 7 | Immobility/stupor,Mutism,Staring,Echopraxia/echolalia,Stereotypy,Negativism,Withdrawal | 97.5 | 96.0 | 98.9 |
| 7 | Immobility/stupor,Mutism,Staring,Echopraxia/echolalia,Stereotypy,Rigidity,Negativism | 97.5 | 96.0 | 98.9 |
| 7 | Immobility/stupor,Mutism,Staring,Echopraxia/echolalia,Stereotypy,Rigidity,Withdrawal | 97.5 | 96.0 | 98.9 |
| 7 | Immobility/stupor,Mutism,Staring,Grimacing,Echopraxia/echolalia,Mannerisms,Rigidity | 97.5 | 96.0 | 98.9 |
| 7 | Immobility/stupor,Mutism,Staring,Grimacing,Stereotypy,Verbigeration,Negativism | 97.5 | 96.0 | 98.9 |
| 7 | Immobility/stupor,Mutism,Staring,Grimacing,Verbigeration,Rigidity,Negativism | 97.5 | 96.0 | 98.9 |
| 7 | Immobility/stupor,Mutism,Staring,Mannerisms,Verbigeration,Rigidity,Waxy flexibility | 97.5 | 96.0 | 98.9 |
| 7 | Immobility/stupor,Mutism,Staring,Posturing/catalepsy,Echopraxia/echolalia,Mannerisms,Withdrawal | 97.5 | 96.0 | 98.9 |
| 7 | Immobility/stupor,Mutism,Staring,Posturing/catalepsy,Echopraxia/echolalia,Rigidity,Negativism | 97.5 | 96.0 | 98.9 |
| 7 | Immobility/stupor,Mutism,Staring,Posturing/catalepsy,Echopraxia/echolalia,Stereotypy,Negativism | 97.5 | 96.0 | 98.9 |
| 7 | Immobility/stupor,Mutism,Staring,Posturing/catalepsy,Echopraxia/echolalia,Verbigeration,Negativism | 97.5 | 96.0 | 98.9 |
| 7 | Immobility/stupor,Mutism,Staring,Posturing/catalepsy,Grimacing,Mannerisms,Rigidity | 97.5 | 96.0 | 98.9 |
| 7 | Immobility/stupor,Mutism,Staring,Posturing/catalepsy,Grimacing,Negativism,Withdrawal | 97.5 | 96.0 | 98.9 |
| 7 | Immobility/stupor,Mutism,Staring,Posturing/catalepsy,Mannerisms,Negativism,Waxy flexibility | 97.5 | 96.0 | 98.9 |
| 7 | Immobility/stupor,Mutism,Staring,Posturing/catalepsy,Rigidity,Negativism,Withdrawal | 97.5 | 96.0 | 98.9 |
| 7 | Immobility/stupor,Mutism,Staring,Posturing/catalepsy,Stereotypy,Mannerisms,Withdrawal | 97.5 | 96.0 | 98.9 |
| 7 | Immobility/stupor,Mutism,Staring,Posturing/catalepsy,Verbigeration,Rigidity,Negativism | 97.5 | 96.0 | 98.9 |
| 7 | Immobility/stupor,Mutism,Staring,Stereotypy,Verbigeration,Rigidity,Waxy flexibility | 97.5 | 96.0 | 98.9 |
| 7 | Immobility/stupor,Posturing/catalepsy,Echopraxia/echolalia,Mannerisms,Verbigeration,Rigidity,Negativism | 97.5 | 96.0 | 98.9 |
| 7 | Immobility/stupor,Posturing/catalepsy,Grimacing,Mannerisms,Verbigeration,Rigidity,Negativism | 97.5 | 96.0 | 98.9 |
| 7 | Immobility/stupor,Posturing/catalepsy,Grimacing,Stereotypy,Mannerisms,Rigidity,Negativism | 97.5 | 96.0 | 98.9 |
| 7 | Immobility/stupor,Staring,Echopraxia/echolalia,Mannerisms,Verbigeration,Negativism,Waxy flexibility | 97.5 | 96.0 | 98.9 |
| 7 | Immobility/stupor,Staring,Echopraxia/echolalia,Stereotypy,Verbigeration,Negativism,Withdrawal | 97.5 | 96.0 | 98.9 |
| 7 | Immobility/stupor,Staring,Grimacing,Echopraxia/echolalia,Mannerisms,Verbigeration,Negativism | 97.5 | 96.0 | 98.9 |
| 7 | Immobility/stupor,Staring,Grimacing,Echopraxia/echolalia,Stereotypy,Mannerisms,Rigidity | 97.5 | 96.0 | 98.9 |
| 7 | Immobility/stupor,Staring,Grimacing,Echopraxia/echolalia,Stereotypy,Mannerisms,Withdrawal | 97.5 | 96.0 | 98.9 |
| 7 | Immobility/stupor,Staring,Grimacing,Echopraxia/echolalia,Stereotypy,Rigidity,Negativism | 97.5 | 96.0 | 98.9 |
| 7 | Immobility/stupor,Staring,Grimacing,Echopraxia/echolalia,Stereotypy,Rigidity,Withdrawal | 97.5 | 96.0 | 98.9 |
| 7 | Immobility/stupor,Staring,Grimacing,Stereotypy,Verbigeration,Rigidity,Waxy flexibility | 97.5 | 96.0 | 98.9 |
| 7 | Immobility/stupor,Staring,Posturing/catalepsy,Echopraxia/echolalia,Stereotypy,Verbigeration,Negativism | 97.5 | 96.0 | 98.9 |
| 7 | Immobility/stupor,Staring,Posturing/catalepsy,Echopraxia/echolalia,Verbigeration,Negativism,Withdrawal | 97.5 | 96.0 | 98.9 |
| 7 | Immobility/stupor,Staring,Posturing/catalepsy,Grimacing,Mannerisms,Verbigeration,Rigidity | 97.5 | 96.0 | 98.9 |
| 7 | Immobility/stupor,Staring,Posturing/catalepsy,Grimacing,Rigidity,Negativism,Withdrawal | 97.5 | 96.0 | 98.9 |
| 7 | Immobility/stupor,Staring,Posturing/catalepsy,Grimacing,Stereotypy,Mannerisms,Withdrawal | 97.5 | 96.0 | 98.9 |
| 7 | Immobility/stupor,Staring,Posturing/catalepsy,Grimacing,Stereotypy,Rigidity,Negativism | 97.5 | 96.0 | 98.9 |
| 7 | Immobility/stupor,Staring,Posturing/catalepsy,Grimacing,Stereotypy,Verbigeration,Rigidity | 97.5 | 96.0 | 98.9 |
| 7 | Immobility/stupor,Staring,Posturing/catalepsy,Grimacing,Verbigeration,Negativism,Withdrawal | 97.5 | 96.0 | 98.9 |
| 7 | Immobility/stupor,Staring,Posturing/catalepsy,Grimacing,Verbigeration,Rigidity,Negativism | 97.5 | 96.0 | 98.9 |
| 7 | Immobility/stupor,Staring,Posturing/catalepsy,Mannerisms,Verbigeration,Negativism,Waxy flexibility | 97.5 | 96.0 | 98.9 |
| 7 | Immobility/stupor,Staring,Posturing/catalepsy,Stereotypy,Verbigeration,Negativism,Waxy flexibility | 97.5 | 96.0 | 98.9 |
| 7 | Immobility/stupor,Staring,Stereotypy,Mannerisms,Verbigeration,Rigidity,Waxy flexibility | 97.5 | 96.0 | 98.9 |
| 7 | Immobility/stupor,Staring,Stereotypy,Verbigeration,Negativism,Waxy flexibility,Withdrawal | 97.5 | 96.0 | 98.9 |
| 7 | Immobility/stupor,Stereotypy,Verbigeration,Rigidity,Negativism,Waxy flexibility,Withdrawal | 97.5 | 96.0 | 98.9 |
| 7 | Mutism,Staring,Echopraxia/echolalia,Mannerisms,Verbigeration,Rigidity,Waxy flexibility | 97.5 | 96.0 | 98.9 |
| 7 | Mutism,Staring,Echopraxia/echolalia,Stereotypy,Mannerisms,Waxy flexibility,Withdrawal | 97.5 | 96.0 | 98.9 |
| 7 | Mutism,Staring,Echopraxia/echolalia,Verbigeration,Negativism,Waxy flexibility,Withdrawal | 97.5 | 96.0 | 98.9 |
| 7 | Mutism,Staring,Grimacing,Echopraxia/echolalia,Mannerisms,Verbigeration,Waxy flexibility | 97.5 | 96.0 | 98.9 |
| 7 | Mutism,Staring,Grimacing,Echopraxia/echolalia,Mannerisms,Verbigeration,Withdrawal | 97.5 | 96.0 | 98.9 |
| 7 | Mutism,Staring,Grimacing,Echopraxia/echolalia,Stereotypy,Negativism,Waxy flexibility | 97.5 | 96.0 | 98.9 |
| 7 | Mutism,Staring,Grimacing,Echopraxia/echolalia,Verbigeration,Rigidity,Waxy flexibility | 97.5 | 96.0 | 98.9 |
| 7 | Mutism,Staring,Grimacing,Mannerisms,Verbigeration,Waxy flexibility,Withdrawal | 97.5 | 96.0 | 98.9 |
| 7 | Mutism,Staring,Grimacing,Stereotypy,Negativism,Waxy flexibility,Withdrawal | 97.5 | 96.0 | 98.9 |
| 7 | Mutism,Staring,Grimacing,Stereotypy,Rigidity,Negativism,Waxy flexibility | 97.5 | 96.0 | 98.9 |
| 7 | Mutism,Staring,Grimacing,Verbigeration,Rigidity,Negativism,Waxy flexibility | 97.5 | 96.0 | 98.9 |
| 7 | Mutism,Staring,Grimacing,Verbigeration,Rigidity,Waxy flexibility,Withdrawal | 97.5 | 96.0 | 98.9 |
| 7 | Mutism,Staring,Posturing/catalepsy,Echopraxia/echolalia,Rigidity,Negativism,Waxy flexibility | 97.5 | 96.0 | 98.9 |
| 7 | Mutism,Staring,Posturing/catalepsy,Grimacing,Echopraxia/echolalia,Stereotypy,Rigidity | 97.5 | 96.0 | 98.9 |
| 7 | Mutism,Staring,Posturing/catalepsy,Grimacing,Negativism,Waxy flexibility,Withdrawal | 97.5 | 96.0 | 98.9 |
| 7 | Mutism,Staring,Posturing/catalepsy,Grimacing,Stereotypy,Mannerisms,Verbigeration | 97.5 | 96.0 | 98.9 |
| 7 | Mutism,Staring,Posturing/catalepsy,Mannerisms,Verbigeration,Waxy flexibility,Withdrawal | 97.5 | 96.0 | 98.9 |
| 7 | Mutism,Staring,Posturing/catalepsy,Rigidity,Negativism,Waxy flexibility,Withdrawal | 97.5 | 96.0 | 98.9 |
| 7 | Mutism,Staring,Posturing/catalepsy,Stereotypy,Mannerisms,Rigidity,Waxy flexibility | 97.5 | 96.0 | 98.9 |
| 7 | Mutism,Staring,Posturing/catalepsy,Stereotypy,Rigidity,Negativism,Waxy flexibility | 97.5 | 96.0 | 98.9 |
| 7 | Mutism,Staring,Posturing/catalepsy,Stereotypy,Rigidity,Waxy flexibility,Withdrawal | 97.5 | 96.0 | 98.9 |
| 7 | Staring,Echopraxia/echolalia,Mannerisms,Verbigeration,Negativism,Waxy flexibility,Withdrawal | 97.5 | 96.0 | 98.9 |
| 7 | Staring,Echopraxia/echolalia,Stereotypy,Mannerisms,Rigidity,Waxy flexibility,Withdrawal | 97.5 | 96.0 | 98.9 |
| 7 | Staring,Echopraxia/echolalia,Stereotypy,Mannerisms,Verbigeration,Rigidity,Negativism | 97.5 | 96.0 | 98.9 |
| 7 | Staring,Echopraxia/echolalia,Verbigeration,Rigidity,Negativism,Waxy flexibility,Withdrawal | 97.5 | 96.0 | 98.9 |
| 7 | Staring,Grimacing,Echopraxia/echolalia,Mannerisms,Verbigeration,Negativism,Withdrawal | 97.5 | 96.0 | 98.9 |
| 7 | Staring,Grimacing,Echopraxia/echolalia,Mannerisms,Verbigeration,Rigidity,Withdrawal | 97.5 | 96.0 | 98.9 |
| 7 | Staring,Grimacing,Echopraxia/echolalia,Stereotypy,Mannerisms,Rigidity,Negativism | 97.5 | 96.0 | 98.9 |
| 7 | Staring,Grimacing,Echopraxia/echolalia,Verbigeration,Rigidity,Negativism,Withdrawal | 97.5 | 96.0 | 98.9 |
| 7 | Staring,Grimacing,Mannerisms,Verbigeration,Negativism,Waxy flexibility,Withdrawal | 97.5 | 96.0 | 98.9 |
| 7 | Staring,Grimacing,Stereotypy,Mannerisms,Rigidity,Negativism,Waxy flexibility | 97.5 | 96.0 | 98.9 |
| 7 | Staring,Grimacing,Stereotypy,Mannerisms,Verbigeration,Rigidity,Negativism | 97.5 | 96.0 | 98.9 |
| 7 | Staring,Grimacing,Stereotypy,Rigidity,Negativism,Waxy flexibility,Withdrawal | 97.5 | 96.0 | 98.9 |
| 7 | Staring,Grimacing,Stereotypy,Verbigeration,Rigidity,Waxy flexibility,Withdrawal | 97.5 | 96.0 | 98.9 |
| 7 | Staring,Posturing/catalepsy,Echopraxia/echolalia,Mannerisms,Verbigeration,Rigidity,Negativism | 97.5 | 96.0 | 98.9 |
| 7 | Staring,Posturing/catalepsy,Echopraxia/echolalia,Rigidity,Negativism,Waxy flexibility,Withdrawal | 97.5 | 96.0 | 98.9 |
| 7 | Staring,Posturing/catalepsy,Grimacing,Echopraxia/echolalia,Stereotypy,Mannerisms,Withdrawal | 97.5 | 96.0 | 98.9 |
| 7 | Staring,Posturing/catalepsy,Grimacing,Echopraxia/echolalia,Stereotypy,Negativism,Withdrawal | 97.5 | 96.0 | 98.9 |
| 7 | Staring,Posturing/catalepsy,Grimacing,Mannerisms,Rigidity,Waxy flexibility,Withdrawal | 97.5 | 96.0 | 98.9 |
| 7 | Staring,Posturing/catalepsy,Grimacing,Stereotypy,Mannerisms,Rigidity,Negativism | 97.5 | 96.0 | 98.9 |
| 7 | Staring,Posturing/catalepsy,Grimacing,Stereotypy,Verbigeration,Rigidity,Withdrawal | 97.5 | 96.0 | 98.9 |
| 7 | Staring,Posturing/catalepsy,Grimacing,Verbigeration,Rigidity,Negativism,Withdrawal | 97.5 | 96.0 | 98.9 |
| 7 | Staring,Posturing/catalepsy,Stereotypy,Mannerisms,Rigidity,Waxy flexibility,Withdrawal | 97.5 | 96.0 | 98.9 |
| 7 | Staring,Posturing/catalepsy,Stereotypy,Rigidity,Negativism,Waxy flexibility,Withdrawal | 97.5 | 96.0 | 98.9 |
| 7 | Staring,Stereotypy,Mannerisms,Verbigeration,Rigidity,Waxy flexibility,Withdrawal | 97.5 | 96.0 | 98.9 |
| 7 | Excitement,Immobility/stupor,Echopraxia/echolalia,Stereotypy,Mannerisms,Negativism,Withdrawal | 97.3 | 95.7 | 98.7 |
| 7 | Excitement,Immobility/stupor,Echopraxia/echolalia,Stereotypy,Negativism,Waxy flexibility,Withdrawal | 97.3 | 95.7 | 98.7 |
| 7 | Excitement,Immobility/stupor,Echopraxia/echolalia,Stereotypy,Verbigeration,Rigidity,Waxy flexibility | 97.3 | 95.7 | 98.7 |
| 7 | Excitement,Immobility/stupor,Grimacing,Echopraxia/echolalia,Stereotypy,Negativism,Withdrawal | 97.3 | 95.7 | 98.7 |
| 7 | Excitement,Immobility/stupor,Grimacing,Echopraxia/echolalia,Verbigeration,Rigidity,Waxy flexibility | 97.3 | 95.7 | 98.7 |
| 7 | Excitement,Immobility/stupor,Grimacing,Stereotypy,Mannerisms,Rigidity,Negativism | 97.3 | 95.7 | 98.7 |
| 7 | Excitement,Immobility/stupor,Grimacing,Stereotypy,Mannerisms,Rigidity,Withdrawal | 97.3 | 95.7 | 98.7 |
| 7 | Excitement,Immobility/stupor,Grimacing,Verbigeration,Rigidity,Waxy flexibility,Withdrawal | 97.3 | 95.7 | 98.7 |
| 7 | Excitement,Immobility/stupor,Mutism,Echopraxia/echolalia,Negativism,Waxy flexibility,Withdrawal | 97.3 | 95.7 | 98.7 |
| 7 | Excitement,Immobility/stupor,Mutism,Posturing/catalepsy,Echopraxia/echolalia,Mannerisms,Negativism | 97.3 | 95.7 | 98.7 |
| 7 | Excitement,Immobility/stupor,Mutism,Posturing/catalepsy,Echopraxia/echolalia,Stereotypy,Withdrawal | 97.3 | 95.7 | 98.7 |
| 7 | Excitement,Immobility/stupor,Mutism,Posturing/catalepsy,Echopraxia/echolalia,Waxy flexibility,Withdrawal | 97.3 | 95.7 | 98.7 |
| 7 | Excitement,Immobility/stupor,Mutism,Posturing/catalepsy,Grimacing,Verbigeration,Withdrawal | 97.3 | 95.7 | 98.7 |
| 7 | Excitement,Immobility/stupor,Mutism,Posturing/catalepsy,Stereotypy,Negativism,Waxy flexibility | 97.3 | 95.7 | 98.7 |
| 7 | Excitement,Immobility/stupor,Mutism,Staring,Echopraxia/echolalia,Mannerisms,Verbigeration | 97.3 | 95.7 | 98.7 |
| 7 | Excitement,Immobility/stupor,Mutism,Staring,Grimacing,Echopraxia/echolalia,Mannerisms | 97.3 | 95.7 | 98.7 |
| 7 | Excitement,Immobility/stupor,Mutism,Staring,Grimacing,Mannerisms,Withdrawal | 97.3 | 95.7 | 98.7 |
| 7 | Excitement,Immobility/stupor,Mutism,Staring,Stereotypy,Mannerisms,Verbigeration | 97.3 | 95.7 | 98.7 |
| 7 | Excitement,Immobility/stupor,Mutism,Stereotypy,Mannerisms,Rigidity,Waxy flexibility | 97.3 | 95.7 | 98.7 |
| 7 | Excitement,Immobility/stupor,Mutism,Stereotypy,Negativism,Waxy flexibility,Withdrawal | 97.3 | 95.7 | 98.7 |
| 7 | Excitement,Immobility/stupor,Mutism,Stereotypy,Verbigeration,Negativism,Waxy flexibility | 97.3 | 95.7 | 98.7 |
| 7 | Excitement,Immobility/stupor,Posturing/catalepsy,Grimacing,Echopraxia/echolalia,Negativism,Waxy flexibility | 97.3 | 95.7 | 98.7 |
| 7 | Excitement,Immobility/stupor,Posturing/catalepsy,Grimacing,Echopraxia/echolalia,Stereotypy,Rigidity | 97.3 | 95.7 | 98.7 |
| 7 | Excitement,Immobility/stupor,Posturing/catalepsy,Grimacing,Echopraxia/echolalia,Verbigeration,Negativism | 97.3 | 95.7 | 98.7 |
| 7 | Excitement,Immobility/stupor,Posturing/catalepsy,Grimacing,Mannerisms,Verbigeration,Negativism | 97.3 | 95.7 | 98.7 |
| 7 | Excitement,Immobility/stupor,Posturing/catalepsy,Grimacing,Negativism,Waxy flexibility,Withdrawal | 97.3 | 95.7 | 98.7 |
| 7 | Excitement,Immobility/stupor,Posturing/catalepsy,Grimacing,Stereotypy,Verbigeration,Rigidity | 97.3 | 95.7 | 98.7 |
| 7 | Excitement,Immobility/stupor,Posturing/catalepsy,Grimacing,Verbigeration,Negativism,Waxy flexibility | 97.3 | 95.7 | 98.7 |
| 7 | Excitement,Immobility/stupor,Posturing/catalepsy,Grimacing,Verbigeration,Rigidity,Waxy flexibility | 97.3 | 95.7 | 98.7 |
| 7 | Excitement,Immobility/stupor,Posturing/catalepsy,Mannerisms,Negativism,Waxy flexibility,Withdrawal | 97.3 | 95.7 | 98.7 |
| 7 | Excitement,Immobility/stupor,Posturing/catalepsy,Stereotypy,Negativism,Waxy flexibility,Withdrawal | 97.3 | 95.7 | 98.7 |
| 7 | Excitement,Immobility/stupor,Staring,Grimacing,Echopraxia/echolalia,Verbigeration,Withdrawal | 97.3 | 95.7 | 98.7 |
| 7 | Excitement,Immobility/stupor,Staring,Grimacing,Mannerisms,Verbigeration,Withdrawal | 97.3 | 95.7 | 98.7 |
| 7 | Excitement,Immobility/stupor,Staring,Grimacing,Stereotypy,Verbigeration,Waxy flexibility | 97.3 | 95.7 | 98.7 |
| 7 | Excitement,Immobility/stupor,Staring,Grimacing,Stereotypy,Verbigeration,Withdrawal | 97.3 | 95.7 | 98.7 |
| 7 | Excitement,Immobility/stupor,Staring,Grimacing,Stereotypy,Waxy flexibility,Withdrawal | 97.3 | 95.7 | 98.7 |
| 7 | Excitement,Immobility/stupor,Staring,Grimacing,Verbigeration,Waxy flexibility,Withdrawal | 97.3 | 95.7 | 98.7 |
| 7 | Excitement,Immobility/stupor,Staring,Posturing/catalepsy,Grimacing,Stereotypy,Mannerisms | 97.3 | 95.7 | 98.7 |
| 7 | Excitement,Immobility/stupor,Staring,Posturing/catalepsy,Grimacing,Stereotypy,Waxy flexibility | 97.3 | 95.7 | 98.7 |
| 7 | Excitement,Immobility/stupor,Staring,Posturing/catalepsy,Stereotypy,Mannerisms,Waxy flexibility | 97.3 | 95.7 | 98.7 |
| 7 | Excitement,Immobility/stupor,Stereotypy,Mannerisms,Verbigeration,Negativism,Withdrawal | 97.3 | 95.7 | 98.7 |
| 7 | Excitement,Mutism,Staring,Grimacing,Echopraxia/echolalia,Mannerisms,Verbigeration | 97.3 | 95.7 | 98.7 |
| 7 | Excitement,Mutism,Staring,Grimacing,Stereotypy,Mannerisms,Verbigeration | 97.3 | 95.7 | 98.7 |
| 7 | Excitement,Staring,Grimacing,Echopraxia/echolalia,Mannerisms,Rigidity,Negativism | 97.3 | 95.7 | 98.7 |
| 7 | Excitement,Staring,Grimacing,Echopraxia/echolalia,Stereotypy,Mannerisms,Rigidity | 97.3 | 95.7 | 98.7 |
| 7 | Excitement,Staring,Grimacing,Echopraxia/echolalia,Stereotypy,Waxy flexibility,Withdrawal | 97.3 | 95.7 | 98.7 |
| 7 | Excitement,Staring,Grimacing,Echopraxia/echolalia,Verbigeration,Rigidity,Waxy flexibility | 97.3 | 95.7 | 98.7 |
| 7 | Excitement,Staring,Grimacing,Mannerisms,Verbigeration,Rigidity,Negativism | 97.3 | 95.7 | 98.7 |
| 7 | Excitement,Staring,Grimacing,Mannerisms,Verbigeration,Waxy flexibility,Withdrawal | 97.3 | 95.7 | 98.7 |
| 7 | Excitement,Staring,Grimacing,Stereotypy,Mannerisms,Rigidity,Negativism | 97.3 | 95.7 | 98.7 |
| 7 | Excitement,Staring,Grimacing,Stereotypy,Mannerisms,Verbigeration,Rigidity | 97.3 | 95.7 | 98.7 |
| 7 | Excitement,Staring,Grimacing,Verbigeration,Rigidity,Negativism,Waxy flexibility | 97.3 | 95.7 | 98.7 |
| 7 | Excitement,Staring,Posturing/catalepsy,Echopraxia/echolalia,Mannerisms,Verbigeration,Negativism | 97.3 | 95.7 | 98.7 |
| 7 | Excitement,Staring,Posturing/catalepsy,Echopraxia/echolalia,Mannerisms,Verbigeration,Rigidity | 97.3 | 95.7 | 98.7 |
| 7 | Excitement,Staring,Posturing/catalepsy,Echopraxia/echolalia,Stereotypy,Mannerisms,Negativism | 97.3 | 95.7 | 98.7 |
| 7 | Excitement,Staring,Posturing/catalepsy,Echopraxia/echolalia,Stereotypy,Negativism,Waxy flexibility | 97.3 | 95.7 | 98.7 |
| 7 | Excitement,Staring,Posturing/catalepsy,Echopraxia/echolalia,Stereotypy,Waxy flexibility,Withdrawal | 97.3 | 95.7 | 98.7 |
| 7 | Excitement,Staring,Posturing/catalepsy,Echopraxia/echolalia,Verbigeration,Negativism,Waxy flexibility | 97.3 | 95.7 | 98.7 |
| 7 | Excitement,Staring,Posturing/catalepsy,Echopraxia/echolalia,Verbigeration,Waxy flexibility,Withdrawal | 97.3 | 95.7 | 98.7 |
| 7 | Excitement,Staring,Posturing/catalepsy,Grimacing,Echopraxia/echolalia,Mannerisms,Negativism | 97.3 | 95.7 | 98.7 |
| 7 | Excitement,Staring,Posturing/catalepsy,Grimacing,Echopraxia/echolalia,Rigidity,Waxy flexibility | 97.3 | 95.7 | 98.7 |
| 7 | Excitement,Staring,Posturing/catalepsy,Grimacing,Mannerisms,Rigidity,Waxy flexibility | 97.3 | 95.7 | 98.7 |
| 7 | Excitement,Staring,Posturing/catalepsy,Grimacing,Mannerisms,Verbigeration,Negativism | 97.3 | 95.7 | 98.7 |
| 7 | Excitement,Staring,Posturing/catalepsy,Grimacing,Stereotypy,Verbigeration,Withdrawal | 97.3 | 95.7 | 98.7 |
| 7 | Excitement,Staring,Posturing/catalepsy,Grimacing,Verbigeration,Waxy flexibility,Withdrawal | 97.3 | 95.7 | 98.7 |
| 7 | Excitement,Staring,Posturing/catalepsy,Stereotypy,Mannerisms,Verbigeration,Negativism | 97.3 | 95.7 | 98.7 |
| 7 | Excitement,Staring,Posturing/catalepsy,Stereotypy,Mannerisms,Verbigeration,Rigidity | 97.3 | 95.7 | 98.7 |
| 7 | Excitement,Staring,Posturing/catalepsy,Stereotypy,Mannerisms,Verbigeration,Withdrawal | 97.3 | 95.7 | 98.7 |
| 7 | Excitement,Staring,Posturing/catalepsy,Stereotypy,Mannerisms,Waxy flexibility,Withdrawal | 97.3 | 95.7 | 98.7 |
| 7 | Excitement,Staring,Posturing/catalepsy,Stereotypy,Verbigeration,Negativism,Waxy flexibility | 97.3 | 95.7 | 98.7 |
| 7 | Immobility/stupor,Grimacing,Echopraxia/echolalia,Mannerisms,Verbigeration,Rigidity,Negativism | 97.3 | 95.7 | 98.7 |
| 7 | Immobility/stupor,Mutism,Echopraxia/echolalia,Stereotypy,Rigidity,Negativism,Withdrawal | 97.3 | 95.7 | 98.7 |
| 7 | Immobility/stupor,Mutism,Grimacing,Mannerisms,Rigidity,Negativism,Waxy flexibility | 97.3 | 95.7 | 98.7 |
| 7 | Immobility/stupor,Mutism,Posturing/catalepsy,Echopraxia/echolalia,Mannerisms,Negativism,Withdrawal | 97.3 | 95.7 | 98.7 |
| 7 | Immobility/stupor,Mutism,Posturing/catalepsy,Grimacing,Mannerisms,Negativism,Withdrawal | 97.3 | 95.7 | 98.7 |
| 7 | Immobility/stupor,Mutism,Staring,Echopraxia/echolalia,Mannerisms,Rigidity,Waxy flexibility | 97.3 | 95.7 | 98.7 |
| 7 | Immobility/stupor,Mutism,Staring,Grimacing,Echopraxia/echolalia,Mannerisms,Withdrawal | 97.3 | 95.7 | 98.7 |
| 7 | Immobility/stupor,Mutism,Staring,Grimacing,Mannerisms,Negativism,Waxy flexibility | 97.3 | 95.7 | 98.7 |
| 7 | Immobility/stupor,Mutism,Staring,Grimacing,Mannerisms,Rigidity,Waxy flexibility | 97.3 | 95.7 | 98.7 |
| 7 | Immobility/stupor,Mutism,Staring,Grimacing,Mannerisms,Verbigeration,Negativism | 97.3 | 95.7 | 98.7 |
| 7 | Immobility/stupor,Mutism,Staring,Grimacing,Stereotypy,Mannerisms,Withdrawal | 97.3 | 95.7 | 98.7 |
| 7 | Immobility/stupor,Mutism,Staring,Grimacing,Stereotypy,Negativism,Withdrawal | 97.3 | 95.7 | 98.7 |
| 7 | Immobility/stupor,Mutism,Staring,Grimacing,Stereotypy,Rigidity,Negativism | 97.3 | 95.7 | 98.7 |
| 7 | Immobility/stupor,Mutism,Staring,Grimacing,Stereotypy,Rigidity,Withdrawal | 97.3 | 95.7 | 98.7 |
| 7 | Immobility/stupor,Mutism,Staring,Grimacing,Verbigeration,Rigidity,Withdrawal | 97.3 | 95.7 | 98.7 |
| 7 | Immobility/stupor,Mutism,Staring,Mannerisms,Verbigeration,Negativism,Waxy flexibility | 97.3 | 95.7 | 98.7 |
| 7 | Immobility/stupor,Mutism,Staring,Posturing/catalepsy,Echopraxia/echolalia,Rigidity,Withdrawal | 97.3 | 95.7 | 98.7 |
| 7 | Immobility/stupor,Mutism,Staring,Posturing/catalepsy,Echopraxia/echolalia,Verbigeration,Rigidity | 97.3 | 95.7 | 98.7 |
| 7 | Immobility/stupor,Mutism,Staring,Posturing/catalepsy,Grimacing,Stereotypy,Negativism | 97.3 | 95.7 | 98.7 |
| 7 | Immobility/stupor,Mutism,Staring,Posturing/catalepsy,Grimacing,Verbigeration,Negativism | 97.3 | 95.7 | 98.7 |
| 7 | Immobility/stupor,Mutism,Staring,Posturing/catalepsy,Mannerisms,Verbigeration,Withdrawal | 97.3 | 95.7 | 98.7 |
| 7 | Immobility/stupor,Mutism,Staring,Posturing/catalepsy,Stereotypy,Rigidity,Withdrawal | 97.3 | 95.7 | 98.7 |
| 7 | Immobility/stupor,Mutism,Staring,Stereotypy,Rigidity,Negativism,Waxy flexibility | 97.3 | 95.7 | 98.7 |
| 7 | Immobility/stupor,Mutism,Staring,Stereotypy,Verbigeration,Negativism,Waxy flexibility | 97.3 | 95.7 | 98.7 |
| 7 | Immobility/stupor,Mutism,Stereotypy,Verbigeration,Rigidity,Negativism,Waxy flexibility | 97.3 | 95.7 | 98.7 |
| 7 | Immobility/stupor,Posturing/catalepsy,Echopraxia/echolalia,Mannerisms,Rigidity,Negativism,Waxy flexibility | 97.3 | 95.7 | 98.7 |
| 7 | Immobility/stupor,Posturing/catalepsy,Echopraxia/echolalia,Stereotypy,Mannerisms,Negativism,Withdrawal | 97.3 | 95.7 | 98.7 |
| 7 | Immobility/stupor,Posturing/catalepsy,Echopraxia/echolalia,Stereotypy,Mannerisms,Rigidity,Negativism | 97.3 | 95.7 | 98.7 |
| 7 | Immobility/stupor,Posturing/catalepsy,Echopraxia/echolalia,Stereotypy,Rigidity,Negativism,Withdrawal | 97.3 | 95.7 | 98.7 |
| 7 | Immobility/stupor,Posturing/catalepsy,Grimacing,Mannerisms,Rigidity,Negativism,Waxy flexibility | 97.3 | 95.7 | 98.7 |
| 7 | Immobility/stupor,Posturing/catalepsy,Grimacing,Stereotypy,Rigidity,Negativism,Withdrawal | 97.3 | 95.7 | 98.7 |
| 7 | Immobility/stupor,Posturing/catalepsy,Grimacing,Stereotypy,Verbigeration,Rigidity,Negativism | 97.3 | 95.7 | 98.7 |
| 7 | Immobility/stupor,Posturing/catalepsy,Mannerisms,Verbigeration,Rigidity,Negativism,Waxy flexibility | 97.3 | 95.7 | 98.7 |
| 7 | Immobility/stupor,Posturing/catalepsy,Stereotypy,Mannerisms,Rigidity,Negativism,Waxy flexibility | 97.3 | 95.7 | 98.7 |
| 7 | Immobility/stupor,Posturing/catalepsy,Stereotypy,Mannerisms,Verbigeration,Negativism,Withdrawal | 97.3 | 95.7 | 98.7 |
| 7 | Immobility/stupor,Staring,Echopraxia/echolalia,Mannerisms,Rigidity,Waxy flexibility,Withdrawal | 97.3 | 95.7 | 98.7 |
| 7 | Immobility/stupor,Staring,Echopraxia/echolalia,Mannerisms,Verbigeration,Rigidity,Withdrawal | 97.3 | 95.7 | 98.7 |
| 7 | Immobility/stupor,Staring,Echopraxia/echolalia,Stereotypy,Negativism,Waxy flexibility,Withdrawal | 97.3 | 95.7 | 98.7 |
| 7 | Immobility/stupor,Staring,Echopraxia/echolalia,Stereotypy,Rigidity,Negativism,Waxy flexibility | 97.3 | 95.7 | 98.7 |
| 7 | Immobility/stupor,Staring,Echopraxia/echolalia,Verbigeration,Rigidity,Negativism,Waxy flexibility | 97.3 | 95.7 | 98.7 |
| 7 | Immobility/stupor,Staring,Grimacing,Echopraxia/echolalia,Mannerisms,Verbigeration,Rigidity | 97.3 | 95.7 | 98.7 |
| 7 | Immobility/stupor,Staring,Grimacing,Echopraxia/echolalia,Stereotypy,Verbigeration,Negativism | 97.3 | 95.7 | 98.7 |
| 7 | Immobility/stupor,Staring,Grimacing,Mannerisms,Verbigeration,Negativism,Waxy flexibility | 97.3 | 95.7 | 98.7 |
| 7 | Immobility/stupor,Staring,Grimacing,Mannerisms,Verbigeration,Rigidity,Waxy flexibility | 97.3 | 95.7 | 98.7 |
| 7 | Immobility/stupor,Staring,Grimacing,Stereotypy,Rigidity,Negativism,Waxy flexibility | 97.3 | 95.7 | 98.7 |
| 7 | Immobility/stupor,Staring,Grimacing,Stereotypy,Verbigeration,Negativism,Waxy flexibility | 97.3 | 95.7 | 98.7 |
| 7 | Immobility/stupor,Staring,Grimacing,Verbigeration,Rigidity,Negativism,Waxy flexibility | 97.3 | 95.7 | 98.7 |
| 7 | Immobility/stupor,Staring,Mannerisms,Verbigeration,Rigidity,Waxy flexibility,Withdrawal | 97.3 | 95.7 | 98.7 |
| 7 | Immobility/stupor,Staring,Posturing/catalepsy,Echopraxia/echolalia,Mannerisms,Verbigeration,Rigidity | 97.3 | 95.7 | 98.7 |
| 7 | Immobility/stupor,Staring,Posturing/catalepsy,Echopraxia/echolalia,Negativism,Waxy flexibility,Withdrawal | 97.3 | 95.7 | 98.7 |
| 7 | Immobility/stupor,Staring,Posturing/catalepsy,Echopraxia/echolalia,Stereotypy,Rigidity,Withdrawal | 97.3 | 95.7 | 98.7 |
| 7 | Immobility/stupor,Staring,Posturing/catalepsy,Grimacing,Echopraxia/echolalia,Mannerisms,Rigidity | 97.3 | 95.7 | 98.7 |
| 7 | Immobility/stupor,Staring,Posturing/catalepsy,Grimacing,Echopraxia/echolalia,Mannerisms,Withdrawal | 97.3 | 95.7 | 98.7 |
| 7 | Immobility/stupor,Staring,Posturing/catalepsy,Grimacing,Echopraxia/echolalia,Rigidity,Negativism | 97.3 | 95.7 | 98.7 |
| 7 | Immobility/stupor,Staring,Posturing/catalepsy,Grimacing,Echopraxia/echolalia,Stereotypy,Negativism | 97.3 | 95.7 | 98.7 |
| 7 | Immobility/stupor,Staring,Posturing/catalepsy,Grimacing,Echopraxia/echolalia,Verbigeration,Negativism | 97.3 | 95.7 | 98.7 |
| 7 | Immobility/stupor,Staring,Posturing/catalepsy,Grimacing,Mannerisms,Negativism,Waxy flexibility | 97.3 | 95.7 | 98.7 |
| 7 | Immobility/stupor,Staring,Posturing/catalepsy,Grimacing,Stereotypy,Rigidity,Withdrawal | 97.3 | 95.7 | 98.7 |
| 7 | Immobility/stupor,Staring,Posturing/catalepsy,Mannerisms,Rigidity,Waxy flexibility,Withdrawal | 97.3 | 95.7 | 98.7 |
| 7 | Immobility/stupor,Staring,Posturing/catalepsy,Rigidity,Negativism,Waxy flexibility,Withdrawal | 97.3 | 95.7 | 98.7 |
| 7 | Immobility/stupor,Staring,Posturing/catalepsy,Stereotypy,Negativism,Waxy flexibility,Withdrawal | 97.3 | 95.7 | 98.7 |
| 7 | Immobility/stupor,Staring,Posturing/catalepsy,Stereotypy,Rigidity,Negativism,Waxy flexibility | 97.3 | 95.7 | 98.7 |
| 7 | Immobility/stupor,Staring,Posturing/catalepsy,Stereotypy,Verbigeration,Rigidity,Withdrawal | 97.3 | 95.7 | 98.7 |
| 7 | Immobility/stupor,Staring,Stereotypy,Verbigeration,Rigidity,Waxy flexibility,Withdrawal | 97.3 | 95.7 | 98.7 |
| 7 | Mutism,Staring,Echopraxia/echolalia,Stereotypy,Verbigeration,Rigidity,Waxy flexibility | 97.3 | 95.7 | 98.7 |
| 7 | Mutism,Staring,Grimacing,Echopraxia/echolalia,Negativism,Waxy flexibility,Withdrawal | 97.3 | 95.7 | 98.7 |
| 7 | Mutism,Staring,Grimacing,Echopraxia/echolalia,Rigidity,Negativism,Waxy flexibility | 97.3 | 95.7 | 98.7 |
| 7 | Mutism,Staring,Grimacing,Echopraxia/echolalia,Rigidity,Waxy flexibility,Withdrawal | 97.3 | 95.7 | 98.7 |
| 7 | Mutism,Staring,Grimacing,Echopraxia/echolalia,Stereotypy,Mannerisms,Verbigeration | 97.3 | 95.7 | 98.7 |
| 7 | Mutism,Staring,Grimacing,Echopraxia/echolalia,Stereotypy,Rigidity,Waxy flexibility | 97.3 | 95.7 | 98.7 |
| 7 | Mutism,Staring,Grimacing,Echopraxia/echolalia,Stereotypy,Verbigeration,Waxy flexibility | 97.3 | 95.7 | 98.7 |
| 7 | Mutism,Staring,Grimacing,Echopraxia/echolalia,Stereotypy,Verbigeration,Withdrawal | 97.3 | 95.7 | 98.7 |
| 7 | Mutism,Staring,Grimacing,Echopraxia/echolalia,Stereotypy,Waxy flexibility,Withdrawal | 97.3 | 95.7 | 98.7 |
| 7 | Mutism,Staring,Grimacing,Rigidity,Negativism,Waxy flexibility,Withdrawal | 97.3 | 95.7 | 98.7 |
| 7 | Mutism,Staring,Grimacing,Stereotypy,Mannerisms,Verbigeration,Waxy flexibility | 97.3 | 95.7 | 98.7 |
| 7 | Mutism,Staring,Grimacing,Stereotypy,Verbigeration,Waxy flexibility,Withdrawal | 97.3 | 95.7 | 98.7 |
| 7 | Mutism,Staring,Grimacing,Verbigeration,Negativism,Waxy flexibility,Withdrawal | 97.3 | 95.7 | 98.7 |
| 7 | Mutism,Staring,Posturing/catalepsy,Echopraxia/echolalia,Rigidity,Waxy flexibility,Withdrawal | 97.3 | 95.7 | 98.7 |
| 7 | Mutism,Staring,Posturing/catalepsy,Echopraxia/echolalia,Stereotypy,Mannerisms,Verbigeration | 97.3 | 95.7 | 98.7 |
| 7 | Mutism,Staring,Posturing/catalepsy,Echopraxia/echolalia,Stereotypy,Mannerisms,Waxy flexibility | 97.3 | 95.7 | 98.7 |
| 7 | Mutism,Staring,Posturing/catalepsy,Echopraxia/echolalia,Verbigeration,Rigidity,Waxy flexibility | 97.3 | 95.7 | 98.7 |
| 7 | Mutism,Staring,Posturing/catalepsy,Grimacing,Echopraxia/echolalia,Mannerisms,Waxy flexibility | 97.3 | 95.7 | 98.7 |
| 7 | Mutism,Staring,Posturing/catalepsy,Grimacing,Echopraxia/echolalia,Negativism,Waxy flexibility | 97.3 | 95.7 | 98.7 |
| 7 | Mutism,Staring,Posturing/catalepsy,Grimacing,Echopraxia/echolalia,Stereotypy,Verbigeration | 97.3 | 95.7 | 98.7 |
| 7 | Mutism,Staring,Posturing/catalepsy,Grimacing,Echopraxia/echolalia,Verbigeration,Withdrawal | 97.3 | 95.7 | 98.7 |
| 7 | Mutism,Staring,Posturing/catalepsy,Grimacing,Mannerisms,Rigidity,Waxy flexibility | 97.3 | 95.7 | 98.7 |
| 7 | Mutism,Staring,Posturing/catalepsy,Grimacing,Stereotypy,Negativism,Waxy flexibility | 97.3 | 95.7 | 98.7 |
| 7 | Mutism,Staring,Posturing/catalepsy,Grimacing,Verbigeration,Negativism,Waxy flexibility | 97.3 | 95.7 | 98.7 |
| 7 | Mutism,Staring,Posturing/catalepsy,Verbigeration,Rigidity,Negativism,Waxy flexibility | 97.3 | 95.7 | 98.7 |
| 7 | Mutism,Staring,Posturing/catalepsy,Verbigeration,Rigidity,Waxy flexibility,Withdrawal | 97.3 | 95.7 | 98.7 |
| 7 | Staring,Echopraxia/echolalia,Mannerisms,Verbigeration,Rigidity,Negativism,Waxy flexibility | 97.3 | 95.7 | 98.7 |
| 7 | Staring,Echopraxia/echolalia,Stereotypy,Mannerisms,Verbigeration,Rigidity,Withdrawal | 97.3 | 95.7 | 98.7 |
| 7 | Staring,Grimacing,Echopraxia/echolalia,Mannerisms,Rigidity,Negativism,Waxy flexibility | 97.3 | 95.7 | 98.7 |
| 7 | Staring,Grimacing,Echopraxia/echolalia,Rigidity,Negativism,Waxy flexibility,Withdrawal | 97.3 | 95.7 | 98.7 |
| 7 | Staring,Grimacing,Echopraxia/echolalia,Stereotypy,Mannerisms,Waxy flexibility,Withdrawal | 97.3 | 95.7 | 98.7 |
| 7 | Staring,Grimacing,Echopraxia/echolalia,Stereotypy,Verbigeration,Negativism,Withdrawal | 97.3 | 95.7 | 98.7 |
| 7 | Staring,Grimacing,Echopraxia/echolalia,Stereotypy,Verbigeration,Rigidity,Withdrawal | 97.3 | 95.7 | 98.7 |
| 7 | Staring,Grimacing,Verbigeration,Rigidity,Negativism,Waxy flexibility,Withdrawal | 97.3 | 95.7 | 98.7 |
| 7 | Staring,Posturing/catalepsy,Echopraxia/echolalia,Mannerisms,Rigidity,Negativism,Waxy flexibility | 97.3 | 95.7 | 98.7 |
| 7 | Staring,Posturing/catalepsy,Echopraxia/echolalia,Mannerisms,Verbigeration,Rigidity,Withdrawal | 97.3 | 95.7 | 98.7 |
| 7 | Staring,Posturing/catalepsy,Grimacing,Echopraxia/echolalia,Stereotypy,Rigidity,Withdrawal | 97.3 | 95.7 | 98.7 |
| 7 | Staring,Posturing/catalepsy,Grimacing,Stereotypy,Mannerisms,Verbigeration,Withdrawal | 97.3 | 95.7 | 98.7 |
| 7 | Staring,Posturing/catalepsy,Stereotypy,Mannerisms,Rigidity,Negativism,Waxy flexibility | 97.3 | 95.7 | 98.7 |
| 7 | Staring,Posturing/catalepsy,Stereotypy,Verbigeration,Negativism,Waxy flexibility,Withdrawal | 97.3 | 95.7 | 98.7 |
| 7 | Staring,Posturing/catalepsy,Verbigeration,Rigidity,Negativism,Waxy flexibility,Withdrawal | 97.3 | 95.7 | 98.7 |
| 7 | Excitement,Immobility/stupor,Echopraxia/echolalia,Mannerisms,Verbigeration,Negativism,Withdrawal | 97.1 | 95.3 | 98.7 |
| 7 | Excitement,Immobility/stupor,Echopraxia/echolalia,Verbigeration,Negativism,Waxy flexibility,Withdrawal | 97.1 | 95.5 | 98.7 |
| 7 | Excitement,Immobility/stupor,Grimacing,Echopraxia/echolalia,Mannerisms,Rigidity,Waxy flexibility | 97.1 | 95.5 | 98.4 |
| 7 | Excitement,Immobility/stupor,Grimacing,Echopraxia/echolalia,Negativism,Waxy flexibility,Withdrawal | 97.1 | 95.5 | 98.7 |
| 7 | Excitement,Immobility/stupor,Grimacing,Echopraxia/echolalia,Stereotypy,Rigidity,Waxy flexibility | 97.1 | 95.5 | 98.4 |
| 7 | Excitement,Immobility/stupor,Grimacing,Echopraxia/echolalia,Verbigeration,Negativism,Withdrawal | 97.1 | 95.5 | 98.4 |
| 7 | Excitement,Immobility/stupor,Grimacing,Stereotypy,Verbigeration,Negativism,Withdrawal | 97.1 | 95.5 | 98.7 |
| 7 | Excitement,Immobility/stupor,Mutism,Echopraxia/echolalia,Stereotypy,Negativism,Waxy flexibility | 97.1 | 95.5 | 98.7 |
| 7 | Excitement,Immobility/stupor,Mutism,Echopraxia/echolalia,Stereotypy,Verbigeration,Negativism | 97.1 | 95.5 | 98.4 |
| 7 | Excitement,Immobility/stupor,Mutism,Echopraxia/echolalia,Verbigeration,Negativism,Waxy flexibility | 97.1 | 95.5 | 98.4 |
| 7 | Excitement,Immobility/stupor,Mutism,Grimacing,Echopraxia/echolalia,Negativism,Waxy flexibility | 97.1 | 95.5 | 98.4 |
| 7 | Excitement,Immobility/stupor,Mutism,Grimacing,Echopraxia/echolalia,Negativism,Withdrawal | 97.1 | 95.5 | 98.4 |
| 7 | Excitement,Immobility/stupor,Mutism,Grimacing,Stereotypy,Mannerisms,Rigidity | 97.1 | 95.5 | 98.7 |
| 7 | Excitement,Immobility/stupor,Mutism,Grimacing,Stereotypy,Negativism,Withdrawal | 97.1 | 95.5 | 98.4 |
| 7 | Excitement,Immobility/stupor,Mutism,Grimacing,Verbigeration,Negativism,Withdrawal | 97.1 | 95.5 | 98.7 |
| 7 | Excitement,Immobility/stupor,Mutism,Posturing/catalepsy,Echopraxia/echolalia,Mannerisms,Withdrawal | 97.1 | 95.5 | 98.4 |
| 7 | Excitement,Immobility/stupor,Mutism,Posturing/catalepsy,Grimacing,Echopraxia/echolalia,Verbigeration | 97.1 | 95.5 | 98.4 |
| 7 | Excitement,Immobility/stupor,Mutism,Posturing/catalepsy,Grimacing,Echopraxia/echolalia,Waxy flexibility | 97.1 | 95.5 | 98.4 |
| 7 | Excitement,Immobility/stupor,Mutism,Posturing/catalepsy,Grimacing,Mannerisms,Withdrawal | 97.1 | 95.3 | 98.7 |
| 7 | Excitement,Immobility/stupor,Mutism,Posturing/catalepsy,Grimacing,Stereotypy,Withdrawal | 97.1 | 95.3 | 98.4 |
| 7 | Excitement,Immobility/stupor,Mutism,Posturing/catalepsy,Grimacing,Waxy flexibility,Withdrawal | 97.1 | 95.5 | 98.4 |
| 7 | Excitement,Immobility/stupor,Mutism,Posturing/catalepsy,Mannerisms,Negativism,Waxy flexibility | 97.1 | 95.5 | 98.4 |
| 7 | Excitement,Immobility/stupor,Mutism,Posturing/catalepsy,Mannerisms,Verbigeration,Withdrawal | 97.1 | 95.5 | 98.4 |
| 7 | Excitement,Immobility/stupor,Mutism,Posturing/catalepsy,Stereotypy,Mannerisms,Negativism | 97.1 | 95.5 | 98.7 |
| 7 | Excitement,Immobility/stupor,Mutism,Posturing/catalepsy,Stereotypy,Verbigeration,Withdrawal | 97.1 | 95.5 | 98.7 |
| 7 | Excitement,Immobility/stupor,Mutism,Posturing/catalepsy,Verbigeration,Waxy flexibility,Withdrawal | 97.1 | 95.5 | 98.7 |
| 7 | Excitement,Immobility/stupor,Mutism,Staring,Grimacing,Mannerisms,Verbigeration | 97.1 | 95.5 | 98.7 |
| 7 | Excitement,Immobility/stupor,Mutism,Verbigeration,Negativism,Waxy flexibility,Withdrawal | 97.1 | 95.5 | 98.4 |
| 7 | Excitement,Immobility/stupor,Posturing/catalepsy,Echopraxia/echolalia,Verbigeration,Negativism,Waxy flexibility | 97.1 | 95.5 | 98.4 |
| 7 | Excitement,Immobility/stupor,Posturing/catalepsy,Grimacing,Echopraxia/echolalia,Mannerisms,Negativism | 97.1 | 95.5 | 98.4 |
| 7 | Excitement,Immobility/stupor,Posturing/catalepsy,Grimacing,Echopraxia/echolalia,Mannerisms,Withdrawal | 97.1 | 95.5 | 98.4 |
| 7 | Excitement,Immobility/stupor,Posturing/catalepsy,Grimacing,Mannerisms,Rigidity,Waxy flexibility | 97.1 | 95.3 | 98.4 |
| 7 | Excitement,Immobility/stupor,Posturing/catalepsy,Grimacing,Stereotypy,Verbigeration,Negativism | 97.1 | 95.5 | 98.4 |
| 7 | Excitement,Immobility/stupor,Posturing/catalepsy,Mannerisms,Verbigeration,Negativism,Waxy flexibility | 97.1 | 95.5 | 98.4 |
| 7 | Excitement,Immobility/stupor,Posturing/catalepsy,Mannerisms,Verbigeration,Rigidity,Waxy flexibility | 97.1 | 95.3 | 98.4 |
| 7 | Excitement,Immobility/stupor,Posturing/catalepsy,Stereotypy,Mannerisms,Verbigeration,Rigidity | 97.1 | 95.3 | 98.4 |
| 7 | Excitement,Immobility/stupor,Posturing/catalepsy,Stereotypy,Verbigeration,Negativism,Waxy flexibility | 97.1 | 95.5 | 98.7 |
| 7 | Excitement,Immobility/stupor,Posturing/catalepsy,Stereotypy,Verbigeration,Rigidity,Waxy flexibility | 97.1 | 95.5 | 98.7 |
| 7 | Excitement,Immobility/stupor,Staring,Echopraxia/echolalia,Stereotypy,Mannerisms,Verbigeration | 97.1 | 95.5 | 98.7 |
| 7 | Excitement,Immobility/stupor,Staring,Grimacing,Echopraxia/echolalia,Stereotypy,Mannerisms | 97.1 | 95.5 | 98.4 |
| 7 | Excitement,Immobility/stupor,Staring,Grimacing,Stereotypy,Mannerisms,Waxy flexibility | 97.1 | 95.5 | 98.7 |
| 7 | Excitement,Immobility/stupor,Staring,Grimacing,Stereotypy,Mannerisms,Withdrawal | 97.1 | 95.5 | 98.4 |
| 7 | Excitement,Immobility/stupor,Stereotypy,Verbigeration,Negativism,Waxy flexibility,Withdrawal | 97.1 | 95.5 | 98.4 |
| 7 | Excitement,Staring,Echopraxia/echolalia,Mannerisms,Verbigeration,Waxy flexibility,Withdrawal | 97.1 | 95.3 | 98.4 |
| 7 | Excitement,Staring,Echopraxia/echolalia,Stereotypy,Mannerisms,Negativism,Waxy flexibility | 97.1 | 95.3 | 98.7 |
| 7 | Excitement,Staring,Grimacing,Echopraxia/echolalia,Stereotypy,Verbigeration,Rigidity | 97.1 | 95.5 | 98.4 |
| 7 | Excitement,Staring,Grimacing,Echopraxia/echolalia,Verbigeration,Rigidity,Negativism | 97.1 | 95.5 | 98.7 |
| 7 | Excitement,Staring,Grimacing,Echopraxia/echolalia,Verbigeration,Waxy flexibility,Withdrawal | 97.1 | 95.5 | 98.4 |
| 7 | Excitement,Staring,Grimacing,Stereotypy,Mannerisms,Verbigeration,Withdrawal | 97.1 | 95.5 | 98.4 |
| 7 | Excitement,Staring,Grimacing,Stereotypy,Mannerisms,Waxy flexibility,Withdrawal | 97.1 | 95.5 | 98.4 |
| 7 | Excitement,Staring,Posturing/catalepsy,Echopraxia/echolalia,Stereotypy,Mannerisms,Rigidity | 97.1 | 95.5 | 98.4 |
| 7 | Excitement,Staring,Posturing/catalepsy,Echopraxia/echolalia,Stereotypy,Verbigeration,Negativism | 97.1 | 95.5 | 98.4 |
| 7 | Excitement,Staring,Posturing/catalepsy,Echopraxia/echolalia,Stereotypy,Verbigeration,Rigidity | 97.1 | 95.5 | 98.4 |
| 7 | Excitement,Staring,Posturing/catalepsy,Grimacing,Echopraxia/echolalia,Negativism,Waxy flexibility | 97.1 | 95.5 | 98.7 |
| 7 | Excitement,Staring,Posturing/catalepsy,Grimacing,Echopraxia/echolalia,Stereotypy,Negativism | 97.1 | 95.5 | 98.4 |
| 7 | Excitement,Staring,Posturing/catalepsy,Grimacing,Echopraxia/echolalia,Verbigeration,Negativism | 97.1 | 95.5 | 98.7 |
| 7 | Excitement,Staring,Posturing/catalepsy,Grimacing,Rigidity,Negativism,Waxy flexibility | 97.1 | 95.5 | 98.4 |
| 7 | Excitement,Staring,Posturing/catalepsy,Grimacing,Stereotypy,Rigidity,Waxy flexibility | 97.1 | 95.5 | 98.4 |
| 7 | Excitement,Staring,Posturing/catalepsy,Grimacing,Stereotypy,Verbigeration,Negativism | 97.1 | 95.5 | 98.4 |
| 7 | Excitement,Staring,Posturing/catalepsy,Grimacing,Stereotypy,Waxy flexibility,Withdrawal | 97.1 | 95.5 | 98.4 |
| 7 | Excitement,Staring,Posturing/catalepsy,Grimacing,Verbigeration,Rigidity,Waxy flexibility | 97.1 | 95.5 | 98.7 |
| 7 | Excitement,Staring,Posturing/catalepsy,Mannerisms,Verbigeration,Rigidity,Waxy flexibility | 97.1 | 95.5 | 98.4 |
| 7 | Excitement,Staring,Posturing/catalepsy,Stereotypy,Mannerisms,Negativism,Waxy flexibility | 97.1 | 95.5 | 98.4 |
| 7 | Excitement,Staring,Posturing/catalepsy,Stereotypy,Verbigeration,Waxy flexibility,Withdrawal | 97.1 | 95.3 | 98.7 |
| 7 | Excitement,Staring,Stereotypy,Mannerisms,Verbigeration,Rigidity,Waxy flexibility | 97.1 | 95.5 | 98.7 |
| 7 | Excitement,Staring,Stereotypy,Mannerisms,Verbigeration,Waxy flexibility,Withdrawal | 97.1 | 95.5 | 98.4 |
| 7 | Immobility/stupor,Echopraxia/echolalia,Mannerisms,Verbigeration,Rigidity,Negativism,Waxy flexibility | 97.1 | 95.5 | 98.4 |
| 7 | Immobility/stupor,Echopraxia/echolalia,Stereotypy,Mannerisms,Verbigeration,Negativism,Withdrawal | 97.1 | 95.3 | 98.4 |
| 7 | Immobility/stupor,Echopraxia/echolalia,Stereotypy,Rigidity,Negativism,Waxy flexibility,Withdrawal | 97.1 | 95.3 | 98.4 |
| 7 | Immobility/stupor,Grimacing,Echopraxia/echolalia,Stereotypy,Mannerisms,Negativism,Withdrawal | 97.1 | 95.5 | 98.4 |
| 7 | Immobility/stupor,Grimacing,Echopraxia/echolalia,Verbigeration,Rigidity,Negativism,Withdrawal | 97.1 | 95.5 | 98.7 |
| 7 | Immobility/stupor,Grimacing,Mannerisms,Verbigeration,Rigidity,Negativism,Waxy flexibility | 97.1 | 95.5 | 98.4 |
| 7 | Immobility/stupor,Grimacing,Stereotypy,Verbigeration,Rigidity,Negativism,Waxy flexibility | 97.1 | 95.5 | 98.7 |
| 7 | Immobility/stupor,Mutism,Echopraxia/echolalia,Stereotypy,Verbigeration,Rigidity,Negativism | 97.1 | 95.5 | 98.7 |
| 7 | Immobility/stupor,Mutism,Echopraxia/echolalia,Verbigeration,Rigidity,Negativism,Withdrawal | 97.1 | 95.5 | 98.4 |
| 7 | Immobility/stupor,Mutism,Grimacing,Echopraxia/echolalia,Mannerisms,Rigidity,Withdrawal | 97.1 | 95.5 | 98.4 |
| 7 | Immobility/stupor,Mutism,Grimacing,Stereotypy,Rigidity,Negativism,Withdrawal | 97.1 | 95.5 | 98.7 |
| 7 | Immobility/stupor,Mutism,Grimacing,Verbigeration,Rigidity,Negativism,Withdrawal | 97.1 | 95.3 | 98.7 |
| 7 | Immobility/stupor,Mutism,Posturing/catalepsy,Echopraxia/echolalia,Verbigeration,Rigidity,Negativism | 97.1 | 95.5 | 98.4 |
| 7 | Immobility/stupor,Mutism,Posturing/catalepsy,Mannerisms,Rigidity,Negativism,Waxy flexibility | 97.1 | 95.5 | 98.7 |
| 7 | Immobility/stupor,Mutism,Posturing/catalepsy,Mannerisms,Verbigeration,Negativism,Withdrawal | 97.1 | 95.5 | 98.4 |
| 7 | Immobility/stupor,Mutism,Posturing/catalepsy,Stereotypy,Rigidity,Negativism,Withdrawal | 97.1 | 95.3 | 98.4 |
| 7 | Immobility/stupor,Mutism,Staring,Echopraxia/echolalia,Stereotypy,Mannerisms,Withdrawal | 97.1 | 95.5 | 98.4 |
| 7 | Immobility/stupor,Mutism,Staring,Echopraxia/echolalia,Stereotypy,Verbigeration,Negativism | 97.1 | 95.5 | 98.4 |
| 7 | Immobility/stupor,Mutism,Staring,Echopraxia/echolalia,Stereotypy,Verbigeration,Rigidity | 97.1 | 95.5 | 98.7 |
| 7 | Immobility/stupor,Mutism,Staring,Echopraxia/echolalia,Verbigeration,Negativism,Withdrawal | 97.1 | 95.5 | 98.4 |
| 7 | Immobility/stupor,Mutism,Staring,Echopraxia/echolalia,Verbigeration,Rigidity,Withdrawal | 97.1 | 95.5 | 98.7 |
| 7 | Immobility/stupor,Mutism,Staring,Grimacing,Echopraxia/echolalia,Negativism,Withdrawal | 97.1 | 95.5 | 98.4 |
| 7 | Immobility/stupor,Mutism,Staring,Grimacing,Echopraxia/echolalia,Rigidity,Negativism | 97.1 | 95.5 | 98.4 |
| 7 | Immobility/stupor,Mutism,Staring,Grimacing,Echopraxia/echolalia,Rigidity,Withdrawal | 97.1 | 95.5 | 98.4 |
| 7 | Immobility/stupor,Mutism,Staring,Grimacing,Echopraxia/echolalia,Stereotypy,Negativism | 97.1 | 95.5 | 98.7 |
| 7 | Immobility/stupor,Mutism,Staring,Grimacing,Echopraxia/echolalia,Verbigeration,Negativism | 97.1 | 95.5 | 98.4 |
| 7 | Immobility/stupor,Mutism,Staring,Grimacing,Echopraxia/echolalia,Verbigeration,Rigidity | 97.1 | 95.5 | 98.4 |
| 7 | Immobility/stupor,Mutism,Staring,Grimacing,Mannerisms,Waxy flexibility,Withdrawal | 97.1 | 95.5 | 98.7 |
| 7 | Immobility/stupor,Mutism,Staring,Grimacing,Rigidity,Negativism,Withdrawal | 97.1 | 95.5 | 98.7 |
| 7 | Immobility/stupor,Mutism,Staring,Grimacing,Verbigeration,Negativism,Withdrawal | 97.1 | 95.3 | 98.4 |
| 7 | Immobility/stupor,Mutism,Staring,Posturing/catalepsy,Echopraxia/echolalia,Stereotypy,Rigidity | 97.1 | 95.5 | 98.4 |
| 7 | Immobility/stupor,Mutism,Staring,Posturing/catalepsy,Grimacing,Echopraxia/echolalia,Mannerisms | 97.1 | 95.5 | 98.7 |
| 7 | Immobility/stupor,Mutism,Staring,Posturing/catalepsy,Grimacing,Echopraxia/echolalia,Negativism | 97.1 | 95.5 | 98.4 |
| 7 | Immobility/stupor,Mutism,Staring,Posturing/catalepsy,Grimacing,Mannerisms,Verbigeration | 97.1 | 95.3 | 98.4 |
| 7 | Immobility/stupor,Mutism,Staring,Posturing/catalepsy,Grimacing,Verbigeration,Rigidity | 97.1 | 95.5 | 98.7 |
| 7 | Immobility/stupor,Mutism,Staring,Posturing/catalepsy,Mannerisms,Waxy flexibility,Withdrawal | 97.1 | 95.5 | 98.7 |
| 7 | Immobility/stupor,Mutism,Staring,Posturing/catalepsy,Negativism,Waxy flexibility,Withdrawal | 97.1 | 95.5 | 98.4 |
| 7 | Immobility/stupor,Mutism,Staring,Posturing/catalepsy,Stereotypy,Negativism,Waxy flexibility | 97.1 | 95.5 | 98.7 |
| 7 | Immobility/stupor,Mutism,Staring,Posturing/catalepsy,Verbigeration,Rigidity,Withdrawal | 97.1 | 95.5 | 98.4 |
| 7 | Immobility/stupor,Mutism,Staring,Rigidity,Negativism,Waxy flexibility,Withdrawal | 97.1 | 95.5 | 98.4 |
| 7 | Immobility/stupor,Mutism,Staring,Stereotypy,Negativism,Waxy flexibility,Withdrawal | 97.1 | 95.3 | 98.7 |
| 7 | Immobility/stupor,Mutism,Staring,Stereotypy,Rigidity,Waxy flexibility,Withdrawal | 97.1 | 95.5 | 98.4 |
| 7 | Immobility/stupor,Mutism,Staring,Verbigeration,Rigidity,Negativism,Waxy flexibility | 97.1 | 95.5 | 98.7 |
| 7 | Immobility/stupor,Posturing/catalepsy,Echopraxia/echolalia,Verbigeration,Rigidity,Negativism,Withdrawal | 97.1 | 95.5 | 98.4 |
| 7 | Immobility/stupor,Posturing/catalepsy,Grimacing,Echopraxia/echolalia,Mannerisms,Negativism,Withdrawal | 97.1 | 95.5 | 98.4 |
| 7 | Immobility/stupor,Posturing/catalepsy,Grimacing,Echopraxia/echolalia,Rigidity,Negativism,Withdrawal | 97.1 | 95.5 | 98.7 |
| 7 | Immobility/stupor,Posturing/catalepsy,Grimacing,Stereotypy,Mannerisms,Negativism,Withdrawal | 97.1 | 95.5 | 98.4 |
| 7 | Immobility/stupor,Posturing/catalepsy,Grimacing,Verbigeration,Rigidity,Negativism,Withdrawal | 97.1 | 95.5 | 98.4 |
| 7 | Immobility/stupor,Posturing/catalepsy,Stereotypy,Verbigeration,Rigidity,Negativism,Waxy flexibility | 97.1 | 95.5 | 98.4 |
| 7 | Immobility/stupor,Staring,Echopraxia/echolalia,Stereotypy,Mannerisms,Verbigeration,Rigidity | 97.1 | 95.5 | 98.4 |
| 7 | Immobility/stupor,Staring,Echopraxia/echolalia,Stereotypy,Verbigeration,Negativism,Waxy flexibility | 97.1 | 95.5 | 98.7 |
| 7 | Immobility/stupor,Staring,Echopraxia/echolalia,Stereotypy,Verbigeration,Rigidity,Withdrawal | 97.1 | 95.3 | 98.4 |
| 7 | Immobility/stupor,Staring,Grimacing,Echopraxia/echolalia,Mannerisms,Rigidity,Waxy flexibility | 97.1 | 95.5 | 98.4 |
| 7 | Immobility/stupor,Staring,Grimacing,Echopraxia/echolalia,Stereotypy,Verbigeration,Rigidity | 97.1 | 95.5 | 98.4 |
| 7 | Immobility/stupor,Staring,Grimacing,Echopraxia/echolalia,Verbigeration,Negativism,Withdrawal | 97.1 | 95.5 | 98.7 |
| 7 | Immobility/stupor,Staring,Grimacing,Rigidity,Negativism,Waxy flexibility,Withdrawal | 97.1 | 95.3 | 98.4 |
| 7 | Immobility/stupor,Staring,Grimacing,Stereotypy,Mannerisms,Verbigeration,Withdrawal | 97.1 | 95.5 | 98.4 |
| 7 | Immobility/stupor,Staring,Grimacing,Stereotypy,Mannerisms,Waxy flexibility,Withdrawal | 97.1 | 95.5 | 98.4 |
| 7 | Immobility/stupor,Staring,Grimacing,Stereotypy,Rigidity,Waxy flexibility,Withdrawal | 97.1 | 95.5 | 98.7 |
| 7 | Immobility/stupor,Staring,Posturing/catalepsy,Echopraxia/echolalia,Stereotypy,Mannerisms,Rigidity | 97.1 | 95.5 | 98.4 |
| 7 | Immobility/stupor,Staring,Posturing/catalepsy,Echopraxia/echolalia,Stereotypy,Mannerisms,Withdrawal | 97.1 | 95.5 | 98.4 |
| 7 | Immobility/stupor,Staring,Posturing/catalepsy,Echopraxia/echolalia,Stereotypy,Negativism,Waxy flexibility | 97.1 | 95.5 | 98.4 |
| 7 | Immobility/stupor,Staring,Posturing/catalepsy,Grimacing,Mannerisms,Verbigeration,Withdrawal | 97.1 | 95.5 | 98.7 |
| 7 | Immobility/stupor,Staring,Posturing/catalepsy,Verbigeration,Negativism,Waxy flexibility,Withdrawal | 97.1 | 95.5 | 98.7 |
| 7 | Mutism,Staring,Echopraxia/echolalia,Stereotypy,Mannerisms,Verbigeration,Withdrawal | 97.1 | 95.5 | 98.4 |
| 7 | Mutism,Staring,Echopraxia/echolalia,Verbigeration,Rigidity,Waxy flexibility,Withdrawal | 97.1 | 95.5 | 98.4 |
| 7 | Mutism,Staring,Posturing/catalepsy,Echopraxia/echolalia,Mannerisms,Verbigeration,Waxy flexibility | 97.1 | 95.5 | 98.4 |
| 7 | Mutism,Staring,Posturing/catalepsy,Echopraxia/echolalia,Stereotypy,Rigidity,Waxy flexibility | 97.1 | 95.5 | 98.7 |
| 7 | Mutism,Staring,Posturing/catalepsy,Echopraxia/echolalia,Stereotypy,Verbigeration,Withdrawal | 97.1 | 95.3 | 98.4 |
| 7 | Mutism,Staring,Posturing/catalepsy,Echopraxia/echolalia,Stereotypy,Waxy flexibility,Withdrawal | 97.1 | 95.5 | 98.4 |
| 7 | Mutism,Staring,Posturing/catalepsy,Grimacing,Echopraxia/echolalia,Waxy flexibility,Withdrawal | 97.1 | 95.5 | 98.4 |
| 7 | Mutism,Staring,Posturing/catalepsy,Grimacing,Mannerisms,Verbigeration,Waxy flexibility | 97.1 | 95.5 | 98.4 |
| 7 | Mutism,Staring,Posturing/catalepsy,Grimacing,Stereotypy,Mannerisms,Waxy flexibility | 97.1 | 95.5 | 98.4 |
| 7 | Mutism,Staring,Posturing/catalepsy,Grimacing,Stereotypy,Verbigeration,Waxy flexibility | 97.1 | 95.5 | 98.4 |
| 7 | Mutism,Staring,Posturing/catalepsy,Grimacing,Stereotypy,Waxy flexibility,Withdrawal | 97.1 | 95.3 | 98.4 |
| 7 | Mutism,Staring,Posturing/catalepsy,Stereotypy,Mannerisms,Verbigeration,Waxy flexibility | 97.1 | 95.5 | 98.4 |
| 7 | Mutism,Staring,Posturing/catalepsy,Stereotypy,Verbigeration,Waxy flexibility,Withdrawal | 97.1 | 95.5 | 98.7 |
| 7 | Mutism,Staring,Stereotypy,Mannerisms,Verbigeration,Waxy flexibility,Withdrawal | 97.1 | 95.5 | 98.4 |
| 7 | Staring,Grimacing,Echopraxia/echolalia,Stereotypy,Rigidity,Waxy flexibility,Withdrawal | 97.1 | 95.5 | 98.4 |
| 7 | Staring,Grimacing,Stereotypy,Verbigeration,Negativism,Waxy flexibility,Withdrawal | 97.1 | 95.5 | 98.4 |
| 7 | Staring,Posturing/catalepsy,Echopraxia/echolalia,Mannerisms,Rigidity,Waxy flexibility,Withdrawal | 97.1 | 95.5 | 98.4 |
| 7 | Staring,Posturing/catalepsy,Echopraxia/echolalia,Stereotypy,Negativism,Waxy flexibility,Withdrawal | 97.1 | 95.5 | 98.4 |
| 7 | Staring,Posturing/catalepsy,Echopraxia/echolalia,Stereotypy,Verbigeration,Negativism,Withdrawal | 97.1 | 95.5 | 98.7 |
| 7 | Staring,Posturing/catalepsy,Grimacing,Echopraxia/echolalia,Stereotypy,Mannerisms,Negativism | 97.1 | 95.5 | 98.4 |
| 7 | Staring,Posturing/catalepsy,Grimacing,Echopraxia/echolalia,Verbigeration,Negativism,Withdrawal | 97.1 | 95.5 | 98.4 |
| 7 | Staring,Posturing/catalepsy,Grimacing,Mannerisms,Verbigeration,Rigidity,Negativism | 97.1 | 95.5 | 98.4 |
| 7 | Staring,Posturing/catalepsy,Grimacing,Stereotypy,Mannerisms,Waxy flexibility,Withdrawal | 97.1 | 95.5 | 98.4 |
| 7 | Staring,Posturing/catalepsy,Mannerisms,Verbigeration,Rigidity,Waxy flexibility,Withdrawal | 97.1 | 95.3 | 98.4 |
| 7 | Excitement,Immobility/stupor,Echopraxia/echolalia,Mannerisms,Negativism,Waxy flexibility,Withdrawal | 96.9 | 95.1 | 98.4 |
| 7 | Excitement,Immobility/stupor,Echopraxia/echolalia,Mannerisms,Verbigeration,Rigidity,Waxy flexibility | 96.9 | 95.1 | 98.4 |
| 7 | Excitement,Immobility/stupor,Echopraxia/echolalia,Stereotypy,Mannerisms,Verbigeration,Rigidity | 96.9 | 95.1 | 98.4 |
| 7 | Excitement,Immobility/stupor,Grimacing,Echopraxia/echolalia,Mannerisms,Verbigeration,Rigidity | 96.9 | 95.1 | 98.4 |
| 7 | Excitement,Immobility/stupor,Grimacing,Echopraxia/echolalia,Stereotypy,Verbigeration,Rigidity | 96.9 | 95.1 | 98.4 |
| 7 | Excitement,Immobility/stupor,Grimacing,Mannerisms,Verbigeration,Rigidity,Waxy flexibility | 96.9 | 95.3 | 98.4 |
| 7 | Excitement,Immobility/stupor,Grimacing,Stereotypy,Verbigeration,Rigidity,Waxy flexibility | 96.9 | 95.1 | 98.4 |
| 7 | Excitement,Immobility/stupor,Mutism,Echopraxia/echolalia,Stereotypy,Waxy flexibility,Withdrawal | 96.9 | 95.1 | 98.4 |
| 7 | Excitement,Immobility/stupor,Mutism,Grimacing,Echopraxia/echolalia,Waxy flexibility,Withdrawal | 96.9 | 95.3 | 98.4 |
| 7 | Excitement,Immobility/stupor,Mutism,Grimacing,Negativism,Waxy flexibility,Withdrawal | 96.9 | 95.1 | 98.4 |
| 7 | Excitement,Immobility/stupor,Mutism,Grimacing,Stereotypy,Negativism,Waxy flexibility | 96.9 | 95.1 | 98.4 |
| 7 | Excitement,Immobility/stupor,Mutism,Grimacing,Stereotypy,Verbigeration,Negativism | 96.9 | 95.1 | 98.4 |
| 7 | Excitement,Immobility/stupor,Mutism,Grimacing,Verbigeration,Negativism,Waxy flexibility | 96.9 | 95.1 | 98.4 |
| 7 | Excitement,Immobility/stupor,Mutism,Mannerisms,Verbigeration,Negativism,Withdrawal | 96.9 | 95.1 | 98.4 |
| 7 | Excitement,Immobility/stupor,Mutism,Posturing/catalepsy,Echopraxia/echolalia,Verbigeration,Waxy flexibility | 96.9 | 95.1 | 98.4 |
| 7 | Excitement,Immobility/stupor,Mutism,Posturing/catalepsy,Grimacing,Verbigeration,Waxy flexibility | 96.9 | 95.1 | 98.4 |
| 7 | Excitement,Immobility/stupor,Mutism,Posturing/catalepsy,Stereotypy,Waxy flexibility,Withdrawal | 96.9 | 95.3 | 98.4 |
| 7 | Excitement,Immobility/stupor,Mutism,Staring,Grimacing,Stereotypy,Mannerisms | 96.9 | 95.1 | 98.4 |
| 7 | Excitement,Immobility/stupor,Mutism,Stereotypy,Mannerisms,Negativism,Withdrawal | 96.9 | 95.1 | 98.4 |
| 7 | Excitement,Immobility/stupor,Posturing/catalepsy,Echopraxia/echolalia,Mannerisms,Rigidity,Waxy flexibility | 96.9 | 95.3 | 98.4 |
| 7 | Excitement,Immobility/stupor,Posturing/catalepsy,Echopraxia/echolalia,Mannerisms,Verbigeration,Negativism | 96.9 | 95.1 | 98.4 |
| 7 | Excitement,Immobility/stupor,Posturing/catalepsy,Echopraxia/echolalia,Mannerisms,Verbigeration,Withdrawal | 96.9 | 95.1 | 98.4 |
| 7 | Excitement,Immobility/stupor,Posturing/catalepsy,Echopraxia/echolalia,Stereotypy,Mannerisms,Rigidity | 96.9 | 95.1 | 98.4 |
| 7 | Excitement,Immobility/stupor,Posturing/catalepsy,Echopraxia/echolalia,Stereotypy,Rigidity,Waxy flexibility | 96.9 | 95.1 | 98.4 |
| 7 | Excitement,Immobility/stupor,Posturing/catalepsy,Echopraxia/echolalia,Stereotypy,Verbigeration,Negativism | 96.9 | 95.1 | 98.4 |
| 7 | Excitement,Immobility/stupor,Posturing/catalepsy,Grimacing,Echopraxia/echolalia,Stereotypy,Negativism | 96.9 | 95.1 | 98.4 |
| 7 | Excitement,Immobility/stupor,Posturing/catalepsy,Grimacing,Echopraxia/echolalia,Verbigeration,Withdrawal | 96.9 | 95.1 | 98.4 |
| 7 | Excitement,Immobility/stupor,Posturing/catalepsy,Grimacing,Echopraxia/echolalia,Waxy flexibility,Withdrawal | 96.9 | 95.1 | 98.4 |
| 7 | Excitement,Immobility/stupor,Posturing/catalepsy,Grimacing,Mannerisms,Negativism,Waxy flexibility | 96.9 | 95.1 | 98.4 |
| 7 | Excitement,Immobility/stupor,Posturing/catalepsy,Grimacing,Stereotypy,Mannerisms,Rigidity | 96.9 | 95.1 | 98.4 |
| 7 | Excitement,Immobility/stupor,Posturing/catalepsy,Grimacing,Stereotypy,Rigidity,Waxy flexibility | 96.9 | 95.1 | 98.4 |
| 7 | Excitement,Immobility/stupor,Posturing/catalepsy,Stereotypy,Mannerisms,Verbigeration,Negativism | 96.9 | 95.1 | 98.4 |
| 7 | Excitement,Immobility/stupor,Staring,Grimacing,Echopraxia/echolalia,Mannerisms,Verbigeration | 96.9 | 95.1 | 98.4 |
| 7 | Excitement,Immobility/stupor,Staring,Grimacing,Echopraxia/echolalia,Stereotypy,Verbigeration | 96.9 | 95.1 | 98.4 |
| 7 | Excitement,Immobility/stupor,Staring,Grimacing,Stereotypy,Mannerisms,Verbigeration | 96.9 | 95.1 | 98.4 |
| 7 | Excitement,Staring,Echopraxia/echolalia,Mannerisms,Verbigeration,Negativism,Waxy flexibility | 96.9 | 95.1 | 98.4 |
| 7 | Excitement,Staring,Echopraxia/echolalia,Stereotypy,Mannerisms,Rigidity,Waxy flexibility | 96.9 | 95.1 | 98.4 |
| 7 | Excitement,Staring,Echopraxia/echolalia,Stereotypy,Verbigeration,Negativism,Waxy flexibility | 96.9 | 95.1 | 98.4 |
| 7 | Excitement,Staring,Echopraxia/echolalia,Stereotypy,Verbigeration,Waxy flexibility,Withdrawal | 96.9 | 95.1 | 98.4 |
| 7 | Excitement,Staring,Grimacing,Echopraxia/echolalia,Mannerisms,Verbigeration,Withdrawal | 96.9 | 95.3 | 98.4 |
| 7 | Excitement,Staring,Grimacing,Echopraxia/echolalia,Stereotypy,Verbigeration,Withdrawal | 96.9 | 95.1 | 98.4 |
| 7 | Excitement,Staring,Grimacing,Stereotypy,Verbigeration,Waxy flexibility,Withdrawal | 96.9 | 95.3 | 98.4 |
| 7 | Excitement,Staring,Posturing/catalepsy,Echopraxia/echolalia,Mannerisms,Rigidity,Waxy flexibility | 96.9 | 95.1 | 98.4 |
| 7 | Excitement,Staring,Posturing/catalepsy,Echopraxia/echolalia,Stereotypy,Verbigeration,Withdrawal | 96.9 | 95.1 | 98.4 |
| 7 | Excitement,Staring,Posturing/catalepsy,Echopraxia/echolalia,Verbigeration,Rigidity,Waxy flexibility | 96.9 | 95.1 | 98.4 |
| 7 | Excitement,Staring,Posturing/catalepsy,Grimacing,Mannerisms,Negativism,Waxy flexibility | 96.9 | 95.1 | 98.4 |
| 7 | Excitement,Staring,Posturing/catalepsy,Grimacing,Stereotypy,Mannerisms,Negativism | 96.9 | 95.1 | 98.4 |
| 7 | Excitement,Staring,Posturing/catalepsy,Grimacing,Verbigeration,Negativism,Waxy flexibility | 96.9 | 95.1 | 98.4 |
| 7 | Excitement,Staring,Posturing/catalepsy,Stereotypy,Verbigeration,Rigidity,Waxy flexibility | 96.9 | 95.1 | 98.4 |
| 7 | Excitement,Staring,Stereotypy,Mannerisms,Verbigeration,Negativism,Waxy flexibility | 96.9 | 95.1 | 98.4 |
| 7 | Immobility/stupor,Echopraxia/echolalia,Stereotypy,Mannerisms,Negativism,Waxy flexibility,Withdrawal | 96.9 | 95.1 | 98.4 |
| 7 | Immobility/stupor,Echopraxia/echolalia,Verbigeration,Rigidity,Negativism,Waxy flexibility,Withdrawal | 96.9 | 95.3 | 98.4 |
| 7 | Immobility/stupor,Grimacing,Echopraxia/echolalia,Stereotypy,Mannerisms,Rigidity,Withdrawal | 96.9 | 95.3 | 98.4 |
| 7 | Immobility/stupor,Grimacing,Echopraxia/echolalia,Stereotypy,Verbigeration,Rigidity,Negativism | 96.9 | 95.1 | 98.4 |
| 7 | Immobility/stupor,Grimacing,Stereotypy,Rigidity,Negativism,Waxy flexibility,Withdrawal | 96.9 | 95.3 | 98.4 |
| 7 | Immobility/stupor,Mutism,Echopraxia/echolalia,Stereotypy,Mannerisms,Negativism,Withdrawal | 96.9 | 95.1 | 98.4 |
| 7 | Immobility/stupor,Mutism,Grimacing,Echopraxia/echolalia,Verbigeration,Rigidity,Negativism | 96.9 | 95.1 | 98.4 |
| 7 | Immobility/stupor,Mutism,Grimacing,Mannerisms,Verbigeration,Rigidity,Withdrawal | 96.9 | 95.1 | 98.4 |
| 7 | Immobility/stupor,Mutism,Grimacing,Stereotypy,Mannerisms,Rigidity,Withdrawal | 96.9 | 95.1 | 98.4 |
| 7 | Immobility/stupor,Mutism,Grimacing,Stereotypy,Verbigeration,Rigidity,Withdrawal | 96.9 | 95.1 | 98.4 |
| 7 | Immobility/stupor,Mutism,Posturing/catalepsy,Grimacing,Echopraxia/echolalia,Mannerisms,Negativism | 96.9 | 95.1 | 98.4 |
| 7 | Immobility/stupor,Mutism,Posturing/catalepsy,Grimacing,Mannerisms,Rigidity,Withdrawal | 96.9 | 95.1 | 98.4 |
| 7 | Immobility/stupor,Mutism,Posturing/catalepsy,Grimacing,Verbigeration,Rigidity,Negativism | 96.9 | 95.1 | 98.4 |
| 7 | Immobility/stupor,Mutism,Posturing/catalepsy,Stereotypy,Mannerisms,Negativism,Withdrawal | 96.9 | 95.1 | 98.4 |
| 7 | Immobility/stupor,Mutism,Posturing/catalepsy,Verbigeration,Rigidity,Negativism,Withdrawal | 96.9 | 95.1 | 98.4 |
| 7 | Immobility/stupor,Mutism,Staring,Echopraxia/echolalia,Mannerisms,Waxy flexibility,Withdrawal | 96.9 | 95.1 | 98.4 |
| 7 | Immobility/stupor,Mutism,Staring,Echopraxia/echolalia,Negativism,Waxy flexibility,Withdrawal | 96.9 | 95.1 | 98.4 |
| 7 | Immobility/stupor,Mutism,Staring,Echopraxia/echolalia,Rigidity,Negativism,Waxy flexibility | 96.9 | 95.1 | 98.4 |
| 7 | Immobility/stupor,Mutism,Staring,Echopraxia/echolalia,Stereotypy,Negativism,Waxy flexibility | 96.9 | 95.3 | 98.4 |
| 7 | Immobility/stupor,Mutism,Staring,Echopraxia/echolalia,Verbigeration,Negativism,Waxy flexibility | 96.9 | 95.1 | 98.4 |
| 7 | Immobility/stupor,Mutism,Staring,Grimacing,Echopraxia/echolalia,Mannerisms,Waxy flexibility | 96.9 | 95.1 | 98.4 |
| 7 | Immobility/stupor,Mutism,Staring,Grimacing,Echopraxia/echolalia,Stereotypy,Mannerisms | 96.9 | 95.3 | 98.4 |
| 7 | Immobility/stupor,Mutism,Staring,Grimacing,Echopraxia/echolalia,Stereotypy,Rigidity | 96.9 | 95.1 | 98.4 |
| 7 | Immobility/stupor,Mutism,Staring,Grimacing,Echopraxia/echolalia,Stereotypy,Withdrawal | 96.9 | 95.1 | 98.4 |
| 7 | Immobility/stupor,Mutism,Staring,Grimacing,Mannerisms,Verbigeration,Withdrawal | 96.9 | 95.1 | 98.4 |
| 7 | Immobility/stupor,Mutism,Staring,Grimacing,Stereotypy,Verbigeration,Withdrawal | 96.9 | 95.1 | 98.4 |
| 7 | Immobility/stupor,Mutism,Staring,Grimacing,Verbigeration,Rigidity,Waxy flexibility | 96.9 | 95.1 | 98.4 |
| 7 | Immobility/stupor,Mutism,Staring,Posturing/catalepsy,Echopraxia/echolalia,Mannerisms,Verbigeration | 96.9 | 95.1 | 98.4 |
| 7 | Immobility/stupor,Mutism,Staring,Posturing/catalepsy,Echopraxia/echolalia,Stereotypy,Mannerisms | 96.9 | 95.1 | 98.4 |
| 7 | Immobility/stupor,Mutism,Staring,Posturing/catalepsy,Echopraxia/echolalia,Stereotypy,Withdrawal | 96.9 | 95.1 | 98.4 |
| 7 | Immobility/stupor,Mutism,Staring,Posturing/catalepsy,Grimacing,Echopraxia/echolalia,Withdrawal | 96.9 | 95.1 | 98.4 |
| 7 | Immobility/stupor,Mutism,Staring,Posturing/catalepsy,Grimacing,Stereotypy,Mannerisms | 96.9 | 95.1 | 98.4 |
| 7 | Immobility/stupor,Mutism,Staring,Posturing/catalepsy,Grimacing,Stereotypy,Rigidity | 96.9 | 95.1 | 98.4 |
| 7 | Immobility/stupor,Mutism,Staring,Posturing/catalepsy,Grimacing,Stereotypy,Verbigeration | 96.9 | 95.1 | 98.4 |
| 7 | Immobility/stupor,Mutism,Staring,Posturing/catalepsy,Grimacing,Stereotypy,Withdrawal | 96.9 | 95.1 | 98.4 |
| 7 | Immobility/stupor,Mutism,Staring,Posturing/catalepsy,Mannerisms,Rigidity,Waxy flexibility | 96.9 | 95.1 | 98.4 |
| 7 | Immobility/stupor,Mutism,Staring,Posturing/catalepsy,Stereotypy,Mannerisms,Verbigeration | 96.9 | 95.1 | 98.4 |
| 7 | Immobility/stupor,Mutism,Staring,Stereotypy,Mannerisms,Waxy flexibility,Withdrawal | 96.9 | 95.1 | 98.4 |
| 7 | Immobility/stupor,Mutism,Staring,Verbigeration,Negativism,Waxy flexibility,Withdrawal | 96.9 | 95.1 | 98.4 |
| 7 | Immobility/stupor,Mutism,Staring,Verbigeration,Rigidity,Waxy flexibility,Withdrawal | 96.9 | 95.1 | 98.4 |
| 7 | Immobility/stupor,Mutism,Stereotypy,Mannerisms,Verbigeration,Negativism,Withdrawal | 96.9 | 95.1 | 98.4 |
| 7 | Immobility/stupor,Mutism,Stereotypy,Rigidity,Negativism,Waxy flexibility,Withdrawal | 96.9 | 95.1 | 98.4 |
| 7 | Immobility/stupor,Mutism,Verbigeration,Rigidity,Negativism,Waxy flexibility,Withdrawal | 96.9 | 95.1 | 98.4 |
| 7 | Immobility/stupor,Posturing/catalepsy,Echopraxia/echolalia,Mannerisms,Verbigeration,Negativism,Withdrawal | 96.9 | 95.3 | 98.4 |
| 7 | Immobility/stupor,Posturing/catalepsy,Echopraxia/echolalia,Stereotypy,Verbigeration,Rigidity,Negativism | 96.9 | 95.1 | 98.4 |
| 7 | Immobility/stupor,Posturing/catalepsy,Grimacing,Echopraxia/echolalia,Verbigeration,Rigidity,Negativism | 96.9 | 95.1 | 98.4 |
| 7 | Immobility/stupor,Posturing/catalepsy,Grimacing,Mannerisms,Verbigeration,Negativism,Withdrawal | 96.9 | 95.3 | 98.4 |
| 7 | Immobility/stupor,Posturing/catalepsy,Stereotypy,Rigidity,Negativism,Waxy flexibility,Withdrawal | 96.9 | 95.1 | 98.4 |
| 7 | Immobility/stupor,Posturing/catalepsy,Verbigeration,Rigidity,Negativism,Waxy flexibility,Withdrawal | 96.9 | 95.3 | 98.4 |
| 7 | Immobility/stupor,Staring,Echopraxia/echolalia,Mannerisms,Verbigeration,Rigidity,Waxy flexibility | 96.9 | 95.1 | 98.4 |
| 7 | Immobility/stupor,Staring,Echopraxia/echolalia,Stereotypy,Mannerisms,Rigidity,Waxy flexibility | 96.9 | 95.3 | 98.4 |
| 7 | Immobility/stupor,Staring,Echopraxia/echolalia,Stereotypy,Rigidity,Waxy flexibility,Withdrawal | 96.9 | 95.1 | 98.4 |
| 7 | Immobility/stupor,Staring,Grimacing,Echopraxia/echolalia,Mannerisms,Waxy flexibility,Withdrawal | 96.9 | 95.1 | 98.4 |
| 7 | Immobility/stupor,Staring,Grimacing,Echopraxia/echolalia,Rigidity,Negativism,Waxy flexibility | 96.9 | 95.1 | 98.4 |
| 7 | Immobility/stupor,Staring,Grimacing,Echopraxia/echolalia,Stereotypy,Mannerisms,Waxy flexibility | 96.9 | 95.1 | 98.4 |
| 7 | Immobility/stupor,Staring,Grimacing,Echopraxia/echolalia,Stereotypy,Negativism,Waxy flexibility | 96.9 | 95.1 | 98.4 |
| 7 | Immobility/stupor,Staring,Grimacing,Echopraxia/echolalia,Verbigeration,Negativism,Waxy flexibility | 96.9 | 95.1 | 98.4 |
| 7 | Immobility/stupor,Staring,Grimacing,Echopraxia/echolalia,Verbigeration,Rigidity,Withdrawal | 96.9 | 95.1 | 98.4 |
| 7 | Immobility/stupor,Staring,Grimacing,Stereotypy,Negativism,Waxy flexibility,Withdrawal | 96.9 | 95.1 | 98.4 |
| 7 | Immobility/stupor,Staring,Posturing/catalepsy,Echopraxia/echolalia,Rigidity,Negativism,Waxy flexibility | 96.9 | 95.1 | 98.4 |
| 7 | Immobility/stupor,Staring,Posturing/catalepsy,Echopraxia/echolalia,Stereotypy,Verbigeration,Rigidity | 96.9 | 95.1 | 98.4 |
| 7 | Immobility/stupor,Staring,Posturing/catalepsy,Grimacing,Echopraxia/echolalia,Rigidity,Withdrawal | 96.9 | 95.1 | 98.4 |
| 7 | Immobility/stupor,Staring,Posturing/catalepsy,Grimacing,Echopraxia/echolalia,Stereotypy,Mannerisms | 96.9 | 95.1 | 98.4 |
| 7 | Immobility/stupor,Staring,Posturing/catalepsy,Grimacing,Stereotypy,Mannerisms,Verbigeration | 96.9 | 95.1 | 98.4 |
| 7 | Immobility/stupor,Staring,Posturing/catalepsy,Mannerisms,Verbigeration,Rigidity,Waxy flexibility | 96.9 | 95.1 | 98.4 |
| 7 | Immobility/stupor,Staring,Posturing/catalepsy,Stereotypy,Mannerisms,Verbigeration,Withdrawal | 96.9 | 95.1 | 98.4 |
| 7 | Immobility/stupor,Staring,Posturing/catalepsy,Stereotypy,Verbigeration,Rigidity,Waxy flexibility | 96.9 | 95.1 | 98.4 |
| 7 | Mutism,Staring,Echopraxia/echolalia,Mannerisms,Verbigeration,Waxy flexibility,Withdrawal | 96.9 | 95.1 | 98.4 |
| 7 | Mutism,Staring,Echopraxia/echolalia,Stereotypy,Mannerisms,Verbigeration,Waxy flexibility | 96.9 | 95.3 | 98.4 |
| 7 | Mutism,Staring,Grimacing,Echopraxia/echolalia,Verbigeration,Waxy flexibility,Withdrawal | 96.9 | 95.1 | 98.4 |
| 7 | Mutism,Staring,Posturing/catalepsy,Grimacing,Echopraxia/echolalia,Stereotypy,Waxy flexibility | 96.9 | 95.1 | 98.4 |
| 7 | Mutism,Staring,Posturing/catalepsy,Grimacing,Echopraxia/echolalia,Verbigeration,Waxy flexibility | 96.9 | 95.1 | 98.4 |
| 7 | Mutism,Staring,Posturing/catalepsy,Grimacing,Verbigeration,Rigidity,Waxy flexibility | 96.9 | 95.1 | 98.4 |
| 7 | Mutism,Staring,Posturing/catalepsy,Grimacing,Verbigeration,Waxy flexibility,Withdrawal | 96.9 | 95.3 | 98.4 |
| 7 | Staring,Echopraxia/echolalia,Stereotypy,Verbigeration,Negativism,Waxy flexibility,Withdrawal | 96.9 | 95.1 | 98.4 |
| 7 | Staring,Grimacing,Echopraxia/echolalia,Mannerisms,Verbigeration,Rigidity,Negativism | 96.9 | 95.3 | 98.4 |
| 7 | Staring,Grimacing,Echopraxia/echolalia,Stereotypy,Mannerisms,Verbigeration,Withdrawal | 96.9 | 95.1 | 98.4 |
| 7 | Staring,Grimacing,Echopraxia/echolalia,Stereotypy,Negativism,Waxy flexibility,Withdrawal | 96.9 | 95.1 | 98.4 |
| 7 | Staring,Grimacing,Mannerisms,Verbigeration,Rigidity,Negativism,Waxy flexibility | 96.9 | 95.3 | 98.4 |
| 7 | Staring,Grimacing,Stereotypy,Mannerisms,Verbigeration,Rigidity,Waxy flexibility | 96.9 | 95.1 | 98.4 |
| 7 | Staring,Grimacing,Stereotypy,Verbigeration,Rigidity,Negativism,Waxy flexibility | 96.9 | 95.1 | 98.4 |
| 7 | Staring,Posturing/catalepsy,Echopraxia/echolalia,Stereotypy,Mannerisms,Negativism,Waxy flexibility | 96.9 | 95.1 | 98.4 |
| 7 | Staring,Posturing/catalepsy,Echopraxia/echolalia,Stereotypy,Verbigeration,Rigidity,Negativism | 96.9 | 95.1 | 98.4 |
| 7 | Staring,Posturing/catalepsy,Grimacing,Echopraxia/echolalia,Mannerisms,Verbigeration,Withdrawal | 96.9 | 95.1 | 98.4 |
| 7 | Staring,Posturing/catalepsy,Grimacing,Echopraxia/echolalia,Mannerisms,Waxy flexibility,Withdrawal | 96.9 | 95.1 | 98.4 |
| 7 | Staring,Posturing/catalepsy,Grimacing,Echopraxia/echolalia,Negativism,Waxy flexibility,Withdrawal | 96.9 | 95.1 | 98.4 |
| 7 | Staring,Posturing/catalepsy,Grimacing,Rigidity,Negativism,Waxy flexibility,Withdrawal | 96.9 | 95.1 | 98.4 |
| 7 | Staring,Posturing/catalepsy,Grimacing,Stereotypy,Mannerisms,Verbigeration,Negativism | 96.9 | 95.1 | 98.4 |
| 7 | Staring,Posturing/catalepsy,Grimacing,Stereotypy,Mannerisms,Verbigeration,Rigidity | 96.9 | 95.3 | 98.4 |
| 7 | Staring,Posturing/catalepsy,Grimacing,Stereotypy,Negativism,Waxy flexibility,Withdrawal | 96.9 | 95.1 | 98.4 |
| 7 | Staring,Posturing/catalepsy,Grimacing,Stereotypy,Verbigeration,Rigidity,Negativism | 96.9 | 95.1 | 98.4 |
| 7 | Staring,Posturing/catalepsy,Mannerisms,Verbigeration,Rigidity,Negativism,Waxy flexibility | 96.9 | 95.1 | 98.4 |
| 7 | Excitement,Immobility/stupor,Mutism,Echopraxia/echolalia,Stereotypy,Verbigeration,Withdrawal | 96.8 | 95.1 | 98.4 |
| 7 | Excitement,Staring,Echopraxia/echolalia,Stereotypy,Verbigeration,Rigidity,Waxy flexibility | 96.8 | 95.1 | 98.4 |
| 7 | Excitement,Staring,Grimacing,Echopraxia/echolalia,Mannerisms,Verbigeration,Rigidity | 96.8 | 95.1 | 98.4 |
| 7 | Immobility/stupor,Staring,Echopraxia/echolalia,Verbigeration,Negativism,Waxy flexibility,Withdrawal | 96.8 | 95.1 | 98.4 |
| 7 | Immobility/stupor,Staring,Posturing/catalepsy,Grimacing,Verbigeration,Rigidity,Withdrawal | 96.8 | 95.3 | 98.4 |
| 7 | Mutism,Staring,Posturing/catalepsy,Echopraxia/echolalia,Stereotypy,Verbigeration,Waxy flexibility | 96.8 | 95.1 | 98.4 |
| 7 | Staring,Grimacing,Stereotypy,Mannerisms,Verbigeration,Waxy flexibility,Withdrawal | 96.8 | 95.3 | 98.4 |
| 7 | Excitement,Staring,Echopraxia/echolalia,Mannerisms,Verbigeration,Rigidity,Waxy flexibility | 96.7 | 94.8 | 98.2 |
| 7 | Immobility/stupor,Mutism,Staring,Posturing/catalepsy,Verbigeration,Negativism,Waxy flexibility | 96.7 | 94.8 | 98.2 |
| 7 | Immobility/stupor,Staring,Grimacing,Verbigeration,Negativism,Waxy flexibility,Withdrawal | 96.7 | 94.8 | 98.2 |
| 7 | Immobility/stupor,Staring,Posturing/catalepsy,Grimacing,Echopraxia/echolalia,Verbigeration,Rigidity | 96.7 | 94.8 | 98.2 |
| 7 | Mutism,Staring,Echopraxia/echolalia,Stereotypy,Verbigeration,Waxy flexibility,Withdrawal | 96.7 | 94.8 | 98.2 |
| 7 | Staring,Posturing/catalepsy,Echopraxia/echolalia,Stereotypy,Verbigeration,Rigidity,Withdrawal | 96.7 | 94.8 | 98.2 |
| 7 | Excitement,Immobility/stupor,Echopraxia/echolalia,Stereotypy,Mannerisms,Rigidity,Waxy flexibility | 96.6 | 94.8 | 98.2 |
| 7 | Excitement,Immobility/stupor,Echopraxia/echolalia,Stereotypy,Verbigeration,Negativism,Waxy flexibility | 96.6 | 94.8 | 98.2 |
| 7 | Excitement,Immobility/stupor,Grimacing,Echopraxia/echolalia,Mannerisms,Negativism,Withdrawal | 96.6 | 94.8 | 98.2 |
| 7 | Excitement,Immobility/stupor,Grimacing,Echopraxia/echolalia,Stereotypy,Mannerisms,Rigidity | 96.6 | 94.8 | 98.2 |
| 7 | Excitement,Immobility/stupor,Grimacing,Echopraxia/echolalia,Stereotypy,Negativism,Waxy flexibility | 96.6 | 94.8 | 98.2 |
| 7 | Excitement,Immobility/stupor,Grimacing,Echopraxia/echolalia,Verbigeration,Negativism,Waxy flexibility | 96.6 | 94.8 | 98.2 |
| 7 | Excitement,Immobility/stupor,Grimacing,Mannerisms,Verbigeration,Negativism,Withdrawal | 96.6 | 94.8 | 98.2 |
| 7 | Excitement,Immobility/stupor,Mannerisms,Verbigeration,Negativism,Waxy flexibility,Withdrawal | 96.6 | 94.8 | 98.2 |
| 7 | Excitement,Immobility/stupor,Mutism,Echopraxia/echolalia,Mannerisms,Negativism,Withdrawal | 96.6 | 94.8 | 98.2 |
| 7 | Excitement,Immobility/stupor,Mutism,Echopraxia/echolalia,Verbigeration,Waxy flexibility,Withdrawal | 96.6 | 94.8 | 98.2 |
| 7 | Excitement,Immobility/stupor,Mutism,Grimacing,Echopraxia/echolalia,Stereotypy,Negativism | 96.6 | 94.8 | 98.2 |
| 7 | Excitement,Immobility/stupor,Mutism,Grimacing,Echopraxia/echolalia,Stereotypy,Withdrawal | 96.6 | 94.8 | 98.2 |
| 7 | Excitement,Immobility/stupor,Mutism,Grimacing,Echopraxia/echolalia,Verbigeration,Negativism | 96.6 | 94.8 | 98.2 |
| 7 | Excitement,Immobility/stupor,Mutism,Mannerisms,Negativism,Waxy flexibility,Withdrawal | 96.6 | 94.8 | 98.2 |
| 7 | Excitement,Immobility/stupor,Mutism,Mannerisms,Verbigeration,Negativism,Waxy flexibility | 96.6 | 94.8 | 98.2 |
| 7 | Excitement,Immobility/stupor,Mutism,Posturing/catalepsy,Echopraxia/echolalia,Stereotypy,Verbigeration | 96.6 | 94.8 | 98.2 |
| 7 | Excitement,Immobility/stupor,Mutism,Posturing/catalepsy,Grimacing,Echopraxia/echolalia,Mannerisms | 96.6 | 94.8 | 98.2 |
| 7 | Excitement,Immobility/stupor,Mutism,Posturing/catalepsy,Grimacing,Echopraxia/echolalia,Stereotypy | 96.6 | 94.8 | 98.2 |
| 7 | Excitement,Immobility/stupor,Mutism,Posturing/catalepsy,Grimacing,Mannerisms,Verbigeration | 96.6 | 94.8 | 98.2 |
| 7 | Excitement,Immobility/stupor,Mutism,Posturing/catalepsy,Grimacing,Stereotypy,Verbigeration | 96.6 | 94.8 | 98.2 |
| 7 | Excitement,Immobility/stupor,Mutism,Posturing/catalepsy,Mannerisms,Waxy flexibility,Withdrawal | 96.6 | 94.8 | 98.2 |
| 7 | Excitement,Immobility/stupor,Mutism,Posturing/catalepsy,Stereotypy,Mannerisms,Withdrawal | 96.6 | 94.8 | 98.2 |
| 7 | Excitement,Immobility/stupor,Mutism,Stereotypy,Mannerisms,Verbigeration,Negativism | 96.6 | 94.8 | 98.2 |
| 7 | Excitement,Immobility/stupor,Mutism,Stereotypy,Verbigeration,Waxy flexibility,Withdrawal | 96.6 | 94.8 | 98.2 |
| 7 | Excitement,Immobility/stupor,Posturing/catalepsy,Echopraxia/echolalia,Mannerisms,Negativism,Waxy flexibility | 96.6 | 94.8 | 98.2 |
| 7 | Excitement,Immobility/stupor,Posturing/catalepsy,Echopraxia/echolalia,Stereotypy,Negativism,Waxy flexibility | 96.6 | 94.8 | 98.2 |
| 7 | Excitement,Immobility/stupor,Posturing/catalepsy,Echopraxia/echolalia,Stereotypy,Verbigeration,Withdrawal | 96.6 | 94.8 | 98.2 |
| 7 | Excitement,Immobility/stupor,Posturing/catalepsy,Echopraxia/echolalia,Verbigeration,Waxy flexibility,Withdrawal | 96.6 | 94.8 | 98.2 |
| 7 | Excitement,Immobility/stupor,Posturing/catalepsy,Grimacing,Echopraxia/echolalia,Stereotypy,Withdrawal | 96.6 | 94.8 | 98.2 |
| 7 | Excitement,Immobility/stupor,Posturing/catalepsy,Grimacing,Mannerisms,Verbigeration,Withdrawal | 96.6 | 94.8 | 98.2 |
| 7 | Excitement,Immobility/stupor,Posturing/catalepsy,Grimacing,Stereotypy,Negativism,Waxy flexibility | 96.6 | 94.8 | 98.2 |
| 7 | Excitement,Immobility/stupor,Stereotypy,Mannerisms,Verbigeration,Rigidity,Waxy flexibility | 96.6 | 94.8 | 98.2 |
| 7 | Excitement,Staring,Echopraxia/echolalia,Stereotypy,Mannerisms,Verbigeration,Rigidity | 96.6 | 94.8 | 98.2 |
| 7 | Excitement,Staring,Echopraxia/echolalia,Stereotypy,Mannerisms,Verbigeration,Withdrawal | 96.6 | 94.8 | 98.2 |
| 7 | Excitement,Staring,Grimacing,Echopraxia/echolalia,Mannerisms,Negativism,Waxy flexibility | 96.6 | 94.8 | 98.2 |
| 7 | Excitement,Staring,Grimacing,Echopraxia/echolalia,Stereotypy,Negativism,Waxy flexibility | 96.6 | 94.8 | 98.2 |
| 7 | Excitement,Staring,Posturing/catalepsy,Echopraxia/echolalia,Stereotypy,Rigidity,Waxy flexibility | 96.6 | 94.8 | 98.2 |
| 7 | Excitement,Staring,Posturing/catalepsy,Grimacing,Stereotypy,Negativism,Waxy flexibility | 96.6 | 94.8 | 98.2 |
| 7 | Excitement,Staring,Posturing/catalepsy,Stereotypy,Mannerisms,Rigidity,Waxy flexibility | 96.6 | 94.8 | 98.2 |
| 7 | Immobility/stupor,Echopraxia/echolalia,Stereotypy,Verbigeration,Rigidity,Negativism,Waxy flexibility | 96.6 | 94.8 | 98.2 |
| 7 | Immobility/stupor,Grimacing,Echopraxia/echolalia,Rigidity,Negativism,Waxy flexibility,Withdrawal | 96.6 | 94.8 | 98.2 |
| 7 | Immobility/stupor,Grimacing,Echopraxia/echolalia,Verbigeration,Rigidity,Negativism,Waxy flexibility | 96.6 | 94.8 | 98.2 |
| 7 | Immobility/stupor,Grimacing,Stereotypy,Mannerisms,Verbigeration,Negativism,Withdrawal | 96.6 | 94.8 | 98.2 |
| 7 | Immobility/stupor,Mutism,Echopraxia/echolalia,Verbigeration,Rigidity,Negativism,Waxy flexibility | 96.6 | 94.8 | 98.2 |
| 7 | Immobility/stupor,Mutism,Grimacing,Echopraxia/echolalia,Rigidity,Negativism,Withdrawal | 96.6 | 94.8 | 98.2 |
| 7 | Immobility/stupor,Mutism,Grimacing,Echopraxia/echolalia,Stereotypy,Rigidity,Withdrawal | 96.6 | 94.8 | 98.2 |
| 7 | Immobility/stupor,Mutism,Grimacing,Mannerisms,Rigidity,Waxy flexibility,Withdrawal | 96.6 | 94.8 | 98.2 |
| 7 | Immobility/stupor,Mutism,Grimacing,Verbigeration,Rigidity,Negativism,Waxy flexibility | 96.6 | 94.8 | 98.2 |
| 7 | Immobility/stupor,Mutism,Posturing/catalepsy,Echopraxia/echolalia,Rigidity,Negativism,Withdrawal | 96.6 | 94.8 | 98.2 |
| 7 | Immobility/stupor,Mutism,Posturing/catalepsy,Echopraxia/echolalia,Stereotypy,Rigidity,Negativism | 96.6 | 94.8 | 98.2 |
| 7 | Immobility/stupor,Mutism,Posturing/catalepsy,Grimacing,Stereotypy,Rigidity,Negativism | 96.6 | 94.8 | 98.2 |
| 7 | Immobility/stupor,Mutism,Posturing/catalepsy,Mannerisms,Negativism,Waxy flexibility,Withdrawal | 96.6 | 94.8 | 98.2 |
| 7 | Immobility/stupor,Mutism,Posturing/catalepsy,Stereotypy,Verbigeration,Negativism,Withdrawal | 96.6 | 94.8 | 98.2 |
| 7 | Immobility/stupor,Mutism,Staring,Echopraxia/echolalia,Rigidity,Waxy flexibility,Withdrawal | 96.6 | 94.8 | 98.2 |
| 7 | Immobility/stupor,Mutism,Staring,Echopraxia/echolalia,Stereotypy,Mannerisms,Waxy flexibility | 96.6 | 94.8 | 98.2 |
| 7 | Immobility/stupor,Mutism,Staring,Echopraxia/echolalia,Verbigeration,Rigidity,Waxy flexibility | 96.6 | 94.8 | 98.2 |
| 7 | Immobility/stupor,Mutism,Staring,Grimacing,Echopraxia/echolalia,Mannerisms,Verbigeration | 96.6 | 94.8 | 98.2 |
| 7 | Immobility/stupor,Mutism,Staring,Grimacing,Mannerisms,Verbigeration,Waxy flexibility | 96.6 | 94.8 | 98.2 |
| 7 | Immobility/stupor,Mutism,Staring,Grimacing,Stereotypy,Mannerisms,Verbigeration | 96.6 | 94.8 | 98.2 |
| 7 | Immobility/stupor,Mutism,Staring,Grimacing,Stereotypy,Mannerisms,Waxy flexibility | 96.6 | 94.8 | 98.2 |
| 7 | Immobility/stupor,Mutism,Staring,Grimacing,Stereotypy,Negativism,Waxy flexibility | 96.6 | 94.8 | 98.2 |
| 7 | Immobility/stupor,Mutism,Staring,Grimacing,Stereotypy,Rigidity,Waxy flexibility | 96.6 | 94.8 | 98.2 |
| 7 | Immobility/stupor,Mutism,Staring,Grimacing,Stereotypy,Verbigeration,Waxy flexibility | 96.6 | 94.8 | 98.2 |
| 7 | Immobility/stupor,Mutism,Staring,Grimacing,Verbigeration,Negativism,Waxy flexibility | 96.6 | 94.8 | 98.2 |
| 7 | Immobility/stupor,Mutism,Staring,Posturing/catalepsy,Echopraxia/echolalia,Negativism,Waxy flexibility | 96.6 | 94.8 | 98.2 |
| 7 | Immobility/stupor,Mutism,Staring,Posturing/catalepsy,Grimacing,Echopraxia/echolalia,Verbigeration | 96.6 | 94.8 | 98.2 |
| 7 | Immobility/stupor,Mutism,Staring,Posturing/catalepsy,Grimacing,Rigidity,Withdrawal | 96.6 | 94.8 | 98.2 |
| 7 | Immobility/stupor,Mutism,Staring,Posturing/catalepsy,Grimacing,Verbigeration,Withdrawal | 96.6 | 94.8 | 98.2 |
| 7 | Immobility/stupor,Mutism,Staring,Posturing/catalepsy,Stereotypy,Verbigeration,Withdrawal | 96.6 | 94.8 | 98.2 |
| 7 | Immobility/stupor,Mutism,Staring,Stereotypy,Mannerisms,Verbigeration,Withdrawal | 96.6 | 94.8 | 98.2 |
| 7 | Immobility/stupor,Posturing/catalepsy,Grimacing,Echopraxia/echolalia,Mannerisms,Rigidity,Withdrawal | 96.6 | 94.8 | 98.2 |
| 7 | Immobility/stupor,Posturing/catalepsy,Grimacing,Echopraxia/echolalia,Stereotypy,Negativism,Withdrawal | 96.6 | 94.8 | 98.2 |
| 7 | Immobility/stupor,Posturing/catalepsy,Grimacing,Stereotypy,Verbigeration,Negativism,Withdrawal | 96.6 | 94.8 | 98.2 |
| 7 | Immobility/stupor,Posturing/catalepsy,Grimacing,Verbigeration,Rigidity,Negativism,Waxy flexibility | 96.6 | 94.8 | 98.2 |
| 7 | Immobility/stupor,Posturing/catalepsy,Stereotypy,Mannerisms,Negativism,Waxy flexibility,Withdrawal | 96.6 | 94.8 | 98.2 |
| 7 | Immobility/stupor,Staring,Echopraxia/echolalia,Stereotypy,Mannerisms,Waxy flexibility,Withdrawal | 96.6 | 94.8 | 98.2 |
| 7 | Immobility/stupor,Staring,Grimacing,Echopraxia/echolalia,Negativism,Waxy flexibility,Withdrawal | 96.6 | 94.8 | 98.2 |
| 7 | Immobility/stupor,Staring,Grimacing,Stereotypy,Mannerisms,Verbigeration,Waxy flexibility | 96.6 | 94.8 | 98.2 |
| 7 | Immobility/stupor,Staring,Grimacing,Verbigeration,Rigidity,Waxy flexibility,Withdrawal | 96.6 | 94.8 | 98.2 |
| 7 | Immobility/stupor,Staring,Posturing/catalepsy,Echopraxia/echolalia,Verbigeration,Negativism,Waxy flexibility | 96.6 | 94.8 | 98.2 |
| 7 | Immobility/stupor,Staring,Posturing/catalepsy,Echopraxia/echolalia,Verbigeration,Rigidity,Withdrawal | 96.6 | 94.8 | 98.2 |
| 7 | Immobility/stupor,Staring,Posturing/catalepsy,Grimacing,Echopraxia/echolalia,Mannerisms,Verbigeration | 96.6 | 94.8 | 98.2 |
| 7 | Immobility/stupor,Staring,Posturing/catalepsy,Grimacing,Echopraxia/echolalia,Stereotypy,Rigidity | 96.6 | 94.8 | 98.2 |
| 7 | Immobility/stupor,Staring,Posturing/catalepsy,Grimacing,Echopraxia/echolalia,Stereotypy,Withdrawal | 96.6 | 94.8 | 98.2 |
| 7 | Immobility/stupor,Staring,Posturing/catalepsy,Grimacing,Mannerisms,Rigidity,Waxy flexibility | 96.6 | 94.8 | 98.2 |
| 7 | Immobility/stupor,Staring,Posturing/catalepsy,Grimacing,Mannerisms,Waxy flexibility,Withdrawal | 96.6 | 94.8 | 98.2 |
| 7 | Immobility/stupor,Staring,Posturing/catalepsy,Grimacing,Negativism,Waxy flexibility,Withdrawal | 96.6 | 94.8 | 98.2 |
| 7 | Immobility/stupor,Staring,Posturing/catalepsy,Grimacing,Stereotypy,Negativism,Waxy flexibility | 96.6 | 94.8 | 98.2 |
| 7 | Immobility/stupor,Staring,Posturing/catalepsy,Grimacing,Stereotypy,Verbigeration,Withdrawal | 96.6 | 94.8 | 98.2 |
| 7 | Immobility/stupor,Staring,Posturing/catalepsy,Grimacing,Verbigeration,Negativism,Waxy flexibility | 96.6 | 94.8 | 98.2 |
| 7 | Immobility/stupor,Staring,Posturing/catalepsy,Stereotypy,Mannerisms,Rigidity,Waxy flexibility | 96.6 | 94.8 | 98.2 |
| 7 | Immobility/stupor,Staring,Posturing/catalepsy,Stereotypy,Mannerisms,Waxy flexibility,Withdrawal | 96.6 | 94.8 | 98.2 |
| 7 | Immobility/stupor,Staring,Posturing/catalepsy,Stereotypy,Rigidity,Waxy flexibility,Withdrawal | 96.6 | 94.8 | 98.2 |
| 7 | Immobility/stupor,Staring,Posturing/catalepsy,Verbigeration,Rigidity,Negativism,Waxy flexibility | 96.6 | 94.8 | 98.2 |
| 7 | Mutism,Staring,Posturing/catalepsy,Echopraxia/echolalia,Verbigeration,Waxy flexibility,Withdrawal | 96.6 | 94.8 | 98.2 |
| 7 | Mutism,Staring,Posturing/catalepsy,Grimacing,Rigidity,Waxy flexibility,Withdrawal | 96.6 | 94.8 | 98.2 |
| 7 | Mutism,Staring,Posturing/catalepsy,Grimacing,Stereotypy,Rigidity,Waxy flexibility | 96.6 | 94.8 | 98.2 |
| 7 | Staring,Echopraxia/echolalia,Mannerisms,Verbigeration,Rigidity,Waxy flexibility,Withdrawal | 96.6 | 94.8 | 98.2 |
| 7 | Staring,Echopraxia/echolalia,Stereotypy,Verbigeration,Rigidity,Negativism,Waxy flexibility | 96.6 | 94.8 | 98.2 |
| 7 | Staring,Grimacing,Echopraxia/echolalia,Stereotypy,Mannerisms,Negativism,Waxy flexibility | 96.6 | 94.8 | 98.2 |
| 7 | Staring,Grimacing,Echopraxia/echolalia,Stereotypy,Mannerisms,Rigidity,Waxy flexibility | 96.6 | 94.8 | 98.2 |
| 7 | Staring,Grimacing,Echopraxia/echolalia,Stereotypy,Verbigeration,Rigidity,Negativism | 96.6 | 94.8 | 98.2 |
| 7 | Staring,Posturing/catalepsy,Echopraxia/echolalia,Stereotypy,Mannerisms,Verbigeration,Negativism | 96.6 | 94.8 | 98.2 |
| 7 | Staring,Posturing/catalepsy,Echopraxia/echolalia,Verbigeration,Negativism,Waxy flexibility,Withdrawal | 96.6 | 94.8 | 98.2 |
| 7 | Staring,Posturing/catalepsy,Grimacing,Echopraxia/echolalia,Mannerisms,Verbigeration,Negativism | 96.6 | 94.8 | 98.2 |
| 7 | Staring,Posturing/catalepsy,Grimacing,Echopraxia/echolalia,Stereotypy,Mannerisms,Rigidity | 96.6 | 94.8 | 98.2 |
| 7 | Staring,Posturing/catalepsy,Grimacing,Echopraxia/echolalia,Verbigeration,Rigidity,Withdrawal | 96.6 | 94.8 | 98.2 |
| 7 | Staring,Posturing/catalepsy,Grimacing,Mannerisms,Rigidity,Negativism,Waxy flexibility | 96.6 | 94.8 | 98.2 |
| 7 | Staring,Posturing/catalepsy,Grimacing,Mannerisms,Verbigeration,Waxy flexibility,Withdrawal | 96.6 | 94.8 | 98.2 |
| 7 | Staring,Posturing/catalepsy,Grimacing,Stereotypy,Rigidity,Waxy flexibility,Withdrawal | 96.6 | 94.8 | 98.2 |
| 7 | Staring,Posturing/catalepsy,Grimacing,Verbigeration,Negativism,Waxy flexibility,Withdrawal | 96.6 | 94.8 | 98.2 |
| 7 | Staring,Posturing/catalepsy,Stereotypy,Mannerisms,Verbigeration,Negativism,Waxy flexibility | 96.6 | 94.8 | 98.2 |
| 7 | Staring,Posturing/catalepsy,Stereotypy,Verbigeration,Rigidity,Negativism,Waxy flexibility | 96.6 | 94.8 | 98.2 |
| 7 | Staring,Posturing/catalepsy,Stereotypy,Verbigeration,Rigidity,Waxy flexibility,Withdrawal | 96.6 | 94.8 | 98.2 |
| 7 | Excitement,Immobility/stupor,Grimacing,Echopraxia/echolalia,Mannerisms,Negativism,Waxy flexibility | 96.4 | 94.6 | 98.0 |
| 7 | Excitement,Immobility/stupor,Grimacing,Stereotypy,Mannerisms,Rigidity,Waxy flexibility | 96.4 | 94.6 | 98.0 |
| 7 | Excitement,Immobility/stupor,Grimacing,Stereotypy,Mannerisms,Verbigeration,Rigidity | 96.4 | 94.6 | 98.0 |
| 7 | Excitement,Immobility/stupor,Grimacing,Stereotypy,Negativism,Waxy flexibility,Withdrawal | 96.4 | 94.6 | 98.0 |
| 7 | Excitement,Immobility/stupor,Grimacing,Stereotypy,Verbigeration,Negativism,Waxy flexibility | 96.4 | 94.6 | 98.0 |
| 7 | Excitement,Immobility/stupor,Mutism,Echopraxia/echolalia,Mannerisms,Negativism,Waxy flexibility | 96.4 | 94.6 | 98.0 |
| 7 | Excitement,Immobility/stupor,Mutism,Grimacing,Echopraxia/echolalia,Verbigeration,Withdrawal | 96.4 | 94.6 | 98.0 |
| 7 | Excitement,Immobility/stupor,Mutism,Grimacing,Mannerisms,Negativism,Waxy flexibility | 96.4 | 94.6 | 98.0 |
| 7 | Excitement,Immobility/stupor,Mutism,Grimacing,Stereotypy,Verbigeration,Withdrawal | 96.4 | 94.6 | 98.0 |
| 7 | Excitement,Immobility/stupor,Mutism,Grimacing,Stereotypy,Waxy flexibility,Withdrawal | 96.4 | 94.6 | 98.0 |
| 7 | Excitement,Immobility/stupor,Mutism,Posturing/catalepsy,Echopraxia/echolalia,Mannerisms,Verbigeration | 96.4 | 94.6 | 98.0 |
| 7 | Excitement,Immobility/stupor,Mutism,Posturing/catalepsy,Echopraxia/echolalia,Stereotypy,Waxy flexibility | 96.4 | 94.6 | 98.0 |
| 7 | Excitement,Immobility/stupor,Mutism,Posturing/catalepsy,Grimacing,Mannerisms,Waxy flexibility | 96.4 | 94.6 | 98.0 |
| 7 | Excitement,Immobility/stupor,Mutism,Posturing/catalepsy,Grimacing,Stereotypy,Waxy flexibility | 96.4 | 94.6 | 98.0 |
| 7 | Excitement,Immobility/stupor,Mutism,Posturing/catalepsy,Stereotypy,Verbigeration,Waxy flexibility | 96.4 | 94.6 | 98.0 |
| 7 | Excitement,Immobility/stupor,Mutism,Stereotypy,Mannerisms,Negativism,Waxy flexibility | 96.4 | 94.6 | 98.0 |
| 7 | Excitement,Immobility/stupor,Posturing/catalepsy,Echopraxia/echolalia,Mannerisms,Waxy flexibility,Withdrawal | 96.4 | 94.6 | 98.0 |
| 7 | Excitement,Immobility/stupor,Posturing/catalepsy,Echopraxia/echolalia,Stereotypy,Mannerisms,Negativism | 96.4 | 94.6 | 98.0 |
| 7 | Excitement,Immobility/stupor,Posturing/catalepsy,Echopraxia/echolalia,Stereotypy,Mannerisms,Withdrawal | 96.4 | 94.6 | 98.0 |
| 7 | Excitement,Immobility/stupor,Posturing/catalepsy,Grimacing,Stereotypy,Mannerisms,Negativism | 96.4 | 94.6 | 98.0 |
| 7 | Excitement,Immobility/stupor,Posturing/catalepsy,Mannerisms,Verbigeration,Waxy flexibility,Withdrawal | 96.4 | 94.6 | 98.0 |
| 7 | Excitement,Immobility/stupor,Posturing/catalepsy,Stereotypy,Mannerisms,Verbigeration,Withdrawal | 96.4 | 94.6 | 98.0 |
| 7 | Excitement,Immobility/stupor,Stereotypy,Mannerisms,Negativism,Waxy flexibility,Withdrawal | 96.4 | 94.6 | 98.0 |
| 7 | Excitement,Mutism,Posturing/catalepsy,Verbigeration,Rigidity,Negativism,Withdrawal | 96.4 | 94.6 | 98.0 |
| 7 | Excitement,Staring,Echopraxia/echolalia,Stereotypy,Mannerisms,Verbigeration,Negativism | 96.4 | 94.6 | 98.0 |
| 7 | Excitement,Staring,Grimacing,Echopraxia/echolalia,Stereotypy,Mannerisms,Negativism | 96.4 | 94.6 | 98.0 |
| 7 | Excitement,Staring,Grimacing,Echopraxia/echolalia,Verbigeration,Negativism,Waxy flexibility | 96.4 | 94.6 | 98.0 |
| 7 | Excitement,Staring,Grimacing,Mannerisms,Verbigeration,Negativism,Waxy flexibility | 96.4 | 94.6 | 98.0 |
| 7 | Excitement,Staring,Grimacing,Stereotypy,Verbigeration,Negativism,Waxy flexibility | 96.4 | 94.6 | 98.0 |
| 7 | Excitement,Staring,Posturing/catalepsy,Grimacing,Echopraxia/echolalia,Mannerisms,Waxy flexibility | 96.4 | 94.6 | 98.0 |
| 7 | Excitement,Staring,Posturing/catalepsy,Grimacing,Echopraxia/echolalia,Stereotypy,Mannerisms | 96.4 | 94.6 | 98.0 |
| 7 | Immobility/stupor,Grimacing,Echopraxia/echolalia,Stereotypy,Mannerisms,Negativism,Waxy flexibility | 96.4 | 94.6 | 98.0 |
| 7 | Immobility/stupor,Grimacing,Echopraxia/echolalia,Stereotypy,Rigidity,Negativism,Waxy flexibility | 96.4 | 94.6 | 98.0 |
| 7 | Immobility/stupor,Grimacing,Echopraxia/echolalia,Stereotypy,Verbigeration,Negativism,Withdrawal | 96.4 | 94.6 | 98.0 |
| 7 | Immobility/stupor,Grimacing,Verbigeration,Rigidity,Negativism,Waxy flexibility,Withdrawal | 96.4 | 94.6 | 98.0 |
| 7 | Immobility/stupor,Mutism,Echopraxia/echolalia,Mannerisms,Negativism,Waxy flexibility,Withdrawal | 96.4 | 94.6 | 98.0 |
| 7 | Immobility/stupor,Mutism,Echopraxia/echolalia,Mannerisms,Verbigeration,Negativism,Withdrawal | 96.4 | 94.6 | 98.0 |
| 7 | Immobility/stupor,Mutism,Echopraxia/echolalia,Rigidity,Negativism,Waxy flexibility,Withdrawal | 96.4 | 94.6 | 98.0 |
| 7 | Immobility/stupor,Mutism,Echopraxia/echolalia,Stereotypy,Mannerisms,Rigidity,Withdrawal | 96.4 | 94.6 | 98.0 |
| 7 | Immobility/stupor,Mutism,Grimacing,Echopraxia/echolalia,Mannerisms,Negativism,Withdrawal | 96.4 | 94.6 | 98.0 |
| 7 | Immobility/stupor,Mutism,Grimacing,Echopraxia/echolalia,Stereotypy,Rigidity,Negativism | 96.4 | 94.6 | 98.0 |
| 7 | Immobility/stupor,Mutism,Grimacing,Stereotypy,Mannerisms,Negativism,Withdrawal | 96.4 | 94.6 | 98.0 |
| 7 | Immobility/stupor,Mutism,Grimacing,Stereotypy,Rigidity,Negativism,Waxy flexibility | 96.4 | 94.6 | 98.0 |
| 7 | Immobility/stupor,Mutism,Posturing/catalepsy,Echopraxia/echolalia,Mannerisms,Rigidity,Withdrawal | 96.4 | 94.6 | 98.0 |
| 7 | Immobility/stupor,Mutism,Posturing/catalepsy,Echopraxia/echolalia,Mannerisms,Verbigeration,Negativism | 96.4 | 94.6 | 98.0 |
| 7 | Immobility/stupor,Mutism,Posturing/catalepsy,Echopraxia/echolalia,Stereotypy,Mannerisms,Negativism | 96.4 | 94.6 | 98.0 |
| 7 | Immobility/stupor,Mutism,Posturing/catalepsy,Echopraxia/echolalia,Stereotypy,Negativism,Withdrawal | 96.4 | 94.6 | 98.0 |
| 7 | Immobility/stupor,Mutism,Posturing/catalepsy,Grimacing,Echopraxia/echolalia,Negativism,Withdrawal | 96.4 | 94.6 | 98.0 |
| 7 | Immobility/stupor,Mutism,Posturing/catalepsy,Grimacing,Mannerisms,Verbigeration,Negativism | 96.4 | 94.6 | 98.0 |
| 7 | Immobility/stupor,Mutism,Posturing/catalepsy,Grimacing,Rigidity,Negativism,Withdrawal | 96.4 | 94.6 | 98.0 |
| 7 | Immobility/stupor,Mutism,Posturing/catalepsy,Grimacing,Stereotypy,Mannerisms,Negativism | 96.4 | 94.6 | 98.0 |
| 7 | Immobility/stupor,Mutism,Posturing/catalepsy,Grimacing,Stereotypy,Negativism,Withdrawal | 96.4 | 94.6 | 98.0 |
| 7 | Immobility/stupor,Mutism,Posturing/catalepsy,Grimacing,Stereotypy,Verbigeration,Rigidity | 96.4 | 94.6 | 98.0 |
| 7 | Immobility/stupor,Mutism,Posturing/catalepsy,Mannerisms,Verbigeration,Rigidity,Withdrawal | 96.4 | 94.6 | 98.0 |
| 7 | Immobility/stupor,Mutism,Posturing/catalepsy,Stereotypy,Mannerisms,Rigidity,Withdrawal | 96.4 | 94.6 | 98.0 |
| 7 | Immobility/stupor,Mutism,Posturing/catalepsy,Stereotypy,Mannerisms,Verbigeration,Negativism | 96.4 | 94.6 | 98.0 |
| 7 | Immobility/stupor,Mutism,Posturing/catalepsy,Stereotypy,Rigidity,Negativism,Waxy flexibility | 96.4 | 94.6 | 98.0 |
| 7 | Immobility/stupor,Mutism,Posturing/catalepsy,Verbigeration,Rigidity,Negativism,Waxy flexibility | 96.4 | 94.6 | 98.0 |
| 7 | Immobility/stupor,Mutism,Staring,Echopraxia/echolalia,Mannerisms,Verbigeration,Waxy flexibility | 96.4 | 94.6 | 98.0 |
| 7 | Immobility/stupor,Mutism,Staring,Echopraxia/echolalia,Mannerisms,Verbigeration,Withdrawal | 96.4 | 94.6 | 98.0 |
| 7 | Immobility/stupor,Mutism,Staring,Echopraxia/echolalia,Stereotypy,Rigidity,Waxy flexibility | 96.4 | 94.6 | 98.0 |
| 7 | Immobility/stupor,Mutism,Staring,Grimacing,Echopraxia/echolalia,Stereotypy,Verbigeration | 96.4 | 94.6 | 98.0 |
| 7 | Immobility/stupor,Mutism,Staring,Grimacing,Echopraxia/echolalia,Verbigeration,Waxy flexibility | 96.4 | 94.6 | 98.0 |
| 7 | Immobility/stupor,Mutism,Staring,Mannerisms,Verbigeration,Waxy flexibility,Withdrawal | 96.4 | 94.6 | 98.0 |
| 7 | Immobility/stupor,Mutism,Staring,Posturing/catalepsy,Echopraxia/echolalia,Mannerisms,Waxy flexibility | 96.4 | 94.6 | 98.0 |
| 7 | Immobility/stupor,Mutism,Staring,Posturing/catalepsy,Echopraxia/echolalia,Stereotypy,Verbigeration | 96.4 | 94.6 | 98.0 |
| 7 | Immobility/stupor,Mutism,Staring,Posturing/catalepsy,Echopraxia/echolalia,Verbigeration,Withdrawal | 96.4 | 94.6 | 98.0 |
| 7 | Immobility/stupor,Mutism,Staring,Posturing/catalepsy,Grimacing,Echopraxia/echolalia,Rigidity | 96.4 | 94.6 | 98.0 |
| 7 | Immobility/stupor,Mutism,Staring,Posturing/catalepsy,Grimacing,Echopraxia/echolalia,Stereotypy | 96.4 | 94.6 | 98.0 |
| 7 | Immobility/stupor,Mutism,Staring,Posturing/catalepsy,Grimacing,Rigidity,Negativism | 96.4 | 94.6 | 98.0 |
| 7 | Immobility/stupor,Mutism,Staring,Posturing/catalepsy,Mannerisms,Verbigeration,Waxy flexibility | 96.4 | 94.6 | 98.0 |
| 7 | Immobility/stupor,Mutism,Staring,Posturing/catalepsy,Stereotypy,Verbigeration,Waxy flexibility | 96.4 | 94.6 | 98.0 |
| 7 | Immobility/stupor,Mutism,Staring,Posturing/catalepsy,Verbigeration,Rigidity,Waxy flexibility | 96.4 | 94.6 | 98.0 |
| 7 | Immobility/stupor,Mutism,Staring,Stereotypy,Mannerisms,Verbigeration,Waxy flexibility | 96.4 | 94.6 | 98.0 |
| 7 | Immobility/stupor,Mutism,Stereotypy,Mannerisms,Negativism,Waxy flexibility,Withdrawal | 96.4 | 94.6 | 98.0 |
| 7 | Immobility/stupor,Mutism,Stereotypy,Mannerisms,Verbigeration,Rigidity,Withdrawal | 96.4 | 94.6 | 98.0 |
| 7 | Immobility/stupor,Posturing/catalepsy,Echopraxia/echolalia,Mannerisms,Negativism,Waxy flexibility,Withdrawal | 96.4 | 94.6 | 98.0 |
| 7 | Immobility/stupor,Posturing/catalepsy,Echopraxia/echolalia,Rigidity,Negativism,Waxy flexibility,Withdrawal | 96.4 | 94.6 | 98.0 |
| 7 | Immobility/stupor,Posturing/catalepsy,Echopraxia/echolalia,Stereotypy,Verbigeration,Negativism,Withdrawal | 96.4 | 94.6 | 98.0 |
| 7 | Immobility/stupor,Posturing/catalepsy,Echopraxia/echolalia,Verbigeration,Rigidity,Negativism,Waxy flexibility | 96.4 | 94.6 | 98.0 |
| 7 | Immobility/stupor,Posturing/catalepsy,Grimacing,Echopraxia/echolalia,Stereotypy,Mannerisms,Negativism | 96.4 | 94.6 | 98.0 |
| 7 | Immobility/stupor,Posturing/catalepsy,Grimacing,Echopraxia/echolalia,Stereotypy,Rigidity,Negativism | 96.4 | 94.6 | 98.0 |
| 7 | Immobility/stupor,Posturing/catalepsy,Grimacing,Rigidity,Negativism,Waxy flexibility,Withdrawal | 96.4 | 94.6 | 98.0 |
| 7 | Immobility/stupor,Posturing/catalepsy,Grimacing,Stereotypy,Mannerisms,Rigidity,Withdrawal | 96.4 | 94.6 | 98.0 |
| 7 | Immobility/stupor,Posturing/catalepsy,Grimacing,Stereotypy,Rigidity,Negativism,Waxy flexibility | 96.4 | 94.6 | 98.0 |
| 7 | Immobility/stupor,Posturing/catalepsy,Mannerisms,Verbigeration,Negativism,Waxy flexibility,Withdrawal | 96.4 | 94.6 | 98.0 |
| 7 | Immobility/stupor,Staring,Echopraxia/echolalia,Stereotypy,Verbigeration,Rigidity,Waxy flexibility | 96.4 | 94.6 | 98.0 |
| 7 | Immobility/stupor,Staring,Grimacing,Echopraxia/echolalia,Mannerisms,Verbigeration,Waxy flexibility | 96.4 | 94.6 | 98.0 |
| 7 | Immobility/stupor,Staring,Grimacing,Echopraxia/echolalia,Mannerisms,Verbigeration,Withdrawal | 96.4 | 94.6 | 98.0 |
| 7 | Immobility/stupor,Staring,Grimacing,Echopraxia/echolalia,Rigidity,Waxy flexibility,Withdrawal | 96.4 | 94.6 | 98.0 |
| 7 | Immobility/stupor,Staring,Grimacing,Echopraxia/echolalia,Stereotypy,Mannerisms,Verbigeration | 96.4 | 94.6 | 98.0 |
| 7 | Immobility/stupor,Staring,Grimacing,Echopraxia/echolalia,Stereotypy,Rigidity,Waxy flexibility | 96.4 | 94.6 | 98.0 |
| 7 | Immobility/stupor,Staring,Grimacing,Echopraxia/echolalia,Verbigeration,Rigidity,Waxy flexibility | 96.4 | 94.6 | 98.0 |
| 7 | Immobility/stupor,Staring,Grimacing,Mannerisms,Verbigeration,Waxy flexibility,Withdrawal | 96.4 | 94.6 | 98.0 |
| 7 | Immobility/stupor,Staring,Posturing/catalepsy,Echopraxia/echolalia,Mannerisms,Rigidity,Waxy flexibility | 96.4 | 94.6 | 98.0 |
| 7 | Immobility/stupor,Staring,Posturing/catalepsy,Echopraxia/echolalia,Mannerisms,Verbigeration,Withdrawal | 96.4 | 94.6 | 98.0 |
| 7 | Immobility/stupor,Staring,Posturing/catalepsy,Echopraxia/echolalia,Mannerisms,Waxy flexibility,Withdrawal | 96.4 | 94.6 | 98.0 |
| 7 | Immobility/stupor,Staring,Posturing/catalepsy,Grimacing,Echopraxia/echolalia,Negativism,Waxy flexibility | 96.4 | 94.6 | 98.0 |
| 7 | Immobility/stupor,Stereotypy,Mannerisms,Verbigeration,Negativism,Waxy flexibility,Withdrawal | 96.4 | 94.6 | 98.0 |
| 7 | Mutism,Staring,Posturing/catalepsy,Grimacing,Echopraxia/echolalia,Rigidity,Waxy flexibility | 96.4 | 94.6 | 98.0 |
| 7 | Mutism,Staring,Posturing/catalepsy,Grimacing,Rigidity,Negativism,Waxy flexibility | 96.4 | 94.6 | 98.0 |
| 7 | Staring,Echopraxia/echolalia,Stereotypy,Mannerisms,Verbigeration,Negativism,Waxy flexibility | 96.4 | 94.6 | 98.0 |
| 7 | Staring,Echopraxia/echolalia,Stereotypy,Verbigeration,Rigidity,Waxy flexibility,Withdrawal | 96.4 | 94.6 | 98.0 |
| 7 | Staring,Grimacing,Echopraxia/echolalia,Mannerisms,Verbigeration,Waxy flexibility,Withdrawal | 96.4 | 94.6 | 98.0 |
| 7 | Staring,Grimacing,Echopraxia/echolalia,Stereotypy,Mannerisms,Verbigeration,Rigidity | 96.4 | 94.6 | 98.0 |
| 7 | Staring,Grimacing,Echopraxia/echolalia,Verbigeration,Negativism,Waxy flexibility,Withdrawal | 96.4 | 94.6 | 98.0 |
| 7 | Staring,Grimacing,Echopraxia/echolalia,Verbigeration,Rigidity,Waxy flexibility,Withdrawal | 96.4 | 94.6 | 98.0 |
| 7 | Staring,Posturing/catalepsy,Echopraxia/echolalia,Stereotypy,Mannerisms,Waxy flexibility,Withdrawal | 96.4 | 94.6 | 98.0 |
| 7 | Staring,Posturing/catalepsy,Echopraxia/echolalia,Stereotypy,Rigidity,Waxy flexibility,Withdrawal | 96.4 | 94.6 | 98.0 |
| 7 | Staring,Posturing/catalepsy,Grimacing,Echopraxia/echolalia,Mannerisms,Negativism,Waxy flexibility | 96.4 | 94.6 | 98.0 |
| 7 | Staring,Posturing/catalepsy,Grimacing,Echopraxia/echolalia,Mannerisms,Verbigeration,Rigidity | 96.4 | 94.6 | 98.0 |
| 7 | Staring,Posturing/catalepsy,Grimacing,Echopraxia/echolalia,Stereotypy,Rigidity,Negativism | 96.4 | 94.6 | 98.0 |
| 7 | Staring,Posturing/catalepsy,Grimacing,Echopraxia/echolalia,Verbigeration,Rigidity,Negativism | 96.4 | 94.6 | 98.0 |
| 7 | Staring,Posturing/catalepsy,Grimacing,Stereotypy,Mannerisms,Negativism,Waxy flexibility | 96.4 | 94.6 | 98.0 |
| 7 | Excitement,Immobility/stupor,Echopraxia/echolalia,Stereotypy,Mannerisms,Negativism,Waxy flexibility | 96.2 | 94.4 | 97.8 |
| 7 | Excitement,Immobility/stupor,Echopraxia/echolalia,Stereotypy,Verbigeration,Waxy flexibility,Withdrawal | 96.2 | 94.4 | 98.0 |
| 7 | Excitement,Immobility/stupor,Grimacing,Echopraxia/echolalia,Mannerisms,Waxy flexibility,Withdrawal | 96.2 | 94.4 | 97.8 |
| 7 | Excitement,Immobility/stupor,Grimacing,Echopraxia/echolalia,Stereotypy,Verbigeration,Negativism | 96.2 | 94.4 | 97.8 |
| 7 | Excitement,Immobility/stupor,Grimacing,Echopraxia/echolalia,Stereotypy,Waxy flexibility,Withdrawal | 96.2 | 94.4 | 97.8 |
| 7 | Excitement,Immobility/stupor,Grimacing,Mannerisms,Negativism,Waxy flexibility,Withdrawal | 96.2 | 94.4 | 97.8 |
| 7 | Excitement,Immobility/stupor,Grimacing,Mannerisms,Verbigeration,Negativism,Waxy flexibility | 96.2 | 94.4 | 97.8 |
| 7 | Excitement,Immobility/stupor,Grimacing,Stereotypy,Mannerisms,Negativism,Withdrawal | 96.2 | 94.4 | 97.8 |
| 7 | Excitement,Immobility/stupor,Grimacing,Verbigeration,Negativism,Waxy flexibility,Withdrawal | 96.2 | 94.4 | 97.8 |
| 7 | Excitement,Immobility/stupor,Mutism,Echopraxia/echolalia,Mannerisms,Verbigeration,Negativism | 96.2 | 94.4 | 98.0 |
| 7 | Excitement,Immobility/stupor,Mutism,Echopraxia/echolalia,Mannerisms,Waxy flexibility,Withdrawal | 96.2 | 94.4 | 97.8 |
| 7 | Excitement,Immobility/stupor,Mutism,Echopraxia/echolalia,Stereotypy,Mannerisms,Negativism | 96.2 | 94.4 | 97.8 |
| 7 | Excitement,Immobility/stupor,Mutism,Echopraxia/echolalia,Stereotypy,Mannerisms,Withdrawal | 96.2 | 94.4 | 97.8 |
| 7 | Excitement,Immobility/stupor,Mutism,Echopraxia/echolalia,Stereotypy,Verbigeration,Waxy flexibility | 96.2 | 94.4 | 97.8 |
| 7 | Excitement,Immobility/stupor,Mutism,Grimacing,Echopraxia/echolalia,Stereotypy,Waxy flexibility | 96.2 | 94.4 | 97.8 |
| 7 | Excitement,Immobility/stupor,Mutism,Grimacing,Echopraxia/echolalia,Verbigeration,Waxy flexibility | 96.2 | 94.2 | 97.8 |
| 7 | Excitement,Immobility/stupor,Mutism,Grimacing,Mannerisms,Negativism,Withdrawal | 96.2 | 94.4 | 98.0 |
| 7 | Excitement,Immobility/stupor,Mutism,Grimacing,Mannerisms,Verbigeration,Negativism | 96.2 | 94.4 | 97.8 |
| 7 | Excitement,Immobility/stupor,Mutism,Grimacing,Verbigeration,Waxy flexibility,Withdrawal | 96.2 | 94.4 | 97.8 |
| 7 | Excitement,Immobility/stupor,Mutism,Posturing/catalepsy,Echopraxia/echolalia,Mannerisms,Waxy flexibility | 96.2 | 94.4 | 97.8 |
| 7 | Excitement,Immobility/stupor,Mutism,Posturing/catalepsy,Mannerisms,Verbigeration,Waxy flexibility | 96.2 | 94.4 | 98.0 |
| 7 | Excitement,Immobility/stupor,Mutism,Stereotypy,Mannerisms,Verbigeration,Withdrawal | 96.2 | 94.4 | 97.8 |
| 7 | Excitement,Immobility/stupor,Posturing/catalepsy,Echopraxia/echolalia,Stereotypy,Waxy flexibility,Withdrawal | 96.2 | 94.4 | 98.0 |
| 7 | Excitement,Immobility/stupor,Posturing/catalepsy,Grimacing,Mannerisms,Waxy flexibility,Withdrawal | 96.2 | 94.4 | 97.8 |
| 7 | Excitement,Immobility/stupor,Posturing/catalepsy,Grimacing,Stereotypy,Verbigeration,Withdrawal | 96.2 | 94.4 | 98.0 |
| 7 | Excitement,Immobility/stupor,Posturing/catalepsy,Grimacing,Verbigeration,Waxy flexibility,Withdrawal | 96.2 | 94.4 | 98.0 |
| 7 | Excitement,Immobility/stupor,Posturing/catalepsy,Stereotypy,Mannerisms,Negativism,Waxy flexibility | 96.2 | 94.4 | 97.8 |
| 7 | Excitement,Immobility/stupor,Posturing/catalepsy,Stereotypy,Mannerisms,Rigidity,Waxy flexibility | 96.2 | 94.4 | 97.8 |
| 7 | Excitement,Immobility/stupor,Posturing/catalepsy,Stereotypy,Verbigeration,Waxy flexibility,Withdrawal | 96.2 | 94.4 | 97.8 |
| 7 | Excitement,Immobility/stupor,Stereotypy,Mannerisms,Verbigeration,Negativism,Waxy flexibility | 96.2 | 94.4 | 97.8 |
| 7 | Excitement,Mutism,Posturing/catalepsy,Echopraxia/echolalia,Rigidity,Negativism,Withdrawal | 96.2 | 94.4 | 97.8 |
| 7 | Excitement,Staring,Grimacing,Echopraxia/echolalia,Stereotypy,Mannerisms,Waxy flexibility | 96.2 | 94.4 | 97.8 |
| 7 | Excitement,Staring,Grimacing,Echopraxia/echolalia,Stereotypy,Verbigeration,Negativism | 96.2 | 94.4 | 97.8 |
| 7 | Excitement,Staring,Grimacing,Stereotypy,Mannerisms,Negativism,Waxy flexibility | 96.2 | 94.4 | 97.8 |
| 7 | Excitement,Staring,Posturing/catalepsy,Grimacing,Echopraxia/echolalia,Mannerisms,Verbigeration | 96.2 | 94.4 | 97.8 |
| 7 | Excitement,Staring,Posturing/catalepsy,Grimacing,Echopraxia/echolalia,Stereotypy,Waxy flexibility | 96.2 | 94.4 | 97.8 |
| 7 | Excitement,Staring,Posturing/catalepsy,Grimacing,Mannerisms,Verbigeration,Waxy flexibility | 96.2 | 94.4 | 98.0 |
| 7 | Excitement,Staring,Posturing/catalepsy,Grimacing,Stereotypy,Mannerisms,Verbigeration | 96.2 | 94.4 | 98.0 |
| 7 | Immobility/stupor,Grimacing,Echopraxia/echolalia,Mannerisms,Rigidity,Waxy flexibility,Withdrawal | 96.2 | 94.4 | 97.8 |
| 7 | Immobility/stupor,Grimacing,Echopraxia/echolalia,Mannerisms,Verbigeration,Rigidity,Withdrawal | 96.2 | 94.4 | 98.0 |
| 7 | Immobility/stupor,Grimacing,Echopraxia/echolalia,Stereotypy,Verbigeration,Rigidity,Withdrawal | 96.2 | 94.4 | 97.8 |
| 7 | Immobility/stupor,Grimacing,Stereotypy,Mannerisms,Negativism,Waxy flexibility,Withdrawal | 96.2 | 94.4 | 97.8 |
| 7 | Immobility/stupor,Grimacing,Stereotypy,Mannerisms,Rigidity,Waxy flexibility,Withdrawal | 96.2 | 94.4 | 98.0 |
| 7 | Immobility/stupor,Grimacing,Stereotypy,Mannerisms,Verbigeration,Rigidity,Withdrawal | 96.2 | 94.2 | 97.8 |
| 7 | Immobility/stupor,Mutism,Echopraxia/echolalia,Mannerisms,Verbigeration,Rigidity,Withdrawal | 96.2 | 94.4 | 98.0 |
| 7 | Immobility/stupor,Mutism,Echopraxia/echolalia,Stereotypy,Mannerisms,Negativism,Waxy flexibility | 96.2 | 94.4 | 97.8 |
| 7 | Immobility/stupor,Mutism,Echopraxia/echolalia,Stereotypy,Rigidity,Negativism,Waxy flexibility | 96.2 | 94.4 | 98.0 |
| 7 | Immobility/stupor,Mutism,Echopraxia/echolalia,Stereotypy,Verbigeration,Negativism,Withdrawal | 96.2 | 94.4 | 97.8 |
| 7 | Immobility/stupor,Mutism,Grimacing,Echopraxia/echolalia,Mannerisms,Negativism,Waxy flexibility | 96.2 | 94.4 | 97.8 |
| 7 | Immobility/stupor,Mutism,Grimacing,Echopraxia/echolalia,Stereotypy,Negativism,Withdrawal | 96.2 | 94.4 | 98.0 |
| 7 | Immobility/stupor,Mutism,Grimacing,Echopraxia/echolalia,Verbigeration,Rigidity,Withdrawal | 96.2 | 94.4 | 97.8 |
| 7 | Immobility/stupor,Mutism,Grimacing,Mannerisms,Negativism,Waxy flexibility,Withdrawal | 96.2 | 94.4 | 97.8 |
| 7 | Immobility/stupor,Mutism,Grimacing,Mannerisms,Verbigeration,Negativism,Withdrawal | 96.2 | 94.4 | 97.8 |
| 7 | Immobility/stupor,Mutism,Grimacing,Stereotypy,Mannerisms,Verbigeration,Rigidity | 96.2 | 94.4 | 97.8 |
| 7 | Immobility/stupor,Mutism,Grimacing,Stereotypy,Verbigeration,Negativism,Withdrawal | 96.2 | 94.4 | 98.0 |
| 7 | Immobility/stupor,Mutism,Posturing/catalepsy,Echopraxia/echolalia,Verbigeration,Negativism,Withdrawal | 96.2 | 94.4 | 97.8 |
| 7 | Immobility/stupor,Mutism,Posturing/catalepsy,Grimacing,Echopraxia/echolalia,Mannerisms,Rigidity | 96.2 | 94.4 | 97.8 |
| 7 | Immobility/stupor,Mutism,Posturing/catalepsy,Grimacing,Echopraxia/echolalia,Mannerisms,Withdrawal | 96.2 | 94.4 | 98.0 |
| 7 | Immobility/stupor,Mutism,Posturing/catalepsy,Grimacing,Echopraxia/echolalia,Rigidity,Negativism | 96.2 | 94.4 | 97.8 |
| 7 | Immobility/stupor,Mutism,Posturing/catalepsy,Grimacing,Echopraxia/echolalia,Verbigeration,Negativism | 96.2 | 94.4 | 97.8 |
| 7 | Immobility/stupor,Mutism,Posturing/catalepsy,Grimacing,Mannerisms,Verbigeration,Rigidity | 96.2 | 94.4 | 98.0 |
| 7 | Immobility/stupor,Mutism,Posturing/catalepsy,Grimacing,Stereotypy,Mannerisms,Rigidity | 96.2 | 94.4 | 98.0 |
| 7 | Immobility/stupor,Mutism,Posturing/catalepsy,Grimacing,Stereotypy,Rigidity,Withdrawal | 96.2 | 94.4 | 97.8 |
| 7 | Immobility/stupor,Mutism,Posturing/catalepsy,Grimacing,Stereotypy,Verbigeration,Negativism | 96.2 | 94.4 | 97.8 |
| 7 | Immobility/stupor,Mutism,Posturing/catalepsy,Grimacing,Verbigeration,Negativism,Withdrawal | 96.2 | 94.4 | 98.0 |
| 7 | Immobility/stupor,Mutism,Staring,Echopraxia/echolalia,Stereotypy,Mannerisms,Verbigeration | 96.2 | 94.4 | 97.8 |
| 7 | Immobility/stupor,Mutism,Staring,Echopraxia/echolalia,Stereotypy,Waxy flexibility,Withdrawal | 96.2 | 94.4 | 97.8 |
| 7 | Immobility/stupor,Mutism,Staring,Grimacing,Echopraxia/echolalia,Negativism,Waxy flexibility | 96.2 | 94.4 | 98.0 |
| 7 | Immobility/stupor,Mutism,Staring,Grimacing,Echopraxia/echolalia,Stereotypy,Waxy flexibility | 96.2 | 94.4 | 97.8 |
| 7 | Immobility/stupor,Mutism,Staring,Grimacing,Echopraxia/echolalia,Verbigeration,Withdrawal | 96.2 | 94.4 | 98.0 |
| 7 | Immobility/stupor,Mutism,Staring,Grimacing,Negativism,Waxy flexibility,Withdrawal | 96.2 | 94.4 | 97.8 |
| 7 | Immobility/stupor,Mutism,Staring,Grimacing,Rigidity,Negativism,Waxy flexibility | 96.2 | 94.4 | 97.8 |
| 7 | Immobility/stupor,Mutism,Staring,Grimacing,Rigidity,Waxy flexibility,Withdrawal | 96.2 | 94.4 | 97.8 |
| 7 | Immobility/stupor,Mutism,Staring,Grimacing,Stereotypy,Waxy flexibility,Withdrawal | 96.2 | 94.4 | 97.8 |
| 7 | Immobility/stupor,Mutism,Staring,Posturing/catalepsy,Echopraxia/echolalia,Waxy flexibility,Withdrawal | 96.2 | 94.4 | 98.0 |
| 7 | Immobility/stupor,Mutism,Staring,Posturing/catalepsy,Grimacing,Mannerisms,Waxy flexibility | 96.2 | 94.2 | 98.0 |
| 7 | Immobility/stupor,Mutism,Staring,Posturing/catalepsy,Rigidity,Negativism,Waxy flexibility | 96.2 | 94.4 | 97.8 |
| 7 | Immobility/stupor,Mutism,Staring,Posturing/catalepsy,Rigidity,Waxy flexibility,Withdrawal | 96.2 | 94.4 | 97.8 |
| 7 | Immobility/stupor,Mutism,Staring,Posturing/catalepsy,Stereotypy,Mannerisms,Waxy flexibility | 96.2 | 94.4 | 98.0 |
| 7 | Immobility/stupor,Mutism,Staring,Posturing/catalepsy,Stereotypy,Rigidity,Waxy flexibility | 96.2 | 94.4 | 98.0 |
| 7 | Immobility/stupor,Mutism,Staring,Posturing/catalepsy,Stereotypy,Waxy flexibility,Withdrawal | 96.2 | 94.4 | 97.8 |
| 7 | Immobility/stupor,Mutism,Staring,Stereotypy,Verbigeration,Waxy flexibility,Withdrawal | 96.2 | 94.4 | 97.8 |
| 7 | Immobility/stupor,Mutism,Stereotypy,Mannerisms,Rigidity,Waxy flexibility,Withdrawal | 96.2 | 94.4 | 97.8 |
| 7 | Immobility/stupor,Posturing/catalepsy,Echopraxia/echolalia,Stereotypy,Rigidity,Negativism,Waxy flexibility | 96.2 | 94.4 | 97.8 |
| 7 | Immobility/stupor,Posturing/catalepsy,Grimacing,Echopraxia/echolalia,Mannerisms,Verbigeration,Negativism | 96.2 | 94.4 | 97.8 |
| 7 | Immobility/stupor,Posturing/catalepsy,Grimacing,Echopraxia/echolalia,Rigidity,Negativism,Waxy flexibility | 96.2 | 94.4 | 98.0 |
| 7 | Immobility/stupor,Posturing/catalepsy,Grimacing,Echopraxia/echolalia,Stereotypy,Rigidity,Withdrawal | 96.2 | 94.4 | 97.8 |
| 7 | Immobility/stupor,Posturing/catalepsy,Grimacing,Mannerisms,Negativism,Waxy flexibility,Withdrawal | 96.2 | 94.4 | 97.8 |
| 7 | Immobility/stupor,Posturing/catalepsy,Grimacing,Mannerisms,Verbigeration,Rigidity,Withdrawal | 96.2 | 94.2 | 97.8 |
| 7 | Immobility/stupor,Posturing/catalepsy,Grimacing,Stereotypy,Mannerisms,Verbigeration,Negativism | 96.2 | 94.4 | 97.8 |
| 7 | Immobility/stupor,Posturing/catalepsy,Stereotypy,Verbigeration,Negativism,Waxy flexibility,Withdrawal | 96.2 | 94.4 | 97.8 |
| 7 | Immobility/stupor,Staring,Echopraxia/echolalia,Stereotypy,Mannerisms,Verbigeration,Withdrawal | 96.2 | 94.4 | 97.8 |
| 7 | Immobility/stupor,Staring,Echopraxia/echolalia,Verbigeration,Rigidity,Waxy flexibility,Withdrawal | 96.2 | 94.4 | 97.8 |
| 7 | Immobility/stupor,Staring,Grimacing,Echopraxia/echolalia,Stereotypy,Verbigeration,Withdrawal | 96.2 | 94.4 | 97.8 |
| 7 | Immobility/stupor,Staring,Grimacing,Echopraxia/echolalia,Stereotypy,Waxy flexibility,Withdrawal | 96.2 | 94.4 | 97.8 |
| 7 | Immobility/stupor,Staring,Grimacing,Stereotypy,Verbigeration,Waxy flexibility,Withdrawal | 96.2 | 94.4 | 97.8 |
| 7 | Immobility/stupor,Staring,Posturing/catalepsy,Echopraxia/echolalia,Rigidity,Waxy flexibility,Withdrawal | 96.2 | 94.2 | 97.8 |
| 7 | Immobility/stupor,Staring,Posturing/catalepsy,Echopraxia/echolalia,Stereotypy,Mannerisms,Verbigeration | 96.2 | 94.4 | 97.8 |
| 7 | Immobility/stupor,Staring,Posturing/catalepsy,Grimacing,Echopraxia/echolalia,Mannerisms,Waxy flexibility | 96.2 | 94.4 | 97.8 |
| 7 | Immobility/stupor,Staring,Posturing/catalepsy,Grimacing,Mannerisms,Verbigeration,Waxy flexibility | 96.2 | 94.4 | 97.8 |
| 7 | Immobility/stupor,Staring,Posturing/catalepsy,Grimacing,Rigidity,Negativism,Waxy flexibility | 96.2 | 94.4 | 97.8 |
| 7 | Immobility/stupor,Staring,Posturing/catalepsy,Grimacing,Stereotypy,Mannerisms,Waxy flexibility | 96.2 | 94.4 | 97.8 |
| 7 | Immobility/stupor,Staring,Posturing/catalepsy,Mannerisms,Verbigeration,Waxy flexibility,Withdrawal | 96.2 | 94.4 | 97.8 |
| 7 | Immobility/stupor,Staring,Posturing/catalepsy,Stereotypy,Mannerisms,Verbigeration,Waxy flexibility | 96.2 | 94.4 | 97.8 |
| 7 | Immobility/stupor,Staring,Posturing/catalepsy,Verbigeration,Rigidity,Waxy flexibility,Withdrawal | 96.2 | 94.4 | 97.8 |
| 7 | Immobility/stupor,Staring,Stereotypy,Mannerisms,Verbigeration,Waxy flexibility,Withdrawal | 96.2 | 94.4 | 97.8 |
| 7 | Staring,Grimacing,Echopraxia/echolalia,Mannerisms,Verbigeration,Rigidity,Waxy flexibility | 96.2 | 94.4 | 97.8 |
| 7 | Staring,Grimacing,Echopraxia/echolalia,Stereotypy,Mannerisms,Verbigeration,Negativism | 96.2 | 94.4 | 97.8 |
| 7 | Staring,Grimacing,Echopraxia/echolalia,Stereotypy,Rigidity,Negativism,Waxy flexibility | 96.2 | 94.4 | 97.8 |
| 7 | Staring,Grimacing,Echopraxia/echolalia,Verbigeration,Rigidity,Negativism,Waxy flexibility | 96.2 | 94.4 | 97.8 |
| 7 | Staring,Grimacing,Stereotypy,Mannerisms,Verbigeration,Negativism,Waxy flexibility | 96.2 | 94.4 | 97.8 |
| 7 | Staring,Posturing/catalepsy,Echopraxia/echolalia,Mannerisms,Verbigeration,Negativism,Waxy flexibility | 96.2 | 94.4 | 98.0 |
| 7 | Staring,Posturing/catalepsy,Echopraxia/echolalia,Stereotypy,Rigidity,Negativism,Waxy flexibility | 96.2 | 94.4 | 97.8 |
| 7 | Staring,Posturing/catalepsy,Grimacing,Echopraxia/echolalia,Rigidity,Waxy flexibility,Withdrawal | 96.2 | 94.4 | 97.8 |
| 7 | Staring,Posturing/catalepsy,Grimacing,Stereotypy,Mannerisms,Rigidity,Waxy flexibility | 96.2 | 94.4 | 97.8 |
| 7 | Staring,Posturing/catalepsy,Grimacing,Verbigeration,Rigidity,Waxy flexibility,Withdrawal | 96.2 | 94.4 | 97.8 |
| 7 | Staring,Posturing/catalepsy,Stereotypy,Mannerisms,Verbigeration,Waxy flexibility,Withdrawal | 96.2 | 94.4 | 98.0 |
| 7 | Excitement,Immobility/stupor,Echopraxia/echolalia,Mannerisms,Verbigeration,Negativism,Waxy flexibility | 96.0 | 94.2 | 97.8 |
| 7 | Excitement,Immobility/stupor,Echopraxia/echolalia,Mannerisms,Verbigeration,Waxy flexibility,Withdrawal | 96.0 | 94.2 | 97.8 |
| 7 | Excitement,Immobility/stupor,Echopraxia/echolalia,Stereotypy,Mannerisms,Verbigeration,Negativism | 96.0 | 94.2 | 97.8 |
| 7 | Excitement,Immobility/stupor,Echopraxia/echolalia,Stereotypy,Mannerisms,Verbigeration,Withdrawal | 96.0 | 94.2 | 97.8 |
| 7 | Excitement,Immobility/stupor,Echopraxia/echolalia,Stereotypy,Mannerisms,Waxy flexibility,Withdrawal | 96.0 | 94.2 | 97.8 |
| 7 | Excitement,Immobility/stupor,Grimacing,Echopraxia/echolalia,Verbigeration,Waxy flexibility,Withdrawal | 96.0 | 93.9 | 97.8 |
| 7 | Excitement,Immobility/stupor,Mutism,Echopraxia/echolalia,Mannerisms,Verbigeration,Withdrawal | 96.0 | 94.2 | 97.8 |
| 7 | Excitement,Immobility/stupor,Mutism,Grimacing,Mannerisms,Waxy flexibility,Withdrawal | 96.0 | 93.9 | 97.8 |
| 7 | Excitement,Immobility/stupor,Mutism,Grimacing,Stereotypy,Verbigeration,Waxy flexibility | 96.0 | 93.9 | 97.8 |
| 7 | Excitement,Immobility/stupor,Mutism,Mannerisms,Verbigeration,Waxy flexibility,Withdrawal | 96.0 | 93.9 | 97.8 |
| 7 | Excitement,Immobility/stupor,Mutism,Posturing/catalepsy,Echopraxia/echolalia,Stereotypy,Mannerisms | 96.0 | 93.9 | 97.8 |
| 7 | Excitement,Immobility/stupor,Mutism,Posturing/catalepsy,Grimacing,Stereotypy,Mannerisms | 96.0 | 94.2 | 97.8 |
| 7 | Excitement,Immobility/stupor,Mutism,Posturing/catalepsy,Stereotypy,Mannerisms,Verbigeration | 96.0 | 94.2 | 97.8 |
| 7 | Excitement,Immobility/stupor,Posturing/catalepsy,Grimacing,Echopraxia/echolalia,Mannerisms,Waxy flexibility | 96.0 | 94.2 | 97.8 |
| 7 | Excitement,Immobility/stupor,Posturing/catalepsy,Grimacing,Echopraxia/echolalia,Verbigeration,Waxy flexibility | 96.0 | 93.9 | 97.8 |
| 7 | Excitement,Mutism,Posturing/catalepsy,Grimacing,Rigidity,Negativism,Withdrawal | 96.0 | 93.9 | 97.8 |
| 7 | Excitement,Mutism,Posturing/catalepsy,Mannerisms,Rigidity,Negativism,Withdrawal | 96.0 | 93.9 | 97.8 |
| 7 | Excitement,Mutism,Posturing/catalepsy,Stereotypy,Rigidity,Negativism,Withdrawal | 96.0 | 93.9 | 97.8 |
| 7 | Excitement,Staring,Grimacing,Stereotypy,Mannerisms,Verbigeration,Negativism | 96.0 | 93.9 | 97.8 |
| 7 | Excitement,Staring,Posturing/catalepsy,Grimacing,Echopraxia/echolalia,Stereotypy,Verbigeration | 96.0 | 93.9 | 97.8 |
| 7 | Excitement,Staring,Posturing/catalepsy,Grimacing,Echopraxia/echolalia,Verbigeration,Waxy flexibility | 96.0 | 93.9 | 97.8 |
| 7 | Excitement,Staring,Posturing/catalepsy,Grimacing,Stereotypy,Mannerisms,Waxy flexibility | 96.0 | 93.9 | 97.8 |
| 7 | Excitement,Staring,Posturing/catalepsy,Grimacing,Stereotypy,Verbigeration,Waxy flexibility | 96.0 | 93.9 | 97.8 |
| 7 | Immobility/stupor,Echopraxia/echolalia,Stereotypy,Mannerisms,Verbigeration,Negativism,Waxy flexibility | 96.0 | 94.2 | 97.8 |
| 7 | Immobility/stupor,Echopraxia/echolalia,Stereotypy,Verbigeration,Negativism,Waxy flexibility,Withdrawal | 96.0 | 94.2 | 97.8 |
| 7 | Immobility/stupor,Grimacing,Echopraxia/echolalia,Mannerisms,Negativism,Waxy flexibility,Withdrawal | 96.0 | 93.9 | 97.8 |
| 7 | Immobility/stupor,Grimacing,Echopraxia/echolalia,Mannerisms,Verbigeration,Negativism,Withdrawal | 96.0 | 93.9 | 97.8 |
| 7 | Immobility/stupor,Grimacing,Echopraxia/echolalia,Stereotypy,Negativism,Waxy flexibility,Withdrawal | 96.0 | 94.2 | 97.8 |
| 7 | Immobility/stupor,Mutism,Echopraxia/echolalia,Mannerisms,Rigidity,Waxy flexibility,Withdrawal | 96.0 | 94.2 | 97.8 |
| 7 | Immobility/stupor,Mutism,Echopraxia/echolalia,Stereotypy,Mannerisms,Verbigeration,Negativism | 96.0 | 93.9 | 97.8 |
| 7 | Immobility/stupor,Mutism,Echopraxia/echolalia,Stereotypy,Verbigeration,Rigidity,Withdrawal | 96.0 | 93.9 | 97.8 |
| 7 | Immobility/stupor,Mutism,Grimacing,Echopraxia/echolalia,Mannerisms,Verbigeration,Rigidity | 96.0 | 93.9 | 97.8 |
| 7 | Immobility/stupor,Mutism,Grimacing,Echopraxia/echolalia,Rigidity,Negativism,Waxy flexibility | 96.0 | 93.9 | 97.8 |
| 7 | Immobility/stupor,Mutism,Grimacing,Echopraxia/echolalia,Stereotypy,Mannerisms,Negativism | 96.0 | 93.9 | 97.8 |
| 7 | Immobility/stupor,Mutism,Grimacing,Echopraxia/echolalia,Stereotypy,Mannerisms,Rigidity | 96.0 | 93.9 | 97.8 |
| 7 | Immobility/stupor,Mutism,Grimacing,Rigidity,Negativism,Waxy flexibility,Withdrawal | 96.0 | 94.2 | 97.8 |
| 7 | Immobility/stupor,Mutism,Grimacing,Stereotypy,Mannerisms,Negativism,Waxy flexibility | 96.0 | 94.2 | 97.8 |
| 7 | Immobility/stupor,Mutism,Grimacing,Stereotypy,Mannerisms,Rigidity,Waxy flexibility | 96.0 | 94.2 | 97.8 |
| 7 | Immobility/stupor,Mutism,Grimacing,Stereotypy,Rigidity,Waxy flexibility,Withdrawal | 96.0 | 93.9 | 97.8 |
| 7 | Immobility/stupor,Mutism,Grimacing,Stereotypy,Verbigeration,Rigidity,Waxy flexibility | 96.0 | 94.2 | 97.8 |
| 7 | Immobility/stupor,Mutism,Grimacing,Verbigeration,Rigidity,Waxy flexibility,Withdrawal | 96.0 | 93.9 | 97.8 |
| 7 | Immobility/stupor,Mutism,Mannerisms,Verbigeration,Negativism,Waxy flexibility,Withdrawal | 96.0 | 94.2 | 97.8 |
| 7 | Immobility/stupor,Mutism,Mannerisms,Verbigeration,Rigidity,Waxy flexibility,Withdrawal | 96.0 | 93.9 | 97.8 |
| 7 | Immobility/stupor,Mutism,Posturing/catalepsy,Echopraxia/echolalia,Mannerisms,Negativism,Waxy flexibility | 96.0 | 94.2 | 97.8 |
| 7 | Immobility/stupor,Mutism,Posturing/catalepsy,Echopraxia/echolalia,Mannerisms,Verbigeration,Rigidity | 96.0 | 94.2 | 97.8 |
| 7 | Immobility/stupor,Mutism,Posturing/catalepsy,Echopraxia/echolalia,Rigidity,Negativism,Waxy flexibility | 96.0 | 94.2 | 97.8 |
| 7 | Immobility/stupor,Mutism,Posturing/catalepsy,Echopraxia/echolalia,Stereotypy,Rigidity,Withdrawal | 96.0 | 94.2 | 97.8 |
| 7 | Immobility/stupor,Mutism,Posturing/catalepsy,Echopraxia/echolalia,Stereotypy,Verbigeration,Negativism | 96.0 | 93.9 | 97.8 |
| 7 | Immobility/stupor,Mutism,Posturing/catalepsy,Grimacing,Echopraxia/echolalia,Rigidity,Withdrawal | 96.0 | 93.9 | 97.8 |
| 7 | Immobility/stupor,Mutism,Posturing/catalepsy,Grimacing,Mannerisms,Negativism,Waxy flexibility | 96.0 | 93.9 | 97.8 |
| 7 | Immobility/stupor,Mutism,Posturing/catalepsy,Grimacing,Verbigeration,Rigidity,Withdrawal | 96.0 | 93.9 | 97.8 |
| 7 | Immobility/stupor,Mutism,Posturing/catalepsy,Stereotypy,Mannerisms,Negativism,Waxy flexibility | 96.0 | 94.2 | 97.8 |
| 7 | Immobility/stupor,Mutism,Posturing/catalepsy,Stereotypy,Mannerisms,Verbigeration,Rigidity | 96.0 | 93.9 | 97.8 |
| 7 | Immobility/stupor,Mutism,Posturing/catalepsy,Stereotypy,Verbigeration,Rigidity,Withdrawal | 96.0 | 93.9 | 97.8 |
| 7 | Immobility/stupor,Mutism,Staring,Echopraxia/echolalia,Stereotypy,Verbigeration,Waxy flexibility | 96.0 | 93.9 | 97.8 |
| 7 | Immobility/stupor,Mutism,Staring,Echopraxia/echolalia,Stereotypy,Verbigeration,Withdrawal | 96.0 | 94.2 | 97.8 |
| 7 | Immobility/stupor,Mutism,Staring,Grimacing,Echopraxia/echolalia,Rigidity,Waxy flexibility | 96.0 | 94.2 | 97.8 |
| 7 | Immobility/stupor,Mutism,Staring,Grimacing,Echopraxia/echolalia,Waxy flexibility,Withdrawal | 96.0 | 94.2 | 97.8 |
| 7 | Immobility/stupor,Mutism,Staring,Grimacing,Verbigeration,Waxy flexibility,Withdrawal | 96.0 | 93.9 | 97.8 |
| 7 | Immobility/stupor,Mutism,Staring,Posturing/catalepsy,Echopraxia/echolalia,Rigidity,Waxy flexibility | 96.0 | 93.9 | 97.8 |
| 7 | Immobility/stupor,Mutism,Staring,Posturing/catalepsy,Echopraxia/echolalia,Verbigeration,Waxy flexibility | 96.0 | 93.9 | 97.8 |
| 7 | Immobility/stupor,Mutism,Staring,Posturing/catalepsy,Grimacing,Verbigeration,Waxy flexibility | 96.0 | 93.9 | 97.8 |
| 7 | Immobility/stupor,Mutism,Staring,Posturing/catalepsy,Verbigeration,Waxy flexibility,Withdrawal | 96.0 | 94.2 | 97.8 |
| 7 | Immobility/stupor,Mutism,Stereotypy,Mannerisms,Verbigeration,Negativism,Waxy flexibility | 96.0 | 93.9 | 97.8 |
| 7 | Immobility/stupor,Mutism,Stereotypy,Verbigeration,Rigidity,Waxy flexibility,Withdrawal | 96.0 | 93.9 | 97.8 |
| 7 | Immobility/stupor,Posturing/catalepsy,Echopraxia/echolalia,Stereotypy,Mannerisms,Negativism,Waxy flexibility | 96.0 | 93.9 | 97.5 |
| 7 | Immobility/stupor,Posturing/catalepsy,Echopraxia/echolalia,Stereotypy,Mannerisms,Rigidity,Withdrawal | 96.0 | 93.9 | 97.8 |
| 7 | Immobility/stupor,Posturing/catalepsy,Echopraxia/echolalia,Stereotypy,Negativism,Waxy flexibility,Withdrawal | 96.0 | 93.9 | 97.8 |
| 7 | Immobility/stupor,Posturing/catalepsy,Grimacing,Echopraxia/echolalia,Mannerisms,Negativism,Waxy flexibility | 96.0 | 93.9 | 97.8 |
| 7 | Immobility/stupor,Posturing/catalepsy,Grimacing,Echopraxia/echolalia,Verbigeration,Negativism,Withdrawal | 96.0 | 93.9 | 97.8 |
| 7 | Immobility/stupor,Posturing/catalepsy,Grimacing,Stereotypy,Mannerisms,Negativism,Waxy flexibility | 96.0 | 93.9 | 97.8 |
| 7 | Immobility/stupor,Posturing/catalepsy,Grimacing,Stereotypy,Verbigeration,Rigidity,Withdrawal | 96.0 | 93.9 | 97.8 |
| 7 | Immobility/stupor,Posturing/catalepsy,Stereotypy,Mannerisms,Verbigeration,Negativism,Waxy flexibility | 96.0 | 94.2 | 97.8 |
| 7 | Immobility/stupor,Staring,Echopraxia/echolalia,Stereotypy,Mannerisms,Verbigeration,Waxy flexibility | 96.0 | 94.2 | 97.8 |
| 7 | Immobility/stupor,Staring,Grimacing,Echopraxia/echolalia,Stereotypy,Verbigeration,Waxy flexibility | 96.0 | 93.9 | 97.8 |
| 7 | Immobility/stupor,Staring,Posturing/catalepsy,Echopraxia/echolalia,Stereotypy,Mannerisms,Waxy flexibility | 96.0 | 94.2 | 97.8 |
| 7 | Immobility/stupor,Staring,Posturing/catalepsy,Grimacing,Echopraxia/echolalia,Stereotypy,Verbigeration | 96.0 | 94.2 | 97.8 |
| 7 | Immobility/stupor,Staring,Posturing/catalepsy,Grimacing,Stereotypy,Rigidity,Waxy flexibility | 96.0 | 93.9 | 97.8 |
| 7 | Immobility/stupor,Staring,Posturing/catalepsy,Grimacing,Stereotypy,Verbigeration,Waxy flexibility | 96.0 | 93.9 | 97.8 |
| 7 | Immobility/stupor,Staring,Posturing/catalepsy,Grimacing,Verbigeration,Rigidity,Waxy flexibility | 96.0 | 94.2 | 97.8 |
| 7 | Staring,Grimacing,Echopraxia/echolalia,Mannerisms,Verbigeration,Negativism,Waxy flexibility | 96.0 | 93.9 | 97.8 |
| 7 | Staring,Grimacing,Echopraxia/echolalia,Stereotypy,Verbigeration,Rigidity,Waxy flexibility | 96.0 | 94.2 | 97.8 |
| 7 | Staring,Posturing/catalepsy,Echopraxia/echolalia,Verbigeration,Rigidity,Negativism,Waxy flexibility | 96.0 | 93.9 | 97.8 |
| 7 | Staring,Posturing/catalepsy,Grimacing,Echopraxia/echolalia,Stereotypy,Verbigeration,Negativism | 96.0 | 93.9 | 97.8 |
| 7 | Staring,Posturing/catalepsy,Grimacing,Echopraxia/echolalia,Stereotypy,Verbigeration,Rigidity | 96.0 | 93.9 | 97.8 |
| 7 | Staring,Posturing/catalepsy,Grimacing,Echopraxia/echolalia,Stereotypy,Verbigeration,Withdrawal | 96.0 | 94.2 | 97.8 |
| 7 | Staring,Posturing/catalepsy,Grimacing,Echopraxia/echolalia,Stereotypy,Waxy flexibility,Withdrawal | 96.0 | 94.2 | 97.8 |
| 7 | Staring,Posturing/catalepsy,Grimacing,Mannerisms,Verbigeration,Negativism,Waxy flexibility | 96.0 | 93.9 | 97.8 |
| 7 | Staring,Posturing/catalepsy,Grimacing,Stereotypy,Verbigeration,Waxy flexibility,Withdrawal | 96.0 | 94.2 | 97.8 |
| 7 | Excitement,Immobility/stupor,Mutism,Stereotypy,Mannerisms,Waxy flexibility,Withdrawal | 95.9 | 93.9 | 97.8 |
| 7 | Excitement,Immobility/stupor,Posturing/catalepsy,Grimacing,Stereotypy,Mannerisms,Withdrawal | 95.9 | 93.9 | 97.8 |
| 7 | Excitement,Mutism,Stereotypy,Verbigeration,Rigidity,Negativism,Withdrawal | 95.9 | 93.9 | 97.8 |
| 7 | Immobility/stupor,Grimacing,Echopraxia/echolalia,Stereotypy,Rigidity,Waxy flexibility,Withdrawal | 95.9 | 93.9 | 97.8 |
| 7 | Immobility/stupor,Mutism,Staring,Posturing/catalepsy,Echopraxia/echolalia,Stereotypy,Waxy flexibility | 95.9 | 93.9 | 97.5 |
| 7 | Immobility/stupor,Mutism,Staring,Posturing/catalepsy,Grimacing,Negativism,Waxy flexibility | 95.9 | 93.9 | 97.8 |
| 7 | Immobility/stupor,Posturing/catalepsy,Echopraxia/echolalia,Stereotypy,Mannerisms,Verbigeration,Negativism | 95.9 | 93.9 | 97.8 |
| 7 | Staring,Posturing/catalepsy,Echopraxia/echolalia,Stereotypy,Mannerisms,Verbigeration,Withdrawal | 95.9 | 93.9 | 97.8 |
| 7 | Excitement,Immobility/stupor,Mutism,Grimacing,Echopraxia/echolalia,Mannerisms,Negativism | 95.8 | 93.9 | 97.5 |
| 7 | Excitement,Immobility/stupor,Posturing/catalepsy,Grimacing,Stereotypy,Waxy flexibility,Withdrawal | 95.8 | 93.7 | 97.5 |
| 7 | Excitement,Staring,Grimacing,Echopraxia/echolalia,Mannerisms,Verbigeration,Waxy flexibility | 95.8 | 93.7 | 97.5 |
| 7 | Immobility/stupor,Echopraxia/echolalia,Stereotypy,Mannerisms,Verbigeration,Rigidity,Withdrawal | 95.8 | 93.7 | 97.5 |
| 7 | Immobility/stupor,Mutism,Grimacing,Echopraxia/echolalia,Mannerisms,Rigidity,Waxy flexibility | 95.8 | 93.7 | 97.5 |
| 7 | Immobility/stupor,Mutism,Grimacing,Echopraxia/echolalia,Stereotypy,Verbigeration,Rigidity | 95.8 | 93.7 | 97.5 |
| 7 | Immobility/stupor,Mutism,Posturing/catalepsy,Rigidity,Negativism,Waxy flexibility,Withdrawal | 95.8 | 93.7 | 97.5 |
| 7 | Immobility/stupor,Posturing/catalepsy,Grimacing,Echopraxia/echolalia,Stereotypy,Verbigeration,Negativism | 95.8 | 93.7 | 97.5 |
| 7 | Immobility/stupor,Posturing/catalepsy,Grimacing,Mannerisms,Verbigeration,Negativism,Waxy flexibility | 95.8 | 93.7 | 97.5 |
| 7 | Immobility/stupor,Staring,Posturing/catalepsy,Echopraxia/echolalia,Verbigeration,Rigidity,Waxy flexibility | 95.8 | 93.7 | 97.5 |
| 7 | Staring,Posturing/catalepsy,Grimacing,Stereotypy,Rigidity,Negativism,Waxy flexibility | 95.8 | 93.7 | 97.5 |
| 7 | Staring,Posturing/catalepsy,Grimacing,Stereotypy,Verbigeration,Rigidity,Waxy flexibility | 95.8 | 93.7 | 97.5 |
| 7 | Excitement,Immobility/stupor,Grimacing,Echopraxia/echolalia,Mannerisms,Verbigeration,Negativism | 95.7 | 93.7 | 97.5 |
| 7 | Excitement,Immobility/stupor,Grimacing,Echopraxia/echolalia,Stereotypy,Mannerisms,Negativism | 95.7 | 93.7 | 97.5 |
| 7 | Excitement,Immobility/stupor,Grimacing,Echopraxia/echolalia,Stereotypy,Mannerisms,Withdrawal | 95.7 | 93.7 | 97.5 |
| 7 | Excitement,Immobility/stupor,Grimacing,Echopraxia/echolalia,Stereotypy,Verbigeration,Withdrawal | 95.7 | 93.7 | 97.5 |
| 7 | Excitement,Immobility/stupor,Grimacing,Stereotypy,Mannerisms,Negativism,Waxy flexibility | 95.7 | 93.7 | 97.5 |
| 7 | Excitement,Immobility/stupor,Grimacing,Stereotypy,Mannerisms,Verbigeration,Negativism | 95.7 | 93.7 | 97.5 |
| 7 | Excitement,Immobility/stupor,Mutism,Grimacing,Echopraxia/echolalia,Mannerisms,Waxy flexibility | 95.7 | 93.7 | 97.5 |
| 7 | Excitement,Immobility/stupor,Mutism,Grimacing,Echopraxia/echolalia,Mannerisms,Withdrawal | 95.7 | 93.7 | 97.5 |
| 7 | Excitement,Immobility/stupor,Mutism,Grimacing,Mannerisms,Verbigeration,Withdrawal | 95.7 | 93.7 | 97.5 |
| 7 | Excitement,Immobility/stupor,Mutism,Grimacing,Stereotypy,Mannerisms,Negativism | 95.7 | 93.7 | 97.5 |
| 7 | Excitement,Immobility/stupor,Posturing/catalepsy,Grimacing,Echopraxia/echolalia,Mannerisms,Verbigeration | 95.7 | 93.7 | 97.5 |
| 7 | Excitement,Immobility/stupor,Posturing/catalepsy,Grimacing,Mannerisms,Verbigeration,Waxy flexibility | 95.7 | 93.7 | 97.5 |
| 7 | Excitement,Mutism,Echopraxia/echolalia,Stereotypy,Rigidity,Negativism,Withdrawal | 95.7 | 93.7 | 97.5 |
| 7 | Excitement,Mutism,Posturing/catalepsy,Echopraxia/echolalia,Verbigeration,Rigidity,Negativism | 95.7 | 93.7 | 97.5 |
| 7 | Excitement,Mutism,Posturing/catalepsy,Rigidity,Negativism,Waxy flexibility,Withdrawal | 95.7 | 93.7 | 97.5 |
| 7 | Excitement,Mutism,Posturing/catalepsy,Verbigeration,Rigidity,Waxy flexibility,Withdrawal | 95.7 | 93.7 | 97.5 |
| 7 | Excitement,Mutism,Stereotypy,Rigidity,Negativism,Waxy flexibility,Withdrawal | 95.7 | 93.9 | 97.5 |
| 7 | Excitement,Staring,Grimacing,Echopraxia/echolalia,Mannerisms,Verbigeration,Negativism | 95.7 | 93.7 | 97.5 |
| 7 | Excitement,Staring,Grimacing,Echopraxia/echolalia,Stereotypy,Verbigeration,Waxy flexibility | 95.7 | 93.7 | 97.5 |
| 7 | Excitement,Staring,Grimacing,Stereotypy,Mannerisms,Verbigeration,Waxy flexibility | 95.7 | 93.7 | 97.5 |
| 7 | Excitement,Staring,Posturing/catalepsy,Echopraxia/echolalia,Mannerisms,Verbigeration,Waxy flexibility | 95.7 | 93.7 | 97.5 |
| 7 | Excitement,Staring,Posturing/catalepsy,Echopraxia/echolalia,Stereotypy,Mannerisms,Waxy flexibility | 95.7 | 93.7 | 97.5 |
| 7 | Excitement,Staring,Posturing/catalepsy,Stereotypy,Mannerisms,Verbigeration,Waxy flexibility | 95.7 | 93.7 | 97.5 |
| 7 | Immobility/stupor,Echopraxia/echolalia,Mannerisms,Verbigeration,Negativism,Waxy flexibility,Withdrawal | 95.7 | 93.7 | 97.5 |
| 7 | Immobility/stupor,Echopraxia/echolalia,Stereotypy,Mannerisms,Rigidity,Waxy flexibility,Withdrawal | 95.7 | 93.7 | 97.5 |
| 7 | Immobility/stupor,Grimacing,Echopraxia/echolalia,Stereotypy,Mannerisms,Verbigeration,Negativism | 95.7 | 93.7 | 97.5 |
| 7 | Immobility/stupor,Grimacing,Stereotypy,Mannerisms,Verbigeration,Negativism,Waxy flexibility | 95.7 | 93.7 | 97.5 |
| 7 | Immobility/stupor,Grimacing,Stereotypy,Verbigeration,Rigidity,Waxy flexibility,Withdrawal | 95.7 | 93.7 | 97.5 |
| 7 | Immobility/stupor,Mutism,Echopraxia/echolalia,Mannerisms,Verbigeration,Negativism,Waxy flexibility | 95.7 | 93.7 | 97.5 |
| 7 | Immobility/stupor,Mutism,Echopraxia/echolalia,Stereotypy,Negativism,Waxy flexibility,Withdrawal | 95.7 | 93.7 | 97.5 |
| 7 | Immobility/stupor,Mutism,Grimacing,Echopraxia/echolalia,Mannerisms,Verbigeration,Negativism | 95.7 | 93.7 | 97.5 |
| 7 | Immobility/stupor,Mutism,Grimacing,Echopraxia/echolalia,Rigidity,Waxy flexibility,Withdrawal | 95.7 | 93.7 | 97.5 |
| 7 | Immobility/stupor,Mutism,Grimacing,Echopraxia/echolalia,Verbigeration,Negativism,Withdrawal | 95.7 | 93.7 | 97.5 |
| 7 | Immobility/stupor,Mutism,Grimacing,Mannerisms,Verbigeration,Rigidity,Waxy flexibility | 95.7 | 93.7 | 97.5 |
| 7 | Immobility/stupor,Mutism,Grimacing,Stereotypy,Mannerisms,Verbigeration,Negativism | 95.7 | 93.7 | 97.5 |
| 7 | Immobility/stupor,Mutism,Posturing/catalepsy,Echopraxia/echolalia,Negativism,Waxy flexibility,Withdrawal | 95.7 | 93.7 | 97.5 |
| 7 | Immobility/stupor,Mutism,Posturing/catalepsy,Echopraxia/echolalia,Stereotypy,Mannerisms,Rigidity | 95.7 | 93.7 | 97.5 |
| 7 | Immobility/stupor,Mutism,Posturing/catalepsy,Echopraxia/echolalia,Stereotypy,Verbigeration,Rigidity | 95.7 | 93.7 | 97.5 |
| 7 | Immobility/stupor,Mutism,Posturing/catalepsy,Grimacing,Echopraxia/echolalia,Stereotypy,Negativism | 95.7 | 93.7 | 97.5 |
| 7 | Immobility/stupor,Mutism,Posturing/catalepsy,Grimacing,Echopraxia/echolalia,Verbigeration,Rigidity | 95.7 | 93.7 | 97.5 |
| 7 | Immobility/stupor,Mutism,Posturing/catalepsy,Mannerisms,Rigidity,Waxy flexibility,Withdrawal | 95.7 | 93.7 | 97.5 |
| 7 | Immobility/stupor,Mutism,Posturing/catalepsy,Mannerisms,Verbigeration,Negativism,Waxy flexibility | 95.7 | 93.7 | 97.5 |
| 7 | Immobility/stupor,Mutism,Posturing/catalepsy,Stereotypy,Negativism,Waxy flexibility,Withdrawal | 95.7 | 93.7 | 97.5 |
| 7 | Immobility/stupor,Mutism,Posturing/catalepsy,Stereotypy,Verbigeration,Negativism,Waxy flexibility | 95.7 | 93.7 | 97.5 |
| 7 | Immobility/stupor,Mutism,Posturing/catalepsy,Verbigeration,Negativism,Waxy flexibility,Withdrawal | 95.7 | 93.7 | 97.5 |
| 7 | Immobility/stupor,Mutism,Staring,Echopraxia/echolalia,Verbigeration,Waxy flexibility,Withdrawal | 95.7 | 93.7 | 97.5 |
| 7 | Immobility/stupor,Mutism,Staring,Posturing/catalepsy,Grimacing,Waxy flexibility,Withdrawal | 95.7 | 93.7 | 97.5 |
| 7 | Immobility/stupor,Mutism,Stereotypy,Verbigeration,Negativism,Waxy flexibility,Withdrawal | 95.7 | 93.7 | 97.5 |
| 7 | Immobility/stupor,Posturing/catalepsy,Echopraxia/echolalia,Mannerisms,Verbigeration,Rigidity,Withdrawal | 95.7 | 93.7 | 97.5 |
| 7 | Immobility/stupor,Posturing/catalepsy,Grimacing,Mannerisms,Rigidity,Waxy flexibility,Withdrawal | 95.7 | 93.7 | 97.5 |
| 7 | Immobility/stupor,Posturing/catalepsy,Grimacing,Stereotypy,Negativism,Waxy flexibility,Withdrawal | 95.7 | 93.7 | 97.5 |
| 7 | Immobility/stupor,Posturing/catalepsy,Grimacing,Stereotypy,Verbigeration,Negativism,Waxy flexibility | 95.7 | 93.7 | 97.5 |
| 7 | Immobility/stupor,Posturing/catalepsy,Stereotypy,Mannerisms,Verbigeration,Rigidity,Withdrawal | 95.7 | 93.7 | 97.5 |
| 7 | Immobility/stupor,Staring,Echopraxia/echolalia,Mannerisms,Verbigeration,Waxy flexibility,Withdrawal | 95.7 | 93.7 | 97.5 |
| 7 | Immobility/stupor,Staring,Posturing/catalepsy,Echopraxia/echolalia,Mannerisms,Verbigeration,Waxy flexibility | 95.7 | 93.7 | 97.5 |
| 7 | Immobility/stupor,Staring,Posturing/catalepsy,Echopraxia/echolalia,Stereotypy,Rigidity,Waxy flexibility | 95.7 | 93.7 | 97.5 |
| 7 | Immobility/stupor,Staring,Posturing/catalepsy,Echopraxia/echolalia,Stereotypy,Verbigeration,Withdrawal | 95.7 | 93.7 | 97.5 |
| 7 | Immobility/stupor,Staring,Posturing/catalepsy,Echopraxia/echolalia,Stereotypy,Waxy flexibility,Withdrawal | 95.7 | 93.7 | 97.5 |
| 7 | Immobility/stupor,Staring,Posturing/catalepsy,Grimacing,Echopraxia/echolalia,Verbigeration,Withdrawal | 95.7 | 93.7 | 97.5 |
| 7 | Immobility/stupor,Staring,Posturing/catalepsy,Grimacing,Rigidity,Waxy flexibility,Withdrawal | 95.7 | 93.7 | 97.5 |
| 7 | Immobility/stupor,Staring,Posturing/catalepsy,Grimacing,Stereotypy,Waxy flexibility,Withdrawal | 95.7 | 93.7 | 97.5 |
| 7 | Immobility/stupor,Staring,Posturing/catalepsy,Stereotypy,Verbigeration,Waxy flexibility,Withdrawal | 95.7 | 93.7 | 97.5 |
| 7 | Mutism,Posturing/catalepsy,Mannerisms,Verbigeration,Rigidity,Negativism,Withdrawal | 95.7 | 93.7 | 97.5 |
| 7 | Staring,Echopraxia/echolalia,Stereotypy,Mannerisms,Verbigeration,Waxy flexibility,Withdrawal | 95.7 | 93.7 | 97.5 |
| 7 | Staring,Grimacing,Echopraxia/echolalia,Stereotypy,Verbigeration,Waxy flexibility,Withdrawal | 95.7 | 93.7 | 97.5 |
| 7 | Staring,Posturing/catalepsy,Echopraxia/echolalia,Mannerisms,Verbigeration,Waxy flexibility,Withdrawal | 95.7 | 93.7 | 97.5 |
| 7 | Staring,Posturing/catalepsy,Echopraxia/echolalia,Stereotypy,Mannerisms,Verbigeration,Rigidity | 95.7 | 93.7 | 97.5 |
| 7 | Staring,Posturing/catalepsy,Echopraxia/echolalia,Stereotypy,Verbigeration,Negativism,Waxy flexibility | 95.7 | 93.7 | 97.5 |
| 7 | Staring,Posturing/catalepsy,Echopraxia/echolalia,Verbigeration,Rigidity,Waxy flexibility,Withdrawal | 95.7 | 93.7 | 97.5 |
| 7 | Staring,Posturing/catalepsy,Grimacing,Echopraxia/echolalia,Mannerisms,Rigidity,Waxy flexibility | 95.7 | 93.7 | 97.5 |
| 7 | Staring,Posturing/catalepsy,Grimacing,Mannerisms,Verbigeration,Rigidity,Waxy flexibility | 95.7 | 93.7 | 97.5 |
| 7 | Staring,Posturing/catalepsy,Grimacing,Stereotypy,Verbigeration,Negativism,Waxy flexibility | 95.7 | 93.7 | 97.5 |
| 7 | Excitement,Immobility/stupor,Grimacing,Echopraxia/echolalia,Mannerisms,Verbigeration,Withdrawal | 95.5 | 93.5 | 97.3 |
| 7 | Excitement,Immobility/stupor,Mutism,Echopraxia/echolalia,Mannerisms,Verbigeration,Waxy flexibility | 95.5 | 93.5 | 97.3 |
| 7 | Excitement,Immobility/stupor,Mutism,Echopraxia/echolalia,Stereotypy,Mannerisms,Waxy flexibility | 95.5 | 93.5 | 97.3 |
| 7 | Excitement,Immobility/stupor,Mutism,Grimacing,Echopraxia/echolalia,Stereotypy,Verbigeration | 95.5 | 93.5 | 97.3 |
| 7 | Excitement,Immobility/stupor,Mutism,Grimacing,Mannerisms,Verbigeration,Waxy flexibility | 95.5 | 93.5 | 97.3 |
| 7 | Excitement,Immobility/stupor,Mutism,Grimacing,Stereotypy,Mannerisms,Withdrawal | 95.5 | 93.5 | 97.3 |
| 7 | Excitement,Immobility/stupor,Mutism,Posturing/catalepsy,Stereotypy,Mannerisms,Waxy flexibility | 95.5 | 93.5 | 97.3 |
| 7 | Excitement,Immobility/stupor,Posturing/catalepsy,Echopraxia/echolalia,Mannerisms,Verbigeration,Waxy flexibility | 95.5 | 93.5 | 97.3 |
| 7 | Excitement,Immobility/stupor,Posturing/catalepsy,Grimacing,Echopraxia/echolalia,Stereotypy,Waxy flexibility | 95.5 | 93.5 | 97.3 |
| 7 | Excitement,Immobility/stupor,Posturing/catalepsy,Stereotypy,Mannerisms,Waxy flexibility,Withdrawal | 95.5 | 93.5 | 97.3 |
| 7 | Excitement,Immobility/stupor,Stereotypy,Mannerisms,Verbigeration,Waxy flexibility,Withdrawal | 95.5 | 93.5 | 97.3 |
| 7 | Excitement,Mutism,Echopraxia/echolalia,Verbigeration,Rigidity,Negativism,Withdrawal | 95.5 | 93.5 | 97.3 |
| 7 | Excitement,Mutism,Grimacing,Echopraxia/echolalia,Rigidity,Negativism,Withdrawal | 95.5 | 93.5 | 97.3 |
| 7 | Excitement,Mutism,Grimacing,Verbigeration,Rigidity,Negativism,Withdrawal | 95.5 | 93.5 | 97.3 |
| 7 | Excitement,Mutism,Posturing/catalepsy,Echopraxia/echolalia,Rigidity,Waxy flexibility,Withdrawal | 95.5 | 93.5 | 97.3 |
| 7 | Excitement,Mutism,Posturing/catalepsy,Grimacing,Echopraxia/echolalia,Rigidity,Negativism | 95.5 | 93.5 | 97.3 |
| 7 | Excitement,Mutism,Posturing/catalepsy,Grimacing,Verbigeration,Rigidity,Negativism | 95.5 | 93.5 | 97.3 |
| 7 | Excitement,Mutism,Posturing/catalepsy,Mannerisms,Verbigeration,Rigidity,Negativism | 95.5 | 93.5 | 97.3 |
| 7 | Excitement,Mutism,Posturing/catalepsy,Stereotypy,Verbigeration,Rigidity,Negativism | 95.5 | 93.5 | 97.3 |
| 7 | Excitement,Mutism,Verbigeration,Rigidity,Negativism,Waxy flexibility,Withdrawal | 95.5 | 93.5 | 97.3 |
| 7 | Excitement,Staring,Posturing/catalepsy,Echopraxia/echolalia,Stereotypy,Mannerisms,Verbigeration | 95.5 | 93.5 | 97.3 |
| 7 | Excitement,Staring,Posturing/catalepsy,Echopraxia/echolalia,Stereotypy,Verbigeration,Waxy flexibility | 95.5 | 93.5 | 97.3 |
| 7 | Immobility/stupor,Grimacing,Echopraxia/echolalia,Mannerisms,Verbigeration,Negativism,Waxy flexibility | 95.5 | 93.5 | 97.3 |
| 7 | Immobility/stupor,Grimacing,Echopraxia/echolalia,Stereotypy,Verbigeration,Negativism,Waxy flexibility | 95.5 | 93.5 | 97.3 |
| 7 | Immobility/stupor,Grimacing,Mannerisms,Verbigeration,Rigidity,Waxy flexibility,Withdrawal | 95.5 | 93.5 | 97.3 |
| 7 | Immobility/stupor,Grimacing,Stereotypy,Verbigeration,Negativism,Waxy flexibility,Withdrawal | 95.5 | 93.5 | 97.3 |
| 7 | Immobility/stupor,Mutism,Echopraxia/echolalia,Stereotypy,Mannerisms,Verbigeration,Rigidity | 95.5 | 93.5 | 97.3 |
| 7 | Immobility/stupor,Mutism,Echopraxia/echolalia,Stereotypy,Rigidity,Waxy flexibility,Withdrawal | 95.5 | 93.5 | 97.3 |
| 7 | Immobility/stupor,Mutism,Echopraxia/echolalia,Verbigeration,Negativism,Waxy flexibility,Withdrawal | 95.5 | 93.5 | 97.3 |
| 7 | Immobility/stupor,Mutism,Grimacing,Echopraxia/echolalia,Stereotypy,Verbigeration,Negativism | 95.5 | 93.5 | 97.3 |
| 7 | Immobility/stupor,Mutism,Grimacing,Echopraxia/echolalia,Verbigeration,Negativism,Waxy flexibility | 95.5 | 93.5 | 97.3 |
| 7 | Immobility/stupor,Mutism,Posturing/catalepsy,Echopraxia/echolalia,Verbigeration,Negativism,Waxy flexibility | 95.5 | 93.5 | 97.3 |
| 7 | Immobility/stupor,Mutism,Posturing/catalepsy,Echopraxia/echolalia,Verbigeration,Rigidity,Withdrawal | 95.5 | 93.5 | 97.3 |
| 7 | Immobility/stupor,Mutism,Posturing/catalepsy,Grimacing,Echopraxia/echolalia,Stereotypy,Rigidity | 95.5 | 93.5 | 97.3 |
| 7 | Immobility/stupor,Mutism,Posturing/catalepsy,Grimacing,Echopraxia/echolalia,Stereotypy,Withdrawal | 95.5 | 93.5 | 97.3 |
| 7 | Immobility/stupor,Mutism,Posturing/catalepsy,Grimacing,Mannerisms,Verbigeration,Withdrawal | 95.5 | 93.5 | 97.3 |
| 7 | Immobility/stupor,Mutism,Posturing/catalepsy,Grimacing,Negativism,Waxy flexibility,Withdrawal | 95.5 | 93.5 | 97.3 |
| 7 | Immobility/stupor,Mutism,Posturing/catalepsy,Grimacing,Rigidity,Negativism,Waxy flexibility | 95.5 | 93.5 | 97.3 |
| 7 | Immobility/stupor,Mutism,Posturing/catalepsy,Grimacing,Stereotypy,Mannerisms,Withdrawal | 95.5 | 93.5 | 97.3 |
| 7 | Immobility/stupor,Mutism,Posturing/catalepsy,Grimacing,Verbigeration,Negativism,Waxy flexibility | 95.5 | 93.5 | 97.3 |
| 7 | Immobility/stupor,Mutism,Staring,Posturing/catalepsy,Grimacing,Echopraxia/echolalia,Waxy flexibility | 95.5 | 93.5 | 97.3 |
| 7 | Immobility/stupor,Mutism,Staring,Posturing/catalepsy,Grimacing,Stereotypy,Waxy flexibility | 95.5 | 93.5 | 97.3 |
| 7 | Immobility/stupor,Mutism,Stereotypy,Mannerisms,Verbigeration,Rigidity,Waxy flexibility | 95.5 | 93.5 | 97.3 |
| 7 | Immobility/stupor,Posturing/catalepsy,Echopraxia/echolalia,Mannerisms,Verbigeration,Negativism,Waxy flexibility | 95.5 | 93.5 | 97.3 |
| 7 | Immobility/stupor,Posturing/catalepsy,Echopraxia/echolalia,Verbigeration,Negativism,Waxy flexibility,Withdrawal | 95.5 | 93.5 | 97.3 |
| 7 | Immobility/stupor,Posturing/catalepsy,Grimacing,Echopraxia/echolalia,Negativism,Waxy flexibility,Withdrawal | 95.5 | 93.5 | 97.3 |
| 7 | Immobility/stupor,Posturing/catalepsy,Grimacing,Echopraxia/echolalia,Stereotypy,Mannerisms,Withdrawal | 95.5 | 93.5 | 97.3 |
| 7 | Immobility/stupor,Posturing/catalepsy,Grimacing,Echopraxia/echolalia,Stereotypy,Negativism,Waxy flexibility | 95.5 | 93.5 | 97.3 |
| 7 | Immobility/stupor,Posturing/catalepsy,Grimacing,Echopraxia/echolalia,Verbigeration,Negativism,Waxy flexibility | 95.5 | 93.5 | 97.3 |
| 7 | Immobility/stupor,Posturing/catalepsy,Grimacing,Echopraxia/echolalia,Verbigeration,Rigidity,Withdrawal | 95.5 | 93.5 | 97.3 |
| 7 | Immobility/stupor,Posturing/catalepsy,Grimacing,Verbigeration,Negativism,Waxy flexibility,Withdrawal | 95.5 | 93.5 | 97.3 |
| 7 | Immobility/stupor,Staring,Posturing/catalepsy,Grimacing,Echopraxia/echolalia,Stereotypy,Waxy flexibility | 95.5 | 93.5 | 97.3 |
| 7 | Immobility/stupor,Staring,Posturing/catalepsy,Grimacing,Echopraxia/echolalia,Waxy flexibility,Withdrawal | 95.5 | 93.5 | 97.3 |
| 7 | Mutism,Posturing/catalepsy,Echopraxia/echolalia,Mannerisms,Rigidity,Negativism,Withdrawal | 95.5 | 93.5 | 97.3 |
| 7 | Mutism,Posturing/catalepsy,Grimacing,Mannerisms,Rigidity,Negativism,Withdrawal | 95.5 | 93.5 | 97.3 |
| 7 | Mutism,Posturing/catalepsy,Stereotypy,Mannerisms,Rigidity,Negativism,Withdrawal | 95.5 | 93.5 | 97.3 |
| 7 | Mutism,Stereotypy,Mannerisms,Rigidity,Negativism,Waxy flexibility,Withdrawal | 95.5 | 93.5 | 97.3 |
| 7 | Mutism,Stereotypy,Mannerisms,Verbigeration,Rigidity,Negativism,Withdrawal | 95.5 | 93.5 | 97.3 |
| 7 | Staring,Grimacing,Echopraxia/echolalia,Stereotypy,Verbigeration,Negativism,Waxy flexibility | 95.5 | 93.5 | 97.3 |
| 7 | Staring,Posturing/catalepsy,Grimacing,Echopraxia/echolalia,Rigidity,Negativism,Waxy flexibility | 95.5 | 93.5 | 97.3 |
| 7 | Staring,Posturing/catalepsy,Grimacing,Echopraxia/echolalia,Stereotypy,Negativism,Waxy flexibility | 95.5 | 93.5 | 97.3 |
| 7 | Staring,Posturing/catalepsy,Grimacing,Verbigeration,Rigidity,Negativism,Waxy flexibility | 95.5 | 93.5 | 97.3 |
| 7 | Staring,Posturing/catalepsy,Stereotypy,Mannerisms,Verbigeration,Rigidity,Waxy flexibility | 95.5 | 93.5 | 97.3 |
| 7 | Excitement,Immobility/stupor,Grimacing,Mannerisms,Verbigeration,Waxy flexibility,Withdrawal | 95.3 | 93.3 | 97.1 |
| 7 | Excitement,Immobility/stupor,Grimacing,Stereotypy,Verbigeration,Waxy flexibility,Withdrawal | 95.3 | 93.3 | 97.1 |
| 7 | Excitement,Immobility/stupor,Mutism,Grimacing,Stereotypy,Mannerisms,Waxy flexibility | 95.3 | 93.3 | 97.1 |
| 7 | Excitement,Immobility/stupor,Mutism,Stereotypy,Mannerisms,Verbigeration,Waxy flexibility | 95.3 | 93.3 | 97.1 |
| 7 | Excitement,Immobility/stupor,Posturing/catalepsy,Echopraxia/echolalia,Stereotypy,Verbigeration,Waxy flexibility | 95.3 | 93.3 | 97.1 |
| 7 | Excitement,Immobility/stupor,Posturing/catalepsy,Grimacing,Echopraxia/echolalia,Stereotypy,Mannerisms | 95.3 | 93.3 | 97.1 |
| 7 | Excitement,Immobility/stupor,Posturing/catalepsy,Grimacing,Echopraxia/echolalia,Stereotypy,Verbigeration | 95.3 | 93.3 | 97.1 |
| 7 | Excitement,Immobility/stupor,Posturing/catalepsy,Grimacing,Stereotypy,Verbigeration,Waxy flexibility | 95.3 | 93.3 | 97.1 |
| 7 | Excitement,Mutism,Echopraxia/echolalia,Rigidity,Negativism,Waxy flexibility,Withdrawal | 95.3 | 93.3 | 97.1 |
| 7 | Excitement,Mutism,Grimacing,Rigidity,Negativism,Waxy flexibility,Withdrawal | 95.3 | 93.3 | 97.1 |
| 7 | Excitement,Mutism,Grimacing,Stereotypy,Rigidity,Negativism,Withdrawal | 95.3 | 93.3 | 97.1 |
| 7 | Excitement,Mutism,Mannerisms,Verbigeration,Rigidity,Negativism,Withdrawal | 95.3 | 93.3 | 97.1 |
| 7 | Excitement,Mutism,Posturing/catalepsy,Echopraxia/echolalia,Verbigeration,Rigidity,Withdrawal | 95.3 | 93.3 | 97.1 |
| 7 | Excitement,Mutism,Posturing/catalepsy,Grimacing,Rigidity,Waxy flexibility,Withdrawal | 95.3 | 93.3 | 97.1 |
| 7 | Excitement,Mutism,Posturing/catalepsy,Mannerisms,Rigidity,Waxy flexibility,Withdrawal | 95.3 | 93.3 | 97.1 |
| 7 | Excitement,Mutism,Posturing/catalepsy,Mannerisms,Verbigeration,Rigidity,Withdrawal | 95.3 | 93.3 | 97.1 |
| 7 | Excitement,Mutism,Posturing/catalepsy,Stereotypy,Rigidity,Waxy flexibility,Withdrawal | 95.3 | 93.3 | 97.1 |
| 7 | Excitement,Mutism,Posturing/catalepsy,Verbigeration,Rigidity,Negativism,Waxy flexibility | 95.3 | 93.3 | 97.1 |
| 7 | Excitement,Mutism,Stereotypy,Verbigeration,Rigidity,Waxy flexibility,Withdrawal | 95.3 | 93.3 | 97.1 |
| 7 | Excitement,Staring,Echopraxia/echolalia,Stereotypy,Mannerisms,Verbigeration,Waxy flexibility | 95.3 | 93.3 | 97.1 |
| 7 | Immobility/stupor,Grimacing,Echopraxia/echolalia,Stereotypy,Mannerisms,Rigidity,Waxy flexibility | 95.3 | 93.3 | 97.1 |
| 7 | Immobility/stupor,Grimacing,Echopraxia/echolalia,Verbigeration,Rigidity,Waxy flexibility,Withdrawal | 95.3 | 93.3 | 97.1 |
| 7 | Immobility/stupor,Grimacing,Mannerisms,Verbigeration,Negativism,Waxy flexibility,Withdrawal | 95.3 | 93.3 | 97.1 |
| 7 | Immobility/stupor,Mutism,Echopraxia/echolalia,Mannerisms,Verbigeration,Rigidity,Waxy flexibility | 95.3 | 93.3 | 97.1 |
| 7 | Immobility/stupor,Mutism,Echopraxia/echolalia,Stereotypy,Mannerisms,Rigidity,Waxy flexibility | 95.3 | 93.3 | 97.1 |
| 7 | Immobility/stupor,Mutism,Echopraxia/echolalia,Stereotypy,Verbigeration,Negativism,Waxy flexibility | 95.3 | 93.3 | 97.1 |
| 7 | Immobility/stupor,Mutism,Echopraxia/echolalia,Verbigeration,Rigidity,Waxy flexibility,Withdrawal | 95.3 | 93.3 | 97.1 |
| 7 | Immobility/stupor,Mutism,Grimacing,Echopraxia/echolalia,Mannerisms,Waxy flexibility,Withdrawal | 95.3 | 93.3 | 97.1 |
| 7 | Immobility/stupor,Mutism,Grimacing,Echopraxia/echolalia,Negativism,Waxy flexibility,Withdrawal | 95.3 | 93.3 | 97.1 |
| 7 | Immobility/stupor,Mutism,Grimacing,Echopraxia/echolalia,Stereotypy,Mannerisms,Withdrawal | 95.3 | 93.3 | 97.1 |
| 7 | Immobility/stupor,Mutism,Grimacing,Echopraxia/echolalia,Stereotypy,Negativism,Waxy flexibility | 95.3 | 93.3 | 97.1 |
| 7 | Immobility/stupor,Mutism,Grimacing,Echopraxia/echolalia,Verbigeration,Rigidity,Waxy flexibility | 95.3 | 93.3 | 97.1 |
| 7 | Immobility/stupor,Mutism,Grimacing,Mannerisms,Verbigeration,Negativism,Waxy flexibility | 95.3 | 93.3 | 97.1 |
| 7 | Immobility/stupor,Mutism,Grimacing,Stereotypy,Negativism,Waxy flexibility,Withdrawal | 95.3 | 93.3 | 97.1 |
| 7 | Immobility/stupor,Mutism,Grimacing,Stereotypy,Verbigeration,Negativism,Waxy flexibility | 95.3 | 93.3 | 97.1 |
| 7 | Immobility/stupor,Mutism,Posturing/catalepsy,Echopraxia/echolalia,Stereotypy,Negativism,Waxy flexibility | 95.3 | 93.3 | 97.1 |
| 7 | Immobility/stupor,Mutism,Posturing/catalepsy,Grimacing,Echopraxia/echolalia,Negativism,Waxy flexibility | 95.3 | 93.3 | 97.1 |
| 7 | Immobility/stupor,Mutism,Posturing/catalepsy,Grimacing,Mannerisms,Rigidity,Waxy flexibility | 95.3 | 93.3 | 97.1 |
| 7 | Immobility/stupor,Mutism,Posturing/catalepsy,Grimacing,Mannerisms,Waxy flexibility,Withdrawal | 95.3 | 93.3 | 97.1 |
| 7 | Immobility/stupor,Mutism,Posturing/catalepsy,Grimacing,Stereotypy,Negativism,Waxy flexibility | 95.3 | 93.3 | 97.1 |
| 7 | Immobility/stupor,Mutism,Posturing/catalepsy,Stereotypy,Verbigeration,Rigidity,Waxy flexibility | 95.3 | 93.3 | 97.1 |
| 7 | Immobility/stupor,Posturing/catalepsy,Echopraxia/echolalia,Mannerisms,Rigidity,Waxy flexibility,Withdrawal | 95.3 | 93.3 | 97.1 |
| 7 | Immobility/stupor,Posturing/catalepsy,Echopraxia/echolalia,Stereotypy,Verbigeration,Negativism,Waxy flexibility | 95.3 | 93.3 | 97.1 |
| 7 | Immobility/stupor,Posturing/catalepsy,Echopraxia/echolalia,Stereotypy,Verbigeration,Rigidity,Withdrawal | 95.3 | 93.3 | 97.1 |
| 7 | Immobility/stupor,Posturing/catalepsy,Grimacing,Echopraxia/echolalia,Mannerisms,Verbigeration,Rigidity | 95.3 | 93.3 | 97.1 |
| 7 | Immobility/stupor,Posturing/catalepsy,Grimacing,Echopraxia/echolalia,Stereotypy,Mannerisms,Rigidity | 95.3 | 93.3 | 97.1 |
| 7 | Immobility/stupor,Posturing/catalepsy,Grimacing,Stereotypy,Mannerisms,Verbigeration,Rigidity | 95.3 | 93.3 | 97.1 |
| 7 | Immobility/stupor,Posturing/catalepsy,Stereotypy,Mannerisms,Rigidity,Waxy flexibility,Withdrawal | 95.3 | 93.3 | 97.1 |
| 7 | Immobility/stupor,Staring,Echopraxia/echolalia,Stereotypy,Verbigeration,Waxy flexibility,Withdrawal | 95.3 | 93.3 | 97.1 |
| 7 | Immobility/stupor,Staring,Grimacing,Echopraxia/echolalia,Verbigeration,Waxy flexibility,Withdrawal | 95.3 | 93.3 | 97.1 |
| 7 | Immobility/stupor,Staring,Posturing/catalepsy,Echopraxia/echolalia,Stereotypy,Verbigeration,Waxy flexibility | 95.3 | 93.3 | 97.1 |
| 7 | Immobility/stupor,Staring,Posturing/catalepsy,Grimacing,Echopraxia/echolalia,Rigidity,Waxy flexibility | 95.3 | 93.3 | 97.1 |
| 7 | Immobility/stupor,Staring,Posturing/catalepsy,Grimacing,Echopraxia/echolalia,Verbigeration,Waxy flexibility | 95.3 | 93.3 | 97.1 |
| 7 | Immobility/stupor,Staring,Posturing/catalepsy,Grimacing,Verbigeration,Waxy flexibility,Withdrawal | 95.3 | 93.3 | 97.1 |
| 7 | Immobility/stupor,Stereotypy,Mannerisms,Verbigeration,Rigidity,Waxy flexibility,Withdrawal | 95.3 | 93.3 | 97.1 |
| 7 | Mutism,Echopraxia/echolalia,Stereotypy,Mannerisms,Rigidity,Negativism,Withdrawal | 95.3 | 93.3 | 97.1 |
| 7 | Mutism,Posturing/catalepsy,Mannerisms,Rigidity,Negativism,Waxy flexibility,Withdrawal | 95.3 | 93.3 | 97.1 |
| 7 | Mutism,Posturing/catalepsy,Stereotypy,Verbigeration,Rigidity,Negativism,Withdrawal | 95.3 | 93.3 | 97.1 |
| 7 | Mutism,Stereotypy,Verbigeration,Rigidity,Negativism,Waxy flexibility,Withdrawal | 95.3 | 93.3 | 97.1 |
| 7 | Staring,Echopraxia/echolalia,Stereotypy,Mannerisms,Verbigeration,Rigidity,Waxy flexibility | 95.3 | 93.3 | 97.1 |
| 7 | Staring,Posturing/catalepsy,Grimacing,Echopraxia/echolalia,Stereotypy,Mannerisms,Verbigeration | 95.3 | 93.3 | 97.1 |
| 7 | Staring,Posturing/catalepsy,Grimacing,Echopraxia/echolalia,Stereotypy,Mannerisms,Waxy flexibility | 95.3 | 93.3 | 97.1 |
| 7 | Staring,Posturing/catalepsy,Grimacing,Echopraxia/echolalia,Verbigeration,Negativism,Waxy flexibility | 95.3 | 93.3 | 97.1 |
| 7 | Excitement,Immobility/stupor,Grimacing,Echopraxia/echolalia,Stereotypy,Mannerisms,Waxy flexibility | 95.1 | 92.8 | 97.1 |
| 7 | Excitement,Immobility/stupor,Grimacing,Stereotypy,Mannerisms,Verbigeration,Withdrawal | 95.1 | 93.0 | 96.9 |
| 7 | Excitement,Immobility/stupor,Grimacing,Stereotypy,Mannerisms,Waxy flexibility,Withdrawal | 95.1 | 92.8 | 96.9 |
| 7 | Excitement,Immobility/stupor,Mutism,Echopraxia/echolalia,Stereotypy,Mannerisms,Verbigeration | 95.1 | 93.0 | 96.9 |
| 7 | Excitement,Immobility/stupor,Posturing/catalepsy,Echopraxia/echolalia,Stereotypy,Mannerisms,Verbigeration | 95.1 | 93.0 | 97.1 |
| 7 | Excitement,Immobility/stupor,Posturing/catalepsy,Grimacing,Stereotypy,Mannerisms,Verbigeration | 95.1 | 93.0 | 97.1 |
| 7 | Excitement,Mutism,Echopraxia/echolalia,Mannerisms,Rigidity,Negativism,Withdrawal | 95.1 | 93.0 | 97.1 |
| 7 | Excitement,Mutism,Grimacing,Mannerisms,Rigidity,Negativism,Withdrawal | 95.1 | 93.0 | 97.1 |
| 7 | Excitement,Mutism,Mannerisms,Rigidity,Negativism,Waxy flexibility,Withdrawal | 95.1 | 93.0 | 96.9 |
| 7 | Excitement,Mutism,Posturing/catalepsy,Echopraxia/echolalia,Mannerisms,Rigidity,Negativism | 95.1 | 93.0 | 97.1 |
| 7 | Excitement,Mutism,Posturing/catalepsy,Echopraxia/echolalia,Rigidity,Negativism,Waxy flexibility | 95.1 | 93.0 | 96.9 |
| 7 | Excitement,Mutism,Posturing/catalepsy,Echopraxia/echolalia,Stereotypy,Rigidity,Negativism | 95.1 | 93.0 | 96.9 |
| 7 | Excitement,Mutism,Posturing/catalepsy,Grimacing,Echopraxia/echolalia,Rigidity,Withdrawal | 95.1 | 93.0 | 97.1 |
| 7 | Excitement,Mutism,Posturing/catalepsy,Grimacing,Verbigeration,Rigidity,Withdrawal | 95.1 | 93.0 | 96.9 |
| 7 | Excitement,Mutism,Posturing/catalepsy,Stereotypy,Verbigeration,Rigidity,Withdrawal | 95.1 | 92.8 | 96.9 |
| 7 | Excitement,Mutism,Stereotypy,Mannerisms,Rigidity,Negativism,Withdrawal | 95.1 | 93.0 | 96.9 |
| 7 | Excitement,Mutism,Stereotypy,Verbigeration,Rigidity,Negativism,Waxy flexibility | 95.1 | 93.0 | 96.9 |
| 7 | Excitement,Staring,Grimacing,Echopraxia/echolalia,Stereotypy,Mannerisms,Verbigeration | 95.1 | 93.0 | 96.9 |
| 7 | Immobility/stupor,Echopraxia/echolalia,Mannerisms,Verbigeration,Rigidity,Waxy flexibility,Withdrawal | 95.1 | 93.0 | 97.1 |
| 7 | Immobility/stupor,Echopraxia/echolalia,Stereotypy,Verbigeration,Rigidity,Waxy flexibility,Withdrawal | 95.1 | 93.0 | 97.1 |
| 7 | Immobility/stupor,Grimacing,Echopraxia/echolalia,Stereotypy,Mannerisms,Verbigeration,Rigidity | 95.1 | 93.0 | 96.9 |
| 7 | Immobility/stupor,Grimacing,Echopraxia/echolalia,Stereotypy,Mannerisms,Waxy flexibility,Withdrawal | 95.1 | 93.0 | 97.1 |
| 7 | Immobility/stupor,Grimacing,Echopraxia/echolalia,Verbigeration,Negativism,Waxy flexibility,Withdrawal | 95.1 | 92.8 | 96.9 |
| 7 | Immobility/stupor,Grimacing,Stereotypy,Mannerisms,Verbigeration,Rigidity,Waxy flexibility | 95.1 | 92.8 | 97.1 |
| 7 | Immobility/stupor,Mutism,Grimacing,Verbigeration,Negativism,Waxy flexibility,Withdrawal | 95.1 | 92.8 | 96.9 |
| 7 | Immobility/stupor,Mutism,Posturing/catalepsy,Echopraxia/echolalia,Mannerisms,Verbigeration,Withdrawal | 95.1 | 93.0 | 97.1 |
| 7 | Immobility/stupor,Mutism,Posturing/catalepsy,Echopraxia/echolalia,Stereotypy,Mannerisms,Withdrawal | 95.1 | 92.8 | 96.9 |
| 7 | Immobility/stupor,Mutism,Posturing/catalepsy,Grimacing,Echopraxia/echolalia,Mannerisms,Verbigeration | 95.1 | 93.0 | 96.9 |
| 7 | Immobility/stupor,Mutism,Posturing/catalepsy,Grimacing,Stereotypy,Verbigeration,Withdrawal | 95.1 | 93.0 | 96.9 |
| 7 | Immobility/stupor,Mutism,Posturing/catalepsy,Mannerisms,Verbigeration,Rigidity,Waxy flexibility | 95.1 | 93.0 | 96.9 |
| 7 | Immobility/stupor,Mutism,Posturing/catalepsy,Stereotypy,Rigidity,Waxy flexibility,Withdrawal | 95.1 | 93.0 | 97.1 |
| 7 | Immobility/stupor,Mutism,Posturing/catalepsy,Verbigeration,Rigidity,Waxy flexibility,Withdrawal | 95.1 | 93.0 | 97.1 |
| 7 | Immobility/stupor,Posturing/catalepsy,Grimacing,Echopraxia/echolalia,Rigidity,Waxy flexibility,Withdrawal | 95.1 | 93.0 | 97.1 |
| 7 | Immobility/stupor,Posturing/catalepsy,Grimacing,Echopraxia/echolalia,Stereotypy,Verbigeration,Rigidity | 95.1 | 93.0 | 97.1 |
| 7 | Immobility/stupor,Posturing/catalepsy,Grimacing,Stereotypy,Rigidity,Waxy flexibility,Withdrawal | 95.1 | 93.0 | 97.1 |
| 7 | Immobility/stupor,Posturing/catalepsy,Mannerisms,Verbigeration,Rigidity,Waxy flexibility,Withdrawal | 95.1 | 93.0 | 96.9 |
| 7 | Mutism,Grimacing,Echopraxia/echolalia,Mannerisms,Rigidity,Negativism,Withdrawal | 95.1 | 93.0 | 96.9 |
| 7 | Mutism,Grimacing,Mannerisms,Rigidity,Negativism,Waxy flexibility,Withdrawal | 95.1 | 93.0 | 97.1 |
| 7 | Mutism,Grimacing,Mannerisms,Verbigeration,Rigidity,Negativism,Withdrawal | 95.1 | 93.0 | 97.1 |
| 7 | Mutism,Grimacing,Stereotypy,Mannerisms,Rigidity,Negativism,Withdrawal | 95.1 | 92.8 | 97.1 |
| 7 | Mutism,Grimacing,Stereotypy,Verbigeration,Rigidity,Negativism,Withdrawal | 95.1 | 93.0 | 97.1 |
| 7 | Mutism,Mannerisms,Verbigeration,Rigidity,Negativism,Waxy flexibility,Withdrawal | 95.1 | 93.0 | 97.1 |
| 7 | Staring,Grimacing,Echopraxia/echolalia,Stereotypy,Mannerisms,Verbigeration,Waxy flexibility | 95.1 | 92.8 | 96.9 |
| 7 | Staring,Posturing/catalepsy,Echopraxia/echolalia,Mannerisms,Verbigeration,Rigidity,Waxy flexibility | 95.1 | 93.0 | 97.1 |
| 7 | Staring,Posturing/catalepsy,Echopraxia/echolalia,Stereotypy,Mannerisms,Rigidity,Waxy flexibility | 95.1 | 93.0 | 96.9 |
| 7 | Staring,Posturing/catalepsy,Grimacing,Echopraxia/echolalia,Stereotypy,Rigidity,Waxy flexibility | 95.1 | 93.0 | 97.1 |
| 7 | Staring,Posturing/catalepsy,Grimacing,Echopraxia/echolalia,Verbigeration,Waxy flexibility,Withdrawal | 95.1 | 93.0 | 96.9 |
| 7 | Staring,Posturing/catalepsy,Grimacing,Stereotypy,Mannerisms,Verbigeration,Waxy flexibility | 95.1 | 92.8 | 97.1 |
| 7 | Excitement,Immobility/stupor,Grimacing,Echopraxia/echolalia,Mannerisms,Verbigeration,Waxy flexibility | 95.0 | 93.0 | 96.9 |
| 7 | Excitement,Immobility/stupor,Grimacing,Echopraxia/echolalia,Stereotypy,Verbigeration,Waxy flexibility | 95.0 | 93.0 | 96.9 |
| 7 | Immobility/stupor,Mutism,Grimacing,Echopraxia/echolalia,Stereotypy,Rigidity,Waxy flexibility | 95.0 | 93.0 | 97.1 |
| 7 | Immobility/stupor,Mutism,Posturing/catalepsy,Echopraxia/echolalia,Mannerisms,Rigidity,Waxy flexibility | 95.0 | 93.0 | 96.9 |
| 7 | Immobility/stupor,Mutism,Posturing/catalepsy,Grimacing,Echopraxia/echolalia,Verbigeration,Withdrawal | 95.0 | 93.0 | 96.9 |
| 7 | Immobility/stupor,Mutism,Posturing/catalepsy,Grimacing,Verbigeration,Rigidity,Waxy flexibility | 95.0 | 92.8 | 96.9 |
| 7 | Excitement,Immobility/stupor,Posturing/catalepsy,Stereotypy,Mannerisms,Verbigeration,Waxy flexibility | 94.9 | 92.6 | 96.9 |
| 7 | Excitement,Mutism,Posturing/catalepsy,Grimacing,Mannerisms,Rigidity,Negativism | 94.9 | 92.6 | 96.9 |
| 7 | Excitement,Mutism,Posturing/catalepsy,Grimacing,Mannerisms,Rigidity,Withdrawal | 94.9 | 92.6 | 96.9 |
| 7 | Excitement,Mutism,Posturing/catalepsy,Stereotypy,Verbigeration,Negativism,Withdrawal | 94.9 | 92.8 | 96.9 |
| 7 | Immobility/stupor,Mutism,Grimacing,Echopraxia/echolalia,Stereotypy,Verbigeration,Withdrawal | 94.9 | 92.6 | 96.9 |
| 7 | Immobility/stupor,Mutism,Posturing/catalepsy,Echopraxia/echolalia,Mannerisms,Waxy flexibility,Withdrawal | 94.9 | 92.8 | 96.9 |
| 7 | Immobility/stupor,Mutism,Posturing/catalepsy,Echopraxia/echolalia,Rigidity,Waxy flexibility,Withdrawal | 94.9 | 92.8 | 96.9 |
| 7 | Immobility/stupor,Posturing/catalepsy,Echopraxia/echolalia,Stereotypy,Rigidity,Waxy flexibility,Withdrawal | 94.9 | 92.8 | 96.9 |
| 7 | Immobility/stupor,Posturing/catalepsy,Grimacing,Echopraxia/echolalia,Mannerisms,Rigidity,Waxy flexibility | 94.9 | 92.6 | 96.9 |
| 7 | Immobility/stupor,Posturing/catalepsy,Stereotypy,Verbigeration,Rigidity,Waxy flexibility,Withdrawal | 94.9 | 92.6 | 96.9 |
| 7 | Mutism,Echopraxia/echolalia,Mannerisms,Rigidity,Negativism,Waxy flexibility,Withdrawal | 94.9 | 92.6 | 96.9 |
| 7 | Mutism,Echopraxia/echolalia,Stereotypy,Verbigeration,Rigidity,Negativism,Withdrawal | 94.9 | 92.8 | 96.6 |
| 7 | Mutism,Posturing/catalepsy,Stereotypy,Mannerisms,Verbigeration,Rigidity,Negativism | 94.9 | 92.8 | 96.9 |
| 7 | Staring,Posturing/catalepsy,Grimacing,Echopraxia/echolalia,Verbigeration,Rigidity,Waxy flexibility | 94.9 | 92.8 | 96.9 |
| 7 | Excitement,Immobility/stupor,Echopraxia/echolalia,Stereotypy,Mannerisms,Verbigeration,Waxy flexibility | 94.8 | 92.6 | 96.9 |
| 7 | Excitement,Immobility/stupor,Mutism,Grimacing,Echopraxia/echolalia,Mannerisms,Verbigeration | 94.8 | 92.8 | 96.9 |
| 7 | Excitement,Immobility/stupor,Mutism,Grimacing,Echopraxia/echolalia,Stereotypy,Mannerisms | 94.8 | 92.8 | 96.9 |
| 7 | Excitement,Immobility/stupor,Mutism,Grimacing,Stereotypy,Mannerisms,Verbigeration | 94.8 | 92.8 | 96.9 |
| 7 | Excitement,Immobility/stupor,Posturing/catalepsy,Grimacing,Stereotypy,Mannerisms,Waxy flexibility | 94.8 | 92.6 | 96.9 |
| 7 | Excitement,Mutism,Echopraxia/echolalia,Stereotypy,Rigidity,Waxy flexibility,Withdrawal | 94.8 | 92.8 | 96.6 |
| 7 | Excitement,Mutism,Posturing/catalepsy,Echopraxia/echolalia,Mannerisms,Rigidity,Withdrawal | 94.8 | 92.8 | 96.9 |
| 7 | Excitement,Mutism,Posturing/catalepsy,Echopraxia/echolalia,Stereotypy,Rigidity,Withdrawal | 94.8 | 92.6 | 96.9 |
| 7 | Immobility/stupor,Grimacing,Echopraxia/echolalia,Mannerisms,Verbigeration,Rigidity,Waxy flexibility | 94.8 | 92.6 | 96.9 |
| 7 | Immobility/stupor,Grimacing,Echopraxia/echolalia,Stereotypy,Verbigeration,Rigidity,Waxy flexibility | 94.8 | 92.8 | 96.9 |
| 7 | Immobility/stupor,Mutism,Echopraxia/echolalia,Stereotypy,Verbigeration,Rigidity,Waxy flexibility | 94.8 | 92.6 | 96.9 |
| 7 | Immobility/stupor,Mutism,Grimacing,Echopraxia/echolalia,Mannerisms,Verbigeration,Withdrawal | 94.8 | 92.8 | 96.9 |
| 7 | Immobility/stupor,Mutism,Grimacing,Echopraxia/echolalia,Stereotypy,Waxy flexibility,Withdrawal | 94.8 | 92.8 | 96.9 |
| 7 | Immobility/stupor,Mutism,Posturing/catalepsy,Grimacing,Echopraxia/echolalia,Waxy flexibility,Withdrawal | 94.8 | 92.8 | 96.9 |
| 7 | Immobility/stupor,Mutism,Posturing/catalepsy,Grimacing,Rigidity,Waxy flexibility,Withdrawal | 94.8 | 92.6 | 96.9 |
| 7 | Immobility/stupor,Mutism,Posturing/catalepsy,Grimacing,Stereotypy,Rigidity,Waxy flexibility | 94.8 | 92.6 | 96.9 |
| 7 | Immobility/stupor,Mutism,Posturing/catalepsy,Stereotypy,Mannerisms,Rigidity,Waxy flexibility | 94.8 | 92.8 | 96.9 |
| 7 | Immobility/stupor,Mutism,Staring,Posturing/catalepsy,Grimacing,Rigidity,Waxy flexibility | 94.8 | 92.6 | 96.9 |
| 7 | Immobility/stupor,Posturing/catalepsy,Grimacing,Echopraxia/echolalia,Mannerisms,Verbigeration,Withdrawal | 94.8 | 92.6 | 96.9 |
| 7 | Immobility/stupor,Posturing/catalepsy,Grimacing,Stereotypy,Mannerisms,Rigidity,Waxy flexibility | 94.8 | 92.8 | 96.9 |
| 7 | Immobility/stupor,Posturing/catalepsy,Grimacing,Stereotypy,Verbigeration,Rigidity,Waxy flexibility | 94.8 | 92.6 | 96.9 |
| 7 | Immobility/stupor,Posturing/catalepsy,Grimacing,Verbigeration,Rigidity,Waxy flexibility,Withdrawal | 94.8 | 92.6 | 96.9 |
| 7 | Immobility/stupor,Staring,Posturing/catalepsy,Echopraxia/echolalia,Verbigeration,Waxy flexibility,Withdrawal | 94.8 | 92.6 | 96.6 |
| 7 | Mutism,Echopraxia/echolalia,Mannerisms,Verbigeration,Rigidity,Negativism,Withdrawal | 94.8 | 92.6 | 96.9 |
| 7 | Mutism,Posturing/catalepsy,Echopraxia/echolalia,Mannerisms,Verbigeration,Rigidity,Negativism | 94.8 | 92.6 | 96.9 |
| 7 | Mutism,Posturing/catalepsy,Grimacing,Echopraxia/echolalia,Mannerisms,Rigidity,Negativism | 94.8 | 92.6 | 96.9 |
| 7 | Staring,Posturing/catalepsy,Echopraxia/echolalia,Stereotypy,Verbigeration,Waxy flexibility,Withdrawal | 94.8 | 92.6 | 96.6 |
| 7 | Excitement,Immobility/stupor,Posturing/catalepsy,Echopraxia/echolalia,Stereotypy,Mannerisms,Waxy flexibility | 94.6 | 92.4 | 96.6 |
| 7 | Excitement,Mutism,Echopraxia/echolalia,Stereotypy,Rigidity,Negativism,Waxy flexibility | 94.6 | 92.4 | 96.6 |
| 7 | Excitement,Mutism,Echopraxia/echolalia,Stereotypy,Verbigeration,Rigidity,Negativism | 94.6 | 92.4 | 96.6 |
| 7 | Excitement,Mutism,Grimacing,Echopraxia/echolalia,Rigidity,Waxy flexibility,Withdrawal | 94.6 | 92.4 | 96.6 |
| 7 | Excitement,Mutism,Grimacing,Stereotypy,Rigidity,Waxy flexibility,Withdrawal | 94.6 | 92.4 | 96.6 |
| 7 | Excitement,Mutism,Grimacing,Verbigeration,Rigidity,Waxy flexibility,Withdrawal | 94.6 | 92.4 | 96.6 |
| 7 | Excitement,Mutism,Posturing/catalepsy,Echopraxia/echolalia,Verbigeration,Negativism,Withdrawal | 94.6 | 92.4 | 96.6 |
| 7 | Excitement,Mutism,Posturing/catalepsy,Echopraxia/echolalia,Verbigeration,Rigidity,Waxy flexibility | 94.6 | 92.4 | 96.6 |
| 7 | Excitement,Mutism,Posturing/catalepsy,Grimacing,Rigidity,Negativism,Waxy flexibility | 94.6 | 92.4 | 96.6 |
| 7 | Excitement,Mutism,Posturing/catalepsy,Grimacing,Stereotypy,Rigidity,Negativism | 94.6 | 92.4 | 96.6 |
| 7 | Excitement,Mutism,Posturing/catalepsy,Grimacing,Stereotypy,Rigidity,Withdrawal | 94.6 | 92.4 | 96.6 |
| 7 | Excitement,Mutism,Posturing/catalepsy,Mannerisms,Verbigeration,Negativism,Withdrawal | 94.6 | 92.4 | 96.6 |
| 7 | Excitement,Mutism,Posturing/catalepsy,Stereotypy,Mannerisms,Rigidity,Withdrawal | 94.6 | 92.4 | 96.6 |
| 7 | Excitement,Mutism,Stereotypy,Mannerisms,Rigidity,Waxy flexibility,Withdrawal | 94.6 | 92.4 | 96.6 |
| 7 | Immobility/stupor,Mutism,Grimacing,Stereotypy,Mannerisms,Verbigeration,Withdrawal | 94.6 | 92.4 | 96.6 |
| 7 | Immobility/stupor,Mutism,Grimacing,Stereotypy,Mannerisms,Waxy flexibility,Withdrawal | 94.6 | 92.4 | 96.6 |
| 7 | Immobility/stupor,Mutism,Posturing/catalepsy,Echopraxia/echolalia,Stereotypy,Rigidity,Waxy flexibility | 94.6 | 92.4 | 96.6 |
| 7 | Immobility/stupor,Mutism,Posturing/catalepsy,Echopraxia/echolalia,Verbigeration,Rigidity,Waxy flexibility | 94.6 | 92.4 | 96.6 |
| 7 | Immobility/stupor,Mutism,Posturing/catalepsy,Grimacing,Echopraxia/echolalia,Stereotypy,Mannerisms | 94.6 | 92.4 | 96.6 |
| 7 | Immobility/stupor,Mutism,Posturing/catalepsy,Grimacing,Echopraxia/echolalia,Stereotypy,Verbigeration | 94.6 | 92.4 | 96.6 |
| 7 | Immobility/stupor,Mutism,Posturing/catalepsy,Stereotypy,Mannerisms,Verbigeration,Withdrawal | 94.6 | 92.4 | 96.6 |
| 7 | Immobility/stupor,Posturing/catalepsy,Echopraxia/echolalia,Stereotypy,Mannerisms,Verbigeration,Rigidity | 94.6 | 92.4 | 96.6 |
| 7 | Immobility/stupor,Posturing/catalepsy,Grimacing,Echopraxia/echolalia,Mannerisms,Waxy flexibility,Withdrawal | 94.6 | 92.4 | 96.6 |
| 7 | Immobility/stupor,Posturing/catalepsy,Grimacing,Mannerisms,Verbigeration,Rigidity,Waxy flexibility | 94.6 | 92.4 | 96.6 |
| 7 | Immobility/stupor,Posturing/catalepsy,Grimacing,Stereotypy,Mannerisms,Verbigeration,Withdrawal | 94.6 | 92.4 | 96.6 |
| 7 | Mutism,Posturing/catalepsy,Echopraxia/echolalia,Stereotypy,Rigidity,Negativism,Withdrawal | 94.6 | 92.4 | 96.6 |
| 7 | Mutism,Posturing/catalepsy,Echopraxia/echolalia,Verbigeration,Rigidity,Negativism,Withdrawal | 94.6 | 92.4 | 96.6 |
| 7 | Mutism,Posturing/catalepsy,Grimacing,Mannerisms,Verbigeration,Rigidity,Negativism | 94.6 | 92.4 | 96.6 |
| 7 | Mutism,Posturing/catalepsy,Grimacing,Stereotypy,Rigidity,Negativism,Withdrawal | 94.6 | 92.4 | 96.6 |
| 7 | Mutism,Posturing/catalepsy,Grimacing,Verbigeration,Rigidity,Negativism,Withdrawal | 94.6 | 92.4 | 96.6 |
| 7 | Mutism,Posturing/catalepsy,Verbigeration,Rigidity,Negativism,Waxy flexibility,Withdrawal | 94.6 | 92.4 | 96.6 |
| 7 | Mutism,Stereotypy,Mannerisms,Verbigeration,Rigidity,Negativism,Waxy flexibility | 94.6 | 92.4 | 96.6 |
| 7 | Staring,Posturing/catalepsy,Echopraxia/echolalia,Stereotypy,Verbigeration,Rigidity,Waxy flexibility | 94.6 | 92.4 | 96.6 |
| 7 | Staring,Posturing/catalepsy,Grimacing,Echopraxia/echolalia,Mannerisms,Verbigeration,Waxy flexibility | 94.6 | 92.4 | 96.6 |
| 7 | Excitement,Immobility/stupor,Grimacing,Stereotypy,Mannerisms,Verbigeration,Waxy flexibility | 94.4 | 92.2 | 96.4 |
| 7 | Excitement,Mutism,Echopraxia/echolalia,Verbigeration,Rigidity,Waxy flexibility,Withdrawal | 94.4 | 92.2 | 96.4 |
| 7 | Excitement,Mutism,Grimacing,Echopraxia/echolalia,Rigidity,Negativism,Waxy flexibility | 94.4 | 92.2 | 96.4 |
| 7 | Excitement,Mutism,Grimacing,Mannerisms,Rigidity,Waxy flexibility,Withdrawal | 94.4 | 92.2 | 96.4 |
| 7 | Excitement,Mutism,Grimacing,Stereotypy,Verbigeration,Rigidity,Negativism | 94.4 | 92.2 | 96.4 |
| 7 | Excitement,Mutism,Mannerisms,Verbigeration,Rigidity,Waxy flexibility,Withdrawal | 94.4 | 92.2 | 96.4 |
| 7 | Excitement,Mutism,Posturing/catalepsy,Echopraxia/echolalia,Stereotypy,Negativism,Withdrawal | 94.4 | 92.2 | 96.4 |
| 7 | Excitement,Mutism,Posturing/catalepsy,Grimacing,Echopraxia/echolalia,Negativism,Withdrawal | 94.4 | 92.2 | 96.4 |
| 7 | Excitement,Mutism,Posturing/catalepsy,Grimacing,Echopraxia/echolalia,Rigidity,Waxy flexibility | 94.4 | 92.2 | 96.4 |
| 7 | Excitement,Mutism,Posturing/catalepsy,Grimacing,Verbigeration,Negativism,Withdrawal | 94.4 | 92.2 | 96.4 |
| 7 | Excitement,Mutism,Posturing/catalepsy,Grimacing,Verbigeration,Rigidity,Waxy flexibility | 94.4 | 92.2 | 96.4 |
| 7 | Excitement,Mutism,Posturing/catalepsy,Mannerisms,Rigidity,Negativism,Waxy flexibility | 94.4 | 92.2 | 96.4 |
| 7 | Excitement,Mutism,Posturing/catalepsy,Stereotypy,Mannerisms,Rigidity,Negativism | 94.4 | 92.2 | 96.4 |
| 7 | Excitement,Mutism,Posturing/catalepsy,Stereotypy,Rigidity,Negativism,Waxy flexibility | 94.4 | 92.2 | 96.4 |
| 7 | Excitement,Mutism,Posturing/catalepsy,Verbigeration,Negativism,Waxy flexibility,Withdrawal | 94.4 | 92.2 | 96.4 |
| 7 | Excitement,Posturing/catalepsy,Echopraxia/echolalia,Verbigeration,Rigidity,Negativism,Withdrawal | 94.4 | 92.2 | 96.4 |
| 7 | Excitement,Posturing/catalepsy,Mannerisms,Verbigeration,Rigidity,Negativism,Withdrawal | 94.4 | 92.2 | 96.4 |
| 7 | Excitement,Posturing/catalepsy,Stereotypy,Verbigeration,Rigidity,Negativism,Withdrawal | 94.4 | 92.2 | 96.4 |
| 7 | Immobility/stupor,Echopraxia/echolalia,Stereotypy,Mannerisms,Verbigeration,Rigidity,Waxy flexibility | 94.4 | 92.2 | 96.4 |
| 7 | Immobility/stupor,Grimacing,Echopraxia/echolalia,Stereotypy,Mannerisms,Verbigeration,Withdrawal | 94.4 | 92.2 | 96.4 |
| 7 | Immobility/stupor,Mutism,Echopraxia/echolalia,Stereotypy,Mannerisms,Waxy flexibility,Withdrawal | 94.4 | 92.2 | 96.4 |
| 7 | Immobility/stupor,Mutism,Grimacing,Echopraxia/echolalia,Verbigeration,Waxy flexibility,Withdrawal | 94.4 | 92.2 | 96.4 |
| 7 | Immobility/stupor,Mutism,Grimacing,Mannerisms,Verbigeration,Waxy flexibility,Withdrawal | 94.4 | 92.2 | 96.4 |
| 7 | Immobility/stupor,Mutism,Posturing/catalepsy,Echopraxia/echolalia,Stereotypy,Verbigeration,Withdrawal | 94.4 | 92.2 | 96.4 |
| 7 | Immobility/stupor,Mutism,Posturing/catalepsy,Grimacing,Echopraxia/echolalia,Mannerisms,Waxy flexibility | 94.4 | 92.2 | 96.4 |
| 7 | Immobility/stupor,Mutism,Posturing/catalepsy,Grimacing,Echopraxia/echolalia,Rigidity,Waxy flexibility | 94.4 | 92.2 | 96.4 |
| 7 | Immobility/stupor,Mutism,Posturing/catalepsy,Grimacing,Echopraxia/echolalia,Verbigeration,Waxy flexibility | 94.4 | 92.2 | 96.4 |
| 7 | Immobility/stupor,Mutism,Posturing/catalepsy,Grimacing,Stereotypy,Mannerisms,Verbigeration | 94.4 | 92.2 | 96.4 |
| 7 | Immobility/stupor,Mutism,Posturing/catalepsy,Grimacing,Stereotypy,Waxy flexibility,Withdrawal | 94.4 | 92.2 | 96.4 |
| 7 | Immobility/stupor,Mutism,Posturing/catalepsy,Grimacing,Verbigeration,Waxy flexibility,Withdrawal | 94.4 | 92.2 | 96.4 |
| 7 | Immobility/stupor,Mutism,Posturing/catalepsy,Mannerisms,Verbigeration,Waxy flexibility,Withdrawal | 94.4 | 92.2 | 96.4 |
| 7 | Immobility/stupor,Mutism,Posturing/catalepsy,Stereotypy,Mannerisms,Waxy flexibility,Withdrawal | 94.4 | 92.2 | 96.4 |
| 7 | Immobility/stupor,Posturing/catalepsy,Echopraxia/echolalia,Verbigeration,Rigidity,Waxy flexibility,Withdrawal | 94.4 | 92.2 | 96.4 |
| 7 | Immobility/stupor,Posturing/catalepsy,Grimacing,Echopraxia/echolalia,Stereotypy,Rigidity,Waxy flexibility | 94.4 | 92.2 | 96.4 |
| 7 | Immobility/stupor,Posturing/catalepsy,Grimacing,Echopraxia/echolalia,Stereotypy,Verbigeration,Withdrawal | 94.4 | 92.2 | 96.4 |
| 7 | Immobility/stupor,Posturing/catalepsy,Grimacing,Echopraxia/echolalia,Stereotypy,Waxy flexibility,Withdrawal | 94.4 | 92.2 | 96.4 |
| 7 | Immobility/stupor,Posturing/catalepsy,Grimacing,Echopraxia/echolalia,Verbigeration,Rigidity,Waxy flexibility | 94.4 | 92.2 | 96.4 |
| 7 | Immobility/stupor,Posturing/catalepsy,Grimacing,Stereotypy,Mannerisms,Waxy flexibility,Withdrawal | 94.4 | 92.2 | 96.4 |
| 7 | Mutism,Echopraxia/echolalia,Stereotypy,Rigidity,Negativism,Waxy flexibility,Withdrawal | 94.4 | 92.2 | 96.4 |
| 7 | Mutism,Grimacing,Echopraxia/echolalia,Stereotypy,Rigidity,Negativism,Withdrawal | 94.4 | 92.2 | 96.4 |
| 7 | Mutism,Grimacing,Stereotypy,Rigidity,Negativism,Waxy flexibility,Withdrawal | 94.4 | 92.2 | 96.4 |
| 7 | Mutism,Grimacing,Verbigeration,Rigidity,Negativism,Waxy flexibility,Withdrawal | 94.4 | 92.2 | 96.4 |
| 7 | Mutism,Posturing/catalepsy,Echopraxia/echolalia,Mannerisms,Rigidity,Negativism,Waxy flexibility | 94.4 | 92.2 | 96.4 |
| 7 | Mutism,Posturing/catalepsy,Echopraxia/echolalia,Stereotypy,Mannerisms,Rigidity,Negativism | 94.4 | 92.2 | 96.4 |
| 7 | Mutism,Posturing/catalepsy,Grimacing,Echopraxia/echolalia,Rigidity,Negativism,Withdrawal | 94.4 | 92.2 | 96.4 |
| 7 | Mutism,Posturing/catalepsy,Grimacing,Stereotypy,Verbigeration,Rigidity,Negativism | 94.4 | 92.2 | 96.4 |
| 7 | Mutism,Posturing/catalepsy,Mannerisms,Verbigeration,Rigidity,Negativism,Waxy flexibility | 94.4 | 92.2 | 96.4 |
| 7 | Mutism,Posturing/catalepsy,Stereotypy,Rigidity,Negativism,Waxy flexibility,Withdrawal | 94.4 | 92.2 | 96.4 |
| 7 | Posturing/catalepsy,Stereotypy,Mannerisms,Verbigeration,Rigidity,Negativism,Withdrawal | 94.4 | 92.2 | 96.4 |
| 7 | Excitement,Mutism,Echopraxia/echolalia,Verbigeration,Rigidity,Negativism,Waxy flexibility | 94.2 | 91.9 | 96.2 |
| 7 | Excitement,Mutism,Grimacing,Echopraxia/echolalia,Stereotypy,Rigidity,Negativism | 94.2 | 91.9 | 96.2 |
| 7 | Excitement,Mutism,Grimacing,Echopraxia/echolalia,Verbigeration,Rigidity,Negativism | 94.2 | 91.9 | 96.2 |
| 7 | Excitement,Mutism,Grimacing,Stereotypy,Rigidity,Negativism,Waxy flexibility | 94.2 | 91.9 | 96.2 |
| 7 | Excitement,Mutism,Grimacing,Verbigeration,Rigidity,Negativism,Waxy flexibility | 94.2 | 91.9 | 96.2 |
| 7 | Excitement,Mutism,Posturing/catalepsy,Echopraxia/echolalia,Mannerisms,Negativism,Withdrawal | 94.2 | 91.9 | 96.2 |
| 7 | Excitement,Mutism,Posturing/catalepsy,Echopraxia/echolalia,Negativism,Waxy flexibility,Withdrawal | 94.2 | 91.9 | 96.2 |
| 7 | Excitement,Mutism,Posturing/catalepsy,Grimacing,Echopraxia/echolalia,Verbigeration,Rigidity | 94.2 | 91.9 | 96.2 |
| 7 | Excitement,Mutism,Posturing/catalepsy,Grimacing,Mannerisms,Negativism,Withdrawal | 94.2 | 91.9 | 96.2 |
| 7 | Excitement,Mutism,Posturing/catalepsy,Grimacing,Stereotypy,Negativism,Withdrawal | 94.2 | 91.9 | 96.2 |
| 7 | Excitement,Mutism,Posturing/catalepsy,Mannerisms,Verbigeration,Rigidity,Waxy flexibility | 94.2 | 91.9 | 96.2 |
| 7 | Excitement,Mutism,Posturing/catalepsy,Stereotypy,Mannerisms,Negativism,Withdrawal | 94.2 | 91.9 | 96.2 |
| 7 | Excitement,Mutism,Posturing/catalepsy,Stereotypy,Negativism,Waxy flexibility,Withdrawal | 94.2 | 91.9 | 96.2 |
| 7 | Excitement,Mutism,Posturing/catalepsy,Stereotypy,Verbigeration,Rigidity,Waxy flexibility | 94.2 | 91.9 | 96.2 |
| 7 | Excitement,Mutism,Stereotypy,Mannerisms,Verbigeration,Rigidity,Negativism | 94.2 | 91.9 | 96.2 |
| 7 | Immobility/stupor,Mutism,Echopraxia/echolalia,Stereotypy,Mannerisms,Verbigeration,Withdrawal | 94.2 | 91.9 | 96.2 |
| 7 | Immobility/stupor,Mutism,Grimacing,Echopraxia/echolalia,Mannerisms,Verbigeration,Waxy flexibility | 94.2 | 91.9 | 96.2 |
| 7 | Immobility/stupor,Mutism,Grimacing,Echopraxia/echolalia,Stereotypy,Mannerisms,Waxy flexibility | 94.2 | 91.9 | 96.2 |
| 7 | Immobility/stupor,Mutism,Grimacing,Stereotypy,Verbigeration,Waxy flexibility,Withdrawal | 94.2 | 91.9 | 96.2 |
| 7 | Immobility/stupor,Mutism,Posturing/catalepsy,Echopraxia/echolalia,Stereotypy,Waxy flexibility,Withdrawal | 94.2 | 91.9 | 96.2 |
| 7 | Immobility/stupor,Mutism,Posturing/catalepsy,Grimacing,Mannerisms,Verbigeration,Waxy flexibility | 94.2 | 91.9 | 96.2 |
| 7 | Immobility/stupor,Mutism,Posturing/catalepsy,Grimacing,Stereotypy,Verbigeration,Waxy flexibility | 94.2 | 91.9 | 96.2 |
| 7 | Immobility/stupor,Posturing/catalepsy,Echopraxia/echolalia,Stereotypy,Mannerisms,Verbigeration,Withdrawal | 94.2 | 91.9 | 96.2 |
| 7 | Immobility/stupor,Posturing/catalepsy,Stereotypy,Mannerisms,Verbigeration,Rigidity,Waxy flexibility | 94.2 | 91.9 | 96.2 |
| 7 | Mutism,Echopraxia/echolalia,Stereotypy,Mannerisms,Rigidity,Negativism,Waxy flexibility | 94.2 | 91.9 | 96.4 |
| 7 | Mutism,Echopraxia/echolalia,Verbigeration,Rigidity,Negativism,Waxy flexibility,Withdrawal | 94.2 | 91.9 | 96.2 |
| 7 | Mutism,Grimacing,Echopraxia/echolalia,Verbigeration,Rigidity,Negativism,Withdrawal | 94.2 | 91.9 | 96.2 |
| 7 | Mutism,Posturing/catalepsy,Echopraxia/echolalia,Rigidity,Negativism,Waxy flexibility,Withdrawal | 94.2 | 91.9 | 96.2 |
| 7 | Mutism,Posturing/catalepsy,Echopraxia/echolalia,Stereotypy,Verbigeration,Rigidity,Negativism | 94.2 | 91.9 | 96.2 |
| 7 | Mutism,Posturing/catalepsy,Grimacing,Echopraxia/echolalia,Verbigeration,Rigidity,Negativism | 94.2 | 91.9 | 96.2 |
| 7 | Mutism,Posturing/catalepsy,Grimacing,Stereotypy,Mannerisms,Rigidity,Negativism | 94.2 | 91.9 | 96.2 |
| 7 | Mutism,Posturing/catalepsy,Stereotypy,Verbigeration,Rigidity,Negativism,Waxy flexibility | 94.2 | 91.9 | 96.2 |
| 7 | Staring,Posturing/catalepsy,Grimacing,Echopraxia/echolalia,Stereotypy,Verbigeration,Waxy flexibility | 94.2 | 91.9 | 96.2 |
| 7 | Excitement,Immobility/stupor,Grimacing,Echopraxia/echolalia,Stereotypy,Mannerisms,Verbigeration | 94.0 | 91.7 | 96.2 |
| 7 | Excitement,Mutism,Echopraxia/echolalia,Stereotypy,Mannerisms,Rigidity,Negativism | 94.0 | 91.7 | 96.0 |
| 7 | Excitement,Mutism,Posturing/catalepsy,Grimacing,Mannerisms,Verbigeration,Rigidity | 94.0 | 91.7 | 96.0 |
| 7 | Excitement,Mutism,Posturing/catalepsy,Mannerisms,Negativism,Waxy flexibility,Withdrawal | 94.0 | 91.7 | 96.0 |
| 7 | Excitement,Posturing/catalepsy,Echopraxia/echolalia,Mannerisms,Rigidity,Negativism,Withdrawal | 94.0 | 91.7 | 96.2 |
| 7 | Excitement,Posturing/catalepsy,Echopraxia/echolalia,Stereotypy,Rigidity,Negativism,Withdrawal | 94.0 | 91.7 | 96.0 |
| 7 | Excitement,Posturing/catalepsy,Grimacing,Echopraxia/echolalia,Rigidity,Negativism,Withdrawal | 94.0 | 91.7 | 96.2 |
| 7 | Excitement,Posturing/catalepsy,Verbigeration,Rigidity,Negativism,Waxy flexibility,Withdrawal | 94.0 | 91.7 | 96.0 |
| 7 | Immobility/stupor,Grimacing,Echopraxia/echolalia,Stereotypy,Verbigeration,Waxy flexibility,Withdrawal | 94.0 | 91.7 | 96.2 |
| 7 | Immobility/stupor,Mutism,Echopraxia/echolalia,Mannerisms,Verbigeration,Waxy flexibility,Withdrawal | 94.0 | 91.7 | 96.2 |
| 7 | Immobility/stupor,Mutism,Posturing/catalepsy,Echopraxia/echolalia,Verbigeration,Waxy flexibility,Withdrawal | 94.0 | 91.7 | 96.0 |
| 7 | Immobility/stupor,Mutism,Posturing/catalepsy,Stereotypy,Verbigeration,Waxy flexibility,Withdrawal | 94.0 | 91.7 | 96.0 |
| 7 | Immobility/stupor,Posturing/catalepsy,Echopraxia/echolalia,Stereotypy,Mannerisms,Rigidity,Waxy flexibility | 94.0 | 91.7 | 96.2 |
| 7 | Immobility/stupor,Posturing/catalepsy,Echopraxia/echolalia,Stereotypy,Mannerisms,Waxy flexibility,Withdrawal | 94.0 | 91.7 | 96.2 |
| 7 | Mutism,Grimacing,Echopraxia/echolalia,Rigidity,Negativism,Waxy flexibility,Withdrawal | 94.0 | 91.7 | 96.2 |
| 7 | Mutism,Grimacing,Stereotypy,Mannerisms,Rigidity,Negativism,Waxy flexibility | 94.0 | 91.7 | 96.0 |
| 7 | Mutism,Posturing/catalepsy,Grimacing,Mannerisms,Rigidity,Negativism,Waxy flexibility | 94.0 | 91.7 | 96.0 |
| 7 | Mutism,Posturing/catalepsy,Grimacing,Mannerisms,Rigidity,Waxy flexibility,Withdrawal | 94.0 | 91.7 | 96.0 |
| 7 | Mutism,Posturing/catalepsy,Stereotypy,Mannerisms,Rigidity,Negativism,Waxy flexibility | 94.0 | 91.7 | 96.0 |
| 7 | Posturing/catalepsy,Echopraxia/echolalia,Mannerisms,Verbigeration,Rigidity,Negativism,Withdrawal | 94.0 | 91.7 | 96.2 |
| 7 | Posturing/catalepsy,Echopraxia/echolalia,Stereotypy,Mannerisms,Rigidity,Negativism,Withdrawal | 94.0 | 91.7 | 96.0 |
| 7 | Posturing/catalepsy,Grimacing,Stereotypy,Mannerisms,Rigidity,Negativism,Withdrawal | 94.0 | 91.7 | 96.0 |
| 7 | Posturing/catalepsy,Stereotypy,Mannerisms,Rigidity,Negativism,Waxy flexibility,Withdrawal | 94.0 | 91.7 | 96.0 |
| 7 | Staring,Posturing/catalepsy,Echopraxia/echolalia,Stereotypy,Mannerisms,Verbigeration,Waxy flexibility | 94.0 | 91.7 | 96.0 |
| 7 | Excitement,Mutism,Echopraxia/echolalia,Mannerisms,Rigidity,Waxy flexibility,Withdrawal | 93.9 | 91.7 | 96.0 |
| 7 | Excitement,Mutism,Posturing/catalepsy,Echopraxia/echolalia,Mannerisms,Rigidity,Waxy flexibility | 93.9 | 91.7 | 96.0 |
| 7 | Excitement,Mutism,Posturing/catalepsy,Echopraxia/echolalia,Mannerisms,Verbigeration,Rigidity | 93.9 | 91.7 | 96.0 |
| 7 | Excitement,Mutism,Posturing/catalepsy,Echopraxia/echolalia,Stereotypy,Rigidity,Waxy flexibility | 93.9 | 91.7 | 96.0 |
| 7 | Excitement,Mutism,Posturing/catalepsy,Echopraxia/echolalia,Stereotypy,Verbigeration,Rigidity | 93.9 | 91.7 | 96.0 |
| 7 | Excitement,Mutism,Posturing/catalepsy,Grimacing,Negativism,Waxy flexibility,Withdrawal | 93.9 | 91.7 | 96.0 |
| 7 | Excitement,Mutism,Stereotypy,Mannerisms,Rigidity,Negativism,Waxy flexibility | 93.9 | 91.7 | 96.0 |
| 7 | Excitement,Posturing/catalepsy,Grimacing,Verbigeration,Rigidity,Negativism,Withdrawal | 93.9 | 91.7 | 96.0 |
| 7 | Excitement,Posturing/catalepsy,Stereotypy,Mannerisms,Rigidity,Negativism,Withdrawal | 93.9 | 91.7 | 96.0 |
| 7 | Immobility/stupor,Mutism,Grimacing,Echopraxia/echolalia,Stereotypy,Verbigeration,Waxy flexibility | 93.9 | 91.7 | 96.0 |
| 7 | Immobility/stupor,Mutism,Posturing/catalepsy,Echopraxia/echolalia,Stereotypy,Mannerisms,Verbigeration | 93.9 | 91.7 | 96.0 |
| 7 | Immobility/stupor,Mutism,Posturing/catalepsy,Grimacing,Echopraxia/echolalia,Stereotypy,Waxy flexibility | 93.9 | 91.7 | 96.0 |
| 7 | Immobility/stupor,Posturing/catalepsy,Echopraxia/echolalia,Mannerisms,Verbigeration,Rigidity,Waxy flexibility | 93.9 | 91.7 | 96.0 |
| 7 | Immobility/stupor,Posturing/catalepsy,Echopraxia/echolalia,Stereotypy,Verbigeration,Rigidity,Waxy flexibility | 93.9 | 91.7 | 96.0 |
| 7 | Immobility/stupor,Posturing/catalepsy,Grimacing,Mannerisms,Verbigeration,Waxy flexibility,Withdrawal | 93.9 | 91.7 | 96.2 |
| 7 | Mutism,Echopraxia/echolalia,Stereotypy,Mannerisms,Verbigeration,Rigidity,Negativism | 93.9 | 91.7 | 96.0 |
| 7 | Mutism,Grimacing,Echopraxia/echolalia,Mannerisms,Rigidity,Negativism,Waxy flexibility | 93.9 | 91.7 | 96.0 |
| 7 | Mutism,Grimacing,Stereotypy,Mannerisms,Verbigeration,Rigidity,Negativism | 93.9 | 91.7 | 96.0 |
| 7 | Mutism,Grimacing,Stereotypy,Verbigeration,Rigidity,Negativism,Waxy flexibility | 93.9 | 91.7 | 96.2 |
| 7 | Mutism,Posturing/catalepsy,Echopraxia/echolalia,Verbigeration,Rigidity,Negativism,Waxy flexibility | 93.9 | 91.7 | 96.0 |
| 7 | Mutism,Posturing/catalepsy,Grimacing,Echopraxia/echolalia,Mannerisms,Negativism,Withdrawal | 93.9 | 91.7 | 96.0 |
| 7 | Mutism,Posturing/catalepsy,Grimacing,Rigidity,Negativism,Waxy flexibility,Withdrawal | 93.9 | 91.7 | 96.2 |
| 7 | Mutism,Posturing/catalepsy,Stereotypy,Mannerisms,Verbigeration,Negativism,Withdrawal | 93.9 | 91.7 | 96.0 |
| 7 | Excitement,Mutism,Echopraxia/echolalia,Stereotypy,Verbigeration,Rigidity,Withdrawal | 93.7 | 91.3 | 96.0 |
| 7 | Excitement,Mutism,Grimacing,Echopraxia/echolalia,Mannerisms,Rigidity,Negativism | 93.7 | 91.3 | 95.7 |
| 7 | Excitement,Mutism,Grimacing,Mannerisms,Verbigeration,Rigidity,Negativism | 93.7 | 91.5 | 95.7 |
| 7 | Excitement,Mutism,Mannerisms,Verbigeration,Rigidity,Negativism,Waxy flexibility | 93.7 | 91.5 | 96.0 |
| 7 | Excitement,Mutism,Posturing/catalepsy,Grimacing,Echopraxia/echolalia,Mannerisms,Rigidity | 93.7 | 91.3 | 95.7 |
| 7 | Excitement,Mutism,Posturing/catalepsy,Grimacing,Mannerisms,Rigidity,Waxy flexibility | 93.7 | 91.3 | 96.0 |
| 7 | Excitement,Mutism,Posturing/catalepsy,Grimacing,Stereotypy,Verbigeration,Rigidity | 93.7 | 91.5 | 96.0 |
| 7 | Excitement,Posturing/catalepsy,Echopraxia/echolalia,Rigidity,Negativism,Waxy flexibility,Withdrawal | 93.7 | 91.3 | 96.0 |
| 7 | Excitement,Posturing/catalepsy,Grimacing,Mannerisms,Rigidity,Negativism,Withdrawal | 93.7 | 91.3 | 96.0 |
| 7 | Excitement,Posturing/catalepsy,Grimacing,Stereotypy,Rigidity,Negativism,Withdrawal | 93.7 | 91.3 | 96.0 |
| 7 | Excitement,Posturing/catalepsy,Stereotypy,Rigidity,Negativism,Waxy flexibility,Withdrawal | 93.7 | 91.3 | 96.0 |
| 7 | Immobility/stupor,Grimacing,Echopraxia/echolalia,Mannerisms,Verbigeration,Waxy flexibility,Withdrawal | 93.7 | 91.5 | 96.0 |
| 7 | Immobility/stupor,Grimacing,Stereotypy,Mannerisms,Verbigeration,Waxy flexibility,Withdrawal | 93.7 | 91.5 | 95.7 |
| 7 | Immobility/stupor,Mutism,Echopraxia/echolalia,Stereotypy,Verbigeration,Waxy flexibility,Withdrawal | 93.7 | 91.3 | 96.0 |
| 7 | Immobility/stupor,Mutism,Grimacing,Echopraxia/echolalia,Stereotypy,Mannerisms,Verbigeration | 93.7 | 91.5 | 96.0 |
| 7 | Immobility/stupor,Mutism,Grimacing,Stereotypy,Mannerisms,Verbigeration,Waxy flexibility | 93.7 | 91.5 | 96.0 |
| 7 | Immobility/stupor,Mutism,Posturing/catalepsy,Echopraxia/echolalia,Mannerisms,Verbigeration,Waxy flexibility | 93.7 | 91.5 | 96.0 |
| 7 | Immobility/stupor,Mutism,Posturing/catalepsy,Grimacing,Stereotypy,Mannerisms,Waxy flexibility | 93.7 | 91.5 | 96.0 |
| 7 | Immobility/stupor,Mutism,Stereotypy,Mannerisms,Verbigeration,Waxy flexibility,Withdrawal | 93.7 | 91.3 | 96.0 |
| 7 | Immobility/stupor,Posturing/catalepsy,Grimacing,Stereotypy,Verbigeration,Waxy flexibility,Withdrawal | 93.7 | 91.5 | 95.7 |
| 7 | Mutism,Grimacing,Echopraxia/echolalia,Stereotypy,Mannerisms,Rigidity,Negativism | 93.7 | 91.5 | 96.0 |
| 7 | Mutism,Grimacing,Stereotypy,Mannerisms,Rigidity,Waxy flexibility,Withdrawal | 93.7 | 91.5 | 96.0 |
| 7 | Mutism,Posturing/catalepsy,Echopraxia/echolalia,Mannerisms,Verbigeration,Negativism,Withdrawal | 93.7 | 91.3 | 96.0 |
| 7 | Mutism,Posturing/catalepsy,Echopraxia/echolalia,Stereotypy,Mannerisms,Negativism,Withdrawal | 93.7 | 91.5 | 96.0 |
| 7 | Mutism,Posturing/catalepsy,Grimacing,Echopraxia/echolalia,Mannerisms,Rigidity,Withdrawal | 93.7 | 91.5 | 95.7 |
| 7 | Mutism,Posturing/catalepsy,Grimacing,Mannerisms,Verbigeration,Negativism,Withdrawal | 93.7 | 91.5 | 96.0 |
| 7 | Mutism,Posturing/catalepsy,Grimacing,Mannerisms,Verbigeration,Rigidity,Withdrawal | 93.7 | 91.3 | 96.0 |
| 7 | Mutism,Posturing/catalepsy,Grimacing,Stereotypy,Mannerisms,Negativism,Withdrawal | 93.7 | 91.5 | 96.0 |
| 7 | Mutism,Posturing/catalepsy,Grimacing,Verbigeration,Rigidity,Negativism,Waxy flexibility | 93.7 | 91.5 | 96.0 |
| 7 | Mutism,Posturing/catalepsy,Mannerisms,Verbigeration,Rigidity,Waxy flexibility,Withdrawal | 93.7 | 91.3 | 96.0 |
| 7 | Posturing/catalepsy,Grimacing,Echopraxia/echolalia,Mannerisms,Rigidity,Negativism,Withdrawal | 93.7 | 91.5 | 96.0 |
| 7 | Posturing/catalepsy,Grimacing,Mannerisms,Verbigeration,Rigidity,Negativism,Withdrawal | 93.7 | 91.5 | 96.0 |
| 7 | Posturing/catalepsy,Mannerisms,Verbigeration,Rigidity,Negativism,Waxy flexibility,Withdrawal | 93.7 | 91.5 | 96.0 |
| 7 | Excitement,Mutism,Echopraxia/echolalia,Mannerisms,Rigidity,Negativism,Waxy flexibility | 93.5 | 91.0 | 95.7 |
| 7 | Excitement,Mutism,Echopraxia/echolalia,Mannerisms,Verbigeration,Rigidity,Negativism | 93.5 | 91.0 | 95.7 |
| 7 | Excitement,Mutism,Grimacing,Mannerisms,Rigidity,Negativism,Waxy flexibility | 93.5 | 91.0 | 95.7 |
| 7 | Excitement,Mutism,Grimacing,Stereotypy,Verbigeration,Rigidity,Withdrawal | 93.5 | 91.3 | 95.7 |
| 7 | Excitement,Mutism,Posturing/catalepsy,Grimacing,Echopraxia/echolalia,Stereotypy,Rigidity | 93.5 | 91.0 | 95.7 |
| 7 | Excitement,Mutism,Posturing/catalepsy,Grimacing,Stereotypy,Rigidity,Waxy flexibility | 93.5 | 91.0 | 95.7 |
| 7 | Excitement,Mutism,Posturing/catalepsy,Stereotypy,Mannerisms,Verbigeration,Rigidity | 93.5 | 91.3 | 95.7 |
| 7 | Excitement,Mutism,Stereotypy,Mannerisms,Verbigeration,Rigidity,Withdrawal | 93.5 | 91.0 | 95.7 |
| 7 | Excitement,Posturing/catalepsy,Mannerisms,Rigidity,Negativism,Waxy flexibility,Withdrawal | 93.5 | 91.0 | 95.7 |
| 7 | Immobility/stupor,Mutism,Posturing/catalepsy,Echopraxia/echolalia,Stereotypy,Mannerisms,Waxy flexibility | 93.5 | 91.0 | 95.7 |
| 7 | Immobility/stupor,Mutism,Posturing/catalepsy,Echopraxia/echolalia,Stereotypy,Verbigeration,Waxy flexibility | 93.5 | 91.0 | 95.7 |
| 7 | Immobility/stupor,Posturing/catalepsy,Echopraxia/echolalia,Mannerisms,Verbigeration,Waxy flexibility,Withdrawal | 93.5 | 91.0 | 95.7 |
| 7 | Immobility/stupor,Posturing/catalepsy,Grimacing,Echopraxia/echolalia,Stereotypy,Mannerisms,Verbigeration | 93.5 | 91.0 | 95.7 |
| 7 | Immobility/stupor,Posturing/catalepsy,Grimacing,Echopraxia/echolalia,Stereotypy,Mannerisms,Waxy flexibility | 93.5 | 91.0 | 95.7 |
| 7 | Immobility/stupor,Posturing/catalepsy,Grimacing,Echopraxia/echolalia,Verbigeration,Waxy flexibility,Withdrawal | 93.5 | 91.0 | 95.7 |
| 7 | Immobility/stupor,Posturing/catalepsy,Stereotypy,Mannerisms,Verbigeration,Waxy flexibility,Withdrawal | 93.5 | 91.0 | 95.7 |
| 7 | Mutism,Echopraxia/echolalia,Mannerisms,Verbigeration,Rigidity,Negativism,Waxy flexibility | 93.5 | 91.0 | 95.7 |
| 7 | Mutism,Echopraxia/echolalia,Stereotypy,Verbigeration,Rigidity,Negativism,Waxy flexibility | 93.5 | 91.0 | 95.7 |
| 7 | Mutism,Grimacing,Echopraxia/echolalia,Mannerisms,Rigidity,Waxy flexibility,Withdrawal | 93.5 | 91.3 | 95.7 |
| 7 | Mutism,Grimacing,Echopraxia/echolalia,Mannerisms,Verbigeration,Rigidity,Negativism | 93.5 | 91.0 | 95.5 |
| 7 | Mutism,Grimacing,Mannerisms,Verbigeration,Rigidity,Negativism,Waxy flexibility | 93.5 | 91.0 | 95.7 |
| 7 | Mutism,Grimacing,Mannerisms,Verbigeration,Rigidity,Waxy flexibility,Withdrawal | 93.5 | 91.3 | 95.7 |
| 7 | Mutism,Grimacing,Stereotypy,Verbigeration,Rigidity,Waxy flexibility,Withdrawal | 93.5 | 91.0 | 95.7 |
| 7 | Mutism,Posturing/catalepsy,Echopraxia/echolalia,Mannerisms,Negativism,Waxy flexibility,Withdrawal | 93.5 | 91.0 | 95.7 |
| 7 | Mutism,Posturing/catalepsy,Echopraxia/echolalia,Mannerisms,Rigidity,Waxy flexibility,Withdrawal | 93.5 | 91.0 | 95.7 |
| 7 | Mutism,Posturing/catalepsy,Grimacing,Echopraxia/echolalia,Stereotypy,Rigidity,Negativism | 93.5 | 91.0 | 95.7 |
| 7 | Mutism,Posturing/catalepsy,Grimacing,Mannerisms,Negativism,Waxy flexibility,Withdrawal | 93.5 | 91.3 | 95.7 |
| 7 | Mutism,Posturing/catalepsy,Grimacing,Stereotypy,Mannerisms,Rigidity,Withdrawal | 93.5 | 91.0 | 95.7 |
| 7 | Mutism,Posturing/catalepsy,Mannerisms,Verbigeration,Negativism,Waxy flexibility,Withdrawal | 93.5 | 91.3 | 95.7 |
| 7 | Mutism,Posturing/catalepsy,Stereotypy,Mannerisms,Negativism,Waxy flexibility,Withdrawal | 93.5 | 91.0 | 95.7 |
| 7 | Mutism,Posturing/catalepsy,Stereotypy,Mannerisms,Rigidity,Waxy flexibility,Withdrawal | 93.5 | 91.0 | 95.7 |
| 7 | Posturing/catalepsy,Echopraxia/echolalia,Mannerisms,Rigidity,Negativism,Waxy flexibility,Withdrawal | 93.5 | 91.0 | 95.7 |
| 7 | Posturing/catalepsy,Grimacing,Stereotypy,Verbigeration,Rigidity,Negativism,Withdrawal | 93.5 | 91.3 | 95.7 |
| 7 | Posturing/catalepsy,Stereotypy,Verbigeration,Rigidity,Negativism,Waxy flexibility,Withdrawal | 93.5 | 91.0 | 95.7 |
| 7 | Excitement,Mutism,Echopraxia/echolalia,Stereotypy,Mannerisms,Rigidity,Withdrawal | 93.3 | 90.8 | 95.5 |
| 7 | Excitement,Mutism,Echopraxia/echolalia,Stereotypy,Verbigeration,Rigidity,Waxy flexibility | 93.3 | 90.8 | 95.5 |
| 7 | Excitement,Mutism,Grimacing,Echopraxia/echolalia,Stereotypy,Rigidity,Withdrawal | 93.3 | 91.0 | 95.5 |
| 7 | Excitement,Mutism,Grimacing,Echopraxia/echolalia,Verbigeration,Rigidity,Withdrawal | 93.3 | 90.8 | 95.5 |
| 7 | Excitement,Mutism,Grimacing,Mannerisms,Verbigeration,Rigidity,Withdrawal | 93.3 | 90.8 | 95.5 |
| 7 | Excitement,Mutism,Grimacing,Stereotypy,Mannerisms,Rigidity,Negativism | 93.3 | 90.8 | 95.5 |
| 7 | Excitement,Mutism,Posturing/catalepsy,Echopraxia/echolalia,Stereotypy,Mannerisms,Rigidity | 93.3 | 90.8 | 95.5 |
| 7 | Excitement,Mutism,Posturing/catalepsy,Echopraxia/echolalia,Verbigeration,Waxy flexibility,Withdrawal | 93.3 | 90.8 | 95.5 |
| 7 | Excitement,Mutism,Posturing/catalepsy,Grimacing,Echopraxia/echolalia,Verbigeration,Negativism | 93.3 | 90.8 | 95.5 |
| 7 | Excitement,Mutism,Posturing/catalepsy,Grimacing,Echopraxia/echolalia,Waxy flexibility,Withdrawal | 93.3 | 90.8 | 95.5 |
| 7 | Excitement,Mutism,Posturing/catalepsy,Stereotypy,Verbigeration,Waxy flexibility,Withdrawal | 93.3 | 90.8 | 95.5 |
| 7 | Excitement,Posturing/catalepsy,Echopraxia/echolalia,Verbigeration,Rigidity,Waxy flexibility,Withdrawal | 93.3 | 90.8 | 95.5 |
| 7 | Excitement,Posturing/catalepsy,Grimacing,Rigidity,Negativism,Waxy flexibility,Withdrawal | 93.3 | 90.8 | 95.5 |
| 7 | Immobility/stupor,Echopraxia/echolalia,Stereotypy,Mannerisms,Verbigeration,Waxy flexibility,Withdrawal | 93.3 | 90.8 | 95.5 |
| 7 | Immobility/stupor,Grimacing,Echopraxia/echolalia,Stereotypy,Mannerisms,Verbigeration,Waxy flexibility | 93.3 | 90.8 | 95.5 |
| 7 | Immobility/stupor,Mutism,Echopraxia/echolalia,Stereotypy,Mannerisms,Verbigeration,Waxy flexibility | 93.3 | 90.8 | 95.5 |
| 7 | Immobility/stupor,Mutism,Posturing/catalepsy,Stereotypy,Mannerisms,Verbigeration,Waxy flexibility | 93.3 | 90.8 | 95.5 |
| 7 | Immobility/stupor,Posturing/catalepsy,Grimacing,Echopraxia/echolalia,Mannerisms,Verbigeration,Waxy flexibility | 93.3 | 90.8 | 95.5 |
| 7 | Mutism,Grimacing,Echopraxia/echolalia,Stereotypy,Verbigeration,Rigidity,Negativism | 93.3 | 90.8 | 95.5 |
| 7 | Mutism,Grimacing,Echopraxia/echolalia,Verbigeration,Rigidity,Negativism,Waxy flexibility | 93.3 | 90.8 | 95.5 |
| 7 | Mutism,Posturing/catalepsy,Echopraxia/echolalia,Mannerisms,Verbigeration,Rigidity,Withdrawal | 93.3 | 90.8 | 95.5 |
| 7 | Mutism,Posturing/catalepsy,Echopraxia/echolalia,Stereotypy,Rigidity,Negativism,Waxy flexibility | 93.3 | 91.0 | 95.5 |
| 7 | Mutism,Posturing/catalepsy,Grimacing,Echopraxia/echolalia,Rigidity,Negativism,Waxy flexibility | 93.3 | 90.8 | 95.5 |
| 7 | Mutism,Posturing/catalepsy,Grimacing,Stereotypy,Verbigeration,Negativism,Withdrawal | 93.3 | 90.8 | 95.5 |
| 7 | Mutism,Posturing/catalepsy,Grimacing,Stereotypy,Verbigeration,Rigidity,Withdrawal | 93.3 | 90.8 | 95.5 |
| 7 | Mutism,Posturing/catalepsy,Grimacing,Verbigeration,Rigidity,Waxy flexibility,Withdrawal | 93.3 | 90.8 | 95.5 |
| 7 | Mutism,Posturing/catalepsy,Stereotypy,Mannerisms,Verbigeration,Rigidity,Withdrawal | 93.3 | 90.8 | 95.5 |
| 7 | Mutism,Posturing/catalepsy,Stereotypy,Verbigeration,Rigidity,Waxy flexibility,Withdrawal | 93.3 | 90.8 | 95.5 |
| 7 | Mutism,Stereotypy,Mannerisms,Verbigeration,Rigidity,Waxy flexibility,Withdrawal | 93.3 | 90.8 | 95.5 |
| 7 | Posturing/catalepsy,Echopraxia/echolalia,Stereotypy,Verbigeration,Rigidity,Negativism,Withdrawal | 93.3 | 90.8 | 95.5 |
| 7 | Posturing/catalepsy,Grimacing,Mannerisms,Rigidity,Negativism,Waxy flexibility,Withdrawal | 93.3 | 90.8 | 95.5 |
| 7 | Excitement,Mutism,Echopraxia/echolalia,Stereotypy,Negativism,Waxy flexibility,Withdrawal | 93.1 | 90.6 | 95.3 |
| 7 | Excitement,Mutism,Echopraxia/echolalia,Stereotypy,Verbigeration,Negativism,Withdrawal | 93.1 | 90.6 | 95.3 |
| 7 | Excitement,Mutism,Grimacing,Echopraxia/echolalia,Verbigeration,Rigidity,Waxy flexibility | 93.1 | 90.6 | 95.3 |
| 7 | Excitement,Mutism,Grimacing,Stereotypy,Verbigeration,Rigidity,Waxy flexibility | 93.1 | 90.6 | 95.3 |
| 7 | Excitement,Mutism,Posturing/catalepsy,Echopraxia/echolalia,Verbigeration,Negativism,Waxy flexibility | 93.1 | 90.6 | 95.3 |
| 7 | Excitement,Mutism,Posturing/catalepsy,Grimacing,Echopraxia/echolalia,Negativism,Waxy flexibility | 93.1 | 90.6 | 95.3 |
| 7 | Excitement,Mutism,Posturing/catalepsy,Grimacing,Stereotypy,Verbigeration,Negativism | 93.1 | 90.6 | 95.3 |
| 7 | Excitement,Mutism,Posturing/catalepsy,Stereotypy,Mannerisms,Rigidity,Waxy flexibility | 93.1 | 90.6 | 95.3 |
| 7 | Immobility/stupor,Posturing/catalepsy,Grimacing,Echopraxia/echolalia,Stereotypy,Verbigeration,Waxy flexibility | 93.1 | 90.6 | 95.3 |
| 7 | Mutism,Posturing/catalepsy,Echopraxia/echolalia,Stereotypy,Mannerisms,Rigidity,Withdrawal | 93.1 | 90.6 | 95.3 |
| 7 | Mutism,Posturing/catalepsy,Echopraxia/echolalia,Stereotypy,Verbigeration,Negativism,Withdrawal | 93.1 | 90.6 | 95.3 |
| 7 | Mutism,Posturing/catalepsy,Grimacing,Echopraxia/echolalia,Rigidity,Waxy flexibility,Withdrawal | 93.1 | 90.6 | 95.3 |
| 7 | Mutism,Posturing/catalepsy,Grimacing,Stereotypy,Rigidity,Negativism,Waxy flexibility | 93.1 | 90.6 | 95.3 |
| 7 | Mutism,Posturing/catalepsy,Grimacing,Stereotypy,Rigidity,Waxy flexibility,Withdrawal | 93.1 | 90.6 | 95.3 |
| 7 | Excitement,Mutism,Grimacing,Echopraxia/echolalia,Mannerisms,Rigidity,Withdrawal | 93.0 | 90.6 | 95.3 |
| 7 | Excitement,Mutism,Grimacing,Echopraxia/echolalia,Stereotypy,Rigidity,Waxy flexibility | 93.0 | 90.6 | 95.3 |
| 7 | Excitement,Mutism,Posturing/catalepsy,Echopraxia/echolalia,Stereotypy,Verbigeration,Negativism | 93.0 | 90.6 | 95.3 |
| 7 | Excitement,Mutism,Posturing/catalepsy,Echopraxia/echolalia,Stereotypy,Waxy flexibility,Withdrawal | 93.0 | 90.6 | 95.3 |
| 7 | Excitement,Mutism,Posturing/catalepsy,Grimacing,Verbigeration,Negativism,Waxy flexibility | 93.0 | 90.6 | 95.3 |
| 7 | Excitement,Mutism,Posturing/catalepsy,Grimacing,Verbigeration,Waxy flexibility,Withdrawal | 93.0 | 90.6 | 95.3 |
| 7 | Excitement,Mutism,Posturing/catalepsy,Mannerisms,Verbigeration,Waxy flexibility,Withdrawal | 93.0 | 90.6 | 95.3 |
| 7 | Excitement,Mutism,Posturing/catalepsy,Stereotypy,Verbigeration,Negativism,Waxy flexibility | 93.0 | 90.6 | 95.3 |
| 7 | Excitement,Mutism,Stereotypy,Verbigeration,Negativism,Waxy flexibility,Withdrawal | 93.0 | 90.6 | 95.3 |
| 7 | Excitement,Posturing/catalepsy,Mannerisms,Verbigeration,Rigidity,Waxy flexibility,Withdrawal | 93.0 | 90.6 | 95.3 |
| 7 | Immobility/stupor,Posturing/catalepsy,Echopraxia/echolalia,Stereotypy,Verbigeration,Waxy flexibility,Withdrawal | 93.0 | 90.6 | 95.3 |
| 7 | Immobility/stupor,Posturing/catalepsy,Grimacing,Stereotypy,Mannerisms,Verbigeration,Waxy flexibility | 93.0 | 90.6 | 95.3 |
| 7 | Mutism,Echopraxia/echolalia,Stereotypy,Mannerisms,Rigidity,Waxy flexibility,Withdrawal | 93.0 | 90.6 | 95.3 |
| 7 | Mutism,Grimacing,Echopraxia/echolalia,Stereotypy,Rigidity,Negativism,Waxy flexibility | 93.0 | 90.6 | 95.3 |
| 7 | Mutism,Grimacing,Echopraxia/echolalia,Stereotypy,Rigidity,Waxy flexibility,Withdrawal | 93.0 | 90.6 | 95.3 |
| 7 | Mutism,Posturing/catalepsy,Grimacing,Echopraxia/echolalia,Stereotypy,Negativism,Withdrawal | 93.0 | 90.6 | 95.3 |
| 7 | Mutism,Posturing/catalepsy,Grimacing,Echopraxia/echolalia,Stereotypy,Rigidity,Withdrawal | 93.0 | 90.6 | 95.3 |
| 7 | Mutism,Posturing/catalepsy,Stereotypy,Verbigeration,Negativism,Waxy flexibility,Withdrawal | 93.0 | 90.6 | 95.3 |
| 7 | Posturing/catalepsy,Grimacing,Echopraxia/echolalia,Stereotypy,Rigidity,Negativism,Withdrawal | 93.0 | 90.6 | 95.3 |
| 7 | Excitement,Mutism,Posturing/catalepsy,Stereotypy,Mannerisms,Verbigeration,Negativism | 92.9 | 90.4 | 95.1 |
| 7 | Excitement,Mutism,Echopraxia/echolalia,Mannerisms,Verbigeration,Rigidity,Withdrawal | 92.8 | 90.4 | 95.1 |
| 7 | Excitement,Mutism,Grimacing,Stereotypy,Mannerisms,Rigidity,Withdrawal | 92.8 | 90.4 | 95.1 |
| 7 | Excitement,Mutism,Posturing/catalepsy,Echopraxia/echolalia,Mannerisms,Verbigeration,Negativism | 92.8 | 90.4 | 95.1 |
| 7 | Excitement,Mutism,Posturing/catalepsy,Echopraxia/echolalia,Mannerisms,Waxy flexibility,Withdrawal | 92.8 | 90.4 | 95.1 |
| 7 | Excitement,Mutism,Posturing/catalepsy,Grimacing,Echopraxia/echolalia,Mannerisms,Negativism | 92.8 | 90.4 | 95.1 |
| 7 | Excitement,Mutism,Posturing/catalepsy,Grimacing,Echopraxia/echolalia,Stereotypy,Negativism | 92.8 | 90.4 | 95.1 |
| 7 | Excitement,Mutism,Posturing/catalepsy,Grimacing,Mannerisms,Verbigeration,Negativism | 92.8 | 90.4 | 95.1 |
| 7 | Excitement,Mutism,Posturing/catalepsy,Grimacing,Mannerisms,Waxy flexibility,Withdrawal | 92.8 | 90.4 | 95.1 |
| 7 | Excitement,Mutism,Posturing/catalepsy,Grimacing,Stereotypy,Mannerisms,Rigidity | 92.8 | 90.4 | 95.1 |
| 7 | Excitement,Mutism,Posturing/catalepsy,Grimacing,Stereotypy,Waxy flexibility,Withdrawal | 92.8 | 90.4 | 95.1 |
| 7 | Excitement,Posturing/catalepsy,Echopraxia/echolalia,Mannerisms,Rigidity,Waxy flexibility,Withdrawal | 92.8 | 90.4 | 95.1 |
| 7 | Excitement,Posturing/catalepsy,Echopraxia/echolalia,Mannerisms,Verbigeration,Rigidity,Withdrawal | 92.8 | 90.4 | 95.1 |
| 7 | Excitement,Posturing/catalepsy,Grimacing,Echopraxia/echolalia,Rigidity,Waxy flexibility,Withdrawal | 92.8 | 90.4 | 95.1 |
| 7 | Excitement,Posturing/catalepsy,Stereotypy,Verbigeration,Rigidity,Waxy flexibility,Withdrawal | 92.8 | 90.4 | 95.1 |
| 7 | Mutism,Grimacing,Echopraxia/echolalia,Verbigeration,Rigidity,Waxy flexibility,Withdrawal | 92.8 | 90.4 | 95.1 |
| 7 | Mutism,Posturing/catalepsy,Echopraxia/echolalia,Stereotypy,Rigidity,Waxy flexibility,Withdrawal | 92.8 | 90.4 | 95.1 |
| 7 | Mutism,Posturing/catalepsy,Echopraxia/echolalia,Verbigeration,Rigidity,Waxy flexibility,Withdrawal | 92.8 | 90.4 | 95.1 |
| 7 | Mutism,Posturing/catalepsy,Grimacing,Echopraxia/echolalia,Verbigeration,Negativism,Withdrawal | 92.8 | 90.4 | 95.1 |
| 7 | Mutism,Posturing/catalepsy,Grimacing,Echopraxia/echolalia,Verbigeration,Rigidity,Withdrawal | 92.8 | 90.4 | 95.1 |
| 7 | Posturing/catalepsy,Echopraxia/echolalia,Stereotypy,Rigidity,Negativism,Waxy flexibility,Withdrawal | 92.8 | 90.4 | 95.1 |
| 7 | Posturing/catalepsy,Echopraxia/echolalia,Verbigeration,Rigidity,Negativism,Waxy flexibility,Withdrawal | 92.8 | 90.4 | 95.1 |
| 7 | Posturing/catalepsy,Grimacing,Echopraxia/echolalia,Verbigeration,Rigidity,Negativism,Withdrawal | 92.8 | 90.4 | 95.1 |
| 7 | Posturing/catalepsy,Grimacing,Stereotypy,Rigidity,Negativism,Waxy flexibility,Withdrawal | 92.8 | 90.4 | 95.3 |
| 7 | Excitement,Mutism,Echopraxia/echolalia,Stereotypy,Mannerisms,Rigidity,Waxy flexibility | 92.6 | 90.1 | 94.8 |
| 7 | Excitement,Mutism,Posturing/catalepsy,Echopraxia/echolalia,Stereotypy,Negativism,Waxy flexibility | 92.6 | 90.1 | 94.8 |
| 7 | Excitement,Mutism,Posturing/catalepsy,Echopraxia/echolalia,Stereotypy,Verbigeration,Withdrawal | 92.6 | 90.1 | 94.8 |
| 7 | Excitement,Mutism,Posturing/catalepsy,Grimacing,Echopraxia/echolalia,Verbigeration,Withdrawal | 92.6 | 90.1 | 95.1 |
| 7 | Excitement,Mutism,Posturing/catalepsy,Mannerisms,Verbigeration,Negativism,Waxy flexibility | 92.6 | 90.1 | 94.8 |
| 7 | Excitement,Mutism,Posturing/catalepsy,Stereotypy,Mannerisms,Waxy flexibility,Withdrawal | 92.6 | 89.9 | 95.1 |
| 7 | Excitement,Mutism,Stereotypy,Mannerisms,Verbigeration,Rigidity,Waxy flexibility | 92.6 | 90.1 | 94.8 |
| 7 | Excitement,Posturing/catalepsy,Echopraxia/echolalia,Stereotypy,Rigidity,Waxy flexibility,Withdrawal | 92.6 | 90.1 | 95.1 |
| 7 | Excitement,Posturing/catalepsy,Grimacing,Echopraxia/echolalia,Verbigeration,Rigidity,Withdrawal | 92.6 | 90.1 | 94.8 |
| 7 | Excitement,Posturing/catalepsy,Grimacing,Verbigeration,Rigidity,Waxy flexibility,Withdrawal | 92.6 | 90.1 | 94.8 |
| 7 | Mutism,Echopraxia/echolalia,Stereotypy,Mannerisms,Negativism,Waxy flexibility,Withdrawal | 92.6 | 90.1 | 94.8 |
| 7 | Mutism,Echopraxia/echolalia,Stereotypy,Verbigeration,Rigidity,Waxy flexibility,Withdrawal | 92.6 | 89.9 | 94.8 |
| 7 | Mutism,Posturing/catalepsy,Echopraxia/echolalia,Stereotypy,Negativism,Waxy flexibility,Withdrawal | 92.6 | 90.1 | 94.8 |
| 7 | Mutism,Posturing/catalepsy,Echopraxia/echolalia,Stereotypy,Verbigeration,Rigidity,Withdrawal | 92.6 | 90.1 | 94.8 |
| 7 | Mutism,Posturing/catalepsy,Echopraxia/echolalia,Verbigeration,Negativism,Waxy flexibility,Withdrawal | 92.6 | 90.1 | 94.8 |
| 7 | Mutism,Posturing/catalepsy,Grimacing,Echopraxia/echolalia,Mannerisms,Verbigeration,Negativism | 92.6 | 90.1 | 94.8 |
| 7 | Mutism,Posturing/catalepsy,Grimacing,Echopraxia/echolalia,Negativism,Waxy flexibility,Withdrawal | 92.6 | 90.1 | 94.8 |
| 7 | Mutism,Posturing/catalepsy,Grimacing,Stereotypy,Negativism,Waxy flexibility,Withdrawal | 92.6 | 90.1 | 95.1 |
| 7 | Mutism,Posturing/catalepsy,Grimacing,Verbigeration,Negativism,Waxy flexibility,Withdrawal | 92.6 | 90.1 | 94.8 |
| 7 | Posturing/catalepsy,Grimacing,Verbigeration,Rigidity,Negativism,Waxy flexibility,Withdrawal | 92.6 | 90.1 | 94.8 |
| 7 | Echopraxia/echolalia,Stereotypy,Mannerisms,Rigidity,Negativism,Waxy flexibility,Withdrawal | 92.4 | 89.9 | 94.6 |
| 7 | Excitement,Mutism,Echopraxia/echolalia,Stereotypy,Mannerisms,Negativism,Withdrawal | 92.4 | 89.9 | 94.8 |
| 7 | Excitement,Mutism,Grimacing,Echopraxia/echolalia,Mannerisms,Rigidity,Waxy flexibility | 92.4 | 89.9 | 94.8 |
| 7 | Excitement,Mutism,Grimacing,Echopraxia/echolalia,Stereotypy,Negativism,Withdrawal | 92.4 | 89.9 | 94.8 |
| 7 | Excitement,Mutism,Grimacing,Mannerisms,Verbigeration,Rigidity,Waxy flexibility | 92.4 | 89.7 | 94.6 |
| 7 | Excitement,Mutism,Grimacing,Stereotypy,Negativism,Waxy flexibility,Withdrawal | 92.4 | 89.9 | 94.6 |
| 7 | Excitement,Mutism,Grimacing,Stereotypy,Verbigeration,Negativism,Withdrawal | 92.4 | 89.7 | 94.8 |
| 7 | Excitement,Mutism,Posturing/catalepsy,Echopraxia/echolalia,Mannerisms,Negativism,Waxy flexibility | 92.4 | 89.9 | 94.8 |
| 7 | Excitement,Mutism,Posturing/catalepsy,Echopraxia/echolalia,Mannerisms,Verbigeration,Withdrawal | 92.4 | 89.9 | 94.8 |
| 7 | Excitement,Mutism,Posturing/catalepsy,Echopraxia/echolalia,Stereotypy,Mannerisms,Negativism | 92.4 | 89.7 | 94.8 |
| 7 | Excitement,Mutism,Posturing/catalepsy,Grimacing,Echopraxia/echolalia,Mannerisms,Withdrawal | 92.4 | 89.9 | 94.6 |
| 7 | Excitement,Mutism,Posturing/catalepsy,Grimacing,Echopraxia/echolalia,Stereotypy,Withdrawal | 92.4 | 89.9 | 94.8 |
| 7 | Excitement,Mutism,Posturing/catalepsy,Grimacing,Mannerisms,Negativism,Waxy flexibility | 92.4 | 89.9 | 94.8 |
| 7 | Excitement,Mutism,Posturing/catalepsy,Grimacing,Mannerisms,Verbigeration,Withdrawal | 92.4 | 89.9 | 94.6 |
| 7 | Excitement,Mutism,Posturing/catalepsy,Grimacing,Stereotypy,Negativism,Waxy flexibility | 92.4 | 89.9 | 94.8 |
| 7 | Excitement,Mutism,Posturing/catalepsy,Grimacing,Stereotypy,Verbigeration,Withdrawal | 92.4 | 89.9 | 94.8 |
| 7 | Excitement,Mutism,Posturing/catalepsy,Stereotypy,Mannerisms,Verbigeration,Withdrawal | 92.4 | 89.9 | 94.8 |
| 7 | Excitement,Mutism,Stereotypy,Mannerisms,Negativism,Waxy flexibility,Withdrawal | 92.4 | 89.9 | 94.8 |
| 7 | Excitement,Mutism,Stereotypy,Mannerisms,Verbigeration,Negativism,Withdrawal | 92.4 | 89.9 | 94.8 |
| 7 | Excitement,Posturing/catalepsy,Echopraxia/echolalia,Stereotypy,Verbigeration,Rigidity,Withdrawal | 92.4 | 89.9 | 94.8 |
| 7 | Excitement,Posturing/catalepsy,Grimacing,Echopraxia/echolalia,Mannerisms,Rigidity,Withdrawal | 92.4 | 89.9 | 94.8 |
| 7 | Excitement,Posturing/catalepsy,Grimacing,Mannerisms,Rigidity,Waxy flexibility,Withdrawal | 92.4 | 89.9 | 94.6 |
| 7 | Excitement,Posturing/catalepsy,Grimacing,Mannerisms,Verbigeration,Rigidity,Withdrawal | 92.4 | 89.7 | 94.8 |
| 7 | Excitement,Posturing/catalepsy,Stereotypy,Mannerisms,Rigidity,Waxy flexibility,Withdrawal | 92.4 | 89.9 | 94.6 |
| 7 | Excitement,Posturing/catalepsy,Stereotypy,Mannerisms,Verbigeration,Negativism,Withdrawal | 92.4 | 89.9 | 94.6 |
| 7 | Excitement,Posturing/catalepsy,Stereotypy,Mannerisms,Verbigeration,Rigidity,Withdrawal | 92.4 | 89.9 | 94.8 |
| 7 | Excitement,Posturing/catalepsy,Stereotypy,Verbigeration,Negativism,Waxy flexibility,Withdrawal | 92.4 | 89.9 | 94.6 |
| 7 | Immobility/stupor,Posturing/catalepsy,Echopraxia/echolalia,Stereotypy,Mannerisms,Verbigeration,Waxy flexibility | 92.4 | 89.9 | 94.8 |
| 7 | Mutism,Echopraxia/echolalia,Mannerisms,Verbigeration,Rigidity,Waxy flexibility,Withdrawal | 92.4 | 89.9 | 94.8 |
| 7 | Mutism,Echopraxia/echolalia,Stereotypy,Mannerisms,Verbigeration,Negativism,Withdrawal | 92.4 | 89.7 | 94.8 |
| 7 | Mutism,Grimacing,Echopraxia/echolalia,Stereotypy,Mannerisms,Rigidity,Withdrawal | 92.4 | 89.9 | 94.8 |
| 7 | Mutism,Grimacing,Stereotypy,Mannerisms,Verbigeration,Rigidity,Withdrawal | 92.4 | 89.7 | 94.8 |
| 7 | Mutism,Posturing/catalepsy,Grimacing,Echopraxia/echolalia,Mannerisms,Negativism,Waxy flexibility | 92.4 | 89.7 | 94.6 |
| 7 | Mutism,Posturing/catalepsy,Grimacing,Echopraxia/echolalia,Mannerisms,Rigidity,Waxy flexibility | 92.4 | 89.9 | 94.8 |
| 7 | Mutism,Posturing/catalepsy,Grimacing,Echopraxia/echolalia,Mannerisms,Verbigeration,Rigidity | 92.4 | 89.9 | 94.8 |
| 7 | Mutism,Posturing/catalepsy,Grimacing,Echopraxia/echolalia,Stereotypy,Mannerisms,Negativism | 92.4 | 89.9 | 94.6 |
| 7 | Mutism,Posturing/catalepsy,Grimacing,Mannerisms,Verbigeration,Rigidity,Waxy flexibility | 92.4 | 89.9 | 94.8 |
| 7 | Mutism,Posturing/catalepsy,Grimacing,Stereotypy,Verbigeration,Rigidity,Waxy flexibility | 92.4 | 89.9 | 94.8 |
| 7 | Mutism,Stereotypy,Mannerisms,Verbigeration,Negativism,Waxy flexibility,Withdrawal | 92.4 | 89.9 | 94.8 |
| 7 | Posturing/catalepsy,Grimacing,Echopraxia/echolalia,Rigidity,Negativism,Waxy flexibility,Withdrawal | 92.4 | 89.9 | 94.6 |
| 7 | Stereotypy,Mannerisms,Verbigeration,Rigidity,Negativism,Waxy flexibility,Withdrawal | 92.4 | 89.9 | 94.8 |
| 7 | Echopraxia/echolalia,Stereotypy,Mannerisms,Verbigeration,Rigidity,Negativism,Withdrawal | 92.2 | 89.7 | 94.6 |
| 7 | Excitement,Mutism,Echopraxia/echolalia,Verbigeration,Negativism,Waxy flexibility,Withdrawal | 92.2 | 89.5 | 94.6 |
| 7 | Excitement,Mutism,Grimacing,Stereotypy,Mannerisms,Rigidity,Waxy flexibility | 92.2 | 89.5 | 94.6 |
| 7 | Excitement,Mutism,Posturing/catalepsy,Echopraxia/echolalia,Stereotypy,Mannerisms,Withdrawal | 92.2 | 89.7 | 94.6 |
| 7 | Excitement,Mutism,Posturing/catalepsy,Grimacing,Stereotypy,Mannerisms,Negativism | 92.2 | 89.5 | 94.6 |
| 7 | Excitement,Posturing/catalepsy,Grimacing,Echopraxia/echolalia,Verbigeration,Negativism,Withdrawal | 92.2 | 89.5 | 94.6 |
| 7 | Excitement,Posturing/catalepsy,Grimacing,Stereotypy,Rigidity,Waxy flexibility,Withdrawal | 92.2 | 89.7 | 94.6 |
| 7 | Excitement,Posturing/catalepsy,Grimacing,Stereotypy,Verbigeration,Negativism,Withdrawal | 92.2 | 89.5 | 94.6 |
| 7 | Grimacing,Echopraxia/echolalia,Stereotypy,Mannerisms,Rigidity,Negativism,Withdrawal | 92.2 | 89.7 | 94.4 |
| 7 | Mutism,Grimacing,Echopraxia/echolalia,Mannerisms,Verbigeration,Rigidity,Withdrawal | 92.2 | 89.7 | 94.6 |
| 7 | Mutism,Grimacing,Echopraxia/echolalia,Stereotypy,Mannerisms,Negativism,Withdrawal | 92.2 | 89.7 | 94.6 |
| 7 | Mutism,Posturing/catalepsy,Echopraxia/echolalia,Mannerisms,Verbigeration,Rigidity,Waxy flexibility | 92.2 | 89.5 | 94.6 |
| 7 | Mutism,Posturing/catalepsy,Echopraxia/echolalia,Stereotypy,Mannerisms,Verbigeration,Negativism | 92.2 | 89.7 | 94.6 |
| 7 | Mutism,Posturing/catalepsy,Grimacing,Echopraxia/echolalia,Verbigeration,Rigidity,Waxy flexibility | 92.2 | 89.7 | 94.4 |
| 7 | Mutism,Posturing/catalepsy,Grimacing,Stereotypy,Mannerisms,Verbigeration,Negativism | 92.2 | 89.5 | 94.6 |
| 7 | Posturing/catalepsy,Grimacing,Stereotypy,Mannerisms,Verbigeration,Negativism,Withdrawal | 92.2 | 89.7 | 94.6 |
| 7 | Excitement,Mutism,Grimacing,Echopraxia/echolalia,Negativism,Waxy flexibility,Withdrawal | 92.1 | 89.5 | 94.4 |
| 7 | Excitement,Posturing/catalepsy,Echopraxia/echolalia,Stereotypy,Mannerisms,Rigidity,Withdrawal | 92.1 | 89.7 | 94.6 |
| 7 | Excitement,Posturing/catalepsy,Echopraxia/echolalia,Stereotypy,Verbigeration,Negativism,Withdrawal | 92.1 | 89.5 | 94.4 |
| 7 | Excitement,Posturing/catalepsy,Grimacing,Mannerisms,Verbigeration,Negativism,Withdrawal | 92.1 | 89.5 | 94.4 |
| 7 | Grimacing,Stereotypy,Mannerisms,Rigidity,Negativism,Waxy flexibility,Withdrawal | 92.1 | 89.5 | 94.6 |
| 7 | Mutism,Grimacing,Stereotypy,Mannerisms,Negativism,Waxy flexibility,Withdrawal | 92.1 | 89.7 | 94.6 |
| 7 | Mutism,Posturing/catalepsy,Grimacing,Stereotypy,Mannerisms,Verbigeration,Rigidity | 92.1 | 89.5 | 94.4 |
| 7 | Posturing/catalepsy,Grimacing,Echopraxia/echolalia,Stereotypy,Mannerisms,Negativism,Withdrawal | 92.1 | 89.5 | 94.6 |
| 7 | Posturing/catalepsy,Stereotypy,Mannerisms,Verbigeration,Negativism,Waxy flexibility,Withdrawal | 92.1 | 89.5 | 94.4 |
| 7 | Excitement,Echopraxia/echolalia,Stereotypy,Mannerisms,Rigidity,Negativism,Withdrawal | 92.0 | 89.5 | 94.4 |
| 7 | Excitement,Stereotypy,Verbigeration,Rigidity,Negativism,Waxy flexibility,Withdrawal | 92.0 | 89.5 | 94.4 |
| 7 | Posturing/catalepsy,Echopraxia/echolalia,Stereotypy,Mannerisms,Verbigeration,Negativism,Withdrawal | 92.0 | 89.2 | 94.4 |
| 7 | Excitement,Echopraxia/echolalia,Stereotypy,Rigidity,Negativism,Waxy flexibility,Withdrawal | 91.9 | 89.2 | 94.4 |
| 7 | Excitement,Echopraxia/echolalia,Stereotypy,Verbigeration,Rigidity,Negativism,Withdrawal | 91.9 | 89.5 | 94.4 |
| 7 | Excitement,Mutism,Echopraxia/echolalia,Mannerisms,Verbigeration,Rigidity,Waxy flexibility | 91.9 | 89.2 | 94.4 |
| 7 | Excitement,Mutism,Posturing/catalepsy,Grimacing,Stereotypy,Mannerisms,Withdrawal | 91.9 | 89.2 | 94.4 |
| 7 | Excitement,Mutism,Posturing/catalepsy,Stereotypy,Mannerisms,Negativism,Waxy flexibility | 91.9 | 89.2 | 94.4 |
| 7 | Excitement,Posturing/catalepsy,Echopraxia/echolalia,Mannerisms,Verbigeration,Negativism,Withdrawal | 91.9 | 89.5 | 94.4 |
| 7 | Excitement,Posturing/catalepsy,Echopraxia/echolalia,Stereotypy,Mannerisms,Negativism,Withdrawal | 91.9 | 89.5 | 94.4 |
| 7 | Excitement,Posturing/catalepsy,Echopraxia/echolalia,Stereotypy,Negativism,Waxy flexibility,Withdrawal | 91.9 | 89.2 | 94.4 |
| 7 | Excitement,Posturing/catalepsy,Echopraxia/echolalia,Verbigeration,Negativism,Waxy flexibility,Withdrawal | 91.9 | 89.2 | 94.4 |
| 7 | Excitement,Posturing/catalepsy,Grimacing,Echopraxia/echolalia,Mannerisms,Negativism,Withdrawal | 91.9 | 89.2 | 94.4 |
| 7 | Excitement,Posturing/catalepsy,Grimacing,Echopraxia/echolalia,Stereotypy,Negativism,Withdrawal | 91.9 | 89.2 | 94.4 |
| 7 | Excitement,Posturing/catalepsy,Grimacing,Echopraxia/echolalia,Stereotypy,Rigidity,Withdrawal | 91.9 | 89.2 | 94.4 |
| 7 | Excitement,Posturing/catalepsy,Grimacing,Stereotypy,Verbigeration,Rigidity,Withdrawal | 91.9 | 89.2 | 94.4 |
| 7 | Excitement,Posturing/catalepsy,Mannerisms,Verbigeration,Negativism,Waxy flexibility,Withdrawal | 91.9 | 89.2 | 94.4 |
| 7 | Grimacing,Stereotypy,Mannerisms,Verbigeration,Rigidity,Negativism,Withdrawal | 91.9 | 89.2 | 94.2 |
| 7 | Mutism,Echopraxia/echolalia,Stereotypy,Verbigeration,Negativism,Waxy flexibility,Withdrawal | 91.9 | 89.2 | 94.4 |
| 7 | Mutism,Grimacing,Echopraxia/echolalia,Mannerisms,Negativism,Waxy flexibility,Withdrawal | 91.9 | 89.2 | 94.4 |
| 7 | Mutism,Grimacing,Echopraxia/echolalia,Stereotypy,Verbigeration,Rigidity,Withdrawal | 91.9 | 89.2 | 94.4 |
| 7 | Mutism,Grimacing,Stereotypy,Mannerisms,Verbigeration,Negativism,Withdrawal | 91.9 | 89.2 | 94.4 |
| 7 | Mutism,Posturing/catalepsy,Echopraxia/echolalia,Mannerisms,Verbigeration,Negativism,Waxy flexibility | 91.9 | 89.2 | 94.4 |
| 7 | Mutism,Posturing/catalepsy,Echopraxia/echolalia,Stereotypy,Mannerisms,Negativism,Waxy flexibility | 91.9 | 89.2 | 94.4 |
| 7 | Mutism,Posturing/catalepsy,Grimacing,Echopraxia/echolalia,Mannerisms,Waxy flexibility,Withdrawal | 91.9 | 89.2 | 94.4 |
| 7 | Mutism,Posturing/catalepsy,Grimacing,Echopraxia/echolalia,Stereotypy,Mannerisms,Rigidity | 91.9 | 89.2 | 94.4 |
| 7 | Mutism,Posturing/catalepsy,Grimacing,Echopraxia/echolalia,Stereotypy,Verbigeration,Negativism | 91.9 | 89.2 | 94.4 |
| 7 | Mutism,Posturing/catalepsy,Grimacing,Echopraxia/echolalia,Stereotypy,Verbigeration,Rigidity | 91.9 | 89.2 | 94.4 |
| 7 | Mutism,Posturing/catalepsy,Grimacing,Echopraxia/echolalia,Verbigeration,Negativism,Waxy flexibility | 91.9 | 89.2 | 94.4 |
| 7 | Mutism,Posturing/catalepsy,Grimacing,Mannerisms,Verbigeration,Negativism,Waxy flexibility | 91.9 | 89.2 | 94.4 |
| 7 | Mutism,Posturing/catalepsy,Grimacing,Stereotypy,Mannerisms,Rigidity,Waxy flexibility | 91.9 | 89.2 | 94.4 |
| 7 | Mutism,Posturing/catalepsy,Stereotypy,Mannerisms,Verbigeration,Negativism,Waxy flexibility | 91.9 | 89.2 | 94.4 |
| 7 | Mutism,Posturing/catalepsy,Stereotypy,Mannerisms,Verbigeration,Rigidity,Waxy flexibility | 91.9 | 89.2 | 94.4 |
| 7 | Posturing/catalepsy,Echopraxia/echolalia,Stereotypy,Mannerisms,Negativism,Waxy flexibility,Withdrawal | 91.9 | 89.2 | 94.4 |
| 7 | Excitement,Grimacing,Echopraxia/echolalia,Stereotypy,Rigidity,Negativism,Withdrawal | 91.7 | 89.0 | 94.2 |
| 7 | Excitement,Mutism,Echopraxia/echolalia,Mannerisms,Negativism,Waxy flexibility,Withdrawal | 91.7 | 89.0 | 94.2 |
| 7 | Excitement,Mutism,Grimacing,Echopraxia/echolalia,Verbigeration,Negativism,Withdrawal | 91.7 | 89.0 | 94.2 |
| 7 | Excitement,Mutism,Grimacing,Verbigeration,Negativism,Waxy flexibility,Withdrawal | 91.7 | 89.0 | 94.2 |
| 7 | Excitement,Mutism,Mannerisms,Verbigeration,Negativism,Waxy flexibility,Withdrawal | 91.7 | 89.0 | 94.2 |
| 7 | Excitement,Posturing/catalepsy,Grimacing,Echopraxia/echolalia,Negativism,Waxy flexibility,Withdrawal | 91.7 | 89.0 | 94.2 |
| 7 | Excitement,Posturing/catalepsy,Grimacing,Stereotypy,Mannerisms,Negativism,Withdrawal | 91.7 | 89.0 | 94.2 |
| 7 | Excitement,Posturing/catalepsy,Grimacing,Stereotypy,Mannerisms,Rigidity,Withdrawal | 91.7 | 89.0 | 94.2 |
| 7 | Excitement,Posturing/catalepsy,Stereotypy,Mannerisms,Negativism,Waxy flexibility,Withdrawal | 91.7 | 89.2 | 94.2 |
| 7 | Excitement,Stereotypy,Mannerisms,Verbigeration,Rigidity,Negativism,Withdrawal | 91.7 | 89.0 | 94.2 |
| 7 | Mutism,Echopraxia/echolalia,Stereotypy,Mannerisms,Verbigeration,Rigidity,Withdrawal | 91.7 | 89.0 | 94.2 |
| 7 | Mutism,Grimacing,Echopraxia/echolalia,Stereotypy,Negativism,Waxy flexibility,Withdrawal | 91.7 | 89.0 | 94.2 |
| 7 | Mutism,Grimacing,Echopraxia/echolalia,Stereotypy,Verbigeration,Negativism,Withdrawal | 91.7 | 89.0 | 94.2 |
| 7 | Mutism,Posturing/catalepsy,Echopraxia/echolalia,Stereotypy,Mannerisms,Rigidity,Waxy flexibility | 91.7 | 89.0 | 94.2 |
| 7 | Mutism,Posturing/catalepsy,Echopraxia/echolalia,Stereotypy,Mannerisms,Verbigeration,Rigidity | 91.7 | 89.2 | 94.2 |
| 7 | Mutism,Posturing/catalepsy,Echopraxia/echolalia,Stereotypy,Verbigeration,Rigidity,Waxy flexibility | 91.7 | 89.0 | 94.2 |
| 7 | Mutism,Posturing/catalepsy,Grimacing,Echopraxia/echolalia,Stereotypy,Rigidity,Waxy flexibility | 91.7 | 89.0 | 94.2 |
| 7 | Mutism,Posturing/catalepsy,Grimacing,Mannerisms,Verbigeration,Waxy flexibility,Withdrawal | 91.7 | 89.0 | 94.2 |
| 7 | Mutism,Posturing/catalepsy,Grimacing,Stereotypy,Mannerisms,Negativism,Waxy flexibility | 91.7 | 89.0 | 94.2 |
| 7 | Mutism,Posturing/catalepsy,Grimacing,Stereotypy,Mannerisms,Waxy flexibility,Withdrawal | 91.7 | 89.0 | 94.2 |
| 7 | Mutism,Posturing/catalepsy,Grimacing,Stereotypy,Verbigeration,Negativism,Waxy flexibility | 91.7 | 89.0 | 94.2 |
| 7 | Posturing/catalepsy,Grimacing,Stereotypy,Mannerisms,Negativism,Waxy flexibility,Withdrawal | 91.7 | 89.0 | 94.2 |
| 7 | Echopraxia/echolalia,Stereotypy,Verbigeration,Rigidity,Negativism,Waxy flexibility,Withdrawal | 91.5 | 88.8 | 93.9 |
| 7 | Excitement,Grimacing,Echopraxia/echolalia,Rigidity,Negativism,Waxy flexibility,Withdrawal | 91.5 | 88.8 | 93.9 |
| 7 | Excitement,Grimacing,Stereotypy,Rigidity,Negativism,Waxy flexibility,Withdrawal | 91.5 | 88.8 | 93.9 |
| 7 | Excitement,Grimacing,Stereotypy,Verbigeration,Rigidity,Negativism,Withdrawal | 91.5 | 88.8 | 93.9 |
| 7 | Excitement,Mutism,Echopraxia/echolalia,Mannerisms,Verbigeration,Negativism,Withdrawal | 91.5 | 88.8 | 93.9 |
| 7 | Excitement,Mutism,Grimacing,Echopraxia/echolalia,Stereotypy,Verbigeration,Rigidity | 91.5 | 88.8 | 93.9 |
| 7 | Excitement,Mutism,Grimacing,Mannerisms,Negativism,Waxy flexibility,Withdrawal | 91.5 | 88.8 | 93.9 |
| 7 | Excitement,Mutism,Grimacing,Stereotypy,Mannerisms,Negativism,Withdrawal | 91.5 | 88.8 | 93.9 |
| 7 | Excitement,Posturing/catalepsy,Echopraxia/echolalia,Mannerisms,Negativism,Waxy flexibility,Withdrawal | 91.5 | 88.8 | 93.9 |
| 7 | Excitement,Posturing/catalepsy,Grimacing,Stereotypy,Negativism,Waxy flexibility,Withdrawal | 91.5 | 88.8 | 93.9 |
| 7 | Excitement,Posturing/catalepsy,Grimacing,Verbigeration,Negativism,Waxy flexibility,Withdrawal | 91.5 | 88.8 | 93.9 |
| 7 | Excitement,Stereotypy,Mannerisms,Rigidity,Negativism,Waxy flexibility,Withdrawal | 91.5 | 88.8 | 93.9 |
| 7 | Grimacing,Echopraxia/echolalia,Mannerisms,Rigidity,Negativism,Waxy flexibility,Withdrawal | 91.5 | 88.8 | 93.9 |
| 7 | Grimacing,Stereotypy,Verbigeration,Rigidity,Negativism,Waxy flexibility,Withdrawal | 91.5 | 88.8 | 93.9 |
| 7 | Mutism,Echopraxia/echolalia,Mannerisms,Verbigeration,Negativism,Waxy flexibility,Withdrawal | 91.5 | 88.8 | 93.9 |
| 7 | Mutism,Grimacing,Echopraxia/echolalia,Stereotypy,Mannerisms,Rigidity,Waxy flexibility | 91.5 | 88.8 | 93.9 |
| 7 | Mutism,Grimacing,Stereotypy,Mannerisms,Verbigeration,Rigidity,Waxy flexibility | 91.5 | 88.8 | 93.9 |
| 7 | Mutism,Grimacing,Stereotypy,Verbigeration,Negativism,Waxy flexibility,Withdrawal | 91.5 | 88.8 | 93.9 |
| 7 | Mutism,Posturing/catalepsy,Echopraxia/echolalia,Stereotypy,Verbigeration,Negativism,Waxy flexibility | 91.5 | 88.8 | 93.9 |
| 7 | Mutism,Posturing/catalepsy,Grimacing,Echopraxia/echolalia,Mannerisms,Verbigeration,Withdrawal | 91.5 | 88.8 | 93.9 |
| 7 | Mutism,Posturing/catalepsy,Grimacing,Echopraxia/echolalia,Stereotypy,Mannerisms,Withdrawal | 91.5 | 88.8 | 93.9 |
| 7 | Mutism,Posturing/catalepsy,Grimacing,Echopraxia/echolalia,Stereotypy,Negativism,Waxy flexibility | 91.5 | 88.8 | 93.9 |
| 7 | Mutism,Posturing/catalepsy,Grimacing,Echopraxia/echolalia,Stereotypy,Waxy flexibility,Withdrawal | 91.5 | 88.8 | 93.9 |
| 7 | Posturing/catalepsy,Grimacing,Echopraxia/echolalia,Mannerisms,Verbigeration,Negativism,Withdrawal | 91.5 | 88.8 | 93.9 |
| 7 | Excitement,Mutism,Echopraxia/echolalia,Stereotypy,Verbigeration,Waxy flexibility,Withdrawal | 91.4 | 88.8 | 93.9 |
| 7 | Excitement,Grimacing,Echopraxia/echolalia,Mannerisms,Rigidity,Negativism,Withdrawal | 91.3 | 88.6 | 93.7 |
| 7 | Excitement,Grimacing,Echopraxia/echolalia,Verbigeration,Rigidity,Negativism,Withdrawal | 91.3 | 88.6 | 93.7 |
| 7 | Excitement,Mutism,Echopraxia/echolalia,Stereotypy,Mannerisms,Verbigeration,Rigidity | 91.3 | 88.6 | 93.7 |
| 7 | Excitement,Mutism,Echopraxia/echolalia,Stereotypy,Verbigeration,Negativism,Waxy flexibility | 91.3 | 88.6 | 93.7 |
| 7 | Excitement,Mutism,Grimacing,Echopraxia/echolalia,Stereotypy,Waxy flexibility,Withdrawal | 91.3 | 88.6 | 93.9 |
| 7 | Excitement,Mutism,Grimacing,Mannerisms,Verbigeration,Negativism,Withdrawal | 91.3 | 88.6 | 93.7 |
| 7 | Grimacing,Echopraxia/echolalia,Stereotypy,Rigidity,Negativism,Waxy flexibility,Withdrawal | 91.3 | 88.6 | 93.7 |
| 7 | Grimacing,Echopraxia/echolalia,Stereotypy,Verbigeration,Rigidity,Negativism,Withdrawal | 91.3 | 88.6 | 93.7 |
| 7 | Mutism,Grimacing,Echopraxia/echolalia,Mannerisms,Verbigeration,Rigidity,Waxy flexibility | 91.3 | 88.6 | 93.7 |
| 7 | Mutism,Grimacing,Echopraxia/echolalia,Stereotypy,Verbigeration,Rigidity,Waxy flexibility | 91.3 | 88.6 | 93.7 |
| 7 | Mutism,Grimacing,Mannerisms,Verbigeration,Negativism,Waxy flexibility,Withdrawal | 91.3 | 88.6 | 93.7 |
| 7 | Mutism,Posturing/catalepsy,Echopraxia/echolalia,Mannerisms,Verbigeration,Waxy flexibility,Withdrawal | 91.3 | 88.6 | 93.7 |
| 7 | Mutism,Posturing/catalepsy,Echopraxia/echolalia,Stereotypy,Mannerisms,Waxy flexibility,Withdrawal | 91.3 | 88.6 | 93.7 |
| 7 | Mutism,Posturing/catalepsy,Grimacing,Echopraxia/echolalia,Verbigeration,Waxy flexibility,Withdrawal | 91.3 | 88.6 | 93.7 |
| 7 | Posturing/catalepsy,Grimacing,Echopraxia/echolalia,Mannerisms,Verbigeration,Rigidity,Withdrawal | 91.3 | 88.6 | 93.7 |
| 7 | Posturing/catalepsy,Grimacing,Echopraxia/echolalia,Stereotypy,Verbigeration,Negativism,Withdrawal | 91.3 | 88.6 | 93.7 |
| 7 | Posturing/catalepsy,Grimacing,Mannerisms,Verbigeration,Rigidity,Waxy flexibility,Withdrawal | 91.3 | 88.6 | 93.7 |
| 7 | Posturing/catalepsy,Grimacing,Stereotypy,Verbigeration,Negativism,Waxy flexibility,Withdrawal | 91.3 | 88.6 | 93.7 |
| 7 | Excitement,Echopraxia/echolalia,Verbigeration,Rigidity,Negativism,Waxy flexibility,Withdrawal | 91.2 | 88.6 | 93.7 |
| 7 | Excitement,Grimacing,Stereotypy,Mannerisms,Rigidity,Negativism,Withdrawal | 91.2 | 88.6 | 93.7 |
| 7 | Excitement,Posturing/catalepsy,Grimacing,Mannerisms,Negativism,Waxy flexibility,Withdrawal | 91.2 | 88.6 | 93.7 |
| 7 | Mutism,Grimacing,Echopraxia/echolalia,Mannerisms,Verbigeration,Negativism,Withdrawal | 91.2 | 88.6 | 93.7 |
| 7 | Mutism,Posturing/catalepsy,Grimacing,Stereotypy,Verbigeration,Waxy flexibility,Withdrawal | 91.2 | 88.6 | 93.7 |
| 7 | Posturing/catalepsy,Grimacing,Echopraxia/echolalia,Mannerisms,Rigidity,Waxy flexibility,Withdrawal | 91.2 | 88.6 | 93.7 |
| 7 | Posturing/catalepsy,Grimacing,Stereotypy,Mannerisms,Rigidity,Waxy flexibility,Withdrawal | 91.2 | 88.6 | 93.7 |
| 7 | Echopraxia/echolalia,Mannerisms,Verbigeration,Rigidity,Negativism,Waxy flexibility,Withdrawal | 91.1 | 88.3 | 93.5 |
| 7 | Excitement,Echopraxia/echolalia,Mannerisms,Rigidity,Negativism,Waxy flexibility,Withdrawal | 91.0 | 88.3 | 93.5 |
| 7 | Excitement,Echopraxia/echolalia,Mannerisms,Verbigeration,Rigidity,Negativism,Withdrawal | 91.0 | 88.3 | 93.7 |
| 7 | Excitement,Grimacing,Mannerisms,Verbigeration,Rigidity,Negativism,Withdrawal | 91.0 | 88.3 | 93.5 |
| 7 | Excitement,Mannerisms,Verbigeration,Rigidity,Negativism,Waxy flexibility,Withdrawal | 91.0 | 88.3 | 93.5 |
| 7 | Excitement,Mutism,Echopraxia/echolalia,Stereotypy,Mannerisms,Waxy flexibility,Withdrawal | 91.0 | 88.3 | 93.7 |
| 7 | Excitement,Mutism,Grimacing,Echopraxia/echolalia,Mannerisms,Negativism,Withdrawal | 91.0 | 88.3 | 93.5 |
| 7 | Excitement,Mutism,Grimacing,Echopraxia/echolalia,Mannerisms,Verbigeration,Rigidity | 91.0 | 88.3 | 93.7 |
| 7 | Excitement,Mutism,Grimacing,Echopraxia/echolalia,Stereotypy,Mannerisms,Rigidity | 91.0 | 88.3 | 93.5 |
| 7 | Excitement,Mutism,Grimacing,Echopraxia/echolalia,Stereotypy,Negativism,Waxy flexibility | 91.0 | 88.3 | 93.5 |
| 7 | Excitement,Mutism,Grimacing,Stereotypy,Mannerisms,Verbigeration,Rigidity | 91.0 | 88.3 | 93.5 |
| 7 | Grimacing,Echopraxia/echolalia,Mannerisms,Verbigeration,Rigidity,Negativism,Withdrawal | 91.0 | 88.3 | 93.7 |
| 7 | Grimacing,Mannerisms,Verbigeration,Rigidity,Negativism,Waxy flexibility,Withdrawal | 91.0 | 88.3 | 93.5 |
| 7 | Mutism,Grimacing,Echopraxia/echolalia,Stereotypy,Mannerisms,Negativism,Waxy flexibility | 91.0 | 88.3 | 93.5 |
| 7 | Mutism,Grimacing,Echopraxia/echolalia,Verbigeration,Negativism,Waxy flexibility,Withdrawal | 91.0 | 88.3 | 93.5 |
| 7 | Mutism,Posturing/catalepsy,Grimacing,Stereotypy,Mannerisms,Verbigeration,Withdrawal | 91.0 | 88.3 | 93.5 |
| 7 | Mutism,Posturing/catalepsy,Stereotypy,Mannerisms,Verbigeration,Waxy flexibility,Withdrawal | 91.0 | 88.3 | 93.7 |
| 7 | Posturing/catalepsy,Echopraxia/echolalia,Mannerisms,Verbigeration,Negativism,Waxy flexibility,Withdrawal | 91.0 | 88.3 | 93.5 |
| 7 | Posturing/catalepsy,Echopraxia/echolalia,Mannerisms,Verbigeration,Rigidity,Waxy flexibility,Withdrawal | 91.0 | 88.3 | 93.5 |
| 7 | Posturing/catalepsy,Echopraxia/echolalia,Stereotypy,Verbigeration,Negativism,Waxy flexibility,Withdrawal | 91.0 | 88.3 | 93.7 |
| 7 | Posturing/catalepsy,Grimacing,Echopraxia/echolalia,Mannerisms,Negativism,Waxy flexibility,Withdrawal | 91.0 | 88.3 | 93.5 |
| 7 | Posturing/catalepsy,Grimacing,Echopraxia/echolalia,Stereotypy,Mannerisms,Rigidity,Withdrawal | 91.0 | 88.3 | 93.5 |
| 7 | Posturing/catalepsy,Grimacing,Echopraxia/echolalia,Stereotypy,Negativism,Waxy flexibility,Withdrawal | 91.0 | 88.3 | 93.5 |
| 7 | Posturing/catalepsy,Grimacing,Mannerisms,Verbigeration,Negativism,Waxy flexibility,Withdrawal | 91.0 | 88.3 | 93.5 |
| 7 | Posturing/catalepsy,Grimacing,Stereotypy,Mannerisms,Verbigeration,Rigidity,Withdrawal | 91.0 | 88.3 | 93.5 |
| 7 | Posturing/catalepsy,Stereotypy,Mannerisms,Verbigeration,Rigidity,Waxy flexibility,Withdrawal | 91.0 | 88.3 | 93.5 |
| 7 | Excitement,Grimacing,Mannerisms,Rigidity,Negativism,Waxy flexibility,Withdrawal | 90.8 | 88.1 | 93.3 |
| 7 | Excitement,Grimacing,Verbigeration,Rigidity,Negativism,Waxy flexibility,Withdrawal | 90.8 | 88.1 | 93.5 |
| 7 | Excitement,Mutism,Echopraxia/echolalia,Stereotypy,Mannerisms,Negativism,Waxy flexibility | 90.8 | 87.9 | 93.5 |
| 7 | Excitement,Mutism,Grimacing,Stereotypy,Verbigeration,Negativism,Waxy flexibility | 90.8 | 88.1 | 93.5 |
| 7 | Excitement,Mutism,Grimacing,Stereotypy,Verbigeration,Waxy flexibility,Withdrawal | 90.8 | 88.1 | 93.5 |
| 7 | Excitement,Mutism,Posturing/catalepsy,Grimacing,Echopraxia/echolalia,Verbigeration,Waxy flexibility | 90.8 | 88.1 | 93.3 |
| 7 | Excitement,Mutism,Stereotypy,Mannerisms,Verbigeration,Negativism,Waxy flexibility | 90.8 | 88.1 | 93.5 |
| 7 | Excitement,Mutism,Stereotypy,Mannerisms,Verbigeration,Waxy flexibility,Withdrawal | 90.8 | 88.1 | 93.5 |
| 7 | Mutism,Echopraxia/echolalia,Stereotypy,Mannerisms,Verbigeration,Negativism,Waxy flexibility | 90.8 | 88.1 | 93.3 |
| 7 | Mutism,Echopraxia/echolalia,Stereotypy,Mannerisms,Verbigeration,Rigidity,Waxy flexibility | 90.8 | 88.1 | 93.3 |
| 7 | Mutism,Posturing/catalepsy,Grimacing,Echopraxia/echolalia,Stereotypy,Verbigeration,Withdrawal | 90.8 | 88.1 | 93.5 |
| 7 | Posturing/catalepsy,Echopraxia/echolalia,Stereotypy,Mannerisms,Rigidity,Waxy flexibility,Withdrawal | 90.8 | 88.1 | 93.5 |
| 7 | Posturing/catalepsy,Grimacing,Echopraxia/echolalia,Stereotypy,Rigidity,Waxy flexibility,Withdrawal | 90.8 | 87.9 | 93.3 |
| 7 | Posturing/catalepsy,Grimacing,Echopraxia/echolalia,Verbigeration,Rigidity,Waxy flexibility,Withdrawal | 90.8 | 88.1 | 93.5 |
| 7 | Posturing/catalepsy,Grimacing,Stereotypy,Verbigeration,Rigidity,Waxy flexibility,Withdrawal | 90.8 | 88.1 | 93.5 |
| 7 | Excitement,Echopraxia/echolalia,Stereotypy,Mannerisms,Rigidity,Waxy flexibility,Withdrawal | 90.6 | 87.9 | 93.3 |
| 7 | Excitement,Echopraxia/echolalia,Stereotypy,Verbigeration,Rigidity,Waxy flexibility,Withdrawal | 90.6 | 87.9 | 93.3 |
| 7 | Excitement,Mutism,Grimacing,Echopraxia/echolalia,Verbigeration,Waxy flexibility,Withdrawal | 90.6 | 87.7 | 93.3 |
| 7 | Excitement,Mutism,Posturing/catalepsy,Echopraxia/echolalia,Stereotypy,Verbigeration,Waxy flexibility | 90.6 | 87.7 | 93.3 |
| 7 | Excitement,Mutism,Posturing/catalepsy,Grimacing,Echopraxia/echolalia,Stereotypy,Waxy flexibility | 90.6 | 87.9 | 93.3 |
| 7 | Excitement,Mutism,Posturing/catalepsy,Grimacing,Stereotypy,Verbigeration,Waxy flexibility | 90.6 | 87.7 | 93.3 |
| 7 | Excitement,Posturing/catalepsy,Grimacing,Echopraxia/echolalia,Mannerisms,Waxy flexibility,Withdrawal | 90.6 | 87.9 | 93.3 |
| 7 | Excitement,Posturing/catalepsy,Grimacing,Echopraxia/echolalia,Verbigeration,Waxy flexibility,Withdrawal | 90.6 | 87.9 | 93.3 |
| 7 | Grimacing,Echopraxia/echolalia,Verbigeration,Rigidity,Negativism,Waxy flexibility,Withdrawal | 90.6 | 87.9 | 93.3 |
| 7 | Mutism,Posturing/catalepsy,Echopraxia/echolalia,Stereotypy,Mannerisms,Verbigeration,Withdrawal | 90.6 | 87.9 | 93.3 |
| 7 | Mutism,Posturing/catalepsy,Echopraxia/echolalia,Stereotypy,Verbigeration,Waxy flexibility,Withdrawal | 90.6 | 87.9 | 93.3 |
| 7 | Posturing/catalepsy,Echopraxia/echolalia,Stereotypy,Mannerisms,Verbigeration,Rigidity,Withdrawal | 90.6 | 87.9 | 93.3 |
| 7 | Posturing/catalepsy,Grimacing,Echopraxia/echolalia,Stereotypy,Verbigeration,Rigidity,Withdrawal | 90.6 | 87.7 | 93.3 |
| 7 | Excitement,Mutism,Grimacing,Echopraxia/echolalia,Mannerisms,Waxy flexibility,Withdrawal | 90.4 | 87.4 | 93.0 |
| 7 | Excitement,Mutism,Grimacing,Echopraxia/echolalia,Verbigeration,Negativism,Waxy flexibility | 90.4 | 87.4 | 93.0 |
| 7 | Excitement,Mutism,Posturing/catalepsy,Grimacing,Echopraxia/echolalia,Mannerisms,Waxy flexibility | 90.4 | 87.7 | 93.0 |
| 7 | Excitement,Stereotypy,Mannerisms,Verbigeration,Rigidity,Waxy flexibility,Withdrawal | 90.4 | 87.4 | 93.0 |
| 7 | Mutism,Grimacing,Echopraxia/echolalia,Stereotypy,Mannerisms,Waxy flexibility,Withdrawal | 90.4 | 87.7 | 93.0 |
| 7 | Mutism,Grimacing,Stereotypy,Mannerisms,Verbigeration,Negativism,Waxy flexibility | 90.4 | 87.7 | 93.0 |
| 7 | Posturing/catalepsy,Echopraxia/echolalia,Stereotypy,Verbigeration,Rigidity,Waxy flexibility,Withdrawal | 90.4 | 87.7 | 93.0 |
| 7 | Posturing/catalepsy,Grimacing,Echopraxia/echolalia,Verbigeration,Negativism,Waxy flexibility,Withdrawal | 90.4 | 87.4 | 93.0 |
| 7 | Excitement,Grimacing,Echopraxia/echolalia,Stereotypy,Rigidity,Waxy flexibility,Withdrawal | 90.3 | 87.7 | 93.0 |
| 7 | Excitement,Mutism,Grimacing,Stereotypy,Mannerisms,Waxy flexibility,Withdrawal | 90.3 | 87.4 | 92.8 |
| 7 | Excitement,Mutism,Posturing/catalepsy,Grimacing,Mannerisms,Verbigeration,Waxy flexibility | 90.3 | 87.7 | 93.0 |
| 7 | Excitement,Grimacing,Echopraxia/echolalia,Verbigeration,Rigidity,Waxy flexibility,Withdrawal | 90.1 | 87.2 | 92.8 |
| 7 | Excitement,Mutism,Echopraxia/echolalia,Stereotypy,Mannerisms,Verbigeration,Negativism | 90.1 | 87.2 | 92.8 |
| 7 | Excitement,Mutism,Grimacing,Echopraxia/echolalia,Mannerisms,Negativism,Waxy flexibility | 90.1 | 87.2 | 92.8 |
| 7 | Excitement,Mutism,Grimacing,Echopraxia/echolalia,Stereotypy,Verbigeration,Negativism | 90.1 | 87.2 | 92.8 |
| 7 | Excitement,Mutism,Grimacing,Stereotypy,Mannerisms,Negativism,Waxy flexibility | 90.1 | 87.4 | 92.8 |
| 7 | Excitement,Mutism,Posturing/catalepsy,Echopraxia/echolalia,Mannerisms,Verbigeration,Waxy flexibility | 90.1 | 87.2 | 92.8 |
| 7 | Excitement,Posturing/catalepsy,Echopraxia/echolalia,Mannerisms,Verbigeration,Waxy flexibility,Withdrawal | 90.1 | 87.2 | 92.8 |
| 7 | Excitement,Posturing/catalepsy,Grimacing,Echopraxia/echolalia,Mannerisms,Verbigeration,Withdrawal | 90.1 | 87.2 | 92.8 |
| 7 | Excitement,Posturing/catalepsy,Grimacing,Echopraxia/echolalia,Stereotypy,Waxy flexibility,Withdrawal | 90.1 | 87.2 | 92.8 |
| 7 | Excitement,Posturing/catalepsy,Grimacing,Mannerisms,Verbigeration,Waxy flexibility,Withdrawal | 90.1 | 87.2 | 92.8 |
| 7 | Mutism,Grimacing,Echopraxia/echolalia,Mannerisms,Verbigeration,Negativism,Waxy flexibility | 90.1 | 87.2 | 92.8 |
| 7 | Mutism,Grimacing,Echopraxia/echolalia,Stereotypy,Verbigeration,Negativism,Waxy flexibility | 90.1 | 87.2 | 92.8 |
| 7 | Excitement,Grimacing,Stereotypy,Mannerisms,Rigidity,Waxy flexibility,Withdrawal | 89.9 | 87.0 | 92.6 |
| 7 | Excitement,Grimacing,Stereotypy,Verbigeration,Rigidity,Waxy flexibility,Withdrawal | 89.9 | 87.0 | 92.6 |
| 7 | Excitement,Mutism,Echopraxia/echolalia,Mannerisms,Verbigeration,Waxy flexibility,Withdrawal | 89.9 | 87.0 | 92.6 |
| 7 | Excitement,Mutism,Grimacing,Mannerisms,Verbigeration,Waxy flexibility,Withdrawal | 89.9 | 87.0 | 92.6 |
| 7 | Excitement,Mutism,Posturing/catalepsy,Echopraxia/echolalia,Stereotypy,Mannerisms,Waxy flexibility | 89.9 | 87.0 | 92.6 |
| 7 | Excitement,Mutism,Posturing/catalepsy,Grimacing,Echopraxia/echolalia,Stereotypy,Verbigeration | 89.9 | 87.0 | 92.6 |
| 7 | Excitement,Mutism,Posturing/catalepsy,Stereotypy,Mannerisms,Verbigeration,Waxy flexibility | 89.9 | 87.0 | 92.6 |
| 7 | Excitement,Posturing/catalepsy,Echopraxia/echolalia,Stereotypy,Verbigeration,Waxy flexibility,Withdrawal | 89.9 | 87.0 | 92.6 |
| 7 | Excitement,Posturing/catalepsy,Grimacing,Stereotypy,Verbigeration,Waxy flexibility,Withdrawal | 89.9 | 87.0 | 92.6 |
| 7 | Excitement,Posturing/catalepsy,Stereotypy,Mannerisms,Verbigeration,Waxy flexibility,Withdrawal | 89.9 | 87.0 | 92.6 |
| 7 | Grimacing,Echopraxia/echolalia,Stereotypy,Mannerisms,Rigidity,Waxy flexibility,Withdrawal | 89.9 | 87.2 | 92.6 |
| 7 | Mutism,Grimacing,Echopraxia/echolalia,Stereotypy,Mannerisms,Verbigeration,Negativism | 89.9 | 87.0 | 92.6 |
| 7 | Mutism,Grimacing,Echopraxia/echolalia,Stereotypy,Mannerisms,Verbigeration,Rigidity | 89.9 | 87.0 | 92.6 |
| 7 | Excitement,Echopraxia/echolalia,Mannerisms,Verbigeration,Rigidity,Waxy flexibility,Withdrawal | 89.7 | 86.8 | 92.4 |
| 7 | Excitement,Grimacing,Echopraxia/echolalia,Mannerisms,Rigidity,Waxy flexibility,Withdrawal | 89.7 | 86.8 | 92.4 |
| 7 | Excitement,Grimacing,Mannerisms,Verbigeration,Rigidity,Waxy flexibility,Withdrawal | 89.7 | 86.8 | 92.4 |
| 7 | Excitement,Mutism,Grimacing,Echopraxia/echolalia,Stereotypy,Mannerisms,Negativism | 89.7 | 86.8 | 92.4 |
| 7 | Excitement,Mutism,Grimacing,Mannerisms,Verbigeration,Negativism,Waxy flexibility | 89.7 | 86.8 | 92.4 |
| 7 | Excitement,Mutism,Grimacing,Stereotypy,Mannerisms,Verbigeration,Negativism | 89.7 | 86.8 | 92.4 |
| 7 | Excitement,Mutism,Posturing/catalepsy,Grimacing,Echopraxia/echolalia,Mannerisms,Verbigeration | 89.7 | 86.8 | 92.4 |
| 7 | Excitement,Mutism,Posturing/catalepsy,Grimacing,Stereotypy,Mannerisms,Waxy flexibility | 89.7 | 86.8 | 92.4 |
| 7 | Excitement,Posturing/catalepsy,Echopraxia/echolalia,Stereotypy,Mannerisms,Waxy flexibility,Withdrawal | 89.7 | 86.8 | 92.4 |
| 7 | Excitement,Posturing/catalepsy,Grimacing,Echopraxia/echolalia,Stereotypy,Mannerisms,Withdrawal | 89.7 | 86.8 | 92.4 |
| 7 | Excitement,Posturing/catalepsy,Grimacing,Echopraxia/echolalia,Verbigeration,Rigidity,Negativism | 89.7 | 86.8 | 92.4 |
| 7 | Excitement,Posturing/catalepsy,Grimacing,Stereotypy,Mannerisms,Waxy flexibility,Withdrawal | 89.7 | 86.8 | 92.4 |
| 7 | Mutism,Grimacing,Echopraxia/echolalia,Stereotypy,Verbigeration,Waxy flexibility,Withdrawal | 89.7 | 86.8 | 92.4 |
| 7 | Posturing/catalepsy,Grimacing,Echopraxia/echolalia,Stereotypy,Mannerisms,Waxy flexibility,Withdrawal | 89.7 | 86.8 | 92.4 |
| 7 | Excitement,Mutism,Echopraxia/echolalia,Mannerisms,Verbigeration,Negativism,Waxy flexibility | 89.6 | 86.8 | 92.4 |
| 7 | Excitement,Mutism,Echopraxia/echolalia,Stereotypy,Mannerisms,Verbigeration,Withdrawal | 89.5 | 86.5 | 92.2 |
| 7 | Excitement,Mutism,Grimacing,Echopraxia/echolalia,Stereotypy,Verbigeration,Withdrawal | 89.5 | 86.5 | 92.4 |
| 7 | Excitement,Mutism,Posturing/catalepsy,Echopraxia/echolalia,Stereotypy,Mannerisms,Verbigeration | 89.5 | 86.5 | 92.2 |
| 7 | Excitement,Mutism,Posturing/catalepsy,Grimacing,Echopraxia/echolalia,Stereotypy,Mannerisms | 89.5 | 86.5 | 92.2 |
| 7 | Excitement,Mutism,Posturing/catalepsy,Grimacing,Stereotypy,Mannerisms,Verbigeration | 89.5 | 86.5 | 92.4 |
| 7 | Excitement,Posturing/catalepsy,Grimacing,Echopraxia/echolalia,Rigidity,Negativism,Waxy flexibility | 89.5 | 86.5 | 92.4 |
| 7 | Excitement,Posturing/catalepsy,Grimacing,Echopraxia/echolalia,Stereotypy,Verbigeration,Withdrawal | 89.5 | 86.5 | 92.2 |
| 7 | Excitement,Posturing/catalepsy,Grimacing,Mannerisms,Verbigeration,Rigidity,Negativism | 89.5 | 86.5 | 92.2 |
| 7 | Excitement,Posturing/catalepsy,Grimacing,Stereotypy,Mannerisms,Verbigeration,Withdrawal | 89.5 | 86.5 | 92.2 |
| 7 | Excitement,Posturing/catalepsy,Grimacing,Verbigeration,Rigidity,Negativism,Waxy flexibility | 89.5 | 86.5 | 92.2 |
| 7 | Mutism,Grimacing,Echopraxia/echolalia,Mannerisms,Verbigeration,Waxy flexibility,Withdrawal | 89.5 | 86.5 | 92.4 |
| 7 | Excitement,Posturing/catalepsy,Grimacing,Stereotypy,Verbigeration,Rigidity,Negativism | 89.4 | 86.5 | 92.2 |
| 7 | Grimacing,Stereotypy,Mannerisms,Verbigeration,Rigidity,Waxy flexibility,Withdrawal | 89.4 | 86.5 | 92.2 |
| 7 | Mutism,Grimacing,Stereotypy,Mannerisms,Verbigeration,Waxy flexibility,Withdrawal | 89.4 | 86.5 | 92.2 |
| 7 | Excitement,Posturing/catalepsy,Grimacing,Echopraxia/echolalia,Mannerisms,Rigidity,Negativism | 89.3 | 86.3 | 92.2 |
| 7 | Excitement,Posturing/catalepsy,Grimacing,Echopraxia/echolalia,Stereotypy,Rigidity,Negativism | 89.3 | 86.3 | 91.9 |
| 7 | Grimacing,Echopraxia/echolalia,Stereotypy,Mannerisms,Negativism,Waxy flexibility,Withdrawal | 89.3 | 86.3 | 91.9 |
| 7 | Mutism,Echopraxia/echolalia,Stereotypy,Mannerisms,Verbigeration,Waxy flexibility,Withdrawal | 89.3 | 86.3 | 92.2 |
| 7 | Posturing/catalepsy,Grimacing,Stereotypy,Mannerisms,Verbigeration,Rigidity,Negativism | 89.3 | 86.3 | 92.2 |
| 7 | Excitement,Echopraxia/echolalia,Stereotypy,Mannerisms,Verbigeration,Rigidity,Withdrawal | 89.2 | 86.3 | 92.2 |
| 7 | Excitement,Posturing/catalepsy,Echopraxia/echolalia,Stereotypy,Mannerisms,Verbigeration,Withdrawal | 89.2 | 86.3 | 92.2 |
| 7 | Excitement,Posturing/catalepsy,Echopraxia/echolalia,Stereotypy,Verbigeration,Rigidity,Negativism | 89.2 | 86.1 | 92.2 |
| 7 | Excitement,Posturing/catalepsy,Stereotypy,Verbigeration,Rigidity,Negativism,Waxy flexibility | 89.2 | 86.3 | 91.9 |
| 7 | Grimacing,Echopraxia/echolalia,Stereotypy,Verbigeration,Rigidity,Waxy flexibility,Withdrawal | 89.2 | 86.3 | 91.9 |
| 7 | Mutism,Posturing/catalepsy,Grimacing,Echopraxia/echolalia,Mannerisms,Verbigeration,Waxy flexibility | 89.2 | 86.3 | 91.9 |
| 7 | Posturing/catalepsy,Grimacing,Echopraxia/echolalia,Mannerisms,Verbigeration,Waxy flexibility,Withdrawal | 89.2 | 86.3 | 91.9 |
| 7 | Posturing/catalepsy,Grimacing,Stereotypy,Mannerisms,Verbigeration,Waxy flexibility,Withdrawal | 89.2 | 86.3 | 91.9 |
| 7 | Echopraxia/echolalia,Stereotypy,Mannerisms,Verbigeration,Negativism,Waxy flexibility,Withdrawal | 89.0 | 86.1 | 91.9 |
| 7 | Echopraxia/echolalia,Stereotypy,Mannerisms,Verbigeration,Rigidity,Waxy flexibility,Withdrawal | 89.0 | 85.9 | 91.7 |
| 7 | Excitement,Grimacing,Echopraxia/echolalia,Stereotypy,Mannerisms,Rigidity,Withdrawal | 89.0 | 86.1 | 91.7 |
| 7 | Excitement,Mutism,Grimacing,Echopraxia/echolalia,Mannerisms,Verbigeration,Negativism | 89.0 | 86.1 | 91.7 |
| 7 | Excitement,Mutism,Grimacing,Echopraxia/echolalia,Stereotypy,Mannerisms,Withdrawal | 89.0 | 86.1 | 91.9 |
| 7 | Excitement,Posturing/catalepsy,Echopraxia/echolalia,Mannerisms,Verbigeration,Rigidity,Negativism | 89.0 | 86.1 | 91.9 |
| 7 | Excitement,Posturing/catalepsy,Echopraxia/echolalia,Stereotypy,Rigidity,Negativism,Waxy flexibility | 89.0 | 86.1 | 91.7 |
| 7 | Excitement,Posturing/catalepsy,Echopraxia/echolalia,Verbigeration,Rigidity,Negativism,Waxy flexibility | 89.0 | 86.1 | 91.9 |
| 7 | Excitement,Posturing/catalepsy,Grimacing,Stereotypy,Rigidity,Negativism,Waxy flexibility | 89.0 | 86.1 | 91.9 |
| 7 | Excitement,Posturing/catalepsy,Stereotypy,Mannerisms,Verbigeration,Rigidity,Negativism | 89.0 | 86.1 | 91.7 |
| 7 | Grimacing,Echopraxia/echolalia,Mannerisms,Verbigeration,Rigidity,Waxy flexibility,Withdrawal | 89.0 | 86.1 | 91.9 |
| 7 | Mutism,Posturing/catalepsy,Grimacing,Echopraxia/echolalia,Stereotypy,Mannerisms,Waxy flexibility | 89.0 | 85.9 | 91.7 |
| 7 | Mutism,Posturing/catalepsy,Grimacing,Echopraxia/echolalia,Stereotypy,Verbigeration,Waxy flexibility | 89.0 | 86.1 | 91.9 |
| 7 | Posturing/catalepsy,Grimacing,Echopraxia/echolalia,Mannerisms,Rigidity,Negativism,Waxy flexibility | 89.0 | 86.1 | 91.9 |
| 7 | Posturing/catalepsy,Grimacing,Echopraxia/echolalia,Mannerisms,Verbigeration,Rigidity,Negativism | 89.0 | 86.1 | 91.9 |
| 7 | Posturing/catalepsy,Grimacing,Echopraxia/echolalia,Stereotypy,Mannerisms,Rigidity,Negativism | 89.0 | 86.1 | 91.9 |
| 7 | Posturing/catalepsy,Grimacing,Echopraxia/echolalia,Stereotypy,Mannerisms,Verbigeration,Withdrawal | 89.0 | 86.1 | 91.7 |
| 7 | Posturing/catalepsy,Grimacing,Stereotypy,Mannerisms,Rigidity,Negativism,Waxy flexibility | 89.0 | 85.9 | 91.7 |
| 7 | Posturing/catalepsy,Grimacing,Stereotypy,Verbigeration,Rigidity,Negativism,Waxy flexibility | 89.0 | 86.1 | 91.7 |
| 7 | Posturing/catalepsy,Stereotypy,Mannerisms,Verbigeration,Rigidity,Negativism,Waxy flexibility | 89.0 | 86.1 | 91.7 |
| 7 | Excitement,Echopraxia/echolalia,Stereotypy,Mannerisms,Negativism,Waxy flexibility,Withdrawal | 88.8 | 85.7 | 91.7 |
| 7 | Excitement,Echopraxia/echolalia,Stereotypy,Verbigeration,Negativism,Waxy flexibility,Withdrawal | 88.8 | 85.9 | 91.7 |
| 7 | Excitement,Grimacing,Echopraxia/echolalia,Stereotypy,Verbigeration,Rigidity,Withdrawal | 88.8 | 85.9 | 91.7 |
| 7 | Excitement,Mutism,Grimacing,Stereotypy,Mannerisms,Verbigeration,Withdrawal | 88.8 | 85.9 | 91.7 |
| 7 | Excitement,Posturing/catalepsy,Echopraxia/echolalia,Stereotypy,Mannerisms,Rigidity,Negativism | 88.8 | 85.7 | 91.7 |
| 7 | Excitement,Posturing/catalepsy,Grimacing,Mannerisms,Rigidity,Negativism,Waxy flexibility | 88.8 | 85.9 | 91.7 |
| 7 | Excitement,Posturing/catalepsy,Grimacing,Stereotypy,Mannerisms,Rigidity,Negativism | 88.8 | 85.9 | 91.7 |
| 7 | Excitement,Posturing/catalepsy,Mannerisms,Verbigeration,Rigidity,Negativism,Waxy flexibility | 88.8 | 85.9 | 91.7 |
| 7 | Mutism,Posturing/catalepsy,Grimacing,Stereotypy,Mannerisms,Verbigeration,Waxy flexibility | 88.8 | 85.9 | 91.7 |
| 7 | Posturing/catalepsy,Echopraxia/echolalia,Stereotypy,Mannerisms,Rigidity,Negativism,Waxy flexibility | 88.8 | 85.9 | 91.7 |
| 7 | Posturing/catalepsy,Echopraxia/echolalia,Stereotypy,Mannerisms,Verbigeration,Rigidity,Negativism | 88.8 | 85.9 | 91.7 |
| 7 | Posturing/catalepsy,Grimacing,Echopraxia/echolalia,Stereotypy,Verbigeration,Waxy flexibility,Withdrawal | 88.8 | 85.7 | 91.7 |
| 7 | Posturing/catalepsy,Grimacing,Mannerisms,Verbigeration,Rigidity,Negativism,Waxy flexibility | 88.8 | 85.9 | 91.7 |
| 7 | Excitement,Grimacing,Echopraxia/echolalia,Mannerisms,Verbigeration,Rigidity,Withdrawal | 88.6 | 85.4 | 91.5 |
| 7 | Excitement,Grimacing,Echopraxia/echolalia,Stereotypy,Negativism,Waxy flexibility,Withdrawal | 88.6 | 85.7 | 91.5 |
| 7 | Excitement,Grimacing,Stereotypy,Mannerisms,Verbigeration,Rigidity,Withdrawal | 88.6 | 85.4 | 91.5 |
| 7 | Excitement,Posturing/catalepsy,Echopraxia/echolalia,Mannerisms,Rigidity,Negativism,Waxy flexibility | 88.6 | 85.7 | 91.5 |
| 7 | Posturing/catalepsy,Echopraxia/echolalia,Mannerisms,Verbigeration,Rigidity,Negativism,Waxy flexibility | 88.6 | 85.4 | 91.5 |
| 7 | Posturing/catalepsy,Grimacing,Echopraxia/echolalia,Stereotypy,Verbigeration,Rigidity,Negativism | 88.6 | 85.7 | 91.5 |
| 7 | Posturing/catalepsy,Grimacing,Echopraxia/echolalia,Verbigeration,Rigidity,Negativism,Waxy flexibility | 88.6 | 85.7 | 91.5 |
| 7 | Mutism,Posturing/catalepsy,Grimacing,Echopraxia/echolalia,Stereotypy,Mannerisms,Verbigeration | 88.5 | 85.4 | 91.5 |
| 7 | Mutism,Grimacing,Echopraxia/echolalia,Stereotypy,Mannerisms,Verbigeration,Withdrawal | 88.4 | 85.4 | 91.3 |
| 7 | Excitement,Mutism,Grimacing,Echopraxia/echolalia,Mannerisms,Verbigeration,Withdrawal | 88.3 | 85.2 | 91.3 |
| 7 | Excitement,Posturing/catalepsy,Stereotypy,Mannerisms,Rigidity,Negativism,Waxy flexibility | 88.3 | 85.4 | 91.3 |
| 7 | Excitement,Stereotypy,Mannerisms,Verbigeration,Negativism,Waxy flexibility,Withdrawal | 88.3 | 85.2 | 91.3 |
| 7 | Grimacing,Echopraxia/echolalia,Stereotypy,Mannerisms,Verbigeration,Rigidity,Withdrawal | 88.3 | 85.2 | 91.3 |
| 7 | Grimacing,Stereotypy,Mannerisms,Verbigeration,Negativism,Waxy flexibility,Withdrawal | 88.3 | 85.2 | 91.3 |
| 7 | Mutism,Posturing/catalepsy,Echopraxia/echolalia,Stereotypy,Mannerisms,Verbigeration,Waxy flexibility | 88.3 | 85.2 | 91.3 |
| 7 | Posturing/catalepsy,Echopraxia/echolalia,Stereotypy,Mannerisms,Verbigeration,Waxy flexibility,Withdrawal | 88.3 | 85.2 | 91.3 |
| 7 | Posturing/catalepsy,Echopraxia/echolalia,Stereotypy,Verbigeration,Rigidity,Negativism,Waxy flexibility | 88.3 | 85.2 | 91.3 |
| 7 | Posturing/catalepsy,Grimacing,Echopraxia/echolalia,Stereotypy,Rigidity,Negativism,Waxy flexibility | 88.3 | 85.2 | 91.3 |
| 7 | Excitement,Mutism,Grimacing,Echopraxia/echolalia,Stereotypy,Verbigeration,Waxy flexibility | 88.1 | 85.0 | 91.0 |
| 7 | Grimacing,Echopraxia/echolalia,Stereotypy,Mannerisms,Verbigeration,Negativism,Withdrawal | 88.1 | 85.0 | 91.0 |
| 7 | Grimacing,Echopraxia/echolalia,Stereotypy,Verbigeration,Negativism,Waxy flexibility,Withdrawal | 88.1 | 85.0 | 91.0 |
| 7 | Excitement,Echopraxia/echolalia,Stereotypy,Mannerisms,Verbigeration,Negativism,Withdrawal | 87.9 | 84.8 | 90.8 |
| 7 | Excitement,Grimacing,Stereotypy,Verbigeration,Negativism,Waxy flexibility,Withdrawal | 87.9 | 84.8 | 90.8 |
| 7 | Excitement,Mutism,Grimacing,Echopraxia/echolalia,Stereotypy,Mannerisms,Waxy flexibility | 87.9 | 84.8 | 90.8 |
| 7 | Excitement,Grimacing,Echopraxia/echolalia,Mannerisms,Negativism,Waxy flexibility,Withdrawal | 87.7 | 84.5 | 90.6 |
| 7 | Excitement,Grimacing,Echopraxia/echolalia,Stereotypy,Verbigeration,Negativism,Withdrawal | 87.7 | 84.5 | 90.6 |
| 7 | Excitement,Grimacing,Echopraxia/echolalia,Verbigeration,Negativism,Waxy flexibility,Withdrawal | 87.7 | 84.5 | 90.6 |
| 7 | Excitement,Grimacing,Stereotypy,Mannerisms,Negativism,Waxy flexibility,Withdrawal | 87.7 | 84.5 | 90.6 |
| 7 | Excitement,Mutism,Echopraxia/echolalia,Stereotypy,Mannerisms,Verbigeration,Waxy flexibility | 87.7 | 84.8 | 90.8 |
| 7 | Excitement,Grimacing,Echopraxia/echolalia,Stereotypy,Mannerisms,Negativism,Withdrawal | 87.6 | 84.5 | 90.6 |
| 7 | Excitement,Echopraxia/echolalia,Mannerisms,Verbigeration,Negativism,Waxy flexibility,Withdrawal | 87.2 | 84.1 | 90.4 |
| 7 | Excitement,Grimacing,Stereotypy,Mannerisms,Verbigeration,Negativism,Withdrawal | 87.2 | 84.1 | 90.4 |
| 7 | Excitement,Mutism,Grimacing,Stereotypy,Mannerisms,Verbigeration,Waxy flexibility | 87.2 | 84.1 | 90.4 |
| 7 | Grimacing,Echopraxia/echolalia,Mannerisms,Verbigeration,Negativism,Waxy flexibility,Withdrawal | 87.2 | 84.1 | 90.4 |
| 7 | Excitement,Grimacing,Mannerisms,Verbigeration,Negativism,Waxy flexibility,Withdrawal | 87.0 | 83.9 | 90.1 |
| 7 | Excitement,Mutism,Grimacing,Echopraxia/echolalia,Mannerisms,Verbigeration,Waxy flexibility | 87.0 | 83.9 | 89.9 |
| 7 | Mutism,Grimacing,Echopraxia/echolalia,Stereotypy,Mannerisms,Verbigeration,Waxy flexibility | 86.8 | 83.6 | 89.9 |
| 7 | Excitement,Posturing/catalepsy,Grimacing,Echopraxia/echolalia,Verbigeration,Negativism,Waxy flexibility | 86.6 | 83.2 | 89.7 |
| 7 | Excitement,Posturing/catalepsy,Grimacing,Echopraxia/echolalia,Verbigeration,Rigidity,Waxy flexibility | 86.6 | 83.4 | 89.7 |
| 7 | Excitement,Posturing/catalepsy,Grimacing,Stereotypy,Verbigeration,Negativism,Waxy flexibility | 86.6 | 83.4 | 89.7 |
| 7 | Excitement,Grimacing,Echopraxia/echolalia,Mannerisms,Verbigeration,Negativism,Withdrawal | 86.5 | 83.2 | 89.7 |
| 7 | Excitement,Posturing/catalepsy,Grimacing,Echopraxia/echolalia,Stereotypy,Negativism,Waxy flexibility | 86.3 | 83.0 | 89.5 |
| 7 | Excitement,Posturing/catalepsy,Grimacing,Mannerisms,Verbigeration,Negativism,Waxy flexibility | 86.3 | 83.2 | 89.5 |
| 7 | Posturing/catalepsy,Grimacing,Echopraxia/echolalia,Stereotypy,Mannerisms,Negativism,Waxy flexibility | 86.3 | 83.0 | 89.5 |
| 7 | Excitement,Grimacing,Echopraxia/echolalia,Stereotypy,Mannerisms,Waxy flexibility,Withdrawal | 86.1 | 82.7 | 89.2 |
| 7 | Excitement,Posturing/catalepsy,Grimacing,Echopraxia/echolalia,Mannerisms,Negativism,Waxy flexibility | 86.1 | 82.7 | 89.2 |
| 7 | Excitement,Posturing/catalepsy,Grimacing,Echopraxia/echolalia,Mannerisms,Verbigeration,Negativism | 86.1 | 82.7 | 89.2 |
| 7 | Excitement,Posturing/catalepsy,Grimacing,Echopraxia/echolalia,Stereotypy,Verbigeration,Negativism | 86.1 | 82.7 | 89.2 |
| 7 | Excitement,Posturing/catalepsy,Grimacing,Stereotypy,Mannerisms,Verbigeration,Negativism | 86.1 | 82.7 | 89.2 |
| 7 | Posturing/catalepsy,Grimacing,Stereotypy,Mannerisms,Verbigeration,Negativism,Waxy flexibility | 86.1 | 83.0 | 89.2 |
| 7 | Excitement,Echopraxia/echolalia,Stereotypy,Mannerisms,Verbigeration,Waxy flexibility,Withdrawal | 85.9 | 82.5 | 89.0 |
| 7 | Excitement,Grimacing,Echopraxia/echolalia,Stereotypy,Rigidity,Negativism,Waxy flexibility | 85.9 | 82.5 | 89.0 |
| 7 | Excitement,Grimacing,Echopraxia/echolalia,Stereotypy,Verbigeration,Waxy flexibility,Withdrawal | 85.9 | 82.5 | 89.0 |
| 7 | Excitement,Posturing/catalepsy,Grimacing,Echopraxia/echolalia,Mannerisms,Rigidity,Waxy flexibility | 85.9 | 82.5 | 89.0 |
| 7 | Excitement,Posturing/catalepsy,Grimacing,Echopraxia/echolalia,Stereotypy,Mannerisms,Negativism | 85.9 | 82.5 | 89.0 |
| 7 | Grimacing,Echopraxia/echolalia,Stereotypy,Mannerisms,Rigidity,Negativism,Waxy flexibility | 85.9 | 82.5 | 89.0 |
| 7 | Excitement,Posturing/catalepsy,Echopraxia/echolalia,Stereotypy,Verbigeration,Negativism,Waxy flexibility | 85.7 | 82.3 | 88.8 |
| 7 | Excitement,Posturing/catalepsy,Grimacing,Echopraxia/echolalia,Stereotypy,Rigidity,Waxy flexibility | 85.7 | 82.3 | 89.0 |
| 7 | Excitement,Posturing/catalepsy,Grimacing,Stereotypy,Mannerisms,Negativism,Waxy flexibility | 85.7 | 82.3 | 88.8 |
| 7 | Excitement,Posturing/catalepsy,Grimacing,Stereotypy,Verbigeration,Rigidity,Waxy flexibility | 85.7 | 82.3 | 88.8 |
| 7 | Grimacing,Echopraxia/echolalia,Stereotypy,Mannerisms,Verbigeration,Waxy flexibility,Withdrawal | 85.7 | 82.3 | 88.8 |
| 7 | Grimacing,Stereotypy,Mannerisms,Verbigeration,Rigidity,Negativism,Waxy flexibility | 85.7 | 82.5 | 88.8 |
| 7 | Posturing/catalepsy,Grimacing,Echopraxia/echolalia,Mannerisms,Verbigeration,Negativism,Waxy flexibility | 85.7 | 82.3 | 88.8 |
| 7 | Posturing/catalepsy,Grimacing,Echopraxia/echolalia,Stereotypy,Mannerisms,Verbigeration,Negativism | 85.7 | 82.5 | 88.8 |
| 7 | Posturing/catalepsy,Grimacing,Echopraxia/echolalia,Stereotypy,Verbigeration,Negativism,Waxy flexibility | 85.7 | 82.3 | 88.8 |
| 7 | Excitement,Grimacing,Stereotypy,Verbigeration,Rigidity,Negativism,Waxy flexibility | 85.6 | 82.3 | 88.8 |
| 7 | Excitement,Posturing/catalepsy,Grimacing,Mannerisms,Verbigeration,Rigidity,Waxy flexibility | 85.6 | 82.3 | 88.8 |
| 7 | Excitement,Posturing/catalepsy,Grimacing,Echopraxia/echolalia,Mannerisms,Verbigeration,Rigidity | 85.5 | 82.1 | 88.6 |
| 7 | Excitement,Grimacing,Echopraxia/echolalia,Mannerisms,Verbigeration,Waxy flexibility,Withdrawal | 85.4 | 82.1 | 88.6 |
| 7 | Excitement,Mutism,Grimacing,Echopraxia/echolalia,Stereotypy,Mannerisms,Verbigeration | 85.4 | 82.1 | 88.6 |
| 7 | Excitement,Posturing/catalepsy,Stereotypy,Mannerisms,Verbigeration,Negativism,Waxy flexibility | 85.4 | 82.1 | 88.6 |
| 7 | Echopraxia/echolalia,Stereotypy,Mannerisms,Verbigeration,Rigidity,Negativism,Waxy flexibility | 85.2 | 81.8 | 88.6 |
| 7 | Excitement,Echopraxia/echolalia,Stereotypy,Mannerisms,Rigidity,Negativism,Waxy flexibility | 85.2 | 81.8 | 88.3 |
| 7 | Excitement,Echopraxia/echolalia,Stereotypy,Verbigeration,Rigidity,Negativism,Waxy flexibility | 85.2 | 81.8 | 88.3 |
| 7 | Excitement,Grimacing,Stereotypy,Mannerisms,Rigidity,Negativism,Waxy flexibility | 85.2 | 81.8 | 88.3 |
| 7 | Excitement,Posturing/catalepsy,Echopraxia/echolalia,Stereotypy,Mannerisms,Negativism,Waxy flexibility | 85.2 | 81.8 | 88.6 |
| 7 | Excitement,Posturing/catalepsy,Echopraxia/echolalia,Mannerisms,Verbigeration,Negativism,Waxy flexibility | 85.0 | 81.6 | 88.1 |
| 7 | Excitement,Posturing/catalepsy,Echopraxia/echolalia,Stereotypy,Mannerisms,Verbigeration,Negativism | 85.0 | 81.6 | 88.1 |
| 7 | Excitement,Posturing/catalepsy,Echopraxia/echolalia,Stereotypy,Verbigeration,Rigidity,Waxy flexibility | 85.0 | 81.6 | 88.3 |
| 7 | Excitement,Posturing/catalepsy,Grimacing,Echopraxia/echolalia,Stereotypy,Verbigeration,Rigidity | 85.0 | 81.6 | 88.3 |
| 7 | Excitement,Stereotypy,Mannerisms,Verbigeration,Rigidity,Negativism,Waxy flexibility | 85.0 | 81.6 | 88.1 |
| 7 | Posturing/catalepsy,Echopraxia/echolalia,Stereotypy,Mannerisms,Verbigeration,Negativism,Waxy flexibility | 85.0 | 81.6 | 88.1 |
| 7 | Grimacing,Echopraxia/echolalia,Stereotypy,Verbigeration,Rigidity,Negativism,Waxy flexibility | 84.9 | 81.6 | 88.1 |
| 7 | Excitement,Posturing/catalepsy,Echopraxia/echolalia,Mannerisms,Verbigeration,Rigidity,Waxy flexibility | 84.8 | 81.4 | 88.1 |
| 7 | Excitement,Grimacing,Echopraxia/echolalia,Verbigeration,Rigidity,Negativism,Waxy flexibility | 84.7 | 81.4 | 88.1 |
| 7 | Excitement,Grimacing,Stereotypy,Mannerisms,Verbigeration,Waxy flexibility,Withdrawal | 84.7 | 81.4 | 88.1 |
| 7 | Excitement,Grimacing,Echopraxia/echolalia,Mannerisms,Rigidity,Negativism,Waxy flexibility | 84.5 | 80.9 | 87.9 |
| 7 | Excitement,Posturing/catalepsy,Grimacing,Echopraxia/echolalia,Stereotypy,Mannerisms,Rigidity | 84.5 | 81.2 | 87.9 |
| 7 | Excitement,Posturing/catalepsy,Grimacing,Stereotypy,Mannerisms,Rigidity,Waxy flexibility | 84.5 | 81.2 | 87.9 |
| 7 | Excitement,Posturing/catalepsy,Grimacing,Stereotypy,Mannerisms,Verbigeration,Rigidity | 84.5 | 81.2 | 87.7 |
| 7 | Posturing/catalepsy,Grimacing,Echopraxia/echolalia,Mannerisms,Verbigeration,Rigidity,Waxy flexibility | 84.5 | 80.9 | 87.9 |
| 7 | Excitement,Grimacing,Echopraxia/echolalia,Stereotypy,Mannerisms,Rigidity,Negativism | 84.3 | 80.9 | 87.7 |
| 7 | Excitement,Grimacing,Echopraxia/echolalia,Stereotypy,Verbigeration,Rigidity,Negativism | 84.3 | 80.7 | 87.7 |
| 7 | Posturing/catalepsy,Grimacing,Echopraxia/echolalia,Stereotypy,Verbigeration,Rigidity,Waxy flexibility | 84.3 | 80.9 | 87.7 |
| 7 | Posturing/catalepsy,Grimacing,Stereotypy,Mannerisms,Verbigeration,Rigidity,Waxy flexibility | 84.3 | 80.9 | 87.7 |
| 7 | Excitement,Grimacing,Mannerisms,Verbigeration,Rigidity,Negativism,Waxy flexibility | 84.1 | 80.7 | 87.4 |
| 7 | Excitement,Grimacing,Stereotypy,Mannerisms,Verbigeration,Rigidity,Negativism | 84.1 | 80.7 | 87.4 |
| 7 | Excitement,Posturing/catalepsy,Echopraxia/echolalia,Stereotypy,Mannerisms,Rigidity,Waxy flexibility | 84.1 | 80.7 | 87.4 |
| 7 | Excitement,Posturing/catalepsy,Stereotypy,Mannerisms,Verbigeration,Rigidity,Waxy flexibility | 84.1 | 80.5 | 87.4 |
| 7 | Grimacing,Echopraxia/echolalia,Mannerisms,Verbigeration,Rigidity,Negativism,Waxy flexibility | 84.1 | 80.7 | 87.4 |
| 7 | Grimacing,Echopraxia/echolalia,Stereotypy,Mannerisms,Verbigeration,Rigidity,Negativism | 84.1 | 80.7 | 87.4 |
| 7 | Posturing/catalepsy,Grimacing,Echopraxia/echolalia,Stereotypy,Mannerisms,Rigidity,Waxy flexibility | 84.1 | 80.7 | 87.4 |
| 7 | Excitement,Posturing/catalepsy,Echopraxia/echolalia,Stereotypy,Mannerisms,Verbigeration,Rigidity | 83.8 | 80.3 | 87.2 |
| 7 | Posturing/catalepsy,Grimacing,Echopraxia/echolalia,Stereotypy,Mannerisms,Verbigeration,Rigidity | 83.7 | 80.3 | 87.0 |
| 7 | Excitement,Echopraxia/echolalia,Stereotypy,Mannerisms,Verbigeration,Rigidity,Negativism | 83.6 | 80.0 | 87.0 |
| 7 | Excitement,Grimacing,Echopraxia/echolalia,Stereotypy,Mannerisms,Verbigeration,Withdrawal | 83.4 | 79.8 | 86.8 |
| 7 | Excitement,Grimacing,Echopraxia/echolalia,Mannerisms,Verbigeration,Rigidity,Negativism | 83.0 | 79.4 | 86.3 |
| 7 | Excitement,Echopraxia/echolalia,Mannerisms,Verbigeration,Rigidity,Negativism,Waxy flexibility | 82.7 | 79.1 | 86.1 |
| 7 | Posturing/catalepsy,Echopraxia/echolalia,Stereotypy,Mannerisms,Verbigeration,Rigidity,Waxy flexibility | 82.7 | 79.1 | 86.1 |
| 7 | Excitement,Posturing/catalepsy,Grimacing,Echopraxia/echolalia,Stereotypy,Verbigeration,Waxy flexibility | 81.2 | 77.6 | 84.8 |
| 7 | Excitement,Posturing/catalepsy,Grimacing,Echopraxia/echolalia,Mannerisms,Verbigeration,Waxy flexibility | 81.1 | 77.4 | 84.8 |
| 7 | Excitement,Grimacing,Echopraxia/echolalia,Stereotypy,Verbigeration,Rigidity,Waxy flexibility | 80.9 | 77.1 | 84.5 |
| 7 | Excitement,Grimacing,Echopraxia/echolalia,Stereotypy,Mannerisms,Rigidity,Waxy flexibility | 80.7 | 76.9 | 84.3 |
| 7 | Excitement,Posturing/catalepsy,Grimacing,Echopraxia/echolalia,Stereotypy,Mannerisms,Waxy flexibility | 80.7 | 76.9 | 84.3 |
| 7 | Excitement,Posturing/catalepsy,Grimacing,Stereotypy,Mannerisms,Verbigeration,Waxy flexibility | 80.7 | 76.9 | 84.3 |
| 7 | Excitement,Grimacing,Echopraxia/echolalia,Stereotypy,Mannerisms,Negativism,Waxy flexibility | 80.1 | 76.2 | 83.6 |
| 7 | Excitement,Grimacing,Stereotypy,Mannerisms,Verbigeration,Rigidity,Waxy flexibility | 80.1 | 76.5 | 83.6 |
| 7 | Grimacing,Echopraxia/echolalia,Stereotypy,Mannerisms,Verbigeration,Negativism,Waxy flexibility | 80.1 | 76.2 | 83.6 |
| 7 | Excitement,Grimacing,Echopraxia/echolalia,Mannerisms,Verbigeration,Rigidity,Waxy flexibility | 79.8 | 76.0 | 83.4 |
| 7 | Excitement,Grimacing,Echopraxia/echolalia,Stereotypy,Verbigeration,Negativism,Waxy flexibility | 79.8 | 76.0 | 83.4 |
| 7 | Excitement,Posturing/catalepsy,Grimacing,Echopraxia/echolalia,Stereotypy,Mannerisms,Verbigeration | 79.8 | 76.0 | 83.4 |
| 7 | Posturing/catalepsy,Grimacing,Echopraxia/echolalia,Stereotypy,Mannerisms,Verbigeration,Waxy flexibility | 79.8 | 76.0 | 83.4 |
| 7 | Grimacing,Echopraxia/echolalia,Stereotypy,Mannerisms,Verbigeration,Rigidity,Waxy flexibility | 79.6 | 75.8 | 83.2 |
| 7 | Excitement,Grimacing,Stereotypy,Mannerisms,Verbigeration,Negativism,Waxy flexibility | 79.4 | 75.6 | 83.0 |
| 7 | Excitement,Echopraxia/echolalia,Stereotypy,Mannerisms,Verbigeration,Negativism,Waxy flexibility | 78.5 | 74.7 | 82.1 |
| 7 | Excitement,Echopraxia/echolalia,Stereotypy,Mannerisms,Verbigeration,Rigidity,Waxy flexibility | 78.5 | 74.7 | 82.1 |
| 7 | Excitement,Posturing/catalepsy,Echopraxia/echolalia,Stereotypy,Mannerisms,Verbigeration,Waxy flexibility | 78.5 | 74.7 | 82.3 |
| 7 | Excitement,Grimacing,Echopraxia/echolalia,Mannerisms,Verbigeration,Negativism,Waxy flexibility | 78.0 | 74.2 | 81.8 |
| 7 | Excitement,Grimacing,Echopraxia/echolalia,Stereotypy,Mannerisms,Verbigeration,Rigidity | 77.8 | 73.8 | 81.6 |
| 7 | Excitement,Grimacing,Echopraxia/echolalia,Stereotypy,Mannerisms,Verbigeration,Negativism | 77.1 | 73.1 | 80.9 |
| 7 | Excitement,Grimacing,Echopraxia/echolalia,Stereotypy,Mannerisms,Verbigeration,Waxy flexibility | 72.0 | 67.7 | 76.2 |

Table S8: sensitivity (relative to the full BFCSI) of all eight question combinations, and the 95% confidence interval derived from bootstrapping, sorted by decreasing sensitivity.

|  |  |  | 95% CI | |
| --- | --- | --- | --- | --- |
| # of Qs | Combination | Sensitivity | Lower | Upper |
| 8 | Excitement,Immobility/stupor,Mutism,Staring,Posturing/catalepsy,Echopraxia/echolalia,Negativism,Withdrawal | 100.0 | 100.0 | 100.0 |
| 8 | Excitement,Immobility/stupor,Mutism,Staring,Posturing/catalepsy,Echopraxia/echolalia,Rigidity,Withdrawal | 100.0 | 100.0 | 100.0 |
| 8 | Excitement,Mutism,Staring,Echopraxia/echolalia,Stereotypy,Rigidity,Negativism,Withdrawal | 100.0 | 100.0 | 100.0 |
| 8 | Excitement,Mutism,Staring,Echopraxia/echolalia,Stereotypy,Rigidity,Waxy flexibility,Withdrawal | 100.0 | 100.0 | 100.0 |
| 8 | Excitement,Mutism,Staring,Posturing/catalepsy,Echopraxia/echolalia,Mannerisms,Negativism,Withdrawal | 100.0 | 100.0 | 100.0 |
| 8 | Excitement,Mutism,Staring,Posturing/catalepsy,Echopraxia/echolalia,Mannerisms,Rigidity,Withdrawal | 100.0 | 100.0 | 100.0 |
| 8 | Excitement,Mutism,Staring,Posturing/catalepsy,Echopraxia/echolalia,Negativism,Waxy flexibility,Withdrawal | 100.0 | 100.0 | 100.0 |
| 8 | Excitement,Mutism,Staring,Posturing/catalepsy,Echopraxia/echolalia,Rigidity,Negativism,Withdrawal | 100.0 | 100.0 | 100.0 |
| 8 | Excitement,Mutism,Staring,Posturing/catalepsy,Echopraxia/echolalia,Rigidity,Waxy flexibility,Withdrawal | 100.0 | 100.0 | 100.0 |
| 8 | Excitement,Mutism,Staring,Posturing/catalepsy,Echopraxia/echolalia,Stereotypy,Negativism,Withdrawal | 100.0 | 100.0 | 100.0 |
| 8 | Excitement,Mutism,Staring,Posturing/catalepsy,Echopraxia/echolalia,Stereotypy,Rigidity,Withdrawal | 100.0 | 100.0 | 100.0 |
| 8 | Excitement,Mutism,Staring,Posturing/catalepsy,Echopraxia/echolalia,Verbigeration,Negativism,Waxy flexibility | 100.0 | 100.0 | 100.0 |
| 8 | Excitement,Mutism,Staring,Posturing/catalepsy,Echopraxia/echolalia,Verbigeration,Negativism,Withdrawal | 100.0 | 100.0 | 100.0 |
| 8 | Excitement,Mutism,Staring,Posturing/catalepsy,Echopraxia/echolalia,Verbigeration,Rigidity,Negativism | 100.0 | 100.0 | 100.0 |
| 8 | Excitement,Mutism,Staring,Posturing/catalepsy,Echopraxia/echolalia,Verbigeration,Rigidity,Withdrawal | 100.0 | 100.0 | 100.0 |
| 8 | Excitement,Mutism,Staring,Posturing/catalepsy,Grimacing,Echopraxia/echolalia,Negativism,Withdrawal | 100.0 | 100.0 | 100.0 |
| 8 | Excitement,Mutism,Staring,Posturing/catalepsy,Grimacing,Echopraxia/echolalia,Rigidity,Withdrawal | 100.0 | 100.0 | 100.0 |
| 8 | Excitement,Mutism,Staring,Posturing/catalepsy,Mannerisms,Verbigeration,Negativism,Withdrawal | 100.0 | 100.0 | 100.0 |
| 8 | Excitement,Mutism,Staring,Posturing/catalepsy,Mannerisms,Verbigeration,Rigidity,Withdrawal | 100.0 | 100.0 | 100.0 |
| 8 | Excitement,Mutism,Staring,Posturing/catalepsy,Stereotypy,Verbigeration,Negativism,Withdrawal | 100.0 | 100.0 | 100.0 |
| 8 | Excitement,Mutism,Staring,Stereotypy,Verbigeration,Rigidity,Negativism,Withdrawal | 100.0 | 100.0 | 100.0 |
| 8 | Excitement,Mutism,Staring,Stereotypy,Verbigeration,Rigidity,Waxy flexibility,Withdrawal | 100.0 | 100.0 | 100.0 |
| 8 | Excitement,Immobility/stupor,Mutism,Staring,Echopraxia/echolalia,Rigidity,Negativism,Withdrawal | 99.8 | 99.3 | 100.0 |
| 8 | Excitement,Immobility/stupor,Mutism,Staring,Echopraxia/echolalia,Rigidity,Waxy flexibility,Withdrawal | 99.8 | 99.3 | 100.0 |
| 8 | Excitement,Immobility/stupor,Mutism,Staring,Echopraxia/echolalia,Stereotypy,Rigidity,Withdrawal | 99.8 | 99.3 | 100.0 |
| 8 | Excitement,Immobility/stupor,Mutism,Staring,Posturing/catalepsy,Echopraxia/echolalia,Negativism,Waxy flexibility | 99.8 | 99.3 | 100.0 |
| 8 | Excitement,Immobility/stupor,Mutism,Staring,Posturing/catalepsy,Echopraxia/echolalia,Rigidity,Negativism | 99.8 | 99.3 | 100.0 |
| 8 | Excitement,Immobility/stupor,Mutism,Staring,Posturing/catalepsy,Echopraxia/echolalia,Verbigeration,Negativism | 99.8 | 99.3 | 100.0 |
| 8 | Excitement,Immobility/stupor,Mutism,Staring,Posturing/catalepsy,Echopraxia/echolalia,Verbigeration,Rigidity | 99.8 | 99.3 | 100.0 |
| 8 | Excitement,Immobility/stupor,Mutism,Staring,Posturing/catalepsy,Mannerisms,Negativism,Withdrawal | 99.8 | 99.3 | 100.0 |
| 8 | Excitement,Immobility/stupor,Mutism,Staring,Posturing/catalepsy,Mannerisms,Rigidity,Withdrawal | 99.8 | 99.3 | 100.0 |
| 8 | Excitement,Immobility/stupor,Mutism,Staring,Posturing/catalepsy,Mannerisms,Verbigeration,Negativism | 99.8 | 99.3 | 100.0 |
| 8 | Excitement,Immobility/stupor,Mutism,Staring,Posturing/catalepsy,Mannerisms,Verbigeration,Rigidity | 99.8 | 99.3 | 100.0 |
| 8 | Excitement,Immobility/stupor,Mutism,Staring,Posturing/catalepsy,Stereotypy,Negativism,Withdrawal | 99.8 | 99.3 | 100.0 |
| 8 | Excitement,Immobility/stupor,Mutism,Staring,Posturing/catalepsy,Stereotypy,Verbigeration,Negativism | 99.8 | 99.3 | 100.0 |
| 8 | Excitement,Immobility/stupor,Mutism,Staring,Posturing/catalepsy,Verbigeration,Negativism,Waxy flexibility | 99.8 | 99.3 | 100.0 |
| 8 | Excitement,Immobility/stupor,Mutism,Staring,Posturing/catalepsy,Verbigeration,Negativism,Withdrawal | 99.8 | 99.3 | 100.0 |
| 8 | Excitement,Immobility/stupor,Mutism,Staring,Posturing/catalepsy,Verbigeration,Rigidity,Negativism | 99.8 | 99.3 | 100.0 |
| 8 | Excitement,Immobility/stupor,Mutism,Staring,Stereotypy,Rigidity,Waxy flexibility,Withdrawal | 99.8 | 99.3 | 100.0 |
| 8 | Excitement,Immobility/stupor,Mutism,Staring,Stereotypy,Verbigeration,Rigidity,Negativism | 99.8 | 99.3 | 100.0 |
| 8 | Excitement,Immobility/stupor,Mutism,Staring,Stereotypy,Verbigeration,Rigidity,Waxy flexibility | 99.8 | 99.3 | 100.0 |
| 8 | Excitement,Immobility/stupor,Staring,Echopraxia/echolalia,Mannerisms,Rigidity,Negativism,Withdrawal | 99.8 | 99.3 | 100.0 |
| 8 | Excitement,Immobility/stupor,Staring,Echopraxia/echolalia,Stereotypy,Rigidity,Negativism,Withdrawal | 99.8 | 99.3 | 100.0 |
| 8 | Excitement,Immobility/stupor,Staring,Mannerisms,Verbigeration,Rigidity,Negativism,Withdrawal | 99.8 | 99.3 | 100.0 |
| 8 | Excitement,Immobility/stupor,Staring,Posturing/catalepsy,Echopraxia/echolalia,Mannerisms,Negativism,Withdrawal | 99.8 | 99.3 | 100.0 |
| 8 | Excitement,Immobility/stupor,Staring,Posturing/catalepsy,Echopraxia/echolalia,Mannerisms,Rigidity,Withdrawal | 99.8 | 99.3 | 100.0 |
| 8 | Excitement,Immobility/stupor,Staring,Posturing/catalepsy,Echopraxia/echolalia,Rigidity,Negativism,Withdrawal | 99.8 | 99.3 | 100.0 |
| 8 | Excitement,Immobility/stupor,Staring,Posturing/catalepsy,Mannerisms,Rigidity,Negativism,Withdrawal | 99.8 | 99.3 | 100.0 |
| 8 | Excitement,Immobility/stupor,Staring,Posturing/catalepsy,Mannerisms,Verbigeration,Negativism,Withdrawal | 99.8 | 99.3 | 100.0 |
| 8 | Excitement,Immobility/stupor,Staring,Posturing/catalepsy,Mannerisms,Verbigeration,Rigidity,Negativism | 99.8 | 99.3 | 100.0 |
| 8 | Excitement,Immobility/stupor,Staring,Posturing/catalepsy,Mannerisms,Verbigeration,Rigidity,Withdrawal | 99.8 | 99.3 | 100.0 |
| 8 | Excitement,Immobility/stupor,Staring,Stereotypy,Verbigeration,Rigidity,Negativism,Withdrawal | 99.8 | 99.3 | 100.0 |
| 8 | Excitement,Mutism,Staring,Echopraxia/echolalia,Mannerisms,Rigidity,Negativism,Withdrawal | 99.8 | 99.3 | 100.0 |
| 8 | Excitement,Mutism,Staring,Echopraxia/echolalia,Rigidity,Negativism,Waxy flexibility,Withdrawal | 99.8 | 99.3 | 100.0 |
| 8 | Excitement,Mutism,Staring,Echopraxia/echolalia,Stereotypy,Mannerisms,Rigidity,Withdrawal | 99.8 | 99.3 | 100.0 |
| 8 | Excitement,Mutism,Staring,Echopraxia/echolalia,Stereotypy,Rigidity,Negativism,Waxy flexibility | 99.8 | 99.3 | 100.0 |
| 8 | Excitement,Mutism,Staring,Echopraxia/echolalia,Stereotypy,Verbigeration,Rigidity,Negativism | 99.8 | 99.3 | 100.0 |
| 8 | Excitement,Mutism,Staring,Echopraxia/echolalia,Stereotypy,Verbigeration,Rigidity,Withdrawal | 99.8 | 99.3 | 100.0 |
| 8 | Excitement,Mutism,Staring,Echopraxia/echolalia,Verbigeration,Rigidity,Negativism,Waxy flexibility | 99.8 | 99.3 | 100.0 |
| 8 | Excitement,Mutism,Staring,Echopraxia/echolalia,Verbigeration,Rigidity,Negativism,Withdrawal | 99.8 | 99.3 | 100.0 |
| 8 | Excitement,Mutism,Staring,Grimacing,Echopraxia/echolalia,Rigidity,Negativism,Withdrawal | 99.8 | 99.3 | 100.0 |
| 8 | Excitement,Mutism,Staring,Grimacing,Echopraxia/echolalia,Rigidity,Waxy flexibility,Withdrawal | 99.8 | 99.3 | 100.0 |
| 8 | Excitement,Mutism,Staring,Grimacing,Echopraxia/echolalia,Stereotypy,Rigidity,Withdrawal | 99.8 | 99.3 | 100.0 |
| 8 | Excitement,Mutism,Staring,Grimacing,Stereotypy,Rigidity,Waxy flexibility,Withdrawal | 99.8 | 99.3 | 100.0 |
| 8 | Excitement,Mutism,Staring,Grimacing,Stereotypy,Verbigeration,Rigidity,Withdrawal | 99.8 | 99.3 | 100.0 |
| 8 | Excitement,Mutism,Staring,Mannerisms,Verbigeration,Rigidity,Negativism,Withdrawal | 99.8 | 99.3 | 100.0 |
| 8 | Excitement,Mutism,Staring,Posturing/catalepsy,Echopraxia/echolalia,Mannerisms,Negativism,Waxy flexibility | 99.8 | 99.3 | 100.0 |
| 8 | Excitement,Mutism,Staring,Posturing/catalepsy,Echopraxia/echolalia,Mannerisms,Rigidity,Negativism | 99.8 | 99.3 | 100.0 |
| 8 | Excitement,Mutism,Staring,Posturing/catalepsy,Echopraxia/echolalia,Mannerisms,Verbigeration,Negativism | 99.8 | 99.3 | 100.0 |
| 8 | Excitement,Mutism,Staring,Posturing/catalepsy,Echopraxia/echolalia,Mannerisms,Verbigeration,Rigidity | 99.8 | 99.3 | 100.0 |
| 8 | Excitement,Mutism,Staring,Posturing/catalepsy,Echopraxia/echolalia,Rigidity,Negativism,Waxy flexibility | 99.8 | 99.3 | 100.0 |
| 8 | Excitement,Mutism,Staring,Posturing/catalepsy,Echopraxia/echolalia,Stereotypy,Negativism,Waxy flexibility | 99.8 | 99.3 | 100.0 |
| 8 | Excitement,Mutism,Staring,Posturing/catalepsy,Echopraxia/echolalia,Stereotypy,Rigidity,Negativism | 99.8 | 99.3 | 100.0 |
| 8 | Excitement,Mutism,Staring,Posturing/catalepsy,Echopraxia/echolalia,Stereotypy,Verbigeration,Negativism | 99.8 | 99.3 | 100.0 |
| 8 | Excitement,Mutism,Staring,Posturing/catalepsy,Echopraxia/echolalia,Stereotypy,Verbigeration,Rigidity | 99.8 | 99.3 | 100.0 |
| 8 | Excitement,Mutism,Staring,Posturing/catalepsy,Echopraxia/echolalia,Verbigeration,Rigidity,Waxy flexibility | 99.8 | 99.3 | 100.0 |
| 8 | Excitement,Mutism,Staring,Posturing/catalepsy,Grimacing,Echopraxia/echolalia,Negativism,Waxy flexibility | 99.8 | 99.3 | 100.0 |
| 8 | Excitement,Mutism,Staring,Posturing/catalepsy,Grimacing,Echopraxia/echolalia,Rigidity,Negativism | 99.8 | 99.3 | 100.0 |
| 8 | Excitement,Mutism,Staring,Posturing/catalepsy,Grimacing,Echopraxia/echolalia,Verbigeration,Negativism | 99.8 | 99.3 | 100.0 |
| 8 | Excitement,Mutism,Staring,Posturing/catalepsy,Grimacing,Echopraxia/echolalia,Verbigeration,Rigidity | 99.8 | 99.3 | 100.0 |
| 8 | Excitement,Mutism,Staring,Posturing/catalepsy,Grimacing,Mannerisms,Negativism,Withdrawal | 99.8 | 99.3 | 100.0 |
| 8 | Excitement,Mutism,Staring,Posturing/catalepsy,Grimacing,Mannerisms,Rigidity,Withdrawal | 99.8 | 99.3 | 100.0 |
| 8 | Excitement,Mutism,Staring,Posturing/catalepsy,Grimacing,Stereotypy,Negativism,Withdrawal | 99.8 | 99.3 | 100.0 |
| 8 | Excitement,Mutism,Staring,Posturing/catalepsy,Grimacing,Stereotypy,Rigidity,Withdrawal | 99.8 | 99.3 | 100.0 |
| 8 | Excitement,Mutism,Staring,Posturing/catalepsy,Grimacing,Verbigeration,Negativism,Withdrawal | 99.8 | 99.3 | 100.0 |
| 8 | Excitement,Mutism,Staring,Posturing/catalepsy,Grimacing,Verbigeration,Rigidity,Withdrawal | 99.8 | 99.3 | 100.0 |
| 8 | Excitement,Mutism,Staring,Posturing/catalepsy,Mannerisms,Negativism,Waxy flexibility,Withdrawal | 99.8 | 99.3 | 100.0 |
| 8 | Excitement,Mutism,Staring,Posturing/catalepsy,Mannerisms,Rigidity,Negativism,Withdrawal | 99.8 | 99.3 | 100.0 |
| 8 | Excitement,Mutism,Staring,Posturing/catalepsy,Mannerisms,Rigidity,Waxy flexibility,Withdrawal | 99.8 | 99.3 | 100.0 |
| 8 | Excitement,Mutism,Staring,Posturing/catalepsy,Mannerisms,Verbigeration,Negativism,Waxy flexibility | 99.8 | 99.3 | 100.0 |
| 8 | Excitement,Mutism,Staring,Posturing/catalepsy,Mannerisms,Verbigeration,Rigidity,Negativism | 99.8 | 99.3 | 100.0 |
| 8 | Excitement,Mutism,Staring,Posturing/catalepsy,Stereotypy,Mannerisms,Negativism,Withdrawal | 99.8 | 99.3 | 100.0 |
| 8 | Excitement,Mutism,Staring,Posturing/catalepsy,Stereotypy,Mannerisms,Rigidity,Withdrawal | 99.8 | 99.3 | 100.0 |
| 8 | Excitement,Mutism,Staring,Posturing/catalepsy,Stereotypy,Negativism,Waxy flexibility,Withdrawal | 99.8 | 99.3 | 100.0 |
| 8 | Excitement,Mutism,Staring,Posturing/catalepsy,Stereotypy,Rigidity,Negativism,Withdrawal | 99.8 | 99.3 | 100.0 |
| 8 | Excitement,Mutism,Staring,Posturing/catalepsy,Stereotypy,Rigidity,Waxy flexibility,Withdrawal | 99.8 | 99.3 | 100.0 |
| 8 | Excitement,Mutism,Staring,Posturing/catalepsy,Stereotypy,Verbigeration,Negativism,Waxy flexibility | 99.8 | 99.3 | 100.0 |
| 8 | Excitement,Mutism,Staring,Posturing/catalepsy,Stereotypy,Verbigeration,Rigidity,Negativism | 99.8 | 99.3 | 100.0 |
| 8 | Excitement,Mutism,Staring,Posturing/catalepsy,Stereotypy,Verbigeration,Rigidity,Withdrawal | 99.8 | 99.3 | 100.0 |
| 8 | Excitement,Mutism,Staring,Posturing/catalepsy,Verbigeration,Negativism,Waxy flexibility,Withdrawal | 99.8 | 99.3 | 100.0 |
| 8 | Excitement,Mutism,Staring,Posturing/catalepsy,Verbigeration,Rigidity,Negativism,Withdrawal | 99.8 | 99.3 | 100.0 |
| 8 | Excitement,Mutism,Staring,Posturing/catalepsy,Verbigeration,Rigidity,Waxy flexibility,Withdrawal | 99.8 | 99.3 | 100.0 |
| 8 | Excitement,Mutism,Staring,Stereotypy,Mannerisms,Rigidity,Waxy flexibility,Withdrawal | 99.8 | 99.3 | 100.0 |
| 8 | Excitement,Mutism,Staring,Stereotypy,Mannerisms,Verbigeration,Rigidity,Withdrawal | 99.8 | 99.3 | 100.0 |
| 8 | Excitement,Mutism,Staring,Stereotypy,Rigidity,Negativism,Waxy flexibility,Withdrawal | 99.8 | 99.3 | 100.0 |
| 8 | Excitement,Mutism,Staring,Stereotypy,Verbigeration,Rigidity,Negativism,Waxy flexibility | 99.8 | 99.3 | 100.0 |
| 8 | Immobility/stupor,Staring,Echopraxia/echolalia,Stereotypy,Mannerisms,Rigidity,Negativism,Withdrawal | 99.8 | 99.3 | 100.0 |
| 8 | Immobility/stupor,Staring,Stereotypy,Mannerisms,Verbigeration,Rigidity,Negativism,Withdrawal | 99.8 | 99.3 | 100.0 |
| 8 | Mutism,Staring,Echopraxia/echolalia,Stereotypy,Mannerisms,Rigidity,Negativism,Withdrawal | 99.8 | 99.3 | 100.0 |
| 8 | Mutism,Staring,Posturing/catalepsy,Grimacing,Echopraxia/echolalia,Mannerisms,Negativism,Withdrawal | 99.8 | 99.3 | 100.0 |
| 8 | Mutism,Staring,Posturing/catalepsy,Grimacing,Stereotypy,Mannerisms,Negativism,Withdrawal | 99.8 | 99.3 | 100.0 |
| 8 | Mutism,Staring,Stereotypy,Mannerisms,Verbigeration,Rigidity,Negativism,Withdrawal | 99.8 | 99.3 | 100.0 |
| 8 | Excitement,Immobility/stupor,Mutism,Posturing/catalepsy,Echopraxia/echolalia,Verbigeration,Rigidity,Negativism | 99.6 | 98.9 | 100.0 |
| 8 | Excitement,Immobility/stupor,Mutism,Posturing/catalepsy,Stereotypy,Verbigeration,Rigidity,Negativism | 99.6 | 98.9 | 100.0 |
| 8 | Excitement,Immobility/stupor,Mutism,Staring,Echopraxia/echolalia,Mannerisms,Rigidity,Withdrawal | 99.6 | 98.9 | 100.0 |
| 8 | Excitement,Immobility/stupor,Mutism,Staring,Echopraxia/echolalia,Rigidity,Negativism,Waxy flexibility | 99.6 | 98.9 | 100.0 |
| 8 | Excitement,Immobility/stupor,Mutism,Staring,Echopraxia/echolalia,Stereotypy,Negativism,Withdrawal | 99.6 | 98.9 | 100.0 |
| 8 | Excitement,Immobility/stupor,Mutism,Staring,Echopraxia/echolalia,Stereotypy,Rigidity,Negativism | 99.6 | 98.9 | 100.0 |
| 8 | Excitement,Immobility/stupor,Mutism,Staring,Echopraxia/echolalia,Stereotypy,Rigidity,Waxy flexibility | 99.6 | 98.9 | 100.0 |
| 8 | Excitement,Immobility/stupor,Mutism,Staring,Echopraxia/echolalia,Verbigeration,Rigidity,Negativism | 99.6 | 98.9 | 100.0 |
| 8 | Excitement,Immobility/stupor,Mutism,Staring,Echopraxia/echolalia,Verbigeration,Rigidity,Waxy flexibility | 99.6 | 98.9 | 100.0 |
| 8 | Excitement,Immobility/stupor,Mutism,Staring,Echopraxia/echolalia,Verbigeration,Rigidity,Withdrawal | 99.6 | 98.9 | 100.0 |
| 8 | Excitement,Immobility/stupor,Mutism,Staring,Grimacing,Echopraxia/echolalia,Rigidity,Withdrawal | 99.6 | 98.9 | 100.0 |
| 8 | Excitement,Immobility/stupor,Mutism,Staring,Mannerisms,Rigidity,Waxy flexibility,Withdrawal | 99.6 | 98.9 | 100.0 |
| 8 | Excitement,Immobility/stupor,Mutism,Staring,Mannerisms,Verbigeration,Rigidity,Negativism | 99.6 | 98.9 | 100.0 |
| 8 | Excitement,Immobility/stupor,Mutism,Staring,Mannerisms,Verbigeration,Rigidity,Waxy flexibility | 99.6 | 98.9 | 100.0 |
| 8 | Excitement,Immobility/stupor,Mutism,Staring,Mannerisms,Verbigeration,Rigidity,Withdrawal | 99.6 | 98.9 | 100.0 |
| 8 | Excitement,Immobility/stupor,Mutism,Staring,Posturing/catalepsy,Echopraxia/echolalia,Mannerisms,Rigidity | 99.6 | 98.9 | 100.0 |
| 8 | Excitement,Immobility/stupor,Mutism,Staring,Posturing/catalepsy,Echopraxia/echolalia,Stereotypy,Negativism | 99.6 | 98.9 | 100.0 |
| 8 | Excitement,Immobility/stupor,Mutism,Staring,Posturing/catalepsy,Grimacing,Echopraxia/echolalia,Negativism | 99.6 | 98.9 | 100.0 |
| 8 | Excitement,Immobility/stupor,Mutism,Staring,Posturing/catalepsy,Grimacing,Echopraxia/echolalia,Rigidity | 99.6 | 98.9 | 100.0 |
| 8 | Excitement,Immobility/stupor,Mutism,Staring,Posturing/catalepsy,Grimacing,Negativism,Withdrawal | 99.6 | 98.9 | 100.0 |
| 8 | Excitement,Immobility/stupor,Mutism,Staring,Posturing/catalepsy,Grimacing,Rigidity,Withdrawal | 99.6 | 98.9 | 100.0 |
| 8 | Excitement,Immobility/stupor,Mutism,Staring,Posturing/catalepsy,Grimacing,Verbigeration,Rigidity | 99.6 | 98.9 | 100.0 |
| 8 | Excitement,Immobility/stupor,Mutism,Staring,Posturing/catalepsy,Mannerisms,Negativism,Waxy flexibility | 99.6 | 98.9 | 100.0 |
| 8 | Excitement,Immobility/stupor,Mutism,Staring,Posturing/catalepsy,Mannerisms,Rigidity,Negativism | 99.6 | 98.9 | 100.0 |
| 8 | Excitement,Immobility/stupor,Mutism,Staring,Posturing/catalepsy,Negativism,Waxy flexibility,Withdrawal | 99.6 | 98.9 | 100.0 |
| 8 | Excitement,Immobility/stupor,Mutism,Staring,Posturing/catalepsy,Rigidity,Negativism,Withdrawal | 99.6 | 98.9 | 100.0 |
| 8 | Excitement,Immobility/stupor,Mutism,Staring,Posturing/catalepsy,Rigidity,Waxy flexibility,Withdrawal | 99.6 | 98.9 | 100.0 |
| 8 | Excitement,Immobility/stupor,Mutism,Staring,Posturing/catalepsy,Stereotypy,Negativism,Waxy flexibility | 99.6 | 98.9 | 100.0 |
| 8 | Excitement,Immobility/stupor,Mutism,Staring,Posturing/catalepsy,Stereotypy,Rigidity,Withdrawal | 99.6 | 98.9 | 100.0 |
| 8 | Excitement,Immobility/stupor,Mutism,Staring,Posturing/catalepsy,Verbigeration,Rigidity,Waxy flexibility | 99.6 | 98.9 | 100.0 |
| 8 | Excitement,Immobility/stupor,Mutism,Staring,Posturing/catalepsy,Verbigeration,Rigidity,Withdrawal | 99.6 | 98.9 | 100.0 |
| 8 | Excitement,Immobility/stupor,Mutism,Staring,Stereotypy,Rigidity,Negativism,Waxy flexibility | 99.6 | 98.9 | 100.0 |
| 8 | Excitement,Immobility/stupor,Mutism,Staring,Stereotypy,Rigidity,Negativism,Withdrawal | 99.6 | 98.9 | 100.0 |
| 8 | Excitement,Immobility/stupor,Mutism,Staring,Stereotypy,Verbigeration,Rigidity,Withdrawal | 99.6 | 98.9 | 100.0 |
| 8 | Excitement,Immobility/stupor,Mutism,Staring,Verbigeration,Rigidity,Negativism,Waxy flexibility | 99.6 | 98.9 | 100.0 |
| 8 | Excitement,Immobility/stupor,Mutism,Staring,Verbigeration,Rigidity,Negativism,Withdrawal | 99.6 | 98.9 | 100.0 |
| 8 | Excitement,Immobility/stupor,Mutism,Stereotypy,Verbigeration,Rigidity,Negativism,Withdrawal | 99.6 | 98.9 | 100.0 |
| 8 | Excitement,Immobility/stupor,Staring,Echopraxia/echolalia,Mannerisms,Rigidity,Waxy flexibility,Withdrawal | 99.6 | 98.9 | 100.0 |
| 8 | Excitement,Immobility/stupor,Staring,Echopraxia/echolalia,Rigidity,Negativism,Waxy flexibility,Withdrawal | 99.6 | 98.9 | 100.0 |
| 8 | Excitement,Immobility/stupor,Staring,Echopraxia/echolalia,Stereotypy,Rigidity,Waxy flexibility,Withdrawal | 99.6 | 98.9 | 100.0 |
| 8 | Excitement,Immobility/stupor,Staring,Echopraxia/echolalia,Verbigeration,Rigidity,Negativism,Withdrawal | 99.6 | 98.9 | 100.0 |
| 8 | Excitement,Immobility/stupor,Staring,Grimacing,Echopraxia/echolalia,Rigidity,Negativism,Withdrawal | 99.6 | 98.9 | 100.0 |
| 8 | Excitement,Immobility/stupor,Staring,Mannerisms,Rigidity,Negativism,Waxy flexibility,Withdrawal | 99.6 | 98.9 | 100.0 |
| 8 | Excitement,Immobility/stupor,Staring,Mannerisms,Verbigeration,Rigidity,Negativism,Waxy flexibility | 99.6 | 98.9 | 100.0 |
| 8 | Excitement,Immobility/stupor,Staring,Mannerisms,Verbigeration,Rigidity,Waxy flexibility,Withdrawal | 99.6 | 98.9 | 100.0 |
| 8 | Excitement,Immobility/stupor,Staring,Posturing/catalepsy,Echopraxia/echolalia,Mannerisms,Rigidity,Negativism | 99.6 | 98.9 | 100.0 |
| 8 | Excitement,Immobility/stupor,Staring,Posturing/catalepsy,Echopraxia/echolalia,Rigidity,Waxy flexibility,Withdrawal | 99.6 | 98.9 | 100.0 |
| 8 | Excitement,Immobility/stupor,Staring,Posturing/catalepsy,Echopraxia/echolalia,Stereotypy,Negativism,Withdrawal | 99.6 | 98.9 | 100.0 |
| 8 | Excitement,Immobility/stupor,Staring,Posturing/catalepsy,Echopraxia/echolalia,Stereotypy,Rigidity,Withdrawal | 99.6 | 98.9 | 100.0 |
| 8 | Excitement,Immobility/stupor,Staring,Posturing/catalepsy,Echopraxia/echolalia,Verbigeration,Rigidity,Negativism | 99.6 | 98.9 | 100.0 |
| 8 | Excitement,Immobility/stupor,Staring,Posturing/catalepsy,Grimacing,Echopraxia/echolalia,Negativism,Withdrawal | 99.6 | 98.9 | 100.0 |
| 8 | Excitement,Immobility/stupor,Staring,Posturing/catalepsy,Grimacing,Echopraxia/echolalia,Rigidity,Withdrawal | 99.6 | 98.9 | 100.0 |
| 8 | Excitement,Immobility/stupor,Staring,Posturing/catalepsy,Mannerisms,Rigidity,Waxy flexibility,Withdrawal | 99.6 | 98.9 | 100.0 |
| 8 | Excitement,Immobility/stupor,Staring,Posturing/catalepsy,Stereotypy,Mannerisms,Rigidity,Withdrawal | 99.6 | 98.9 | 100.0 |
| 8 | Excitement,Immobility/stupor,Staring,Posturing/catalepsy,Stereotypy,Rigidity,Negativism,Withdrawal | 99.6 | 98.9 | 100.0 |
| 8 | Excitement,Immobility/stupor,Staring,Posturing/catalepsy,Stereotypy,Verbigeration,Negativism,Withdrawal | 99.6 | 98.9 | 100.0 |
| 8 | Excitement,Immobility/stupor,Staring,Posturing/catalepsy,Stereotypy,Verbigeration,Rigidity,Negativism | 99.6 | 98.9 | 100.0 |
| 8 | Excitement,Immobility/stupor,Staring,Posturing/catalepsy,Verbigeration,Rigidity,Negativism,Withdrawal | 99.6 | 98.9 | 100.0 |
| 8 | Excitement,Immobility/stupor,Staring,Stereotypy,Mannerisms,Rigidity,Negativism,Withdrawal | 99.6 | 98.9 | 100.0 |
| 8 | Excitement,Immobility/stupor,Staring,Stereotypy,Mannerisms,Verbigeration,Rigidity,Negativism | 99.6 | 98.9 | 100.0 |
| 8 | Excitement,Immobility/stupor,Staring,Stereotypy,Mannerisms,Verbigeration,Rigidity,Withdrawal | 99.6 | 98.9 | 100.0 |
| 8 | Excitement,Immobility/stupor,Staring,Stereotypy,Rigidity,Negativism,Waxy flexibility,Withdrawal | 99.6 | 98.9 | 100.0 |
| 8 | Excitement,Immobility/stupor,Staring,Stereotypy,Verbigeration,Rigidity,Negativism,Waxy flexibility | 99.6 | 98.9 | 100.0 |
| 8 | Excitement,Immobility/stupor,Staring,Stereotypy,Verbigeration,Rigidity,Waxy flexibility,Withdrawal | 99.6 | 98.9 | 100.0 |
| 8 | Excitement,Mutism,Staring,Echopraxia/echolalia,Mannerisms,Rigidity,Waxy flexibility,Withdrawal | 99.6 | 98.9 | 100.0 |
| 8 | Excitement,Mutism,Staring,Echopraxia/echolalia,Mannerisms,Verbigeration,Rigidity,Negativism | 99.6 | 98.9 | 100.0 |
| 8 | Excitement,Mutism,Staring,Echopraxia/echolalia,Stereotypy,Mannerisms,Negativism,Withdrawal | 99.6 | 98.9 | 100.0 |
| 8 | Excitement,Mutism,Staring,Echopraxia/echolalia,Stereotypy,Mannerisms,Rigidity,Negativism | 99.6 | 98.9 | 100.0 |
| 8 | Excitement,Mutism,Staring,Echopraxia/echolalia,Stereotypy,Negativism,Waxy flexibility,Withdrawal | 99.6 | 98.9 | 100.0 |
| 8 | Excitement,Mutism,Staring,Echopraxia/echolalia,Stereotypy,Verbigeration,Negativism,Waxy flexibility | 99.6 | 98.9 | 100.0 |
| 8 | Excitement,Mutism,Staring,Grimacing,Echopraxia/echolalia,Mannerisms,Rigidity,Withdrawal | 99.6 | 98.9 | 100.0 |
| 8 | Excitement,Mutism,Staring,Grimacing,Echopraxia/echolalia,Rigidity,Negativism,Waxy flexibility | 99.6 | 98.9 | 100.0 |
| 8 | Excitement,Mutism,Staring,Grimacing,Echopraxia/echolalia,Stereotypy,Rigidity,Negativism | 99.6 | 98.9 | 100.0 |
| 8 | Excitement,Mutism,Staring,Grimacing,Echopraxia/echolalia,Stereotypy,Rigidity,Waxy flexibility | 99.6 | 98.9 | 100.0 |
| 8 | Excitement,Mutism,Staring,Grimacing,Echopraxia/echolalia,Verbigeration,Rigidity,Withdrawal | 99.6 | 98.9 | 100.0 |
| 8 | Excitement,Mutism,Staring,Grimacing,Mannerisms,Verbigeration,Rigidity,Withdrawal | 99.6 | 98.9 | 100.0 |
| 8 | Excitement,Mutism,Staring,Grimacing,Stereotypy,Rigidity,Negativism,Withdrawal | 99.6 | 98.9 | 100.0 |
| 8 | Excitement,Mutism,Staring,Grimacing,Stereotypy,Verbigeration,Rigidity,Waxy flexibility | 99.6 | 98.9 | 100.0 |
| 8 | Excitement,Mutism,Staring,Grimacing,Verbigeration,Rigidity,Negativism,Withdrawal | 99.6 | 98.9 | 100.0 |
| 8 | Excitement,Mutism,Staring,Grimacing,Verbigeration,Rigidity,Waxy flexibility,Withdrawal | 99.6 | 98.9 | 100.0 |
| 8 | Excitement,Mutism,Staring,Mannerisms,Rigidity,Negativism,Waxy flexibility,Withdrawal | 99.6 | 98.9 | 100.0 |
| 8 | Excitement,Mutism,Staring,Mannerisms,Verbigeration,Rigidity,Negativism,Waxy flexibility | 99.6 | 98.9 | 100.0 |
| 8 | Excitement,Mutism,Staring,Mannerisms,Verbigeration,Rigidity,Waxy flexibility,Withdrawal | 99.6 | 98.9 | 100.0 |
| 8 | Excitement,Mutism,Staring,Posturing/catalepsy,Echopraxia/echolalia,Stereotypy,Mannerisms,Rigidity | 99.6 | 98.9 | 100.0 |
| 8 | Excitement,Mutism,Staring,Posturing/catalepsy,Echopraxia/echolalia,Stereotypy,Rigidity,Waxy flexibility | 99.6 | 98.9 | 100.0 |
| 8 | Excitement,Mutism,Staring,Posturing/catalepsy,Grimacing,Echopraxia/echolalia,Stereotypy,Rigidity | 99.6 | 98.9 | 100.0 |
| 8 | Excitement,Mutism,Staring,Posturing/catalepsy,Grimacing,Echopraxia/echolalia,Waxy flexibility,Withdrawal | 99.6 | 98.9 | 100.0 |
| 8 | Excitement,Mutism,Staring,Posturing/catalepsy,Grimacing,Mannerisms,Verbigeration,Negativism | 99.6 | 98.9 | 100.0 |
| 8 | Excitement,Mutism,Staring,Posturing/catalepsy,Grimacing,Negativism,Waxy flexibility,Withdrawal | 99.6 | 98.9 | 100.0 |
| 8 | Excitement,Mutism,Staring,Posturing/catalepsy,Grimacing,Stereotypy,Verbigeration,Negativism | 99.6 | 98.9 | 100.0 |
| 8 | Excitement,Mutism,Staring,Posturing/catalepsy,Grimacing,Stereotypy,Verbigeration,Rigidity | 99.6 | 98.9 | 100.0 |
| 8 | Excitement,Mutism,Staring,Posturing/catalepsy,Grimacing,Verbigeration,Negativism,Waxy flexibility | 99.6 | 98.9 | 100.0 |
| 8 | Excitement,Mutism,Staring,Posturing/catalepsy,Grimacing,Verbigeration,Rigidity,Negativism | 99.6 | 98.9 | 100.0 |
| 8 | Excitement,Mutism,Staring,Posturing/catalepsy,Stereotypy,Mannerisms,Verbigeration,Negativism | 99.6 | 98.9 | 100.0 |
| 8 | Excitement,Mutism,Staring,Posturing/catalepsy,Stereotypy,Mannerisms,Verbigeration,Rigidity | 99.6 | 98.9 | 100.0 |
| 8 | Excitement,Mutism,Staring,Posturing/catalepsy,Stereotypy,Verbigeration,Rigidity,Waxy flexibility | 99.6 | 98.9 | 100.0 |
| 8 | Excitement,Mutism,Staring,Posturing/catalepsy,Verbigeration,Rigidity,Negativism,Waxy flexibility | 99.6 | 98.9 | 100.0 |
| 8 | Excitement,Mutism,Staring,Stereotypy,Mannerisms,Rigidity,Negativism,Withdrawal | 99.6 | 98.9 | 100.0 |
| 8 | Excitement,Mutism,Staring,Verbigeration,Rigidity,Negativism,Waxy flexibility,Withdrawal | 99.6 | 98.9 | 100.0 |
| 8 | Excitement,Staring,Echopraxia/echolalia,Stereotypy,Mannerisms,Rigidity,Negativism,Withdrawal | 99.6 | 98.9 | 100.0 |
| 8 | Excitement,Staring,Posturing/catalepsy,Mannerisms,Verbigeration,Rigidity,Negativism,Withdrawal | 99.6 | 98.9 | 100.0 |
| 8 | Immobility/stupor,Staring,Grimacing,Echopraxia/echolalia,Mannerisms,Rigidity,Negativism,Withdrawal | 99.6 | 98.9 | 100.0 |
| 8 | Immobility/stupor,Staring,Grimacing,Stereotypy,Mannerisms,Rigidity,Negativism,Withdrawal | 99.6 | 98.9 | 100.0 |
| 8 | Mutism,Staring,Grimacing,Echopraxia/echolalia,Mannerisms,Rigidity,Negativism,Withdrawal | 99.6 | 98.9 | 100.0 |
| 8 | Mutism,Staring,Grimacing,Echopraxia/echolalia,Stereotypy,Mannerisms,Negativism,Withdrawal | 99.6 | 98.9 | 100.0 |
| 8 | Mutism,Staring,Posturing/catalepsy,Echopraxia/echolalia,Mannerisms,Rigidity,Negativism,Withdrawal | 99.6 | 98.9 | 100.0 |
| 8 | Mutism,Staring,Posturing/catalepsy,Stereotypy,Mannerisms,Rigidity,Negativism,Withdrawal | 99.6 | 98.9 | 100.0 |
| 8 | Excitement,Immobility/stupor,Mutism,Staring,Posturing/catalepsy,Echopraxia/echolalia,Mannerisms,Negativism | 99.5 | 98.9 | 100.0 |
| 8 | Excitement,Immobility/stupor,Mutism,Staring,Posturing/catalepsy,Echopraxia/echolalia,Rigidity,Waxy flexibility | 99.5 | 98.9 | 100.0 |
| 8 | Excitement,Immobility/stupor,Mutism,Staring,Posturing/catalepsy,Echopraxia/echolalia,Stereotypy,Rigidity | 99.5 | 98.9 | 100.0 |
| 8 | Excitement,Immobility/stupor,Mutism,Staring,Posturing/catalepsy,Grimacing,Verbigeration,Negativism | 99.5 | 98.9 | 100.0 |
| 8 | Excitement,Immobility/stupor,Mutism,Staring,Posturing/catalepsy,Stereotypy,Rigidity,Negativism | 99.5 | 98.9 | 100.0 |
| 8 | Excitement,Immobility/stupor,Mutism,Staring,Posturing/catalepsy,Stereotypy,Verbigeration,Rigidity | 99.5 | 98.9 | 100.0 |
| 8 | Excitement,Immobility/stupor,Mutism,Staring,Verbigeration,Rigidity,Waxy flexibility,Withdrawal | 99.5 | 98.9 | 100.0 |
| 8 | Excitement,Immobility/stupor,Staring,Echopraxia/echolalia,Stereotypy,Mannerisms,Rigidity,Withdrawal | 99.5 | 98.9 | 100.0 |
| 8 | Excitement,Immobility/stupor,Staring,Posturing/catalepsy,Echopraxia/echolalia,Negativism,Waxy flexibility,Withdrawal | 99.5 | 98.9 | 100.0 |
| 8 | Excitement,Immobility/stupor,Staring,Posturing/catalepsy,Echopraxia/echolalia,Verbigeration,Negativism,Withdrawal | 99.5 | 98.9 | 100.0 |
| 8 | Excitement,Immobility/stupor,Staring,Posturing/catalepsy,Echopraxia/echolalia,Verbigeration,Rigidity,Withdrawal | 99.5 | 98.9 | 100.0 |
| 8 | Excitement,Immobility/stupor,Staring,Posturing/catalepsy,Grimacing,Mannerisms,Rigidity,Withdrawal | 99.5 | 98.9 | 100.0 |
| 8 | Excitement,Immobility/stupor,Staring,Posturing/catalepsy,Mannerisms,Verbigeration,Negativism,Waxy flexibility | 99.5 | 98.9 | 100.0 |
| 8 | Excitement,Immobility/stupor,Staring,Stereotypy,Mannerisms,Rigidity,Waxy flexibility,Withdrawal | 99.5 | 98.9 | 100.0 |
| 8 | Excitement,Mutism,Staring,Echopraxia/echolalia,Mannerisms,Rigidity,Negativism,Waxy flexibility | 99.5 | 98.9 | 100.0 |
| 8 | Excitement,Mutism,Staring,Echopraxia/echolalia,Stereotypy,Verbigeration,Negativism,Withdrawal | 99.5 | 98.9 | 100.0 |
| 8 | Excitement,Mutism,Staring,Echopraxia/echolalia,Stereotypy,Verbigeration,Rigidity,Waxy flexibility | 99.5 | 98.9 | 100.0 |
| 8 | Excitement,Mutism,Staring,Echopraxia/echolalia,Verbigeration,Rigidity,Waxy flexibility,Withdrawal | 99.5 | 98.9 | 100.0 |
| 8 | Excitement,Mutism,Staring,Grimacing,Echopraxia/echolalia,Stereotypy,Negativism,Withdrawal | 99.5 | 98.9 | 100.0 |
| 8 | Excitement,Mutism,Staring,Grimacing,Echopraxia/echolalia,Verbigeration,Rigidity,Negativism | 99.5 | 98.9 | 100.0 |
| 8 | Excitement,Mutism,Staring,Grimacing,Echopraxia/echolalia,Verbigeration,Rigidity,Waxy flexibility | 99.5 | 98.9 | 100.0 |
| 8 | Excitement,Mutism,Staring,Grimacing,Mannerisms,Rigidity,Waxy flexibility,Withdrawal | 99.5 | 98.9 | 100.0 |
| 8 | Excitement,Mutism,Staring,Grimacing,Stereotypy,Verbigeration,Rigidity,Negativism | 99.5 | 98.9 | 100.0 |
| 8 | Excitement,Mutism,Staring,Posturing/catalepsy,Echopraxia/echolalia,Mannerisms,Rigidity,Waxy flexibility | 99.5 | 98.9 | 100.0 |
| 8 | Excitement,Mutism,Staring,Posturing/catalepsy,Echopraxia/echolalia,Stereotypy,Mannerisms,Negativism | 99.5 | 98.9 | 100.0 |
| 8 | Excitement,Mutism,Staring,Posturing/catalepsy,Grimacing,Echopraxia/echolalia,Mannerisms,Negativism | 99.5 | 98.9 | 100.0 |
| 8 | Excitement,Mutism,Staring,Posturing/catalepsy,Grimacing,Echopraxia/echolalia,Mannerisms,Rigidity | 99.5 | 98.9 | 100.0 |
| 8 | Excitement,Mutism,Staring,Posturing/catalepsy,Grimacing,Echopraxia/echolalia,Rigidity,Waxy flexibility | 99.5 | 98.9 | 100.0 |
| 8 | Excitement,Mutism,Staring,Posturing/catalepsy,Grimacing,Echopraxia/echolalia,Stereotypy,Negativism | 99.5 | 98.9 | 100.0 |
| 8 | Excitement,Mutism,Staring,Posturing/catalepsy,Grimacing,Mannerisms,Verbigeration,Rigidity | 99.5 | 98.9 | 100.0 |
| 8 | Excitement,Mutism,Staring,Posturing/catalepsy,Grimacing,Rigidity,Negativism,Withdrawal | 99.5 | 98.9 | 100.0 |
| 8 | Excitement,Mutism,Staring,Posturing/catalepsy,Grimacing,Rigidity,Waxy flexibility,Withdrawal | 99.5 | 98.9 | 100.0 |
| 8 | Excitement,Mutism,Staring,Posturing/catalepsy,Mannerisms,Verbigeration,Rigidity,Waxy flexibility | 99.5 | 98.9 | 100.0 |
| 8 | Excitement,Mutism,Staring,Posturing/catalepsy,Rigidity,Negativism,Waxy flexibility,Withdrawal | 99.5 | 98.9 | 100.0 |
| 8 | Excitement,Mutism,Staring,Stereotypy,Mannerisms,Verbigeration,Rigidity,Negativism | 99.5 | 98.9 | 100.0 |
| 8 | Excitement,Staring,Posturing/catalepsy,Echopraxia/echolalia,Mannerisms,Rigidity,Negativism,Withdrawal | 99.5 | 98.9 | 100.0 |
| 8 | Excitement,Staring,Stereotypy,Mannerisms,Verbigeration,Rigidity,Negativism,Withdrawal | 99.5 | 98.9 | 100.0 |
| 8 | Immobility/stupor,Staring,Posturing/catalepsy,Echopraxia/echolalia,Mannerisms,Rigidity,Negativism,Withdrawal | 99.5 | 98.9 | 100.0 |
| 8 | Immobility/stupor,Staring,Posturing/catalepsy,Stereotypy,Mannerisms,Rigidity,Negativism,Withdrawal | 99.5 | 98.9 | 100.0 |
| 8 | Immobility/stupor,Staring,Stereotypy,Mannerisms,Rigidity,Negativism,Waxy flexibility,Withdrawal | 99.5 | 98.9 | 100.0 |
| 8 | Mutism,Staring,Grimacing,Stereotypy,Mannerisms,Rigidity,Negativism,Withdrawal | 99.5 | 98.9 | 100.0 |
| 8 | Mutism,Staring,Posturing/catalepsy,Echopraxia/echolalia,Stereotypy,Mannerisms,Negativism,Withdrawal | 99.5 | 98.9 | 100.0 |
| 8 | Mutism,Staring,Posturing/catalepsy,Grimacing,Mannerisms,Verbigeration,Negativism,Withdrawal | 99.5 | 98.9 | 100.0 |
| 8 | Mutism,Staring,Posturing/catalepsy,Stereotypy,Mannerisms,Verbigeration,Negativism,Withdrawal | 99.5 | 98.9 | 100.0 |
| 8 | Mutism,Staring,Stereotypy,Mannerisms,Rigidity,Negativism,Waxy flexibility,Withdrawal | 99.5 | 98.9 | 100.0 |
| 8 | Staring,Grimacing,Echopraxia/echolalia,Stereotypy,Mannerisms,Rigidity,Negativism,Withdrawal | 99.5 | 98.9 | 100.0 |
| 8 | Staring,Grimacing,Stereotypy,Mannerisms,Verbigeration,Rigidity,Negativism,Withdrawal | 99.5 | 98.9 | 100.0 |
| 8 | Excitement,Immobility/stupor,Echopraxia/echolalia,Stereotypy,Verbigeration,Rigidity,Negativism,Withdrawal | 99.3 | 98.4 | 100.0 |
| 8 | Excitement,Immobility/stupor,Mutism,Echopraxia/echolalia,Stereotypy,Rigidity,Negativism,Withdrawal | 99.3 | 98.4 | 100.0 |
| 8 | Excitement,Immobility/stupor,Mutism,Echopraxia/echolalia,Stereotypy,Verbigeration,Rigidity,Negativism | 99.3 | 98.4 | 100.0 |
| 8 | Excitement,Immobility/stupor,Mutism,Echopraxia/echolalia,Verbigeration,Rigidity,Negativism,Withdrawal | 99.3 | 98.4 | 100.0 |
| 8 | Excitement,Immobility/stupor,Mutism,Grimacing,Echopraxia/echolalia,Rigidity,Negativism,Withdrawal | 99.3 | 98.4 | 100.0 |
| 8 | Excitement,Immobility/stupor,Mutism,Posturing/catalepsy,Echopraxia/echolalia,Rigidity,Negativism,Withdrawal | 99.3 | 98.4 | 100.0 |
| 8 | Excitement,Immobility/stupor,Mutism,Posturing/catalepsy,Echopraxia/echolalia,Verbigeration,Rigidity,Withdrawal | 99.3 | 98.4 | 100.0 |
| 8 | Excitement,Immobility/stupor,Mutism,Posturing/catalepsy,Grimacing,Echopraxia/echolalia,Rigidity,Negativism | 99.3 | 98.4 | 100.0 |
| 8 | Excitement,Immobility/stupor,Mutism,Posturing/catalepsy,Grimacing,Echopraxia/echolalia,Rigidity,Withdrawal | 99.3 | 98.4 | 100.0 |
| 8 | Excitement,Immobility/stupor,Mutism,Posturing/catalepsy,Grimacing,Verbigeration,Rigidity,Negativism | 99.3 | 98.4 | 100.0 |
| 8 | Excitement,Immobility/stupor,Mutism,Posturing/catalepsy,Mannerisms,Verbigeration,Rigidity,Negativism | 99.3 | 98.4 | 100.0 |
| 8 | Excitement,Immobility/stupor,Mutism,Posturing/catalepsy,Verbigeration,Rigidity,Negativism,Withdrawal | 99.3 | 98.4 | 100.0 |
| 8 | Excitement,Immobility/stupor,Mutism,Staring,Echopraxia/echolalia,Mannerisms,Rigidity,Negativism | 99.3 | 98.4 | 100.0 |
| 8 | Excitement,Immobility/stupor,Mutism,Staring,Echopraxia/echolalia,Mannerisms,Rigidity,Waxy flexibility | 99.3 | 98.4 | 100.0 |
| 8 | Excitement,Immobility/stupor,Mutism,Staring,Echopraxia/echolalia,Negativism,Waxy flexibility,Withdrawal | 99.3 | 98.4 | 100.0 |
| 8 | Excitement,Immobility/stupor,Mutism,Staring,Echopraxia/echolalia,Stereotypy,Negativism,Waxy flexibility | 99.3 | 98.4 | 100.0 |
| 8 | Excitement,Immobility/stupor,Mutism,Staring,Echopraxia/echolalia,Stereotypy,Verbigeration,Rigidity | 99.3 | 98.4 | 100.0 |
| 8 | Excitement,Immobility/stupor,Mutism,Staring,Echopraxia/echolalia,Verbigeration,Negativism,Waxy flexibility | 99.3 | 98.4 | 100.0 |
| 8 | Excitement,Immobility/stupor,Mutism,Staring,Grimacing,Echopraxia/echolalia,Rigidity,Negativism | 99.3 | 98.4 | 100.0 |
| 8 | Excitement,Immobility/stupor,Mutism,Staring,Grimacing,Echopraxia/echolalia,Rigidity,Waxy flexibility | 99.3 | 98.4 | 100.0 |
| 8 | Excitement,Immobility/stupor,Mutism,Staring,Grimacing,Rigidity,Waxy flexibility,Withdrawal | 99.3 | 98.4 | 100.0 |
| 8 | Excitement,Immobility/stupor,Mutism,Staring,Grimacing,Stereotypy,Rigidity,Waxy flexibility | 99.3 | 98.4 | 100.0 |
| 8 | Excitement,Immobility/stupor,Mutism,Staring,Grimacing,Stereotypy,Rigidity,Withdrawal | 99.3 | 98.4 | 100.0 |
| 8 | Excitement,Immobility/stupor,Mutism,Staring,Grimacing,Stereotypy,Verbigeration,Rigidity | 99.3 | 98.4 | 100.0 |
| 8 | Excitement,Immobility/stupor,Mutism,Staring,Grimacing,Verbigeration,Rigidity,Negativism | 99.3 | 98.4 | 100.0 |
| 8 | Excitement,Immobility/stupor,Mutism,Staring,Grimacing,Verbigeration,Rigidity,Waxy flexibility | 99.3 | 98.4 | 100.0 |
| 8 | Excitement,Immobility/stupor,Mutism,Staring,Grimacing,Verbigeration,Rigidity,Withdrawal | 99.3 | 98.4 | 100.0 |
| 8 | Excitement,Immobility/stupor,Mutism,Staring,Mannerisms,Rigidity,Negativism,Waxy flexibility | 99.3 | 98.4 | 100.0 |
| 8 | Excitement,Immobility/stupor,Mutism,Staring,Mannerisms,Rigidity,Negativism,Withdrawal | 99.3 | 98.4 | 100.0 |
| 8 | Excitement,Immobility/stupor,Mutism,Staring,Posturing/catalepsy,Echopraxia/echolalia,Waxy flexibility,Withdrawal | 99.3 | 98.4 | 100.0 |
| 8 | Excitement,Immobility/stupor,Mutism,Staring,Posturing/catalepsy,Grimacing,Echopraxia/echolalia,Withdrawal | 99.3 | 98.4 | 100.0 |
| 8 | Excitement,Immobility/stupor,Mutism,Staring,Posturing/catalepsy,Grimacing,Mannerisms,Negativism | 99.3 | 98.4 | 100.0 |
| 8 | Excitement,Immobility/stupor,Mutism,Staring,Posturing/catalepsy,Grimacing,Mannerisms,Rigidity | 99.3 | 98.4 | 100.0 |
| 8 | Excitement,Immobility/stupor,Mutism,Staring,Posturing/catalepsy,Grimacing,Negativism,Waxy flexibility | 99.3 | 98.4 | 100.0 |
| 8 | Excitement,Immobility/stupor,Mutism,Staring,Posturing/catalepsy,Grimacing,Rigidity,Negativism | 99.3 | 98.4 | 100.0 |
| 8 | Excitement,Immobility/stupor,Mutism,Staring,Posturing/catalepsy,Grimacing,Stereotypy,Negativism | 99.3 | 98.4 | 100.0 |
| 8 | Excitement,Immobility/stupor,Mutism,Staring,Posturing/catalepsy,Grimacing,Stereotypy,Rigidity | 99.3 | 98.4 | 100.0 |
| 8 | Excitement,Immobility/stupor,Mutism,Staring,Posturing/catalepsy,Mannerisms,Rigidity,Waxy flexibility | 99.3 | 98.4 | 100.0 |
| 8 | Excitement,Immobility/stupor,Mutism,Staring,Posturing/catalepsy,Rigidity,Negativism,Waxy flexibility | 99.3 | 98.4 | 100.0 |
| 8 | Excitement,Immobility/stupor,Mutism,Staring,Posturing/catalepsy,Stereotypy,Mannerisms,Negativism | 99.3 | 98.4 | 100.0 |
| 8 | Excitement,Immobility/stupor,Mutism,Staring,Posturing/catalepsy,Stereotypy,Mannerisms,Rigidity | 99.3 | 98.4 | 100.0 |
| 8 | Excitement,Immobility/stupor,Mutism,Staring,Posturing/catalepsy,Stereotypy,Rigidity,Waxy flexibility | 99.3 | 98.4 | 100.0 |
| 8 | Excitement,Immobility/stupor,Mutism,Staring,Rigidity,Negativism,Waxy flexibility,Withdrawal | 99.3 | 98.4 | 100.0 |
| 8 | Excitement,Immobility/stupor,Mutism,Staring,Stereotypy,Mannerisms,Rigidity,Waxy flexibility | 99.3 | 98.4 | 100.0 |
| 8 | Excitement,Immobility/stupor,Mutism,Staring,Stereotypy,Mannerisms,Rigidity,Withdrawal | 99.3 | 98.4 | 100.0 |
| 8 | Excitement,Immobility/stupor,Mutism,Staring,Stereotypy,Mannerisms,Verbigeration,Rigidity | 99.3 | 98.4 | 100.0 |
| 8 | Excitement,Immobility/stupor,Mutism,Staring,Stereotypy,Verbigeration,Negativism,Waxy flexibility | 99.3 | 98.4 | 100.0 |
| 8 | Excitement,Immobility/stupor,Mutism,Staring,Stereotypy,Verbigeration,Negativism,Withdrawal | 99.3 | 98.4 | 100.0 |
| 8 | Excitement,Immobility/stupor,Mutism,Stereotypy,Verbigeration,Rigidity,Negativism,Waxy flexibility | 99.3 | 98.4 | 100.0 |
| 8 | Excitement,Immobility/stupor,Posturing/catalepsy,Echopraxia/echolalia,Verbigeration,Rigidity,Negativism,Withdrawal | 99.3 | 98.4 | 100.0 |
| 8 | Excitement,Immobility/stupor,Posturing/catalepsy,Grimacing,Echopraxia/echolalia,Rigidity,Negativism,Withdrawal | 99.3 | 98.4 | 100.0 |
| 8 | Excitement,Immobility/stupor,Posturing/catalepsy,Mannerisms,Verbigeration,Rigidity,Negativism,Withdrawal | 99.3 | 98.4 | 100.0 |
| 8 | Excitement,Immobility/stupor,Posturing/catalepsy,Stereotypy,Verbigeration,Rigidity,Negativism,Withdrawal | 99.3 | 98.4 | 100.0 |
| 8 | Excitement,Immobility/stupor,Staring,Echopraxia/echolalia,Mannerisms,Rigidity,Negativism,Waxy flexibility | 99.3 | 98.4 | 100.0 |
| 8 | Excitement,Immobility/stupor,Staring,Echopraxia/echolalia,Mannerisms,Verbigeration,Rigidity,Negativism | 99.3 | 98.4 | 100.0 |
| 8 | Excitement,Immobility/stupor,Staring,Echopraxia/echolalia,Mannerisms,Verbigeration,Rigidity,Withdrawal | 99.3 | 98.4 | 100.0 |
| 8 | Excitement,Immobility/stupor,Staring,Echopraxia/echolalia,Stereotypy,Mannerisms,Negativism,Withdrawal | 99.3 | 98.4 | 100.0 |
| 8 | Excitement,Immobility/stupor,Staring,Echopraxia/echolalia,Stereotypy,Mannerisms,Rigidity,Negativism | 99.3 | 98.4 | 100.0 |
| 8 | Excitement,Immobility/stupor,Staring,Echopraxia/echolalia,Stereotypy,Rigidity,Negativism,Waxy flexibility | 99.3 | 98.4 | 100.0 |
| 8 | Excitement,Immobility/stupor,Staring,Echopraxia/echolalia,Stereotypy,Verbigeration,Rigidity,Negativism | 99.3 | 98.4 | 100.0 |
| 8 | Excitement,Immobility/stupor,Staring,Echopraxia/echolalia,Stereotypy,Verbigeration,Rigidity,Withdrawal | 99.3 | 98.4 | 100.0 |
| 8 | Excitement,Immobility/stupor,Staring,Echopraxia/echolalia,Verbigeration,Rigidity,Negativism,Waxy flexibility | 99.3 | 98.4 | 100.0 |
| 8 | Excitement,Immobility/stupor,Staring,Echopraxia/echolalia,Verbigeration,Rigidity,Waxy flexibility,Withdrawal | 99.3 | 98.4 | 100.0 |
| 8 | Excitement,Immobility/stupor,Staring,Grimacing,Echopraxia/echolalia,Mannerisms,Rigidity,Withdrawal | 99.3 | 98.4 | 100.0 |
| 8 | Excitement,Immobility/stupor,Staring,Grimacing,Echopraxia/echolalia,Rigidity,Waxy flexibility,Withdrawal | 99.3 | 98.4 | 100.0 |
| 8 | Excitement,Immobility/stupor,Staring,Grimacing,Echopraxia/echolalia,Stereotypy,Rigidity,Withdrawal | 99.3 | 98.4 | 100.0 |
| 8 | Excitement,Immobility/stupor,Staring,Grimacing,Mannerisms,Rigidity,Negativism,Withdrawal | 99.3 | 98.4 | 100.0 |
| 8 | Excitement,Immobility/stupor,Staring,Grimacing,Mannerisms,Rigidity,Waxy flexibility,Withdrawal | 99.3 | 98.4 | 100.0 |
| 8 | Excitement,Immobility/stupor,Staring,Grimacing,Mannerisms,Verbigeration,Rigidity,Negativism | 99.3 | 98.4 | 100.0 |
| 8 | Excitement,Immobility/stupor,Staring,Grimacing,Mannerisms,Verbigeration,Rigidity,Withdrawal | 99.3 | 98.4 | 100.0 |
| 8 | Excitement,Immobility/stupor,Staring,Grimacing,Stereotypy,Rigidity,Negativism,Withdrawal | 99.3 | 98.4 | 100.0 |
| 8 | Excitement,Immobility/stupor,Staring,Grimacing,Stereotypy,Rigidity,Waxy flexibility,Withdrawal | 99.3 | 98.4 | 100.0 |
| 8 | Excitement,Immobility/stupor,Staring,Grimacing,Stereotypy,Verbigeration,Rigidity,Negativism | 99.3 | 98.4 | 100.0 |
| 8 | Excitement,Immobility/stupor,Staring,Grimacing,Stereotypy,Verbigeration,Rigidity,Withdrawal | 99.3 | 98.4 | 100.0 |
| 8 | Excitement,Immobility/stupor,Staring,Grimacing,Verbigeration,Rigidity,Negativism,Withdrawal | 99.3 | 98.4 | 100.0 |
| 8 | Excitement,Immobility/stupor,Staring,Posturing/catalepsy,Echopraxia/echolalia,Mannerisms,Negativism,Waxy flexibility | 99.3 | 98.4 | 100.0 |
| 8 | Excitement,Immobility/stupor,Staring,Posturing/catalepsy,Echopraxia/echolalia,Mannerisms,Verbigeration,Negativism | 99.3 | 98.4 | 100.0 |
| 8 | Excitement,Immobility/stupor,Staring,Posturing/catalepsy,Echopraxia/echolalia,Mannerisms,Verbigeration,Rigidity | 99.3 | 98.4 | 100.0 |
| 8 | Excitement,Immobility/stupor,Staring,Posturing/catalepsy,Echopraxia/echolalia,Rigidity,Negativism,Waxy flexibility | 99.3 | 98.4 | 100.0 |
| 8 | Excitement,Immobility/stupor,Staring,Posturing/catalepsy,Echopraxia/echolalia,Stereotypy,Rigidity,Negativism | 99.3 | 98.4 | 100.0 |
| 8 | Excitement,Immobility/stupor,Staring,Posturing/catalepsy,Echopraxia/echolalia,Verbigeration,Negativism,Waxy flexibility | 99.3 | 98.4 | 100.0 |
| 8 | Excitement,Immobility/stupor,Staring,Posturing/catalepsy,Grimacing,Echopraxia/echolalia,Rigidity,Negativism | 99.3 | 98.4 | 100.0 |
| 8 | Excitement,Immobility/stupor,Staring,Posturing/catalepsy,Grimacing,Mannerisms,Negativism,Withdrawal | 99.3 | 98.4 | 100.0 |
| 8 | Excitement,Immobility/stupor,Staring,Posturing/catalepsy,Grimacing,Mannerisms,Rigidity,Negativism | 99.3 | 98.4 | 100.0 |
| 8 | Excitement,Immobility/stupor,Staring,Posturing/catalepsy,Grimacing,Mannerisms,Verbigeration,Negativism | 99.3 | 98.4 | 100.0 |
| 8 | Excitement,Immobility/stupor,Staring,Posturing/catalepsy,Grimacing,Mannerisms,Verbigeration,Rigidity | 99.3 | 98.4 | 100.0 |
| 8 | Excitement,Immobility/stupor,Staring,Posturing/catalepsy,Grimacing,Rigidity,Negativism,Withdrawal | 99.3 | 98.4 | 100.0 |
| 8 | Excitement,Immobility/stupor,Staring,Posturing/catalepsy,Grimacing,Stereotypy,Rigidity,Withdrawal | 99.3 | 98.4 | 100.0 |
| 8 | Excitement,Immobility/stupor,Staring,Posturing/catalepsy,Grimacing,Verbigeration,Negativism,Withdrawal | 99.3 | 98.4 | 100.0 |
| 8 | Excitement,Immobility/stupor,Staring,Posturing/catalepsy,Grimacing,Verbigeration,Rigidity,Negativism | 99.3 | 98.4 | 100.0 |
| 8 | Excitement,Immobility/stupor,Staring,Posturing/catalepsy,Grimacing,Verbigeration,Rigidity,Withdrawal | 99.3 | 98.4 | 100.0 |
| 8 | Excitement,Immobility/stupor,Staring,Posturing/catalepsy,Mannerisms,Negativism,Waxy flexibility,Withdrawal | 99.3 | 98.4 | 100.0 |
| 8 | Excitement,Immobility/stupor,Staring,Posturing/catalepsy,Mannerisms,Rigidity,Negativism,Waxy flexibility | 99.3 | 98.4 | 100.0 |
| 8 | Excitement,Immobility/stupor,Staring,Posturing/catalepsy,Mannerisms,Verbigeration,Rigidity,Waxy flexibility | 99.3 | 98.4 | 100.0 |
| 8 | Excitement,Immobility/stupor,Staring,Posturing/catalepsy,Rigidity,Negativism,Waxy flexibility,Withdrawal | 99.3 | 98.4 | 100.0 |
| 8 | Excitement,Immobility/stupor,Staring,Posturing/catalepsy,Stereotypy,Mannerisms,Negativism,Withdrawal | 99.3 | 98.4 | 100.0 |
| 8 | Excitement,Immobility/stupor,Staring,Posturing/catalepsy,Stereotypy,Mannerisms,Rigidity,Negativism | 99.3 | 98.4 | 100.0 |
| 8 | Excitement,Immobility/stupor,Staring,Posturing/catalepsy,Stereotypy,Mannerisms,Verbigeration,Negativism | 99.3 | 98.4 | 100.0 |
| 8 | Excitement,Immobility/stupor,Staring,Posturing/catalepsy,Stereotypy,Mannerisms,Verbigeration,Rigidity | 99.3 | 98.4 | 100.0 |
| 8 | Excitement,Immobility/stupor,Staring,Posturing/catalepsy,Stereotypy,Rigidity,Waxy flexibility,Withdrawal | 99.3 | 98.4 | 100.0 |
| 8 | Excitement,Immobility/stupor,Staring,Posturing/catalepsy,Stereotypy,Verbigeration,Negativism,Waxy flexibility | 99.3 | 98.4 | 100.0 |
| 8 | Excitement,Immobility/stupor,Staring,Posturing/catalepsy,Stereotypy,Verbigeration,Rigidity,Withdrawal | 99.3 | 98.4 | 100.0 |
| 8 | Excitement,Immobility/stupor,Staring,Posturing/catalepsy,Verbigeration,Negativism,Waxy flexibility,Withdrawal | 99.3 | 98.4 | 100.0 |
| 8 | Excitement,Immobility/stupor,Staring,Posturing/catalepsy,Verbigeration,Rigidity,Negativism,Waxy flexibility | 99.3 | 98.4 | 100.0 |
| 8 | Excitement,Immobility/stupor,Staring,Posturing/catalepsy,Verbigeration,Rigidity,Waxy flexibility,Withdrawal | 99.3 | 98.4 | 100.0 |
| 8 | Excitement,Immobility/stupor,Staring,Stereotypy,Mannerisms,Rigidity,Negativism,Waxy flexibility | 99.3 | 98.4 | 100.0 |
| 8 | Excitement,Immobility/stupor,Staring,Stereotypy,Mannerisms,Verbigeration,Rigidity,Waxy flexibility | 99.3 | 98.4 | 100.0 |
| 8 | Excitement,Immobility/stupor,Staring,Verbigeration,Rigidity,Negativism,Waxy flexibility,Withdrawal | 99.3 | 98.4 | 100.0 |
| 8 | Excitement,Mutism,Staring,Echopraxia/echolalia,Mannerisms,Negativism,Waxy flexibility,Withdrawal | 99.3 | 98.4 | 100.0 |
| 8 | Excitement,Mutism,Staring,Echopraxia/echolalia,Mannerisms,Verbigeration,Negativism,Waxy flexibility | 99.3 | 98.4 | 100.0 |
| 8 | Excitement,Mutism,Staring,Echopraxia/echolalia,Mannerisms,Verbigeration,Rigidity,Withdrawal | 99.3 | 98.4 | 100.0 |
| 8 | Excitement,Mutism,Staring,Echopraxia/echolalia,Stereotypy,Mannerisms,Negativism,Waxy flexibility | 99.3 | 98.4 | 100.0 |
| 8 | Excitement,Mutism,Staring,Echopraxia/echolalia,Stereotypy,Mannerisms,Rigidity,Waxy flexibility | 99.3 | 98.4 | 100.0 |
| 8 | Excitement,Mutism,Staring,Echopraxia/echolalia,Verbigeration,Negativism,Waxy flexibility,Withdrawal | 99.3 | 98.4 | 100.0 |
| 8 | Excitement,Mutism,Staring,Grimacing,Echopraxia/echolalia,Mannerisms,Rigidity,Negativism | 99.3 | 98.4 | 100.0 |
| 8 | Excitement,Mutism,Staring,Grimacing,Echopraxia/echolalia,Mannerisms,Rigidity,Waxy flexibility | 99.3 | 98.4 | 100.0 |
| 8 | Excitement,Mutism,Staring,Grimacing,Echopraxia/echolalia,Negativism,Waxy flexibility,Withdrawal | 99.3 | 98.4 | 100.0 |
| 8 | Excitement,Mutism,Staring,Grimacing,Echopraxia/echolalia,Stereotypy,Negativism,Waxy flexibility | 99.3 | 98.4 | 100.0 |
| 8 | Excitement,Mutism,Staring,Grimacing,Echopraxia/echolalia,Stereotypy,Verbigeration,Rigidity | 99.3 | 98.4 | 100.0 |
| 8 | Excitement,Mutism,Staring,Grimacing,Echopraxia/echolalia,Verbigeration,Negativism,Waxy flexibility | 99.3 | 98.4 | 100.0 |
| 8 | Excitement,Mutism,Staring,Grimacing,Mannerisms,Rigidity,Negativism,Withdrawal | 99.3 | 98.4 | 100.0 |
| 8 | Excitement,Mutism,Staring,Grimacing,Mannerisms,Verbigeration,Rigidity,Negativism | 99.3 | 98.4 | 100.0 |
| 8 | Excitement,Mutism,Staring,Grimacing,Mannerisms,Verbigeration,Rigidity,Waxy flexibility | 99.3 | 98.4 | 100.0 |
| 8 | Excitement,Mutism,Staring,Grimacing,Rigidity,Negativism,Waxy flexibility,Withdrawal | 99.3 | 98.4 | 100.0 |
| 8 | Excitement,Mutism,Staring,Grimacing,Stereotypy,Mannerisms,Rigidity,Withdrawal | 99.3 | 98.4 | 100.0 |
| 8 | Excitement,Mutism,Staring,Grimacing,Stereotypy,Rigidity,Negativism,Waxy flexibility | 99.3 | 98.4 | 100.0 |
| 8 | Excitement,Mutism,Staring,Grimacing,Stereotypy,Verbigeration,Negativism,Withdrawal | 99.3 | 98.4 | 100.0 |
| 8 | Excitement,Mutism,Staring,Grimacing,Verbigeration,Rigidity,Negativism,Waxy flexibility | 99.3 | 98.4 | 100.0 |
| 8 | Excitement,Mutism,Staring,Posturing/catalepsy,Echopraxia/echolalia,Mannerisms,Waxy flexibility,Withdrawal | 99.3 | 98.4 | 100.0 |
| 8 | Excitement,Mutism,Staring,Posturing/catalepsy,Echopraxia/echolalia,Stereotypy,Waxy flexibility,Withdrawal | 99.3 | 98.4 | 100.0 |
| 8 | Excitement,Mutism,Staring,Posturing/catalepsy,Echopraxia/echolalia,Verbigeration,Waxy flexibility,Withdrawal | 99.3 | 98.4 | 100.0 |
| 8 | Excitement,Mutism,Staring,Posturing/catalepsy,Grimacing,Echopraxia/echolalia,Mannerisms,Withdrawal | 99.3 | 98.4 | 100.0 |
| 8 | Excitement,Mutism,Staring,Posturing/catalepsy,Grimacing,Echopraxia/echolalia,Stereotypy,Withdrawal | 99.3 | 98.4 | 100.0 |
| 8 | Excitement,Mutism,Staring,Posturing/catalepsy,Grimacing,Echopraxia/echolalia,Verbigeration,Withdrawal | 99.3 | 98.4 | 100.0 |
| 8 | Excitement,Mutism,Staring,Posturing/catalepsy,Grimacing,Mannerisms,Negativism,Waxy flexibility | 99.3 | 98.4 | 100.0 |
| 8 | Excitement,Mutism,Staring,Posturing/catalepsy,Grimacing,Mannerisms,Rigidity,Negativism | 99.3 | 98.4 | 100.0 |
| 8 | Excitement,Mutism,Staring,Posturing/catalepsy,Grimacing,Stereotypy,Negativism,Waxy flexibility | 99.3 | 98.4 | 100.0 |
| 8 | Excitement,Mutism,Staring,Posturing/catalepsy,Grimacing,Stereotypy,Rigidity,Negativism | 99.3 | 98.4 | 100.0 |
| 8 | Excitement,Mutism,Staring,Posturing/catalepsy,Grimacing,Verbigeration,Rigidity,Waxy flexibility | 99.3 | 98.4 | 100.0 |
| 8 | Excitement,Mutism,Staring,Posturing/catalepsy,Mannerisms,Rigidity,Negativism,Waxy flexibility | 99.3 | 98.4 | 100.0 |
| 8 | Excitement,Mutism,Staring,Posturing/catalepsy,Stereotypy,Mannerisms,Negativism,Waxy flexibility | 99.3 | 98.4 | 100.0 |
| 8 | Excitement,Mutism,Staring,Posturing/catalepsy,Stereotypy,Mannerisms,Rigidity,Negativism | 99.3 | 98.4 | 100.0 |
| 8 | Excitement,Mutism,Staring,Posturing/catalepsy,Stereotypy,Rigidity,Negativism,Waxy flexibility | 99.3 | 98.4 | 100.0 |
| 8 | Excitement,Mutism,Staring,Stereotypy,Mannerisms,Rigidity,Negativism,Waxy flexibility | 99.3 | 98.4 | 100.0 |
| 8 | Excitement,Mutism,Staring,Stereotypy,Mannerisms,Verbigeration,Negativism,Withdrawal | 99.3 | 98.4 | 100.0 |
| 8 | Excitement,Mutism,Staring,Stereotypy,Mannerisms,Verbigeration,Rigidity,Waxy flexibility | 99.3 | 98.4 | 100.0 |
| 8 | Excitement,Mutism,Staring,Stereotypy,Verbigeration,Negativism,Waxy flexibility,Withdrawal | 99.3 | 98.4 | 100.0 |
| 8 | Excitement,Staring,Echopraxia/echolalia,Mannerisms,Rigidity,Negativism,Waxy flexibility,Withdrawal | 99.3 | 98.4 | 100.0 |
| 8 | Excitement,Staring,Mannerisms,Verbigeration,Rigidity,Negativism,Waxy flexibility,Withdrawal | 99.3 | 98.4 | 100.0 |
| 8 | Excitement,Staring,Posturing/catalepsy,Grimacing,Echopraxia/echolalia,Mannerisms,Rigidity,Withdrawal | 99.3 | 98.4 | 100.0 |
| 8 | Excitement,Staring,Posturing/catalepsy,Grimacing,Mannerisms,Rigidity,Negativism,Withdrawal | 99.3 | 98.4 | 100.0 |
| 8 | Excitement,Staring,Posturing/catalepsy,Grimacing,Mannerisms,Verbigeration,Rigidity,Withdrawal | 99.3 | 98.4 | 100.0 |
| 8 | Excitement,Staring,Posturing/catalepsy,Mannerisms,Rigidity,Negativism,Waxy flexibility,Withdrawal | 99.3 | 98.4 | 100.0 |
| 8 | Excitement,Staring,Posturing/catalepsy,Stereotypy,Mannerisms,Rigidity,Negativism,Withdrawal | 99.3 | 98.4 | 100.0 |
| 8 | Excitement,Staring,Stereotypy,Mannerisms,Rigidity,Negativism,Waxy flexibility,Withdrawal | 99.3 | 98.4 | 100.0 |
| 8 | Immobility/stupor,Mutism,Staring,Echopraxia/echolalia,Mannerisms,Rigidity,Negativism,Withdrawal | 99.3 | 98.4 | 100.0 |
| 8 | Immobility/stupor,Mutism,Staring,Posturing/catalepsy,Echopraxia/echolalia,Mannerisms,Negativism,Withdrawal | 99.3 | 98.4 | 100.0 |
| 8 | Immobility/stupor,Mutism,Staring,Posturing/catalepsy,Grimacing,Mannerisms,Negativism,Withdrawal | 99.3 | 98.4 | 100.0 |
| 8 | Immobility/stupor,Mutism,Staring,Posturing/catalepsy,Stereotypy,Mannerisms,Negativism,Withdrawal | 99.3 | 98.4 | 100.0 |
| 8 | Immobility/stupor,Mutism,Staring,Stereotypy,Mannerisms,Rigidity,Negativism,Withdrawal | 99.3 | 98.4 | 100.0 |
| 8 | Immobility/stupor,Mutism,Staring,Stereotypy,Mannerisms,Verbigeration,Rigidity,Negativism | 99.3 | 98.4 | 100.0 |
| 8 | Immobility/stupor,Staring,Echopraxia/echolalia,Mannerisms,Rigidity,Negativism,Waxy flexibility,Withdrawal | 99.3 | 98.4 | 100.0 |
| 8 | Immobility/stupor,Staring,Echopraxia/echolalia,Mannerisms,Verbigeration,Rigidity,Negativism,Withdrawal | 99.3 | 98.4 | 100.0 |
| 8 | Immobility/stupor,Staring,Grimacing,Echopraxia/echolalia,Stereotypy,Mannerisms,Negativism,Withdrawal | 99.3 | 98.4 | 100.0 |
| 8 | Immobility/stupor,Staring,Grimacing,Mannerisms,Verbigeration,Rigidity,Negativism,Withdrawal | 99.3 | 98.4 | 100.0 |
| 8 | Immobility/stupor,Staring,Grimacing,Stereotypy,Mannerisms,Verbigeration,Rigidity,Negativism | 99.3 | 98.4 | 100.0 |
| 8 | Immobility/stupor,Staring,Posturing/catalepsy,Echopraxia/echolalia,Stereotypy,Mannerisms,Negativism,Withdrawal | 99.3 | 98.4 | 100.0 |
| 8 | Immobility/stupor,Staring,Posturing/catalepsy,Grimacing,Echopraxia/echolalia,Mannerisms,Negativism,Withdrawal | 99.3 | 98.4 | 100.0 |
| 8 | Immobility/stupor,Staring,Posturing/catalepsy,Grimacing,Mannerisms,Rigidity,Negativism,Withdrawal | 99.3 | 98.4 | 100.0 |
| 8 | Immobility/stupor,Staring,Posturing/catalepsy,Grimacing,Stereotypy,Mannerisms,Negativism,Withdrawal | 99.3 | 98.4 | 100.0 |
| 8 | Immobility/stupor,Staring,Posturing/catalepsy,Mannerisms,Verbigeration,Rigidity,Negativism,Withdrawal | 99.3 | 98.4 | 100.0 |
| 8 | Immobility/stupor,Staring,Posturing/catalepsy,Stereotypy,Mannerisms,Verbigeration,Negativism,Withdrawal | 99.3 | 98.4 | 100.0 |
| 8 | Immobility/stupor,Staring,Posturing/catalepsy,Stereotypy,Mannerisms,Verbigeration,Rigidity,Negativism | 99.3 | 98.4 | 100.0 |
| 8 | Immobility/stupor,Staring,Stereotypy,Mannerisms,Verbigeration,Rigidity,Negativism,Waxy flexibility | 99.3 | 98.4 | 100.0 |
| 8 | Mutism,Staring,Echopraxia/echolalia,Mannerisms,Rigidity,Negativism,Waxy flexibility,Withdrawal | 99.3 | 98.4 | 100.0 |
| 8 | Mutism,Staring,Echopraxia/echolalia,Mannerisms,Verbigeration,Rigidity,Negativism,Withdrawal | 99.3 | 98.4 | 100.0 |
| 8 | Mutism,Staring,Echopraxia/echolalia,Stereotypy,Mannerisms,Rigidity,Negativism,Waxy flexibility | 99.3 | 98.4 | 100.0 |
| 8 | Mutism,Staring,Echopraxia/echolalia,Stereotypy,Mannerisms,Verbigeration,Rigidity,Negativism | 99.3 | 98.4 | 100.0 |
| 8 | Mutism,Staring,Grimacing,Echopraxia/echolalia,Stereotypy,Mannerisms,Rigidity,Negativism | 99.3 | 98.4 | 100.0 |
| 8 | Mutism,Staring,Grimacing,Echopraxia/echolalia,Stereotypy,Mannerisms,Rigidity,Withdrawal | 99.3 | 98.4 | 100.0 |
| 8 | Mutism,Staring,Grimacing,Mannerisms,Verbigeration,Rigidity,Negativism,Withdrawal | 99.3 | 98.4 | 100.0 |
| 8 | Mutism,Staring,Grimacing,Stereotypy,Mannerisms,Rigidity,Waxy flexibility,Withdrawal | 99.3 | 98.4 | 100.0 |
| 8 | Mutism,Staring,Grimacing,Stereotypy,Mannerisms,Verbigeration,Negativism,Withdrawal | 99.3 | 98.4 | 100.0 |
| 8 | Mutism,Staring,Grimacing,Stereotypy,Mannerisms,Verbigeration,Rigidity,Negativism | 99.3 | 98.4 | 100.0 |
| 8 | Mutism,Staring,Grimacing,Stereotypy,Mannerisms,Verbigeration,Rigidity,Withdrawal | 99.3 | 98.4 | 100.0 |
| 8 | Mutism,Staring,Grimacing,Stereotypy,Verbigeration,Rigidity,Negativism,Withdrawal | 99.3 | 98.4 | 100.0 |
| 8 | Mutism,Staring,Posturing/catalepsy,Echopraxia/echolalia,Mannerisms,Negativism,Waxy flexibility,Withdrawal | 99.3 | 98.4 | 100.0 |
| 8 | Mutism,Staring,Posturing/catalepsy,Echopraxia/echolalia,Mannerisms,Verbigeration,Negativism,Withdrawal | 99.3 | 98.4 | 100.0 |
| 8 | Mutism,Staring,Posturing/catalepsy,Echopraxia/echolalia,Mannerisms,Verbigeration,Rigidity,Negativism | 99.3 | 98.4 | 100.0 |
| 8 | Mutism,Staring,Posturing/catalepsy,Echopraxia/echolalia,Stereotypy,Mannerisms,Rigidity,Negativism | 99.3 | 98.4 | 100.0 |
| 8 | Mutism,Staring,Posturing/catalepsy,Grimacing,Echopraxia/echolalia,Mannerisms,Rigidity,Negativism | 99.3 | 98.4 | 100.0 |
| 8 | Mutism,Staring,Posturing/catalepsy,Grimacing,Echopraxia/echolalia,Mannerisms,Verbigeration,Negativism | 99.3 | 98.4 | 100.0 |
| 8 | Mutism,Staring,Posturing/catalepsy,Grimacing,Echopraxia/echolalia,Stereotypy,Mannerisms,Negativism | 99.3 | 98.4 | 100.0 |
| 8 | Mutism,Staring,Posturing/catalepsy,Grimacing,Mannerisms,Negativism,Waxy flexibility,Withdrawal | 99.3 | 98.4 | 100.0 |
| 8 | Mutism,Staring,Posturing/catalepsy,Grimacing,Mannerisms,Rigidity,Negativism,Withdrawal | 99.3 | 98.4 | 100.0 |
| 8 | Mutism,Staring,Posturing/catalepsy,Grimacing,Stereotypy,Mannerisms,Rigidity,Withdrawal | 99.3 | 98.4 | 100.0 |
| 8 | Mutism,Staring,Posturing/catalepsy,Mannerisms,Verbigeration,Rigidity,Negativism,Withdrawal | 99.3 | 98.4 | 100.0 |
| 8 | Mutism,Staring,Posturing/catalepsy,Stereotypy,Mannerisms,Negativism,Waxy flexibility,Withdrawal | 99.3 | 98.4 | 100.0 |
| 8 | Mutism,Staring,Posturing/catalepsy,Stereotypy,Mannerisms,Verbigeration,Rigidity,Negativism | 99.3 | 98.4 | 100.0 |
| 8 | Mutism,Staring,Stereotypy,Mannerisms,Verbigeration,Rigidity,Negativism,Waxy flexibility | 99.3 | 98.4 | 100.0 |
| 8 | Staring,Echopraxia/echolalia,Stereotypy,Mannerisms,Rigidity,Negativism,Waxy flexibility,Withdrawal | 99.3 | 98.4 | 100.0 |
| 8 | Staring,Echopraxia/echolalia,Stereotypy,Mannerisms,Verbigeration,Rigidity,Negativism,Withdrawal | 99.3 | 98.4 | 100.0 |
| 8 | Staring,Grimacing,Stereotypy,Mannerisms,Rigidity,Negativism,Waxy flexibility,Withdrawal | 99.3 | 98.4 | 100.0 |
| 8 | Staring,Posturing/catalepsy,Echopraxia/echolalia,Stereotypy,Mannerisms,Rigidity,Negativism,Withdrawal | 99.3 | 98.4 | 100.0 |
| 8 | Staring,Posturing/catalepsy,Grimacing,Echopraxia/echolalia,Mannerisms,Rigidity,Negativism,Withdrawal | 99.3 | 98.4 | 100.0 |
| 8 | Staring,Posturing/catalepsy,Grimacing,Stereotypy,Mannerisms,Rigidity,Negativism,Withdrawal | 99.3 | 98.4 | 100.0 |
| 8 | Staring,Posturing/catalepsy,Stereotypy,Mannerisms,Verbigeration,Rigidity,Negativism,Withdrawal | 99.3 | 98.4 | 100.0 |
| 8 | Staring,Stereotypy,Mannerisms,Verbigeration,Rigidity,Negativism,Waxy flexibility,Withdrawal | 99.3 | 98.4 | 100.0 |
| 8 | Excitement,Immobility/stupor,Echopraxia/echolalia,Mannerisms,Verbigeration,Rigidity,Negativism,Withdrawal | 99.1 | 98.2 | 99.8 |
| 8 | Excitement,Immobility/stupor,Echopraxia/echolalia,Stereotypy,Mannerisms,Rigidity,Negativism,Withdrawal | 99.1 | 98.2 | 99.8 |
| 8 | Excitement,Immobility/stupor,Grimacing,Echopraxia/echolalia,Mannerisms,Rigidity,Negativism,Withdrawal | 99.1 | 98.2 | 99.8 |
| 8 | Excitement,Immobility/stupor,Grimacing,Echopraxia/echolalia,Stereotypy,Rigidity,Negativism,Withdrawal | 99.1 | 98.2 | 99.8 |
| 8 | Excitement,Immobility/stupor,Grimacing,Echopraxia/echolalia,Verbigeration,Rigidity,Negativism,Withdrawal | 99.1 | 98.2 | 99.8 |
| 8 | Excitement,Immobility/stupor,Mutism,Echopraxia/echolalia,Stereotypy,Verbigeration,Rigidity,Withdrawal | 99.1 | 98.2 | 99.8 |
| 8 | Excitement,Immobility/stupor,Mutism,Echopraxia/echolalia,Verbigeration,Rigidity,Negativism,Waxy flexibility | 99.1 | 98.2 | 99.8 |
| 8 | Excitement,Immobility/stupor,Mutism,Grimacing,Echopraxia/echolalia,Verbigeration,Rigidity,Negativism | 99.1 | 98.2 | 99.8 |
| 8 | Excitement,Immobility/stupor,Mutism,Grimacing,Stereotypy,Verbigeration,Rigidity,Negativism | 99.1 | 98.2 | 99.8 |
| 8 | Excitement,Immobility/stupor,Mutism,Grimacing,Verbigeration,Rigidity,Negativism,Withdrawal | 99.1 | 98.2 | 99.8 |
| 8 | Excitement,Immobility/stupor,Mutism,Mannerisms,Verbigeration,Rigidity,Negativism,Withdrawal | 99.1 | 98.2 | 99.8 |
| 8 | Excitement,Immobility/stupor,Mutism,Posturing/catalepsy,Echopraxia/echolalia,Stereotypy,Rigidity,Negativism | 99.1 | 98.2 | 99.8 |
| 8 | Excitement,Immobility/stupor,Mutism,Posturing/catalepsy,Echopraxia/echolalia,Stereotypy,Rigidity,Withdrawal | 99.1 | 98.2 | 99.8 |
| 8 | Excitement,Immobility/stupor,Mutism,Posturing/catalepsy,Echopraxia/echolalia,Stereotypy,Verbigeration,Rigidity | 99.1 | 98.2 | 99.8 |
| 8 | Excitement,Immobility/stupor,Mutism,Posturing/catalepsy,Grimacing,Echopraxia/echolalia,Verbigeration,Rigidity | 99.1 | 98.2 | 99.8 |
| 8 | Excitement,Immobility/stupor,Mutism,Posturing/catalepsy,Grimacing,Rigidity,Negativism,Withdrawal | 99.1 | 98.2 | 99.8 |
| 8 | Excitement,Immobility/stupor,Mutism,Posturing/catalepsy,Grimacing,Verbigeration,Rigidity,Withdrawal | 99.1 | 98.2 | 99.8 |
| 8 | Excitement,Immobility/stupor,Mutism,Posturing/catalepsy,Mannerisms,Verbigeration,Rigidity,Withdrawal | 99.1 | 98.2 | 99.8 |
| 8 | Excitement,Immobility/stupor,Mutism,Posturing/catalepsy,Stereotypy,Rigidity,Negativism,Withdrawal | 99.1 | 98.2 | 99.8 |
| 8 | Excitement,Immobility/stupor,Mutism,Posturing/catalepsy,Stereotypy,Verbigeration,Rigidity,Withdrawal | 99.1 | 98.2 | 99.8 |
| 8 | Excitement,Immobility/stupor,Mutism,Posturing/catalepsy,Verbigeration,Rigidity,Negativism,Waxy flexibility | 99.1 | 98.2 | 99.8 |
| 8 | Excitement,Immobility/stupor,Mutism,Staring,Echopraxia/echolalia,Mannerisms,Negativism,Waxy flexibility | 99.1 | 98.2 | 99.8 |
| 8 | Excitement,Immobility/stupor,Mutism,Staring,Echopraxia/echolalia,Mannerisms,Negativism,Withdrawal | 99.1 | 98.2 | 99.8 |
| 8 | Excitement,Immobility/stupor,Mutism,Staring,Echopraxia/echolalia,Mannerisms,Verbigeration,Rigidity | 99.1 | 98.2 | 99.8 |
| 8 | Excitement,Immobility/stupor,Mutism,Staring,Echopraxia/echolalia,Stereotypy,Mannerisms,Rigidity | 99.1 | 98.2 | 99.8 |
| 8 | Excitement,Immobility/stupor,Mutism,Staring,Echopraxia/echolalia,Stereotypy,Verbigeration,Negativism | 99.1 | 98.2 | 99.8 |
| 8 | Excitement,Immobility/stupor,Mutism,Staring,Echopraxia/echolalia,Verbigeration,Negativism,Withdrawal | 99.1 | 98.2 | 99.8 |
| 8 | Excitement,Immobility/stupor,Mutism,Staring,Grimacing,Echopraxia/echolalia,Negativism,Waxy flexibility | 99.1 | 98.2 | 99.8 |
| 8 | Excitement,Immobility/stupor,Mutism,Staring,Grimacing,Echopraxia/echolalia,Negativism,Withdrawal | 99.1 | 98.2 | 99.8 |
| 8 | Excitement,Immobility/stupor,Mutism,Staring,Grimacing,Echopraxia/echolalia,Stereotypy,Rigidity | 99.1 | 98.2 | 99.8 |
| 8 | Excitement,Immobility/stupor,Mutism,Staring,Grimacing,Echopraxia/echolalia,Verbigeration,Rigidity | 99.1 | 98.2 | 99.8 |
| 8 | Excitement,Immobility/stupor,Mutism,Staring,Grimacing,Mannerisms,Rigidity,Waxy flexibility | 99.1 | 98.2 | 99.8 |
| 8 | Excitement,Immobility/stupor,Mutism,Staring,Grimacing,Mannerisms,Rigidity,Withdrawal | 99.1 | 98.2 | 99.8 |
| 8 | Excitement,Immobility/stupor,Mutism,Staring,Grimacing,Mannerisms,Verbigeration,Rigidity | 99.1 | 98.2 | 99.8 |
| 8 | Excitement,Immobility/stupor,Mutism,Staring,Grimacing,Rigidity,Negativism,Waxy flexibility | 99.1 | 98.2 | 99.8 |
| 8 | Excitement,Immobility/stupor,Mutism,Staring,Grimacing,Rigidity,Negativism,Withdrawal | 99.1 | 98.2 | 99.8 |
| 8 | Excitement,Immobility/stupor,Mutism,Staring,Grimacing,Stereotypy,Rigidity,Negativism | 99.1 | 98.2 | 99.8 |
| 8 | Excitement,Immobility/stupor,Mutism,Staring,Mannerisms,Verbigeration,Negativism,Waxy flexibility | 99.1 | 98.2 | 99.8 |
| 8 | Excitement,Immobility/stupor,Mutism,Staring,Posturing/catalepsy,Echopraxia/echolalia,Mannerisms,Withdrawal | 99.1 | 98.2 | 99.8 |
| 8 | Excitement,Immobility/stupor,Mutism,Staring,Posturing/catalepsy,Echopraxia/echolalia,Stereotypy,Withdrawal | 99.1 | 98.2 | 99.8 |
| 8 | Excitement,Immobility/stupor,Mutism,Staring,Posturing/catalepsy,Echopraxia/echolalia,Verbigeration,Waxy flexibility | 99.1 | 98.2 | 99.8 |
| 8 | Excitement,Immobility/stupor,Mutism,Staring,Posturing/catalepsy,Echopraxia/echolalia,Verbigeration,Withdrawal | 99.1 | 98.2 | 99.8 |
| 8 | Excitement,Immobility/stupor,Mutism,Staring,Posturing/catalepsy,Grimacing,Echopraxia/echolalia,Waxy flexibility | 99.1 | 98.2 | 99.8 |
| 8 | Excitement,Immobility/stupor,Mutism,Staring,Stereotypy,Mannerisms,Rigidity,Negativism | 99.1 | 98.2 | 99.8 |
| 8 | Excitement,Immobility/stupor,Mutism,Staring,Stereotypy,Negativism,Waxy flexibility,Withdrawal | 99.1 | 98.2 | 99.8 |
| 8 | Excitement,Immobility/stupor,Mutism,Stereotypy,Mannerisms,Verbigeration,Rigidity,Negativism | 99.1 | 98.2 | 99.8 |
| 8 | Excitement,Immobility/stupor,Mutism,Stereotypy,Verbigeration,Rigidity,Waxy flexibility,Withdrawal | 99.1 | 98.2 | 99.8 |
| 8 | Excitement,Immobility/stupor,Posturing/catalepsy,Echopraxia/echolalia,Mannerisms,Rigidity,Negativism,Withdrawal | 99.1 | 98.2 | 99.8 |
| 8 | Excitement,Immobility/stupor,Posturing/catalepsy,Echopraxia/echolalia,Stereotypy,Rigidity,Negativism,Withdrawal | 99.1 | 98.2 | 99.8 |
| 8 | Excitement,Immobility/stupor,Posturing/catalepsy,Grimacing,Mannerisms,Rigidity,Negativism,Withdrawal | 99.1 | 98.2 | 99.8 |
| 8 | Excitement,Immobility/stupor,Posturing/catalepsy,Grimacing,Verbigeration,Rigidity,Negativism,Withdrawal | 99.1 | 98.2 | 99.8 |
| 8 | Excitement,Immobility/stupor,Staring,Echopraxia/echolalia,Mannerisms,Negativism,Waxy flexibility,Withdrawal | 99.1 | 98.2 | 99.8 |
| 8 | Excitement,Immobility/stupor,Staring,Echopraxia/echolalia,Mannerisms,Verbigeration,Rigidity,Waxy flexibility | 99.1 | 98.2 | 99.8 |
| 8 | Excitement,Immobility/stupor,Staring,Echopraxia/echolalia,Stereotypy,Negativism,Waxy flexibility,Withdrawal | 99.1 | 98.2 | 99.8 |
| 8 | Excitement,Immobility/stupor,Staring,Echopraxia/echolalia,Stereotypy,Verbigeration,Negativism,Withdrawal | 99.1 | 98.2 | 99.8 |
| 8 | Excitement,Immobility/stupor,Staring,Echopraxia/echolalia,Stereotypy,Verbigeration,Rigidity,Waxy flexibility | 99.1 | 98.2 | 99.8 |
| 8 | Excitement,Immobility/stupor,Staring,Grimacing,Echopraxia/echolalia,Mannerisms,Rigidity,Negativism | 99.1 | 98.2 | 99.8 |
| 8 | Excitement,Immobility/stupor,Staring,Grimacing,Echopraxia/echolalia,Rigidity,Negativism,Waxy flexibility | 99.1 | 98.2 | 99.8 |
| 8 | Excitement,Immobility/stupor,Staring,Grimacing,Echopraxia/echolalia,Stereotypy,Negativism,Withdrawal | 99.1 | 98.2 | 99.8 |
| 8 | Excitement,Immobility/stupor,Staring,Grimacing,Echopraxia/echolalia,Stereotypy,Rigidity,Negativism | 99.1 | 98.2 | 99.8 |
| 8 | Excitement,Immobility/stupor,Staring,Grimacing,Echopraxia/echolalia,Verbigeration,Rigidity,Negativism | 99.1 | 98.2 | 99.8 |
| 8 | Excitement,Immobility/stupor,Staring,Grimacing,Echopraxia/echolalia,Verbigeration,Rigidity,Withdrawal | 99.1 | 98.2 | 99.8 |
| 8 | Excitement,Immobility/stupor,Staring,Grimacing,Mannerisms,Rigidity,Negativism,Waxy flexibility | 99.1 | 98.2 | 99.8 |
| 8 | Excitement,Immobility/stupor,Staring,Grimacing,Mannerisms,Verbigeration,Rigidity,Waxy flexibility | 99.1 | 98.2 | 99.8 |
| 8 | Excitement,Immobility/stupor,Staring,Grimacing,Rigidity,Negativism,Waxy flexibility,Withdrawal | 99.1 | 98.2 | 99.8 |
| 8 | Excitement,Immobility/stupor,Staring,Grimacing,Stereotypy,Mannerisms,Rigidity,Withdrawal | 99.1 | 98.2 | 99.8 |
| 8 | Excitement,Immobility/stupor,Staring,Grimacing,Stereotypy,Rigidity,Negativism,Waxy flexibility | 99.1 | 98.2 | 99.8 |
| 8 | Excitement,Immobility/stupor,Staring,Grimacing,Stereotypy,Verbigeration,Rigidity,Waxy flexibility | 99.1 | 98.2 | 99.8 |
| 8 | Excitement,Immobility/stupor,Staring,Grimacing,Verbigeration,Rigidity,Negativism,Waxy flexibility | 99.1 | 98.2 | 99.8 |
| 8 | Excitement,Immobility/stupor,Staring,Grimacing,Verbigeration,Rigidity,Waxy flexibility,Withdrawal | 99.1 | 98.2 | 99.8 |
| 8 | Excitement,Immobility/stupor,Staring,Posturing/catalepsy,Echopraxia/echolalia,Stereotypy,Mannerisms,Negativism | 99.1 | 98.2 | 99.8 |
| 8 | Excitement,Immobility/stupor,Staring,Posturing/catalepsy,Echopraxia/echolalia,Stereotypy,Negativism,Waxy flexibility | 99.1 | 98.2 | 99.8 |
| 8 | Excitement,Immobility/stupor,Staring,Posturing/catalepsy,Echopraxia/echolalia,Stereotypy,Verbigeration,Negativism | 99.1 | 98.2 | 99.8 |
| 8 | Excitement,Immobility/stupor,Staring,Posturing/catalepsy,Echopraxia/echolalia,Stereotypy,Verbigeration,Rigidity | 99.1 | 98.2 | 99.8 |
| 8 | Excitement,Immobility/stupor,Staring,Posturing/catalepsy,Echopraxia/echolalia,Verbigeration,Rigidity,Waxy flexibility | 99.1 | 98.2 | 99.8 |
| 8 | Excitement,Immobility/stupor,Staring,Posturing/catalepsy,Grimacing,Echopraxia/echolalia,Mannerisms,Negativism | 99.1 | 98.2 | 99.8 |
| 8 | Excitement,Immobility/stupor,Staring,Posturing/catalepsy,Grimacing,Echopraxia/echolalia,Mannerisms,Rigidity | 99.1 | 98.2 | 99.8 |
| 8 | Excitement,Immobility/stupor,Staring,Posturing/catalepsy,Grimacing,Echopraxia/echolalia,Negativism,Waxy flexibility | 99.1 | 98.2 | 99.8 |
| 8 | Excitement,Immobility/stupor,Staring,Posturing/catalepsy,Grimacing,Echopraxia/echolalia,Verbigeration,Negativism | 99.1 | 98.2 | 99.8 |
| 8 | Excitement,Immobility/stupor,Staring,Posturing/catalepsy,Grimacing,Echopraxia/echolalia,Verbigeration,Rigidity | 99.1 | 98.2 | 99.8 |
| 8 | Excitement,Immobility/stupor,Staring,Posturing/catalepsy,Grimacing,Rigidity,Waxy flexibility,Withdrawal | 99.1 | 98.2 | 99.8 |
| 8 | Excitement,Immobility/stupor,Staring,Posturing/catalepsy,Grimacing,Stereotypy,Negativism,Withdrawal | 99.1 | 98.2 | 99.8 |
| 8 | Excitement,Immobility/stupor,Staring,Posturing/catalepsy,Grimacing,Stereotypy,Rigidity,Negativism | 99.1 | 98.2 | 99.8 |
| 8 | Excitement,Immobility/stupor,Staring,Posturing/catalepsy,Grimacing,Stereotypy,Verbigeration,Negativism | 99.1 | 98.2 | 99.8 |
| 8 | Excitement,Immobility/stupor,Staring,Posturing/catalepsy,Grimacing,Stereotypy,Verbigeration,Rigidity | 99.1 | 98.2 | 99.8 |
| 8 | Excitement,Immobility/stupor,Staring,Posturing/catalepsy,Grimacing,Verbigeration,Negativism,Waxy flexibility | 99.1 | 98.2 | 99.8 |
| 8 | Excitement,Immobility/stupor,Staring,Posturing/catalepsy,Stereotypy,Negativism,Waxy flexibility,Withdrawal | 99.1 | 98.2 | 99.8 |
| 8 | Excitement,Immobility/stupor,Staring,Posturing/catalepsy,Stereotypy,Rigidity,Negativism,Waxy flexibility | 99.1 | 98.2 | 99.8 |
| 8 | Excitement,Immobility/stupor,Staring,Posturing/catalepsy,Stereotypy,Verbigeration,Rigidity,Waxy flexibility | 99.1 | 98.2 | 99.8 |
| 8 | Excitement,Immobility/stupor,Staring,Stereotypy,Mannerisms,Verbigeration,Negativism,Withdrawal | 99.1 | 98.2 | 99.8 |
| 8 | Excitement,Immobility/stupor,Stereotypy,Mannerisms,Verbigeration,Rigidity,Negativism,Withdrawal | 99.1 | 98.2 | 99.8 |
| 8 | Excitement,Mutism,Staring,Echopraxia/echolalia,Mannerisms,Verbigeration,Negativism,Withdrawal | 99.1 | 98.2 | 99.8 |
| 8 | Excitement,Mutism,Staring,Echopraxia/echolalia,Mannerisms,Verbigeration,Rigidity,Waxy flexibility | 99.1 | 98.2 | 99.8 |
| 8 | Excitement,Mutism,Staring,Echopraxia/echolalia,Stereotypy,Mannerisms,Verbigeration,Negativism | 99.1 | 98.2 | 99.8 |
| 8 | Excitement,Mutism,Staring,Echopraxia/echolalia,Stereotypy,Mannerisms,Verbigeration,Rigidity | 99.1 | 98.2 | 99.8 |
| 8 | Excitement,Mutism,Staring,Grimacing,Echopraxia/echolalia,Mannerisms,Negativism,Waxy flexibility | 99.1 | 98.2 | 99.8 |
| 8 | Excitement,Mutism,Staring,Grimacing,Echopraxia/echolalia,Mannerisms,Negativism,Withdrawal | 99.1 | 98.2 | 99.8 |
| 8 | Excitement,Mutism,Staring,Grimacing,Echopraxia/echolalia,Mannerisms,Verbigeration,Rigidity | 99.1 | 98.2 | 99.8 |
| 8 | Excitement,Mutism,Staring,Grimacing,Echopraxia/echolalia,Stereotypy,Mannerisms,Rigidity | 99.1 | 98.2 | 99.8 |
| 8 | Excitement,Mutism,Staring,Grimacing,Echopraxia/echolalia,Stereotypy,Verbigeration,Negativism | 99.1 | 98.2 | 99.8 |
| 8 | Excitement,Mutism,Staring,Grimacing,Echopraxia/echolalia,Stereotypy,Waxy flexibility,Withdrawal | 99.1 | 98.2 | 99.8 |
| 8 | Excitement,Mutism,Staring,Grimacing,Echopraxia/echolalia,Verbigeration,Negativism,Withdrawal | 99.1 | 98.2 | 99.8 |
| 8 | Excitement,Mutism,Staring,Grimacing,Mannerisms,Rigidity,Negativism,Waxy flexibility | 99.1 | 98.2 | 99.8 |
| 8 | Excitement,Mutism,Staring,Grimacing,Stereotypy,Mannerisms,Rigidity,Waxy flexibility | 99.1 | 98.2 | 99.8 |
| 8 | Excitement,Mutism,Staring,Grimacing,Stereotypy,Mannerisms,Verbigeration,Rigidity | 99.1 | 98.2 | 99.8 |
| 8 | Excitement,Mutism,Staring,Grimacing,Stereotypy,Negativism,Waxy flexibility,Withdrawal | 99.1 | 98.2 | 99.8 |
| 8 | Excitement,Mutism,Staring,Grimacing,Stereotypy,Verbigeration,Negativism,Waxy flexibility | 99.1 | 98.2 | 99.8 |
| 8 | Excitement,Mutism,Staring,Mannerisms,Verbigeration,Negativism,Waxy flexibility,Withdrawal | 99.1 | 98.2 | 99.8 |
| 8 | Excitement,Mutism,Staring,Posturing/catalepsy,Echopraxia/echolalia,Mannerisms,Verbigeration,Withdrawal | 99.1 | 98.2 | 99.8 |
| 8 | Excitement,Mutism,Staring,Posturing/catalepsy,Echopraxia/echolalia,Stereotypy,Mannerisms,Withdrawal | 99.1 | 98.2 | 99.8 |
| 8 | Excitement,Mutism,Staring,Posturing/catalepsy,Echopraxia/echolalia,Stereotypy,Verbigeration,Waxy flexibility | 99.1 | 98.2 | 99.8 |
| 8 | Excitement,Mutism,Staring,Posturing/catalepsy,Echopraxia/echolalia,Stereotypy,Verbigeration,Withdrawal | 99.1 | 98.2 | 99.8 |
| 8 | Excitement,Mutism,Staring,Posturing/catalepsy,Grimacing,Echopraxia/echolalia,Stereotypy,Waxy flexibility | 99.1 | 98.2 | 99.8 |
| 8 | Excitement,Mutism,Staring,Posturing/catalepsy,Grimacing,Echopraxia/echolalia,Verbigeration,Waxy flexibility | 99.1 | 98.2 | 99.8 |
| 8 | Excitement,Mutism,Staring,Posturing/catalepsy,Grimacing,Mannerisms,Rigidity,Waxy flexibility | 99.1 | 98.2 | 99.8 |
| 8 | Excitement,Mutism,Staring,Posturing/catalepsy,Grimacing,Mannerisms,Verbigeration,Withdrawal | 99.1 | 98.2 | 99.8 |
| 8 | Excitement,Mutism,Staring,Posturing/catalepsy,Grimacing,Mannerisms,Waxy flexibility,Withdrawal | 99.1 | 98.2 | 99.8 |
| 8 | Excitement,Mutism,Staring,Posturing/catalepsy,Grimacing,Rigidity,Negativism,Waxy flexibility | 99.1 | 98.2 | 99.8 |
| 8 | Excitement,Mutism,Staring,Posturing/catalepsy,Grimacing,Stereotypy,Mannerisms,Negativism | 99.1 | 98.2 | 99.8 |
| 8 | Excitement,Mutism,Staring,Posturing/catalepsy,Grimacing,Stereotypy,Mannerisms,Rigidity | 99.1 | 98.2 | 99.8 |
| 8 | Excitement,Mutism,Staring,Posturing/catalepsy,Grimacing,Stereotypy,Rigidity,Waxy flexibility | 99.1 | 98.2 | 99.8 |
| 8 | Excitement,Mutism,Staring,Posturing/catalepsy,Grimacing,Stereotypy,Verbigeration,Withdrawal | 99.1 | 98.2 | 99.8 |
| 8 | Excitement,Mutism,Staring,Posturing/catalepsy,Grimacing,Stereotypy,Waxy flexibility,Withdrawal | 99.1 | 98.2 | 99.8 |
| 8 | Excitement,Mutism,Staring,Posturing/catalepsy,Grimacing,Verbigeration,Waxy flexibility,Withdrawal | 99.1 | 98.2 | 99.8 |
| 8 | Excitement,Mutism,Staring,Posturing/catalepsy,Mannerisms,Verbigeration,Waxy flexibility,Withdrawal | 99.1 | 98.2 | 99.8 |
| 8 | Excitement,Mutism,Staring,Posturing/catalepsy,Stereotypy,Mannerisms,Rigidity,Waxy flexibility | 99.1 | 98.2 | 99.8 |
| 8 | Excitement,Mutism,Staring,Posturing/catalepsy,Stereotypy,Verbigeration,Waxy flexibility,Withdrawal | 99.1 | 98.2 | 99.8 |
| 8 | Excitement,Mutism,Staring,Stereotypy,Mannerisms,Negativism,Waxy flexibility,Withdrawal | 99.1 | 98.2 | 99.8 |
| 8 | Excitement,Mutism,Staring,Stereotypy,Mannerisms,Verbigeration,Negativism,Waxy flexibility | 99.1 | 98.2 | 99.8 |
| 8 | Excitement,Staring,Echopraxia/echolalia,Mannerisms,Verbigeration,Rigidity,Negativism,Withdrawal | 99.1 | 98.2 | 99.8 |
| 8 | Excitement,Staring,Echopraxia/echolalia,Stereotypy,Mannerisms,Rigidity,Waxy flexibility,Withdrawal | 99.1 | 98.2 | 99.8 |
| 8 | Excitement,Staring,Echopraxia/echolalia,Stereotypy,Rigidity,Negativism,Waxy flexibility,Withdrawal | 99.1 | 98.2 | 99.8 |
| 8 | Excitement,Staring,Echopraxia/echolalia,Stereotypy,Verbigeration,Rigidity,Negativism,Withdrawal | 99.1 | 98.2 | 99.8 |
| 8 | Excitement,Staring,Grimacing,Echopraxia/echolalia,Mannerisms,Rigidity,Negativism,Withdrawal | 99.1 | 98.2 | 99.8 |
| 8 | Excitement,Staring,Grimacing,Echopraxia/echolalia,Mannerisms,Rigidity,Waxy flexibility,Withdrawal | 99.1 | 98.2 | 99.8 |
| 8 | Excitement,Staring,Grimacing,Echopraxia/echolalia,Stereotypy,Mannerisms,Rigidity,Withdrawal | 99.1 | 98.2 | 99.8 |
| 8 | Excitement,Staring,Grimacing,Echopraxia/echolalia,Stereotypy,Rigidity,Negativism,Withdrawal | 99.1 | 98.2 | 99.8 |
| 8 | Excitement,Staring,Grimacing,Mannerisms,Rigidity,Negativism,Waxy flexibility,Withdrawal | 99.1 | 98.2 | 99.8 |
| 8 | Excitement,Staring,Grimacing,Mannerisms,Verbigeration,Rigidity,Negativism,Withdrawal | 99.1 | 98.2 | 99.8 |
| 8 | Excitement,Staring,Grimacing,Mannerisms,Verbigeration,Rigidity,Waxy flexibility,Withdrawal | 99.1 | 98.2 | 99.8 |
| 8 | Excitement,Staring,Grimacing,Stereotypy,Mannerisms,Rigidity,Negativism,Withdrawal | 99.1 | 98.2 | 99.8 |
| 8 | Excitement,Staring,Grimacing,Stereotypy,Mannerisms,Rigidity,Waxy flexibility,Withdrawal | 99.1 | 98.2 | 99.8 |
| 8 | Excitement,Staring,Grimacing,Stereotypy,Mannerisms,Verbigeration,Rigidity,Withdrawal | 99.1 | 98.2 | 99.8 |
| 8 | Excitement,Staring,Grimacing,Stereotypy,Verbigeration,Rigidity,Negativism,Withdrawal | 99.1 | 98.2 | 99.8 |
| 8 | Excitement,Staring,Posturing/catalepsy,Echopraxia/echolalia,Mannerisms,Negativism,Waxy flexibility,Withdrawal | 99.1 | 98.2 | 99.8 |
| 8 | Excitement,Staring,Posturing/catalepsy,Echopraxia/echolalia,Mannerisms,Rigidity,Waxy flexibility,Withdrawal | 99.1 | 98.2 | 99.8 |
| 8 | Excitement,Staring,Posturing/catalepsy,Echopraxia/echolalia,Mannerisms,Verbigeration,Rigidity,Withdrawal | 99.1 | 98.2 | 99.8 |
| 8 | Excitement,Staring,Posturing/catalepsy,Echopraxia/echolalia,Rigidity,Negativism,Waxy flexibility,Withdrawal | 99.1 | 98.2 | 99.8 |
| 8 | Excitement,Staring,Posturing/catalepsy,Echopraxia/echolalia,Stereotypy,Mannerisms,Rigidity,Withdrawal | 99.1 | 98.2 | 99.8 |
| 8 | Excitement,Staring,Posturing/catalepsy,Echopraxia/echolalia,Stereotypy,Rigidity,Negativism,Withdrawal | 99.1 | 98.2 | 99.8 |
| 8 | Excitement,Staring,Posturing/catalepsy,Echopraxia/echolalia,Verbigeration,Rigidity,Negativism,Withdrawal | 99.1 | 98.2 | 99.8 |
| 8 | Excitement,Staring,Posturing/catalepsy,Grimacing,Echopraxia/echolalia,Mannerisms,Negativism,Withdrawal | 99.1 | 98.2 | 99.8 |
| 8 | Excitement,Staring,Posturing/catalepsy,Grimacing,Echopraxia/echolalia,Rigidity,Negativism,Withdrawal | 99.1 | 98.2 | 99.8 |
| 8 | Excitement,Staring,Posturing/catalepsy,Grimacing,Mannerisms,Rigidity,Waxy flexibility,Withdrawal | 99.1 | 98.2 | 99.8 |
| 8 | Excitement,Staring,Posturing/catalepsy,Grimacing,Mannerisms,Verbigeration,Negativism,Withdrawal | 99.1 | 98.2 | 99.8 |
| 8 | Excitement,Staring,Posturing/catalepsy,Grimacing,Stereotypy,Mannerisms,Rigidity,Withdrawal | 99.1 | 98.2 | 99.8 |
| 8 | Excitement,Staring,Posturing/catalepsy,Mannerisms,Verbigeration,Negativism,Waxy flexibility,Withdrawal | 99.1 | 98.2 | 99.8 |
| 8 | Excitement,Staring,Posturing/catalepsy,Mannerisms,Verbigeration,Rigidity,Waxy flexibility,Withdrawal | 99.1 | 98.2 | 99.8 |
| 8 | Excitement,Staring,Posturing/catalepsy,Stereotypy,Mannerisms,Verbigeration,Rigidity,Withdrawal | 99.1 | 98.2 | 99.8 |
| 8 | Excitement,Staring,Posturing/catalepsy,Stereotypy,Verbigeration,Rigidity,Negativism,Withdrawal | 99.1 | 98.2 | 99.8 |
| 8 | Excitement,Staring,Stereotypy,Mannerisms,Verbigeration,Rigidity,Waxy flexibility,Withdrawal | 99.1 | 98.2 | 99.8 |
| 8 | Excitement,Staring,Stereotypy,Verbigeration,Rigidity,Negativism,Waxy flexibility,Withdrawal | 99.1 | 98.2 | 99.8 |
| 8 | Immobility/stupor,Mutism,Staring,Echopraxia/echolalia,Stereotypy,Mannerisms,Negativism,Withdrawal | 99.1 | 98.2 | 99.8 |
| 8 | Immobility/stupor,Mutism,Staring,Echopraxia/echolalia,Stereotypy,Mannerisms,Rigidity,Negativism | 99.1 | 98.2 | 99.8 |
| 8 | Immobility/stupor,Mutism,Staring,Mannerisms,Verbigeration,Rigidity,Negativism,Withdrawal | 99.1 | 98.2 | 99.8 |
| 8 | Immobility/stupor,Mutism,Staring,Posturing/catalepsy,Echopraxia/echolalia,Mannerisms,Rigidity,Negativism | 99.1 | 98.2 | 99.8 |
| 8 | Immobility/stupor,Mutism,Staring,Posturing/catalepsy,Mannerisms,Rigidity,Negativism,Withdrawal | 99.1 | 98.2 | 99.8 |
| 8 | Immobility/stupor,Mutism,Staring,Posturing/catalepsy,Mannerisms,Verbigeration,Negativism,Withdrawal | 99.1 | 98.2 | 99.8 |
| 8 | Immobility/stupor,Mutism,Staring,Posturing/catalepsy,Stereotypy,Mannerisms,Rigidity,Negativism | 99.1 | 98.2 | 99.8 |
| 8 | Immobility/stupor,Mutism,Staring,Stereotypy,Mannerisms,Rigidity,Negativism,Waxy flexibility | 99.1 | 98.2 | 99.8 |
| 8 | Immobility/stupor,Mutism,Staring,Stereotypy,Verbigeration,Rigidity,Negativism,Withdrawal | 99.1 | 98.2 | 99.8 |
| 8 | Immobility/stupor,Staring,Echopraxia/echolalia,Stereotypy,Mannerisms,Rigidity,Negativism,Waxy flexibility | 99.1 | 98.2 | 99.8 |
| 8 | Immobility/stupor,Staring,Echopraxia/echolalia,Stereotypy,Mannerisms,Verbigeration,Rigidity,Negativism | 99.1 | 98.2 | 99.8 |
| 8 | Immobility/stupor,Staring,Grimacing,Echopraxia/echolalia,Stereotypy,Mannerisms,Rigidity,Negativism | 99.1 | 98.2 | 99.8 |
| 8 | Immobility/stupor,Staring,Grimacing,Mannerisms,Rigidity,Negativism,Waxy flexibility,Withdrawal | 99.1 | 98.2 | 99.8 |
| 8 | Immobility/stupor,Staring,Grimacing,Stereotypy,Mannerisms,Rigidity,Negativism,Waxy flexibility | 99.1 | 98.2 | 99.8 |
| 8 | Immobility/stupor,Staring,Grimacing,Stereotypy,Mannerisms,Verbigeration,Negativism,Withdrawal | 99.1 | 98.2 | 99.8 |
| 8 | Immobility/stupor,Staring,Grimacing,Stereotypy,Verbigeration,Rigidity,Negativism,Withdrawal | 99.1 | 98.2 | 99.8 |
| 8 | Immobility/stupor,Staring,Mannerisms,Verbigeration,Rigidity,Negativism,Waxy flexibility,Withdrawal | 99.1 | 98.2 | 99.8 |
| 8 | Immobility/stupor,Staring,Posturing/catalepsy,Echopraxia/echolalia,Mannerisms,Verbigeration,Rigidity,Negativism | 99.1 | 98.2 | 99.8 |
| 8 | Immobility/stupor,Staring,Posturing/catalepsy,Echopraxia/echolalia,Stereotypy,Mannerisms,Rigidity,Negativism | 99.1 | 98.2 | 99.8 |
| 8 | Immobility/stupor,Staring,Posturing/catalepsy,Grimacing,Echopraxia/echolalia,Mannerisms,Rigidity,Negativism | 99.1 | 98.2 | 99.8 |
| 8 | Immobility/stupor,Staring,Posturing/catalepsy,Grimacing,Mannerisms,Verbigeration,Negativism,Withdrawal | 99.1 | 98.2 | 99.8 |
| 8 | Immobility/stupor,Staring,Posturing/catalepsy,Grimacing,Stereotypy,Mannerisms,Rigidity,Negativism | 99.1 | 98.2 | 99.8 |
| 8 | Immobility/stupor,Staring,Posturing/catalepsy,Mannerisms,Rigidity,Negativism,Waxy flexibility,Withdrawal | 99.1 | 98.2 | 99.8 |
| 8 | Mutism,Staring,Echopraxia/echolalia,Mannerisms,Verbigeration,Rigidity,Negativism,Waxy flexibility | 99.1 | 98.2 | 99.8 |
| 8 | Mutism,Staring,Echopraxia/echolalia,Stereotypy,Mannerisms,Negativism,Waxy flexibility,Withdrawal | 99.1 | 98.2 | 99.8 |
| 8 | Mutism,Staring,Echopraxia/echolalia,Stereotypy,Mannerisms,Verbigeration,Negativism,Withdrawal | 99.1 | 98.2 | 99.8 |
| 8 | Mutism,Staring,Echopraxia/echolalia,Stereotypy,Verbigeration,Rigidity,Negativism,Withdrawal | 99.1 | 98.2 | 99.8 |
| 8 | Mutism,Staring,Grimacing,Echopraxia/echolalia,Mannerisms,Negativism,Waxy flexibility,Withdrawal | 99.1 | 98.2 | 99.8 |
| 8 | Mutism,Staring,Grimacing,Echopraxia/echolalia,Mannerisms,Rigidity,Negativism,Waxy flexibility | 99.1 | 98.2 | 99.8 |
| 8 | Mutism,Staring,Grimacing,Echopraxia/echolalia,Mannerisms,Verbigeration,Rigidity,Negativism | 99.1 | 98.2 | 99.8 |
| 8 | Mutism,Staring,Grimacing,Echopraxia/echolalia,Stereotypy,Mannerisms,Negativism,Waxy flexibility | 99.1 | 98.2 | 99.8 |
| 8 | Mutism,Staring,Grimacing,Mannerisms,Rigidity,Negativism,Waxy flexibility,Withdrawal | 99.1 | 98.2 | 99.8 |
| 8 | Mutism,Staring,Grimacing,Stereotypy,Mannerisms,Negativism,Waxy flexibility,Withdrawal | 99.1 | 98.2 | 99.8 |
| 8 | Mutism,Staring,Grimacing,Stereotypy,Mannerisms,Rigidity,Negativism,Waxy flexibility | 99.1 | 98.2 | 99.8 |
| 8 | Mutism,Staring,Mannerisms,Verbigeration,Rigidity,Negativism,Waxy flexibility,Withdrawal | 99.1 | 98.2 | 99.8 |
| 8 | Mutism,Staring,Posturing/catalepsy,Echopraxia/echolalia,Mannerisms,Rigidity,Negativism,Waxy flexibility | 99.1 | 98.2 | 99.8 |
| 8 | Mutism,Staring,Posturing/catalepsy,Echopraxia/echolalia,Stereotypy,Mannerisms,Negativism,Waxy flexibility | 99.1 | 98.2 | 99.8 |
| 8 | Mutism,Staring,Posturing/catalepsy,Echopraxia/echolalia,Stereotypy,Mannerisms,Rigidity,Withdrawal | 99.1 | 98.2 | 99.8 |
| 8 | Mutism,Staring,Posturing/catalepsy,Echopraxia/echolalia,Stereotypy,Mannerisms,Verbigeration,Negativism | 99.1 | 98.2 | 99.8 |
| 8 | Mutism,Staring,Posturing/catalepsy,Grimacing,Echopraxia/echolalia,Mannerisms,Negativism,Waxy flexibility | 99.1 | 98.2 | 99.8 |
| 8 | Mutism,Staring,Posturing/catalepsy,Grimacing,Echopraxia/echolalia,Mannerisms,Rigidity,Withdrawal | 99.1 | 98.2 | 99.8 |
| 8 | Mutism,Staring,Posturing/catalepsy,Grimacing,Mannerisms,Verbigeration,Rigidity,Withdrawal | 99.1 | 98.2 | 99.8 |
| 8 | Mutism,Staring,Posturing/catalepsy,Grimacing,Stereotypy,Mannerisms,Rigidity,Negativism | 99.1 | 98.2 | 99.8 |
| 8 | Mutism,Staring,Posturing/catalepsy,Grimacing,Stereotypy,Mannerisms,Verbigeration,Negativism | 99.1 | 98.2 | 99.8 |
| 8 | Mutism,Staring,Posturing/catalepsy,Grimacing,Stereotypy,Verbigeration,Negativism,Withdrawal | 99.1 | 98.2 | 99.8 |
| 8 | Mutism,Staring,Posturing/catalepsy,Mannerisms,Rigidity,Negativism,Waxy flexibility,Withdrawal | 99.1 | 98.2 | 99.8 |
| 8 | Mutism,Staring,Posturing/catalepsy,Mannerisms,Verbigeration,Negativism,Waxy flexibility,Withdrawal | 99.1 | 98.2 | 99.8 |
| 8 | Mutism,Staring,Posturing/catalepsy,Stereotypy,Mannerisms,Verbigeration,Rigidity,Withdrawal | 99.1 | 98.2 | 99.8 |
| 8 | Mutism,Staring,Posturing/catalepsy,Stereotypy,Verbigeration,Rigidity,Negativism,Withdrawal | 99.1 | 98.2 | 99.8 |
| 8 | Mutism,Staring,Stereotypy,Verbigeration,Rigidity,Negativism,Waxy flexibility,Withdrawal | 99.1 | 98.2 | 99.8 |
| 8 | Staring,Grimacing,Echopraxia/echolalia,Mannerisms,Rigidity,Negativism,Waxy flexibility,Withdrawal | 99.1 | 98.2 | 99.8 |
| 8 | Staring,Posturing/catalepsy,Echopraxia/echolalia,Mannerisms,Rigidity,Negativism,Waxy flexibility,Withdrawal | 99.1 | 98.2 | 99.8 |
| 8 | Staring,Posturing/catalepsy,Echopraxia/echolalia,Mannerisms,Verbigeration,Rigidity,Negativism,Withdrawal | 99.1 | 98.2 | 99.8 |
| 8 | Staring,Posturing/catalepsy,Grimacing,Echopraxia/echolalia,Stereotypy,Mannerisms,Negativism,Withdrawal | 99.1 | 98.2 | 99.8 |
| 8 | Staring,Posturing/catalepsy,Grimacing,Mannerisms,Verbigeration,Rigidity,Negativism,Withdrawal | 99.1 | 98.2 | 99.8 |
| 8 | Staring,Posturing/catalepsy,Grimacing,Stereotypy,Mannerisms,Verbigeration,Negativism,Withdrawal | 99.1 | 98.2 | 99.8 |
| 8 | Staring,Posturing/catalepsy,Stereotypy,Mannerisms,Rigidity,Negativism,Waxy flexibility,Withdrawal | 99.1 | 98.2 | 99.8 |
| 8 | Excitement,Immobility/stupor,Echopraxia/echolalia,Stereotypy,Rigidity,Negativism,Waxy flexibility,Withdrawal | 98.9 | 97.8 | 99.8 |
| 8 | Excitement,Immobility/stupor,Echopraxia/echolalia,Verbigeration,Rigidity,Negativism,Waxy flexibility,Withdrawal | 98.9 | 97.8 | 99.8 |
| 8 | Excitement,Immobility/stupor,Grimacing,Echopraxia/echolalia,Rigidity,Negativism,Waxy flexibility,Withdrawal | 98.9 | 97.8 | 99.8 |
| 8 | Excitement,Immobility/stupor,Grimacing,Mannerisms,Verbigeration,Rigidity,Negativism,Withdrawal | 98.9 | 97.8 | 99.8 |
| 8 | Excitement,Immobility/stupor,Grimacing,Stereotypy,Verbigeration,Rigidity,Negativism,Withdrawal | 98.9 | 97.8 | 99.8 |
| 8 | Excitement,Immobility/stupor,Mutism,Echopraxia/echolalia,Mannerisms,Rigidity,Negativism,Withdrawal | 98.9 | 97.8 | 99.8 |
| 8 | Excitement,Immobility/stupor,Mutism,Echopraxia/echolalia,Mannerisms,Verbigeration,Rigidity,Negativism | 98.9 | 97.8 | 99.8 |
| 8 | Excitement,Immobility/stupor,Mutism,Echopraxia/echolalia,Rigidity,Negativism,Waxy flexibility,Withdrawal | 98.9 | 97.8 | 99.8 |
| 8 | Excitement,Immobility/stupor,Mutism,Echopraxia/echolalia,Stereotypy,Rigidity,Negativism,Waxy flexibility | 98.9 | 97.8 | 99.8 |
| 8 | Excitement,Immobility/stupor,Mutism,Echopraxia/echolalia,Stereotypy,Rigidity,Waxy flexibility,Withdrawal | 98.9 | 97.8 | 99.8 |
| 8 | Excitement,Immobility/stupor,Mutism,Echopraxia/echolalia,Verbigeration,Rigidity,Waxy flexibility,Withdrawal | 98.9 | 97.8 | 99.8 |
| 8 | Excitement,Immobility/stupor,Mutism,Grimacing,Echopraxia/echolalia,Rigidity,Negativism,Waxy flexibility | 98.9 | 97.8 | 99.8 |
| 8 | Excitement,Immobility/stupor,Mutism,Grimacing,Echopraxia/echolalia,Rigidity,Waxy flexibility,Withdrawal | 98.9 | 97.8 | 99.8 |
| 8 | Excitement,Immobility/stupor,Mutism,Grimacing,Echopraxia/echolalia,Stereotypy,Rigidity,Negativism | 98.9 | 97.8 | 99.8 |
| 8 | Excitement,Immobility/stupor,Mutism,Grimacing,Echopraxia/echolalia,Stereotypy,Rigidity,Withdrawal | 98.9 | 97.8 | 99.8 |
| 8 | Excitement,Immobility/stupor,Mutism,Grimacing,Echopraxia/echolalia,Verbigeration,Rigidity,Withdrawal | 98.9 | 97.8 | 99.8 |
| 8 | Excitement,Immobility/stupor,Mutism,Grimacing,Mannerisms,Verbigeration,Rigidity,Negativism | 98.9 | 97.8 | 99.8 |
| 8 | Excitement,Immobility/stupor,Mutism,Grimacing,Stereotypy,Rigidity,Negativism,Withdrawal | 98.9 | 97.8 | 99.8 |
| 8 | Excitement,Immobility/stupor,Mutism,Grimacing,Stereotypy,Verbigeration,Rigidity,Withdrawal | 98.9 | 97.8 | 99.8 |
| 8 | Excitement,Immobility/stupor,Mutism,Grimacing,Verbigeration,Rigidity,Negativism,Waxy flexibility | 98.9 | 97.8 | 99.8 |
| 8 | Excitement,Immobility/stupor,Mutism,Mannerisms,Verbigeration,Rigidity,Negativism,Waxy flexibility | 98.9 | 97.8 | 99.8 |
| 8 | Excitement,Immobility/stupor,Mutism,Posturing/catalepsy,Echopraxia/echolalia,Mannerisms,Rigidity,Negativism | 98.9 | 97.8 | 99.8 |
| 8 | Excitement,Immobility/stupor,Mutism,Posturing/catalepsy,Echopraxia/echolalia,Mannerisms,Rigidity,Withdrawal | 98.9 | 97.8 | 99.8 |
| 8 | Excitement,Immobility/stupor,Mutism,Posturing/catalepsy,Echopraxia/echolalia,Mannerisms,Verbigeration,Rigidity | 98.9 | 97.8 | 99.8 |
| 8 | Excitement,Immobility/stupor,Mutism,Posturing/catalepsy,Echopraxia/echolalia,Rigidity,Negativism,Waxy flexibility | 98.9 | 97.8 | 99.8 |
| 8 | Excitement,Immobility/stupor,Mutism,Posturing/catalepsy,Echopraxia/echolalia,Rigidity,Waxy flexibility,Withdrawal | 98.9 | 97.8 | 99.8 |
| 8 | Excitement,Immobility/stupor,Mutism,Posturing/catalepsy,Echopraxia/echolalia,Verbigeration,Negativism,Withdrawal | 98.9 | 97.8 | 99.8 |
| 8 | Excitement,Immobility/stupor,Mutism,Posturing/catalepsy,Echopraxia/echolalia,Verbigeration,Rigidity,Waxy flexibility | 98.9 | 97.8 | 99.8 |
| 8 | Excitement,Immobility/stupor,Mutism,Posturing/catalepsy,Grimacing,Echopraxia/echolalia,Negativism,Withdrawal | 98.9 | 97.8 | 99.8 |
| 8 | Excitement,Immobility/stupor,Mutism,Posturing/catalepsy,Grimacing,Mannerisms,Rigidity,Negativism | 98.9 | 97.8 | 99.8 |
| 8 | Excitement,Immobility/stupor,Mutism,Posturing/catalepsy,Grimacing,Mannerisms,Rigidity,Withdrawal | 98.9 | 97.8 | 99.8 |
| 8 | Excitement,Immobility/stupor,Mutism,Posturing/catalepsy,Grimacing,Mannerisms,Verbigeration,Rigidity | 98.9 | 97.8 | 99.8 |
| 8 | Excitement,Immobility/stupor,Mutism,Posturing/catalepsy,Grimacing,Stereotypy,Rigidity,Negativism | 98.9 | 97.8 | 99.8 |
| 8 | Excitement,Immobility/stupor,Mutism,Posturing/catalepsy,Grimacing,Stereotypy,Rigidity,Withdrawal | 98.9 | 97.8 | 99.8 |
| 8 | Excitement,Immobility/stupor,Mutism,Posturing/catalepsy,Grimacing,Stereotypy,Verbigeration,Rigidity | 98.9 | 97.8 | 99.8 |
| 8 | Excitement,Immobility/stupor,Mutism,Posturing/catalepsy,Mannerisms,Rigidity,Negativism,Withdrawal | 98.9 | 97.8 | 99.8 |
| 8 | Excitement,Immobility/stupor,Mutism,Posturing/catalepsy,Stereotypy,Verbigeration,Negativism,Withdrawal | 98.9 | 97.8 | 99.8 |
| 8 | Excitement,Immobility/stupor,Mutism,Posturing/catalepsy,Verbigeration,Rigidity,Waxy flexibility,Withdrawal | 98.9 | 97.8 | 99.8 |
| 8 | Excitement,Immobility/stupor,Mutism,Staring,Echopraxia/echolalia,Stereotypy,Mannerisms,Negativism | 98.9 | 97.8 | 99.8 |
| 8 | Excitement,Immobility/stupor,Mutism,Staring,Echopraxia/echolalia,Stereotypy,Waxy flexibility,Withdrawal | 98.9 | 97.8 | 99.8 |
| 8 | Excitement,Immobility/stupor,Mutism,Staring,Grimacing,Echopraxia/echolalia,Mannerisms,Rigidity | 98.9 | 97.8 | 99.8 |
| 8 | Excitement,Immobility/stupor,Mutism,Staring,Grimacing,Echopraxia/echolalia,Stereotypy,Negativism | 98.9 | 97.8 | 99.8 |
| 8 | Excitement,Immobility/stupor,Mutism,Staring,Grimacing,Echopraxia/echolalia,Waxy flexibility,Withdrawal | 98.9 | 97.8 | 99.8 |
| 8 | Excitement,Immobility/stupor,Mutism,Staring,Grimacing,Mannerisms,Rigidity,Negativism | 98.9 | 97.8 | 99.8 |
| 8 | Excitement,Immobility/stupor,Mutism,Staring,Grimacing,Stereotypy,Negativism,Waxy flexibility | 98.9 | 97.8 | 99.8 |
| 8 | Excitement,Immobility/stupor,Mutism,Staring,Grimacing,Stereotypy,Negativism,Withdrawal | 98.9 | 97.8 | 99.8 |
| 8 | Excitement,Immobility/stupor,Mutism,Staring,Grimacing,Stereotypy,Verbigeration,Negativism | 98.9 | 97.8 | 99.8 |
| 8 | Excitement,Immobility/stupor,Mutism,Staring,Grimacing,Verbigeration,Negativism,Waxy flexibility | 98.9 | 97.8 | 99.8 |
| 8 | Excitement,Immobility/stupor,Mutism,Staring,Mannerisms,Negativism,Waxy flexibility,Withdrawal | 98.9 | 97.8 | 99.8 |
| 8 | Excitement,Immobility/stupor,Mutism,Staring,Mannerisms,Verbigeration,Negativism,Withdrawal | 98.9 | 97.8 | 99.8 |
| 8 | Excitement,Immobility/stupor,Mutism,Staring,Posturing/catalepsy,Echopraxia/echolalia,Mannerisms,Waxy flexibility | 98.9 | 97.8 | 99.8 |
| 8 | Excitement,Immobility/stupor,Mutism,Staring,Posturing/catalepsy,Echopraxia/echolalia,Stereotypy,Waxy flexibility | 98.9 | 97.8 | 99.8 |
| 8 | Excitement,Immobility/stupor,Mutism,Staring,Posturing/catalepsy,Grimacing,Echopraxia/echolalia,Verbigeration | 98.9 | 97.8 | 99.8 |
| 8 | Excitement,Immobility/stupor,Mutism,Staring,Posturing/catalepsy,Grimacing,Mannerisms,Withdrawal | 98.9 | 97.8 | 99.8 |
| 8 | Excitement,Immobility/stupor,Mutism,Staring,Posturing/catalepsy,Grimacing,Rigidity,Waxy flexibility | 98.9 | 97.8 | 99.8 |
| 8 | Excitement,Immobility/stupor,Mutism,Staring,Posturing/catalepsy,Grimacing,Stereotypy,Withdrawal | 98.9 | 97.8 | 99.8 |
| 8 | Excitement,Immobility/stupor,Mutism,Staring,Posturing/catalepsy,Grimacing,Verbigeration,Waxy flexibility | 98.9 | 97.8 | 99.8 |
| 8 | Excitement,Immobility/stupor,Mutism,Staring,Posturing/catalepsy,Grimacing,Verbigeration,Withdrawal | 98.9 | 97.8 | 99.8 |
| 8 | Excitement,Immobility/stupor,Mutism,Staring,Posturing/catalepsy,Grimacing,Waxy flexibility,Withdrawal | 98.9 | 97.8 | 99.8 |
| 8 | Excitement,Immobility/stupor,Mutism,Staring,Posturing/catalepsy,Mannerisms,Verbigeration,Waxy flexibility | 98.9 | 97.8 | 99.8 |
| 8 | Excitement,Immobility/stupor,Mutism,Staring,Posturing/catalepsy,Mannerisms,Verbigeration,Withdrawal | 98.9 | 97.8 | 99.8 |
| 8 | Excitement,Immobility/stupor,Mutism,Staring,Posturing/catalepsy,Mannerisms,Waxy flexibility,Withdrawal | 98.9 | 97.8 | 99.8 |
| 8 | Excitement,Immobility/stupor,Mutism,Staring,Posturing/catalepsy,Stereotypy,Verbigeration,Waxy flexibility | 98.9 | 97.8 | 99.8 |
| 8 | Excitement,Immobility/stupor,Mutism,Staring,Posturing/catalepsy,Stereotypy,Waxy flexibility,Withdrawal | 98.9 | 97.8 | 99.8 |
| 8 | Excitement,Immobility/stupor,Mutism,Staring,Posturing/catalepsy,Verbigeration,Waxy flexibility,Withdrawal | 98.9 | 97.8 | 99.8 |
| 8 | Excitement,Immobility/stupor,Mutism,Staring,Stereotypy,Mannerisms,Negativism,Waxy flexibility | 98.9 | 97.8 | 99.8 |
| 8 | Excitement,Immobility/stupor,Mutism,Staring,Stereotypy,Mannerisms,Negativism,Withdrawal | 98.9 | 97.8 | 99.8 |
| 8 | Excitement,Immobility/stupor,Mutism,Staring,Stereotypy,Mannerisms,Verbigeration,Negativism | 98.9 | 97.8 | 99.8 |
| 8 | Excitement,Immobility/stupor,Mutism,Staring,Verbigeration,Negativism,Waxy flexibility,Withdrawal | 98.9 | 97.8 | 99.8 |
| 8 | Excitement,Immobility/stupor,Mutism,Stereotypy,Mannerisms,Verbigeration,Rigidity,Withdrawal | 98.9 | 97.8 | 99.8 |
| 8 | Excitement,Immobility/stupor,Mutism,Stereotypy,Rigidity,Negativism,Waxy flexibility,Withdrawal | 98.9 | 97.8 | 99.8 |
| 8 | Excitement,Immobility/stupor,Mutism,Verbigeration,Rigidity,Negativism,Waxy flexibility,Withdrawal | 98.9 | 97.8 | 99.8 |
| 8 | Excitement,Immobility/stupor,Posturing/catalepsy,Echopraxia/echolalia,Mannerisms,Verbigeration,Rigidity,Withdrawal | 98.9 | 97.8 | 99.8 |
| 8 | Excitement,Immobility/stupor,Posturing/catalepsy,Echopraxia/echolalia,Rigidity,Negativism,Waxy flexibility,Withdrawal | 98.9 | 97.8 | 99.8 |
| 8 | Excitement,Immobility/stupor,Posturing/catalepsy,Echopraxia/echolalia,Stereotypy,Verbigeration,Rigidity,Withdrawal | 98.9 | 97.8 | 99.8 |
| 8 | Excitement,Immobility/stupor,Posturing/catalepsy,Grimacing,Echopraxia/echolalia,Mannerisms,Rigidity,Withdrawal | 98.9 | 97.8 | 99.8 |
| 8 | Excitement,Immobility/stupor,Posturing/catalepsy,Grimacing,Echopraxia/echolalia,Verbigeration,Rigidity,Negativism | 98.9 | 97.8 | 99.8 |
| 8 | Excitement,Immobility/stupor,Posturing/catalepsy,Grimacing,Echopraxia/echolalia,Verbigeration,Rigidity,Withdrawal | 98.9 | 97.8 | 99.8 |
| 8 | Excitement,Immobility/stupor,Posturing/catalepsy,Grimacing,Mannerisms,Verbigeration,Rigidity,Negativism | 98.9 | 97.8 | 99.8 |
| 8 | Excitement,Immobility/stupor,Posturing/catalepsy,Grimacing,Stereotypy,Rigidity,Negativism,Withdrawal | 98.9 | 97.8 | 99.8 |
| 8 | Excitement,Immobility/stupor,Posturing/catalepsy,Stereotypy,Mannerisms,Rigidity,Negativism,Withdrawal | 98.9 | 97.8 | 99.8 |
| 8 | Excitement,Immobility/stupor,Posturing/catalepsy,Verbigeration,Rigidity,Negativism,Waxy flexibility,Withdrawal | 98.9 | 97.8 | 99.8 |
| 8 | Excitement,Immobility/stupor,Staring,Echopraxia/echolalia,Mannerisms,Verbigeration,Negativism,Waxy flexibility | 98.9 | 97.8 | 99.8 |
| 8 | Excitement,Immobility/stupor,Staring,Echopraxia/echolalia,Mannerisms,Verbigeration,Negativism,Withdrawal | 98.9 | 97.8 | 99.8 |
| 8 | Excitement,Immobility/stupor,Staring,Echopraxia/echolalia,Stereotypy,Mannerisms,Negativism,Waxy flexibility | 98.9 | 97.8 | 99.8 |
| 8 | Excitement,Immobility/stupor,Staring,Echopraxia/echolalia,Stereotypy,Mannerisms,Rigidity,Waxy flexibility | 98.9 | 97.8 | 99.8 |
| 8 | Excitement,Immobility/stupor,Staring,Echopraxia/echolalia,Stereotypy,Mannerisms,Verbigeration,Rigidity | 98.9 | 97.8 | 99.8 |
| 8 | Excitement,Immobility/stupor,Staring,Echopraxia/echolalia,Stereotypy,Verbigeration,Negativism,Waxy flexibility | 98.9 | 97.8 | 99.8 |
| 8 | Excitement,Immobility/stupor,Staring,Echopraxia/echolalia,Verbigeration,Negativism,Waxy flexibility,Withdrawal | 98.9 | 97.8 | 99.8 |
| 8 | Excitement,Immobility/stupor,Staring,Grimacing,Echopraxia/echolalia,Mannerisms,Negativism,Withdrawal | 98.9 | 97.8 | 99.8 |
| 8 | Excitement,Immobility/stupor,Staring,Grimacing,Echopraxia/echolalia,Mannerisms,Rigidity,Waxy flexibility | 98.9 | 97.8 | 99.8 |
| 8 | Excitement,Immobility/stupor,Staring,Grimacing,Echopraxia/echolalia,Negativism,Waxy flexibility,Withdrawal | 98.9 | 97.8 | 99.8 |
| 8 | Excitement,Immobility/stupor,Staring,Grimacing,Echopraxia/echolalia,Stereotypy,Rigidity,Waxy flexibility | 98.9 | 97.8 | 99.8 |
| 8 | Excitement,Immobility/stupor,Staring,Grimacing,Echopraxia/echolalia,Verbigeration,Rigidity,Waxy flexibility | 98.9 | 97.8 | 99.8 |
| 8 | Excitement,Immobility/stupor,Staring,Grimacing,Stereotypy,Mannerisms,Rigidity,Negativism | 98.9 | 97.8 | 99.8 |
| 8 | Excitement,Immobility/stupor,Staring,Grimacing,Stereotypy,Mannerisms,Rigidity,Waxy flexibility | 98.9 | 97.8 | 99.8 |
| 8 | Excitement,Immobility/stupor,Staring,Grimacing,Stereotypy,Mannerisms,Verbigeration,Rigidity | 98.9 | 97.8 | 99.8 |
| 8 | Excitement,Immobility/stupor,Staring,Grimacing,Stereotypy,Verbigeration,Negativism,Withdrawal | 98.9 | 97.8 | 99.8 |
| 8 | Excitement,Immobility/stupor,Staring,Mannerisms,Verbigeration,Negativism,Waxy flexibility,Withdrawal | 98.9 | 97.8 | 99.8 |
| 8 | Excitement,Immobility/stupor,Staring,Posturing/catalepsy,Echopraxia/echolalia,Mannerisms,Rigidity,Waxy flexibility | 98.9 | 97.8 | 99.8 |
| 8 | Excitement,Immobility/stupor,Staring,Posturing/catalepsy,Echopraxia/echolalia,Mannerisms,Waxy flexibility,Withdrawal | 98.9 | 97.8 | 99.8 |
| 8 | Excitement,Immobility/stupor,Staring,Posturing/catalepsy,Echopraxia/echolalia,Stereotypy,Mannerisms,Rigidity | 98.9 | 97.8 | 99.8 |
| 8 | Excitement,Immobility/stupor,Staring,Posturing/catalepsy,Grimacing,Echopraxia/echolalia,Mannerisms,Withdrawal | 98.9 | 97.8 | 99.8 |
| 8 | Excitement,Immobility/stupor,Staring,Posturing/catalepsy,Grimacing,Echopraxia/echolalia,Rigidity,Waxy flexibility | 98.9 | 97.8 | 99.8 |
| 8 | Excitement,Immobility/stupor,Staring,Posturing/catalepsy,Grimacing,Echopraxia/echolalia,Stereotypy,Negativism | 98.9 | 97.8 | 99.8 |
| 8 | Excitement,Immobility/stupor,Staring,Posturing/catalepsy,Grimacing,Echopraxia/echolalia,Stereotypy,Rigidity | 98.9 | 97.8 | 99.8 |
| 8 | Excitement,Immobility/stupor,Staring,Posturing/catalepsy,Grimacing,Mannerisms,Negativism,Waxy flexibility | 98.9 | 97.8 | 99.8 |
| 8 | Excitement,Immobility/stupor,Staring,Posturing/catalepsy,Grimacing,Mannerisms,Rigidity,Waxy flexibility | 98.9 | 97.8 | 99.8 |
| 8 | Excitement,Immobility/stupor,Staring,Posturing/catalepsy,Grimacing,Negativism,Waxy flexibility,Withdrawal | 98.9 | 97.8 | 99.8 |
| 8 | Excitement,Immobility/stupor,Staring,Posturing/catalepsy,Grimacing,Rigidity,Negativism,Waxy flexibility | 98.9 | 97.8 | 99.8 |
| 8 | Excitement,Immobility/stupor,Staring,Posturing/catalepsy,Grimacing,Stereotypy,Mannerisms,Rigidity | 98.9 | 97.8 | 99.8 |
| 8 | Excitement,Immobility/stupor,Staring,Posturing/catalepsy,Grimacing,Verbigeration,Rigidity,Waxy flexibility | 98.9 | 97.8 | 99.8 |
| 8 | Excitement,Immobility/stupor,Staring,Posturing/catalepsy,Stereotypy,Mannerisms,Negativism,Waxy flexibility | 98.9 | 97.8 | 99.8 |
| 8 | Excitement,Immobility/stupor,Staring,Stereotypy,Mannerisms,Verbigeration,Negativism,Waxy flexibility | 98.9 | 97.8 | 99.8 |
| 8 | Excitement,Immobility/stupor,Staring,Stereotypy,Verbigeration,Negativism,Waxy flexibility,Withdrawal | 98.9 | 97.8 | 99.8 |
| 8 | Excitement,Immobility/stupor,Stereotypy,Verbigeration,Rigidity,Negativism,Waxy flexibility,Withdrawal | 98.9 | 97.8 | 99.8 |
| 8 | Excitement,Mutism,Staring,Echopraxia/echolalia,Stereotypy,Mannerisms,Waxy flexibility,Withdrawal | 98.9 | 97.8 | 99.8 |
| 8 | Excitement,Mutism,Staring,Echopraxia/echolalia,Stereotypy,Verbigeration,Waxy flexibility,Withdrawal | 98.9 | 97.8 | 99.8 |
| 8 | Excitement,Mutism,Staring,Grimacing,Echopraxia/echolalia,Mannerisms,Waxy flexibility,Withdrawal | 98.9 | 97.8 | 99.8 |
| 8 | Excitement,Mutism,Staring,Grimacing,Echopraxia/echolalia,Stereotypy,Mannerisms,Negativism | 98.9 | 97.8 | 99.8 |
| 8 | Excitement,Mutism,Staring,Grimacing,Echopraxia/echolalia,Stereotypy,Verbigeration,Waxy flexibility | 98.9 | 97.8 | 99.8 |
| 8 | Excitement,Mutism,Staring,Grimacing,Echopraxia/echolalia,Verbigeration,Waxy flexibility,Withdrawal | 98.9 | 97.8 | 99.8 |
| 8 | Excitement,Mutism,Staring,Grimacing,Mannerisms,Negativism,Waxy flexibility,Withdrawal | 98.9 | 97.8 | 99.8 |
| 8 | Excitement,Mutism,Staring,Grimacing,Mannerisms,Verbigeration,Negativism,Waxy flexibility | 98.9 | 97.8 | 99.8 |
| 8 | Excitement,Mutism,Staring,Grimacing,Mannerisms,Verbigeration,Negativism,Withdrawal | 98.9 | 97.8 | 99.8 |
| 8 | Excitement,Mutism,Staring,Grimacing,Stereotypy,Mannerisms,Negativism,Withdrawal | 98.9 | 97.8 | 99.8 |
| 8 | Excitement,Mutism,Staring,Grimacing,Stereotypy,Mannerisms,Rigidity,Negativism | 98.9 | 97.8 | 99.8 |
| 8 | Excitement,Mutism,Staring,Grimacing,Verbigeration,Negativism,Waxy flexibility,Withdrawal | 98.9 | 97.8 | 99.8 |
| 8 | Excitement,Mutism,Staring,Posturing/catalepsy,Echopraxia/echolalia,Mannerisms,Verbigeration,Waxy flexibility | 98.9 | 97.8 | 99.8 |
| 8 | Excitement,Mutism,Staring,Posturing/catalepsy,Echopraxia/echolalia,Stereotypy,Mannerisms,Waxy flexibility | 98.9 | 97.8 | 99.8 |
| 8 | Excitement,Mutism,Staring,Posturing/catalepsy,Grimacing,Echopraxia/echolalia,Mannerisms,Waxy flexibility | 98.9 | 97.8 | 99.8 |
| 8 | Excitement,Mutism,Staring,Posturing/catalepsy,Grimacing,Echopraxia/echolalia,Stereotypy,Verbigeration | 98.9 | 97.8 | 99.8 |
| 8 | Excitement,Mutism,Staring,Posturing/catalepsy,Grimacing,Stereotypy,Mannerisms,Withdrawal | 98.9 | 97.8 | 99.8 |
| 8 | Excitement,Mutism,Staring,Posturing/catalepsy,Grimacing,Stereotypy,Verbigeration,Waxy flexibility | 98.9 | 97.8 | 99.8 |
| 8 | Excitement,Mutism,Staring,Posturing/catalepsy,Stereotypy,Mannerisms,Verbigeration,Withdrawal | 98.9 | 97.8 | 99.8 |
| 8 | Excitement,Mutism,Staring,Posturing/catalepsy,Stereotypy,Mannerisms,Waxy flexibility,Withdrawal | 98.9 | 97.8 | 99.8 |
| 8 | Excitement,Staring,Echopraxia/echolalia,Stereotypy,Mannerisms,Verbigeration,Rigidity,Withdrawal | 98.9 | 97.8 | 99.8 |
| 8 | Excitement,Staring,Echopraxia/echolalia,Verbigeration,Rigidity,Negativism,Waxy flexibility,Withdrawal | 98.9 | 97.8 | 99.8 |
| 8 | Excitement,Staring,Grimacing,Echopraxia/echolalia,Rigidity,Negativism,Waxy flexibility,Withdrawal | 98.9 | 97.8 | 99.8 |
| 8 | Excitement,Staring,Grimacing,Echopraxia/echolalia,Stereotypy,Rigidity,Waxy flexibility,Withdrawal | 98.9 | 97.8 | 99.8 |
| 8 | Excitement,Staring,Grimacing,Stereotypy,Rigidity,Negativism,Waxy flexibility,Withdrawal | 98.9 | 97.8 | 99.8 |
| 8 | Excitement,Staring,Grimacing,Stereotypy,Verbigeration,Rigidity,Waxy flexibility,Withdrawal | 98.9 | 97.8 | 99.8 |
| 8 | Excitement,Staring,Posturing/catalepsy,Echopraxia/echolalia,Mannerisms,Verbigeration,Negativism,Withdrawal | 98.9 | 97.8 | 99.8 |
| 8 | Excitement,Staring,Posturing/catalepsy,Echopraxia/echolalia,Stereotypy,Mannerisms,Negativism,Withdrawal | 98.9 | 97.8 | 99.8 |
| 8 | Excitement,Staring,Posturing/catalepsy,Grimacing,Echopraxia/echolalia,Rigidity,Waxy flexibility,Withdrawal | 98.9 | 97.8 | 99.8 |
| 8 | Excitement,Staring,Posturing/catalepsy,Grimacing,Echopraxia/echolalia,Stereotypy,Rigidity,Withdrawal | 98.9 | 97.8 | 99.8 |
| 8 | Excitement,Staring,Posturing/catalepsy,Grimacing,Echopraxia/echolalia,Verbigeration,Rigidity,Withdrawal | 98.9 | 97.8 | 99.8 |
| 8 | Excitement,Staring,Posturing/catalepsy,Grimacing,Stereotypy,Rigidity,Negativism,Withdrawal | 98.9 | 97.8 | 99.8 |
| 8 | Excitement,Staring,Posturing/catalepsy,Grimacing,Stereotypy,Verbigeration,Rigidity,Withdrawal | 98.9 | 97.8 | 99.8 |
| 8 | Excitement,Staring,Posturing/catalepsy,Grimacing,Verbigeration,Rigidity,Negativism,Withdrawal | 98.9 | 97.8 | 99.8 |
| 8 | Excitement,Staring,Posturing/catalepsy,Stereotypy,Mannerisms,Rigidity,Waxy flexibility,Withdrawal | 98.9 | 97.8 | 99.8 |
| 8 | Excitement,Staring,Posturing/catalepsy,Stereotypy,Mannerisms,Verbigeration,Negativism,Withdrawal | 98.9 | 97.8 | 99.8 |
| 8 | Excitement,Staring,Posturing/catalepsy,Stereotypy,Rigidity,Negativism,Waxy flexibility,Withdrawal | 98.9 | 97.8 | 99.8 |
| 8 | Excitement,Staring,Posturing/catalepsy,Verbigeration,Rigidity,Negativism,Waxy flexibility,Withdrawal | 98.9 | 97.8 | 99.8 |
| 8 | Immobility/stupor,Echopraxia/echolalia,Stereotypy,Mannerisms,Verbigeration,Rigidity,Negativism,Withdrawal | 98.9 | 97.8 | 99.8 |
| 8 | Immobility/stupor,Grimacing,Echopraxia/echolalia,Stereotypy,Mannerisms,Rigidity,Negativism,Withdrawal | 98.9 | 97.8 | 99.8 |
| 8 | Immobility/stupor,Mutism,Staring,Echopraxia/echolalia,Mannerisms,Rigidity,Negativism,Waxy flexibility | 98.9 | 97.8 | 99.8 |
| 8 | Immobility/stupor,Mutism,Staring,Echopraxia/echolalia,Mannerisms,Verbigeration,Rigidity,Negativism | 98.9 | 97.8 | 99.8 |
| 8 | Immobility/stupor,Mutism,Staring,Grimacing,Echopraxia/echolalia,Mannerisms,Negativism,Withdrawal | 98.9 | 97.8 | 99.8 |
| 8 | Immobility/stupor,Mutism,Staring,Grimacing,Echopraxia/echolalia,Mannerisms,Rigidity,Negativism | 98.9 | 97.8 | 99.8 |
| 8 | Immobility/stupor,Mutism,Staring,Grimacing,Mannerisms,Rigidity,Negativism,Withdrawal | 98.9 | 97.8 | 99.8 |
| 8 | Immobility/stupor,Mutism,Staring,Grimacing,Stereotypy,Mannerisms,Negativism,Withdrawal | 98.9 | 97.8 | 99.8 |
| 8 | Immobility/stupor,Mutism,Staring,Grimacing,Stereotypy,Mannerisms,Rigidity,Negativism | 98.9 | 97.8 | 99.8 |
| 8 | Immobility/stupor,Mutism,Staring,Grimacing,Stereotypy,Mannerisms,Rigidity,Withdrawal | 98.9 | 97.8 | 99.8 |
| 8 | Immobility/stupor,Mutism,Staring,Grimacing,Stereotypy,Verbigeration,Rigidity,Negativism | 98.9 | 97.8 | 99.8 |
| 8 | Immobility/stupor,Mutism,Staring,Mannerisms,Rigidity,Negativism,Waxy flexibility,Withdrawal | 98.9 | 97.8 | 99.8 |
| 8 | Immobility/stupor,Mutism,Staring,Posturing/catalepsy,Echopraxia/echolalia,Stereotypy,Mannerisms,Negativism | 98.9 | 97.8 | 99.8 |
| 8 | Immobility/stupor,Mutism,Staring,Posturing/catalepsy,Grimacing,Echopraxia/echolalia,Mannerisms,Negativism | 98.9 | 97.8 | 99.8 |
| 8 | Immobility/stupor,Mutism,Staring,Posturing/catalepsy,Grimacing,Stereotypy,Mannerisms,Negativism | 98.9 | 97.8 | 99.8 |
| 8 | Immobility/stupor,Mutism,Staring,Posturing/catalepsy,Mannerisms,Negativism,Waxy flexibility,Withdrawal | 98.9 | 97.8 | 99.8 |
| 8 | Immobility/stupor,Mutism,Staring,Posturing/catalepsy,Mannerisms,Verbigeration,Rigidity,Negativism | 98.9 | 97.8 | 99.8 |
| 8 | Immobility/stupor,Mutism,Staring,Posturing/catalepsy,Stereotypy,Mannerisms,Rigidity,Withdrawal | 98.9 | 97.8 | 99.8 |
| 8 | Immobility/stupor,Mutism,Staring,Posturing/catalepsy,Stereotypy,Mannerisms,Verbigeration,Negativism | 98.9 | 97.8 | 99.8 |
| 8 | Immobility/stupor,Mutism,Staring,Posturing/catalepsy,Stereotypy,Verbigeration,Rigidity,Negativism | 98.9 | 97.8 | 99.8 |
| 8 | Immobility/stupor,Mutism,Staring,Stereotypy,Mannerisms,Verbigeration,Negativism,Withdrawal | 98.9 | 97.8 | 99.8 |
| 8 | Immobility/stupor,Mutism,Stereotypy,Mannerisms,Verbigeration,Rigidity,Negativism,Withdrawal | 98.9 | 97.8 | 99.8 |
| 8 | Immobility/stupor,Posturing/catalepsy,Grimacing,Echopraxia/echolalia,Mannerisms,Rigidity,Negativism,Withdrawal | 98.9 | 97.8 | 99.8 |
| 8 | Immobility/stupor,Posturing/catalepsy,Stereotypy,Mannerisms,Verbigeration,Rigidity,Negativism,Withdrawal | 98.9 | 97.8 | 99.8 |
| 8 | Immobility/stupor,Staring,Echopraxia/echolalia,Mannerisms,Verbigeration,Rigidity,Negativism,Waxy flexibility | 98.9 | 97.8 | 99.8 |
| 8 | Immobility/stupor,Staring,Echopraxia/echolalia,Stereotypy,Mannerisms,Negativism,Waxy flexibility,Withdrawal | 98.9 | 97.8 | 99.8 |
| 8 | Immobility/stupor,Staring,Echopraxia/echolalia,Stereotypy,Mannerisms,Verbigeration,Negativism,Withdrawal | 98.9 | 97.8 | 99.8 |
| 8 | Immobility/stupor,Staring,Echopraxia/echolalia,Stereotypy,Verbigeration,Rigidity,Negativism,Withdrawal | 98.9 | 97.8 | 99.8 |
| 8 | Immobility/stupor,Staring,Grimacing,Echopraxia/echolalia,Mannerisms,Rigidity,Negativism,Waxy flexibility | 98.9 | 97.8 | 99.8 |
| 8 | Immobility/stupor,Staring,Grimacing,Echopraxia/echolalia,Mannerisms,Verbigeration,Rigidity,Negativism | 98.9 | 97.8 | 99.8 |
| 8 | Immobility/stupor,Staring,Grimacing,Echopraxia/echolalia,Stereotypy,Mannerisms,Rigidity,Withdrawal | 98.9 | 97.8 | 99.8 |
| 8 | Immobility/stupor,Staring,Grimacing,Stereotypy,Mannerisms,Rigidity,Waxy flexibility,Withdrawal | 98.9 | 97.8 | 99.8 |
| 8 | Immobility/stupor,Staring,Grimacing,Stereotypy,Mannerisms,Verbigeration,Rigidity,Withdrawal | 98.9 | 97.8 | 99.8 |
| 8 | Immobility/stupor,Staring,Posturing/catalepsy,Echopraxia/echolalia,Mannerisms,Negativism,Waxy flexibility,Withdrawal | 98.9 | 97.8 | 99.8 |
| 8 | Immobility/stupor,Staring,Posturing/catalepsy,Echopraxia/echolalia,Mannerisms,Rigidity,Negativism,Waxy flexibility | 98.9 | 97.8 | 99.8 |
| 8 | Immobility/stupor,Staring,Posturing/catalepsy,Echopraxia/echolalia,Mannerisms,Verbigeration,Negativism,Withdrawal | 98.9 | 97.8 | 99.8 |
| 8 | Immobility/stupor,Staring,Posturing/catalepsy,Grimacing,Echopraxia/echolalia,Stereotypy,Mannerisms,Negativism | 98.9 | 97.8 | 99.8 |
| 8 | Immobility/stupor,Staring,Posturing/catalepsy,Grimacing,Mannerisms,Verbigeration,Rigidity,Negativism | 98.9 | 97.8 | 99.8 |
| 8 | Immobility/stupor,Staring,Posturing/catalepsy,Grimacing,Stereotypy,Mannerisms,Rigidity,Withdrawal | 98.9 | 97.8 | 99.8 |
| 8 | Immobility/stupor,Staring,Posturing/catalepsy,Grimacing,Stereotypy,Mannerisms,Verbigeration,Negativism | 98.9 | 97.8 | 99.8 |
| 8 | Immobility/stupor,Staring,Posturing/catalepsy,Stereotypy,Mannerisms,Negativism,Waxy flexibility,Withdrawal | 98.9 | 97.8 | 99.8 |
| 8 | Immobility/stupor,Staring,Posturing/catalepsy,Stereotypy,Mannerisms,Rigidity,Negativism,Waxy flexibility | 98.9 | 97.8 | 99.8 |
| 8 | Immobility/stupor,Staring,Posturing/catalepsy,Stereotypy,Verbigeration,Rigidity,Negativism,Withdrawal | 98.9 | 97.8 | 99.8 |
| 8 | Immobility/stupor,Staring,Stereotypy,Verbigeration,Rigidity,Negativism,Waxy flexibility,Withdrawal | 98.9 | 97.8 | 99.8 |
| 8 | Mutism,Staring,Echopraxia/echolalia,Stereotypy,Mannerisms,Rigidity,Waxy flexibility,Withdrawal | 98.9 | 97.8 | 99.8 |
| 8 | Mutism,Staring,Echopraxia/echolalia,Stereotypy,Mannerisms,Verbigeration,Negativism,Waxy flexibility | 98.9 | 97.8 | 99.8 |
| 8 | Mutism,Staring,Grimacing,Echopraxia/echolalia,Mannerisms,Rigidity,Waxy flexibility,Withdrawal | 98.9 | 97.8 | 99.8 |
| 8 | Mutism,Staring,Grimacing,Echopraxia/echolalia,Mannerisms,Verbigeration,Negativism,Waxy flexibility | 98.9 | 97.8 | 99.8 |
| 8 | Mutism,Staring,Grimacing,Echopraxia/echolalia,Mannerisms,Verbigeration,Negativism,Withdrawal | 98.9 | 97.8 | 99.8 |
| 8 | Mutism,Staring,Grimacing,Echopraxia/echolalia,Mannerisms,Verbigeration,Rigidity,Withdrawal | 98.9 | 97.8 | 99.8 |
| 8 | Mutism,Staring,Grimacing,Echopraxia/echolalia,Stereotypy,Mannerisms,Verbigeration,Negativism | 98.9 | 97.8 | 99.8 |
| 8 | Mutism,Staring,Grimacing,Echopraxia/echolalia,Stereotypy,Rigidity,Negativism,Withdrawal | 98.9 | 97.8 | 99.8 |
| 8 | Mutism,Staring,Grimacing,Echopraxia/echolalia,Stereotypy,Verbigeration,Rigidity,Negativism | 98.9 | 97.8 | 99.8 |
| 8 | Mutism,Staring,Grimacing,Mannerisms,Verbigeration,Rigidity,Waxy flexibility,Withdrawal | 98.9 | 97.8 | 99.8 |
| 8 | Mutism,Staring,Grimacing,Stereotypy,Mannerisms,Verbigeration,Rigidity,Waxy flexibility | 98.9 | 97.8 | 99.8 |
| 8 | Mutism,Staring,Grimacing,Stereotypy,Verbigeration,Rigidity,Negativism,Waxy flexibility | 98.9 | 97.8 | 99.8 |
| 8 | Mutism,Staring,Grimacing,Stereotypy,Verbigeration,Rigidity,Waxy flexibility,Withdrawal | 98.9 | 97.8 | 99.8 |
| 8 | Mutism,Staring,Posturing/catalepsy,Echopraxia/echolalia,Mannerisms,Verbigeration,Negativism,Waxy flexibility | 98.9 | 97.8 | 99.8 |
| 8 | Mutism,Staring,Posturing/catalepsy,Echopraxia/echolalia,Mannerisms,Verbigeration,Rigidity,Withdrawal | 98.9 | 97.8 | 99.8 |
| 8 | Mutism,Staring,Posturing/catalepsy,Echopraxia/echolalia,Stereotypy,Rigidity,Negativism,Withdrawal | 98.9 | 97.8 | 99.8 |
| 8 | Mutism,Staring,Posturing/catalepsy,Echopraxia/echolalia,Stereotypy,Verbigeration,Rigidity,Negativism | 98.9 | 97.8 | 99.8 |
| 8 | Mutism,Staring,Posturing/catalepsy,Grimacing,Echopraxia/echolalia,Mannerisms,Verbigeration,Rigidity | 98.9 | 97.8 | 99.8 |
| 8 | Mutism,Staring,Posturing/catalepsy,Grimacing,Echopraxia/echolalia,Stereotypy,Mannerisms,Withdrawal | 98.9 | 97.8 | 99.8 |
| 8 | Mutism,Staring,Posturing/catalepsy,Grimacing,Echopraxia/echolalia,Stereotypy,Negativism,Withdrawal | 98.9 | 97.8 | 99.8 |
| 8 | Mutism,Staring,Posturing/catalepsy,Grimacing,Mannerisms,Verbigeration,Rigidity,Negativism | 98.9 | 97.8 | 99.8 |
| 8 | Mutism,Staring,Posturing/catalepsy,Grimacing,Stereotypy,Mannerisms,Negativism,Waxy flexibility | 98.9 | 97.8 | 99.8 |
| 8 | Mutism,Staring,Posturing/catalepsy,Grimacing,Stereotypy,Mannerisms,Verbigeration,Rigidity | 98.9 | 97.8 | 99.8 |
| 8 | Mutism,Staring,Posturing/catalepsy,Grimacing,Stereotypy,Verbigeration,Rigidity,Negativism | 98.9 | 97.8 | 99.8 |
| 8 | Mutism,Staring,Posturing/catalepsy,Grimacing,Stereotypy,Verbigeration,Rigidity,Withdrawal | 98.9 | 97.8 | 99.8 |
| 8 | Mutism,Staring,Posturing/catalepsy,Stereotypy,Mannerisms,Rigidity,Negativism,Waxy flexibility | 98.9 | 97.8 | 99.8 |
| 8 | Mutism,Staring,Posturing/catalepsy,Stereotypy,Mannerisms,Rigidity,Waxy flexibility,Withdrawal | 98.9 | 97.8 | 99.8 |
| 8 | Mutism,Staring,Posturing/catalepsy,Stereotypy,Mannerisms,Verbigeration,Negativism,Waxy flexibility | 98.9 | 97.8 | 99.8 |
| 8 | Mutism,Staring,Stereotypy,Mannerisms,Verbigeration,Negativism,Waxy flexibility,Withdrawal | 98.9 | 97.8 | 99.8 |
| 8 | Mutism,Staring,Stereotypy,Mannerisms,Verbigeration,Rigidity,Waxy flexibility,Withdrawal | 98.9 | 97.8 | 99.8 |
| 8 | Staring,Echopraxia/echolalia,Mannerisms,Verbigeration,Rigidity,Negativism,Waxy flexibility,Withdrawal | 98.9 | 97.8 | 99.8 |
| 8 | Staring,Grimacing,Echopraxia/echolalia,Mannerisms,Verbigeration,Rigidity,Negativism,Withdrawal | 98.9 | 97.8 | 99.8 |
| 8 | Staring,Grimacing,Mannerisms,Verbigeration,Rigidity,Negativism,Waxy flexibility,Withdrawal | 98.9 | 97.8 | 99.8 |
| 8 | Staring,Posturing/catalepsy,Grimacing,Mannerisms,Rigidity,Negativism,Waxy flexibility,Withdrawal | 98.9 | 97.8 | 99.8 |
| 8 | Staring,Posturing/catalepsy,Mannerisms,Verbigeration,Rigidity,Negativism,Waxy flexibility,Withdrawal | 98.9 | 97.8 | 99.8 |
| 8 | Excitement,Immobility/stupor,Echopraxia/echolalia,Mannerisms,Rigidity,Negativism,Waxy flexibility,Withdrawal | 98.7 | 97.5 | 99.6 |
| 8 | Excitement,Immobility/stupor,Echopraxia/echolalia,Stereotypy,Verbigeration,Rigidity,Negativism,Waxy flexibility | 98.7 | 97.5 | 99.6 |
| 8 | Excitement,Immobility/stupor,Grimacing,Echopraxia/echolalia,Verbigeration,Rigidity,Negativism,Waxy flexibility | 98.7 | 97.5 | 99.6 |
| 8 | Excitement,Immobility/stupor,Mannerisms,Verbigeration,Rigidity,Negativism,Waxy flexibility,Withdrawal | 98.7 | 97.5 | 99.6 |
| 8 | Excitement,Immobility/stupor,Mutism,Echopraxia/echolalia,Mannerisms,Verbigeration,Rigidity,Withdrawal | 98.7 | 97.5 | 99.6 |
| 8 | Excitement,Immobility/stupor,Mutism,Echopraxia/echolalia,Stereotypy,Mannerisms,Rigidity,Negativism | 98.7 | 97.5 | 99.6 |
| 8 | Excitement,Immobility/stupor,Mutism,Echopraxia/echolalia,Stereotypy,Mannerisms,Rigidity,Withdrawal | 98.7 | 97.5 | 99.6 |
| 8 | Excitement,Immobility/stupor,Mutism,Echopraxia/echolalia,Stereotypy,Verbigeration,Rigidity,Waxy flexibility | 98.7 | 97.5 | 99.6 |
| 8 | Excitement,Immobility/stupor,Mutism,Grimacing,Echopraxia/echolalia,Mannerisms,Rigidity,Negativism | 98.7 | 97.5 | 99.6 |
| 8 | Excitement,Immobility/stupor,Mutism,Grimacing,Echopraxia/echolalia,Mannerisms,Rigidity,Withdrawal | 98.7 | 97.5 | 99.6 |
| 8 | Excitement,Immobility/stupor,Mutism,Grimacing,Mannerisms,Verbigeration,Rigidity,Withdrawal | 98.7 | 97.5 | 99.6 |
| 8 | Excitement,Immobility/stupor,Mutism,Grimacing,Stereotypy,Rigidity,Negativism,Waxy flexibility | 98.7 | 97.5 | 99.6 |
| 8 | Excitement,Immobility/stupor,Mutism,Grimacing,Verbigeration,Rigidity,Waxy flexibility,Withdrawal | 98.7 | 97.5 | 99.6 |
| 8 | Excitement,Immobility/stupor,Mutism,Posturing/catalepsy,Echopraxia/echolalia,Stereotypy,Negativism,Withdrawal | 98.7 | 97.5 | 99.6 |
| 8 | Excitement,Immobility/stupor,Mutism,Posturing/catalepsy,Grimacing,Echopraxia/echolalia,Mannerisms,Rigidity | 98.7 | 97.5 | 99.6 |
| 8 | Excitement,Immobility/stupor,Mutism,Posturing/catalepsy,Grimacing,Echopraxia/echolalia,Rigidity,Waxy flexibility | 98.7 | 97.5 | 99.6 |
| 8 | Excitement,Immobility/stupor,Mutism,Posturing/catalepsy,Grimacing,Echopraxia/echolalia,Stereotypy,Rigidity | 98.7 | 97.5 | 99.6 |
| 8 | Excitement,Immobility/stupor,Mutism,Posturing/catalepsy,Grimacing,Rigidity,Waxy flexibility,Withdrawal | 98.7 | 97.5 | 99.6 |
| 8 | Excitement,Immobility/stupor,Mutism,Posturing/catalepsy,Rigidity,Negativism,Waxy flexibility,Withdrawal | 98.7 | 97.5 | 99.6 |
| 8 | Excitement,Immobility/stupor,Mutism,Posturing/catalepsy,Stereotypy,Mannerisms,Rigidity,Negativism | 98.7 | 97.5 | 99.6 |
| 8 | Excitement,Immobility/stupor,Mutism,Posturing/catalepsy,Stereotypy,Mannerisms,Rigidity,Withdrawal | 98.7 | 97.5 | 99.6 |
| 8 | Excitement,Immobility/stupor,Mutism,Posturing/catalepsy,Stereotypy,Mannerisms,Verbigeration,Rigidity | 98.7 | 97.5 | 99.6 |
| 8 | Excitement,Immobility/stupor,Mutism,Posturing/catalepsy,Stereotypy,Rigidity,Negativism,Waxy flexibility | 98.7 | 97.5 | 99.6 |
| 8 | Excitement,Immobility/stupor,Mutism,Posturing/catalepsy,Stereotypy,Rigidity,Waxy flexibility,Withdrawal | 98.7 | 97.5 | 99.6 |
| 8 | Excitement,Immobility/stupor,Mutism,Posturing/catalepsy,Stereotypy,Verbigeration,Rigidity,Waxy flexibility | 98.7 | 97.5 | 99.6 |
| 8 | Excitement,Immobility/stupor,Mutism,Staring,Echopraxia/echolalia,Mannerisms,Waxy flexibility,Withdrawal | 98.7 | 97.5 | 99.6 |
| 8 | Excitement,Immobility/stupor,Mutism,Staring,Echopraxia/echolalia,Verbigeration,Waxy flexibility,Withdrawal | 98.7 | 97.5 | 99.6 |
| 8 | Excitement,Immobility/stupor,Mutism,Staring,Grimacing,Echopraxia/echolalia,Stereotypy,Waxy flexibility | 98.7 | 97.5 | 99.6 |
| 8 | Excitement,Immobility/stupor,Mutism,Staring,Grimacing,Echopraxia/echolalia,Stereotypy,Withdrawal | 98.7 | 97.5 | 99.6 |
| 8 | Excitement,Immobility/stupor,Mutism,Staring,Grimacing,Echopraxia/echolalia,Verbigeration,Waxy flexibility | 98.7 | 97.5 | 99.6 |
| 8 | Excitement,Immobility/stupor,Mutism,Staring,Grimacing,Negativism,Waxy flexibility,Withdrawal | 98.7 | 97.5 | 99.6 |
| 8 | Excitement,Immobility/stupor,Mutism,Staring,Grimacing,Stereotypy,Mannerisms,Rigidity | 98.7 | 97.5 | 99.6 |
| 8 | Excitement,Immobility/stupor,Mutism,Staring,Grimacing,Verbigeration,Negativism,Withdrawal | 98.7 | 97.5 | 99.6 |
| 8 | Excitement,Immobility/stupor,Mutism,Staring,Posturing/catalepsy,Echopraxia/echolalia,Mannerisms,Verbigeration | 98.7 | 97.5 | 99.6 |
| 8 | Excitement,Immobility/stupor,Mutism,Staring,Posturing/catalepsy,Echopraxia/echolalia,Stereotypy,Verbigeration | 98.7 | 97.5 | 99.6 |
| 8 | Excitement,Immobility/stupor,Mutism,Staring,Posturing/catalepsy,Grimacing,Echopraxia/echolalia,Mannerisms | 98.7 | 97.5 | 99.6 |
| 8 | Excitement,Immobility/stupor,Mutism,Staring,Posturing/catalepsy,Grimacing,Echopraxia/echolalia,Stereotypy | 98.7 | 97.5 | 99.6 |
| 8 | Excitement,Immobility/stupor,Mutism,Staring,Posturing/catalepsy,Grimacing,Mannerisms,Verbigeration | 98.7 | 97.5 | 99.6 |
| 8 | Excitement,Immobility/stupor,Mutism,Staring,Posturing/catalepsy,Grimacing,Mannerisms,Waxy flexibility | 98.7 | 97.5 | 99.6 |
| 8 | Excitement,Immobility/stupor,Mutism,Staring,Posturing/catalepsy,Grimacing,Stereotypy,Verbigeration | 98.7 | 97.5 | 99.6 |
| 8 | Excitement,Immobility/stupor,Mutism,Staring,Posturing/catalepsy,Grimacing,Stereotypy,Waxy flexibility | 98.7 | 97.5 | 99.6 |
| 8 | Excitement,Immobility/stupor,Mutism,Staring,Posturing/catalepsy,Stereotypy,Mannerisms,Withdrawal | 98.7 | 97.5 | 99.6 |
| 8 | Excitement,Immobility/stupor,Mutism,Staring,Posturing/catalepsy,Stereotypy,Verbigeration,Withdrawal | 98.7 | 97.5 | 99.6 |
| 8 | Excitement,Immobility/stupor,Mutism,Stereotypy,Mannerisms,Rigidity,Negativism,Withdrawal | 98.7 | 97.5 | 99.6 |
| 8 | Excitement,Immobility/stupor,Posturing/catalepsy,Echopraxia/echolalia,Mannerisms,Verbigeration,Rigidity,Negativism | 98.7 | 97.5 | 99.6 |
| 8 | Excitement,Immobility/stupor,Posturing/catalepsy,Echopraxia/echolalia,Stereotypy,Mannerisms,Rigidity,Withdrawal | 98.7 | 97.5 | 99.6 |
| 8 | Excitement,Immobility/stupor,Posturing/catalepsy,Echopraxia/echolalia,Stereotypy,Verbigeration,Rigidity,Negativism | 98.7 | 97.5 | 99.6 |
| 8 | Excitement,Immobility/stupor,Posturing/catalepsy,Echopraxia/echolalia,Verbigeration,Rigidity,Negativism,Waxy flexibility | 98.7 | 97.5 | 99.6 |
| 8 | Excitement,Immobility/stupor,Posturing/catalepsy,Echopraxia/echolalia,Verbigeration,Rigidity,Waxy flexibility,Withdrawal | 98.7 | 97.5 | 99.6 |
| 8 | Excitement,Immobility/stupor,Posturing/catalepsy,Grimacing,Echopraxia/echolalia,Mannerisms,Rigidity,Negativism | 98.7 | 97.5 | 99.6 |
| 8 | Excitement,Immobility/stupor,Posturing/catalepsy,Grimacing,Echopraxia/echolalia,Rigidity,Negativism,Waxy flexibility | 98.7 | 97.5 | 99.6 |
| 8 | Excitement,Immobility/stupor,Posturing/catalepsy,Grimacing,Echopraxia/echolalia,Rigidity,Waxy flexibility,Withdrawal | 98.7 | 97.5 | 99.6 |
| 8 | Excitement,Immobility/stupor,Posturing/catalepsy,Grimacing,Echopraxia/echolalia,Stereotypy,Rigidity,Withdrawal | 98.7 | 97.5 | 99.6 |
| 8 | Excitement,Immobility/stupor,Posturing/catalepsy,Grimacing,Rigidity,Negativism,Waxy flexibility,Withdrawal | 98.7 | 97.5 | 99.6 |
| 8 | Excitement,Immobility/stupor,Posturing/catalepsy,Grimacing,Stereotypy,Verbigeration,Rigidity,Negativism | 98.7 | 97.5 | 99.6 |
| 8 | Excitement,Immobility/stupor,Posturing/catalepsy,Grimacing,Verbigeration,Rigidity,Negativism,Waxy flexibility | 98.7 | 97.5 | 99.6 |
| 8 | Excitement,Immobility/stupor,Posturing/catalepsy,Mannerisms,Rigidity,Negativism,Waxy flexibility,Withdrawal | 98.7 | 97.5 | 99.6 |
| 8 | Excitement,Immobility/stupor,Posturing/catalepsy,Mannerisms,Verbigeration,Rigidity,Negativism,Waxy flexibility | 98.7 | 97.5 | 99.6 |
| 8 | Excitement,Immobility/stupor,Posturing/catalepsy,Stereotypy,Mannerisms,Verbigeration,Rigidity,Negativism | 98.7 | 97.5 | 99.6 |
| 8 | Excitement,Immobility/stupor,Posturing/catalepsy,Stereotypy,Mannerisms,Verbigeration,Rigidity,Withdrawal | 98.7 | 97.5 | 99.6 |
| 8 | Excitement,Immobility/stupor,Posturing/catalepsy,Stereotypy,Rigidity,Negativism,Waxy flexibility,Withdrawal | 98.7 | 97.5 | 99.6 |
| 8 | Excitement,Immobility/stupor,Posturing/catalepsy,Stereotypy,Verbigeration,Rigidity,Negativism,Waxy flexibility | 98.7 | 97.5 | 99.6 |
| 8 | Excitement,Immobility/stupor,Staring,Echopraxia/echolalia,Stereotypy,Mannerisms,Verbigeration,Negativism | 98.7 | 97.5 | 99.6 |
| 8 | Excitement,Immobility/stupor,Staring,Grimacing,Echopraxia/echolalia,Mannerisms,Verbigeration,Rigidity | 98.7 | 97.5 | 99.6 |
| 8 | Excitement,Immobility/stupor,Staring,Grimacing,Echopraxia/echolalia,Stereotypy,Mannerisms,Rigidity | 98.7 | 97.5 | 99.6 |
| 8 | Excitement,Immobility/stupor,Staring,Grimacing,Echopraxia/echolalia,Stereotypy,Negativism,Waxy flexibility | 98.7 | 97.5 | 99.6 |
| 8 | Excitement,Immobility/stupor,Staring,Grimacing,Echopraxia/echolalia,Stereotypy,Verbigeration,Rigidity | 98.7 | 97.5 | 99.6 |
| 8 | Excitement,Immobility/stupor,Staring,Grimacing,Echopraxia/echolalia,Verbigeration,Negativism,Waxy flexibility | 98.7 | 97.5 | 99.6 |
| 8 | Excitement,Immobility/stupor,Staring,Grimacing,Echopraxia/echolalia,Verbigeration,Negativism,Withdrawal | 98.7 | 97.5 | 99.6 |
| 8 | Excitement,Immobility/stupor,Staring,Grimacing,Mannerisms,Verbigeration,Negativism,Waxy flexibility | 98.7 | 97.5 | 99.6 |
| 8 | Excitement,Immobility/stupor,Staring,Grimacing,Mannerisms,Verbigeration,Negativism,Withdrawal | 98.7 | 97.5 | 99.6 |
| 8 | Excitement,Immobility/stupor,Staring,Posturing/catalepsy,Echopraxia/echolalia,Mannerisms,Verbigeration,Withdrawal | 98.7 | 97.5 | 99.6 |
| 8 | Excitement,Immobility/stupor,Staring,Posturing/catalepsy,Echopraxia/echolalia,Stereotypy,Mannerisms,Withdrawal | 98.7 | 97.5 | 99.6 |
| 8 | Excitement,Immobility/stupor,Staring,Posturing/catalepsy,Echopraxia/echolalia,Stereotypy,Rigidity,Waxy flexibility | 98.7 | 97.5 | 99.6 |
| 8 | Excitement,Immobility/stupor,Staring,Posturing/catalepsy,Grimacing,Echopraxia/echolalia,Waxy flexibility,Withdrawal | 98.7 | 97.5 | 99.6 |
| 8 | Excitement,Immobility/stupor,Staring,Posturing/catalepsy,Grimacing,Mannerisms,Verbigeration,Withdrawal | 98.7 | 97.5 | 99.6 |
| 8 | Excitement,Immobility/stupor,Staring,Posturing/catalepsy,Grimacing,Stereotypy,Mannerisms,Negativism | 98.7 | 97.5 | 99.6 |
| 8 | Excitement,Immobility/stupor,Staring,Posturing/catalepsy,Grimacing,Stereotypy,Negativism,Waxy flexibility | 98.7 | 97.5 | 99.6 |
| 8 | Excitement,Immobility/stupor,Staring,Posturing/catalepsy,Grimacing,Stereotypy,Rigidity,Waxy flexibility | 98.7 | 97.5 | 99.6 |
| 8 | Excitement,Immobility/stupor,Staring,Posturing/catalepsy,Mannerisms,Verbigeration,Waxy flexibility,Withdrawal | 98.7 | 97.5 | 99.6 |
| 8 | Excitement,Immobility/stupor,Staring,Posturing/catalepsy,Stereotypy,Mannerisms,Rigidity,Waxy flexibility | 98.7 | 97.5 | 99.6 |
| 8 | Excitement,Immobility/stupor,Staring,Stereotypy,Mannerisms,Negativism,Waxy flexibility,Withdrawal | 98.7 | 97.5 | 99.6 |
| 8 | Excitement,Mutism,Staring,Grimacing,Echopraxia/echolalia,Mannerisms,Verbigeration,Negativism | 98.7 | 97.5 | 99.6 |
| 8 | Excitement,Mutism,Staring,Grimacing,Echopraxia/echolalia,Stereotypy,Mannerisms,Waxy flexibility | 98.7 | 97.5 | 99.6 |
| 8 | Excitement,Mutism,Staring,Grimacing,Echopraxia/echolalia,Stereotypy,Verbigeration,Withdrawal | 98.7 | 97.5 | 99.6 |
| 8 | Excitement,Mutism,Staring,Grimacing,Stereotypy,Mannerisms,Verbigeration,Negativism | 98.7 | 97.5 | 99.6 |
| 8 | Excitement,Mutism,Staring,Grimacing,Stereotypy,Verbigeration,Waxy flexibility,Withdrawal | 98.7 | 97.5 | 99.6 |
| 8 | Excitement,Mutism,Staring,Posturing/catalepsy,Grimacing,Echopraxia/echolalia,Mannerisms,Verbigeration | 98.7 | 97.5 | 99.6 |
| 8 | Excitement,Mutism,Staring,Posturing/catalepsy,Grimacing,Echopraxia/echolalia,Stereotypy,Mannerisms | 98.7 | 97.5 | 99.6 |
| 8 | Excitement,Mutism,Staring,Posturing/catalepsy,Stereotypy,Mannerisms,Verbigeration,Waxy flexibility | 98.7 | 97.5 | 99.6 |
| 8 | Excitement,Staring,Echopraxia/echolalia,Mannerisms,Verbigeration,Rigidity,Waxy flexibility,Withdrawal | 98.7 | 97.5 | 99.6 |
| 8 | Excitement,Staring,Echopraxia/echolalia,Stereotypy,Mannerisms,Negativism,Waxy flexibility,Withdrawal | 98.7 | 97.5 | 99.6 |
| 8 | Excitement,Staring,Grimacing,Echopraxia/echolalia,Mannerisms,Verbigeration,Rigidity,Withdrawal | 98.7 | 97.5 | 99.6 |
| 8 | Excitement,Staring,Grimacing,Echopraxia/echolalia,Stereotypy,Mannerisms,Negativism,Withdrawal | 98.7 | 97.5 | 99.6 |
| 8 | Excitement,Staring,Grimacing,Echopraxia/echolalia,Stereotypy,Verbigeration,Rigidity,Withdrawal | 98.7 | 97.5 | 99.6 |
| 8 | Excitement,Staring,Grimacing,Echopraxia/echolalia,Verbigeration,Rigidity,Negativism,Withdrawal | 98.7 | 97.5 | 99.6 |
| 8 | Excitement,Staring,Grimacing,Echopraxia/echolalia,Verbigeration,Rigidity,Waxy flexibility,Withdrawal | 98.7 | 97.5 | 99.6 |
| 8 | Excitement,Staring,Posturing/catalepsy,Echopraxia/echolalia,Stereotypy,Negativism,Waxy flexibility,Withdrawal | 98.7 | 97.5 | 99.6 |
| 8 | Excitement,Staring,Posturing/catalepsy,Echopraxia/echolalia,Stereotypy,Verbigeration,Rigidity,Withdrawal | 98.7 | 97.5 | 99.6 |
| 8 | Excitement,Staring,Posturing/catalepsy,Echopraxia/echolalia,Verbigeration,Negativism,Waxy flexibility,Withdrawal | 98.7 | 97.5 | 99.6 |
| 8 | Excitement,Staring,Posturing/catalepsy,Grimacing,Echopraxia/echolalia,Negativism,Waxy flexibility,Withdrawal | 98.7 | 97.5 | 99.6 |
| 8 | Excitement,Staring,Posturing/catalepsy,Grimacing,Echopraxia/echolalia,Stereotypy,Negativism,Withdrawal | 98.7 | 97.5 | 99.6 |
| 8 | Excitement,Staring,Posturing/catalepsy,Grimacing,Echopraxia/echolalia,Verbigeration,Negativism,Withdrawal | 98.7 | 97.5 | 99.6 |
| 8 | Excitement,Staring,Posturing/catalepsy,Grimacing,Rigidity,Negativism,Waxy flexibility,Withdrawal | 98.7 | 97.5 | 99.6 |
| 8 | Excitement,Staring,Posturing/catalepsy,Grimacing,Stereotypy,Mannerisms,Negativism,Withdrawal | 98.7 | 97.5 | 99.6 |
| 8 | Excitement,Staring,Posturing/catalepsy,Grimacing,Stereotypy,Rigidity,Waxy flexibility,Withdrawal | 98.7 | 97.5 | 99.6 |
| 8 | Excitement,Staring,Posturing/catalepsy,Grimacing,Stereotypy,Verbigeration,Negativism,Withdrawal | 98.7 | 97.5 | 99.6 |
| 8 | Excitement,Staring,Posturing/catalepsy,Stereotypy,Mannerisms,Negativism,Waxy flexibility,Withdrawal | 98.7 | 97.5 | 99.6 |
| 8 | Excitement,Staring,Posturing/catalepsy,Stereotypy,Verbigeration,Negativism,Waxy flexibility,Withdrawal | 98.7 | 97.5 | 99.6 |
| 8 | Excitement,Staring,Posturing/catalepsy,Stereotypy,Verbigeration,Rigidity,Waxy flexibility,Withdrawal | 98.7 | 97.5 | 99.6 |
| 8 | Immobility/stupor,Echopraxia/echolalia,Stereotypy,Mannerisms,Rigidity,Negativism,Waxy flexibility,Withdrawal | 98.7 | 97.5 | 99.6 |
| 8 | Immobility/stupor,Mutism,Echopraxia/echolalia,Stereotypy,Mannerisms,Rigidity,Negativism,Withdrawal | 98.7 | 97.5 | 99.6 |
| 8 | Immobility/stupor,Mutism,Grimacing,Echopraxia/echolalia,Mannerisms,Rigidity,Negativism,Withdrawal | 98.7 | 97.5 | 99.6 |
| 8 | Immobility/stupor,Mutism,Posturing/catalepsy,Stereotypy,Mannerisms,Verbigeration,Rigidity,Negativism | 98.7 | 97.5 | 99.6 |
| 8 | Immobility/stupor,Mutism,Staring,Echopraxia/echolalia,Stereotypy,Mannerisms,Negativism,Waxy flexibility | 98.7 | 97.5 | 99.6 |
| 8 | Immobility/stupor,Mutism,Staring,Echopraxia/echolalia,Stereotypy,Mannerisms,Rigidity,Withdrawal | 98.7 | 97.5 | 99.6 |
| 8 | Immobility/stupor,Mutism,Staring,Echopraxia/echolalia,Stereotypy,Rigidity,Negativism,Withdrawal | 98.7 | 97.5 | 99.6 |
| 8 | Immobility/stupor,Mutism,Staring,Grimacing,Echopraxia/echolalia,Stereotypy,Mannerisms,Negativism | 98.7 | 97.5 | 99.6 |
| 8 | Immobility/stupor,Mutism,Staring,Grimacing,Mannerisms,Verbigeration,Rigidity,Negativism | 98.7 | 97.5 | 99.6 |
| 8 | Immobility/stupor,Mutism,Staring,Grimacing,Mannerisms,Verbigeration,Rigidity,Withdrawal | 98.7 | 97.5 | 99.6 |
| 8 | Immobility/stupor,Mutism,Staring,Grimacing,Stereotypy,Mannerisms,Verbigeration,Rigidity | 98.7 | 97.5 | 99.6 |
| 8 | Immobility/stupor,Mutism,Staring,Mannerisms,Verbigeration,Rigidity,Negativism,Waxy flexibility | 98.7 | 97.5 | 99.6 |
| 8 | Immobility/stupor,Mutism,Staring,Posturing/catalepsy,Echopraxia/echolalia,Mannerisms,Verbigeration,Negativism | 98.7 | 97.5 | 99.6 |
| 8 | Immobility/stupor,Mutism,Staring,Posturing/catalepsy,Grimacing,Mannerisms,Rigidity,Withdrawal | 98.7 | 97.5 | 99.6 |
| 8 | Immobility/stupor,Mutism,Staring,Posturing/catalepsy,Grimacing,Mannerisms,Verbigeration,Negativism | 98.7 | 97.5 | 99.6 |
| 8 | Immobility/stupor,Mutism,Staring,Posturing/catalepsy,Mannerisms,Verbigeration,Rigidity,Withdrawal | 98.7 | 97.5 | 99.6 |
| 8 | Immobility/stupor,Mutism,Staring,Posturing/catalepsy,Stereotypy,Mannerisms,Negativism,Waxy flexibility | 98.7 | 97.5 | 99.6 |
| 8 | Immobility/stupor,Mutism,Staring,Posturing/catalepsy,Stereotypy,Verbigeration,Negativism,Withdrawal | 98.7 | 97.5 | 99.6 |
| 8 | Immobility/stupor,Mutism,Staring,Stereotypy,Mannerisms,Negativism,Waxy flexibility,Withdrawal | 98.7 | 97.5 | 99.6 |
| 8 | Immobility/stupor,Mutism,Staring,Stereotypy,Mannerisms,Rigidity,Waxy flexibility,Withdrawal | 98.7 | 97.5 | 99.6 |
| 8 | Immobility/stupor,Mutism,Staring,Stereotypy,Mannerisms,Verbigeration,Rigidity,Withdrawal | 98.7 | 97.5 | 99.6 |
| 8 | Immobility/stupor,Mutism,Staring,Stereotypy,Verbigeration,Rigidity,Negativism,Waxy flexibility | 98.7 | 97.5 | 99.6 |
| 8 | Immobility/stupor,Posturing/catalepsy,Echopraxia/echolalia,Mannerisms,Verbigeration,Rigidity,Negativism,Withdrawal | 98.7 | 97.5 | 99.6 |
| 8 | Immobility/stupor,Posturing/catalepsy,Echopraxia/echolalia,Stereotypy,Mannerisms,Rigidity,Negativism,Withdrawal | 98.7 | 97.5 | 99.6 |
| 8 | Immobility/stupor,Posturing/catalepsy,Grimacing,Stereotypy,Mannerisms,Rigidity,Negativism,Withdrawal | 98.7 | 97.5 | 99.6 |
| 8 | Immobility/stupor,Staring,Grimacing,Echopraxia/echolalia,Mannerisms,Negativism,Waxy flexibility,Withdrawal | 98.7 | 97.5 | 99.6 |
| 8 | Immobility/stupor,Staring,Grimacing,Echopraxia/echolalia,Stereotypy,Mannerisms,Negativism,Waxy flexibility | 98.7 | 97.5 | 99.6 |
| 8 | Immobility/stupor,Staring,Grimacing,Echopraxia/echolalia,Stereotypy,Rigidity,Negativism,Withdrawal | 98.7 | 97.5 | 99.6 |
| 8 | Immobility/stupor,Staring,Grimacing,Mannerisms,Verbigeration,Rigidity,Negativism,Waxy flexibility | 98.7 | 97.5 | 99.6 |
| 8 | Immobility/stupor,Staring,Grimacing,Stereotypy,Mannerisms,Negativism,Waxy flexibility,Withdrawal | 98.7 | 97.5 | 99.6 |
| 8 | Immobility/stupor,Staring,Posturing/catalepsy,Echopraxia/echolalia,Stereotypy,Mannerisms,Negativism,Waxy flexibility | 98.7 | 97.5 | 99.6 |
| 8 | Immobility/stupor,Staring,Posturing/catalepsy,Echopraxia/echolalia,Stereotypy,Mannerisms,Verbigeration,Negativism | 98.7 | 97.5 | 99.6 |
| 8 | Immobility/stupor,Staring,Posturing/catalepsy,Grimacing,Echopraxia/echolalia,Mannerisms,Verbigeration,Negativism | 98.7 | 97.5 | 99.6 |
| 8 | Immobility/stupor,Staring,Posturing/catalepsy,Grimacing,Mannerisms,Negativism,Waxy flexibility,Withdrawal | 98.7 | 97.5 | 99.6 |
| 8 | Immobility/stupor,Staring,Posturing/catalepsy,Grimacing,Stereotypy,Verbigeration,Negativism,Withdrawal | 98.7 | 97.5 | 99.6 |
| 8 | Immobility/stupor,Staring,Posturing/catalepsy,Grimacing,Stereotypy,Verbigeration,Rigidity,Negativism | 98.7 | 97.5 | 99.6 |
| 8 | Immobility/stupor,Staring,Posturing/catalepsy,Mannerisms,Verbigeration,Negativism,Waxy flexibility,Withdrawal | 98.7 | 97.5 | 99.6 |
| 8 | Immobility/stupor,Staring,Posturing/catalepsy,Mannerisms,Verbigeration,Rigidity,Negativism,Waxy flexibility | 98.7 | 97.5 | 99.6 |
| 8 | Immobility/stupor,Staring,Posturing/catalepsy,Stereotypy,Mannerisms,Verbigeration,Negativism,Waxy flexibility | 98.7 | 97.5 | 99.6 |
| 8 | Immobility/stupor,Stereotypy,Mannerisms,Verbigeration,Rigidity,Negativism,Waxy flexibility,Withdrawal | 98.7 | 97.5 | 99.6 |
| 8 | Mutism,Staring,Echopraxia/echolalia,Mannerisms,Verbigeration,Negativism,Waxy flexibility,Withdrawal | 98.7 | 97.5 | 99.6 |
| 8 | Mutism,Staring,Echopraxia/echolalia,Stereotypy,Mannerisms,Verbigeration,Rigidity,Withdrawal | 98.7 | 97.5 | 99.6 |
| 8 | Mutism,Staring,Echopraxia/echolalia,Stereotypy,Rigidity,Negativism,Waxy flexibility,Withdrawal | 98.7 | 97.5 | 99.6 |
| 8 | Mutism,Staring,Echopraxia/echolalia,Stereotypy,Verbigeration,Rigidity,Negativism,Waxy flexibility | 98.7 | 97.5 | 99.6 |
| 8 | Mutism,Staring,Grimacing,Echopraxia/echolalia,Mannerisms,Verbigeration,Rigidity,Waxy flexibility | 98.7 | 97.5 | 99.6 |
| 8 | Mutism,Staring,Grimacing,Echopraxia/echolalia,Stereotypy,Mannerisms,Rigidity,Waxy flexibility | 98.7 | 97.5 | 99.6 |
| 8 | Mutism,Staring,Grimacing,Echopraxia/echolalia,Stereotypy,Mannerisms,Verbigeration,Rigidity | 98.7 | 97.5 | 99.6 |
| 8 | Mutism,Staring,Grimacing,Echopraxia/echolalia,Stereotypy,Mannerisms,Waxy flexibility,Withdrawal | 98.7 | 97.5 | 99.6 |
| 8 | Mutism,Staring,Grimacing,Echopraxia/echolalia,Stereotypy,Verbigeration,Negativism,Withdrawal | 98.7 | 97.5 | 99.6 |
| 8 | Mutism,Staring,Grimacing,Echopraxia/echolalia,Stereotypy,Verbigeration,Rigidity,Withdrawal | 98.7 | 97.5 | 99.6 |
| 8 | Mutism,Staring,Grimacing,Echopraxia/echolalia,Verbigeration,Rigidity,Negativism,Withdrawal | 98.7 | 97.5 | 99.6 |
| 8 | Mutism,Staring,Grimacing,Mannerisms,Verbigeration,Negativism,Waxy flexibility,Withdrawal | 98.7 | 97.5 | 99.6 |
| 8 | Mutism,Staring,Grimacing,Mannerisms,Verbigeration,Rigidity,Negativism,Waxy flexibility | 98.7 | 97.5 | 99.6 |
| 8 | Mutism,Staring,Posturing/catalepsy,Echopraxia/echolalia,Mannerisms,Rigidity,Waxy flexibility,Withdrawal | 98.7 | 97.5 | 99.6 |
| 8 | Mutism,Staring,Posturing/catalepsy,Echopraxia/echolalia,Stereotypy,Verbigeration,Negativism,Withdrawal | 98.7 | 97.5 | 99.6 |
| 8 | Mutism,Staring,Posturing/catalepsy,Echopraxia/echolalia,Verbigeration,Rigidity,Negativism,Withdrawal | 98.7 | 97.5 | 99.6 |
| 8 | Mutism,Staring,Posturing/catalepsy,Grimacing,Echopraxia/echolalia,Mannerisms,Verbigeration,Withdrawal | 98.7 | 97.5 | 99.6 |
| 8 | Mutism,Staring,Posturing/catalepsy,Grimacing,Echopraxia/echolalia,Mannerisms,Waxy flexibility,Withdrawal | 98.7 | 97.5 | 99.6 |
| 8 | Mutism,Staring,Posturing/catalepsy,Grimacing,Echopraxia/echolalia,Rigidity,Negativism,Withdrawal | 98.7 | 97.5 | 99.6 |
| 8 | Mutism,Staring,Posturing/catalepsy,Grimacing,Echopraxia/echolalia,Stereotypy,Mannerisms,Rigidity | 98.7 | 97.5 | 99.6 |
| 8 | Mutism,Staring,Posturing/catalepsy,Grimacing,Echopraxia/echolalia,Stereotypy,Rigidity,Withdrawal | 98.7 | 97.5 | 99.6 |
| 8 | Mutism,Staring,Posturing/catalepsy,Grimacing,Echopraxia/echolalia,Stereotypy,Verbigeration,Negativism | 98.7 | 97.5 | 99.6 |
| 8 | Mutism,Staring,Posturing/catalepsy,Grimacing,Echopraxia/echolalia,Verbigeration,Rigidity,Negativism | 98.7 | 97.5 | 99.6 |
| 8 | Mutism,Staring,Posturing/catalepsy,Grimacing,Mannerisms,Rigidity,Waxy flexibility,Withdrawal | 98.7 | 97.5 | 99.6 |
| 8 | Mutism,Staring,Posturing/catalepsy,Grimacing,Stereotypy,Mannerisms,Verbigeration,Withdrawal | 98.7 | 97.5 | 99.6 |
| 8 | Mutism,Staring,Posturing/catalepsy,Grimacing,Stereotypy,Mannerisms,Waxy flexibility,Withdrawal | 98.7 | 97.5 | 99.6 |
| 8 | Mutism,Staring,Posturing/catalepsy,Grimacing,Stereotypy,Rigidity,Negativism,Withdrawal | 98.7 | 97.5 | 99.6 |
| 8 | Mutism,Staring,Posturing/catalepsy,Mannerisms,Verbigeration,Rigidity,Negativism,Waxy flexibility | 98.7 | 97.5 | 99.6 |
| 8 | Mutism,Staring,Posturing/catalepsy,Mannerisms,Verbigeration,Rigidity,Waxy flexibility,Withdrawal | 98.7 | 97.5 | 99.6 |
| 8 | Mutism,Staring,Posturing/catalepsy,Stereotypy,Verbigeration,Negativism,Waxy flexibility,Withdrawal | 98.7 | 97.5 | 99.6 |
| 8 | Staring,Grimacing,Echopraxia/echolalia,Stereotypy,Mannerisms,Negativism,Waxy flexibility,Withdrawal | 98.7 | 97.5 | 99.6 |
| 8 | Staring,Grimacing,Echopraxia/echolalia,Stereotypy,Mannerisms,Rigidity,Waxy flexibility,Withdrawal | 98.7 | 97.5 | 99.6 |
| 8 | Staring,Grimacing,Echopraxia/echolalia,Stereotypy,Mannerisms,Verbigeration,Negativism,Withdrawal | 98.7 | 97.5 | 99.6 |
| 8 | Staring,Posturing/catalepsy,Grimacing,Echopraxia/echolalia,Mannerisms,Negativism,Waxy flexibility,Withdrawal | 98.7 | 97.5 | 99.6 |
| 8 | Staring,Posturing/catalepsy,Grimacing,Echopraxia/echolalia,Mannerisms,Verbigeration,Negativism,Withdrawal | 98.7 | 97.5 | 99.6 |
| 8 | Staring,Posturing/catalepsy,Grimacing,Echopraxia/echolalia,Stereotypy,Mannerisms,Rigidity,Withdrawal | 98.7 | 97.5 | 99.6 |
| 8 | Staring,Posturing/catalepsy,Grimacing,Stereotypy,Mannerisms,Negativism,Waxy flexibility,Withdrawal | 98.7 | 97.5 | 99.6 |
| 8 | Staring,Posturing/catalepsy,Grimacing,Stereotypy,Mannerisms,Verbigeration,Rigidity,Withdrawal | 98.7 | 97.5 | 99.6 |
| 8 | Staring,Posturing/catalepsy,Stereotypy,Mannerisms,Verbigeration,Negativism,Waxy flexibility,Withdrawal | 98.7 | 97.5 | 99.6 |
| 8 | Excitement,Immobility/stupor,Echopraxia/echolalia,Stereotypy,Mannerisms,Verbigeration,Rigidity,Withdrawal | 98.6 | 97.5 | 99.6 |
| 8 | Excitement,Immobility/stupor,Echopraxia/echolalia,Stereotypy,Verbigeration,Rigidity,Waxy flexibility,Withdrawal | 98.6 | 97.5 | 99.6 |
| 8 | Excitement,Immobility/stupor,Mutism,Grimacing,Mannerisms,Rigidity,Negativism,Withdrawal | 98.6 | 97.5 | 99.6 |
| 8 | Excitement,Immobility/stupor,Mutism,Grimacing,Rigidity,Negativism,Waxy flexibility,Withdrawal | 98.6 | 97.5 | 99.6 |
| 8 | Excitement,Immobility/stupor,Mutism,Grimacing,Stereotypy,Rigidity,Waxy flexibility,Withdrawal | 98.6 | 97.5 | 99.6 |
| 8 | Excitement,Immobility/stupor,Mutism,Mannerisms,Verbigeration,Rigidity,Waxy flexibility,Withdrawal | 98.6 | 97.5 | 99.6 |
| 8 | Excitement,Immobility/stupor,Mutism,Posturing/catalepsy,Grimacing,Rigidity,Negativism,Waxy flexibility | 98.6 | 97.5 | 99.6 |
| 8 | Excitement,Immobility/stupor,Mutism,Posturing/catalepsy,Grimacing,Verbigeration,Negativism,Withdrawal | 98.6 | 97.5 | 99.6 |
| 8 | Excitement,Immobility/stupor,Mutism,Posturing/catalepsy,Grimacing,Verbigeration,Rigidity,Waxy flexibility | 98.6 | 97.5 | 99.6 |
| 8 | Excitement,Immobility/stupor,Mutism,Posturing/catalepsy,Mannerisms,Verbigeration,Negativism,Withdrawal | 98.6 | 97.5 | 99.6 |
| 8 | Excitement,Immobility/stupor,Mutism,Staring,Echopraxia/echolalia,Mannerisms,Verbigeration,Negativism | 98.6 | 97.5 | 99.6 |
| 8 | Excitement,Immobility/stupor,Mutism,Staring,Echopraxia/echolalia,Stereotypy,Verbigeration,Waxy flexibility | 98.6 | 97.5 | 99.6 |
| 8 | Excitement,Immobility/stupor,Mutism,Staring,Grimacing,Echopraxia/echolalia,Verbigeration,Negativism | 98.6 | 97.5 | 99.6 |
| 8 | Excitement,Immobility/stupor,Mutism,Staring,Grimacing,Mannerisms,Negativism,Waxy flexibility | 98.6 | 97.5 | 99.6 |
| 8 | Excitement,Immobility/stupor,Posturing/catalepsy,Grimacing,Mannerisms,Verbigeration,Rigidity,Withdrawal | 98.6 | 97.5 | 99.6 |
| 8 | Excitement,Immobility/stupor,Staring,Grimacing,Echopraxia/echolalia,Mannerisms,Negativism,Waxy flexibility | 98.6 | 97.5 | 99.6 |
| 8 | Excitement,Immobility/stupor,Staring,Grimacing,Stereotypy,Verbigeration,Negativism,Waxy flexibility | 98.6 | 97.5 | 99.6 |
| 8 | Excitement,Mutism,Staring,Grimacing,Echopraxia/echolalia,Stereotypy,Mannerisms,Withdrawal | 98.6 | 97.5 | 99.6 |
| 8 | Excitement,Mutism,Staring,Grimacing,Stereotypy,Mannerisms,Negativism,Waxy flexibility | 98.6 | 97.5 | 99.6 |
| 8 | Excitement,Mutism,Staring,Posturing/catalepsy,Echopraxia/echolalia,Stereotypy,Mannerisms,Verbigeration | 98.6 | 97.5 | 99.6 |
| 8 | Excitement,Mutism,Staring,Posturing/catalepsy,Grimacing,Mannerisms,Verbigeration,Waxy flexibility | 98.6 | 97.5 | 99.6 |
| 8 | Excitement,Staring,Echopraxia/echolalia,Stereotypy,Verbigeration,Rigidity,Waxy flexibility,Withdrawal | 98.6 | 97.5 | 99.6 |
| 8 | Excitement,Staring,Grimacing,Verbigeration,Rigidity,Negativism,Waxy flexibility,Withdrawal | 98.6 | 97.5 | 99.6 |
| 8 | Excitement,Staring,Posturing/catalepsy,Echopraxia/echolalia,Stereotypy,Rigidity,Waxy flexibility,Withdrawal | 98.6 | 97.5 | 99.6 |
| 8 | Excitement,Staring,Posturing/catalepsy,Echopraxia/echolalia,Verbigeration,Rigidity,Waxy flexibility,Withdrawal | 98.6 | 97.5 | 99.6 |
| 8 | Excitement,Staring,Posturing/catalepsy,Grimacing,Mannerisms,Negativism,Waxy flexibility,Withdrawal | 98.6 | 97.5 | 99.6 |
| 8 | Excitement,Staring,Posturing/catalepsy,Grimacing,Verbigeration,Rigidity,Waxy flexibility,Withdrawal | 98.6 | 97.5 | 99.6 |
| 8 | Immobility/stupor,Grimacing,Stereotypy,Mannerisms,Verbigeration,Rigidity,Negativism,Withdrawal | 98.6 | 97.5 | 99.6 |
| 8 | Immobility/stupor,Mutism,Staring,Echopraxia/echolalia,Mannerisms,Negativism,Waxy flexibility,Withdrawal | 98.6 | 97.5 | 99.6 |
| 8 | Immobility/stupor,Mutism,Staring,Grimacing,Echopraxia/echolalia,Mannerisms,Rigidity,Withdrawal | 98.6 | 97.5 | 99.6 |
| 8 | Immobility/stupor,Mutism,Staring,Grimacing,Stereotypy,Verbigeration,Rigidity,Withdrawal | 98.6 | 97.5 | 99.6 |
| 8 | Immobility/stupor,Mutism,Staring,Posturing/catalepsy,Echopraxia/echolalia,Mannerisms,Rigidity,Withdrawal | 98.6 | 97.5 | 99.6 |
| 8 | Immobility/stupor,Mutism,Staring,Posturing/catalepsy,Grimacing,Mannerisms,Rigidity,Negativism | 98.6 | 97.5 | 99.6 |
| 8 | Immobility/stupor,Mutism,Staring,Posturing/catalepsy,Stereotypy,Mannerisms,Verbigeration,Rigidity | 98.6 | 97.5 | 99.6 |
| 8 | Immobility/stupor,Mutism,Stereotypy,Mannerisms,Verbigeration,Rigidity,Negativism,Waxy flexibility | 98.6 | 97.5 | 99.6 |
| 8 | Immobility/stupor,Posturing/catalepsy,Grimacing,Mannerisms,Verbigeration,Rigidity,Negativism,Withdrawal | 98.6 | 97.5 | 99.6 |
| 8 | Immobility/stupor,Staring,Grimacing,Stereotypy,Verbigeration,Rigidity,Negativism,Waxy flexibility | 98.6 | 97.5 | 99.6 |
| 8 | Immobility/stupor,Staring,Posturing/catalepsy,Echopraxia/echolalia,Stereotypy,Rigidity,Negativism,Withdrawal | 98.6 | 97.5 | 99.6 |
| 8 | Immobility/stupor,Staring,Stereotypy,Mannerisms,Verbigeration,Negativism,Waxy flexibility,Withdrawal | 98.6 | 97.5 | 99.6 |
| 8 | Mutism,Staring,Grimacing,Stereotypy,Mannerisms,Verbigeration,Negativism,Waxy flexibility | 98.6 | 97.5 | 99.6 |
| 8 | Mutism,Staring,Posturing/catalepsy,Echopraxia/echolalia,Stereotypy,Mannerisms,Verbigeration,Rigidity | 98.6 | 97.5 | 99.6 |
| 8 | Mutism,Staring,Posturing/catalepsy,Grimacing,Echopraxia/echolalia,Verbigeration,Negativism,Withdrawal | 98.6 | 97.5 | 99.6 |
| 8 | Mutism,Staring,Posturing/catalepsy,Grimacing,Mannerisms,Verbigeration,Negativism,Waxy flexibility | 98.6 | 97.5 | 99.6 |
| 8 | Mutism,Staring,Posturing/catalepsy,Stereotypy,Verbigeration,Rigidity,Negativism,Waxy flexibility | 98.6 | 97.5 | 99.6 |
| 8 | Staring,Grimacing,Stereotypy,Mannerisms,Verbigeration,Rigidity,Waxy flexibility,Withdrawal | 98.6 | 97.5 | 99.6 |
| 8 | Staring,Posturing/catalepsy,Echopraxia/echolalia,Stereotypy,Mannerisms,Negativism,Waxy flexibility,Withdrawal | 98.6 | 97.5 | 99.6 |
| 8 | Excitement,Immobility/stupor,Echopraxia/echolalia,Mannerisms,Verbigeration,Rigidity,Waxy flexibility,Withdrawal | 98.4 | 97.3 | 99.6 |
| 8 | Excitement,Immobility/stupor,Echopraxia/echolalia,Stereotypy,Mannerisms,Rigidity,Waxy flexibility,Withdrawal | 98.4 | 97.1 | 99.6 |
| 8 | Excitement,Immobility/stupor,Echopraxia/echolalia,Stereotypy,Mannerisms,Verbigeration,Rigidity,Negativism | 98.4 | 97.1 | 99.6 |
| 8 | Excitement,Immobility/stupor,Grimacing,Echopraxia/echolalia,Mannerisms,Rigidity,Negativism,Waxy flexibility | 98.4 | 97.3 | 99.6 |
| 8 | Excitement,Immobility/stupor,Grimacing,Echopraxia/echolalia,Mannerisms,Rigidity,Waxy flexibility,Withdrawal | 98.4 | 97.1 | 99.6 |
| 8 | Excitement,Immobility/stupor,Grimacing,Echopraxia/echolalia,Mannerisms,Verbigeration,Rigidity,Negativism | 98.4 | 97.3 | 99.6 |
| 8 | Excitement,Immobility/stupor,Grimacing,Echopraxia/echolalia,Mannerisms,Verbigeration,Rigidity,Withdrawal | 98.4 | 97.1 | 99.6 |
| 8 | Excitement,Immobility/stupor,Grimacing,Echopraxia/echolalia,Stereotypy,Mannerisms,Rigidity,Withdrawal | 98.4 | 97.1 | 99.6 |
| 8 | Excitement,Immobility/stupor,Grimacing,Echopraxia/echolalia,Stereotypy,Rigidity,Negativism,Waxy flexibility | 98.4 | 97.3 | 99.6 |
| 8 | Excitement,Immobility/stupor,Grimacing,Echopraxia/echolalia,Stereotypy,Rigidity,Waxy flexibility,Withdrawal | 98.4 | 97.1 | 99.6 |
| 8 | Excitement,Immobility/stupor,Grimacing,Echopraxia/echolalia,Stereotypy,Verbigeration,Rigidity,Negativism | 98.4 | 97.1 | 99.6 |
| 8 | Excitement,Immobility/stupor,Grimacing,Echopraxia/echolalia,Stereotypy,Verbigeration,Rigidity,Withdrawal | 98.4 | 97.1 | 99.6 |
| 8 | Excitement,Immobility/stupor,Grimacing,Echopraxia/echolalia,Verbigeration,Rigidity,Waxy flexibility,Withdrawal | 98.4 | 97.1 | 99.6 |
| 8 | Excitement,Immobility/stupor,Grimacing,Mannerisms,Rigidity,Negativism,Waxy flexibility,Withdrawal | 98.4 | 97.1 | 99.6 |
| 8 | Excitement,Immobility/stupor,Grimacing,Mannerisms,Verbigeration,Rigidity,Negativism,Waxy flexibility | 98.4 | 97.1 | 99.6 |
| 8 | Excitement,Immobility/stupor,Grimacing,Stereotypy,Mannerisms,Rigidity,Negativism,Withdrawal | 98.4 | 97.1 | 99.6 |
| 8 | Excitement,Immobility/stupor,Grimacing,Stereotypy,Rigidity,Negativism,Waxy flexibility,Withdrawal | 98.4 | 97.3 | 99.6 |
| 8 | Excitement,Immobility/stupor,Grimacing,Stereotypy,Verbigeration,Rigidity,Negativism,Waxy flexibility | 98.4 | 97.3 | 99.6 |
| 8 | Excitement,Immobility/stupor,Mutism,Echopraxia/echolalia,Mannerisms,Rigidity,Negativism,Waxy flexibility | 98.4 | 97.3 | 99.6 |
| 8 | Excitement,Immobility/stupor,Mutism,Echopraxia/echolalia,Mannerisms,Rigidity,Waxy flexibility,Withdrawal | 98.4 | 97.1 | 99.6 |
| 8 | Excitement,Immobility/stupor,Mutism,Echopraxia/echolalia,Stereotypy,Verbigeration,Negativism,Withdrawal | 98.4 | 97.1 | 99.6 |
| 8 | Excitement,Immobility/stupor,Mutism,Grimacing,Echopraxia/echolalia,Verbigeration,Rigidity,Waxy flexibility | 98.4 | 97.1 | 99.6 |
| 8 | Excitement,Immobility/stupor,Mutism,Grimacing,Mannerisms,Rigidity,Negativism,Waxy flexibility | 98.4 | 97.3 | 99.6 |
| 8 | Excitement,Immobility/stupor,Mutism,Grimacing,Mannerisms,Rigidity,Waxy flexibility,Withdrawal | 98.4 | 97.1 | 99.6 |
| 8 | Excitement,Immobility/stupor,Mutism,Grimacing,Stereotypy,Verbigeration,Rigidity,Waxy flexibility | 98.4 | 97.1 | 99.6 |
| 8 | Excitement,Immobility/stupor,Mutism,Mannerisms,Rigidity,Negativism,Waxy flexibility,Withdrawal | 98.4 | 97.1 | 99.6 |
| 8 | Excitement,Immobility/stupor,Mutism,Posturing/catalepsy,Echopraxia/echolalia,Mannerisms,Negativism,Withdrawal | 98.4 | 97.1 | 99.6 |
| 8 | Excitement,Immobility/stupor,Mutism,Posturing/catalepsy,Echopraxia/echolalia,Negativism,Waxy flexibility,Withdrawal | 98.4 | 97.1 | 99.6 |
| 8 | Excitement,Immobility/stupor,Mutism,Posturing/catalepsy,Echopraxia/echolalia,Stereotypy,Mannerisms,Rigidity | 98.4 | 97.3 | 99.6 |
| 8 | Excitement,Immobility/stupor,Mutism,Posturing/catalepsy,Echopraxia/echolalia,Stereotypy,Rigidity,Waxy flexibility | 98.4 | 97.1 | 99.6 |
| 8 | Excitement,Immobility/stupor,Mutism,Posturing/catalepsy,Grimacing,Echopraxia/echolalia,Verbigeration,Negativism | 98.4 | 97.1 | 99.6 |
| 8 | Excitement,Immobility/stupor,Mutism,Posturing/catalepsy,Grimacing,Mannerisms,Negativism,Withdrawal | 98.4 | 97.1 | 99.6 |
| 8 | Excitement,Immobility/stupor,Mutism,Posturing/catalepsy,Grimacing,Stereotypy,Negativism,Withdrawal | 98.4 | 97.3 | 99.6 |
| 8 | Excitement,Immobility/stupor,Mutism,Posturing/catalepsy,Mannerisms,Rigidity,Negativism,Waxy flexibility | 98.4 | 97.1 | 99.6 |
| 8 | Excitement,Immobility/stupor,Mutism,Posturing/catalepsy,Mannerisms,Rigidity,Waxy flexibility,Withdrawal | 98.4 | 97.3 | 99.6 |
| 8 | Excitement,Immobility/stupor,Mutism,Posturing/catalepsy,Mannerisms,Verbigeration,Rigidity,Waxy flexibility | 98.4 | 97.1 | 99.6 |
| 8 | Excitement,Immobility/stupor,Mutism,Posturing/catalepsy,Verbigeration,Negativism,Waxy flexibility,Withdrawal | 98.4 | 97.3 | 99.6 |
| 8 | Excitement,Immobility/stupor,Mutism,Staring,Echopraxia/echolalia,Mannerisms,Verbigeration,Waxy flexibility | 98.4 | 97.3 | 99.6 |
| 8 | Excitement,Immobility/stupor,Mutism,Staring,Echopraxia/echolalia,Stereotypy,Mannerisms,Waxy flexibility | 98.4 | 97.3 | 99.6 |
| 8 | Excitement,Immobility/stupor,Mutism,Staring,Echopraxia/echolalia,Stereotypy,Mannerisms,Withdrawal | 98.4 | 97.3 | 99.6 |
| 8 | Excitement,Immobility/stupor,Mutism,Staring,Echopraxia/echolalia,Stereotypy,Verbigeration,Withdrawal | 98.4 | 97.1 | 99.6 |
| 8 | Excitement,Immobility/stupor,Mutism,Staring,Grimacing,Echopraxia/echolalia,Mannerisms,Negativism | 98.4 | 97.1 | 99.6 |
| 8 | Excitement,Immobility/stupor,Mutism,Staring,Grimacing,Echopraxia/echolalia,Mannerisms,Waxy flexibility | 98.4 | 97.1 | 99.6 |
| 8 | Excitement,Immobility/stupor,Mutism,Staring,Grimacing,Mannerisms,Negativism,Withdrawal | 98.4 | 97.1 | 99.6 |
| 8 | Excitement,Immobility/stupor,Mutism,Staring,Grimacing,Mannerisms,Verbigeration,Negativism | 98.4 | 97.1 | 99.6 |
| 8 | Excitement,Immobility/stupor,Mutism,Staring,Grimacing,Stereotypy,Verbigeration,Waxy flexibility | 98.4 | 97.1 | 99.6 |
| 8 | Excitement,Immobility/stupor,Mutism,Staring,Grimacing,Stereotypy,Waxy flexibility,Withdrawal | 98.4 | 97.1 | 99.6 |
| 8 | Excitement,Immobility/stupor,Mutism,Staring,Posturing/catalepsy,Echopraxia/echolalia,Stereotypy,Mannerisms | 98.4 | 97.1 | 99.6 |
| 8 | Excitement,Immobility/stupor,Mutism,Staring,Posturing/catalepsy,Stereotypy,Mannerisms,Verbigeration | 98.4 | 97.1 | 99.6 |
| 8 | Excitement,Immobility/stupor,Mutism,Staring,Posturing/catalepsy,Stereotypy,Mannerisms,Waxy flexibility | 98.4 | 97.1 | 99.6 |
| 8 | Excitement,Immobility/stupor,Mutism,Staring,Stereotypy,Verbigeration,Waxy flexibility,Withdrawal | 98.4 | 97.1 | 99.6 |
| 8 | Excitement,Immobility/stupor,Mutism,Stereotypy,Mannerisms,Rigidity,Negativism,Waxy flexibility | 98.4 | 97.3 | 99.6 |
| 8 | Excitement,Immobility/stupor,Mutism,Stereotypy,Mannerisms,Rigidity,Waxy flexibility,Withdrawal | 98.4 | 97.1 | 99.6 |
| 8 | Excitement,Immobility/stupor,Posturing/catalepsy,Echopraxia/echolalia,Mannerisms,Rigidity,Waxy flexibility,Withdrawal | 98.4 | 97.3 | 99.6 |
| 8 | Excitement,Immobility/stupor,Posturing/catalepsy,Echopraxia/echolalia,Mannerisms,Verbigeration,Negativism,Withdrawal | 98.4 | 97.3 | 99.6 |
| 8 | Excitement,Immobility/stupor,Posturing/catalepsy,Echopraxia/echolalia,Stereotypy,Rigidity,Waxy flexibility,Withdrawal | 98.4 | 97.1 | 99.6 |
| 8 | Excitement,Immobility/stupor,Posturing/catalepsy,Echopraxia/echolalia,Stereotypy,Verbigeration,Negativism,Withdrawal | 98.4 | 97.1 | 99.6 |
| 8 | Excitement,Immobility/stupor,Posturing/catalepsy,Grimacing,Echopraxia/echolalia,Mannerisms,Negativism,Withdrawal | 98.4 | 97.1 | 99.6 |
| 8 | Excitement,Immobility/stupor,Posturing/catalepsy,Grimacing,Echopraxia/echolalia,Stereotypy,Rigidity,Negativism | 98.4 | 97.1 | 99.6 |
| 8 | Excitement,Immobility/stupor,Posturing/catalepsy,Grimacing,Echopraxia/echolalia,Verbigeration,Negativism,Withdrawal | 98.4 | 97.1 | 99.6 |
| 8 | Excitement,Immobility/stupor,Posturing/catalepsy,Grimacing,Mannerisms,Rigidity,Negativism,Waxy flexibility | 98.4 | 97.3 | 99.6 |
| 8 | Excitement,Immobility/stupor,Posturing/catalepsy,Grimacing,Mannerisms,Verbigeration,Negativism,Withdrawal | 98.4 | 97.1 | 99.6 |
| 8 | Excitement,Immobility/stupor,Posturing/catalepsy,Grimacing,Stereotypy,Verbigeration,Rigidity,Withdrawal | 98.4 | 97.1 | 99.6 |
| 8 | Excitement,Immobility/stupor,Posturing/catalepsy,Mannerisms,Verbigeration,Rigidity,Waxy flexibility,Withdrawal | 98.4 | 97.1 | 99.6 |
| 8 | Excitement,Immobility/stupor,Posturing/catalepsy,Stereotypy,Mannerisms,Verbigeration,Negativism,Withdrawal | 98.4 | 97.1 | 99.6 |
| 8 | Excitement,Immobility/stupor,Posturing/catalepsy,Stereotypy,Verbigeration,Rigidity,Waxy flexibility,Withdrawal | 98.4 | 97.1 | 99.6 |
| 8 | Excitement,Immobility/stupor,Staring,Echopraxia/echolalia,Stereotypy,Mannerisms,Waxy flexibility,Withdrawal | 98.4 | 97.1 | 99.6 |
| 8 | Excitement,Immobility/stupor,Staring,Grimacing,Echopraxia/echolalia,Mannerisms,Waxy flexibility,Withdrawal | 98.4 | 97.1 | 99.6 |
| 8 | Excitement,Immobility/stupor,Staring,Grimacing,Echopraxia/echolalia,Stereotypy,Mannerisms,Negativism | 98.4 | 97.3 | 99.6 |
| 8 | Excitement,Immobility/stupor,Staring,Grimacing,Echopraxia/echolalia,Stereotypy,Verbigeration,Negativism | 98.4 | 97.1 | 99.6 |
| 8 | Excitement,Immobility/stupor,Staring,Grimacing,Mannerisms,Negativism,Waxy flexibility,Withdrawal | 98.4 | 97.1 | 99.6 |
| 8 | Excitement,Immobility/stupor,Staring,Grimacing,Stereotypy,Mannerisms,Negativism,Withdrawal | 98.4 | 97.1 | 99.6 |
| 8 | Excitement,Immobility/stupor,Staring,Grimacing,Stereotypy,Mannerisms,Verbigeration,Negativism | 98.4 | 97.3 | 99.6 |
| 8 | Excitement,Immobility/stupor,Staring,Grimacing,Stereotypy,Negativism,Waxy flexibility,Withdrawal | 98.4 | 97.1 | 99.6 |
| 8 | Excitement,Immobility/stupor,Staring,Grimacing,Verbigeration,Negativism,Waxy flexibility,Withdrawal | 98.4 | 97.1 | 99.6 |
| 8 | Excitement,Immobility/stupor,Staring,Posturing/catalepsy,Echopraxia/echolalia,Mannerisms,Verbigeration,Waxy flexibility | 98.4 | 97.3 | 99.6 |
| 8 | Excitement,Immobility/stupor,Staring,Posturing/catalepsy,Echopraxia/echolalia,Stereotypy,Waxy flexibility,Withdrawal | 98.4 | 97.3 | 99.6 |
| 8 | Excitement,Immobility/stupor,Staring,Posturing/catalepsy,Echopraxia/echolalia,Verbigeration,Waxy flexibility,Withdrawal | 98.4 | 97.1 | 99.6 |
| 8 | Excitement,Immobility/stupor,Staring,Posturing/catalepsy,Grimacing,Echopraxia/echolalia,Mannerisms,Waxy flexibility | 98.4 | 97.1 | 99.6 |
| 8 | Excitement,Immobility/stupor,Staring,Posturing/catalepsy,Grimacing,Echopraxia/echolalia,Stereotypy,Withdrawal | 98.4 | 97.1 | 99.6 |
| 8 | Excitement,Immobility/stupor,Staring,Posturing/catalepsy,Grimacing,Echopraxia/echolalia,Verbigeration,Withdrawal | 98.4 | 97.3 | 99.6 |
| 8 | Excitement,Immobility/stupor,Staring,Posturing/catalepsy,Grimacing,Mannerisms,Verbigeration,Waxy flexibility | 98.4 | 97.3 | 99.6 |
| 8 | Excitement,Immobility/stupor,Staring,Posturing/catalepsy,Grimacing,Mannerisms,Waxy flexibility,Withdrawal | 98.4 | 97.3 | 99.6 |
| 8 | Excitement,Immobility/stupor,Staring,Posturing/catalepsy,Stereotypy,Mannerisms,Verbigeration,Withdrawal | 98.4 | 97.1 | 99.3 |
| 8 | Excitement,Immobility/stupor,Stereotypy,Mannerisms,Rigidity,Negativism,Waxy flexibility,Withdrawal | 98.4 | 97.1 | 99.6 |
| 8 | Excitement,Immobility/stupor,Stereotypy,Mannerisms,Verbigeration,Rigidity,Negativism,Waxy flexibility | 98.4 | 97.1 | 99.6 |
| 8 | Excitement,Mutism,Staring,Echopraxia/echolalia,Mannerisms,Verbigeration,Waxy flexibility,Withdrawal | 98.4 | 97.1 | 99.6 |
| 8 | Excitement,Mutism,Staring,Echopraxia/echolalia,Stereotypy,Mannerisms,Verbigeration,Waxy flexibility | 98.4 | 97.1 | 99.6 |
| 8 | Excitement,Mutism,Staring,Echopraxia/echolalia,Stereotypy,Mannerisms,Verbigeration,Withdrawal | 98.4 | 97.1 | 99.6 |
| 8 | Excitement,Mutism,Staring,Grimacing,Echopraxia/echolalia,Mannerisms,Verbigeration,Waxy flexibility | 98.4 | 97.1 | 99.6 |
| 8 | Excitement,Mutism,Staring,Grimacing,Mannerisms,Verbigeration,Waxy flexibility,Withdrawal | 98.4 | 97.1 | 99.6 |
| 8 | Excitement,Mutism,Staring,Grimacing,Stereotypy,Mannerisms,Waxy flexibility,Withdrawal | 98.4 | 97.1 | 99.6 |
| 8 | Excitement,Mutism,Staring,Posturing/catalepsy,Grimacing,Stereotypy,Mannerisms,Verbigeration | 98.4 | 97.1 | 99.6 |
| 8 | Excitement,Mutism,Staring,Posturing/catalepsy,Grimacing,Stereotypy,Mannerisms,Waxy flexibility | 98.4 | 97.1 | 99.6 |
| 8 | Excitement,Mutism,Staring,Stereotypy,Mannerisms,Verbigeration,Waxy flexibility,Withdrawal | 98.4 | 97.1 | 99.6 |
| 8 | Excitement,Staring,Echopraxia/echolalia,Mannerisms,Verbigeration,Negativism,Waxy flexibility,Withdrawal | 98.4 | 97.1 | 99.6 |
| 8 | Excitement,Staring,Echopraxia/echolalia,Stereotypy,Mannerisms,Rigidity,Negativism,Waxy flexibility | 98.4 | 97.3 | 99.6 |
| 8 | Excitement,Staring,Echopraxia/echolalia,Stereotypy,Mannerisms,Verbigeration,Negativism,Withdrawal | 98.4 | 97.3 | 99.6 |
| 8 | Excitement,Staring,Grimacing,Echopraxia/echolalia,Mannerisms,Negativism,Waxy flexibility,Withdrawal | 98.4 | 97.1 | 99.6 |
| 8 | Excitement,Staring,Grimacing,Stereotypy,Mannerisms,Verbigeration,Negativism,Withdrawal | 98.4 | 97.1 | 99.3 |
| 8 | Excitement,Staring,Posturing/catalepsy,Echopraxia/echolalia,Mannerisms,Verbigeration,Rigidity,Negativism | 98.4 | 97.1 | 99.6 |
| 8 | Excitement,Staring,Posturing/catalepsy,Echopraxia/echolalia,Stereotypy,Mannerisms,Rigidity,Negativism | 98.4 | 97.1 | 99.6 |
| 8 | Excitement,Staring,Posturing/catalepsy,Echopraxia/echolalia,Stereotypy,Verbigeration,Negativism,Withdrawal | 98.4 | 97.1 | 99.6 |
| 8 | Excitement,Staring,Posturing/catalepsy,Grimacing,Echopraxia/echolalia,Mannerisms,Waxy flexibility,Withdrawal | 98.4 | 97.1 | 99.6 |
| 8 | Excitement,Staring,Posturing/catalepsy,Grimacing,Verbigeration,Negativism,Waxy flexibility,Withdrawal | 98.4 | 97.1 | 99.6 |
| 8 | Excitement,Staring,Posturing/catalepsy,Stereotypy,Mannerisms,Verbigeration,Rigidity,Negativism | 98.4 | 97.1 | 99.6 |
| 8 | Excitement,Staring,Stereotypy,Mannerisms,Verbigeration,Negativism,Waxy flexibility,Withdrawal | 98.4 | 97.1 | 99.6 |
| 8 | Excitement,Staring,Stereotypy,Mannerisms,Verbigeration,Rigidity,Negativism,Waxy flexibility | 98.4 | 97.1 | 99.6 |
| 8 | Immobility/stupor,Grimacing,Echopraxia/echolalia,Mannerisms,Rigidity,Negativism,Waxy flexibility,Withdrawal | 98.4 | 97.1 | 99.6 |
| 8 | Immobility/stupor,Grimacing,Echopraxia/echolalia,Mannerisms,Verbigeration,Rigidity,Negativism,Withdrawal | 98.4 | 97.1 | 99.6 |
| 8 | Immobility/stupor,Grimacing,Stereotypy,Mannerisms,Rigidity,Negativism,Waxy flexibility,Withdrawal | 98.4 | 97.3 | 99.6 |
| 8 | Immobility/stupor,Mutism,Echopraxia/echolalia,Mannerisms,Verbigeration,Rigidity,Negativism,Withdrawal | 98.4 | 97.1 | 99.6 |
| 8 | Immobility/stupor,Mutism,Echopraxia/echolalia,Stereotypy,Mannerisms,Verbigeration,Rigidity,Negativism | 98.4 | 97.1 | 99.6 |
| 8 | Immobility/stupor,Mutism,Grimacing,Mannerisms,Verbigeration,Rigidity,Negativism,Withdrawal | 98.4 | 97.1 | 99.6 |
| 8 | Immobility/stupor,Mutism,Grimacing,Stereotypy,Mannerisms,Rigidity,Negativism,Withdrawal | 98.4 | 97.1 | 99.6 |
| 8 | Immobility/stupor,Mutism,Grimacing,Stereotypy,Mannerisms,Verbigeration,Rigidity,Negativism | 98.4 | 97.1 | 99.6 |
| 8 | Immobility/stupor,Mutism,Grimacing,Stereotypy,Verbigeration,Rigidity,Negativism,Withdrawal | 98.4 | 97.3 | 99.6 |
| 8 | Immobility/stupor,Mutism,Posturing/catalepsy,Echopraxia/echolalia,Mannerisms,Rigidity,Negativism,Withdrawal | 98.4 | 97.3 | 99.6 |
| 8 | Immobility/stupor,Mutism,Posturing/catalepsy,Echopraxia/echolalia,Mannerisms,Verbigeration,Rigidity,Negativism | 98.4 | 97.1 | 99.6 |
| 8 | Immobility/stupor,Mutism,Posturing/catalepsy,Grimacing,Echopraxia/echolalia,Mannerisms,Rigidity,Negativism | 98.4 | 97.1 | 99.6 |
| 8 | Immobility/stupor,Mutism,Posturing/catalepsy,Grimacing,Mannerisms,Rigidity,Negativism,Withdrawal | 98.4 | 97.1 | 99.6 |
| 8 | Immobility/stupor,Mutism,Posturing/catalepsy,Mannerisms,Verbigeration,Rigidity,Negativism,Withdrawal | 98.4 | 97.1 | 99.6 |
| 8 | Immobility/stupor,Mutism,Posturing/catalepsy,Stereotypy,Mannerisms,Rigidity,Negativism,Withdrawal | 98.4 | 97.1 | 99.6 |
| 8 | Immobility/stupor,Mutism,Staring,Echopraxia/echolalia,Mannerisms,Verbigeration,Negativism,Withdrawal | 98.4 | 97.1 | 99.6 |
| 8 | Immobility/stupor,Mutism,Staring,Echopraxia/echolalia,Stereotypy,Mannerisms,Verbigeration,Negativism | 98.4 | 97.1 | 99.6 |
| 8 | Immobility/stupor,Mutism,Staring,Echopraxia/echolalia,Stereotypy,Verbigeration,Rigidity,Negativism | 98.4 | 97.3 | 99.6 |
| 8 | Immobility/stupor,Mutism,Staring,Echopraxia/echolalia,Verbigeration,Rigidity,Negativism,Withdrawal | 98.4 | 97.1 | 99.6 |
| 8 | Immobility/stupor,Mutism,Staring,Grimacing,Echopraxia/echolalia,Mannerisms,Negativism,Waxy flexibility | 98.4 | 97.1 | 99.6 |
| 8 | Immobility/stupor,Mutism,Staring,Grimacing,Mannerisms,Negativism,Waxy flexibility,Withdrawal | 98.4 | 97.3 | 99.6 |
| 8 | Immobility/stupor,Mutism,Staring,Grimacing,Mannerisms,Rigidity,Negativism,Waxy flexibility | 98.4 | 97.3 | 99.6 |
| 8 | Immobility/stupor,Mutism,Staring,Grimacing,Mannerisms,Rigidity,Waxy flexibility,Withdrawal | 98.4 | 97.1 | 99.6 |
| 8 | Immobility/stupor,Mutism,Staring,Grimacing,Mannerisms,Verbigeration,Negativism,Withdrawal | 98.4 | 97.1 | 99.6 |
| 8 | Immobility/stupor,Mutism,Staring,Grimacing,Stereotypy,Mannerisms,Negativism,Waxy flexibility | 98.4 | 97.3 | 99.6 |
| 8 | Immobility/stupor,Mutism,Staring,Grimacing,Stereotypy,Mannerisms,Rigidity,Waxy flexibility | 98.4 | 97.1 | 99.6 |
| 8 | Immobility/stupor,Mutism,Staring,Grimacing,Stereotypy,Mannerisms,Verbigeration,Negativism | 98.4 | 97.1 | 99.6 |
| 8 | Immobility/stupor,Mutism,Staring,Grimacing,Stereotypy,Verbigeration,Negativism,Withdrawal | 98.4 | 97.1 | 99.6 |
| 8 | Immobility/stupor,Mutism,Staring,Grimacing,Stereotypy,Verbigeration,Rigidity,Waxy flexibility | 98.4 | 97.1 | 99.6 |
| 8 | Immobility/stupor,Mutism,Staring,Posturing/catalepsy,Echopraxia/echolalia,Mannerisms,Negativism,Waxy flexibility | 98.4 | 97.1 | 99.6 |
| 8 | Immobility/stupor,Mutism,Staring,Posturing/catalepsy,Echopraxia/echolalia,Mannerisms,Verbigeration,Rigidity | 98.4 | 97.3 | 99.6 |
| 8 | Immobility/stupor,Mutism,Staring,Posturing/catalepsy,Echopraxia/echolalia,Rigidity,Negativism,Withdrawal | 98.4 | 97.1 | 99.6 |
| 8 | Immobility/stupor,Mutism,Staring,Posturing/catalepsy,Echopraxia/echolalia,Stereotypy,Negativism,Withdrawal | 98.4 | 97.1 | 99.6 |
| 8 | Immobility/stupor,Mutism,Staring,Posturing/catalepsy,Echopraxia/echolalia,Verbigeration,Rigidity,Negativism | 98.4 | 97.3 | 99.6 |
| 8 | Immobility/stupor,Mutism,Staring,Posturing/catalepsy,Grimacing,Echopraxia/echolalia,Mannerisms,Withdrawal | 98.4 | 97.1 | 99.6 |
| 8 | Immobility/stupor,Mutism,Staring,Posturing/catalepsy,Grimacing,Echopraxia/echolalia,Negativism,Withdrawal | 98.4 | 97.1 | 99.6 |
| 8 | Immobility/stupor,Mutism,Staring,Posturing/catalepsy,Grimacing,Mannerisms,Verbigeration,Rigidity | 98.4 | 97.1 | 99.6 |
| 8 | Immobility/stupor,Mutism,Staring,Posturing/catalepsy,Grimacing,Stereotypy,Mannerisms,Rigidity | 98.4 | 97.1 | 99.6 |
| 8 | Immobility/stupor,Mutism,Staring,Posturing/catalepsy,Grimacing,Stereotypy,Mannerisms,Withdrawal | 98.4 | 97.3 | 99.6 |
| 8 | Immobility/stupor,Mutism,Staring,Posturing/catalepsy,Grimacing,Stereotypy,Negativism,Withdrawal | 98.4 | 97.1 | 99.6 |
| 8 | Immobility/stupor,Mutism,Staring,Posturing/catalepsy,Grimacing,Stereotypy,Verbigeration,Negativism | 98.4 | 97.3 | 99.6 |
| 8 | Immobility/stupor,Mutism,Staring,Posturing/catalepsy,Grimacing,Stereotypy,Verbigeration,Rigidity | 98.4 | 97.1 | 99.6 |
| 8 | Immobility/stupor,Mutism,Staring,Posturing/catalepsy,Mannerisms,Rigidity,Negativism,Waxy flexibility | 98.4 | 97.1 | 99.6 |
| 8 | Immobility/stupor,Mutism,Staring,Posturing/catalepsy,Stereotypy,Rigidity,Negativism,Withdrawal | 98.4 | 97.1 | 99.6 |
| 8 | Immobility/stupor,Mutism,Staring,Stereotypy,Mannerisms,Verbigeration,Negativism,Waxy flexibility | 98.4 | 97.1 | 99.6 |
| 8 | Immobility/stupor,Mutism,Staring,Stereotypy,Mannerisms,Verbigeration,Rigidity,Waxy flexibility | 98.4 | 97.1 | 99.6 |
| 8 | Immobility/stupor,Mutism,Stereotypy,Mannerisms,Rigidity,Negativism,Waxy flexibility,Withdrawal | 98.4 | 97.1 | 99.6 |
| 8 | Immobility/stupor,Posturing/catalepsy,Echopraxia/echolalia,Mannerisms,Rigidity,Negativism,Waxy flexibility,Withdrawal | 98.4 | 97.1 | 99.6 |
| 8 | Immobility/stupor,Posturing/catalepsy,Grimacing,Mannerisms,Rigidity,Negativism,Waxy flexibility,Withdrawal | 98.4 | 97.1 | 99.6 |
| 8 | Immobility/stupor,Posturing/catalepsy,Mannerisms,Verbigeration,Rigidity,Negativism,Waxy flexibility,Withdrawal | 98.4 | 97.1 | 99.6 |
| 8 | Immobility/stupor,Posturing/catalepsy,Stereotypy,Mannerisms,Rigidity,Negativism,Waxy flexibility,Withdrawal | 98.4 | 97.1 | 99.6 |
| 8 | Immobility/stupor,Staring,Echopraxia/echolalia,Stereotypy,Mannerisms,Verbigeration,Negativism,Waxy flexibility | 98.4 | 97.1 | 99.6 |
| 8 | Immobility/stupor,Staring,Echopraxia/echolalia,Stereotypy,Rigidity,Negativism,Waxy flexibility,Withdrawal | 98.4 | 97.1 | 99.6 |
| 8 | Immobility/stupor,Staring,Grimacing,Echopraxia/echolalia,Mannerisms,Verbigeration,Negativism,Withdrawal | 98.4 | 97.1 | 99.6 |
| 8 | Immobility/stupor,Staring,Grimacing,Echopraxia/echolalia,Stereotypy,Mannerisms,Verbigeration,Negativism | 98.4 | 97.1 | 99.6 |
| 8 | Immobility/stupor,Staring,Grimacing,Echopraxia/echolalia,Stereotypy,Verbigeration,Rigidity,Negativism | 98.4 | 97.1 | 99.6 |
| 8 | Immobility/stupor,Staring,Grimacing,Echopraxia/echolalia,Verbigeration,Rigidity,Negativism,Withdrawal | 98.4 | 97.1 | 99.6 |
| 8 | Immobility/stupor,Staring,Grimacing,Stereotypy,Mannerisms,Verbigeration,Negativism,Waxy flexibility | 98.4 | 97.1 | 99.6 |
| 8 | Immobility/stupor,Staring,Grimacing,Stereotypy,Mannerisms,Verbigeration,Rigidity,Waxy flexibility | 98.4 | 97.1 | 99.6 |
| 8 | Immobility/stupor,Staring,Posturing/catalepsy,Echopraxia/echolalia,Stereotypy,Mannerisms,Rigidity,Withdrawal | 98.4 | 97.3 | 99.6 |
| 8 | Immobility/stupor,Staring,Posturing/catalepsy,Echopraxia/echolalia,Stereotypy,Verbigeration,Rigidity,Negativism | 98.4 | 97.3 | 99.6 |
| 8 | Immobility/stupor,Staring,Posturing/catalepsy,Echopraxia/echolalia,Verbigeration,Rigidity,Negativism,Withdrawal | 98.4 | 97.1 | 99.6 |
| 8 | Immobility/stupor,Staring,Posturing/catalepsy,Grimacing,Echopraxia/echolalia,Mannerisms,Negativism,Waxy flexibility | 98.4 | 97.1 | 99.6 |
| 8 | Immobility/stupor,Staring,Posturing/catalepsy,Grimacing,Echopraxia/echolalia,Mannerisms,Rigidity,Withdrawal | 98.4 | 97.1 | 99.6 |
| 8 | Immobility/stupor,Staring,Posturing/catalepsy,Grimacing,Echopraxia/echolalia,Rigidity,Negativism,Withdrawal | 98.4 | 97.1 | 99.6 |
| 8 | Immobility/stupor,Staring,Posturing/catalepsy,Grimacing,Echopraxia/echolalia,Stereotypy,Negativism,Withdrawal | 98.4 | 97.1 | 99.6 |
| 8 | Immobility/stupor,Staring,Posturing/catalepsy,Grimacing,Mannerisms,Rigidity,Negativism,Waxy flexibility | 98.4 | 97.3 | 99.6 |
| 8 | Immobility/stupor,Staring,Posturing/catalepsy,Grimacing,Mannerisms,Verbigeration,Rigidity,Withdrawal | 98.4 | 97.1 | 99.6 |
| 8 | Immobility/stupor,Staring,Posturing/catalepsy,Grimacing,Stereotypy,Mannerisms,Negativism,Waxy flexibility | 98.4 | 97.1 | 99.6 |
| 8 | Immobility/stupor,Staring,Posturing/catalepsy,Grimacing,Stereotypy,Mannerisms,Verbigeration,Rigidity | 98.4 | 97.1 | 99.6 |
| 8 | Immobility/stupor,Staring,Posturing/catalepsy,Grimacing,Stereotypy,Rigidity,Negativism,Withdrawal | 98.4 | 97.1 | 99.6 |
| 8 | Immobility/stupor,Staring,Posturing/catalepsy,Stereotypy,Mannerisms,Verbigeration,Rigidity,Withdrawal | 98.4 | 97.1 | 99.6 |
| 8 | Immobility/stupor,Staring,Posturing/catalepsy,Stereotypy,Verbigeration,Rigidity,Negativism,Waxy flexibility | 98.4 | 97.3 | 99.6 |
| 8 | Mutism,Staring,Echopraxia/echolalia,Verbigeration,Rigidity,Negativism,Waxy flexibility,Withdrawal | 98.4 | 97.1 | 99.6 |
| 8 | Mutism,Staring,Grimacing,Echopraxia/echolalia,Stereotypy,Rigidity,Waxy flexibility,Withdrawal | 98.4 | 97.1 | 99.6 |
| 8 | Mutism,Staring,Grimacing,Echopraxia/echolalia,Stereotypy,Verbigeration,Negativism,Waxy flexibility | 98.4 | 97.3 | 99.6 |
| 8 | Mutism,Staring,Grimacing,Echopraxia/echolalia,Stereotypy,Verbigeration,Rigidity,Waxy flexibility | 98.4 | 97.1 | 99.6 |
| 8 | Mutism,Staring,Grimacing,Echopraxia/echolalia,Verbigeration,Rigidity,Negativism,Waxy flexibility | 98.4 | 97.3 | 99.6 |
| 8 | Mutism,Staring,Grimacing,Stereotypy,Rigidity,Negativism,Waxy flexibility,Withdrawal | 98.4 | 97.1 | 99.6 |
| 8 | Mutism,Staring,Grimacing,Stereotypy,Verbigeration,Negativism,Waxy flexibility,Withdrawal | 98.4 | 97.1 | 99.6 |
| 8 | Mutism,Staring,Posturing/catalepsy,Echopraxia/echolalia,Mannerisms,Verbigeration,Rigidity,Waxy flexibility | 98.4 | 97.3 | 99.6 |
| 8 | Mutism,Staring,Posturing/catalepsy,Echopraxia/echolalia,Rigidity,Negativism,Waxy flexibility,Withdrawal | 98.4 | 97.1 | 99.6 |
| 8 | Mutism,Staring,Posturing/catalepsy,Echopraxia/echolalia,Stereotypy,Negativism,Waxy flexibility,Withdrawal | 98.4 | 97.3 | 99.6 |
| 8 | Mutism,Staring,Posturing/catalepsy,Echopraxia/echolalia,Stereotypy,Verbigeration,Negativism,Waxy flexibility | 98.4 | 97.1 | 99.6 |
| 8 | Mutism,Staring,Posturing/catalepsy,Echopraxia/echolalia,Stereotypy,Verbigeration,Rigidity,Withdrawal | 98.4 | 97.1 | 99.6 |
| 8 | Mutism,Staring,Posturing/catalepsy,Echopraxia/echolalia,Verbigeration,Rigidity,Negativism,Waxy flexibility | 98.4 | 97.1 | 99.6 |
| 8 | Mutism,Staring,Posturing/catalepsy,Grimacing,Echopraxia/echolalia,Negativism,Waxy flexibility,Withdrawal | 98.4 | 97.1 | 99.6 |
| 8 | Mutism,Staring,Posturing/catalepsy,Grimacing,Echopraxia/echolalia,Stereotypy,Rigidity,Negativism | 98.4 | 97.1 | 99.6 |
| 8 | Mutism,Staring,Posturing/catalepsy,Grimacing,Echopraxia/echolalia,Stereotypy,Verbigeration,Rigidity | 98.4 | 97.1 | 99.6 |
| 8 | Mutism,Staring,Posturing/catalepsy,Grimacing,Echopraxia/echolalia,Verbigeration,Negativism,Waxy flexibility | 98.4 | 97.1 | 99.6 |
| 8 | Mutism,Staring,Posturing/catalepsy,Grimacing,Mannerisms,Rigidity,Negativism,Waxy flexibility | 98.4 | 97.3 | 99.6 |
| 8 | Mutism,Staring,Posturing/catalepsy,Grimacing,Mannerisms,Verbigeration,Waxy flexibility,Withdrawal | 98.4 | 97.1 | 99.6 |
| 8 | Mutism,Staring,Posturing/catalepsy,Grimacing,Stereotypy,Negativism,Waxy flexibility,Withdrawal | 98.4 | 97.3 | 99.6 |
| 8 | Mutism,Staring,Posturing/catalepsy,Grimacing,Stereotypy,Verbigeration,Negativism,Waxy flexibility | 98.4 | 97.3 | 99.6 |
| 8 | Mutism,Staring,Posturing/catalepsy,Grimacing,Verbigeration,Rigidity,Negativism,Withdrawal | 98.4 | 97.1 | 99.6 |
| 8 | Mutism,Staring,Posturing/catalepsy,Stereotypy,Mannerisms,Verbigeration,Rigidity,Waxy flexibility | 98.4 | 97.1 | 99.6 |
| 8 | Mutism,Staring,Posturing/catalepsy,Stereotypy,Rigidity,Negativism,Waxy flexibility,Withdrawal | 98.4 | 97.1 | 99.6 |
| 8 | Mutism,Staring,Posturing/catalepsy,Stereotypy,Verbigeration,Rigidity,Waxy flexibility,Withdrawal | 98.4 | 97.1 | 99.6 |
| 8 | Staring,Grimacing,Echopraxia/echolalia,Stereotypy,Mannerisms,Verbigeration,Rigidity,Withdrawal | 98.4 | 97.1 | 99.6 |
| 8 | Staring,Grimacing,Echopraxia/echolalia,Stereotypy,Verbigeration,Rigidity,Negativism,Withdrawal | 98.4 | 97.3 | 99.6 |
| 8 | Staring,Grimacing,Stereotypy,Mannerisms,Verbigeration,Negativism,Waxy flexibility,Withdrawal | 98.4 | 97.1 | 99.6 |
| 8 | Staring,Grimacing,Stereotypy,Verbigeration,Rigidity,Negativism,Waxy flexibility,Withdrawal | 98.4 | 97.3 | 99.6 |
| 8 | Staring,Posturing/catalepsy,Echopraxia/echolalia,Stereotypy,Mannerisms,Verbigeration,Negativism,Withdrawal | 98.4 | 97.3 | 99.6 |
| 8 | Staring,Posturing/catalepsy,Grimacing,Mannerisms,Verbigeration,Negativism,Waxy flexibility,Withdrawal | 98.4 | 97.3 | 99.6 |
| 8 | Staring,Posturing/catalepsy,Grimacing,Stereotypy,Mannerisms,Rigidity,Waxy flexibility,Withdrawal | 98.4 | 97.1 | 99.6 |
| 8 | Staring,Posturing/catalepsy,Grimacing,Stereotypy,Verbigeration,Rigidity,Negativism,Withdrawal | 98.4 | 97.3 | 99.6 |
| 8 | Excitement,Immobility/stupor,Echopraxia/echolalia,Mannerisms,Verbigeration,Rigidity,Negativism,Waxy flexibility | 98.2 | 96.9 | 99.3 |
| 8 | Excitement,Immobility/stupor,Echopraxia/echolalia,Stereotypy,Mannerisms,Rigidity,Negativism,Waxy flexibility | 98.2 | 96.9 | 99.3 |
| 8 | Excitement,Immobility/stupor,Grimacing,Echopraxia/echolalia,Stereotypy,Mannerisms,Rigidity,Negativism | 98.2 | 96.9 | 99.3 |
| 8 | Excitement,Immobility/stupor,Grimacing,Stereotypy,Mannerisms,Verbigeration,Rigidity,Negativism | 98.2 | 96.9 | 99.3 |
| 8 | Excitement,Immobility/stupor,Grimacing,Verbigeration,Rigidity,Negativism,Waxy flexibility,Withdrawal | 98.2 | 96.9 | 99.3 |
| 8 | Excitement,Immobility/stupor,Mutism,Echopraxia/echolalia,Mannerisms,Verbigeration,Rigidity,Waxy flexibility | 98.2 | 96.9 | 99.3 |
| 8 | Excitement,Immobility/stupor,Mutism,Echopraxia/echolalia,Stereotypy,Mannerisms,Verbigeration,Rigidity | 98.2 | 96.9 | 99.3 |
| 8 | Excitement,Immobility/stupor,Mutism,Grimacing,Echopraxia/echolalia,Stereotypy,Rigidity,Waxy flexibility | 98.2 | 96.9 | 99.3 |
| 8 | Excitement,Immobility/stupor,Mutism,Grimacing,Echopraxia/echolalia,Stereotypy,Verbigeration,Rigidity | 98.2 | 96.9 | 99.3 |
| 8 | Excitement,Immobility/stupor,Mutism,Grimacing,Mannerisms,Verbigeration,Rigidity,Waxy flexibility | 98.2 | 96.9 | 99.3 |
| 8 | Excitement,Immobility/stupor,Mutism,Grimacing,Stereotypy,Mannerisms,Rigidity,Negativism | 98.2 | 96.9 | 99.3 |
| 8 | Excitement,Immobility/stupor,Mutism,Grimacing,Stereotypy,Mannerisms,Rigidity,Withdrawal | 98.2 | 96.9 | 99.3 |
| 8 | Excitement,Immobility/stupor,Mutism,Posturing/catalepsy,Echopraxia/echolalia,Mannerisms,Rigidity,Waxy flexibility | 98.2 | 96.9 | 99.3 |
| 8 | Excitement,Immobility/stupor,Mutism,Posturing/catalepsy,Echopraxia/echolalia,Stereotypy,Verbigeration,Negativism | 98.2 | 96.9 | 99.3 |
| 8 | Excitement,Immobility/stupor,Mutism,Posturing/catalepsy,Echopraxia/echolalia,Verbigeration,Negativism,Waxy flexibility | 98.2 | 96.9 | 99.3 |
| 8 | Excitement,Immobility/stupor,Mutism,Posturing/catalepsy,Grimacing,Echopraxia/echolalia,Negativism,Waxy flexibility | 98.2 | 96.9 | 99.3 |
| 8 | Excitement,Immobility/stupor,Mutism,Posturing/catalepsy,Grimacing,Mannerisms,Rigidity,Waxy flexibility | 98.2 | 96.9 | 99.3 |
| 8 | Excitement,Immobility/stupor,Mutism,Posturing/catalepsy,Grimacing,Mannerisms,Verbigeration,Negativism | 98.2 | 96.9 | 99.3 |
| 8 | Excitement,Immobility/stupor,Mutism,Posturing/catalepsy,Grimacing,Negativism,Waxy flexibility,Withdrawal | 98.2 | 96.9 | 99.3 |
| 8 | Excitement,Immobility/stupor,Mutism,Posturing/catalepsy,Grimacing,Stereotypy,Mannerisms,Rigidity | 98.2 | 96.9 | 99.3 |
| 8 | Excitement,Immobility/stupor,Mutism,Posturing/catalepsy,Grimacing,Stereotypy,Rigidity,Waxy flexibility | 98.2 | 96.9 | 99.3 |
| 8 | Excitement,Immobility/stupor,Mutism,Posturing/catalepsy,Grimacing,Stereotypy,Verbigeration,Negativism | 98.2 | 96.9 | 99.3 |
| 8 | Excitement,Immobility/stupor,Mutism,Posturing/catalepsy,Grimacing,Verbigeration,Negativism,Waxy flexibility | 98.2 | 96.9 | 99.3 |
| 8 | Excitement,Immobility/stupor,Mutism,Posturing/catalepsy,Stereotypy,Mannerisms,Negativism,Withdrawal | 98.2 | 96.9 | 99.3 |
| 8 | Excitement,Immobility/stupor,Mutism,Posturing/catalepsy,Stereotypy,Negativism,Waxy flexibility,Withdrawal | 98.2 | 96.9 | 99.3 |
| 8 | Excitement,Immobility/stupor,Mutism,Posturing/catalepsy,Stereotypy,Verbigeration,Negativism,Waxy flexibility | 98.2 | 96.9 | 99.3 |
| 8 | Excitement,Immobility/stupor,Mutism,Staring,Grimacing,Echopraxia/echolalia,Mannerisms,Withdrawal | 98.2 | 96.9 | 99.3 |
| 8 | Excitement,Immobility/stupor,Mutism,Staring,Grimacing,Echopraxia/echolalia,Verbigeration,Withdrawal | 98.2 | 96.9 | 99.3 |
| 8 | Excitement,Immobility/stupor,Mutism,Staring,Grimacing,Mannerisms,Verbigeration,Waxy flexibility | 98.2 | 96.9 | 99.3 |
| 8 | Excitement,Immobility/stupor,Mutism,Staring,Grimacing,Mannerisms,Waxy flexibility,Withdrawal | 98.2 | 96.9 | 99.3 |
| 8 | Excitement,Immobility/stupor,Mutism,Staring,Grimacing,Stereotypy,Mannerisms,Negativism | 98.2 | 96.9 | 99.3 |
| 8 | Excitement,Immobility/stupor,Mutism,Staring,Grimacing,Stereotypy,Verbigeration,Withdrawal | 98.2 | 96.9 | 99.3 |
| 8 | Excitement,Immobility/stupor,Mutism,Staring,Grimacing,Verbigeration,Waxy flexibility,Withdrawal | 98.2 | 96.9 | 99.3 |
| 8 | Excitement,Immobility/stupor,Mutism,Staring,Mannerisms,Verbigeration,Waxy flexibility,Withdrawal | 98.2 | 96.9 | 99.3 |
| 8 | Excitement,Immobility/stupor,Mutism,Staring,Posturing/catalepsy,Grimacing,Stereotypy,Mannerisms | 98.2 | 96.9 | 99.3 |
| 8 | Excitement,Immobility/stupor,Mutism,Staring,Stereotypy,Mannerisms,Verbigeration,Waxy flexibility | 98.2 | 96.9 | 99.3 |
| 8 | Excitement,Immobility/stupor,Mutism,Staring,Stereotypy,Mannerisms,Waxy flexibility,Withdrawal | 98.2 | 96.9 | 99.3 |
| 8 | Excitement,Immobility/stupor,Mutism,Stereotypy,Mannerisms,Verbigeration,Rigidity,Waxy flexibility | 98.2 | 96.9 | 99.3 |
| 8 | Excitement,Immobility/stupor,Posturing/catalepsy,Echopraxia/echolalia,Mannerisms,Rigidity,Negativism,Waxy flexibility | 98.2 | 96.9 | 99.3 |
| 8 | Excitement,Immobility/stupor,Posturing/catalepsy,Echopraxia/echolalia,Stereotypy,Mannerisms,Negativism,Withdrawal | 98.2 | 96.9 | 99.3 |
| 8 | Excitement,Immobility/stupor,Posturing/catalepsy,Echopraxia/echolalia,Stereotypy,Mannerisms,Rigidity,Negativism | 98.2 | 96.9 | 99.3 |
| 8 | Excitement,Immobility/stupor,Posturing/catalepsy,Echopraxia/echolalia,Stereotypy,Rigidity,Negativism,Waxy flexibility | 98.2 | 96.9 | 99.3 |
| 8 | Excitement,Immobility/stupor,Posturing/catalepsy,Echopraxia/echolalia,Verbigeration,Negativism,Waxy flexibility,Withdrawal | 98.2 | 96.9 | 99.3 |
| 8 | Excitement,Immobility/stupor,Posturing/catalepsy,Grimacing,Echopraxia/echolalia,Negativism,Waxy flexibility,Withdrawal | 98.2 | 96.9 | 99.3 |
| 8 | Excitement,Immobility/stupor,Posturing/catalepsy,Grimacing,Echopraxia/echolalia,Stereotypy,Negativism,Withdrawal | 98.2 | 96.9 | 99.3 |
| 8 | Excitement,Immobility/stupor,Posturing/catalepsy,Grimacing,Mannerisms,Rigidity,Waxy flexibility,Withdrawal | 98.2 | 96.9 | 99.3 |
| 8 | Excitement,Immobility/stupor,Posturing/catalepsy,Grimacing,Stereotypy,Mannerisms,Rigidity,Negativism | 98.2 | 96.9 | 99.3 |
| 8 | Excitement,Immobility/stupor,Posturing/catalepsy,Grimacing,Stereotypy,Mannerisms,Rigidity,Withdrawal | 98.2 | 96.9 | 99.3 |
| 8 | Excitement,Immobility/stupor,Posturing/catalepsy,Grimacing,Stereotypy,Rigidity,Negativism,Waxy flexibility | 98.2 | 96.9 | 99.3 |
| 8 | Excitement,Immobility/stupor,Posturing/catalepsy,Grimacing,Stereotypy,Verbigeration,Negativism,Withdrawal | 98.2 | 96.9 | 99.3 |
| 8 | Excitement,Immobility/stupor,Posturing/catalepsy,Grimacing,Verbigeration,Rigidity,Waxy flexibility,Withdrawal | 98.2 | 96.9 | 99.3 |
| 8 | Excitement,Immobility/stupor,Posturing/catalepsy,Mannerisms,Verbigeration,Negativism,Waxy flexibility,Withdrawal | 98.2 | 96.9 | 99.3 |
| 8 | Excitement,Immobility/stupor,Posturing/catalepsy,Stereotypy,Verbigeration,Negativism,Waxy flexibility,Withdrawal | 98.2 | 96.9 | 99.3 |
| 8 | Excitement,Immobility/stupor,Staring,Echopraxia/echolalia,Mannerisms,Verbigeration,Waxy flexibility,Withdrawal | 98.2 | 96.9 | 99.3 |
| 8 | Excitement,Immobility/stupor,Staring,Grimacing,Echopraxia/echolalia,Mannerisms,Verbigeration,Negativism | 98.2 | 96.9 | 99.3 |
| 8 | Excitement,Immobility/stupor,Staring,Grimacing,Echopraxia/echolalia,Stereotypy,Mannerisms,Withdrawal | 98.2 | 96.9 | 99.3 |
| 8 | Excitement,Immobility/stupor,Staring,Grimacing,Echopraxia/echolalia,Stereotypy,Waxy flexibility,Withdrawal | 98.2 | 96.9 | 99.3 |
| 8 | Excitement,Immobility/stupor,Staring,Grimacing,Stereotypy,Mannerisms,Negativism,Waxy flexibility | 98.2 | 96.9 | 99.3 |
| 8 | Excitement,Immobility/stupor,Staring,Posturing/catalepsy,Echopraxia/echolalia,Stereotypy,Verbigeration,Withdrawal | 98.2 | 96.9 | 99.3 |
| 8 | Excitement,Immobility/stupor,Staring,Posturing/catalepsy,Grimacing,Echopraxia/echolalia,Mannerisms,Verbigeration | 98.2 | 96.9 | 99.3 |
| 8 | Excitement,Immobility/stupor,Staring,Posturing/catalepsy,Grimacing,Echopraxia/echolalia,Verbigeration,Waxy flexibility | 98.2 | 96.9 | 99.3 |
| 8 | Excitement,Immobility/stupor,Staring,Posturing/catalepsy,Grimacing,Stereotypy,Mannerisms,Withdrawal | 98.2 | 96.9 | 99.3 |
| 8 | Excitement,Immobility/stupor,Staring,Posturing/catalepsy,Grimacing,Stereotypy,Verbigeration,Withdrawal | 98.2 | 96.9 | 99.3 |
| 8 | Excitement,Immobility/stupor,Staring,Posturing/catalepsy,Grimacing,Verbigeration,Waxy flexibility,Withdrawal | 98.2 | 96.9 | 99.3 |
| 8 | Excitement,Immobility/stupor,Staring,Posturing/catalepsy,Stereotypy,Mannerisms,Verbigeration,Waxy flexibility | 98.2 | 96.9 | 99.3 |
| 8 | Excitement,Immobility/stupor,Staring,Posturing/catalepsy,Stereotypy,Mannerisms,Waxy flexibility,Withdrawal | 98.2 | 96.9 | 99.3 |
| 8 | Excitement,Immobility/stupor,Staring,Posturing/catalepsy,Stereotypy,Verbigeration,Waxy flexibility,Withdrawal | 98.2 | 96.9 | 99.3 |
| 8 | Excitement,Immobility/stupor,Stereotypy,Mannerisms,Verbigeration,Rigidity,Waxy flexibility,Withdrawal | 98.2 | 96.9 | 99.3 |
| 8 | Excitement,Mutism,Staring,Grimacing,Echopraxia/echolalia,Mannerisms,Verbigeration,Withdrawal | 98.2 | 96.9 | 99.3 |
| 8 | Excitement,Mutism,Staring,Grimacing,Stereotypy,Mannerisms,Verbigeration,Waxy flexibility | 98.2 | 96.9 | 99.3 |
| 8 | Excitement,Mutism,Staring,Grimacing,Stereotypy,Mannerisms,Verbigeration,Withdrawal | 98.2 | 96.9 | 99.3 |
| 8 | Excitement,Staring,Echopraxia/echolalia,Mannerisms,Verbigeration,Rigidity,Negativism,Waxy flexibility | 98.2 | 96.9 | 99.3 |
| 8 | Excitement,Staring,Echopraxia/echolalia,Stereotypy,Mannerisms,Verbigeration,Rigidity,Negativism | 98.2 | 96.9 | 99.3 |
| 8 | Excitement,Staring,Echopraxia/echolalia,Stereotypy,Verbigeration,Negativism,Waxy flexibility,Withdrawal | 98.2 | 96.9 | 99.3 |
| 8 | Excitement,Staring,Echopraxia/echolalia,Stereotypy,Verbigeration,Rigidity,Negativism,Waxy flexibility | 98.2 | 96.9 | 99.3 |
| 8 | Excitement,Staring,Grimacing,Echopraxia/echolalia,Stereotypy,Negativism,Waxy flexibility,Withdrawal | 98.2 | 96.9 | 99.3 |
| 8 | Excitement,Staring,Grimacing,Echopraxia/echolalia,Stereotypy,Verbigeration,Negativism,Withdrawal | 98.2 | 96.9 | 99.3 |
| 8 | Excitement,Staring,Grimacing,Mannerisms,Verbigeration,Negativism,Waxy flexibility,Withdrawal | 98.2 | 96.9 | 99.3 |
| 8 | Excitement,Staring,Posturing/catalepsy,Echopraxia/echolalia,Mannerisms,Rigidity,Negativism,Waxy flexibility | 98.2 | 96.9 | 99.3 |
| 8 | Excitement,Staring,Posturing/catalepsy,Echopraxia/echolalia,Stereotypy,Verbigeration,Rigidity,Negativism | 98.2 | 96.9 | 99.3 |
| 8 | Excitement,Staring,Posturing/catalepsy,Grimacing,Echopraxia/echolalia,Mannerisms,Rigidity,Negativism | 98.2 | 96.9 | 99.3 |
| 8 | Excitement,Staring,Posturing/catalepsy,Grimacing,Echopraxia/echolalia,Mannerisms,Verbigeration,Withdrawal | 98.2 | 96.9 | 99.3 |
| 8 | Excitement,Staring,Posturing/catalepsy,Grimacing,Echopraxia/echolalia,Stereotypy,Mannerisms,Withdrawal | 98.2 | 96.9 | 99.3 |
| 8 | Excitement,Staring,Posturing/catalepsy,Grimacing,Mannerisms,Verbigeration,Rigidity,Negativism | 98.2 | 96.9 | 99.3 |
| 8 | Excitement,Staring,Posturing/catalepsy,Grimacing,Mannerisms,Verbigeration,Waxy flexibility,Withdrawal | 98.2 | 96.9 | 99.3 |
| 8 | Excitement,Staring,Posturing/catalepsy,Grimacing,Stereotypy,Negativism,Waxy flexibility,Withdrawal | 98.2 | 96.9 | 99.3 |
| 8 | Excitement,Staring,Posturing/catalepsy,Mannerisms,Verbigeration,Rigidity,Negativism,Waxy flexibility | 98.2 | 96.9 | 99.3 |
| 8 | Immobility/stupor,Echopraxia/echolalia,Mannerisms,Verbigeration,Rigidity,Negativism,Waxy flexibility,Withdrawal | 98.2 | 96.9 | 99.3 |
| 8 | Immobility/stupor,Echopraxia/echolalia,Stereotypy,Mannerisms,Verbigeration,Rigidity,Negativism,Waxy flexibility | 98.2 | 96.9 | 99.3 |
| 8 | Immobility/stupor,Grimacing,Echopraxia/echolalia,Stereotypy,Mannerisms,Rigidity,Negativism,Waxy flexibility | 98.2 | 96.9 | 99.3 |
| 8 | Immobility/stupor,Grimacing,Echopraxia/echolalia,Stereotypy,Verbigeration,Rigidity,Negativism,Withdrawal | 98.2 | 96.9 | 99.3 |
| 8 | Immobility/stupor,Grimacing,Stereotypy,Mannerisms,Verbigeration,Rigidity,Negativism,Waxy flexibility | 98.2 | 96.9 | 99.3 |
| 8 | Immobility/stupor,Mutism,Echopraxia/echolalia,Mannerisms,Rigidity,Negativism,Waxy flexibility,Withdrawal | 98.2 | 96.9 | 99.3 |
| 8 | Immobility/stupor,Mutism,Echopraxia/echolalia,Mannerisms,Verbigeration,Rigidity,Negativism,Waxy flexibility | 98.2 | 96.9 | 99.3 |
| 8 | Immobility/stupor,Mutism,Echopraxia/echolalia,Stereotypy,Mannerisms,Rigidity,Negativism,Waxy flexibility | 98.2 | 96.9 | 99.3 |
| 8 | Immobility/stupor,Mutism,Echopraxia/echolalia,Stereotypy,Verbigeration,Rigidity,Negativism,Withdrawal | 98.2 | 96.9 | 99.3 |
| 8 | Immobility/stupor,Mutism,Grimacing,Echopraxia/echolalia,Mannerisms,Rigidity,Negativism,Waxy flexibility | 98.2 | 96.9 | 99.3 |
| 8 | Immobility/stupor,Mutism,Grimacing,Echopraxia/echolalia,Mannerisms,Verbigeration,Rigidity,Negativism | 98.2 | 96.9 | 99.3 |
| 8 | Immobility/stupor,Mutism,Grimacing,Echopraxia/echolalia,Stereotypy,Mannerisms,Rigidity,Negativism | 98.2 | 96.9 | 99.3 |
| 8 | Immobility/stupor,Mutism,Grimacing,Mannerisms,Rigidity,Negativism,Waxy flexibility,Withdrawal | 98.2 | 96.9 | 99.3 |
| 8 | Immobility/stupor,Mutism,Grimacing,Stereotypy,Mannerisms,Rigidity,Negativism,Waxy flexibility | 98.2 | 96.9 | 99.3 |
| 8 | Immobility/stupor,Mutism,Mannerisms,Verbigeration,Rigidity,Negativism,Waxy flexibility,Withdrawal | 98.2 | 96.9 | 99.3 |
| 8 | Immobility/stupor,Mutism,Posturing/catalepsy,Echopraxia/echolalia,Stereotypy,Mannerisms,Rigidity,Negativism | 98.2 | 96.9 | 99.3 |
| 8 | Immobility/stupor,Mutism,Posturing/catalepsy,Grimacing,Echopraxia/echolalia,Mannerisms,Negativism,Withdrawal | 98.2 | 96.9 | 99.3 |
| 8 | Immobility/stupor,Mutism,Posturing/catalepsy,Grimacing,Mannerisms,Verbigeration,Rigidity,Negativism | 98.2 | 96.9 | 99.3 |
| 8 | Immobility/stupor,Mutism,Posturing/catalepsy,Grimacing,Stereotypy,Mannerisms,Rigidity,Negativism | 98.2 | 96.9 | 99.3 |
| 8 | Immobility/stupor,Mutism,Posturing/catalepsy,Grimacing,Stereotypy,Verbigeration,Rigidity,Negativism | 98.2 | 96.9 | 99.3 |
| 8 | Immobility/stupor,Mutism,Posturing/catalepsy,Stereotypy,Verbigeration,Rigidity,Negativism,Withdrawal | 98.2 | 96.9 | 99.3 |
| 8 | Immobility/stupor,Mutism,Staring,Echopraxia/echolalia,Mannerisms,Rigidity,Waxy flexibility,Withdrawal | 98.2 | 96.9 | 99.3 |
| 8 | Immobility/stupor,Mutism,Staring,Echopraxia/echolalia,Mannerisms,Verbigeration,Negativism,Waxy flexibility | 98.2 | 96.9 | 99.3 |
| 8 | Immobility/stupor,Mutism,Staring,Echopraxia/echolalia,Mannerisms,Verbigeration,Rigidity,Withdrawal | 98.2 | 96.9 | 99.3 |
| 8 | Immobility/stupor,Mutism,Staring,Grimacing,Echopraxia/echolalia,Mannerisms,Verbigeration,Negativism | 98.2 | 96.9 | 99.3 |
| 8 | Immobility/stupor,Mutism,Staring,Grimacing,Echopraxia/echolalia,Mannerisms,Verbigeration,Rigidity | 98.2 | 96.9 | 99.3 |
| 8 | Immobility/stupor,Mutism,Staring,Grimacing,Echopraxia/echolalia,Rigidity,Negativism,Withdrawal | 98.2 | 96.9 | 99.3 |
| 8 | Immobility/stupor,Mutism,Staring,Grimacing,Echopraxia/echolalia,Stereotypy,Mannerisms,Rigidity | 98.2 | 96.9 | 99.3 |
| 8 | Immobility/stupor,Mutism,Staring,Grimacing,Echopraxia/echolalia,Stereotypy,Mannerisms,Withdrawal | 98.2 | 96.9 | 99.3 |
| 8 | Immobility/stupor,Mutism,Staring,Grimacing,Echopraxia/echolalia,Stereotypy,Negativism,Withdrawal | 98.2 | 96.9 | 99.3 |
| 8 | Immobility/stupor,Mutism,Staring,Grimacing,Echopraxia/echolalia,Stereotypy,Rigidity,Withdrawal | 98.2 | 96.9 | 99.3 |
| 8 | Immobility/stupor,Mutism,Staring,Grimacing,Echopraxia/echolalia,Verbigeration,Rigidity,Negativism | 98.2 | 96.9 | 99.3 |
| 8 | Immobility/stupor,Mutism,Staring,Grimacing,Mannerisms,Verbigeration,Rigidity,Waxy flexibility | 98.2 | 96.9 | 99.3 |
| 8 | Immobility/stupor,Mutism,Staring,Grimacing,Stereotypy,Rigidity,Negativism,Withdrawal | 98.2 | 96.9 | 99.3 |
| 8 | Immobility/stupor,Mutism,Staring,Grimacing,Verbigeration,Rigidity,Negativism,Withdrawal | 98.2 | 96.9 | 99.3 |
| 8 | Immobility/stupor,Mutism,Staring,Mannerisms,Verbigeration,Negativism,Waxy flexibility,Withdrawal | 98.2 | 96.9 | 99.3 |
| 8 | Immobility/stupor,Mutism,Staring,Mannerisms,Verbigeration,Rigidity,Waxy flexibility,Withdrawal | 98.2 | 96.9 | 99.3 |
| 8 | Immobility/stupor,Mutism,Staring,Posturing/catalepsy,Echopraxia/echolalia,Stereotypy,Mannerisms,Rigidity | 98.2 | 96.9 | 99.3 |
| 8 | Immobility/stupor,Mutism,Staring,Posturing/catalepsy,Echopraxia/echolalia,Stereotypy,Rigidity,Negativism | 98.2 | 96.9 | 99.3 |
| 8 | Immobility/stupor,Mutism,Staring,Posturing/catalepsy,Echopraxia/echolalia,Stereotypy,Rigidity,Withdrawal | 98.2 | 96.9 | 99.3 |
| 8 | Immobility/stupor,Mutism,Staring,Posturing/catalepsy,Echopraxia/echolalia,Stereotypy,Verbigeration,Negativism | 98.2 | 96.9 | 99.3 |
| 8 | Immobility/stupor,Mutism,Staring,Posturing/catalepsy,Echopraxia/echolalia,Verbigeration,Negativism,Withdrawal | 98.2 | 96.9 | 99.3 |
| 8 | Immobility/stupor,Mutism,Staring,Posturing/catalepsy,Grimacing,Echopraxia/echolalia,Mannerisms,Rigidity | 98.2 | 96.9 | 99.3 |
| 8 | Immobility/stupor,Mutism,Staring,Posturing/catalepsy,Grimacing,Echopraxia/echolalia,Verbigeration,Negativism | 98.2 | 96.9 | 99.3 |
| 8 | Immobility/stupor,Mutism,Staring,Posturing/catalepsy,Grimacing,Mannerisms,Negativism,Waxy flexibility | 98.2 | 96.9 | 99.3 |
| 8 | Immobility/stupor,Mutism,Staring,Posturing/catalepsy,Grimacing,Mannerisms,Verbigeration,Withdrawal | 98.2 | 96.9 | 99.3 |
| 8 | Immobility/stupor,Mutism,Staring,Posturing/catalepsy,Grimacing,Verbigeration,Negativism,Withdrawal | 98.2 | 96.9 | 99.3 |
| 8 | Immobility/stupor,Mutism,Staring,Posturing/catalepsy,Mannerisms,Rigidity,Waxy flexibility,Withdrawal | 98.2 | 96.9 | 99.3 |
| 8 | Immobility/stupor,Mutism,Staring,Posturing/catalepsy,Mannerisms,Verbigeration,Negativism,Waxy flexibility | 98.2 | 96.9 | 99.3 |
| 8 | Immobility/stupor,Mutism,Staring,Posturing/catalepsy,Stereotypy,Verbigeration,Negativism,Waxy flexibility | 98.2 | 96.9 | 99.3 |
| 8 | Immobility/stupor,Mutism,Staring,Posturing/catalepsy,Stereotypy,Verbigeration,Rigidity,Withdrawal | 98.2 | 96.9 | 99.3 |
| 8 | Immobility/stupor,Mutism,Staring,Posturing/catalepsy,Verbigeration,Rigidity,Negativism,Withdrawal | 98.2 | 96.9 | 99.3 |
| 8 | Immobility/stupor,Mutism,Staring,Stereotypy,Rigidity,Negativism,Waxy flexibility,Withdrawal | 98.2 | 96.9 | 99.3 |
| 8 | Immobility/stupor,Mutism,Staring,Stereotypy,Verbigeration,Rigidity,Waxy flexibility,Withdrawal | 98.2 | 96.9 | 99.3 |
| 8 | Immobility/stupor,Mutism,Stereotypy,Verbigeration,Rigidity,Negativism,Waxy flexibility,Withdrawal | 98.2 | 96.9 | 99.3 |
| 8 | Immobility/stupor,Posturing/catalepsy,Grimacing,Echopraxia/echolalia,Mannerisms,Rigidity,Negativism,Waxy flexibility | 98.2 | 96.9 | 99.3 |
| 8 | Immobility/stupor,Posturing/catalepsy,Grimacing,Echopraxia/echolalia,Mannerisms,Verbigeration,Rigidity,Negativism | 98.2 | 96.9 | 99.3 |
| 8 | Immobility/stupor,Posturing/catalepsy,Grimacing,Stereotypy,Mannerisms,Verbigeration,Rigidity,Negativism | 98.2 | 96.9 | 99.3 |
| 8 | Immobility/stupor,Posturing/catalepsy,Grimacing,Stereotypy,Verbigeration,Rigidity,Negativism,Withdrawal | 98.2 | 96.9 | 99.3 |
| 8 | Immobility/stupor,Posturing/catalepsy,Stereotypy,Mannerisms,Verbigeration,Rigidity,Negativism,Waxy flexibility | 98.2 | 96.9 | 99.3 |
| 8 | Immobility/stupor,Staring,Echopraxia/echolalia,Mannerisms,Verbigeration,Negativism,Waxy flexibility,Withdrawal | 98.2 | 96.9 | 99.3 |
| 8 | Immobility/stupor,Staring,Echopraxia/echolalia,Stereotypy,Mannerisms,Rigidity,Waxy flexibility,Withdrawal | 98.2 | 96.9 | 99.3 |
| 8 | Immobility/stupor,Staring,Echopraxia/echolalia,Stereotypy,Verbigeration,Rigidity,Negativism,Waxy flexibility | 98.2 | 96.9 | 99.3 |
| 8 | Immobility/stupor,Staring,Echopraxia/echolalia,Verbigeration,Rigidity,Negativism,Waxy flexibility,Withdrawal | 98.2 | 96.9 | 99.3 |
| 8 | Immobility/stupor,Staring,Grimacing,Echopraxia/echolalia,Mannerisms,Rigidity,Waxy flexibility,Withdrawal | 98.2 | 96.9 | 99.3 |
| 8 | Immobility/stupor,Staring,Grimacing,Echopraxia/echolalia,Mannerisms,Verbigeration,Negativism,Waxy flexibility | 98.2 | 96.9 | 99.3 |
| 8 | Immobility/stupor,Staring,Grimacing,Echopraxia/echolalia,Mannerisms,Verbigeration,Rigidity,Withdrawal | 98.2 | 96.9 | 99.3 |
| 8 | Immobility/stupor,Staring,Grimacing,Echopraxia/echolalia,Stereotypy,Verbigeration,Negativism,Withdrawal | 98.2 | 96.9 | 99.3 |
| 8 | Immobility/stupor,Staring,Grimacing,Mannerisms,Verbigeration,Negativism,Waxy flexibility,Withdrawal | 98.2 | 96.9 | 99.3 |
| 8 | Immobility/stupor,Staring,Grimacing,Mannerisms,Verbigeration,Rigidity,Waxy flexibility,Withdrawal | 98.2 | 96.9 | 99.3 |
| 8 | Immobility/stupor,Staring,Grimacing,Stereotypy,Rigidity,Negativism,Waxy flexibility,Withdrawal | 98.2 | 96.9 | 99.3 |
| 8 | Immobility/stupor,Staring,Grimacing,Stereotypy,Verbigeration,Rigidity,Waxy flexibility,Withdrawal | 98.2 | 96.9 | 99.3 |
| 8 | Immobility/stupor,Staring,Posturing/catalepsy,Echopraxia/echolalia,Mannerisms,Verbigeration,Negativism,Waxy flexibility | 98.2 | 96.9 | 99.3 |
| 8 | Immobility/stupor,Staring,Posturing/catalepsy,Echopraxia/echolalia,Rigidity,Negativism,Waxy flexibility,Withdrawal | 98.2 | 96.9 | 99.3 |
| 8 | Immobility/stupor,Staring,Posturing/catalepsy,Echopraxia/echolalia,Stereotypy,Verbigeration,Negativism,Withdrawal | 98.2 | 96.9 | 99.3 |
| 8 | Immobility/stupor,Staring,Posturing/catalepsy,Grimacing,Echopraxia/echolalia,Stereotypy,Mannerisms,Withdrawal | 98.2 | 96.9 | 99.3 |
| 8 | Immobility/stupor,Staring,Posturing/catalepsy,Grimacing,Echopraxia/echolalia,Verbigeration,Rigidity,Negativism | 98.2 | 96.9 | 99.3 |
| 8 | Immobility/stupor,Staring,Posturing/catalepsy,Grimacing,Mannerisms,Verbigeration,Negativism,Waxy flexibility | 98.2 | 96.9 | 99.3 |
| 8 | Immobility/stupor,Staring,Posturing/catalepsy,Grimacing,Stereotypy,Verbigeration,Rigidity,Withdrawal | 98.2 | 96.9 | 99.3 |
| 8 | Immobility/stupor,Staring,Posturing/catalepsy,Grimacing,Verbigeration,Rigidity,Negativism,Withdrawal | 98.2 | 96.9 | 99.3 |
| 8 | Immobility/stupor,Staring,Posturing/catalepsy,Stereotypy,Mannerisms,Rigidity,Waxy flexibility,Withdrawal | 98.2 | 96.9 | 99.3 |
| 8 | Immobility/stupor,Staring,Posturing/catalepsy,Stereotypy,Rigidity,Negativism,Waxy flexibility,Withdrawal | 98.2 | 96.9 | 99.3 |
| 8 | Immobility/stupor,Staring,Posturing/catalepsy,Stereotypy,Verbigeration,Negativism,Waxy flexibility,Withdrawal | 98.2 | 96.9 | 99.3 |
| 8 | Immobility/stupor,Staring,Stereotypy,Mannerisms,Verbigeration,Rigidity,Waxy flexibility,Withdrawal | 98.2 | 96.9 | 99.3 |
| 8 | Mutism,Staring,Echopraxia/echolalia,Mannerisms,Verbigeration,Rigidity,Waxy flexibility,Withdrawal | 98.2 | 96.9 | 99.3 |
| 8 | Mutism,Staring,Echopraxia/echolalia,Stereotypy,Mannerisms,Verbigeration,Rigidity,Waxy flexibility | 98.2 | 96.9 | 99.3 |
| 8 | Mutism,Staring,Echopraxia/echolalia,Stereotypy,Verbigeration,Negativism,Waxy flexibility,Withdrawal | 98.2 | 96.9 | 99.3 |
| 8 | Mutism,Staring,Echopraxia/echolalia,Stereotypy,Verbigeration,Rigidity,Waxy flexibility,Withdrawal | 98.2 | 96.9 | 99.3 |
| 8 | Mutism,Staring,Grimacing,Echopraxia/echolalia,Mannerisms,Verbigeration,Waxy flexibility,Withdrawal | 98.2 | 96.9 | 99.3 |
| 8 | Mutism,Staring,Grimacing,Echopraxia/echolalia,Rigidity,Negativism,Waxy flexibility,Withdrawal | 98.2 | 96.9 | 99.3 |
| 8 | Mutism,Staring,Grimacing,Echopraxia/echolalia,Stereotypy,Mannerisms,Verbigeration,Waxy flexibility | 98.2 | 96.9 | 99.3 |
| 8 | Mutism,Staring,Grimacing,Echopraxia/echolalia,Stereotypy,Mannerisms,Verbigeration,Withdrawal | 98.2 | 96.9 | 99.3 |
| 8 | Mutism,Staring,Grimacing,Echopraxia/echolalia,Stereotypy,Negativism,Waxy flexibility,Withdrawal | 98.2 | 96.9 | 99.3 |
| 8 | Mutism,Staring,Grimacing,Echopraxia/echolalia,Stereotypy,Rigidity,Negativism,Waxy flexibility | 98.2 | 96.9 | 99.3 |
| 8 | Mutism,Staring,Grimacing,Stereotypy,Mannerisms,Verbigeration,Waxy flexibility,Withdrawal | 98.2 | 96.9 | 99.3 |
| 8 | Mutism,Staring,Grimacing,Verbigeration,Rigidity,Negativism,Waxy flexibility,Withdrawal | 98.2 | 96.9 | 99.3 |
| 8 | Mutism,Staring,Posturing/catalepsy,Echopraxia/echolalia,Stereotypy,Mannerisms,Rigidity,Waxy flexibility | 98.2 | 96.9 | 99.3 |
| 8 | Mutism,Staring,Posturing/catalepsy,Echopraxia/echolalia,Stereotypy,Mannerisms,Waxy flexibility,Withdrawal | 98.2 | 96.9 | 99.3 |
| 8 | Mutism,Staring,Posturing/catalepsy,Echopraxia/echolalia,Stereotypy,Rigidity,Negativism,Waxy flexibility | 98.2 | 96.9 | 99.3 |
| 8 | Mutism,Staring,Posturing/catalepsy,Echopraxia/echolalia,Stereotypy,Rigidity,Waxy flexibility,Withdrawal | 98.2 | 96.9 | 99.3 |
| 8 | Mutism,Staring,Posturing/catalepsy,Echopraxia/echolalia,Verbigeration,Negativism,Waxy flexibility,Withdrawal | 98.2 | 96.9 | 99.3 |
| 8 | Mutism,Staring,Posturing/catalepsy,Grimacing,Echopraxia/echolalia,Mannerisms,Rigidity,Waxy flexibility | 98.2 | 96.9 | 99.3 |
| 8 | Mutism,Staring,Posturing/catalepsy,Grimacing,Echopraxia/echolalia,Mannerisms,Verbigeration,Waxy flexibility | 98.2 | 96.9 | 99.3 |
| 8 | Mutism,Staring,Posturing/catalepsy,Grimacing,Echopraxia/echolalia,Stereotypy,Mannerisms,Verbigeration | 98.2 | 96.9 | 99.3 |
| 8 | Mutism,Staring,Posturing/catalepsy,Grimacing,Echopraxia/echolalia,Stereotypy,Mannerisms,Waxy flexibility | 98.2 | 96.9 | 99.3 |
| 8 | Mutism,Staring,Posturing/catalepsy,Grimacing,Echopraxia/echolalia,Stereotypy,Negativism,Waxy flexibility | 98.2 | 96.9 | 99.3 |
| 8 | Mutism,Staring,Posturing/catalepsy,Grimacing,Echopraxia/echolalia,Verbigeration,Rigidity,Withdrawal | 98.2 | 96.9 | 99.3 |
| 8 | Mutism,Staring,Posturing/catalepsy,Grimacing,Mannerisms,Verbigeration,Rigidity,Waxy flexibility | 98.2 | 96.9 | 99.3 |
| 8 | Mutism,Staring,Posturing/catalepsy,Grimacing,Stereotypy,Mannerisms,Rigidity,Waxy flexibility | 98.2 | 96.9 | 99.3 |
| 8 | Mutism,Staring,Posturing/catalepsy,Grimacing,Stereotypy,Verbigeration,Rigidity,Waxy flexibility | 98.2 | 96.9 | 99.3 |
| 8 | Mutism,Staring,Posturing/catalepsy,Grimacing,Verbigeration,Negativism,Waxy flexibility,Withdrawal | 98.2 | 96.9 | 99.3 |
| 8 | Mutism,Staring,Posturing/catalepsy,Verbigeration,Rigidity,Negativism,Waxy flexibility,Withdrawal | 98.2 | 96.9 | 99.3 |
| 8 | Staring,Echopraxia/echolalia,Stereotypy,Mannerisms,Verbigeration,Negativism,Waxy flexibility,Withdrawal | 98.2 | 96.9 | 99.3 |
| 8 | Staring,Echopraxia/echolalia,Stereotypy,Verbigeration,Rigidity,Negativism,Waxy flexibility,Withdrawal | 98.2 | 96.9 | 99.3 |
| 8 | Staring,Posturing/catalepsy,Echopraxia/echolalia,Mannerisms,Verbigeration,Negativism,Waxy flexibility,Withdrawal | 98.2 | 96.9 | 99.3 |
| 8 | Staring,Posturing/catalepsy,Echopraxia/echolalia,Stereotypy,Verbigeration,Rigidity,Negativism,Withdrawal | 98.2 | 96.9 | 99.3 |
| 8 | Staring,Posturing/catalepsy,Grimacing,Echopraxia/echolalia,Mannerisms,Verbigeration,Rigidity,Withdrawal | 98.2 | 96.9 | 99.3 |
| 8 | Staring,Posturing/catalepsy,Grimacing,Echopraxia/echolalia,Stereotypy,Rigidity,Negativism,Withdrawal | 98.2 | 96.9 | 99.3 |
| 8 | Staring,Posturing/catalepsy,Stereotypy,Verbigeration,Rigidity,Negativism,Waxy flexibility,Withdrawal | 98.2 | 96.9 | 99.3 |
| 8 | Excitement,Immobility/stupor,Echopraxia/echolalia,Stereotypy,Mannerisms,Verbigeration,Negativism,Withdrawal | 98.0 | 96.6 | 99.1 |
| 8 | Excitement,Immobility/stupor,Grimacing,Mannerisms,Verbigeration,Rigidity,Waxy flexibility,Withdrawal | 98.0 | 96.6 | 99.1 |
| 8 | Excitement,Immobility/stupor,Grimacing,Stereotypy,Mannerisms,Rigidity,Negativism,Waxy flexibility | 98.0 | 96.6 | 99.1 |
| 8 | Excitement,Immobility/stupor,Grimacing,Stereotypy,Mannerisms,Verbigeration,Rigidity,Withdrawal | 98.0 | 96.6 | 99.1 |
| 8 | Excitement,Immobility/stupor,Grimacing,Stereotypy,Verbigeration,Rigidity,Waxy flexibility,Withdrawal | 98.0 | 96.6 | 99.1 |
| 8 | Excitement,Immobility/stupor,Mutism,Echopraxia/echolalia,Stereotypy,Mannerisms,Rigidity,Waxy flexibility | 98.0 | 96.6 | 99.1 |
| 8 | Excitement,Immobility/stupor,Mutism,Echopraxia/echolalia,Stereotypy,Negativism,Waxy flexibility,Withdrawal | 98.0 | 96.6 | 99.1 |
| 8 | Excitement,Immobility/stupor,Mutism,Grimacing,Echopraxia/echolalia,Mannerisms,Rigidity,Waxy flexibility | 98.0 | 96.6 | 99.1 |
| 8 | Excitement,Immobility/stupor,Mutism,Grimacing,Echopraxia/echolalia,Mannerisms,Verbigeration,Rigidity | 98.0 | 96.6 | 99.1 |
| 8 | Excitement,Immobility/stupor,Mutism,Grimacing,Echopraxia/echolalia,Stereotypy,Negativism,Withdrawal | 98.0 | 96.6 | 99.1 |
| 8 | Excitement,Immobility/stupor,Mutism,Grimacing,Stereotypy,Mannerisms,Verbigeration,Rigidity | 98.0 | 96.6 | 99.1 |
| 8 | Excitement,Immobility/stupor,Mutism,Grimacing,Stereotypy,Verbigeration,Negativism,Withdrawal | 98.0 | 96.6 | 99.1 |
| 8 | Excitement,Immobility/stupor,Mutism,Posturing/catalepsy,Echopraxia/echolalia,Mannerisms,Verbigeration,Negativism | 98.0 | 96.6 | 99.1 |
| 8 | Excitement,Immobility/stupor,Mutism,Posturing/catalepsy,Grimacing,Echopraxia/echolalia,Mannerisms,Negativism | 98.0 | 96.6 | 99.1 |
| 8 | Excitement,Immobility/stupor,Mutism,Posturing/catalepsy,Grimacing,Echopraxia/echolalia,Stereotypy,Negativism | 98.0 | 96.6 | 99.1 |
| 8 | Excitement,Immobility/stupor,Mutism,Posturing/catalepsy,Grimacing,Echopraxia/echolalia,Verbigeration,Withdrawal | 98.0 | 96.6 | 99.1 |
| 8 | Excitement,Immobility/stupor,Mutism,Posturing/catalepsy,Grimacing,Echopraxia/echolalia,Waxy flexibility,Withdrawal | 98.0 | 96.6 | 99.1 |
| 8 | Excitement,Immobility/stupor,Mutism,Posturing/catalepsy,Mannerisms,Negativism,Waxy flexibility,Withdrawal | 98.0 | 96.6 | 99.1 |
| 8 | Excitement,Immobility/stupor,Mutism,Posturing/catalepsy,Mannerisms,Verbigeration,Negativism,Waxy flexibility | 98.0 | 96.6 | 99.1 |
| 8 | Excitement,Immobility/stupor,Mutism,Posturing/catalepsy,Stereotypy,Mannerisms,Verbigeration,Negativism | 98.0 | 96.6 | 99.1 |
| 8 | Excitement,Immobility/stupor,Mutism,Staring,Echopraxia/echolalia,Mannerisms,Verbigeration,Withdrawal | 98.0 | 96.6 | 99.1 |
| 8 | Excitement,Immobility/stupor,Mutism,Staring,Grimacing,Echopraxia/echolalia,Stereotypy,Verbigeration | 98.0 | 96.6 | 99.1 |
| 8 | Excitement,Immobility/stupor,Mutism,Staring,Grimacing,Stereotypy,Mannerisms,Waxy flexibility | 98.0 | 96.6 | 99.1 |
| 8 | Excitement,Immobility/stupor,Mutism,Staring,Stereotypy,Mannerisms,Verbigeration,Withdrawal | 98.0 | 96.6 | 99.1 |
| 8 | Excitement,Immobility/stupor,Mutism,Stereotypy,Mannerisms,Verbigeration,Negativism,Withdrawal | 98.0 | 96.6 | 99.1 |
| 8 | Excitement,Immobility/stupor,Mutism,Stereotypy,Verbigeration,Negativism,Waxy flexibility,Withdrawal | 98.0 | 96.6 | 99.1 |
| 8 | Excitement,Immobility/stupor,Posturing/catalepsy,Echopraxia/echolalia,Mannerisms,Negativism,Waxy flexibility,Withdrawal | 98.0 | 96.6 | 99.1 |
| 8 | Excitement,Immobility/stupor,Posturing/catalepsy,Echopraxia/echolalia,Stereotypy,Negativism,Waxy flexibility,Withdrawal | 98.0 | 96.6 | 99.1 |
| 8 | Excitement,Immobility/stupor,Posturing/catalepsy,Grimacing,Echopraxia/echolalia,Mannerisms,Verbigeration,Rigidity | 98.0 | 96.6 | 99.1 |
| 8 | Excitement,Immobility/stupor,Posturing/catalepsy,Grimacing,Echopraxia/echolalia,Verbigeration,Rigidity,Waxy flexibility | 98.0 | 96.6 | 99.1 |
| 8 | Excitement,Immobility/stupor,Posturing/catalepsy,Grimacing,Stereotypy,Rigidity,Waxy flexibility,Withdrawal | 98.0 | 96.6 | 99.1 |
| 8 | Excitement,Immobility/stupor,Posturing/catalepsy,Grimacing,Verbigeration,Negativism,Waxy flexibility,Withdrawal | 98.0 | 96.6 | 99.1 |
| 8 | Excitement,Immobility/stupor,Posturing/catalepsy,Stereotypy,Mannerisms,Rigidity,Negativism,Waxy flexibility | 98.0 | 96.6 | 99.1 |
| 8 | Excitement,Immobility/stupor,Posturing/catalepsy,Stereotypy,Mannerisms,Rigidity,Waxy flexibility,Withdrawal | 98.0 | 96.6 | 99.1 |
| 8 | Excitement,Immobility/stupor,Staring,Echopraxia/echolalia,Stereotypy,Mannerisms,Verbigeration,Waxy flexibility | 98.0 | 96.6 | 99.1 |
| 8 | Excitement,Immobility/stupor,Staring,Echopraxia/echolalia,Stereotypy,Mannerisms,Verbigeration,Withdrawal | 98.0 | 96.6 | 99.1 |
| 8 | Excitement,Immobility/stupor,Staring,Echopraxia/echolalia,Stereotypy,Verbigeration,Waxy flexibility,Withdrawal | 98.0 | 96.6 | 99.1 |
| 8 | Excitement,Immobility/stupor,Staring,Grimacing,Echopraxia/echolalia,Mannerisms,Verbigeration,Waxy flexibility | 98.0 | 96.6 | 99.1 |
| 8 | Excitement,Immobility/stupor,Staring,Grimacing,Echopraxia/echolalia,Stereotypy,Mannerisms,Waxy flexibility | 98.0 | 96.6 | 99.1 |
| 8 | Excitement,Immobility/stupor,Staring,Grimacing,Echopraxia/echolalia,Verbigeration,Waxy flexibility,Withdrawal | 98.0 | 96.6 | 99.1 |
| 8 | Excitement,Immobility/stupor,Staring,Grimacing,Mannerisms,Verbigeration,Waxy flexibility,Withdrawal | 98.0 | 96.6 | 99.1 |
| 8 | Excitement,Immobility/stupor,Staring,Posturing/catalepsy,Echopraxia/echolalia,Stereotypy,Mannerisms,Verbigeration | 98.0 | 96.6 | 99.1 |
| 8 | Excitement,Immobility/stupor,Staring,Posturing/catalepsy,Echopraxia/echolalia,Stereotypy,Mannerisms,Waxy flexibility | 98.0 | 96.6 | 99.1 |
| 8 | Excitement,Immobility/stupor,Staring,Posturing/catalepsy,Echopraxia/echolalia,Stereotypy,Verbigeration,Waxy flexibility | 98.0 | 96.6 | 99.1 |
| 8 | Excitement,Immobility/stupor,Staring,Posturing/catalepsy,Grimacing,Echopraxia/echolalia,Stereotypy,Mannerisms | 98.0 | 96.6 | 99.1 |
| 8 | Excitement,Immobility/stupor,Staring,Posturing/catalepsy,Grimacing,Echopraxia/echolalia,Stereotypy,Waxy flexibility | 98.0 | 96.6 | 99.1 |
| 8 | Excitement,Immobility/stupor,Staring,Posturing/catalepsy,Grimacing,Stereotypy,Mannerisms,Verbigeration | 98.0 | 96.6 | 99.1 |
| 8 | Excitement,Immobility/stupor,Staring,Posturing/catalepsy,Grimacing,Stereotypy,Verbigeration,Waxy flexibility | 98.0 | 96.6 | 99.1 |
| 8 | Excitement,Immobility/stupor,Staring,Posturing/catalepsy,Grimacing,Stereotypy,Waxy flexibility,Withdrawal | 98.0 | 96.6 | 99.1 |
| 8 | Excitement,Immobility/stupor,Staring,Stereotypy,Mannerisms,Verbigeration,Waxy flexibility,Withdrawal | 98.0 | 96.6 | 99.1 |
| 8 | Excitement,Mutism,Staring,Grimacing,Echopraxia/echolalia,Stereotypy,Mannerisms,Verbigeration | 98.0 | 96.6 | 99.1 |
| 8 | Excitement,Staring,Grimacing,Echopraxia/echolalia,Mannerisms,Rigidity,Negativism,Waxy flexibility | 98.0 | 96.6 | 99.1 |
| 8 | Excitement,Staring,Grimacing,Echopraxia/echolalia,Mannerisms,Verbigeration,Negativism,Withdrawal | 98.0 | 96.6 | 99.1 |
| 8 | Excitement,Staring,Grimacing,Echopraxia/echolalia,Stereotypy,Mannerisms,Rigidity,Negativism | 98.0 | 96.6 | 99.1 |
| 8 | Excitement,Staring,Grimacing,Echopraxia/echolalia,Stereotypy,Mannerisms,Rigidity,Waxy flexibility | 98.0 | 96.6 | 99.1 |
| 8 | Excitement,Staring,Grimacing,Echopraxia/echolalia,Stereotypy,Mannerisms,Waxy flexibility,Withdrawal | 98.0 | 96.6 | 99.1 |
| 8 | Excitement,Staring,Grimacing,Echopraxia/echolalia,Stereotypy,Rigidity,Negativism,Waxy flexibility | 98.0 | 96.6 | 99.1 |
| 8 | Excitement,Staring,Grimacing,Echopraxia/echolalia,Verbigeration,Negativism,Waxy flexibility,Withdrawal | 98.0 | 96.6 | 99.1 |
| 8 | Excitement,Staring,Grimacing,Mannerisms,Verbigeration,Rigidity,Negativism,Waxy flexibility | 98.0 | 96.6 | 99.1 |
| 8 | Excitement,Staring,Grimacing,Stereotypy,Mannerisms,Negativism,Waxy flexibility,Withdrawal | 98.0 | 96.6 | 99.1 |
| 8 | Excitement,Staring,Grimacing,Stereotypy,Mannerisms,Rigidity,Negativism,Waxy flexibility | 98.0 | 96.6 | 99.1 |
| 8 | Excitement,Staring,Grimacing,Stereotypy,Mannerisms,Verbigeration,Rigidity,Negativism | 98.0 | 96.6 | 99.1 |
| 8 | Excitement,Staring,Grimacing,Stereotypy,Mannerisms,Verbigeration,Rigidity,Waxy flexibility | 98.0 | 96.6 | 99.1 |
| 8 | Excitement,Staring,Grimacing,Stereotypy,Verbigeration,Negativism,Waxy flexibility,Withdrawal | 98.0 | 96.6 | 99.1 |
| 8 | Excitement,Staring,Grimacing,Stereotypy,Verbigeration,Rigidity,Negativism,Waxy flexibility | 98.0 | 96.6 | 99.1 |
| 8 | Excitement,Staring,Posturing/catalepsy,Echopraxia/echolalia,Mannerisms,Verbigeration,Waxy flexibility,Withdrawal | 98.0 | 96.6 | 99.1 |
| 8 | Excitement,Staring,Posturing/catalepsy,Echopraxia/echolalia,Stereotypy,Mannerisms,Waxy flexibility,Withdrawal | 98.0 | 96.6 | 99.1 |
| 8 | Excitement,Staring,Posturing/catalepsy,Echopraxia/echolalia,Stereotypy,Rigidity,Negativism,Waxy flexibility | 98.0 | 96.6 | 99.1 |
| 8 | Excitement,Staring,Posturing/catalepsy,Echopraxia/echolalia,Verbigeration,Rigidity,Negativism,Waxy flexibility | 98.0 | 96.6 | 99.1 |
| 8 | Excitement,Staring,Posturing/catalepsy,Grimacing,Echopraxia/echolalia,Mannerisms,Verbigeration,Rigidity | 98.0 | 96.6 | 99.1 |
| 8 | Excitement,Staring,Posturing/catalepsy,Grimacing,Echopraxia/echolalia,Stereotypy,Mannerisms,Rigidity | 98.0 | 96.6 | 99.1 |
| 8 | Excitement,Staring,Posturing/catalepsy,Grimacing,Echopraxia/echolalia,Stereotypy,Rigidity,Negativism | 98.0 | 96.6 | 99.1 |
| 8 | Excitement,Staring,Posturing/catalepsy,Grimacing,Echopraxia/echolalia,Verbigeration,Rigidity,Negativism | 98.0 | 96.6 | 99.1 |
| 8 | Excitement,Staring,Posturing/catalepsy,Grimacing,Stereotypy,Mannerisms,Rigidity,Negativism | 98.0 | 96.6 | 99.1 |
| 8 | Excitement,Staring,Posturing/catalepsy,Grimacing,Stereotypy,Mannerisms,Verbigeration,Rigidity | 98.0 | 96.6 | 99.1 |
| 8 | Excitement,Staring,Posturing/catalepsy,Grimacing,Stereotypy,Mannerisms,Verbigeration,Withdrawal | 98.0 | 96.6 | 99.1 |
| 8 | Excitement,Staring,Posturing/catalepsy,Grimacing,Stereotypy,Verbigeration,Rigidity,Negativism | 98.0 | 96.6 | 99.1 |
| 8 | Excitement,Staring,Posturing/catalepsy,Stereotypy,Mannerisms,Rigidity,Negativism,Waxy flexibility | 98.0 | 96.6 | 99.1 |
| 8 | Excitement,Staring,Posturing/catalepsy,Stereotypy,Verbigeration,Rigidity,Negativism,Waxy flexibility | 98.0 | 96.6 | 99.1 |
| 8 | Immobility/stupor,Echopraxia/echolalia,Stereotypy,Verbigeration,Rigidity,Negativism,Waxy flexibility,Withdrawal | 98.0 | 96.6 | 99.1 |
| 8 | Immobility/stupor,Grimacing,Echopraxia/echolalia,Mannerisms,Verbigeration,Rigidity,Negativism,Waxy flexibility | 98.0 | 96.6 | 99.1 |
| 8 | Immobility/stupor,Grimacing,Echopraxia/echolalia,Stereotypy,Mannerisms,Verbigeration,Rigidity,Negativism | 98.0 | 96.6 | 99.1 |
| 8 | Immobility/stupor,Grimacing,Mannerisms,Verbigeration,Rigidity,Negativism,Waxy flexibility,Withdrawal | 98.0 | 96.6 | 99.1 |
| 8 | Immobility/stupor,Grimacing,Stereotypy,Verbigeration,Rigidity,Negativism,Waxy flexibility,Withdrawal | 98.0 | 96.6 | 99.1 |
| 8 | Immobility/stupor,Mutism,Grimacing,Echopraxia/echolalia,Stereotypy,Rigidity,Negativism,Withdrawal | 98.0 | 96.6 | 99.1 |
| 8 | Immobility/stupor,Mutism,Grimacing,Mannerisms,Verbigeration,Rigidity,Negativism,Waxy flexibility | 98.0 | 96.6 | 99.1 |
| 8 | Immobility/stupor,Mutism,Grimacing,Stereotypy,Verbigeration,Rigidity,Negativism,Waxy flexibility | 98.0 | 96.6 | 99.1 |
| 8 | Immobility/stupor,Mutism,Posturing/catalepsy,Echopraxia/echolalia,Mannerisms,Rigidity,Negativism,Waxy flexibility | 98.0 | 96.6 | 99.1 |
| 8 | Immobility/stupor,Mutism,Posturing/catalepsy,Echopraxia/echolalia,Stereotypy,Verbigeration,Rigidity,Negativism | 98.0 | 96.6 | 99.1 |
| 8 | Immobility/stupor,Mutism,Posturing/catalepsy,Mannerisms,Rigidity,Negativism,Waxy flexibility,Withdrawal | 98.0 | 96.6 | 99.1 |
| 8 | Immobility/stupor,Mutism,Posturing/catalepsy,Mannerisms,Verbigeration,Rigidity,Negativism,Waxy flexibility | 98.0 | 96.6 | 99.1 |
| 8 | Immobility/stupor,Mutism,Posturing/catalepsy,Stereotypy,Mannerisms,Rigidity,Negativism,Waxy flexibility | 98.0 | 96.6 | 99.1 |
| 8 | Immobility/stupor,Mutism,Posturing/catalepsy,Stereotypy,Verbigeration,Rigidity,Negativism,Waxy flexibility | 98.0 | 96.6 | 99.1 |
| 8 | Immobility/stupor,Mutism,Staring,Echopraxia/echolalia,Mannerisms,Verbigeration,Rigidity,Waxy flexibility | 98.0 | 96.6 | 99.1 |
| 8 | Immobility/stupor,Mutism,Staring,Echopraxia/echolalia,Rigidity,Negativism,Waxy flexibility,Withdrawal | 98.0 | 96.6 | 99.1 |
| 8 | Immobility/stupor,Mutism,Staring,Echopraxia/echolalia,Stereotypy,Mannerisms,Rigidity,Waxy flexibility | 98.0 | 96.6 | 99.1 |
| 8 | Immobility/stupor,Mutism,Staring,Echopraxia/echolalia,Stereotypy,Mannerisms,Verbigeration,Rigidity | 98.0 | 96.6 | 99.1 |
| 8 | Immobility/stupor,Mutism,Staring,Echopraxia/echolalia,Stereotypy,Verbigeration,Negativism,Withdrawal | 98.0 | 96.6 | 99.1 |
| 8 | Immobility/stupor,Mutism,Staring,Echopraxia/echolalia,Stereotypy,Verbigeration,Rigidity,Withdrawal | 98.0 | 96.6 | 99.1 |
| 8 | Immobility/stupor,Mutism,Staring,Echopraxia/echolalia,Verbigeration,Rigidity,Negativism,Waxy flexibility | 98.0 | 96.6 | 99.1 |
| 8 | Immobility/stupor,Mutism,Staring,Grimacing,Echopraxia/echolalia,Mannerisms,Rigidity,Waxy flexibility | 98.0 | 96.6 | 99.1 |
| 8 | Immobility/stupor,Mutism,Staring,Grimacing,Echopraxia/echolalia,Mannerisms,Waxy flexibility,Withdrawal | 98.0 | 96.6 | 99.1 |
| 8 | Immobility/stupor,Mutism,Staring,Grimacing,Echopraxia/echolalia,Stereotypy,Rigidity,Negativism | 98.0 | 96.6 | 99.1 |
| 8 | Immobility/stupor,Mutism,Staring,Grimacing,Echopraxia/echolalia,Stereotypy,Verbigeration,Negativism | 98.0 | 96.6 | 99.1 |
| 8 | Immobility/stupor,Mutism,Staring,Grimacing,Echopraxia/echolalia,Stereotypy,Verbigeration,Rigidity | 98.0 | 96.6 | 99.1 |
| 8 | Immobility/stupor,Mutism,Staring,Grimacing,Mannerisms,Verbigeration,Negativism,Waxy flexibility | 98.0 | 96.6 | 99.1 |
| 8 | Immobility/stupor,Mutism,Staring,Grimacing,Stereotypy,Mannerisms,Waxy flexibility,Withdrawal | 98.0 | 96.6 | 99.1 |
| 8 | Immobility/stupor,Mutism,Staring,Grimacing,Stereotypy,Verbigeration,Negativism,Waxy flexibility | 98.0 | 96.6 | 99.1 |
| 8 | Immobility/stupor,Mutism,Staring,Posturing/catalepsy,Echopraxia/echolalia,Negativism,Waxy flexibility,Withdrawal | 98.0 | 96.6 | 99.1 |
| 8 | Immobility/stupor,Mutism,Staring,Posturing/catalepsy,Echopraxia/echolalia,Stereotypy,Mannerisms,Withdrawal | 98.0 | 96.6 | 99.1 |
| 8 | Immobility/stupor,Mutism,Staring,Posturing/catalepsy,Echopraxia/echolalia,Stereotypy,Verbigeration,Rigidity | 98.0 | 96.6 | 99.1 |
| 8 | Immobility/stupor,Mutism,Staring,Posturing/catalepsy,Grimacing,Echopraxia/echolalia,Mannerisms,Verbigeration | 98.0 | 96.6 | 99.1 |
| 8 | Immobility/stupor,Mutism,Staring,Posturing/catalepsy,Grimacing,Echopraxia/echolalia,Stereotypy,Negativism | 98.0 | 96.6 | 99.1 |
| 8 | Immobility/stupor,Mutism,Staring,Posturing/catalepsy,Grimacing,Mannerisms,Waxy flexibility,Withdrawal | 98.0 | 96.4 | 99.1 |
| 8 | Immobility/stupor,Mutism,Staring,Posturing/catalepsy,Grimacing,Stereotypy,Rigidity,Negativism | 98.0 | 96.6 | 99.1 |
| 8 | Immobility/stupor,Mutism,Staring,Posturing/catalepsy,Grimacing,Stereotypy,Rigidity,Withdrawal | 98.0 | 96.6 | 99.1 |
| 8 | Immobility/stupor,Mutism,Staring,Posturing/catalepsy,Grimacing,Verbigeration,Rigidity,Negativism | 98.0 | 96.6 | 99.1 |
| 8 | Immobility/stupor,Mutism,Staring,Posturing/catalepsy,Mannerisms,Verbigeration,Rigidity,Waxy flexibility | 98.0 | 96.6 | 99.1 |
| 8 | Immobility/stupor,Mutism,Staring,Posturing/catalepsy,Stereotypy,Negativism,Waxy flexibility,Withdrawal | 98.0 | 96.6 | 99.1 |
| 8 | Immobility/stupor,Mutism,Staring,Posturing/catalepsy,Stereotypy,Verbigeration,Rigidity,Waxy flexibility | 98.0 | 96.6 | 99.1 |
| 8 | Immobility/stupor,Mutism,Staring,Stereotypy,Verbigeration,Negativism,Waxy flexibility,Withdrawal | 98.0 | 96.6 | 99.1 |
| 8 | Immobility/stupor,Mutism,Staring,Verbigeration,Rigidity,Negativism,Waxy flexibility,Withdrawal | 98.0 | 96.6 | 99.1 |
| 8 | Immobility/stupor,Posturing/catalepsy,Echopraxia/echolalia,Mannerisms,Verbigeration,Rigidity,Negativism,Waxy flexibility | 98.0 | 96.6 | 99.1 |
| 8 | Immobility/stupor,Posturing/catalepsy,Echopraxia/echolalia,Stereotypy,Mannerisms,Verbigeration,Rigidity,Negativism | 98.0 | 96.6 | 99.1 |
| 8 | Immobility/stupor,Posturing/catalepsy,Echopraxia/echolalia,Stereotypy,Verbigeration,Rigidity,Negativism,Withdrawal | 98.0 | 96.6 | 99.1 |
| 8 | Immobility/stupor,Posturing/catalepsy,Grimacing,Echopraxia/echolalia,Stereotypy,Mannerisms,Negativism,Withdrawal | 98.0 | 96.6 | 99.1 |
| 8 | Immobility/stupor,Posturing/catalepsy,Grimacing,Echopraxia/echolalia,Stereotypy,Mannerisms,Rigidity,Negativism | 98.0 | 96.6 | 99.1 |
| 8 | Immobility/stupor,Posturing/catalepsy,Grimacing,Echopraxia/echolalia,Stereotypy,Rigidity,Negativism,Withdrawal | 98.0 | 96.6 | 99.1 |
| 8 | Immobility/stupor,Posturing/catalepsy,Grimacing,Mannerisms,Verbigeration,Rigidity,Negativism,Waxy flexibility | 98.0 | 96.6 | 99.1 |
| 8 | Immobility/stupor,Posturing/catalepsy,Grimacing,Stereotypy,Mannerisms,Rigidity,Negativism,Waxy flexibility | 98.0 | 96.6 | 99.1 |
| 8 | Immobility/stupor,Posturing/catalepsy,Stereotypy,Verbigeration,Rigidity,Negativism,Waxy flexibility,Withdrawal | 98.0 | 96.6 | 99.1 |
| 8 | Immobility/stupor,Staring,Echopraxia/echolalia,Stereotypy,Mannerisms,Verbigeration,Rigidity,Withdrawal | 98.0 | 96.6 | 99.1 |
| 8 | Immobility/stupor,Staring,Grimacing,Echopraxia/echolalia,Rigidity,Negativism,Waxy flexibility,Withdrawal | 98.0 | 96.6 | 99.1 |
| 8 | Immobility/stupor,Staring,Grimacing,Echopraxia/echolalia,Stereotypy,Mannerisms,Rigidity,Waxy flexibility | 98.0 | 96.6 | 99.1 |
| 8 | Immobility/stupor,Staring,Grimacing,Echopraxia/echolalia,Stereotypy,Mannerisms,Verbigeration,Rigidity | 98.0 | 96.6 | 99.1 |
| 8 | Immobility/stupor,Staring,Grimacing,Echopraxia/echolalia,Stereotypy,Mannerisms,Waxy flexibility,Withdrawal | 98.0 | 96.6 | 99.1 |
| 8 | Immobility/stupor,Staring,Grimacing,Echopraxia/echolalia,Stereotypy,Verbigeration,Rigidity,Withdrawal | 98.0 | 96.6 | 99.1 |
| 8 | Immobility/stupor,Staring,Grimacing,Echopraxia/echolalia,Verbigeration,Rigidity,Negativism,Waxy flexibility | 98.0 | 96.6 | 99.1 |
| 8 | Immobility/stupor,Staring,Grimacing,Stereotypy,Verbigeration,Negativism,Waxy flexibility,Withdrawal | 98.0 | 96.6 | 99.1 |
| 8 | Immobility/stupor,Staring,Grimacing,Verbigeration,Rigidity,Negativism,Waxy flexibility,Withdrawal | 98.0 | 96.6 | 99.1 |
| 8 | Immobility/stupor,Staring,Posturing/catalepsy,Echopraxia/echolalia,Mannerisms,Verbigeration,Rigidity,Withdrawal | 98.0 | 96.6 | 99.1 |
| 8 | Immobility/stupor,Staring,Posturing/catalepsy,Echopraxia/echolalia,Stereotypy,Negativism,Waxy flexibility,Withdrawal | 98.0 | 96.6 | 99.1 |
| 8 | Immobility/stupor,Staring,Posturing/catalepsy,Grimacing,Echopraxia/echolalia,Mannerisms,Verbigeration,Rigidity | 98.0 | 96.6 | 99.1 |
| 8 | Immobility/stupor,Staring,Posturing/catalepsy,Grimacing,Echopraxia/echolalia,Stereotypy,Mannerisms,Rigidity | 98.0 | 96.6 | 99.1 |
| 8 | Immobility/stupor,Staring,Posturing/catalepsy,Grimacing,Echopraxia/echolalia,Stereotypy,Rigidity,Negativism | 98.0 | 96.6 | 99.1 |
| 8 | Immobility/stupor,Staring,Posturing/catalepsy,Grimacing,Echopraxia/echolalia,Stereotypy,Rigidity,Withdrawal | 98.0 | 96.6 | 99.1 |
| 8 | Immobility/stupor,Staring,Posturing/catalepsy,Grimacing,Echopraxia/echolalia,Stereotypy,Verbigeration,Negativism | 98.0 | 96.6 | 99.1 |
| 8 | Immobility/stupor,Staring,Posturing/catalepsy,Grimacing,Echopraxia/echolalia,Verbigeration,Negativism,Withdrawal | 98.0 | 96.6 | 99.1 |
| 8 | Immobility/stupor,Staring,Posturing/catalepsy,Grimacing,Mannerisms,Rigidity,Waxy flexibility,Withdrawal | 98.0 | 96.6 | 99.1 |
| 8 | Immobility/stupor,Staring,Posturing/catalepsy,Grimacing,Stereotypy,Mannerisms,Verbigeration,Withdrawal | 98.0 | 96.6 | 99.1 |
| 8 | Immobility/stupor,Staring,Posturing/catalepsy,Grimacing,Stereotypy,Verbigeration,Negativism,Waxy flexibility | 98.0 | 96.6 | 99.1 |
| 8 | Immobility/stupor,Staring,Posturing/catalepsy,Verbigeration,Rigidity,Negativism,Waxy flexibility,Withdrawal | 98.0 | 96.6 | 99.1 |
| 8 | Mutism,Staring,Grimacing,Echopraxia/echolalia,Verbigeration,Negativism,Waxy flexibility,Withdrawal | 98.0 | 96.6 | 99.1 |
| 8 | Mutism,Staring,Grimacing,Echopraxia/echolalia,Verbigeration,Rigidity,Waxy flexibility,Withdrawal | 98.0 | 96.6 | 99.1 |
| 8 | Mutism,Staring,Posturing/catalepsy,Echopraxia/echolalia,Mannerisms,Verbigeration,Waxy flexibility,Withdrawal | 98.0 | 96.6 | 99.1 |
| 8 | Mutism,Staring,Posturing/catalepsy,Echopraxia/echolalia,Stereotypy,Mannerisms,Verbigeration,Withdrawal | 98.0 | 96.6 | 99.1 |
| 8 | Mutism,Staring,Posturing/catalepsy,Echopraxia/echolalia,Stereotypy,Verbigeration,Rigidity,Waxy flexibility | 98.0 | 96.6 | 99.1 |
| 8 | Mutism,Staring,Posturing/catalepsy,Grimacing,Echopraxia/echolalia,Stereotypy,Verbigeration,Withdrawal | 98.0 | 96.6 | 99.1 |
| 8 | Mutism,Staring,Posturing/catalepsy,Grimacing,Echopraxia/echolalia,Stereotypy,Waxy flexibility,Withdrawal | 98.0 | 96.6 | 99.1 |
| 8 | Mutism,Staring,Posturing/catalepsy,Grimacing,Stereotypy,Mannerisms,Verbigeration,Waxy flexibility | 98.0 | 96.6 | 99.1 |
| 8 | Mutism,Staring,Posturing/catalepsy,Grimacing,Stereotypy,Rigidity,Waxy flexibility,Withdrawal | 98.0 | 96.6 | 99.1 |
| 8 | Mutism,Staring,Posturing/catalepsy,Grimacing,Stereotypy,Verbigeration,Waxy flexibility,Withdrawal | 98.0 | 96.6 | 99.1 |
| 8 | Mutism,Staring,Posturing/catalepsy,Stereotypy,Mannerisms,Verbigeration,Waxy flexibility,Withdrawal | 98.0 | 96.6 | 99.1 |
| 8 | Staring,Echopraxia/echolalia,Stereotypy,Mannerisms,Verbigeration,Rigidity,Negativism,Waxy flexibility | 98.0 | 96.6 | 99.1 |
| 8 | Staring,Grimacing,Echopraxia/echolalia,Mannerisms,Verbigeration,Negativism,Waxy flexibility,Withdrawal | 98.0 | 96.6 | 99.1 |
| 8 | Staring,Grimacing,Echopraxia/echolalia,Mannerisms,Verbigeration,Rigidity,Waxy flexibility,Withdrawal | 98.0 | 96.6 | 99.1 |
| 8 | Staring,Grimacing,Echopraxia/echolalia,Stereotypy,Mannerisms,Rigidity,Negativism,Waxy flexibility | 98.0 | 96.6 | 99.1 |
| 8 | Staring,Grimacing,Echopraxia/echolalia,Stereotypy,Rigidity,Negativism,Waxy flexibility,Withdrawal | 98.0 | 96.6 | 99.1 |
| 8 | Staring,Grimacing,Stereotypy,Mannerisms,Verbigeration,Rigidity,Negativism,Waxy flexibility | 98.0 | 96.6 | 99.1 |
| 8 | Staring,Posturing/catalepsy,Echopraxia/echolalia,Stereotypy,Mannerisms,Verbigeration,Rigidity,Negativism | 98.0 | 96.6 | 99.1 |
| 8 | Staring,Posturing/catalepsy,Echopraxia/echolalia,Stereotypy,Rigidity,Negativism,Waxy flexibility,Withdrawal | 98.0 | 96.6 | 99.1 |
| 8 | Staring,Posturing/catalepsy,Grimacing,Echopraxia/echolalia,Mannerisms,Rigidity,Waxy flexibility,Withdrawal | 98.0 | 96.6 | 99.1 |
| 8 | Staring,Posturing/catalepsy,Grimacing,Echopraxia/echolalia,Stereotypy,Mannerisms,Rigidity,Negativism | 98.0 | 96.6 | 99.1 |
| 8 | Staring,Posturing/catalepsy,Grimacing,Echopraxia/echolalia,Verbigeration,Rigidity,Negativism,Withdrawal | 98.0 | 96.6 | 99.1 |
| 8 | Staring,Posturing/catalepsy,Grimacing,Mannerisms,Verbigeration,Rigidity,Waxy flexibility,Withdrawal | 98.0 | 96.6 | 99.1 |
| 8 | Staring,Posturing/catalepsy,Grimacing,Stereotypy,Mannerisms,Verbigeration,Rigidity,Negativism | 98.0 | 96.6 | 99.1 |
| 8 | Excitement,Immobility/stupor,Echopraxia/echolalia,Stereotypy,Verbigeration,Negativism,Waxy flexibility,Withdrawal | 97.8 | 96.2 | 99.1 |
| 8 | Excitement,Immobility/stupor,Grimacing,Echopraxia/echolalia,Stereotypy,Verbigeration,Negativism,Withdrawal | 97.8 | 96.4 | 99.1 |
| 8 | Excitement,Immobility/stupor,Mutism,Echopraxia/echolalia,Stereotypy,Mannerisms,Negativism,Withdrawal | 97.8 | 96.2 | 99.1 |
| 8 | Excitement,Immobility/stupor,Mutism,Echopraxia/echolalia,Stereotypy,Verbigeration,Negativism,Waxy flexibility | 97.8 | 96.4 | 98.9 |
| 8 | Excitement,Immobility/stupor,Mutism,Echopraxia/echolalia,Verbigeration,Negativism,Waxy flexibility,Withdrawal | 97.8 | 96.2 | 99.1 |
| 8 | Excitement,Immobility/stupor,Mutism,Grimacing,Echopraxia/echolalia,Negativism,Waxy flexibility,Withdrawal | 97.8 | 96.4 | 99.1 |
| 8 | Excitement,Immobility/stupor,Mutism,Grimacing,Echopraxia/echolalia,Verbigeration,Negativism,Withdrawal | 97.8 | 96.4 | 99.1 |
| 8 | Excitement,Immobility/stupor,Mutism,Grimacing,Stereotypy,Mannerisms,Rigidity,Waxy flexibility | 97.8 | 96.2 | 99.1 |
| 8 | Excitement,Immobility/stupor,Mutism,Posturing/catalepsy,Echopraxia/echolalia,Stereotypy,Negativism,Waxy flexibility | 97.8 | 96.4 | 99.1 |
| 8 | Excitement,Immobility/stupor,Mutism,Posturing/catalepsy,Echopraxia/echolalia,Stereotypy,Verbigeration,Withdrawal | 97.8 | 96.4 | 99.1 |
| 8 | Excitement,Immobility/stupor,Mutism,Posturing/catalepsy,Echopraxia/echolalia,Verbigeration,Waxy flexibility,Withdrawal | 97.8 | 96.2 | 99.1 |
| 8 | Excitement,Immobility/stupor,Mutism,Posturing/catalepsy,Grimacing,Mannerisms,Negativism,Waxy flexibility | 97.8 | 96.2 | 99.1 |
| 8 | Excitement,Immobility/stupor,Mutism,Posturing/catalepsy,Grimacing,Stereotypy,Negativism,Waxy flexibility | 97.8 | 96.2 | 99.1 |
| 8 | Excitement,Immobility/stupor,Mutism,Posturing/catalepsy,Stereotypy,Mannerisms,Rigidity,Waxy flexibility | 97.8 | 96.2 | 99.1 |
| 8 | Excitement,Immobility/stupor,Mutism,Staring,Echopraxia/echolalia,Stereotypy,Mannerisms,Verbigeration | 97.8 | 96.4 | 99.1 |
| 8 | Excitement,Immobility/stupor,Mutism,Staring,Grimacing,Echopraxia/echolalia,Stereotypy,Mannerisms | 97.8 | 96.4 | 99.1 |
| 8 | Excitement,Immobility/stupor,Mutism,Staring,Grimacing,Mannerisms,Verbigeration,Withdrawal | 97.8 | 96.2 | 99.1 |
| 8 | Excitement,Immobility/stupor,Mutism,Staring,Grimacing,Stereotypy,Mannerisms,Withdrawal | 97.8 | 96.4 | 99.1 |
| 8 | Excitement,Immobility/stupor,Posturing/catalepsy,Echopraxia/echolalia,Mannerisms,Verbigeration,Rigidity,Waxy flexibility | 97.8 | 96.2 | 99.1 |
| 8 | Excitement,Immobility/stupor,Posturing/catalepsy,Echopraxia/echolalia,Stereotypy,Mannerisms,Verbigeration,Rigidity | 97.8 | 96.4 | 99.1 |
| 8 | Excitement,Immobility/stupor,Posturing/catalepsy,Grimacing,Echopraxia/echolalia,Mannerisms,Rigidity,Waxy flexibility | 97.8 | 96.4 | 99.1 |
| 8 | Excitement,Immobility/stupor,Posturing/catalepsy,Grimacing,Echopraxia/echolalia,Verbigeration,Negativism,Waxy flexibility | 97.8 | 96.2 | 99.1 |
| 8 | Excitement,Immobility/stupor,Posturing/catalepsy,Grimacing,Mannerisms,Negativism,Waxy flexibility,Withdrawal | 97.8 | 96.4 | 99.1 |
| 8 | Excitement,Immobility/stupor,Posturing/catalepsy,Grimacing,Mannerisms,Verbigeration,Negativism,Waxy flexibility | 97.8 | 96.2 | 99.1 |
| 8 | Excitement,Immobility/stupor,Posturing/catalepsy,Grimacing,Mannerisms,Verbigeration,Rigidity,Waxy flexibility | 97.8 | 96.4 | 99.1 |
| 8 | Excitement,Immobility/stupor,Posturing/catalepsy,Grimacing,Stereotypy,Mannerisms,Negativism,Withdrawal | 97.8 | 96.2 | 99.1 |
| 8 | Excitement,Immobility/stupor,Staring,Grimacing,Echopraxia/echolalia,Mannerisms,Verbigeration,Withdrawal | 97.8 | 96.2 | 99.1 |
| 8 | Excitement,Immobility/stupor,Staring,Grimacing,Echopraxia/echolalia,Stereotypy,Verbigeration,Waxy flexibility | 97.8 | 96.2 | 99.1 |
| 8 | Excitement,Immobility/stupor,Staring,Grimacing,Stereotypy,Mannerisms,Verbigeration,Waxy flexibility | 97.8 | 96.4 | 99.1 |
| 8 | Excitement,Immobility/stupor,Staring,Grimacing,Stereotypy,Mannerisms,Verbigeration,Withdrawal | 97.8 | 96.2 | 99.1 |
| 8 | Excitement,Immobility/stupor,Staring,Grimacing,Stereotypy,Mannerisms,Waxy flexibility,Withdrawal | 97.8 | 96.2 | 99.1 |
| 8 | Excitement,Immobility/stupor,Staring,Grimacing,Stereotypy,Verbigeration,Waxy flexibility,Withdrawal | 97.8 | 96.4 | 99.1 |
| 8 | Excitement,Immobility/stupor,Staring,Posturing/catalepsy,Grimacing,Stereotypy,Mannerisms,Waxy flexibility | 97.8 | 96.4 | 99.1 |
| 8 | Excitement,Staring,Grimacing,Echopraxia/echolalia,Mannerisms,Verbigeration,Rigidity,Waxy flexibility | 97.8 | 96.4 | 99.1 |
| 8 | Excitement,Staring,Grimacing,Echopraxia/echolalia,Mannerisms,Verbigeration,Waxy flexibility,Withdrawal | 97.8 | 96.4 | 99.1 |
| 8 | Excitement,Staring,Grimacing,Echopraxia/echolalia,Stereotypy,Verbigeration,Rigidity,Negativism | 97.8 | 96.2 | 99.1 |
| 8 | Excitement,Staring,Grimacing,Echopraxia/echolalia,Stereotypy,Verbigeration,Rigidity,Waxy flexibility | 97.8 | 96.4 | 99.1 |
| 8 | Excitement,Staring,Grimacing,Echopraxia/echolalia,Verbigeration,Rigidity,Negativism,Waxy flexibility | 97.8 | 96.2 | 99.1 |
| 8 | Excitement,Staring,Posturing/catalepsy,Echopraxia/echolalia,Mannerisms,Verbigeration,Negativism,Waxy flexibility | 97.8 | 96.2 | 99.1 |
| 8 | Excitement,Staring,Posturing/catalepsy,Echopraxia/echolalia,Stereotypy,Mannerisms,Negativism,Waxy flexibility | 97.8 | 96.2 | 99.1 |
| 8 | Excitement,Staring,Posturing/catalepsy,Grimacing,Echopraxia/echolalia,Mannerisms,Rigidity,Waxy flexibility | 97.8 | 96.4 | 99.1 |
| 8 | Excitement,Staring,Posturing/catalepsy,Grimacing,Echopraxia/echolalia,Rigidity,Negativism,Waxy flexibility | 97.8 | 96.2 | 99.1 |
| 8 | Excitement,Staring,Posturing/catalepsy,Grimacing,Echopraxia/echolalia,Stereotypy,Verbigeration,Rigidity | 97.8 | 96.4 | 99.1 |
| 8 | Excitement,Staring,Posturing/catalepsy,Grimacing,Echopraxia/echolalia,Stereotypy,Waxy flexibility,Withdrawal | 97.8 | 96.4 | 99.1 |
| 8 | Excitement,Staring,Posturing/catalepsy,Grimacing,Echopraxia/echolalia,Verbigeration,Waxy flexibility,Withdrawal | 97.8 | 96.2 | 99.1 |
| 8 | Excitement,Staring,Posturing/catalepsy,Grimacing,Mannerisms,Rigidity,Negativism,Waxy flexibility | 97.8 | 96.2 | 99.1 |
| 8 | Excitement,Staring,Posturing/catalepsy,Grimacing,Mannerisms,Verbigeration,Rigidity,Waxy flexibility | 97.8 | 96.2 | 99.1 |
| 8 | Excitement,Staring,Posturing/catalepsy,Stereotypy,Mannerisms,Verbigeration,Waxy flexibility,Withdrawal | 97.8 | 96.2 | 99.1 |
| 8 | Immobility/stupor,Grimacing,Echopraxia/echolalia,Stereotypy,Rigidity,Negativism,Waxy flexibility,Withdrawal | 97.8 | 96.4 | 99.1 |
| 8 | Immobility/stupor,Mutism,Grimacing,Echopraxia/echolalia,Stereotypy,Mannerisms,Rigidity,Withdrawal | 97.8 | 96.2 | 99.1 |
| 8 | Immobility/stupor,Mutism,Grimacing,Echopraxia/echolalia,Stereotypy,Verbigeration,Rigidity,Negativism | 97.8 | 96.2 | 99.1 |
| 8 | Immobility/stupor,Mutism,Grimacing,Echopraxia/echolalia,Verbigeration,Rigidity,Negativism,Withdrawal | 97.8 | 96.2 | 99.1 |
| 8 | Immobility/stupor,Mutism,Posturing/catalepsy,Echopraxia/echolalia,Mannerisms,Verbigeration,Negativism,Withdrawal | 97.8 | 96.4 | 99.1 |
| 8 | Immobility/stupor,Mutism,Posturing/catalepsy,Echopraxia/echolalia,Stereotypy,Mannerisms,Negativism,Withdrawal | 97.8 | 96.2 | 99.1 |
| 8 | Immobility/stupor,Mutism,Posturing/catalepsy,Echopraxia/echolalia,Stereotypy,Rigidity,Negativism,Withdrawal | 97.8 | 96.4 | 99.1 |
| 8 | Immobility/stupor,Mutism,Posturing/catalepsy,Grimacing,Echopraxia/echolalia,Mannerisms,Rigidity,Withdrawal | 97.8 | 96.2 | 99.1 |
| 8 | Immobility/stupor,Mutism,Posturing/catalepsy,Grimacing,Echopraxia/echolalia,Verbigeration,Rigidity,Negativism | 97.8 | 96.2 | 99.1 |
| 8 | Immobility/stupor,Mutism,Posturing/catalepsy,Grimacing,Mannerisms,Rigidity,Negativism,Waxy flexibility | 97.8 | 96.2 | 99.1 |
| 8 | Immobility/stupor,Mutism,Posturing/catalepsy,Grimacing,Mannerisms,Verbigeration,Negativism,Withdrawal | 97.8 | 96.2 | 99.1 |
| 8 | Immobility/stupor,Mutism,Posturing/catalepsy,Grimacing,Stereotypy,Mannerisms,Negativism,Withdrawal | 97.8 | 96.4 | 99.1 |
| 8 | Immobility/stupor,Mutism,Posturing/catalepsy,Grimacing,Stereotypy,Rigidity,Negativism,Withdrawal | 97.8 | 96.4 | 99.1 |
| 8 | Immobility/stupor,Mutism,Posturing/catalepsy,Stereotypy,Mannerisms,Verbigeration,Negativism,Withdrawal | 97.8 | 96.4 | 99.1 |
| 8 | Immobility/stupor,Mutism,Staring,Echopraxia/echolalia,Stereotypy,Negativism,Waxy flexibility,Withdrawal | 97.8 | 96.4 | 99.1 |
| 8 | Immobility/stupor,Mutism,Staring,Echopraxia/echolalia,Stereotypy,Rigidity,Negativism,Waxy flexibility | 97.8 | 96.2 | 99.1 |
| 8 | Immobility/stupor,Mutism,Staring,Echopraxia/echolalia,Stereotypy,Rigidity,Waxy flexibility,Withdrawal | 97.8 | 96.4 | 99.1 |
| 8 | Immobility/stupor,Mutism,Staring,Echopraxia/echolalia,Stereotypy,Verbigeration,Negativism,Waxy flexibility | 97.8 | 96.4 | 99.1 |
| 8 | Immobility/stupor,Mutism,Staring,Grimacing,Echopraxia/echolalia,Mannerisms,Verbigeration,Waxy flexibility | 97.8 | 96.2 | 99.1 |
| 8 | Immobility/stupor,Mutism,Staring,Grimacing,Echopraxia/echolalia,Stereotypy,Mannerisms,Waxy flexibility | 97.8 | 96.2 | 99.1 |
| 8 | Immobility/stupor,Mutism,Staring,Grimacing,Echopraxia/echolalia,Verbigeration,Negativism,Waxy flexibility | 97.8 | 96.2 | 99.1 |
| 8 | Immobility/stupor,Mutism,Staring,Grimacing,Echopraxia/echolalia,Verbigeration,Negativism,Withdrawal | 97.8 | 96.2 | 99.1 |
| 8 | Immobility/stupor,Mutism,Staring,Grimacing,Echopraxia/echolalia,Verbigeration,Rigidity,Withdrawal | 97.8 | 96.4 | 99.1 |
| 8 | Immobility/stupor,Mutism,Staring,Grimacing,Stereotypy,Rigidity,Negativism,Waxy flexibility | 97.8 | 96.2 | 99.1 |
| 8 | Immobility/stupor,Mutism,Staring,Grimacing,Stereotypy,Rigidity,Waxy flexibility,Withdrawal | 97.8 | 96.2 | 99.1 |
| 8 | Immobility/stupor,Mutism,Staring,Grimacing,Verbigeration,Rigidity,Negativism,Waxy flexibility | 97.8 | 96.2 | 99.1 |
| 8 | Immobility/stupor,Mutism,Staring,Posturing/catalepsy,Echopraxia/echolalia,Mannerisms,Rigidity,Waxy flexibility | 97.8 | 96.4 | 99.1 |
| 8 | Immobility/stupor,Mutism,Staring,Posturing/catalepsy,Echopraxia/echolalia,Mannerisms,Verbigeration,Withdrawal | 97.8 | 96.4 | 99.1 |
| 8 | Immobility/stupor,Mutism,Staring,Posturing/catalepsy,Echopraxia/echolalia,Mannerisms,Waxy flexibility,Withdrawal | 97.8 | 96.2 | 99.1 |
| 8 | Immobility/stupor,Mutism,Staring,Posturing/catalepsy,Echopraxia/echolalia,Stereotypy,Negativism,Waxy flexibility | 97.8 | 96.2 | 99.1 |
| 8 | Immobility/stupor,Mutism,Staring,Posturing/catalepsy,Echopraxia/echolalia,Verbigeration,Rigidity,Withdrawal | 97.8 | 96.4 | 99.1 |
| 8 | Immobility/stupor,Mutism,Staring,Posturing/catalepsy,Grimacing,Echopraxia/echolalia,Rigidity,Negativism | 97.8 | 96.2 | 99.1 |
| 8 | Immobility/stupor,Mutism,Staring,Posturing/catalepsy,Grimacing,Echopraxia/echolalia,Rigidity,Withdrawal | 97.8 | 96.4 | 99.1 |
| 8 | Immobility/stupor,Mutism,Staring,Posturing/catalepsy,Grimacing,Echopraxia/echolalia,Stereotypy,Mannerisms | 97.8 | 96.4 | 99.1 |
| 8 | Immobility/stupor,Mutism,Staring,Posturing/catalepsy,Grimacing,Echopraxia/echolalia,Stereotypy,Withdrawal | 97.8 | 96.2 | 99.1 |
| 8 | Immobility/stupor,Mutism,Staring,Posturing/catalepsy,Grimacing,Echopraxia/echolalia,Verbigeration,Rigidity | 97.8 | 96.2 | 99.1 |
| 8 | Immobility/stupor,Mutism,Staring,Posturing/catalepsy,Grimacing,Stereotypy,Mannerisms,Verbigeration | 97.8 | 96.2 | 99.1 |
| 8 | Immobility/stupor,Mutism,Staring,Posturing/catalepsy,Grimacing,Stereotypy,Verbigeration,Withdrawal | 97.8 | 96.4 | 99.1 |
| 8 | Immobility/stupor,Mutism,Staring,Posturing/catalepsy,Grimacing,Verbigeration,Rigidity,Withdrawal | 97.8 | 96.2 | 99.1 |
| 8 | Immobility/stupor,Mutism,Staring,Posturing/catalepsy,Stereotypy,Mannerisms,Rigidity,Waxy flexibility | 97.8 | 96.2 | 99.1 |
| 8 | Immobility/stupor,Mutism,Staring,Posturing/catalepsy,Stereotypy,Mannerisms,Verbigeration,Withdrawal | 97.8 | 96.4 | 99.1 |
| 8 | Immobility/stupor,Mutism,Staring,Posturing/catalepsy,Stereotypy,Mannerisms,Waxy flexibility,Withdrawal | 97.8 | 96.2 | 99.1 |
| 8 | Immobility/stupor,Mutism,Staring,Posturing/catalepsy,Stereotypy,Rigidity,Negativism,Waxy flexibility | 97.8 | 96.2 | 99.1 |
| 8 | Immobility/stupor,Posturing/catalepsy,Echopraxia/echolalia,Stereotypy,Mannerisms,Verbigeration,Negativism,Withdrawal | 97.8 | 96.2 | 99.1 |
| 8 | Immobility/stupor,Posturing/catalepsy,Grimacing,Echopraxia/echolalia,Verbigeration,Rigidity,Negativism,Withdrawal | 97.8 | 96.2 | 99.1 |
| 8 | Immobility/stupor,Posturing/catalepsy,Grimacing,Stereotypy,Mannerisms,Verbigeration,Negativism,Withdrawal | 97.8 | 96.2 | 99.1 |
| 8 | Immobility/stupor,Posturing/catalepsy,Grimacing,Stereotypy,Verbigeration,Rigidity,Negativism,Waxy flexibility | 97.8 | 96.2 | 99.1 |
| 8 | Immobility/stupor,Staring,Echopraxia/echolalia,Stereotypy,Verbigeration,Negativism,Waxy flexibility,Withdrawal | 97.8 | 96.2 | 99.1 |
| 8 | Immobility/stupor,Staring,Grimacing,Echopraxia/echolalia,Stereotypy,Negativism,Waxy flexibility,Withdrawal | 97.8 | 96.4 | 99.1 |
| 8 | Immobility/stupor,Staring,Grimacing,Echopraxia/echolalia,Stereotypy,Rigidity,Negativism,Waxy flexibility | 97.8 | 96.2 | 99.1 |
| 8 | Immobility/stupor,Staring,Grimacing,Echopraxia/echolalia,Stereotypy,Rigidity,Waxy flexibility,Withdrawal | 97.8 | 96.2 | 99.1 |
| 8 | Immobility/stupor,Staring,Grimacing,Echopraxia/echolalia,Stereotypy,Verbigeration,Negativism,Waxy flexibility | 97.8 | 96.2 | 99.1 |
| 8 | Immobility/stupor,Staring,Posturing/catalepsy,Echopraxia/echolalia,Stereotypy,Mannerisms,Verbigeration,Rigidity | 97.8 | 96.4 | 99.1 |
| 8 | Immobility/stupor,Staring,Posturing/catalepsy,Echopraxia/echolalia,Stereotypy,Rigidity,Negativism,Waxy flexibility | 97.8 | 96.2 | 99.1 |
| 8 | Immobility/stupor,Staring,Posturing/catalepsy,Echopraxia/echolalia,Stereotypy,Verbigeration,Negativism,Waxy flexibility | 97.8 | 96.4 | 99.1 |
| 8 | Immobility/stupor,Staring,Posturing/catalepsy,Grimacing,Echopraxia/echolalia,Negativism,Waxy flexibility,Withdrawal | 97.8 | 96.4 | 99.1 |
| 8 | Immobility/stupor,Staring,Posturing/catalepsy,Grimacing,Stereotypy,Mannerisms,Rigidity,Waxy flexibility | 97.8 | 96.4 | 99.1 |
| 8 | Immobility/stupor,Staring,Posturing/catalepsy,Grimacing,Stereotypy,Negativism,Waxy flexibility,Withdrawal | 97.8 | 96.2 | 99.1 |
| 8 | Immobility/stupor,Staring,Posturing/catalepsy,Mannerisms,Verbigeration,Rigidity,Waxy flexibility,Withdrawal | 97.8 | 96.4 | 99.1 |
| 8 | Immobility/stupor,Staring,Posturing/catalepsy,Stereotypy,Mannerisms,Verbigeration,Rigidity,Waxy flexibility | 97.8 | 96.2 | 99.1 |
| 8 | Mutism,Staring,Grimacing,Echopraxia/echolalia,Stereotypy,Verbigeration,Waxy flexibility,Withdrawal | 97.8 | 96.2 | 99.1 |
| 8 | Mutism,Staring,Posturing/catalepsy,Echopraxia/echolalia,Stereotypy,Mannerisms,Verbigeration,Waxy flexibility | 97.8 | 96.4 | 99.1 |
| 8 | Mutism,Staring,Posturing/catalepsy,Echopraxia/echolalia,Verbigeration,Rigidity,Waxy flexibility,Withdrawal | 97.8 | 96.2 | 99.1 |
| 8 | Mutism,Staring,Posturing/catalepsy,Grimacing,Echopraxia/echolalia,Rigidity,Negativism,Waxy flexibility | 97.8 | 96.4 | 99.1 |
| 8 | Mutism,Staring,Posturing/catalepsy,Grimacing,Echopraxia/echolalia,Rigidity,Waxy flexibility,Withdrawal | 97.8 | 96.4 | 99.1 |
| 8 | Mutism,Staring,Posturing/catalepsy,Grimacing,Echopraxia/echolalia,Stereotypy,Verbigeration,Waxy flexibility | 97.8 | 96.2 | 99.1 |
| 8 | Mutism,Staring,Posturing/catalepsy,Grimacing,Echopraxia/echolalia,Verbigeration,Rigidity,Waxy flexibility | 97.8 | 96.2 | 99.1 |
| 8 | Mutism,Staring,Posturing/catalepsy,Grimacing,Rigidity,Negativism,Waxy flexibility,Withdrawal | 97.8 | 96.4 | 99.1 |
| 8 | Mutism,Staring,Posturing/catalepsy,Grimacing,Stereotypy,Rigidity,Negativism,Waxy flexibility | 97.8 | 96.4 | 99.1 |
| 8 | Mutism,Staring,Posturing/catalepsy,Grimacing,Verbigeration,Rigidity,Negativism,Waxy flexibility | 97.8 | 96.2 | 99.1 |
| 8 | Mutism,Staring,Posturing/catalepsy,Grimacing,Verbigeration,Rigidity,Waxy flexibility,Withdrawal | 97.8 | 96.4 | 99.1 |
| 8 | Staring,Grimacing,Echopraxia/echolalia,Stereotypy,Mannerisms,Verbigeration,Rigidity,Negativism | 97.8 | 96.2 | 99.1 |
| 8 | Staring,Grimacing,Echopraxia/echolalia,Verbigeration,Rigidity,Negativism,Waxy flexibility,Withdrawal | 97.8 | 96.4 | 99.1 |
| 8 | Staring,Posturing/catalepsy,Echopraxia/echolalia,Stereotypy,Mannerisms,Rigidity,Negativism,Waxy flexibility | 97.8 | 96.2 | 99.1 |
| 8 | Staring,Posturing/catalepsy,Echopraxia/echolalia,Stereotypy,Mannerisms,Rigidity,Waxy flexibility,Withdrawal | 97.8 | 96.4 | 99.1 |
| 8 | Staring,Posturing/catalepsy,Echopraxia/echolalia,Verbigeration,Rigidity,Negativism,Waxy flexibility,Withdrawal | 97.8 | 96.4 | 99.1 |
| 8 | Staring,Posturing/catalepsy,Grimacing,Echopraxia/echolalia,Mannerisms,Verbigeration,Rigidity,Negativism | 97.8 | 96.2 | 99.1 |
| 8 | Staring,Posturing/catalepsy,Grimacing,Echopraxia/echolalia,Rigidity,Negativism,Waxy flexibility,Withdrawal | 97.8 | 96.2 | 99.1 |
| 8 | Staring,Posturing/catalepsy,Grimacing,Echopraxia/echolalia,Stereotypy,Mannerisms,Waxy flexibility,Withdrawal | 97.8 | 96.4 | 99.1 |
| 8 | Staring,Posturing/catalepsy,Grimacing,Echopraxia/echolalia,Stereotypy,Verbigeration,Negativism,Withdrawal | 97.8 | 96.2 | 99.1 |
| 8 | Staring,Posturing/catalepsy,Grimacing,Stereotypy,Rigidity,Negativism,Waxy flexibility,Withdrawal | 97.8 | 96.2 | 99.1 |
| 8 | Staring,Posturing/catalepsy,Grimacing,Stereotypy,Verbigeration,Negativism,Waxy flexibility,Withdrawal | 97.8 | 96.4 | 99.1 |
| 8 | Staring,Posturing/catalepsy,Stereotypy,Mannerisms,Verbigeration,Rigidity,Negativism,Waxy flexibility | 97.8 | 96.4 | 99.1 |
| 8 | Staring,Posturing/catalepsy,Stereotypy,Mannerisms,Verbigeration,Rigidity,Waxy flexibility,Withdrawal | 97.8 | 96.2 | 99.1 |
| 8 | Excitement,Immobility/stupor,Grimacing,Stereotypy,Mannerisms,Rigidity,Waxy flexibility,Withdrawal | 97.7 | 96.2 | 98.9 |
| 8 | Excitement,Immobility/stupor,Mutism,Grimacing,Echopraxia/echolalia,Stereotypy,Mannerisms,Rigidity | 97.7 | 96.2 | 99.1 |
| 8 | Excitement,Immobility/stupor,Mutism,Posturing/catalepsy,Grimacing,Echopraxia/echolalia,Mannerisms,Withdrawal | 97.7 | 96.2 | 99.1 |
| 8 | Excitement,Immobility/stupor,Mutism,Posturing/catalepsy,Grimacing,Echopraxia/echolalia,Stereotypy,Withdrawal | 97.7 | 96.2 | 99.1 |
| 8 | Excitement,Immobility/stupor,Posturing/catalepsy,Echopraxia/echolalia,Stereotypy,Verbigeration,Rigidity,Waxy flexibility | 97.7 | 96.2 | 99.1 |
| 8 | Excitement,Immobility/stupor,Posturing/catalepsy,Grimacing,Echopraxia/echolalia,Stereotypy,Verbigeration,Rigidity | 97.7 | 96.2 | 99.1 |
| 8 | Excitement,Immobility/stupor,Staring,Grimacing,Echopraxia/echolalia,Stereotypy,Verbigeration,Withdrawal | 97.7 | 96.2 | 99.1 |
| 8 | Excitement,Immobility/stupor,Staring,Posturing/catalepsy,Grimacing,Echopraxia/echolalia,Stereotypy,Verbigeration | 97.7 | 96.2 | 99.1 |
| 8 | Excitement,Staring,Posturing/catalepsy,Grimacing,Stereotypy,Mannerisms,Waxy flexibility,Withdrawal | 97.7 | 96.2 | 99.1 |
| 8 | Excitement,Staring,Posturing/catalepsy,Stereotypy,Mannerisms,Verbigeration,Negativism,Waxy flexibility | 97.7 | 96.2 | 99.1 |
| 8 | Immobility/stupor,Mutism,Staring,Grimacing,Stereotypy,Mannerisms,Verbigeration,Withdrawal | 97.7 | 96.2 | 99.1 |
| 8 | Immobility/stupor,Mutism,Staring,Posturing/catalepsy,Echopraxia/echolalia,Verbigeration,Negativism,Waxy flexibility | 97.7 | 96.4 | 98.9 |
| 8 | Immobility/stupor,Mutism,Staring,Posturing/catalepsy,Grimacing,Rigidity,Negativism,Withdrawal | 97.7 | 96.2 | 99.1 |
| 8 | Immobility/stupor,Mutism,Staring,Posturing/catalepsy,Verbigeration,Negativism,Waxy flexibility,Withdrawal | 97.7 | 96.2 | 99.1 |
| 8 | Immobility/stupor,Posturing/catalepsy,Echopraxia/echolalia,Stereotypy,Mannerisms,Rigidity,Negativism,Waxy flexibility | 97.7 | 96.2 | 99.1 |
| 8 | Immobility/stupor,Staring,Grimacing,Echopraxia/echolalia,Mannerisms,Verbigeration,Rigidity,Waxy flexibility | 97.7 | 96.2 | 99.1 |
| 8 | Immobility/stupor,Staring,Posturing/catalepsy,Echopraxia/echolalia,Mannerisms,Rigidity,Waxy flexibility,Withdrawal | 97.7 | 96.2 | 99.1 |
| 8 | Immobility/stupor,Staring,Posturing/catalepsy,Echopraxia/echolalia,Verbigeration,Rigidity,Negativism,Waxy flexibility | 97.7 | 96.2 | 99.1 |
| 8 | Immobility/stupor,Staring,Posturing/catalepsy,Grimacing,Stereotypy,Mannerisms,Waxy flexibility,Withdrawal | 97.7 | 96.2 | 99.1 |
| 8 | Staring,Posturing/catalepsy,Echopraxia/echolalia,Stereotypy,Mannerisms,Verbigeration,Rigidity,Withdrawal | 97.7 | 96.4 | 99.1 |
| 8 | Excitement,Immobility/stupor,Grimacing,Echopraxia/echolalia,Stereotypy,Negativism,Waxy flexibility,Withdrawal | 97.6 | 96.0 | 98.9 |
| 8 | Immobility/stupor,Mutism,Posturing/catalepsy,Echopraxia/echolalia,Verbigeration,Rigidity,Negativism,Withdrawal | 97.6 | 96.0 | 98.9 |
| 8 | Excitement,Immobility/stupor,Echopraxia/echolalia,Stereotypy,Mannerisms,Negativism,Waxy flexibility,Withdrawal | 97.5 | 96.0 | 98.9 |
| 8 | Excitement,Immobility/stupor,Echopraxia/echolalia,Stereotypy,Mannerisms,Verbigeration,Rigidity,Waxy flexibility | 97.5 | 96.0 | 98.9 |
| 8 | Excitement,Immobility/stupor,Grimacing,Echopraxia/echolalia,Mannerisms,Verbigeration,Rigidity,Waxy flexibility | 97.5 | 96.0 | 98.9 |
| 8 | Excitement,Immobility/stupor,Grimacing,Echopraxia/echolalia,Stereotypy,Mannerisms,Negativism,Withdrawal | 97.5 | 96.0 | 98.9 |
| 8 | Excitement,Immobility/stupor,Grimacing,Echopraxia/echolalia,Stereotypy,Verbigeration,Rigidity,Waxy flexibility | 97.5 | 96.0 | 98.9 |
| 8 | Excitement,Immobility/stupor,Mutism,Echopraxia/echolalia,Mannerisms,Verbigeration,Negativism,Withdrawal | 97.5 | 96.0 | 98.9 |
| 8 | Excitement,Immobility/stupor,Mutism,Grimacing,Echopraxia/echolalia,Stereotypy,Negativism,Waxy flexibility | 97.5 | 96.0 | 98.9 |
| 8 | Excitement,Immobility/stupor,Mutism,Grimacing,Echopraxia/echolalia,Verbigeration,Negativism,Waxy flexibility | 97.5 | 96.0 | 98.9 |
| 8 | Excitement,Immobility/stupor,Mutism,Grimacing,Stereotypy,Negativism,Waxy flexibility,Withdrawal | 97.5 | 96.0 | 98.9 |
| 8 | Excitement,Immobility/stupor,Mutism,Grimacing,Stereotypy,Verbigeration,Negativism,Waxy flexibility | 97.5 | 96.0 | 98.9 |
| 8 | Excitement,Immobility/stupor,Mutism,Posturing/catalepsy,Echopraxia/echolalia,Mannerisms,Negativism,Waxy flexibility | 97.5 | 96.0 | 98.9 |
| 8 | Excitement,Immobility/stupor,Mutism,Posturing/catalepsy,Echopraxia/echolalia,Mannerisms,Verbigeration,Withdrawal | 97.5 | 96.0 | 98.9 |
| 8 | Excitement,Immobility/stupor,Mutism,Posturing/catalepsy,Echopraxia/echolalia,Stereotypy,Mannerisms,Negativism | 97.5 | 96.0 | 98.9 |
| 8 | Excitement,Immobility/stupor,Mutism,Posturing/catalepsy,Echopraxia/echolalia,Stereotypy,Waxy flexibility,Withdrawal | 97.5 | 96.0 | 98.9 |
| 8 | Excitement,Immobility/stupor,Mutism,Posturing/catalepsy,Grimacing,Echopraxia/echolalia,Verbigeration,Waxy flexibility | 97.5 | 96.0 | 98.9 |
| 8 | Excitement,Immobility/stupor,Mutism,Posturing/catalepsy,Grimacing,Mannerisms,Verbigeration,Withdrawal | 97.5 | 96.0 | 98.9 |
| 8 | Excitement,Immobility/stupor,Mutism,Posturing/catalepsy,Grimacing,Stereotypy,Mannerisms,Negativism | 97.5 | 96.0 | 98.9 |
| 8 | Excitement,Immobility/stupor,Mutism,Posturing/catalepsy,Grimacing,Stereotypy,Verbigeration,Withdrawal | 97.5 | 96.0 | 98.9 |
| 8 | Excitement,Immobility/stupor,Mutism,Posturing/catalepsy,Grimacing,Verbigeration,Waxy flexibility,Withdrawal | 97.5 | 96.0 | 98.9 |
| 8 | Excitement,Immobility/stupor,Mutism,Posturing/catalepsy,Stereotypy,Verbigeration,Waxy flexibility,Withdrawal | 97.5 | 96.0 | 98.9 |
| 8 | Excitement,Immobility/stupor,Mutism,Staring,Grimacing,Echopraxia/echolalia,Mannerisms,Verbigeration | 97.5 | 96.0 | 98.9 |
| 8 | Excitement,Immobility/stupor,Mutism,Staring,Grimacing,Stereotypy,Mannerisms,Verbigeration | 97.5 | 96.0 | 98.9 |
| 8 | Excitement,Immobility/stupor,Posturing/catalepsy,Grimacing,Echopraxia/echolalia,Mannerisms,Negativism,Waxy flexibility | 97.5 | 96.0 | 98.9 |
| 8 | Excitement,Immobility/stupor,Posturing/catalepsy,Grimacing,Echopraxia/echolalia,Mannerisms,Verbigeration,Negativism | 97.5 | 96.0 | 98.9 |
| 8 | Excitement,Immobility/stupor,Posturing/catalepsy,Grimacing,Echopraxia/echolalia,Stereotypy,Mannerisms,Rigidity | 97.5 | 96.0 | 98.9 |
| 8 | Excitement,Immobility/stupor,Posturing/catalepsy,Grimacing,Echopraxia/echolalia,Stereotypy,Rigidity,Waxy flexibility | 97.5 | 96.0 | 98.9 |
| 8 | Excitement,Immobility/stupor,Posturing/catalepsy,Grimacing,Stereotypy,Mannerisms,Verbigeration,Rigidity | 97.5 | 96.0 | 98.9 |
| 8 | Excitement,Immobility/stupor,Posturing/catalepsy,Grimacing,Stereotypy,Negativism,Waxy flexibility,Withdrawal | 97.5 | 96.0 | 98.9 |
| 8 | Excitement,Immobility/stupor,Posturing/catalepsy,Grimacing,Stereotypy,Verbigeration,Negativism,Waxy flexibility | 97.5 | 96.0 | 98.9 |
| 8 | Excitement,Immobility/stupor,Posturing/catalepsy,Grimacing,Stereotypy,Verbigeration,Rigidity,Waxy flexibility | 97.5 | 96.0 | 98.9 |
| 8 | Excitement,Immobility/stupor,Posturing/catalepsy,Stereotypy,Mannerisms,Negativism,Waxy flexibility,Withdrawal | 97.5 | 96.0 | 98.9 |
| 8 | Excitement,Staring,Echopraxia/echolalia,Stereotypy,Mannerisms,Verbigeration,Waxy flexibility,Withdrawal | 97.5 | 96.0 | 98.9 |
| 8 | Excitement,Staring,Grimacing,Echopraxia/echolalia,Mannerisms,Verbigeration,Rigidity,Negativism | 97.5 | 96.0 | 98.9 |
| 8 | Excitement,Staring,Grimacing,Echopraxia/echolalia,Stereotypy,Mannerisms,Verbigeration,Rigidity | 97.5 | 96.0 | 98.9 |
| 8 | Excitement,Staring,Grimacing,Echopraxia/echolalia,Stereotypy,Mannerisms,Verbigeration,Withdrawal | 97.5 | 96.0 | 98.9 |
| 8 | Excitement,Staring,Grimacing,Stereotypy,Mannerisms,Verbigeration,Waxy flexibility,Withdrawal | 97.5 | 96.0 | 98.9 |
| 8 | Excitement,Staring,Posturing/catalepsy,Echopraxia/echolalia,Stereotypy,Mannerisms,Verbigeration,Negativism | 97.5 | 96.0 | 98.9 |
| 8 | Excitement,Staring,Posturing/catalepsy,Echopraxia/echolalia,Stereotypy,Mannerisms,Verbigeration,Rigidity | 97.5 | 96.0 | 98.9 |
| 8 | Excitement,Staring,Posturing/catalepsy,Echopraxia/echolalia,Stereotypy,Mannerisms,Verbigeration,Withdrawal | 97.5 | 96.0 | 98.9 |
| 8 | Excitement,Staring,Posturing/catalepsy,Echopraxia/echolalia,Stereotypy,Verbigeration,Negativism,Waxy flexibility | 97.5 | 96.0 | 98.9 |
| 8 | Excitement,Staring,Posturing/catalepsy,Grimacing,Echopraxia/echolalia,Mannerisms,Negativism,Waxy flexibility | 97.5 | 96.0 | 98.9 |
| 8 | Excitement,Staring,Posturing/catalepsy,Grimacing,Echopraxia/echolalia,Mannerisms,Verbigeration,Negativism | 97.5 | 96.0 | 98.9 |
| 8 | Excitement,Staring,Posturing/catalepsy,Grimacing,Echopraxia/echolalia,Stereotypy,Mannerisms,Negativism | 97.5 | 96.0 | 98.9 |
| 8 | Excitement,Staring,Posturing/catalepsy,Grimacing,Echopraxia/echolalia,Stereotypy,Rigidity,Waxy flexibility | 97.5 | 96.0 | 98.9 |
| 8 | Excitement,Staring,Posturing/catalepsy,Grimacing,Echopraxia/echolalia,Stereotypy,Verbigeration,Withdrawal | 97.5 | 96.0 | 98.9 |
| 8 | Excitement,Staring,Posturing/catalepsy,Grimacing,Echopraxia/echolalia,Verbigeration,Rigidity,Waxy flexibility | 97.5 | 96.0 | 98.9 |
| 8 | Excitement,Staring,Posturing/catalepsy,Grimacing,Mannerisms,Verbigeration,Negativism,Waxy flexibility | 97.5 | 96.0 | 98.9 |
| 8 | Excitement,Staring,Posturing/catalepsy,Grimacing,Stereotypy,Mannerisms,Rigidity,Waxy flexibility | 97.5 | 96.0 | 98.9 |
| 8 | Excitement,Staring,Posturing/catalepsy,Grimacing,Stereotypy,Mannerisms,Verbigeration,Negativism | 97.5 | 96.0 | 98.9 |
| 8 | Excitement,Staring,Posturing/catalepsy,Grimacing,Stereotypy,Rigidity,Negativism,Waxy flexibility | 97.5 | 96.0 | 98.9 |
| 8 | Excitement,Staring,Posturing/catalepsy,Grimacing,Stereotypy,Verbigeration,Rigidity,Waxy flexibility | 97.5 | 96.0 | 98.9 |
| 8 | Excitement,Staring,Posturing/catalepsy,Grimacing,Stereotypy,Verbigeration,Waxy flexibility,Withdrawal | 97.5 | 96.0 | 98.9 |
| 8 | Excitement,Staring,Posturing/catalepsy,Grimacing,Verbigeration,Rigidity,Negativism,Waxy flexibility | 97.5 | 96.0 | 98.9 |
| 8 | Immobility/stupor,Grimacing,Echopraxia/echolalia,Stereotypy,Mannerisms,Negativism,Waxy flexibility,Withdrawal | 97.5 | 96.0 | 98.9 |
| 8 | Immobility/stupor,Grimacing,Echopraxia/echolalia,Stereotypy,Mannerisms,Verbigeration,Negativism,Withdrawal | 97.5 | 96.0 | 98.9 |
| 8 | Immobility/stupor,Grimacing,Echopraxia/echolalia,Stereotypy,Verbigeration,Rigidity,Negativism,Waxy flexibility | 97.5 | 96.0 | 98.9 |
| 8 | Immobility/stupor,Grimacing,Echopraxia/echolalia,Verbigeration,Rigidity,Negativism,Waxy flexibility,Withdrawal | 97.5 | 96.0 | 98.9 |
| 8 | Immobility/stupor,Mutism,Echopraxia/echolalia,Stereotypy,Mannerisms,Verbigeration,Negativism,Withdrawal | 97.5 | 96.0 | 98.9 |
| 8 | Immobility/stupor,Mutism,Echopraxia/echolalia,Stereotypy,Rigidity,Negativism,Waxy flexibility,Withdrawal | 97.5 | 96.0 | 98.9 |
| 8 | Immobility/stupor,Mutism,Echopraxia/echolalia,Stereotypy,Verbigeration,Rigidity,Negativism,Waxy flexibility | 97.5 | 96.0 | 98.9 |
| 8 | Immobility/stupor,Mutism,Echopraxia/echolalia,Verbigeration,Rigidity,Negativism,Waxy flexibility,Withdrawal | 97.5 | 96.0 | 98.9 |
| 8 | Immobility/stupor,Mutism,Grimacing,Echopraxia/echolalia,Mannerisms,Rigidity,Waxy flexibility,Withdrawal | 97.5 | 96.0 | 98.9 |
| 8 | Immobility/stupor,Mutism,Grimacing,Echopraxia/echolalia,Mannerisms,Verbigeration,Rigidity,Withdrawal | 97.5 | 96.0 | 98.9 |
| 8 | Immobility/stupor,Mutism,Grimacing,Echopraxia/echolalia,Stereotypy,Mannerisms,Negativism,Withdrawal | 97.5 | 96.0 | 98.9 |
| 8 | Immobility/stupor,Mutism,Grimacing,Echopraxia/echolalia,Verbigeration,Rigidity,Negativism,Waxy flexibility | 97.5 | 96.0 | 98.9 |
| 8 | Immobility/stupor,Mutism,Grimacing,Stereotypy,Mannerisms,Rigidity,Waxy flexibility,Withdrawal | 97.5 | 96.0 | 98.9 |
| 8 | Immobility/stupor,Mutism,Grimacing,Stereotypy,Mannerisms,Verbigeration,Rigidity,Withdrawal | 97.5 | 96.0 | 98.9 |
| 8 | Immobility/stupor,Mutism,Grimacing,Stereotypy,Rigidity,Negativism,Waxy flexibility,Withdrawal | 97.5 | 96.0 | 98.9 |
| 8 | Immobility/stupor,Mutism,Grimacing,Verbigeration,Rigidity,Negativism,Waxy flexibility,Withdrawal | 97.5 | 96.0 | 98.9 |
| 8 | Immobility/stupor,Mutism,Posturing/catalepsy,Echopraxia/echolalia,Mannerisms,Negativism,Waxy flexibility,Withdrawal | 97.5 | 96.0 | 98.9 |
| 8 | Immobility/stupor,Mutism,Posturing/catalepsy,Echopraxia/echolalia,Verbigeration,Rigidity,Negativism,Waxy flexibility | 97.5 | 96.0 | 98.9 |
| 8 | Immobility/stupor,Mutism,Posturing/catalepsy,Grimacing,Echopraxia/echolalia,Mannerisms,Verbigeration,Negativism | 97.5 | 96.0 | 98.9 |
| 8 | Immobility/stupor,Mutism,Posturing/catalepsy,Grimacing,Echopraxia/echolalia,Rigidity,Negativism,Withdrawal | 97.5 | 96.0 | 98.9 |
| 8 | Immobility/stupor,Mutism,Posturing/catalepsy,Grimacing,Mannerisms,Negativism,Waxy flexibility,Withdrawal | 97.5 | 96.0 | 98.9 |
| 8 | Immobility/stupor,Mutism,Posturing/catalepsy,Grimacing,Mannerisms,Verbigeration,Rigidity,Withdrawal | 97.5 | 96.0 | 98.9 |
| 8 | Immobility/stupor,Mutism,Posturing/catalepsy,Grimacing,Stereotypy,Mannerisms,Rigidity,Withdrawal | 97.5 | 96.0 | 98.9 |
| 8 | Immobility/stupor,Mutism,Posturing/catalepsy,Grimacing,Verbigeration,Rigidity,Negativism,Withdrawal | 97.5 | 96.0 | 98.9 |
| 8 | Immobility/stupor,Mutism,Staring,Echopraxia/echolalia,Stereotypy,Mannerisms,Waxy flexibility,Withdrawal | 97.5 | 96.0 | 98.9 |
| 8 | Immobility/stupor,Mutism,Staring,Echopraxia/echolalia,Stereotypy,Verbigeration,Rigidity,Waxy flexibility | 97.5 | 96.0 | 98.9 |
| 8 | Immobility/stupor,Mutism,Staring,Echopraxia/echolalia,Verbigeration,Negativism,Waxy flexibility,Withdrawal | 97.5 | 96.0 | 98.9 |
| 8 | Immobility/stupor,Mutism,Staring,Grimacing,Echopraxia/echolalia,Mannerisms,Verbigeration,Withdrawal | 97.5 | 96.0 | 98.9 |
| 8 | Immobility/stupor,Mutism,Staring,Grimacing,Echopraxia/echolalia,Stereotypy,Negativism,Waxy flexibility | 97.5 | 96.0 | 98.9 |
| 8 | Immobility/stupor,Mutism,Staring,Grimacing,Echopraxia/echolalia,Verbigeration,Rigidity,Waxy flexibility | 97.5 | 96.0 | 98.9 |
| 8 | Immobility/stupor,Mutism,Staring,Grimacing,Mannerisms,Verbigeration,Waxy flexibility,Withdrawal | 97.5 | 96.0 | 98.9 |
| 8 | Immobility/stupor,Mutism,Staring,Grimacing,Stereotypy,Mannerisms,Verbigeration,Waxy flexibility | 97.5 | 96.0 | 98.9 |
| 8 | Immobility/stupor,Mutism,Staring,Grimacing,Stereotypy,Negativism,Waxy flexibility,Withdrawal | 97.5 | 96.0 | 98.9 |
| 8 | Immobility/stupor,Mutism,Staring,Grimacing,Verbigeration,Rigidity,Waxy flexibility,Withdrawal | 97.5 | 96.0 | 98.9 |
| 8 | Immobility/stupor,Mutism,Staring,Posturing/catalepsy,Echopraxia/echolalia,Rigidity,Negativism,Waxy flexibility | 97.5 | 96.0 | 98.9 |
| 8 | Immobility/stupor,Mutism,Staring,Posturing/catalepsy,Grimacing,Echopraxia/echolalia,Mannerisms,Waxy flexibility | 97.5 | 96.0 | 98.9 |
| 8 | Immobility/stupor,Mutism,Staring,Posturing/catalepsy,Grimacing,Echopraxia/echolalia,Stereotypy,Rigidity | 97.5 | 96.0 | 98.9 |
| 8 | Immobility/stupor,Mutism,Staring,Posturing/catalepsy,Grimacing,Mannerisms,Rigidity,Waxy flexibility | 97.5 | 96.0 | 98.9 |
| 8 | Immobility/stupor,Mutism,Staring,Posturing/catalepsy,Grimacing,Mannerisms,Verbigeration,Waxy flexibility | 97.5 | 96.0 | 98.9 |
| 8 | Immobility/stupor,Mutism,Staring,Posturing/catalepsy,Grimacing,Negativism,Waxy flexibility,Withdrawal | 97.5 | 96.0 | 98.9 |
| 8 | Immobility/stupor,Mutism,Staring,Posturing/catalepsy,Grimacing,Stereotypy,Negativism,Waxy flexibility | 97.5 | 96.0 | 98.9 |
| 8 | Immobility/stupor,Mutism,Staring,Posturing/catalepsy,Grimacing,Verbigeration,Negativism,Waxy flexibility | 97.5 | 96.0 | 98.9 |
| 8 | Immobility/stupor,Mutism,Staring,Posturing/catalepsy,Mannerisms,Verbigeration,Waxy flexibility,Withdrawal | 97.5 | 96.0 | 98.9 |
| 8 | Immobility/stupor,Mutism,Staring,Posturing/catalepsy,Rigidity,Negativism,Waxy flexibility,Withdrawal | 97.5 | 96.0 | 98.9 |
| 8 | Immobility/stupor,Mutism,Staring,Posturing/catalepsy,Stereotypy,Rigidity,Waxy flexibility,Withdrawal | 97.5 | 96.0 | 98.9 |
| 8 | Immobility/stupor,Mutism,Staring,Posturing/catalepsy,Verbigeration,Rigidity,Negativism,Waxy flexibility | 97.5 | 96.0 | 98.9 |
| 8 | Immobility/stupor,Posturing/catalepsy,Echopraxia/echolalia,Stereotypy,Mannerisms,Negativism,Waxy flexibility,Withdrawal | 97.5 | 96.0 | 98.9 |
| 8 | Immobility/stupor,Posturing/catalepsy,Echopraxia/echolalia,Stereotypy,Rigidity,Negativism,Waxy flexibility,Withdrawal | 97.5 | 96.0 | 98.9 |
| 8 | Immobility/stupor,Posturing/catalepsy,Echopraxia/echolalia,Verbigeration,Rigidity,Negativism,Waxy flexibility,Withdrawal | 97.5 | 96.0 | 98.9 |
| 8 | Immobility/stupor,Posturing/catalepsy,Grimacing,Echopraxia/echolalia,Mannerisms,Verbigeration,Negativism,Withdrawal | 97.5 | 96.0 | 98.9 |
| 8 | Immobility/stupor,Posturing/catalepsy,Grimacing,Echopraxia/echolalia,Rigidity,Negativism,Waxy flexibility,Withdrawal | 97.5 | 96.0 | 98.9 |
| 8 | Immobility/stupor,Posturing/catalepsy,Grimacing,Echopraxia/echolalia,Stereotypy,Verbigeration,Rigidity,Negativism | 97.5 | 96.0 | 98.9 |
| 8 | Immobility/stupor,Posturing/catalepsy,Grimacing,Echopraxia/echolalia,Verbigeration,Rigidity,Negativism,Waxy flexibility | 97.5 | 96.0 | 98.9 |
| 8 | Immobility/stupor,Posturing/catalepsy,Grimacing,Stereotypy,Rigidity,Negativism,Waxy flexibility,Withdrawal | 97.5 | 96.0 | 98.9 |
| 8 | Immobility/stupor,Posturing/catalepsy,Grimacing,Verbigeration,Rigidity,Negativism,Waxy flexibility,Withdrawal | 97.5 | 96.0 | 98.9 |
| 8 | Immobility/stupor,Posturing/catalepsy,Stereotypy,Mannerisms,Verbigeration,Negativism,Waxy flexibility,Withdrawal | 97.5 | 96.0 | 98.9 |
| 8 | Immobility/stupor,Staring,Echopraxia/echolalia,Mannerisms,Verbigeration,Rigidity,Waxy flexibility,Withdrawal | 97.5 | 96.0 | 98.9 |
| 8 | Immobility/stupor,Staring,Echopraxia/echolalia,Stereotypy,Mannerisms,Verbigeration,Rigidity,Waxy flexibility | 97.5 | 96.0 | 98.9 |
| 8 | Immobility/stupor,Staring,Grimacing,Echopraxia/echolalia,Stereotypy,Mannerisms,Verbigeration,Withdrawal | 97.5 | 96.0 | 98.9 |
| 8 | Immobility/stupor,Staring,Grimacing,Echopraxia/echolalia,Stereotypy,Verbigeration,Rigidity,Waxy flexibility | 97.5 | 96.0 | 98.9 |
| 8 | Immobility/stupor,Staring,Grimacing,Stereotypy,Mannerisms,Verbigeration,Waxy flexibility,Withdrawal | 97.5 | 96.0 | 98.9 |
| 8 | Immobility/stupor,Staring,Posturing/catalepsy,Echopraxia/echolalia,Stereotypy,Verbigeration,Rigidity,Withdrawal | 97.5 | 96.0 | 98.9 |
| 8 | Immobility/stupor,Staring,Posturing/catalepsy,Echopraxia/echolalia,Verbigeration,Negativism,Waxy flexibility,Withdrawal | 97.5 | 96.0 | 98.9 |
| 8 | Immobility/stupor,Staring,Posturing/catalepsy,Grimacing,Echopraxia/echolalia,Mannerisms,Verbigeration,Withdrawal | 97.5 | 96.0 | 98.9 |
| 8 | Immobility/stupor,Staring,Posturing/catalepsy,Grimacing,Echopraxia/echolalia,Mannerisms,Waxy flexibility,Withdrawal | 97.5 | 96.0 | 98.9 |
| 8 | Immobility/stupor,Staring,Posturing/catalepsy,Grimacing,Echopraxia/echolalia,Stereotypy,Negativism,Waxy flexibility | 97.5 | 96.0 | 98.9 |
| 8 | Immobility/stupor,Staring,Posturing/catalepsy,Grimacing,Echopraxia/echolalia,Stereotypy,Verbigeration,Rigidity | 97.5 | 96.0 | 98.9 |
| 8 | Immobility/stupor,Staring,Posturing/catalepsy,Grimacing,Echopraxia/echolalia,Verbigeration,Negativism,Waxy flexibility | 97.5 | 96.0 | 98.9 |
| 8 | Immobility/stupor,Staring,Posturing/catalepsy,Grimacing,Mannerisms,Verbigeration,Rigidity,Waxy flexibility | 97.5 | 96.0 | 98.9 |
| 8 | Immobility/stupor,Staring,Posturing/catalepsy,Grimacing,Rigidity,Negativism,Waxy flexibility,Withdrawal | 97.5 | 96.0 | 98.9 |
| 8 | Immobility/stupor,Staring,Posturing/catalepsy,Grimacing,Stereotypy,Rigidity,Negativism,Waxy flexibility | 97.5 | 96.0 | 98.9 |
| 8 | Immobility/stupor,Staring,Posturing/catalepsy,Grimacing,Stereotypy,Verbigeration,Rigidity,Waxy flexibility | 97.5 | 96.0 | 98.9 |
| 8 | Immobility/stupor,Staring,Posturing/catalepsy,Grimacing,Verbigeration,Negativism,Waxy flexibility,Withdrawal | 97.5 | 96.0 | 98.9 |
| 8 | Immobility/stupor,Staring,Posturing/catalepsy,Grimacing,Verbigeration,Rigidity,Negativism,Waxy flexibility | 97.5 | 96.0 | 98.9 |
| 8 | Immobility/stupor,Staring,Posturing/catalepsy,Stereotypy,Verbigeration,Rigidity,Waxy flexibility,Withdrawal | 97.5 | 96.0 | 98.9 |
| 8 | Mutism,Staring,Echopraxia/echolalia,Stereotypy,Mannerisms,Verbigeration,Waxy flexibility,Withdrawal | 97.5 | 96.0 | 98.9 |
| 8 | Mutism,Staring,Posturing/catalepsy,Grimacing,Echopraxia/echolalia,Stereotypy,Rigidity,Waxy flexibility | 97.5 | 96.0 | 98.9 |
| 8 | Mutism,Staring,Posturing/catalepsy,Grimacing,Echopraxia/echolalia,Verbigeration,Waxy flexibility,Withdrawal | 97.5 | 96.0 | 98.9 |
| 8 | Staring,Echopraxia/echolalia,Stereotypy,Mannerisms,Verbigeration,Rigidity,Waxy flexibility,Withdrawal | 97.5 | 96.0 | 98.9 |
| 8 | Staring,Grimacing,Echopraxia/echolalia,Mannerisms,Verbigeration,Rigidity,Negativism,Waxy flexibility | 97.5 | 96.0 | 98.9 |
| 8 | Staring,Grimacing,Echopraxia/echolalia,Stereotypy,Verbigeration,Rigidity,Waxy flexibility,Withdrawal | 97.5 | 96.0 | 98.9 |
| 8 | Staring,Posturing/catalepsy,Echopraxia/echolalia,Mannerisms,Verbigeration,Rigidity,Negativism,Waxy flexibility | 97.5 | 96.0 | 98.9 |
| 8 | Staring,Posturing/catalepsy,Grimacing,Echopraxia/echolalia,Mannerisms,Rigidity,Negativism,Waxy flexibility | 97.5 | 96.0 | 98.9 |
| 8 | Staring,Posturing/catalepsy,Grimacing,Echopraxia/echolalia,Stereotypy,Mannerisms,Verbigeration,Withdrawal | 97.5 | 96.0 | 98.9 |
| 8 | Staring,Posturing/catalepsy,Grimacing,Echopraxia/echolalia,Stereotypy,Negativism,Waxy flexibility,Withdrawal | 97.5 | 96.0 | 98.9 |
| 8 | Staring,Posturing/catalepsy,Grimacing,Echopraxia/echolalia,Stereotypy,Verbigeration,Rigidity,Withdrawal | 97.5 | 96.0 | 98.9 |
| 8 | Staring,Posturing/catalepsy,Grimacing,Stereotypy,Mannerisms,Rigidity,Negativism,Waxy flexibility | 97.5 | 96.0 | 98.9 |
| 8 | Staring,Posturing/catalepsy,Grimacing,Stereotypy,Mannerisms,Verbigeration,Waxy flexibility,Withdrawal | 97.5 | 96.0 | 98.9 |
| 8 | Staring,Posturing/catalepsy,Grimacing,Stereotypy,Verbigeration,Rigidity,Waxy flexibility,Withdrawal | 97.5 | 96.0 | 98.9 |
| 8 | Staring,Posturing/catalepsy,Grimacing,Verbigeration,Rigidity,Negativism,Waxy flexibility,Withdrawal | 97.5 | 96.0 | 98.9 |
| 8 | Excitement,Immobility/stupor,Echopraxia/echolalia,Mannerisms,Verbigeration,Negativism,Waxy flexibility,Withdrawal | 97.3 | 95.7 | 98.7 |
| 8 | Excitement,Immobility/stupor,Grimacing,Echopraxia/echolalia,Mannerisms,Negativism,Waxy flexibility,Withdrawal | 97.3 | 95.7 | 98.7 |
| 8 | Excitement,Immobility/stupor,Grimacing,Echopraxia/echolalia,Mannerisms,Verbigeration,Negativism,Withdrawal | 97.3 | 95.7 | 98.7 |
| 8 | Excitement,Immobility/stupor,Grimacing,Echopraxia/echolalia,Stereotypy,Mannerisms,Rigidity,Waxy flexibility | 97.3 | 95.7 | 98.7 |
| 8 | Excitement,Immobility/stupor,Grimacing,Echopraxia/echolalia,Verbigeration,Negativism,Waxy flexibility,Withdrawal | 97.3 | 95.7 | 98.7 |
| 8 | Excitement,Immobility/stupor,Grimacing,Stereotypy,Mannerisms,Verbigeration,Negativism,Withdrawal | 97.3 | 95.7 | 98.7 |
| 8 | Excitement,Immobility/stupor,Mutism,Echopraxia/echolalia,Mannerisms,Negativism,Waxy flexibility,Withdrawal | 97.3 | 95.7 | 98.7 |
| 8 | Excitement,Immobility/stupor,Mutism,Echopraxia/echolalia,Stereotypy,Verbigeration,Waxy flexibility,Withdrawal | 97.3 | 95.7 | 98.7 |
| 8 | Excitement,Immobility/stupor,Mutism,Grimacing,Echopraxia/echolalia,Stereotypy,Verbigeration,Negativism | 97.3 | 95.7 | 98.7 |
| 8 | Excitement,Immobility/stupor,Mutism,Grimacing,Echopraxia/echolalia,Stereotypy,Waxy flexibility,Withdrawal | 97.3 | 95.7 | 98.7 |
| 8 | Excitement,Immobility/stupor,Mutism,Grimacing,Mannerisms,Verbigeration,Negativism,Withdrawal | 97.3 | 95.7 | 98.7 |
| 8 | Excitement,Immobility/stupor,Mutism,Grimacing,Verbigeration,Negativism,Waxy flexibility,Withdrawal | 97.3 | 95.7 | 98.7 |
| 8 | Excitement,Immobility/stupor,Mutism,Mannerisms,Verbigeration,Negativism,Waxy flexibility,Withdrawal | 97.3 | 95.7 | 98.7 |
| 8 | Excitement,Immobility/stupor,Mutism,Posturing/catalepsy,Echopraxia/echolalia,Mannerisms,Waxy flexibility,Withdrawal | 97.3 | 95.7 | 98.7 |
| 8 | Excitement,Immobility/stupor,Mutism,Posturing/catalepsy,Echopraxia/echolalia,Stereotypy,Mannerisms,Withdrawal | 97.3 | 95.7 | 98.7 |
| 8 | Excitement,Immobility/stupor,Mutism,Posturing/catalepsy,Grimacing,Mannerisms,Waxy flexibility,Withdrawal | 97.3 | 95.7 | 98.7 |
| 8 | Excitement,Immobility/stupor,Mutism,Posturing/catalepsy,Grimacing,Stereotypy,Waxy flexibility,Withdrawal | 97.3 | 95.7 | 98.7 |
| 8 | Excitement,Immobility/stupor,Mutism,Posturing/catalepsy,Mannerisms,Verbigeration,Waxy flexibility,Withdrawal | 97.3 | 95.7 | 98.7 |
| 8 | Excitement,Immobility/stupor,Mutism,Posturing/catalepsy,Stereotypy,Mannerisms,Negativism,Waxy flexibility | 97.3 | 95.7 | 98.7 |
| 8 | Excitement,Immobility/stupor,Mutism,Posturing/catalepsy,Stereotypy,Mannerisms,Verbigeration,Withdrawal | 97.3 | 95.7 | 98.7 |
| 8 | Excitement,Immobility/stupor,Mutism,Stereotypy,Mannerisms,Negativism,Waxy flexibility,Withdrawal | 97.3 | 95.7 | 98.7 |
| 8 | Excitement,Immobility/stupor,Mutism,Stereotypy,Mannerisms,Verbigeration,Negativism,Waxy flexibility | 97.3 | 95.7 | 98.7 |
| 8 | Excitement,Immobility/stupor,Posturing/catalepsy,Echopraxia/echolalia,Mannerisms,Verbigeration,Negativism,Waxy flexibility | 97.3 | 95.7 | 98.7 |
| 8 | Excitement,Immobility/stupor,Posturing/catalepsy,Echopraxia/echolalia,Stereotypy,Verbigeration,Negativism,Waxy flexibility | 97.3 | 95.7 | 98.7 |
| 8 | Excitement,Immobility/stupor,Posturing/catalepsy,Grimacing,Echopraxia/echolalia,Mannerisms,Verbigeration,Withdrawal | 97.3 | 95.7 | 98.7 |
| 8 | Excitement,Immobility/stupor,Posturing/catalepsy,Grimacing,Echopraxia/echolalia,Mannerisms,Waxy flexibility,Withdrawal | 97.3 | 95.7 | 98.7 |
| 8 | Excitement,Immobility/stupor,Posturing/catalepsy,Grimacing,Echopraxia/echolalia,Stereotypy,Negativism,Waxy flexibility | 97.3 | 95.7 | 98.7 |
| 8 | Excitement,Immobility/stupor,Posturing/catalepsy,Grimacing,Echopraxia/echolalia,Stereotypy,Verbigeration,Negativism | 97.3 | 95.7 | 98.7 |
| 8 | Excitement,Immobility/stupor,Posturing/catalepsy,Grimacing,Stereotypy,Mannerisms,Verbigeration,Negativism | 97.3 | 95.7 | 98.7 |
| 8 | Excitement,Immobility/stupor,Posturing/catalepsy,Stereotypy,Mannerisms,Verbigeration,Negativism,Waxy flexibility | 97.3 | 95.7 | 98.7 |
| 8 | Excitement,Immobility/stupor,Posturing/catalepsy,Stereotypy,Mannerisms,Verbigeration,Rigidity,Waxy flexibility | 97.3 | 95.7 | 98.7 |
| 8 | Excitement,Immobility/stupor,Staring,Grimacing,Echopraxia/echolalia,Stereotypy,Mannerisms,Verbigeration | 97.3 | 95.7 | 98.7 |
| 8 | Excitement,Immobility/stupor,Stereotypy,Mannerisms,Verbigeration,Negativism,Waxy flexibility,Withdrawal | 97.3 | 95.7 | 98.7 |
| 8 | Excitement,Staring,Echopraxia/echolalia,Stereotypy,Mannerisms,Verbigeration,Negativism,Waxy flexibility | 97.3 | 95.7 | 98.7 |
| 8 | Excitement,Staring,Echopraxia/echolalia,Stereotypy,Mannerisms,Verbigeration,Rigidity,Waxy flexibility | 97.3 | 95.7 | 98.7 |
| 8 | Excitement,Staring,Grimacing,Echopraxia/echolalia,Stereotypy,Verbigeration,Waxy flexibility,Withdrawal | 97.3 | 95.7 | 98.7 |
| 8 | Excitement,Staring,Posturing/catalepsy,Echopraxia/echolalia,Mannerisms,Verbigeration,Rigidity,Waxy flexibility | 97.3 | 95.7 | 98.7 |
| 8 | Excitement,Staring,Posturing/catalepsy,Echopraxia/echolalia,Stereotypy,Verbigeration,Waxy flexibility,Withdrawal | 97.3 | 95.7 | 98.7 |
| 8 | Excitement,Staring,Posturing/catalepsy,Grimacing,Echopraxia/echolalia,Stereotypy,Negativism,Waxy flexibility | 97.3 | 95.7 | 98.7 |
| 8 | Excitement,Staring,Posturing/catalepsy,Grimacing,Echopraxia/echolalia,Stereotypy,Verbigeration,Negativism | 97.3 | 95.7 | 98.7 |
| 8 | Excitement,Staring,Posturing/catalepsy,Grimacing,Echopraxia/echolalia,Verbigeration,Negativism,Waxy flexibility | 97.3 | 95.7 | 98.7 |
| 8 | Excitement,Staring,Posturing/catalepsy,Grimacing,Stereotypy,Verbigeration,Negativism,Waxy flexibility | 97.3 | 95.7 | 98.7 |
| 8 | Excitement,Staring,Posturing/catalepsy,Stereotypy,Mannerisms,Verbigeration,Rigidity,Waxy flexibility | 97.3 | 95.7 | 98.7 |
| 8 | Immobility/stupor,Echopraxia/echolalia,Stereotypy,Mannerisms,Verbigeration,Negativism,Waxy flexibility,Withdrawal | 97.3 | 95.7 | 98.7 |
| 8 | Immobility/stupor,Grimacing,Echopraxia/echolalia,Stereotypy,Mannerisms,Rigidity,Waxy flexibility,Withdrawal | 97.3 | 95.7 | 98.7 |
| 8 | Immobility/stupor,Mutism,Echopraxia/echolalia,Stereotypy,Mannerisms,Negativism,Waxy flexibility,Withdrawal | 97.3 | 95.7 | 98.7 |
| 8 | Immobility/stupor,Mutism,Grimacing,Echopraxia/echolalia,Mannerisms,Negativism,Waxy flexibility,Withdrawal | 97.3 | 95.7 | 98.7 |
| 8 | Immobility/stupor,Mutism,Grimacing,Echopraxia/echolalia,Rigidity,Negativism,Waxy flexibility,Withdrawal | 97.3 | 95.7 | 98.7 |
| 8 | Immobility/stupor,Mutism,Grimacing,Echopraxia/echolalia,Stereotypy,Verbigeration,Rigidity,Withdrawal | 97.3 | 95.7 | 98.7 |
| 8 | Immobility/stupor,Mutism,Grimacing,Mannerisms,Verbigeration,Rigidity,Waxy flexibility,Withdrawal | 97.3 | 95.7 | 98.7 |
| 8 | Immobility/stupor,Mutism,Grimacing,Stereotypy,Mannerisms,Verbigeration,Negativism,Withdrawal | 97.3 | 95.7 | 98.7 |
| 8 | Immobility/stupor,Mutism,Grimacing,Stereotypy,Verbigeration,Rigidity,Waxy flexibility,Withdrawal | 97.3 | 95.7 | 98.7 |
| 8 | Immobility/stupor,Mutism,Posturing/catalepsy,Grimacing,Echopraxia/echolalia,Mannerisms,Negativism,Waxy flexibility | 97.3 | 95.7 | 98.7 |
| 8 | Immobility/stupor,Mutism,Posturing/catalepsy,Grimacing,Echopraxia/echolalia,Stereotypy,Mannerisms,Negativism | 97.3 | 95.7 | 98.7 |
| 8 | Immobility/stupor,Mutism,Posturing/catalepsy,Grimacing,Echopraxia/echolalia,Stereotypy,Negativism,Withdrawal | 97.3 | 95.7 | 98.7 |
| 8 | Immobility/stupor,Mutism,Posturing/catalepsy,Grimacing,Echopraxia/echolalia,Stereotypy,Rigidity,Negativism | 97.3 | 95.7 | 98.7 |
| 8 | Immobility/stupor,Mutism,Posturing/catalepsy,Grimacing,Echopraxia/echolalia,Stereotypy,Rigidity,Withdrawal | 97.3 | 95.7 | 98.7 |
| 8 | Immobility/stupor,Mutism,Posturing/catalepsy,Grimacing,Stereotypy,Verbigeration,Negativism,Withdrawal | 97.3 | 95.7 | 98.7 |
| 8 | Immobility/stupor,Mutism,Posturing/catalepsy,Grimacing,Stereotypy,Verbigeration,Rigidity,Withdrawal | 97.3 | 95.7 | 98.7 |
| 8 | Immobility/stupor,Mutism,Posturing/catalepsy,Grimacing,Verbigeration,Rigidity,Negativism,Waxy flexibility | 97.3 | 95.7 | 98.7 |
| 8 | Immobility/stupor,Mutism,Posturing/catalepsy,Mannerisms,Verbigeration,Negativism,Waxy flexibility,Withdrawal | 97.3 | 95.7 | 98.7 |
| 8 | Immobility/stupor,Mutism,Posturing/catalepsy,Stereotypy,Mannerisms,Negativism,Waxy flexibility,Withdrawal | 97.3 | 95.7 | 98.7 |
| 8 | Immobility/stupor,Mutism,Posturing/catalepsy,Stereotypy,Rigidity,Negativism,Waxy flexibility,Withdrawal | 97.3 | 95.7 | 98.7 |
| 8 | Immobility/stupor,Mutism,Posturing/catalepsy,Verbigeration,Rigidity,Negativism,Waxy flexibility,Withdrawal | 97.3 | 95.7 | 98.7 |
| 8 | Immobility/stupor,Mutism,Staring,Echopraxia/echolalia,Verbigeration,Rigidity,Waxy flexibility,Withdrawal | 97.3 | 95.7 | 98.7 |
| 8 | Immobility/stupor,Mutism,Staring,Grimacing,Echopraxia/echolalia,Negativism,Waxy flexibility,Withdrawal | 97.3 | 95.7 | 98.7 |
| 8 | Immobility/stupor,Mutism,Staring,Grimacing,Echopraxia/echolalia,Rigidity,Negativism,Waxy flexibility | 97.3 | 95.7 | 98.7 |
| 8 | Immobility/stupor,Mutism,Staring,Grimacing,Echopraxia/echolalia,Rigidity,Waxy flexibility,Withdrawal | 97.3 | 95.7 | 98.7 |
| 8 | Immobility/stupor,Mutism,Staring,Grimacing,Echopraxia/echolalia,Stereotypy,Mannerisms,Verbigeration | 97.3 | 95.7 | 98.7 |
| 8 | Immobility/stupor,Mutism,Staring,Grimacing,Echopraxia/echolalia,Stereotypy,Rigidity,Waxy flexibility | 97.3 | 95.7 | 98.7 |
| 8 | Immobility/stupor,Mutism,Staring,Grimacing,Echopraxia/echolalia,Stereotypy,Verbigeration,Waxy flexibility | 97.3 | 95.7 | 98.7 |
| 8 | Immobility/stupor,Mutism,Staring,Grimacing,Echopraxia/echolalia,Stereotypy,Verbigeration,Withdrawal | 97.3 | 95.7 | 98.7 |
| 8 | Immobility/stupor,Mutism,Staring,Grimacing,Echopraxia/echolalia,Stereotypy,Waxy flexibility,Withdrawal | 97.3 | 95.7 | 98.7 |
| 8 | Immobility/stupor,Mutism,Staring,Grimacing,Rigidity,Negativism,Waxy flexibility,Withdrawal | 97.3 | 95.7 | 98.7 |
| 8 | Immobility/stupor,Mutism,Staring,Grimacing,Stereotypy,Verbigeration,Waxy flexibility,Withdrawal | 97.3 | 95.7 | 98.7 |
| 8 | Immobility/stupor,Mutism,Staring,Grimacing,Verbigeration,Negativism,Waxy flexibility,Withdrawal | 97.3 | 95.7 | 98.7 |
| 8 | Immobility/stupor,Mutism,Staring,Posturing/catalepsy,Echopraxia/echolalia,Mannerisms,Verbigeration,Waxy flexibility | 97.3 | 95.7 | 98.7 |
| 8 | Immobility/stupor,Mutism,Staring,Posturing/catalepsy,Echopraxia/echolalia,Rigidity,Waxy flexibility,Withdrawal | 97.3 | 95.7 | 98.7 |
| 8 | Immobility/stupor,Mutism,Staring,Posturing/catalepsy,Echopraxia/echolalia,Stereotypy,Mannerisms,Verbigeration | 97.3 | 95.7 | 98.7 |
| 8 | Immobility/stupor,Mutism,Staring,Posturing/catalepsy,Echopraxia/echolalia,Stereotypy,Mannerisms,Waxy flexibility | 97.3 | 95.7 | 98.7 |
| 8 | Immobility/stupor,Mutism,Staring,Posturing/catalepsy,Echopraxia/echolalia,Verbigeration,Rigidity,Waxy flexibility | 97.3 | 95.7 | 98.7 |
| 8 | Immobility/stupor,Mutism,Staring,Posturing/catalepsy,Grimacing,Echopraxia/echolalia,Negativism,Waxy flexibility | 97.3 | 95.7 | 98.7 |
| 8 | Immobility/stupor,Mutism,Staring,Posturing/catalepsy,Grimacing,Echopraxia/echolalia,Stereotypy,Verbigeration | 97.3 | 95.7 | 98.7 |
| 8 | Immobility/stupor,Mutism,Staring,Posturing/catalepsy,Grimacing,Echopraxia/echolalia,Verbigeration,Withdrawal | 97.3 | 95.7 | 98.7 |
| 8 | Immobility/stupor,Mutism,Staring,Posturing/catalepsy,Grimacing,Stereotypy,Mannerisms,Waxy flexibility | 97.3 | 95.7 | 98.7 |
| 8 | Immobility/stupor,Mutism,Staring,Posturing/catalepsy,Grimacing,Stereotypy,Verbigeration,Waxy flexibility | 97.3 | 95.7 | 98.7 |
| 8 | Immobility/stupor,Mutism,Staring,Posturing/catalepsy,Stereotypy,Mannerisms,Verbigeration,Waxy flexibility | 97.3 | 95.7 | 98.7 |
| 8 | Immobility/stupor,Mutism,Staring,Posturing/catalepsy,Verbigeration,Rigidity,Waxy flexibility,Withdrawal | 97.3 | 95.7 | 98.7 |
| 8 | Immobility/stupor,Posturing/catalepsy,Echopraxia/echolalia,Stereotypy,Verbigeration,Rigidity,Negativism,Waxy flexibility | 97.3 | 95.7 | 98.7 |
| 8 | Immobility/stupor,Posturing/catalepsy,Grimacing,Echopraxia/echolalia,Mannerisms,Negativism,Waxy flexibility,Withdrawal | 97.3 | 95.7 | 98.7 |
| 8 | Immobility/stupor,Posturing/catalepsy,Grimacing,Echopraxia/echolalia,Stereotypy,Mannerisms,Rigidity,Withdrawal | 97.3 | 95.7 | 98.7 |
| 8 | Immobility/stupor,Posturing/catalepsy,Grimacing,Stereotypy,Mannerisms,Negativism,Waxy flexibility,Withdrawal | 97.3 | 95.7 | 98.7 |
| 8 | Immobility/stupor,Staring,Echopraxia/echolalia,Stereotypy,Verbigeration,Rigidity,Waxy flexibility,Withdrawal | 97.3 | 95.7 | 98.7 |
| 8 | Immobility/stupor,Staring,Grimacing,Echopraxia/echolalia,Stereotypy,Mannerisms,Verbigeration,Waxy flexibility | 97.3 | 95.7 | 98.7 |
| 8 | Immobility/stupor,Staring,Grimacing,Echopraxia/echolalia,Verbigeration,Negativism,Waxy flexibility,Withdrawal | 97.3 | 95.7 | 98.7 |
| 8 | Immobility/stupor,Staring,Posturing/catalepsy,Echopraxia/echolalia,Mannerisms,Verbigeration,Rigidity,Waxy flexibility | 97.3 | 95.7 | 98.7 |
| 8 | Immobility/stupor,Staring,Posturing/catalepsy,Echopraxia/echolalia,Stereotypy,Mannerisms,Waxy flexibility,Withdrawal | 97.3 | 95.7 | 98.7 |
| 8 | Immobility/stupor,Staring,Posturing/catalepsy,Echopraxia/echolalia,Stereotypy,Rigidity,Waxy flexibility,Withdrawal | 97.3 | 95.7 | 98.7 |
| 8 | Immobility/stupor,Staring,Posturing/catalepsy,Grimacing,Echopraxia/echolalia,Mannerisms,Rigidity,Waxy flexibility | 97.3 | 95.7 | 98.7 |
| 8 | Immobility/stupor,Staring,Posturing/catalepsy,Grimacing,Echopraxia/echolalia,Rigidity,Negativism,Waxy flexibility | 97.3 | 95.7 | 98.7 |
| 8 | Immobility/stupor,Staring,Posturing/catalepsy,Grimacing,Echopraxia/echolalia,Stereotypy,Mannerisms,Verbigeration | 97.3 | 95.7 | 98.7 |
| 8 | Immobility/stupor,Staring,Posturing/catalepsy,Grimacing,Echopraxia/echolalia,Stereotypy,Mannerisms,Waxy flexibility | 97.3 | 95.7 | 98.7 |
| 8 | Immobility/stupor,Staring,Posturing/catalepsy,Grimacing,Echopraxia/echolalia,Verbigeration,Rigidity,Withdrawal | 97.3 | 95.7 | 98.7 |
| 8 | Immobility/stupor,Staring,Posturing/catalepsy,Grimacing,Mannerisms,Verbigeration,Waxy flexibility,Withdrawal | 97.3 | 95.7 | 98.7 |
| 8 | Immobility/stupor,Staring,Posturing/catalepsy,Grimacing,Stereotypy,Mannerisms,Verbigeration,Waxy flexibility | 97.3 | 95.7 | 98.7 |
| 8 | Immobility/stupor,Staring,Posturing/catalepsy,Grimacing,Stereotypy,Rigidity,Waxy flexibility,Withdrawal | 97.3 | 95.7 | 98.7 |
| 8 | Mutism,Staring,Posturing/catalepsy,Echopraxia/echolalia,Stereotypy,Verbigeration,Waxy flexibility,Withdrawal | 97.3 | 95.7 | 98.7 |
| 8 | Staring,Grimacing,Echopraxia/echolalia,Stereotypy,Mannerisms,Verbigeration,Waxy flexibility,Withdrawal | 97.3 | 95.7 | 98.7 |
| 8 | Staring,Grimacing,Echopraxia/echolalia,Stereotypy,Verbigeration,Negativism,Waxy flexibility,Withdrawal | 97.3 | 95.7 | 98.7 |
| 8 | Staring,Posturing/catalepsy,Echopraxia/echolalia,Mannerisms,Verbigeration,Rigidity,Waxy flexibility,Withdrawal | 97.3 | 95.7 | 98.7 |
| 8 | Staring,Posturing/catalepsy,Echopraxia/echolalia,Stereotypy,Verbigeration,Negativism,Waxy flexibility,Withdrawal | 97.3 | 95.7 | 98.7 |
| 8 | Staring,Posturing/catalepsy,Grimacing,Echopraxia/echolalia,Stereotypy,Mannerisms,Negativism,Waxy flexibility | 97.3 | 95.7 | 98.7 |
| 8 | Staring,Posturing/catalepsy,Grimacing,Echopraxia/echolalia,Stereotypy,Mannerisms,Verbigeration,Negativism | 97.3 | 95.7 | 98.7 |
| 8 | Staring,Posturing/catalepsy,Grimacing,Echopraxia/echolalia,Stereotypy,Rigidity,Waxy flexibility,Withdrawal | 97.3 | 95.7 | 98.7 |
| 8 | Excitement,Immobility/stupor,Grimacing,Echopraxia/echolalia,Stereotypy,Mannerisms,Verbigeration,Rigidity | 97.1 | 95.5 | 98.7 |
| 8 | Excitement,Immobility/stupor,Grimacing,Echopraxia/echolalia,Stereotypy,Verbigeration,Negativism,Waxy flexibility | 97.1 | 95.3 | 98.4 |
| 8 | Excitement,Immobility/stupor,Grimacing,Stereotypy,Mannerisms,Verbigeration,Rigidity,Waxy flexibility | 97.1 | 95.5 | 98.4 |
| 8 | Excitement,Immobility/stupor,Grimacing,Stereotypy,Verbigeration,Negativism,Waxy flexibility,Withdrawal | 97.1 | 95.5 | 98.7 |
| 8 | Excitement,Immobility/stupor,Mutism,Echopraxia/echolalia,Mannerisms,Verbigeration,Negativism,Waxy flexibility | 97.1 | 95.5 | 98.4 |
| 8 | Excitement,Immobility/stupor,Mutism,Echopraxia/echolalia,Stereotypy,Mannerisms,Negativism,Waxy flexibility | 97.1 | 95.5 | 98.4 |
| 8 | Excitement,Immobility/stupor,Mutism,Echopraxia/echolalia,Stereotypy,Mannerisms,Verbigeration,Negativism | 97.1 | 95.5 | 98.7 |
| 8 | Excitement,Immobility/stupor,Mutism,Grimacing,Echopraxia/echolalia,Mannerisms,Negativism,Waxy flexibility | 97.1 | 95.3 | 98.4 |
| 8 | Excitement,Immobility/stupor,Mutism,Grimacing,Echopraxia/echolalia,Mannerisms,Negativism,Withdrawal | 97.1 | 95.5 | 98.4 |
| 8 | Excitement,Immobility/stupor,Mutism,Grimacing,Echopraxia/echolalia,Stereotypy,Verbigeration,Withdrawal | 97.1 | 95.5 | 98.7 |
| 8 | Excitement,Immobility/stupor,Mutism,Grimacing,Echopraxia/echolalia,Verbigeration,Waxy flexibility,Withdrawal | 97.1 | 95.3 | 98.4 |
| 8 | Excitement,Immobility/stupor,Mutism,Grimacing,Mannerisms,Negativism,Waxy flexibility,Withdrawal | 97.1 | 95.5 | 98.7 |
| 8 | Excitement,Immobility/stupor,Mutism,Grimacing,Mannerisms,Verbigeration,Negativism,Waxy flexibility | 97.1 | 95.5 | 98.4 |
| 8 | Excitement,Immobility/stupor,Mutism,Grimacing,Stereotypy,Mannerisms,Negativism,Withdrawal | 97.1 | 95.5 | 98.7 |
| 8 | Excitement,Immobility/stupor,Mutism,Posturing/catalepsy,Echopraxia/echolalia,Stereotypy,Verbigeration,Waxy flexibility | 97.1 | 95.3 | 98.4 |
| 8 | Excitement,Immobility/stupor,Mutism,Posturing/catalepsy,Grimacing,Echopraxia/echolalia,Mannerisms,Verbigeration | 97.1 | 95.5 | 98.7 |
| 8 | Excitement,Immobility/stupor,Mutism,Posturing/catalepsy,Grimacing,Echopraxia/echolalia,Mannerisms,Waxy flexibility | 97.1 | 95.5 | 98.4 |
| 8 | Excitement,Immobility/stupor,Mutism,Posturing/catalepsy,Grimacing,Echopraxia/echolalia,Stereotypy,Verbigeration | 97.1 | 95.5 | 98.4 |
| 8 | Excitement,Immobility/stupor,Mutism,Posturing/catalepsy,Grimacing,Echopraxia/echolalia,Stereotypy,Waxy flexibility | 97.1 | 95.5 | 98.4 |
| 8 | Excitement,Immobility/stupor,Mutism,Posturing/catalepsy,Grimacing,Mannerisms,Verbigeration,Waxy flexibility | 97.1 | 95.5 | 98.4 |
| 8 | Excitement,Immobility/stupor,Mutism,Posturing/catalepsy,Grimacing,Stereotypy,Mannerisms,Withdrawal | 97.1 | 95.5 | 98.4 |
| 8 | Excitement,Immobility/stupor,Mutism,Posturing/catalepsy,Grimacing,Stereotypy,Verbigeration,Waxy flexibility | 97.1 | 95.5 | 98.7 |
| 8 | Excitement,Immobility/stupor,Posturing/catalepsy,Echopraxia/echolalia,Mannerisms,Verbigeration,Waxy flexibility,Withdrawal | 97.1 | 95.5 | 98.4 |
| 8 | Excitement,Immobility/stupor,Posturing/catalepsy,Echopraxia/echolalia,Stereotypy,Mannerisms,Rigidity,Waxy flexibility | 97.1 | 95.3 | 98.4 |
| 8 | Excitement,Immobility/stupor,Posturing/catalepsy,Echopraxia/echolalia,Stereotypy,Mannerisms,Verbigeration,Negativism | 97.1 | 95.5 | 98.4 |
| 8 | Excitement,Immobility/stupor,Posturing/catalepsy,Echopraxia/echolalia,Stereotypy,Mannerisms,Verbigeration,Withdrawal | 97.1 | 95.5 | 98.7 |
| 8 | Excitement,Immobility/stupor,Posturing/catalepsy,Grimacing,Echopraxia/echolalia,Stereotypy,Mannerisms,Negativism | 97.1 | 95.5 | 98.4 |
| 8 | Excitement,Immobility/stupor,Posturing/catalepsy,Grimacing,Echopraxia/echolalia,Stereotypy,Mannerisms,Withdrawal | 97.1 | 95.5 | 98.4 |
| 8 | Excitement,Immobility/stupor,Posturing/catalepsy,Grimacing,Echopraxia/echolalia,Verbigeration,Waxy flexibility,Withdrawal | 97.1 | 95.5 | 98.7 |
| 8 | Excitement,Immobility/stupor,Posturing/catalepsy,Grimacing,Stereotypy,Mannerisms,Rigidity,Waxy flexibility | 97.1 | 95.3 | 98.4 |
| 8 | Excitement,Staring,Grimacing,Echopraxia/echolalia,Stereotypy,Mannerisms,Negativism,Waxy flexibility | 97.1 | 95.5 | 98.4 |
| 8 | Excitement,Staring,Posturing/catalepsy,Echopraxia/echolalia,Stereotypy,Mannerisms,Rigidity,Waxy flexibility | 97.1 | 95.5 | 98.4 |
| 8 | Excitement,Staring,Posturing/catalepsy,Echopraxia/echolalia,Stereotypy,Verbigeration,Rigidity,Waxy flexibility | 97.1 | 95.5 | 98.4 |
| 8 | Excitement,Staring,Posturing/catalepsy,Grimacing,Stereotypy,Mannerisms,Negativism,Waxy flexibility | 97.1 | 95.5 | 98.4 |
| 8 | Immobility/stupor,Grimacing,Echopraxia/echolalia,Stereotypy,Mannerisms,Verbigeration,Rigidity,Withdrawal | 97.1 | 95.5 | 98.4 |
| 8 | Immobility/stupor,Mutism,Grimacing,Echopraxia/echolalia,Mannerisms,Verbigeration,Negativism,Withdrawal | 97.1 | 95.5 | 98.4 |
| 8 | Immobility/stupor,Mutism,Grimacing,Echopraxia/echolalia,Stereotypy,Mannerisms,Negativism,Waxy flexibility | 97.1 | 95.3 | 98.4 |
| 8 | Immobility/stupor,Mutism,Grimacing,Echopraxia/echolalia,Stereotypy,Rigidity,Negativism,Waxy flexibility | 97.1 | 95.5 | 98.7 |
| 8 | Immobility/stupor,Mutism,Grimacing,Echopraxia/echolalia,Stereotypy,Rigidity,Waxy flexibility,Withdrawal | 97.1 | 95.3 | 98.4 |
| 8 | Immobility/stupor,Mutism,Grimacing,Echopraxia/echolalia,Stereotypy,Verbigeration,Negativism,Withdrawal | 97.1 | 95.3 | 98.7 |
| 8 | Immobility/stupor,Mutism,Grimacing,Stereotypy,Mannerisms,Negativism,Waxy flexibility,Withdrawal | 97.1 | 95.5 | 98.7 |
| 8 | Immobility/stupor,Mutism,Posturing/catalepsy,Echopraxia/echolalia,Mannerisms,Verbigeration,Rigidity,Withdrawal | 97.1 | 95.5 | 98.7 |
| 8 | Immobility/stupor,Mutism,Posturing/catalepsy,Echopraxia/echolalia,Rigidity,Negativism,Waxy flexibility,Withdrawal | 97.1 | 95.5 | 98.4 |
| 8 | Immobility/stupor,Mutism,Posturing/catalepsy,Echopraxia/echolalia,Stereotypy,Mannerisms,Rigidity,Withdrawal | 97.1 | 95.5 | 98.4 |
| 8 | Immobility/stupor,Mutism,Posturing/catalepsy,Echopraxia/echolalia,Stereotypy,Mannerisms,Verbigeration,Negativism | 97.1 | 95.5 | 98.4 |
| 8 | Immobility/stupor,Mutism,Posturing/catalepsy,Echopraxia/echolalia,Stereotypy,Rigidity,Negativism,Waxy flexibility | 97.1 | 95.5 | 98.7 |
| 8 | Immobility/stupor,Mutism,Posturing/catalepsy,Echopraxia/echolalia,Stereotypy,Verbigeration,Negativism,Withdrawal | 97.1 | 95.5 | 98.4 |
| 8 | Immobility/stupor,Mutism,Posturing/catalepsy,Grimacing,Echopraxia/echolalia,Mannerisms,Verbigeration,Rigidity | 97.1 | 95.5 | 98.4 |
| 8 | Immobility/stupor,Mutism,Posturing/catalepsy,Grimacing,Echopraxia/echolalia,Verbigeration,Negativism,Withdrawal | 97.1 | 95.5 | 98.4 |
| 8 | Immobility/stupor,Mutism,Posturing/catalepsy,Grimacing,Mannerisms,Rigidity,Waxy flexibility,Withdrawal | 97.1 | 95.5 | 98.4 |
| 8 | Immobility/stupor,Mutism,Posturing/catalepsy,Grimacing,Stereotypy,Mannerisms,Verbigeration,Negativism | 97.1 | 95.5 | 98.4 |
| 8 | Immobility/stupor,Mutism,Posturing/catalepsy,Grimacing,Stereotypy,Mannerisms,Verbigeration,Rigidity | 97.1 | 95.5 | 98.4 |
| 8 | Immobility/stupor,Mutism,Posturing/catalepsy,Grimacing,Stereotypy,Rigidity,Negativism,Waxy flexibility | 97.1 | 95.3 | 98.7 |
| 8 | Immobility/stupor,Mutism,Posturing/catalepsy,Stereotypy,Mannerisms,Verbigeration,Rigidity,Withdrawal | 97.1 | 95.5 | 98.4 |
| 8 | Immobility/stupor,Mutism,Staring,Echopraxia/echolalia,Mannerisms,Verbigeration,Waxy flexibility,Withdrawal | 97.1 | 95.3 | 98.4 |
| 8 | Immobility/stupor,Mutism,Staring,Echopraxia/echolalia,Stereotypy,Mannerisms,Verbigeration,Waxy flexibility | 97.1 | 95.5 | 98.4 |
| 8 | Immobility/stupor,Mutism,Staring,Echopraxia/echolalia,Stereotypy,Mannerisms,Verbigeration,Withdrawal | 97.1 | 95.5 | 98.4 |
| 8 | Immobility/stupor,Mutism,Staring,Posturing/catalepsy,Echopraxia/echolalia,Stereotypy,Rigidity,Waxy flexibility | 97.1 | 95.5 | 98.4 |
| 8 | Immobility/stupor,Mutism,Staring,Posturing/catalepsy,Echopraxia/echolalia,Stereotypy,Verbigeration,Withdrawal | 97.1 | 95.5 | 98.4 |
| 8 | Immobility/stupor,Mutism,Staring,Posturing/catalepsy,Echopraxia/echolalia,Stereotypy,Waxy flexibility,Withdrawal | 97.1 | 95.5 | 98.4 |
| 8 | Immobility/stupor,Mutism,Staring,Posturing/catalepsy,Grimacing,Echopraxia/echolalia,Verbigeration,Waxy flexibility | 97.1 | 95.5 | 98.4 |
| 8 | Immobility/stupor,Mutism,Staring,Posturing/catalepsy,Grimacing,Echopraxia/echolalia,Waxy flexibility,Withdrawal | 97.1 | 95.5 | 98.4 |
| 8 | Immobility/stupor,Mutism,Staring,Posturing/catalepsy,Grimacing,Stereotypy,Waxy flexibility,Withdrawal | 97.1 | 95.5 | 98.4 |
| 8 | Immobility/stupor,Mutism,Staring,Posturing/catalepsy,Grimacing,Verbigeration,Rigidity,Waxy flexibility | 97.1 | 95.5 | 98.4 |
| 8 | Immobility/stupor,Mutism,Staring,Posturing/catalepsy,Stereotypy,Verbigeration,Waxy flexibility,Withdrawal | 97.1 | 95.5 | 98.4 |
| 8 | Immobility/stupor,Mutism,Staring,Stereotypy,Mannerisms,Verbigeration,Waxy flexibility,Withdrawal | 97.1 | 95.5 | 98.4 |
| 8 | Immobility/stupor,Mutism,Stereotypy,Mannerisms,Verbigeration,Negativism,Waxy flexibility,Withdrawal | 97.1 | 95.5 | 98.4 |
| 8 | Immobility/stupor,Posturing/catalepsy,Echopraxia/echolalia,Mannerisms,Verbigeration,Negativism,Waxy flexibility,Withdrawal | 97.1 | 95.5 | 98.4 |
| 8 | Immobility/stupor,Posturing/catalepsy,Grimacing,Echopraxia/echolalia,Mannerisms,Verbigeration,Rigidity,Withdrawal | 97.1 | 95.5 | 98.4 |
| 8 | Immobility/stupor,Posturing/catalepsy,Grimacing,Echopraxia/echolalia,Stereotypy,Mannerisms,Negativism,Waxy flexibility | 97.1 | 95.5 | 98.4 |
| 8 | Immobility/stupor,Posturing/catalepsy,Grimacing,Echopraxia/echolalia,Stereotypy,Rigidity,Negativism,Waxy flexibility | 97.1 | 95.5 | 98.4 |
| 8 | Immobility/stupor,Posturing/catalepsy,Grimacing,Echopraxia/echolalia,Stereotypy,Verbigeration,Negativism,Withdrawal | 97.1 | 95.5 | 98.4 |
| 8 | Immobility/stupor,Posturing/catalepsy,Grimacing,Mannerisms,Verbigeration,Negativism,Waxy flexibility,Withdrawal | 97.1 | 95.5 | 98.4 |
| 8 | Immobility/stupor,Staring,Grimacing,Echopraxia/echolalia,Mannerisms,Verbigeration,Waxy flexibility,Withdrawal | 97.1 | 95.5 | 98.4 |
| 8 | Immobility/stupor,Staring,Grimacing,Echopraxia/echolalia,Verbigeration,Rigidity,Waxy flexibility,Withdrawal | 97.1 | 95.5 | 98.4 |
| 8 | Immobility/stupor,Staring,Posturing/catalepsy,Echopraxia/echolalia,Stereotypy,Mannerisms,Rigidity,Waxy flexibility | 97.1 | 95.5 | 98.7 |
| 8 | Immobility/stupor,Staring,Posturing/catalepsy,Echopraxia/echolalia,Stereotypy,Mannerisms,Verbigeration,Withdrawal | 97.1 | 95.5 | 98.7 |
| 8 | Immobility/stupor,Staring,Posturing/catalepsy,Grimacing,Echopraxia/echolalia,Mannerisms,Verbigeration,Waxy flexibility | 97.1 | 95.5 | 98.4 |
| 8 | Immobility/stupor,Staring,Posturing/catalepsy,Stereotypy,Mannerisms,Verbigeration,Waxy flexibility,Withdrawal | 97.1 | 95.5 | 98.7 |
| 8 | Staring,Grimacing,Echopraxia/echolalia,Stereotypy,Mannerisms,Verbigeration,Rigidity,Waxy flexibility | 97.1 | 95.5 | 98.4 |
| 8 | Staring,Grimacing,Echopraxia/echolalia,Stereotypy,Verbigeration,Rigidity,Negativism,Waxy flexibility | 97.1 | 95.5 | 98.7 |
| 8 | Staring,Posturing/catalepsy,Echopraxia/echolalia,Stereotypy,Mannerisms,Verbigeration,Negativism,Waxy flexibility | 97.1 | 95.5 | 98.4 |
| 8 | Staring,Posturing/catalepsy,Grimacing,Echopraxia/echolalia,Mannerisms,Verbigeration,Waxy flexibility,Withdrawal | 97.1 | 95.5 | 98.7 |
| 8 | Staring,Posturing/catalepsy,Grimacing,Echopraxia/echolalia,Stereotypy,Mannerisms,Verbigeration,Rigidity | 97.1 | 95.5 | 98.4 |
| 8 | Staring,Posturing/catalepsy,Grimacing,Echopraxia/echolalia,Stereotypy,Verbigeration,Rigidity,Negativism | 97.1 | 95.5 | 98.4 |
| 8 | Staring,Posturing/catalepsy,Grimacing,Echopraxia/echolalia,Verbigeration,Negativism,Waxy flexibility,Withdrawal | 97.1 | 95.5 | 98.7 |
| 8 | Staring,Posturing/catalepsy,Grimacing,Mannerisms,Verbigeration,Rigidity,Negativism,Waxy flexibility | 97.1 | 95.5 | 98.7 |
| 8 | Staring,Posturing/catalepsy,Grimacing,Stereotypy,Mannerisms,Verbigeration,Negativism,Waxy flexibility | 97.1 | 95.3 | 98.4 |
| 8 | Excitement,Immobility/stupor,Grimacing,Echopraxia/echolalia,Mannerisms,Verbigeration,Negativism,Waxy flexibility | 96.9 | 95.1 | 98.4 |
| 8 | Excitement,Immobility/stupor,Grimacing,Echopraxia/echolalia,Stereotypy,Mannerisms,Negativism,Waxy flexibility | 96.9 | 95.3 | 98.4 |
| 8 | Excitement,Immobility/stupor,Mutism,Echopraxia/echolalia,Stereotypy,Mannerisms,Verbigeration,Withdrawal | 96.9 | 95.1 | 98.4 |
| 8 | Excitement,Immobility/stupor,Mutism,Echopraxia/echolalia,Stereotypy,Mannerisms,Waxy flexibility,Withdrawal | 96.9 | 95.1 | 98.4 |
| 8 | Excitement,Immobility/stupor,Mutism,Grimacing,Stereotypy,Mannerisms,Negativism,Waxy flexibility | 96.9 | 95.1 | 98.4 |
| 8 | Excitement,Immobility/stupor,Mutism,Grimacing,Stereotypy,Mannerisms,Verbigeration,Negativism | 96.9 | 95.1 | 98.4 |
| 8 | Excitement,Immobility/stupor,Mutism,Grimacing,Stereotypy,Verbigeration,Waxy flexibility,Withdrawal | 96.9 | 95.1 | 98.4 |
| 8 | Excitement,Immobility/stupor,Mutism,Posturing/catalepsy,Echopraxia/echolalia,Mannerisms,Verbigeration,Waxy flexibility | 96.9 | 95.1 | 98.4 |
| 8 | Excitement,Immobility/stupor,Mutism,Posturing/catalepsy,Stereotypy,Mannerisms,Waxy flexibility,Withdrawal | 96.9 | 95.1 | 98.4 |
| 8 | Excitement,Immobility/stupor,Posturing/catalepsy,Echopraxia/echolalia,Stereotypy,Mannerisms,Negativism,Waxy flexibility | 96.9 | 95.3 | 98.4 |
| 8 | Excitement,Immobility/stupor,Posturing/catalepsy,Echopraxia/echolalia,Stereotypy,Verbigeration,Waxy flexibility,Withdrawal | 96.9 | 95.1 | 98.4 |
| 8 | Excitement,Immobility/stupor,Posturing/catalepsy,Grimacing,Echopraxia/echolalia,Stereotypy,Verbigeration,Withdrawal | 96.9 | 95.1 | 98.4 |
| 8 | Excitement,Immobility/stupor,Posturing/catalepsy,Grimacing,Echopraxia/echolalia,Stereotypy,Waxy flexibility,Withdrawal | 96.9 | 95.1 | 98.4 |
| 8 | Excitement,Immobility/stupor,Posturing/catalepsy,Grimacing,Mannerisms,Verbigeration,Waxy flexibility,Withdrawal | 96.9 | 95.1 | 98.4 |
| 8 | Excitement,Immobility/stupor,Posturing/catalepsy,Grimacing,Stereotypy,Mannerisms,Negativism,Waxy flexibility | 96.9 | 95.3 | 98.4 |
| 8 | Excitement,Mutism,Posturing/catalepsy,Echopraxia/echolalia,Verbigeration,Rigidity,Negativism,Withdrawal | 96.9 | 95.1 | 98.4 |
| 8 | Excitement,Mutism,Posturing/catalepsy,Stereotypy,Verbigeration,Rigidity,Negativism,Withdrawal | 96.9 | 95.1 | 98.4 |
| 8 | Excitement,Staring,Grimacing,Echopraxia/echolalia,Mannerisms,Verbigeration,Negativism,Waxy flexibility | 96.9 | 95.1 | 98.4 |
| 8 | Excitement,Staring,Grimacing,Echopraxia/echolalia,Stereotypy,Verbigeration,Negativism,Waxy flexibility | 96.9 | 95.3 | 98.4 |
| 8 | Excitement,Staring,Grimacing,Stereotypy,Mannerisms,Verbigeration,Negativism,Waxy flexibility | 96.9 | 95.3 | 98.4 |
| 8 | Excitement,Staring,Posturing/catalepsy,Grimacing,Echopraxia/echolalia,Stereotypy,Mannerisms,Waxy flexibility | 96.9 | 95.1 | 98.4 |
| 8 | Immobility/stupor,Grimacing,Echopraxia/echolalia,Stereotypy,Mannerisms,Verbigeration,Negativism,Waxy flexibility | 96.9 | 95.1 | 98.4 |
| 8 | Immobility/stupor,Grimacing,Stereotypy,Mannerisms,Verbigeration,Negativism,Waxy flexibility,Withdrawal | 96.9 | 95.1 | 98.4 |
| 8 | Immobility/stupor,Mutism,Echopraxia/echolalia,Mannerisms,Verbigeration,Negativism,Waxy flexibility,Withdrawal | 96.9 | 95.1 | 98.4 |
| 8 | Immobility/stupor,Mutism,Echopraxia/echolalia,Stereotypy,Mannerisms,Rigidity,Waxy flexibility,Withdrawal | 96.9 | 95.1 | 98.4 |
| 8 | Immobility/stupor,Mutism,Echopraxia/echolalia,Stereotypy,Mannerisms,Verbigeration,Negativism,Waxy flexibility | 96.9 | 95.1 | 98.4 |
| 8 | Immobility/stupor,Mutism,Grimacing,Echopraxia/echolalia,Verbigeration,Rigidity,Waxy flexibility,Withdrawal | 96.9 | 95.1 | 98.4 |
| 8 | Immobility/stupor,Mutism,Posturing/catalepsy,Echopraxia/echolalia,Mannerisms,Verbigeration,Negativism,Waxy flexibility | 96.9 | 95.1 | 98.4 |
| 8 | Immobility/stupor,Mutism,Posturing/catalepsy,Echopraxia/echolalia,Stereotypy,Mannerisms,Negativism,Waxy flexibility | 96.9 | 95.1 | 98.4 |
| 8 | Immobility/stupor,Mutism,Posturing/catalepsy,Grimacing,Echopraxia/echolalia,Negativism,Waxy flexibility,Withdrawal | 96.9 | 95.3 | 98.4 |
| 8 | Immobility/stupor,Mutism,Posturing/catalepsy,Grimacing,Echopraxia/echolalia,Rigidity,Negativism,Waxy flexibility | 96.9 | 95.1 | 98.4 |
| 8 | Immobility/stupor,Mutism,Posturing/catalepsy,Grimacing,Echopraxia/echolalia,Stereotypy,Mannerisms,Rigidity | 96.9 | 95.1 | 98.4 |
| 8 | Immobility/stupor,Mutism,Posturing/catalepsy,Grimacing,Echopraxia/echolalia,Stereotypy,Verbigeration,Negativism | 96.9 | 95.1 | 98.4 |
| 8 | Immobility/stupor,Mutism,Posturing/catalepsy,Grimacing,Echopraxia/echolalia,Stereotypy,Verbigeration,Rigidity | 96.9 | 95.1 | 98.4 |
| 8 | Immobility/stupor,Mutism,Posturing/catalepsy,Grimacing,Echopraxia/echolalia,Verbigeration,Negativism,Waxy flexibility | 96.9 | 95.3 | 98.4 |
| 8 | Immobility/stupor,Mutism,Posturing/catalepsy,Grimacing,Echopraxia/echolalia,Verbigeration,Rigidity,Withdrawal | 96.9 | 95.1 | 98.4 |
| 8 | Immobility/stupor,Mutism,Posturing/catalepsy,Grimacing,Mannerisms,Verbigeration,Negativism,Waxy flexibility | 96.9 | 95.1 | 98.4 |
| 8 | Immobility/stupor,Mutism,Posturing/catalepsy,Grimacing,Rigidity,Negativism,Waxy flexibility,Withdrawal | 96.9 | 95.1 | 98.4 |
| 8 | Immobility/stupor,Mutism,Posturing/catalepsy,Grimacing,Stereotypy,Mannerisms,Negativism,Waxy flexibility | 96.9 | 95.1 | 98.4 |
| 8 | Immobility/stupor,Mutism,Posturing/catalepsy,Stereotypy,Mannerisms,Verbigeration,Negativism,Waxy flexibility | 96.9 | 95.1 | 98.4 |
| 8 | Immobility/stupor,Mutism,Posturing/catalepsy,Stereotypy,Verbigeration,Negativism,Waxy flexibility,Withdrawal | 96.9 | 95.1 | 98.4 |
| 8 | Immobility/stupor,Mutism,Staring,Grimacing,Echopraxia/echolalia,Verbigeration,Waxy flexibility,Withdrawal | 96.9 | 95.3 | 98.4 |
| 8 | Immobility/stupor,Mutism,Staring,Posturing/catalepsy,Echopraxia/echolalia,Stereotypy,Verbigeration,Waxy flexibility | 96.9 | 95.1 | 98.4 |
| 8 | Immobility/stupor,Mutism,Staring,Posturing/catalepsy,Grimacing,Echopraxia/echolalia,Stereotypy,Waxy flexibility | 96.9 | 95.1 | 98.4 |
| 8 | Immobility/stupor,Mutism,Staring,Posturing/catalepsy,Grimacing,Stereotypy,Rigidity,Waxy flexibility | 96.9 | 95.3 | 98.4 |
| 8 | Immobility/stupor,Mutism,Staring,Posturing/catalepsy,Grimacing,Verbigeration,Waxy flexibility,Withdrawal | 96.9 | 95.1 | 98.4 |
| 8 | Immobility/stupor,Mutism,Stereotypy,Mannerisms,Verbigeration,Rigidity,Waxy flexibility,Withdrawal | 96.9 | 95.1 | 98.4 |
| 8 | Immobility/stupor,Posturing/catalepsy,Grimacing,Echopraxia/echolalia,Mannerisms,Rigidity,Waxy flexibility,Withdrawal | 96.9 | 95.1 | 98.4 |
| 8 | Immobility/stupor,Posturing/catalepsy,Grimacing,Echopraxia/echolalia,Mannerisms,Verbigeration,Negativism,Waxy flexibility | 96.9 | 95.1 | 98.4 |
| 8 | Immobility/stupor,Posturing/catalepsy,Grimacing,Echopraxia/echolalia,Stereotypy,Mannerisms,Verbigeration,Negativism | 96.9 | 95.1 | 98.4 |
| 8 | Immobility/stupor,Posturing/catalepsy,Grimacing,Echopraxia/echolalia,Stereotypy,Negativism,Waxy flexibility,Withdrawal | 96.9 | 95.3 | 98.2 |
| 8 | Immobility/stupor,Posturing/catalepsy,Grimacing,Stereotypy,Mannerisms,Verbigeration,Negativism,Waxy flexibility | 96.9 | 95.1 | 98.4 |
| 8 | Immobility/stupor,Posturing/catalepsy,Grimacing,Stereotypy,Mannerisms,Verbigeration,Rigidity,Withdrawal | 96.9 | 95.1 | 98.4 |
| 8 | Immobility/stupor,Posturing/catalepsy,Grimacing,Stereotypy,Verbigeration,Negativism,Waxy flexibility,Withdrawal | 96.9 | 95.1 | 98.4 |
| 8 | Immobility/stupor,Staring,Posturing/catalepsy,Echopraxia/echolalia,Stereotypy,Verbigeration,Rigidity,Waxy flexibility | 96.9 | 95.1 | 98.4 |
| 8 | Immobility/stupor,Staring,Posturing/catalepsy,Grimacing,Echopraxia/echolalia,Rigidity,Waxy flexibility,Withdrawal | 96.9 | 95.1 | 98.4 |
| 8 | Immobility/stupor,Staring,Posturing/catalepsy,Grimacing,Echopraxia/echolalia,Stereotypy,Verbigeration,Withdrawal | 96.9 | 95.1 | 98.4 |
| 8 | Immobility/stupor,Staring,Posturing/catalepsy,Grimacing,Echopraxia/echolalia,Stereotypy,Waxy flexibility,Withdrawal | 96.9 | 95.1 | 98.4 |
| 8 | Immobility/stupor,Staring,Posturing/catalepsy,Grimacing,Stereotypy,Verbigeration,Waxy flexibility,Withdrawal | 96.9 | 95.1 | 98.4 |
| 8 | Immobility/stupor,Staring,Posturing/catalepsy,Grimacing,Verbigeration,Rigidity,Waxy flexibility,Withdrawal | 96.9 | 95.1 | 98.4 |
| 8 | Staring,Grimacing,Echopraxia/echolalia,Stereotypy,Mannerisms,Verbigeration,Negativism,Waxy flexibility | 96.9 | 95.1 | 98.4 |
| 8 | Staring,Posturing/catalepsy,Echopraxia/echolalia,Stereotypy,Verbigeration,Rigidity,Negativism,Waxy flexibility | 96.9 | 95.1 | 98.4 |
| 8 | Staring,Posturing/catalepsy,Grimacing,Echopraxia/echolalia,Mannerisms,Verbigeration,Negativism,Waxy flexibility | 96.9 | 95.1 | 98.4 |
| 8 | Staring,Posturing/catalepsy,Grimacing,Stereotypy,Mannerisms,Verbigeration,Rigidity,Waxy flexibility | 96.9 | 95.1 | 98.4 |
| 8 | Staring,Posturing/catalepsy,Grimacing,Stereotypy,Verbigeration,Rigidity,Negativism,Waxy flexibility | 96.9 | 95.1 | 98.4 |
| 8 | Excitement,Immobility/stupor,Echopraxia/echolalia,Stereotypy,Mannerisms,Verbigeration,Negativism,Waxy flexibility | 96.8 | 95.1 | 98.4 |
| 8 | Excitement,Immobility/stupor,Grimacing,Mannerisms,Verbigeration,Negativism,Waxy flexibility,Withdrawal | 96.8 | 95.1 | 98.4 |
| 8 | Excitement,Immobility/stupor,Mutism,Grimacing,Echopraxia/echolalia,Mannerisms,Waxy flexibility,Withdrawal | 96.8 | 95.1 | 98.4 |
| 8 | Excitement,Mutism,Posturing/catalepsy,Mannerisms,Verbigeration,Rigidity,Negativism,Withdrawal | 96.8 | 95.1 | 98.4 |
| 8 | Immobility/stupor,Mutism,Echopraxia/echolalia,Stereotypy,Mannerisms,Verbigeration,Rigidity,Withdrawal | 96.8 | 95.1 | 98.4 |
| 8 | Immobility/stupor,Mutism,Grimacing,Echopraxia/echolalia,Mannerisms,Verbigeration,Negativism,Waxy flexibility | 96.8 | 95.1 | 98.4 |
| 8 | Immobility/stupor,Mutism,Grimacing,Stereotypy,Mannerisms,Verbigeration,Rigidity,Waxy flexibility | 96.8 | 95.1 | 98.4 |
| 8 | Excitement,Immobility/stupor,Grimacing,Stereotypy,Mannerisms,Negativism,Waxy flexibility,Withdrawal | 96.7 | 94.8 | 98.2 |
| 8 | Excitement,Immobility/stupor,Mutism,Grimacing,Echopraxia/echolalia,Stereotypy,Mannerisms,Negativism | 96.7 | 94.8 | 98.2 |
| 8 | Excitement,Mutism,Posturing/catalepsy,Grimacing,Echopraxia/echolalia,Rigidity,Negativism,Withdrawal | 96.7 | 94.8 | 98.2 |
| 8 | Excitement,Staring,Posturing/catalepsy,Grimacing,Echopraxia/echolalia,Mannerisms,Verbigeration,Waxy flexibility | 96.7 | 94.8 | 98.2 |
| 8 | Immobility/stupor,Grimacing,Echopraxia/echolalia,Stereotypy,Verbigeration,Negativism,Waxy flexibility,Withdrawal | 96.7 | 94.8 | 98.2 |
| 8 | Immobility/stupor,Mutism,Grimacing,Echopraxia/echolalia,Stereotypy,Mannerisms,Verbigeration,Rigidity | 96.7 | 94.8 | 98.2 |
| 8 | Immobility/stupor,Mutism,Posturing/catalepsy,Stereotypy,Mannerisms,Rigidity,Waxy flexibility,Withdrawal | 96.7 | 94.8 | 98.2 |
| 8 | Immobility/stupor,Posturing/catalepsy,Echopraxia/echolalia,Stereotypy,Mannerisms,Verbigeration,Negativism,Waxy flexibility | 96.7 | 94.8 | 98.2 |
| 8 | Immobility/stupor,Posturing/catalepsy,Grimacing,Stereotypy,Mannerisms,Rigidity,Waxy flexibility,Withdrawal | 96.7 | 94.8 | 98.2 |
| 8 | Immobility/stupor,Staring,Grimacing,Echopraxia/echolalia,Stereotypy,Verbigeration,Waxy flexibility,Withdrawal | 96.7 | 94.8 | 98.2 |
| 8 | Immobility/stupor,Staring,Posturing/catalepsy,Grimacing,Echopraxia/echolalia,Verbigeration,Rigidity,Waxy flexibility | 96.7 | 94.8 | 98.2 |
| 8 | Excitement,Immobility/stupor,Echopraxia/echolalia,Stereotypy,Mannerisms,Verbigeration,Waxy flexibility,Withdrawal | 96.6 | 94.8 | 98.2 |
| 8 | Excitement,Immobility/stupor,Grimacing,Echopraxia/echolalia,Stereotypy,Mannerisms,Waxy flexibility,Withdrawal | 96.6 | 94.8 | 98.2 |
| 8 | Excitement,Immobility/stupor,Grimacing,Stereotypy,Mannerisms,Verbigeration,Negativism,Waxy flexibility | 96.6 | 94.8 | 98.2 |
| 8 | Excitement,Immobility/stupor,Mutism,Echopraxia/echolalia,Mannerisms,Verbigeration,Waxy flexibility,Withdrawal | 96.6 | 94.8 | 98.2 |
| 8 | Excitement,Immobility/stupor,Mutism,Grimacing,Echopraxia/echolalia,Mannerisms,Verbigeration,Negativism | 96.6 | 94.8 | 98.2 |
| 8 | Excitement,Immobility/stupor,Mutism,Grimacing,Echopraxia/echolalia,Stereotypy,Mannerisms,Withdrawal | 96.6 | 94.8 | 98.2 |
| 8 | Excitement,Immobility/stupor,Mutism,Grimacing,Echopraxia/echolalia,Stereotypy,Verbigeration,Waxy flexibility | 96.6 | 94.8 | 98.2 |
| 8 | Excitement,Immobility/stupor,Mutism,Posturing/catalepsy,Echopraxia/echolalia,Stereotypy,Mannerisms,Verbigeration | 96.6 | 94.8 | 98.2 |
| 8 | Excitement,Immobility/stupor,Mutism,Posturing/catalepsy,Grimacing,Echopraxia/echolalia,Stereotypy,Mannerisms | 96.6 | 94.8 | 98.2 |
| 8 | Excitement,Immobility/stupor,Mutism,Posturing/catalepsy,Grimacing,Stereotypy,Mannerisms,Verbigeration | 96.6 | 94.8 | 98.2 |
| 8 | Excitement,Immobility/stupor,Mutism,Stereotypy,Mannerisms,Verbigeration,Waxy flexibility,Withdrawal | 96.6 | 94.8 | 98.2 |
| 8 | Excitement,Immobility/stupor,Posturing/catalepsy,Echopraxia/echolalia,Stereotypy,Mannerisms,Waxy flexibility,Withdrawal | 96.6 | 94.8 | 98.2 |
| 8 | Excitement,Immobility/stupor,Posturing/catalepsy,Grimacing,Stereotypy,Mannerisms,Verbigeration,Withdrawal | 96.6 | 94.8 | 98.2 |
| 8 | Excitement,Immobility/stupor,Posturing/catalepsy,Stereotypy,Mannerisms,Verbigeration,Waxy flexibility,Withdrawal | 96.6 | 94.8 | 98.2 |
| 8 | Excitement,Mutism,Posturing/catalepsy,Grimacing,Verbigeration,Rigidity,Negativism,Withdrawal | 96.6 | 94.8 | 98.2 |
| 8 | Excitement,Mutism,Posturing/catalepsy,Verbigeration,Rigidity,Negativism,Waxy flexibility,Withdrawal | 96.6 | 94.8 | 98.2 |
| 8 | Excitement,Mutism,Stereotypy,Verbigeration,Rigidity,Negativism,Waxy flexibility,Withdrawal | 96.6 | 94.8 | 98.2 |
| 8 | Excitement,Staring,Grimacing,Echopraxia/echolalia,Stereotypy,Mannerisms,Verbigeration,Negativism | 96.6 | 94.8 | 98.2 |
| 8 | Excitement,Staring,Posturing/catalepsy,Grimacing,Echopraxia/echolalia,Stereotypy,Mannerisms,Verbigeration | 96.6 | 94.8 | 98.2 |
| 8 | Excitement,Staring,Posturing/catalepsy,Grimacing,Stereotypy,Mannerisms,Verbigeration,Waxy flexibility | 96.6 | 94.8 | 98.2 |
| 8 | Immobility/stupor,Grimacing,Echopraxia/echolalia,Mannerisms,Verbigeration,Rigidity,Waxy flexibility,Withdrawal | 96.6 | 94.8 | 98.2 |
| 8 | Immobility/stupor,Grimacing,Echopraxia/echolalia,Stereotypy,Verbigeration,Rigidity,Waxy flexibility,Withdrawal | 96.6 | 94.8 | 98.2 |
| 8 | Immobility/stupor,Grimacing,Stereotypy,Mannerisms,Verbigeration,Rigidity,Waxy flexibility,Withdrawal | 96.6 | 94.8 | 98.2 |
| 8 | Immobility/stupor,Mutism,Echopraxia/echolalia,Mannerisms,Verbigeration,Rigidity,Waxy flexibility,Withdrawal | 96.6 | 94.8 | 98.2 |
| 8 | Immobility/stupor,Mutism,Echopraxia/echolalia,Stereotypy,Verbigeration,Negativism,Waxy flexibility,Withdrawal | 96.6 | 94.8 | 98.2 |
| 8 | Immobility/stupor,Mutism,Grimacing,Echopraxia/echolalia,Mannerisms,Verbigeration,Rigidity,Waxy flexibility | 96.6 | 94.8 | 98.2 |
| 8 | Immobility/stupor,Mutism,Grimacing,Echopraxia/echolalia,Stereotypy,Mannerisms,Rigidity,Waxy flexibility | 96.6 | 94.8 | 98.2 |
| 8 | Immobility/stupor,Mutism,Grimacing,Echopraxia/echolalia,Stereotypy,Mannerisms,Verbigeration,Negativism | 96.6 | 94.8 | 98.2 |
| 8 | Immobility/stupor,Mutism,Grimacing,Echopraxia/echolalia,Stereotypy,Negativism,Waxy flexibility,Withdrawal | 96.6 | 94.8 | 98.2 |
| 8 | Immobility/stupor,Mutism,Grimacing,Mannerisms,Verbigeration,Negativism,Waxy flexibility,Withdrawal | 96.6 | 94.8 | 98.2 |
| 8 | Immobility/stupor,Mutism,Grimacing,Stereotypy,Mannerisms,Verbigeration,Negativism,Waxy flexibility | 96.6 | 94.8 | 98.2 |
| 8 | Immobility/stupor,Mutism,Posturing/catalepsy,Echopraxia/echolalia,Mannerisms,Rigidity,Waxy flexibility,Withdrawal | 96.6 | 94.8 | 98.2 |
| 8 | Immobility/stupor,Mutism,Posturing/catalepsy,Echopraxia/echolalia,Stereotypy,Mannerisms,Verbigeration,Rigidity | 96.6 | 94.8 | 98.2 |
| 8 | Immobility/stupor,Mutism,Posturing/catalepsy,Echopraxia/echolalia,Stereotypy,Negativism,Waxy flexibility,Withdrawal | 96.6 | 94.8 | 98.2 |
| 8 | Immobility/stupor,Mutism,Posturing/catalepsy,Echopraxia/echolalia,Stereotypy,Verbigeration,Rigidity,Withdrawal | 96.6 | 94.8 | 98.2 |
| 8 | Immobility/stupor,Mutism,Posturing/catalepsy,Echopraxia/echolalia,Verbigeration,Negativism,Waxy flexibility,Withdrawal | 96.6 | 94.8 | 98.2 |
| 8 | Immobility/stupor,Mutism,Posturing/catalepsy,Grimacing,Echopraxia/echolalia,Mannerisms,Verbigeration,Withdrawal | 96.6 | 94.8 | 98.2 |
| 8 | Immobility/stupor,Mutism,Posturing/catalepsy,Grimacing,Echopraxia/echolalia,Mannerisms,Waxy flexibility,Withdrawal | 96.6 | 94.8 | 98.2 |
| 8 | Immobility/stupor,Mutism,Posturing/catalepsy,Grimacing,Echopraxia/echolalia,Stereotypy,Mannerisms,Withdrawal | 96.6 | 94.8 | 98.2 |
| 8 | Immobility/stupor,Mutism,Posturing/catalepsy,Grimacing,Stereotypy,Negativism,Waxy flexibility,Withdrawal | 96.6 | 94.8 | 98.2 |
| 8 | Immobility/stupor,Mutism,Posturing/catalepsy,Grimacing,Stereotypy,Verbigeration,Negativism,Waxy flexibility | 96.6 | 94.8 | 98.2 |
| 8 | Immobility/stupor,Mutism,Posturing/catalepsy,Grimacing,Stereotypy,Verbigeration,Rigidity,Waxy flexibility | 96.6 | 94.8 | 98.2 |
| 8 | Immobility/stupor,Mutism,Posturing/catalepsy,Grimacing,Verbigeration,Negativism,Waxy flexibility,Withdrawal | 96.6 | 94.8 | 98.2 |
| 8 | Immobility/stupor,Mutism,Posturing/catalepsy,Mannerisms,Verbigeration,Rigidity,Waxy flexibility,Withdrawal | 96.6 | 94.8 | 98.2 |
| 8 | Immobility/stupor,Mutism,Staring,Echopraxia/echolalia,Stereotypy,Verbigeration,Waxy flexibility,Withdrawal | 96.6 | 94.8 | 98.2 |
| 8 | Immobility/stupor,Mutism,Staring,Posturing/catalepsy,Echopraxia/echolalia,Verbigeration,Waxy flexibility,Withdrawal | 96.6 | 94.8 | 98.2 |
| 8 | Immobility/stupor,Mutism,Staring,Posturing/catalepsy,Grimacing,Rigidity,Negativism,Waxy flexibility | 96.6 | 94.8 | 98.2 |
| 8 | Immobility/stupor,Mutism,Staring,Posturing/catalepsy,Grimacing,Rigidity,Waxy flexibility,Withdrawal | 96.6 | 94.8 | 98.2 |
| 8 | Immobility/stupor,Posturing/catalepsy,Echopraxia/echolalia,Stereotypy,Verbigeration,Negativism,Waxy flexibility,Withdrawal | 96.6 | 94.8 | 98.2 |
| 8 | Immobility/stupor,Posturing/catalepsy,Grimacing,Echopraxia/echolalia,Stereotypy,Verbigeration,Rigidity,Withdrawal | 96.6 | 94.8 | 98.2 |
| 8 | Immobility/stupor,Staring,Echopraxia/echolalia,Stereotypy,Mannerisms,Verbigeration,Waxy flexibility,Withdrawal | 96.6 | 94.8 | 98.2 |
| 8 | Immobility/stupor,Staring,Posturing/catalepsy,Echopraxia/echolalia,Mannerisms,Verbigeration,Waxy flexibility,Withdrawal | 96.6 | 94.8 | 98.2 |
| 8 | Immobility/stupor,Staring,Posturing/catalepsy,Echopraxia/echolalia,Stereotypy,Mannerisms,Verbigeration,Waxy flexibility | 96.6 | 94.8 | 98.2 |
| 8 | Immobility/stupor,Staring,Posturing/catalepsy,Echopraxia/echolalia,Verbigeration,Rigidity,Waxy flexibility,Withdrawal | 96.6 | 94.8 | 98.2 |
| 8 | Immobility/stupor,Staring,Posturing/catalepsy,Grimacing,Echopraxia/echolalia,Stereotypy,Rigidity,Waxy flexibility | 96.6 | 94.8 | 98.2 |
| 8 | Staring,Posturing/catalepsy,Echopraxia/echolalia,Stereotypy,Verbigeration,Rigidity,Waxy flexibility,Withdrawal | 96.6 | 94.8 | 98.2 |
| 8 | Staring,Posturing/catalepsy,Grimacing,Echopraxia/echolalia,Stereotypy,Mannerisms,Rigidity,Waxy flexibility | 96.6 | 94.8 | 98.2 |
| 8 | Staring,Posturing/catalepsy,Grimacing,Echopraxia/echolalia,Verbigeration,Rigidity,Waxy flexibility,Withdrawal | 96.6 | 94.8 | 98.2 |
| 8 | Excitement,Immobility/stupor,Grimacing,Echopraxia/echolalia,Mannerisms,Verbigeration,Waxy flexibility,Withdrawal | 96.4 | 94.6 | 98.0 |
| 8 | Excitement,Immobility/stupor,Grimacing,Echopraxia/echolalia,Stereotypy,Mannerisms,Verbigeration,Negativism | 96.4 | 94.6 | 98.0 |
| 8 | Excitement,Immobility/stupor,Grimacing,Echopraxia/echolalia,Stereotypy,Verbigeration,Waxy flexibility,Withdrawal | 96.4 | 94.6 | 98.0 |
| 8 | Excitement,Immobility/stupor,Mutism,Grimacing,Echopraxia/echolalia,Mannerisms,Verbigeration,Withdrawal | 96.4 | 94.6 | 98.0 |
| 8 | Excitement,Immobility/stupor,Mutism,Grimacing,Mannerisms,Verbigeration,Waxy flexibility,Withdrawal | 96.4 | 94.6 | 98.0 |
| 8 | Excitement,Immobility/stupor,Mutism,Grimacing,Stereotypy,Mannerisms,Verbigeration,Withdrawal | 96.4 | 94.6 | 98.0 |
| 8 | Excitement,Immobility/stupor,Mutism,Grimacing,Stereotypy,Mannerisms,Waxy flexibility,Withdrawal | 96.4 | 94.6 | 98.0 |
| 8 | Excitement,Immobility/stupor,Mutism,Posturing/catalepsy,Echopraxia/echolalia,Stereotypy,Mannerisms,Waxy flexibility | 96.4 | 94.6 | 98.0 |
| 8 | Excitement,Immobility/stupor,Mutism,Posturing/catalepsy,Grimacing,Stereotypy,Mannerisms,Waxy flexibility | 96.4 | 94.6 | 98.0 |
| 8 | Excitement,Immobility/stupor,Mutism,Posturing/catalepsy,Stereotypy,Mannerisms,Verbigeration,Waxy flexibility | 96.4 | 94.6 | 98.0 |
| 8 | Excitement,Immobility/stupor,Posturing/catalepsy,Grimacing,Echopraxia/echolalia,Mannerisms,Verbigeration,Waxy flexibility | 96.4 | 94.6 | 98.0 |
| 8 | Excitement,Immobility/stupor,Posturing/catalepsy,Grimacing,Stereotypy,Verbigeration,Waxy flexibility,Withdrawal | 96.4 | 94.6 | 98.0 |
| 8 | Excitement,Mutism,Echopraxia/echolalia,Stereotypy,Verbigeration,Rigidity,Negativism,Withdrawal | 96.4 | 94.6 | 98.0 |
| 8 | Excitement,Mutism,Posturing/catalepsy,Echopraxia/echolalia,Mannerisms,Rigidity,Negativism,Withdrawal | 96.4 | 94.6 | 98.0 |
| 8 | Excitement,Mutism,Posturing/catalepsy,Echopraxia/echolalia,Rigidity,Negativism,Waxy flexibility,Withdrawal | 96.4 | 94.6 | 98.0 |
| 8 | Excitement,Mutism,Posturing/catalepsy,Echopraxia/echolalia,Stereotypy,Rigidity,Negativism,Withdrawal | 96.4 | 94.6 | 98.0 |
| 8 | Excitement,Mutism,Posturing/catalepsy,Grimacing,Mannerisms,Rigidity,Negativism,Withdrawal | 96.4 | 94.6 | 98.0 |
| 8 | Excitement,Staring,Grimacing,Echopraxia/echolalia,Stereotypy,Mannerisms,Verbigeration,Waxy flexibility | 96.4 | 94.6 | 98.0 |
| 8 | Excitement,Staring,Posturing/catalepsy,Grimacing,Echopraxia/echolalia,Stereotypy,Verbigeration,Waxy flexibility | 96.4 | 94.6 | 98.0 |
| 8 | Immobility/stupor,Grimacing,Echopraxia/echolalia,Mannerisms,Verbigeration,Negativism,Waxy flexibility,Withdrawal | 96.4 | 94.6 | 98.0 |
| 8 | Immobility/stupor,Mutism,Echopraxia/echolalia,Stereotypy,Verbigeration,Rigidity,Waxy flexibility,Withdrawal | 96.4 | 94.6 | 98.0 |
| 8 | Immobility/stupor,Mutism,Grimacing,Echopraxia/echolalia,Stereotypy,Verbigeration,Negativism,Waxy flexibility | 96.4 | 94.6 | 98.0 |
| 8 | Immobility/stupor,Mutism,Grimacing,Echopraxia/echolalia,Stereotypy,Verbigeration,Rigidity,Waxy flexibility | 96.4 | 94.6 | 98.0 |
| 8 | Immobility/stupor,Mutism,Grimacing,Echopraxia/echolalia,Verbigeration,Negativism,Waxy flexibility,Withdrawal | 96.4 | 94.6 | 98.0 |
| 8 | Immobility/stupor,Mutism,Grimacing,Stereotypy,Verbigeration,Negativism,Waxy flexibility,Withdrawal | 96.4 | 94.6 | 98.0 |
| 8 | Immobility/stupor,Mutism,Posturing/catalepsy,Echopraxia/echolalia,Stereotypy,Verbigeration,Negativism,Waxy flexibility | 96.4 | 94.6 | 98.0 |
| 8 | Immobility/stupor,Mutism,Posturing/catalepsy,Grimacing,Echopraxia/echolalia,Mannerisms,Rigidity,Waxy flexibility | 96.4 | 94.6 | 98.0 |
| 8 | Immobility/stupor,Mutism,Posturing/catalepsy,Grimacing,Echopraxia/echolalia,Rigidity,Waxy flexibility,Withdrawal | 96.4 | 94.6 | 98.0 |
| 8 | Immobility/stupor,Mutism,Posturing/catalepsy,Grimacing,Echopraxia/echolalia,Stereotypy,Negativism,Waxy flexibility | 96.4 | 94.6 | 98.0 |
| 8 | Immobility/stupor,Mutism,Posturing/catalepsy,Grimacing,Mannerisms,Verbigeration,Rigidity,Waxy flexibility | 96.4 | 94.6 | 98.0 |
| 8 | Immobility/stupor,Mutism,Posturing/catalepsy,Grimacing,Stereotypy,Mannerisms,Rigidity,Waxy flexibility | 96.4 | 94.6 | 98.0 |
| 8 | Immobility/stupor,Mutism,Posturing/catalepsy,Grimacing,Stereotypy,Rigidity,Waxy flexibility,Withdrawal | 96.4 | 94.6 | 98.0 |
| 8 | Immobility/stupor,Mutism,Posturing/catalepsy,Grimacing,Verbigeration,Rigidity,Waxy flexibility,Withdrawal | 96.4 | 94.6 | 98.0 |
| 8 | Immobility/stupor,Mutism,Posturing/catalepsy,Stereotypy,Verbigeration,Rigidity,Waxy flexibility,Withdrawal | 96.4 | 94.6 | 98.0 |
| 8 | Immobility/stupor,Mutism,Staring,Posturing/catalepsy,Grimacing,Echopraxia/echolalia,Rigidity,Waxy flexibility | 96.4 | 94.6 | 98.0 |
| 8 | Immobility/stupor,Posturing/catalepsy,Echopraxia/echolalia,Stereotypy,Mannerisms,Verbigeration,Rigidity,Withdrawal | 96.4 | 94.6 | 98.0 |
| 8 | Immobility/stupor,Posturing/catalepsy,Grimacing,Echopraxia/echolalia,Stereotypy,Rigidity,Waxy flexibility,Withdrawal | 96.4 | 94.6 | 98.0 |
| 8 | Immobility/stupor,Posturing/catalepsy,Grimacing,Echopraxia/echolalia,Stereotypy,Verbigeration,Negativism,Waxy flexibility | 96.4 | 94.6 | 98.0 |
| 8 | Immobility/stupor,Posturing/catalepsy,Grimacing,Echopraxia/echolalia,Verbigeration,Negativism,Waxy flexibility,Withdrawal | 96.4 | 94.6 | 98.0 |
| 8 | Immobility/stupor,Posturing/catalepsy,Grimacing,Mannerisms,Verbigeration,Rigidity,Waxy flexibility,Withdrawal | 96.4 | 94.6 | 98.0 |
| 8 | Immobility/stupor,Staring,Posturing/catalepsy,Grimacing,Echopraxia/echolalia,Stereotypy,Verbigeration,Waxy flexibility | 96.4 | 94.6 | 98.0 |
| 8 | Mutism,Posturing/catalepsy,Stereotypy,Mannerisms,Verbigeration,Rigidity,Negativism,Withdrawal | 96.4 | 94.6 | 98.0 |
| 8 | Mutism,Stereotypy,Mannerisms,Verbigeration,Rigidity,Negativism,Waxy flexibility,Withdrawal | 96.4 | 94.6 | 98.0 |
| 8 | Staring,Posturing/catalepsy,Echopraxia/echolalia,Stereotypy,Mannerisms,Verbigeration,Waxy flexibility,Withdrawal | 96.4 | 94.6 | 98.0 |
| 8 | Staring,Posturing/catalepsy,Grimacing,Echopraxia/echolalia,Mannerisms,Verbigeration,Rigidity,Waxy flexibility | 96.4 | 94.6 | 98.0 |
| 8 | Staring,Posturing/catalepsy,Grimacing,Echopraxia/echolalia,Stereotypy,Rigidity,Negativism,Waxy flexibility | 96.4 | 94.6 | 98.0 |
| 8 | Staring,Posturing/catalepsy,Grimacing,Echopraxia/echolalia,Verbigeration,Rigidity,Negativism,Waxy flexibility | 96.4 | 94.6 | 98.0 |
| 8 | Excitement,Immobility/stupor,Grimacing,Echopraxia/echolalia,Stereotypy,Mannerisms,Verbigeration,Withdrawal | 96.2 | 94.4 | 97.8 |
| 8 | Excitement,Immobility/stupor,Mutism,Echopraxia/echolalia,Stereotypy,Mannerisms,Verbigeration,Waxy flexibility | 96.2 | 94.4 | 97.8 |
| 8 | Excitement,Immobility/stupor,Mutism,Grimacing,Echopraxia/echolalia,Mannerisms,Verbigeration,Waxy flexibility | 96.2 | 94.4 | 97.8 |
| 8 | Excitement,Immobility/stupor,Mutism,Grimacing,Echopraxia/echolalia,Stereotypy,Mannerisms,Waxy flexibility | 96.2 | 94.4 | 98.0 |
| 8 | Excitement,Immobility/stupor,Posturing/catalepsy,Grimacing,Stereotypy,Mannerisms,Waxy flexibility,Withdrawal | 96.2 | 94.4 | 97.8 |
| 8 | Excitement,Mutism,Echopraxia/echolalia,Stereotypy,Rigidity,Negativism,Waxy flexibility,Withdrawal | 96.2 | 94.4 | 97.8 |
| 8 | Excitement,Mutism,Grimacing,Stereotypy,Verbigeration,Rigidity,Negativism,Withdrawal | 96.2 | 94.4 | 97.8 |
| 8 | Excitement,Mutism,Posturing/catalepsy,Echopraxia/echolalia,Verbigeration,Rigidity,Waxy flexibility,Withdrawal | 96.2 | 94.4 | 97.8 |
| 8 | Excitement,Mutism,Posturing/catalepsy,Grimacing,Echopraxia/echolalia,Verbigeration,Rigidity,Negativism | 96.2 | 94.4 | 97.8 |
| 8 | Excitement,Mutism,Posturing/catalepsy,Grimacing,Rigidity,Negativism,Waxy flexibility,Withdrawal | 96.2 | 94.4 | 97.8 |
| 8 | Excitement,Mutism,Posturing/catalepsy,Grimacing,Stereotypy,Rigidity,Negativism,Withdrawal | 96.2 | 94.4 | 97.8 |
| 8 | Excitement,Mutism,Posturing/catalepsy,Mannerisms,Rigidity,Negativism,Waxy flexibility,Withdrawal | 96.2 | 94.2 | 97.8 |
| 8 | Excitement,Mutism,Posturing/catalepsy,Mannerisms,Verbigeration,Rigidity,Waxy flexibility,Withdrawal | 96.2 | 94.4 | 97.8 |
| 8 | Excitement,Mutism,Posturing/catalepsy,Stereotypy,Mannerisms,Rigidity,Negativism,Withdrawal | 96.2 | 94.4 | 97.8 |
| 8 | Excitement,Mutism,Posturing/catalepsy,Stereotypy,Rigidity,Negativism,Waxy flexibility,Withdrawal | 96.2 | 94.4 | 98.0 |
| 8 | Excitement,Mutism,Posturing/catalepsy,Stereotypy,Verbigeration,Rigidity,Waxy flexibility,Withdrawal | 96.2 | 94.4 | 98.0 |
| 8 | Excitement,Mutism,Stereotypy,Mannerisms,Verbigeration,Rigidity,Negativism,Withdrawal | 96.2 | 94.2 | 97.8 |
| 8 | Excitement,Staring,Posturing/catalepsy,Echopraxia/echolalia,Stereotypy,Mannerisms,Verbigeration,Waxy flexibility | 96.2 | 94.4 | 98.0 |
| 8 | Immobility/stupor,Echopraxia/echolalia,Stereotypy,Mannerisms,Verbigeration,Rigidity,Waxy flexibility,Withdrawal | 96.2 | 94.4 | 97.8 |
| 8 | Immobility/stupor,Mutism,Echopraxia/echolalia,Stereotypy,Mannerisms,Verbigeration,Rigidity,Waxy flexibility | 96.2 | 94.4 | 97.8 |
| 8 | Immobility/stupor,Mutism,Grimacing,Echopraxia/echolalia,Stereotypy,Mannerisms,Waxy flexibility,Withdrawal | 96.2 | 94.2 | 97.8 |
| 8 | Immobility/stupor,Mutism,Posturing/catalepsy,Echopraxia/echolalia,Mannerisms,Verbigeration,Rigidity,Waxy flexibility | 96.2 | 94.4 | 98.0 |
| 8 | Immobility/stupor,Mutism,Posturing/catalepsy,Echopraxia/echolalia,Stereotypy,Rigidity,Waxy flexibility,Withdrawal | 96.2 | 94.4 | 97.8 |
| 8 | Immobility/stupor,Mutism,Posturing/catalepsy,Grimacing,Echopraxia/echolalia,Verbigeration,Rigidity,Waxy flexibility | 96.2 | 94.4 | 98.0 |
| 8 | Immobility/stupor,Mutism,Posturing/catalepsy,Stereotypy,Mannerisms,Verbigeration,Rigidity,Waxy flexibility | 96.2 | 94.4 | 97.8 |
| 8 | Immobility/stupor,Posturing/catalepsy,Echopraxia/echolalia,Stereotypy,Mannerisms,Rigidity,Waxy flexibility,Withdrawal | 96.2 | 94.4 | 97.8 |
| 8 | Immobility/stupor,Posturing/catalepsy,Grimacing,Stereotypy,Verbigeration,Rigidity,Waxy flexibility,Withdrawal | 96.2 | 94.4 | 98.0 |
| 8 | Mutism,Posturing/catalepsy,Echopraxia/echolalia,Mannerisms,Verbigeration,Rigidity,Negativism,Withdrawal | 96.2 | 94.4 | 97.8 |
| 8 | Mutism,Posturing/catalepsy,Grimacing,Echopraxia/echolalia,Mannerisms,Rigidity,Negativism,Withdrawal | 96.2 | 94.4 | 97.8 |
| 8 | Mutism,Posturing/catalepsy,Grimacing,Mannerisms,Verbigeration,Rigidity,Negativism,Withdrawal | 96.2 | 94.4 | 97.8 |
| 8 | Mutism,Posturing/catalepsy,Mannerisms,Verbigeration,Rigidity,Negativism,Waxy flexibility,Withdrawal | 96.2 | 94.2 | 97.8 |
| 8 | Staring,Posturing/catalepsy,Grimacing,Echopraxia/echolalia,Stereotypy,Verbigeration,Negativism,Waxy flexibility | 96.2 | 94.4 | 97.8 |
| 8 | Staring,Posturing/catalepsy,Grimacing,Echopraxia/echolalia,Stereotypy,Verbigeration,Waxy flexibility,Withdrawal | 96.2 | 94.4 | 97.8 |
| 8 | Excitement,Immobility/stupor,Posturing/catalepsy,Grimacing,Echopraxia/echolalia,Stereotypy,Mannerisms,Waxy flexibility | 96.0 | 93.9 | 97.8 |
| 8 | Excitement,Immobility/stupor,Posturing/catalepsy,Grimacing,Echopraxia/echolalia,Stereotypy,Verbigeration,Waxy flexibility | 96.0 | 94.2 | 97.8 |
| 8 | Excitement,Mutism,Echopraxia/echolalia,Stereotypy,Mannerisms,Rigidity,Negativism,Withdrawal | 96.0 | 94.2 | 97.8 |
| 8 | Excitement,Mutism,Echopraxia/echolalia,Verbigeration,Rigidity,Negativism,Waxy flexibility,Withdrawal | 96.0 | 94.2 | 97.8 |
| 8 | Excitement,Mutism,Grimacing,Echopraxia/echolalia,Rigidity,Negativism,Waxy flexibility,Withdrawal | 96.0 | 94.2 | 97.8 |
| 8 | Excitement,Mutism,Grimacing,Echopraxia/echolalia,Stereotypy,Rigidity,Negativism,Withdrawal | 96.0 | 94.2 | 97.8 |
| 8 | Excitement,Mutism,Grimacing,Echopraxia/echolalia,Verbigeration,Rigidity,Negativism,Withdrawal | 96.0 | 93.9 | 97.8 |
| 8 | Excitement,Mutism,Grimacing,Mannerisms,Verbigeration,Rigidity,Negativism,Withdrawal | 96.0 | 94.2 | 97.8 |
| 8 | Excitement,Mutism,Grimacing,Stereotypy,Rigidity,Negativism,Waxy flexibility,Withdrawal | 96.0 | 93.9 | 97.8 |
| 8 | Excitement,Mutism,Grimacing,Verbigeration,Rigidity,Negativism,Waxy flexibility,Withdrawal | 96.0 | 94.2 | 97.8 |
| 8 | Excitement,Mutism,Mannerisms,Verbigeration,Rigidity,Negativism,Waxy flexibility,Withdrawal | 96.0 | 93.9 | 97.8 |
| 8 | Excitement,Mutism,Posturing/catalepsy,Echopraxia/echolalia,Mannerisms,Verbigeration,Rigidity,Negativism | 96.0 | 93.9 | 97.8 |
| 8 | Excitement,Mutism,Posturing/catalepsy,Echopraxia/echolalia,Stereotypy,Verbigeration,Rigidity,Negativism | 96.0 | 93.9 | 97.8 |
| 8 | Excitement,Mutism,Posturing/catalepsy,Echopraxia/echolalia,Verbigeration,Rigidity,Negativism,Waxy flexibility | 96.0 | 93.9 | 97.8 |
| 8 | Excitement,Mutism,Posturing/catalepsy,Grimacing,Echopraxia/echolalia,Rigidity,Waxy flexibility,Withdrawal | 96.0 | 93.9 | 97.8 |
| 8 | Excitement,Mutism,Posturing/catalepsy,Grimacing,Mannerisms,Verbigeration,Rigidity,Negativism | 96.0 | 93.9 | 97.5 |
| 8 | Excitement,Mutism,Posturing/catalepsy,Grimacing,Verbigeration,Rigidity,Waxy flexibility,Withdrawal | 96.0 | 93.9 | 97.8 |
| 8 | Excitement,Mutism,Stereotypy,Mannerisms,Rigidity,Negativism,Waxy flexibility,Withdrawal | 96.0 | 94.2 | 97.8 |
| 8 | Immobility/stupor,Grimacing,Echopraxia/echolalia,Stereotypy,Mannerisms,Verbigeration,Rigidity,Waxy flexibility | 96.0 | 94.2 | 97.8 |
| 8 | Immobility/stupor,Mutism,Posturing/catalepsy,Echopraxia/echolalia,Stereotypy,Mannerisms,Rigidity,Waxy flexibility | 96.0 | 94.2 | 97.5 |
| 8 | Immobility/stupor,Mutism,Posturing/catalepsy,Echopraxia/echolalia,Stereotypy,Verbigeration,Rigidity,Waxy flexibility | 96.0 | 93.9 | 97.8 |
| 8 | Immobility/stupor,Mutism,Posturing/catalepsy,Echopraxia/echolalia,Verbigeration,Rigidity,Waxy flexibility,Withdrawal | 96.0 | 93.9 | 97.8 |
| 8 | Immobility/stupor,Mutism,Posturing/catalepsy,Grimacing,Echopraxia/echolalia,Stereotypy,Rigidity,Waxy flexibility | 96.0 | 93.9 | 97.8 |
| 8 | Immobility/stupor,Mutism,Posturing/catalepsy,Grimacing,Echopraxia/echolalia,Stereotypy,Verbigeration,Withdrawal | 96.0 | 93.9 | 97.8 |
| 8 | Immobility/stupor,Mutism,Posturing/catalepsy,Grimacing,Echopraxia/echolalia,Stereotypy,Waxy flexibility,Withdrawal | 96.0 | 93.9 | 97.8 |
| 8 | Immobility/stupor,Mutism,Posturing/catalepsy,Grimacing,Mannerisms,Verbigeration,Waxy flexibility,Withdrawal | 96.0 | 93.9 | 97.8 |
| 8 | Immobility/stupor,Mutism,Posturing/catalepsy,Grimacing,Stereotypy,Mannerisms,Verbigeration,Withdrawal | 96.0 | 94.2 | 97.8 |
| 8 | Immobility/stupor,Mutism,Posturing/catalepsy,Grimacing,Stereotypy,Mannerisms,Waxy flexibility,Withdrawal | 96.0 | 93.9 | 97.8 |
| 8 | Immobility/stupor,Posturing/catalepsy,Echopraxia/echolalia,Mannerisms,Verbigeration,Rigidity,Waxy flexibility,Withdrawal | 96.0 | 93.9 | 97.8 |
| 8 | Immobility/stupor,Posturing/catalepsy,Grimacing,Echopraxia/echolalia,Stereotypy,Mannerisms,Verbigeration,Rigidity | 96.0 | 93.9 | 97.8 |
| 8 | Immobility/stupor,Posturing/catalepsy,Grimacing,Echopraxia/echolalia,Stereotypy,Mannerisms,Waxy flexibility,Withdrawal | 96.0 | 93.9 | 97.8 |
| 8 | Immobility/stupor,Posturing/catalepsy,Grimacing,Echopraxia/echolalia,Verbigeration,Rigidity,Waxy flexibility,Withdrawal | 96.0 | 93.9 | 97.8 |
| 8 | Immobility/stupor,Posturing/catalepsy,Stereotypy,Mannerisms,Verbigeration,Rigidity,Waxy flexibility,Withdrawal | 96.0 | 93.9 | 97.8 |
| 8 | Immobility/stupor,Staring,Posturing/catalepsy,Echopraxia/echolalia,Stereotypy,Verbigeration,Waxy flexibility,Withdrawal | 96.0 | 94.2 | 97.8 |
| 8 | Immobility/stupor,Staring,Posturing/catalepsy,Grimacing,Echopraxia/echolalia,Verbigeration,Waxy flexibility,Withdrawal | 96.0 | 93.9 | 97.8 |
| 8 | Mutism,Echopraxia/echolalia,Stereotypy,Mannerisms,Rigidity,Negativism,Waxy flexibility,Withdrawal | 96.0 | 93.9 | 97.8 |
| 8 | Mutism,Echopraxia/echolalia,Stereotypy,Mannerisms,Verbigeration,Rigidity,Negativism,Withdrawal | 96.0 | 93.9 | 97.8 |
| 8 | Mutism,Grimacing,Stereotypy,Mannerisms,Rigidity,Negativism,Waxy flexibility,Withdrawal | 96.0 | 93.9 | 97.8 |
| 8 | Mutism,Posturing/catalepsy,Echopraxia/echolalia,Mannerisms,Rigidity,Negativism,Waxy flexibility,Withdrawal | 96.0 | 93.9 | 97.8 |
| 8 | Mutism,Posturing/catalepsy,Echopraxia/echolalia,Stereotypy,Mannerisms,Rigidity,Negativism,Withdrawal | 96.0 | 93.9 | 97.8 |
| 8 | Mutism,Posturing/catalepsy,Grimacing,Mannerisms,Rigidity,Negativism,Waxy flexibility,Withdrawal | 96.0 | 93.9 | 97.8 |
| 8 | Mutism,Posturing/catalepsy,Grimacing,Stereotypy,Mannerisms,Rigidity,Negativism,Withdrawal | 96.0 | 93.9 | 97.8 |
| 8 | Mutism,Posturing/catalepsy,Stereotypy,Mannerisms,Rigidity,Negativism,Waxy flexibility,Withdrawal | 96.0 | 93.9 | 97.8 |
| 8 | Staring,Posturing/catalepsy,Grimacing,Echopraxia/echolalia,Stereotypy,Verbigeration,Rigidity,Waxy flexibility | 96.0 | 94.2 | 97.8 |
| 8 | Excitement,Immobility/stupor,Mutism,Grimacing,Stereotypy,Mannerisms,Verbigeration,Waxy flexibility | 95.9 | 93.9 | 97.8 |
| 8 | Mutism,Grimacing,Stereotypy,Mannerisms,Verbigeration,Rigidity,Negativism,Withdrawal | 95.9 | 93.9 | 97.8 |
| 8 | Excitement,Mutism,Posturing/catalepsy,Echopraxia/echolalia,Mannerisms,Rigidity,Waxy flexibility,Withdrawal | 95.8 | 93.7 | 97.5 |
| 8 | Immobility/stupor,Mutism,Grimacing,Echopraxia/echolalia,Mannerisms,Verbigeration,Waxy flexibility,Withdrawal | 95.8 | 93.7 | 97.5 |
| 8 | Immobility/stupor,Mutism,Posturing/catalepsy,Grimacing,Echopraxia/echolalia,Mannerisms,Verbigeration,Waxy flexibility | 95.8 | 93.7 | 97.5 |
| 8 | Mutism,Grimacing,Echopraxia/echolalia,Mannerisms,Rigidity,Negativism,Waxy flexibility,Withdrawal | 95.8 | 93.7 | 97.5 |
| 8 | Mutism,Grimacing,Mannerisms,Verbigeration,Rigidity,Negativism,Waxy flexibility,Withdrawal | 95.8 | 93.7 | 97.5 |
| 8 | Mutism,Grimacing,Stereotypy,Verbigeration,Rigidity,Negativism,Waxy flexibility,Withdrawal | 95.8 | 93.7 | 97.5 |
| 8 | Excitement,Immobility/stupor,Grimacing,Stereotypy,Mannerisms,Verbigeration,Waxy flexibility,Withdrawal | 95.7 | 93.7 | 97.5 |
| 8 | Excitement,Immobility/stupor,Posturing/catalepsy,Echopraxia/echolalia,Stereotypy,Mannerisms,Verbigeration,Waxy flexibility | 95.7 | 93.7 | 97.5 |
| 8 | Excitement,Immobility/stupor,Posturing/catalepsy,Grimacing,Echopraxia/echolalia,Stereotypy,Mannerisms,Verbigeration | 95.7 | 93.7 | 97.5 |
| 8 | Excitement,Immobility/stupor,Posturing/catalepsy,Grimacing,Stereotypy,Mannerisms,Verbigeration,Waxy flexibility | 95.7 | 93.7 | 97.5 |
| 8 | Excitement,Mutism,Echopraxia/echolalia,Mannerisms,Verbigeration,Rigidity,Negativism,Withdrawal | 95.7 | 93.7 | 97.5 |
| 8 | Excitement,Mutism,Grimacing,Echopraxia/echolalia,Mannerisms,Rigidity,Negativism,Withdrawal | 95.7 | 93.7 | 97.5 |
| 8 | Excitement,Mutism,Grimacing,Mannerisms,Rigidity,Negativism,Waxy flexibility,Withdrawal | 95.7 | 93.7 | 97.5 |
| 8 | Excitement,Mutism,Posturing/catalepsy,Echopraxia/echolalia,Stereotypy,Rigidity,Waxy flexibility,Withdrawal | 95.7 | 93.7 | 97.5 |
| 8 | Excitement,Mutism,Posturing/catalepsy,Grimacing,Echopraxia/echolalia,Mannerisms,Rigidity,Negativism | 95.7 | 93.7 | 97.5 |
| 8 | Excitement,Mutism,Posturing/catalepsy,Grimacing,Echopraxia/echolalia,Rigidity,Negativism,Waxy flexibility | 95.7 | 93.7 | 97.5 |
| 8 | Excitement,Mutism,Posturing/catalepsy,Grimacing,Mannerisms,Rigidity,Waxy flexibility,Withdrawal | 95.7 | 93.7 | 97.5 |
| 8 | Excitement,Mutism,Posturing/catalepsy,Grimacing,Stereotypy,Verbigeration,Rigidity,Negativism | 95.7 | 93.7 | 97.5 |
| 8 | Excitement,Mutism,Posturing/catalepsy,Grimacing,Verbigeration,Rigidity,Negativism,Waxy flexibility | 95.7 | 93.7 | 97.5 |
| 8 | Excitement,Mutism,Posturing/catalepsy,Mannerisms,Verbigeration,Rigidity,Negativism,Waxy flexibility | 95.7 | 93.7 | 97.5 |
| 8 | Excitement,Mutism,Posturing/catalepsy,Stereotypy,Mannerisms,Verbigeration,Rigidity,Negativism | 95.7 | 93.7 | 97.5 |
| 8 | Excitement,Mutism,Posturing/catalepsy,Stereotypy,Verbigeration,Rigidity,Negativism,Waxy flexibility | 95.7 | 93.7 | 97.5 |
| 8 | Immobility/stupor,Mutism,Grimacing,Echopraxia/echolalia,Stereotypy,Mannerisms,Verbigeration,Withdrawal | 95.7 | 93.7 | 97.5 |
| 8 | Immobility/stupor,Mutism,Posturing/catalepsy,Grimacing,Echopraxia/echolalia,Verbigeration,Waxy flexibility,Withdrawal | 95.7 | 93.7 | 97.5 |
| 8 | Immobility/stupor,Posturing/catalepsy,Grimacing,Echopraxia/echolalia,Mannerisms,Verbigeration,Rigidity,Waxy flexibility | 95.7 | 93.7 | 97.5 |
| 8 | Immobility/stupor,Posturing/catalepsy,Grimacing,Echopraxia/echolalia,Stereotypy,Mannerisms,Rigidity,Waxy flexibility | 95.7 | 93.7 | 97.5 |
| 8 | Immobility/stupor,Posturing/catalepsy,Grimacing,Echopraxia/echolalia,Stereotypy,Mannerisms,Verbigeration,Withdrawal | 95.7 | 93.7 | 97.5 |
| 8 | Immobility/stupor,Posturing/catalepsy,Grimacing,Stereotypy,Mannerisms,Verbigeration,Rigidity,Waxy flexibility | 95.7 | 93.7 | 97.5 |
| 8 | Mutism,Grimacing,Echopraxia/echolalia,Stereotypy,Mannerisms,Rigidity,Negativism,Withdrawal | 95.7 | 93.7 | 97.5 |
| 8 | Mutism,Posturing/catalepsy,Grimacing,Stereotypy,Verbigeration,Rigidity,Negativism,Withdrawal | 95.7 | 93.7 | 97.5 |
| 8 | Mutism,Posturing/catalepsy,Stereotypy,Verbigeration,Rigidity,Negativism,Waxy flexibility,Withdrawal | 95.7 | 93.7 | 97.5 |
| 8 | Staring,Posturing/catalepsy,Echopraxia/echolalia,Stereotypy,Mannerisms,Verbigeration,Rigidity,Waxy flexibility | 95.7 | 93.7 | 97.5 |
| 8 | Staring,Posturing/catalepsy,Grimacing,Echopraxia/echolalia,Stereotypy,Mannerisms,Verbigeration,Waxy flexibility | 95.7 | 93.7 | 97.5 |
| 8 | Excitement,Immobility/stupor,Grimacing,Echopraxia/echolalia,Stereotypy,Mannerisms,Verbigeration,Waxy flexibility | 95.5 | 93.5 | 97.3 |
| 8 | Excitement,Immobility/stupor,Mutism,Grimacing,Echopraxia/echolalia,Stereotypy,Mannerisms,Verbigeration | 95.5 | 93.5 | 97.3 |
| 8 | Excitement,Mutism,Echopraxia/echolalia,Mannerisms,Rigidity,Negativism,Waxy flexibility,Withdrawal | 95.5 | 93.5 | 97.3 |
| 8 | Excitement,Mutism,Echopraxia/echolalia,Stereotypy,Verbigeration,Rigidity,Negativism,Waxy flexibility | 95.5 | 93.5 | 97.3 |
| 8 | Excitement,Mutism,Echopraxia/echolalia,Stereotypy,Verbigeration,Rigidity,Waxy flexibility,Withdrawal | 95.5 | 93.5 | 97.3 |
| 8 | Excitement,Mutism,Grimacing,Stereotypy,Mannerisms,Rigidity,Negativism,Withdrawal | 95.5 | 93.5 | 97.3 |
| 8 | Excitement,Mutism,Posturing/catalepsy,Echopraxia/echolalia,Mannerisms,Verbigeration,Rigidity,Withdrawal | 95.5 | 93.5 | 97.3 |
| 8 | Excitement,Mutism,Posturing/catalepsy,Echopraxia/echolalia,Stereotypy,Verbigeration,Rigidity,Withdrawal | 95.5 | 93.5 | 97.3 |
| 8 | Excitement,Mutism,Posturing/catalepsy,Grimacing,Echopraxia/echolalia,Stereotypy,Rigidity,Negativism | 95.5 | 93.5 | 97.3 |
| 8 | Excitement,Mutism,Posturing/catalepsy,Grimacing,Echopraxia/echolalia,Verbigeration,Rigidity,Withdrawal | 95.5 | 93.5 | 97.3 |
| 8 | Excitement,Mutism,Posturing/catalepsy,Grimacing,Mannerisms,Verbigeration,Rigidity,Withdrawal | 95.5 | 93.5 | 97.3 |
| 8 | Excitement,Mutism,Posturing/catalepsy,Grimacing,Stereotypy,Rigidity,Waxy flexibility,Withdrawal | 95.5 | 93.5 | 97.3 |
| 8 | Excitement,Mutism,Posturing/catalepsy,Stereotypy,Mannerisms,Rigidity,Waxy flexibility,Withdrawal | 95.5 | 93.5 | 97.3 |
| 8 | Excitement,Mutism,Posturing/catalepsy,Stereotypy,Mannerisms,Verbigeration,Rigidity,Withdrawal | 95.5 | 93.5 | 97.3 |
| 8 | Excitement,Mutism,Stereotypy,Mannerisms,Verbigeration,Rigidity,Waxy flexibility,Withdrawal | 95.5 | 93.5 | 97.3 |
| 8 | Immobility/stupor,Mutism,Grimacing,Echopraxia/echolalia,Stereotypy,Verbigeration,Waxy flexibility,Withdrawal | 95.5 | 93.5 | 97.3 |
| 8 | Immobility/stupor,Mutism,Posturing/catalepsy,Echopraxia/echolalia,Mannerisms,Verbigeration,Waxy flexibility,Withdrawal | 95.5 | 93.5 | 97.3 |
| 8 | Immobility/stupor,Mutism,Posturing/catalepsy,Echopraxia/echolalia,Stereotypy,Mannerisms,Verbigeration,Withdrawal | 95.5 | 93.5 | 97.3 |
| 8 | Immobility/stupor,Mutism,Posturing/catalepsy,Echopraxia/echolalia,Stereotypy,Mannerisms,Waxy flexibility,Withdrawal | 95.5 | 93.5 | 97.3 |
| 8 | Immobility/stupor,Mutism,Posturing/catalepsy,Grimacing,Echopraxia/echolalia,Stereotypy,Mannerisms,Verbigeration | 95.5 | 93.5 | 97.3 |
| 8 | Immobility/stupor,Mutism,Posturing/catalepsy,Grimacing,Stereotypy,Verbigeration,Waxy flexibility,Withdrawal | 95.5 | 93.5 | 97.3 |
| 8 | Immobility/stupor,Posturing/catalepsy,Echopraxia/echolalia,Stereotypy,Verbigeration,Rigidity,Waxy flexibility,Withdrawal | 95.5 | 93.5 | 97.3 |
| 8 | Immobility/stupor,Posturing/catalepsy,Grimacing,Echopraxia/echolalia,Stereotypy,Verbigeration,Rigidity,Waxy flexibility | 95.5 | 93.5 | 97.3 |
| 8 | Mutism,Echopraxia/echolalia,Mannerisms,Verbigeration,Rigidity,Negativism,Waxy flexibility,Withdrawal | 95.5 | 93.5 | 97.3 |
| 8 | Mutism,Echopraxia/echolalia,Stereotypy,Verbigeration,Rigidity,Negativism,Waxy flexibility,Withdrawal | 95.5 | 93.5 | 97.3 |
| 8 | Mutism,Grimacing,Echopraxia/echolalia,Mannerisms,Verbigeration,Rigidity,Negativism,Withdrawal | 95.5 | 93.5 | 97.3 |
| 8 | Mutism,Posturing/catalepsy,Echopraxia/echolalia,Stereotypy,Verbigeration,Rigidity,Negativism,Withdrawal | 95.5 | 93.5 | 97.3 |
| 8 | Mutism,Posturing/catalepsy,Grimacing,Echopraxia/echolalia,Mannerisms,Verbigeration,Rigidity,Negativism | 95.5 | 93.5 | 97.3 |
| 8 | Excitement,Mutism,Grimacing,Stereotypy,Verbigeration,Rigidity,Negativism,Waxy flexibility | 95.3 | 93.3 | 97.1 |
| 8 | Excitement,Mutism,Grimacing,Stereotypy,Verbigeration,Rigidity,Waxy flexibility,Withdrawal | 95.3 | 93.3 | 97.1 |
| 8 | Excitement,Mutism,Posturing/catalepsy,Echopraxia/echolalia,Mannerisms,Rigidity,Negativism,Waxy flexibility | 95.3 | 93.3 | 97.1 |
| 8 | Excitement,Mutism,Posturing/catalepsy,Echopraxia/echolalia,Stereotypy,Mannerisms,Rigidity,Negativism | 95.3 | 93.3 | 97.1 |
| 8 | Excitement,Mutism,Posturing/catalepsy,Echopraxia/echolalia,Stereotypy,Rigidity,Negativism,Waxy flexibility | 95.3 | 93.3 | 97.1 |
| 8 | Excitement,Mutism,Posturing/catalepsy,Grimacing,Echopraxia/echolalia,Mannerisms,Rigidity,Withdrawal | 95.3 | 93.3 | 97.1 |
| 8 | Excitement,Mutism,Posturing/catalepsy,Grimacing,Stereotypy,Verbigeration,Rigidity,Withdrawal | 95.3 | 93.3 | 97.1 |
| 8 | Excitement,Mutism,Stereotypy,Mannerisms,Verbigeration,Rigidity,Negativism,Waxy flexibility | 95.3 | 93.3 | 97.1 |
| 8 | Immobility/stupor,Grimacing,Echopraxia/echolalia,Stereotypy,Mannerisms,Verbigeration,Waxy flexibility,Withdrawal | 95.3 | 93.3 | 97.1 |
| 8 | Immobility/stupor,Mutism,Grimacing,Stereotypy,Mannerisms,Verbigeration,Waxy flexibility,Withdrawal | 95.3 | 93.3 | 97.1 |
| 8 | Immobility/stupor,Mutism,Posturing/catalepsy,Grimacing,Echopraxia/echolalia,Stereotypy,Mannerisms,Waxy flexibility | 95.3 | 93.3 | 97.1 |
| 8 | Immobility/stupor,Mutism,Posturing/catalepsy,Grimacing,Echopraxia/echolalia,Stereotypy,Verbigeration,Waxy flexibility | 95.3 | 93.3 | 97.1 |
| 8 | Immobility/stupor,Posturing/catalepsy,Grimacing,Echopraxia/echolalia,Mannerisms,Verbigeration,Waxy flexibility,Withdrawal | 95.3 | 93.3 | 97.1 |
| 8 | Mutism,Grimacing,Echopraxia/echolalia,Stereotypy,Verbigeration,Rigidity,Negativism,Withdrawal | 95.3 | 93.3 | 97.1 |
| 8 | Mutism,Posturing/catalepsy,Echopraxia/echolalia,Mannerisms,Verbigeration,Rigidity,Negativism,Waxy flexibility | 95.3 | 93.3 | 97.1 |
| 8 | Mutism,Posturing/catalepsy,Echopraxia/echolalia,Stereotypy,Mannerisms,Verbigeration,Rigidity,Negativism | 95.3 | 93.3 | 97.1 |
| 8 | Mutism,Posturing/catalepsy,Echopraxia/echolalia,Verbigeration,Rigidity,Negativism,Waxy flexibility,Withdrawal | 95.3 | 93.3 | 97.1 |
| 8 | Mutism,Posturing/catalepsy,Grimacing,Echopraxia/echolalia,Mannerisms,Rigidity,Negativism,Waxy flexibility | 95.3 | 93.3 | 97.1 |
| 8 | Mutism,Posturing/catalepsy,Grimacing,Echopraxia/echolalia,Stereotypy,Rigidity,Negativism,Withdrawal | 95.3 | 93.3 | 97.1 |
| 8 | Mutism,Posturing/catalepsy,Grimacing,Echopraxia/echolalia,Verbigeration,Rigidity,Negativism,Withdrawal | 95.3 | 93.3 | 97.1 |
| 8 | Mutism,Posturing/catalepsy,Grimacing,Stereotypy,Mannerisms,Verbigeration,Rigidity,Negativism | 95.3 | 93.3 | 97.1 |
| 8 | Mutism,Posturing/catalepsy,Grimacing,Verbigeration,Rigidity,Negativism,Waxy flexibility,Withdrawal | 95.3 | 93.3 | 97.1 |
| 8 | Mutism,Posturing/catalepsy,Stereotypy,Mannerisms,Verbigeration,Rigidity,Negativism,Waxy flexibility | 95.3 | 93.3 | 97.1 |
| 8 | Excitement,Mutism,Echopraxia/echolalia,Stereotypy,Mannerisms,Rigidity,Waxy flexibility,Withdrawal | 95.1 | 93.0 | 96.9 |
| 8 | Excitement,Mutism,Grimacing,Echopraxia/echolalia,Stereotypy,Rigidity,Negativism,Waxy flexibility | 95.1 | 93.0 | 96.9 |
| 8 | Excitement,Mutism,Grimacing,Echopraxia/echolalia,Stereotypy,Rigidity,Waxy flexibility,Withdrawal | 95.1 | 93.0 | 97.1 |
| 8 | Excitement,Mutism,Grimacing,Echopraxia/echolalia,Verbigeration,Rigidity,Negativism,Waxy flexibility | 95.1 | 93.0 | 96.9 |
| 8 | Excitement,Mutism,Grimacing,Mannerisms,Verbigeration,Rigidity,Waxy flexibility,Withdrawal | 95.1 | 93.0 | 96.9 |
| 8 | Excitement,Mutism,Posturing/catalepsy,Echopraxia/echolalia,Stereotypy,Mannerisms,Rigidity,Withdrawal | 95.1 | 93.0 | 96.9 |
| 8 | Excitement,Mutism,Posturing/catalepsy,Echopraxia/echolalia,Stereotypy,Verbigeration,Negativism,Withdrawal | 95.1 | 93.0 | 97.1 |
| 8 | Excitement,Mutism,Posturing/catalepsy,Grimacing,Echopraxia/echolalia,Stereotypy,Rigidity,Withdrawal | 95.1 | 93.0 | 97.1 |
| 8 | Excitement,Mutism,Posturing/catalepsy,Grimacing,Echopraxia/echolalia,Verbigeration,Rigidity,Waxy flexibility | 95.1 | 93.0 | 97.1 |
| 8 | Excitement,Mutism,Posturing/catalepsy,Grimacing,Mannerisms,Rigidity,Negativism,Waxy flexibility | 95.1 | 93.0 | 96.9 |
| 8 | Excitement,Mutism,Posturing/catalepsy,Stereotypy,Mannerisms,Verbigeration,Negativism,Withdrawal | 95.1 | 93.0 | 97.1 |
| 8 | Excitement,Mutism,Posturing/catalepsy,Stereotypy,Verbigeration,Negativism,Waxy flexibility,Withdrawal | 95.1 | 93.0 | 97.1 |
| 8 | Excitement,Posturing/catalepsy,Echopraxia/echolalia,Mannerisms,Verbigeration,Rigidity,Negativism,Withdrawal | 95.1 | 93.0 | 96.9 |
| 8 | Excitement,Posturing/catalepsy,Stereotypy,Mannerisms,Verbigeration,Rigidity,Negativism,Withdrawal | 95.1 | 93.0 | 96.9 |
| 8 | Immobility/stupor,Mutism,Grimacing,Echopraxia/echolalia,Stereotypy,Mannerisms,Verbigeration,Waxy flexibility | 95.1 | 93.0 | 97.1 |
| 8 | Immobility/stupor,Mutism,Posturing/catalepsy,Grimacing,Stereotypy,Mannerisms,Verbigeration,Waxy flexibility | 95.1 | 93.0 | 97.1 |
| 8 | Immobility/stupor,Mutism,Posturing/catalepsy,Stereotypy,Mannerisms,Verbigeration,Waxy flexibility,Withdrawal | 95.1 | 92.8 | 97.1 |
| 8 | Immobility/stupor,Posturing/catalepsy,Echopraxia/echolalia,Stereotypy,Mannerisms,Verbigeration,Rigidity,Waxy flexibility | 95.1 | 93.0 | 97.1 |
| 8 | Mutism,Echopraxia/echolalia,Stereotypy,Mannerisms,Verbigeration,Rigidity,Negativism,Waxy flexibility | 95.1 | 93.0 | 96.9 |
| 8 | Mutism,Grimacing,Echopraxia/echolalia,Stereotypy,Rigidity,Negativism,Waxy flexibility,Withdrawal | 95.1 | 93.0 | 97.1 |
| 8 | Mutism,Grimacing,Echopraxia/echolalia,Verbigeration,Rigidity,Negativism,Waxy flexibility,Withdrawal | 95.1 | 93.0 | 97.1 |
| 8 | Mutism,Grimacing,Stereotypy,Mannerisms,Verbigeration,Rigidity,Negativism,Waxy flexibility | 95.1 | 93.0 | 96.9 |
| 8 | Mutism,Posturing/catalepsy,Echopraxia/echolalia,Stereotypy,Rigidity,Negativism,Waxy flexibility,Withdrawal | 95.1 | 92.8 | 96.9 |
| 8 | Mutism,Posturing/catalepsy,Grimacing,Echopraxia/echolalia,Rigidity,Negativism,Waxy flexibility,Withdrawal | 95.1 | 93.0 | 96.9 |
| 8 | Mutism,Posturing/catalepsy,Grimacing,Echopraxia/echolalia,Stereotypy,Mannerisms,Rigidity,Negativism | 95.1 | 92.8 | 96.9 |
| 8 | Mutism,Posturing/catalepsy,Grimacing,Mannerisms,Verbigeration,Rigidity,Negativism,Waxy flexibility | 95.1 | 93.0 | 97.1 |
| 8 | Mutism,Posturing/catalepsy,Grimacing,Stereotypy,Rigidity,Negativism,Waxy flexibility,Withdrawal | 95.1 | 93.0 | 96.9 |
| 8 | Posturing/catalepsy,Stereotypy,Mannerisms,Verbigeration,Rigidity,Negativism,Waxy flexibility,Withdrawal | 95.1 | 93.0 | 96.9 |
| 8 | Excitement,Mutism,Grimacing,Echopraxia/echolalia,Verbigeration,Rigidity,Waxy flexibility,Withdrawal | 95.0 | 92.8 | 96.9 |
| 8 | Immobility/stupor,Posturing/catalepsy,Grimacing,Stereotypy,Mannerisms,Verbigeration,Waxy flexibility,Withdrawal | 95.0 | 92.8 | 97.1 |
| 8 | Excitement,Mutism,Echopraxia/echolalia,Stereotypy,Mannerisms,Rigidity,Negativism,Waxy flexibility | 94.9 | 92.8 | 96.9 |
| 8 | Excitement,Mutism,Echopraxia/echolalia,Stereotypy,Mannerisms,Verbigeration,Rigidity,Negativism | 94.9 | 92.8 | 96.9 |
| 8 | Excitement,Mutism,Grimacing,Stereotypy,Mannerisms,Rigidity,Waxy flexibility,Withdrawal | 94.9 | 92.8 | 96.9 |
| 8 | Excitement,Mutism,Posturing/catalepsy,Grimacing,Stereotypy,Mannerisms,Rigidity,Withdrawal | 94.9 | 92.8 | 96.9 |
| 8 | Excitement,Mutism,Posturing/catalepsy,Mannerisms,Verbigeration,Negativism,Waxy flexibility,Withdrawal | 94.9 | 92.8 | 96.9 |
| 8 | Excitement,Posturing/catalepsy,Echopraxia/echolalia,Stereotypy,Verbigeration,Rigidity,Negativism,Withdrawal | 94.9 | 92.6 | 96.9 |
| 8 | Immobility/stupor,Mutism,Echopraxia/echolalia,Stereotypy,Mannerisms,Verbigeration,Waxy flexibility,Withdrawal | 94.9 | 92.8 | 96.9 |
| 8 | Immobility/stupor,Mutism,Posturing/catalepsy,Echopraxia/echolalia,Stereotypy,Verbigeration,Waxy flexibility,Withdrawal | 94.9 | 92.8 | 96.9 |
| 8 | Immobility/stupor,Posturing/catalepsy,Grimacing,Echopraxia/echolalia,Stereotypy,Verbigeration,Waxy flexibility,Withdrawal | 94.9 | 92.8 | 96.9 |
| 8 | Mutism,Posturing/catalepsy,Grimacing,Echopraxia/echolalia,Verbigeration,Rigidity,Negativism,Waxy flexibility | 94.9 | 92.6 | 96.9 |
| 8 | Excitement,Mutism,Grimacing,Echopraxia/echolalia,Mannerisms,Rigidity,Waxy flexibility,Withdrawal | 94.8 | 92.6 | 96.9 |
| 8 | Excitement,Mutism,Grimacing,Echopraxia/echolalia,Stereotypy,Verbigeration,Rigidity,Negativism | 94.8 | 92.6 | 96.9 |
| 8 | Excitement,Mutism,Posturing/catalepsy,Echopraxia/echolalia,Mannerisms,Verbigeration,Negativism,Withdrawal | 94.8 | 92.6 | 96.9 |
| 8 | Excitement,Mutism,Posturing/catalepsy,Echopraxia/echolalia,Mannerisms,Verbigeration,Rigidity,Waxy flexibility | 94.8 | 92.6 | 96.9 |
| 8 | Excitement,Mutism,Posturing/catalepsy,Echopraxia/echolalia,Stereotypy,Verbigeration,Rigidity,Waxy flexibility | 94.8 | 92.6 | 96.9 |
| 8 | Excitement,Mutism,Posturing/catalepsy,Echopraxia/echolalia,Verbigeration,Negativism,Waxy flexibility,Withdrawal | 94.8 | 92.8 | 96.9 |
| 8 | Excitement,Mutism,Posturing/catalepsy,Grimacing,Echopraxia/echolalia,Verbigeration,Negativism,Withdrawal | 94.8 | 92.6 | 96.9 |
| 8 | Excitement,Mutism,Posturing/catalepsy,Grimacing,Mannerisms,Verbigeration,Negativism,Withdrawal | 94.8 | 92.6 | 96.9 |
| 8 | Excitement,Mutism,Posturing/catalepsy,Grimacing,Mannerisms,Verbigeration,Rigidity,Waxy flexibility | 94.8 | 92.8 | 96.9 |
| 8 | Excitement,Mutism,Posturing/catalepsy,Grimacing,Stereotypy,Mannerisms,Rigidity,Negativism | 94.8 | 92.6 | 96.9 |
| 8 | Excitement,Mutism,Posturing/catalepsy,Grimacing,Stereotypy,Rigidity,Negativism,Waxy flexibility | 94.8 | 92.6 | 96.9 |
| 8 | Excitement,Mutism,Posturing/catalepsy,Grimacing,Stereotypy,Verbigeration,Negativism,Withdrawal | 94.8 | 92.8 | 96.9 |
| 8 | Excitement,Posturing/catalepsy,Echopraxia/echolalia,Verbigeration,Rigidity,Negativism,Waxy flexibility,Withdrawal | 94.8 | 92.6 | 96.9 |
| 8 | Excitement,Posturing/catalepsy,Grimacing,Echopraxia/echolalia,Verbigeration,Rigidity,Negativism,Withdrawal | 94.8 | 92.6 | 96.9 |
| 8 | Excitement,Posturing/catalepsy,Grimacing,Mannerisms,Verbigeration,Rigidity,Negativism,Withdrawal | 94.8 | 92.8 | 96.9 |
| 8 | Excitement,Posturing/catalepsy,Mannerisms,Verbigeration,Rigidity,Negativism,Waxy flexibility,Withdrawal | 94.8 | 92.6 | 96.9 |
| 8 | Excitement,Posturing/catalepsy,Stereotypy,Verbigeration,Rigidity,Negativism,Waxy flexibility,Withdrawal | 94.8 | 92.6 | 96.9 |
| 8 | Mutism,Grimacing,Echopraxia/echolalia,Stereotypy,Mannerisms,Rigidity,Negativism,Waxy flexibility | 94.8 | 92.6 | 96.9 |
| 8 | Mutism,Posturing/catalepsy,Echopraxia/echolalia,Stereotypy,Mannerisms,Rigidity,Negativism,Waxy flexibility | 94.8 | 92.6 | 96.9 |
| 8 | Mutism,Posturing/catalepsy,Grimacing,Echopraxia/echolalia,Mannerisms,Rigidity,Waxy flexibility,Withdrawal | 94.8 | 92.6 | 96.9 |
| 8 | Mutism,Posturing/catalepsy,Grimacing,Echopraxia/echolalia,Stereotypy,Verbigeration,Rigidity,Negativism | 94.8 | 92.6 | 96.9 |
| 8 | Mutism,Posturing/catalepsy,Grimacing,Mannerisms,Verbigeration,Rigidity,Waxy flexibility,Withdrawal | 94.8 | 92.6 | 96.9 |
| 8 | Mutism,Posturing/catalepsy,Grimacing,Stereotypy,Verbigeration,Rigidity,Negativism,Waxy flexibility | 94.8 | 92.6 | 96.6 |
| 8 | Posturing/catalepsy,Echopraxia/echolalia,Stereotypy,Mannerisms,Verbigeration,Rigidity,Negativism,Withdrawal | 94.8 | 92.8 | 96.9 |
| 8 | Posturing/catalepsy,Grimacing,Stereotypy,Mannerisms,Verbigeration,Rigidity,Negativism,Withdrawal | 94.8 | 92.6 | 96.9 |
| 8 | Immobility/stupor,Posturing/catalepsy,Echopraxia/echolalia,Stereotypy,Mannerisms,Verbigeration,Waxy flexibility,Withdrawal | 94.7 | 92.6 | 96.6 |
| 8 | Excitement,Mutism,Echopraxia/echolalia,Mannerisms,Verbigeration,Rigidity,Waxy flexibility,Withdrawal | 94.6 | 92.4 | 96.6 |
| 8 | Excitement,Mutism,Grimacing,Echopraxia/echolalia,Mannerisms,Rigidity,Negativism,Waxy flexibility | 94.6 | 92.4 | 96.6 |
| 8 | Excitement,Mutism,Grimacing,Mannerisms,Verbigeration,Rigidity,Negativism,Waxy flexibility | 94.6 | 92.4 | 96.6 |
| 8 | Excitement,Mutism,Grimacing,Stereotypy,Mannerisms,Verbigeration,Rigidity,Negativism | 94.6 | 92.4 | 96.6 |
| 8 | Excitement,Mutism,Posturing/catalepsy,Echopraxia/echolalia,Stereotypy,Mannerisms,Negativism,Withdrawal | 94.6 | 92.4 | 96.6 |
| 8 | Excitement,Mutism,Posturing/catalepsy,Echopraxia/echolalia,Stereotypy,Negativism,Waxy flexibility,Withdrawal | 94.6 | 92.4 | 96.6 |
| 8 | Excitement,Mutism,Posturing/catalepsy,Grimacing,Echopraxia/echolalia,Mannerisms,Negativism,Withdrawal | 94.6 | 92.4 | 96.6 |
| 8 | Excitement,Mutism,Posturing/catalepsy,Grimacing,Echopraxia/echolalia,Mannerisms,Rigidity,Waxy flexibility | 94.6 | 92.4 | 96.6 |
| 8 | Excitement,Mutism,Posturing/catalepsy,Grimacing,Echopraxia/echolalia,Negativism,Waxy flexibility,Withdrawal | 94.6 | 92.4 | 96.6 |
| 8 | Excitement,Mutism,Posturing/catalepsy,Grimacing,Echopraxia/echolalia,Stereotypy,Negativism,Withdrawal | 94.6 | 92.6 | 96.6 |
| 8 | Excitement,Mutism,Posturing/catalepsy,Grimacing,Stereotypy,Verbigeration,Rigidity,Waxy flexibility | 94.6 | 92.4 | 96.6 |
| 8 | Excitement,Mutism,Posturing/catalepsy,Grimacing,Verbigeration,Negativism,Waxy flexibility,Withdrawal | 94.6 | 92.4 | 96.6 |
| 8 | Excitement,Mutism,Posturing/catalepsy,Stereotypy,Mannerisms,Rigidity,Negativism,Waxy flexibility | 94.6 | 92.6 | 96.6 |
| 8 | Excitement,Posturing/catalepsy,Echopraxia/echolalia,Stereotypy,Mannerisms,Rigidity,Negativism,Withdrawal | 94.6 | 92.4 | 96.6 |
| 8 | Excitement,Posturing/catalepsy,Grimacing,Echopraxia/echolalia,Mannerisms,Rigidity,Negativism,Withdrawal | 94.6 | 92.4 | 96.6 |
| 8 | Excitement,Posturing/catalepsy,Grimacing,Stereotypy,Verbigeration,Rigidity,Negativism,Withdrawal | 94.6 | 92.4 | 96.6 |
| 8 | Immobility/stupor,Mutism,Posturing/catalepsy,Echopraxia/echolalia,Stereotypy,Mannerisms,Verbigeration,Waxy flexibility | 94.6 | 92.4 | 96.6 |
| 8 | Mutism,Grimacing,Echopraxia/echolalia,Mannerisms,Verbigeration,Rigidity,Negativism,Waxy flexibility | 94.6 | 92.4 | 96.6 |
| 8 | Mutism,Posturing/catalepsy,Echopraxia/echolalia,Stereotypy,Verbigeration,Rigidity,Negativism,Waxy flexibility | 94.6 | 92.4 | 96.6 |
| 8 | Mutism,Posturing/catalepsy,Grimacing,Stereotypy,Mannerisms,Rigidity,Negativism,Waxy flexibility | 94.6 | 92.4 | 96.6 |
| 8 | Mutism,Posturing/catalepsy,Grimacing,Stereotypy,Mannerisms,Rigidity,Waxy flexibility,Withdrawal | 94.6 | 92.4 | 96.6 |
| 8 | Posturing/catalepsy,Echopraxia/echolalia,Mannerisms,Verbigeration,Rigidity,Negativism,Waxy flexibility,Withdrawal | 94.6 | 92.6 | 96.6 |
| 8 | Posturing/catalepsy,Echopraxia/echolalia,Stereotypy,Mannerisms,Rigidity,Negativism,Waxy flexibility,Withdrawal | 94.6 | 92.4 | 96.6 |
| 8 | Posturing/catalepsy,Grimacing,Echopraxia/echolalia,Mannerisms,Verbigeration,Rigidity,Negativism,Withdrawal | 94.6 | 92.4 | 96.6 |
| 8 | Posturing/catalepsy,Grimacing,Echopraxia/echolalia,Stereotypy,Mannerisms,Rigidity,Negativism,Withdrawal | 94.6 | 92.4 | 96.6 |
| 8 | Posturing/catalepsy,Grimacing,Stereotypy,Mannerisms,Rigidity,Negativism,Waxy flexibility,Withdrawal | 94.6 | 92.4 | 96.6 |
| 8 | Excitement,Mutism,Echopraxia/echolalia,Mannerisms,Verbigeration,Rigidity,Negativism,Waxy flexibility | 94.4 | 92.2 | 96.4 |
| 8 | Excitement,Mutism,Grimacing,Echopraxia/echolalia,Mannerisms,Verbigeration,Rigidity,Negativism | 94.4 | 92.2 | 96.4 |
| 8 | Excitement,Mutism,Grimacing,Echopraxia/echolalia,Stereotypy,Mannerisms,Rigidity,Negativism | 94.4 | 92.2 | 96.4 |
| 8 | Excitement,Mutism,Grimacing,Stereotypy,Mannerisms,Rigidity,Negativism,Waxy flexibility | 94.4 | 92.2 | 96.4 |
| 8 | Excitement,Mutism,Posturing/catalepsy,Echopraxia/echolalia,Mannerisms,Negativism,Waxy flexibility,Withdrawal | 94.4 | 92.2 | 96.4 |
| 8 | Excitement,Mutism,Posturing/catalepsy,Grimacing,Echopraxia/echolalia,Mannerisms,Verbigeration,Rigidity | 94.4 | 92.2 | 96.4 |
| 8 | Excitement,Mutism,Posturing/catalepsy,Grimacing,Echopraxia/echolalia,Stereotypy,Rigidity,Waxy flexibility | 94.4 | 92.2 | 96.4 |
| 8 | Excitement,Mutism,Posturing/catalepsy,Grimacing,Mannerisms,Negativism,Waxy flexibility,Withdrawal | 94.4 | 92.2 | 96.4 |
| 8 | Excitement,Mutism,Posturing/catalepsy,Grimacing,Stereotypy,Mannerisms,Negativism,Withdrawal | 94.4 | 92.2 | 96.4 |
| 8 | Excitement,Mutism,Posturing/catalepsy,Grimacing,Stereotypy,Negativism,Waxy flexibility,Withdrawal | 94.4 | 92.2 | 96.4 |
| 8 | Excitement,Mutism,Posturing/catalepsy,Stereotypy,Mannerisms,Negativism,Waxy flexibility,Withdrawal | 94.4 | 92.2 | 96.4 |
| 8 | Excitement,Mutism,Posturing/catalepsy,Stereotypy,Mannerisms,Verbigeration,Rigidity,Waxy flexibility | 94.4 | 92.2 | 96.4 |
| 8 | Excitement,Posturing/catalepsy,Echopraxia/echolalia,Mannerisms,Rigidity,Negativism,Waxy flexibility,Withdrawal | 94.4 | 92.2 | 96.4 |
| 8 | Excitement,Posturing/catalepsy,Echopraxia/echolalia,Stereotypy,Rigidity,Negativism,Waxy flexibility,Withdrawal | 94.4 | 92.2 | 96.4 |
| 8 | Excitement,Posturing/catalepsy,Grimacing,Echopraxia/echolalia,Rigidity,Negativism,Waxy flexibility,Withdrawal | 94.4 | 92.2 | 96.4 |
| 8 | Excitement,Posturing/catalepsy,Grimacing,Echopraxia/echolalia,Stereotypy,Rigidity,Negativism,Withdrawal | 94.4 | 92.2 | 96.4 |
| 8 | Excitement,Posturing/catalepsy,Grimacing,Stereotypy,Mannerisms,Rigidity,Negativism,Withdrawal | 94.4 | 92.2 | 96.4 |
| 8 | Excitement,Posturing/catalepsy,Grimacing,Verbigeration,Rigidity,Negativism,Waxy flexibility,Withdrawal | 94.4 | 92.2 | 96.4 |
| 8 | Excitement,Posturing/catalepsy,Stereotypy,Mannerisms,Rigidity,Negativism,Waxy flexibility,Withdrawal | 94.4 | 92.2 | 96.4 |
| 8 | Immobility/stupor,Posturing/catalepsy,Grimacing,Echopraxia/echolalia,Stereotypy,Mannerisms,Verbigeration,Waxy flexibility | 94.4 | 92.2 | 96.4 |
| 8 | Mutism,Grimacing,Echopraxia/echolalia,Stereotypy,Mannerisms,Rigidity,Waxy flexibility,Withdrawal | 94.4 | 92.2 | 96.4 |
| 8 | Mutism,Grimacing,Echopraxia/echolalia,Stereotypy,Mannerisms,Verbigeration,Rigidity,Negativism | 94.4 | 92.2 | 96.4 |
| 8 | Mutism,Grimacing,Echopraxia/echolalia,Stereotypy,Verbigeration,Rigidity,Negativism,Waxy flexibility | 94.4 | 92.2 | 96.4 |
| 8 | Mutism,Grimacing,Stereotypy,Mannerisms,Verbigeration,Rigidity,Waxy flexibility,Withdrawal | 94.4 | 92.2 | 96.4 |
| 8 | Mutism,Posturing/catalepsy,Echopraxia/echolalia,Mannerisms,Verbigeration,Rigidity,Waxy flexibility,Withdrawal | 94.4 | 92.2 | 96.4 |
| 8 | Mutism,Posturing/catalepsy,Echopraxia/echolalia,Stereotypy,Mannerisms,Verbigeration,Negativism,Withdrawal | 94.4 | 92.2 | 96.4 |
| 8 | Mutism,Posturing/catalepsy,Grimacing,Echopraxia/echolalia,Mannerisms,Negativism,Waxy flexibility,Withdrawal | 94.4 | 92.2 | 96.4 |
| 8 | Mutism,Posturing/catalepsy,Grimacing,Echopraxia/echolalia,Mannerisms,Verbigeration,Negativism,Withdrawal | 94.4 | 92.2 | 96.4 |
| 8 | Mutism,Posturing/catalepsy,Grimacing,Echopraxia/echolalia,Mannerisms,Verbigeration,Rigidity,Withdrawal | 94.4 | 92.2 | 96.4 |
| 8 | Mutism,Posturing/catalepsy,Grimacing,Echopraxia/echolalia,Stereotypy,Mannerisms,Negativism,Withdrawal | 94.4 | 92.2 | 96.4 |
| 8 | Mutism,Posturing/catalepsy,Grimacing,Stereotypy,Mannerisms,Verbigeration,Negativism,Withdrawal | 94.4 | 92.2 | 96.4 |
| 8 | Mutism,Posturing/catalepsy,Grimacing,Stereotypy,Verbigeration,Rigidity,Waxy flexibility,Withdrawal | 94.4 | 92.2 | 96.4 |
| 8 | Mutism,Posturing/catalepsy,Stereotypy,Mannerisms,Verbigeration,Negativism,Waxy flexibility,Withdrawal | 94.4 | 92.2 | 96.4 |
| 8 | Mutism,Posturing/catalepsy,Stereotypy,Mannerisms,Verbigeration,Rigidity,Waxy flexibility,Withdrawal | 94.4 | 92.2 | 96.4 |
| 8 | Posturing/catalepsy,Grimacing,Echopraxia/echolalia,Mannerisms,Rigidity,Negativism,Waxy flexibility,Withdrawal | 94.4 | 92.2 | 96.4 |
| 8 | Posturing/catalepsy,Grimacing,Mannerisms,Verbigeration,Rigidity,Negativism,Waxy flexibility,Withdrawal | 94.4 | 92.2 | 96.4 |
| 8 | Excitement,Mutism,Posturing/catalepsy,Echopraxia/echolalia,Stereotypy,Mannerisms,Rigidity,Waxy flexibility | 94.2 | 91.9 | 96.2 |
| 8 | Excitement,Mutism,Posturing/catalepsy,Echopraxia/echolalia,Stereotypy,Mannerisms,Verbigeration,Rigidity | 94.2 | 91.9 | 96.2 |
| 8 | Excitement,Mutism,Posturing/catalepsy,Grimacing,Echopraxia/echolalia,Stereotypy,Verbigeration,Rigidity | 94.2 | 91.9 | 96.2 |
| 8 | Excitement,Posturing/catalepsy,Echopraxia/echolalia,Mannerisms,Verbigeration,Rigidity,Waxy flexibility,Withdrawal | 94.2 | 91.9 | 96.2 |
| 8 | Excitement,Posturing/catalepsy,Grimacing,Mannerisms,Rigidity,Negativism,Waxy flexibility,Withdrawal | 94.2 | 91.9 | 96.2 |
| 8 | Excitement,Posturing/catalepsy,Grimacing,Stereotypy,Rigidity,Negativism,Waxy flexibility,Withdrawal | 94.2 | 91.9 | 96.2 |
| 8 | Mutism,Grimacing,Echopraxia/echolalia,Mannerisms,Verbigeration,Rigidity,Waxy flexibility,Withdrawal | 94.2 | 91.9 | 96.2 |
| 8 | Mutism,Posturing/catalepsy,Echopraxia/echolalia,Mannerisms,Verbigeration,Negativism,Waxy flexibility,Withdrawal | 94.2 | 91.9 | 96.2 |
| 8 | Mutism,Posturing/catalepsy,Echopraxia/echolalia,Stereotypy,Mannerisms,Negativism,Waxy flexibility,Withdrawal | 94.2 | 91.9 | 96.2 |
| 8 | Mutism,Posturing/catalepsy,Echopraxia/echolalia,Stereotypy,Mannerisms,Rigidity,Waxy flexibility,Withdrawal | 94.2 | 91.9 | 96.2 |
| 8 | Mutism,Posturing/catalepsy,Grimacing,Echopraxia/echolalia,Stereotypy,Mannerisms,Rigidity,Withdrawal | 94.2 | 91.9 | 96.2 |
| 8 | Mutism,Posturing/catalepsy,Grimacing,Echopraxia/echolalia,Stereotypy,Rigidity,Negativism,Waxy flexibility | 94.2 | 91.9 | 96.2 |
| 8 | Mutism,Posturing/catalepsy,Grimacing,Echopraxia/echolalia,Stereotypy,Rigidity,Waxy flexibility,Withdrawal | 94.2 | 91.9 | 96.2 |
| 8 | Mutism,Posturing/catalepsy,Grimacing,Echopraxia/echolalia,Verbigeration,Rigidity,Waxy flexibility,Withdrawal | 94.2 | 91.9 | 96.2 |
| 8 | Mutism,Posturing/catalepsy,Grimacing,Mannerisms,Verbigeration,Negativism,Waxy flexibility,Withdrawal | 94.2 | 91.9 | 96.2 |
| 8 | Mutism,Posturing/catalepsy,Grimacing,Stereotypy,Mannerisms,Negativism,Waxy flexibility,Withdrawal | 94.2 | 91.9 | 96.2 |
| 8 | Mutism,Posturing/catalepsy,Grimacing,Stereotypy,Mannerisms,Verbigeration,Rigidity,Withdrawal | 94.2 | 91.9 | 96.2 |
| 8 | Posturing/catalepsy,Grimacing,Stereotypy,Verbigeration,Rigidity,Negativism,Waxy flexibility,Withdrawal | 94.2 | 91.9 | 96.2 |
| 8 | Excitement,Mutism,Echopraxia/echolalia,Stereotypy,Mannerisms,Verbigeration,Rigidity,Withdrawal | 94.0 | 91.7 | 96.0 |
| 8 | Excitement,Mutism,Posturing/catalepsy,Grimacing,Stereotypy,Mannerisms,Verbigeration,Rigidity | 94.0 | 91.7 | 96.0 |
| 8 | Excitement,Posturing/catalepsy,Grimacing,Echopraxia/echolalia,Verbigeration,Rigidity,Waxy flexibility,Withdrawal | 94.0 | 91.7 | 96.2 |
| 8 | Mutism,Grimacing,Echopraxia/echolalia,Stereotypy,Verbigeration,Rigidity,Waxy flexibility,Withdrawal | 93.9 | 91.7 | 96.0 |
| 8 | Posturing/catalepsy,Echopraxia/echolalia,Stereotypy,Verbigeration,Rigidity,Negativism,Waxy flexibility,Withdrawal | 93.9 | 91.7 | 96.0 |
| 8 | Posturing/catalepsy,Grimacing,Echopraxia/echolalia,Stereotypy,Verbigeration,Rigidity,Negativism,Withdrawal | 93.9 | 91.7 | 96.0 |
| 8 | Excitement,Mutism,Echopraxia/echolalia,Stereotypy,Verbigeration,Negativism,Waxy flexibility,Withdrawal | 93.7 | 91.5 | 96.0 |
| 8 | Excitement,Mutism,Grimacing,Echopraxia/echolalia,Stereotypy,Verbigeration,Rigidity,Waxy flexibility | 93.7 | 91.5 | 96.0 |
| 8 | Excitement,Mutism,Grimacing,Echopraxia/echolalia,Stereotypy,Verbigeration,Rigidity,Withdrawal | 93.7 | 91.5 | 96.0 |
| 8 | Excitement,Mutism,Grimacing,Stereotypy,Mannerisms,Verbigeration,Rigidity,Withdrawal | 93.7 | 91.5 | 96.0 |
| 8 | Excitement,Mutism,Posturing/catalepsy,Echopraxia/echolalia,Stereotypy,Verbigeration,Waxy flexibility,Withdrawal | 93.7 | 91.5 | 96.0 |
| 8 | Excitement,Mutism,Posturing/catalepsy,Grimacing,Echopraxia/echolalia,Stereotypy,Mannerisms,Rigidity | 93.7 | 91.5 | 96.0 |
| 8 | Excitement,Mutism,Posturing/catalepsy,Grimacing,Echopraxia/echolalia,Verbigeration,Negativism,Waxy flexibility | 93.7 | 91.5 | 95.7 |
| 8 | Excitement,Mutism,Posturing/catalepsy,Grimacing,Echopraxia/echolalia,Verbigeration,Waxy flexibility,Withdrawal | 93.7 | 91.5 | 96.0 |
| 8 | Excitement,Mutism,Posturing/catalepsy,Grimacing,Stereotypy,Mannerisms,Rigidity,Waxy flexibility | 93.7 | 91.5 | 96.0 |
| 8 | Excitement,Posturing/catalepsy,Echopraxia/echolalia,Stereotypy,Verbigeration,Rigidity,Waxy flexibility,Withdrawal | 93.7 | 91.5 | 95.7 |
| 8 | Excitement,Posturing/catalepsy,Grimacing,Echopraxia/echolalia,Mannerisms,Rigidity,Waxy flexibility,Withdrawal | 93.7 | 91.5 | 96.0 |
| 8 | Excitement,Posturing/catalepsy,Grimacing,Mannerisms,Verbigeration,Rigidity,Waxy flexibility,Withdrawal | 93.7 | 91.5 | 96.0 |
| 8 | Excitement,Posturing/catalepsy,Stereotypy,Mannerisms,Verbigeration,Rigidity,Waxy flexibility,Withdrawal | 93.7 | 91.5 | 96.0 |
| 8 | Mutism,Echopraxia/echolalia,Stereotypy,Mannerisms,Verbigeration,Rigidity,Waxy flexibility,Withdrawal | 93.7 | 91.5 | 96.0 |
| 8 | Mutism,Posturing/catalepsy,Echopraxia/echolalia,Stereotypy,Mannerisms,Verbigeration,Rigidity,Withdrawal | 93.7 | 91.5 | 96.0 |
| 8 | Mutism,Posturing/catalepsy,Echopraxia/echolalia,Stereotypy,Verbigeration,Rigidity,Waxy flexibility,Withdrawal | 93.7 | 91.5 | 96.0 |
| 8 | Mutism,Posturing/catalepsy,Grimacing,Echopraxia/echolalia,Stereotypy,Verbigeration,Negativism,Withdrawal | 93.7 | 91.3 | 95.7 |
| 8 | Mutism,Posturing/catalepsy,Grimacing,Echopraxia/echolalia,Stereotypy,Verbigeration,Rigidity,Withdrawal | 93.7 | 91.3 | 96.0 |
| 8 | Mutism,Posturing/catalepsy,Grimacing,Stereotypy,Verbigeration,Negativism,Waxy flexibility,Withdrawal | 93.7 | 91.3 | 96.0 |
| 8 | Posturing/catalepsy,Grimacing,Echopraxia/echolalia,Stereotypy,Rigidity,Negativism,Waxy flexibility,Withdrawal | 93.7 | 91.5 | 96.0 |
| 8 | Posturing/catalepsy,Grimacing,Echopraxia/echolalia,Verbigeration,Rigidity,Negativism,Waxy flexibility,Withdrawal | 93.7 | 91.3 | 96.0 |
| 8 | Excitement,Mutism,Echopraxia/echolalia,Stereotypy,Mannerisms,Verbigeration,Rigidity,Waxy flexibility | 93.5 | 91.3 | 95.7 |
| 8 | Excitement,Mutism,Grimacing,Echopraxia/echolalia,Mannerisms,Verbigeration,Rigidity,Withdrawal | 93.5 | 91.0 | 95.7 |
| 8 | Excitement,Mutism,Grimacing,Echopraxia/echolalia,Stereotypy,Mannerisms,Rigidity,Withdrawal | 93.5 | 91.0 | 95.7 |
| 8 | Excitement,Mutism,Posturing/catalepsy,Echopraxia/echolalia,Mannerisms,Verbigeration,Waxy flexibility,Withdrawal | 93.5 | 91.0 | 95.7 |
| 8 | Excitement,Mutism,Posturing/catalepsy,Echopraxia/echolalia,Stereotypy,Verbigeration,Negativism,Waxy flexibility | 93.5 | 91.3 | 95.7 |
| 8 | Excitement,Mutism,Posturing/catalepsy,Grimacing,Echopraxia/echolalia,Mannerisms,Verbigeration,Negativism | 93.5 | 91.0 | 95.5 |
| 8 | Excitement,Mutism,Posturing/catalepsy,Grimacing,Echopraxia/echolalia,Mannerisms,Waxy flexibility,Withdrawal | 93.5 | 91.3 | 95.7 |
| 8 | Excitement,Mutism,Posturing/catalepsy,Grimacing,Echopraxia/echolalia,Stereotypy,Verbigeration,Negativism | 93.5 | 91.3 | 95.7 |
| 8 | Excitement,Mutism,Posturing/catalepsy,Grimacing,Echopraxia/echolalia,Stereotypy,Waxy flexibility,Withdrawal | 93.5 | 91.0 | 95.7 |
| 8 | Excitement,Mutism,Posturing/catalepsy,Grimacing,Mannerisms,Verbigeration,Negativism,Waxy flexibility | 93.5 | 91.0 | 95.7 |
| 8 | Excitement,Mutism,Posturing/catalepsy,Grimacing,Mannerisms,Verbigeration,Waxy flexibility,Withdrawal | 93.5 | 91.0 | 95.7 |
| 8 | Excitement,Mutism,Posturing/catalepsy,Grimacing,Stereotypy,Verbigeration,Negativism,Waxy flexibility | 93.5 | 91.0 | 95.7 |
| 8 | Excitement,Mutism,Posturing/catalepsy,Grimacing,Stereotypy,Verbigeration,Waxy flexibility,Withdrawal | 93.5 | 91.0 | 95.7 |
| 8 | Excitement,Mutism,Posturing/catalepsy,Stereotypy,Mannerisms,Verbigeration,Waxy flexibility,Withdrawal | 93.5 | 91.0 | 95.7 |
| 8 | Excitement,Posturing/catalepsy,Echopraxia/echolalia,Stereotypy,Mannerisms,Rigidity,Waxy flexibility,Withdrawal | 93.5 | 91.0 | 95.7 |
| 8 | Excitement,Posturing/catalepsy,Grimacing,Echopraxia/echolalia,Mannerisms,Verbigeration,Rigidity,Withdrawal | 93.5 | 91.0 | 95.7 |
| 8 | Mutism,Posturing/catalepsy,Echopraxia/echolalia,Stereotypy,Verbigeration,Negativism,Waxy flexibility,Withdrawal | 93.5 | 91.0 | 95.7 |
| 8 | Mutism,Posturing/catalepsy,Grimacing,Echopraxia/echolalia,Mannerisms,Verbigeration,Rigidity,Waxy flexibility | 93.5 | 91.3 | 95.7 |
| 8 | Mutism,Posturing/catalepsy,Grimacing,Echopraxia/echolalia,Stereotypy,Negativism,Waxy flexibility,Withdrawal | 93.5 | 91.0 | 95.7 |
| 8 | Mutism,Posturing/catalepsy,Grimacing,Echopraxia/echolalia,Verbigeration,Negativism,Waxy flexibility,Withdrawal | 93.5 | 91.3 | 95.7 |
| 8 | Echopraxia/echolalia,Stereotypy,Mannerisms,Verbigeration,Rigidity,Negativism,Waxy flexibility,Withdrawal | 93.3 | 90.8 | 95.5 |
| 8 | Excitement,Mutism,Echopraxia/echolalia,Stereotypy,Mannerisms,Negativism,Waxy flexibility,Withdrawal | 93.3 | 90.8 | 95.5 |
| 8 | Excitement,Mutism,Echopraxia/echolalia,Stereotypy,Mannerisms,Verbigeration,Negativism,Withdrawal | 93.3 | 90.8 | 95.5 |
| 8 | Excitement,Mutism,Grimacing,Echopraxia/echolalia,Mannerisms,Verbigeration,Rigidity,Waxy flexibility | 93.3 | 90.8 | 95.5 |
| 8 | Excitement,Mutism,Grimacing,Echopraxia/echolalia,Stereotypy,Mannerisms,Rigidity,Waxy flexibility | 93.3 | 90.8 | 95.5 |
| 8 | Excitement,Mutism,Grimacing,Echopraxia/echolalia,Stereotypy,Negativism,Waxy flexibility,Withdrawal | 93.3 | 90.8 | 95.5 |
| 8 | Excitement,Mutism,Grimacing,Stereotypy,Mannerisms,Verbigeration,Rigidity,Waxy flexibility | 93.3 | 90.8 | 95.5 |
| 8 | Excitement,Mutism,Posturing/catalepsy,Echopraxia/echolalia,Mannerisms,Verbigeration,Negativism,Waxy flexibility | 93.3 | 90.8 | 95.5 |
| 8 | Excitement,Mutism,Posturing/catalepsy,Echopraxia/echolalia,Stereotypy,Mannerisms,Verbigeration,Negativism | 93.3 | 90.8 | 95.5 |
| 8 | Excitement,Mutism,Posturing/catalepsy,Echopraxia/echolalia,Stereotypy,Mannerisms,Waxy flexibility,Withdrawal | 93.3 | 91.0 | 95.5 |
| 8 | Excitement,Mutism,Posturing/catalepsy,Grimacing,Echopraxia/echolalia,Mannerisms,Negativism,Waxy flexibility | 93.3 | 90.8 | 95.5 |
| 8 | Excitement,Mutism,Posturing/catalepsy,Grimacing,Echopraxia/echolalia,Stereotypy,Negativism,Waxy flexibility | 93.3 | 90.8 | 95.5 |
| 8 | Excitement,Mutism,Posturing/catalepsy,Grimacing,Stereotypy,Mannerisms,Verbigeration,Negativism | 93.3 | 90.8 | 95.5 |
| 8 | Excitement,Mutism,Posturing/catalepsy,Stereotypy,Mannerisms,Verbigeration,Negativism,Waxy flexibility | 93.3 | 90.8 | 95.5 |
| 8 | Excitement,Mutism,Stereotypy,Mannerisms,Verbigeration,Negativism,Waxy flexibility,Withdrawal | 93.3 | 90.8 | 95.5 |
| 8 | Excitement,Posturing/catalepsy,Echopraxia/echolalia,Stereotypy,Mannerisms,Verbigeration,Rigidity,Withdrawal | 93.3 | 91.0 | 95.5 |
| 8 | Excitement,Posturing/catalepsy,Grimacing,Echopraxia/echolalia,Stereotypy,Rigidity,Waxy flexibility,Withdrawal | 93.3 | 90.8 | 95.5 |
| 8 | Excitement,Posturing/catalepsy,Grimacing,Stereotypy,Verbigeration,Rigidity,Waxy flexibility,Withdrawal | 93.3 | 90.8 | 95.5 |
| 8 | Grimacing,Echopraxia/echolalia,Stereotypy,Mannerisms,Rigidity,Negativism,Waxy flexibility,Withdrawal | 93.3 | 90.8 | 95.5 |
| 8 | Mutism,Echopraxia/echolalia,Stereotypy,Mannerisms,Verbigeration,Negativism,Waxy flexibility,Withdrawal | 93.3 | 90.8 | 95.5 |
| 8 | Mutism,Grimacing,Echopraxia/echolalia,Stereotypy,Mannerisms,Negativism,Waxy flexibility,Withdrawal | 93.3 | 90.8 | 95.5 |
| 8 | Mutism,Posturing/catalepsy,Grimacing,Echopraxia/echolalia,Mannerisms,Verbigeration,Negativism,Waxy flexibility | 93.3 | 90.8 | 95.5 |
| 8 | Mutism,Posturing/catalepsy,Grimacing,Stereotypy,Mannerisms,Verbigeration,Rigidity,Waxy flexibility | 93.3 | 90.8 | 95.5 |
| 8 | Excitement,Mutism,Grimacing,Echopraxia/echolalia,Stereotypy,Verbigeration,Negativism,Withdrawal | 93.1 | 90.6 | 95.3 |
| 8 | Excitement,Mutism,Posturing/catalepsy,Grimacing,Echopraxia/echolalia,Stereotypy,Mannerisms,Negativism | 93.1 | 90.6 | 95.3 |
| 8 | Excitement,Mutism,Posturing/catalepsy,Grimacing,Stereotypy,Mannerisms,Waxy flexibility,Withdrawal | 93.1 | 90.6 | 95.3 |
| 8 | Grimacing,Stereotypy,Mannerisms,Verbigeration,Rigidity,Negativism,Waxy flexibility,Withdrawal | 93.1 | 90.6 | 95.3 |
| 8 | Mutism,Posturing/catalepsy,Grimacing,Echopraxia/echolalia,Stereotypy,Mannerisms,Negativism,Waxy flexibility | 93.1 | 90.6 | 95.3 |
| 8 | Mutism,Posturing/catalepsy,Grimacing,Echopraxia/echolalia,Stereotypy,Mannerisms,Rigidity,Waxy flexibility | 93.1 | 90.6 | 95.3 |
| 8 | Mutism,Posturing/catalepsy,Grimacing,Echopraxia/echolalia,Stereotypy,Mannerisms,Verbigeration,Negativism | 93.1 | 90.6 | 95.3 |
| 8 | Excitement,Mutism,Grimacing,Stereotypy,Verbigeration,Negativism,Waxy flexibility,Withdrawal | 93.0 | 90.6 | 95.3 |
| 8 | Excitement,Posturing/catalepsy,Grimacing,Stereotypy,Mannerisms,Rigidity,Waxy flexibility,Withdrawal | 93.0 | 90.6 | 95.3 |
| 8 | Excitement,Posturing/catalepsy,Stereotypy,Mannerisms,Verbigeration,Negativism,Waxy flexibility,Withdrawal | 93.0 | 90.6 | 95.3 |
| 8 | Mutism,Posturing/catalepsy,Grimacing,Echopraxia/echolalia,Stereotypy,Verbigeration,Rigidity,Waxy flexibility | 93.0 | 90.6 | 95.3 |
| 8 | Excitement,Echopraxia/echolalia,Stereotypy,Mannerisms,Rigidity,Negativism,Waxy flexibility,Withdrawal | 92.8 | 90.4 | 95.1 |
| 8 | Excitement,Echopraxia/echolalia,Stereotypy,Mannerisms,Verbigeration,Rigidity,Negativism,Withdrawal | 92.8 | 90.4 | 95.1 |
| 8 | Excitement,Echopraxia/echolalia,Stereotypy,Verbigeration,Rigidity,Negativism,Waxy flexibility,Withdrawal | 92.8 | 90.4 | 95.1 |
| 8 | Excitement,Mutism,Posturing/catalepsy,Echopraxia/echolalia,Stereotypy,Mannerisms,Negativism,Waxy flexibility | 92.8 | 90.4 | 95.1 |
| 8 | Excitement,Mutism,Posturing/catalepsy,Echopraxia/echolalia,Stereotypy,Mannerisms,Verbigeration,Withdrawal | 92.8 | 90.4 | 95.1 |
| 8 | Excitement,Mutism,Posturing/catalepsy,Grimacing,Echopraxia/echolalia,Mannerisms,Verbigeration,Withdrawal | 92.8 | 90.4 | 95.1 |
| 8 | Excitement,Mutism,Posturing/catalepsy,Grimacing,Echopraxia/echolalia,Stereotypy,Verbigeration,Withdrawal | 92.8 | 90.4 | 95.1 |
| 8 | Excitement,Posturing/catalepsy,Echopraxia/echolalia,Stereotypy,Mannerisms,Verbigeration,Negativism,Withdrawal | 92.8 | 90.4 | 95.1 |
| 8 | Excitement,Posturing/catalepsy,Echopraxia/echolalia,Stereotypy,Verbigeration,Negativism,Waxy flexibility,Withdrawal | 92.8 | 90.4 | 95.1 |
| 8 | Excitement,Posturing/catalepsy,Grimacing,Echopraxia/echolalia,Mannerisms,Verbigeration,Negativism,Withdrawal | 92.8 | 90.4 | 95.1 |
| 8 | Excitement,Posturing/catalepsy,Grimacing,Echopraxia/echolalia,Stereotypy,Mannerisms,Rigidity,Withdrawal | 92.8 | 90.4 | 95.1 |
| 8 | Excitement,Posturing/catalepsy,Grimacing,Echopraxia/echolalia,Stereotypy,Verbigeration,Rigidity,Withdrawal | 92.8 | 90.4 | 95.1 |
| 8 | Excitement,Posturing/catalepsy,Grimacing,Stereotypy,Mannerisms,Verbigeration,Negativism,Withdrawal | 92.8 | 90.4 | 95.1 |
| 8 | Excitement,Posturing/catalepsy,Grimacing,Stereotypy,Mannerisms,Verbigeration,Rigidity,Withdrawal | 92.8 | 90.4 | 95.3 |
| 8 | Excitement,Stereotypy,Mannerisms,Verbigeration,Rigidity,Negativism,Waxy flexibility,Withdrawal | 92.8 | 90.4 | 95.1 |
| 8 | Grimacing,Echopraxia/echolalia,Stereotypy,Mannerisms,Verbigeration,Rigidity,Negativism,Withdrawal | 92.8 | 90.4 | 95.1 |
| 8 | Mutism,Grimacing,Echopraxia/echolalia,Stereotypy,Mannerisms,Verbigeration,Negativism,Withdrawal | 92.8 | 90.4 | 95.1 |
| 8 | Mutism,Grimacing,Echopraxia/echolalia,Stereotypy,Mannerisms,Verbigeration,Rigidity,Withdrawal | 92.8 | 90.4 | 95.1 |
| 8 | Mutism,Grimacing,Stereotypy,Mannerisms,Verbigeration,Negativism,Waxy flexibility,Withdrawal | 92.8 | 90.4 | 95.1 |
| 8 | Mutism,Posturing/catalepsy,Echopraxia/echolalia,Stereotypy,Mannerisms,Verbigeration,Negativism,Waxy flexibility | 92.8 | 90.4 | 95.1 |
| 8 | Mutism,Posturing/catalepsy,Echopraxia/echolalia,Stereotypy,Mannerisms,Verbigeration,Rigidity,Waxy flexibility | 92.8 | 90.4 | 95.1 |
| 8 | Mutism,Posturing/catalepsy,Grimacing,Echopraxia/echolalia,Mannerisms,Verbigeration,Waxy flexibility,Withdrawal | 92.8 | 90.4 | 95.1 |
| 8 | Mutism,Posturing/catalepsy,Grimacing,Echopraxia/echolalia,Stereotypy,Mannerisms,Verbigeration,Rigidity | 92.8 | 90.4 | 95.1 |
| 8 | Mutism,Posturing/catalepsy,Grimacing,Echopraxia/echolalia,Stereotypy,Mannerisms,Waxy flexibility,Withdrawal | 92.8 | 90.4 | 95.1 |
| 8 | Mutism,Posturing/catalepsy,Grimacing,Stereotypy,Mannerisms,Verbigeration,Negativism,Waxy flexibility | 92.8 | 90.4 | 95.1 |
| 8 | Posturing/catalepsy,Echopraxia/echolalia,Stereotypy,Mannerisms,Verbigeration,Negativism,Waxy flexibility,Withdrawal | 92.8 | 90.4 | 95.1 |
| 8 | Posturing/catalepsy,Grimacing,Echopraxia/echolalia,Mannerisms,Verbigeration,Rigidity,Waxy flexibility,Withdrawal | 92.8 | 90.4 | 95.1 |
| 8 | Posturing/catalepsy,Grimacing,Echopraxia/echolalia,Stereotypy,Mannerisms,Negativism,Waxy flexibility,Withdrawal | 92.8 | 90.4 | 95.1 |
| 8 | Posturing/catalepsy,Grimacing,Echopraxia/echolalia,Stereotypy,Mannerisms,Verbigeration,Negativism,Withdrawal | 92.8 | 90.4 | 95.1 |
| 8 | Posturing/catalepsy,Grimacing,Stereotypy,Mannerisms,Verbigeration,Negativism,Waxy flexibility,Withdrawal | 92.8 | 90.4 | 95.1 |
| 8 | Excitement,Grimacing,Echopraxia/echolalia,Stereotypy,Mannerisms,Rigidity,Negativism,Withdrawal | 92.6 | 90.1 | 94.8 |
| 8 | Excitement,Grimacing,Echopraxia/echolalia,Stereotypy,Rigidity,Negativism,Waxy flexibility,Withdrawal | 92.6 | 90.1 | 94.8 |
| 8 | Excitement,Mutism,Grimacing,Echopraxia/echolalia,Stereotypy,Mannerisms,Negativism,Withdrawal | 92.6 | 90.1 | 94.8 |
| 8 | Excitement,Mutism,Grimacing,Echopraxia/echolalia,Verbigeration,Negativism,Waxy flexibility,Withdrawal | 92.6 | 90.1 | 95.1 |
| 8 | Excitement,Mutism,Grimacing,Stereotypy,Mannerisms,Negativism,Waxy flexibility,Withdrawal | 92.6 | 90.1 | 94.8 |
| 8 | Excitement,Mutism,Grimacing,Stereotypy,Mannerisms,Verbigeration,Negativism,Withdrawal | 92.6 | 90.1 | 94.8 |
| 8 | Excitement,Mutism,Posturing/catalepsy,Grimacing,Echopraxia/echolalia,Stereotypy,Mannerisms,Withdrawal | 92.6 | 90.1 | 94.8 |
| 8 | Excitement,Mutism,Posturing/catalepsy,Grimacing,Stereotypy,Mannerisms,Negativism,Waxy flexibility | 92.6 | 90.1 | 94.8 |
| 8 | Excitement,Mutism,Posturing/catalepsy,Grimacing,Stereotypy,Mannerisms,Verbigeration,Withdrawal | 92.6 | 90.1 | 95.1 |
| 8 | Excitement,Posturing/catalepsy,Echopraxia/echolalia,Mannerisms,Verbigeration,Negativism,Waxy flexibility,Withdrawal | 92.6 | 90.1 | 94.8 |
| 8 | Excitement,Posturing/catalepsy,Echopraxia/echolalia,Stereotypy,Mannerisms,Negativism,Waxy flexibility,Withdrawal | 92.6 | 89.9 | 94.8 |
| 8 | Excitement,Posturing/catalepsy,Grimacing,Echopraxia/echolalia,Stereotypy,Mannerisms,Negativism,Withdrawal | 92.6 | 90.1 | 94.8 |
| 8 | Excitement,Posturing/catalepsy,Grimacing,Echopraxia/echolalia,Stereotypy,Verbigeration,Negativism,Withdrawal | 92.6 | 90.1 | 94.8 |
| 8 | Excitement,Posturing/catalepsy,Grimacing,Echopraxia/echolalia,Verbigeration,Negativism,Waxy flexibility,Withdrawal | 92.6 | 90.1 | 94.8 |
| 8 | Excitement,Posturing/catalepsy,Grimacing,Mannerisms,Verbigeration,Negativism,Waxy flexibility,Withdrawal | 92.6 | 90.1 | 95.1 |
| 8 | Excitement,Posturing/catalepsy,Grimacing,Stereotypy,Verbigeration,Negativism,Waxy flexibility,Withdrawal | 92.6 | 90.1 | 94.8 |
| 8 | Mutism,Grimacing,Echopraxia/echolalia,Stereotypy,Verbigeration,Negativism,Waxy flexibility,Withdrawal | 92.6 | 90.1 | 94.8 |
| 8 | Mutism,Posturing/catalepsy,Grimacing,Echopraxia/echolalia,Stereotypy,Verbigeration,Negativism,Waxy flexibility | 92.6 | 90.1 | 94.8 |
| 8 | Posturing/catalepsy,Grimacing,Echopraxia/echolalia,Stereotypy,Mannerisms,Rigidity,Waxy flexibility,Withdrawal | 92.6 | 90.1 | 94.8 |
| 8 | Posturing/catalepsy,Grimacing,Stereotypy,Mannerisms,Verbigeration,Rigidity,Waxy flexibility,Withdrawal | 92.6 | 90.1 | 94.8 |
| 8 | Excitement,Grimacing,Echopraxia/echolalia,Mannerisms,Rigidity,Negativism,Waxy flexibility,Withdrawal | 92.4 | 89.9 | 94.8 |
| 8 | Excitement,Grimacing,Echopraxia/echolalia,Stereotypy,Verbigeration,Rigidity,Negativism,Withdrawal | 92.4 | 89.9 | 94.6 |
| 8 | Excitement,Grimacing,Echopraxia/echolalia,Verbigeration,Rigidity,Negativism,Waxy flexibility,Withdrawal | 92.4 | 89.9 | 94.8 |
| 8 | Excitement,Grimacing,Stereotypy,Mannerisms,Rigidity,Negativism,Waxy flexibility,Withdrawal | 92.4 | 89.9 | 94.8 |
| 8 | Excitement,Grimacing,Stereotypy,Mannerisms,Verbigeration,Rigidity,Negativism,Withdrawal | 92.4 | 89.9 | 94.8 |
| 8 | Excitement,Grimacing,Stereotypy,Verbigeration,Rigidity,Negativism,Waxy flexibility,Withdrawal | 92.4 | 89.9 | 94.8 |
| 8 | Excitement,Mutism,Echopraxia/echolalia,Mannerisms,Verbigeration,Negativism,Waxy flexibility,Withdrawal | 92.4 | 89.9 | 94.8 |
| 8 | Excitement,Mutism,Grimacing,Echopraxia/echolalia,Mannerisms,Negativism,Waxy flexibility,Withdrawal | 92.4 | 89.9 | 94.8 |
| 8 | Excitement,Posturing/catalepsy,Grimacing,Echopraxia/echolalia,Mannerisms,Negativism,Waxy flexibility,Withdrawal | 92.4 | 89.9 | 94.8 |
| 8 | Excitement,Posturing/catalepsy,Grimacing,Echopraxia/echolalia,Stereotypy,Negativism,Waxy flexibility,Withdrawal | 92.4 | 89.9 | 94.8 |
| 8 | Grimacing,Echopraxia/echolalia,Mannerisms,Verbigeration,Rigidity,Negativism,Waxy flexibility,Withdrawal | 92.4 | 89.9 | 94.8 |
| 8 | Mutism,Grimacing,Echopraxia/echolalia,Mannerisms,Verbigeration,Negativism,Waxy flexibility,Withdrawal | 92.4 | 89.9 | 94.8 |
| 8 | Mutism,Grimacing,Echopraxia/echolalia,Stereotypy,Mannerisms,Verbigeration,Rigidity,Waxy flexibility | 92.4 | 89.9 | 94.8 |
| 8 | Mutism,Posturing/catalepsy,Grimacing,Stereotypy,Mannerisms,Verbigeration,Waxy flexibility,Withdrawal | 92.4 | 89.9 | 94.8 |
| 8 | Grimacing,Echopraxia/echolalia,Stereotypy,Verbigeration,Rigidity,Negativism,Waxy flexibility,Withdrawal | 92.3 | 89.9 | 94.6 |
| 8 | Excitement,Echopraxia/echolalia,Mannerisms,Verbigeration,Rigidity,Negativism,Waxy flexibility,Withdrawal | 92.2 | 89.7 | 94.6 |
| 8 | Excitement,Grimacing,Echopraxia/echolalia,Mannerisms,Verbigeration,Rigidity,Negativism,Withdrawal | 92.2 | 89.7 | 94.6 |
| 8 | Excitement,Grimacing,Mannerisms,Verbigeration,Rigidity,Negativism,Waxy flexibility,Withdrawal | 92.2 | 89.5 | 94.6 |
| 8 | Excitement,Mutism,Grimacing,Mannerisms,Verbigeration,Negativism,Waxy flexibility,Withdrawal | 92.2 | 89.7 | 94.6 |
| 8 | Excitement,Posturing/catalepsy,Grimacing,Stereotypy,Mannerisms,Negativism,Waxy flexibility,Withdrawal | 92.2 | 89.7 | 94.4 |
| 8 | Mutism,Posturing/catalepsy,Grimacing,Echopraxia/echolalia,Stereotypy,Verbigeration,Waxy flexibility,Withdrawal | 92.2 | 89.7 | 94.6 |
| 8 | Posturing/catalepsy,Grimacing,Echopraxia/echolalia,Mannerisms,Verbigeration,Negativism,Waxy flexibility,Withdrawal | 92.2 | 89.7 | 94.6 |
| 8 | Posturing/catalepsy,Grimacing,Echopraxia/echolalia,Stereotypy,Verbigeration,Rigidity,Waxy flexibility,Withdrawal | 92.2 | 89.7 | 94.6 |
| 8 | Posturing/catalepsy,Echopraxia/echolalia,Stereotypy,Mannerisms,Verbigeration,Rigidity,Waxy flexibility,Withdrawal | 92.1 | 89.5 | 94.6 |
| 8 | Posturing/catalepsy,Grimacing,Echopraxia/echolalia,Stereotypy,Mannerisms,Verbigeration,Rigidity,Withdrawal | 92.1 | 89.7 | 94.6 |
| 8 | Excitement,Mutism,Grimacing,Echopraxia/echolalia,Mannerisms,Verbigeration,Negativism,Withdrawal | 91.9 | 89.2 | 94.4 |
| 8 | Mutism,Posturing/catalepsy,Echopraxia/echolalia,Stereotypy,Mannerisms,Verbigeration,Waxy flexibility,Withdrawal | 91.9 | 89.5 | 94.4 |
| 8 | Mutism,Posturing/catalepsy,Grimacing,Echopraxia/echolalia,Stereotypy,Mannerisms,Verbigeration,Withdrawal | 91.9 | 89.2 | 94.4 |
| 8 | Posturing/catalepsy,Grimacing,Echopraxia/echolalia,Stereotypy,Verbigeration,Negativism,Waxy flexibility,Withdrawal | 91.9 | 89.2 | 94.4 |
| 8 | Excitement,Echopraxia/echolalia,Stereotypy,Mannerisms,Verbigeration,Rigidity,Waxy flexibility,Withdrawal | 91.7 | 89.0 | 94.2 |
| 8 | Excitement,Mutism,Echopraxia/echolalia,Stereotypy,Mannerisms,Verbigeration,Negativism,Waxy flexibility | 91.7 | 89.0 | 94.2 |
| 8 | Excitement,Mutism,Echopraxia/echolalia,Stereotypy,Mannerisms,Verbigeration,Waxy flexibility,Withdrawal | 91.7 | 89.0 | 94.2 |
| 8 | Excitement,Mutism,Grimacing,Echopraxia/echolalia,Stereotypy,Mannerisms,Verbigeration,Rigidity | 91.7 | 89.0 | 94.2 |
| 8 | Excitement,Mutism,Grimacing,Echopraxia/echolalia,Stereotypy,Verbigeration,Negativism,Waxy flexibility | 91.7 | 89.0 | 94.2 |
| 8 | Excitement,Mutism,Grimacing,Echopraxia/echolalia,Stereotypy,Verbigeration,Waxy flexibility,Withdrawal | 91.7 | 89.0 | 94.2 |
| 8 | Excitement,Posturing/catalepsy,Grimacing,Echopraxia/echolalia,Mannerisms,Verbigeration,Waxy flexibility,Withdrawal | 91.7 | 89.0 | 94.2 |
| 8 | Mutism,Grimacing,Echopraxia/echolalia,Stereotypy,Mannerisms,Verbigeration,Negativism,Waxy flexibility | 91.7 | 89.0 | 94.2 |
| 8 | Excitement,Grimacing,Echopraxia/echolalia,Stereotypy,Mannerisms,Rigidity,Waxy flexibility,Withdrawal | 91.5 | 88.8 | 93.9 |
| 8 | Excitement,Mutism,Grimacing,Echopraxia/echolalia,Stereotypy,Mannerisms,Negativism,Waxy flexibility | 91.5 | 88.8 | 93.9 |
| 8 | Excitement,Mutism,Grimacing,Echopraxia/echolalia,Stereotypy,Mannerisms,Waxy flexibility,Withdrawal | 91.5 | 88.8 | 93.9 |
| 8 | Excitement,Grimacing,Echopraxia/echolalia,Stereotypy,Verbigeration,Rigidity,Waxy flexibility,Withdrawal | 91.3 | 88.6 | 93.7 |
| 8 | Excitement,Mutism,Grimacing,Stereotypy,Mannerisms,Verbigeration,Negativism,Waxy flexibility | 91.3 | 88.6 | 93.7 |
| 8 | Excitement,Posturing/catalepsy,Grimacing,Echopraxia/echolalia,Stereotypy,Mannerisms,Waxy flexibility,Withdrawal | 91.3 | 88.6 | 93.9 |
| 8 | Excitement,Grimacing,Echopraxia/echolalia,Mannerisms,Verbigeration,Rigidity,Waxy flexibility,Withdrawal | 91.2 | 88.6 | 93.7 |
| 8 | Excitement,Mutism,Posturing/catalepsy,Grimacing,Echopraxia/echolalia,Stereotypy,Verbigeration,Waxy flexibility | 91.2 | 88.6 | 93.7 |
| 8 | Excitement,Grimacing,Stereotypy,Mannerisms,Verbigeration,Rigidity,Waxy flexibility,Withdrawal | 91.0 | 88.3 | 93.5 |
| 8 | Excitement,Mutism,Grimacing,Stereotypy,Mannerisms,Verbigeration,Waxy flexibility,Withdrawal | 91.0 | 88.3 | 93.5 |
| 8 | Excitement,Mutism,Posturing/catalepsy,Grimacing,Echopraxia/echolalia,Mannerisms,Verbigeration,Waxy flexibility | 91.0 | 88.3 | 93.5 |
| 8 | Excitement,Posturing/catalepsy,Echopraxia/echolalia,Stereotypy,Mannerisms,Verbigeration,Waxy flexibility,Withdrawal | 91.0 | 88.3 | 93.5 |
| 8 | Excitement,Posturing/catalepsy,Grimacing,Echopraxia/echolalia,Stereotypy,Verbigeration,Waxy flexibility,Withdrawal | 91.0 | 88.3 | 93.5 |
| 8 | Excitement,Posturing/catalepsy,Grimacing,Stereotypy,Mannerisms,Verbigeration,Waxy flexibility,Withdrawal | 91.0 | 88.3 | 93.5 |
| 8 | Grimacing,Echopraxia/echolalia,Stereotypy,Mannerisms,Verbigeration,Rigidity,Waxy flexibility,Withdrawal | 91.0 | 88.3 | 93.5 |
| 8 | Excitement,Mutism,Grimacing,Echopraxia/echolalia,Mannerisms,Verbigeration,Negativism,Waxy flexibility | 90.8 | 88.1 | 93.5 |
| 8 | Excitement,Mutism,Grimacing,Echopraxia/echolalia,Mannerisms,Verbigeration,Waxy flexibility,Withdrawal | 90.8 | 88.1 | 93.5 |
| 8 | Excitement,Mutism,Posturing/catalepsy,Echopraxia/echolalia,Stereotypy,Mannerisms,Verbigeration,Waxy flexibility | 90.8 | 88.1 | 93.5 |
| 8 | Excitement,Mutism,Posturing/catalepsy,Grimacing,Echopraxia/echolalia,Stereotypy,Mannerisms,Waxy flexibility | 90.8 | 88.1 | 93.5 |
| 8 | Excitement,Mutism,Posturing/catalepsy,Grimacing,Stereotypy,Mannerisms,Verbigeration,Waxy flexibility | 90.8 | 87.9 | 93.5 |
| 8 | Mutism,Grimacing,Echopraxia/echolalia,Stereotypy,Mannerisms,Verbigeration,Waxy flexibility,Withdrawal | 90.8 | 88.1 | 93.5 |
| 8 | Posturing/catalepsy,Grimacing,Echopraxia/echolalia,Stereotypy,Mannerisms,Verbigeration,Waxy flexibility,Withdrawal | 90.8 | 88.1 | 93.5 |
| 8 | Excitement,Mutism,Grimacing,Echopraxia/echolalia,Stereotypy,Mannerisms,Verbigeration,Negativism | 90.6 | 87.9 | 93.3 |
| 8 | Excitement,Posturing/catalepsy,Grimacing,Echopraxia/echolalia,Stereotypy,Mannerisms,Verbigeration,Withdrawal | 90.6 | 87.9 | 93.0 |
| 8 | Excitement,Posturing/catalepsy,Grimacing,Echopraxia/echolalia,Verbigeration,Rigidity,Negativism,Waxy flexibility | 90.6 | 87.7 | 93.3 |
| 8 | Excitement,Posturing/catalepsy,Grimacing,Mannerisms,Verbigeration,Rigidity,Negativism,Waxy flexibility | 90.4 | 87.4 | 93.0 |
| 8 | Posturing/catalepsy,Grimacing,Stereotypy,Mannerisms,Verbigeration,Rigidity,Negativism,Waxy flexibility | 90.4 | 87.4 | 93.0 |
| 8 | Excitement,Posturing/catalepsy,Grimacing,Echopraxia/echolalia,Mannerisms,Verbigeration,Rigidity,Negativism | 90.3 | 87.4 | 93.0 |
| 8 | Excitement,Posturing/catalepsy,Grimacing,Stereotypy,Verbigeration,Rigidity,Negativism,Waxy flexibility | 90.3 | 87.4 | 93.0 |
| 8 | Excitement,Posturing/catalepsy,Grimacing,Echopraxia/echolalia,Mannerisms,Rigidity,Negativism,Waxy flexibility | 90.2 | 87.2 | 92.8 |
| 8 | Excitement,Posturing/catalepsy,Grimacing,Stereotypy,Mannerisms,Verbigeration,Rigidity,Negativism | 90.2 | 87.2 | 92.8 |
| 8 | Excitement,Mutism,Posturing/catalepsy,Grimacing,Echopraxia/echolalia,Stereotypy,Mannerisms,Verbigeration | 90.1 | 87.2 | 92.8 |
| 8 | Excitement,Posturing/catalepsy,Echopraxia/echolalia,Stereotypy,Verbigeration,Rigidity,Negativism,Waxy flexibility | 90.1 | 87.4 | 92.8 |
| 8 | Excitement,Posturing/catalepsy,Grimacing,Echopraxia/echolalia,Stereotypy,Rigidity,Negativism,Waxy flexibility | 90.1 | 87.2 | 92.8 |
| 8 | Excitement,Posturing/catalepsy,Grimacing,Echopraxia/echolalia,Stereotypy,Verbigeration,Rigidity,Negativism | 90.1 | 87.2 | 92.8 |
| 8 | Mutism,Posturing/catalepsy,Grimacing,Echopraxia/echolalia,Stereotypy,Mannerisms,Verbigeration,Waxy flexibility | 90.1 | 87.2 | 92.8 |
| 8 | Posturing/catalepsy,Grimacing,Echopraxia/echolalia,Mannerisms,Verbigeration,Rigidity,Negativism,Waxy flexibility | 90.1 | 87.2 | 92.8 |
| 8 | Posturing/catalepsy,Grimacing,Echopraxia/echolalia,Stereotypy,Mannerisms,Rigidity,Negativism,Waxy flexibility | 90.1 | 87.2 | 92.8 |
| 8 | Excitement,Grimacing,Echopraxia/echolalia,Stereotypy,Mannerisms,Verbigeration,Rigidity,Withdrawal | 89.9 | 87.0 | 92.6 |
| 8 | Excitement,Posturing/catalepsy,Echopraxia/echolalia,Mannerisms,Verbigeration,Rigidity,Negativism,Waxy flexibility | 89.9 | 87.0 | 92.6 |
| 8 | Excitement,Posturing/catalepsy,Echopraxia/echolalia,Stereotypy,Mannerisms,Verbigeration,Rigidity,Negativism | 89.9 | 87.0 | 92.6 |
| 8 | Excitement,Posturing/catalepsy,Grimacing,Echopraxia/echolalia,Stereotypy,Mannerisms,Rigidity,Negativism | 89.9 | 87.0 | 92.6 |
| 8 | Excitement,Posturing/catalepsy,Stereotypy,Mannerisms,Verbigeration,Rigidity,Negativism,Waxy flexibility | 89.9 | 87.0 | 92.6 |
| 8 | Grimacing,Echopraxia/echolalia,Stereotypy,Mannerisms,Verbigeration,Negativism,Waxy flexibility,Withdrawal | 89.9 | 87.0 | 92.6 |
| 8 | Posturing/catalepsy,Echopraxia/echolalia,Stereotypy,Mannerisms,Verbigeration,Rigidity,Negativism,Waxy flexibility | 89.9 | 87.0 | 92.6 |
| 8 | Posturing/catalepsy,Grimacing,Echopraxia/echolalia,Stereotypy,Mannerisms,Verbigeration,Rigidity,Negativism | 89.9 | 87.0 | 92.6 |
| 8 | Excitement,Echopraxia/echolalia,Stereotypy,Mannerisms,Verbigeration,Negativism,Waxy flexibility,Withdrawal | 89.7 | 86.8 | 92.4 |
| 8 | Excitement,Mutism,Grimacing,Echopraxia/echolalia,Stereotypy,Mannerisms,Verbigeration,Withdrawal | 89.7 | 86.8 | 92.4 |
| 8 | Excitement,Posturing/catalepsy,Echopraxia/echolalia,Stereotypy,Mannerisms,Rigidity,Negativism,Waxy flexibility | 89.7 | 86.8 | 92.4 |
| 8 | Excitement,Posturing/catalepsy,Grimacing,Stereotypy,Mannerisms,Rigidity,Negativism,Waxy flexibility | 89.7 | 86.8 | 92.4 |
| 8 | Posturing/catalepsy,Grimacing,Echopraxia/echolalia,Stereotypy,Verbigeration,Rigidity,Negativism,Waxy flexibility | 89.7 | 86.8 | 92.4 |
| 8 | Excitement,Grimacing,Echopraxia/echolalia,Stereotypy,Mannerisms,Negativism,Waxy flexibility,Withdrawal | 89.5 | 86.5 | 92.2 |
| 8 | Excitement,Grimacing,Echopraxia/echolalia,Stereotypy,Verbigeration,Negativism,Waxy flexibility,Withdrawal | 89.2 | 86.3 | 91.9 |
| 8 | Excitement,Grimacing,Stereotypy,Mannerisms,Verbigeration,Negativism,Waxy flexibility,Withdrawal | 88.8 | 85.7 | 91.7 |
| 8 | Excitement,Grimacing,Echopraxia/echolalia,Mannerisms,Verbigeration,Negativism,Waxy flexibility,Withdrawal | 88.6 | 85.4 | 91.5 |
| 8 | Excitement,Grimacing,Echopraxia/echolalia,Stereotypy,Mannerisms,Verbigeration,Negativism,Withdrawal | 88.6 | 85.4 | 91.5 |
| 8 | Excitement,Mutism,Grimacing,Echopraxia/echolalia,Stereotypy,Mannerisms,Verbigeration,Waxy flexibility | 88.5 | 85.4 | 91.5 |
| 8 | Excitement,Posturing/catalepsy,Grimacing,Echopraxia/echolalia,Mannerisms,Verbigeration,Rigidity,Waxy flexibility | 87.4 | 84.3 | 90.4 |
| 8 | Excitement,Grimacing,Echopraxia/echolalia,Stereotypy,Mannerisms,Verbigeration,Waxy flexibility,Withdrawal | 87.2 | 84.1 | 90.1 |
| 8 | Excitement,Posturing/catalepsy,Grimacing,Echopraxia/echolalia,Mannerisms,Verbigeration,Negativism,Waxy flexibility | 87.2 | 84.1 | 90.1 |
| 8 | Excitement,Posturing/catalepsy,Grimacing,Echopraxia/echolalia,Stereotypy,Verbigeration,Negativism,Waxy flexibility | 87.2 | 84.1 | 90.1 |
| 8 | Excitement,Posturing/catalepsy,Grimacing,Stereotypy,Mannerisms,Verbigeration,Negativism,Waxy flexibility | 87.2 | 84.1 | 90.4 |
| 8 | Posturing/catalepsy,Grimacing,Echopraxia/echolalia,Stereotypy,Mannerisms,Verbigeration,Negativism,Waxy flexibility | 87.2 | 84.1 | 90.4 |
| 8 | Excitement,Posturing/catalepsy,Grimacing,Echopraxia/echolalia,Stereotypy,Mannerisms,Negativism,Waxy flexibility | 87.0 | 83.9 | 90.1 |
| 8 | Excitement,Posturing/catalepsy,Grimacing,Echopraxia/echolalia,Stereotypy,Verbigeration,Rigidity,Waxy flexibility | 87.0 | 83.9 | 89.9 |
| 8 | Excitement,Grimacing,Echopraxia/echolalia,Stereotypy,Mannerisms,Rigidity,Negativism,Waxy flexibility | 86.8 | 83.6 | 89.7 |
| 8 | Excitement,Grimacing,Echopraxia/echolalia,Stereotypy,Verbigeration,Rigidity,Negativism,Waxy flexibility | 86.8 | 83.6 | 89.9 |
| 8 | Grimacing,Echopraxia/echolalia,Stereotypy,Mannerisms,Verbigeration,Rigidity,Negativism,Waxy flexibility | 86.8 | 83.6 | 89.9 |
| 8 | Excitement,Posturing/catalepsy,Grimacing,Echopraxia/echolalia,Stereotypy,Mannerisms,Verbigeration,Negativism | 86.7 | 83.4 | 89.7 |
| 8 | Excitement,Grimacing,Stereotypy,Mannerisms,Verbigeration,Rigidity,Negativism,Waxy flexibility | 86.6 | 83.4 | 89.5 |
| 8 | Excitement,Posturing/catalepsy,Grimacing,Stereotypy,Mannerisms,Verbigeration,Rigidity,Waxy flexibility | 86.6 | 83.4 | 89.7 |
| 8 | Excitement,Posturing/catalepsy,Grimacing,Echopraxia/echolalia,Stereotypy,Mannerisms,Rigidity,Waxy flexibility | 86.5 | 83.2 | 89.7 |
| 8 | Excitement,Echopraxia/echolalia,Stereotypy,Mannerisms,Verbigeration,Rigidity,Negativism,Waxy flexibility | 86.3 | 83.2 | 89.5 |
| 8 | Excitement,Posturing/catalepsy,Echopraxia/echolalia,Stereotypy,Mannerisms,Verbigeration,Negativism,Waxy flexibility | 86.3 | 83.0 | 89.5 |
| 8 | Excitement,Posturing/catalepsy,Echopraxia/echolalia,Stereotypy,Mannerisms,Verbigeration,Rigidity,Waxy flexibility | 85.9 | 82.5 | 89.0 |
| 8 | Excitement,Posturing/catalepsy,Grimacing,Echopraxia/echolalia,Stereotypy,Mannerisms,Verbigeration,Rigidity | 85.9 | 82.5 | 89.0 |
| 8 | Posturing/catalepsy,Grimacing,Echopraxia/echolalia,Stereotypy,Mannerisms,Verbigeration,Rigidity,Waxy flexibility | 85.9 | 82.5 | 89.0 |
| 8 | Excitement,Grimacing,Echopraxia/echolalia,Mannerisms,Verbigeration,Rigidity,Negativism,Waxy flexibility | 85.7 | 82.3 | 88.8 |
| 8 | Excitement,Grimacing,Echopraxia/echolalia,Stereotypy,Mannerisms,Verbigeration,Rigidity,Negativism | 85.2 | 81.8 | 88.3 |
| 8 | Excitement,Posturing/catalepsy,Grimacing,Echopraxia/echolalia,Stereotypy,Mannerisms,Verbigeration,Waxy flexibility | 82.3 | 78.7 | 85.9 |
| 8 | Excitement,Grimacing,Echopraxia/echolalia,Stereotypy,Mannerisms,Verbigeration,Rigidity,Waxy flexibility | 82.1 | 78.5 | 85.7 |
| 8 | Excitement,Grimacing,Echopraxia/echolalia,Stereotypy,Mannerisms,Verbigeration,Negativism,Waxy flexibility | 80.9 | 77.4 | 84.5 |

Table S9: sensitivity (relative to the full BFCSI) of all nine question combinations, and the 95% confidence interval derived from bootstrapping, sorted by decreasing sensitivity.

|  |  |  | 95% CI | |
| --- | --- | --- | --- | --- |
| # of Qs | Combination | Sensitivity | Lower | Upper |
| 9 | Excitement,Immobility/stupor,Mutism,Staring,Echopraxia/echolalia,Stereotypy,Rigidity,Negativism,Withdrawal | 100.0 | 100.0 | 100.0 |
| 9 | Excitement,Immobility/stupor,Mutism,Staring,Echopraxia/echolalia,Stereotypy,Rigidity,Waxy flexibility,Withdrawal | 100.0 | 100.0 | 100.0 |
| 9 | Excitement,Immobility/stupor,Mutism,Staring,Posturing/catalepsy,Echopraxia/echolalia,Mannerisms,Negativism,Withdrawal | 100.0 | 100.0 | 100.0 |
| 9 | Excitement,Immobility/stupor,Mutism,Staring,Posturing/catalepsy,Echopraxia/echolalia,Mannerisms,Rigidity,Withdrawal | 100.0 | 100.0 | 100.0 |
| 9 | Excitement,Immobility/stupor,Mutism,Staring,Posturing/catalepsy,Echopraxia/echolalia,Negativism,Waxy flexibility,Withdrawal | 100.0 | 100.0 | 100.0 |
| 9 | Excitement,Immobility/stupor,Mutism,Staring,Posturing/catalepsy,Echopraxia/echolalia,Rigidity,Negativism,Withdrawal | 100.0 | 100.0 | 100.0 |
| 9 | Excitement,Immobility/stupor,Mutism,Staring,Posturing/catalepsy,Echopraxia/echolalia,Rigidity,Waxy flexibility,Withdrawal | 100.0 | 100.0 | 100.0 |
| 9 | Excitement,Immobility/stupor,Mutism,Staring,Posturing/catalepsy,Echopraxia/echolalia,Stereotypy,Negativism,Withdrawal | 100.0 | 100.0 | 100.0 |
| 9 | Excitement,Immobility/stupor,Mutism,Staring,Posturing/catalepsy,Echopraxia/echolalia,Stereotypy,Rigidity,Withdrawal | 100.0 | 100.0 | 100.0 |
| 9 | Excitement,Immobility/stupor,Mutism,Staring,Posturing/catalepsy,Echopraxia/echolalia,Verbigeration,Negativism,Waxy flexibility | 100.0 | 100.0 | 100.0 |
| 9 | Excitement,Immobility/stupor,Mutism,Staring,Posturing/catalepsy,Echopraxia/echolalia,Verbigeration,Negativism,Withdrawal | 100.0 | 100.0 | 100.0 |
| 9 | Excitement,Immobility/stupor,Mutism,Staring,Posturing/catalepsy,Echopraxia/echolalia,Verbigeration,Rigidity,Negativism | 100.0 | 100.0 | 100.0 |
| 9 | Excitement,Immobility/stupor,Mutism,Staring,Posturing/catalepsy,Echopraxia/echolalia,Verbigeration,Rigidity,Withdrawal | 100.0 | 100.0 | 100.0 |
| 9 | Excitement,Immobility/stupor,Mutism,Staring,Posturing/catalepsy,Grimacing,Echopraxia/echolalia,Negativism,Withdrawal | 100.0 | 100.0 | 100.0 |
| 9 | Excitement,Immobility/stupor,Mutism,Staring,Posturing/catalepsy,Grimacing,Echopraxia/echolalia,Rigidity,Withdrawal | 100.0 | 100.0 | 100.0 |
| 9 | Excitement,Immobility/stupor,Mutism,Staring,Posturing/catalepsy,Mannerisms,Verbigeration,Negativism,Waxy flexibility | 100.0 | 100.0 | 100.0 |
| 9 | Excitement,Immobility/stupor,Mutism,Staring,Posturing/catalepsy,Mannerisms,Verbigeration,Negativism,Withdrawal | 100.0 | 100.0 | 100.0 |
| 9 | Excitement,Immobility/stupor,Mutism,Staring,Posturing/catalepsy,Mannerisms,Verbigeration,Rigidity,Negativism | 100.0 | 100.0 | 100.0 |
| 9 | Excitement,Immobility/stupor,Mutism,Staring,Posturing/catalepsy,Mannerisms,Verbigeration,Rigidity,Withdrawal | 100.0 | 100.0 | 100.0 |
| 9 | Excitement,Immobility/stupor,Mutism,Staring,Posturing/catalepsy,Stereotypy,Verbigeration,Negativism,Waxy flexibility | 100.0 | 100.0 | 100.0 |
| 9 | Excitement,Immobility/stupor,Mutism,Staring,Posturing/catalepsy,Stereotypy,Verbigeration,Negativism,Withdrawal | 100.0 | 100.0 | 100.0 |
| 9 | Excitement,Immobility/stupor,Mutism,Staring,Posturing/catalepsy,Stereotypy,Verbigeration,Rigidity,Negativism | 100.0 | 100.0 | 100.0 |
| 9 | Excitement,Immobility/stupor,Mutism,Staring,Stereotypy,Verbigeration,Rigidity,Negativism,Waxy flexibility | 100.0 | 100.0 | 100.0 |
| 9 | Excitement,Immobility/stupor,Mutism,Staring,Stereotypy,Verbigeration,Rigidity,Negativism,Withdrawal | 100.0 | 100.0 | 100.0 |
| 9 | Excitement,Immobility/stupor,Mutism,Staring,Stereotypy,Verbigeration,Rigidity,Waxy flexibility,Withdrawal | 100.0 | 100.0 | 100.0 |
| 9 | Excitement,Immobility/stupor,Staring,Echopraxia/echolalia,Stereotypy,Mannerisms,Rigidity,Negativism,Withdrawal | 100.0 | 100.0 | 100.0 |
| 9 | Excitement,Immobility/stupor,Staring,Posturing/catalepsy,Echopraxia/echolalia,Mannerisms,Rigidity,Negativism,Withdrawal | 100.0 | 100.0 | 100.0 |
| 9 | Excitement,Immobility/stupor,Staring,Posturing/catalepsy,Mannerisms,Verbigeration,Rigidity,Negativism,Withdrawal | 100.0 | 100.0 | 100.0 |
| 9 | Excitement,Immobility/stupor,Staring,Stereotypy,Mannerisms,Verbigeration,Rigidity,Negativism,Withdrawal | 100.0 | 100.0 | 100.0 |
| 9 | Excitement,Mutism,Staring,Echopraxia/echolalia,Stereotypy,Mannerisms,Rigidity,Negativism,Withdrawal | 100.0 | 100.0 | 100.0 |
| 9 | Excitement,Mutism,Staring,Echopraxia/echolalia,Stereotypy,Mannerisms,Rigidity,Waxy flexibility,Withdrawal | 100.0 | 100.0 | 100.0 |
| 9 | Excitement,Mutism,Staring,Echopraxia/echolalia,Stereotypy,Rigidity,Negativism,Waxy flexibility,Withdrawal | 100.0 | 100.0 | 100.0 |
| 9 | Excitement,Mutism,Staring,Echopraxia/echolalia,Stereotypy,Verbigeration,Rigidity,Negativism,Waxy flexibility | 100.0 | 100.0 | 100.0 |
| 9 | Excitement,Mutism,Staring,Echopraxia/echolalia,Stereotypy,Verbigeration,Rigidity,Negativism,Withdrawal | 100.0 | 100.0 | 100.0 |
| 9 | Excitement,Mutism,Staring,Echopraxia/echolalia,Stereotypy,Verbigeration,Rigidity,Waxy flexibility,Withdrawal | 100.0 | 100.0 | 100.0 |
| 9 | Excitement,Mutism,Staring,Grimacing,Echopraxia/echolalia,Stereotypy,Rigidity,Negativism,Withdrawal | 100.0 | 100.0 | 100.0 |
| 9 | Excitement,Mutism,Staring,Grimacing,Echopraxia/echolalia,Stereotypy,Rigidity,Waxy flexibility,Withdrawal | 100.0 | 100.0 | 100.0 |
| 9 | Excitement,Mutism,Staring,Grimacing,Stereotypy,Verbigeration,Rigidity,Negativism,Withdrawal | 100.0 | 100.0 | 100.0 |
| 9 | Excitement,Mutism,Staring,Grimacing,Stereotypy,Verbigeration,Rigidity,Waxy flexibility,Withdrawal | 100.0 | 100.0 | 100.0 |
| 9 | Excitement,Mutism,Staring,Posturing/catalepsy,Echopraxia/echolalia,Mannerisms,Negativism,Waxy flexibility,Withdrawal | 100.0 | 100.0 | 100.0 |
| 9 | Excitement,Mutism,Staring,Posturing/catalepsy,Echopraxia/echolalia,Mannerisms,Rigidity,Negativism,Withdrawal | 100.0 | 100.0 | 100.0 |
| 9 | Excitement,Mutism,Staring,Posturing/catalepsy,Echopraxia/echolalia,Mannerisms,Rigidity,Waxy flexibility,Withdrawal | 100.0 | 100.0 | 100.0 |
| 9 | Excitement,Mutism,Staring,Posturing/catalepsy,Echopraxia/echolalia,Mannerisms,Verbigeration,Negativism,Waxy flexibility | 100.0 | 100.0 | 100.0 |
| 9 | Excitement,Mutism,Staring,Posturing/catalepsy,Echopraxia/echolalia,Mannerisms,Verbigeration,Negativism,Withdrawal | 100.0 | 100.0 | 100.0 |
| 9 | Excitement,Mutism,Staring,Posturing/catalepsy,Echopraxia/echolalia,Mannerisms,Verbigeration,Rigidity,Negativism | 100.0 | 100.0 | 100.0 |
| 9 | Excitement,Mutism,Staring,Posturing/catalepsy,Echopraxia/echolalia,Mannerisms,Verbigeration,Rigidity,Withdrawal | 100.0 | 100.0 | 100.0 |
| 9 | Excitement,Mutism,Staring,Posturing/catalepsy,Echopraxia/echolalia,Rigidity,Negativism,Waxy flexibility,Withdrawal | 100.0 | 100.0 | 100.0 |
| 9 | Excitement,Mutism,Staring,Posturing/catalepsy,Echopraxia/echolalia,Stereotypy,Mannerisms,Negativism,Withdrawal | 100.0 | 100.0 | 100.0 |
| 9 | Excitement,Mutism,Staring,Posturing/catalepsy,Echopraxia/echolalia,Stereotypy,Mannerisms,Rigidity,Withdrawal | 100.0 | 100.0 | 100.0 |
| 9 | Excitement,Mutism,Staring,Posturing/catalepsy,Echopraxia/echolalia,Stereotypy,Negativism,Waxy flexibility,Withdrawal | 100.0 | 100.0 | 100.0 |
| 9 | Excitement,Mutism,Staring,Posturing/catalepsy,Echopraxia/echolalia,Stereotypy,Rigidity,Negativism,Withdrawal | 100.0 | 100.0 | 100.0 |
| 9 | Excitement,Mutism,Staring,Posturing/catalepsy,Echopraxia/echolalia,Stereotypy,Rigidity,Waxy flexibility,Withdrawal | 100.0 | 100.0 | 100.0 |
| 9 | Excitement,Mutism,Staring,Posturing/catalepsy,Echopraxia/echolalia,Stereotypy,Verbigeration,Negativism,Waxy flexibility | 100.0 | 100.0 | 100.0 |
| 9 | Excitement,Mutism,Staring,Posturing/catalepsy,Echopraxia/echolalia,Stereotypy,Verbigeration,Negativism,Withdrawal | 100.0 | 100.0 | 100.0 |
| 9 | Excitement,Mutism,Staring,Posturing/catalepsy,Echopraxia/echolalia,Stereotypy,Verbigeration,Rigidity,Negativism | 100.0 | 100.0 | 100.0 |
| 9 | Excitement,Mutism,Staring,Posturing/catalepsy,Echopraxia/echolalia,Stereotypy,Verbigeration,Rigidity,Withdrawal | 100.0 | 100.0 | 100.0 |
| 9 | Excitement,Mutism,Staring,Posturing/catalepsy,Echopraxia/echolalia,Verbigeration,Negativism,Waxy flexibility,Withdrawal | 100.0 | 100.0 | 100.0 |
| 9 | Excitement,Mutism,Staring,Posturing/catalepsy,Echopraxia/echolalia,Verbigeration,Rigidity,Negativism,Waxy flexibility | 100.0 | 100.0 | 100.0 |
| 9 | Excitement,Mutism,Staring,Posturing/catalepsy,Echopraxia/echolalia,Verbigeration,Rigidity,Negativism,Withdrawal | 100.0 | 100.0 | 100.0 |
| 9 | Excitement,Mutism,Staring,Posturing/catalepsy,Echopraxia/echolalia,Verbigeration,Rigidity,Waxy flexibility,Withdrawal | 100.0 | 100.0 | 100.0 |
| 9 | Excitement,Mutism,Staring,Posturing/catalepsy,Grimacing,Echopraxia/echolalia,Mannerisms,Negativism,Withdrawal | 100.0 | 100.0 | 100.0 |
| 9 | Excitement,Mutism,Staring,Posturing/catalepsy,Grimacing,Echopraxia/echolalia,Mannerisms,Rigidity,Withdrawal | 100.0 | 100.0 | 100.0 |
| 9 | Excitement,Mutism,Staring,Posturing/catalepsy,Grimacing,Echopraxia/echolalia,Negativism,Waxy flexibility,Withdrawal | 100.0 | 100.0 | 100.0 |
| 9 | Excitement,Mutism,Staring,Posturing/catalepsy,Grimacing,Echopraxia/echolalia,Rigidity,Negativism,Withdrawal | 100.0 | 100.0 | 100.0 |
| 9 | Excitement,Mutism,Staring,Posturing/catalepsy,Grimacing,Echopraxia/echolalia,Rigidity,Waxy flexibility,Withdrawal | 100.0 | 100.0 | 100.0 |
| 9 | Excitement,Mutism,Staring,Posturing/catalepsy,Grimacing,Echopraxia/echolalia,Stereotypy,Negativism,Withdrawal | 100.0 | 100.0 | 100.0 |
| 9 | Excitement,Mutism,Staring,Posturing/catalepsy,Grimacing,Echopraxia/echolalia,Stereotypy,Rigidity,Withdrawal | 100.0 | 100.0 | 100.0 |
| 9 | Excitement,Mutism,Staring,Posturing/catalepsy,Grimacing,Echopraxia/echolalia,Verbigeration,Negativism,Waxy flexibility | 100.0 | 100.0 | 100.0 |
| 9 | Excitement,Mutism,Staring,Posturing/catalepsy,Grimacing,Echopraxia/echolalia,Verbigeration,Negativism,Withdrawal | 100.0 | 100.0 | 100.0 |
| 9 | Excitement,Mutism,Staring,Posturing/catalepsy,Grimacing,Echopraxia/echolalia,Verbigeration,Rigidity,Negativism | 100.0 | 100.0 | 100.0 |
| 9 | Excitement,Mutism,Staring,Posturing/catalepsy,Grimacing,Echopraxia/echolalia,Verbigeration,Rigidity,Withdrawal | 100.0 | 100.0 | 100.0 |
| 9 | Excitement,Mutism,Staring,Posturing/catalepsy,Grimacing,Mannerisms,Verbigeration,Negativism,Withdrawal | 100.0 | 100.0 | 100.0 |
| 9 | Excitement,Mutism,Staring,Posturing/catalepsy,Grimacing,Mannerisms,Verbigeration,Rigidity,Withdrawal | 100.0 | 100.0 | 100.0 |
| 9 | Excitement,Mutism,Staring,Posturing/catalepsy,Grimacing,Stereotypy,Verbigeration,Negativism,Withdrawal | 100.0 | 100.0 | 100.0 |
| 9 | Excitement,Mutism,Staring,Posturing/catalepsy,Grimacing,Stereotypy,Verbigeration,Rigidity,Withdrawal | 100.0 | 100.0 | 100.0 |
| 9 | Excitement,Mutism,Staring,Posturing/catalepsy,Mannerisms,Verbigeration,Negativism,Waxy flexibility,Withdrawal | 100.0 | 100.0 | 100.0 |
| 9 | Excitement,Mutism,Staring,Posturing/catalepsy,Mannerisms,Verbigeration,Rigidity,Negativism,Withdrawal | 100.0 | 100.0 | 100.0 |
| 9 | Excitement,Mutism,Staring,Posturing/catalepsy,Mannerisms,Verbigeration,Rigidity,Waxy flexibility,Withdrawal | 100.0 | 100.0 | 100.0 |
| 9 | Excitement,Mutism,Staring,Posturing/catalepsy,Stereotypy,Mannerisms,Verbigeration,Negativism,Withdrawal | 100.0 | 100.0 | 100.0 |
| 9 | Excitement,Mutism,Staring,Posturing/catalepsy,Stereotypy,Mannerisms,Verbigeration,Rigidity,Withdrawal | 100.0 | 100.0 | 100.0 |
| 9 | Excitement,Mutism,Staring,Posturing/catalepsy,Stereotypy,Verbigeration,Negativism,Waxy flexibility,Withdrawal | 100.0 | 100.0 | 100.0 |
| 9 | Excitement,Mutism,Staring,Posturing/catalepsy,Stereotypy,Verbigeration,Rigidity,Negativism,Withdrawal | 100.0 | 100.0 | 100.0 |
| 9 | Excitement,Mutism,Staring,Posturing/catalepsy,Stereotypy,Verbigeration,Rigidity,Waxy flexibility,Withdrawal | 100.0 | 100.0 | 100.0 |
| 9 | Excitement,Mutism,Staring,Stereotypy,Mannerisms,Verbigeration,Rigidity,Negativism,Withdrawal | 100.0 | 100.0 | 100.0 |
| 9 | Excitement,Mutism,Staring,Stereotypy,Mannerisms,Verbigeration,Rigidity,Waxy flexibility,Withdrawal | 100.0 | 100.0 | 100.0 |
| 9 | Excitement,Mutism,Staring,Stereotypy,Verbigeration,Rigidity,Negativism,Waxy flexibility,Withdrawal | 100.0 | 100.0 | 100.0 |
| 9 | Immobility/stupor,Staring,Grimacing,Echopraxia/echolalia,Stereotypy,Mannerisms,Rigidity,Negativism,Withdrawal | 100.0 | 100.0 | 100.0 |
| 9 | Immobility/stupor,Staring,Grimacing,Stereotypy,Mannerisms,Verbigeration,Rigidity,Negativism,Withdrawal | 100.0 | 100.0 | 100.0 |
| 9 | Mutism,Staring,Grimacing,Echopraxia/echolalia,Stereotypy,Mannerisms,Rigidity,Negativism,Withdrawal | 100.0 | 100.0 | 100.0 |
| 9 | Mutism,Staring,Grimacing,Stereotypy,Mannerisms,Verbigeration,Rigidity,Negativism,Withdrawal | 100.0 | 100.0 | 100.0 |
| 9 | Mutism,Staring,Posturing/catalepsy,Grimacing,Echopraxia/echolalia,Stereotypy,Mannerisms,Negativism,Withdrawal | 100.0 | 100.0 | 100.0 |
| 9 | Mutism,Staring,Posturing/catalepsy,Grimacing,Stereotypy,Mannerisms,Verbigeration,Negativism,Withdrawal | 100.0 | 100.0 | 100.0 |
| 9 | Excitement,Immobility/stupor,Mutism,Echopraxia/echolalia,Stereotypy,Verbigeration,Rigidity,Negativism,Withdrawal | 99.8 | 99.3 | 100.0 |
| 9 | Excitement,Immobility/stupor,Mutism,Posturing/catalepsy,Echopraxia/echolalia,Stereotypy,Verbigeration,Rigidity,Negativism | 99.8 | 99.3 | 100.0 |
| 9 | Excitement,Immobility/stupor,Mutism,Posturing/catalepsy,Echopraxia/echolalia,Verbigeration,Rigidity,Negativism,Withdrawal | 99.8 | 99.3 | 100.0 |
| 9 | Excitement,Immobility/stupor,Mutism,Posturing/catalepsy,Grimacing,Echopraxia/echolalia,Rigidity,Negativism,Withdrawal | 99.8 | 99.3 | 100.0 |
| 9 | Excitement,Immobility/stupor,Mutism,Posturing/catalepsy,Grimacing,Echopraxia/echolalia,Verbigeration,Rigidity,Negativism | 99.8 | 99.3 | 100.0 |
| 9 | Excitement,Immobility/stupor,Mutism,Posturing/catalepsy,Stereotypy,Verbigeration,Rigidity,Negativism,Withdrawal | 99.8 | 99.3 | 100.0 |
| 9 | Excitement,Immobility/stupor,Mutism,Staring,Echopraxia/echolalia,Mannerisms,Rigidity,Negativism,Withdrawal | 99.8 | 99.3 | 100.0 |
| 9 | Excitement,Immobility/stupor,Mutism,Staring,Echopraxia/echolalia,Mannerisms,Rigidity,Waxy flexibility,Withdrawal | 99.8 | 99.3 | 100.0 |
| 9 | Excitement,Immobility/stupor,Mutism,Staring,Echopraxia/echolalia,Rigidity,Negativism,Waxy flexibility,Withdrawal | 99.8 | 99.3 | 100.0 |
| 9 | Excitement,Immobility/stupor,Mutism,Staring,Echopraxia/echolalia,Stereotypy,Mannerisms,Rigidity,Withdrawal | 99.8 | 99.3 | 100.0 |
| 9 | Excitement,Immobility/stupor,Mutism,Staring,Echopraxia/echolalia,Stereotypy,Rigidity,Negativism,Waxy flexibility | 99.8 | 99.3 | 100.0 |
| 9 | Excitement,Immobility/stupor,Mutism,Staring,Echopraxia/echolalia,Stereotypy,Verbigeration,Rigidity,Negativism | 99.8 | 99.3 | 100.0 |
| 9 | Excitement,Immobility/stupor,Mutism,Staring,Echopraxia/echolalia,Stereotypy,Verbigeration,Rigidity,Waxy flexibility | 99.8 | 99.3 | 100.0 |
| 9 | Excitement,Immobility/stupor,Mutism,Staring,Echopraxia/echolalia,Stereotypy,Verbigeration,Rigidity,Withdrawal | 99.8 | 99.3 | 100.0 |
| 9 | Excitement,Immobility/stupor,Mutism,Staring,Echopraxia/echolalia,Verbigeration,Rigidity,Negativism,Waxy flexibility | 99.8 | 99.3 | 100.0 |
| 9 | Excitement,Immobility/stupor,Mutism,Staring,Echopraxia/echolalia,Verbigeration,Rigidity,Negativism,Withdrawal | 99.8 | 99.3 | 100.0 |
| 9 | Excitement,Immobility/stupor,Mutism,Staring,Echopraxia/echolalia,Verbigeration,Rigidity,Waxy flexibility,Withdrawal | 99.8 | 99.3 | 100.0 |
| 9 | Excitement,Immobility/stupor,Mutism,Staring,Grimacing,Echopraxia/echolalia,Rigidity,Negativism,Withdrawal | 99.8 | 99.3 | 100.0 |
| 9 | Excitement,Immobility/stupor,Mutism,Staring,Grimacing,Echopraxia/echolalia,Rigidity,Waxy flexibility,Withdrawal | 99.8 | 99.3 | 100.0 |
| 9 | Excitement,Immobility/stupor,Mutism,Staring,Grimacing,Echopraxia/echolalia,Stereotypy,Rigidity,Withdrawal | 99.8 | 99.3 | 100.0 |
| 9 | Excitement,Immobility/stupor,Mutism,Staring,Grimacing,Stereotypy,Rigidity,Waxy flexibility,Withdrawal | 99.8 | 99.3 | 100.0 |
| 9 | Excitement,Immobility/stupor,Mutism,Staring,Grimacing,Stereotypy,Verbigeration,Rigidity,Negativism | 99.8 | 99.3 | 100.0 |
| 9 | Excitement,Immobility/stupor,Mutism,Staring,Grimacing,Stereotypy,Verbigeration,Rigidity,Waxy flexibility | 99.8 | 99.3 | 100.0 |
| 9 | Excitement,Immobility/stupor,Mutism,Staring,Grimacing,Stereotypy,Verbigeration,Rigidity,Withdrawal | 99.8 | 99.3 | 100.0 |
| 9 | Excitement,Immobility/stupor,Mutism,Staring,Mannerisms,Verbigeration,Rigidity,Negativism,Waxy flexibility | 99.8 | 99.3 | 100.0 |
| 9 | Excitement,Immobility/stupor,Mutism,Staring,Mannerisms,Verbigeration,Rigidity,Negativism,Withdrawal | 99.8 | 99.3 | 100.0 |
| 9 | Excitement,Immobility/stupor,Mutism,Staring,Mannerisms,Verbigeration,Rigidity,Waxy flexibility,Withdrawal | 99.8 | 99.3 | 100.0 |
| 9 | Excitement,Immobility/stupor,Mutism,Staring,Posturing/catalepsy,Echopraxia/echolalia,Mannerisms,Negativism,Waxy flexibility | 99.8 | 99.3 | 100.0 |
| 9 | Excitement,Immobility/stupor,Mutism,Staring,Posturing/catalepsy,Echopraxia/echolalia,Mannerisms,Rigidity,Negativism | 99.8 | 99.3 | 100.0 |
| 9 | Excitement,Immobility/stupor,Mutism,Staring,Posturing/catalepsy,Echopraxia/echolalia,Mannerisms,Verbigeration,Negativism | 99.8 | 99.3 | 100.0 |
| 9 | Excitement,Immobility/stupor,Mutism,Staring,Posturing/catalepsy,Echopraxia/echolalia,Mannerisms,Verbigeration,Rigidity | 99.8 | 99.3 | 100.0 |
| 9 | Excitement,Immobility/stupor,Mutism,Staring,Posturing/catalepsy,Echopraxia/echolalia,Rigidity,Negativism,Waxy flexibility | 99.8 | 99.3 | 100.0 |
| 9 | Excitement,Immobility/stupor,Mutism,Staring,Posturing/catalepsy,Echopraxia/echolalia,Stereotypy,Negativism,Waxy flexibility | 99.8 | 99.3 | 100.0 |
| 9 | Excitement,Immobility/stupor,Mutism,Staring,Posturing/catalepsy,Echopraxia/echolalia,Stereotypy,Rigidity,Negativism | 99.8 | 99.3 | 100.0 |
| 9 | Excitement,Immobility/stupor,Mutism,Staring,Posturing/catalepsy,Echopraxia/echolalia,Stereotypy,Verbigeration,Negativism | 99.8 | 99.3 | 100.0 |
| 9 | Excitement,Immobility/stupor,Mutism,Staring,Posturing/catalepsy,Echopraxia/echolalia,Stereotypy,Verbigeration,Rigidity | 99.8 | 99.3 | 100.0 |
| 9 | Excitement,Immobility/stupor,Mutism,Staring,Posturing/catalepsy,Echopraxia/echolalia,Verbigeration,Rigidity,Waxy flexibility | 99.8 | 99.3 | 100.0 |
| 9 | Excitement,Immobility/stupor,Mutism,Staring,Posturing/catalepsy,Grimacing,Echopraxia/echolalia,Negativism,Waxy flexibility | 99.8 | 99.3 | 100.0 |
| 9 | Excitement,Immobility/stupor,Mutism,Staring,Posturing/catalepsy,Grimacing,Echopraxia/echolalia,Rigidity,Negativism | 99.8 | 99.3 | 100.0 |
| 9 | Excitement,Immobility/stupor,Mutism,Staring,Posturing/catalepsy,Grimacing,Echopraxia/echolalia,Verbigeration,Negativism | 99.8 | 99.3 | 100.0 |
| 9 | Excitement,Immobility/stupor,Mutism,Staring,Posturing/catalepsy,Grimacing,Echopraxia/echolalia,Verbigeration,Rigidity | 99.8 | 99.3 | 100.0 |
| 9 | Excitement,Immobility/stupor,Mutism,Staring,Posturing/catalepsy,Grimacing,Mannerisms,Negativism,Withdrawal | 99.8 | 99.3 | 100.0 |
| 9 | Excitement,Immobility/stupor,Mutism,Staring,Posturing/catalepsy,Grimacing,Mannerisms,Rigidity,Withdrawal | 99.8 | 99.3 | 100.0 |
| 9 | Excitement,Immobility/stupor,Mutism,Staring,Posturing/catalepsy,Grimacing,Mannerisms,Verbigeration,Negativism | 99.8 | 99.3 | 100.0 |
| 9 | Excitement,Immobility/stupor,Mutism,Staring,Posturing/catalepsy,Grimacing,Mannerisms,Verbigeration,Rigidity | 99.8 | 99.3 | 100.0 |
| 9 | Excitement,Immobility/stupor,Mutism,Staring,Posturing/catalepsy,Grimacing,Stereotypy,Negativism,Withdrawal | 99.8 | 99.3 | 100.0 |
| 9 | Excitement,Immobility/stupor,Mutism,Staring,Posturing/catalepsy,Grimacing,Stereotypy,Rigidity,Withdrawal | 99.8 | 99.3 | 100.0 |
| 9 | Excitement,Immobility/stupor,Mutism,Staring,Posturing/catalepsy,Grimacing,Stereotypy,Verbigeration,Negativism | 99.8 | 99.3 | 100.0 |
| 9 | Excitement,Immobility/stupor,Mutism,Staring,Posturing/catalepsy,Grimacing,Stereotypy,Verbigeration,Rigidity | 99.8 | 99.3 | 100.0 |
| 9 | Excitement,Immobility/stupor,Mutism,Staring,Posturing/catalepsy,Grimacing,Verbigeration,Negativism,Waxy flexibility | 99.8 | 99.3 | 100.0 |
| 9 | Excitement,Immobility/stupor,Mutism,Staring,Posturing/catalepsy,Grimacing,Verbigeration,Negativism,Withdrawal | 99.8 | 99.3 | 100.0 |
| 9 | Excitement,Immobility/stupor,Mutism,Staring,Posturing/catalepsy,Grimacing,Verbigeration,Rigidity,Negativism | 99.8 | 99.3 | 100.0 |
| 9 | Excitement,Immobility/stupor,Mutism,Staring,Posturing/catalepsy,Grimacing,Verbigeration,Rigidity,Withdrawal | 99.8 | 99.3 | 100.0 |
| 9 | Excitement,Immobility/stupor,Mutism,Staring,Posturing/catalepsy,Mannerisms,Negativism,Waxy flexibility,Withdrawal | 99.8 | 99.3 | 100.0 |
| 9 | Excitement,Immobility/stupor,Mutism,Staring,Posturing/catalepsy,Mannerisms,Rigidity,Negativism,Withdrawal | 99.8 | 99.3 | 100.0 |
| 9 | Excitement,Immobility/stupor,Mutism,Staring,Posturing/catalepsy,Mannerisms,Rigidity,Waxy flexibility,Withdrawal | 99.8 | 99.3 | 100.0 |
| 9 | Excitement,Immobility/stupor,Mutism,Staring,Posturing/catalepsy,Mannerisms,Verbigeration,Rigidity,Waxy flexibility | 99.8 | 99.3 | 100.0 |
| 9 | Excitement,Immobility/stupor,Mutism,Staring,Posturing/catalepsy,Stereotypy,Mannerisms,Negativism,Withdrawal | 99.8 | 99.3 | 100.0 |
| 9 | Excitement,Immobility/stupor,Mutism,Staring,Posturing/catalepsy,Stereotypy,Mannerisms,Rigidity,Withdrawal | 99.8 | 99.3 | 100.0 |
| 9 | Excitement,Immobility/stupor,Mutism,Staring,Posturing/catalepsy,Stereotypy,Mannerisms,Verbigeration,Negativism | 99.8 | 99.3 | 100.0 |
| 9 | Excitement,Immobility/stupor,Mutism,Staring,Posturing/catalepsy,Stereotypy,Mannerisms,Verbigeration,Rigidity | 99.8 | 99.3 | 100.0 |
| 9 | Excitement,Immobility/stupor,Mutism,Staring,Posturing/catalepsy,Stereotypy,Negativism,Waxy flexibility,Withdrawal | 99.8 | 99.3 | 100.0 |
| 9 | Excitement,Immobility/stupor,Mutism,Staring,Posturing/catalepsy,Stereotypy,Rigidity,Negativism,Withdrawal | 99.8 | 99.3 | 100.0 |
| 9 | Excitement,Immobility/stupor,Mutism,Staring,Posturing/catalepsy,Stereotypy,Rigidity,Waxy flexibility,Withdrawal | 99.8 | 99.3 | 100.0 |
| 9 | Excitement,Immobility/stupor,Mutism,Staring,Posturing/catalepsy,Stereotypy,Verbigeration,Rigidity,Waxy flexibility | 99.8 | 99.3 | 100.0 |
| 9 | Excitement,Immobility/stupor,Mutism,Staring,Posturing/catalepsy,Stereotypy,Verbigeration,Rigidity,Withdrawal | 99.8 | 99.3 | 100.0 |
| 9 | Excitement,Immobility/stupor,Mutism,Staring,Posturing/catalepsy,Verbigeration,Negativism,Waxy flexibility,Withdrawal | 99.8 | 99.3 | 100.0 |
| 9 | Excitement,Immobility/stupor,Mutism,Staring,Posturing/catalepsy,Verbigeration,Rigidity,Negativism,Waxy flexibility | 99.8 | 99.3 | 100.0 |
| 9 | Excitement,Immobility/stupor,Mutism,Staring,Posturing/catalepsy,Verbigeration,Rigidity,Negativism,Withdrawal | 99.8 | 99.3 | 100.0 |
| 9 | Excitement,Immobility/stupor,Mutism,Staring,Posturing/catalepsy,Verbigeration,Rigidity,Waxy flexibility,Withdrawal | 99.8 | 99.3 | 100.0 |
| 9 | Excitement,Immobility/stupor,Mutism,Staring,Stereotypy,Mannerisms,Rigidity,Waxy flexibility,Withdrawal | 99.8 | 99.3 | 100.0 |
| 9 | Excitement,Immobility/stupor,Mutism,Staring,Stereotypy,Mannerisms,Verbigeration,Rigidity,Negativism | 99.8 | 99.3 | 100.0 |
| 9 | Excitement,Immobility/stupor,Mutism,Staring,Stereotypy,Mannerisms,Verbigeration,Rigidity,Waxy flexibility | 99.8 | 99.3 | 100.0 |
| 9 | Excitement,Immobility/stupor,Mutism,Staring,Stereotypy,Mannerisms,Verbigeration,Rigidity,Withdrawal | 99.8 | 99.3 | 100.0 |
| 9 | Excitement,Immobility/stupor,Mutism,Staring,Stereotypy,Rigidity,Negativism,Waxy flexibility,Withdrawal | 99.8 | 99.3 | 100.0 |
| 9 | Excitement,Immobility/stupor,Staring,Echopraxia/echolalia,Mannerisms,Rigidity,Negativism,Waxy flexibility,Withdrawal | 99.8 | 99.3 | 100.0 |
| 9 | Excitement,Immobility/stupor,Staring,Echopraxia/echolalia,Mannerisms,Verbigeration,Rigidity,Negativism,Withdrawal | 99.8 | 99.3 | 100.0 |
| 9 | Excitement,Immobility/stupor,Staring,Echopraxia/echolalia,Stereotypy,Mannerisms,Rigidity,Waxy flexibility,Withdrawal | 99.8 | 99.3 | 100.0 |
| 9 | Excitement,Immobility/stupor,Staring,Echopraxia/echolalia,Stereotypy,Rigidity,Negativism,Waxy flexibility,Withdrawal | 99.8 | 99.3 | 100.0 |
| 9 | Excitement,Immobility/stupor,Staring,Echopraxia/echolalia,Stereotypy,Verbigeration,Rigidity,Negativism,Withdrawal | 99.8 | 99.3 | 100.0 |
| 9 | Excitement,Immobility/stupor,Staring,Grimacing,Echopraxia/echolalia,Mannerisms,Rigidity,Negativism,Withdrawal | 99.8 | 99.3 | 100.0 |
| 9 | Excitement,Immobility/stupor,Staring,Grimacing,Echopraxia/echolalia,Stereotypy,Rigidity,Negativism,Withdrawal | 99.8 | 99.3 | 100.0 |
| 9 | Excitement,Immobility/stupor,Staring,Grimacing,Mannerisms,Verbigeration,Rigidity,Negativism,Withdrawal | 99.8 | 99.3 | 100.0 |
| 9 | Excitement,Immobility/stupor,Staring,Grimacing,Stereotypy,Verbigeration,Rigidity,Negativism,Withdrawal | 99.8 | 99.3 | 100.0 |
| 9 | Excitement,Immobility/stupor,Staring,Mannerisms,Verbigeration,Rigidity,Negativism,Waxy flexibility,Withdrawal | 99.8 | 99.3 | 100.0 |
| 9 | Excitement,Immobility/stupor,Staring,Posturing/catalepsy,Echopraxia/echolalia,Mannerisms,Negativism,Waxy flexibility,Withdrawal | 99.8 | 99.3 | 100.0 |
| 9 | Excitement,Immobility/stupor,Staring,Posturing/catalepsy,Echopraxia/echolalia,Mannerisms,Rigidity,Waxy flexibility,Withdrawal | 99.8 | 99.3 | 100.0 |
| 9 | Excitement,Immobility/stupor,Staring,Posturing/catalepsy,Echopraxia/echolalia,Mannerisms,Verbigeration,Negativism,Withdrawal | 99.8 | 99.3 | 100.0 |
| 9 | Excitement,Immobility/stupor,Staring,Posturing/catalepsy,Echopraxia/echolalia,Mannerisms,Verbigeration,Rigidity,Negativism | 99.8 | 99.3 | 100.0 |
| 9 | Excitement,Immobility/stupor,Staring,Posturing/catalepsy,Echopraxia/echolalia,Mannerisms,Verbigeration,Rigidity,Withdrawal | 99.8 | 99.3 | 100.0 |
| 9 | Excitement,Immobility/stupor,Staring,Posturing/catalepsy,Echopraxia/echolalia,Rigidity,Negativism,Waxy flexibility,Withdrawal | 99.8 | 99.3 | 100.0 |
| 9 | Excitement,Immobility/stupor,Staring,Posturing/catalepsy,Echopraxia/echolalia,Stereotypy,Mannerisms,Negativism,Withdrawal | 99.8 | 99.3 | 100.0 |
| 9 | Excitement,Immobility/stupor,Staring,Posturing/catalepsy,Echopraxia/echolalia,Stereotypy,Mannerisms,Rigidity,Withdrawal | 99.8 | 99.3 | 100.0 |
| 9 | Excitement,Immobility/stupor,Staring,Posturing/catalepsy,Echopraxia/echolalia,Stereotypy,Rigidity,Negativism,Withdrawal | 99.8 | 99.3 | 100.0 |
| 9 | Excitement,Immobility/stupor,Staring,Posturing/catalepsy,Echopraxia/echolalia,Verbigeration,Rigidity,Negativism,Withdrawal | 99.8 | 99.3 | 100.0 |
| 9 | Excitement,Immobility/stupor,Staring,Posturing/catalepsy,Grimacing,Echopraxia/echolalia,Mannerisms,Negativism,Withdrawal | 99.8 | 99.3 | 100.0 |
| 9 | Excitement,Immobility/stupor,Staring,Posturing/catalepsy,Grimacing,Echopraxia/echolalia,Mannerisms,Rigidity,Withdrawal | 99.8 | 99.3 | 100.0 |
| 9 | Excitement,Immobility/stupor,Staring,Posturing/catalepsy,Grimacing,Echopraxia/echolalia,Rigidity,Negativism,Withdrawal | 99.8 | 99.3 | 100.0 |
| 9 | Excitement,Immobility/stupor,Staring,Posturing/catalepsy,Grimacing,Mannerisms,Rigidity,Negativism,Withdrawal | 99.8 | 99.3 | 100.0 |
| 9 | Excitement,Immobility/stupor,Staring,Posturing/catalepsy,Grimacing,Mannerisms,Verbigeration,Negativism,Withdrawal | 99.8 | 99.3 | 100.0 |
| 9 | Excitement,Immobility/stupor,Staring,Posturing/catalepsy,Grimacing,Mannerisms,Verbigeration,Rigidity,Negativism | 99.8 | 99.3 | 100.0 |
| 9 | Excitement,Immobility/stupor,Staring,Posturing/catalepsy,Grimacing,Mannerisms,Verbigeration,Rigidity,Withdrawal | 99.8 | 99.3 | 100.0 |
| 9 | Excitement,Immobility/stupor,Staring,Posturing/catalepsy,Mannerisms,Rigidity,Negativism,Waxy flexibility,Withdrawal | 99.8 | 99.3 | 100.0 |
| 9 | Excitement,Immobility/stupor,Staring,Posturing/catalepsy,Mannerisms,Verbigeration,Negativism,Waxy flexibility,Withdrawal | 99.8 | 99.3 | 100.0 |
| 9 | Excitement,Immobility/stupor,Staring,Posturing/catalepsy,Mannerisms,Verbigeration,Rigidity,Negativism,Waxy flexibility | 99.8 | 99.3 | 100.0 |
| 9 | Excitement,Immobility/stupor,Staring,Posturing/catalepsy,Mannerisms,Verbigeration,Rigidity,Waxy flexibility,Withdrawal | 99.8 | 99.3 | 100.0 |
| 9 | Excitement,Immobility/stupor,Staring,Posturing/catalepsy,Stereotypy,Mannerisms,Rigidity,Negativism,Withdrawal | 99.8 | 99.3 | 100.0 |
| 9 | Excitement,Immobility/stupor,Staring,Posturing/catalepsy,Stereotypy,Mannerisms,Verbigeration,Negativism,Withdrawal | 99.8 | 99.3 | 100.0 |
| 9 | Excitement,Immobility/stupor,Staring,Posturing/catalepsy,Stereotypy,Mannerisms,Verbigeration,Rigidity,Negativism | 99.8 | 99.3 | 100.0 |
| 9 | Excitement,Immobility/stupor,Staring,Posturing/catalepsy,Stereotypy,Mannerisms,Verbigeration,Rigidity,Withdrawal | 99.8 | 99.3 | 100.0 |
| 9 | Excitement,Immobility/stupor,Staring,Posturing/catalepsy,Stereotypy,Verbigeration,Rigidity,Negativism,Withdrawal | 99.8 | 99.3 | 100.0 |
| 9 | Excitement,Immobility/stupor,Staring,Stereotypy,Mannerisms,Rigidity,Negativism,Waxy flexibility,Withdrawal | 99.8 | 99.3 | 100.0 |
| 9 | Excitement,Immobility/stupor,Staring,Stereotypy,Mannerisms,Verbigeration,Rigidity,Negativism,Waxy flexibility | 99.8 | 99.3 | 100.0 |
| 9 | Excitement,Immobility/stupor,Staring,Stereotypy,Mannerisms,Verbigeration,Rigidity,Waxy flexibility,Withdrawal | 99.8 | 99.3 | 100.0 |
| 9 | Excitement,Immobility/stupor,Staring,Stereotypy,Verbigeration,Rigidity,Negativism,Waxy flexibility,Withdrawal | 99.8 | 99.3 | 100.0 |
| 9 | Excitement,Mutism,Staring,Echopraxia/echolalia,Mannerisms,Rigidity,Negativism,Waxy flexibility,Withdrawal | 99.8 | 99.3 | 100.0 |
| 9 | Excitement,Mutism,Staring,Echopraxia/echolalia,Mannerisms,Verbigeration,Rigidity,Negativism,Waxy flexibility | 99.8 | 99.3 | 100.0 |
| 9 | Excitement,Mutism,Staring,Echopraxia/echolalia,Mannerisms,Verbigeration,Rigidity,Negativism,Withdrawal | 99.8 | 99.3 | 100.0 |
| 9 | Excitement,Mutism,Staring,Echopraxia/echolalia,Stereotypy,Mannerisms,Rigidity,Negativism,Waxy flexibility | 99.8 | 99.3 | 100.0 |
| 9 | Excitement,Mutism,Staring,Echopraxia/echolalia,Stereotypy,Mannerisms,Verbigeration,Rigidity,Negativism | 99.8 | 99.3 | 100.0 |
| 9 | Excitement,Mutism,Staring,Echopraxia/echolalia,Stereotypy,Mannerisms,Verbigeration,Rigidity,Withdrawal | 99.8 | 99.3 | 100.0 |
| 9 | Excitement,Mutism,Staring,Echopraxia/echolalia,Verbigeration,Rigidity,Negativism,Waxy flexibility,Withdrawal | 99.8 | 99.3 | 100.0 |
| 9 | Excitement,Mutism,Staring,Grimacing,Echopraxia/echolalia,Mannerisms,Rigidity,Negativism,Withdrawal | 99.8 | 99.3 | 100.0 |
| 9 | Excitement,Mutism,Staring,Grimacing,Echopraxia/echolalia,Mannerisms,Rigidity,Waxy flexibility,Withdrawal | 99.8 | 99.3 | 100.0 |
| 9 | Excitement,Mutism,Staring,Grimacing,Echopraxia/echolalia,Rigidity,Negativism,Waxy flexibility,Withdrawal | 99.8 | 99.3 | 100.0 |
| 9 | Excitement,Mutism,Staring,Grimacing,Echopraxia/echolalia,Stereotypy,Mannerisms,Rigidity,Withdrawal | 99.8 | 99.3 | 100.0 |
| 9 | Excitement,Mutism,Staring,Grimacing,Echopraxia/echolalia,Stereotypy,Rigidity,Negativism,Waxy flexibility | 99.8 | 99.3 | 100.0 |
| 9 | Excitement,Mutism,Staring,Grimacing,Echopraxia/echolalia,Stereotypy,Verbigeration,Rigidity,Negativism | 99.8 | 99.3 | 100.0 |
| 9 | Excitement,Mutism,Staring,Grimacing,Echopraxia/echolalia,Stereotypy,Verbigeration,Rigidity,Waxy flexibility | 99.8 | 99.3 | 100.0 |
| 9 | Excitement,Mutism,Staring,Grimacing,Echopraxia/echolalia,Stereotypy,Verbigeration,Rigidity,Withdrawal | 99.8 | 99.3 | 100.0 |
| 9 | Excitement,Mutism,Staring,Grimacing,Echopraxia/echolalia,Verbigeration,Rigidity,Negativism,Waxy flexibility | 99.8 | 99.3 | 100.0 |
| 9 | Excitement,Mutism,Staring,Grimacing,Echopraxia/echolalia,Verbigeration,Rigidity,Negativism,Withdrawal | 99.8 | 99.3 | 100.0 |
| 9 | Excitement,Mutism,Staring,Grimacing,Echopraxia/echolalia,Verbigeration,Rigidity,Waxy flexibility,Withdrawal | 99.8 | 99.3 | 100.0 |
| 9 | Excitement,Mutism,Staring,Grimacing,Mannerisms,Verbigeration,Rigidity,Negativism,Withdrawal | 99.8 | 99.3 | 100.0 |
| 9 | Excitement,Mutism,Staring,Grimacing,Mannerisms,Verbigeration,Rigidity,Waxy flexibility,Withdrawal | 99.8 | 99.3 | 100.0 |
| 9 | Excitement,Mutism,Staring,Grimacing,Stereotypy,Mannerisms,Rigidity,Waxy flexibility,Withdrawal | 99.8 | 99.3 | 100.0 |
| 9 | Excitement,Mutism,Staring,Grimacing,Stereotypy,Mannerisms,Verbigeration,Rigidity,Withdrawal | 99.8 | 99.3 | 100.0 |
| 9 | Excitement,Mutism,Staring,Grimacing,Stereotypy,Rigidity,Negativism,Waxy flexibility,Withdrawal | 99.8 | 99.3 | 100.0 |
| 9 | Excitement,Mutism,Staring,Grimacing,Stereotypy,Verbigeration,Rigidity,Negativism,Waxy flexibility | 99.8 | 99.3 | 100.0 |
| 9 | Excitement,Mutism,Staring,Mannerisms,Verbigeration,Rigidity,Negativism,Waxy flexibility,Withdrawal | 99.8 | 99.3 | 100.0 |
| 9 | Excitement,Mutism,Staring,Posturing/catalepsy,Echopraxia/echolalia,Mannerisms,Rigidity,Negativism,Waxy flexibility | 99.8 | 99.3 | 100.0 |
| 9 | Excitement,Mutism,Staring,Posturing/catalepsy,Echopraxia/echolalia,Mannerisms,Verbigeration,Rigidity,Waxy flexibility | 99.8 | 99.3 | 100.0 |
| 9 | Excitement,Mutism,Staring,Posturing/catalepsy,Echopraxia/echolalia,Stereotypy,Mannerisms,Negativism,Waxy flexibility | 99.8 | 99.3 | 100.0 |
| 9 | Excitement,Mutism,Staring,Posturing/catalepsy,Echopraxia/echolalia,Stereotypy,Mannerisms,Rigidity,Negativism | 99.8 | 99.3 | 100.0 |
| 9 | Excitement,Mutism,Staring,Posturing/catalepsy,Echopraxia/echolalia,Stereotypy,Mannerisms,Verbigeration,Negativism | 99.8 | 99.3 | 100.0 |
| 9 | Excitement,Mutism,Staring,Posturing/catalepsy,Echopraxia/echolalia,Stereotypy,Mannerisms,Verbigeration,Rigidity | 99.8 | 99.3 | 100.0 |
| 9 | Excitement,Mutism,Staring,Posturing/catalepsy,Echopraxia/echolalia,Stereotypy,Rigidity,Negativism,Waxy flexibility | 99.8 | 99.3 | 100.0 |
| 9 | Excitement,Mutism,Staring,Posturing/catalepsy,Echopraxia/echolalia,Stereotypy,Verbigeration,Rigidity,Waxy flexibility | 99.8 | 99.3 | 100.0 |
| 9 | Excitement,Mutism,Staring,Posturing/catalepsy,Grimacing,Echopraxia/echolalia,Mannerisms,Negativism,Waxy flexibility | 99.8 | 99.3 | 100.0 |
| 9 | Excitement,Mutism,Staring,Posturing/catalepsy,Grimacing,Echopraxia/echolalia,Mannerisms,Rigidity,Negativism | 99.8 | 99.3 | 100.0 |
| 9 | Excitement,Mutism,Staring,Posturing/catalepsy,Grimacing,Echopraxia/echolalia,Mannerisms,Verbigeration,Negativism | 99.8 | 99.3 | 100.0 |
| 9 | Excitement,Mutism,Staring,Posturing/catalepsy,Grimacing,Echopraxia/echolalia,Mannerisms,Verbigeration,Rigidity | 99.8 | 99.3 | 100.0 |
| 9 | Excitement,Mutism,Staring,Posturing/catalepsy,Grimacing,Echopraxia/echolalia,Rigidity,Negativism,Waxy flexibility | 99.8 | 99.3 | 100.0 |
| 9 | Excitement,Mutism,Staring,Posturing/catalepsy,Grimacing,Echopraxia/echolalia,Stereotypy,Negativism,Waxy flexibility | 99.8 | 99.3 | 100.0 |
| 9 | Excitement,Mutism,Staring,Posturing/catalepsy,Grimacing,Echopraxia/echolalia,Stereotypy,Rigidity,Negativism | 99.8 | 99.3 | 100.0 |
| 9 | Excitement,Mutism,Staring,Posturing/catalepsy,Grimacing,Echopraxia/echolalia,Stereotypy,Verbigeration,Negativism | 99.8 | 99.3 | 100.0 |
| 9 | Excitement,Mutism,Staring,Posturing/catalepsy,Grimacing,Echopraxia/echolalia,Stereotypy,Verbigeration,Rigidity | 99.8 | 99.3 | 100.0 |
| 9 | Excitement,Mutism,Staring,Posturing/catalepsy,Grimacing,Echopraxia/echolalia,Verbigeration,Rigidity,Waxy flexibility | 99.8 | 99.3 | 100.0 |
| 9 | Excitement,Mutism,Staring,Posturing/catalepsy,Grimacing,Mannerisms,Negativism,Waxy flexibility,Withdrawal | 99.8 | 99.3 | 100.0 |
| 9 | Excitement,Mutism,Staring,Posturing/catalepsy,Grimacing,Mannerisms,Rigidity,Negativism,Withdrawal | 99.8 | 99.3 | 100.0 |
| 9 | Excitement,Mutism,Staring,Posturing/catalepsy,Grimacing,Mannerisms,Rigidity,Waxy flexibility,Withdrawal | 99.8 | 99.3 | 100.0 |
| 9 | Excitement,Mutism,Staring,Posturing/catalepsy,Grimacing,Mannerisms,Verbigeration,Negativism,Waxy flexibility | 99.8 | 99.3 | 100.0 |
| 9 | Excitement,Mutism,Staring,Posturing/catalepsy,Grimacing,Mannerisms,Verbigeration,Rigidity,Negativism | 99.8 | 99.3 | 100.0 |
| 9 | Excitement,Mutism,Staring,Posturing/catalepsy,Grimacing,Stereotypy,Mannerisms,Negativism,Withdrawal | 99.8 | 99.3 | 100.0 |
| 9 | Excitement,Mutism,Staring,Posturing/catalepsy,Grimacing,Stereotypy,Mannerisms,Rigidity,Withdrawal | 99.8 | 99.3 | 100.0 |
| 9 | Excitement,Mutism,Staring,Posturing/catalepsy,Grimacing,Stereotypy,Negativism,Waxy flexibility,Withdrawal | 99.8 | 99.3 | 100.0 |
| 9 | Excitement,Mutism,Staring,Posturing/catalepsy,Grimacing,Stereotypy,Rigidity,Negativism,Withdrawal | 99.8 | 99.3 | 100.0 |
| 9 | Excitement,Mutism,Staring,Posturing/catalepsy,Grimacing,Stereotypy,Rigidity,Waxy flexibility,Withdrawal | 99.8 | 99.3 | 100.0 |
| 9 | Excitement,Mutism,Staring,Posturing/catalepsy,Grimacing,Stereotypy,Verbigeration,Negativism,Waxy flexibility | 99.8 | 99.3 | 100.0 |
| 9 | Excitement,Mutism,Staring,Posturing/catalepsy,Grimacing,Stereotypy,Verbigeration,Rigidity,Negativism | 99.8 | 99.3 | 100.0 |
| 9 | Excitement,Mutism,Staring,Posturing/catalepsy,Grimacing,Verbigeration,Negativism,Waxy flexibility,Withdrawal | 99.8 | 99.3 | 100.0 |
| 9 | Excitement,Mutism,Staring,Posturing/catalepsy,Grimacing,Verbigeration,Rigidity,Negativism,Withdrawal | 99.8 | 99.3 | 100.0 |
| 9 | Excitement,Mutism,Staring,Posturing/catalepsy,Grimacing,Verbigeration,Rigidity,Waxy flexibility,Withdrawal | 99.8 | 99.3 | 100.0 |
| 9 | Excitement,Mutism,Staring,Posturing/catalepsy,Mannerisms,Rigidity,Negativism,Waxy flexibility,Withdrawal | 99.8 | 99.3 | 100.0 |
| 9 | Excitement,Mutism,Staring,Posturing/catalepsy,Mannerisms,Verbigeration,Rigidity,Negativism,Waxy flexibility | 99.8 | 99.3 | 100.0 |
| 9 | Excitement,Mutism,Staring,Posturing/catalepsy,Stereotypy,Mannerisms,Negativism,Waxy flexibility,Withdrawal | 99.8 | 99.3 | 100.0 |
| 9 | Excitement,Mutism,Staring,Posturing/catalepsy,Stereotypy,Mannerisms,Rigidity,Negativism,Withdrawal | 99.8 | 99.3 | 100.0 |
| 9 | Excitement,Mutism,Staring,Posturing/catalepsy,Stereotypy,Mannerisms,Rigidity,Waxy flexibility,Withdrawal | 99.8 | 99.3 | 100.0 |
| 9 | Excitement,Mutism,Staring,Posturing/catalepsy,Stereotypy,Mannerisms,Verbigeration,Negativism,Waxy flexibility | 99.8 | 99.3 | 100.0 |
| 9 | Excitement,Mutism,Staring,Posturing/catalepsy,Stereotypy,Mannerisms,Verbigeration,Rigidity,Negativism | 99.8 | 99.3 | 100.0 |
| 9 | Excitement,Mutism,Staring,Posturing/catalepsy,Stereotypy,Rigidity,Negativism,Waxy flexibility,Withdrawal | 99.8 | 99.3 | 100.0 |
| 9 | Excitement,Mutism,Staring,Posturing/catalepsy,Stereotypy,Verbigeration,Rigidity,Negativism,Waxy flexibility | 99.8 | 99.3 | 100.0 |
| 9 | Excitement,Mutism,Staring,Posturing/catalepsy,Verbigeration,Rigidity,Negativism,Waxy flexibility,Withdrawal | 99.8 | 99.3 | 100.0 |
| 9 | Excitement,Mutism,Staring,Stereotypy,Mannerisms,Rigidity,Negativism,Waxy flexibility,Withdrawal | 99.8 | 99.3 | 100.0 |
| 9 | Excitement,Mutism,Staring,Stereotypy,Mannerisms,Verbigeration,Rigidity,Negativism,Waxy flexibility | 99.8 | 99.3 | 100.0 |
| 9 | Immobility/stupor,Mutism,Staring,Echopraxia/echolalia,Stereotypy,Mannerisms,Rigidity,Negativism,Withdrawal | 99.8 | 99.3 | 100.0 |
| 9 | Immobility/stupor,Mutism,Staring,Posturing/catalepsy,Grimacing,Echopraxia/echolalia,Mannerisms,Negativism,Withdrawal | 99.8 | 99.3 | 100.0 |
| 9 | Immobility/stupor,Mutism,Staring,Posturing/catalepsy,Grimacing,Stereotypy,Mannerisms,Negativism,Withdrawal | 99.8 | 99.3 | 100.0 |
| 9 | Immobility/stupor,Mutism,Staring,Stereotypy,Mannerisms,Verbigeration,Rigidity,Negativism,Withdrawal | 99.8 | 99.3 | 100.0 |
| 9 | Immobility/stupor,Staring,Echopraxia/echolalia,Stereotypy,Mannerisms,Rigidity,Negativism,Waxy flexibility,Withdrawal | 99.8 | 99.3 | 100.0 |
| 9 | Immobility/stupor,Staring,Echopraxia/echolalia,Stereotypy,Mannerisms,Verbigeration,Rigidity,Negativism,Withdrawal | 99.8 | 99.3 | 100.0 |
| 9 | Immobility/stupor,Staring,Grimacing,Stereotypy,Mannerisms,Rigidity,Negativism,Waxy flexibility,Withdrawal | 99.8 | 99.3 | 100.0 |
| 9 | Immobility/stupor,Staring,Posturing/catalepsy,Echopraxia/echolalia,Stereotypy,Mannerisms,Rigidity,Negativism,Withdrawal | 99.8 | 99.3 | 100.0 |
| 9 | Immobility/stupor,Staring,Posturing/catalepsy,Grimacing,Echopraxia/echolalia,Mannerisms,Rigidity,Negativism,Withdrawal | 99.8 | 99.3 | 100.0 |
| 9 | Immobility/stupor,Staring,Posturing/catalepsy,Grimacing,Echopraxia/echolalia,Stereotypy,Mannerisms,Negativism,Withdrawal | 99.8 | 99.3 | 100.0 |
| 9 | Immobility/stupor,Staring,Posturing/catalepsy,Grimacing,Stereotypy,Mannerisms,Rigidity,Negativism,Withdrawal | 99.8 | 99.3 | 100.0 |
| 9 | Immobility/stupor,Staring,Posturing/catalepsy,Grimacing,Stereotypy,Mannerisms,Verbigeration,Negativism,Withdrawal | 99.8 | 99.3 | 100.0 |
| 9 | Immobility/stupor,Staring,Posturing/catalepsy,Stereotypy,Mannerisms,Verbigeration,Rigidity,Negativism,Withdrawal | 99.8 | 99.3 | 100.0 |
| 9 | Immobility/stupor,Staring,Stereotypy,Mannerisms,Verbigeration,Rigidity,Negativism,Waxy flexibility,Withdrawal | 99.8 | 99.3 | 100.0 |
| 9 | Mutism,Staring,Echopraxia/echolalia,Stereotypy,Mannerisms,Rigidity,Negativism,Waxy flexibility,Withdrawal | 99.8 | 99.3 | 100.0 |
| 9 | Mutism,Staring,Echopraxia/echolalia,Stereotypy,Mannerisms,Verbigeration,Rigidity,Negativism,Withdrawal | 99.8 | 99.3 | 100.0 |
| 9 | Mutism,Staring,Grimacing,Stereotypy,Mannerisms,Rigidity,Negativism,Waxy flexibility,Withdrawal | 99.8 | 99.3 | 100.0 |
| 9 | Mutism,Staring,Posturing/catalepsy,Echopraxia/echolalia,Stereotypy,Mannerisms,Rigidity,Negativism,Withdrawal | 99.8 | 99.3 | 100.0 |
| 9 | Mutism,Staring,Posturing/catalepsy,Grimacing,Echopraxia/echolalia,Mannerisms,Negativism,Waxy flexibility,Withdrawal | 99.8 | 99.3 | 100.0 |
| 9 | Mutism,Staring,Posturing/catalepsy,Grimacing,Echopraxia/echolalia,Mannerisms,Rigidity,Negativism,Withdrawal | 99.8 | 99.3 | 100.0 |
| 9 | Mutism,Staring,Posturing/catalepsy,Grimacing,Echopraxia/echolalia,Mannerisms,Verbigeration,Negativism,Withdrawal | 99.8 | 99.3 | 100.0 |
| 9 | Mutism,Staring,Posturing/catalepsy,Grimacing,Stereotypy,Mannerisms,Negativism,Waxy flexibility,Withdrawal | 99.8 | 99.3 | 100.0 |
| 9 | Mutism,Staring,Posturing/catalepsy,Grimacing,Stereotypy,Mannerisms,Rigidity,Negativism,Withdrawal | 99.8 | 99.3 | 100.0 |
| 9 | Mutism,Staring,Posturing/catalepsy,Stereotypy,Mannerisms,Verbigeration,Rigidity,Negativism,Withdrawal | 99.8 | 99.3 | 100.0 |
| 9 | Mutism,Staring,Stereotypy,Mannerisms,Verbigeration,Rigidity,Negativism,Waxy flexibility,Withdrawal | 99.8 | 99.3 | 100.0 |
| 9 | Excitement,Immobility/stupor,Echopraxia/echolalia,Stereotypy,Mannerisms,Verbigeration,Rigidity,Negativism,Withdrawal | 99.6 | 98.9 | 100.0 |
| 9 | Excitement,Immobility/stupor,Mutism,Echopraxia/echolalia,Stereotypy,Verbigeration,Rigidity,Negativism,Waxy flexibility | 99.6 | 98.9 | 100.0 |
| 9 | Excitement,Immobility/stupor,Mutism,Grimacing,Echopraxia/echolalia,Stereotypy,Rigidity,Negativism,Withdrawal | 99.6 | 98.9 | 100.0 |
| 9 | Excitement,Immobility/stupor,Mutism,Grimacing,Stereotypy,Verbigeration,Rigidity,Negativism,Withdrawal | 99.6 | 98.9 | 100.0 |
| 9 | Excitement,Immobility/stupor,Mutism,Posturing/catalepsy,Echopraxia/echolalia,Stereotypy,Rigidity,Negativism,Withdrawal | 99.6 | 98.9 | 100.0 |
| 9 | Excitement,Immobility/stupor,Mutism,Posturing/catalepsy,Echopraxia/echolalia,Stereotypy,Verbigeration,Rigidity,Withdrawal | 99.6 | 98.9 | 100.0 |
| 9 | Excitement,Immobility/stupor,Mutism,Posturing/catalepsy,Echopraxia/echolalia,Verbigeration,Rigidity,Negativism,Waxy flexibility | 99.6 | 98.9 | 100.0 |
| 9 | Excitement,Immobility/stupor,Mutism,Posturing/catalepsy,Grimacing,Echopraxia/echolalia,Verbigeration,Rigidity,Withdrawal | 99.6 | 98.9 | 100.0 |
| 9 | Excitement,Immobility/stupor,Mutism,Posturing/catalepsy,Grimacing,Mannerisms,Verbigeration,Rigidity,Negativism | 99.6 | 98.9 | 100.0 |
| 9 | Excitement,Immobility/stupor,Mutism,Posturing/catalepsy,Grimacing,Stereotypy,Verbigeration,Rigidity,Negativism | 99.6 | 98.9 | 100.0 |
| 9 | Excitement,Immobility/stupor,Mutism,Posturing/catalepsy,Grimacing,Verbigeration,Rigidity,Negativism,Withdrawal | 99.6 | 98.9 | 100.0 |
| 9 | Excitement,Immobility/stupor,Mutism,Posturing/catalepsy,Mannerisms,Verbigeration,Rigidity,Negativism,Withdrawal | 99.6 | 98.9 | 100.0 |
| 9 | Excitement,Immobility/stupor,Mutism,Posturing/catalepsy,Stereotypy,Mannerisms,Verbigeration,Rigidity,Negativism | 99.6 | 98.9 | 100.0 |
| 9 | Excitement,Immobility/stupor,Mutism,Posturing/catalepsy,Stereotypy,Verbigeration,Rigidity,Negativism,Waxy flexibility | 99.6 | 98.9 | 100.0 |
| 9 | Excitement,Immobility/stupor,Mutism,Staring,Echopraxia/echolalia,Mannerisms,Rigidity,Negativism,Waxy flexibility | 99.6 | 98.9 | 100.0 |
| 9 | Excitement,Immobility/stupor,Mutism,Staring,Echopraxia/echolalia,Mannerisms,Verbigeration,Rigidity,Waxy flexibility | 99.6 | 98.9 | 100.0 |
| 9 | Excitement,Immobility/stupor,Mutism,Staring,Echopraxia/echolalia,Mannerisms,Verbigeration,Rigidity,Withdrawal | 99.6 | 98.9 | 100.0 |
| 9 | Excitement,Immobility/stupor,Mutism,Staring,Echopraxia/echolalia,Stereotypy,Mannerisms,Rigidity,Waxy flexibility | 99.6 | 98.9 | 100.0 |
| 9 | Excitement,Immobility/stupor,Mutism,Staring,Echopraxia/echolalia,Stereotypy,Verbigeration,Negativism,Waxy flexibility | 99.6 | 98.9 | 100.0 |
| 9 | Excitement,Immobility/stupor,Mutism,Staring,Echopraxia/echolalia,Stereotypy,Verbigeration,Negativism,Withdrawal | 99.6 | 98.9 | 100.0 |
| 9 | Excitement,Immobility/stupor,Mutism,Staring,Grimacing,Echopraxia/echolalia,Mannerisms,Rigidity,Withdrawal | 99.6 | 98.9 | 100.0 |
| 9 | Excitement,Immobility/stupor,Mutism,Staring,Grimacing,Echopraxia/echolalia,Stereotypy,Negativism,Withdrawal | 99.6 | 98.9 | 100.0 |
| 9 | Excitement,Immobility/stupor,Mutism,Staring,Grimacing,Echopraxia/echolalia,Stereotypy,Rigidity,Negativism | 99.6 | 98.9 | 100.0 |
| 9 | Excitement,Immobility/stupor,Mutism,Staring,Grimacing,Echopraxia/echolalia,Stereotypy,Rigidity,Waxy flexibility | 99.6 | 98.9 | 100.0 |
| 9 | Excitement,Immobility/stupor,Mutism,Staring,Grimacing,Echopraxia/echolalia,Verbigeration,Rigidity,Negativism | 99.6 | 98.9 | 100.0 |
| 9 | Excitement,Immobility/stupor,Mutism,Staring,Grimacing,Echopraxia/echolalia,Verbigeration,Rigidity,Waxy flexibility | 99.6 | 98.9 | 100.0 |
| 9 | Excitement,Immobility/stupor,Mutism,Staring,Grimacing,Echopraxia/echolalia,Verbigeration,Rigidity,Withdrawal | 99.6 | 98.9 | 100.0 |
| 9 | Excitement,Immobility/stupor,Mutism,Staring,Grimacing,Mannerisms,Rigidity,Waxy flexibility,Withdrawal | 99.6 | 98.9 | 100.0 |
| 9 | Excitement,Immobility/stupor,Mutism,Staring,Grimacing,Mannerisms,Verbigeration,Rigidity,Negativism | 99.6 | 98.9 | 100.0 |
| 9 | Excitement,Immobility/stupor,Mutism,Staring,Grimacing,Stereotypy,Rigidity,Negativism,Waxy flexibility | 99.6 | 98.9 | 100.0 |
| 9 | Excitement,Immobility/stupor,Mutism,Staring,Grimacing,Stereotypy,Rigidity,Negativism,Withdrawal | 99.6 | 98.9 | 100.0 |
| 9 | Excitement,Immobility/stupor,Mutism,Staring,Grimacing,Verbigeration,Rigidity,Negativism,Waxy flexibility | 99.6 | 98.9 | 100.0 |
| 9 | Excitement,Immobility/stupor,Mutism,Staring,Grimacing,Verbigeration,Rigidity,Negativism,Withdrawal | 99.6 | 98.9 | 100.0 |
| 9 | Excitement,Immobility/stupor,Mutism,Staring,Mannerisms,Rigidity,Negativism,Waxy flexibility,Withdrawal | 99.6 | 98.9 | 100.0 |
| 9 | Excitement,Immobility/stupor,Mutism,Staring,Posturing/catalepsy,Echopraxia/echolalia,Mannerisms,Rigidity,Waxy flexibility | 99.6 | 98.9 | 100.0 |
| 9 | Excitement,Immobility/stupor,Mutism,Staring,Posturing/catalepsy,Echopraxia/echolalia,Stereotypy,Mannerisms,Rigidity | 99.6 | 98.9 | 100.0 |
| 9 | Excitement,Immobility/stupor,Mutism,Staring,Posturing/catalepsy,Echopraxia/echolalia,Stereotypy,Rigidity,Waxy flexibility | 99.6 | 98.9 | 100.0 |
| 9 | Excitement,Immobility/stupor,Mutism,Staring,Posturing/catalepsy,Grimacing,Echopraxia/echolalia,Mannerisms,Negativism | 99.6 | 98.9 | 100.0 |
| 9 | Excitement,Immobility/stupor,Mutism,Staring,Posturing/catalepsy,Grimacing,Echopraxia/echolalia,Rigidity,Waxy flexibility | 99.6 | 98.9 | 100.0 |
| 9 | Excitement,Immobility/stupor,Mutism,Staring,Posturing/catalepsy,Grimacing,Echopraxia/echolalia,Stereotypy,Negativism | 99.6 | 98.9 | 100.0 |
| 9 | Excitement,Immobility/stupor,Mutism,Staring,Posturing/catalepsy,Grimacing,Echopraxia/echolalia,Stereotypy,Rigidity | 99.6 | 98.9 | 100.0 |
| 9 | Excitement,Immobility/stupor,Mutism,Staring,Posturing/catalepsy,Grimacing,Echopraxia/echolalia,Waxy flexibility,Withdrawal | 99.6 | 98.9 | 100.0 |
| 9 | Excitement,Immobility/stupor,Mutism,Staring,Posturing/catalepsy,Grimacing,Mannerisms,Negativism,Waxy flexibility | 99.6 | 98.9 | 100.0 |
| 9 | Excitement,Immobility/stupor,Mutism,Staring,Posturing/catalepsy,Grimacing,Mannerisms,Rigidity,Negativism | 99.6 | 98.9 | 100.0 |
| 9 | Excitement,Immobility/stupor,Mutism,Staring,Posturing/catalepsy,Grimacing,Rigidity,Negativism,Withdrawal | 99.6 | 98.9 | 100.0 |
| 9 | Excitement,Immobility/stupor,Mutism,Staring,Posturing/catalepsy,Grimacing,Rigidity,Waxy flexibility,Withdrawal | 99.6 | 98.9 | 100.0 |
| 9 | Excitement,Immobility/stupor,Mutism,Staring,Posturing/catalepsy,Grimacing,Verbigeration,Rigidity,Waxy flexibility | 99.6 | 98.9 | 100.0 |
| 9 | Excitement,Immobility/stupor,Mutism,Staring,Posturing/catalepsy,Mannerisms,Rigidity,Negativism,Waxy flexibility | 99.6 | 98.9 | 100.0 |
| 9 | Excitement,Immobility/stupor,Mutism,Staring,Posturing/catalepsy,Rigidity,Negativism,Waxy flexibility,Withdrawal | 99.6 | 98.9 | 100.0 |
| 9 | Excitement,Immobility/stupor,Mutism,Staring,Posturing/catalepsy,Stereotypy,Mannerisms,Negativism,Waxy flexibility | 99.6 | 98.9 | 100.0 |
| 9 | Excitement,Immobility/stupor,Mutism,Staring,Stereotypy,Mannerisms,Rigidity,Negativism,Waxy flexibility | 99.6 | 98.9 | 100.0 |
| 9 | Excitement,Immobility/stupor,Mutism,Staring,Stereotypy,Mannerisms,Rigidity,Negativism,Withdrawal | 99.6 | 98.9 | 100.0 |
| 9 | Excitement,Immobility/stupor,Mutism,Stereotypy,Mannerisms,Verbigeration,Rigidity,Negativism,Withdrawal | 99.6 | 98.9 | 100.0 |
| 9 | Excitement,Immobility/stupor,Posturing/catalepsy,Grimacing,Echopraxia/echolalia,Mannerisms,Rigidity,Negativism,Withdrawal | 99.6 | 98.9 | 100.0 |
| 9 | Excitement,Immobility/stupor,Posturing/catalepsy,Stereotypy,Mannerisms,Verbigeration,Rigidity,Negativism,Withdrawal | 99.6 | 98.9 | 100.0 |
| 9 | Excitement,Immobility/stupor,Staring,Echopraxia/echolalia,Mannerisms,Verbigeration,Rigidity,Negativism,Waxy flexibility | 99.6 | 98.9 | 100.0 |
| 9 | Excitement,Immobility/stupor,Staring,Echopraxia/echolalia,Mannerisms,Verbigeration,Rigidity,Waxy flexibility,Withdrawal | 99.6 | 98.9 | 100.0 |
| 9 | Excitement,Immobility/stupor,Staring,Echopraxia/echolalia,Stereotypy,Mannerisms,Verbigeration,Rigidity,Negativism | 99.6 | 98.9 | 100.0 |
| 9 | Excitement,Immobility/stupor,Staring,Echopraxia/echolalia,Stereotypy,Mannerisms,Verbigeration,Rigidity,Withdrawal | 99.6 | 98.9 | 100.0 |
| 9 | Excitement,Immobility/stupor,Staring,Echopraxia/echolalia,Stereotypy,Verbigeration,Rigidity,Negativism,Waxy flexibility | 99.6 | 98.9 | 100.0 |
| 9 | Excitement,Immobility/stupor,Staring,Echopraxia/echolalia,Verbigeration,Rigidity,Negativism,Waxy flexibility,Withdrawal | 99.6 | 98.9 | 100.0 |
| 9 | Excitement,Immobility/stupor,Staring,Grimacing,Echopraxia/echolalia,Mannerisms,Rigidity,Waxy flexibility,Withdrawal | 99.6 | 98.9 | 100.0 |
| 9 | Excitement,Immobility/stupor,Staring,Grimacing,Echopraxia/echolalia,Rigidity,Negativism,Waxy flexibility,Withdrawal | 99.6 | 98.9 | 100.0 |
| 9 | Excitement,Immobility/stupor,Staring,Grimacing,Mannerisms,Rigidity,Negativism,Waxy flexibility,Withdrawal | 99.6 | 98.9 | 100.0 |
| 9 | Excitement,Immobility/stupor,Staring,Grimacing,Mannerisms,Verbigeration,Rigidity,Waxy flexibility,Withdrawal | 99.6 | 98.9 | 100.0 |
| 9 | Excitement,Immobility/stupor,Staring,Grimacing,Stereotypy,Mannerisms,Rigidity,Negativism,Withdrawal | 99.6 | 98.9 | 100.0 |
| 9 | Excitement,Immobility/stupor,Staring,Grimacing,Stereotypy,Mannerisms,Verbigeration,Rigidity,Negativism | 99.6 | 98.9 | 100.0 |
| 9 | Excitement,Immobility/stupor,Staring,Grimacing,Stereotypy,Mannerisms,Verbigeration,Rigidity,Withdrawal | 99.6 | 98.9 | 100.0 |
| 9 | Excitement,Immobility/stupor,Staring,Grimacing,Stereotypy,Rigidity,Negativism,Waxy flexibility,Withdrawal | 99.6 | 98.9 | 100.0 |
| 9 | Excitement,Immobility/stupor,Staring,Grimacing,Stereotypy,Verbigeration,Rigidity,Negativism,Waxy flexibility | 99.6 | 98.9 | 100.0 |
| 9 | Excitement,Immobility/stupor,Staring,Posturing/catalepsy,Echopraxia/echolalia,Mannerisms,Rigidity,Negativism,Waxy flexibility | 99.6 | 98.9 | 100.0 |
| 9 | Excitement,Immobility/stupor,Staring,Posturing/catalepsy,Echopraxia/echolalia,Stereotypy,Mannerisms,Rigidity,Negativism | 99.6 | 98.9 | 100.0 |
| 9 | Excitement,Immobility/stupor,Staring,Posturing/catalepsy,Echopraxia/echolalia,Stereotypy,Negativism,Waxy flexibility,Withdrawal | 99.6 | 98.9 | 100.0 |
| 9 | Excitement,Immobility/stupor,Staring,Posturing/catalepsy,Echopraxia/echolalia,Stereotypy,Rigidity,Waxy flexibility,Withdrawal | 99.6 | 98.9 | 100.0 |
| 9 | Excitement,Immobility/stupor,Staring,Posturing/catalepsy,Echopraxia/echolalia,Stereotypy,Verbigeration,Negativism,Withdrawal | 99.6 | 98.9 | 100.0 |
| 9 | Excitement,Immobility/stupor,Staring,Posturing/catalepsy,Echopraxia/echolalia,Verbigeration,Rigidity,Negativism,Waxy flexibility | 99.6 | 98.9 | 100.0 |
| 9 | Excitement,Immobility/stupor,Staring,Posturing/catalepsy,Echopraxia/echolalia,Verbigeration,Rigidity,Waxy flexibility,Withdrawal | 99.6 | 98.9 | 100.0 |
| 9 | Excitement,Immobility/stupor,Staring,Posturing/catalepsy,Grimacing,Echopraxia/echolalia,Mannerisms,Rigidity,Negativism | 99.6 | 98.9 | 100.0 |
| 9 | Excitement,Immobility/stupor,Staring,Posturing/catalepsy,Grimacing,Echopraxia/echolalia,Negativism,Waxy flexibility,Withdrawal | 99.6 | 98.9 | 100.0 |
| 9 | Excitement,Immobility/stupor,Staring,Posturing/catalepsy,Grimacing,Echopraxia/echolalia,Stereotypy,Negativism,Withdrawal | 99.6 | 98.9 | 100.0 |
| 9 | Excitement,Immobility/stupor,Staring,Posturing/catalepsy,Grimacing,Echopraxia/echolalia,Verbigeration,Rigidity,Negativism | 99.6 | 98.9 | 100.0 |
| 9 | Excitement,Immobility/stupor,Staring,Posturing/catalepsy,Grimacing,Echopraxia/echolalia,Verbigeration,Rigidity,Withdrawal | 99.6 | 98.9 | 100.0 |
| 9 | Excitement,Immobility/stupor,Staring,Posturing/catalepsy,Grimacing,Mannerisms,Rigidity,Waxy flexibility,Withdrawal | 99.6 | 98.9 | 100.0 |
| 9 | Excitement,Immobility/stupor,Staring,Posturing/catalepsy,Grimacing,Stereotypy,Rigidity,Negativism,Withdrawal | 99.6 | 98.9 | 100.0 |
| 9 | Excitement,Immobility/stupor,Staring,Posturing/catalepsy,Grimacing,Stereotypy,Verbigeration,Negativism,Withdrawal | 99.6 | 98.9 | 100.0 |
| 9 | Excitement,Immobility/stupor,Staring,Posturing/catalepsy,Grimacing,Stereotypy,Verbigeration,Rigidity,Withdrawal | 99.6 | 98.9 | 100.0 |
| 9 | Excitement,Immobility/stupor,Staring,Posturing/catalepsy,Stereotypy,Mannerisms,Rigidity,Waxy flexibility,Withdrawal | 99.6 | 98.9 | 100.0 |
| 9 | Excitement,Immobility/stupor,Staring,Posturing/catalepsy,Stereotypy,Mannerisms,Verbigeration,Negativism,Waxy flexibility | 99.6 | 98.9 | 100.0 |
| 9 | Excitement,Immobility/stupor,Staring,Posturing/catalepsy,Stereotypy,Rigidity,Negativism,Waxy flexibility,Withdrawal | 99.6 | 98.9 | 100.0 |
| 9 | Excitement,Immobility/stupor,Staring,Posturing/catalepsy,Stereotypy,Verbigeration,Negativism,Waxy flexibility,Withdrawal | 99.6 | 98.9 | 100.0 |
| 9 | Excitement,Immobility/stupor,Staring,Posturing/catalepsy,Stereotypy,Verbigeration,Rigidity,Negativism,Waxy flexibility | 99.6 | 98.9 | 100.0 |
| 9 | Excitement,Immobility/stupor,Staring,Posturing/catalepsy,Stereotypy,Verbigeration,Rigidity,Waxy flexibility,Withdrawal | 99.6 | 98.9 | 100.0 |
| 9 | Excitement,Mutism,Staring,Echopraxia/echolalia,Mannerisms,Verbigeration,Rigidity,Waxy flexibility,Withdrawal | 99.6 | 98.9 | 100.0 |
| 9 | Excitement,Mutism,Staring,Echopraxia/echolalia,Stereotypy,Mannerisms,Verbigeration,Rigidity,Waxy flexibility | 99.6 | 98.9 | 100.0 |
| 9 | Excitement,Mutism,Staring,Echopraxia/echolalia,Stereotypy,Verbigeration,Negativism,Waxy flexibility,Withdrawal | 99.6 | 98.9 | 100.0 |
| 9 | Excitement,Mutism,Staring,Grimacing,Echopraxia/echolalia,Mannerisms,Rigidity,Negativism,Waxy flexibility | 99.6 | 98.9 | 100.0 |
| 9 | Excitement,Mutism,Staring,Grimacing,Echopraxia/echolalia,Mannerisms,Verbigeration,Rigidity,Negativism | 99.6 | 98.9 | 100.0 |
| 9 | Excitement,Mutism,Staring,Grimacing,Echopraxia/echolalia,Mannerisms,Verbigeration,Rigidity,Waxy flexibility | 99.6 | 98.9 | 100.0 |
| 9 | Excitement,Mutism,Staring,Grimacing,Echopraxia/echolalia,Mannerisms,Verbigeration,Rigidity,Withdrawal | 99.6 | 98.9 | 100.0 |
| 9 | Excitement,Mutism,Staring,Grimacing,Echopraxia/echolalia,Stereotypy,Mannerisms,Negativism,Withdrawal | 99.6 | 98.9 | 100.0 |
| 9 | Excitement,Mutism,Staring,Grimacing,Echopraxia/echolalia,Stereotypy,Mannerisms,Rigidity,Negativism | 99.6 | 98.9 | 100.0 |
| 9 | Excitement,Mutism,Staring,Grimacing,Echopraxia/echolalia,Stereotypy,Negativism,Waxy flexibility,Withdrawal | 99.6 | 98.9 | 100.0 |
| 9 | Excitement,Mutism,Staring,Grimacing,Echopraxia/echolalia,Stereotypy,Verbigeration,Negativism,Withdrawal | 99.6 | 98.9 | 100.0 |
| 9 | Excitement,Mutism,Staring,Grimacing,Mannerisms,Rigidity,Negativism,Waxy flexibility,Withdrawal | 99.6 | 98.9 | 100.0 |
| 9 | Excitement,Mutism,Staring,Grimacing,Stereotypy,Mannerisms,Rigidity,Negativism,Withdrawal | 99.6 | 98.9 | 100.0 |
| 9 | Excitement,Mutism,Staring,Grimacing,Stereotypy,Mannerisms,Verbigeration,Rigidity,Negativism | 99.6 | 98.9 | 100.0 |
| 9 | Excitement,Mutism,Staring,Grimacing,Stereotypy,Mannerisms,Verbigeration,Rigidity,Waxy flexibility | 99.6 | 98.9 | 100.0 |
| 9 | Excitement,Mutism,Staring,Grimacing,Verbigeration,Rigidity,Negativism,Waxy flexibility,Withdrawal | 99.6 | 98.9 | 100.0 |
| 9 | Excitement,Mutism,Staring,Posturing/catalepsy,Echopraxia/echolalia,Stereotypy,Mannerisms,Rigidity,Waxy flexibility | 99.6 | 98.9 | 100.0 |
| 9 | Excitement,Mutism,Staring,Posturing/catalepsy,Grimacing,Echopraxia/echolalia,Mannerisms,Rigidity,Waxy flexibility | 99.6 | 98.9 | 100.0 |
| 9 | Excitement,Mutism,Staring,Posturing/catalepsy,Grimacing,Echopraxia/echolalia,Mannerisms,Waxy flexibility,Withdrawal | 99.6 | 98.9 | 100.0 |
| 9 | Excitement,Mutism,Staring,Posturing/catalepsy,Grimacing,Echopraxia/echolalia,Stereotypy,Mannerisms,Negativism | 99.6 | 98.9 | 100.0 |
| 9 | Excitement,Mutism,Staring,Posturing/catalepsy,Grimacing,Echopraxia/echolalia,Stereotypy,Mannerisms,Rigidity | 99.6 | 98.9 | 100.0 |
| 9 | Excitement,Mutism,Staring,Posturing/catalepsy,Grimacing,Echopraxia/echolalia,Stereotypy,Rigidity,Waxy flexibility | 99.6 | 98.9 | 100.0 |
| 9 | Excitement,Mutism,Staring,Posturing/catalepsy,Grimacing,Echopraxia/echolalia,Stereotypy,Waxy flexibility,Withdrawal | 99.6 | 98.9 | 100.0 |
| 9 | Excitement,Mutism,Staring,Posturing/catalepsy,Grimacing,Mannerisms,Verbigeration,Rigidity,Waxy flexibility | 99.6 | 98.9 | 100.0 |
| 9 | Excitement,Mutism,Staring,Posturing/catalepsy,Grimacing,Stereotypy,Mannerisms,Verbigeration,Negativism | 99.6 | 98.9 | 100.0 |
| 9 | Excitement,Mutism,Staring,Posturing/catalepsy,Grimacing,Stereotypy,Mannerisms,Verbigeration,Rigidity | 99.6 | 98.9 | 100.0 |
| 9 | Excitement,Mutism,Staring,Posturing/catalepsy,Grimacing,Verbigeration,Rigidity,Negativism,Waxy flexibility | 99.6 | 98.9 | 100.0 |
| 9 | Excitement,Staring,Echopraxia/echolalia,Stereotypy,Mannerisms,Rigidity,Negativism,Waxy flexibility,Withdrawal | 99.6 | 98.9 | 100.0 |
| 9 | Excitement,Staring,Echopraxia/echolalia,Stereotypy,Mannerisms,Verbigeration,Rigidity,Negativism,Withdrawal | 99.6 | 98.9 | 100.0 |
| 9 | Excitement,Staring,Grimacing,Echopraxia/echolalia,Stereotypy,Mannerisms,Rigidity,Negativism,Withdrawal | 99.6 | 98.9 | 100.0 |
| 9 | Excitement,Staring,Grimacing,Stereotypy,Mannerisms,Verbigeration,Rigidity,Negativism,Withdrawal | 99.6 | 98.9 | 100.0 |
| 9 | Excitement,Staring,Posturing/catalepsy,Echopraxia/echolalia,Mannerisms,Rigidity,Negativism,Waxy flexibility,Withdrawal | 99.6 | 98.9 | 100.0 |
| 9 | Excitement,Staring,Posturing/catalepsy,Echopraxia/echolalia,Mannerisms,Verbigeration,Rigidity,Negativism,Withdrawal | 99.6 | 98.9 | 100.0 |
| 9 | Excitement,Staring,Posturing/catalepsy,Grimacing,Echopraxia/echolalia,Mannerisms,Rigidity,Negativism,Withdrawal | 99.6 | 98.9 | 100.0 |
| 9 | Excitement,Staring,Posturing/catalepsy,Grimacing,Mannerisms,Verbigeration,Rigidity,Negativism,Withdrawal | 99.6 | 98.9 | 100.0 |
| 9 | Excitement,Staring,Posturing/catalepsy,Mannerisms,Verbigeration,Rigidity,Negativism,Waxy flexibility,Withdrawal | 99.6 | 98.9 | 100.0 |
| 9 | Excitement,Staring,Posturing/catalepsy,Stereotypy,Mannerisms,Verbigeration,Rigidity,Negativism,Withdrawal | 99.6 | 98.9 | 100.0 |
| 9 | Excitement,Staring,Stereotypy,Mannerisms,Verbigeration,Rigidity,Negativism,Waxy flexibility,Withdrawal | 99.6 | 98.9 | 100.0 |
| 9 | Immobility/stupor,Mutism,Staring,Grimacing,Echopraxia/echolalia,Stereotypy,Mannerisms,Negativism,Withdrawal | 99.6 | 98.9 | 100.0 |
| 9 | Immobility/stupor,Mutism,Staring,Posturing/catalepsy,Echopraxia/echolalia,Mannerisms,Rigidity,Negativism,Withdrawal | 99.6 | 98.9 | 100.0 |
| 9 | Immobility/stupor,Mutism,Staring,Posturing/catalepsy,Echopraxia/echolalia,Stereotypy,Mannerisms,Negativism,Withdrawal | 99.6 | 98.9 | 100.0 |
| 9 | Immobility/stupor,Mutism,Staring,Posturing/catalepsy,Grimacing,Mannerisms,Verbigeration,Negativism,Withdrawal | 99.6 | 98.9 | 100.0 |
| 9 | Immobility/stupor,Mutism,Staring,Posturing/catalepsy,Stereotypy,Mannerisms,Rigidity,Negativism,Withdrawal | 99.6 | 98.9 | 100.0 |
| 9 | Immobility/stupor,Mutism,Staring,Posturing/catalepsy,Stereotypy,Mannerisms,Verbigeration,Negativism,Withdrawal | 99.6 | 98.9 | 100.0 |
| 9 | Immobility/stupor,Mutism,Staring,Stereotypy,Mannerisms,Verbigeration,Rigidity,Negativism,Waxy flexibility | 99.6 | 98.9 | 100.0 |
| 9 | Immobility/stupor,Staring,Grimacing,Echopraxia/echolalia,Mannerisms,Rigidity,Negativism,Waxy flexibility,Withdrawal | 99.6 | 98.9 | 100.0 |
| 9 | Immobility/stupor,Staring,Grimacing,Echopraxia/echolalia,Mannerisms,Verbigeration,Rigidity,Negativism,Withdrawal | 99.6 | 98.9 | 100.0 |
| 9 | Immobility/stupor,Staring,Posturing/catalepsy,Echopraxia/echolalia,Mannerisms,Rigidity,Negativism,Waxy flexibility,Withdrawal | 99.6 | 98.9 | 100.0 |
| 9 | Immobility/stupor,Staring,Posturing/catalepsy,Echopraxia/echolalia,Mannerisms,Verbigeration,Rigidity,Negativism,Withdrawal | 99.6 | 98.9 | 100.0 |
| 9 | Immobility/stupor,Staring,Posturing/catalepsy,Grimacing,Mannerisms,Verbigeration,Rigidity,Negativism,Withdrawal | 99.6 | 98.9 | 100.0 |
| 9 | Immobility/stupor,Staring,Posturing/catalepsy,Stereotypy,Mannerisms,Rigidity,Negativism,Waxy flexibility,Withdrawal | 99.6 | 98.9 | 100.0 |
| 9 | Mutism,Staring,Echopraxia/echolalia,Stereotypy,Mannerisms,Verbigeration,Rigidity,Negativism,Waxy flexibility | 99.6 | 98.9 | 100.0 |
| 9 | Mutism,Staring,Grimacing,Echopraxia/echolalia,Mannerisms,Rigidity,Negativism,Waxy flexibility,Withdrawal | 99.6 | 98.9 | 100.0 |
| 9 | Mutism,Staring,Grimacing,Echopraxia/echolalia,Mannerisms,Verbigeration,Rigidity,Negativism,Withdrawal | 99.6 | 98.9 | 100.0 |
| 9 | Mutism,Staring,Grimacing,Echopraxia/echolalia,Stereotypy,Mannerisms,Negativism,Waxy flexibility,Withdrawal | 99.6 | 98.9 | 100.0 |
| 9 | Mutism,Staring,Grimacing,Echopraxia/echolalia,Stereotypy,Mannerisms,Rigidity,Negativism,Waxy flexibility | 99.6 | 98.9 | 100.0 |
| 9 | Mutism,Staring,Grimacing,Echopraxia/echolalia,Stereotypy,Mannerisms,Rigidity,Waxy flexibility,Withdrawal | 99.6 | 98.9 | 100.0 |
| 9 | Mutism,Staring,Grimacing,Echopraxia/echolalia,Stereotypy,Mannerisms,Verbigeration,Negativism,Withdrawal | 99.6 | 98.9 | 100.0 |
| 9 | Mutism,Staring,Grimacing,Echopraxia/echolalia,Stereotypy,Mannerisms,Verbigeration,Rigidity,Negativism | 99.6 | 98.9 | 100.0 |
| 9 | Mutism,Staring,Grimacing,Stereotypy,Mannerisms,Verbigeration,Rigidity,Negativism,Waxy flexibility | 99.6 | 98.9 | 100.0 |
| 9 | Mutism,Staring,Posturing/catalepsy,Echopraxia/echolalia,Stereotypy,Mannerisms,Negativism,Waxy flexibility,Withdrawal | 99.6 | 98.9 | 100.0 |
| 9 | Mutism,Staring,Posturing/catalepsy,Echopraxia/echolalia,Stereotypy,Mannerisms,Verbigeration,Negativism,Withdrawal | 99.6 | 98.9 | 100.0 |
| 9 | Mutism,Staring,Posturing/catalepsy,Grimacing,Echopraxia/echolalia,Mannerisms,Verbigeration,Negativism,Waxy flexibility | 99.6 | 98.9 | 100.0 |
| 9 | Mutism,Staring,Posturing/catalepsy,Grimacing,Echopraxia/echolalia,Stereotypy,Mannerisms,Negativism,Waxy flexibility | 99.6 | 98.9 | 100.0 |
| 9 | Mutism,Staring,Posturing/catalepsy,Grimacing,Echopraxia/echolalia,Stereotypy,Mannerisms,Rigidity,Negativism | 99.6 | 98.9 | 100.0 |
| 9 | Mutism,Staring,Posturing/catalepsy,Grimacing,Echopraxia/echolalia,Stereotypy,Mannerisms,Rigidity,Withdrawal | 99.6 | 98.9 | 100.0 |
| 9 | Mutism,Staring,Posturing/catalepsy,Grimacing,Echopraxia/echolalia,Stereotypy,Mannerisms,Verbigeration,Negativism | 99.6 | 98.9 | 100.0 |
| 9 | Mutism,Staring,Posturing/catalepsy,Grimacing,Mannerisms,Verbigeration,Negativism,Waxy flexibility,Withdrawal | 99.6 | 98.9 | 100.0 |
| 9 | Mutism,Staring,Posturing/catalepsy,Grimacing,Stereotypy,Mannerisms,Verbigeration,Rigidity,Negativism | 99.6 | 98.9 | 100.0 |
| 9 | Mutism,Staring,Posturing/catalepsy,Grimacing,Stereotypy,Mannerisms,Verbigeration,Rigidity,Withdrawal | 99.6 | 98.9 | 100.0 |
| 9 | Mutism,Staring,Posturing/catalepsy,Stereotypy,Mannerisms,Rigidity,Negativism,Waxy flexibility,Withdrawal | 99.6 | 98.9 | 100.0 |
| 9 | Staring,Grimacing,Echopraxia/echolalia,Stereotypy,Mannerisms,Rigidity,Negativism,Waxy flexibility,Withdrawal | 99.6 | 98.9 | 100.0 |
| 9 | Staring,Grimacing,Echopraxia/echolalia,Stereotypy,Mannerisms,Verbigeration,Rigidity,Negativism,Withdrawal | 99.6 | 98.9 | 100.0 |
| 9 | Staring,Grimacing,Stereotypy,Mannerisms,Verbigeration,Rigidity,Negativism,Waxy flexibility,Withdrawal | 99.6 | 98.9 | 100.0 |
| 9 | Staring,Posturing/catalepsy,Grimacing,Stereotypy,Mannerisms,Verbigeration,Rigidity,Negativism,Withdrawal | 99.6 | 98.9 | 100.0 |
| 9 | Excitement,Immobility/stupor,Mutism,Grimacing,Echopraxia/echolalia,Verbigeration,Rigidity,Negativism,Withdrawal | 99.5 | 98.9 | 100.0 |
| 9 | Excitement,Immobility/stupor,Mutism,Posturing/catalepsy,Echopraxia/echolalia,Mannerisms,Verbigeration,Rigidity,Negativism | 99.5 | 98.9 | 100.0 |
| 9 | Excitement,Immobility/stupor,Mutism,Staring,Echopraxia/echolalia,Mannerisms,Verbigeration,Rigidity,Negativism | 99.5 | 98.9 | 100.0 |
| 9 | Excitement,Immobility/stupor,Mutism,Staring,Echopraxia/echolalia,Stereotypy,Mannerisms,Negativism,Withdrawal | 99.5 | 98.9 | 100.0 |
| 9 | Excitement,Immobility/stupor,Mutism,Staring,Echopraxia/echolalia,Stereotypy,Mannerisms,Rigidity,Negativism | 99.5 | 98.9 | 100.0 |
| 9 | Excitement,Immobility/stupor,Mutism,Staring,Echopraxia/echolalia,Stereotypy,Negativism,Waxy flexibility,Withdrawal | 99.5 | 98.9 | 100.0 |
| 9 | Excitement,Immobility/stupor,Mutism,Staring,Grimacing,Echopraxia/echolalia,Rigidity,Negativism,Waxy flexibility | 99.5 | 98.9 | 100.0 |
| 9 | Excitement,Immobility/stupor,Mutism,Staring,Grimacing,Mannerisms,Verbigeration,Rigidity,Waxy flexibility | 99.5 | 98.9 | 100.0 |
| 9 | Excitement,Immobility/stupor,Mutism,Staring,Grimacing,Mannerisms,Verbigeration,Rigidity,Withdrawal | 99.5 | 98.9 | 100.0 |
| 9 | Excitement,Immobility/stupor,Mutism,Staring,Grimacing,Verbigeration,Rigidity,Waxy flexibility,Withdrawal | 99.5 | 98.9 | 100.0 |
| 9 | Excitement,Immobility/stupor,Mutism,Staring,Posturing/catalepsy,Echopraxia/echolalia,Stereotypy,Mannerisms,Negativism | 99.5 | 98.9 | 100.0 |
| 9 | Excitement,Immobility/stupor,Mutism,Staring,Posturing/catalepsy,Grimacing,Echopraxia/echolalia,Mannerisms,Rigidity | 99.5 | 98.9 | 100.0 |
| 9 | Excitement,Immobility/stupor,Mutism,Staring,Posturing/catalepsy,Grimacing,Negativism,Waxy flexibility,Withdrawal | 99.5 | 98.9 | 100.0 |
| 9 | Excitement,Immobility/stupor,Mutism,Staring,Posturing/catalepsy,Grimacing,Stereotypy,Negativism,Waxy flexibility | 99.5 | 98.9 | 100.0 |
| 9 | Excitement,Immobility/stupor,Mutism,Staring,Posturing/catalepsy,Grimacing,Stereotypy,Rigidity,Negativism | 99.5 | 98.9 | 100.0 |
| 9 | Excitement,Immobility/stupor,Mutism,Staring,Posturing/catalepsy,Stereotypy,Mannerisms,Rigidity,Negativism | 99.5 | 98.9 | 100.0 |
| 9 | Excitement,Immobility/stupor,Mutism,Staring,Posturing/catalepsy,Stereotypy,Rigidity,Negativism,Waxy flexibility | 99.5 | 98.9 | 100.0 |
| 9 | Excitement,Immobility/stupor,Mutism,Staring,Verbigeration,Rigidity,Negativism,Waxy flexibility,Withdrawal | 99.5 | 98.9 | 100.0 |
| 9 | Excitement,Immobility/stupor,Mutism,Stereotypy,Verbigeration,Rigidity,Negativism,Waxy flexibility,Withdrawal | 99.5 | 98.9 | 100.0 |
| 9 | Excitement,Immobility/stupor,Posturing/catalepsy,Echopraxia/echolalia,Mannerisms,Verbigeration,Rigidity,Negativism,Withdrawal | 99.5 | 98.9 | 100.0 |
| 9 | Excitement,Immobility/stupor,Posturing/catalepsy,Echopraxia/echolalia,Stereotypy,Verbigeration,Rigidity,Negativism,Withdrawal | 99.5 | 98.9 | 100.0 |
| 9 | Excitement,Immobility/stupor,Posturing/catalepsy,Grimacing,Echopraxia/echolalia,Verbigeration,Rigidity,Negativism,Withdrawal | 99.5 | 98.9 | 100.0 |
| 9 | Excitement,Immobility/stupor,Posturing/catalepsy,Grimacing,Mannerisms,Verbigeration,Rigidity,Negativism,Withdrawal | 99.5 | 98.9 | 100.0 |
| 9 | Excitement,Immobility/stupor,Staring,Echopraxia/echolalia,Stereotypy,Mannerisms,Rigidity,Negativism,Waxy flexibility | 99.5 | 98.9 | 100.0 |
| 9 | Excitement,Immobility/stupor,Staring,Echopraxia/echolalia,Stereotypy,Verbigeration,Rigidity,Waxy flexibility,Withdrawal | 99.5 | 98.9 | 100.0 |
| 9 | Excitement,Immobility/stupor,Staring,Grimacing,Echopraxia/echolalia,Stereotypy,Mannerisms,Rigidity,Withdrawal | 99.5 | 98.9 | 100.0 |
| 9 | Excitement,Immobility/stupor,Staring,Grimacing,Echopraxia/echolalia,Stereotypy,Rigidity,Waxy flexibility,Withdrawal | 99.5 | 98.9 | 100.0 |
| 9 | Excitement,Immobility/stupor,Staring,Grimacing,Echopraxia/echolalia,Verbigeration,Rigidity,Negativism,Withdrawal | 99.5 | 98.9 | 100.0 |
| 9 | Excitement,Immobility/stupor,Staring,Grimacing,Mannerisms,Verbigeration,Rigidity,Negativism,Waxy flexibility | 99.5 | 98.9 | 100.0 |
| 9 | Excitement,Immobility/stupor,Staring,Grimacing,Stereotypy,Mannerisms,Rigidity,Waxy flexibility,Withdrawal | 99.5 | 98.9 | 100.0 |
| 9 | Excitement,Immobility/stupor,Staring,Grimacing,Stereotypy,Verbigeration,Rigidity,Waxy flexibility,Withdrawal | 99.5 | 98.9 | 100.0 |
| 9 | Excitement,Immobility/stupor,Staring,Posturing/catalepsy,Echopraxia/echolalia,Mannerisms,Verbigeration,Negativism,Waxy flexibility | 99.5 | 98.9 | 100.0 |
| 9 | Excitement,Immobility/stupor,Staring,Posturing/catalepsy,Echopraxia/echolalia,Stereotypy,Verbigeration,Rigidity,Negativism | 99.5 | 98.9 | 100.0 |
| 9 | Excitement,Immobility/stupor,Staring,Posturing/catalepsy,Echopraxia/echolalia,Stereotypy,Verbigeration,Rigidity,Withdrawal | 99.5 | 98.9 | 100.0 |
| 9 | Excitement,Immobility/stupor,Staring,Posturing/catalepsy,Echopraxia/echolalia,Verbigeration,Negativism,Waxy flexibility,Withdrawal | 99.5 | 98.9 | 100.0 |
| 9 | Excitement,Immobility/stupor,Staring,Posturing/catalepsy,Grimacing,Echopraxia/echolalia,Rigidity,Waxy flexibility,Withdrawal | 99.5 | 98.9 | 100.0 |
| 9 | Excitement,Immobility/stupor,Staring,Posturing/catalepsy,Grimacing,Echopraxia/echolalia,Stereotypy,Rigidity,Withdrawal | 99.5 | 98.9 | 100.0 |
| 9 | Excitement,Immobility/stupor,Staring,Posturing/catalepsy,Grimacing,Echopraxia/echolalia,Verbigeration,Negativism,Withdrawal | 99.5 | 98.9 | 100.0 |
| 9 | Excitement,Immobility/stupor,Staring,Posturing/catalepsy,Grimacing,Mannerisms,Verbigeration,Negativism,Waxy flexibility | 99.5 | 98.9 | 100.0 |
| 9 | Excitement,Immobility/stupor,Staring,Posturing/catalepsy,Grimacing,Stereotypy,Mannerisms,Rigidity,Withdrawal | 99.5 | 98.9 | 100.0 |
| 9 | Excitement,Immobility/stupor,Staring,Posturing/catalepsy,Grimacing,Stereotypy,Verbigeration,Rigidity,Negativism | 99.5 | 98.9 | 100.0 |
| 9 | Excitement,Immobility/stupor,Staring,Posturing/catalepsy,Grimacing,Verbigeration,Rigidity,Negativism,Withdrawal | 99.5 | 98.9 | 100.0 |
| 9 | Excitement,Immobility/stupor,Staring,Posturing/catalepsy,Verbigeration,Rigidity,Negativism,Waxy flexibility,Withdrawal | 99.5 | 98.9 | 100.0 |
| 9 | Excitement,Mutism,Staring,Echopraxia/echolalia,Stereotypy,Mannerisms,Negativism,Waxy flexibility,Withdrawal | 99.5 | 98.9 | 100.0 |
| 9 | Excitement,Mutism,Staring,Echopraxia/echolalia,Stereotypy,Mannerisms,Verbigeration,Negativism,Waxy flexibility | 99.5 | 98.9 | 100.0 |
| 9 | Excitement,Mutism,Staring,Echopraxia/echolalia,Stereotypy,Mannerisms,Verbigeration,Negativism,Withdrawal | 99.5 | 98.9 | 100.0 |
| 9 | Excitement,Mutism,Staring,Grimacing,Echopraxia/echolalia,Stereotypy,Mannerisms,Rigidity,Waxy flexibility | 99.5 | 98.9 | 100.0 |
| 9 | Excitement,Mutism,Staring,Grimacing,Echopraxia/echolalia,Stereotypy,Verbigeration,Negativism,Waxy flexibility | 99.5 | 98.9 | 100.0 |
| 9 | Excitement,Mutism,Staring,Grimacing,Mannerisms,Verbigeration,Rigidity,Negativism,Waxy flexibility | 99.5 | 98.9 | 100.0 |
| 9 | Excitement,Mutism,Staring,Posturing/catalepsy,Grimacing,Echopraxia/echolalia,Verbigeration,Waxy flexibility,Withdrawal | 99.5 | 98.9 | 100.0 |
| 9 | Excitement,Mutism,Staring,Posturing/catalepsy,Grimacing,Rigidity,Negativism,Waxy flexibility,Withdrawal | 99.5 | 98.9 | 100.0 |
| 9 | Excitement,Mutism,Staring,Posturing/catalepsy,Grimacing,Stereotypy,Verbigeration,Rigidity,Waxy flexibility | 99.5 | 98.9 | 100.0 |
| 9 | Excitement,Mutism,Staring,Posturing/catalepsy,Stereotypy,Mannerisms,Verbigeration,Rigidity,Waxy flexibility | 99.5 | 98.9 | 100.0 |
| 9 | Excitement,Staring,Posturing/catalepsy,Echopraxia/echolalia,Stereotypy,Mannerisms,Rigidity,Negativism,Withdrawal | 99.5 | 98.9 | 100.0 |
| 9 | Immobility/stupor,Mutism,Staring,Grimacing,Echopraxia/echolalia,Mannerisms,Rigidity,Negativism,Withdrawal | 99.5 | 98.9 | 100.0 |
| 9 | Immobility/stupor,Mutism,Staring,Grimacing,Stereotypy,Mannerisms,Rigidity,Negativism,Withdrawal | 99.5 | 98.9 | 100.0 |
| 9 | Immobility/stupor,Mutism,Staring,Grimacing,Stereotypy,Mannerisms,Verbigeration,Rigidity,Negativism | 99.5 | 98.9 | 100.0 |
| 9 | Immobility/stupor,Mutism,Staring,Posturing/catalepsy,Stereotypy,Mannerisms,Verbigeration,Rigidity,Negativism | 99.5 | 98.9 | 100.0 |
| 9 | Immobility/stupor,Mutism,Staring,Stereotypy,Mannerisms,Rigidity,Negativism,Waxy flexibility,Withdrawal | 99.5 | 98.9 | 100.0 |
| 9 | Immobility/stupor,Staring,Grimacing,Stereotypy,Mannerisms,Verbigeration,Rigidity,Negativism,Waxy flexibility | 99.5 | 98.9 | 100.0 |
| 9 | Immobility/stupor,Staring,Posturing/catalepsy,Grimacing,Stereotypy,Mannerisms,Verbigeration,Rigidity,Negativism | 99.5 | 98.9 | 100.0 |
| 9 | Mutism,Staring,Grimacing,Stereotypy,Mannerisms,Verbigeration,Rigidity,Waxy flexibility,Withdrawal | 99.5 | 98.9 | 100.0 |
| 9 | Mutism,Staring,Posturing/catalepsy,Echopraxia/echolalia,Mannerisms,Rigidity,Negativism,Waxy flexibility,Withdrawal | 99.5 | 98.9 | 100.0 |
| 9 | Mutism,Staring,Posturing/catalepsy,Echopraxia/echolalia,Mannerisms,Verbigeration,Rigidity,Negativism,Withdrawal | 99.5 | 98.9 | 100.0 |
| 9 | Mutism,Staring,Posturing/catalepsy,Echopraxia/echolalia,Stereotypy,Mannerisms,Verbigeration,Rigidity,Negativism | 99.5 | 98.9 | 100.0 |
| 9 | Mutism,Staring,Posturing/catalepsy,Grimacing,Echopraxia/echolalia,Mannerisms,Verbigeration,Rigidity,Negativism | 99.5 | 98.9 | 100.0 |
| 9 | Mutism,Staring,Posturing/catalepsy,Grimacing,Mannerisms,Verbigeration,Rigidity,Negativism,Withdrawal | 99.5 | 98.9 | 100.0 |
| 9 | Mutism,Staring,Posturing/catalepsy,Stereotypy,Mannerisms,Verbigeration,Negativism,Waxy flexibility,Withdrawal | 99.5 | 98.9 | 100.0 |
| 9 | Staring,Posturing/catalepsy,Grimacing,Echopraxia/echolalia,Stereotypy,Mannerisms,Rigidity,Negativism,Withdrawal | 99.5 | 98.9 | 100.0 |
| 9 | Excitement,Immobility/stupor,Echopraxia/echolalia,Stereotypy,Verbigeration,Rigidity,Negativism,Waxy flexibility,Withdrawal | 99.3 | 98.4 | 100.0 |
| 9 | Excitement,Immobility/stupor,Grimacing,Echopraxia/echolalia,Mannerisms,Verbigeration,Rigidity,Negativism,Withdrawal | 99.3 | 98.4 | 100.0 |
| 9 | Excitement,Immobility/stupor,Grimacing,Echopraxia/echolalia,Stereotypy,Mannerisms,Rigidity,Negativism,Withdrawal | 99.3 | 98.4 | 100.0 |
| 9 | Excitement,Immobility/stupor,Grimacing,Echopraxia/echolalia,Stereotypy,Verbigeration,Rigidity,Negativism,Withdrawal | 99.3 | 98.4 | 100.0 |
| 9 | Excitement,Immobility/stupor,Mutism,Echopraxia/echolalia,Mannerisms,Verbigeration,Rigidity,Negativism,Withdrawal | 99.3 | 98.4 | 100.0 |
| 9 | Excitement,Immobility/stupor,Mutism,Echopraxia/echolalia,Stereotypy,Mannerisms,Rigidity,Negativism,Withdrawal | 99.3 | 98.4 | 100.0 |
| 9 | Excitement,Immobility/stupor,Mutism,Echopraxia/echolalia,Stereotypy,Mannerisms,Verbigeration,Rigidity,Negativism | 99.3 | 98.4 | 100.0 |
| 9 | Excitement,Immobility/stupor,Mutism,Echopraxia/echolalia,Stereotypy,Rigidity,Negativism,Waxy flexibility,Withdrawal | 99.3 | 98.4 | 100.0 |
| 9 | Excitement,Immobility/stupor,Mutism,Echopraxia/echolalia,Stereotypy,Verbigeration,Rigidity,Waxy flexibility,Withdrawal | 99.3 | 98.4 | 100.0 |
| 9 | Excitement,Immobility/stupor,Mutism,Echopraxia/echolalia,Verbigeration,Rigidity,Negativism,Waxy flexibility,Withdrawal | 99.3 | 98.4 | 100.0 |
| 9 | Excitement,Immobility/stupor,Mutism,Grimacing,Echopraxia/echolalia,Mannerisms,Rigidity,Negativism,Withdrawal | 99.3 | 98.4 | 100.0 |
| 9 | Excitement,Immobility/stupor,Mutism,Grimacing,Echopraxia/echolalia,Rigidity,Negativism,Waxy flexibility,Withdrawal | 99.3 | 98.4 | 100.0 |
| 9 | Excitement,Immobility/stupor,Mutism,Grimacing,Echopraxia/echolalia,Stereotypy,Verbigeration,Rigidity,Negativism | 99.3 | 98.4 | 100.0 |
| 9 | Excitement,Immobility/stupor,Mutism,Grimacing,Echopraxia/echolalia,Verbigeration,Rigidity,Negativism,Waxy flexibility | 99.3 | 98.4 | 100.0 |
| 9 | Excitement,Immobility/stupor,Mutism,Grimacing,Mannerisms,Verbigeration,Rigidity,Negativism,Withdrawal | 99.3 | 98.4 | 100.0 |
| 9 | Excitement,Immobility/stupor,Mutism,Grimacing,Stereotypy,Verbigeration,Rigidity,Negativism,Waxy flexibility | 99.3 | 98.4 | 100.0 |
| 9 | Excitement,Immobility/stupor,Mutism,Posturing/catalepsy,Echopraxia/echolalia,Mannerisms,Rigidity,Negativism,Withdrawal | 99.3 | 98.4 | 100.0 |
| 9 | Excitement,Immobility/stupor,Mutism,Posturing/catalepsy,Echopraxia/echolalia,Mannerisms,Verbigeration,Rigidity,Withdrawal | 99.3 | 98.4 | 100.0 |
| 9 | Excitement,Immobility/stupor,Mutism,Posturing/catalepsy,Echopraxia/echolalia,Rigidity,Negativism,Waxy flexibility,Withdrawal | 99.3 | 98.4 | 100.0 |
| 9 | Excitement,Immobility/stupor,Mutism,Posturing/catalepsy,Echopraxia/echolalia,Verbigeration,Rigidity,Waxy flexibility,Withdrawal | 99.3 | 98.4 | 100.0 |
| 9 | Excitement,Immobility/stupor,Mutism,Posturing/catalepsy,Grimacing,Echopraxia/echolalia,Mannerisms,Rigidity,Negativism | 99.3 | 98.4 | 100.0 |
| 9 | Excitement,Immobility/stupor,Mutism,Posturing/catalepsy,Grimacing,Echopraxia/echolalia,Mannerisms,Rigidity,Withdrawal | 99.3 | 98.4 | 100.0 |
| 9 | Excitement,Immobility/stupor,Mutism,Posturing/catalepsy,Grimacing,Echopraxia/echolalia,Rigidity,Negativism,Waxy flexibility | 99.3 | 98.4 | 100.0 |
| 9 | Excitement,Immobility/stupor,Mutism,Posturing/catalepsy,Grimacing,Echopraxia/echolalia,Rigidity,Waxy flexibility,Withdrawal | 99.3 | 98.4 | 100.0 |
| 9 | Excitement,Immobility/stupor,Mutism,Posturing/catalepsy,Grimacing,Echopraxia/echolalia,Stereotypy,Rigidity,Negativism | 99.3 | 98.4 | 100.0 |
| 9 | Excitement,Immobility/stupor,Mutism,Posturing/catalepsy,Grimacing,Echopraxia/echolalia,Stereotypy,Rigidity,Withdrawal | 99.3 | 98.4 | 100.0 |
| 9 | Excitement,Immobility/stupor,Mutism,Posturing/catalepsy,Grimacing,Mannerisms,Rigidity,Negativism,Withdrawal | 99.3 | 98.4 | 100.0 |
| 9 | Excitement,Immobility/stupor,Mutism,Posturing/catalepsy,Grimacing,Mannerisms,Verbigeration,Rigidity,Withdrawal | 99.3 | 98.4 | 100.0 |
| 9 | Excitement,Immobility/stupor,Mutism,Posturing/catalepsy,Grimacing,Stereotypy,Rigidity,Negativism,Withdrawal | 99.3 | 98.4 | 100.0 |
| 9 | Excitement,Immobility/stupor,Mutism,Posturing/catalepsy,Grimacing,Stereotypy,Verbigeration,Rigidity,Withdrawal | 99.3 | 98.4 | 100.0 |
| 9 | Excitement,Immobility/stupor,Mutism,Posturing/catalepsy,Grimacing,Verbigeration,Rigidity,Negativism,Waxy flexibility | 99.3 | 98.4 | 100.0 |
| 9 | Excitement,Immobility/stupor,Mutism,Posturing/catalepsy,Mannerisms,Verbigeration,Rigidity,Negativism,Waxy flexibility | 99.3 | 98.4 | 100.0 |
| 9 | Excitement,Immobility/stupor,Mutism,Posturing/catalepsy,Stereotypy,Mannerisms,Verbigeration,Rigidity,Withdrawal | 99.3 | 98.4 | 100.0 |
| 9 | Excitement,Immobility/stupor,Mutism,Posturing/catalepsy,Stereotypy,Verbigeration,Rigidity,Waxy flexibility,Withdrawal | 99.3 | 98.4 | 100.0 |
| 9 | Excitement,Immobility/stupor,Mutism,Posturing/catalepsy,Verbigeration,Rigidity,Negativism,Waxy flexibility,Withdrawal | 99.3 | 98.4 | 100.0 |
| 9 | Excitement,Immobility/stupor,Mutism,Staring,Echopraxia/echolalia,Mannerisms,Negativism,Waxy flexibility,Withdrawal | 99.3 | 98.4 | 100.0 |
| 9 | Excitement,Immobility/stupor,Mutism,Staring,Echopraxia/echolalia,Mannerisms,Verbigeration,Negativism,Waxy flexibility | 99.3 | 98.4 | 100.0 |
| 9 | Excitement,Immobility/stupor,Mutism,Staring,Echopraxia/echolalia,Stereotypy,Mannerisms,Negativism,Waxy flexibility | 99.3 | 98.4 | 100.0 |
| 9 | Excitement,Immobility/stupor,Mutism,Staring,Echopraxia/echolalia,Stereotypy,Mannerisms,Verbigeration,Rigidity | 99.3 | 98.4 | 100.0 |
| 9 | Excitement,Immobility/stupor,Mutism,Staring,Echopraxia/echolalia,Verbigeration,Negativism,Waxy flexibility,Withdrawal | 99.3 | 98.4 | 100.0 |
| 9 | Excitement,Immobility/stupor,Mutism,Staring,Grimacing,Echopraxia/echolalia,Mannerisms,Rigidity,Negativism | 99.3 | 98.4 | 100.0 |
| 9 | Excitement,Immobility/stupor,Mutism,Staring,Grimacing,Echopraxia/echolalia,Mannerisms,Rigidity,Waxy flexibility | 99.3 | 98.4 | 100.0 |
| 9 | Excitement,Immobility/stupor,Mutism,Staring,Grimacing,Echopraxia/echolalia,Negativism,Waxy flexibility,Withdrawal | 99.3 | 98.4 | 100.0 |
| 9 | Excitement,Immobility/stupor,Mutism,Staring,Grimacing,Echopraxia/echolalia,Stereotypy,Negativism,Waxy flexibility | 99.3 | 98.4 | 100.0 |
| 9 | Excitement,Immobility/stupor,Mutism,Staring,Grimacing,Echopraxia/echolalia,Stereotypy,Verbigeration,Rigidity | 99.3 | 98.4 | 100.0 |
| 9 | Excitement,Immobility/stupor,Mutism,Staring,Grimacing,Echopraxia/echolalia,Verbigeration,Negativism,Waxy flexibility | 99.3 | 98.4 | 100.0 |
| 9 | Excitement,Immobility/stupor,Mutism,Staring,Grimacing,Mannerisms,Rigidity,Negativism,Waxy flexibility | 99.3 | 98.4 | 100.0 |
| 9 | Excitement,Immobility/stupor,Mutism,Staring,Grimacing,Mannerisms,Rigidity,Negativism,Withdrawal | 99.3 | 98.4 | 100.0 |
| 9 | Excitement,Immobility/stupor,Mutism,Staring,Grimacing,Rigidity,Negativism,Waxy flexibility,Withdrawal | 99.3 | 98.4 | 100.0 |
| 9 | Excitement,Immobility/stupor,Mutism,Staring,Grimacing,Stereotypy,Mannerisms,Rigidity,Waxy flexibility | 99.3 | 98.4 | 100.0 |
| 9 | Excitement,Immobility/stupor,Mutism,Staring,Grimacing,Stereotypy,Mannerisms,Rigidity,Withdrawal | 99.3 | 98.4 | 100.0 |
| 9 | Excitement,Immobility/stupor,Mutism,Staring,Grimacing,Stereotypy,Mannerisms,Verbigeration,Rigidity | 99.3 | 98.4 | 100.0 |
| 9 | Excitement,Immobility/stupor,Mutism,Staring,Grimacing,Stereotypy,Verbigeration,Negativism,Waxy flexibility | 99.3 | 98.4 | 100.0 |
| 9 | Excitement,Immobility/stupor,Mutism,Staring,Grimacing,Stereotypy,Verbigeration,Negativism,Withdrawal | 99.3 | 98.4 | 100.0 |
| 9 | Excitement,Immobility/stupor,Mutism,Staring,Posturing/catalepsy,Echopraxia/echolalia,Mannerisms,Waxy flexibility,Withdrawal | 99.3 | 98.4 | 100.0 |
| 9 | Excitement,Immobility/stupor,Mutism,Staring,Posturing/catalepsy,Echopraxia/echolalia,Stereotypy,Waxy flexibility,Withdrawal | 99.3 | 98.4 | 100.0 |
| 9 | Excitement,Immobility/stupor,Mutism,Staring,Posturing/catalepsy,Echopraxia/echolalia,Verbigeration,Waxy flexibility,Withdrawal | 99.3 | 98.4 | 100.0 |
| 9 | Excitement,Immobility/stupor,Mutism,Staring,Posturing/catalepsy,Grimacing,Echopraxia/echolalia,Mannerisms,Withdrawal | 99.3 | 98.4 | 100.0 |
| 9 | Excitement,Immobility/stupor,Mutism,Staring,Posturing/catalepsy,Grimacing,Echopraxia/echolalia,Stereotypy,Withdrawal | 99.3 | 98.4 | 100.0 |
| 9 | Excitement,Immobility/stupor,Mutism,Staring,Posturing/catalepsy,Grimacing,Echopraxia/echolalia,Verbigeration,Waxy flexibility | 99.3 | 98.4 | 100.0 |
| 9 | Excitement,Immobility/stupor,Mutism,Staring,Posturing/catalepsy,Grimacing,Echopraxia/echolalia,Verbigeration,Withdrawal | 99.3 | 98.4 | 100.0 |
| 9 | Excitement,Immobility/stupor,Mutism,Staring,Posturing/catalepsy,Grimacing,Mannerisms,Rigidity,Waxy flexibility | 99.3 | 98.4 | 100.0 |
| 9 | Excitement,Immobility/stupor,Mutism,Staring,Posturing/catalepsy,Grimacing,Rigidity,Negativism,Waxy flexibility | 99.3 | 98.4 | 100.0 |
| 9 | Excitement,Immobility/stupor,Mutism,Staring,Posturing/catalepsy,Grimacing,Stereotypy,Mannerisms,Negativism | 99.3 | 98.4 | 100.0 |
| 9 | Excitement,Immobility/stupor,Mutism,Staring,Posturing/catalepsy,Grimacing,Stereotypy,Mannerisms,Rigidity | 99.3 | 98.4 | 100.0 |
| 9 | Excitement,Immobility/stupor,Mutism,Staring,Posturing/catalepsy,Grimacing,Stereotypy,Rigidity,Waxy flexibility | 99.3 | 98.4 | 100.0 |
| 9 | Excitement,Immobility/stupor,Mutism,Staring,Posturing/catalepsy,Stereotypy,Mannerisms,Rigidity,Waxy flexibility | 99.3 | 98.4 | 100.0 |
| 9 | Excitement,Immobility/stupor,Mutism,Staring,Stereotypy,Mannerisms,Verbigeration,Negativism,Waxy flexibility | 99.3 | 98.4 | 100.0 |
| 9 | Excitement,Immobility/stupor,Mutism,Staring,Stereotypy,Mannerisms,Verbigeration,Negativism,Withdrawal | 99.3 | 98.4 | 100.0 |
| 9 | Excitement,Immobility/stupor,Mutism,Staring,Stereotypy,Verbigeration,Negativism,Waxy flexibility,Withdrawal | 99.3 | 98.4 | 100.0 |
| 9 | Excitement,Immobility/stupor,Mutism,Stereotypy,Mannerisms,Verbigeration,Rigidity,Negativism,Waxy flexibility | 99.3 | 98.4 | 100.0 |
| 9 | Excitement,Immobility/stupor,Posturing/catalepsy,Echopraxia/echolalia,Stereotypy,Mannerisms,Rigidity,Negativism,Withdrawal | 99.3 | 98.4 | 100.0 |
| 9 | Excitement,Immobility/stupor,Posturing/catalepsy,Echopraxia/echolalia,Verbigeration,Rigidity,Negativism,Waxy flexibility,Withdrawal | 99.3 | 98.4 | 100.0 |
| 9 | Excitement,Immobility/stupor,Posturing/catalepsy,Grimacing,Echopraxia/echolalia,Rigidity,Negativism,Waxy flexibility,Withdrawal | 99.3 | 98.4 | 100.0 |
| 9 | Excitement,Immobility/stupor,Posturing/catalepsy,Grimacing,Echopraxia/echolalia,Stereotypy,Rigidity,Negativism,Withdrawal | 99.3 | 98.4 | 100.0 |
| 9 | Excitement,Immobility/stupor,Posturing/catalepsy,Grimacing,Stereotypy,Verbigeration,Rigidity,Negativism,Withdrawal | 99.3 | 98.4 | 100.0 |
| 9 | Excitement,Immobility/stupor,Posturing/catalepsy,Mannerisms,Verbigeration,Rigidity,Negativism,Waxy flexibility,Withdrawal | 99.3 | 98.4 | 100.0 |
| 9 | Excitement,Immobility/stupor,Posturing/catalepsy,Stereotypy,Verbigeration,Rigidity,Negativism,Waxy flexibility,Withdrawal | 99.3 | 98.4 | 100.0 |
| 9 | Excitement,Immobility/stupor,Staring,Echopraxia/echolalia,Stereotypy,Mannerisms,Negativism,Waxy flexibility,Withdrawal | 99.3 | 98.4 | 100.0 |
| 9 | Excitement,Immobility/stupor,Staring,Echopraxia/echolalia,Stereotypy,Mannerisms,Verbigeration,Negativism,Withdrawal | 99.3 | 98.4 | 100.0 |
| 9 | Excitement,Immobility/stupor,Staring,Echopraxia/echolalia,Stereotypy,Mannerisms,Verbigeration,Rigidity,Waxy flexibility | 99.3 | 98.4 | 100.0 |
| 9 | Excitement,Immobility/stupor,Staring,Grimacing,Echopraxia/echolalia,Mannerisms,Rigidity,Negativism,Waxy flexibility | 99.3 | 98.4 | 100.0 |
| 9 | Excitement,Immobility/stupor,Staring,Grimacing,Echopraxia/echolalia,Mannerisms,Verbigeration,Rigidity,Negativism | 99.3 | 98.4 | 100.0 |
| 9 | Excitement,Immobility/stupor,Staring,Grimacing,Echopraxia/echolalia,Mannerisms,Verbigeration,Rigidity,Withdrawal | 99.3 | 98.4 | 100.0 |
| 9 | Excitement,Immobility/stupor,Staring,Grimacing,Echopraxia/echolalia,Stereotypy,Mannerisms,Negativism,Withdrawal | 99.3 | 98.4 | 100.0 |
| 9 | Excitement,Immobility/stupor,Staring,Grimacing,Echopraxia/echolalia,Stereotypy,Mannerisms,Rigidity,Negativism | 99.3 | 98.4 | 100.0 |
| 9 | Excitement,Immobility/stupor,Staring,Grimacing,Echopraxia/echolalia,Stereotypy,Rigidity,Negativism,Waxy flexibility | 99.3 | 98.4 | 100.0 |
| 9 | Excitement,Immobility/stupor,Staring,Grimacing,Echopraxia/echolalia,Stereotypy,Verbigeration,Rigidity,Negativism | 99.3 | 98.4 | 100.0 |
| 9 | Excitement,Immobility/stupor,Staring,Grimacing,Echopraxia/echolalia,Stereotypy,Verbigeration,Rigidity,Withdrawal | 99.3 | 98.4 | 100.0 |
| 9 | Excitement,Immobility/stupor,Staring,Grimacing,Echopraxia/echolalia,Verbigeration,Rigidity,Negativism,Waxy flexibility | 99.3 | 98.4 | 100.0 |
| 9 | Excitement,Immobility/stupor,Staring,Grimacing,Echopraxia/echolalia,Verbigeration,Rigidity,Waxy flexibility,Withdrawal | 99.3 | 98.4 | 100.0 |
| 9 | Excitement,Immobility/stupor,Staring,Grimacing,Stereotypy,Mannerisms,Rigidity,Negativism,Waxy flexibility | 99.3 | 98.4 | 100.0 |
| 9 | Excitement,Immobility/stupor,Staring,Grimacing,Stereotypy,Mannerisms,Verbigeration,Rigidity,Waxy flexibility | 99.3 | 98.4 | 100.0 |
| 9 | Excitement,Immobility/stupor,Staring,Grimacing,Verbigeration,Rigidity,Negativism,Waxy flexibility,Withdrawal | 99.3 | 98.4 | 100.0 |
| 9 | Excitement,Immobility/stupor,Staring,Posturing/catalepsy,Echopraxia/echolalia,Mannerisms,Verbigeration,Rigidity,Waxy flexibility | 99.3 | 98.4 | 100.0 |
| 9 | Excitement,Immobility/stupor,Staring,Posturing/catalepsy,Echopraxia/echolalia,Stereotypy,Mannerisms,Negativism,Waxy flexibility | 99.3 | 98.4 | 100.0 |
| 9 | Excitement,Immobility/stupor,Staring,Posturing/catalepsy,Echopraxia/echolalia,Stereotypy,Mannerisms,Verbigeration,Negativism | 99.3 | 98.4 | 100.0 |
| 9 | Excitement,Immobility/stupor,Staring,Posturing/catalepsy,Echopraxia/echolalia,Stereotypy,Mannerisms,Verbigeration,Rigidity | 99.3 | 98.4 | 100.0 |
| 9 | Excitement,Immobility/stupor,Staring,Posturing/catalepsy,Echopraxia/echolalia,Stereotypy,Rigidity,Negativism,Waxy flexibility | 99.3 | 98.4 | 100.0 |
| 9 | Excitement,Immobility/stupor,Staring,Posturing/catalepsy,Echopraxia/echolalia,Stereotypy,Verbigeration,Negativism,Waxy flexibility | 99.3 | 98.4 | 100.0 |
| 9 | Excitement,Immobility/stupor,Staring,Posturing/catalepsy,Grimacing,Echopraxia/echolalia,Mannerisms,Negativism,Waxy flexibility | 99.3 | 98.4 | 100.0 |
| 9 | Excitement,Immobility/stupor,Staring,Posturing/catalepsy,Grimacing,Echopraxia/echolalia,Mannerisms,Verbigeration,Negativism | 99.3 | 98.4 | 100.0 |
| 9 | Excitement,Immobility/stupor,Staring,Posturing/catalepsy,Grimacing,Echopraxia/echolalia,Mannerisms,Verbigeration,Rigidity | 99.3 | 98.4 | 100.0 |
| 9 | Excitement,Immobility/stupor,Staring,Posturing/catalepsy,Grimacing,Echopraxia/echolalia,Rigidity,Negativism,Waxy flexibility | 99.3 | 98.4 | 100.0 |
| 9 | Excitement,Immobility/stupor,Staring,Posturing/catalepsy,Grimacing,Echopraxia/echolalia,Stereotypy,Rigidity,Negativism | 99.3 | 98.4 | 100.0 |
| 9 | Excitement,Immobility/stupor,Staring,Posturing/catalepsy,Grimacing,Echopraxia/echolalia,Verbigeration,Negativism,Waxy flexibility | 99.3 | 98.4 | 100.0 |
| 9 | Excitement,Immobility/stupor,Staring,Posturing/catalepsy,Grimacing,Mannerisms,Negativism,Waxy flexibility,Withdrawal | 99.3 | 98.4 | 100.0 |
| 9 | Excitement,Immobility/stupor,Staring,Posturing/catalepsy,Grimacing,Mannerisms,Rigidity,Negativism,Waxy flexibility | 99.3 | 98.4 | 100.0 |
| 9 | Excitement,Immobility/stupor,Staring,Posturing/catalepsy,Grimacing,Mannerisms,Verbigeration,Rigidity,Waxy flexibility | 99.3 | 98.4 | 100.0 |
| 9 | Excitement,Immobility/stupor,Staring,Posturing/catalepsy,Grimacing,Rigidity,Negativism,Waxy flexibility,Withdrawal | 99.3 | 98.4 | 100.0 |
| 9 | Excitement,Immobility/stupor,Staring,Posturing/catalepsy,Grimacing,Stereotypy,Mannerisms,Negativism,Withdrawal | 99.3 | 98.4 | 100.0 |
| 9 | Excitement,Immobility/stupor,Staring,Posturing/catalepsy,Grimacing,Stereotypy,Mannerisms,Rigidity,Negativism | 99.3 | 98.4 | 100.0 |
| 9 | Excitement,Immobility/stupor,Staring,Posturing/catalepsy,Grimacing,Stereotypy,Mannerisms,Verbigeration,Negativism | 99.3 | 98.4 | 100.0 |
| 9 | Excitement,Immobility/stupor,Staring,Posturing/catalepsy,Grimacing,Stereotypy,Mannerisms,Verbigeration,Rigidity | 99.3 | 98.4 | 100.0 |
| 9 | Excitement,Immobility/stupor,Staring,Posturing/catalepsy,Grimacing,Stereotypy,Rigidity,Waxy flexibility,Withdrawal | 99.3 | 98.4 | 100.0 |
| 9 | Excitement,Immobility/stupor,Staring,Posturing/catalepsy,Grimacing,Stereotypy,Verbigeration,Negativism,Waxy flexibility | 99.3 | 98.4 | 100.0 |
| 9 | Excitement,Immobility/stupor,Staring,Posturing/catalepsy,Grimacing,Verbigeration,Negativism,Waxy flexibility,Withdrawal | 99.3 | 98.4 | 100.0 |
| 9 | Excitement,Immobility/stupor,Staring,Posturing/catalepsy,Grimacing,Verbigeration,Rigidity,Negativism,Waxy flexibility | 99.3 | 98.4 | 100.0 |
| 9 | Excitement,Immobility/stupor,Staring,Posturing/catalepsy,Grimacing,Verbigeration,Rigidity,Waxy flexibility,Withdrawal | 99.3 | 98.4 | 100.0 |
| 9 | Excitement,Immobility/stupor,Staring,Posturing/catalepsy,Stereotypy,Mannerisms,Negativism,Waxy flexibility,Withdrawal | 99.3 | 98.4 | 100.0 |
| 9 | Excitement,Immobility/stupor,Staring,Posturing/catalepsy,Stereotypy,Mannerisms,Rigidity,Negativism,Waxy flexibility | 99.3 | 98.4 | 100.0 |
| 9 | Excitement,Immobility/stupor,Staring,Posturing/catalepsy,Stereotypy,Mannerisms,Verbigeration,Rigidity,Waxy flexibility | 99.3 | 98.4 | 100.0 |
| 9 | Excitement,Mutism,Staring,Echopraxia/echolalia,Mannerisms,Verbigeration,Negativism,Waxy flexibility,Withdrawal | 99.3 | 98.4 | 100.0 |
| 9 | Excitement,Mutism,Staring,Grimacing,Echopraxia/echolalia,Mannerisms,Negativism,Waxy flexibility,Withdrawal | 99.3 | 98.4 | 100.0 |
| 9 | Excitement,Mutism,Staring,Grimacing,Echopraxia/echolalia,Mannerisms,Verbigeration,Negativism,Waxy flexibility | 99.3 | 98.4 | 100.0 |
| 9 | Excitement,Mutism,Staring,Grimacing,Echopraxia/echolalia,Stereotypy,Mannerisms,Negativism,Waxy flexibility | 99.3 | 98.4 | 100.0 |
| 9 | Excitement,Mutism,Staring,Grimacing,Echopraxia/echolalia,Stereotypy,Mannerisms,Verbigeration,Rigidity | 99.3 | 98.4 | 100.0 |
| 9 | Excitement,Mutism,Staring,Grimacing,Echopraxia/echolalia,Verbigeration,Negativism,Waxy flexibility,Withdrawal | 99.3 | 98.4 | 100.0 |
| 9 | Excitement,Mutism,Staring,Grimacing,Stereotypy,Mannerisms,Rigidity,Negativism,Waxy flexibility | 99.3 | 98.4 | 100.0 |
| 9 | Excitement,Mutism,Staring,Grimacing,Stereotypy,Mannerisms,Verbigeration,Negativism,Withdrawal | 99.3 | 98.4 | 100.0 |
| 9 | Excitement,Mutism,Staring,Grimacing,Stereotypy,Verbigeration,Negativism,Waxy flexibility,Withdrawal | 99.3 | 98.4 | 100.0 |
| 9 | Excitement,Mutism,Staring,Posturing/catalepsy,Echopraxia/echolalia,Mannerisms,Verbigeration,Waxy flexibility,Withdrawal | 99.3 | 98.4 | 100.0 |
| 9 | Excitement,Mutism,Staring,Posturing/catalepsy,Echopraxia/echolalia,Stereotypy,Mannerisms,Waxy flexibility,Withdrawal | 99.3 | 98.4 | 100.0 |
| 9 | Excitement,Mutism,Staring,Posturing/catalepsy,Echopraxia/echolalia,Stereotypy,Verbigeration,Waxy flexibility,Withdrawal | 99.3 | 98.4 | 100.0 |
| 9 | Excitement,Mutism,Staring,Posturing/catalepsy,Grimacing,Echopraxia/echolalia,Mannerisms,Verbigeration,Withdrawal | 99.3 | 98.4 | 100.0 |
| 9 | Excitement,Mutism,Staring,Posturing/catalepsy,Grimacing,Echopraxia/echolalia,Stereotypy,Mannerisms,Withdrawal | 99.3 | 98.4 | 100.0 |
| 9 | Excitement,Mutism,Staring,Posturing/catalepsy,Grimacing,Echopraxia/echolalia,Stereotypy,Verbigeration,Waxy flexibility | 99.3 | 98.4 | 100.0 |
| 9 | Excitement,Mutism,Staring,Posturing/catalepsy,Grimacing,Echopraxia/echolalia,Stereotypy,Verbigeration,Withdrawal | 99.3 | 98.4 | 100.0 |
| 9 | Excitement,Mutism,Staring,Posturing/catalepsy,Grimacing,Mannerisms,Rigidity,Negativism,Waxy flexibility | 99.3 | 98.4 | 100.0 |
| 9 | Excitement,Mutism,Staring,Posturing/catalepsy,Grimacing,Mannerisms,Verbigeration,Waxy flexibility,Withdrawal | 99.3 | 98.4 | 100.0 |
| 9 | Excitement,Mutism,Staring,Posturing/catalepsy,Grimacing,Stereotypy,Mannerisms,Negativism,Waxy flexibility | 99.3 | 98.4 | 100.0 |
| 9 | Excitement,Mutism,Staring,Posturing/catalepsy,Grimacing,Stereotypy,Mannerisms,Rigidity,Negativism | 99.3 | 98.4 | 100.0 |
| 9 | Excitement,Mutism,Staring,Posturing/catalepsy,Grimacing,Stereotypy,Rigidity,Negativism,Waxy flexibility | 99.3 | 98.4 | 100.0 |
| 9 | Excitement,Mutism,Staring,Posturing/catalepsy,Grimacing,Stereotypy,Verbigeration,Waxy flexibility,Withdrawal | 99.3 | 98.4 | 100.0 |
| 9 | Excitement,Mutism,Staring,Posturing/catalepsy,Stereotypy,Mannerisms,Rigidity,Negativism,Waxy flexibility | 99.3 | 98.4 | 100.0 |
| 9 | Excitement,Mutism,Staring,Stereotypy,Mannerisms,Verbigeration,Negativism,Waxy flexibility,Withdrawal | 99.3 | 98.4 | 100.0 |
| 9 | Excitement,Staring,Echopraxia/echolalia,Mannerisms,Verbigeration,Rigidity,Negativism,Waxy flexibility,Withdrawal | 99.3 | 98.4 | 100.0 |
| 9 | Excitement,Staring,Grimacing,Echopraxia/echolalia,Mannerisms,Rigidity,Negativism,Waxy flexibility,Withdrawal | 99.3 | 98.4 | 100.0 |
| 9 | Excitement,Staring,Grimacing,Echopraxia/echolalia,Stereotypy,Mannerisms,Rigidity,Waxy flexibility,Withdrawal | 99.3 | 98.4 | 100.0 |
| 9 | Excitement,Staring,Grimacing,Mannerisms,Verbigeration,Rigidity,Negativism,Waxy flexibility,Withdrawal | 99.3 | 98.4 | 100.0 |
| 9 | Excitement,Staring,Grimacing,Stereotypy,Mannerisms,Rigidity,Negativism,Waxy flexibility,Withdrawal | 99.3 | 98.4 | 100.0 |
| 9 | Excitement,Staring,Grimacing,Stereotypy,Mannerisms,Verbigeration,Rigidity,Waxy flexibility,Withdrawal | 99.3 | 98.4 | 100.0 |
| 9 | Excitement,Staring,Posturing/catalepsy,Grimacing,Echopraxia/echolalia,Mannerisms,Rigidity,Waxy flexibility,Withdrawal | 99.3 | 98.4 | 100.0 |
| 9 | Excitement,Staring,Posturing/catalepsy,Grimacing,Echopraxia/echolalia,Mannerisms,Verbigeration,Rigidity,Withdrawal | 99.3 | 98.4 | 100.0 |
| 9 | Excitement,Staring,Posturing/catalepsy,Grimacing,Echopraxia/echolalia,Stereotypy,Mannerisms,Rigidity,Withdrawal | 99.3 | 98.4 | 100.0 |
| 9 | Excitement,Staring,Posturing/catalepsy,Grimacing,Mannerisms,Rigidity,Negativism,Waxy flexibility,Withdrawal | 99.3 | 98.4 | 100.0 |
| 9 | Excitement,Staring,Posturing/catalepsy,Grimacing,Mannerisms,Verbigeration,Rigidity,Waxy flexibility,Withdrawal | 99.3 | 98.4 | 100.0 |
| 9 | Excitement,Staring,Posturing/catalepsy,Grimacing,Stereotypy,Mannerisms,Rigidity,Negativism,Withdrawal | 99.3 | 98.4 | 100.0 |
| 9 | Excitement,Staring,Posturing/catalepsy,Grimacing,Stereotypy,Mannerisms,Verbigeration,Rigidity,Withdrawal | 99.3 | 98.4 | 100.0 |
| 9 | Excitement,Staring,Posturing/catalepsy,Stereotypy,Mannerisms,Rigidity,Negativism,Waxy flexibility,Withdrawal | 99.3 | 98.4 | 100.0 |
| 9 | Immobility/stupor,Mutism,Staring,Echopraxia/echolalia,Mannerisms,Rigidity,Negativism,Waxy flexibility,Withdrawal | 99.3 | 98.4 | 100.0 |
| 9 | Immobility/stupor,Mutism,Staring,Echopraxia/echolalia,Mannerisms,Verbigeration,Rigidity,Negativism,Withdrawal | 99.3 | 98.4 | 100.0 |
| 9 | Immobility/stupor,Mutism,Staring,Echopraxia/echolalia,Stereotypy,Mannerisms,Rigidity,Negativism,Waxy flexibility | 99.3 | 98.4 | 100.0 |
| 9 | Immobility/stupor,Mutism,Staring,Echopraxia/echolalia,Stereotypy,Mannerisms,Verbigeration,Rigidity,Negativism | 99.3 | 98.4 | 100.0 |
| 9 | Immobility/stupor,Mutism,Staring,Grimacing,Echopraxia/echolalia,Stereotypy,Mannerisms,Rigidity,Negativism | 99.3 | 98.4 | 100.0 |
| 9 | Immobility/stupor,Mutism,Staring,Grimacing,Echopraxia/echolalia,Stereotypy,Mannerisms,Rigidity,Withdrawal | 99.3 | 98.4 | 100.0 |
| 9 | Immobility/stupor,Mutism,Staring,Grimacing,Mannerisms,Verbigeration,Rigidity,Negativism,Withdrawal | 99.3 | 98.4 | 100.0 |
| 9 | Immobility/stupor,Mutism,Staring,Grimacing,Stereotypy,Mannerisms,Rigidity,Negativism,Waxy flexibility | 99.3 | 98.4 | 100.0 |
| 9 | Immobility/stupor,Mutism,Staring,Grimacing,Stereotypy,Mannerisms,Rigidity,Waxy flexibility,Withdrawal | 99.3 | 98.4 | 100.0 |
| 9 | Immobility/stupor,Mutism,Staring,Grimacing,Stereotypy,Mannerisms,Verbigeration,Negativism,Withdrawal | 99.3 | 98.4 | 100.0 |
| 9 | Immobility/stupor,Mutism,Staring,Grimacing,Stereotypy,Mannerisms,Verbigeration,Rigidity,Withdrawal | 99.3 | 98.4 | 100.0 |
| 9 | Immobility/stupor,Mutism,Staring,Grimacing,Stereotypy,Verbigeration,Rigidity,Negativism,Withdrawal | 99.3 | 98.4 | 100.0 |
| 9 | Immobility/stupor,Mutism,Staring,Posturing/catalepsy,Echopraxia/echolalia,Mannerisms,Negativism,Waxy flexibility,Withdrawal | 99.3 | 98.4 | 100.0 |
| 9 | Immobility/stupor,Mutism,Staring,Posturing/catalepsy,Echopraxia/echolalia,Mannerisms,Verbigeration,Negativism,Withdrawal | 99.3 | 98.4 | 100.0 |
| 9 | Immobility/stupor,Mutism,Staring,Posturing/catalepsy,Echopraxia/echolalia,Mannerisms,Verbigeration,Rigidity,Negativism | 99.3 | 98.4 | 100.0 |
| 9 | Immobility/stupor,Mutism,Staring,Posturing/catalepsy,Echopraxia/echolalia,Stereotypy,Mannerisms,Rigidity,Negativism | 99.3 | 98.4 | 100.0 |
| 9 | Immobility/stupor,Mutism,Staring,Posturing/catalepsy,Grimacing,Echopraxia/echolalia,Mannerisms,Rigidity,Negativism | 99.3 | 98.4 | 100.0 |
| 9 | Immobility/stupor,Mutism,Staring,Posturing/catalepsy,Grimacing,Echopraxia/echolalia,Mannerisms,Verbigeration,Negativism | 99.3 | 98.4 | 100.0 |
| 9 | Immobility/stupor,Mutism,Staring,Posturing/catalepsy,Grimacing,Echopraxia/echolalia,Stereotypy,Mannerisms,Negativism | 99.3 | 98.4 | 100.0 |
| 9 | Immobility/stupor,Mutism,Staring,Posturing/catalepsy,Grimacing,Mannerisms,Negativism,Waxy flexibility,Withdrawal | 99.3 | 98.4 | 100.0 |
| 9 | Immobility/stupor,Mutism,Staring,Posturing/catalepsy,Grimacing,Mannerisms,Rigidity,Negativism,Withdrawal | 99.3 | 98.4 | 100.0 |
| 9 | Immobility/stupor,Mutism,Staring,Posturing/catalepsy,Grimacing,Stereotypy,Mannerisms,Rigidity,Negativism | 99.3 | 98.4 | 100.0 |
| 9 | Immobility/stupor,Mutism,Staring,Posturing/catalepsy,Grimacing,Stereotypy,Mannerisms,Rigidity,Withdrawal | 99.3 | 98.4 | 100.0 |
| 9 | Immobility/stupor,Mutism,Staring,Posturing/catalepsy,Grimacing,Stereotypy,Mannerisms,Verbigeration,Negativism | 99.3 | 98.4 | 100.0 |
| 9 | Immobility/stupor,Mutism,Staring,Posturing/catalepsy,Mannerisms,Verbigeration,Rigidity,Negativism,Withdrawal | 99.3 | 98.4 | 100.0 |
| 9 | Immobility/stupor,Mutism,Staring,Posturing/catalepsy,Stereotypy,Mannerisms,Negativism,Waxy flexibility,Withdrawal | 99.3 | 98.4 | 100.0 |
| 9 | Immobility/stupor,Staring,Echopraxia/echolalia,Mannerisms,Verbigeration,Rigidity,Negativism,Waxy flexibility,Withdrawal | 99.3 | 98.4 | 100.0 |
| 9 | Immobility/stupor,Staring,Echopraxia/echolalia,Stereotypy,Mannerisms,Verbigeration,Rigidity,Negativism,Waxy flexibility | 99.3 | 98.4 | 100.0 |
| 9 | Immobility/stupor,Staring,Grimacing,Echopraxia/echolalia,Stereotypy,Mannerisms,Negativism,Waxy flexibility,Withdrawal | 99.3 | 98.4 | 100.0 |
| 9 | Immobility/stupor,Staring,Grimacing,Echopraxia/echolalia,Stereotypy,Mannerisms,Rigidity,Negativism,Waxy flexibility | 99.3 | 98.4 | 100.0 |
| 9 | Immobility/stupor,Staring,Grimacing,Echopraxia/echolalia,Stereotypy,Mannerisms,Verbigeration,Negativism,Withdrawal | 99.3 | 98.4 | 100.0 |
| 9 | Immobility/stupor,Staring,Grimacing,Echopraxia/echolalia,Stereotypy,Mannerisms,Verbigeration,Rigidity,Negativism | 99.3 | 98.4 | 100.0 |
| 9 | Immobility/stupor,Staring,Grimacing,Mannerisms,Verbigeration,Rigidity,Negativism,Waxy flexibility,Withdrawal | 99.3 | 98.4 | 100.0 |
| 9 | Immobility/stupor,Staring,Posturing/catalepsy,Echopraxia/echolalia,Stereotypy,Mannerisms,Negativism,Waxy flexibility,Withdrawal | 99.3 | 98.4 | 100.0 |
| 9 | Immobility/stupor,Staring,Posturing/catalepsy,Echopraxia/echolalia,Stereotypy,Mannerisms,Verbigeration,Negativism,Withdrawal | 99.3 | 98.4 | 100.0 |
| 9 | Immobility/stupor,Staring,Posturing/catalepsy,Echopraxia/echolalia,Stereotypy,Mannerisms,Verbigeration,Rigidity,Negativism | 99.3 | 98.4 | 100.0 |
| 9 | Immobility/stupor,Staring,Posturing/catalepsy,Grimacing,Echopraxia/echolalia,Mannerisms,Negativism,Waxy flexibility,Withdrawal | 99.3 | 98.4 | 100.0 |
| 9 | Immobility/stupor,Staring,Posturing/catalepsy,Grimacing,Echopraxia/echolalia,Mannerisms,Verbigeration,Negativism,Withdrawal | 99.3 | 98.4 | 100.0 |
| 9 | Immobility/stupor,Staring,Posturing/catalepsy,Grimacing,Echopraxia/echolalia,Mannerisms,Verbigeration,Rigidity,Negativism | 99.3 | 98.4 | 100.0 |
| 9 | Immobility/stupor,Staring,Posturing/catalepsy,Grimacing,Echopraxia/echolalia,Stereotypy,Mannerisms,Rigidity,Negativism | 99.3 | 98.4 | 100.0 |
| 9 | Immobility/stupor,Staring,Posturing/catalepsy,Grimacing,Mannerisms,Rigidity,Negativism,Waxy flexibility,Withdrawal | 99.3 | 98.4 | 100.0 |
| 9 | Immobility/stupor,Staring,Posturing/catalepsy,Grimacing,Stereotypy,Mannerisms,Negativism,Waxy flexibility,Withdrawal | 99.3 | 98.4 | 100.0 |
| 9 | Immobility/stupor,Staring,Posturing/catalepsy,Mannerisms,Verbigeration,Rigidity,Negativism,Waxy flexibility,Withdrawal | 99.3 | 98.4 | 100.0 |
| 9 | Immobility/stupor,Staring,Posturing/catalepsy,Stereotypy,Mannerisms,Verbigeration,Negativism,Waxy flexibility,Withdrawal | 99.3 | 98.4 | 100.0 |
| 9 | Immobility/stupor,Staring,Posturing/catalepsy,Stereotypy,Mannerisms,Verbigeration,Rigidity,Negativism,Waxy flexibility | 99.3 | 98.4 | 100.0 |
| 9 | Mutism,Staring,Echopraxia/echolalia,Mannerisms,Verbigeration,Rigidity,Negativism,Waxy flexibility,Withdrawal | 99.3 | 98.4 | 100.0 |
| 9 | Mutism,Staring,Grimacing,Echopraxia/echolalia,Mannerisms,Verbigeration,Rigidity,Negativism,Waxy flexibility | 99.3 | 98.4 | 100.0 |
| 9 | Mutism,Staring,Grimacing,Echopraxia/echolalia,Stereotypy,Mannerisms,Verbigeration,Negativism,Waxy flexibility | 99.3 | 98.4 | 100.0 |
| 9 | Mutism,Staring,Grimacing,Echopraxia/echolalia,Stereotypy,Mannerisms,Verbigeration,Rigidity,Withdrawal | 99.3 | 98.4 | 100.0 |
| 9 | Mutism,Staring,Grimacing,Echopraxia/echolalia,Stereotypy,Verbigeration,Rigidity,Negativism,Withdrawal | 99.3 | 98.4 | 100.0 |
| 9 | Mutism,Staring,Grimacing,Mannerisms,Verbigeration,Rigidity,Negativism,Waxy flexibility,Withdrawal | 99.3 | 98.4 | 100.0 |
| 9 | Mutism,Staring,Grimacing,Stereotypy,Mannerisms,Verbigeration,Negativism,Waxy flexibility,Withdrawal | 99.3 | 98.4 | 100.0 |
| 9 | Mutism,Staring,Grimacing,Stereotypy,Verbigeration,Rigidity,Negativism,Waxy flexibility,Withdrawal | 99.3 | 98.4 | 100.0 |
| 9 | Mutism,Staring,Posturing/catalepsy,Echopraxia/echolalia,Mannerisms,Verbigeration,Negativism,Waxy flexibility,Withdrawal | 99.3 | 98.4 | 100.0 |
| 9 | Mutism,Staring,Posturing/catalepsy,Echopraxia/echolalia,Mannerisms,Verbigeration,Rigidity,Negativism,Waxy flexibility | 99.3 | 98.4 | 100.0 |
| 9 | Mutism,Staring,Posturing/catalepsy,Echopraxia/echolalia,Stereotypy,Mannerisms,Rigidity,Negativism,Waxy flexibility | 99.3 | 98.4 | 100.0 |
| 9 | Mutism,Staring,Posturing/catalepsy,Echopraxia/echolalia,Stereotypy,Mannerisms,Verbigeration,Negativism,Waxy flexibility | 99.3 | 98.4 | 100.0 |
| 9 | Mutism,Staring,Posturing/catalepsy,Grimacing,Echopraxia/echolalia,Mannerisms,Rigidity,Negativism,Waxy flexibility | 99.3 | 98.4 | 100.0 |
| 9 | Mutism,Staring,Posturing/catalepsy,Grimacing,Echopraxia/echolalia,Mannerisms,Verbigeration,Rigidity,Withdrawal | 99.3 | 98.4 | 100.0 |
| 9 | Mutism,Staring,Posturing/catalepsy,Grimacing,Mannerisms,Rigidity,Negativism,Waxy flexibility,Withdrawal | 99.3 | 98.4 | 100.0 |
| 9 | Mutism,Staring,Posturing/catalepsy,Grimacing,Stereotypy,Mannerisms,Rigidity,Waxy flexibility,Withdrawal | 99.3 | 98.4 | 100.0 |
| 9 | Mutism,Staring,Posturing/catalepsy,Grimacing,Stereotypy,Mannerisms,Verbigeration,Negativism,Waxy flexibility | 99.3 | 98.4 | 100.0 |
| 9 | Mutism,Staring,Posturing/catalepsy,Grimacing,Stereotypy,Verbigeration,Rigidity,Negativism,Withdrawal | 99.3 | 98.4 | 100.0 |
| 9 | Mutism,Staring,Posturing/catalepsy,Mannerisms,Verbigeration,Rigidity,Negativism,Waxy flexibility,Withdrawal | 99.3 | 98.4 | 100.0 |
| 9 | Mutism,Staring,Posturing/catalepsy,Stereotypy,Mannerisms,Verbigeration,Rigidity,Negativism,Waxy flexibility | 99.3 | 98.4 | 100.0 |
| 9 | Staring,Echopraxia/echolalia,Stereotypy,Mannerisms,Verbigeration,Rigidity,Negativism,Waxy flexibility,Withdrawal | 99.3 | 98.4 | 100.0 |
| 9 | Staring,Posturing/catalepsy,Echopraxia/echolalia,Stereotypy,Mannerisms,Rigidity,Negativism,Waxy flexibility,Withdrawal | 99.3 | 98.4 | 100.0 |
| 9 | Staring,Posturing/catalepsy,Echopraxia/echolalia,Stereotypy,Mannerisms,Verbigeration,Rigidity,Negativism,Withdrawal | 99.3 | 98.4 | 100.0 |
| 9 | Staring,Posturing/catalepsy,Grimacing,Echopraxia/echolalia,Mannerisms,Rigidity,Negativism,Waxy flexibility,Withdrawal | 99.3 | 98.4 | 100.0 |
| 9 | Staring,Posturing/catalepsy,Grimacing,Echopraxia/echolalia,Mannerisms,Verbigeration,Rigidity,Negativism,Withdrawal | 99.3 | 98.4 | 100.0 |
| 9 | Staring,Posturing/catalepsy,Grimacing,Stereotypy,Mannerisms,Rigidity,Negativism,Waxy flexibility,Withdrawal | 99.3 | 98.4 | 100.0 |
| 9 | Staring,Posturing/catalepsy,Stereotypy,Mannerisms,Verbigeration,Rigidity,Negativism,Waxy flexibility,Withdrawal | 99.3 | 98.4 | 100.0 |
| 9 | Excitement,Immobility/stupor,Echopraxia/echolalia,Mannerisms,Verbigeration,Rigidity,Negativism,Waxy flexibility,Withdrawal | 99.1 | 98.2 | 99.8 |
| 9 | Excitement,Immobility/stupor,Echopraxia/echolalia,Stereotypy,Mannerisms,Rigidity,Negativism,Waxy flexibility,Withdrawal | 99.1 | 98.2 | 99.8 |
| 9 | Excitement,Immobility/stupor,Grimacing,Echopraxia/echolalia,Mannerisms,Rigidity,Negativism,Waxy flexibility,Withdrawal | 99.1 | 98.2 | 99.8 |
| 9 | Excitement,Immobility/stupor,Grimacing,Echopraxia/echolalia,Stereotypy,Rigidity,Negativism,Waxy flexibility,Withdrawal | 99.1 | 98.2 | 99.8 |
| 9 | Excitement,Immobility/stupor,Grimacing,Echopraxia/echolalia,Verbigeration,Rigidity,Negativism,Waxy flexibility,Withdrawal | 99.1 | 98.2 | 99.8 |
| 9 | Excitement,Immobility/stupor,Grimacing,Stereotypy,Mannerisms,Verbigeration,Rigidity,Negativism,Withdrawal | 99.1 | 98.2 | 99.8 |
| 9 | Excitement,Immobility/stupor,Mutism,Echopraxia/echolalia,Mannerisms,Verbigeration,Rigidity,Negativism,Waxy flexibility | 99.1 | 98.2 | 99.8 |
| 9 | Excitement,Immobility/stupor,Mutism,Echopraxia/echolalia,Stereotypy,Mannerisms,Verbigeration,Rigidity,Withdrawal | 99.1 | 98.2 | 99.8 |
| 9 | Excitement,Immobility/stupor,Mutism,Grimacing,Echopraxia/echolalia,Mannerisms,Verbigeration,Rigidity,Negativism | 99.1 | 98.2 | 99.8 |
| 9 | Excitement,Immobility/stupor,Mutism,Grimacing,Echopraxia/echolalia,Stereotypy,Rigidity,Negativism,Waxy flexibility | 99.1 | 98.2 | 99.8 |
| 9 | Excitement,Immobility/stupor,Mutism,Grimacing,Echopraxia/echolalia,Stereotypy,Rigidity,Waxy flexibility,Withdrawal | 99.1 | 98.2 | 99.8 |
| 9 | Excitement,Immobility/stupor,Mutism,Grimacing,Echopraxia/echolalia,Stereotypy,Verbigeration,Rigidity,Withdrawal | 99.1 | 98.2 | 99.8 |
| 9 | Excitement,Immobility/stupor,Mutism,Grimacing,Echopraxia/echolalia,Verbigeration,Rigidity,Waxy flexibility,Withdrawal | 99.1 | 98.2 | 99.8 |
| 9 | Excitement,Immobility/stupor,Mutism,Grimacing,Mannerisms,Verbigeration,Rigidity,Negativism,Waxy flexibility | 99.1 | 98.2 | 99.8 |
| 9 | Excitement,Immobility/stupor,Mutism,Grimacing,Stereotypy,Mannerisms,Verbigeration,Rigidity,Negativism | 99.1 | 98.2 | 99.8 |
| 9 | Excitement,Immobility/stupor,Mutism,Grimacing,Stereotypy,Rigidity,Negativism,Waxy flexibility,Withdrawal | 99.1 | 98.2 | 99.8 |
| 9 | Excitement,Immobility/stupor,Mutism,Grimacing,Stereotypy,Verbigeration,Rigidity,Waxy flexibility,Withdrawal | 99.1 | 98.2 | 99.8 |
| 9 | Excitement,Immobility/stupor,Mutism,Grimacing,Verbigeration,Rigidity,Negativism,Waxy flexibility,Withdrawal | 99.1 | 98.2 | 99.8 |
| 9 | Excitement,Immobility/stupor,Mutism,Mannerisms,Verbigeration,Rigidity,Negativism,Waxy flexibility,Withdrawal | 99.1 | 98.2 | 99.8 |
| 9 | Excitement,Immobility/stupor,Mutism,Posturing/catalepsy,Echopraxia/echolalia,Stereotypy,Mannerisms,Rigidity,Negativism | 99.1 | 98.2 | 99.8 |
| 9 | Excitement,Immobility/stupor,Mutism,Posturing/catalepsy,Echopraxia/echolalia,Stereotypy,Mannerisms,Rigidity,Withdrawal | 99.1 | 98.2 | 99.8 |
| 9 | Excitement,Immobility/stupor,Mutism,Posturing/catalepsy,Echopraxia/echolalia,Stereotypy,Mannerisms,Verbigeration,Rigidity | 99.1 | 98.2 | 99.8 |
| 9 | Excitement,Immobility/stupor,Mutism,Posturing/catalepsy,Echopraxia/echolalia,Stereotypy,Rigidity,Negativism,Waxy flexibility | 99.1 | 98.2 | 99.8 |
| 9 | Excitement,Immobility/stupor,Mutism,Posturing/catalepsy,Echopraxia/echolalia,Stereotypy,Rigidity,Waxy flexibility,Withdrawal | 99.1 | 98.2 | 99.8 |
| 9 | Excitement,Immobility/stupor,Mutism,Posturing/catalepsy,Echopraxia/echolalia,Stereotypy,Verbigeration,Negativism,Withdrawal | 99.1 | 98.2 | 99.8 |
| 9 | Excitement,Immobility/stupor,Mutism,Posturing/catalepsy,Echopraxia/echolalia,Stereotypy,Verbigeration,Rigidity,Waxy flexibility | 99.1 | 98.2 | 99.8 |
| 9 | Excitement,Immobility/stupor,Mutism,Posturing/catalepsy,Grimacing,Echopraxia/echolalia,Mannerisms,Verbigeration,Rigidity | 99.1 | 98.2 | 99.8 |
| 9 | Excitement,Immobility/stupor,Mutism,Posturing/catalepsy,Grimacing,Echopraxia/echolalia,Stereotypy,Verbigeration,Rigidity | 99.1 | 98.2 | 99.8 |
| 9 | Excitement,Immobility/stupor,Mutism,Posturing/catalepsy,Grimacing,Echopraxia/echolalia,Verbigeration,Negativism,Withdrawal | 99.1 | 98.2 | 99.8 |
| 9 | Excitement,Immobility/stupor,Mutism,Posturing/catalepsy,Grimacing,Echopraxia/echolalia,Verbigeration,Rigidity,Waxy flexibility | 99.1 | 98.2 | 99.8 |
| 9 | Excitement,Immobility/stupor,Mutism,Posturing/catalepsy,Grimacing,Rigidity,Negativism,Waxy flexibility,Withdrawal | 99.1 | 98.2 | 99.8 |
| 9 | Excitement,Immobility/stupor,Mutism,Posturing/catalepsy,Grimacing,Verbigeration,Rigidity,Waxy flexibility,Withdrawal | 99.1 | 98.2 | 99.8 |
| 9 | Excitement,Immobility/stupor,Mutism,Posturing/catalepsy,Mannerisms,Verbigeration,Rigidity,Waxy flexibility,Withdrawal | 99.1 | 98.2 | 99.8 |
| 9 | Excitement,Immobility/stupor,Mutism,Posturing/catalepsy,Stereotypy,Mannerisms,Rigidity,Negativism,Withdrawal | 99.1 | 98.2 | 99.8 |
| 9 | Excitement,Immobility/stupor,Mutism,Posturing/catalepsy,Stereotypy,Rigidity,Negativism,Waxy flexibility,Withdrawal | 99.1 | 98.2 | 99.8 |
| 9 | Excitement,Immobility/stupor,Mutism,Staring,Echopraxia/echolalia,Mannerisms,Verbigeration,Negativism,Withdrawal | 99.1 | 98.2 | 99.8 |
| 9 | Excitement,Immobility/stupor,Mutism,Staring,Echopraxia/echolalia,Stereotypy,Mannerisms,Verbigeration,Negativism | 99.1 | 98.2 | 99.8 |
| 9 | Excitement,Immobility/stupor,Mutism,Staring,Grimacing,Echopraxia/echolalia,Mannerisms,Negativism,Waxy flexibility | 99.1 | 98.2 | 99.8 |
| 9 | Excitement,Immobility/stupor,Mutism,Staring,Grimacing,Echopraxia/echolalia,Mannerisms,Negativism,Withdrawal | 99.1 | 98.2 | 99.8 |
| 9 | Excitement,Immobility/stupor,Mutism,Staring,Grimacing,Echopraxia/echolalia,Mannerisms,Verbigeration,Rigidity | 99.1 | 98.2 | 99.8 |
| 9 | Excitement,Immobility/stupor,Mutism,Staring,Grimacing,Echopraxia/echolalia,Stereotypy,Mannerisms,Rigidity | 99.1 | 98.2 | 99.8 |
| 9 | Excitement,Immobility/stupor,Mutism,Staring,Grimacing,Echopraxia/echolalia,Stereotypy,Verbigeration,Negativism | 99.1 | 98.2 | 99.8 |
| 9 | Excitement,Immobility/stupor,Mutism,Staring,Grimacing,Echopraxia/echolalia,Stereotypy,Waxy flexibility,Withdrawal | 99.1 | 98.2 | 99.8 |
| 9 | Excitement,Immobility/stupor,Mutism,Staring,Grimacing,Echopraxia/echolalia,Verbigeration,Negativism,Withdrawal | 99.1 | 98.2 | 99.8 |
| 9 | Excitement,Immobility/stupor,Mutism,Staring,Grimacing,Mannerisms,Verbigeration,Negativism,Waxy flexibility | 99.1 | 98.2 | 99.8 |
| 9 | Excitement,Immobility/stupor,Mutism,Staring,Grimacing,Stereotypy,Mannerisms,Rigidity,Negativism | 99.1 | 98.2 | 99.8 |
| 9 | Excitement,Immobility/stupor,Mutism,Staring,Grimacing,Stereotypy,Negativism,Waxy flexibility,Withdrawal | 99.1 | 98.2 | 99.8 |
| 9 | Excitement,Immobility/stupor,Mutism,Staring,Mannerisms,Verbigeration,Negativism,Waxy flexibility,Withdrawal | 99.1 | 98.2 | 99.8 |
| 9 | Excitement,Immobility/stupor,Mutism,Staring,Posturing/catalepsy,Echopraxia/echolalia,Mannerisms,Verbigeration,Waxy flexibility | 99.1 | 98.2 | 99.8 |
| 9 | Excitement,Immobility/stupor,Mutism,Staring,Posturing/catalepsy,Echopraxia/echolalia,Mannerisms,Verbigeration,Withdrawal | 99.1 | 98.2 | 99.8 |
| 9 | Excitement,Immobility/stupor,Mutism,Staring,Posturing/catalepsy,Echopraxia/echolalia,Stereotypy,Mannerisms,Withdrawal | 99.1 | 98.2 | 99.8 |
| 9 | Excitement,Immobility/stupor,Mutism,Staring,Posturing/catalepsy,Echopraxia/echolalia,Stereotypy,Verbigeration,Waxy flexibility | 99.1 | 98.2 | 99.8 |
| 9 | Excitement,Immobility/stupor,Mutism,Staring,Posturing/catalepsy,Echopraxia/echolalia,Stereotypy,Verbigeration,Withdrawal | 99.1 | 98.2 | 99.8 |
| 9 | Excitement,Immobility/stupor,Mutism,Staring,Posturing/catalepsy,Grimacing,Echopraxia/echolalia,Mannerisms,Waxy flexibility | 99.1 | 98.2 | 99.8 |
| 9 | Excitement,Immobility/stupor,Mutism,Staring,Posturing/catalepsy,Grimacing,Echopraxia/echolalia,Stereotypy,Waxy flexibility | 99.1 | 98.2 | 99.8 |
| 9 | Excitement,Immobility/stupor,Mutism,Staring,Posturing/catalepsy,Grimacing,Mannerisms,Verbigeration,Waxy flexibility | 99.1 | 98.2 | 99.8 |
| 9 | Excitement,Immobility/stupor,Mutism,Staring,Posturing/catalepsy,Grimacing,Mannerisms,Verbigeration,Withdrawal | 99.1 | 98.2 | 99.8 |
| 9 | Excitement,Immobility/stupor,Mutism,Staring,Posturing/catalepsy,Grimacing,Mannerisms,Waxy flexibility,Withdrawal | 99.1 | 98.2 | 99.8 |
| 9 | Excitement,Immobility/stupor,Mutism,Staring,Posturing/catalepsy,Grimacing,Stereotypy,Verbigeration,Waxy flexibility | 99.1 | 98.2 | 99.8 |
| 9 | Excitement,Immobility/stupor,Mutism,Staring,Posturing/catalepsy,Grimacing,Stereotypy,Verbigeration,Withdrawal | 99.1 | 98.2 | 99.8 |
| 9 | Excitement,Immobility/stupor,Mutism,Staring,Posturing/catalepsy,Grimacing,Stereotypy,Waxy flexibility,Withdrawal | 99.1 | 98.2 | 99.8 |
| 9 | Excitement,Immobility/stupor,Mutism,Staring,Posturing/catalepsy,Grimacing,Verbigeration,Waxy flexibility,Withdrawal | 99.1 | 98.2 | 99.8 |
| 9 | Excitement,Immobility/stupor,Mutism,Staring,Posturing/catalepsy,Mannerisms,Verbigeration,Waxy flexibility,Withdrawal | 99.1 | 98.2 | 99.8 |
| 9 | Excitement,Immobility/stupor,Mutism,Staring,Posturing/catalepsy,Stereotypy,Verbigeration,Waxy flexibility,Withdrawal | 99.1 | 98.2 | 99.8 |
| 9 | Excitement,Immobility/stupor,Mutism,Staring,Stereotypy,Mannerisms,Negativism,Waxy flexibility,Withdrawal | 99.1 | 98.2 | 99.8 |
| 9 | Excitement,Immobility/stupor,Mutism,Stereotypy,Mannerisms,Verbigeration,Rigidity,Waxy flexibility,Withdrawal | 99.1 | 98.2 | 99.8 |
| 9 | Excitement,Immobility/stupor,Posturing/catalepsy,Echopraxia/echolalia,Mannerisms,Rigidity,Negativism,Waxy flexibility,Withdrawal | 99.1 | 98.2 | 99.8 |
| 9 | Excitement,Immobility/stupor,Posturing/catalepsy,Echopraxia/echolalia,Stereotypy,Mannerisms,Verbigeration,Rigidity,Withdrawal | 99.1 | 98.2 | 99.8 |
| 9 | Excitement,Immobility/stupor,Posturing/catalepsy,Echopraxia/echolalia,Stereotypy,Rigidity,Negativism,Waxy flexibility,Withdrawal | 99.1 | 98.2 | 99.8 |
| 9 | Excitement,Immobility/stupor,Posturing/catalepsy,Grimacing,Echopraxia/echolalia,Mannerisms,Verbigeration,Rigidity,Negativism | 99.1 | 98.2 | 99.8 |
| 9 | Excitement,Immobility/stupor,Posturing/catalepsy,Grimacing,Echopraxia/echolalia,Mannerisms,Verbigeration,Rigidity,Withdrawal | 99.1 | 98.2 | 99.8 |
| 9 | Excitement,Immobility/stupor,Posturing/catalepsy,Grimacing,Echopraxia/echolalia,Verbigeration,Rigidity,Negativism,Waxy flexibility | 99.1 | 98.2 | 99.8 |
| 9 | Excitement,Immobility/stupor,Posturing/catalepsy,Grimacing,Mannerisms,Rigidity,Negativism,Waxy flexibility,Withdrawal | 99.1 | 98.2 | 99.8 |
| 9 | Excitement,Immobility/stupor,Posturing/catalepsy,Grimacing,Mannerisms,Verbigeration,Rigidity,Negativism,Waxy flexibility | 99.1 | 98.2 | 99.8 |
| 9 | Excitement,Immobility/stupor,Posturing/catalepsy,Grimacing,Stereotypy,Mannerisms,Rigidity,Negativism,Withdrawal | 99.1 | 98.2 | 99.8 |
| 9 | Excitement,Immobility/stupor,Posturing/catalepsy,Grimacing,Verbigeration,Rigidity,Negativism,Waxy flexibility,Withdrawal | 99.1 | 98.2 | 99.8 |
| 9 | Excitement,Immobility/stupor,Staring,Echopraxia/echolalia,Mannerisms,Verbigeration,Negativism,Waxy flexibility,Withdrawal | 99.1 | 98.2 | 99.8 |
| 9 | Excitement,Immobility/stupor,Staring,Echopraxia/echolalia,Stereotypy,Mannerisms,Verbigeration,Negativism,Waxy flexibility | 99.1 | 98.2 | 99.8 |
| 9 | Excitement,Immobility/stupor,Staring,Echopraxia/echolalia,Stereotypy,Verbigeration,Negativism,Waxy flexibility,Withdrawal | 99.1 | 98.2 | 99.8 |
| 9 | Excitement,Immobility/stupor,Staring,Grimacing,Echopraxia/echolalia,Mannerisms,Negativism,Waxy flexibility,Withdrawal | 99.1 | 98.2 | 99.8 |
| 9 | Excitement,Immobility/stupor,Staring,Grimacing,Echopraxia/echolalia,Mannerisms,Verbigeration,Rigidity,Waxy flexibility | 99.1 | 98.2 | 99.8 |
| 9 | Excitement,Immobility/stupor,Staring,Grimacing,Echopraxia/echolalia,Stereotypy,Mannerisms,Rigidity,Waxy flexibility | 99.1 | 98.2 | 99.8 |
| 9 | Excitement,Immobility/stupor,Staring,Grimacing,Echopraxia/echolalia,Stereotypy,Negativism,Waxy flexibility,Withdrawal | 99.1 | 98.2 | 99.8 |
| 9 | Excitement,Immobility/stupor,Staring,Grimacing,Echopraxia/echolalia,Stereotypy,Verbigeration,Negativism,Withdrawal | 99.1 | 98.2 | 99.8 |
| 9 | Excitement,Immobility/stupor,Staring,Grimacing,Echopraxia/echolalia,Stereotypy,Verbigeration,Rigidity,Waxy flexibility | 99.1 | 98.2 | 99.8 |
| 9 | Excitement,Immobility/stupor,Staring,Grimacing,Stereotypy,Mannerisms,Verbigeration,Negativism,Withdrawal | 99.1 | 98.2 | 99.8 |
| 9 | Excitement,Immobility/stupor,Staring,Posturing/catalepsy,Echopraxia/echolalia,Stereotypy,Verbigeration,Rigidity,Waxy flexibility | 99.1 | 98.2 | 99.8 |
| 9 | Excitement,Immobility/stupor,Staring,Posturing/catalepsy,Grimacing,Echopraxia/echolalia,Mannerisms,Rigidity,Waxy flexibility | 99.1 | 98.2 | 99.8 |
| 9 | Excitement,Immobility/stupor,Staring,Posturing/catalepsy,Grimacing,Echopraxia/echolalia,Mannerisms,Waxy flexibility,Withdrawal | 99.1 | 98.2 | 99.8 |
| 9 | Excitement,Immobility/stupor,Staring,Posturing/catalepsy,Grimacing,Echopraxia/echolalia,Stereotypy,Mannerisms,Negativism | 99.1 | 98.2 | 99.8 |
| 9 | Excitement,Immobility/stupor,Staring,Posturing/catalepsy,Grimacing,Echopraxia/echolalia,Stereotypy,Mannerisms,Rigidity | 99.1 | 98.2 | 99.8 |
| 9 | Excitement,Immobility/stupor,Staring,Posturing/catalepsy,Grimacing,Echopraxia/echolalia,Stereotypy,Negativism,Waxy flexibility | 99.1 | 98.2 | 99.8 |
| 9 | Excitement,Immobility/stupor,Staring,Posturing/catalepsy,Grimacing,Echopraxia/echolalia,Stereotypy,Verbigeration,Negativism | 99.1 | 98.2 | 99.8 |
| 9 | Excitement,Immobility/stupor,Staring,Posturing/catalepsy,Grimacing,Echopraxia/echolalia,Stereotypy,Verbigeration,Rigidity | 99.1 | 98.2 | 99.8 |
| 9 | Excitement,Immobility/stupor,Staring,Posturing/catalepsy,Grimacing,Echopraxia/echolalia,Verbigeration,Rigidity,Waxy flexibility | 99.1 | 98.2 | 99.8 |
| 9 | Excitement,Immobility/stupor,Staring,Posturing/catalepsy,Grimacing,Stereotypy,Negativism,Waxy flexibility,Withdrawal | 99.1 | 98.2 | 99.8 |
| 9 | Excitement,Immobility/stupor,Staring,Posturing/catalepsy,Grimacing,Stereotypy,Rigidity,Negativism,Waxy flexibility | 99.1 | 98.2 | 99.8 |
| 9 | Excitement,Immobility/stupor,Staring,Posturing/catalepsy,Grimacing,Stereotypy,Verbigeration,Rigidity,Waxy flexibility | 99.1 | 98.2 | 99.8 |
| 9 | Excitement,Immobility/stupor,Staring,Stereotypy,Mannerisms,Verbigeration,Negativism,Waxy flexibility,Withdrawal | 99.1 | 98.2 | 99.8 |
| 9 | Excitement,Immobility/stupor,Stereotypy,Mannerisms,Verbigeration,Rigidity,Negativism,Waxy flexibility,Withdrawal | 99.1 | 98.2 | 99.8 |
| 9 | Excitement,Mutism,Staring,Grimacing,Echopraxia/echolalia,Mannerisms,Verbigeration,Negativism,Withdrawal | 99.1 | 98.2 | 99.8 |
| 9 | Excitement,Mutism,Staring,Grimacing,Echopraxia/echolalia,Stereotypy,Mannerisms,Verbigeration,Negativism | 99.1 | 98.2 | 99.8 |
| 9 | Excitement,Mutism,Staring,Grimacing,Echopraxia/echolalia,Stereotypy,Mannerisms,Waxy flexibility,Withdrawal | 99.1 | 98.2 | 99.8 |
| 9 | Excitement,Mutism,Staring,Grimacing,Echopraxia/echolalia,Stereotypy,Verbigeration,Waxy flexibility,Withdrawal | 99.1 | 98.2 | 99.8 |
| 9 | Excitement,Mutism,Staring,Grimacing,Mannerisms,Verbigeration,Negativism,Waxy flexibility,Withdrawal | 99.1 | 98.2 | 99.8 |
| 9 | Excitement,Mutism,Staring,Grimacing,Stereotypy,Mannerisms,Negativism,Waxy flexibility,Withdrawal | 99.1 | 98.2 | 99.8 |
| 9 | Excitement,Mutism,Staring,Grimacing,Stereotypy,Mannerisms,Verbigeration,Negativism,Waxy flexibility | 99.1 | 98.2 | 99.8 |
| 9 | Excitement,Mutism,Staring,Posturing/catalepsy,Echopraxia/echolalia,Stereotypy,Mannerisms,Verbigeration,Waxy flexibility | 99.1 | 98.2 | 99.8 |
| 9 | Excitement,Mutism,Staring,Posturing/catalepsy,Echopraxia/echolalia,Stereotypy,Mannerisms,Verbigeration,Withdrawal | 99.1 | 98.2 | 99.8 |
| 9 | Excitement,Mutism,Staring,Posturing/catalepsy,Grimacing,Echopraxia/echolalia,Mannerisms,Verbigeration,Waxy flexibility | 99.1 | 98.2 | 99.8 |
| 9 | Excitement,Mutism,Staring,Posturing/catalepsy,Grimacing,Echopraxia/echolalia,Stereotypy,Mannerisms,Waxy flexibility | 99.1 | 98.2 | 99.8 |
| 9 | Excitement,Mutism,Staring,Posturing/catalepsy,Grimacing,Stereotypy,Mannerisms,Rigidity,Waxy flexibility | 99.1 | 98.2 | 99.8 |
| 9 | Excitement,Mutism,Staring,Posturing/catalepsy,Grimacing,Stereotypy,Mannerisms,Verbigeration,Withdrawal | 99.1 | 98.2 | 99.8 |
| 9 | Excitement,Mutism,Staring,Posturing/catalepsy,Grimacing,Stereotypy,Mannerisms,Waxy flexibility,Withdrawal | 99.1 | 98.2 | 99.8 |
| 9 | Excitement,Mutism,Staring,Posturing/catalepsy,Stereotypy,Mannerisms,Verbigeration,Waxy flexibility,Withdrawal | 99.1 | 98.2 | 99.8 |
| 9 | Excitement,Staring,Echopraxia/echolalia,Stereotypy,Mannerisms,Verbigeration,Rigidity,Waxy flexibility,Withdrawal | 99.1 | 98.2 | 99.8 |
| 9 | Excitement,Staring,Echopraxia/echolalia,Stereotypy,Verbigeration,Rigidity,Negativism,Waxy flexibility,Withdrawal | 99.1 | 98.2 | 99.8 |
| 9 | Excitement,Staring,Grimacing,Echopraxia/echolalia,Mannerisms,Verbigeration,Rigidity,Negativism,Withdrawal | 99.1 | 98.2 | 99.8 |
| 9 | Excitement,Staring,Grimacing,Echopraxia/echolalia,Mannerisms,Verbigeration,Rigidity,Waxy flexibility,Withdrawal | 99.1 | 98.2 | 99.8 |
| 9 | Excitement,Staring,Grimacing,Echopraxia/echolalia,Stereotypy,Mannerisms,Verbigeration,Rigidity,Withdrawal | 99.1 | 98.2 | 99.8 |
| 9 | Excitement,Staring,Grimacing,Echopraxia/echolalia,Stereotypy,Rigidity,Negativism,Waxy flexibility,Withdrawal | 99.1 | 98.2 | 99.8 |
| 9 | Excitement,Staring,Grimacing,Echopraxia/echolalia,Stereotypy,Verbigeration,Rigidity,Negativism,Withdrawal | 99.1 | 98.2 | 99.8 |
| 9 | Excitement,Staring,Grimacing,Stereotypy,Verbigeration,Rigidity,Negativism,Waxy flexibility,Withdrawal | 99.1 | 98.2 | 99.8 |
| 9 | Excitement,Staring,Posturing/catalepsy,Echopraxia/echolalia,Mannerisms,Verbigeration,Negativism,Waxy flexibility,Withdrawal | 99.1 | 98.2 | 99.8 |
| 9 | Excitement,Staring,Posturing/catalepsy,Echopraxia/echolalia,Mannerisms,Verbigeration,Rigidity,Waxy flexibility,Withdrawal | 99.1 | 98.2 | 99.8 |
| 9 | Excitement,Staring,Posturing/catalepsy,Echopraxia/echolalia,Stereotypy,Mannerisms,Negativism,Waxy flexibility,Withdrawal | 99.1 | 98.2 | 99.8 |
| 9 | Excitement,Staring,Posturing/catalepsy,Echopraxia/echolalia,Stereotypy,Mannerisms,Rigidity,Waxy flexibility,Withdrawal | 99.1 | 98.2 | 99.8 |
| 9 | Excitement,Staring,Posturing/catalepsy,Echopraxia/echolalia,Stereotypy,Mannerisms,Verbigeration,Rigidity,Withdrawal | 99.1 | 98.2 | 99.8 |
| 9 | Excitement,Staring,Posturing/catalepsy,Echopraxia/echolalia,Stereotypy,Rigidity,Negativism,Waxy flexibility,Withdrawal | 99.1 | 98.2 | 99.8 |
| 9 | Excitement,Staring,Posturing/catalepsy,Echopraxia/echolalia,Stereotypy,Verbigeration,Rigidity,Negativism,Withdrawal | 99.1 | 98.2 | 99.8 |
| 9 | Excitement,Staring,Posturing/catalepsy,Echopraxia/echolalia,Verbigeration,Rigidity,Negativism,Waxy flexibility,Withdrawal | 99.1 | 98.2 | 99.8 |
| 9 | Excitement,Staring,Posturing/catalepsy,Grimacing,Echopraxia/echolalia,Mannerisms,Negativism,Waxy flexibility,Withdrawal | 99.1 | 98.2 | 99.8 |
| 9 | Excitement,Staring,Posturing/catalepsy,Grimacing,Echopraxia/echolalia,Mannerisms,Verbigeration,Negativism,Withdrawal | 99.1 | 98.2 | 99.8 |
| 9 | Excitement,Staring,Posturing/catalepsy,Grimacing,Echopraxia/echolalia,Rigidity,Negativism,Waxy flexibility,Withdrawal | 99.1 | 98.2 | 99.8 |
| 9 | Excitement,Staring,Posturing/catalepsy,Grimacing,Echopraxia/echolalia,Stereotypy,Mannerisms,Negativism,Withdrawal | 99.1 | 98.2 | 99.8 |
| 9 | Excitement,Staring,Posturing/catalepsy,Grimacing,Echopraxia/echolalia,Stereotypy,Rigidity,Negativism,Withdrawal | 99.1 | 98.2 | 99.8 |
| 9 | Excitement,Staring,Posturing/catalepsy,Grimacing,Echopraxia/echolalia,Verbigeration,Rigidity,Negativism,Withdrawal | 99.1 | 98.2 | 99.8 |
| 9 | Excitement,Staring,Posturing/catalepsy,Grimacing,Mannerisms,Verbigeration,Negativism,Waxy flexibility,Withdrawal | 99.1 | 98.2 | 99.8 |
| 9 | Excitement,Staring,Posturing/catalepsy,Grimacing,Stereotypy,Mannerisms,Rigidity,Waxy flexibility,Withdrawal | 99.1 | 98.2 | 99.8 |
| 9 | Excitement,Staring,Posturing/catalepsy,Grimacing,Stereotypy,Mannerisms,Verbigeration,Negativism,Withdrawal | 99.1 | 98.2 | 99.8 |
| 9 | Excitement,Staring,Posturing/catalepsy,Grimacing,Stereotypy,Verbigeration,Rigidity,Negativism,Withdrawal | 99.1 | 98.2 | 99.8 |
| 9 | Excitement,Staring,Posturing/catalepsy,Stereotypy,Mannerisms,Verbigeration,Negativism,Waxy flexibility,Withdrawal | 99.1 | 98.2 | 99.8 |
| 9 | Excitement,Staring,Posturing/catalepsy,Stereotypy,Mannerisms,Verbigeration,Rigidity,Waxy flexibility,Withdrawal | 99.1 | 98.2 | 99.8 |
| 9 | Excitement,Staring,Posturing/catalepsy,Stereotypy,Verbigeration,Rigidity,Negativism,Waxy flexibility,Withdrawal | 99.1 | 98.2 | 99.8 |
| 9 | Immobility/stupor,Echopraxia/echolalia,Stereotypy,Mannerisms,Verbigeration,Rigidity,Negativism,Waxy flexibility,Withdrawal | 99.1 | 98.2 | 99.8 |
| 9 | Immobility/stupor,Grimacing,Echopraxia/echolalia,Stereotypy,Mannerisms,Rigidity,Negativism,Waxy flexibility,Withdrawal | 99.1 | 98.2 | 99.8 |
| 9 | Immobility/stupor,Grimacing,Echopraxia/echolalia,Stereotypy,Mannerisms,Verbigeration,Rigidity,Negativism,Withdrawal | 99.1 | 98.2 | 99.8 |
| 9 | Immobility/stupor,Mutism,Echopraxia/echolalia,Stereotypy,Mannerisms,Verbigeration,Rigidity,Negativism,Withdrawal | 99.1 | 98.2 | 99.8 |
| 9 | Immobility/stupor,Mutism,Grimacing,Echopraxia/echolalia,Stereotypy,Mannerisms,Rigidity,Negativism,Withdrawal | 99.1 | 98.2 | 99.8 |
| 9 | Immobility/stupor,Mutism,Grimacing,Stereotypy,Mannerisms,Verbigeration,Rigidity,Negativism,Withdrawal | 99.1 | 98.2 | 99.8 |
| 9 | Immobility/stupor,Mutism,Posturing/catalepsy,Grimacing,Echopraxia/echolalia,Mannerisms,Rigidity,Negativism,Withdrawal | 99.1 | 98.2 | 99.8 |
| 9 | Immobility/stupor,Mutism,Posturing/catalepsy,Stereotypy,Mannerisms,Verbigeration,Rigidity,Negativism,Withdrawal | 99.1 | 98.2 | 99.8 |
| 9 | Immobility/stupor,Mutism,Staring,Echopraxia/echolalia,Mannerisms,Verbigeration,Rigidity,Negativism,Waxy flexibility | 99.1 | 98.2 | 99.8 |
| 9 | Immobility/stupor,Mutism,Staring,Echopraxia/echolalia,Stereotypy,Mannerisms,Negativism,Waxy flexibility,Withdrawal | 99.1 | 98.2 | 99.8 |
| 9 | Immobility/stupor,Mutism,Staring,Echopraxia/echolalia,Stereotypy,Mannerisms,Verbigeration,Negativism,Withdrawal | 99.1 | 98.2 | 99.8 |
| 9 | Immobility/stupor,Mutism,Staring,Echopraxia/echolalia,Stereotypy,Verbigeration,Rigidity,Negativism,Withdrawal | 99.1 | 98.2 | 99.8 |
| 9 | Immobility/stupor,Mutism,Staring,Grimacing,Echopraxia/echolalia,Mannerisms,Negativism,Waxy flexibility,Withdrawal | 99.1 | 98.2 | 99.8 |
| 9 | Immobility/stupor,Mutism,Staring,Grimacing,Echopraxia/echolalia,Mannerisms,Rigidity,Negativism,Waxy flexibility | 99.1 | 98.2 | 99.8 |
| 9 | Immobility/stupor,Mutism,Staring,Grimacing,Echopraxia/echolalia,Mannerisms,Verbigeration,Rigidity,Negativism | 99.1 | 98.2 | 99.8 |
| 9 | Immobility/stupor,Mutism,Staring,Grimacing,Echopraxia/echolalia,Stereotypy,Mannerisms,Negativism,Waxy flexibility | 99.1 | 98.2 | 99.8 |
| 9 | Immobility/stupor,Mutism,Staring,Grimacing,Mannerisms,Rigidity,Negativism,Waxy flexibility,Withdrawal | 99.1 | 98.2 | 99.8 |
| 9 | Immobility/stupor,Mutism,Staring,Grimacing,Stereotypy,Mannerisms,Negativism,Waxy flexibility,Withdrawal | 99.1 | 98.2 | 99.8 |
| 9 | Immobility/stupor,Mutism,Staring,Grimacing,Stereotypy,Mannerisms,Verbigeration,Rigidity,Waxy flexibility | 99.1 | 98.2 | 99.8 |
| 9 | Immobility/stupor,Mutism,Staring,Grimacing,Stereotypy,Verbigeration,Rigidity,Negativism,Waxy flexibility | 99.1 | 98.2 | 99.8 |
| 9 | Immobility/stupor,Mutism,Staring,Mannerisms,Verbigeration,Rigidity,Negativism,Waxy flexibility,Withdrawal | 99.1 | 98.2 | 99.8 |
| 9 | Immobility/stupor,Mutism,Staring,Posturing/catalepsy,Echopraxia/echolalia,Mannerisms,Rigidity,Negativism,Waxy flexibility | 99.1 | 98.2 | 99.8 |
| 9 | Immobility/stupor,Mutism,Staring,Posturing/catalepsy,Echopraxia/echolalia,Stereotypy,Mannerisms,Negativism,Waxy flexibility | 99.1 | 98.2 | 99.8 |
| 9 | Immobility/stupor,Mutism,Staring,Posturing/catalepsy,Echopraxia/echolalia,Stereotypy,Mannerisms,Rigidity,Withdrawal | 99.1 | 98.2 | 99.8 |
| 9 | Immobility/stupor,Mutism,Staring,Posturing/catalepsy,Echopraxia/echolalia,Stereotypy,Mannerisms,Verbigeration,Negativism | 99.1 | 98.2 | 99.8 |
| 9 | Immobility/stupor,Mutism,Staring,Posturing/catalepsy,Grimacing,Echopraxia/echolalia,Mannerisms,Negativism,Waxy flexibility | 99.1 | 98.2 | 99.8 |
| 9 | Immobility/stupor,Mutism,Staring,Posturing/catalepsy,Grimacing,Echopraxia/echolalia,Mannerisms,Rigidity,Withdrawal | 99.1 | 98.2 | 99.8 |
| 9 | Immobility/stupor,Mutism,Staring,Posturing/catalepsy,Grimacing,Mannerisms,Verbigeration,Rigidity,Negativism | 99.1 | 98.2 | 99.8 |
| 9 | Immobility/stupor,Mutism,Staring,Posturing/catalepsy,Grimacing,Mannerisms,Verbigeration,Rigidity,Withdrawal | 99.1 | 98.2 | 99.8 |
| 9 | Immobility/stupor,Mutism,Staring,Posturing/catalepsy,Grimacing,Stereotypy,Mannerisms,Negativism,Waxy flexibility | 99.1 | 98.2 | 99.8 |
| 9 | Immobility/stupor,Mutism,Staring,Posturing/catalepsy,Grimacing,Stereotypy,Mannerisms,Verbigeration,Rigidity | 99.1 | 98.2 | 99.8 |
| 9 | Immobility/stupor,Mutism,Staring,Posturing/catalepsy,Grimacing,Stereotypy,Verbigeration,Negativism,Withdrawal | 99.1 | 98.2 | 99.8 |
| 9 | Immobility/stupor,Mutism,Staring,Posturing/catalepsy,Grimacing,Stereotypy,Verbigeration,Rigidity,Negativism | 99.1 | 98.2 | 99.8 |
| 9 | Immobility/stupor,Mutism,Staring,Posturing/catalepsy,Mannerisms,Rigidity,Negativism,Waxy flexibility,Withdrawal | 99.1 | 98.2 | 99.8 |
| 9 | Immobility/stupor,Mutism,Staring,Posturing/catalepsy,Mannerisms,Verbigeration,Negativism,Waxy flexibility,Withdrawal | 99.1 | 98.2 | 99.8 |
| 9 | Immobility/stupor,Mutism,Staring,Posturing/catalepsy,Stereotypy,Mannerisms,Rigidity,Negativism,Waxy flexibility | 99.1 | 98.2 | 99.8 |
| 9 | Immobility/stupor,Mutism,Staring,Posturing/catalepsy,Stereotypy,Mannerisms,Verbigeration,Negativism,Waxy flexibility | 99.1 | 98.2 | 99.8 |
| 9 | Immobility/stupor,Mutism,Staring,Posturing/catalepsy,Stereotypy,Mannerisms,Verbigeration,Rigidity,Withdrawal | 99.1 | 98.2 | 99.8 |
| 9 | Immobility/stupor,Mutism,Staring,Posturing/catalepsy,Stereotypy,Verbigeration,Rigidity,Negativism,Withdrawal | 99.1 | 98.2 | 99.8 |
| 9 | Immobility/stupor,Mutism,Staring,Stereotypy,Verbigeration,Rigidity,Negativism,Waxy flexibility,Withdrawal | 99.1 | 98.2 | 99.8 |
| 9 | Immobility/stupor,Mutism,Stereotypy,Mannerisms,Verbigeration,Rigidity,Negativism,Waxy flexibility,Withdrawal | 99.1 | 98.2 | 99.8 |
| 9 | Immobility/stupor,Posturing/catalepsy,Echopraxia/echolalia,Stereotypy,Mannerisms,Verbigeration,Rigidity,Negativism,Withdrawal | 99.1 | 98.2 | 99.8 |
| 9 | Immobility/stupor,Posturing/catalepsy,Grimacing,Echopraxia/echolalia,Mannerisms,Rigidity,Negativism,Waxy flexibility,Withdrawal | 99.1 | 98.2 | 99.8 |
| 9 | Immobility/stupor,Posturing/catalepsy,Grimacing,Echopraxia/echolalia,Mannerisms,Verbigeration,Rigidity,Negativism,Withdrawal | 99.1 | 98.2 | 99.8 |
| 9 | Immobility/stupor,Posturing/catalepsy,Grimacing,Echopraxia/echolalia,Stereotypy,Mannerisms,Rigidity,Negativism,Withdrawal | 99.1 | 98.2 | 99.8 |
| 9 | Immobility/stupor,Posturing/catalepsy,Grimacing,Stereotypy,Mannerisms,Verbigeration,Rigidity,Negativism,Withdrawal | 99.1 | 98.2 | 99.8 |
| 9 | Immobility/stupor,Posturing/catalepsy,Stereotypy,Mannerisms,Verbigeration,Rigidity,Negativism,Waxy flexibility,Withdrawal | 99.1 | 98.2 | 99.8 |
| 9 | Immobility/stupor,Staring,Grimacing,Echopraxia/echolalia,Mannerisms,Verbigeration,Rigidity,Negativism,Waxy flexibility | 99.1 | 98.2 | 99.8 |
| 9 | Immobility/stupor,Staring,Grimacing,Echopraxia/echolalia,Stereotypy,Mannerisms,Rigidity,Waxy flexibility,Withdrawal | 99.1 | 98.2 | 99.8 |
| 9 | Immobility/stupor,Staring,Grimacing,Echopraxia/echolalia,Stereotypy,Verbigeration,Rigidity,Negativism,Withdrawal | 99.1 | 98.2 | 99.8 |
| 9 | Immobility/stupor,Staring,Grimacing,Stereotypy,Mannerisms,Verbigeration,Negativism,Waxy flexibility,Withdrawal | 99.1 | 98.2 | 99.8 |
| 9 | Immobility/stupor,Staring,Grimacing,Stereotypy,Mannerisms,Verbigeration,Rigidity,Waxy flexibility,Withdrawal | 99.1 | 98.2 | 99.8 |
| 9 | Immobility/stupor,Staring,Grimacing,Stereotypy,Verbigeration,Rigidity,Negativism,Waxy flexibility,Withdrawal | 99.1 | 98.2 | 99.8 |
| 9 | Immobility/stupor,Staring,Posturing/catalepsy,Echopraxia/echolalia,Mannerisms,Verbigeration,Rigidity,Negativism,Waxy flexibility | 99.1 | 98.2 | 99.8 |
| 9 | Immobility/stupor,Staring,Posturing/catalepsy,Echopraxia/echolalia,Stereotypy,Mannerisms,Rigidity,Negativism,Waxy flexibility | 99.1 | 98.2 | 99.8 |
| 9 | Immobility/stupor,Staring,Posturing/catalepsy,Grimacing,Echopraxia/echolalia,Mannerisms,Rigidity,Negativism,Waxy flexibility | 99.1 | 98.2 | 99.8 |
| 9 | Immobility/stupor,Staring,Posturing/catalepsy,Grimacing,Echopraxia/echolalia,Stereotypy,Mannerisms,Negativism,Waxy flexibility | 99.1 | 98.2 | 99.8 |
| 9 | Immobility/stupor,Staring,Posturing/catalepsy,Grimacing,Echopraxia/echolalia,Stereotypy,Mannerisms,Rigidity,Withdrawal | 99.1 | 98.2 | 99.8 |
| 9 | Immobility/stupor,Staring,Posturing/catalepsy,Grimacing,Echopraxia/echolalia,Stereotypy,Mannerisms,Verbigeration,Negativism | 99.1 | 98.2 | 99.8 |
| 9 | Immobility/stupor,Staring,Posturing/catalepsy,Grimacing,Mannerisms,Verbigeration,Negativism,Waxy flexibility,Withdrawal | 99.1 | 98.2 | 99.8 |
| 9 | Immobility/stupor,Staring,Posturing/catalepsy,Grimacing,Stereotypy,Mannerisms,Rigidity,Negativism,Waxy flexibility | 99.1 | 98.2 | 99.8 |
| 9 | Immobility/stupor,Staring,Posturing/catalepsy,Grimacing,Stereotypy,Mannerisms,Verbigeration,Negativism,Waxy flexibility | 99.1 | 98.2 | 99.8 |
| 9 | Immobility/stupor,Staring,Posturing/catalepsy,Grimacing,Stereotypy,Mannerisms,Verbigeration,Rigidity,Withdrawal | 99.1 | 98.2 | 99.8 |
| 9 | Immobility/stupor,Staring,Posturing/catalepsy,Grimacing,Stereotypy,Verbigeration,Rigidity,Negativism,Withdrawal | 99.1 | 98.2 | 99.8 |
| 9 | Mutism,Staring,Echopraxia/echolalia,Stereotypy,Mannerisms,Verbigeration,Negativism,Waxy flexibility,Withdrawal | 99.1 | 98.2 | 99.8 |
| 9 | Mutism,Staring,Echopraxia/echolalia,Stereotypy,Verbigeration,Rigidity,Negativism,Waxy flexibility,Withdrawal | 99.1 | 98.2 | 99.8 |
| 9 | Mutism,Staring,Grimacing,Echopraxia/echolalia,Mannerisms,Verbigeration,Negativism,Waxy flexibility,Withdrawal | 99.1 | 98.2 | 99.8 |
| 9 | Mutism,Staring,Grimacing,Echopraxia/echolalia,Mannerisms,Verbigeration,Rigidity,Waxy flexibility,Withdrawal | 99.1 | 98.2 | 99.8 |
| 9 | Mutism,Staring,Grimacing,Echopraxia/echolalia,Stereotypy,Mannerisms,Verbigeration,Rigidity,Waxy flexibility | 99.1 | 98.2 | 99.8 |
| 9 | Mutism,Staring,Grimacing,Echopraxia/echolalia,Stereotypy,Verbigeration,Rigidity,Negativism,Waxy flexibility | 99.1 | 98.2 | 99.8 |
| 9 | Mutism,Staring,Posturing/catalepsy,Echopraxia/echolalia,Stereotypy,Mannerisms,Rigidity,Waxy flexibility,Withdrawal | 99.1 | 98.2 | 99.8 |
| 9 | Mutism,Staring,Posturing/catalepsy,Echopraxia/echolalia,Stereotypy,Mannerisms,Verbigeration,Rigidity,Withdrawal | 99.1 | 98.2 | 99.8 |
| 9 | Mutism,Staring,Posturing/catalepsy,Echopraxia/echolalia,Stereotypy,Verbigeration,Rigidity,Negativism,Withdrawal | 99.1 | 98.2 | 99.8 |
| 9 | Mutism,Staring,Posturing/catalepsy,Grimacing,Echopraxia/echolalia,Mannerisms,Rigidity,Waxy flexibility,Withdrawal | 99.1 | 98.2 | 99.8 |
| 9 | Mutism,Staring,Posturing/catalepsy,Grimacing,Echopraxia/echolalia,Stereotypy,Mannerisms,Verbigeration,Rigidity | 99.1 | 98.2 | 99.8 |
| 9 | Mutism,Staring,Posturing/catalepsy,Grimacing,Echopraxia/echolalia,Stereotypy,Mannerisms,Waxy flexibility,Withdrawal | 99.1 | 98.2 | 99.8 |
| 9 | Mutism,Staring,Posturing/catalepsy,Grimacing,Echopraxia/echolalia,Stereotypy,Rigidity,Negativism,Withdrawal | 99.1 | 98.2 | 99.8 |
| 9 | Mutism,Staring,Posturing/catalepsy,Grimacing,Echopraxia/echolalia,Stereotypy,Verbigeration,Negativism,Withdrawal | 99.1 | 98.2 | 99.8 |
| 9 | Mutism,Staring,Posturing/catalepsy,Grimacing,Echopraxia/echolalia,Stereotypy,Verbigeration,Rigidity,Negativism | 99.1 | 98.2 | 99.8 |
| 9 | Mutism,Staring,Posturing/catalepsy,Grimacing,Mannerisms,Verbigeration,Rigidity,Waxy flexibility,Withdrawal | 99.1 | 98.2 | 99.8 |
| 9 | Mutism,Staring,Posturing/catalepsy,Grimacing,Stereotypy,Mannerisms,Rigidity,Negativism,Waxy flexibility | 99.1 | 98.2 | 99.8 |
| 9 | Mutism,Staring,Posturing/catalepsy,Grimacing,Stereotypy,Verbigeration,Negativism,Waxy flexibility,Withdrawal | 99.1 | 98.2 | 99.8 |
| 9 | Mutism,Staring,Posturing/catalepsy,Stereotypy,Mannerisms,Verbigeration,Rigidity,Waxy flexibility,Withdrawal | 99.1 | 98.2 | 99.8 |
| 9 | Mutism,Staring,Posturing/catalepsy,Stereotypy,Verbigeration,Rigidity,Negativism,Waxy flexibility,Withdrawal | 99.1 | 98.2 | 99.8 |
| 9 | Staring,Grimacing,Echopraxia/echolalia,Mannerisms,Verbigeration,Rigidity,Negativism,Waxy flexibility,Withdrawal | 99.1 | 98.2 | 99.8 |
| 9 | Staring,Posturing/catalepsy,Echopraxia/echolalia,Mannerisms,Verbigeration,Rigidity,Negativism,Waxy flexibility,Withdrawal | 99.1 | 98.2 | 99.8 |
| 9 | Staring,Posturing/catalepsy,Grimacing,Echopraxia/echolalia,Stereotypy,Mannerisms,Negativism,Waxy flexibility,Withdrawal | 99.1 | 98.2 | 99.8 |
| 9 | Staring,Posturing/catalepsy,Grimacing,Echopraxia/echolalia,Stereotypy,Mannerisms,Verbigeration,Negativism,Withdrawal | 99.1 | 98.2 | 99.8 |
| 9 | Staring,Posturing/catalepsy,Grimacing,Mannerisms,Verbigeration,Rigidity,Negativism,Waxy flexibility,Withdrawal | 99.1 | 98.2 | 99.8 |
| 9 | Staring,Posturing/catalepsy,Grimacing,Stereotypy,Mannerisms,Verbigeration,Negativism,Waxy flexibility,Withdrawal | 99.1 | 98.2 | 99.8 |
| 9 | Excitement,Immobility/stupor,Echopraxia/echolalia,Stereotypy,Mannerisms,Verbigeration,Rigidity,Negativism,Waxy flexibility | 98.9 | 97.8 | 99.8 |
| 9 | Excitement,Immobility/stupor,Echopraxia/echolalia,Stereotypy,Mannerisms,Verbigeration,Rigidity,Waxy flexibility,Withdrawal | 98.9 | 97.8 | 99.8 |
| 9 | Excitement,Immobility/stupor,Grimacing,Echopraxia/echolalia,Mannerisms,Verbigeration,Rigidity,Negativism,Waxy flexibility | 98.9 | 97.8 | 99.8 |
| 9 | Excitement,Immobility/stupor,Grimacing,Echopraxia/echolalia,Stereotypy,Verbigeration,Rigidity,Negativism,Waxy flexibility | 98.9 | 97.8 | 99.8 |
| 9 | Excitement,Immobility/stupor,Grimacing,Mannerisms,Verbigeration,Rigidity,Negativism,Waxy flexibility,Withdrawal | 98.9 | 97.8 | 99.8 |
| 9 | Excitement,Immobility/stupor,Grimacing,Stereotypy,Verbigeration,Rigidity,Negativism,Waxy flexibility,Withdrawal | 98.9 | 97.8 | 99.8 |
| 9 | Excitement,Immobility/stupor,Mutism,Echopraxia/echolalia,Mannerisms,Rigidity,Negativism,Waxy flexibility,Withdrawal | 98.9 | 97.8 | 99.8 |
| 9 | Excitement,Immobility/stupor,Mutism,Echopraxia/echolalia,Mannerisms,Verbigeration,Rigidity,Waxy flexibility,Withdrawal | 98.9 | 97.8 | 99.8 |
| 9 | Excitement,Immobility/stupor,Mutism,Echopraxia/echolalia,Stereotypy,Mannerisms,Rigidity,Negativism,Waxy flexibility | 98.9 | 97.8 | 99.8 |
| 9 | Excitement,Immobility/stupor,Mutism,Echopraxia/echolalia,Stereotypy,Mannerisms,Rigidity,Waxy flexibility,Withdrawal | 98.9 | 97.8 | 99.8 |
| 9 | Excitement,Immobility/stupor,Mutism,Grimacing,Echopraxia/echolalia,Mannerisms,Rigidity,Negativism,Waxy flexibility | 98.9 | 97.8 | 99.8 |
| 9 | Excitement,Immobility/stupor,Mutism,Grimacing,Echopraxia/echolalia,Mannerisms,Rigidity,Waxy flexibility,Withdrawal | 98.9 | 97.8 | 99.8 |
| 9 | Excitement,Immobility/stupor,Mutism,Grimacing,Echopraxia/echolalia,Mannerisms,Verbigeration,Rigidity,Withdrawal | 98.9 | 97.8 | 99.8 |
| 9 | Excitement,Immobility/stupor,Mutism,Grimacing,Echopraxia/echolalia,Stereotypy,Mannerisms,Rigidity,Negativism | 98.9 | 97.8 | 99.8 |
| 9 | Excitement,Immobility/stupor,Mutism,Grimacing,Echopraxia/echolalia,Stereotypy,Mannerisms,Rigidity,Withdrawal | 98.9 | 97.8 | 99.8 |
| 9 | Excitement,Immobility/stupor,Mutism,Grimacing,Mannerisms,Rigidity,Negativism,Waxy flexibility,Withdrawal | 98.9 | 97.8 | 99.8 |
| 9 | Excitement,Immobility/stupor,Mutism,Grimacing,Mannerisms,Verbigeration,Rigidity,Waxy flexibility,Withdrawal | 98.9 | 97.8 | 99.8 |
| 9 | Excitement,Immobility/stupor,Mutism,Grimacing,Stereotypy,Mannerisms,Rigidity,Negativism,Withdrawal | 98.9 | 97.8 | 99.8 |
| 9 | Excitement,Immobility/stupor,Mutism,Grimacing,Stereotypy,Mannerisms,Verbigeration,Rigidity,Withdrawal | 98.9 | 97.8 | 99.8 |
| 9 | Excitement,Immobility/stupor,Mutism,Posturing/catalepsy,Echopraxia/echolalia,Mannerisms,Rigidity,Negativism,Waxy flexibility | 98.9 | 97.8 | 99.8 |
| 9 | Excitement,Immobility/stupor,Mutism,Posturing/catalepsy,Echopraxia/echolalia,Mannerisms,Rigidity,Waxy flexibility,Withdrawal | 98.9 | 97.8 | 99.8 |
| 9 | Excitement,Immobility/stupor,Mutism,Posturing/catalepsy,Echopraxia/echolalia,Mannerisms,Verbigeration,Negativism,Withdrawal | 98.9 | 97.8 | 99.8 |
| 9 | Excitement,Immobility/stupor,Mutism,Posturing/catalepsy,Echopraxia/echolalia,Mannerisms,Verbigeration,Rigidity,Waxy flexibility | 98.9 | 97.8 | 99.8 |
| 9 | Excitement,Immobility/stupor,Mutism,Posturing/catalepsy,Echopraxia/echolalia,Verbigeration,Negativism,Waxy flexibility,Withdrawal | 98.9 | 97.8 | 99.8 |
| 9 | Excitement,Immobility/stupor,Mutism,Posturing/catalepsy,Grimacing,Echopraxia/echolalia,Mannerisms,Negativism,Withdrawal | 98.9 | 97.8 | 99.8 |
| 9 | Excitement,Immobility/stupor,Mutism,Posturing/catalepsy,Grimacing,Echopraxia/echolalia,Negativism,Waxy flexibility,Withdrawal | 98.9 | 97.8 | 99.8 |
| 9 | Excitement,Immobility/stupor,Mutism,Posturing/catalepsy,Grimacing,Echopraxia/echolalia,Stereotypy,Negativism,Withdrawal | 98.9 | 97.8 | 99.8 |
| 9 | Excitement,Immobility/stupor,Mutism,Posturing/catalepsy,Grimacing,Mannerisms,Rigidity,Negativism,Waxy flexibility | 98.9 | 97.8 | 99.8 |
| 9 | Excitement,Immobility/stupor,Mutism,Posturing/catalepsy,Grimacing,Mannerisms,Rigidity,Waxy flexibility,Withdrawal | 98.9 | 97.8 | 99.8 |
| 9 | Excitement,Immobility/stupor,Mutism,Posturing/catalepsy,Grimacing,Mannerisms,Verbigeration,Negativism,Withdrawal | 98.9 | 97.8 | 99.8 |
| 9 | Excitement,Immobility/stupor,Mutism,Posturing/catalepsy,Grimacing,Mannerisms,Verbigeration,Rigidity,Waxy flexibility | 98.9 | 97.8 | 99.8 |
| 9 | Excitement,Immobility/stupor,Mutism,Posturing/catalepsy,Grimacing,Stereotypy,Mannerisms,Rigidity,Negativism | 98.9 | 97.8 | 99.8 |
| 9 | Excitement,Immobility/stupor,Mutism,Posturing/catalepsy,Grimacing,Stereotypy,Mannerisms,Rigidity,Withdrawal | 98.9 | 97.8 | 99.8 |
| 9 | Excitement,Immobility/stupor,Mutism,Posturing/catalepsy,Grimacing,Stereotypy,Mannerisms,Verbigeration,Rigidity | 98.9 | 97.8 | 99.8 |
| 9 | Excitement,Immobility/stupor,Mutism,Posturing/catalepsy,Grimacing,Stereotypy,Rigidity,Negativism,Waxy flexibility | 98.9 | 97.8 | 99.8 |
| 9 | Excitement,Immobility/stupor,Mutism,Posturing/catalepsy,Grimacing,Stereotypy,Rigidity,Waxy flexibility,Withdrawal | 98.9 | 97.8 | 99.8 |
| 9 | Excitement,Immobility/stupor,Mutism,Posturing/catalepsy,Grimacing,Stereotypy,Verbigeration,Negativism,Withdrawal | 98.9 | 97.8 | 99.8 |
| 9 | Excitement,Immobility/stupor,Mutism,Posturing/catalepsy,Grimacing,Stereotypy,Verbigeration,Rigidity,Waxy flexibility | 98.9 | 97.8 | 99.8 |
| 9 | Excitement,Immobility/stupor,Mutism,Posturing/catalepsy,Mannerisms,Rigidity,Negativism,Waxy flexibility,Withdrawal | 98.9 | 97.8 | 99.8 |
| 9 | Excitement,Immobility/stupor,Mutism,Posturing/catalepsy,Stereotypy,Mannerisms,Verbigeration,Negativism,Withdrawal | 98.9 | 97.8 | 99.8 |
| 9 | Excitement,Immobility/stupor,Mutism,Posturing/catalepsy,Stereotypy,Verbigeration,Negativism,Waxy flexibility,Withdrawal | 98.9 | 97.8 | 99.8 |
| 9 | Excitement,Immobility/stupor,Mutism,Staring,Echopraxia/echolalia,Stereotypy,Mannerisms,Waxy flexibility,Withdrawal | 98.9 | 97.8 | 99.8 |
| 9 | Excitement,Immobility/stupor,Mutism,Staring,Echopraxia/echolalia,Stereotypy,Verbigeration,Waxy flexibility,Withdrawal | 98.9 | 97.8 | 99.8 |
| 9 | Excitement,Immobility/stupor,Mutism,Staring,Grimacing,Echopraxia/echolalia,Mannerisms,Waxy flexibility,Withdrawal | 98.9 | 97.8 | 99.8 |
| 9 | Excitement,Immobility/stupor,Mutism,Staring,Grimacing,Echopraxia/echolalia,Stereotypy,Mannerisms,Negativism | 98.9 | 97.8 | 99.8 |
| 9 | Excitement,Immobility/stupor,Mutism,Staring,Grimacing,Echopraxia/echolalia,Stereotypy,Verbigeration,Waxy flexibility | 98.9 | 97.8 | 99.8 |
| 9 | Excitement,Immobility/stupor,Mutism,Staring,Grimacing,Echopraxia/echolalia,Verbigeration,Waxy flexibility,Withdrawal | 98.9 | 97.8 | 99.8 |
| 9 | Excitement,Immobility/stupor,Mutism,Staring,Grimacing,Mannerisms,Negativism,Waxy flexibility,Withdrawal | 98.9 | 97.8 | 99.8 |
| 9 | Excitement,Immobility/stupor,Mutism,Staring,Grimacing,Mannerisms,Verbigeration,Negativism,Withdrawal | 98.9 | 97.8 | 99.8 |
| 9 | Excitement,Immobility/stupor,Mutism,Staring,Grimacing,Stereotypy,Mannerisms,Negativism,Waxy flexibility | 98.9 | 97.8 | 99.8 |
| 9 | Excitement,Immobility/stupor,Mutism,Staring,Grimacing,Stereotypy,Mannerisms,Negativism,Withdrawal | 98.9 | 97.8 | 99.8 |
| 9 | Excitement,Immobility/stupor,Mutism,Staring,Grimacing,Stereotypy,Mannerisms,Verbigeration,Negativism | 98.9 | 97.8 | 99.8 |
| 9 | Excitement,Immobility/stupor,Mutism,Staring,Grimacing,Verbigeration,Negativism,Waxy flexibility,Withdrawal | 98.9 | 97.8 | 99.8 |
| 9 | Excitement,Immobility/stupor,Mutism,Staring,Posturing/catalepsy,Echopraxia/echolalia,Stereotypy,Mannerisms,Waxy flexibility | 98.9 | 97.8 | 99.8 |
| 9 | Excitement,Immobility/stupor,Mutism,Staring,Posturing/catalepsy,Grimacing,Echopraxia/echolalia,Mannerisms,Verbigeration | 98.9 | 97.8 | 99.8 |
| 9 | Excitement,Immobility/stupor,Mutism,Staring,Posturing/catalepsy,Grimacing,Echopraxia/echolalia,Stereotypy,Verbigeration | 98.9 | 97.8 | 99.8 |
| 9 | Excitement,Immobility/stupor,Mutism,Staring,Posturing/catalepsy,Grimacing,Stereotypy,Mannerisms,Withdrawal | 98.9 | 97.8 | 99.8 |
| 9 | Excitement,Immobility/stupor,Mutism,Staring,Posturing/catalepsy,Stereotypy,Mannerisms,Verbigeration,Waxy flexibility | 98.9 | 97.8 | 99.8 |
| 9 | Excitement,Immobility/stupor,Mutism,Staring,Posturing/catalepsy,Stereotypy,Mannerisms,Verbigeration,Withdrawal | 98.9 | 97.8 | 99.8 |
| 9 | Excitement,Immobility/stupor,Mutism,Staring,Posturing/catalepsy,Stereotypy,Mannerisms,Waxy flexibility,Withdrawal | 98.9 | 97.8 | 99.8 |
| 9 | Excitement,Immobility/stupor,Mutism,Stereotypy,Mannerisms,Rigidity,Negativism,Waxy flexibility,Withdrawal | 98.9 | 97.8 | 99.8 |
| 9 | Excitement,Immobility/stupor,Posturing/catalepsy,Echopraxia/echolalia,Mannerisms,Verbigeration,Rigidity,Negativism,Waxy flexibility | 98.9 | 97.8 | 99.8 |
| 9 | Excitement,Immobility/stupor,Posturing/catalepsy,Echopraxia/echolalia,Mannerisms,Verbigeration,Rigidity,Waxy flexibility,Withdrawal | 98.9 | 97.8 | 99.8 |
| 9 | Excitement,Immobility/stupor,Posturing/catalepsy,Echopraxia/echolalia,Stereotypy,Mannerisms,Verbigeration,Rigidity,Negativism | 98.9 | 97.8 | 99.8 |
| 9 | Excitement,Immobility/stupor,Posturing/catalepsy,Echopraxia/echolalia,Stereotypy,Verbigeration,Rigidity,Negativism,Waxy flexibility | 98.9 | 97.8 | 99.8 |
| 9 | Excitement,Immobility/stupor,Posturing/catalepsy,Echopraxia/echolalia,Stereotypy,Verbigeration,Rigidity,Waxy flexibility,Withdrawal | 98.9 | 97.8 | 99.8 |
| 9 | Excitement,Immobility/stupor,Posturing/catalepsy,Grimacing,Echopraxia/echolalia,Mannerisms,Rigidity,Negativism,Waxy flexibility | 98.9 | 97.8 | 99.8 |
| 9 | Excitement,Immobility/stupor,Posturing/catalepsy,Grimacing,Echopraxia/echolalia,Mannerisms,Rigidity,Waxy flexibility,Withdrawal | 98.9 | 97.8 | 99.8 |
| 9 | Excitement,Immobility/stupor,Posturing/catalepsy,Grimacing,Echopraxia/echolalia,Stereotypy,Mannerisms,Rigidity,Withdrawal | 98.9 | 97.8 | 99.8 |
| 9 | Excitement,Immobility/stupor,Posturing/catalepsy,Grimacing,Echopraxia/echolalia,Stereotypy,Verbigeration,Rigidity,Negativism | 98.9 | 97.8 | 99.8 |
| 9 | Excitement,Immobility/stupor,Posturing/catalepsy,Grimacing,Echopraxia/echolalia,Stereotypy,Verbigeration,Rigidity,Withdrawal | 98.9 | 97.8 | 99.8 |
| 9 | Excitement,Immobility/stupor,Posturing/catalepsy,Grimacing,Echopraxia/echolalia,Verbigeration,Rigidity,Waxy flexibility,Withdrawal | 98.9 | 97.8 | 99.8 |
| 9 | Excitement,Immobility/stupor,Posturing/catalepsy,Grimacing,Stereotypy,Mannerisms,Verbigeration,Rigidity,Negativism | 98.9 | 97.8 | 99.8 |
| 9 | Excitement,Immobility/stupor,Posturing/catalepsy,Grimacing,Stereotypy,Rigidity,Negativism,Waxy flexibility,Withdrawal | 98.9 | 97.8 | 99.8 |
| 9 | Excitement,Immobility/stupor,Posturing/catalepsy,Grimacing,Stereotypy,Verbigeration,Rigidity,Negativism,Waxy flexibility | 98.9 | 97.8 | 99.8 |
| 9 | Excitement,Immobility/stupor,Posturing/catalepsy,Stereotypy,Mannerisms,Rigidity,Negativism,Waxy flexibility,Withdrawal | 98.9 | 97.8 | 99.8 |
| 9 | Excitement,Immobility/stupor,Posturing/catalepsy,Stereotypy,Mannerisms,Verbigeration,Rigidity,Negativism,Waxy flexibility | 98.9 | 97.8 | 99.8 |
| 9 | Excitement,Immobility/stupor,Staring,Grimacing,Echopraxia/echolalia,Mannerisms,Verbigeration,Negativism,Waxy flexibility | 98.9 | 97.8 | 99.8 |
| 9 | Excitement,Immobility/stupor,Staring,Grimacing,Echopraxia/echolalia,Mannerisms,Verbigeration,Negativism,Withdrawal | 98.9 | 97.8 | 99.8 |
| 9 | Excitement,Immobility/stupor,Staring,Grimacing,Echopraxia/echolalia,Stereotypy,Mannerisms,Negativism,Waxy flexibility | 98.9 | 97.8 | 99.8 |
| 9 | Excitement,Immobility/stupor,Staring,Grimacing,Echopraxia/echolalia,Stereotypy,Mannerisms,Verbigeration,Rigidity | 98.9 | 97.8 | 99.8 |
| 9 | Excitement,Immobility/stupor,Staring,Grimacing,Echopraxia/echolalia,Stereotypy,Verbigeration,Negativism,Waxy flexibility | 98.9 | 97.8 | 99.8 |
| 9 | Excitement,Immobility/stupor,Staring,Grimacing,Echopraxia/echolalia,Verbigeration,Negativism,Waxy flexibility,Withdrawal | 98.9 | 97.8 | 99.8 |
| 9 | Excitement,Immobility/stupor,Staring,Grimacing,Mannerisms,Verbigeration,Negativism,Waxy flexibility,Withdrawal | 98.9 | 97.8 | 99.8 |
| 9 | Excitement,Immobility/stupor,Staring,Grimacing,Stereotypy,Mannerisms,Verbigeration,Negativism,Waxy flexibility | 98.9 | 97.8 | 99.8 |
| 9 | Excitement,Immobility/stupor,Staring,Grimacing,Stereotypy,Verbigeration,Negativism,Waxy flexibility,Withdrawal | 98.9 | 97.8 | 99.8 |
| 9 | Excitement,Immobility/stupor,Staring,Posturing/catalepsy,Echopraxia/echolalia,Mannerisms,Verbigeration,Waxy flexibility,Withdrawal | 98.9 | 97.8 | 99.8 |
| 9 | Excitement,Immobility/stupor,Staring,Posturing/catalepsy,Echopraxia/echolalia,Stereotypy,Mannerisms,Rigidity,Waxy flexibility | 98.9 | 97.8 | 99.8 |
| 9 | Excitement,Immobility/stupor,Staring,Posturing/catalepsy,Echopraxia/echolalia,Stereotypy,Mannerisms,Waxy flexibility,Withdrawal | 98.9 | 97.8 | 99.8 |
| 9 | Excitement,Immobility/stupor,Staring,Posturing/catalepsy,Grimacing,Echopraxia/echolalia,Mannerisms,Verbigeration,Withdrawal | 98.9 | 97.8 | 99.8 |
| 9 | Excitement,Immobility/stupor,Staring,Posturing/catalepsy,Grimacing,Echopraxia/echolalia,Stereotypy,Mannerisms,Withdrawal | 98.9 | 97.8 | 99.8 |
| 9 | Excitement,Immobility/stupor,Staring,Posturing/catalepsy,Grimacing,Echopraxia/echolalia,Stereotypy,Rigidity,Waxy flexibility | 98.9 | 97.8 | 99.8 |
| 9 | Excitement,Immobility/stupor,Staring,Posturing/catalepsy,Grimacing,Mannerisms,Verbigeration,Waxy flexibility,Withdrawal | 98.9 | 97.8 | 99.8 |
| 9 | Excitement,Immobility/stupor,Staring,Posturing/catalepsy,Grimacing,Stereotypy,Mannerisms,Negativism,Waxy flexibility | 98.9 | 97.8 | 99.8 |
| 9 | Excitement,Immobility/stupor,Staring,Posturing/catalepsy,Grimacing,Stereotypy,Mannerisms,Rigidity,Waxy flexibility | 98.9 | 97.8 | 99.8 |
| 9 | Excitement,Mutism,Staring,Echopraxia/echolalia,Stereotypy,Mannerisms,Verbigeration,Waxy flexibility,Withdrawal | 98.9 | 97.8 | 99.8 |
| 9 | Excitement,Mutism,Staring,Grimacing,Echopraxia/echolalia,Mannerisms,Verbigeration,Waxy flexibility,Withdrawal | 98.9 | 97.8 | 99.8 |
| 9 | Excitement,Mutism,Staring,Grimacing,Echopraxia/echolalia,Stereotypy,Mannerisms,Verbigeration,Waxy flexibility | 98.9 | 97.8 | 99.8 |
| 9 | Excitement,Mutism,Staring,Posturing/catalepsy,Grimacing,Echopraxia/echolalia,Stereotypy,Mannerisms,Verbigeration | 98.9 | 97.8 | 99.8 |
| 9 | Excitement,Mutism,Staring,Posturing/catalepsy,Grimacing,Stereotypy,Mannerisms,Verbigeration,Waxy flexibility | 98.9 | 97.8 | 99.8 |
| 9 | Excitement,Staring,Grimacing,Echopraxia/echolalia,Stereotypy,Verbigeration,Rigidity,Waxy flexibility,Withdrawal | 98.9 | 97.8 | 99.8 |
| 9 | Excitement,Staring,Grimacing,Echopraxia/echolalia,Verbigeration,Rigidity,Negativism,Waxy flexibility,Withdrawal | 98.9 | 97.8 | 99.8 |
| 9 | Excitement,Staring,Posturing/catalepsy,Echopraxia/echolalia,Stereotypy,Mannerisms,Verbigeration,Negativism,Withdrawal | 98.9 | 97.8 | 99.8 |
| 9 | Excitement,Staring,Posturing/catalepsy,Grimacing,Echopraxia/echolalia,Stereotypy,Rigidity,Waxy flexibility,Withdrawal | 98.9 | 97.8 | 99.8 |
| 9 | Excitement,Staring,Posturing/catalepsy,Grimacing,Echopraxia/echolalia,Stereotypy,Verbigeration,Rigidity,Withdrawal | 98.9 | 97.8 | 99.8 |
| 9 | Excitement,Staring,Posturing/catalepsy,Grimacing,Echopraxia/echolalia,Verbigeration,Rigidity,Waxy flexibility,Withdrawal | 98.9 | 97.8 | 99.8 |
| 9 | Excitement,Staring,Posturing/catalepsy,Grimacing,Stereotypy,Rigidity,Negativism,Waxy flexibility,Withdrawal | 98.9 | 97.8 | 99.8 |
| 9 | Excitement,Staring,Posturing/catalepsy,Grimacing,Stereotypy,Verbigeration,Rigidity,Waxy flexibility,Withdrawal | 98.9 | 97.8 | 99.8 |
| 9 | Excitement,Staring,Posturing/catalepsy,Grimacing,Verbigeration,Rigidity,Negativism,Waxy flexibility,Withdrawal | 98.9 | 97.8 | 99.8 |
| 9 | Immobility/stupor,Grimacing,Stereotypy,Mannerisms,Verbigeration,Rigidity,Negativism,Waxy flexibility,Withdrawal | 98.9 | 97.8 | 99.8 |
| 9 | Immobility/stupor,Mutism,Echopraxia/echolalia,Stereotypy,Mannerisms,Rigidity,Negativism,Waxy flexibility,Withdrawal | 98.9 | 97.8 | 99.8 |
| 9 | Immobility/stupor,Mutism,Echopraxia/echolalia,Stereotypy,Mannerisms,Verbigeration,Rigidity,Negativism,Waxy flexibility | 98.9 | 97.8 | 99.8 |
| 9 | Immobility/stupor,Mutism,Grimacing,Echopraxia/echolalia,Mannerisms,Rigidity,Negativism,Waxy flexibility,Withdrawal | 98.9 | 97.8 | 99.8 |
| 9 | Immobility/stupor,Mutism,Grimacing,Echopraxia/echolalia,Mannerisms,Verbigeration,Rigidity,Negativism,Withdrawal | 98.9 | 97.8 | 99.8 |
| 9 | Immobility/stupor,Mutism,Grimacing,Stereotypy,Mannerisms,Rigidity,Negativism,Waxy flexibility,Withdrawal | 98.9 | 97.8 | 99.8 |
| 9 | Immobility/stupor,Mutism,Grimacing,Stereotypy,Mannerisms,Verbigeration,Rigidity,Negativism,Waxy flexibility | 98.9 | 97.8 | 99.8 |
| 9 | Immobility/stupor,Mutism,Posturing/catalepsy,Echopraxia/echolalia,Mannerisms,Verbigeration,Rigidity,Negativism,Withdrawal | 98.9 | 97.8 | 99.8 |
| 9 | Immobility/stupor,Mutism,Posturing/catalepsy,Echopraxia/echolalia,Stereotypy,Mannerisms,Rigidity,Negativism,Withdrawal | 98.9 | 97.8 | 99.8 |
| 9 | Immobility/stupor,Mutism,Posturing/catalepsy,Echopraxia/echolalia,Stereotypy,Mannerisms,Verbigeration,Rigidity,Negativism | 98.9 | 97.8 | 99.8 |
| 9 | Immobility/stupor,Mutism,Posturing/catalepsy,Grimacing,Echopraxia/echolalia,Mannerisms,Verbigeration,Rigidity,Negativism | 98.9 | 97.8 | 99.8 |
| 9 | Immobility/stupor,Mutism,Posturing/catalepsy,Grimacing,Mannerisms,Verbigeration,Rigidity,Negativism,Withdrawal | 98.9 | 97.8 | 99.8 |
| 9 | Immobility/stupor,Mutism,Posturing/catalepsy,Grimacing,Stereotypy,Mannerisms,Rigidity,Negativism,Withdrawal | 98.9 | 97.8 | 99.8 |
| 9 | Immobility/stupor,Mutism,Posturing/catalepsy,Grimacing,Stereotypy,Mannerisms,Verbigeration,Rigidity,Negativism | 98.9 | 97.8 | 99.8 |
| 9 | Immobility/stupor,Mutism,Posturing/catalepsy,Stereotypy,Mannerisms,Verbigeration,Rigidity,Negativism,Waxy flexibility | 98.9 | 97.8 | 99.8 |
| 9 | Immobility/stupor,Mutism,Staring,Echopraxia/echolalia,Stereotypy,Mannerisms,Rigidity,Waxy flexibility,Withdrawal | 98.9 | 97.8 | 99.8 |
| 9 | Immobility/stupor,Mutism,Staring,Echopraxia/echolalia,Stereotypy,Mannerisms,Verbigeration,Negativism,Waxy flexibility | 98.9 | 97.8 | 99.8 |
| 9 | Immobility/stupor,Mutism,Staring,Grimacing,Echopraxia/echolalia,Mannerisms,Rigidity,Waxy flexibility,Withdrawal | 98.9 | 97.8 | 99.8 |
| 9 | Immobility/stupor,Mutism,Staring,Grimacing,Echopraxia/echolalia,Mannerisms,Verbigeration,Negativism,Waxy flexibility | 98.9 | 97.8 | 99.8 |
| 9 | Immobility/stupor,Mutism,Staring,Grimacing,Echopraxia/echolalia,Mannerisms,Verbigeration,Negativism,Withdrawal | 98.9 | 97.8 | 99.8 |
| 9 | Immobility/stupor,Mutism,Staring,Grimacing,Echopraxia/echolalia,Mannerisms,Verbigeration,Rigidity,Withdrawal | 98.9 | 97.8 | 99.8 |
| 9 | Immobility/stupor,Mutism,Staring,Grimacing,Echopraxia/echolalia,Stereotypy,Mannerisms,Verbigeration,Negativism | 98.9 | 97.8 | 99.8 |
| 9 | Immobility/stupor,Mutism,Staring,Grimacing,Echopraxia/echolalia,Stereotypy,Rigidity,Negativism,Withdrawal | 98.9 | 97.8 | 99.8 |
| 9 | Immobility/stupor,Mutism,Staring,Grimacing,Echopraxia/echolalia,Stereotypy,Verbigeration,Rigidity,Negativism | 98.9 | 97.8 | 99.8 |
| 9 | Immobility/stupor,Mutism,Staring,Grimacing,Mannerisms,Verbigeration,Rigidity,Negativism,Waxy flexibility | 98.9 | 97.8 | 99.8 |
| 9 | Immobility/stupor,Mutism,Staring,Grimacing,Mannerisms,Verbigeration,Rigidity,Waxy flexibility,Withdrawal | 98.9 | 97.8 | 99.8 |
| 9 | Immobility/stupor,Mutism,Staring,Grimacing,Stereotypy,Mannerisms,Verbigeration,Negativism,Waxy flexibility | 98.9 | 97.8 | 99.8 |
| 9 | Immobility/stupor,Mutism,Staring,Grimacing,Stereotypy,Verbigeration,Rigidity,Waxy flexibility,Withdrawal | 98.9 | 97.8 | 99.8 |
| 9 | Immobility/stupor,Mutism,Staring,Posturing/catalepsy,Echopraxia/echolalia,Mannerisms,Verbigeration,Negativism,Waxy flexibility | 98.9 | 97.8 | 99.8 |
| 9 | Immobility/stupor,Mutism,Staring,Posturing/catalepsy,Echopraxia/echolalia,Mannerisms,Verbigeration,Rigidity,Withdrawal | 98.9 | 97.8 | 99.8 |
| 9 | Immobility/stupor,Mutism,Staring,Posturing/catalepsy,Echopraxia/echolalia,Stereotypy,Rigidity,Negativism,Withdrawal | 98.9 | 97.8 | 99.8 |
| 9 | Immobility/stupor,Mutism,Staring,Posturing/catalepsy,Echopraxia/echolalia,Stereotypy,Verbigeration,Rigidity,Negativism | 98.9 | 97.8 | 99.8 |
| 9 | Immobility/stupor,Mutism,Staring,Posturing/catalepsy,Grimacing,Echopraxia/echolalia,Mannerisms,Verbigeration,Rigidity | 98.9 | 97.8 | 99.8 |
| 9 | Immobility/stupor,Mutism,Staring,Posturing/catalepsy,Grimacing,Echopraxia/echolalia,Stereotypy,Mannerisms,Withdrawal | 98.9 | 97.8 | 99.8 |
| 9 | Immobility/stupor,Mutism,Staring,Posturing/catalepsy,Grimacing,Echopraxia/echolalia,Stereotypy,Negativism,Withdrawal | 98.9 | 97.8 | 99.8 |
| 9 | Immobility/stupor,Mutism,Staring,Posturing/catalepsy,Grimacing,Mannerisms,Verbigeration,Negativism,Waxy flexibility | 98.9 | 97.8 | 99.8 |
| 9 | Immobility/stupor,Mutism,Staring,Posturing/catalepsy,Grimacing,Stereotypy,Verbigeration,Rigidity,Withdrawal | 98.9 | 97.8 | 99.8 |
| 9 | Immobility/stupor,Mutism,Staring,Posturing/catalepsy,Mannerisms,Verbigeration,Rigidity,Negativism,Waxy flexibility | 98.9 | 97.8 | 99.8 |
| 9 | Immobility/stupor,Mutism,Staring,Posturing/catalepsy,Stereotypy,Mannerisms,Rigidity,Waxy flexibility,Withdrawal | 98.9 | 97.8 | 99.8 |
| 9 | Immobility/stupor,Mutism,Staring,Posturing/catalepsy,Stereotypy,Verbigeration,Rigidity,Negativism,Waxy flexibility | 98.9 | 97.8 | 99.8 |
| 9 | Immobility/stupor,Mutism,Staring,Stereotypy,Mannerisms,Verbigeration,Negativism,Waxy flexibility,Withdrawal | 98.9 | 97.8 | 99.8 |
| 9 | Immobility/stupor,Mutism,Staring,Stereotypy,Mannerisms,Verbigeration,Rigidity,Waxy flexibility,Withdrawal | 98.9 | 97.8 | 99.8 |
| 9 | Immobility/stupor,Posturing/catalepsy,Echopraxia/echolalia,Mannerisms,Verbigeration,Rigidity,Negativism,Waxy flexibility,Withdrawal | 98.9 | 97.8 | 99.8 |
| 9 | Immobility/stupor,Posturing/catalepsy,Echopraxia/echolalia,Stereotypy,Mannerisms,Rigidity,Negativism,Waxy flexibility,Withdrawal | 98.9 | 97.8 | 99.8 |
| 9 | Immobility/stupor,Posturing/catalepsy,Grimacing,Mannerisms,Verbigeration,Rigidity,Negativism,Waxy flexibility,Withdrawal | 98.9 | 97.8 | 99.8 |
| 9 | Immobility/stupor,Posturing/catalepsy,Grimacing,Stereotypy,Mannerisms,Rigidity,Negativism,Waxy flexibility,Withdrawal | 98.9 | 97.8 | 99.8 |
| 9 | Immobility/stupor,Staring,Echopraxia/echolalia,Stereotypy,Mannerisms,Verbigeration,Negativism,Waxy flexibility,Withdrawal | 98.9 | 97.8 | 99.8 |
| 9 | Immobility/stupor,Staring,Echopraxia/echolalia,Stereotypy,Verbigeration,Rigidity,Negativism,Waxy flexibility,Withdrawal | 98.9 | 97.8 | 99.8 |
| 9 | Immobility/stupor,Staring,Grimacing,Echopraxia/echolalia,Stereotypy,Mannerisms,Verbigeration,Negativism,Waxy flexibility | 98.9 | 97.8 | 99.8 |
| 9 | Immobility/stupor,Staring,Grimacing,Echopraxia/echolalia,Stereotypy,Mannerisms,Verbigeration,Rigidity,Withdrawal | 98.9 | 97.8 | 99.8 |
| 9 | Immobility/stupor,Staring,Posturing/catalepsy,Echopraxia/echolalia,Mannerisms,Verbigeration,Negativism,Waxy flexibility,Withdrawal | 98.9 | 97.8 | 99.8 |
| 9 | Immobility/stupor,Staring,Posturing/catalepsy,Echopraxia/echolalia,Stereotypy,Mannerisms,Verbigeration,Negativism,Waxy flexibility | 98.9 | 97.8 | 99.8 |
| 9 | Immobility/stupor,Staring,Posturing/catalepsy,Echopraxia/echolalia,Stereotypy,Verbigeration,Rigidity,Negativism,Withdrawal | 98.9 | 97.8 | 99.8 |
| 9 | Immobility/stupor,Staring,Posturing/catalepsy,Grimacing,Echopraxia/echolalia,Mannerisms,Verbigeration,Negativism,Waxy flexibility | 98.9 | 97.8 | 99.8 |
| 9 | Immobility/stupor,Staring,Posturing/catalepsy,Grimacing,Echopraxia/echolalia,Stereotypy,Rigidity,Negativism,Withdrawal | 98.9 | 97.8 | 99.8 |
| 9 | Immobility/stupor,Staring,Posturing/catalepsy,Grimacing,Mannerisms,Verbigeration,Rigidity,Negativism,Waxy flexibility | 98.9 | 97.8 | 99.8 |
| 9 | Immobility/stupor,Staring,Posturing/catalepsy,Grimacing,Stereotypy,Mannerisms,Rigidity,Waxy flexibility,Withdrawal | 98.9 | 97.8 | 99.8 |
| 9 | Immobility/stupor,Staring,Posturing/catalepsy,Stereotypy,Verbigeration,Rigidity,Negativism,Waxy flexibility,Withdrawal | 98.9 | 97.8 | 99.8 |
| 9 | Mutism,Staring,Echopraxia/echolalia,Stereotypy,Mannerisms,Verbigeration,Rigidity,Waxy flexibility,Withdrawal | 98.9 | 97.8 | 99.8 |
| 9 | Mutism,Staring,Grimacing,Echopraxia/echolalia,Stereotypy,Rigidity,Negativism,Waxy flexibility,Withdrawal | 98.9 | 97.8 | 99.8 |
| 9 | Mutism,Staring,Grimacing,Echopraxia/echolalia,Stereotypy,Verbigeration,Rigidity,Waxy flexibility,Withdrawal | 98.9 | 97.8 | 99.8 |
| 9 | Mutism,Staring,Posturing/catalepsy,Echopraxia/echolalia,Mannerisms,Verbigeration,Rigidity,Waxy flexibility,Withdrawal | 98.9 | 97.8 | 99.8 |
| 9 | Mutism,Staring,Posturing/catalepsy,Echopraxia/echolalia,Stereotypy,Rigidity,Negativism,Waxy flexibility,Withdrawal | 98.9 | 97.8 | 99.8 |
| 9 | Mutism,Staring,Posturing/catalepsy,Echopraxia/echolalia,Stereotypy,Verbigeration,Rigidity,Negativism,Waxy flexibility | 98.9 | 97.8 | 99.8 |
| 9 | Mutism,Staring,Posturing/catalepsy,Grimacing,Echopraxia/echolalia,Mannerisms,Verbigeration,Rigidity,Waxy flexibility | 98.9 | 97.8 | 99.8 |
| 9 | Mutism,Staring,Posturing/catalepsy,Grimacing,Echopraxia/echolalia,Mannerisms,Verbigeration,Waxy flexibility,Withdrawal | 98.9 | 97.8 | 99.8 |
| 9 | Mutism,Staring,Posturing/catalepsy,Grimacing,Echopraxia/echolalia,Stereotypy,Mannerisms,Verbigeration,Withdrawal | 98.9 | 97.8 | 99.8 |
| 9 | Mutism,Staring,Posturing/catalepsy,Grimacing,Echopraxia/echolalia,Stereotypy,Negativism,Waxy flexibility,Withdrawal | 98.9 | 97.8 | 99.8 |
| 9 | Mutism,Staring,Posturing/catalepsy,Grimacing,Echopraxia/echolalia,Stereotypy,Verbigeration,Negativism,Waxy flexibility | 98.9 | 97.8 | 99.8 |
| 9 | Mutism,Staring,Posturing/catalepsy,Grimacing,Echopraxia/echolalia,Stereotypy,Verbigeration,Rigidity,Withdrawal | 98.9 | 97.8 | 99.8 |
| 9 | Mutism,Staring,Posturing/catalepsy,Grimacing,Echopraxia/echolalia,Verbigeration,Rigidity,Negativism,Withdrawal | 98.9 | 97.8 | 99.8 |
| 9 | Mutism,Staring,Posturing/catalepsy,Grimacing,Mannerisms,Verbigeration,Rigidity,Negativism,Waxy flexibility | 98.9 | 97.8 | 99.8 |
| 9 | Mutism,Staring,Posturing/catalepsy,Grimacing,Stereotypy,Mannerisms,Verbigeration,Rigidity,Waxy flexibility | 98.9 | 97.8 | 99.8 |
| 9 | Mutism,Staring,Posturing/catalepsy,Grimacing,Stereotypy,Mannerisms,Verbigeration,Waxy flexibility,Withdrawal | 98.9 | 97.8 | 99.8 |
| 9 | Mutism,Staring,Posturing/catalepsy,Grimacing,Stereotypy,Verbigeration,Rigidity,Negativism,Waxy flexibility | 98.9 | 97.8 | 99.8 |
| 9 | Mutism,Staring,Posturing/catalepsy,Grimacing,Stereotypy,Verbigeration,Rigidity,Waxy flexibility,Withdrawal | 98.9 | 97.8 | 99.8 |
| 9 | Excitement,Immobility/stupor,Grimacing,Echopraxia/echolalia,Mannerisms,Verbigeration,Rigidity,Waxy flexibility,Withdrawal | 98.7 | 97.5 | 99.6 |
| 9 | Excitement,Immobility/stupor,Grimacing,Echopraxia/echolalia,Stereotypy,Mannerisms,Rigidity,Negativism,Waxy flexibility | 98.7 | 97.5 | 99.6 |
| 9 | Excitement,Immobility/stupor,Grimacing,Echopraxia/echolalia,Stereotypy,Mannerisms,Rigidity,Waxy flexibility,Withdrawal | 98.7 | 97.5 | 99.6 |
| 9 | Excitement,Immobility/stupor,Grimacing,Echopraxia/echolalia,Stereotypy,Mannerisms,Verbigeration,Rigidity,Withdrawal | 98.7 | 97.5 | 99.6 |
| 9 | Excitement,Immobility/stupor,Grimacing,Echopraxia/echolalia,Stereotypy,Verbigeration,Rigidity,Waxy flexibility,Withdrawal | 98.7 | 97.5 | 99.6 |
| 9 | Excitement,Immobility/stupor,Grimacing,Stereotypy,Mannerisms,Rigidity,Negativism,Waxy flexibility,Withdrawal | 98.7 | 97.5 | 99.6 |
| 9 | Excitement,Immobility/stupor,Grimacing,Stereotypy,Mannerisms,Verbigeration,Rigidity,Negativism,Waxy flexibility | 98.7 | 97.5 | 99.6 |
| 9 | Excitement,Immobility/stupor,Mutism,Echopraxia/echolalia,Stereotypy,Mannerisms,Verbigeration,Rigidity,Waxy flexibility | 98.7 | 97.5 | 99.6 |
| 9 | Excitement,Immobility/stupor,Mutism,Grimacing,Echopraxia/echolalia,Stereotypy,Verbigeration,Rigidity,Waxy flexibility | 98.7 | 97.5 | 99.6 |
| 9 | Excitement,Immobility/stupor,Mutism,Grimacing,Stereotypy,Mannerisms,Rigidity,Negativism,Waxy flexibility | 98.7 | 97.5 | 99.6 |
| 9 | Excitement,Immobility/stupor,Mutism,Grimacing,Stereotypy,Mannerisms,Rigidity,Waxy flexibility,Withdrawal | 98.7 | 97.5 | 99.6 |
| 9 | Excitement,Immobility/stupor,Mutism,Posturing/catalepsy,Echopraxia/echolalia,Stereotypy,Mannerisms,Negativism,Withdrawal | 98.7 | 97.5 | 99.6 |
| 9 | Excitement,Immobility/stupor,Mutism,Posturing/catalepsy,Echopraxia/echolalia,Stereotypy,Negativism,Waxy flexibility,Withdrawal | 98.7 | 97.5 | 99.6 |
| 9 | Excitement,Immobility/stupor,Mutism,Posturing/catalepsy,Grimacing,Echopraxia/echolalia,Mannerisms,Rigidity,Waxy flexibility | 98.7 | 97.5 | 99.6 |
| 9 | Excitement,Immobility/stupor,Mutism,Posturing/catalepsy,Grimacing,Echopraxia/echolalia,Stereotypy,Mannerisms,Rigidity | 98.7 | 97.5 | 99.6 |
| 9 | Excitement,Immobility/stupor,Mutism,Posturing/catalepsy,Grimacing,Echopraxia/echolalia,Stereotypy,Rigidity,Waxy flexibility | 98.7 | 97.5 | 99.6 |
| 9 | Excitement,Immobility/stupor,Mutism,Posturing/catalepsy,Grimacing,Echopraxia/echolalia,Verbigeration,Negativism,Waxy flexibility | 98.7 | 97.5 | 99.6 |
| 9 | Excitement,Immobility/stupor,Mutism,Posturing/catalepsy,Grimacing,Verbigeration,Negativism,Waxy flexibility,Withdrawal | 98.7 | 97.5 | 99.6 |
| 9 | Excitement,Immobility/stupor,Mutism,Posturing/catalepsy,Mannerisms,Verbigeration,Negativism,Waxy flexibility,Withdrawal | 98.7 | 97.5 | 99.6 |
| 9 | Excitement,Immobility/stupor,Mutism,Posturing/catalepsy,Stereotypy,Mannerisms,Rigidity,Negativism,Waxy flexibility | 98.7 | 97.5 | 99.6 |
| 9 | Excitement,Immobility/stupor,Mutism,Posturing/catalepsy,Stereotypy,Mannerisms,Rigidity,Waxy flexibility,Withdrawal | 98.7 | 97.5 | 99.6 |
| 9 | Excitement,Immobility/stupor,Mutism,Posturing/catalepsy,Stereotypy,Mannerisms,Verbigeration,Rigidity,Waxy flexibility | 98.7 | 97.5 | 99.6 |
| 9 | Excitement,Immobility/stupor,Mutism,Staring,Echopraxia/echolalia,Mannerisms,Verbigeration,Waxy flexibility,Withdrawal | 98.7 | 97.5 | 99.6 |
| 9 | Excitement,Immobility/stupor,Mutism,Staring,Echopraxia/echolalia,Stereotypy,Mannerisms,Verbigeration,Waxy flexibility | 98.7 | 97.5 | 99.6 |
| 9 | Excitement,Immobility/stupor,Mutism,Staring,Grimacing,Echopraxia/echolalia,Mannerisms,Verbigeration,Negativism | 98.7 | 97.5 | 99.6 |
| 9 | Excitement,Immobility/stupor,Mutism,Staring,Grimacing,Echopraxia/echolalia,Mannerisms,Verbigeration,Waxy flexibility | 98.7 | 97.5 | 99.6 |
| 9 | Excitement,Immobility/stupor,Mutism,Staring,Grimacing,Echopraxia/echolalia,Stereotypy,Mannerisms,Waxy flexibility | 98.7 | 97.5 | 99.6 |
| 9 | Excitement,Immobility/stupor,Mutism,Staring,Grimacing,Echopraxia/echolalia,Stereotypy,Verbigeration,Withdrawal | 98.7 | 97.5 | 99.6 |
| 9 | Excitement,Immobility/stupor,Mutism,Staring,Posturing/catalepsy,Echopraxia/echolalia,Stereotypy,Mannerisms,Verbigeration | 98.7 | 97.5 | 99.6 |
| 9 | Excitement,Immobility/stupor,Mutism,Staring,Posturing/catalepsy,Grimacing,Echopraxia/echolalia,Stereotypy,Mannerisms | 98.7 | 97.5 | 99.6 |
| 9 | Excitement,Immobility/stupor,Mutism,Staring,Posturing/catalepsy,Grimacing,Stereotypy,Mannerisms,Verbigeration | 98.7 | 97.5 | 99.6 |
| 9 | Excitement,Immobility/stupor,Mutism,Staring,Posturing/catalepsy,Grimacing,Stereotypy,Mannerisms,Waxy flexibility | 98.7 | 97.5 | 99.6 |
| 9 | Excitement,Immobility/stupor,Posturing/catalepsy,Echopraxia/echolalia,Stereotypy,Mannerisms,Rigidity,Waxy flexibility,Withdrawal | 98.7 | 97.5 | 99.6 |
| 9 | Excitement,Immobility/stupor,Posturing/catalepsy,Echopraxia/echolalia,Stereotypy,Mannerisms,Verbigeration,Negativism,Withdrawal | 98.7 | 97.5 | 99.6 |
| 9 | Excitement,Immobility/stupor,Posturing/catalepsy,Grimacing,Echopraxia/echolalia,Mannerisms,Verbigeration,Negativism,Withdrawal | 98.7 | 97.5 | 99.6 |
| 9 | Excitement,Immobility/stupor,Posturing/catalepsy,Grimacing,Echopraxia/echolalia,Stereotypy,Mannerisms,Rigidity,Negativism | 98.7 | 97.5 | 99.6 |
| 9 | Excitement,Immobility/stupor,Posturing/catalepsy,Grimacing,Echopraxia/echolalia,Stereotypy,Rigidity,Negativism,Waxy flexibility | 98.7 | 97.5 | 99.6 |
| 9 | Excitement,Immobility/stupor,Posturing/catalepsy,Grimacing,Echopraxia/echolalia,Stereotypy,Rigidity,Waxy flexibility,Withdrawal | 98.7 | 97.5 | 99.6 |
| 9 | Excitement,Immobility/stupor,Posturing/catalepsy,Grimacing,Mannerisms,Verbigeration,Rigidity,Waxy flexibility,Withdrawal | 98.7 | 97.5 | 99.6 |
| 9 | Excitement,Immobility/stupor,Posturing/catalepsy,Grimacing,Stereotypy,Mannerisms,Verbigeration,Rigidity,Withdrawal | 98.7 | 97.5 | 99.6 |
| 9 | Excitement,Immobility/stupor,Posturing/catalepsy,Stereotypy,Mannerisms,Verbigeration,Rigidity,Waxy flexibility,Withdrawal | 98.7 | 97.5 | 99.6 |
| 9 | Excitement,Immobility/stupor,Staring,Grimacing,Echopraxia/echolalia,Stereotypy,Mannerisms,Verbigeration,Negativism | 98.7 | 97.5 | 99.6 |
| 9 | Excitement,Immobility/stupor,Staring,Grimacing,Echopraxia/echolalia,Stereotypy,Mannerisms,Waxy flexibility,Withdrawal | 98.7 | 97.5 | 99.6 |
| 9 | Excitement,Immobility/stupor,Staring,Grimacing,Stereotypy,Mannerisms,Negativism,Waxy flexibility,Withdrawal | 98.7 | 97.5 | 99.6 |
| 9 | Excitement,Immobility/stupor,Staring,Posturing/catalepsy,Echopraxia/echolalia,Stereotypy,Mannerisms,Verbigeration,Withdrawal | 98.7 | 97.5 | 99.6 |
| 9 | Excitement,Immobility/stupor,Staring,Posturing/catalepsy,Grimacing,Echopraxia/echolalia,Mannerisms,Verbigeration,Waxy flexibility | 98.7 | 97.5 | 99.6 |
| 9 | Excitement,Immobility/stupor,Staring,Posturing/catalepsy,Grimacing,Stereotypy,Mannerisms,Verbigeration,Withdrawal | 98.7 | 97.5 | 99.6 |
| 9 | Excitement,Immobility/stupor,Staring,Posturing/catalepsy,Stereotypy,Mannerisms,Verbigeration,Waxy flexibility,Withdrawal | 98.7 | 97.5 | 99.6 |
| 9 | Excitement,Mutism,Staring,Grimacing,Echopraxia/echolalia,Stereotypy,Mannerisms,Verbigeration,Withdrawal | 98.7 | 97.5 | 99.6 |
| 9 | Excitement,Staring,Grimacing,Echopraxia/echolalia,Stereotypy,Mannerisms,Negativism,Waxy flexibility,Withdrawal | 98.7 | 97.5 | 99.6 |
| 9 | Excitement,Staring,Grimacing,Echopraxia/echolalia,Stereotypy,Mannerisms,Verbigeration,Negativism,Withdrawal | 98.7 | 97.5 | 99.6 |
| 9 | Excitement,Staring,Posturing/catalepsy,Echopraxia/echolalia,Stereotypy,Mannerisms,Verbigeration,Rigidity,Negativism | 98.7 | 97.5 | 99.6 |
| 9 | Excitement,Staring,Posturing/catalepsy,Echopraxia/echolalia,Stereotypy,Verbigeration,Negativism,Waxy flexibility,Withdrawal | 98.7 | 97.5 | 99.6 |
| 9 | Excitement,Staring,Posturing/catalepsy,Echopraxia/echolalia,Stereotypy,Verbigeration,Rigidity,Waxy flexibility,Withdrawal | 98.7 | 97.5 | 99.6 |
| 9 | Excitement,Staring,Posturing/catalepsy,Grimacing,Echopraxia/echolalia,Stereotypy,Negativism,Waxy flexibility,Withdrawal | 98.7 | 97.5 | 99.6 |
| 9 | Excitement,Staring,Posturing/catalepsy,Grimacing,Echopraxia/echolalia,Stereotypy,Verbigeration,Negativism,Withdrawal | 98.7 | 97.5 | 99.6 |
| 9 | Excitement,Staring,Posturing/catalepsy,Grimacing,Echopraxia/echolalia,Verbigeration,Negativism,Waxy flexibility,Withdrawal | 98.7 | 97.5 | 99.6 |
| 9 | Excitement,Staring,Posturing/catalepsy,Grimacing,Stereotypy,Verbigeration,Negativism,Waxy flexibility,Withdrawal | 98.7 | 97.5 | 99.6 |
| 9 | Immobility/stupor,Grimacing,Echopraxia/echolalia,Mannerisms,Verbigeration,Rigidity,Negativism,Waxy flexibility,Withdrawal | 98.7 | 97.5 | 99.6 |
| 9 | Immobility/stupor,Mutism,Echopraxia/echolalia,Mannerisms,Verbigeration,Rigidity,Negativism,Waxy flexibility,Withdrawal | 98.7 | 97.5 | 99.6 |
| 9 | Immobility/stupor,Mutism,Grimacing,Echopraxia/echolalia,Mannerisms,Verbigeration,Rigidity,Negativism,Waxy flexibility | 98.7 | 97.5 | 99.6 |
| 9 | Immobility/stupor,Mutism,Grimacing,Echopraxia/echolalia,Stereotypy,Mannerisms,Rigidity,Negativism,Waxy flexibility | 98.7 | 97.5 | 99.6 |
| 9 | Immobility/stupor,Mutism,Grimacing,Echopraxia/echolalia,Stereotypy,Verbigeration,Rigidity,Negativism,Withdrawal | 98.7 | 97.5 | 99.6 |
| 9 | Immobility/stupor,Mutism,Grimacing,Mannerisms,Verbigeration,Rigidity,Negativism,Waxy flexibility,Withdrawal | 98.7 | 97.5 | 99.6 |
| 9 | Immobility/stupor,Mutism,Grimacing,Stereotypy,Verbigeration,Rigidity,Negativism,Waxy flexibility,Withdrawal | 98.7 | 97.5 | 99.6 |
| 9 | Immobility/stupor,Mutism,Posturing/catalepsy,Echopraxia/echolalia,Mannerisms,Rigidity,Negativism,Waxy flexibility,Withdrawal | 98.7 | 97.5 | 99.6 |
| 9 | Immobility/stupor,Mutism,Posturing/catalepsy,Echopraxia/echolalia,Mannerisms,Verbigeration,Rigidity,Negativism,Waxy flexibility | 98.7 | 97.5 | 99.6 |
| 9 | Immobility/stupor,Mutism,Posturing/catalepsy,Grimacing,Echopraxia/echolalia,Stereotypy,Mannerisms,Rigidity,Negativism | 98.7 | 97.5 | 99.6 |
| 9 | Immobility/stupor,Mutism,Posturing/catalepsy,Grimacing,Stereotypy,Verbigeration,Rigidity,Negativism,Withdrawal | 98.7 | 97.5 | 99.6 |
| 9 | Immobility/stupor,Mutism,Posturing/catalepsy,Mannerisms,Verbigeration,Rigidity,Negativism,Waxy flexibility,Withdrawal | 98.7 | 97.5 | 99.6 |
| 9 | Immobility/stupor,Mutism,Posturing/catalepsy,Stereotypy,Mannerisms,Rigidity,Negativism,Waxy flexibility,Withdrawal | 98.7 | 97.5 | 99.6 |
| 9 | Immobility/stupor,Mutism,Staring,Echopraxia/echolalia,Mannerisms,Verbigeration,Negativism,Waxy flexibility,Withdrawal | 98.7 | 97.5 | 99.6 |
| 9 | Immobility/stupor,Mutism,Staring,Echopraxia/echolalia,Stereotypy,Mannerisms,Verbigeration,Rigidity,Withdrawal | 98.7 | 97.5 | 99.6 |
| 9 | Immobility/stupor,Mutism,Staring,Echopraxia/echolalia,Stereotypy,Verbigeration,Rigidity,Negativism,Waxy flexibility | 98.7 | 97.5 | 99.6 |
| 9 | Immobility/stupor,Mutism,Staring,Grimacing,Echopraxia/echolalia,Mannerisms,Verbigeration,Rigidity,Waxy flexibility | 98.7 | 97.5 | 99.6 |
| 9 | Immobility/stupor,Mutism,Staring,Grimacing,Echopraxia/echolalia,Stereotypy,Mannerisms,Rigidity,Waxy flexibility | 98.7 | 97.5 | 99.6 |
| 9 | Immobility/stupor,Mutism,Staring,Grimacing,Echopraxia/echolalia,Stereotypy,Mannerisms,Verbigeration,Rigidity | 98.7 | 97.5 | 99.6 |
| 9 | Immobility/stupor,Mutism,Staring,Grimacing,Echopraxia/echolalia,Stereotypy,Mannerisms,Waxy flexibility,Withdrawal | 98.7 | 97.5 | 99.6 |
| 9 | Immobility/stupor,Mutism,Staring,Grimacing,Echopraxia/echolalia,Stereotypy,Verbigeration,Rigidity,Withdrawal | 98.7 | 97.5 | 99.6 |
| 9 | Immobility/stupor,Mutism,Staring,Grimacing,Mannerisms,Verbigeration,Negativism,Waxy flexibility,Withdrawal | 98.7 | 97.5 | 99.6 |
| 9 | Immobility/stupor,Mutism,Staring,Posturing/catalepsy,Echopraxia/echolalia,Mannerisms,Rigidity,Waxy flexibility,Withdrawal | 98.7 | 97.5 | 99.6 |
| 9 | Immobility/stupor,Mutism,Staring,Posturing/catalepsy,Echopraxia/echolalia,Stereotypy,Mannerisms,Verbigeration,Rigidity | 98.7 | 97.5 | 99.6 |
| 9 | Immobility/stupor,Mutism,Staring,Posturing/catalepsy,Echopraxia/echolalia,Stereotypy,Verbigeration,Negativism,Withdrawal | 98.7 | 97.5 | 99.6 |
| 9 | Immobility/stupor,Mutism,Staring,Posturing/catalepsy,Echopraxia/echolalia,Verbigeration,Rigidity,Negativism,Withdrawal | 98.7 | 97.5 | 99.6 |
| 9 | Immobility/stupor,Mutism,Staring,Posturing/catalepsy,Grimacing,Echopraxia/echolalia,Mannerisms,Verbigeration,Withdrawal | 98.7 | 97.5 | 99.6 |
| 9 | Immobility/stupor,Mutism,Staring,Posturing/catalepsy,Grimacing,Echopraxia/echolalia,Mannerisms,Waxy flexibility,Withdrawal | 98.7 | 97.5 | 99.6 |
| 9 | Immobility/stupor,Mutism,Staring,Posturing/catalepsy,Grimacing,Echopraxia/echolalia,Stereotypy,Rigidity,Withdrawal | 98.7 | 97.5 | 99.6 |
| 9 | Immobility/stupor,Mutism,Staring,Posturing/catalepsy,Grimacing,Echopraxia/echolalia,Stereotypy,Verbigeration,Negativism | 98.7 | 97.5 | 99.6 |
| 9 | Immobility/stupor,Mutism,Staring,Posturing/catalepsy,Grimacing,Echopraxia/echolalia,Verbigeration,Rigidity,Negativism | 98.7 | 97.5 | 99.6 |
| 9 | Immobility/stupor,Mutism,Staring,Posturing/catalepsy,Grimacing,Mannerisms,Rigidity,Waxy flexibility,Withdrawal | 98.7 | 97.5 | 99.6 |
| 9 | Immobility/stupor,Mutism,Staring,Posturing/catalepsy,Grimacing,Stereotypy,Mannerisms,Waxy flexibility,Withdrawal | 98.7 | 97.5 | 99.6 |
| 9 | Immobility/stupor,Mutism,Staring,Posturing/catalepsy,Grimacing,Stereotypy,Rigidity,Negativism,Withdrawal | 98.7 | 97.5 | 99.6 |
| 9 | Immobility/stupor,Mutism,Staring,Posturing/catalepsy,Grimacing,Stereotypy,Verbigeration,Negativism,Waxy flexibility | 98.7 | 97.5 | 99.6 |
| 9 | Immobility/stupor,Mutism,Staring,Posturing/catalepsy,Mannerisms,Verbigeration,Rigidity,Waxy flexibility,Withdrawal | 98.7 | 97.5 | 99.6 |
| 9 | Immobility/stupor,Mutism,Staring,Posturing/catalepsy,Stereotypy,Verbigeration,Negativism,Waxy flexibility,Withdrawal | 98.7 | 97.5 | 99.6 |
| 9 | Immobility/stupor,Posturing/catalepsy,Grimacing,Echopraxia/echolalia,Mannerisms,Verbigeration,Rigidity,Negativism,Waxy flexibility | 98.7 | 97.5 | 99.6 |
| 9 | Immobility/stupor,Staring,Grimacing,Echopraxia/echolalia,Mannerisms,Verbigeration,Negativism,Waxy flexibility,Withdrawal | 98.7 | 97.5 | 99.6 |
| 9 | Immobility/stupor,Staring,Grimacing,Echopraxia/echolalia,Stereotypy,Rigidity,Negativism,Waxy flexibility,Withdrawal | 98.7 | 97.5 | 99.6 |
| 9 | Immobility/stupor,Staring,Grimacing,Echopraxia/echolalia,Stereotypy,Verbigeration,Rigidity,Negativism,Waxy flexibility | 98.7 | 97.5 | 99.6 |
| 9 | Immobility/stupor,Staring,Posturing/catalepsy,Grimacing,Echopraxia/echolalia,Mannerisms,Verbigeration,Rigidity,Withdrawal | 98.7 | 97.5 | 99.6 |
| 9 | Immobility/stupor,Staring,Posturing/catalepsy,Grimacing,Echopraxia/echolalia,Verbigeration,Rigidity,Negativism,Withdrawal | 98.7 | 97.5 | 99.6 |
| 9 | Immobility/stupor,Staring,Posturing/catalepsy,Grimacing,Stereotypy,Verbigeration,Negativism,Waxy flexibility,Withdrawal | 98.7 | 97.5 | 99.6 |
| 9 | Immobility/stupor,Staring,Posturing/catalepsy,Grimacing,Stereotypy,Verbigeration,Rigidity,Negativism,Waxy flexibility | 98.7 | 97.5 | 99.6 |
| 9 | Mutism,Staring,Grimacing,Echopraxia/echolalia,Stereotypy,Verbigeration,Negativism,Waxy flexibility,Withdrawal | 98.7 | 97.5 | 99.6 |
| 9 | Mutism,Staring,Grimacing,Echopraxia/echolalia,Verbigeration,Rigidity,Negativism,Waxy flexibility,Withdrawal | 98.7 | 97.5 | 99.6 |
| 9 | Mutism,Staring,Posturing/catalepsy,Echopraxia/echolalia,Stereotypy,Verbigeration,Negativism,Waxy flexibility,Withdrawal | 98.7 | 97.5 | 99.6 |
| 9 | Mutism,Staring,Posturing/catalepsy,Echopraxia/echolalia,Verbigeration,Rigidity,Negativism,Waxy flexibility,Withdrawal | 98.7 | 97.5 | 99.6 |
| 9 | Mutism,Staring,Posturing/catalepsy,Grimacing,Echopraxia/echolalia,Rigidity,Negativism,Waxy flexibility,Withdrawal | 98.7 | 97.5 | 99.6 |
| 9 | Mutism,Staring,Posturing/catalepsy,Grimacing,Echopraxia/echolalia,Stereotypy,Mannerisms,Rigidity,Waxy flexibility | 98.7 | 97.5 | 99.6 |
| 9 | Mutism,Staring,Posturing/catalepsy,Grimacing,Echopraxia/echolalia,Stereotypy,Rigidity,Waxy flexibility,Withdrawal | 98.7 | 97.5 | 99.6 |
| 9 | Mutism,Staring,Posturing/catalepsy,Grimacing,Echopraxia/echolalia,Verbigeration,Negativism,Waxy flexibility,Withdrawal | 98.7 | 97.5 | 99.6 |
| 9 | Mutism,Staring,Posturing/catalepsy,Grimacing,Echopraxia/echolalia,Verbigeration,Rigidity,Negativism,Waxy flexibility | 98.7 | 97.5 | 99.6 |
| 9 | Staring,Grimacing,Echopraxia/echolalia,Stereotypy,Mannerisms,Verbigeration,Negativism,Waxy flexibility,Withdrawal | 98.7 | 97.5 | 99.6 |
| 9 | Staring,Grimacing,Echopraxia/echolalia,Stereotypy,Mannerisms,Verbigeration,Rigidity,Waxy flexibility,Withdrawal | 98.7 | 97.5 | 99.6 |
| 9 | Staring,Posturing/catalepsy,Echopraxia/echolalia,Stereotypy,Mannerisms,Verbigeration,Negativism,Waxy flexibility,Withdrawal | 98.7 | 97.5 | 99.6 |
| 9 | Staring,Posturing/catalepsy,Grimacing,Echopraxia/echolalia,Mannerisms,Verbigeration,Negativism,Waxy flexibility,Withdrawal | 98.7 | 97.5 | 99.6 |
| 9 | Staring,Posturing/catalepsy,Grimacing,Echopraxia/echolalia,Stereotypy,Mannerisms,Verbigeration,Rigidity,Withdrawal | 98.7 | 97.5 | 99.6 |
| 9 | Staring,Posturing/catalepsy,Grimacing,Stereotypy,Mannerisms,Verbigeration,Rigidity,Waxy flexibility,Withdrawal | 98.7 | 97.5 | 99.6 |
| 9 | Excitement,Immobility/stupor,Grimacing,Echopraxia/echolalia,Stereotypy,Mannerisms,Verbigeration,Rigidity,Negativism | 98.6 | 97.5 | 99.6 |
| 9 | Excitement,Immobility/stupor,Mutism,Staring,Grimacing,Echopraxia/echolalia,Stereotypy,Mannerisms,Withdrawal | 98.6 | 97.5 | 99.6 |
| 9 | Excitement,Immobility/stupor,Mutism,Staring,Grimacing,Stereotypy,Verbigeration,Waxy flexibility,Withdrawal | 98.6 | 97.5 | 99.6 |
| 9 | Excitement,Immobility/stupor,Staring,Posturing/catalepsy,Grimacing,Echopraxia/echolalia,Stereotypy,Waxy flexibility,Withdrawal | 98.6 | 97.5 | 99.6 |
| 9 | Excitement,Immobility/stupor,Staring,Posturing/catalepsy,Grimacing,Echopraxia/echolalia,Verbigeration,Waxy flexibility,Withdrawal | 98.6 | 97.5 | 99.6 |
| 9 | Excitement,Mutism,Staring,Grimacing,Stereotypy,Mannerisms,Verbigeration,Waxy flexibility,Withdrawal | 98.6 | 97.5 | 99.6 |
| 9 | Excitement,Staring,Echopraxia/echolalia,Stereotypy,Mannerisms,Verbigeration,Negativism,Waxy flexibility,Withdrawal | 98.6 | 97.5 | 99.6 |
| 9 | Excitement,Staring,Echopraxia/echolalia,Stereotypy,Mannerisms,Verbigeration,Rigidity,Negativism,Waxy flexibility | 98.6 | 97.5 | 99.6 |
| 9 | Excitement,Staring,Posturing/catalepsy,Grimacing,Stereotypy,Mannerisms,Negativism,Waxy flexibility,Withdrawal | 98.6 | 97.5 | 99.6 |
| 9 | Immobility/stupor,Grimacing,Echopraxia/echolalia,Stereotypy,Mannerisms,Verbigeration,Rigidity,Negativism,Waxy flexibility | 98.6 | 97.5 | 99.6 |
| 9 | Immobility/stupor,Mutism,Grimacing,Echopraxia/echolalia,Stereotypy,Mannerisms,Verbigeration,Rigidity,Negativism | 98.6 | 97.5 | 99.6 |
| 9 | Immobility/stupor,Mutism,Posturing/catalepsy,Grimacing,Echopraxia/echolalia,Mannerisms,Rigidity,Negativism,Waxy flexibility | 98.6 | 97.5 | 99.6 |
| 9 | Immobility/stupor,Mutism,Posturing/catalepsy,Grimacing,Mannerisms,Rigidity,Negativism,Waxy flexibility,Withdrawal | 98.6 | 97.5 | 99.6 |
| 9 | Immobility/stupor,Mutism,Staring,Echopraxia/echolalia,Stereotypy,Rigidity,Negativism,Waxy flexibility,Withdrawal | 98.6 | 97.5 | 99.6 |
| 9 | Immobility/stupor,Mutism,Staring,Grimacing,Echopraxia/echolalia,Stereotypy,Verbigeration,Negativism,Withdrawal | 98.6 | 97.5 | 99.6 |
| 9 | Immobility/stupor,Mutism,Staring,Grimacing,Echopraxia/echolalia,Verbigeration,Rigidity,Negativism,Withdrawal | 98.6 | 97.5 | 99.6 |
| 9 | Immobility/stupor,Mutism,Staring,Posturing/catalepsy,Grimacing,Echopraxia/echolalia,Rigidity,Negativism,Withdrawal | 98.6 | 97.5 | 99.6 |
| 9 | Immobility/stupor,Mutism,Staring,Posturing/catalepsy,Grimacing,Echopraxia/echolalia,Stereotypy,Mannerisms,Rigidity | 98.6 | 97.5 | 99.6 |
| 9 | Immobility/stupor,Mutism,Staring,Posturing/catalepsy,Grimacing,Echopraxia/echolalia,Verbigeration,Negativism,Withdrawal | 98.6 | 97.5 | 99.6 |
| 9 | Immobility/stupor,Mutism,Staring,Posturing/catalepsy,Grimacing,Mannerisms,Rigidity,Negativism,Waxy flexibility | 98.6 | 97.5 | 99.6 |
| 9 | Immobility/stupor,Mutism,Staring,Posturing/catalepsy,Grimacing,Stereotypy,Mannerisms,Verbigeration,Withdrawal | 98.6 | 97.5 | 99.6 |
| 9 | Immobility/stupor,Mutism,Staring,Posturing/catalepsy,Stereotypy,Mannerisms,Verbigeration,Rigidity,Waxy flexibility | 98.6 | 97.5 | 99.6 |
| 9 | Immobility/stupor,Posturing/catalepsy,Grimacing,Stereotypy,Mannerisms,Verbigeration,Rigidity,Negativism,Waxy flexibility | 98.6 | 97.5 | 99.6 |
| 9 | Immobility/stupor,Staring,Posturing/catalepsy,Echopraxia/echolalia,Stereotypy,Rigidity,Negativism,Waxy flexibility,Withdrawal | 98.6 | 97.5 | 99.6 |
| 9 | Immobility/stupor,Staring,Posturing/catalepsy,Grimacing,Echopraxia/echolalia,Stereotypy,Verbigeration,Negativism,Withdrawal | 98.6 | 97.5 | 99.6 |
| 9 | Immobility/stupor,Staring,Posturing/catalepsy,Grimacing,Echopraxia/echolalia,Stereotypy,Verbigeration,Rigidity,Negativism | 98.6 | 97.5 | 99.6 |
| 9 | Mutism,Staring,Grimacing,Echopraxia/echolalia,Stereotypy,Mannerisms,Verbigeration,Waxy flexibility,Withdrawal | 98.6 | 97.5 | 99.6 |
| 9 | Mutism,Staring,Posturing/catalepsy,Echopraxia/echolalia,Stereotypy,Mannerisms,Verbigeration,Rigidity,Waxy flexibility | 98.6 | 97.5 | 99.6 |
| 9 | Mutism,Staring,Posturing/catalepsy,Grimacing,Echopraxia/echolalia,Stereotypy,Mannerisms,Verbigeration,Waxy flexibility | 98.6 | 97.5 | 99.6 |
| 9 | Mutism,Staring,Posturing/catalepsy,Grimacing,Stereotypy,Rigidity,Negativism,Waxy flexibility,Withdrawal | 98.6 | 97.5 | 99.6 |
| 9 | Staring,Posturing/catalepsy,Grimacing,Echopraxia/echolalia,Stereotypy,Mannerisms,Rigidity,Waxy flexibility,Withdrawal | 98.6 | 97.5 | 99.6 |
| 9 | Excitement,Immobility/stupor,Mutism,Echopraxia/echolalia,Stereotypy,Mannerisms,Verbigeration,Negativism,Withdrawal | 98.4 | 97.3 | 99.6 |
| 9 | Excitement,Immobility/stupor,Mutism,Echopraxia/echolalia,Stereotypy,Verbigeration,Negativism,Waxy flexibility,Withdrawal | 98.4 | 97.1 | 99.6 |
| 9 | Excitement,Immobility/stupor,Mutism,Grimacing,Echopraxia/echolalia,Mannerisms,Verbigeration,Rigidity,Waxy flexibility | 98.4 | 97.3 | 99.6 |
| 9 | Excitement,Immobility/stupor,Mutism,Grimacing,Echopraxia/echolalia,Stereotypy,Verbigeration,Negativism,Withdrawal | 98.4 | 97.1 | 99.6 |
| 9 | Excitement,Immobility/stupor,Mutism,Grimacing,Stereotypy,Mannerisms,Verbigeration,Rigidity,Waxy flexibility | 98.4 | 97.3 | 99.6 |
| 9 | Excitement,Immobility/stupor,Mutism,Posturing/catalepsy,Echopraxia/echolalia,Mannerisms,Negativism,Waxy flexibility,Withdrawal | 98.4 | 97.1 | 99.6 |
| 9 | Excitement,Immobility/stupor,Mutism,Posturing/catalepsy,Echopraxia/echolalia,Stereotypy,Mannerisms,Rigidity,Waxy flexibility | 98.4 | 97.1 | 99.6 |
| 9 | Excitement,Immobility/stupor,Mutism,Posturing/catalepsy,Echopraxia/echolalia,Stereotypy,Verbigeration,Negativism,Waxy flexibility | 98.4 | 97.1 | 99.6 |
| 9 | Excitement,Immobility/stupor,Mutism,Posturing/catalepsy,Grimacing,Echopraxia/echolalia,Mannerisms,Verbigeration,Negativism | 98.4 | 97.3 | 99.6 |
| 9 | Excitement,Immobility/stupor,Mutism,Posturing/catalepsy,Grimacing,Echopraxia/echolalia,Stereotypy,Verbigeration,Negativism | 98.4 | 97.1 | 99.6 |
| 9 | Excitement,Immobility/stupor,Mutism,Posturing/catalepsy,Grimacing,Mannerisms,Negativism,Waxy flexibility,Withdrawal | 98.4 | 97.1 | 99.6 |
| 9 | Excitement,Immobility/stupor,Mutism,Posturing/catalepsy,Grimacing,Mannerisms,Verbigeration,Negativism,Waxy flexibility | 98.4 | 97.1 | 99.6 |
| 9 | Excitement,Immobility/stupor,Mutism,Posturing/catalepsy,Grimacing,Stereotypy,Mannerisms,Negativism,Withdrawal | 98.4 | 97.1 | 99.6 |
| 9 | Excitement,Immobility/stupor,Mutism,Posturing/catalepsy,Grimacing,Stereotypy,Negativism,Waxy flexibility,Withdrawal | 98.4 | 97.1 | 99.6 |
| 9 | Excitement,Immobility/stupor,Mutism,Posturing/catalepsy,Grimacing,Stereotypy,Verbigeration,Negativism,Waxy flexibility | 98.4 | 97.1 | 99.6 |
| 9 | Excitement,Immobility/stupor,Mutism,Staring,Echopraxia/echolalia,Stereotypy,Mannerisms,Verbigeration,Withdrawal | 98.4 | 97.1 | 99.6 |
| 9 | Excitement,Immobility/stupor,Mutism,Staring,Grimacing,Mannerisms,Verbigeration,Waxy flexibility,Withdrawal | 98.4 | 97.3 | 99.6 |
| 9 | Excitement,Immobility/stupor,Mutism,Staring,Grimacing,Stereotypy,Mannerisms,Verbigeration,Waxy flexibility | 98.4 | 97.1 | 99.6 |
| 9 | Excitement,Immobility/stupor,Mutism,Staring,Grimacing,Stereotypy,Mannerisms,Waxy flexibility,Withdrawal | 98.4 | 97.3 | 99.6 |
| 9 | Excitement,Immobility/stupor,Mutism,Staring,Stereotypy,Mannerisms,Verbigeration,Waxy flexibility,Withdrawal | 98.4 | 97.1 | 99.6 |
| 9 | Excitement,Immobility/stupor,Posturing/catalepsy,Echopraxia/echolalia,Mannerisms,Verbigeration,Negativism,Waxy flexibility,Withdrawal | 98.4 | 97.1 | 99.6 |
| 9 | Excitement,Immobility/stupor,Posturing/catalepsy,Echopraxia/echolalia,Stereotypy,Mannerisms,Rigidity,Negativism,Waxy flexibility | 98.4 | 97.1 | 99.6 |
| 9 | Excitement,Immobility/stupor,Posturing/catalepsy,Echopraxia/echolalia,Stereotypy,Verbigeration,Negativism,Waxy flexibility,Withdrawal | 98.4 | 97.1 | 99.6 |
| 9 | Excitement,Immobility/stupor,Posturing/catalepsy,Grimacing,Echopraxia/echolalia,Mannerisms,Negativism,Waxy flexibility,Withdrawal | 98.4 | 97.1 | 99.6 |
| 9 | Excitement,Immobility/stupor,Posturing/catalepsy,Grimacing,Echopraxia/echolalia,Stereotypy,Mannerisms,Negativism,Withdrawal | 98.4 | 97.1 | 99.6 |
| 9 | Excitement,Immobility/stupor,Posturing/catalepsy,Grimacing,Echopraxia/echolalia,Stereotypy,Verbigeration,Negativism,Withdrawal | 98.4 | 97.1 | 99.6 |
| 9 | Excitement,Immobility/stupor,Posturing/catalepsy,Grimacing,Echopraxia/echolalia,Verbigeration,Negativism,Waxy flexibility,Withdrawal | 98.4 | 97.3 | 99.6 |
| 9 | Excitement,Immobility/stupor,Posturing/catalepsy,Grimacing,Mannerisms,Verbigeration,Negativism,Waxy flexibility,Withdrawal | 98.4 | 97.1 | 99.6 |
| 9 | Excitement,Immobility/stupor,Posturing/catalepsy,Grimacing,Stereotypy,Mannerisms,Rigidity,Negativism,Waxy flexibility | 98.4 | 97.1 | 99.6 |
| 9 | Excitement,Immobility/stupor,Posturing/catalepsy,Grimacing,Stereotypy,Mannerisms,Verbigeration,Negativism,Withdrawal | 98.4 | 97.1 | 99.6 |
| 9 | Excitement,Immobility/stupor,Posturing/catalepsy,Grimacing,Stereotypy,Verbigeration,Rigidity,Waxy flexibility,Withdrawal | 98.4 | 97.1 | 99.6 |
| 9 | Excitement,Immobility/stupor,Posturing/catalepsy,Stereotypy,Mannerisms,Verbigeration,Negativism,Waxy flexibility,Withdrawal | 98.4 | 97.1 | 99.6 |
| 9 | Excitement,Immobility/stupor,Staring,Echopraxia/echolalia,Stereotypy,Mannerisms,Verbigeration,Waxy flexibility,Withdrawal | 98.4 | 97.1 | 99.6 |
| 9 | Excitement,Immobility/stupor,Staring,Grimacing,Echopraxia/echolalia,Mannerisms,Verbigeration,Waxy flexibility,Withdrawal | 98.4 | 97.3 | 99.6 |
| 9 | Excitement,Immobility/stupor,Staring,Posturing/catalepsy,Echopraxia/echolalia,Stereotypy,Mannerisms,Verbigeration,Waxy flexibility | 98.4 | 97.1 | 99.6 |
| 9 | Excitement,Immobility/stupor,Staring,Posturing/catalepsy,Echopraxia/echolalia,Stereotypy,Verbigeration,Waxy flexibility,Withdrawal | 98.4 | 97.1 | 99.6 |
| 9 | Excitement,Immobility/stupor,Staring,Posturing/catalepsy,Grimacing,Echopraxia/echolalia,Stereotypy,Mannerisms,Waxy flexibility | 98.4 | 97.1 | 99.6 |
| 9 | Excitement,Immobility/stupor,Staring,Posturing/catalepsy,Grimacing,Echopraxia/echolalia,Stereotypy,Verbigeration,Withdrawal | 98.4 | 97.1 | 99.6 |
| 9 | Excitement,Immobility/stupor,Staring,Posturing/catalepsy,Grimacing,Stereotypy,Mannerisms,Verbigeration,Waxy flexibility | 98.4 | 97.1 | 99.6 |
| 9 | Excitement,Immobility/stupor,Staring,Posturing/catalepsy,Grimacing,Stereotypy,Mannerisms,Waxy flexibility,Withdrawal | 98.4 | 97.1 | 99.6 |
| 9 | Excitement,Immobility/stupor,Staring,Posturing/catalepsy,Grimacing,Stereotypy,Verbigeration,Waxy flexibility,Withdrawal | 98.4 | 97.3 | 99.6 |
| 9 | Excitement,Staring,Grimacing,Echopraxia/echolalia,Mannerisms,Verbigeration,Negativism,Waxy flexibility,Withdrawal | 98.4 | 97.1 | 99.6 |
| 9 | Excitement,Staring,Grimacing,Echopraxia/echolalia,Stereotypy,Mannerisms,Rigidity,Negativism,Waxy flexibility | 98.4 | 97.1 | 99.6 |
| 9 | Excitement,Staring,Grimacing,Stereotypy,Mannerisms,Verbigeration,Negativism,Waxy flexibility,Withdrawal | 98.4 | 97.1 | 99.6 |
| 9 | Excitement,Staring,Grimacing,Stereotypy,Mannerisms,Verbigeration,Rigidity,Negativism,Waxy flexibility | 98.4 | 97.3 | 99.6 |
| 9 | Excitement,Staring,Posturing/catalepsy,Echopraxia/echolalia,Mannerisms,Verbigeration,Rigidity,Negativism,Waxy flexibility | 98.4 | 97.1 | 99.6 |
| 9 | Excitement,Staring,Posturing/catalepsy,Echopraxia/echolalia,Stereotypy,Mannerisms,Rigidity,Negativism,Waxy flexibility | 98.4 | 97.1 | 99.6 |
| 9 | Excitement,Staring,Posturing/catalepsy,Grimacing,Echopraxia/echolalia,Mannerisms,Verbigeration,Rigidity,Negativism | 98.4 | 97.1 | 99.6 |
| 9 | Excitement,Staring,Posturing/catalepsy,Grimacing,Echopraxia/echolalia,Mannerisms,Verbigeration,Waxy flexibility,Withdrawal | 98.4 | 97.1 | 99.6 |
| 9 | Excitement,Staring,Posturing/catalepsy,Grimacing,Echopraxia/echolalia,Stereotypy,Mannerisms,Rigidity,Negativism | 98.4 | 97.1 | 99.6 |
| 9 | Excitement,Staring,Posturing/catalepsy,Grimacing,Echopraxia/echolalia,Stereotypy,Mannerisms,Waxy flexibility,Withdrawal | 98.4 | 97.1 | 99.6 |
| 9 | Excitement,Staring,Posturing/catalepsy,Grimacing,Stereotypy,Mannerisms,Verbigeration,Rigidity,Negativism | 98.4 | 97.3 | 99.6 |
| 9 | Excitement,Staring,Posturing/catalepsy,Stereotypy,Mannerisms,Verbigeration,Rigidity,Negativism,Waxy flexibility | 98.4 | 97.1 | 99.6 |
| 9 | Immobility/stupor,Grimacing,Echopraxia/echolalia,Stereotypy,Verbigeration,Rigidity,Negativism,Waxy flexibility,Withdrawal | 98.4 | 97.1 | 99.6 |
| 9 | Immobility/stupor,Mutism,Echopraxia/echolalia,Stereotypy,Verbigeration,Rigidity,Negativism,Waxy flexibility,Withdrawal | 98.4 | 97.3 | 99.6 |
| 9 | Immobility/stupor,Mutism,Posturing/catalepsy,Echopraxia/echolalia,Stereotypy,Mannerisms,Rigidity,Negativism,Waxy flexibility | 98.4 | 97.1 | 99.6 |
| 9 | Immobility/stupor,Mutism,Posturing/catalepsy,Echopraxia/echolalia,Stereotypy,Verbigeration,Rigidity,Negativism,Withdrawal | 98.4 | 97.1 | 99.6 |
| 9 | Immobility/stupor,Mutism,Posturing/catalepsy,Grimacing,Echopraxia/echolalia,Mannerisms,Negativism,Waxy flexibility,Withdrawal | 98.4 | 97.3 | 99.6 |
| 9 | Immobility/stupor,Mutism,Posturing/catalepsy,Grimacing,Echopraxia/echolalia,Mannerisms,Verbigeration,Negativism,Withdrawal | 98.4 | 97.3 | 99.6 |
| 9 | Immobility/stupor,Mutism,Posturing/catalepsy,Grimacing,Echopraxia/echolalia,Stereotypy,Mannerisms,Negativism,Withdrawal | 98.4 | 97.1 | 99.6 |
| 9 | Immobility/stupor,Mutism,Posturing/catalepsy,Grimacing,Echopraxia/echolalia,Stereotypy,Rigidity,Negativism,Withdrawal | 98.4 | 97.1 | 99.6 |
| 9 | Immobility/stupor,Mutism,Posturing/catalepsy,Grimacing,Echopraxia/echolalia,Stereotypy,Verbigeration,Rigidity,Negativism | 98.4 | 97.1 | 99.6 |
| 9 | Immobility/stupor,Mutism,Posturing/catalepsy,Grimacing,Mannerisms,Verbigeration,Rigidity,Negativism,Waxy flexibility | 98.4 | 97.1 | 99.6 |
| 9 | Immobility/stupor,Mutism,Posturing/catalepsy,Grimacing,Stereotypy,Mannerisms,Rigidity,Negativism,Waxy flexibility | 98.4 | 97.1 | 99.6 |
| 9 | Immobility/stupor,Mutism,Posturing/catalepsy,Grimacing,Stereotypy,Verbigeration,Rigidity,Negativism,Waxy flexibility | 98.4 | 97.1 | 99.6 |
| 9 | Immobility/stupor,Mutism,Posturing/catalepsy,Stereotypy,Verbigeration,Rigidity,Negativism,Waxy flexibility,Withdrawal | 98.4 | 97.3 | 99.6 |
| 9 | Immobility/stupor,Mutism,Staring,Echopraxia/echolalia,Mannerisms,Verbigeration,Rigidity,Waxy flexibility,Withdrawal | 98.4 | 97.1 | 99.6 |
| 9 | Immobility/stupor,Mutism,Staring,Echopraxia/echolalia,Stereotypy,Mannerisms,Verbigeration,Rigidity,Waxy flexibility | 98.4 | 97.1 | 99.6 |
| 9 | Immobility/stupor,Mutism,Staring,Echopraxia/echolalia,Verbigeration,Rigidity,Negativism,Waxy flexibility,Withdrawal | 98.4 | 97.3 | 99.6 |
| 9 | Immobility/stupor,Mutism,Staring,Grimacing,Echopraxia/echolalia,Stereotypy,Rigidity,Waxy flexibility,Withdrawal | 98.4 | 97.3 | 99.6 |
| 9 | Immobility/stupor,Mutism,Staring,Grimacing,Echopraxia/echolalia,Stereotypy,Verbigeration,Negativism,Waxy flexibility | 98.4 | 97.1 | 99.6 |
| 9 | Immobility/stupor,Mutism,Staring,Grimacing,Echopraxia/echolalia,Stereotypy,Verbigeration,Rigidity,Waxy flexibility | 98.4 | 97.1 | 99.3 |
| 9 | Immobility/stupor,Mutism,Staring,Grimacing,Echopraxia/echolalia,Verbigeration,Rigidity,Negativism,Waxy flexibility | 98.4 | 97.1 | 99.6 |
| 9 | Immobility/stupor,Mutism,Staring,Grimacing,Stereotypy,Rigidity,Negativism,Waxy flexibility,Withdrawal | 98.4 | 97.1 | 99.6 |
| 9 | Immobility/stupor,Mutism,Staring,Grimacing,Stereotypy,Verbigeration,Negativism,Waxy flexibility,Withdrawal | 98.4 | 97.1 | 99.6 |
| 9 | Immobility/stupor,Mutism,Staring,Posturing/catalepsy,Echopraxia/echolalia,Mannerisms,Verbigeration,Rigidity,Waxy flexibility | 98.4 | 97.1 | 99.6 |
| 9 | Immobility/stupor,Mutism,Staring,Posturing/catalepsy,Echopraxia/echolalia,Rigidity,Negativism,Waxy flexibility,Withdrawal | 98.4 | 97.3 | 99.6 |
| 9 | Immobility/stupor,Mutism,Staring,Posturing/catalepsy,Echopraxia/echolalia,Stereotypy,Negativism,Waxy flexibility,Withdrawal | 98.4 | 97.1 | 99.6 |
| 9 | Immobility/stupor,Mutism,Staring,Posturing/catalepsy,Echopraxia/echolalia,Stereotypy,Verbigeration,Negativism,Waxy flexibility | 98.4 | 97.3 | 99.6 |
| 9 | Immobility/stupor,Mutism,Staring,Posturing/catalepsy,Echopraxia/echolalia,Stereotypy,Verbigeration,Rigidity,Withdrawal | 98.4 | 97.1 | 99.6 |
| 9 | Immobility/stupor,Mutism,Staring,Posturing/catalepsy,Echopraxia/echolalia,Verbigeration,Rigidity,Negativism,Waxy flexibility | 98.4 | 97.1 | 99.6 |
| 9 | Immobility/stupor,Mutism,Staring,Posturing/catalepsy,Grimacing,Echopraxia/echolalia,Mannerisms,Verbigeration,Waxy flexibility | 98.4 | 97.1 | 99.6 |
| 9 | Immobility/stupor,Mutism,Staring,Posturing/catalepsy,Grimacing,Echopraxia/echolalia,Negativism,Waxy flexibility,Withdrawal | 98.4 | 97.1 | 99.6 |
| 9 | Immobility/stupor,Mutism,Staring,Posturing/catalepsy,Grimacing,Echopraxia/echolalia,Stereotypy,Rigidity,Negativism | 98.4 | 97.3 | 99.6 |
| 9 | Immobility/stupor,Mutism,Staring,Posturing/catalepsy,Grimacing,Echopraxia/echolalia,Stereotypy,Verbigeration,Rigidity | 98.4 | 97.1 | 99.6 |
| 9 | Immobility/stupor,Mutism,Staring,Posturing/catalepsy,Grimacing,Echopraxia/echolalia,Verbigeration,Negativism,Waxy flexibility | 98.4 | 97.3 | 99.6 |
| 9 | Immobility/stupor,Mutism,Staring,Posturing/catalepsy,Grimacing,Mannerisms,Verbigeration,Rigidity,Waxy flexibility | 98.4 | 97.1 | 99.6 |
| 9 | Immobility/stupor,Mutism,Staring,Posturing/catalepsy,Grimacing,Mannerisms,Verbigeration,Waxy flexibility,Withdrawal | 98.4 | 97.3 | 99.6 |
| 9 | Immobility/stupor,Mutism,Staring,Posturing/catalepsy,Grimacing,Stereotypy,Mannerisms,Rigidity,Waxy flexibility | 98.4 | 97.1 | 99.6 |
| 9 | Immobility/stupor,Mutism,Staring,Posturing/catalepsy,Grimacing,Stereotypy,Negativism,Waxy flexibility,Withdrawal | 98.4 | 97.1 | 99.6 |
| 9 | Immobility/stupor,Mutism,Staring,Posturing/catalepsy,Grimacing,Stereotypy,Verbigeration,Rigidity,Waxy flexibility | 98.4 | 97.1 | 99.6 |
| 9 | Immobility/stupor,Mutism,Staring,Posturing/catalepsy,Grimacing,Verbigeration,Rigidity,Negativism,Withdrawal | 98.4 | 97.1 | 99.6 |
| 9 | Immobility/stupor,Mutism,Staring,Posturing/catalepsy,Stereotypy,Rigidity,Negativism,Waxy flexibility,Withdrawal | 98.4 | 97.1 | 99.6 |
| 9 | Immobility/stupor,Mutism,Staring,Posturing/catalepsy,Stereotypy,Verbigeration,Rigidity,Waxy flexibility,Withdrawal | 98.4 | 97.3 | 99.6 |
| 9 | Immobility/stupor,Posturing/catalepsy,Echopraxia/echolalia,Stereotypy,Mannerisms,Verbigeration,Rigidity,Negativism,Waxy flexibility | 98.4 | 97.3 | 99.6 |
| 9 | Immobility/stupor,Posturing/catalepsy,Grimacing,Echopraxia/echolalia,Stereotypy,Mannerisms,Rigidity,Negativism,Waxy flexibility | 98.4 | 97.1 | 99.6 |
| 9 | Immobility/stupor,Posturing/catalepsy,Grimacing,Echopraxia/echolalia,Stereotypy,Mannerisms,Verbigeration,Rigidity,Negativism | 98.4 | 97.1 | 99.6 |
| 9 | Immobility/stupor,Posturing/catalepsy,Grimacing,Echopraxia/echolalia,Stereotypy,Verbigeration,Rigidity,Negativism,Withdrawal | 98.4 | 97.1 | 99.6 |
| 9 | Immobility/stupor,Posturing/catalepsy,Grimacing,Stereotypy,Verbigeration,Rigidity,Negativism,Waxy flexibility,Withdrawal | 98.4 | 97.3 | 99.3 |
| 9 | Immobility/stupor,Staring,Grimacing,Echopraxia/echolalia,Mannerisms,Verbigeration,Rigidity,Waxy flexibility,Withdrawal | 98.4 | 97.3 | 99.6 |
| 9 | Immobility/stupor,Staring,Grimacing,Echopraxia/echolalia,Stereotypy,Mannerisms,Verbigeration,Rigidity,Waxy flexibility | 98.4 | 97.1 | 99.6 |
| 9 | Immobility/stupor,Staring,Grimacing,Echopraxia/echolalia,Verbigeration,Rigidity,Negativism,Waxy flexibility,Withdrawal | 98.4 | 97.1 | 99.6 |
| 9 | Immobility/stupor,Staring,Posturing/catalepsy,Echopraxia/echolalia,Stereotypy,Mannerisms,Rigidity,Waxy flexibility,Withdrawal | 98.4 | 97.1 | 99.6 |
| 9 | Immobility/stupor,Staring,Posturing/catalepsy,Echopraxia/echolalia,Stereotypy,Mannerisms,Verbigeration,Rigidity,Withdrawal | 98.4 | 97.1 | 99.6 |
| 9 | Immobility/stupor,Staring,Posturing/catalepsy,Echopraxia/echolalia,Stereotypy,Verbigeration,Rigidity,Negativism,Waxy flexibility | 98.4 | 97.1 | 99.6 |
| 9 | Immobility/stupor,Staring,Posturing/catalepsy,Echopraxia/echolalia,Verbigeration,Rigidity,Negativism,Waxy flexibility,Withdrawal | 98.4 | 97.1 | 99.6 |
| 9 | Immobility/stupor,Staring,Posturing/catalepsy,Grimacing,Echopraxia/echolalia,Mannerisms,Rigidity,Waxy flexibility,Withdrawal | 98.4 | 97.3 | 99.6 |
| 9 | Immobility/stupor,Staring,Posturing/catalepsy,Grimacing,Echopraxia/echolalia,Rigidity,Negativism,Waxy flexibility,Withdrawal | 98.4 | 97.3 | 99.6 |
| 9 | Immobility/stupor,Staring,Posturing/catalepsy,Grimacing,Echopraxia/echolalia,Stereotypy,Mannerisms,Verbigeration,Rigidity | 98.4 | 97.1 | 99.3 |
| 9 | Immobility/stupor,Staring,Posturing/catalepsy,Grimacing,Echopraxia/echolalia,Stereotypy,Mannerisms,Waxy flexibility,Withdrawal | 98.4 | 97.1 | 99.6 |
| 9 | Immobility/stupor,Staring,Posturing/catalepsy,Grimacing,Echopraxia/echolalia,Stereotypy,Negativism,Waxy flexibility,Withdrawal | 98.4 | 97.3 | 99.6 |
| 9 | Immobility/stupor,Staring,Posturing/catalepsy,Grimacing,Mannerisms,Verbigeration,Rigidity,Waxy flexibility,Withdrawal | 98.4 | 97.1 | 99.6 |
| 9 | Immobility/stupor,Staring,Posturing/catalepsy,Grimacing,Stereotypy,Mannerisms,Verbigeration,Rigidity,Waxy flexibility | 98.4 | 97.1 | 99.6 |
| 9 | Immobility/stupor,Staring,Posturing/catalepsy,Grimacing,Stereotypy,Rigidity,Negativism,Waxy flexibility,Withdrawal | 98.4 | 97.1 | 99.6 |
| 9 | Immobility/stupor,Staring,Posturing/catalepsy,Stereotypy,Mannerisms,Verbigeration,Rigidity,Waxy flexibility,Withdrawal | 98.4 | 97.1 | 99.6 |
| 9 | Mutism,Staring,Posturing/catalepsy,Echopraxia/echolalia,Stereotypy,Verbigeration,Rigidity,Waxy flexibility,Withdrawal | 98.4 | 97.1 | 99.6 |
| 9 | Mutism,Staring,Posturing/catalepsy,Grimacing,Echopraxia/echolalia,Stereotypy,Rigidity,Negativism,Waxy flexibility | 98.4 | 97.1 | 99.6 |
| 9 | Mutism,Staring,Posturing/catalepsy,Grimacing,Echopraxia/echolalia,Stereotypy,Verbigeration,Rigidity,Waxy flexibility | 98.4 | 97.1 | 99.6 |
| 9 | Mutism,Staring,Posturing/catalepsy,Grimacing,Verbigeration,Rigidity,Negativism,Waxy flexibility,Withdrawal | 98.4 | 97.1 | 99.6 |
| 9 | Staring,Grimacing,Echopraxia/echolalia,Stereotypy,Verbigeration,Rigidity,Negativism,Waxy flexibility,Withdrawal | 98.4 | 97.1 | 99.6 |
| 9 | Staring,Posturing/catalepsy,Grimacing,Echopraxia/echolalia,Stereotypy,Verbigeration,Rigidity,Negativism,Withdrawal | 98.4 | 97.1 | 99.6 |
| 9 | Staring,Posturing/catalepsy,Grimacing,Stereotypy,Verbigeration,Rigidity,Negativism,Waxy flexibility,Withdrawal | 98.4 | 97.1 | 99.6 |
| 9 | Excitement,Immobility/stupor,Grimacing,Stereotypy,Mannerisms,Verbigeration,Rigidity,Waxy flexibility,Withdrawal | 98.2 | 96.9 | 99.3 |
| 9 | Excitement,Immobility/stupor,Mutism,Grimacing,Echopraxia/echolalia,Stereotypy,Mannerisms,Rigidity,Waxy flexibility | 98.2 | 96.9 | 99.3 |
| 9 | Excitement,Immobility/stupor,Mutism,Grimacing,Echopraxia/echolalia,Stereotypy,Mannerisms,Verbigeration,Rigidity | 98.2 | 96.9 | 99.3 |
| 9 | Excitement,Immobility/stupor,Mutism,Grimacing,Echopraxia/echolalia,Stereotypy,Negativism,Waxy flexibility,Withdrawal | 98.2 | 96.9 | 99.3 |
| 9 | Excitement,Immobility/stupor,Mutism,Posturing/catalepsy,Echopraxia/echolalia,Mannerisms,Verbigeration,Negativism,Waxy flexibility | 98.2 | 96.9 | 99.3 |
| 9 | Excitement,Immobility/stupor,Mutism,Posturing/catalepsy,Echopraxia/echolalia,Stereotypy,Mannerisms,Verbigeration,Negativism | 98.2 | 96.9 | 99.3 |
| 9 | Excitement,Immobility/stupor,Mutism,Posturing/catalepsy,Grimacing,Echopraxia/echolalia,Mannerisms,Negativism,Waxy flexibility | 98.2 | 96.9 | 99.3 |
| 9 | Excitement,Immobility/stupor,Mutism,Posturing/catalepsy,Grimacing,Echopraxia/echolalia,Stereotypy,Negativism,Waxy flexibility | 98.2 | 96.9 | 99.3 |
| 9 | Excitement,Immobility/stupor,Mutism,Posturing/catalepsy,Grimacing,Echopraxia/echolalia,Verbigeration,Waxy flexibility,Withdrawal | 98.2 | 96.9 | 99.3 |
| 9 | Excitement,Immobility/stupor,Mutism,Posturing/catalepsy,Grimacing,Stereotypy,Mannerisms,Rigidity,Waxy flexibility | 98.2 | 96.9 | 99.3 |
| 9 | Excitement,Immobility/stupor,Mutism,Posturing/catalepsy,Grimacing,Stereotypy,Mannerisms,Verbigeration,Negativism | 98.2 | 96.9 | 99.3 |
| 9 | Excitement,Immobility/stupor,Mutism,Posturing/catalepsy,Stereotypy,Mannerisms,Negativism,Waxy flexibility,Withdrawal | 98.2 | 96.9 | 99.3 |
| 9 | Excitement,Immobility/stupor,Mutism,Posturing/catalepsy,Stereotypy,Mannerisms,Verbigeration,Negativism,Waxy flexibility | 98.2 | 96.9 | 99.3 |
| 9 | Excitement,Immobility/stupor,Mutism,Staring,Grimacing,Echopraxia/echolalia,Mannerisms,Verbigeration,Withdrawal | 98.2 | 96.9 | 99.3 |
| 9 | Excitement,Immobility/stupor,Mutism,Staring,Grimacing,Stereotypy,Mannerisms,Verbigeration,Withdrawal | 98.2 | 96.9 | 99.3 |
| 9 | Excitement,Immobility/stupor,Posturing/catalepsy,Echopraxia/echolalia,Stereotypy,Mannerisms,Negativism,Waxy flexibility,Withdrawal | 98.2 | 96.9 | 99.3 |
| 9 | Excitement,Immobility/stupor,Posturing/catalepsy,Grimacing,Echopraxia/echolalia,Mannerisms,Verbigeration,Rigidity,Waxy flexibility | 98.2 | 96.9 | 99.3 |
| 9 | Excitement,Immobility/stupor,Posturing/catalepsy,Grimacing,Echopraxia/echolalia,Stereotypy,Negativism,Waxy flexibility,Withdrawal | 98.2 | 96.9 | 99.3 |
| 9 | Excitement,Immobility/stupor,Posturing/catalepsy,Grimacing,Stereotypy,Mannerisms,Rigidity,Waxy flexibility,Withdrawal | 98.2 | 96.9 | 99.3 |
| 9 | Excitement,Immobility/stupor,Posturing/catalepsy,Grimacing,Stereotypy,Verbigeration,Negativism,Waxy flexibility,Withdrawal | 98.2 | 96.9 | 99.3 |
| 9 | Excitement,Immobility/stupor,Staring,Grimacing,Echopraxia/echolalia,Stereotypy,Mannerisms,Verbigeration,Waxy flexibility | 98.2 | 96.9 | 99.3 |
| 9 | Excitement,Immobility/stupor,Staring,Grimacing,Echopraxia/echolalia,Stereotypy,Mannerisms,Verbigeration,Withdrawal | 98.2 | 96.9 | 99.3 |
| 9 | Excitement,Immobility/stupor,Staring,Grimacing,Echopraxia/echolalia,Stereotypy,Verbigeration,Waxy flexibility,Withdrawal | 98.2 | 96.9 | 99.3 |
| 9 | Excitement,Immobility/stupor,Staring,Grimacing,Stereotypy,Mannerisms,Verbigeration,Waxy flexibility,Withdrawal | 98.2 | 96.9 | 99.3 |
| 9 | Excitement,Immobility/stupor,Staring,Posturing/catalepsy,Grimacing,Echopraxia/echolalia,Stereotypy,Mannerisms,Verbigeration | 98.2 | 96.9 | 99.3 |
| 9 | Excitement,Immobility/stupor,Staring,Posturing/catalepsy,Grimacing,Echopraxia/echolalia,Stereotypy,Verbigeration,Waxy flexibility | 98.2 | 96.9 | 99.3 |
| 9 | Excitement,Staring,Grimacing,Echopraxia/echolalia,Mannerisms,Verbigeration,Rigidity,Negativism,Waxy flexibility | 98.2 | 96.9 | 99.3 |
| 9 | Excitement,Staring,Grimacing,Echopraxia/echolalia,Stereotypy,Mannerisms,Verbigeration,Rigidity,Negativism | 98.2 | 96.9 | 99.3 |
| 9 | Excitement,Staring,Grimacing,Echopraxia/echolalia,Stereotypy,Mannerisms,Verbigeration,Rigidity,Waxy flexibility | 98.2 | 96.9 | 99.3 |
| 9 | Excitement,Staring,Grimacing,Echopraxia/echolalia,Stereotypy,Verbigeration,Negativism,Waxy flexibility,Withdrawal | 98.2 | 96.9 | 99.3 |
| 9 | Excitement,Staring,Grimacing,Echopraxia/echolalia,Stereotypy,Verbigeration,Rigidity,Negativism,Waxy flexibility | 98.2 | 96.9 | 99.3 |
| 9 | Excitement,Staring,Posturing/catalepsy,Echopraxia/echolalia,Stereotypy,Verbigeration,Rigidity,Negativism,Waxy flexibility | 98.2 | 96.9 | 99.3 |
| 9 | Excitement,Staring,Posturing/catalepsy,Grimacing,Echopraxia/echolalia,Mannerisms,Rigidity,Negativism,Waxy flexibility | 98.2 | 96.9 | 99.3 |
| 9 | Excitement,Staring,Posturing/catalepsy,Grimacing,Echopraxia/echolalia,Stereotypy,Mannerisms,Verbigeration,Rigidity | 98.2 | 96.9 | 99.3 |
| 9 | Excitement,Staring,Posturing/catalepsy,Grimacing,Echopraxia/echolalia,Stereotypy,Mannerisms,Verbigeration,Withdrawal | 98.2 | 96.9 | 99.3 |
| 9 | Excitement,Staring,Posturing/catalepsy,Grimacing,Echopraxia/echolalia,Stereotypy,Verbigeration,Rigidity,Negativism | 98.2 | 96.9 | 99.3 |
| 9 | Excitement,Staring,Posturing/catalepsy,Grimacing,Mannerisms,Verbigeration,Rigidity,Negativism,Waxy flexibility | 98.2 | 96.9 | 99.3 |
| 9 | Excitement,Staring,Posturing/catalepsy,Grimacing,Stereotypy,Mannerisms,Verbigeration,Waxy flexibility,Withdrawal | 98.2 | 96.9 | 99.3 |
| 9 | Immobility/stupor,Mutism,Grimacing,Echopraxia/echolalia,Stereotypy,Mannerisms,Rigidity,Waxy flexibility,Withdrawal | 98.2 | 96.9 | 99.3 |
| 9 | Immobility/stupor,Mutism,Grimacing,Echopraxia/echolalia,Stereotypy,Rigidity,Negativism,Waxy flexibility,Withdrawal | 98.2 | 96.9 | 99.3 |
| 9 | Immobility/stupor,Mutism,Grimacing,Echopraxia/echolalia,Stereotypy,Verbigeration,Rigidity,Negativism,Waxy flexibility | 98.2 | 96.9 | 99.3 |
| 9 | Immobility/stupor,Mutism,Grimacing,Echopraxia/echolalia,Verbigeration,Rigidity,Negativism,Waxy flexibility,Withdrawal | 98.2 | 96.9 | 99.3 |
| 9 | Immobility/stupor,Mutism,Posturing/catalepsy,Echopraxia/echolalia,Stereotypy,Mannerisms,Verbigeration,Negativism,Withdrawal | 98.2 | 96.9 | 99.3 |
| 9 | Immobility/stupor,Mutism,Posturing/catalepsy,Echopraxia/echolalia,Stereotypy,Verbigeration,Rigidity,Negativism,Waxy flexibility | 98.2 | 96.9 | 99.3 |
| 9 | Immobility/stupor,Mutism,Posturing/catalepsy,Grimacing,Echopraxia/echolalia,Mannerisms,Verbigeration,Rigidity,Withdrawal | 98.2 | 96.9 | 99.3 |
| 9 | Immobility/stupor,Mutism,Posturing/catalepsy,Grimacing,Echopraxia/echolalia,Stereotypy,Mannerisms,Rigidity,Withdrawal | 98.2 | 96.9 | 99.3 |
| 9 | Immobility/stupor,Mutism,Posturing/catalepsy,Grimacing,Echopraxia/echolalia,Verbigeration,Rigidity,Negativism,Waxy flexibility | 98.2 | 96.9 | 99.3 |
| 9 | Immobility/stupor,Mutism,Posturing/catalepsy,Grimacing,Echopraxia/echolalia,Verbigeration,Rigidity,Negativism,Withdrawal | 98.2 | 96.9 | 99.3 |
| 9 | Immobility/stupor,Mutism,Posturing/catalepsy,Grimacing,Stereotypy,Mannerisms,Verbigeration,Negativism,Withdrawal | 98.2 | 96.9 | 99.3 |
| 9 | Immobility/stupor,Mutism,Staring,Echopraxia/echolalia,Stereotypy,Verbigeration,Negativism,Waxy flexibility,Withdrawal | 98.2 | 96.9 | 99.3 |
| 9 | Immobility/stupor,Mutism,Staring,Echopraxia/echolalia,Stereotypy,Verbigeration,Rigidity,Waxy flexibility,Withdrawal | 98.2 | 96.9 | 99.3 |
| 9 | Immobility/stupor,Mutism,Staring,Grimacing,Echopraxia/echolalia,Mannerisms,Verbigeration,Waxy flexibility,Withdrawal | 98.2 | 96.9 | 99.3 |
| 9 | Immobility/stupor,Mutism,Staring,Grimacing,Echopraxia/echolalia,Rigidity,Negativism,Waxy flexibility,Withdrawal | 98.2 | 96.9 | 99.3 |
| 9 | Immobility/stupor,Mutism,Staring,Grimacing,Echopraxia/echolalia,Stereotypy,Mannerisms,Verbigeration,Waxy flexibility | 98.2 | 96.9 | 99.3 |
| 9 | Immobility/stupor,Mutism,Staring,Grimacing,Echopraxia/echolalia,Stereotypy,Mannerisms,Verbigeration,Withdrawal | 98.2 | 96.9 | 99.3 |
| 9 | Immobility/stupor,Mutism,Staring,Grimacing,Echopraxia/echolalia,Stereotypy,Negativism,Waxy flexibility,Withdrawal | 98.2 | 96.9 | 99.3 |
| 9 | Immobility/stupor,Mutism,Staring,Grimacing,Echopraxia/echolalia,Stereotypy,Rigidity,Negativism,Waxy flexibility | 98.2 | 96.9 | 99.3 |
| 9 | Immobility/stupor,Mutism,Staring,Grimacing,Stereotypy,Mannerisms,Verbigeration,Waxy flexibility,Withdrawal | 98.2 | 96.9 | 99.3 |
| 9 | Immobility/stupor,Mutism,Staring,Grimacing,Verbigeration,Rigidity,Negativism,Waxy flexibility,Withdrawal | 98.2 | 96.9 | 99.3 |
| 9 | Immobility/stupor,Mutism,Staring,Posturing/catalepsy,Echopraxia/echolalia,Stereotypy,Mannerisms,Rigidity,Waxy flexibility | 98.2 | 96.9 | 99.3 |
| 9 | Immobility/stupor,Mutism,Staring,Posturing/catalepsy,Echopraxia/echolalia,Stereotypy,Mannerisms,Waxy flexibility,Withdrawal | 98.2 | 96.9 | 99.3 |
| 9 | Immobility/stupor,Mutism,Staring,Posturing/catalepsy,Echopraxia/echolalia,Stereotypy,Rigidity,Negativism,Waxy flexibility | 98.2 | 96.9 | 99.3 |
| 9 | Immobility/stupor,Mutism,Staring,Posturing/catalepsy,Echopraxia/echolalia,Stereotypy,Rigidity,Waxy flexibility,Withdrawal | 98.2 | 96.9 | 99.3 |
| 9 | Immobility/stupor,Mutism,Staring,Posturing/catalepsy,Echopraxia/echolalia,Verbigeration,Negativism,Waxy flexibility,Withdrawal | 98.2 | 96.9 | 99.3 |
| 9 | Immobility/stupor,Mutism,Staring,Posturing/catalepsy,Grimacing,Echopraxia/echolalia,Mannerisms,Rigidity,Waxy flexibility | 98.2 | 96.9 | 99.3 |
| 9 | Immobility/stupor,Mutism,Staring,Posturing/catalepsy,Grimacing,Echopraxia/echolalia,Stereotypy,Mannerisms,Verbigeration | 98.2 | 96.9 | 99.3 |
| 9 | Immobility/stupor,Mutism,Staring,Posturing/catalepsy,Grimacing,Echopraxia/echolalia,Stereotypy,Mannerisms,Waxy flexibility | 98.2 | 96.9 | 99.3 |
| 9 | Immobility/stupor,Mutism,Staring,Posturing/catalepsy,Grimacing,Echopraxia/echolalia,Stereotypy,Negativism,Waxy flexibility | 98.2 | 96.9 | 99.3 |
| 9 | Immobility/stupor,Mutism,Staring,Posturing/catalepsy,Grimacing,Echopraxia/echolalia,Verbigeration,Rigidity,Withdrawal | 98.2 | 96.9 | 99.3 |
| 9 | Immobility/stupor,Mutism,Staring,Posturing/catalepsy,Grimacing,Stereotypy,Mannerisms,Verbigeration,Waxy flexibility | 98.2 | 96.9 | 99.3 |
| 9 | Immobility/stupor,Mutism,Staring,Posturing/catalepsy,Grimacing,Verbigeration,Negativism,Waxy flexibility,Withdrawal | 98.2 | 96.9 | 99.3 |
| 9 | Immobility/stupor,Mutism,Staring,Posturing/catalepsy,Verbigeration,Rigidity,Negativism,Waxy flexibility,Withdrawal | 98.2 | 96.9 | 99.3 |
| 9 | Immobility/stupor,Posturing/catalepsy,Echopraxia/echolalia,Stereotypy,Verbigeration,Rigidity,Negativism,Waxy flexibility,Withdrawal | 98.2 | 96.9 | 99.3 |
| 9 | Immobility/stupor,Posturing/catalepsy,Grimacing,Echopraxia/echolalia,Stereotypy,Mannerisms,Negativism,Waxy flexibility,Withdrawal | 98.2 | 96.9 | 99.3 |
| 9 | Immobility/stupor,Posturing/catalepsy,Grimacing,Echopraxia/echolalia,Stereotypy,Mannerisms,Verbigeration,Negativism,Withdrawal | 98.2 | 96.9 | 99.3 |
| 9 | Immobility/stupor,Posturing/catalepsy,Grimacing,Echopraxia/echolalia,Stereotypy,Rigidity,Negativism,Waxy flexibility,Withdrawal | 98.2 | 96.9 | 99.3 |
| 9 | Immobility/stupor,Posturing/catalepsy,Grimacing,Echopraxia/echolalia,Verbigeration,Rigidity,Negativism,Waxy flexibility,Withdrawal | 98.2 | 96.9 | 99.3 |
| 9 | Immobility/stupor,Staring,Echopraxia/echolalia,Stereotypy,Mannerisms,Verbigeration,Rigidity,Waxy flexibility,Withdrawal | 98.2 | 96.9 | 99.3 |
| 9 | Immobility/stupor,Staring,Grimacing,Echopraxia/echolalia,Stereotypy,Verbigeration,Negativism,Waxy flexibility,Withdrawal | 98.2 | 96.9 | 99.3 |
| 9 | Immobility/stupor,Staring,Grimacing,Echopraxia/echolalia,Stereotypy,Verbigeration,Rigidity,Waxy flexibility,Withdrawal | 98.2 | 96.9 | 99.3 |
| 9 | Immobility/stupor,Staring,Posturing/catalepsy,Echopraxia/echolalia,Stereotypy,Verbigeration,Negativism,Waxy flexibility,Withdrawal | 98.2 | 96.9 | 99.3 |
| 9 | Immobility/stupor,Staring,Posturing/catalepsy,Grimacing,Echopraxia/echolalia,Stereotypy,Mannerisms,Verbigeration,Withdrawal | 98.2 | 96.9 | 99.3 |
| 9 | Immobility/stupor,Staring,Posturing/catalepsy,Grimacing,Echopraxia/echolalia,Stereotypy,Verbigeration,Negativism,Waxy flexibility | 98.2 | 96.9 | 99.3 |
| 9 | Immobility/stupor,Staring,Posturing/catalepsy,Grimacing,Echopraxia/echolalia,Stereotypy,Verbigeration,Rigidity,Withdrawal | 98.2 | 96.9 | 99.3 |
| 9 | Immobility/stupor,Staring,Posturing/catalepsy,Grimacing,Echopraxia/echolalia,Verbigeration,Rigidity,Negativism,Waxy flexibility | 98.2 | 96.9 | 99.3 |
| 9 | Immobility/stupor,Staring,Posturing/catalepsy,Grimacing,Stereotypy,Mannerisms,Verbigeration,Waxy flexibility,Withdrawal | 98.2 | 96.9 | 99.3 |
| 9 | Immobility/stupor,Staring,Posturing/catalepsy,Grimacing,Stereotypy,Verbigeration,Rigidity,Waxy flexibility,Withdrawal | 98.2 | 96.9 | 99.3 |
| 9 | Immobility/stupor,Staring,Posturing/catalepsy,Grimacing,Verbigeration,Rigidity,Negativism,Waxy flexibility,Withdrawal | 98.2 | 96.9 | 99.3 |
| 9 | Mutism,Staring,Posturing/catalepsy,Echopraxia/echolalia,Stereotypy,Mannerisms,Verbigeration,Waxy flexibility,Withdrawal | 98.2 | 96.9 | 99.3 |
| 9 | Mutism,Staring,Posturing/catalepsy,Grimacing,Echopraxia/echolalia,Stereotypy,Verbigeration,Waxy flexibility,Withdrawal | 98.2 | 96.9 | 99.3 |
| 9 | Mutism,Staring,Posturing/catalepsy,Grimacing,Echopraxia/echolalia,Verbigeration,Rigidity,Waxy flexibility,Withdrawal | 98.2 | 96.9 | 99.3 |
| 9 | Staring,Grimacing,Echopraxia/echolalia,Stereotypy,Mannerisms,Verbigeration,Rigidity,Negativism,Waxy flexibility | 98.2 | 96.9 | 99.3 |
| 9 | Staring,Posturing/catalepsy,Echopraxia/echolalia,Stereotypy,Verbigeration,Rigidity,Negativism,Waxy flexibility,Withdrawal | 98.2 | 96.9 | 99.3 |
| 9 | Staring,Posturing/catalepsy,Grimacing,Echopraxia/echolalia,Mannerisms,Verbigeration,Rigidity,Waxy flexibility,Withdrawal | 98.2 | 96.9 | 99.3 |
| 9 | Staring,Posturing/catalepsy,Grimacing,Echopraxia/echolalia,Stereotypy,Mannerisms,Verbigeration,Rigidity,Negativism | 98.2 | 96.9 | 99.3 |
| 9 | Staring,Posturing/catalepsy,Grimacing,Echopraxia/echolalia,Stereotypy,Rigidity,Negativism,Waxy flexibility,Withdrawal | 98.2 | 96.9 | 99.3 |
| 9 | Excitement,Immobility/stupor,Echopraxia/echolalia,Stereotypy,Mannerisms,Verbigeration,Negativism,Waxy flexibility,Withdrawal | 98.0 | 96.6 | 99.1 |
| 9 | Excitement,Immobility/stupor,Grimacing,Echopraxia/echolalia,Stereotypy,Mannerisms,Verbigeration,Negativism,Withdrawal | 98.0 | 96.6 | 99.1 |
| 9 | Excitement,Immobility/stupor,Mutism,Echopraxia/echolalia,Stereotypy,Mannerisms,Negativism,Waxy flexibility,Withdrawal | 98.0 | 96.6 | 99.1 |
| 9 | Excitement,Immobility/stupor,Mutism,Grimacing,Echopraxia/echolalia,Stereotypy,Mannerisms,Negativism,Withdrawal | 98.0 | 96.6 | 99.1 |
| 9 | Excitement,Immobility/stupor,Mutism,Grimacing,Echopraxia/echolalia,Stereotypy,Verbigeration,Negativism,Waxy flexibility | 98.0 | 96.6 | 99.1 |
| 9 | Excitement,Immobility/stupor,Mutism,Grimacing,Echopraxia/echolalia,Verbigeration,Negativism,Waxy flexibility,Withdrawal | 98.0 | 96.6 | 99.1 |
| 9 | Excitement,Immobility/stupor,Mutism,Grimacing,Stereotypy,Mannerisms,Verbigeration,Negativism,Withdrawal | 98.0 | 96.6 | 99.1 |
| 9 | Excitement,Immobility/stupor,Mutism,Grimacing,Stereotypy,Verbigeration,Negativism,Waxy flexibility,Withdrawal | 98.0 | 96.6 | 99.1 |
| 9 | Excitement,Immobility/stupor,Mutism,Posturing/catalepsy,Echopraxia/echolalia,Stereotypy,Verbigeration,Waxy flexibility,Withdrawal | 98.0 | 96.6 | 99.1 |
| 9 | Excitement,Immobility/stupor,Mutism,Posturing/catalepsy,Grimacing,Echopraxia/echolalia,Mannerisms,Verbigeration,Withdrawal | 98.0 | 96.6 | 99.1 |
| 9 | Excitement,Immobility/stupor,Mutism,Posturing/catalepsy,Grimacing,Echopraxia/echolalia,Mannerisms,Waxy flexibility,Withdrawal | 98.0 | 96.6 | 99.1 |
| 9 | Excitement,Immobility/stupor,Mutism,Posturing/catalepsy,Grimacing,Echopraxia/echolalia,Stereotypy,Mannerisms,Negativism | 98.0 | 96.6 | 99.1 |
| 9 | Excitement,Immobility/stupor,Mutism,Posturing/catalepsy,Grimacing,Echopraxia/echolalia,Stereotypy,Verbigeration,Withdrawal | 98.0 | 96.6 | 99.1 |
| 9 | Excitement,Immobility/stupor,Mutism,Posturing/catalepsy,Grimacing,Echopraxia/echolalia,Stereotypy,Waxy flexibility,Withdrawal | 98.0 | 96.6 | 99.1 |
| 9 | Excitement,Immobility/stupor,Mutism,Staring,Grimacing,Echopraxia/echolalia,Stereotypy,Mannerisms,Verbigeration | 98.0 | 96.6 | 99.1 |
| 9 | Excitement,Immobility/stupor,Mutism,Stereotypy,Mannerisms,Verbigeration,Negativism,Waxy flexibility,Withdrawal | 98.0 | 96.6 | 99.1 |
| 9 | Excitement,Immobility/stupor,Posturing/catalepsy,Echopraxia/echolalia,Stereotypy,Mannerisms,Verbigeration,Rigidity,Waxy flexibility | 98.0 | 96.6 | 99.1 |
| 9 | Excitement,Immobility/stupor,Posturing/catalepsy,Grimacing,Echopraxia/echolalia,Mannerisms,Verbigeration,Negativism,Waxy flexibility | 98.0 | 96.6 | 99.1 |
| 9 | Excitement,Immobility/stupor,Posturing/catalepsy,Grimacing,Echopraxia/echolalia,Stereotypy,Mannerisms,Verbigeration,Rigidity | 98.0 | 96.6 | 99.1 |
| 9 | Excitement,Immobility/stupor,Posturing/catalepsy,Grimacing,Echopraxia/echolalia,Stereotypy,Verbigeration,Rigidity,Waxy flexibility | 98.0 | 96.6 | 99.1 |
| 9 | Excitement,Staring,Grimacing,Echopraxia/echolalia,Stereotypy,Mannerisms,Verbigeration,Waxy flexibility,Withdrawal | 98.0 | 96.6 | 99.1 |
| 9 | Excitement,Staring,Posturing/catalepsy,Echopraxia/echolalia,Stereotypy,Mannerisms,Verbigeration,Negativism,Waxy flexibility | 98.0 | 96.6 | 99.1 |
| 9 | Excitement,Staring,Posturing/catalepsy,Echopraxia/echolalia,Stereotypy,Mannerisms,Verbigeration,Waxy flexibility,Withdrawal | 98.0 | 96.6 | 99.1 |
| 9 | Excitement,Staring,Posturing/catalepsy,Grimacing,Echopraxia/echolalia,Mannerisms,Verbigeration,Rigidity,Waxy flexibility | 98.0 | 96.6 | 99.1 |
| 9 | Excitement,Staring,Posturing/catalepsy,Grimacing,Echopraxia/echolalia,Stereotypy,Mannerisms,Rigidity,Waxy flexibility | 98.0 | 96.6 | 99.1 |
| 9 | Excitement,Staring,Posturing/catalepsy,Grimacing,Echopraxia/echolalia,Stereotypy,Rigidity,Negativism,Waxy flexibility | 98.0 | 96.6 | 99.1 |
| 9 | Excitement,Staring,Posturing/catalepsy,Grimacing,Echopraxia/echolalia,Verbigeration,Rigidity,Negativism,Waxy flexibility | 98.0 | 96.6 | 99.1 |
| 9 | Excitement,Staring,Posturing/catalepsy,Grimacing,Stereotypy,Mannerisms,Rigidity,Negativism,Waxy flexibility | 98.0 | 96.6 | 99.1 |
| 9 | Excitement,Staring,Posturing/catalepsy,Grimacing,Stereotypy,Mannerisms,Verbigeration,Rigidity,Waxy flexibility | 98.0 | 96.6 | 99.1 |
| 9 | Excitement,Staring,Posturing/catalepsy,Grimacing,Stereotypy,Verbigeration,Rigidity,Negativism,Waxy flexibility | 98.0 | 96.6 | 99.1 |
| 9 | Immobility/stupor,Mutism,Grimacing,Echopraxia/echolalia,Mannerisms,Verbigeration,Rigidity,Waxy flexibility,Withdrawal | 98.0 | 96.6 | 99.1 |
| 9 | Immobility/stupor,Mutism,Grimacing,Echopraxia/echolalia,Stereotypy,Mannerisms,Negativism,Waxy flexibility,Withdrawal | 98.0 | 96.6 | 99.1 |
| 9 | Immobility/stupor,Mutism,Grimacing,Echopraxia/echolalia,Stereotypy,Mannerisms,Verbigeration,Negativism,Withdrawal | 98.0 | 96.6 | 99.1 |
| 9 | Immobility/stupor,Mutism,Grimacing,Echopraxia/echolalia,Stereotypy,Mannerisms,Verbigeration,Rigidity,Withdrawal | 98.0 | 96.6 | 99.1 |
| 9 | Immobility/stupor,Mutism,Grimacing,Stereotypy,Mannerisms,Verbigeration,Rigidity,Waxy flexibility,Withdrawal | 98.0 | 96.6 | 99.1 |
| 9 | Immobility/stupor,Mutism,Posturing/catalepsy,Echopraxia/echolalia,Mannerisms,Verbigeration,Negativism,Waxy flexibility,Withdrawal | 98.0 | 96.6 | 99.1 |
| 9 | Immobility/stupor,Mutism,Posturing/catalepsy,Echopraxia/echolalia,Stereotypy,Mannerisms,Negativism,Waxy flexibility,Withdrawal | 98.0 | 96.6 | 99.1 |
| 9 | Immobility/stupor,Mutism,Posturing/catalepsy,Echopraxia/echolalia,Stereotypy,Rigidity,Negativism,Waxy flexibility,Withdrawal | 98.0 | 96.6 | 99.1 |
| 9 | Immobility/stupor,Mutism,Posturing/catalepsy,Echopraxia/echolalia,Verbigeration,Rigidity,Negativism,Waxy flexibility,Withdrawal | 98.0 | 96.6 | 99.1 |
| 9 | Immobility/stupor,Mutism,Posturing/catalepsy,Grimacing,Echopraxia/echolalia,Mannerisms,Rigidity,Waxy flexibility,Withdrawal | 98.0 | 96.6 | 99.1 |
| 9 | Immobility/stupor,Mutism,Posturing/catalepsy,Grimacing,Echopraxia/echolalia,Mannerisms,Verbigeration,Negativism,Waxy flexibility | 98.0 | 96.6 | 99.1 |
| 9 | Immobility/stupor,Mutism,Posturing/catalepsy,Grimacing,Echopraxia/echolalia,Rigidity,Negativism,Waxy flexibility,Withdrawal | 98.0 | 96.6 | 99.1 |
| 9 | Immobility/stupor,Mutism,Posturing/catalepsy,Grimacing,Mannerisms,Verbigeration,Negativism,Waxy flexibility,Withdrawal | 98.0 | 96.6 | 99.1 |
| 9 | Immobility/stupor,Mutism,Posturing/catalepsy,Grimacing,Stereotypy,Mannerisms,Negativism,Waxy flexibility,Withdrawal | 98.0 | 96.6 | 99.1 |
| 9 | Immobility/stupor,Mutism,Posturing/catalepsy,Grimacing,Stereotypy,Mannerisms,Verbigeration,Rigidity,Withdrawal | 98.0 | 96.6 | 99.1 |
| 9 | Immobility/stupor,Mutism,Posturing/catalepsy,Grimacing,Stereotypy,Rigidity,Negativism,Waxy flexibility,Withdrawal | 98.0 | 96.6 | 99.1 |
| 9 | Immobility/stupor,Mutism,Posturing/catalepsy,Grimacing,Verbigeration,Rigidity,Negativism,Waxy flexibility,Withdrawal | 98.0 | 96.6 | 99.1 |
| 9 | Immobility/stupor,Mutism,Posturing/catalepsy,Stereotypy,Mannerisms,Verbigeration,Negativism,Waxy flexibility,Withdrawal | 98.0 | 96.6 | 99.1 |
| 9 | Immobility/stupor,Mutism,Staring,Grimacing,Echopraxia/echolalia,Verbigeration,Negativism,Waxy flexibility,Withdrawal | 98.0 | 96.6 | 99.1 |
| 9 | Immobility/stupor,Mutism,Staring,Grimacing,Echopraxia/echolalia,Verbigeration,Rigidity,Waxy flexibility,Withdrawal | 98.0 | 96.6 | 99.1 |
| 9 | Immobility/stupor,Mutism,Staring,Posturing/catalepsy,Echopraxia/echolalia,Mannerisms,Verbigeration,Waxy flexibility,Withdrawal | 98.0 | 96.6 | 99.1 |
| 9 | Immobility/stupor,Mutism,Staring,Posturing/catalepsy,Echopraxia/echolalia,Stereotypy,Mannerisms,Verbigeration,Withdrawal | 98.0 | 96.6 | 99.1 |
| 9 | Immobility/stupor,Mutism,Staring,Posturing/catalepsy,Echopraxia/echolalia,Stereotypy,Verbigeration,Rigidity,Waxy flexibility | 98.0 | 96.6 | 99.1 |
| 9 | Immobility/stupor,Mutism,Staring,Posturing/catalepsy,Grimacing,Echopraxia/echolalia,Stereotypy,Verbigeration,Withdrawal | 98.0 | 96.6 | 99.1 |
| 9 | Immobility/stupor,Mutism,Staring,Posturing/catalepsy,Grimacing,Echopraxia/echolalia,Stereotypy,Waxy flexibility,Withdrawal | 98.0 | 96.6 | 99.1 |
| 9 | Immobility/stupor,Mutism,Staring,Posturing/catalepsy,Grimacing,Stereotypy,Rigidity,Negativism,Waxy flexibility | 98.0 | 96.6 | 99.1 |
| 9 | Immobility/stupor,Mutism,Staring,Posturing/catalepsy,Grimacing,Stereotypy,Rigidity,Waxy flexibility,Withdrawal | 98.0 | 96.6 | 99.1 |
| 9 | Immobility/stupor,Mutism,Staring,Posturing/catalepsy,Grimacing,Stereotypy,Verbigeration,Waxy flexibility,Withdrawal | 98.0 | 96.6 | 99.1 |
| 9 | Immobility/stupor,Mutism,Staring,Posturing/catalepsy,Grimacing,Verbigeration,Rigidity,Negativism,Waxy flexibility | 98.0 | 96.6 | 99.1 |
| 9 | Immobility/stupor,Mutism,Staring,Posturing/catalepsy,Stereotypy,Mannerisms,Verbigeration,Waxy flexibility,Withdrawal | 98.0 | 96.6 | 99.1 |
| 9 | Immobility/stupor,Posturing/catalepsy,Echopraxia/echolalia,Stereotypy,Mannerisms,Verbigeration,Negativism,Waxy flexibility,Withdrawal | 98.0 | 96.6 | 99.1 |
| 9 | Immobility/stupor,Posturing/catalepsy,Grimacing,Echopraxia/echolalia,Stereotypy,Verbigeration,Rigidity,Negativism,Waxy flexibility | 98.0 | 96.6 | 99.1 |
| 9 | Immobility/stupor,Posturing/catalepsy,Grimacing,Stereotypy,Mannerisms,Verbigeration,Negativism,Waxy flexibility,Withdrawal | 98.0 | 96.6 | 99.1 |
| 9 | Immobility/stupor,Staring,Grimacing,Echopraxia/echolalia,Stereotypy,Mannerisms,Verbigeration,Waxy flexibility,Withdrawal | 98.0 | 96.6 | 99.1 |
| 9 | Immobility/stupor,Staring,Posturing/catalepsy,Echopraxia/echolalia,Mannerisms,Verbigeration,Rigidity,Waxy flexibility,Withdrawal | 98.0 | 96.6 | 99.1 |
| 9 | Immobility/stupor,Staring,Posturing/catalepsy,Grimacing,Echopraxia/echolalia,Mannerisms,Verbigeration,Rigidity,Waxy flexibility | 98.0 | 96.6 | 99.1 |
| 9 | Immobility/stupor,Staring,Posturing/catalepsy,Grimacing,Echopraxia/echolalia,Stereotypy,Mannerisms,Rigidity,Waxy flexibility | 98.0 | 96.6 | 99.1 |
| 9 | Immobility/stupor,Staring,Posturing/catalepsy,Grimacing,Echopraxia/echolalia,Stereotypy,Rigidity,Negativism,Waxy flexibility | 98.0 | 96.6 | 99.1 |
| 9 | Immobility/stupor,Staring,Posturing/catalepsy,Grimacing,Echopraxia/echolalia,Stereotypy,Rigidity,Waxy flexibility,Withdrawal | 98.0 | 96.6 | 99.1 |
| 9 | Immobility/stupor,Staring,Posturing/catalepsy,Grimacing,Echopraxia/echolalia,Verbigeration,Negativism,Waxy flexibility,Withdrawal | 98.0 | 96.6 | 99.1 |
| 9 | Staring,Posturing/catalepsy,Echopraxia/echolalia,Stereotypy,Mannerisms,Verbigeration,Rigidity,Negativism,Waxy flexibility | 98.0 | 96.6 | 99.1 |
| 9 | Staring,Posturing/catalepsy,Grimacing,Echopraxia/echolalia,Stereotypy,Mannerisms,Rigidity,Negativism,Waxy flexibility | 98.0 | 96.6 | 99.1 |
| 9 | Staring,Posturing/catalepsy,Grimacing,Echopraxia/echolalia,Verbigeration,Rigidity,Negativism,Waxy flexibility,Withdrawal | 98.0 | 96.6 | 99.1 |
| 9 | Staring,Posturing/catalepsy,Grimacing,Stereotypy,Mannerisms,Verbigeration,Rigidity,Negativism,Waxy flexibility | 98.0 | 96.6 | 99.1 |
| 9 | Excitement,Immobility/stupor,Grimacing,Echopraxia/echolalia,Stereotypy,Mannerisms,Negativism,Waxy flexibility,Withdrawal | 97.8 | 96.2 | 99.1 |
| 9 | Excitement,Immobility/stupor,Grimacing,Echopraxia/echolalia,Stereotypy,Mannerisms,Verbigeration,Rigidity,Waxy flexibility | 97.8 | 96.2 | 99.1 |
| 9 | Excitement,Immobility/stupor,Grimacing,Echopraxia/echolalia,Stereotypy,Verbigeration,Negativism,Waxy flexibility,Withdrawal | 97.8 | 96.2 | 99.1 |
| 9 | Excitement,Immobility/stupor,Mutism,Echopraxia/echolalia,Mannerisms,Verbigeration,Negativism,Waxy flexibility,Withdrawal | 97.8 | 96.2 | 99.1 |
| 9 | Excitement,Immobility/stupor,Mutism,Echopraxia/echolalia,Stereotypy,Mannerisms,Verbigeration,Negativism,Waxy flexibility | 97.8 | 96.2 | 99.1 |
| 9 | Excitement,Immobility/stupor,Mutism,Grimacing,Echopraxia/echolalia,Mannerisms,Verbigeration,Negativism,Withdrawal | 97.8 | 96.4 | 99.1 |
| 9 | Excitement,Immobility/stupor,Mutism,Posturing/catalepsy,Echopraxia/echolalia,Mannerisms,Verbigeration,Waxy flexibility,Withdrawal | 97.8 | 96.2 | 99.1 |
| 9 | Excitement,Immobility/stupor,Mutism,Posturing/catalepsy,Echopraxia/echolalia,Stereotypy,Mannerisms,Negativism,Waxy flexibility | 97.8 | 96.4 | 99.1 |
| 9 | Excitement,Immobility/stupor,Mutism,Posturing/catalepsy,Echopraxia/echolalia,Stereotypy,Mannerisms,Verbigeration,Withdrawal | 97.8 | 96.4 | 98.9 |
| 9 | Excitement,Immobility/stupor,Mutism,Posturing/catalepsy,Grimacing,Echopraxia/echolalia,Stereotypy,Mannerisms,Withdrawal | 97.8 | 96.2 | 98.9 |
| 9 | Excitement,Immobility/stupor,Mutism,Posturing/catalepsy,Grimacing,Mannerisms,Verbigeration,Waxy flexibility,Withdrawal | 97.8 | 96.2 | 99.1 |
| 9 | Excitement,Immobility/stupor,Mutism,Posturing/catalepsy,Grimacing,Stereotypy,Mannerisms,Negativism,Waxy flexibility | 97.8 | 96.2 | 99.1 |
| 9 | Excitement,Immobility/stupor,Mutism,Posturing/catalepsy,Grimacing,Stereotypy,Verbigeration,Waxy flexibility,Withdrawal | 97.8 | 96.4 | 99.1 |
| 9 | Excitement,Immobility/stupor,Posturing/catalepsy,Grimacing,Echopraxia/echolalia,Stereotypy,Mannerisms,Rigidity,Waxy flexibility | 97.8 | 96.4 | 99.1 |
| 9 | Excitement,Immobility/stupor,Posturing/catalepsy,Grimacing,Stereotypy,Mannerisms,Negativism,Waxy flexibility,Withdrawal | 97.8 | 96.2 | 99.1 |
| 9 | Excitement,Immobility/stupor,Posturing/catalepsy,Grimacing,Stereotypy,Mannerisms,Verbigeration,Negativism,Waxy flexibility | 97.8 | 96.4 | 99.1 |
| 9 | Excitement,Immobility/stupor,Posturing/catalepsy,Grimacing,Stereotypy,Mannerisms,Verbigeration,Rigidity,Waxy flexibility | 97.8 | 96.2 | 99.1 |
| 9 | Excitement,Staring,Posturing/catalepsy,Grimacing,Echopraxia/echolalia,Mannerisms,Verbigeration,Negativism,Waxy flexibility | 97.8 | 96.4 | 99.1 |
| 9 | Excitement,Staring,Posturing/catalepsy,Grimacing,Echopraxia/echolalia,Stereotypy,Mannerisms,Negativism,Waxy flexibility | 97.8 | 96.2 | 99.1 |
| 9 | Excitement,Staring,Posturing/catalepsy,Grimacing,Echopraxia/echolalia,Stereotypy,Mannerisms,Verbigeration,Negativism | 97.8 | 96.4 | 99.1 |
| 9 | Excitement,Staring,Posturing/catalepsy,Grimacing,Echopraxia/echolalia,Stereotypy,Verbigeration,Rigidity,Waxy flexibility | 97.8 | 96.2 | 99.1 |
| 9 | Excitement,Staring,Posturing/catalepsy,Grimacing,Echopraxia/echolalia,Stereotypy,Verbigeration,Waxy flexibility,Withdrawal | 97.8 | 96.2 | 99.1 |
| 9 | Immobility/stupor,Grimacing,Echopraxia/echolalia,Stereotypy,Mannerisms,Verbigeration,Negativism,Waxy flexibility,Withdrawal | 97.8 | 96.2 | 99.1 |
| 9 | Immobility/stupor,Mutism,Echopraxia/echolalia,Stereotypy,Mannerisms,Verbigeration,Negativism,Waxy flexibility,Withdrawal | 97.8 | 96.2 | 99.1 |
| 9 | Immobility/stupor,Mutism,Grimacing,Echopraxia/echolalia,Stereotypy,Verbigeration,Rigidity,Waxy flexibility,Withdrawal | 97.8 | 96.4 | 99.1 |
| 9 | Immobility/stupor,Mutism,Posturing/catalepsy,Grimacing,Echopraxia/echolalia,Stereotypy,Mannerisms,Negativism,Waxy flexibility | 97.8 | 96.2 | 99.1 |
| 9 | Immobility/stupor,Mutism,Posturing/catalepsy,Grimacing,Echopraxia/echolalia,Stereotypy,Mannerisms,Verbigeration,Negativism | 97.8 | 96.2 | 99.1 |
| 9 | Immobility/stupor,Mutism,Posturing/catalepsy,Grimacing,Echopraxia/echolalia,Stereotypy,Rigidity,Negativism,Waxy flexibility | 97.8 | 96.2 | 99.1 |
| 9 | Immobility/stupor,Mutism,Posturing/catalepsy,Grimacing,Echopraxia/echolalia,Stereotypy,Verbigeration,Negativism,Withdrawal | 97.8 | 96.4 | 99.1 |
| 9 | Immobility/stupor,Mutism,Posturing/catalepsy,Grimacing,Echopraxia/echolalia,Stereotypy,Verbigeration,Rigidity,Withdrawal | 97.8 | 96.2 | 99.1 |
| 9 | Immobility/stupor,Mutism,Posturing/catalepsy,Grimacing,Mannerisms,Verbigeration,Rigidity,Waxy flexibility,Withdrawal | 97.8 | 96.2 | 99.1 |
| 9 | Immobility/stupor,Mutism,Posturing/catalepsy,Grimacing,Stereotypy,Mannerisms,Rigidity,Waxy flexibility,Withdrawal | 97.8 | 96.4 | 99.1 |
| 9 | Immobility/stupor,Mutism,Staring,Grimacing,Echopraxia/echolalia,Stereotypy,Verbigeration,Waxy flexibility,Withdrawal | 97.8 | 96.4 | 99.1 |
| 9 | Immobility/stupor,Mutism,Staring,Posturing/catalepsy,Echopraxia/echolalia,Stereotypy,Mannerisms,Verbigeration,Waxy flexibility | 97.8 | 96.4 | 99.1 |
| 9 | Immobility/stupor,Mutism,Staring,Posturing/catalepsy,Echopraxia/echolalia,Verbigeration,Rigidity,Waxy flexibility,Withdrawal | 97.8 | 96.2 | 99.1 |
| 9 | Immobility/stupor,Mutism,Staring,Posturing/catalepsy,Grimacing,Echopraxia/echolalia,Rigidity,Negativism,Waxy flexibility | 97.8 | 96.4 | 99.1 |
| 9 | Immobility/stupor,Mutism,Staring,Posturing/catalepsy,Grimacing,Echopraxia/echolalia,Stereotypy,Verbigeration,Waxy flexibility | 97.8 | 96.4 | 99.1 |
| 9 | Immobility/stupor,Mutism,Staring,Posturing/catalepsy,Grimacing,Echopraxia/echolalia,Verbigeration,Rigidity,Waxy flexibility | 97.8 | 96.2 | 99.1 |
| 9 | Immobility/stupor,Mutism,Staring,Posturing/catalepsy,Grimacing,Rigidity,Negativism,Waxy flexibility,Withdrawal | 97.8 | 96.2 | 99.1 |
| 9 | Immobility/stupor,Mutism,Staring,Posturing/catalepsy,Grimacing,Verbigeration,Rigidity,Waxy flexibility,Withdrawal | 97.8 | 96.4 | 99.1 |
| 9 | Immobility/stupor,Posturing/catalepsy,Grimacing,Echopraxia/echolalia,Mannerisms,Verbigeration,Negativism,Waxy flexibility,Withdrawal | 97.8 | 96.2 | 99.1 |
| 9 | Immobility/stupor,Staring,Posturing/catalepsy,Echopraxia/echolalia,Stereotypy,Mannerisms,Verbigeration,Rigidity,Waxy flexibility | 97.8 | 96.2 | 99.1 |
| 9 | Immobility/stupor,Staring,Posturing/catalepsy,Grimacing,Echopraxia/echolalia,Mannerisms,Verbigeration,Waxy flexibility,Withdrawal | 97.8 | 96.4 | 99.1 |
| 9 | Immobility/stupor,Staring,Posturing/catalepsy,Grimacing,Echopraxia/echolalia,Stereotypy,Mannerisms,Verbigeration,Waxy flexibility | 97.8 | 96.2 | 99.1 |
| 9 | Staring,Posturing/catalepsy,Echopraxia/echolalia,Stereotypy,Mannerisms,Verbigeration,Rigidity,Waxy flexibility,Withdrawal | 97.8 | 96.4 | 99.1 |
| 9 | Staring,Posturing/catalepsy,Grimacing,Echopraxia/echolalia,Mannerisms,Verbigeration,Rigidity,Negativism,Waxy flexibility | 97.8 | 96.2 | 99.1 |
| 9 | Staring,Posturing/catalepsy,Grimacing,Echopraxia/echolalia,Stereotypy,Mannerisms,Verbigeration,Waxy flexibility,Withdrawal | 97.8 | 96.4 | 99.1 |
| 9 | Staring,Posturing/catalepsy,Grimacing,Echopraxia/echolalia,Stereotypy,Verbigeration,Negativism,Waxy flexibility,Withdrawal | 97.8 | 96.4 | 99.1 |
| 9 | Excitement,Immobility/stupor,Mutism,Grimacing,Echopraxia/echolalia,Mannerisms,Negativism,Waxy flexibility,Withdrawal | 97.7 | 96.2 | 99.1 |
| 9 | Excitement,Immobility/stupor,Posturing/catalepsy,Grimacing,Echopraxia/echolalia,Stereotypy,Verbigeration,Negativism,Waxy flexibility | 97.7 | 96.4 | 99.1 |
| 9 | Excitement,Staring,Posturing/catalepsy,Grimacing,Stereotypy,Mannerisms,Verbigeration,Negativism,Waxy flexibility | 97.7 | 96.2 | 99.1 |
| 9 | Immobility/stupor,Mutism,Staring,Posturing/catalepsy,Grimacing,Echopraxia/echolalia,Rigidity,Waxy flexibility,Withdrawal | 97.7 | 96.4 | 99.1 |
| 9 | Excitement,Immobility/stupor,Grimacing,Echopraxia/echolalia,Mannerisms,Verbigeration,Negativism,Waxy flexibility,Withdrawal | 97.5 | 96.0 | 98.9 |
| 9 | Excitement,Immobility/stupor,Mutism,Grimacing,Echopraxia/echolalia,Mannerisms,Verbigeration,Negativism,Waxy flexibility | 97.5 | 96.0 | 98.9 |
| 9 | Excitement,Immobility/stupor,Mutism,Grimacing,Echopraxia/echolalia,Stereotypy,Mannerisms,Negativism,Waxy flexibility | 97.5 | 96.0 | 98.9 |
| 9 | Excitement,Immobility/stupor,Mutism,Grimacing,Echopraxia/echolalia,Stereotypy,Verbigeration,Waxy flexibility,Withdrawal | 97.5 | 96.0 | 98.9 |
| 9 | Excitement,Immobility/stupor,Mutism,Grimacing,Mannerisms,Verbigeration,Negativism,Waxy flexibility,Withdrawal | 97.5 | 96.0 | 98.9 |
| 9 | Excitement,Immobility/stupor,Mutism,Grimacing,Stereotypy,Mannerisms,Negativism,Waxy flexibility,Withdrawal | 97.5 | 96.0 | 98.9 |
| 9 | Excitement,Immobility/stupor,Mutism,Grimacing,Stereotypy,Mannerisms,Verbigeration,Negativism,Waxy flexibility | 97.5 | 96.0 | 98.9 |
| 9 | Excitement,Immobility/stupor,Mutism,Posturing/catalepsy,Echopraxia/echolalia,Stereotypy,Mannerisms,Waxy flexibility,Withdrawal | 97.5 | 96.0 | 98.9 |
| 9 | Excitement,Immobility/stupor,Mutism,Posturing/catalepsy,Grimacing,Echopraxia/echolalia,Mannerisms,Verbigeration,Waxy flexibility | 97.5 | 96.0 | 98.9 |
| 9 | Excitement,Immobility/stupor,Mutism,Posturing/catalepsy,Grimacing,Echopraxia/echolalia,Stereotypy,Verbigeration,Waxy flexibility | 97.5 | 96.0 | 98.9 |
| 9 | Excitement,Immobility/stupor,Mutism,Posturing/catalepsy,Grimacing,Stereotypy,Mannerisms,Verbigeration,Withdrawal | 97.5 | 96.0 | 98.9 |
| 9 | Excitement,Immobility/stupor,Mutism,Posturing/catalepsy,Stereotypy,Mannerisms,Verbigeration,Waxy flexibility,Withdrawal | 97.5 | 96.0 | 98.9 |
| 9 | Excitement,Immobility/stupor,Posturing/catalepsy,Echopraxia/echolalia,Stereotypy,Mannerisms,Verbigeration,Negativism,Waxy flexibility | 97.5 | 96.0 | 98.9 |
| 9 | Excitement,Immobility/stupor,Posturing/catalepsy,Grimacing,Echopraxia/echolalia,Mannerisms,Verbigeration,Waxy flexibility,Withdrawal | 97.5 | 96.0 | 98.9 |
| 9 | Excitement,Immobility/stupor,Posturing/catalepsy,Grimacing,Echopraxia/echolalia,Stereotypy,Mannerisms,Negativism,Waxy flexibility | 97.5 | 96.0 | 98.9 |
| 9 | Excitement,Immobility/stupor,Posturing/catalepsy,Grimacing,Echopraxia/echolalia,Stereotypy,Mannerisms,Verbigeration,Negativism | 97.5 | 96.0 | 98.9 |
| 9 | Excitement,Staring,Posturing/catalepsy,Echopraxia/echolalia,Stereotypy,Mannerisms,Verbigeration,Rigidity,Waxy flexibility | 97.5 | 96.0 | 98.9 |
| 9 | Excitement,Staring,Posturing/catalepsy,Grimacing,Echopraxia/echolalia,Stereotypy,Verbigeration,Negativism,Waxy flexibility | 97.5 | 96.0 | 98.9 |
| 9 | Immobility/stupor,Grimacing,Echopraxia/echolalia,Stereotypy,Mannerisms,Verbigeration,Rigidity,Waxy flexibility,Withdrawal | 97.5 | 96.0 | 98.9 |
| 9 | Immobility/stupor,Mutism,Grimacing,Echopraxia/echolalia,Mannerisms,Verbigeration,Negativism,Waxy flexibility,Withdrawal | 97.5 | 96.0 | 98.9 |
| 9 | Immobility/stupor,Mutism,Grimacing,Echopraxia/echolalia,Stereotypy,Mannerisms,Verbigeration,Negativism,Waxy flexibility | 97.5 | 96.0 | 98.9 |
| 9 | Immobility/stupor,Mutism,Grimacing,Stereotypy,Mannerisms,Verbigeration,Negativism,Waxy flexibility,Withdrawal | 97.5 | 96.0 | 98.9 |
| 9 | Immobility/stupor,Mutism,Posturing/catalepsy,Echopraxia/echolalia,Stereotypy,Mannerisms,Verbigeration,Negativism,Waxy flexibility | 97.5 | 96.0 | 98.9 |
| 9 | Immobility/stupor,Mutism,Posturing/catalepsy,Echopraxia/echolalia,Stereotypy,Mannerisms,Verbigeration,Rigidity,Withdrawal | 97.5 | 96.0 | 98.9 |
| 9 | Immobility/stupor,Mutism,Posturing/catalepsy,Grimacing,Echopraxia/echolalia,Stereotypy,Mannerisms,Verbigeration,Rigidity | 97.5 | 96.0 | 98.9 |
| 9 | Immobility/stupor,Mutism,Posturing/catalepsy,Grimacing,Echopraxia/echolalia,Stereotypy,Negativism,Waxy flexibility,Withdrawal | 97.5 | 96.0 | 98.9 |
| 9 | Immobility/stupor,Mutism,Posturing/catalepsy,Grimacing,Echopraxia/echolalia,Stereotypy,Rigidity,Waxy flexibility,Withdrawal | 97.5 | 96.0 | 98.9 |
| 9 | Immobility/stupor,Mutism,Posturing/catalepsy,Grimacing,Echopraxia/echolalia,Verbigeration,Negativism,Waxy flexibility,Withdrawal | 97.5 | 96.0 | 98.9 |
| 9 | Immobility/stupor,Mutism,Posturing/catalepsy,Grimacing,Stereotypy,Mannerisms,Verbigeration,Negativism,Waxy flexibility | 97.5 | 96.0 | 98.9 |
| 9 | Immobility/stupor,Mutism,Posturing/catalepsy,Grimacing,Stereotypy,Verbigeration,Negativism,Waxy flexibility,Withdrawal | 97.5 | 96.0 | 98.9 |
| 9 | Immobility/stupor,Mutism,Posturing/catalepsy,Grimacing,Stereotypy,Verbigeration,Rigidity,Waxy flexibility,Withdrawal | 97.5 | 96.0 | 98.9 |
| 9 | Immobility/stupor,Mutism,Staring,Echopraxia/echolalia,Stereotypy,Mannerisms,Verbigeration,Waxy flexibility,Withdrawal | 97.5 | 96.0 | 98.9 |
| 9 | Immobility/stupor,Mutism,Staring,Posturing/catalepsy,Grimacing,Echopraxia/echolalia,Stereotypy,Rigidity,Waxy flexibility | 97.5 | 96.0 | 98.9 |
| 9 | Immobility/stupor,Mutism,Staring,Posturing/catalepsy,Grimacing,Echopraxia/echolalia,Verbigeration,Waxy flexibility,Withdrawal | 97.5 | 96.0 | 98.9 |
| 9 | Immobility/stupor,Posturing/catalepsy,Grimacing,Echopraxia/echolalia,Stereotypy,Mannerisms,Rigidity,Waxy flexibility,Withdrawal | 97.5 | 96.0 | 98.9 |
| 9 | Immobility/stupor,Posturing/catalepsy,Grimacing,Echopraxia/echolalia,Stereotypy,Mannerisms,Verbigeration,Negativism,Waxy flexibility | 97.5 | 96.0 | 98.9 |
| 9 | Immobility/stupor,Posturing/catalepsy,Grimacing,Echopraxia/echolalia,Stereotypy,Mannerisms,Verbigeration,Rigidity,Withdrawal | 97.5 | 96.0 | 98.9 |
| 9 | Immobility/stupor,Staring,Posturing/catalepsy,Echopraxia/echolalia,Stereotypy,Verbigeration,Rigidity,Waxy flexibility,Withdrawal | 97.5 | 96.0 | 98.9 |
| 9 | Immobility/stupor,Staring,Posturing/catalepsy,Grimacing,Echopraxia/echolalia,Stereotypy,Verbigeration,Rigidity,Waxy flexibility | 97.5 | 96.0 | 98.9 |
| 9 | Staring,Posturing/catalepsy,Grimacing,Echopraxia/echolalia,Stereotypy,Mannerisms,Verbigeration,Negativism,Waxy flexibility | 97.5 | 96.0 | 98.9 |
| 9 | Staring,Posturing/catalepsy,Grimacing,Echopraxia/echolalia,Stereotypy,Verbigeration,Rigidity,Waxy flexibility,Withdrawal | 97.5 | 96.0 | 98.9 |
| 9 | Excitement,Immobility/stupor,Grimacing,Echopraxia/echolalia,Stereotypy,Mannerisms,Verbigeration,Negativism,Waxy flexibility | 97.3 | 95.7 | 98.7 |
| 9 | Excitement,Immobility/stupor,Grimacing,Stereotypy,Mannerisms,Verbigeration,Negativism,Waxy flexibility,Withdrawal | 97.3 | 95.7 | 98.7 |
| 9 | Excitement,Immobility/stupor,Mutism,Echopraxia/echolalia,Stereotypy,Mannerisms,Verbigeration,Waxy flexibility,Withdrawal | 97.3 | 95.7 | 98.7 |
| 9 | Excitement,Immobility/stupor,Mutism,Grimacing,Echopraxia/echolalia,Stereotypy,Mannerisms,Verbigeration,Negativism | 97.3 | 95.7 | 98.7 |
| 9 | Excitement,Immobility/stupor,Mutism,Grimacing,Echopraxia/echolalia,Stereotypy,Mannerisms,Waxy flexibility,Withdrawal | 97.3 | 95.7 | 98.7 |
| 9 | Excitement,Immobility/stupor,Mutism,Posturing/catalepsy,Grimacing,Stereotypy,Mannerisms,Waxy flexibility,Withdrawal | 97.3 | 95.7 | 98.7 |
| 9 | Excitement,Immobility/stupor,Posturing/catalepsy,Echopraxia/echolalia,Stereotypy,Mannerisms,Verbigeration,Waxy flexibility,Withdrawal | 97.3 | 95.7 | 98.7 |
| 9 | Excitement,Immobility/stupor,Posturing/catalepsy,Grimacing,Echopraxia/echolalia,Stereotypy,Mannerisms,Verbigeration,Withdrawal | 97.3 | 95.7 | 98.7 |
| 9 | Excitement,Immobility/stupor,Posturing/catalepsy,Grimacing,Echopraxia/echolalia,Stereotypy,Mannerisms,Waxy flexibility,Withdrawal | 97.3 | 95.7 | 98.7 |
| 9 | Excitement,Staring,Grimacing,Echopraxia/echolalia,Stereotypy,Mannerisms,Verbigeration,Negativism,Waxy flexibility | 97.3 | 95.7 | 98.7 |
| 9 | Immobility/stupor,Mutism,Echopraxia/echolalia,Stereotypy,Mannerisms,Verbigeration,Rigidity,Waxy flexibility,Withdrawal | 97.3 | 95.7 | 98.7 |
| 9 | Immobility/stupor,Mutism,Grimacing,Echopraxia/echolalia,Stereotypy,Mannerisms,Verbigeration,Rigidity,Waxy flexibility | 97.3 | 95.7 | 98.7 |
| 9 | Immobility/stupor,Mutism,Grimacing,Echopraxia/echolalia,Stereotypy,Verbigeration,Negativism,Waxy flexibility,Withdrawal | 97.3 | 95.7 | 98.7 |
| 9 | Immobility/stupor,Mutism,Posturing/catalepsy,Echopraxia/echolalia,Mannerisms,Verbigeration,Rigidity,Waxy flexibility,Withdrawal | 97.3 | 95.7 | 98.7 |
| 9 | Immobility/stupor,Mutism,Posturing/catalepsy,Echopraxia/echolalia,Stereotypy,Mannerisms,Rigidity,Waxy flexibility,Withdrawal | 97.3 | 95.7 | 98.7 |
| 9 | Immobility/stupor,Mutism,Posturing/catalepsy,Echopraxia/echolalia,Stereotypy,Verbigeration,Negativism,Waxy flexibility,Withdrawal | 97.3 | 95.7 | 98.7 |
| 9 | Immobility/stupor,Mutism,Posturing/catalepsy,Grimacing,Echopraxia/echolalia,Mannerisms,Verbigeration,Rigidity,Waxy flexibility | 97.3 | 95.7 | 98.7 |
| 9 | Immobility/stupor,Mutism,Posturing/catalepsy,Grimacing,Echopraxia/echolalia,Stereotypy,Verbigeration,Negativism,Waxy flexibility | 97.3 | 95.7 | 98.7 |
| 9 | Immobility/stupor,Mutism,Posturing/catalepsy,Grimacing,Echopraxia/echolalia,Verbigeration,Rigidity,Waxy flexibility,Withdrawal | 97.3 | 95.7 | 98.7 |
| 9 | Immobility/stupor,Mutism,Posturing/catalepsy,Grimacing,Stereotypy,Mannerisms,Verbigeration,Rigidity,Waxy flexibility | 97.3 | 95.7 | 98.7 |
| 9 | Immobility/stupor,Mutism,Posturing/catalepsy,Stereotypy,Mannerisms,Verbigeration,Rigidity,Waxy flexibility,Withdrawal | 97.3 | 95.7 | 98.7 |
| 9 | Immobility/stupor,Mutism,Staring,Posturing/catalepsy,Echopraxia/echolalia,Stereotypy,Verbigeration,Waxy flexibility,Withdrawal | 97.3 | 95.7 | 98.7 |
| 9 | Immobility/stupor,Posturing/catalepsy,Grimacing,Echopraxia/echolalia,Mannerisms,Verbigeration,Rigidity,Waxy flexibility,Withdrawal | 97.3 | 95.7 | 98.7 |
| 9 | Immobility/stupor,Posturing/catalepsy,Grimacing,Echopraxia/echolalia,Stereotypy,Verbigeration,Negativism,Waxy flexibility,Withdrawal | 97.3 | 95.7 | 98.7 |
| 9 | Immobility/stupor,Staring,Posturing/catalepsy,Echopraxia/echolalia,Stereotypy,Mannerisms,Verbigeration,Waxy flexibility,Withdrawal | 97.3 | 95.7 | 98.7 |
| 9 | Immobility/stupor,Staring,Posturing/catalepsy,Grimacing,Echopraxia/echolalia,Verbigeration,Rigidity,Waxy flexibility,Withdrawal | 97.3 | 95.7 | 98.7 |
| 9 | Excitement,Immobility/stupor,Mutism,Grimacing,Echopraxia/echolalia,Mannerisms,Verbigeration,Waxy flexibility,Withdrawal | 97.1 | 95.5 | 98.4 |
| 9 | Excitement,Immobility/stupor,Mutism,Grimacing,Echopraxia/echolalia,Stereotypy,Mannerisms,Verbigeration,Withdrawal | 97.1 | 95.5 | 98.4 |
| 9 | Excitement,Immobility/stupor,Mutism,Posturing/catalepsy,Echopraxia/echolalia,Stereotypy,Mannerisms,Verbigeration,Waxy flexibility | 97.1 | 95.5 | 98.7 |
| 9 | Excitement,Immobility/stupor,Mutism,Posturing/catalepsy,Grimacing,Echopraxia/echolalia,Stereotypy,Mannerisms,Verbigeration | 97.1 | 95.5 | 98.7 |
| 9 | Excitement,Immobility/stupor,Mutism,Posturing/catalepsy,Grimacing,Echopraxia/echolalia,Stereotypy,Mannerisms,Waxy flexibility | 97.1 | 95.5 | 98.7 |
| 9 | Excitement,Immobility/stupor,Mutism,Posturing/catalepsy,Grimacing,Stereotypy,Mannerisms,Verbigeration,Waxy flexibility | 97.1 | 95.5 | 98.4 |
| 9 | Excitement,Immobility/stupor,Posturing/catalepsy,Grimacing,Echopraxia/echolalia,Stereotypy,Verbigeration,Waxy flexibility,Withdrawal | 97.1 | 95.5 | 98.7 |
| 9 | Excitement,Mutism,Posturing/catalepsy,Echopraxia/echolalia,Mannerisms,Verbigeration,Rigidity,Negativism,Withdrawal | 97.1 | 95.5 | 98.4 |
| 9 | Excitement,Mutism,Posturing/catalepsy,Echopraxia/echolalia,Stereotypy,Verbigeration,Rigidity,Negativism,Withdrawal | 97.1 | 95.5 | 98.4 |
| 9 | Excitement,Mutism,Posturing/catalepsy,Echopraxia/echolalia,Verbigeration,Rigidity,Negativism,Waxy flexibility,Withdrawal | 97.1 | 95.5 | 98.4 |
| 9 | Excitement,Mutism,Posturing/catalepsy,Grimacing,Echopraxia/echolalia,Verbigeration,Rigidity,Negativism,Withdrawal | 97.1 | 95.5 | 98.4 |
| 9 | Excitement,Mutism,Posturing/catalepsy,Grimacing,Mannerisms,Verbigeration,Rigidity,Negativism,Withdrawal | 97.1 | 95.5 | 98.4 |
| 9 | Excitement,Mutism,Posturing/catalepsy,Mannerisms,Verbigeration,Rigidity,Negativism,Waxy flexibility,Withdrawal | 97.1 | 95.5 | 98.7 |
| 9 | Excitement,Mutism,Posturing/catalepsy,Stereotypy,Mannerisms,Verbigeration,Rigidity,Negativism,Withdrawal | 97.1 | 95.5 | 98.4 |
| 9 | Excitement,Mutism,Posturing/catalepsy,Stereotypy,Verbigeration,Rigidity,Negativism,Waxy flexibility,Withdrawal | 97.1 | 95.5 | 98.4 |
| 9 | Excitement,Staring,Posturing/catalepsy,Grimacing,Echopraxia/echolalia,Stereotypy,Mannerisms,Verbigeration,Waxy flexibility | 97.1 | 95.5 | 98.4 |
| 9 | Immobility/stupor,Mutism,Posturing/catalepsy,Grimacing,Echopraxia/echolalia,Mannerisms,Verbigeration,Waxy flexibility,Withdrawal | 97.1 | 95.5 | 98.7 |
| 9 | Immobility/stupor,Mutism,Posturing/catalepsy,Grimacing,Echopraxia/echolalia,Stereotypy,Mannerisms,Rigidity,Waxy flexibility | 97.1 | 95.5 | 98.7 |
| 9 | Immobility/stupor,Mutism,Posturing/catalepsy,Grimacing,Echopraxia/echolalia,Stereotypy,Mannerisms,Waxy flexibility,Withdrawal | 97.1 | 95.5 | 98.4 |
| 9 | Immobility/stupor,Mutism,Posturing/catalepsy,Grimacing,Echopraxia/echolalia,Stereotypy,Verbigeration,Rigidity,Waxy flexibility | 97.1 | 95.3 | 98.7 |
| 9 | Immobility/stupor,Posturing/catalepsy,Grimacing,Stereotypy,Mannerisms,Verbigeration,Rigidity,Waxy flexibility,Withdrawal | 97.1 | 95.3 | 98.4 |
| 9 | Immobility/stupor,Staring,Posturing/catalepsy,Grimacing,Echopraxia/echolalia,Stereotypy,Verbigeration,Waxy flexibility,Withdrawal | 97.1 | 95.5 | 98.4 |
| 9 | Staring,Posturing/catalepsy,Grimacing,Echopraxia/echolalia,Stereotypy,Mannerisms,Verbigeration,Rigidity,Waxy flexibility | 97.1 | 95.5 | 98.4 |
| 9 | Staring,Posturing/catalepsy,Grimacing,Echopraxia/echolalia,Stereotypy,Verbigeration,Rigidity,Negativism,Waxy flexibility | 97.1 | 95.5 | 98.4 |
| 9 | Excitement,Immobility/stupor,Grimacing,Echopraxia/echolalia,Stereotypy,Mannerisms,Verbigeration,Waxy flexibility,Withdrawal | 96.9 | 95.3 | 98.4 |
| 9 | Excitement,Immobility/stupor,Mutism,Grimacing,Stereotypy,Mannerisms,Verbigeration,Waxy flexibility,Withdrawal | 96.9 | 95.1 | 98.4 |
| 9 | Excitement,Immobility/stupor,Posturing/catalepsy,Grimacing,Stereotypy,Mannerisms,Verbigeration,Waxy flexibility,Withdrawal | 96.9 | 95.1 | 98.4 |
| 9 | Excitement,Mutism,Echopraxia/echolalia,Stereotypy,Verbigeration,Rigidity,Negativism,Waxy flexibility,Withdrawal | 96.9 | 95.1 | 98.4 |
| 9 | Excitement,Mutism,Posturing/catalepsy,Grimacing,Echopraxia/echolalia,Mannerisms,Rigidity,Negativism,Withdrawal | 96.9 | 95.3 | 98.4 |
| 9 | Excitement,Mutism,Posturing/catalepsy,Grimacing,Echopraxia/echolalia,Rigidity,Negativism,Waxy flexibility,Withdrawal | 96.9 | 95.1 | 98.4 |
| 9 | Excitement,Mutism,Posturing/catalepsy,Grimacing,Stereotypy,Verbigeration,Rigidity,Negativism,Withdrawal | 96.9 | 95.1 | 98.4 |
| 9 | Excitement,Mutism,Posturing/catalepsy,Grimacing,Verbigeration,Rigidity,Negativism,Waxy flexibility,Withdrawal | 96.9 | 95.1 | 98.4 |
| 9 | Excitement,Mutism,Stereotypy,Mannerisms,Verbigeration,Rigidity,Negativism,Waxy flexibility,Withdrawal | 96.9 | 95.1 | 98.4 |
| 9 | Immobility/stupor,Mutism,Posturing/catalepsy,Echopraxia/echolalia,Stereotypy,Mannerisms,Verbigeration,Rigidity,Waxy flexibility | 96.9 | 95.1 | 98.4 |
| 9 | Immobility/stupor,Mutism,Posturing/catalepsy,Echopraxia/echolalia,Stereotypy,Verbigeration,Rigidity,Waxy flexibility,Withdrawal | 96.9 | 95.1 | 98.4 |
| 9 | Immobility/stupor,Mutism,Posturing/catalepsy,Grimacing,Echopraxia/echolalia,Stereotypy,Mannerisms,Verbigeration,Withdrawal | 96.9 | 95.1 | 98.4 |
| 9 | Immobility/stupor,Posturing/catalepsy,Grimacing,Echopraxia/echolalia,Stereotypy,Verbigeration,Rigidity,Waxy flexibility,Withdrawal | 96.9 | 95.1 | 98.4 |
| 9 | Mutism,Posturing/catalepsy,Stereotypy,Mannerisms,Verbigeration,Rigidity,Negativism,Waxy flexibility,Withdrawal | 96.9 | 95.1 | 98.4 |
| 9 | Excitement,Mutism,Posturing/catalepsy,Echopraxia/echolalia,Mannerisms,Rigidity,Negativism,Waxy flexibility,Withdrawal | 96.7 | 94.8 | 98.2 |
| 9 | Excitement,Immobility/stupor,Mutism,Grimacing,Echopraxia/echolalia,Stereotypy,Mannerisms,Verbigeration,Waxy flexibility | 96.6 | 94.8 | 98.2 |
| 9 | Excitement,Mutism,Echopraxia/echolalia,Stereotypy,Mannerisms,Verbigeration,Rigidity,Negativism,Withdrawal | 96.6 | 94.8 | 98.2 |
| 9 | Excitement,Mutism,Grimacing,Stereotypy,Verbigeration,Rigidity,Negativism,Waxy flexibility,Withdrawal | 96.6 | 94.8 | 98.2 |
| 9 | Excitement,Mutism,Posturing/catalepsy,Echopraxia/echolalia,Stereotypy,Mannerisms,Rigidity,Negativism,Withdrawal | 96.6 | 94.8 | 98.2 |
| 9 | Excitement,Mutism,Posturing/catalepsy,Echopraxia/echolalia,Stereotypy,Rigidity,Negativism,Waxy flexibility,Withdrawal | 96.6 | 94.8 | 98.2 |
| 9 | Excitement,Mutism,Posturing/catalepsy,Grimacing,Echopraxia/echolalia,Stereotypy,Rigidity,Negativism,Withdrawal | 96.6 | 94.8 | 98.2 |
| 9 | Excitement,Mutism,Posturing/catalepsy,Grimacing,Mannerisms,Rigidity,Negativism,Waxy flexibility,Withdrawal | 96.6 | 94.8 | 98.2 |
| 9 | Immobility/stupor,Posturing/catalepsy,Echopraxia/echolalia,Stereotypy,Mannerisms,Verbigeration,Rigidity,Waxy flexibility,Withdrawal | 96.6 | 94.8 | 98.2 |
| 9 | Mutism,Echopraxia/echolalia,Stereotypy,Mannerisms,Verbigeration,Rigidity,Negativism,Waxy flexibility,Withdrawal | 96.6 | 94.8 | 98.2 |
| 9 | Mutism,Grimacing,Stereotypy,Mannerisms,Verbigeration,Rigidity,Negativism,Waxy flexibility,Withdrawal | 96.6 | 94.8 | 98.2 |
| 9 | Mutism,Posturing/catalepsy,Echopraxia/echolalia,Mannerisms,Verbigeration,Rigidity,Negativism,Waxy flexibility,Withdrawal | 96.6 | 94.8 | 98.2 |
| 9 | Mutism,Posturing/catalepsy,Echopraxia/echolalia,Stereotypy,Mannerisms,Verbigeration,Rigidity,Negativism,Withdrawal | 96.6 | 94.8 | 98.2 |
| 9 | Mutism,Posturing/catalepsy,Grimacing,Echopraxia/echolalia,Mannerisms,Rigidity,Negativism,Waxy flexibility,Withdrawal | 96.6 | 94.8 | 98.2 |
| 9 | Mutism,Posturing/catalepsy,Grimacing,Echopraxia/echolalia,Mannerisms,Verbigeration,Rigidity,Negativism,Withdrawal | 96.6 | 94.8 | 98.2 |
| 9 | Mutism,Posturing/catalepsy,Grimacing,Mannerisms,Verbigeration,Rigidity,Negativism,Waxy flexibility,Withdrawal | 96.6 | 94.8 | 98.2 |
| 9 | Mutism,Posturing/catalepsy,Grimacing,Stereotypy,Mannerisms,Verbigeration,Rigidity,Negativism,Withdrawal | 96.6 | 94.8 | 98.2 |
| 9 | Excitement,Immobility/stupor,Posturing/catalepsy,Grimacing,Echopraxia/echolalia,Stereotypy,Mannerisms,Verbigeration,Waxy flexibility | 96.4 | 94.6 | 98.0 |
| 9 | Excitement,Mutism,Echopraxia/echolalia,Stereotypy,Mannerisms,Rigidity,Negativism,Waxy flexibility,Withdrawal | 96.4 | 94.6 | 98.0 |
| 9 | Excitement,Mutism,Grimacing,Echopraxia/echolalia,Stereotypy,Rigidity,Negativism,Waxy flexibility,Withdrawal | 96.4 | 94.6 | 98.0 |
| 9 | Excitement,Mutism,Grimacing,Echopraxia/echolalia,Stereotypy,Verbigeration,Rigidity,Negativism,Withdrawal | 96.4 | 94.6 | 98.0 |
| 9 | Excitement,Mutism,Grimacing,Echopraxia/echolalia,Verbigeration,Rigidity,Negativism,Waxy flexibility,Withdrawal | 96.4 | 94.6 | 98.0 |
| 9 | Excitement,Mutism,Grimacing,Mannerisms,Verbigeration,Rigidity,Negativism,Waxy flexibility,Withdrawal | 96.4 | 94.6 | 98.0 |
| 9 | Excitement,Mutism,Grimacing,Stereotypy,Mannerisms,Verbigeration,Rigidity,Negativism,Withdrawal | 96.4 | 94.6 | 98.0 |
| 9 | Excitement,Mutism,Posturing/catalepsy,Echopraxia/echolalia,Mannerisms,Verbigeration,Rigidity,Waxy flexibility,Withdrawal | 96.4 | 94.6 | 98.0 |
| 9 | Excitement,Mutism,Posturing/catalepsy,Echopraxia/echolalia,Stereotypy,Verbigeration,Rigidity,Waxy flexibility,Withdrawal | 96.4 | 94.6 | 98.0 |
| 9 | Excitement,Mutism,Posturing/catalepsy,Grimacing,Echopraxia/echolalia,Mannerisms,Verbigeration,Rigidity,Negativism | 96.4 | 94.6 | 98.0 |
| 9 | Excitement,Mutism,Posturing/catalepsy,Grimacing,Echopraxia/echolalia,Verbigeration,Rigidity,Negativism,Waxy flexibility | 96.4 | 94.6 | 98.0 |
| 9 | Excitement,Mutism,Posturing/catalepsy,Grimacing,Echopraxia/echolalia,Verbigeration,Rigidity,Waxy flexibility,Withdrawal | 96.4 | 94.6 | 98.0 |
| 9 | Excitement,Mutism,Posturing/catalepsy,Grimacing,Mannerisms,Verbigeration,Rigidity,Waxy flexibility,Withdrawal | 96.4 | 94.6 | 98.0 |
| 9 | Excitement,Mutism,Posturing/catalepsy,Grimacing,Stereotypy,Mannerisms,Rigidity,Negativism,Withdrawal | 96.4 | 94.6 | 98.0 |
| 9 | Excitement,Mutism,Posturing/catalepsy,Grimacing,Stereotypy,Rigidity,Negativism,Waxy flexibility,Withdrawal | 96.4 | 94.6 | 98.0 |
| 9 | Excitement,Mutism,Posturing/catalepsy,Stereotypy,Mannerisms,Rigidity,Negativism,Waxy flexibility,Withdrawal | 96.4 | 94.6 | 98.0 |
| 9 | Excitement,Mutism,Posturing/catalepsy,Stereotypy,Mannerisms,Verbigeration,Rigidity,Waxy flexibility,Withdrawal | 96.4 | 94.6 | 98.0 |
| 9 | Immobility/stupor,Mutism,Grimacing,Echopraxia/echolalia,Stereotypy,Mannerisms,Verbigeration,Waxy flexibility,Withdrawal | 96.4 | 94.6 | 98.0 |
| 9 | Immobility/stupor,Mutism,Posturing/catalepsy,Grimacing,Echopraxia/echolalia,Stereotypy,Verbigeration,Waxy flexibility,Withdrawal | 96.4 | 94.6 | 98.0 |
| 9 | Immobility/stupor,Mutism,Posturing/catalepsy,Grimacing,Stereotypy,Mannerisms,Verbigeration,Waxy flexibility,Withdrawal | 96.4 | 94.6 | 98.0 |
| 9 | Immobility/stupor,Posturing/catalepsy,Grimacing,Echopraxia/echolalia,Stereotypy,Mannerisms,Verbigeration,Rigidity,Waxy flexibility | 96.4 | 94.6 | 98.0 |
| 9 | Mutism,Grimacing,Echopraxia/echolalia,Stereotypy,Mannerisms,Rigidity,Negativism,Waxy flexibility,Withdrawal | 96.4 | 94.6 | 98.0 |
| 9 | Mutism,Posturing/catalepsy,Echopraxia/echolalia,Stereotypy,Mannerisms,Rigidity,Negativism,Waxy flexibility,Withdrawal | 96.4 | 94.6 | 98.0 |
| 9 | Mutism,Posturing/catalepsy,Grimacing,Echopraxia/echolalia,Stereotypy,Mannerisms,Rigidity,Negativism,Withdrawal | 96.4 | 94.6 | 98.0 |
| 9 | Mutism,Posturing/catalepsy,Grimacing,Stereotypy,Mannerisms,Rigidity,Negativism,Waxy flexibility,Withdrawal | 96.4 | 94.6 | 98.0 |
| 9 | Excitement,Mutism,Echopraxia/echolalia,Mannerisms,Verbigeration,Rigidity,Negativism,Waxy flexibility,Withdrawal | 96.2 | 94.4 | 98.0 |
| 9 | Excitement,Mutism,Grimacing,Echopraxia/echolalia,Mannerisms,Rigidity,Negativism,Waxy flexibility,Withdrawal | 96.2 | 94.4 | 97.8 |
| 9 | Excitement,Mutism,Grimacing,Echopraxia/echolalia,Mannerisms,Verbigeration,Rigidity,Negativism,Withdrawal | 96.2 | 94.4 | 98.0 |
| 9 | Excitement,Mutism,Grimacing,Echopraxia/echolalia,Stereotypy,Mannerisms,Rigidity,Negativism,Withdrawal | 96.2 | 94.4 | 97.8 |
| 9 | Excitement,Mutism,Grimacing,Stereotypy,Mannerisms,Rigidity,Negativism,Waxy flexibility,Withdrawal | 96.2 | 94.4 | 97.8 |
| 9 | Excitement,Mutism,Posturing/catalepsy,Echopraxia/echolalia,Mannerisms,Verbigeration,Rigidity,Negativism,Waxy flexibility | 96.2 | 94.4 | 97.8 |
| 9 | Excitement,Mutism,Posturing/catalepsy,Echopraxia/echolalia,Stereotypy,Mannerisms,Verbigeration,Rigidity,Negativism | 96.2 | 94.4 | 98.0 |
| 9 | Excitement,Mutism,Posturing/catalepsy,Echopraxia/echolalia,Stereotypy,Verbigeration,Rigidity,Negativism,Waxy flexibility | 96.2 | 94.4 | 97.8 |
| 9 | Excitement,Mutism,Posturing/catalepsy,Grimacing,Echopraxia/echolalia,Mannerisms,Rigidity,Waxy flexibility,Withdrawal | 96.2 | 94.4 | 97.8 |
| 9 | Excitement,Mutism,Posturing/catalepsy,Grimacing,Echopraxia/echolalia,Stereotypy,Verbigeration,Rigidity,Negativism | 96.2 | 94.4 | 98.0 |
| 9 | Excitement,Mutism,Posturing/catalepsy,Grimacing,Mannerisms,Verbigeration,Rigidity,Negativism,Waxy flexibility | 96.2 | 94.4 | 97.8 |
| 9 | Excitement,Mutism,Posturing/catalepsy,Grimacing,Stereotypy,Verbigeration,Rigidity,Waxy flexibility,Withdrawal | 96.2 | 94.4 | 97.8 |
| 9 | Immobility/stupor,Mutism,Posturing/catalepsy,Grimacing,Echopraxia/echolalia,Stereotypy,Mannerisms,Verbigeration,Waxy flexibility | 96.2 | 94.4 | 97.8 |
| 9 | Immobility/stupor,Posturing/catalepsy,Grimacing,Echopraxia/echolalia,Stereotypy,Mannerisms,Verbigeration,Waxy flexibility,Withdrawal | 96.2 | 94.4 | 98.0 |
| 9 | Mutism,Grimacing,Echopraxia/echolalia,Mannerisms,Verbigeration,Rigidity,Negativism,Waxy flexibility,Withdrawal | 96.2 | 94.2 | 98.0 |
| 9 | Mutism,Grimacing,Echopraxia/echolalia,Stereotypy,Mannerisms,Verbigeration,Rigidity,Negativism,Withdrawal | 96.2 | 94.4 | 97.8 |
| 9 | Mutism,Posturing/catalepsy,Grimacing,Stereotypy,Verbigeration,Rigidity,Negativism,Waxy flexibility,Withdrawal | 96.2 | 94.4 | 97.8 |
| 9 | Excitement,Mutism,Posturing/catalepsy,Echopraxia/echolalia,Stereotypy,Mannerisms,Rigidity,Waxy flexibility,Withdrawal | 96.0 | 94.2 | 97.8 |
| 9 | Excitement,Mutism,Posturing/catalepsy,Grimacing,Echopraxia/echolalia,Mannerisms,Rigidity,Negativism,Waxy flexibility | 96.0 | 93.9 | 97.8 |
| 9 | Excitement,Mutism,Posturing/catalepsy,Grimacing,Echopraxia/echolalia,Stereotypy,Rigidity,Waxy flexibility,Withdrawal | 96.0 | 93.9 | 97.8 |
| 9 | Excitement,Mutism,Posturing/catalepsy,Grimacing,Stereotypy,Mannerisms,Verbigeration,Rigidity,Negativism | 96.0 | 93.9 | 97.8 |
| 9 | Excitement,Mutism,Posturing/catalepsy,Grimacing,Stereotypy,Verbigeration,Rigidity,Negativism,Waxy flexibility | 96.0 | 93.9 | 97.8 |
| 9 | Excitement,Mutism,Posturing/catalepsy,Stereotypy,Mannerisms,Verbigeration,Rigidity,Negativism,Waxy flexibility | 96.0 | 94.2 | 97.8 |
| 9 | Immobility/stupor,Mutism,Posturing/catalepsy,Echopraxia/echolalia,Stereotypy,Mannerisms,Verbigeration,Waxy flexibility,Withdrawal | 96.0 | 93.9 | 97.8 |
| 9 | Mutism,Grimacing,Echopraxia/echolalia,Stereotypy,Verbigeration,Rigidity,Negativism,Waxy flexibility,Withdrawal | 96.0 | 94.2 | 97.8 |
| 9 | Mutism,Posturing/catalepsy,Echopraxia/echolalia,Stereotypy,Verbigeration,Rigidity,Negativism,Waxy flexibility,Withdrawal | 96.0 | 93.9 | 97.8 |
| 9 | Mutism,Posturing/catalepsy,Grimacing,Echopraxia/echolalia,Mannerisms,Verbigeration,Rigidity,Negativism,Waxy flexibility | 96.0 | 93.9 | 97.8 |
| 9 | Mutism,Posturing/catalepsy,Grimacing,Echopraxia/echolalia,Stereotypy,Verbigeration,Rigidity,Negativism,Withdrawal | 96.0 | 93.9 | 97.8 |
| 9 | Mutism,Posturing/catalepsy,Grimacing,Echopraxia/echolalia,Verbigeration,Rigidity,Negativism,Waxy flexibility,Withdrawal | 96.0 | 93.9 | 97.5 |
| 9 | Excitement,Mutism,Posturing/catalepsy,Grimacing,Echopraxia/echolalia,Stereotypy,Mannerisms,Rigidity,Negativism | 95.8 | 93.7 | 97.5 |
| 9 | Excitement,Mutism,Posturing/catalepsy,Grimacing,Echopraxia/echolalia,Stereotypy,Rigidity,Negativism,Waxy flexibility | 95.8 | 93.7 | 97.5 |
| 9 | Mutism,Posturing/catalepsy,Echopraxia/echolalia,Stereotypy,Mannerisms,Verbigeration,Rigidity,Negativism,Waxy flexibility | 95.8 | 93.7 | 97.5 |
| 9 | Mutism,Posturing/catalepsy,Grimacing,Echopraxia/echolalia,Stereotypy,Rigidity,Negativism,Waxy flexibility,Withdrawal | 95.8 | 93.7 | 97.5 |
| 9 | Excitement,Mutism,Echopraxia/echolalia,Stereotypy,Mannerisms,Verbigeration,Rigidity,Negativism,Waxy flexibility | 95.7 | 93.7 | 97.5 |
| 9 | Excitement,Mutism,Echopraxia/echolalia,Stereotypy,Mannerisms,Verbigeration,Rigidity,Waxy flexibility,Withdrawal | 95.7 | 93.7 | 97.5 |
| 9 | Excitement,Mutism,Grimacing,Echopraxia/echolalia,Stereotypy,Verbigeration,Rigidity,Negativism,Waxy flexibility | 95.7 | 93.7 | 97.5 |
| 9 | Excitement,Mutism,Posturing/catalepsy,Echopraxia/echolalia,Stereotypy,Mannerisms,Verbigeration,Rigidity,Withdrawal | 95.7 | 93.7 | 97.5 |
| 9 | Excitement,Mutism,Posturing/catalepsy,Grimacing,Echopraxia/echolalia,Mannerisms,Verbigeration,Rigidity,Withdrawal | 95.7 | 93.7 | 97.5 |
| 9 | Excitement,Mutism,Posturing/catalepsy,Grimacing,Stereotypy,Mannerisms,Rigidity,Waxy flexibility,Withdrawal | 95.7 | 93.7 | 97.5 |
| 9 | Mutism,Posturing/catalepsy,Grimacing,Echopraxia/echolalia,Stereotypy,Mannerisms,Verbigeration,Rigidity,Negativism | 95.7 | 93.7 | 97.5 |
| 9 | Mutism,Posturing/catalepsy,Grimacing,Stereotypy,Mannerisms,Verbigeration,Rigidity,Negativism,Waxy flexibility | 95.7 | 93.7 | 97.5 |
| 9 | Excitement,Mutism,Grimacing,Echopraxia/echolalia,Stereotypy,Verbigeration,Rigidity,Waxy flexibility,Withdrawal | 95.5 | 93.5 | 97.3 |
| 9 | Excitement,Mutism,Grimacing,Stereotypy,Mannerisms,Verbigeration,Rigidity,Negativism,Waxy flexibility | 95.5 | 93.5 | 97.3 |
| 9 | Excitement,Mutism,Grimacing,Stereotypy,Mannerisms,Verbigeration,Rigidity,Waxy flexibility,Withdrawal | 95.5 | 93.5 | 97.3 |
| 9 | Excitement,Mutism,Posturing/catalepsy,Echopraxia/echolalia,Stereotypy,Mannerisms,Rigidity,Negativism,Waxy flexibility | 95.5 | 93.5 | 97.3 |
| 9 | Excitement,Mutism,Posturing/catalepsy,Grimacing,Echopraxia/echolalia,Stereotypy,Verbigeration,Rigidity,Withdrawal | 95.5 | 93.5 | 97.3 |
| 9 | Excitement,Mutism,Posturing/catalepsy,Grimacing,Stereotypy,Mannerisms,Verbigeration,Rigidity,Withdrawal | 95.5 | 93.5 | 97.3 |
| 9 | Excitement,Posturing/catalepsy,Echopraxia/echolalia,Mannerisms,Verbigeration,Rigidity,Negativism,Waxy flexibility,Withdrawal | 95.5 | 93.5 | 97.3 |
| 9 | Excitement,Posturing/catalepsy,Echopraxia/echolalia,Stereotypy,Mannerisms,Verbigeration,Rigidity,Negativism,Withdrawal | 95.5 | 93.5 | 97.3 |
| 9 | Excitement,Posturing/catalepsy,Grimacing,Echopraxia/echolalia,Mannerisms,Verbigeration,Rigidity,Negativism,Withdrawal | 95.5 | 93.5 | 97.3 |
| 9 | Excitement,Posturing/catalepsy,Stereotypy,Mannerisms,Verbigeration,Rigidity,Negativism,Waxy flexibility,Withdrawal | 95.5 | 93.5 | 97.3 |
| 9 | Mutism,Grimacing,Echopraxia/echolalia,Stereotypy,Mannerisms,Verbigeration,Rigidity,Negativism,Waxy flexibility | 95.5 | 93.5 | 97.3 |
| 9 | Mutism,Posturing/catalepsy,Grimacing,Echopraxia/echolalia,Mannerisms,Verbigeration,Rigidity,Waxy flexibility,Withdrawal | 95.5 | 93.5 | 97.3 |
| 9 | Mutism,Posturing/catalepsy,Grimacing,Echopraxia/echolalia,Stereotypy,Mannerisms,Rigidity,Negativism,Waxy flexibility | 95.5 | 93.5 | 97.3 |
| 9 | Posturing/catalepsy,Echopraxia/echolalia,Stereotypy,Mannerisms,Verbigeration,Rigidity,Negativism,Waxy flexibility,Withdrawal | 95.5 | 93.5 | 97.3 |
| 9 | Posturing/catalepsy,Grimacing,Stereotypy,Mannerisms,Verbigeration,Rigidity,Negativism,Waxy flexibility,Withdrawal | 95.5 | 93.5 | 97.3 |
| 9 | Excitement,Mutism,Grimacing,Echopraxia/echolalia,Mannerisms,Verbigeration,Rigidity,Negativism,Waxy flexibility | 95.3 | 93.3 | 97.1 |
| 9 | Excitement,Mutism,Grimacing,Echopraxia/echolalia,Mannerisms,Verbigeration,Rigidity,Waxy flexibility,Withdrawal | 95.3 | 93.3 | 97.1 |
| 9 | Excitement,Mutism,Grimacing,Echopraxia/echolalia,Stereotypy,Mannerisms,Rigidity,Negativism,Waxy flexibility | 95.3 | 93.3 | 97.1 |
| 9 | Excitement,Mutism,Grimacing,Echopraxia/echolalia,Stereotypy,Mannerisms,Rigidity,Waxy flexibility,Withdrawal | 95.3 | 93.3 | 97.3 |
| 9 | Excitement,Mutism,Posturing/catalepsy,Echopraxia/echolalia,Stereotypy,Mannerisms,Verbigeration,Negativism,Withdrawal | 95.3 | 93.3 | 97.1 |
| 9 | Excitement,Mutism,Posturing/catalepsy,Echopraxia/echolalia,Stereotypy,Verbigeration,Negativism,Waxy flexibility,Withdrawal | 95.3 | 93.3 | 97.1 |
| 9 | Excitement,Mutism,Posturing/catalepsy,Grimacing,Echopraxia/echolalia,Mannerisms,Verbigeration,Rigidity,Waxy flexibility | 95.3 | 93.3 | 97.1 |
| 9 | Excitement,Mutism,Posturing/catalepsy,Grimacing,Echopraxia/echolalia,Stereotypy,Mannerisms,Rigidity,Withdrawal | 95.3 | 93.3 | 97.1 |
| 9 | Excitement,Mutism,Posturing/catalepsy,Stereotypy,Mannerisms,Verbigeration,Negativism,Waxy flexibility,Withdrawal | 95.3 | 93.3 | 97.1 |
| 9 | Excitement,Posturing/catalepsy,Echopraxia/echolalia,Stereotypy,Verbigeration,Rigidity,Negativism,Waxy flexibility,Withdrawal | 95.3 | 93.3 | 97.1 |
| 9 | Excitement,Posturing/catalepsy,Grimacing,Echopraxia/echolalia,Verbigeration,Rigidity,Negativism,Waxy flexibility,Withdrawal | 95.3 | 93.3 | 97.1 |
| 9 | Excitement,Posturing/catalepsy,Grimacing,Mannerisms,Verbigeration,Rigidity,Negativism,Waxy flexibility,Withdrawal | 95.3 | 93.3 | 97.1 |
| 9 | Excitement,Posturing/catalepsy,Grimacing,Stereotypy,Mannerisms,Verbigeration,Rigidity,Negativism,Withdrawal | 95.3 | 93.3 | 97.1 |
| 9 | Mutism,Posturing/catalepsy,Grimacing,Echopraxia/echolalia,Stereotypy,Mannerisms,Rigidity,Waxy flexibility,Withdrawal | 95.3 | 93.3 | 97.1 |
| 9 | Mutism,Posturing/catalepsy,Grimacing,Echopraxia/echolalia,Stereotypy,Verbigeration,Rigidity,Negativism,Waxy flexibility | 95.3 | 93.3 | 97.1 |
| 9 | Mutism,Posturing/catalepsy,Grimacing,Stereotypy,Mannerisms,Verbigeration,Rigidity,Waxy flexibility,Withdrawal | 95.3 | 93.3 | 97.1 |
| 9 | Posturing/catalepsy,Grimacing,Echopraxia/echolalia,Mannerisms,Verbigeration,Rigidity,Negativism,Waxy flexibility,Withdrawal | 95.3 | 93.3 | 97.1 |
| 9 | Posturing/catalepsy,Grimacing,Echopraxia/echolalia,Stereotypy,Mannerisms,Rigidity,Negativism,Waxy flexibility,Withdrawal | 95.3 | 93.3 | 97.1 |
| 9 | Posturing/catalepsy,Grimacing,Echopraxia/echolalia,Stereotypy,Mannerisms,Verbigeration,Rigidity,Negativism,Withdrawal | 95.3 | 93.3 | 97.1 |
| 9 | Excitement,Mutism,Grimacing,Echopraxia/echolalia,Stereotypy,Mannerisms,Verbigeration,Rigidity,Negativism | 95.1 | 93.0 | 97.1 |
| 9 | Excitement,Mutism,Posturing/catalepsy,Echopraxia/echolalia,Mannerisms,Verbigeration,Negativism,Waxy flexibility,Withdrawal | 95.1 | 93.0 | 97.1 |
| 9 | Excitement,Mutism,Posturing/catalepsy,Echopraxia/echolalia,Stereotypy,Mannerisms,Verbigeration,Rigidity,Waxy flexibility | 95.1 | 93.0 | 96.9 |
| 9 | Excitement,Mutism,Posturing/catalepsy,Grimacing,Echopraxia/echolalia,Mannerisms,Verbigeration,Negativism,Withdrawal | 95.1 | 93.0 | 96.9 |
| 9 | Excitement,Mutism,Posturing/catalepsy,Grimacing,Echopraxia/echolalia,Stereotypy,Verbigeration,Negativism,Withdrawal | 95.1 | 93.0 | 97.1 |
| 9 | Excitement,Mutism,Posturing/catalepsy,Grimacing,Echopraxia/echolalia,Stereotypy,Verbigeration,Rigidity,Waxy flexibility | 95.1 | 93.0 | 97.1 |
| 9 | Excitement,Mutism,Posturing/catalepsy,Grimacing,Echopraxia/echolalia,Verbigeration,Negativism,Waxy flexibility,Withdrawal | 95.1 | 92.8 | 96.9 |
| 9 | Excitement,Mutism,Posturing/catalepsy,Grimacing,Mannerisms,Verbigeration,Negativism,Waxy flexibility,Withdrawal | 95.1 | 93.0 | 96.9 |
| 9 | Excitement,Mutism,Posturing/catalepsy,Grimacing,Stereotypy,Mannerisms,Rigidity,Negativism,Waxy flexibility | 95.1 | 92.8 | 97.1 |
| 9 | Excitement,Mutism,Posturing/catalepsy,Grimacing,Stereotypy,Mannerisms,Verbigeration,Negativism,Withdrawal | 95.1 | 93.0 | 96.9 |
| 9 | Excitement,Mutism,Posturing/catalepsy,Grimacing,Stereotypy,Verbigeration,Negativism,Waxy flexibility,Withdrawal | 95.1 | 93.0 | 96.9 |
| 9 | Excitement,Posturing/catalepsy,Echopraxia/echolalia,Stereotypy,Mannerisms,Rigidity,Negativism,Waxy flexibility,Withdrawal | 95.1 | 93.0 | 97.1 |
| 9 | Excitement,Posturing/catalepsy,Grimacing,Echopraxia/echolalia,Mannerisms,Rigidity,Negativism,Waxy flexibility,Withdrawal | 95.1 | 93.0 | 97.1 |
| 9 | Excitement,Posturing/catalepsy,Grimacing,Echopraxia/echolalia,Stereotypy,Mannerisms,Rigidity,Negativism,Withdrawal | 95.1 | 92.8 | 96.9 |
| 9 | Excitement,Posturing/catalepsy,Grimacing,Echopraxia/echolalia,Stereotypy,Verbigeration,Rigidity,Negativism,Withdrawal | 95.1 | 93.0 | 97.1 |
| 9 | Excitement,Posturing/catalepsy,Grimacing,Stereotypy,Verbigeration,Rigidity,Negativism,Waxy flexibility,Withdrawal | 95.1 | 93.0 | 96.9 |
| 9 | Excitement,Mutism,Posturing/catalepsy,Echopraxia/echolalia,Stereotypy,Mannerisms,Negativism,Waxy flexibility,Withdrawal | 94.8 | 92.6 | 96.9 |
| 9 | Excitement,Mutism,Posturing/catalepsy,Grimacing,Echopraxia/echolalia,Mannerisms,Negativism,Waxy flexibility,Withdrawal | 94.8 | 92.6 | 96.9 |
| 9 | Excitement,Mutism,Posturing/catalepsy,Grimacing,Echopraxia/echolalia,Stereotypy,Mannerisms,Negativism,Withdrawal | 94.8 | 92.6 | 96.9 |
| 9 | Excitement,Mutism,Posturing/catalepsy,Grimacing,Echopraxia/echolalia,Stereotypy,Negativism,Waxy flexibility,Withdrawal | 94.8 | 92.8 | 96.9 |
| 9 | Excitement,Mutism,Posturing/catalepsy,Grimacing,Stereotypy,Mannerisms,Verbigeration,Rigidity,Waxy flexibility | 94.8 | 92.6 | 96.9 |
| 9 | Excitement,Posturing/catalepsy,Grimacing,Echopraxia/echolalia,Mannerisms,Verbigeration,Rigidity,Waxy flexibility,Withdrawal | 94.8 | 92.6 | 96.9 |
| 9 | Excitement,Posturing/catalepsy,Grimacing,Echopraxia/echolalia,Stereotypy,Rigidity,Negativism,Waxy flexibility,Withdrawal | 94.8 | 92.8 | 96.9 |
| 9 | Excitement,Posturing/catalepsy,Grimacing,Stereotypy,Mannerisms,Rigidity,Negativism,Waxy flexibility,Withdrawal | 94.8 | 92.6 | 96.9 |
| 9 | Mutism,Grimacing,Echopraxia/echolalia,Stereotypy,Mannerisms,Verbigeration,Rigidity,Waxy flexibility,Withdrawal | 94.8 | 92.6 | 96.9 |
| 9 | Mutism,Posturing/catalepsy,Echopraxia/echolalia,Stereotypy,Mannerisms,Verbigeration,Negativism,Waxy flexibility,Withdrawal | 94.8 | 92.6 | 96.9 |
| 9 | Mutism,Posturing/catalepsy,Echopraxia/echolalia,Stereotypy,Mannerisms,Verbigeration,Rigidity,Waxy flexibility,Withdrawal | 94.8 | 92.6 | 96.9 |
| 9 | Mutism,Posturing/catalepsy,Grimacing,Echopraxia/echolalia,Mannerisms,Verbigeration,Negativism,Waxy flexibility,Withdrawal | 94.8 | 92.6 | 96.9 |
| 9 | Mutism,Posturing/catalepsy,Grimacing,Echopraxia/echolalia,Stereotypy,Mannerisms,Negativism,Waxy flexibility,Withdrawal | 94.8 | 92.6 | 96.9 |
| 9 | Mutism,Posturing/catalepsy,Grimacing,Echopraxia/echolalia,Stereotypy,Mannerisms,Verbigeration,Negativism,Withdrawal | 94.8 | 92.6 | 96.6 |
| 9 | Mutism,Posturing/catalepsy,Grimacing,Echopraxia/echolalia,Stereotypy,Verbigeration,Rigidity,Waxy flexibility,Withdrawal | 94.8 | 92.8 | 96.9 |
| 9 | Mutism,Posturing/catalepsy,Grimacing,Stereotypy,Mannerisms,Verbigeration,Negativism,Waxy flexibility,Withdrawal | 94.8 | 92.6 | 96.9 |
| 9 | Excitement,Mutism,Posturing/catalepsy,Grimacing,Echopraxia/echolalia,Stereotypy,Mannerisms,Rigidity,Waxy flexibility | 94.6 | 92.4 | 96.6 |
| 9 | Excitement,Mutism,Posturing/catalepsy,Grimacing,Stereotypy,Mannerisms,Negativism,Waxy flexibility,Withdrawal | 94.6 | 92.4 | 96.6 |
| 9 | Excitement,Posturing/catalepsy,Echopraxia/echolalia,Stereotypy,Mannerisms,Verbigeration,Rigidity,Waxy flexibility,Withdrawal | 94.6 | 92.4 | 96.6 |
| 9 | Mutism,Posturing/catalepsy,Grimacing,Echopraxia/echolalia,Stereotypy,Mannerisms,Verbigeration,Rigidity,Withdrawal | 94.6 | 92.4 | 96.6 |
| 9 | Posturing/catalepsy,Grimacing,Echopraxia/echolalia,Stereotypy,Verbigeration,Rigidity,Negativism,Waxy flexibility,Withdrawal | 94.6 | 92.4 | 96.6 |
| 9 | Excitement,Mutism,Posturing/catalepsy,Grimacing,Echopraxia/echolalia,Stereotypy,Mannerisms,Verbigeration,Rigidity | 94.4 | 92.2 | 96.4 |
| 9 | Excitement,Posturing/catalepsy,Grimacing,Echopraxia/echolalia,Stereotypy,Mannerisms,Rigidity,Waxy flexibility,Withdrawal | 94.2 | 91.9 | 96.2 |
| 9 | Mutism,Posturing/catalepsy,Grimacing,Echopraxia/echolalia,Stereotypy,Verbigeration,Negativism,Waxy flexibility,Withdrawal | 94.2 | 91.9 | 96.2 |
| 9 | Excitement,Posturing/catalepsy,Grimacing,Echopraxia/echolalia,Stereotypy,Verbigeration,Rigidity,Waxy flexibility,Withdrawal | 94.1 | 91.9 | 96.2 |
| 9 | Excitement,Posturing/catalepsy,Grimacing,Stereotypy,Mannerisms,Verbigeration,Rigidity,Waxy flexibility,Withdrawal | 94.1 | 91.9 | 96.2 |
| 9 | Excitement,Mutism,Posturing/catalepsy,Grimacing,Echopraxia/echolalia,Mannerisms,Verbigeration,Negativism,Waxy flexibility | 94.0 | 91.7 | 96.0 |
| 9 | Excitement,Mutism,Posturing/catalepsy,Grimacing,Echopraxia/echolalia,Mannerisms,Verbigeration,Waxy flexibility,Withdrawal | 94.0 | 91.7 | 96.2 |
| 9 | Excitement,Mutism,Echopraxia/echolalia,Stereotypy,Mannerisms,Verbigeration,Negativism,Waxy flexibility,Withdrawal | 93.9 | 91.7 | 96.0 |
| 9 | Excitement,Mutism,Grimacing,Echopraxia/echolalia,Stereotypy,Mannerisms,Verbigeration,Rigidity,Waxy flexibility | 93.9 | 91.7 | 96.0 |
| 9 | Excitement,Mutism,Grimacing,Echopraxia/echolalia,Stereotypy,Mannerisms,Verbigeration,Rigidity,Withdrawal | 93.9 | 91.7 | 96.0 |
| 9 | Excitement,Mutism,Posturing/catalepsy,Echopraxia/echolalia,Stereotypy,Mannerisms,Verbigeration,Waxy flexibility,Withdrawal | 93.9 | 91.7 | 96.0 |
| 9 | Excitement,Mutism,Posturing/catalepsy,Grimacing,Echopraxia/echolalia,Stereotypy,Verbigeration,Negativism,Waxy flexibility | 93.9 | 91.7 | 96.0 |
| 9 | Excitement,Mutism,Posturing/catalepsy,Grimacing,Echopraxia/echolalia,Stereotypy,Verbigeration,Waxy flexibility,Withdrawal | 93.9 | 91.7 | 96.0 |
| 9 | Grimacing,Echopraxia/echolalia,Stereotypy,Mannerisms,Verbigeration,Rigidity,Negativism,Waxy flexibility,Withdrawal | 93.9 | 91.7 | 96.2 |
| 9 | Mutism,Posturing/catalepsy,Grimacing,Echopraxia/echolalia,Stereotypy,Mannerisms,Verbigeration,Rigidity,Waxy flexibility | 93.9 | 91.7 | 96.0 |
| 9 | Excitement,Echopraxia/echolalia,Stereotypy,Mannerisms,Verbigeration,Rigidity,Negativism,Waxy flexibility,Withdrawal | 93.7 | 91.3 | 96.0 |
| 9 | Excitement,Mutism,Grimacing,Echopraxia/echolalia,Stereotypy,Verbigeration,Negativism,Waxy flexibility,Withdrawal | 93.7 | 91.5 | 96.0 |
| 9 | Excitement,Mutism,Posturing/catalepsy,Echopraxia/echolalia,Stereotypy,Mannerisms,Verbigeration,Negativism,Waxy flexibility | 93.7 | 91.5 | 96.0 |
| 9 | Excitement,Mutism,Posturing/catalepsy,Grimacing,Echopraxia/echolalia,Stereotypy,Mannerisms,Verbigeration,Negativism | 93.7 | 91.5 | 96.0 |
| 9 | Excitement,Mutism,Posturing/catalepsy,Grimacing,Echopraxia/echolalia,Stereotypy,Mannerisms,Waxy flexibility,Withdrawal | 93.7 | 91.3 | 96.0 |
| 9 | Excitement,Mutism,Posturing/catalepsy,Grimacing,Stereotypy,Mannerisms,Verbigeration,Negativism,Waxy flexibility | 93.7 | 91.3 | 96.0 |
| 9 | Excitement,Mutism,Posturing/catalepsy,Grimacing,Stereotypy,Mannerisms,Verbigeration,Waxy flexibility,Withdrawal | 93.7 | 91.5 | 96.0 |
| 9 | Excitement,Posturing/catalepsy,Grimacing,Echopraxia/echolalia,Stereotypy,Mannerisms,Verbigeration,Rigidity,Withdrawal | 93.7 | 91.5 | 96.0 |
| 9 | Mutism,Grimacing,Echopraxia/echolalia,Stereotypy,Mannerisms,Verbigeration,Negativism,Waxy flexibility,Withdrawal | 93.7 | 91.5 | 95.7 |
| 9 | Mutism,Posturing/catalepsy,Grimacing,Echopraxia/echolalia,Stereotypy,Mannerisms,Verbigeration,Negativism,Waxy flexibility | 93.7 | 91.5 | 95.7 |
| 9 | Posturing/catalepsy,Grimacing,Echopraxia/echolalia,Stereotypy,Mannerisms,Verbigeration,Rigidity,Waxy flexibility,Withdrawal | 93.7 | 91.5 | 95.7 |
| 9 | Excitement,Grimacing,Echopraxia/echolalia,Stereotypy,Mannerisms,Rigidity,Negativism,Waxy flexibility,Withdrawal | 93.5 | 91.0 | 95.7 |
| 9 | Excitement,Mutism,Grimacing,Echopraxia/echolalia,Stereotypy,Mannerisms,Negativism,Waxy flexibility,Withdrawal | 93.5 | 91.3 | 95.7 |
| 9 | Excitement,Mutism,Posturing/catalepsy,Grimacing,Echopraxia/echolalia,Stereotypy,Mannerisms,Negativism,Waxy flexibility | 93.5 | 91.0 | 95.7 |
| 9 | Excitement,Posturing/catalepsy,Echopraxia/echolalia,Stereotypy,Mannerisms,Verbigeration,Negativism,Waxy flexibility,Withdrawal | 93.5 | 91.0 | 95.7 |
| 9 | Posturing/catalepsy,Grimacing,Echopraxia/echolalia,Stereotypy,Mannerisms,Verbigeration,Negativism,Waxy flexibility,Withdrawal | 93.5 | 91.0 | 95.7 |
| 9 | Excitement,Grimacing,Echopraxia/echolalia,Mannerisms,Verbigeration,Rigidity,Negativism,Waxy flexibility,Withdrawal | 93.3 | 90.8 | 95.5 |
| 9 | Excitement,Grimacing,Echopraxia/echolalia,Stereotypy,Mannerisms,Verbigeration,Rigidity,Negativism,Withdrawal | 93.3 | 90.8 | 95.5 |
| 9 | Excitement,Grimacing,Echopraxia/echolalia,Stereotypy,Verbigeration,Rigidity,Negativism,Waxy flexibility,Withdrawal | 93.3 | 90.8 | 95.5 |
| 9 | Excitement,Grimacing,Stereotypy,Mannerisms,Verbigeration,Rigidity,Negativism,Waxy flexibility,Withdrawal | 93.3 | 90.8 | 95.5 |
| 9 | Excitement,Mutism,Grimacing,Echopraxia/echolalia,Stereotypy,Mannerisms,Verbigeration,Negativism,Withdrawal | 93.3 | 90.8 | 95.5 |
| 9 | Excitement,Mutism,Grimacing,Stereotypy,Mannerisms,Verbigeration,Negativism,Waxy flexibility,Withdrawal | 93.3 | 90.8 | 95.5 |
| 9 | Excitement,Posturing/catalepsy,Grimacing,Echopraxia/echolalia,Mannerisms,Verbigeration,Negativism,Waxy flexibility,Withdrawal | 93.3 | 90.8 | 95.5 |
| 9 | Excitement,Posturing/catalepsy,Grimacing,Echopraxia/echolalia,Stereotypy,Mannerisms,Verbigeration,Negativism,Withdrawal | 93.3 | 90.8 | 95.5 |
| 9 | Excitement,Posturing/catalepsy,Grimacing,Stereotypy,Mannerisms,Verbigeration,Negativism,Waxy flexibility,Withdrawal | 93.3 | 90.8 | 95.5 |
| 9 | Mutism,Posturing/catalepsy,Grimacing,Echopraxia/echolalia,Stereotypy,Mannerisms,Verbigeration,Waxy flexibility,Withdrawal | 93.3 | 90.8 | 95.5 |
| 9 | Excitement,Mutism,Posturing/catalepsy,Grimacing,Echopraxia/echolalia,Stereotypy,Mannerisms,Verbigeration,Withdrawal | 93.1 | 90.6 | 95.3 |
| 9 | Excitement,Posturing/catalepsy,Grimacing,Echopraxia/echolalia,Stereotypy,Mannerisms,Negativism,Waxy flexibility,Withdrawal | 93.1 | 90.6 | 95.3 |
| 9 | Excitement,Posturing/catalepsy,Grimacing,Echopraxia/echolalia,Stereotypy,Verbigeration,Negativism,Waxy flexibility,Withdrawal | 93.0 | 90.6 | 95.3 |
| 9 | Excitement,Mutism,Grimacing,Echopraxia/echolalia,Mannerisms,Verbigeration,Negativism,Waxy flexibility,Withdrawal | 92.8 | 90.4 | 95.1 |
| 9 | Excitement,Grimacing,Echopraxia/echolalia,Stereotypy,Mannerisms,Verbigeration,Rigidity,Waxy flexibility,Withdrawal | 92.4 | 89.9 | 94.6 |
| 9 | Excitement,Mutism,Grimacing,Echopraxia/echolalia,Stereotypy,Mannerisms,Verbigeration,Negativism,Waxy flexibility | 92.2 | 89.5 | 94.6 |
| 9 | Excitement,Posturing/catalepsy,Grimacing,Echopraxia/echolalia,Stereotypy,Mannerisms,Verbigeration,Waxy flexibility,Withdrawal | 92.2 | 89.7 | 94.4 |
| 9 | Excitement,Mutism,Grimacing,Echopraxia/echolalia,Stereotypy,Mannerisms,Verbigeration,Waxy flexibility,Withdrawal | 91.9 | 89.5 | 94.4 |
| 9 | Excitement,Mutism,Posturing/catalepsy,Grimacing,Echopraxia/echolalia,Stereotypy,Mannerisms,Verbigeration,Waxy flexibility | 91.5 | 88.8 | 93.9 |
| 9 | Excitement,Posturing/catalepsy,Grimacing,Echopraxia/echolalia,Mannerisms,Verbigeration,Rigidity,Negativism,Waxy flexibility | 91.2 | 88.6 | 93.7 |
| 9 | Excitement,Posturing/catalepsy,Grimacing,Echopraxia/echolalia,Stereotypy,Verbigeration,Rigidity,Negativism,Waxy flexibility | 91.0 | 88.3 | 93.7 |
| 9 | Excitement,Posturing/catalepsy,Grimacing,Stereotypy,Mannerisms,Verbigeration,Rigidity,Negativism,Waxy flexibility | 91.0 | 88.3 | 93.5 |
| 9 | Posturing/catalepsy,Grimacing,Echopraxia/echolalia,Stereotypy,Mannerisms,Verbigeration,Rigidity,Negativism,Waxy flexibility | 91.0 | 88.3 | 93.5 |
| 9 | Excitement,Posturing/catalepsy,Echopraxia/echolalia,Stereotypy,Mannerisms,Verbigeration,Rigidity,Negativism,Waxy flexibility | 90.8 | 88.1 | 93.5 |
| 9 | Excitement,Posturing/catalepsy,Grimacing,Echopraxia/echolalia,Stereotypy,Mannerisms,Rigidity,Negativism,Waxy flexibility | 90.8 | 88.1 | 93.5 |
| 9 | Excitement,Posturing/catalepsy,Grimacing,Echopraxia/echolalia,Stereotypy,Mannerisms,Verbigeration,Rigidity,Negativism | 90.8 | 87.9 | 93.5 |
| 9 | Excitement,Grimacing,Echopraxia/echolalia,Stereotypy,Mannerisms,Verbigeration,Negativism,Waxy flexibility,Withdrawal | 90.1 | 87.2 | 92.8 |
| 9 | Excitement,Posturing/catalepsy,Grimacing,Echopraxia/echolalia,Stereotypy,Mannerisms,Verbigeration,Negativism,Waxy flexibility | 87.9 | 84.8 | 90.8 |
| 9 | Excitement,Posturing/catalepsy,Grimacing,Echopraxia/echolalia,Stereotypy,Mannerisms,Verbigeration,Rigidity,Waxy flexibility | 87.9 | 84.8 | 90.8 |
| 9 | Excitement,Grimacing,Echopraxia/echolalia,Stereotypy,Mannerisms,Verbigeration,Rigidity,Negativism,Waxy flexibility | 87.7 | 84.5 | 90.6 |

Table S10: sensitivity (relative to the full BFCSI) of all ten question combinations, and the 95% confidence interval derived from bootstrapping, sorted by decreasing sensitivity.

|  |  |  | 95% CI | |
| --- | --- | --- | --- | --- |
| # of Qs | Combination | Sensitivity | Lower | Upper |
| 10 | Excitement,Immobility/stupor,Mutism,Posturing/catalepsy,Echopraxia/echolalia,Stereotypy,Verbigeration,Rigidity,Negativism,Withdrawal | 100.0 | 100.0 | 100.0 |
| 10 | Excitement,Immobility/stupor,Mutism,Posturing/catalepsy,Grimacing,Echopraxia/echolalia,Verbigeration,Rigidity,Negativism,Withdrawal | 100.0 | 100.0 | 100.0 |
| 10 | Excitement,Immobility/stupor,Mutism,Staring,Echopraxia/echolalia,Stereotypy,Mannerisms,Rigidity,Negativism,Withdrawal | 100.0 | 100.0 | 100.0 |
| 10 | Excitement,Immobility/stupor,Mutism,Staring,Echopraxia/echolalia,Stereotypy,Mannerisms,Rigidity,Waxy flexibility,Withdrawal | 100.0 | 100.0 | 100.0 |
| 10 | Excitement,Immobility/stupor,Mutism,Staring,Echopraxia/echolalia,Stereotypy,Rigidity,Negativism,Waxy flexibility,Withdrawal | 100.0 | 100.0 | 100.0 |
| 10 | Excitement,Immobility/stupor,Mutism,Staring,Echopraxia/echolalia,Stereotypy,Verbigeration,Rigidity,Negativism,Waxy flexibility | 100.0 | 100.0 | 100.0 |
| 10 | Excitement,Immobility/stupor,Mutism,Staring,Echopraxia/echolalia,Stereotypy,Verbigeration,Rigidity,Negativism,Withdrawal | 100.0 | 100.0 | 100.0 |
| 10 | Excitement,Immobility/stupor,Mutism,Staring,Echopraxia/echolalia,Stereotypy,Verbigeration,Rigidity,Waxy flexibility,Withdrawal | 100.0 | 100.0 | 100.0 |
| 10 | Excitement,Immobility/stupor,Mutism,Staring,Grimacing,Echopraxia/echolalia,Stereotypy,Rigidity,Negativism,Withdrawal | 100.0 | 100.0 | 100.0 |
| 10 | Excitement,Immobility/stupor,Mutism,Staring,Grimacing,Echopraxia/echolalia,Stereotypy,Rigidity,Waxy flexibility,Withdrawal | 100.0 | 100.0 | 100.0 |
| 10 | Excitement,Immobility/stupor,Mutism,Staring,Grimacing,Stereotypy,Verbigeration,Rigidity,Negativism,Waxy flexibility | 100.0 | 100.0 | 100.0 |
| 10 | Excitement,Immobility/stupor,Mutism,Staring,Grimacing,Stereotypy,Verbigeration,Rigidity,Negativism,Withdrawal | 100.0 | 100.0 | 100.0 |
| 10 | Excitement,Immobility/stupor,Mutism,Staring,Grimacing,Stereotypy,Verbigeration,Rigidity,Waxy flexibility,Withdrawal | 100.0 | 100.0 | 100.0 |
| 10 | Excitement,Immobility/stupor,Mutism,Staring,Posturing/catalepsy,Echopraxia/echolalia,Mannerisms,Negativism,Waxy flexibility,Withdrawal | 100.0 | 100.0 | 100.0 |
| 10 | Excitement,Immobility/stupor,Mutism,Staring,Posturing/catalepsy,Echopraxia/echolalia,Mannerisms,Rigidity,Negativism,Withdrawal | 100.0 | 100.0 | 100.0 |
| 10 | Excitement,Immobility/stupor,Mutism,Staring,Posturing/catalepsy,Echopraxia/echolalia,Mannerisms,Rigidity,Waxy flexibility,Withdrawal | 100.0 | 100.0 | 100.0 |
| 10 | Excitement,Immobility/stupor,Mutism,Staring,Posturing/catalepsy,Echopraxia/echolalia,Mannerisms,Verbigeration,Negativism,Waxy flexibility | 100.0 | 100.0 | 100.0 |
| 10 | Excitement,Immobility/stupor,Mutism,Staring,Posturing/catalepsy,Echopraxia/echolalia,Mannerisms,Verbigeration,Negativism,Withdrawal | 100.0 | 100.0 | 100.0 |
| 10 | Excitement,Immobility/stupor,Mutism,Staring,Posturing/catalepsy,Echopraxia/echolalia,Mannerisms,Verbigeration,Rigidity,Negativism | 100.0 | 100.0 | 100.0 |
| 10 | Excitement,Immobility/stupor,Mutism,Staring,Posturing/catalepsy,Echopraxia/echolalia,Mannerisms,Verbigeration,Rigidity,Withdrawal | 100.0 | 100.0 | 100.0 |
| 10 | Excitement,Immobility/stupor,Mutism,Staring,Posturing/catalepsy,Echopraxia/echolalia,Rigidity,Negativism,Waxy flexibility,Withdrawal | 100.0 | 100.0 | 100.0 |
| 10 | Excitement,Immobility/stupor,Mutism,Staring,Posturing/catalepsy,Echopraxia/echolalia,Stereotypy,Mannerisms,Negativism,Withdrawal | 100.0 | 100.0 | 100.0 |
| 10 | Excitement,Immobility/stupor,Mutism,Staring,Posturing/catalepsy,Echopraxia/echolalia,Stereotypy,Mannerisms,Rigidity,Withdrawal | 100.0 | 100.0 | 100.0 |
| 10 | Excitement,Immobility/stupor,Mutism,Staring,Posturing/catalepsy,Echopraxia/echolalia,Stereotypy,Negativism,Waxy flexibility,Withdrawal | 100.0 | 100.0 | 100.0 |
| 10 | Excitement,Immobility/stupor,Mutism,Staring,Posturing/catalepsy,Echopraxia/echolalia,Stereotypy,Rigidity,Negativism,Withdrawal | 100.0 | 100.0 | 100.0 |
| 10 | Excitement,Immobility/stupor,Mutism,Staring,Posturing/catalepsy,Echopraxia/echolalia,Stereotypy,Rigidity,Waxy flexibility,Withdrawal | 100.0 | 100.0 | 100.0 |
| 10 | Excitement,Immobility/stupor,Mutism,Staring,Posturing/catalepsy,Echopraxia/echolalia,Stereotypy,Verbigeration,Negativism,Waxy flexibility | 100.0 | 100.0 | 100.0 |
| 10 | Excitement,Immobility/stupor,Mutism,Staring,Posturing/catalepsy,Echopraxia/echolalia,Stereotypy,Verbigeration,Negativism,Withdrawal | 100.0 | 100.0 | 100.0 |
| 10 | Excitement,Immobility/stupor,Mutism,Staring,Posturing/catalepsy,Echopraxia/echolalia,Stereotypy,Verbigeration,Rigidity,Negativism | 100.0 | 100.0 | 100.0 |
| 10 | Excitement,Immobility/stupor,Mutism,Staring,Posturing/catalepsy,Echopraxia/echolalia,Stereotypy,Verbigeration,Rigidity,Withdrawal | 100.0 | 100.0 | 100.0 |
| 10 | Excitement,Immobility/stupor,Mutism,Staring,Posturing/catalepsy,Echopraxia/echolalia,Verbigeration,Negativism,Waxy flexibility,Withdrawal | 100.0 | 100.0 | 100.0 |
| 10 | Excitement,Immobility/stupor,Mutism,Staring,Posturing/catalepsy,Echopraxia/echolalia,Verbigeration,Rigidity,Negativism,Waxy flexibility | 100.0 | 100.0 | 100.0 |
| 10 | Excitement,Immobility/stupor,Mutism,Staring,Posturing/catalepsy,Echopraxia/echolalia,Verbigeration,Rigidity,Negativism,Withdrawal | 100.0 | 100.0 | 100.0 |
| 10 | Excitement,Immobility/stupor,Mutism,Staring,Posturing/catalepsy,Echopraxia/echolalia,Verbigeration,Rigidity,Waxy flexibility,Withdrawal | 100.0 | 100.0 | 100.0 |
| 10 | Excitement,Immobility/stupor,Mutism,Staring,Posturing/catalepsy,Grimacing,Echopraxia/echolalia,Mannerisms,Negativism,Withdrawal | 100.0 | 100.0 | 100.0 |
| 10 | Excitement,Immobility/stupor,Mutism,Staring,Posturing/catalepsy,Grimacing,Echopraxia/echolalia,Mannerisms,Rigidity,Withdrawal | 100.0 | 100.0 | 100.0 |
| 10 | Excitement,Immobility/stupor,Mutism,Staring,Posturing/catalepsy,Grimacing,Echopraxia/echolalia,Negativism,Waxy flexibility,Withdrawal | 100.0 | 100.0 | 100.0 |
| 10 | Excitement,Immobility/stupor,Mutism,Staring,Posturing/catalepsy,Grimacing,Echopraxia/echolalia,Rigidity,Negativism,Withdrawal | 100.0 | 100.0 | 100.0 |
| 10 | Excitement,Immobility/stupor,Mutism,Staring,Posturing/catalepsy,Grimacing,Echopraxia/echolalia,Rigidity,Waxy flexibility,Withdrawal | 100.0 | 100.0 | 100.0 |
| 10 | Excitement,Immobility/stupor,Mutism,Staring,Posturing/catalepsy,Grimacing,Echopraxia/echolalia,Stereotypy,Negativism,Withdrawal | 100.0 | 100.0 | 100.0 |
| 10 | Excitement,Immobility/stupor,Mutism,Staring,Posturing/catalepsy,Grimacing,Echopraxia/echolalia,Stereotypy,Rigidity,Withdrawal | 100.0 | 100.0 | 100.0 |
| 10 | Excitement,Immobility/stupor,Mutism,Staring,Posturing/catalepsy,Grimacing,Echopraxia/echolalia,Verbigeration,Negativism,Waxy flexibility | 100.0 | 100.0 | 100.0 |
| 10 | Excitement,Immobility/stupor,Mutism,Staring,Posturing/catalepsy,Grimacing,Echopraxia/echolalia,Verbigeration,Negativism,Withdrawal | 100.0 | 100.0 | 100.0 |
| 10 | Excitement,Immobility/stupor,Mutism,Staring,Posturing/catalepsy,Grimacing,Echopraxia/echolalia,Verbigeration,Rigidity,Negativism | 100.0 | 100.0 | 100.0 |
| 10 | Excitement,Immobility/stupor,Mutism,Staring,Posturing/catalepsy,Grimacing,Echopraxia/echolalia,Verbigeration,Rigidity,Withdrawal | 100.0 | 100.0 | 100.0 |
| 10 | Excitement,Immobility/stupor,Mutism,Staring,Posturing/catalepsy,Grimacing,Mannerisms,Verbigeration,Negativism,Waxy flexibility | 100.0 | 100.0 | 100.0 |
| 10 | Excitement,Immobility/stupor,Mutism,Staring,Posturing/catalepsy,Grimacing,Mannerisms,Verbigeration,Negativism,Withdrawal | 100.0 | 100.0 | 100.0 |
| 10 | Excitement,Immobility/stupor,Mutism,Staring,Posturing/catalepsy,Grimacing,Mannerisms,Verbigeration,Rigidity,Negativism | 100.0 | 100.0 | 100.0 |
| 10 | Excitement,Immobility/stupor,Mutism,Staring,Posturing/catalepsy,Grimacing,Mannerisms,Verbigeration,Rigidity,Withdrawal | 100.0 | 100.0 | 100.0 |
| 10 | Excitement,Immobility/stupor,Mutism,Staring,Posturing/catalepsy,Grimacing,Stereotypy,Verbigeration,Negativism,Waxy flexibility | 100.0 | 100.0 | 100.0 |
| 10 | Excitement,Immobility/stupor,Mutism,Staring,Posturing/catalepsy,Grimacing,Stereotypy,Verbigeration,Negativism,Withdrawal | 100.0 | 100.0 | 100.0 |
| 10 | Excitement,Immobility/stupor,Mutism,Staring,Posturing/catalepsy,Grimacing,Stereotypy,Verbigeration,Rigidity,Negativism | 100.0 | 100.0 | 100.0 |
| 10 | Excitement,Immobility/stupor,Mutism,Staring,Posturing/catalepsy,Grimacing,Stereotypy,Verbigeration,Rigidity,Withdrawal | 100.0 | 100.0 | 100.0 |
| 10 | Excitement,Immobility/stupor,Mutism,Staring,Posturing/catalepsy,Mannerisms,Verbigeration,Negativism,Waxy flexibility,Withdrawal | 100.0 | 100.0 | 100.0 |
| 10 | Excitement,Immobility/stupor,Mutism,Staring,Posturing/catalepsy,Mannerisms,Verbigeration,Rigidity,Negativism,Waxy flexibility | 100.0 | 100.0 | 100.0 |
| 10 | Excitement,Immobility/stupor,Mutism,Staring,Posturing/catalepsy,Mannerisms,Verbigeration,Rigidity,Negativism,Withdrawal | 100.0 | 100.0 | 100.0 |
| 10 | Excitement,Immobility/stupor,Mutism,Staring,Posturing/catalepsy,Mannerisms,Verbigeration,Rigidity,Waxy flexibility,Withdrawal | 100.0 | 100.0 | 100.0 |
| 10 | Excitement,Immobility/stupor,Mutism,Staring,Posturing/catalepsy,Stereotypy,Mannerisms,Verbigeration,Negativism,Waxy flexibility | 100.0 | 100.0 | 100.0 |
| 10 | Excitement,Immobility/stupor,Mutism,Staring,Posturing/catalepsy,Stereotypy,Mannerisms,Verbigeration,Negativism,Withdrawal | 100.0 | 100.0 | 100.0 |
| 10 | Excitement,Immobility/stupor,Mutism,Staring,Posturing/catalepsy,Stereotypy,Mannerisms,Verbigeration,Rigidity,Negativism | 100.0 | 100.0 | 100.0 |
| 10 | Excitement,Immobility/stupor,Mutism,Staring,Posturing/catalepsy,Stereotypy,Mannerisms,Verbigeration,Rigidity,Withdrawal | 100.0 | 100.0 | 100.0 |
| 10 | Excitement,Immobility/stupor,Mutism,Staring,Posturing/catalepsy,Stereotypy,Verbigeration,Negativism,Waxy flexibility,Withdrawal | 100.0 | 100.0 | 100.0 |
| 10 | Excitement,Immobility/stupor,Mutism,Staring,Posturing/catalepsy,Stereotypy,Verbigeration,Rigidity,Negativism,Waxy flexibility | 100.0 | 100.0 | 100.0 |
| 10 | Excitement,Immobility/stupor,Mutism,Staring,Posturing/catalepsy,Stereotypy,Verbigeration,Rigidity,Negativism,Withdrawal | 100.0 | 100.0 | 100.0 |
| 10 | Excitement,Immobility/stupor,Mutism,Staring,Posturing/catalepsy,Stereotypy,Verbigeration,Rigidity,Waxy flexibility,Withdrawal | 100.0 | 100.0 | 100.0 |
| 10 | Excitement,Immobility/stupor,Mutism,Staring,Stereotypy,Mannerisms,Verbigeration,Rigidity,Negativism,Waxy flexibility | 100.0 | 100.0 | 100.0 |
| 10 | Excitement,Immobility/stupor,Mutism,Staring,Stereotypy,Mannerisms,Verbigeration,Rigidity,Negativism,Withdrawal | 100.0 | 100.0 | 100.0 |
| 10 | Excitement,Immobility/stupor,Mutism,Staring,Stereotypy,Mannerisms,Verbigeration,Rigidity,Waxy flexibility,Withdrawal | 100.0 | 100.0 | 100.0 |
| 10 | Excitement,Immobility/stupor,Mutism,Staring,Stereotypy,Verbigeration,Rigidity,Negativism,Waxy flexibility,Withdrawal | 100.0 | 100.0 | 100.0 |
| 10 | Excitement,Immobility/stupor,Staring,Echopraxia/echolalia,Stereotypy,Mannerisms,Rigidity,Negativism,Waxy flexibility,Withdrawal | 100.0 | 100.0 | 100.0 |
| 10 | Excitement,Immobility/stupor,Staring,Echopraxia/echolalia,Stereotypy,Mannerisms,Verbigeration,Rigidity,Negativism,Withdrawal | 100.0 | 100.0 | 100.0 |
| 10 | Excitement,Immobility/stupor,Staring,Grimacing,Echopraxia/echolalia,Stereotypy,Mannerisms,Rigidity,Negativism,Withdrawal | 100.0 | 100.0 | 100.0 |
| 10 | Excitement,Immobility/stupor,Staring,Grimacing,Stereotypy,Mannerisms,Verbigeration,Rigidity,Negativism,Withdrawal | 100.0 | 100.0 | 100.0 |
| 10 | Excitement,Immobility/stupor,Staring,Posturing/catalepsy,Echopraxia/echolalia,Mannerisms,Rigidity,Negativism,Waxy flexibility,Withdrawal | 100.0 | 100.0 | 100.0 |
| 10 | Excitement,Immobility/stupor,Staring,Posturing/catalepsy,Echopraxia/echolalia,Mannerisms,Verbigeration,Rigidity,Negativism,Withdrawal | 100.0 | 100.0 | 100.0 |
| 10 | Excitement,Immobility/stupor,Staring,Posturing/catalepsy,Echopraxia/echolalia,Stereotypy,Mannerisms,Rigidity,Negativism,Withdrawal | 100.0 | 100.0 | 100.0 |
| 10 | Excitement,Immobility/stupor,Staring,Posturing/catalepsy,Grimacing,Echopraxia/echolalia,Mannerisms,Rigidity,Negativism,Withdrawal | 100.0 | 100.0 | 100.0 |
| 10 | Excitement,Immobility/stupor,Staring,Posturing/catalepsy,Grimacing,Mannerisms,Verbigeration,Rigidity,Negativism,Withdrawal | 100.0 | 100.0 | 100.0 |
| 10 | Excitement,Immobility/stupor,Staring,Posturing/catalepsy,Mannerisms,Verbigeration,Rigidity,Negativism,Waxy flexibility,Withdrawal | 100.0 | 100.0 | 100.0 |
| 10 | Excitement,Immobility/stupor,Staring,Posturing/catalepsy,Stereotypy,Mannerisms,Verbigeration,Rigidity,Negativism,Withdrawal | 100.0 | 100.0 | 100.0 |
| 10 | Excitement,Immobility/stupor,Staring,Stereotypy,Mannerisms,Verbigeration,Rigidity,Negativism,Waxy flexibility,Withdrawal | 100.0 | 100.0 | 100.0 |
| 10 | Excitement,Mutism,Staring,Echopraxia/echolalia,Stereotypy,Mannerisms,Rigidity,Negativism,Waxy flexibility,Withdrawal | 100.0 | 100.0 | 100.0 |
| 10 | Excitement,Mutism,Staring,Echopraxia/echolalia,Stereotypy,Mannerisms,Verbigeration,Rigidity,Negativism,Waxy flexibility | 100.0 | 100.0 | 100.0 |
| 10 | Excitement,Mutism,Staring,Echopraxia/echolalia,Stereotypy,Mannerisms,Verbigeration,Rigidity,Negativism,Withdrawal | 100.0 | 100.0 | 100.0 |
| 10 | Excitement,Mutism,Staring,Echopraxia/echolalia,Stereotypy,Mannerisms,Verbigeration,Rigidity,Waxy flexibility,Withdrawal | 100.0 | 100.0 | 100.0 |
| 10 | Excitement,Mutism,Staring,Echopraxia/echolalia,Stereotypy,Verbigeration,Rigidity,Negativism,Waxy flexibility,Withdrawal | 100.0 | 100.0 | 100.0 |
| 10 | Excitement,Mutism,Staring,Grimacing,Echopraxia/echolalia,Stereotypy,Mannerisms,Rigidity,Negativism,Withdrawal | 100.0 | 100.0 | 100.0 |
| 10 | Excitement,Mutism,Staring,Grimacing,Echopraxia/echolalia,Stereotypy,Mannerisms,Rigidity,Waxy flexibility,Withdrawal | 100.0 | 100.0 | 100.0 |
| 10 | Excitement,Mutism,Staring,Grimacing,Echopraxia/echolalia,Stereotypy,Rigidity,Negativism,Waxy flexibility,Withdrawal | 100.0 | 100.0 | 100.0 |
| 10 | Excitement,Mutism,Staring,Grimacing,Echopraxia/echolalia,Stereotypy,Verbigeration,Rigidity,Negativism,Waxy flexibility | 100.0 | 100.0 | 100.0 |
| 10 | Excitement,Mutism,Staring,Grimacing,Echopraxia/echolalia,Stereotypy,Verbigeration,Rigidity,Negativism,Withdrawal | 100.0 | 100.0 | 100.0 |
| 10 | Excitement,Mutism,Staring,Grimacing,Echopraxia/echolalia,Stereotypy,Verbigeration,Rigidity,Waxy flexibility,Withdrawal | 100.0 | 100.0 | 100.0 |
| 10 | Excitement,Mutism,Staring,Grimacing,Stereotypy,Mannerisms,Verbigeration,Rigidity,Negativism,Withdrawal | 100.0 | 100.0 | 100.0 |
| 10 | Excitement,Mutism,Staring,Grimacing,Stereotypy,Mannerisms,Verbigeration,Rigidity,Waxy flexibility,Withdrawal | 100.0 | 100.0 | 100.0 |
| 10 | Excitement,Mutism,Staring,Grimacing,Stereotypy,Verbigeration,Rigidity,Negativism,Waxy flexibility,Withdrawal | 100.0 | 100.0 | 100.0 |
| 10 | Excitement,Mutism,Staring,Posturing/catalepsy,Echopraxia/echolalia,Mannerisms,Rigidity,Negativism,Waxy flexibility,Withdrawal | 100.0 | 100.0 | 100.0 |
| 10 | Excitement,Mutism,Staring,Posturing/catalepsy,Echopraxia/echolalia,Mannerisms,Verbigeration,Negativism,Waxy flexibility,Withdrawal | 100.0 | 100.0 | 100.0 |
| 10 | Excitement,Mutism,Staring,Posturing/catalepsy,Echopraxia/echolalia,Mannerisms,Verbigeration,Rigidity,Negativism,Waxy flexibility | 100.0 | 100.0 | 100.0 |
| 10 | Excitement,Mutism,Staring,Posturing/catalepsy,Echopraxia/echolalia,Mannerisms,Verbigeration,Rigidity,Negativism,Withdrawal | 100.0 | 100.0 | 100.0 |
| 10 | Excitement,Mutism,Staring,Posturing/catalepsy,Echopraxia/echolalia,Mannerisms,Verbigeration,Rigidity,Waxy flexibility,Withdrawal | 100.0 | 100.0 | 100.0 |
| 10 | Excitement,Mutism,Staring,Posturing/catalepsy,Echopraxia/echolalia,Stereotypy,Mannerisms,Negativism,Waxy flexibility,Withdrawal | 100.0 | 100.0 | 100.0 |
| 10 | Excitement,Mutism,Staring,Posturing/catalepsy,Echopraxia/echolalia,Stereotypy,Mannerisms,Rigidity,Negativism,Withdrawal | 100.0 | 100.0 | 100.0 |
| 10 | Excitement,Mutism,Staring,Posturing/catalepsy,Echopraxia/echolalia,Stereotypy,Mannerisms,Rigidity,Waxy flexibility,Withdrawal | 100.0 | 100.0 | 100.0 |
| 10 | Excitement,Mutism,Staring,Posturing/catalepsy,Echopraxia/echolalia,Stereotypy,Mannerisms,Verbigeration,Negativism,Waxy flexibility | 100.0 | 100.0 | 100.0 |
| 10 | Excitement,Mutism,Staring,Posturing/catalepsy,Echopraxia/echolalia,Stereotypy,Mannerisms,Verbigeration,Negativism,Withdrawal | 100.0 | 100.0 | 100.0 |
| 10 | Excitement,Mutism,Staring,Posturing/catalepsy,Echopraxia/echolalia,Stereotypy,Mannerisms,Verbigeration,Rigidity,Negativism | 100.0 | 100.0 | 100.0 |
| 10 | Excitement,Mutism,Staring,Posturing/catalepsy,Echopraxia/echolalia,Stereotypy,Mannerisms,Verbigeration,Rigidity,Withdrawal | 100.0 | 100.0 | 100.0 |
| 10 | Excitement,Mutism,Staring,Posturing/catalepsy,Echopraxia/echolalia,Stereotypy,Rigidity,Negativism,Waxy flexibility,Withdrawal | 100.0 | 100.0 | 100.0 |
| 10 | Excitement,Mutism,Staring,Posturing/catalepsy,Echopraxia/echolalia,Stereotypy,Verbigeration,Negativism,Waxy flexibility,Withdrawal | 100.0 | 100.0 | 100.0 |
| 10 | Excitement,Mutism,Staring,Posturing/catalepsy,Echopraxia/echolalia,Stereotypy,Verbigeration,Rigidity,Negativism,Waxy flexibility | 100.0 | 100.0 | 100.0 |
| 10 | Excitement,Mutism,Staring,Posturing/catalepsy,Echopraxia/echolalia,Stereotypy,Verbigeration,Rigidity,Negativism,Withdrawal | 100.0 | 100.0 | 100.0 |
| 10 | Excitement,Mutism,Staring,Posturing/catalepsy,Echopraxia/echolalia,Stereotypy,Verbigeration,Rigidity,Waxy flexibility,Withdrawal | 100.0 | 100.0 | 100.0 |
| 10 | Excitement,Mutism,Staring,Posturing/catalepsy,Echopraxia/echolalia,Verbigeration,Rigidity,Negativism,Waxy flexibility,Withdrawal | 100.0 | 100.0 | 100.0 |
| 10 | Excitement,Mutism,Staring,Posturing/catalepsy,Grimacing,Echopraxia/echolalia,Mannerisms,Negativism,Waxy flexibility,Withdrawal | 100.0 | 100.0 | 100.0 |
| 10 | Excitement,Mutism,Staring,Posturing/catalepsy,Grimacing,Echopraxia/echolalia,Mannerisms,Rigidity,Negativism,Withdrawal | 100.0 | 100.0 | 100.0 |
| 10 | Excitement,Mutism,Staring,Posturing/catalepsy,Grimacing,Echopraxia/echolalia,Mannerisms,Rigidity,Waxy flexibility,Withdrawal | 100.0 | 100.0 | 100.0 |
| 10 | Excitement,Mutism,Staring,Posturing/catalepsy,Grimacing,Echopraxia/echolalia,Mannerisms,Verbigeration,Negativism,Waxy flexibility | 100.0 | 100.0 | 100.0 |
| 10 | Excitement,Mutism,Staring,Posturing/catalepsy,Grimacing,Echopraxia/echolalia,Mannerisms,Verbigeration,Negativism,Withdrawal | 100.0 | 100.0 | 100.0 |
| 10 | Excitement,Mutism,Staring,Posturing/catalepsy,Grimacing,Echopraxia/echolalia,Mannerisms,Verbigeration,Rigidity,Negativism | 100.0 | 100.0 | 100.0 |
| 10 | Excitement,Mutism,Staring,Posturing/catalepsy,Grimacing,Echopraxia/echolalia,Mannerisms,Verbigeration,Rigidity,Withdrawal | 100.0 | 100.0 | 100.0 |
| 10 | Excitement,Mutism,Staring,Posturing/catalepsy,Grimacing,Echopraxia/echolalia,Rigidity,Negativism,Waxy flexibility,Withdrawal | 100.0 | 100.0 | 100.0 |
| 10 | Excitement,Mutism,Staring,Posturing/catalepsy,Grimacing,Echopraxia/echolalia,Stereotypy,Mannerisms,Negativism,Withdrawal | 100.0 | 100.0 | 100.0 |
| 10 | Excitement,Mutism,Staring,Posturing/catalepsy,Grimacing,Echopraxia/echolalia,Stereotypy,Mannerisms,Rigidity,Withdrawal | 100.0 | 100.0 | 100.0 |
| 10 | Excitement,Mutism,Staring,Posturing/catalepsy,Grimacing,Echopraxia/echolalia,Stereotypy,Negativism,Waxy flexibility,Withdrawal | 100.0 | 100.0 | 100.0 |
| 10 | Excitement,Mutism,Staring,Posturing/catalepsy,Grimacing,Echopraxia/echolalia,Stereotypy,Rigidity,Negativism,Withdrawal | 100.0 | 100.0 | 100.0 |
| 10 | Excitement,Mutism,Staring,Posturing/catalepsy,Grimacing,Echopraxia/echolalia,Stereotypy,Rigidity,Waxy flexibility,Withdrawal | 100.0 | 100.0 | 100.0 |
| 10 | Excitement,Mutism,Staring,Posturing/catalepsy,Grimacing,Echopraxia/echolalia,Stereotypy,Verbigeration,Negativism,Waxy flexibility | 100.0 | 100.0 | 100.0 |
| 10 | Excitement,Mutism,Staring,Posturing/catalepsy,Grimacing,Echopraxia/echolalia,Stereotypy,Verbigeration,Negativism,Withdrawal | 100.0 | 100.0 | 100.0 |
| 10 | Excitement,Mutism,Staring,Posturing/catalepsy,Grimacing,Echopraxia/echolalia,Stereotypy,Verbigeration,Rigidity,Negativism | 100.0 | 100.0 | 100.0 |
| 10 | Excitement,Mutism,Staring,Posturing/catalepsy,Grimacing,Echopraxia/echolalia,Stereotypy,Verbigeration,Rigidity,Withdrawal | 100.0 | 100.0 | 100.0 |
| 10 | Excitement,Mutism,Staring,Posturing/catalepsy,Grimacing,Echopraxia/echolalia,Verbigeration,Negativism,Waxy flexibility,Withdrawal | 100.0 | 100.0 | 100.0 |
| 10 | Excitement,Mutism,Staring,Posturing/catalepsy,Grimacing,Echopraxia/echolalia,Verbigeration,Rigidity,Negativism,Waxy flexibility | 100.0 | 100.0 | 100.0 |
| 10 | Excitement,Mutism,Staring,Posturing/catalepsy,Grimacing,Echopraxia/echolalia,Verbigeration,Rigidity,Negativism,Withdrawal | 100.0 | 100.0 | 100.0 |
| 10 | Excitement,Mutism,Staring,Posturing/catalepsy,Grimacing,Echopraxia/echolalia,Verbigeration,Rigidity,Waxy flexibility,Withdrawal | 100.0 | 100.0 | 100.0 |
| 10 | Excitement,Mutism,Staring,Posturing/catalepsy,Grimacing,Mannerisms,Verbigeration,Negativism,Waxy flexibility,Withdrawal | 100.0 | 100.0 | 100.0 |
| 10 | Excitement,Mutism,Staring,Posturing/catalepsy,Grimacing,Mannerisms,Verbigeration,Rigidity,Negativism,Withdrawal | 100.0 | 100.0 | 100.0 |
| 10 | Excitement,Mutism,Staring,Posturing/catalepsy,Grimacing,Mannerisms,Verbigeration,Rigidity,Waxy flexibility,Withdrawal | 100.0 | 100.0 | 100.0 |
| 10 | Excitement,Mutism,Staring,Posturing/catalepsy,Grimacing,Stereotypy,Mannerisms,Verbigeration,Negativism,Withdrawal | 100.0 | 100.0 | 100.0 |
| 10 | Excitement,Mutism,Staring,Posturing/catalepsy,Grimacing,Stereotypy,Mannerisms,Verbigeration,Rigidity,Withdrawal | 100.0 | 100.0 | 100.0 |
| 10 | Excitement,Mutism,Staring,Posturing/catalepsy,Grimacing,Stereotypy,Verbigeration,Negativism,Waxy flexibility,Withdrawal | 100.0 | 100.0 | 100.0 |
| 10 | Excitement,Mutism,Staring,Posturing/catalepsy,Grimacing,Stereotypy,Verbigeration,Rigidity,Negativism,Withdrawal | 100.0 | 100.0 | 100.0 |
| 10 | Excitement,Mutism,Staring,Posturing/catalepsy,Grimacing,Stereotypy,Verbigeration,Rigidity,Waxy flexibility,Withdrawal | 100.0 | 100.0 | 100.0 |
| 10 | Excitement,Mutism,Staring,Posturing/catalepsy,Mannerisms,Verbigeration,Rigidity,Negativism,Waxy flexibility,Withdrawal | 100.0 | 100.0 | 100.0 |
| 10 | Excitement,Mutism,Staring,Posturing/catalepsy,Stereotypy,Mannerisms,Verbigeration,Negativism,Waxy flexibility,Withdrawal | 100.0 | 100.0 | 100.0 |
| 10 | Excitement,Mutism,Staring,Posturing/catalepsy,Stereotypy,Mannerisms,Verbigeration,Rigidity,Negativism,Withdrawal | 100.0 | 100.0 | 100.0 |
| 10 | Excitement,Mutism,Staring,Posturing/catalepsy,Stereotypy,Mannerisms,Verbigeration,Rigidity,Waxy flexibility,Withdrawal | 100.0 | 100.0 | 100.0 |
| 10 | Excitement,Mutism,Staring,Posturing/catalepsy,Stereotypy,Verbigeration,Rigidity,Negativism,Waxy flexibility,Withdrawal | 100.0 | 100.0 | 100.0 |
| 10 | Excitement,Mutism,Staring,Stereotypy,Mannerisms,Verbigeration,Rigidity,Negativism,Waxy flexibility,Withdrawal | 100.0 | 100.0 | 100.0 |
| 10 | Immobility/stupor,Mutism,Staring,Grimacing,Echopraxia/echolalia,Stereotypy,Mannerisms,Rigidity,Negativism,Withdrawal | 100.0 | 100.0 | 100.0 |
| 10 | Immobility/stupor,Mutism,Staring,Grimacing,Stereotypy,Mannerisms,Verbigeration,Rigidity,Negativism,Withdrawal | 100.0 | 100.0 | 100.0 |
| 10 | Immobility/stupor,Mutism,Staring,Posturing/catalepsy,Grimacing,Echopraxia/echolalia,Stereotypy,Mannerisms,Negativism,Withdrawal | 100.0 | 100.0 | 100.0 |
| 10 | Immobility/stupor,Mutism,Staring,Posturing/catalepsy,Grimacing,Stereotypy,Mannerisms,Verbigeration,Negativism,Withdrawal | 100.0 | 100.0 | 100.0 |
| 10 | Immobility/stupor,Staring,Grimacing,Echopraxia/echolalia,Stereotypy,Mannerisms,Rigidity,Negativism,Waxy flexibility,Withdrawal | 100.0 | 100.0 | 100.0 |
| 10 | Immobility/stupor,Staring,Grimacing,Echopraxia/echolalia,Stereotypy,Mannerisms,Verbigeration,Rigidity,Negativism,Withdrawal | 100.0 | 100.0 | 100.0 |
| 10 | Immobility/stupor,Staring,Grimacing,Stereotypy,Mannerisms,Verbigeration,Rigidity,Negativism,Waxy flexibility,Withdrawal | 100.0 | 100.0 | 100.0 |
| 10 | Immobility/stupor,Staring,Posturing/catalepsy,Grimacing,Echopraxia/echolalia,Stereotypy,Mannerisms,Rigidity,Negativism,Withdrawal | 100.0 | 100.0 | 100.0 |
| 10 | Immobility/stupor,Staring,Posturing/catalepsy,Grimacing,Stereotypy,Mannerisms,Verbigeration,Rigidity,Negativism,Withdrawal | 100.0 | 100.0 | 100.0 |
| 10 | Mutism,Staring,Grimacing,Echopraxia/echolalia,Stereotypy,Mannerisms,Rigidity,Negativism,Waxy flexibility,Withdrawal | 100.0 | 100.0 | 100.0 |
| 10 | Mutism,Staring,Grimacing,Echopraxia/echolalia,Stereotypy,Mannerisms,Verbigeration,Rigidity,Negativism,Withdrawal | 100.0 | 100.0 | 100.0 |
| 10 | Mutism,Staring,Grimacing,Stereotypy,Mannerisms,Verbigeration,Rigidity,Negativism,Waxy flexibility,Withdrawal | 100.0 | 100.0 | 100.0 |
| 10 | Mutism,Staring,Posturing/catalepsy,Grimacing,Echopraxia/echolalia,Stereotypy,Mannerisms,Negativism,Waxy flexibility,Withdrawal | 100.0 | 100.0 | 100.0 |
| 10 | Mutism,Staring,Posturing/catalepsy,Grimacing,Echopraxia/echolalia,Stereotypy,Mannerisms,Rigidity,Negativism,Withdrawal | 100.0 | 100.0 | 100.0 |
| 10 | Mutism,Staring,Posturing/catalepsy,Grimacing,Echopraxia/echolalia,Stereotypy,Mannerisms,Verbigeration,Negativism,Withdrawal | 100.0 | 100.0 | 100.0 |
| 10 | Mutism,Staring,Posturing/catalepsy,Grimacing,Stereotypy,Mannerisms,Verbigeration,Negativism,Waxy flexibility,Withdrawal | 100.0 | 100.0 | 100.0 |
| 10 | Mutism,Staring,Posturing/catalepsy,Grimacing,Stereotypy,Mannerisms,Verbigeration,Rigidity,Negativism,Withdrawal | 100.0 | 100.0 | 100.0 |
| 10 | Excitement,Immobility/stupor,Mutism,Echopraxia/echolalia,Stereotypy,Mannerisms,Verbigeration,Rigidity,Negativism,Withdrawal | 99.8 | 99.3 | 100.0 |
| 10 | Excitement,Immobility/stupor,Mutism,Echopraxia/echolalia,Stereotypy,Verbigeration,Rigidity,Negativism,Waxy flexibility,Withdrawal | 99.8 | 99.3 | 100.0 |
| 10 | Excitement,Immobility/stupor,Mutism,Grimacing,Echopraxia/echolalia,Stereotypy,Verbigeration,Rigidity,Negativism,Withdrawal | 99.8 | 99.3 | 100.0 |
| 10 | Excitement,Immobility/stupor,Mutism,Posturing/catalepsy,Echopraxia/echolalia,Mannerisms,Verbigeration,Rigidity,Negativism,Withdrawal | 99.8 | 99.3 | 100.0 |
| 10 | Excitement,Immobility/stupor,Mutism,Posturing/catalepsy,Echopraxia/echolalia,Stereotypy,Mannerisms,Verbigeration,Rigidity,Negativism | 99.8 | 99.3 | 100.0 |
| 10 | Excitement,Immobility/stupor,Mutism,Posturing/catalepsy,Echopraxia/echolalia,Stereotypy,Verbigeration,Rigidity,Negativism,Waxy flexibility | 99.8 | 99.3 | 100.0 |
| 10 | Excitement,Immobility/stupor,Mutism,Posturing/catalepsy,Echopraxia/echolalia,Verbigeration,Rigidity,Negativism,Waxy flexibility,Withdrawal | 99.8 | 99.3 | 100.0 |
| 10 | Excitement,Immobility/stupor,Mutism,Posturing/catalepsy,Grimacing,Echopraxia/echolalia,Mannerisms,Rigidity,Negativism,Withdrawal | 99.8 | 99.3 | 100.0 |
| 10 | Excitement,Immobility/stupor,Mutism,Posturing/catalepsy,Grimacing,Echopraxia/echolalia,Mannerisms,Verbigeration,Rigidity,Negativism | 99.8 | 99.3 | 100.0 |
| 10 | Excitement,Immobility/stupor,Mutism,Posturing/catalepsy,Grimacing,Echopraxia/echolalia,Rigidity,Negativism,Waxy flexibility,Withdrawal | 99.8 | 99.3 | 100.0 |
| 10 | Excitement,Immobility/stupor,Mutism,Posturing/catalepsy,Grimacing,Echopraxia/echolalia,Stereotypy,Rigidity,Negativism,Withdrawal | 99.8 | 99.3 | 100.0 |
| 10 | Excitement,Immobility/stupor,Mutism,Posturing/catalepsy,Grimacing,Echopraxia/echolalia,Stereotypy,Verbigeration,Rigidity,Negativism | 99.8 | 99.3 | 100.0 |
| 10 | Excitement,Immobility/stupor,Mutism,Posturing/catalepsy,Grimacing,Echopraxia/echolalia,Verbigeration,Rigidity,Negativism,Waxy flexibility | 99.8 | 99.3 | 100.0 |
| 10 | Excitement,Immobility/stupor,Mutism,Posturing/catalepsy,Grimacing,Mannerisms,Verbigeration,Rigidity,Negativism,Withdrawal | 99.8 | 99.3 | 100.0 |
| 10 | Excitement,Immobility/stupor,Mutism,Posturing/catalepsy,Grimacing,Stereotypy,Verbigeration,Rigidity,Negativism,Withdrawal | 99.8 | 99.3 | 100.0 |
| 10 | Excitement,Immobility/stupor,Mutism,Posturing/catalepsy,Stereotypy,Mannerisms,Verbigeration,Rigidity,Negativism,Withdrawal | 99.8 | 99.3 | 100.0 |
| 10 | Excitement,Immobility/stupor,Mutism,Posturing/catalepsy,Stereotypy,Verbigeration,Rigidity,Negativism,Waxy flexibility,Withdrawal | 99.8 | 99.3 | 100.0 |
| 10 | Excitement,Immobility/stupor,Mutism,Staring,Echopraxia/echolalia,Mannerisms,Rigidity,Negativism,Waxy flexibility,Withdrawal | 99.8 | 99.3 | 100.0 |
| 10 | Excitement,Immobility/stupor,Mutism,Staring,Echopraxia/echolalia,Mannerisms,Verbigeration,Rigidity,Negativism,Waxy flexibility | 99.8 | 99.3 | 100.0 |
| 10 | Excitement,Immobility/stupor,Mutism,Staring,Echopraxia/echolalia,Mannerisms,Verbigeration,Rigidity,Negativism,Withdrawal | 99.8 | 99.3 | 100.0 |
| 10 | Excitement,Immobility/stupor,Mutism,Staring,Echopraxia/echolalia,Mannerisms,Verbigeration,Rigidity,Waxy flexibility,Withdrawal | 99.8 | 99.3 | 100.0 |
| 10 | Excitement,Immobility/stupor,Mutism,Staring,Echopraxia/echolalia,Stereotypy,Mannerisms,Rigidity,Negativism,Waxy flexibility | 99.8 | 99.3 | 100.0 |
| 10 | Excitement,Immobility/stupor,Mutism,Staring,Echopraxia/echolalia,Stereotypy,Mannerisms,Verbigeration,Rigidity,Negativism | 99.8 | 99.3 | 100.0 |
| 10 | Excitement,Immobility/stupor,Mutism,Staring,Echopraxia/echolalia,Stereotypy,Mannerisms,Verbigeration,Rigidity,Waxy flexibility | 99.8 | 99.3 | 100.0 |
| 10 | Excitement,Immobility/stupor,Mutism,Staring,Echopraxia/echolalia,Stereotypy,Mannerisms,Verbigeration,Rigidity,Withdrawal | 99.8 | 99.3 | 100.0 |
| 10 | Excitement,Immobility/stupor,Mutism,Staring,Echopraxia/echolalia,Verbigeration,Rigidity,Negativism,Waxy flexibility,Withdrawal | 99.8 | 99.3 | 100.0 |
| 10 | Excitement,Immobility/stupor,Mutism,Staring,Grimacing,Echopraxia/echolalia,Mannerisms,Rigidity,Negativism,Withdrawal | 99.8 | 99.3 | 100.0 |
| 10 | Excitement,Immobility/stupor,Mutism,Staring,Grimacing,Echopraxia/echolalia,Mannerisms,Rigidity,Waxy flexibility,Withdrawal | 99.8 | 99.3 | 100.0 |
| 10 | Excitement,Immobility/stupor,Mutism,Staring,Grimacing,Echopraxia/echolalia,Rigidity,Negativism,Waxy flexibility,Withdrawal | 99.8 | 99.3 | 100.0 |
| 10 | Excitement,Immobility/stupor,Mutism,Staring,Grimacing,Echopraxia/echolalia,Stereotypy,Mannerisms,Rigidity,Withdrawal | 99.8 | 99.3 | 100.0 |
| 10 | Excitement,Immobility/stupor,Mutism,Staring,Grimacing,Echopraxia/echolalia,Stereotypy,Rigidity,Negativism,Waxy flexibility | 99.8 | 99.3 | 100.0 |
| 10 | Excitement,Immobility/stupor,Mutism,Staring,Grimacing,Echopraxia/echolalia,Stereotypy,Verbigeration,Rigidity,Negativism | 99.8 | 99.3 | 100.0 |
| 10 | Excitement,Immobility/stupor,Mutism,Staring,Grimacing,Echopraxia/echolalia,Stereotypy,Verbigeration,Rigidity,Waxy flexibility | 99.8 | 99.3 | 100.0 |
| 10 | Excitement,Immobility/stupor,Mutism,Staring,Grimacing,Echopraxia/echolalia,Stereotypy,Verbigeration,Rigidity,Withdrawal | 99.8 | 99.3 | 100.0 |
| 10 | Excitement,Immobility/stupor,Mutism,Staring,Grimacing,Echopraxia/echolalia,Verbigeration,Rigidity,Negativism,Waxy flexibility | 99.8 | 99.3 | 100.0 |
| 10 | Excitement,Immobility/stupor,Mutism,Staring,Grimacing,Echopraxia/echolalia,Verbigeration,Rigidity,Negativism,Withdrawal | 99.8 | 99.3 | 100.0 |
| 10 | Excitement,Immobility/stupor,Mutism,Staring,Grimacing,Echopraxia/echolalia,Verbigeration,Rigidity,Waxy flexibility,Withdrawal | 99.8 | 99.3 | 100.0 |
| 10 | Excitement,Immobility/stupor,Mutism,Staring,Grimacing,Mannerisms,Verbigeration,Rigidity,Negativism,Waxy flexibility | 99.8 | 99.3 | 100.0 |
| 10 | Excitement,Immobility/stupor,Mutism,Staring,Grimacing,Mannerisms,Verbigeration,Rigidity,Negativism,Withdrawal | 99.8 | 99.3 | 100.0 |
| 10 | Excitement,Immobility/stupor,Mutism,Staring,Grimacing,Mannerisms,Verbigeration,Rigidity,Waxy flexibility,Withdrawal | 99.8 | 99.3 | 100.0 |
| 10 | Excitement,Immobility/stupor,Mutism,Staring,Grimacing,Stereotypy,Mannerisms,Rigidity,Waxy flexibility,Withdrawal | 99.8 | 99.3 | 100.0 |
| 10 | Excitement,Immobility/stupor,Mutism,Staring,Grimacing,Stereotypy,Mannerisms,Verbigeration,Rigidity,Negativism | 99.8 | 99.3 | 100.0 |
| 10 | Excitement,Immobility/stupor,Mutism,Staring,Grimacing,Stereotypy,Mannerisms,Verbigeration,Rigidity,Waxy flexibility | 99.8 | 99.3 | 100.0 |
| 10 | Excitement,Immobility/stupor,Mutism,Staring,Grimacing,Stereotypy,Mannerisms,Verbigeration,Rigidity,Withdrawal | 99.8 | 99.3 | 100.0 |
| 10 | Excitement,Immobility/stupor,Mutism,Staring,Grimacing,Stereotypy,Rigidity,Negativism,Waxy flexibility,Withdrawal | 99.8 | 99.3 | 100.0 |
| 10 | Excitement,Immobility/stupor,Mutism,Staring,Mannerisms,Verbigeration,Rigidity,Negativism,Waxy flexibility,Withdrawal | 99.8 | 99.3 | 100.0 |
| 10 | Excitement,Immobility/stupor,Mutism,Staring,Posturing/catalepsy,Echopraxia/echolalia,Mannerisms,Rigidity,Negativism,Waxy flexibility | 99.8 | 99.3 | 100.0 |
| 10 | Excitement,Immobility/stupor,Mutism,Staring,Posturing/catalepsy,Echopraxia/echolalia,Mannerisms,Verbigeration,Rigidity,Waxy flexibility | 99.8 | 99.3 | 100.0 |
| 10 | Excitement,Immobility/stupor,Mutism,Staring,Posturing/catalepsy,Echopraxia/echolalia,Stereotypy,Mannerisms,Negativism,Waxy flexibility | 99.8 | 99.3 | 100.0 |
| 10 | Excitement,Immobility/stupor,Mutism,Staring,Posturing/catalepsy,Echopraxia/echolalia,Stereotypy,Mannerisms,Rigidity,Negativism | 99.8 | 99.3 | 100.0 |
| 10 | Excitement,Immobility/stupor,Mutism,Staring,Posturing/catalepsy,Echopraxia/echolalia,Stereotypy,Mannerisms,Verbigeration,Negativism | 99.8 | 99.3 | 100.0 |
| 10 | Excitement,Immobility/stupor,Mutism,Staring,Posturing/catalepsy,Echopraxia/echolalia,Stereotypy,Mannerisms,Verbigeration,Rigidity | 99.8 | 99.3 | 100.0 |
| 10 | Excitement,Immobility/stupor,Mutism,Staring,Posturing/catalepsy,Echopraxia/echolalia,Stereotypy,Rigidity,Negativism,Waxy flexibility | 99.8 | 99.3 | 100.0 |
| 10 | Excitement,Immobility/stupor,Mutism,Staring,Posturing/catalepsy,Echopraxia/echolalia,Stereotypy,Verbigeration,Rigidity,Waxy flexibility | 99.8 | 99.3 | 100.0 |
| 10 | Excitement,Immobility/stupor,Mutism,Staring,Posturing/catalepsy,Grimacing,Echopraxia/echolalia,Mannerisms,Negativism,Waxy flexibility | 99.8 | 99.3 | 100.0 |
| 10 | Excitement,Immobility/stupor,Mutism,Staring,Posturing/catalepsy,Grimacing,Echopraxia/echolalia,Mannerisms,Rigidity,Negativism | 99.8 | 99.3 | 100.0 |
| 10 | Excitement,Immobility/stupor,Mutism,Staring,Posturing/catalepsy,Grimacing,Echopraxia/echolalia,Mannerisms,Verbigeration,Negativism | 99.8 | 99.3 | 100.0 |
| 10 | Excitement,Immobility/stupor,Mutism,Staring,Posturing/catalepsy,Grimacing,Echopraxia/echolalia,Mannerisms,Verbigeration,Rigidity | 99.8 | 99.3 | 100.0 |
| 10 | Excitement,Immobility/stupor,Mutism,Staring,Posturing/catalepsy,Grimacing,Echopraxia/echolalia,Rigidity,Negativism,Waxy flexibility | 99.8 | 99.3 | 100.0 |
| 10 | Excitement,Immobility/stupor,Mutism,Staring,Posturing/catalepsy,Grimacing,Echopraxia/echolalia,Stereotypy,Negativism,Waxy flexibility | 99.8 | 99.3 | 100.0 |
| 10 | Excitement,Immobility/stupor,Mutism,Staring,Posturing/catalepsy,Grimacing,Echopraxia/echolalia,Stereotypy,Rigidity,Negativism | 99.8 | 99.3 | 100.0 |
| 10 | Excitement,Immobility/stupor,Mutism,Staring,Posturing/catalepsy,Grimacing,Echopraxia/echolalia,Stereotypy,Verbigeration,Negativism | 99.8 | 99.3 | 100.0 |
| 10 | Excitement,Immobility/stupor,Mutism,Staring,Posturing/catalepsy,Grimacing,Echopraxia/echolalia,Stereotypy,Verbigeration,Rigidity | 99.8 | 99.3 | 100.0 |
| 10 | Excitement,Immobility/stupor,Mutism,Staring,Posturing/catalepsy,Grimacing,Echopraxia/echolalia,Verbigeration,Rigidity,Waxy flexibility | 99.8 | 99.3 | 100.0 |
| 10 | Excitement,Immobility/stupor,Mutism,Staring,Posturing/catalepsy,Grimacing,Mannerisms,Negativism,Waxy flexibility,Withdrawal | 99.8 | 99.3 | 100.0 |
| 10 | Excitement,Immobility/stupor,Mutism,Staring,Posturing/catalepsy,Grimacing,Mannerisms,Rigidity,Negativism,Withdrawal | 99.8 | 99.3 | 100.0 |
| 10 | Excitement,Immobility/stupor,Mutism,Staring,Posturing/catalepsy,Grimacing,Mannerisms,Rigidity,Waxy flexibility,Withdrawal | 99.8 | 99.3 | 100.0 |
| 10 | Excitement,Immobility/stupor,Mutism,Staring,Posturing/catalepsy,Grimacing,Mannerisms,Verbigeration,Rigidity,Waxy flexibility | 99.8 | 99.3 | 100.0 |
| 10 | Excitement,Immobility/stupor,Mutism,Staring,Posturing/catalepsy,Grimacing,Stereotypy,Mannerisms,Negativism,Withdrawal | 99.8 | 99.3 | 100.0 |
| 10 | Excitement,Immobility/stupor,Mutism,Staring,Posturing/catalepsy,Grimacing,Stereotypy,Mannerisms,Rigidity,Withdrawal | 99.8 | 99.3 | 100.0 |
| 10 | Excitement,Immobility/stupor,Mutism,Staring,Posturing/catalepsy,Grimacing,Stereotypy,Mannerisms,Verbigeration,Negativism | 99.8 | 99.3 | 100.0 |
| 10 | Excitement,Immobility/stupor,Mutism,Staring,Posturing/catalepsy,Grimacing,Stereotypy,Mannerisms,Verbigeration,Rigidity | 99.8 | 99.3 | 100.0 |
| 10 | Excitement,Immobility/stupor,Mutism,Staring,Posturing/catalepsy,Grimacing,Stereotypy,Negativism,Waxy flexibility,Withdrawal | 99.8 | 99.3 | 100.0 |
| 10 | Excitement,Immobility/stupor,Mutism,Staring,Posturing/catalepsy,Grimacing,Stereotypy,Rigidity,Negativism,Withdrawal | 99.8 | 99.3 | 100.0 |
| 10 | Excitement,Immobility/stupor,Mutism,Staring,Posturing/catalepsy,Grimacing,Stereotypy,Rigidity,Waxy flexibility,Withdrawal | 99.8 | 99.3 | 100.0 |
| 10 | Excitement,Immobility/stupor,Mutism,Staring,Posturing/catalepsy,Grimacing,Stereotypy,Verbigeration,Rigidity,Waxy flexibility | 99.8 | 99.3 | 100.0 |
| 10 | Excitement,Immobility/stupor,Mutism,Staring,Posturing/catalepsy,Grimacing,Verbigeration,Negativism,Waxy flexibility,Withdrawal | 99.8 | 99.3 | 100.0 |
| 10 | Excitement,Immobility/stupor,Mutism,Staring,Posturing/catalepsy,Grimacing,Verbigeration,Rigidity,Negativism,Waxy flexibility | 99.8 | 99.3 | 100.0 |
| 10 | Excitement,Immobility/stupor,Mutism,Staring,Posturing/catalepsy,Grimacing,Verbigeration,Rigidity,Negativism,Withdrawal | 99.8 | 99.3 | 100.0 |
| 10 | Excitement,Immobility/stupor,Mutism,Staring,Posturing/catalepsy,Grimacing,Verbigeration,Rigidity,Waxy flexibility,Withdrawal | 99.8 | 99.3 | 100.0 |
| 10 | Excitement,Immobility/stupor,Mutism,Staring,Posturing/catalepsy,Mannerisms,Rigidity,Negativism,Waxy flexibility,Withdrawal | 99.8 | 99.3 | 100.0 |
| 10 | Excitement,Immobility/stupor,Mutism,Staring,Posturing/catalepsy,Stereotypy,Mannerisms,Negativism,Waxy flexibility,Withdrawal | 99.8 | 99.3 | 100.0 |
| 10 | Excitement,Immobility/stupor,Mutism,Staring,Posturing/catalepsy,Stereotypy,Mannerisms,Rigidity,Negativism,Withdrawal | 99.8 | 99.3 | 100.0 |
| 10 | Excitement,Immobility/stupor,Mutism,Staring,Posturing/catalepsy,Stereotypy,Mannerisms,Rigidity,Waxy flexibility,Withdrawal | 99.8 | 99.3 | 100.0 |
| 10 | Excitement,Immobility/stupor,Mutism,Staring,Posturing/catalepsy,Stereotypy,Mannerisms,Verbigeration,Rigidity,Waxy flexibility | 99.8 | 99.3 | 100.0 |
| 10 | Excitement,Immobility/stupor,Mutism,Staring,Posturing/catalepsy,Stereotypy,Rigidity,Negativism,Waxy flexibility,Withdrawal | 99.8 | 99.3 | 100.0 |
| 10 | Excitement,Immobility/stupor,Mutism,Staring,Posturing/catalepsy,Verbigeration,Rigidity,Negativism,Waxy flexibility,Withdrawal | 99.8 | 99.3 | 100.0 |
| 10 | Excitement,Immobility/stupor,Mutism,Staring,Stereotypy,Mannerisms,Rigidity,Negativism,Waxy flexibility,Withdrawal | 99.8 | 99.3 | 100.0 |
| 10 | Excitement,Immobility/stupor,Posturing/catalepsy,Echopraxia/echolalia,Stereotypy,Mannerisms,Verbigeration,Rigidity,Negativism,Withdrawal | 99.8 | 99.3 | 100.0 |
| 10 | Excitement,Immobility/stupor,Posturing/catalepsy,Grimacing,Echopraxia/echolalia,Mannerisms,Verbigeration,Rigidity,Negativism,Withdrawal | 99.8 | 99.3 | 100.0 |
| 10 | Excitement,Immobility/stupor,Staring,Echopraxia/echolalia,Mannerisms,Verbigeration,Rigidity,Negativism,Waxy flexibility,Withdrawal | 99.8 | 99.3 | 100.0 |
| 10 | Excitement,Immobility/stupor,Staring,Echopraxia/echolalia,Stereotypy,Mannerisms,Verbigeration,Rigidity,Negativism,Waxy flexibility | 99.8 | 99.3 | 100.0 |
| 10 | Excitement,Immobility/stupor,Staring,Echopraxia/echolalia,Stereotypy,Mannerisms,Verbigeration,Rigidity,Waxy flexibility,Withdrawal | 99.8 | 99.3 | 100.0 |
| 10 | Excitement,Immobility/stupor,Staring,Echopraxia/echolalia,Stereotypy,Verbigeration,Rigidity,Negativism,Waxy flexibility,Withdrawal | 99.8 | 99.3 | 100.0 |
| 10 | Excitement,Immobility/stupor,Staring,Grimacing,Echopraxia/echolalia,Mannerisms,Rigidity,Negativism,Waxy flexibility,Withdrawal | 99.8 | 99.3 | 100.0 |
| 10 | Excitement,Immobility/stupor,Staring,Grimacing,Echopraxia/echolalia,Mannerisms,Verbigeration,Rigidity,Negativism,Withdrawal | 99.8 | 99.3 | 100.0 |
| 10 | Excitement,Immobility/stupor,Staring,Grimacing,Echopraxia/echolalia,Stereotypy,Mannerisms,Rigidity,Waxy flexibility,Withdrawal | 99.8 | 99.3 | 100.0 |
| 10 | Excitement,Immobility/stupor,Staring,Grimacing,Echopraxia/echolalia,Stereotypy,Rigidity,Negativism,Waxy flexibility,Withdrawal | 99.8 | 99.3 | 100.0 |
| 10 | Excitement,Immobility/stupor,Staring,Grimacing,Echopraxia/echolalia,Stereotypy,Verbigeration,Rigidity,Negativism,Withdrawal | 99.8 | 99.3 | 100.0 |
| 10 | Excitement,Immobility/stupor,Staring,Grimacing,Mannerisms,Verbigeration,Rigidity,Negativism,Waxy flexibility,Withdrawal | 99.8 | 99.3 | 100.0 |
| 10 | Excitement,Immobility/stupor,Staring,Grimacing,Stereotypy,Mannerisms,Rigidity,Negativism,Waxy flexibility,Withdrawal | 99.8 | 99.3 | 100.0 |
| 10 | Excitement,Immobility/stupor,Staring,Grimacing,Stereotypy,Mannerisms,Verbigeration,Rigidity,Negativism,Waxy flexibility | 99.8 | 99.3 | 100.0 |
| 10 | Excitement,Immobility/stupor,Staring,Grimacing,Stereotypy,Mannerisms,Verbigeration,Rigidity,Waxy flexibility,Withdrawal | 99.8 | 99.3 | 100.0 |
| 10 | Excitement,Immobility/stupor,Staring,Grimacing,Stereotypy,Verbigeration,Rigidity,Negativism,Waxy flexibility,Withdrawal | 99.8 | 99.3 | 100.0 |
| 10 | Excitement,Immobility/stupor,Staring,Posturing/catalepsy,Echopraxia/echolalia,Mannerisms,Verbigeration,Negativism,Waxy flexibility,Withdrawal | 99.8 | 99.3 | 100.0 |
| 10 | Excitement,Immobility/stupor,Staring,Posturing/catalepsy,Echopraxia/echolalia,Mannerisms,Verbigeration,Rigidity,Negativism,Waxy flexibility | 99.8 | 99.3 | 100.0 |
| 10 | Excitement,Immobility/stupor,Staring,Posturing/catalepsy,Echopraxia/echolalia,Mannerisms,Verbigeration,Rigidity,Waxy flexibility,Withdrawal | 99.8 | 99.3 | 100.0 |
| 10 | Excitement,Immobility/stupor,Staring,Posturing/catalepsy,Echopraxia/echolalia,Stereotypy,Mannerisms,Negativism,Waxy flexibility,Withdrawal | 99.8 | 99.3 | 100.0 |
| 10 | Excitement,Immobility/stupor,Staring,Posturing/catalepsy,Echopraxia/echolalia,Stereotypy,Mannerisms,Rigidity,Waxy flexibility,Withdrawal | 99.8 | 99.3 | 100.0 |
| 10 | Excitement,Immobility/stupor,Staring,Posturing/catalepsy,Echopraxia/echolalia,Stereotypy,Mannerisms,Verbigeration,Negativism,Withdrawal | 99.8 | 99.3 | 100.0 |
| 10 | Excitement,Immobility/stupor,Staring,Posturing/catalepsy,Echopraxia/echolalia,Stereotypy,Mannerisms,Verbigeration,Rigidity,Negativism | 99.8 | 99.3 | 100.0 |
| 10 | Excitement,Immobility/stupor,Staring,Posturing/catalepsy,Echopraxia/echolalia,Stereotypy,Mannerisms,Verbigeration,Rigidity,Withdrawal | 99.8 | 99.3 | 100.0 |
| 10 | Excitement,Immobility/stupor,Staring,Posturing/catalepsy,Echopraxia/echolalia,Stereotypy,Rigidity,Negativism,Waxy flexibility,Withdrawal | 99.8 | 99.3 | 100.0 |
| 10 | Excitement,Immobility/stupor,Staring,Posturing/catalepsy,Echopraxia/echolalia,Stereotypy,Verbigeration,Rigidity,Negativism,Withdrawal | 99.8 | 99.3 | 100.0 |
| 10 | Excitement,Immobility/stupor,Staring,Posturing/catalepsy,Echopraxia/echolalia,Verbigeration,Rigidity,Negativism,Waxy flexibility,Withdrawal | 99.8 | 99.3 | 100.0 |
| 10 | Excitement,Immobility/stupor,Staring,Posturing/catalepsy,Grimacing,Echopraxia/echolalia,Mannerisms,Negativism,Waxy flexibility,Withdrawal | 99.8 | 99.3 | 100.0 |
| 10 | Excitement,Immobility/stupor,Staring,Posturing/catalepsy,Grimacing,Echopraxia/echolalia,Mannerisms,Rigidity,Waxy flexibility,Withdrawal | 99.8 | 99.3 | 100.0 |
| 10 | Excitement,Immobility/stupor,Staring,Posturing/catalepsy,Grimacing,Echopraxia/echolalia,Mannerisms,Verbigeration,Negativism,Withdrawal | 99.8 | 99.3 | 100.0 |
| 10 | Excitement,Immobility/stupor,Staring,Posturing/catalepsy,Grimacing,Echopraxia/echolalia,Mannerisms,Verbigeration,Rigidity,Negativism | 99.8 | 99.3 | 100.0 |
| 10 | Excitement,Immobility/stupor,Staring,Posturing/catalepsy,Grimacing,Echopraxia/echolalia,Mannerisms,Verbigeration,Rigidity,Withdrawal | 99.8 | 99.3 | 100.0 |
| 10 | Excitement,Immobility/stupor,Staring,Posturing/catalepsy,Grimacing,Echopraxia/echolalia,Rigidity,Negativism,Waxy flexibility,Withdrawal | 99.8 | 99.3 | 100.0 |
| 10 | Excitement,Immobility/stupor,Staring,Posturing/catalepsy,Grimacing,Echopraxia/echolalia,Stereotypy,Mannerisms,Negativism,Withdrawal | 99.8 | 99.3 | 100.0 |
| 10 | Excitement,Immobility/stupor,Staring,Posturing/catalepsy,Grimacing,Echopraxia/echolalia,Stereotypy,Mannerisms,Rigidity,Withdrawal | 99.8 | 99.3 | 100.0 |
| 10 | Excitement,Immobility/stupor,Staring,Posturing/catalepsy,Grimacing,Echopraxia/echolalia,Stereotypy,Rigidity,Negativism,Withdrawal | 99.8 | 99.3 | 100.0 |
| 10 | Excitement,Immobility/stupor,Staring,Posturing/catalepsy,Grimacing,Echopraxia/echolalia,Verbigeration,Rigidity,Negativism,Withdrawal | 99.8 | 99.3 | 100.0 |
| 10 | Excitement,Immobility/stupor,Staring,Posturing/catalepsy,Grimacing,Mannerisms,Rigidity,Negativism,Waxy flexibility,Withdrawal | 99.8 | 99.3 | 100.0 |
| 10 | Excitement,Immobility/stupor,Staring,Posturing/catalepsy,Grimacing,Mannerisms,Verbigeration,Negativism,Waxy flexibility,Withdrawal | 99.8 | 99.3 | 100.0 |
| 10 | Excitement,Immobility/stupor,Staring,Posturing/catalepsy,Grimacing,Mannerisms,Verbigeration,Rigidity,Negativism,Waxy flexibility | 99.8 | 99.3 | 100.0 |
| 10 | Excitement,Immobility/stupor,Staring,Posturing/catalepsy,Grimacing,Mannerisms,Verbigeration,Rigidity,Waxy flexibility,Withdrawal | 99.8 | 99.3 | 100.0 |
| 10 | Excitement,Immobility/stupor,Staring,Posturing/catalepsy,Grimacing,Stereotypy,Mannerisms,Rigidity,Negativism,Withdrawal | 99.8 | 99.3 | 100.0 |
| 10 | Excitement,Immobility/stupor,Staring,Posturing/catalepsy,Grimacing,Stereotypy,Mannerisms,Verbigeration,Negativism,Withdrawal | 99.8 | 99.3 | 100.0 |
| 10 | Excitement,Immobility/stupor,Staring,Posturing/catalepsy,Grimacing,Stereotypy,Mannerisms,Verbigeration,Rigidity,Negativism | 99.8 | 99.3 | 100.0 |
| 10 | Excitement,Immobility/stupor,Staring,Posturing/catalepsy,Grimacing,Stereotypy,Mannerisms,Verbigeration,Rigidity,Withdrawal | 99.8 | 99.3 | 100.0 |
| 10 | Excitement,Immobility/stupor,Staring,Posturing/catalepsy,Grimacing,Stereotypy,Verbigeration,Rigidity,Negativism,Withdrawal | 99.8 | 99.3 | 100.0 |
| 10 | Excitement,Immobility/stupor,Staring,Posturing/catalepsy,Stereotypy,Mannerisms,Rigidity,Negativism,Waxy flexibility,Withdrawal | 99.8 | 99.3 | 100.0 |
| 10 | Excitement,Immobility/stupor,Staring,Posturing/catalepsy,Stereotypy,Mannerisms,Verbigeration,Negativism,Waxy flexibility,Withdrawal | 99.8 | 99.3 | 100.0 |
| 10 | Excitement,Immobility/stupor,Staring,Posturing/catalepsy,Stereotypy,Mannerisms,Verbigeration,Rigidity,Negativism,Waxy flexibility | 99.8 | 99.3 | 100.0 |
| 10 | Excitement,Immobility/stupor,Staring,Posturing/catalepsy,Stereotypy,Mannerisms,Verbigeration,Rigidity,Waxy flexibility,Withdrawal | 99.8 | 99.3 | 100.0 |
| 10 | Excitement,Immobility/stupor,Staring,Posturing/catalepsy,Stereotypy,Verbigeration,Rigidity,Negativism,Waxy flexibility,Withdrawal | 99.8 | 99.3 | 100.0 |
| 10 | Excitement,Mutism,Staring,Echopraxia/echolalia,Mannerisms,Verbigeration,Rigidity,Negativism,Waxy flexibility,Withdrawal | 99.8 | 99.3 | 100.0 |
| 10 | Excitement,Mutism,Staring,Grimacing,Echopraxia/echolalia,Mannerisms,Rigidity,Negativism,Waxy flexibility,Withdrawal | 99.8 | 99.3 | 100.0 |
| 10 | Excitement,Mutism,Staring,Grimacing,Echopraxia/echolalia,Mannerisms,Verbigeration,Rigidity,Negativism,Waxy flexibility | 99.8 | 99.3 | 100.0 |
| 10 | Excitement,Mutism,Staring,Grimacing,Echopraxia/echolalia,Mannerisms,Verbigeration,Rigidity,Negativism,Withdrawal | 99.8 | 99.3 | 100.0 |
| 10 | Excitement,Mutism,Staring,Grimacing,Echopraxia/echolalia,Mannerisms,Verbigeration,Rigidity,Waxy flexibility,Withdrawal | 99.8 | 99.3 | 100.0 |
| 10 | Excitement,Mutism,Staring,Grimacing,Echopraxia/echolalia,Stereotypy,Mannerisms,Rigidity,Negativism,Waxy flexibility | 99.8 | 99.3 | 100.0 |
| 10 | Excitement,Mutism,Staring,Grimacing,Echopraxia/echolalia,Stereotypy,Mannerisms,Verbigeration,Rigidity,Negativism | 99.8 | 99.3 | 100.0 |
| 10 | Excitement,Mutism,Staring,Grimacing,Echopraxia/echolalia,Stereotypy,Mannerisms,Verbigeration,Rigidity,Waxy flexibility | 99.8 | 99.3 | 100.0 |
| 10 | Excitement,Mutism,Staring,Grimacing,Echopraxia/echolalia,Stereotypy,Mannerisms,Verbigeration,Rigidity,Withdrawal | 99.8 | 99.3 | 100.0 |
| 10 | Excitement,Mutism,Staring,Grimacing,Echopraxia/echolalia,Verbigeration,Rigidity,Negativism,Waxy flexibility,Withdrawal | 99.8 | 99.3 | 100.0 |
| 10 | Excitement,Mutism,Staring,Grimacing,Mannerisms,Verbigeration,Rigidity,Negativism,Waxy flexibility,Withdrawal | 99.8 | 99.3 | 100.0 |
| 10 | Excitement,Mutism,Staring,Grimacing,Stereotypy,Mannerisms,Rigidity,Negativism,Waxy flexibility,Withdrawal | 99.8 | 99.3 | 100.0 |
| 10 | Excitement,Mutism,Staring,Grimacing,Stereotypy,Mannerisms,Verbigeration,Rigidity,Negativism,Waxy flexibility | 99.8 | 99.3 | 100.0 |
| 10 | Excitement,Mutism,Staring,Posturing/catalepsy,Echopraxia/echolalia,Stereotypy,Mannerisms,Rigidity,Negativism,Waxy flexibility | 99.8 | 99.3 | 100.0 |
| 10 | Excitement,Mutism,Staring,Posturing/catalepsy,Echopraxia/echolalia,Stereotypy,Mannerisms,Verbigeration,Rigidity,Waxy flexibility | 99.8 | 99.3 | 100.0 |
| 10 | Excitement,Mutism,Staring,Posturing/catalepsy,Grimacing,Echopraxia/echolalia,Mannerisms,Rigidity,Negativism,Waxy flexibility | 99.8 | 99.3 | 100.0 |
| 10 | Excitement,Mutism,Staring,Posturing/catalepsy,Grimacing,Echopraxia/echolalia,Mannerisms,Verbigeration,Rigidity,Waxy flexibility | 99.8 | 99.3 | 100.0 |
| 10 | Excitement,Mutism,Staring,Posturing/catalepsy,Grimacing,Echopraxia/echolalia,Stereotypy,Mannerisms,Negativism,Waxy flexibility | 99.8 | 99.3 | 100.0 |
| 10 | Excitement,Mutism,Staring,Posturing/catalepsy,Grimacing,Echopraxia/echolalia,Stereotypy,Mannerisms,Rigidity,Negativism | 99.8 | 99.3 | 100.0 |
| 10 | Excitement,Mutism,Staring,Posturing/catalepsy,Grimacing,Echopraxia/echolalia,Stereotypy,Mannerisms,Verbigeration,Negativism | 99.8 | 99.3 | 100.0 |
| 10 | Excitement,Mutism,Staring,Posturing/catalepsy,Grimacing,Echopraxia/echolalia,Stereotypy,Mannerisms,Verbigeration,Rigidity | 99.8 | 99.3 | 100.0 |
| 10 | Excitement,Mutism,Staring,Posturing/catalepsy,Grimacing,Echopraxia/echolalia,Stereotypy,Rigidity,Negativism,Waxy flexibility | 99.8 | 99.3 | 100.0 |
| 10 | Excitement,Mutism,Staring,Posturing/catalepsy,Grimacing,Echopraxia/echolalia,Stereotypy,Verbigeration,Rigidity,Waxy flexibility | 99.8 | 99.3 | 100.0 |
| 10 | Excitement,Mutism,Staring,Posturing/catalepsy,Grimacing,Mannerisms,Rigidity,Negativism,Waxy flexibility,Withdrawal | 99.8 | 99.3 | 100.0 |
| 10 | Excitement,Mutism,Staring,Posturing/catalepsy,Grimacing,Mannerisms,Verbigeration,Rigidity,Negativism,Waxy flexibility | 99.8 | 99.3 | 100.0 |
| 10 | Excitement,Mutism,Staring,Posturing/catalepsy,Grimacing,Stereotypy,Mannerisms,Negativism,Waxy flexibility,Withdrawal | 99.8 | 99.3 | 100.0 |
| 10 | Excitement,Mutism,Staring,Posturing/catalepsy,Grimacing,Stereotypy,Mannerisms,Rigidity,Negativism,Withdrawal | 99.8 | 99.3 | 100.0 |
| 10 | Excitement,Mutism,Staring,Posturing/catalepsy,Grimacing,Stereotypy,Mannerisms,Rigidity,Waxy flexibility,Withdrawal | 99.8 | 99.3 | 100.0 |
| 10 | Excitement,Mutism,Staring,Posturing/catalepsy,Grimacing,Stereotypy,Mannerisms,Verbigeration,Negativism,Waxy flexibility | 99.8 | 99.3 | 100.0 |
| 10 | Excitement,Mutism,Staring,Posturing/catalepsy,Grimacing,Stereotypy,Mannerisms,Verbigeration,Rigidity,Negativism | 99.8 | 99.3 | 100.0 |
| 10 | Excitement,Mutism,Staring,Posturing/catalepsy,Grimacing,Stereotypy,Rigidity,Negativism,Waxy flexibility,Withdrawal | 99.8 | 99.3 | 100.0 |
| 10 | Excitement,Mutism,Staring,Posturing/catalepsy,Grimacing,Stereotypy,Verbigeration,Rigidity,Negativism,Waxy flexibility | 99.8 | 99.3 | 100.0 |
| 10 | Excitement,Mutism,Staring,Posturing/catalepsy,Grimacing,Verbigeration,Rigidity,Negativism,Waxy flexibility,Withdrawal | 99.8 | 99.3 | 100.0 |
| 10 | Excitement,Mutism,Staring,Posturing/catalepsy,Stereotypy,Mannerisms,Rigidity,Negativism,Waxy flexibility,Withdrawal | 99.8 | 99.3 | 100.0 |
| 10 | Excitement,Mutism,Staring,Posturing/catalepsy,Stereotypy,Mannerisms,Verbigeration,Rigidity,Negativism,Waxy flexibility | 99.8 | 99.3 | 100.0 |
| 10 | Immobility/stupor,Mutism,Staring,Echopraxia/echolalia,Stereotypy,Mannerisms,Rigidity,Negativism,Waxy flexibility,Withdrawal | 99.8 | 99.3 | 100.0 |
| 10 | Immobility/stupor,Mutism,Staring,Echopraxia/echolalia,Stereotypy,Mannerisms,Verbigeration,Rigidity,Negativism,Withdrawal | 99.8 | 99.3 | 100.0 |
| 10 | Immobility/stupor,Mutism,Staring,Grimacing,Stereotypy,Mannerisms,Rigidity,Negativism,Waxy flexibility,Withdrawal | 99.8 | 99.3 | 100.0 |
| 10 | Immobility/stupor,Mutism,Staring,Grimacing,Stereotypy,Mannerisms,Verbigeration,Rigidity,Negativism,Waxy flexibility | 99.8 | 99.3 | 100.0 |
| 10 | Immobility/stupor,Mutism,Staring,Posturing/catalepsy,Echopraxia/echolalia,Stereotypy,Mannerisms,Rigidity,Negativism,Withdrawal | 99.8 | 99.3 | 100.0 |
| 10 | Immobility/stupor,Mutism,Staring,Posturing/catalepsy,Grimacing,Echopraxia/echolalia,Mannerisms,Negativism,Waxy flexibility,Withdrawal | 99.8 | 99.3 | 100.0 |
| 10 | Immobility/stupor,Mutism,Staring,Posturing/catalepsy,Grimacing,Echopraxia/echolalia,Mannerisms,Rigidity,Negativism,Withdrawal | 99.8 | 99.3 | 100.0 |
| 10 | Immobility/stupor,Mutism,Staring,Posturing/catalepsy,Grimacing,Echopraxia/echolalia,Mannerisms,Verbigeration,Negativism,Withdrawal | 99.8 | 99.3 | 100.0 |
| 10 | Immobility/stupor,Mutism,Staring,Posturing/catalepsy,Grimacing,Stereotypy,Mannerisms,Negativism,Waxy flexibility,Withdrawal | 99.8 | 99.3 | 100.0 |
| 10 | Immobility/stupor,Mutism,Staring,Posturing/catalepsy,Grimacing,Stereotypy,Mannerisms,Rigidity,Negativism,Withdrawal | 99.8 | 99.3 | 100.0 |
| 10 | Immobility/stupor,Mutism,Staring,Posturing/catalepsy,Grimacing,Stereotypy,Mannerisms,Verbigeration,Rigidity,Negativism | 99.8 | 99.3 | 100.0 |
| 10 | Immobility/stupor,Mutism,Staring,Posturing/catalepsy,Stereotypy,Mannerisms,Verbigeration,Rigidity,Negativism,Withdrawal | 99.8 | 99.3 | 100.0 |
| 10 | Immobility/stupor,Mutism,Staring,Stereotypy,Mannerisms,Verbigeration,Rigidity,Negativism,Waxy flexibility,Withdrawal | 99.8 | 99.3 | 100.0 |
| 10 | Immobility/stupor,Staring,Echopraxia/echolalia,Stereotypy,Mannerisms,Verbigeration,Rigidity,Negativism,Waxy flexibility,Withdrawal | 99.8 | 99.3 | 100.0 |
| 10 | Immobility/stupor,Staring,Posturing/catalepsy,Echopraxia/echolalia,Stereotypy,Mannerisms,Rigidity,Negativism,Waxy flexibility,Withdrawal | 99.8 | 99.3 | 100.0 |
| 10 | Immobility/stupor,Staring,Posturing/catalepsy,Echopraxia/echolalia,Stereotypy,Mannerisms,Verbigeration,Rigidity,Negativism,Withdrawal | 99.8 | 99.3 | 100.0 |
| 10 | Immobility/stupor,Staring,Posturing/catalepsy,Grimacing,Echopraxia/echolalia,Mannerisms,Rigidity,Negativism,Waxy flexibility,Withdrawal | 99.8 | 99.3 | 100.0 |
| 10 | Immobility/stupor,Staring,Posturing/catalepsy,Grimacing,Echopraxia/echolalia,Mannerisms,Verbigeration,Rigidity,Negativism,Withdrawal | 99.8 | 99.3 | 100.0 |
| 10 | Immobility/stupor,Staring,Posturing/catalepsy,Grimacing,Echopraxia/echolalia,Stereotypy,Mannerisms,Negativism,Waxy flexibility,Withdrawal | 99.8 | 99.3 | 100.0 |
| 10 | Immobility/stupor,Staring,Posturing/catalepsy,Grimacing,Echopraxia/echolalia,Stereotypy,Mannerisms,Verbigeration,Negativism,Withdrawal | 99.8 | 99.3 | 100.0 |
| 10 | Immobility/stupor,Staring,Posturing/catalepsy,Grimacing,Stereotypy,Mannerisms,Rigidity,Negativism,Waxy flexibility,Withdrawal | 99.8 | 99.3 | 100.0 |
| 10 | Immobility/stupor,Staring,Posturing/catalepsy,Grimacing,Stereotypy,Mannerisms,Verbigeration,Negativism,Waxy flexibility,Withdrawal | 99.8 | 99.3 | 100.0 |
| 10 | Immobility/stupor,Staring,Posturing/catalepsy,Stereotypy,Mannerisms,Verbigeration,Rigidity,Negativism,Waxy flexibility,Withdrawal | 99.8 | 99.3 | 100.0 |
| 10 | Mutism,Staring,Echopraxia/echolalia,Stereotypy,Mannerisms,Verbigeration,Rigidity,Negativism,Waxy flexibility,Withdrawal | 99.8 | 99.3 | 100.0 |
| 10 | Mutism,Staring,Grimacing,Echopraxia/echolalia,Stereotypy,Mannerisms,Verbigeration,Rigidity,Negativism,Waxy flexibility | 99.8 | 99.3 | 100.0 |
| 10 | Mutism,Staring,Posturing/catalepsy,Echopraxia/echolalia,Stereotypy,Mannerisms,Rigidity,Negativism,Waxy flexibility,Withdrawal | 99.8 | 99.3 | 100.0 |
| 10 | Mutism,Staring,Posturing/catalepsy,Echopraxia/echolalia,Stereotypy,Mannerisms,Verbigeration,Rigidity,Negativism,Withdrawal | 99.8 | 99.3 | 100.0 |
| 10 | Mutism,Staring,Posturing/catalepsy,Grimacing,Echopraxia/echolalia,Mannerisms,Rigidity,Negativism,Waxy flexibility,Withdrawal | 99.8 | 99.3 | 100.0 |
| 10 | Mutism,Staring,Posturing/catalepsy,Grimacing,Echopraxia/echolalia,Mannerisms,Verbigeration,Negativism,Waxy flexibility,Withdrawal | 99.8 | 99.3 | 100.0 |
| 10 | Mutism,Staring,Posturing/catalepsy,Grimacing,Echopraxia/echolalia,Mannerisms,Verbigeration,Rigidity,Negativism,Withdrawal | 99.8 | 99.3 | 100.0 |
| 10 | Mutism,Staring,Posturing/catalepsy,Grimacing,Echopraxia/echolalia,Stereotypy,Mannerisms,Verbigeration,Negativism,Waxy flexibility | 99.8 | 99.3 | 100.0 |
| 10 | Mutism,Staring,Posturing/catalepsy,Grimacing,Echopraxia/echolalia,Stereotypy,Mannerisms,Verbigeration,Rigidity,Negativism | 99.8 | 99.3 | 100.0 |
| 10 | Mutism,Staring,Posturing/catalepsy,Grimacing,Stereotypy,Mannerisms,Rigidity,Negativism,Waxy flexibility,Withdrawal | 99.8 | 99.3 | 100.0 |
| 10 | Mutism,Staring,Posturing/catalepsy,Stereotypy,Mannerisms,Verbigeration,Rigidity,Negativism,Waxy flexibility,Withdrawal | 99.8 | 99.3 | 100.0 |
| 10 | Excitement,Immobility/stupor,Echopraxia/echolalia,Stereotypy,Mannerisms,Verbigeration,Rigidity,Negativism,Waxy flexibility,Withdrawal | 99.6 | 98.9 | 100.0 |
| 10 | Excitement,Immobility/stupor,Grimacing,Echopraxia/echolalia,Stereotypy,Mannerisms,Verbigeration,Rigidity,Negativism,Withdrawal | 99.6 | 98.9 | 100.0 |
| 10 | Excitement,Immobility/stupor,Mutism,Echopraxia/echolalia,Stereotypy,Mannerisms,Verbigeration,Rigidity,Negativism,Waxy flexibility | 99.6 | 98.9 | 100.0 |
| 10 | Excitement,Immobility/stupor,Mutism,Grimacing,Echopraxia/echolalia,Mannerisms,Verbigeration,Rigidity,Negativism,Withdrawal | 99.6 | 98.9 | 100.0 |
| 10 | Excitement,Immobility/stupor,Mutism,Grimacing,Echopraxia/echolalia,Stereotypy,Mannerisms,Rigidity,Negativism,Withdrawal | 99.6 | 98.9 | 100.0 |
| 10 | Excitement,Immobility/stupor,Mutism,Grimacing,Echopraxia/echolalia,Stereotypy,Rigidity,Negativism,Waxy flexibility,Withdrawal | 99.6 | 98.9 | 100.0 |
| 10 | Excitement,Immobility/stupor,Mutism,Grimacing,Echopraxia/echolalia,Stereotypy,Verbigeration,Rigidity,Negativism,Waxy flexibility | 99.6 | 98.9 | 100.0 |
| 10 | Excitement,Immobility/stupor,Mutism,Grimacing,Echopraxia/echolalia,Verbigeration,Rigidity,Negativism,Waxy flexibility,Withdrawal | 99.6 | 98.9 | 100.0 |
| 10 | Excitement,Immobility/stupor,Mutism,Grimacing,Stereotypy,Verbigeration,Rigidity,Negativism,Waxy flexibility,Withdrawal | 99.6 | 98.9 | 100.0 |
| 10 | Excitement,Immobility/stupor,Mutism,Posturing/catalepsy,Echopraxia/echolalia,Mannerisms,Verbigeration,Rigidity,Negativism,Waxy flexibility | 99.6 | 98.9 | 100.0 |
| 10 | Excitement,Immobility/stupor,Mutism,Posturing/catalepsy,Echopraxia/echolalia,Stereotypy,Mannerisms,Rigidity,Negativism,Withdrawal | 99.6 | 98.9 | 100.0 |
| 10 | Excitement,Immobility/stupor,Mutism,Posturing/catalepsy,Echopraxia/echolalia,Stereotypy,Rigidity,Negativism,Waxy flexibility,Withdrawal | 99.6 | 98.9 | 100.0 |
| 10 | Excitement,Immobility/stupor,Mutism,Posturing/catalepsy,Echopraxia/echolalia,Stereotypy,Verbigeration,Rigidity,Waxy flexibility,Withdrawal | 99.6 | 98.9 | 100.0 |
| 10 | Excitement,Immobility/stupor,Mutism,Posturing/catalepsy,Grimacing,Echopraxia/echolalia,Mannerisms,Verbigeration,Rigidity,Withdrawal | 99.6 | 98.9 | 100.0 |
| 10 | Excitement,Immobility/stupor,Mutism,Posturing/catalepsy,Grimacing,Echopraxia/echolalia,Stereotypy,Verbigeration,Rigidity,Withdrawal | 99.6 | 98.9 | 100.0 |
| 10 | Excitement,Immobility/stupor,Mutism,Posturing/catalepsy,Grimacing,Echopraxia/echolalia,Verbigeration,Rigidity,Waxy flexibility,Withdrawal | 99.6 | 98.9 | 100.0 |
| 10 | Excitement,Immobility/stupor,Mutism,Posturing/catalepsy,Grimacing,Mannerisms,Verbigeration,Rigidity,Negativism,Waxy flexibility | 99.6 | 98.9 | 100.0 |
| 10 | Excitement,Immobility/stupor,Mutism,Posturing/catalepsy,Grimacing,Stereotypy,Mannerisms,Verbigeration,Rigidity,Negativism | 99.6 | 98.9 | 100.0 |
| 10 | Excitement,Immobility/stupor,Mutism,Posturing/catalepsy,Grimacing,Stereotypy,Verbigeration,Rigidity,Negativism,Waxy flexibility | 99.6 | 98.9 | 100.0 |
| 10 | Excitement,Immobility/stupor,Mutism,Posturing/catalepsy,Grimacing,Verbigeration,Rigidity,Negativism,Waxy flexibility,Withdrawal | 99.6 | 98.9 | 100.0 |
| 10 | Excitement,Immobility/stupor,Mutism,Posturing/catalepsy,Mannerisms,Verbigeration,Rigidity,Negativism,Waxy flexibility,Withdrawal | 99.6 | 98.9 | 100.0 |
| 10 | Excitement,Immobility/stupor,Mutism,Staring,Echopraxia/echolalia,Stereotypy,Mannerisms,Negativism,Waxy flexibility,Withdrawal | 99.6 | 98.9 | 100.0 |
| 10 | Excitement,Immobility/stupor,Mutism,Staring,Grimacing,Echopraxia/echolalia,Mannerisms,Verbigeration,Rigidity,Negativism | 99.6 | 98.9 | 100.0 |
| 10 | Excitement,Immobility/stupor,Mutism,Staring,Grimacing,Echopraxia/echolalia,Mannerisms,Verbigeration,Rigidity,Waxy flexibility | 99.6 | 98.9 | 100.0 |
| 10 | Excitement,Immobility/stupor,Mutism,Staring,Grimacing,Echopraxia/echolalia,Mannerisms,Verbigeration,Rigidity,Withdrawal | 99.6 | 98.9 | 100.0 |
| 10 | Excitement,Immobility/stupor,Mutism,Staring,Grimacing,Echopraxia/echolalia,Stereotypy,Mannerisms,Negativism,Withdrawal | 99.6 | 98.9 | 100.0 |
| 10 | Excitement,Immobility/stupor,Mutism,Staring,Grimacing,Echopraxia/echolalia,Stereotypy,Mannerisms,Rigidity,Negativism | 99.6 | 98.9 | 100.0 |
| 10 | Excitement,Immobility/stupor,Mutism,Staring,Grimacing,Echopraxia/echolalia,Stereotypy,Negativism,Waxy flexibility,Withdrawal | 99.6 | 98.9 | 100.0 |
| 10 | Excitement,Immobility/stupor,Mutism,Staring,Grimacing,Echopraxia/echolalia,Stereotypy,Verbigeration,Negativism,Withdrawal | 99.6 | 98.9 | 100.0 |
| 10 | Excitement,Immobility/stupor,Mutism,Staring,Grimacing,Mannerisms,Rigidity,Negativism,Waxy flexibility,Withdrawal | 99.6 | 98.9 | 100.0 |
| 10 | Excitement,Immobility/stupor,Mutism,Staring,Grimacing,Stereotypy,Mannerisms,Rigidity,Negativism,Waxy flexibility | 99.6 | 98.9 | 100.0 |
| 10 | Excitement,Immobility/stupor,Mutism,Staring,Grimacing,Stereotypy,Mannerisms,Rigidity,Negativism,Withdrawal | 99.6 | 98.9 | 100.0 |
| 10 | Excitement,Immobility/stupor,Mutism,Staring,Grimacing,Verbigeration,Rigidity,Negativism,Waxy flexibility,Withdrawal | 99.6 | 98.9 | 100.0 |
| 10 | Excitement,Immobility/stupor,Mutism,Staring,Posturing/catalepsy,Echopraxia/echolalia,Stereotypy,Mannerisms,Rigidity,Waxy flexibility | 99.6 | 98.9 | 100.0 |
| 10 | Excitement,Immobility/stupor,Mutism,Staring,Posturing/catalepsy,Grimacing,Echopraxia/echolalia,Mannerisms,Rigidity,Waxy flexibility | 99.6 | 98.9 | 100.0 |
| 10 | Excitement,Immobility/stupor,Mutism,Staring,Posturing/catalepsy,Grimacing,Echopraxia/echolalia,Mannerisms,Waxy flexibility,Withdrawal | 99.6 | 98.9 | 100.0 |
| 10 | Excitement,Immobility/stupor,Mutism,Staring,Posturing/catalepsy,Grimacing,Echopraxia/echolalia,Stereotypy,Mannerisms,Negativism | 99.6 | 98.9 | 100.0 |
| 10 | Excitement,Immobility/stupor,Mutism,Staring,Posturing/catalepsy,Grimacing,Echopraxia/echolalia,Stereotypy,Rigidity,Waxy flexibility | 99.6 | 98.9 | 100.0 |
| 10 | Excitement,Immobility/stupor,Mutism,Staring,Posturing/catalepsy,Grimacing,Echopraxia/echolalia,Stereotypy,Waxy flexibility,Withdrawal | 99.6 | 98.9 | 100.0 |
| 10 | Excitement,Immobility/stupor,Mutism,Staring,Posturing/catalepsy,Grimacing,Echopraxia/echolalia,Verbigeration,Waxy flexibility,Withdrawal | 99.6 | 98.9 | 100.0 |
| 10 | Excitement,Immobility/stupor,Mutism,Staring,Posturing/catalepsy,Grimacing,Mannerisms,Rigidity,Negativism,Waxy flexibility | 99.6 | 98.9 | 100.0 |
| 10 | Excitement,Immobility/stupor,Mutism,Staring,Posturing/catalepsy,Grimacing,Rigidity,Negativism,Waxy flexibility,Withdrawal | 99.6 | 98.9 | 100.0 |
| 10 | Excitement,Immobility/stupor,Mutism,Staring,Posturing/catalepsy,Grimacing,Stereotypy,Mannerisms,Negativism,Waxy flexibility | 99.6 | 98.9 | 100.0 |
| 10 | Excitement,Immobility/stupor,Mutism,Staring,Posturing/catalepsy,Grimacing,Stereotypy,Mannerisms,Rigidity,Negativism | 99.6 | 98.9 | 100.0 |
| 10 | Excitement,Immobility/stupor,Mutism,Staring,Posturing/catalepsy,Stereotypy,Mannerisms,Rigidity,Negativism,Waxy flexibility | 99.6 | 98.9 | 100.0 |
| 10 | Excitement,Immobility/stupor,Mutism,Stereotypy,Mannerisms,Verbigeration,Rigidity,Negativism,Waxy flexibility,Withdrawal | 99.6 | 98.9 | 100.0 |
| 10 | Excitement,Immobility/stupor,Posturing/catalepsy,Echopraxia/echolalia,Stereotypy,Verbigeration,Rigidity,Negativism,Waxy flexibility,Withdrawal | 99.6 | 98.9 | 100.0 |
| 10 | Excitement,Immobility/stupor,Posturing/catalepsy,Grimacing,Mannerisms,Verbigeration,Rigidity,Negativism,Waxy flexibility,Withdrawal | 99.6 | 98.9 | 100.0 |
| 10 | Excitement,Immobility/stupor,Posturing/catalepsy,Grimacing,Stereotypy,Mannerisms,Verbigeration,Rigidity,Negativism,Withdrawal | 99.6 | 98.9 | 100.0 |
| 10 | Excitement,Immobility/stupor,Staring,Grimacing,Echopraxia/echolalia,Mannerisms,Verbigeration,Rigidity,Negativism,Waxy flexibility | 99.6 | 98.9 | 100.0 |
| 10 | Excitement,Immobility/stupor,Staring,Grimacing,Echopraxia/echolalia,Mannerisms,Verbigeration,Rigidity,Waxy flexibility,Withdrawal | 99.6 | 98.9 | 100.0 |
| 10 | Excitement,Immobility/stupor,Staring,Grimacing,Echopraxia/echolalia,Stereotypy,Mannerisms,Rigidity,Negativism,Waxy flexibility | 99.6 | 98.9 | 100.0 |
| 10 | Excitement,Immobility/stupor,Staring,Grimacing,Echopraxia/echolalia,Stereotypy,Mannerisms,Verbigeration,Rigidity,Negativism | 99.6 | 98.9 | 100.0 |
| 10 | Excitement,Immobility/stupor,Staring,Grimacing,Echopraxia/echolalia,Stereotypy,Verbigeration,Rigidity,Waxy flexibility,Withdrawal | 99.6 | 98.9 | 100.0 |
| 10 | Excitement,Immobility/stupor,Staring,Posturing/catalepsy,Echopraxia/echolalia,Stereotypy,Mannerisms,Rigidity,Negativism,Waxy flexibility | 99.6 | 98.9 | 100.0 |
| 10 | Excitement,Immobility/stupor,Staring,Posturing/catalepsy,Echopraxia/echolalia,Stereotypy,Mannerisms,Verbigeration,Negativism,Waxy flexibility | 99.6 | 98.9 | 100.0 |
| 10 | Excitement,Immobility/stupor,Staring,Posturing/catalepsy,Echopraxia/echolalia,Stereotypy,Verbigeration,Rigidity,Negativism,Waxy flexibility | 99.6 | 98.9 | 100.0 |
| 10 | Excitement,Immobility/stupor,Staring,Posturing/catalepsy,Grimacing,Echopraxia/echolalia,Stereotypy,Mannerisms,Rigidity,Negativism | 99.6 | 98.9 | 100.0 |
| 10 | Excitement,Immobility/stupor,Staring,Posturing/catalepsy,Grimacing,Echopraxia/echolalia,Stereotypy,Rigidity,Waxy flexibility,Withdrawal | 99.6 | 98.9 | 100.0 |
| 10 | Excitement,Immobility/stupor,Staring,Posturing/catalepsy,Grimacing,Echopraxia/echolalia,Verbigeration,Negativism,Waxy flexibility,Withdrawal | 99.6 | 98.9 | 100.0 |
| 10 | Excitement,Immobility/stupor,Staring,Posturing/catalepsy,Grimacing,Echopraxia/echolalia,Verbigeration,Rigidity,Negativism,Waxy flexibility | 99.6 | 98.9 | 100.0 |
| 10 | Excitement,Immobility/stupor,Staring,Posturing/catalepsy,Grimacing,Echopraxia/echolalia,Verbigeration,Rigidity,Waxy flexibility,Withdrawal | 99.6 | 98.9 | 100.0 |
| 10 | Excitement,Immobility/stupor,Staring,Posturing/catalepsy,Grimacing,Stereotypy,Mannerisms,Rigidity,Waxy flexibility,Withdrawal | 99.6 | 98.9 | 100.0 |
| 10 | Excitement,Immobility/stupor,Staring,Posturing/catalepsy,Grimacing,Stereotypy,Rigidity,Negativism,Waxy flexibility,Withdrawal | 99.6 | 98.9 | 100.0 |
| 10 | Excitement,Immobility/stupor,Staring,Posturing/catalepsy,Grimacing,Stereotypy,Verbigeration,Rigidity,Waxy flexibility,Withdrawal | 99.6 | 98.9 | 100.0 |
| 10 | Excitement,Immobility/stupor,Staring,Posturing/catalepsy,Grimacing,Verbigeration,Rigidity,Negativism,Waxy flexibility,Withdrawal | 99.6 | 98.9 | 100.0 |
| 10 | Excitement,Mutism,Staring,Echopraxia/echolalia,Stereotypy,Mannerisms,Verbigeration,Negativism,Waxy flexibility,Withdrawal | 99.6 | 98.9 | 100.0 |
| 10 | Excitement,Mutism,Staring,Grimacing,Echopraxia/echolalia,Stereotypy,Mannerisms,Negativism,Waxy flexibility,Withdrawal | 99.6 | 98.9 | 100.0 |
| 10 | Excitement,Mutism,Staring,Grimacing,Echopraxia/echolalia,Stereotypy,Mannerisms,Verbigeration,Negativism,Waxy flexibility | 99.6 | 98.9 | 100.0 |
| 10 | Excitement,Mutism,Staring,Grimacing,Echopraxia/echolalia,Stereotypy,Mannerisms,Verbigeration,Negativism,Withdrawal | 99.6 | 98.9 | 100.0 |
| 10 | Excitement,Mutism,Staring,Posturing/catalepsy,Grimacing,Echopraxia/echolalia,Stereotypy,Mannerisms,Rigidity,Waxy flexibility | 99.6 | 98.9 | 100.0 |
| 10 | Excitement,Mutism,Staring,Posturing/catalepsy,Grimacing,Echopraxia/echolalia,Stereotypy,Mannerisms,Waxy flexibility,Withdrawal | 99.6 | 98.9 | 100.0 |
| 10 | Excitement,Mutism,Staring,Posturing/catalepsy,Grimacing,Echopraxia/echolalia,Stereotypy,Verbigeration,Waxy flexibility,Withdrawal | 99.6 | 98.9 | 100.0 |
| 10 | Excitement,Mutism,Staring,Posturing/catalepsy,Grimacing,Stereotypy,Mannerisms,Verbigeration,Rigidity,Waxy flexibility | 99.6 | 98.9 | 100.0 |
| 10 | Excitement,Staring,Echopraxia/echolalia,Stereotypy,Mannerisms,Verbigeration,Rigidity,Negativism,Waxy flexibility,Withdrawal | 99.6 | 98.9 | 100.0 |
| 10 | Excitement,Staring,Grimacing,Stereotypy,Mannerisms,Verbigeration,Rigidity,Negativism,Waxy flexibility,Withdrawal | 99.6 | 98.9 | 100.0 |
| 10 | Excitement,Staring,Posturing/catalepsy,Echopraxia/echolalia,Mannerisms,Verbigeration,Rigidity,Negativism,Waxy flexibility,Withdrawal | 99.6 | 98.9 | 100.0 |
| 10 | Excitement,Staring,Posturing/catalepsy,Grimacing,Echopraxia/echolalia,Mannerisms,Verbigeration,Rigidity,Negativism,Withdrawal | 99.6 | 98.9 | 100.0 |
| 10 | Excitement,Staring,Posturing/catalepsy,Grimacing,Echopraxia/echolalia,Stereotypy,Mannerisms,Rigidity,Negativism,Withdrawal | 99.6 | 98.9 | 100.0 |
| 10 | Excitement,Staring,Posturing/catalepsy,Grimacing,Mannerisms,Verbigeration,Rigidity,Negativism,Waxy flexibility,Withdrawal | 99.6 | 98.9 | 100.0 |
| 10 | Excitement,Staring,Posturing/catalepsy,Grimacing,Stereotypy,Mannerisms,Verbigeration,Rigidity,Negativism,Withdrawal | 99.6 | 98.9 | 100.0 |
| 10 | Excitement,Staring,Posturing/catalepsy,Stereotypy,Mannerisms,Verbigeration,Rigidity,Negativism,Waxy flexibility,Withdrawal | 99.6 | 98.9 | 100.0 |
| 10 | Immobility/stupor,Mutism,Staring,Echopraxia/echolalia,Stereotypy,Mannerisms,Verbigeration,Rigidity,Negativism,Waxy flexibility | 99.6 | 98.9 | 100.0 |
| 10 | Immobility/stupor,Mutism,Staring,Grimacing,Echopraxia/echolalia,Mannerisms,Rigidity,Negativism,Waxy flexibility,Withdrawal | 99.6 | 98.9 | 100.0 |
| 10 | Immobility/stupor,Mutism,Staring,Grimacing,Echopraxia/echolalia,Mannerisms,Verbigeration,Rigidity,Negativism,Withdrawal | 99.6 | 98.9 | 100.0 |
| 10 | Immobility/stupor,Mutism,Staring,Grimacing,Echopraxia/echolalia,Stereotypy,Mannerisms,Negativism,Waxy flexibility,Withdrawal | 99.6 | 98.9 | 100.0 |
| 10 | Immobility/stupor,Mutism,Staring,Grimacing,Echopraxia/echolalia,Stereotypy,Mannerisms,Rigidity,Waxy flexibility,Withdrawal | 99.6 | 98.9 | 100.0 |
| 10 | Immobility/stupor,Mutism,Staring,Grimacing,Echopraxia/echolalia,Stereotypy,Mannerisms,Verbigeration,Negativism,Withdrawal | 99.6 | 98.9 | 100.0 |
| 10 | Immobility/stupor,Mutism,Staring,Grimacing,Echopraxia/echolalia,Stereotypy,Mannerisms,Verbigeration,Rigidity,Negativism | 99.6 | 98.9 | 100.0 |
| 10 | Immobility/stupor,Mutism,Staring,Posturing/catalepsy,Echopraxia/echolalia,Mannerisms,Rigidity,Negativism,Waxy flexibility,Withdrawal | 99.6 | 98.9 | 100.0 |
| 10 | Immobility/stupor,Mutism,Staring,Posturing/catalepsy,Echopraxia/echolalia,Mannerisms,Verbigeration,Rigidity,Negativism,Withdrawal | 99.6 | 98.9 | 100.0 |
| 10 | Immobility/stupor,Mutism,Staring,Posturing/catalepsy,Echopraxia/echolalia,Stereotypy,Mannerisms,Verbigeration,Negativism,Withdrawal | 99.6 | 98.9 | 100.0 |
| 10 | Immobility/stupor,Mutism,Staring,Posturing/catalepsy,Echopraxia/echolalia,Stereotypy,Mannerisms,Verbigeration,Rigidity,Negativism | 99.6 | 98.9 | 100.0 |
| 10 | Immobility/stupor,Mutism,Staring,Posturing/catalepsy,Grimacing,Echopraxia/echolalia,Mannerisms,Verbigeration,Rigidity,Negativism | 99.6 | 98.9 | 100.0 |
| 10 | Immobility/stupor,Mutism,Staring,Posturing/catalepsy,Grimacing,Echopraxia/echolalia,Stereotypy,Mannerisms,Negativism,Waxy flexibility | 99.6 | 98.9 | 100.0 |
| 10 | Immobility/stupor,Mutism,Staring,Posturing/catalepsy,Grimacing,Echopraxia/echolalia,Stereotypy,Mannerisms,Rigidity,Negativism | 99.6 | 98.9 | 100.0 |
| 10 | Immobility/stupor,Mutism,Staring,Posturing/catalepsy,Grimacing,Echopraxia/echolalia,Stereotypy,Mannerisms,Verbigeration,Negativism | 99.6 | 98.9 | 100.0 |
| 10 | Immobility/stupor,Mutism,Staring,Posturing/catalepsy,Grimacing,Mannerisms,Verbigeration,Negativism,Waxy flexibility,Withdrawal | 99.6 | 98.9 | 100.0 |
| 10 | Immobility/stupor,Mutism,Staring,Posturing/catalepsy,Grimacing,Mannerisms,Verbigeration,Rigidity,Negativism,Withdrawal | 99.6 | 98.9 | 100.0 |
| 10 | Immobility/stupor,Mutism,Staring,Posturing/catalepsy,Grimacing,Stereotypy,Mannerisms,Verbigeration,Rigidity,Withdrawal | 99.6 | 98.9 | 100.0 |
| 10 | Immobility/stupor,Mutism,Staring,Posturing/catalepsy,Stereotypy,Mannerisms,Rigidity,Negativism,Waxy flexibility,Withdrawal | 99.6 | 98.9 | 100.0 |
| 10 | Immobility/stupor,Mutism,Staring,Posturing/catalepsy,Stereotypy,Mannerisms,Verbigeration,Negativism,Waxy flexibility,Withdrawal | 99.6 | 98.9 | 100.0 |
| 10 | Immobility/stupor,Staring,Grimacing,Echopraxia/echolalia,Mannerisms,Verbigeration,Rigidity,Negativism,Waxy flexibility,Withdrawal | 99.6 | 98.9 | 100.0 |
| 10 | Immobility/stupor,Staring,Posturing/catalepsy,Echopraxia/echolalia,Mannerisms,Verbigeration,Rigidity,Negativism,Waxy flexibility,Withdrawal | 99.6 | 98.9 | 100.0 |
| 10 | Immobility/stupor,Staring,Posturing/catalepsy,Grimacing,Echopraxia/echolalia,Stereotypy,Mannerisms,Verbigeration,Rigidity,Negativism | 99.6 | 98.9 | 100.0 |
| 10 | Immobility/stupor,Staring,Posturing/catalepsy,Grimacing,Stereotypy,Mannerisms,Verbigeration,Rigidity,Negativism,Waxy flexibility | 99.6 | 98.9 | 100.0 |
| 10 | Mutism,Staring,Grimacing,Echopraxia/echolalia,Mannerisms,Verbigeration,Rigidity,Negativism,Waxy flexibility,Withdrawal | 99.6 | 98.9 | 100.0 |
| 10 | Mutism,Staring,Grimacing,Echopraxia/echolalia,Stereotypy,Mannerisms,Verbigeration,Negativism,Waxy flexibility,Withdrawal | 99.6 | 98.9 | 100.0 |
| 10 | Mutism,Staring,Posturing/catalepsy,Echopraxia/echolalia,Mannerisms,Verbigeration,Rigidity,Negativism,Waxy flexibility,Withdrawal | 99.6 | 98.9 | 100.0 |
| 10 | Mutism,Staring,Posturing/catalepsy,Echopraxia/echolalia,Stereotypy,Mannerisms,Verbigeration,Negativism,Waxy flexibility,Withdrawal | 99.6 | 98.9 | 100.0 |
| 10 | Mutism,Staring,Posturing/catalepsy,Echopraxia/echolalia,Stereotypy,Mannerisms,Verbigeration,Rigidity,Negativism,Waxy flexibility | 99.6 | 98.9 | 100.0 |
| 10 | Mutism,Staring,Posturing/catalepsy,Grimacing,Echopraxia/echolalia,Mannerisms,Verbigeration,Rigidity,Negativism,Waxy flexibility | 99.6 | 98.9 | 100.0 |
| 10 | Mutism,Staring,Posturing/catalepsy,Grimacing,Echopraxia/echolalia,Stereotypy,Mannerisms,Rigidity,Negativism,Waxy flexibility | 99.6 | 98.9 | 100.0 |
| 10 | Mutism,Staring,Posturing/catalepsy,Grimacing,Echopraxia/echolalia,Stereotypy,Mannerisms,Rigidity,Waxy flexibility,Withdrawal | 99.6 | 98.9 | 100.0 |
| 10 | Mutism,Staring,Posturing/catalepsy,Grimacing,Echopraxia/echolalia,Stereotypy,Mannerisms,Verbigeration,Rigidity,Withdrawal | 99.6 | 98.9 | 100.0 |
| 10 | Mutism,Staring,Posturing/catalepsy,Grimacing,Mannerisms,Verbigeration,Rigidity,Negativism,Waxy flexibility,Withdrawal | 99.6 | 98.9 | 100.0 |
| 10 | Mutism,Staring,Posturing/catalepsy,Grimacing,Stereotypy,Mannerisms,Verbigeration,Rigidity,Negativism,Waxy flexibility | 99.6 | 98.9 | 100.0 |
| 10 | Mutism,Staring,Posturing/catalepsy,Grimacing,Stereotypy,Mannerisms,Verbigeration,Rigidity,Waxy flexibility,Withdrawal | 99.6 | 98.9 | 100.0 |
| 10 | Staring,Posturing/catalepsy,Grimacing,Echopraxia/echolalia,Stereotypy,Mannerisms,Rigidity,Negativism,Waxy flexibility,Withdrawal | 99.6 | 98.9 | 100.0 |
| 10 | Staring,Posturing/catalepsy,Grimacing,Echopraxia/echolalia,Stereotypy,Mannerisms,Verbigeration,Rigidity,Negativism,Withdrawal | 99.6 | 98.9 | 100.0 |
| 10 | Staring,Posturing/catalepsy,Grimacing,Stereotypy,Mannerisms,Verbigeration,Rigidity,Negativism,Waxy flexibility,Withdrawal | 99.6 | 98.9 | 100.0 |
| 10 | Excitement,Immobility/stupor,Mutism,Grimacing,Stereotypy,Mannerisms,Verbigeration,Rigidity,Negativism,Withdrawal | 99.5 | 98.9 | 100.0 |
| 10 | Excitement,Immobility/stupor,Mutism,Posturing/catalepsy,Echopraxia/echolalia,Stereotypy,Mannerisms,Verbigeration,Rigidity,Withdrawal | 99.5 | 98.9 | 100.0 |
| 10 | Excitement,Immobility/stupor,Mutism,Posturing/catalepsy,Stereotypy,Mannerisms,Verbigeration,Rigidity,Negativism,Waxy flexibility | 99.5 | 98.9 | 100.0 |
| 10 | Excitement,Immobility/stupor,Mutism,Staring,Echopraxia/echolalia,Stereotypy,Mannerisms,Verbigeration,Negativism,Waxy flexibility | 99.5 | 98.9 | 100.0 |
| 10 | Excitement,Immobility/stupor,Mutism,Staring,Echopraxia/echolalia,Stereotypy,Mannerisms,Verbigeration,Negativism,Withdrawal | 99.5 | 98.9 | 100.0 |
| 10 | Excitement,Immobility/stupor,Mutism,Staring,Echopraxia/echolalia,Stereotypy,Verbigeration,Negativism,Waxy flexibility,Withdrawal | 99.5 | 98.9 | 100.0 |
| 10 | Excitement,Immobility/stupor,Mutism,Staring,Grimacing,Echopraxia/echolalia,Mannerisms,Rigidity,Negativism,Waxy flexibility | 99.5 | 98.9 | 100.0 |
| 10 | Excitement,Immobility/stupor,Mutism,Staring,Grimacing,Echopraxia/echolalia,Stereotypy,Mannerisms,Rigidity,Waxy flexibility | 99.5 | 98.9 | 100.0 |
| 10 | Excitement,Immobility/stupor,Mutism,Staring,Grimacing,Echopraxia/echolalia,Stereotypy,Verbigeration,Negativism,Waxy flexibility | 99.5 | 98.9 | 100.0 |
| 10 | Excitement,Immobility/stupor,Mutism,Staring,Posturing/catalepsy,Grimacing,Echopraxia/echolalia,Stereotypy,Mannerisms,Rigidity | 99.5 | 98.9 | 100.0 |
| 10 | Excitement,Immobility/stupor,Mutism,Staring,Posturing/catalepsy,Grimacing,Stereotypy,Rigidity,Negativism,Waxy flexibility | 99.5 | 98.9 | 100.0 |
| 10 | Excitement,Immobility/stupor,Posturing/catalepsy,Echopraxia/echolalia,Mannerisms,Verbigeration,Rigidity,Negativism,Waxy flexibility,Withdrawal | 99.5 | 98.9 | 100.0 |
| 10 | Excitement,Immobility/stupor,Posturing/catalepsy,Grimacing,Echopraxia/echolalia,Mannerisms,Rigidity,Negativism,Waxy flexibility,Withdrawal | 99.5 | 98.9 | 100.0 |
| 10 | Excitement,Immobility/stupor,Posturing/catalepsy,Grimacing,Echopraxia/echolalia,Stereotypy,Mannerisms,Rigidity,Negativism,Withdrawal | 99.5 | 98.9 | 100.0 |
| 10 | Excitement,Immobility/stupor,Posturing/catalepsy,Grimacing,Echopraxia/echolalia,Stereotypy,Verbigeration,Rigidity,Negativism,Withdrawal | 99.5 | 98.9 | 100.0 |
| 10 | Excitement,Immobility/stupor,Posturing/catalepsy,Grimacing,Echopraxia/echolalia,Verbigeration,Rigidity,Negativism,Waxy flexibility,Withdrawal | 99.5 | 98.9 | 100.0 |
| 10 | Excitement,Immobility/stupor,Posturing/catalepsy,Stereotypy,Mannerisms,Verbigeration,Rigidity,Negativism,Waxy flexibility,Withdrawal | 99.5 | 98.9 | 100.0 |
| 10 | Excitement,Immobility/stupor,Staring,Grimacing,Echopraxia/echolalia,Stereotypy,Mannerisms,Verbigeration,Rigidity,Withdrawal | 99.5 | 98.9 | 100.0 |
| 10 | Excitement,Immobility/stupor,Staring,Grimacing,Echopraxia/echolalia,Stereotypy,Verbigeration,Rigidity,Negativism,Waxy flexibility | 99.5 | 98.9 | 100.0 |
| 10 | Excitement,Immobility/stupor,Staring,Grimacing,Echopraxia/echolalia,Verbigeration,Rigidity,Negativism,Waxy flexibility,Withdrawal | 99.5 | 98.9 | 100.0 |
| 10 | Excitement,Immobility/stupor,Staring,Posturing/catalepsy,Echopraxia/echolalia,Stereotypy,Verbigeration,Negativism,Waxy flexibility,Withdrawal | 99.5 | 98.9 | 100.0 |
| 10 | Excitement,Immobility/stupor,Staring,Posturing/catalepsy,Echopraxia/echolalia,Stereotypy,Verbigeration,Rigidity,Waxy flexibility,Withdrawal | 99.5 | 98.9 | 100.0 |
| 10 | Excitement,Immobility/stupor,Staring,Posturing/catalepsy,Grimacing,Echopraxia/echolalia,Mannerisms,Rigidity,Negativism,Waxy flexibility | 99.5 | 98.9 | 100.0 |
| 10 | Excitement,Immobility/stupor,Staring,Posturing/catalepsy,Grimacing,Echopraxia/echolalia,Mannerisms,Verbigeration,Negativism,Waxy flexibility | 99.5 | 98.9 | 100.0 |
| 10 | Excitement,Immobility/stupor,Staring,Posturing/catalepsy,Grimacing,Echopraxia/echolalia,Stereotypy,Negativism,Waxy flexibility,Withdrawal | 99.5 | 98.9 | 100.0 |
| 10 | Excitement,Immobility/stupor,Staring,Posturing/catalepsy,Grimacing,Echopraxia/echolalia,Stereotypy,Verbigeration,Negativism,Withdrawal | 99.5 | 98.9 | 100.0 |
| 10 | Excitement,Immobility/stupor,Staring,Posturing/catalepsy,Grimacing,Echopraxia/echolalia,Stereotypy,Verbigeration,Rigidity,Negativism | 99.5 | 98.9 | 100.0 |
| 10 | Excitement,Immobility/stupor,Staring,Posturing/catalepsy,Grimacing,Echopraxia/echolalia,Stereotypy,Verbigeration,Rigidity,Withdrawal | 99.5 | 98.9 | 100.0 |
| 10 | Excitement,Immobility/stupor,Staring,Posturing/catalepsy,Grimacing,Stereotypy,Mannerisms,Verbigeration,Negativism,Waxy flexibility | 99.5 | 98.9 | 100.0 |
| 10 | Excitement,Immobility/stupor,Staring,Posturing/catalepsy,Grimacing,Stereotypy,Verbigeration,Negativism,Waxy flexibility,Withdrawal | 99.5 | 98.9 | 100.0 |
| 10 | Excitement,Immobility/stupor,Staring,Posturing/catalepsy,Grimacing,Stereotypy,Verbigeration,Rigidity,Negativism,Waxy flexibility | 99.5 | 98.9 | 100.0 |
| 10 | Excitement,Mutism,Staring,Grimacing,Echopraxia/echolalia,Stereotypy,Verbigeration,Negativism,Waxy flexibility,Withdrawal | 99.5 | 98.9 | 100.0 |
| 10 | Excitement,Mutism,Staring,Posturing/catalepsy,Grimacing,Echopraxia/echolalia,Mannerisms,Verbigeration,Waxy flexibility,Withdrawal | 99.5 | 98.9 | 100.0 |
| 10 | Excitement,Staring,Grimacing,Echopraxia/echolalia,Stereotypy,Mannerisms,Rigidity,Negativism,Waxy flexibility,Withdrawal | 99.5 | 98.9 | 100.0 |
| 10 | Excitement,Staring,Grimacing,Echopraxia/echolalia,Stereotypy,Mannerisms,Verbigeration,Rigidity,Negativism,Withdrawal | 99.5 | 98.9 | 100.0 |
| 10 | Excitement,Staring,Posturing/catalepsy,Echopraxia/echolalia,Stereotypy,Mannerisms,Rigidity,Negativism,Waxy flexibility,Withdrawal | 99.5 | 98.9 | 100.0 |
| 10 | Excitement,Staring,Posturing/catalepsy,Echopraxia/echolalia,Stereotypy,Mannerisms,Verbigeration,Rigidity,Negativism,Withdrawal | 99.5 | 98.9 | 100.0 |
| 10 | Excitement,Staring,Posturing/catalepsy,Grimacing,Echopraxia/echolalia,Mannerisms,Rigidity,Negativism,Waxy flexibility,Withdrawal | 99.5 | 98.9 | 100.0 |
| 10 | Immobility/stupor,Mutism,Staring,Grimacing,Echopraxia/echolalia,Stereotypy,Mannerisms,Rigidity,Negativism,Waxy flexibility | 99.5 | 98.9 | 100.0 |
| 10 | Immobility/stupor,Mutism,Staring,Grimacing,Stereotypy,Mannerisms,Verbigeration,Rigidity,Waxy flexibility,Withdrawal | 99.5 | 98.9 | 100.0 |
| 10 | Immobility/stupor,Mutism,Staring,Posturing/catalepsy,Echopraxia/echolalia,Stereotypy,Mannerisms,Negativism,Waxy flexibility,Withdrawal | 99.5 | 98.9 | 100.0 |
| 10 | Immobility/stupor,Mutism,Staring,Posturing/catalepsy,Grimacing,Echopraxia/echolalia,Mannerisms,Verbigeration,Negativism,Waxy flexibility | 99.5 | 98.9 | 100.0 |
| 10 | Immobility/stupor,Mutism,Staring,Posturing/catalepsy,Grimacing,Echopraxia/echolalia,Stereotypy,Mannerisms,Rigidity,Withdrawal | 99.5 | 98.9 | 100.0 |
| 10 | Immobility/stupor,Mutism,Staring,Posturing/catalepsy,Grimacing,Stereotypy,Mannerisms,Verbigeration,Negativism,Waxy flexibility | 99.5 | 98.9 | 100.0 |
| 10 | Immobility/stupor,Mutism,Staring,Posturing/catalepsy,Stereotypy,Mannerisms,Verbigeration,Rigidity,Negativism,Waxy flexibility | 99.5 | 98.9 | 100.0 |
| 10 | Immobility/stupor,Staring,Grimacing,Echopraxia/echolalia,Stereotypy,Mannerisms,Verbigeration,Rigidity,Negativism,Waxy flexibility | 99.5 | 98.9 | 100.0 |
| 10 | Immobility/stupor,Staring,Posturing/catalepsy,Grimacing,Mannerisms,Verbigeration,Rigidity,Negativism,Waxy flexibility,Withdrawal | 99.5 | 98.9 | 100.0 |
| 10 | Mutism,Staring,Grimacing,Echopraxia/echolalia,Stereotypy,Mannerisms,Verbigeration,Rigidity,Waxy flexibility,Withdrawal | 99.5 | 98.9 | 100.0 |
| 10 | Staring,Grimacing,Echopraxia/echolalia,Stereotypy,Mannerisms,Verbigeration,Rigidity,Negativism,Waxy flexibility,Withdrawal | 99.5 | 98.9 | 100.0 |
| 10 | Excitement,Immobility/stupor,Grimacing,Echopraxia/echolalia,Mannerisms,Verbigeration,Rigidity,Negativism,Waxy flexibility,Withdrawal | 99.3 | 98.4 | 100.0 |
| 10 | Excitement,Immobility/stupor,Grimacing,Echopraxia/echolalia,Stereotypy,Mannerisms,Rigidity,Negativism,Waxy flexibility,Withdrawal | 99.3 | 98.4 | 100.0 |
| 10 | Excitement,Immobility/stupor,Grimacing,Echopraxia/echolalia,Stereotypy,Verbigeration,Rigidity,Negativism,Waxy flexibility,Withdrawal | 99.3 | 98.4 | 100.0 |
| 10 | Excitement,Immobility/stupor,Mutism,Echopraxia/echolalia,Mannerisms,Verbigeration,Rigidity,Negativism,Waxy flexibility,Withdrawal | 99.3 | 98.4 | 100.0 |
| 10 | Excitement,Immobility/stupor,Mutism,Echopraxia/echolalia,Stereotypy,Mannerisms,Rigidity,Negativism,Waxy flexibility,Withdrawal | 99.3 | 98.4 | 100.0 |
| 10 | Excitement,Immobility/stupor,Mutism,Echopraxia/echolalia,Stereotypy,Mannerisms,Verbigeration,Rigidity,Waxy flexibility,Withdrawal | 99.3 | 98.4 | 100.0 |
| 10 | Excitement,Immobility/stupor,Mutism,Grimacing,Echopraxia/echolalia,Mannerisms,Rigidity,Negativism,Waxy flexibility,Withdrawal | 99.3 | 98.4 | 100.0 |
| 10 | Excitement,Immobility/stupor,Mutism,Grimacing,Echopraxia/echolalia,Mannerisms,Verbigeration,Rigidity,Negativism,Waxy flexibility | 99.3 | 98.4 | 100.0 |
| 10 | Excitement,Immobility/stupor,Mutism,Grimacing,Echopraxia/echolalia,Stereotypy,Mannerisms,Verbigeration,Rigidity,Negativism | 99.3 | 98.4 | 100.0 |
| 10 | Excitement,Immobility/stupor,Mutism,Grimacing,Echopraxia/echolalia,Stereotypy,Verbigeration,Rigidity,Waxy flexibility,Withdrawal | 99.3 | 98.4 | 100.0 |
| 10 | Excitement,Immobility/stupor,Mutism,Grimacing,Mannerisms,Verbigeration,Rigidity,Negativism,Waxy flexibility,Withdrawal | 99.3 | 98.4 | 100.0 |
| 10 | Excitement,Immobility/stupor,Mutism,Grimacing,Stereotypy,Mannerisms,Verbigeration,Rigidity,Negativism,Waxy flexibility | 99.3 | 98.4 | 100.0 |
| 10 | Excitement,Immobility/stupor,Mutism,Posturing/catalepsy,Echopraxia/echolalia,Mannerisms,Rigidity,Negativism,Waxy flexibility,Withdrawal | 99.3 | 98.4 | 100.0 |
| 10 | Excitement,Immobility/stupor,Mutism,Posturing/catalepsy,Echopraxia/echolalia,Mannerisms,Verbigeration,Rigidity,Waxy flexibility,Withdrawal | 99.3 | 98.4 | 100.0 |
| 10 | Excitement,Immobility/stupor,Mutism,Posturing/catalepsy,Grimacing,Echopraxia/echolalia,Mannerisms,Rigidity,Negativism,Waxy flexibility | 99.3 | 98.4 | 100.0 |
| 10 | Excitement,Immobility/stupor,Mutism,Posturing/catalepsy,Grimacing,Echopraxia/echolalia,Mannerisms,Rigidity,Waxy flexibility,Withdrawal | 99.3 | 98.4 | 100.0 |
| 10 | Excitement,Immobility/stupor,Mutism,Posturing/catalepsy,Grimacing,Echopraxia/echolalia,Stereotypy,Mannerisms,Rigidity,Negativism | 99.3 | 98.4 | 100.0 |
| 10 | Excitement,Immobility/stupor,Mutism,Posturing/catalepsy,Grimacing,Echopraxia/echolalia,Stereotypy,Mannerisms,Rigidity,Withdrawal | 99.3 | 98.4 | 100.0 |
| 10 | Excitement,Immobility/stupor,Mutism,Posturing/catalepsy,Grimacing,Echopraxia/echolalia,Stereotypy,Rigidity,Negativism,Waxy flexibility | 99.3 | 98.4 | 100.0 |
| 10 | Excitement,Immobility/stupor,Mutism,Posturing/catalepsy,Grimacing,Echopraxia/echolalia,Stereotypy,Rigidity,Waxy flexibility,Withdrawal | 99.3 | 98.4 | 100.0 |
| 10 | Excitement,Immobility/stupor,Mutism,Posturing/catalepsy,Grimacing,Mannerisms,Rigidity,Negativism,Waxy flexibility,Withdrawal | 99.3 | 98.4 | 100.0 |
| 10 | Excitement,Immobility/stupor,Mutism,Posturing/catalepsy,Grimacing,Mannerisms,Verbigeration,Rigidity,Waxy flexibility,Withdrawal | 99.3 | 98.4 | 100.0 |
| 10 | Excitement,Immobility/stupor,Mutism,Posturing/catalepsy,Grimacing,Stereotypy,Mannerisms,Rigidity,Negativism,Withdrawal | 99.3 | 98.4 | 100.0 |
| 10 | Excitement,Immobility/stupor,Mutism,Posturing/catalepsy,Grimacing,Stereotypy,Mannerisms,Verbigeration,Rigidity,Withdrawal | 99.3 | 98.4 | 100.0 |
| 10 | Excitement,Immobility/stupor,Mutism,Posturing/catalepsy,Grimacing,Stereotypy,Rigidity,Negativism,Waxy flexibility,Withdrawal | 99.3 | 98.4 | 100.0 |
| 10 | Excitement,Immobility/stupor,Mutism,Posturing/catalepsy,Grimacing,Stereotypy,Verbigeration,Rigidity,Waxy flexibility,Withdrawal | 99.3 | 98.4 | 100.0 |
| 10 | Excitement,Immobility/stupor,Mutism,Posturing/catalepsy,Stereotypy,Mannerisms,Verbigeration,Rigidity,Waxy flexibility,Withdrawal | 99.3 | 98.4 | 100.0 |
| 10 | Excitement,Immobility/stupor,Mutism,Staring,Echopraxia/echolalia,Mannerisms,Verbigeration,Negativism,Waxy flexibility,Withdrawal | 99.3 | 98.4 | 100.0 |
| 10 | Excitement,Immobility/stupor,Mutism,Staring,Grimacing,Echopraxia/echolalia,Mannerisms,Negativism,Waxy flexibility,Withdrawal | 99.3 | 98.4 | 100.0 |
| 10 | Excitement,Immobility/stupor,Mutism,Staring,Grimacing,Echopraxia/echolalia,Mannerisms,Verbigeration,Negativism,Waxy flexibility | 99.3 | 98.4 | 100.0 |
| 10 | Excitement,Immobility/stupor,Mutism,Staring,Grimacing,Echopraxia/echolalia,Stereotypy,Mannerisms,Negativism,Waxy flexibility | 99.3 | 98.4 | 100.0 |
| 10 | Excitement,Immobility/stupor,Mutism,Staring,Grimacing,Echopraxia/echolalia,Stereotypy,Mannerisms,Verbigeration,Rigidity | 99.3 | 98.4 | 100.0 |
| 10 | Excitement,Immobility/stupor,Mutism,Staring,Grimacing,Echopraxia/echolalia,Verbigeration,Negativism,Waxy flexibility,Withdrawal | 99.3 | 98.4 | 100.0 |
| 10 | Excitement,Immobility/stupor,Mutism,Staring,Grimacing,Stereotypy,Mannerisms,Verbigeration,Negativism,Waxy flexibility | 99.3 | 98.4 | 100.0 |
| 10 | Excitement,Immobility/stupor,Mutism,Staring,Grimacing,Stereotypy,Mannerisms,Verbigeration,Negativism,Withdrawal | 99.3 | 98.4 | 100.0 |
| 10 | Excitement,Immobility/stupor,Mutism,Staring,Grimacing,Stereotypy,Verbigeration,Negativism,Waxy flexibility,Withdrawal | 99.3 | 98.4 | 100.0 |
| 10 | Excitement,Immobility/stupor,Mutism,Staring,Posturing/catalepsy,Echopraxia/echolalia,Mannerisms,Verbigeration,Waxy flexibility,Withdrawal | 99.3 | 98.4 | 100.0 |
| 10 | Excitement,Immobility/stupor,Mutism,Staring,Posturing/catalepsy,Echopraxia/echolalia,Stereotypy,Mannerisms,Waxy flexibility,Withdrawal | 99.3 | 98.4 | 100.0 |
| 10 | Excitement,Immobility/stupor,Mutism,Staring,Posturing/catalepsy,Echopraxia/echolalia,Stereotypy,Verbigeration,Waxy flexibility,Withdrawal | 99.3 | 98.4 | 100.0 |
| 10 | Excitement,Immobility/stupor,Mutism,Staring,Posturing/catalepsy,Grimacing,Echopraxia/echolalia,Mannerisms,Verbigeration,Waxy flexibility | 99.3 | 98.4 | 100.0 |
| 10 | Excitement,Immobility/stupor,Mutism,Staring,Posturing/catalepsy,Grimacing,Echopraxia/echolalia,Mannerisms,Verbigeration,Withdrawal | 99.3 | 98.4 | 100.0 |
| 10 | Excitement,Immobility/stupor,Mutism,Staring,Posturing/catalepsy,Grimacing,Echopraxia/echolalia,Stereotypy,Mannerisms,Withdrawal | 99.3 | 98.4 | 100.0 |
| 10 | Excitement,Immobility/stupor,Mutism,Staring,Posturing/catalepsy,Grimacing,Echopraxia/echolalia,Stereotypy,Verbigeration,Waxy flexibility | 99.3 | 98.4 | 100.0 |
| 10 | Excitement,Immobility/stupor,Mutism,Staring,Posturing/catalepsy,Grimacing,Echopraxia/echolalia,Stereotypy,Verbigeration,Withdrawal | 99.3 | 98.4 | 100.0 |
| 10 | Excitement,Immobility/stupor,Mutism,Staring,Posturing/catalepsy,Grimacing,Mannerisms,Verbigeration,Waxy flexibility,Withdrawal | 99.3 | 98.4 | 100.0 |
| 10 | Excitement,Immobility/stupor,Mutism,Staring,Posturing/catalepsy,Grimacing,Stereotypy,Mannerisms,Rigidity,Waxy flexibility | 99.3 | 98.4 | 100.0 |
| 10 | Excitement,Immobility/stupor,Mutism,Staring,Posturing/catalepsy,Grimacing,Stereotypy,Verbigeration,Waxy flexibility,Withdrawal | 99.3 | 98.4 | 100.0 |
| 10 | Excitement,Immobility/stupor,Mutism,Staring,Stereotypy,Mannerisms,Verbigeration,Negativism,Waxy flexibility,Withdrawal | 99.3 | 98.4 | 100.0 |
| 10 | Excitement,Immobility/stupor,Posturing/catalepsy,Echopraxia/echolalia,Stereotypy,Mannerisms,Rigidity,Negativism,Waxy flexibility,Withdrawal | 99.3 | 98.4 | 100.0 |
| 10 | Excitement,Immobility/stupor,Posturing/catalepsy,Grimacing,Echopraxia/echolalia,Mannerisms,Verbigeration,Rigidity,Negativism,Waxy flexibility | 99.3 | 98.4 | 100.0 |
| 10 | Excitement,Immobility/stupor,Posturing/catalepsy,Grimacing,Echopraxia/echolalia,Stereotypy,Rigidity,Negativism,Waxy flexibility,Withdrawal | 99.3 | 98.4 | 100.0 |
| 10 | Excitement,Immobility/stupor,Posturing/catalepsy,Grimacing,Stereotypy,Verbigeration,Rigidity,Negativism,Waxy flexibility,Withdrawal | 99.3 | 98.4 | 100.0 |
| 10 | Excitement,Immobility/stupor,Staring,Echopraxia/echolalia,Stereotypy,Mannerisms,Verbigeration,Negativism,Waxy flexibility,Withdrawal | 99.3 | 98.4 | 100.0 |
| 10 | Excitement,Immobility/stupor,Staring,Grimacing,Echopraxia/echolalia,Stereotypy,Mannerisms,Negativism,Waxy flexibility,Withdrawal | 99.3 | 98.4 | 100.0 |
| 10 | Excitement,Immobility/stupor,Staring,Grimacing,Echopraxia/echolalia,Stereotypy,Mannerisms,Verbigeration,Negativism,Withdrawal | 99.3 | 98.4 | 100.0 |
| 10 | Excitement,Immobility/stupor,Staring,Grimacing,Echopraxia/echolalia,Stereotypy,Mannerisms,Verbigeration,Rigidity,Waxy flexibility | 99.3 | 98.4 | 100.0 |
| 10 | Excitement,Immobility/stupor,Staring,Posturing/catalepsy,Echopraxia/echolalia,Stereotypy,Mannerisms,Verbigeration,Rigidity,Waxy flexibility | 99.3 | 98.4 | 100.0 |
| 10 | Excitement,Immobility/stupor,Staring,Posturing/catalepsy,Grimacing,Echopraxia/echolalia,Mannerisms,Verbigeration,Rigidity,Waxy flexibility | 99.3 | 98.4 | 100.0 |
| 10 | Excitement,Immobility/stupor,Staring,Posturing/catalepsy,Grimacing,Echopraxia/echolalia,Stereotypy,Mannerisms,Negativism,Waxy flexibility | 99.3 | 98.4 | 100.0 |
| 10 | Excitement,Immobility/stupor,Staring,Posturing/catalepsy,Grimacing,Echopraxia/echolalia,Stereotypy,Mannerisms,Verbigeration,Negativism | 99.3 | 98.4 | 100.0 |
| 10 | Excitement,Immobility/stupor,Staring,Posturing/catalepsy,Grimacing,Echopraxia/echolalia,Stereotypy,Mannerisms,Verbigeration,Rigidity | 99.3 | 98.4 | 100.0 |
| 10 | Excitement,Immobility/stupor,Staring,Posturing/catalepsy,Grimacing,Echopraxia/echolalia,Stereotypy,Rigidity,Negativism,Waxy flexibility | 99.3 | 98.4 | 100.0 |
| 10 | Excitement,Immobility/stupor,Staring,Posturing/catalepsy,Grimacing,Echopraxia/echolalia,Stereotypy,Verbigeration,Negativism,Waxy flexibility | 99.3 | 98.4 | 100.0 |
| 10 | Excitement,Immobility/stupor,Staring,Posturing/catalepsy,Grimacing,Stereotypy,Mannerisms,Negativism,Waxy flexibility,Withdrawal | 99.3 | 98.4 | 100.0 |
| 10 | Excitement,Immobility/stupor,Staring,Posturing/catalepsy,Grimacing,Stereotypy,Mannerisms,Rigidity,Negativism,Waxy flexibility | 99.3 | 98.4 | 100.0 |
| 10 | Excitement,Immobility/stupor,Staring,Posturing/catalepsy,Grimacing,Stereotypy,Mannerisms,Verbigeration,Rigidity,Waxy flexibility | 99.3 | 98.4 | 100.0 |
| 10 | Excitement,Mutism,Staring,Grimacing,Echopraxia/echolalia,Mannerisms,Verbigeration,Negativism,Waxy flexibility,Withdrawal | 99.3 | 98.4 | 100.0 |
| 10 | Excitement,Mutism,Staring,Grimacing,Stereotypy,Mannerisms,Verbigeration,Negativism,Waxy flexibility,Withdrawal | 99.3 | 98.4 | 100.0 |
| 10 | Excitement,Mutism,Staring,Posturing/catalepsy,Echopraxia/echolalia,Stereotypy,Mannerisms,Verbigeration,Waxy flexibility,Withdrawal | 99.3 | 98.4 | 100.0 |
| 10 | Excitement,Mutism,Staring,Posturing/catalepsy,Grimacing,Echopraxia/echolalia,Stereotypy,Mannerisms,Verbigeration,Waxy flexibility | 99.3 | 98.4 | 100.0 |
| 10 | Excitement,Mutism,Staring,Posturing/catalepsy,Grimacing,Echopraxia/echolalia,Stereotypy,Mannerisms,Verbigeration,Withdrawal | 99.3 | 98.4 | 100.0 |
| 10 | Excitement,Mutism,Staring,Posturing/catalepsy,Grimacing,Stereotypy,Mannerisms,Rigidity,Negativism,Waxy flexibility | 99.3 | 98.4 | 100.0 |
| 10 | Excitement,Mutism,Staring,Posturing/catalepsy,Grimacing,Stereotypy,Mannerisms,Verbigeration,Waxy flexibility,Withdrawal | 99.3 | 98.4 | 100.0 |
| 10 | Excitement,Staring,Grimacing,Echopraxia/echolalia,Mannerisms,Verbigeration,Rigidity,Negativism,Waxy flexibility,Withdrawal | 99.3 | 98.4 | 100.0 |
| 10 | Excitement,Staring,Grimacing,Echopraxia/echolalia,Stereotypy,Mannerisms,Verbigeration,Rigidity,Waxy flexibility,Withdrawal | 99.3 | 98.4 | 100.0 |
| 10 | Excitement,Staring,Posturing/catalepsy,Grimacing,Echopraxia/echolalia,Mannerisms,Verbigeration,Rigidity,Waxy flexibility,Withdrawal | 99.3 | 98.4 | 100.0 |
| 10 | Excitement,Staring,Posturing/catalepsy,Grimacing,Echopraxia/echolalia,Stereotypy,Mannerisms,Rigidity,Waxy flexibility,Withdrawal | 99.3 | 98.4 | 100.0 |
| 10 | Excitement,Staring,Posturing/catalepsy,Grimacing,Echopraxia/echolalia,Stereotypy,Mannerisms,Verbigeration,Rigidity,Withdrawal | 99.3 | 98.4 | 100.0 |
| 10 | Excitement,Staring,Posturing/catalepsy,Grimacing,Stereotypy,Mannerisms,Rigidity,Negativism,Waxy flexibility,Withdrawal | 99.3 | 98.4 | 100.0 |
| 10 | Excitement,Staring,Posturing/catalepsy,Grimacing,Stereotypy,Mannerisms,Verbigeration,Rigidity,Waxy flexibility,Withdrawal | 99.3 | 98.4 | 100.0 |
| 10 | Immobility/stupor,Grimacing,Echopraxia/echolalia,Stereotypy,Mannerisms,Verbigeration,Rigidity,Negativism,Waxy flexibility,Withdrawal | 99.3 | 98.4 | 100.0 |
| 10 | Immobility/stupor,Mutism,Echopraxia/echolalia,Stereotypy,Mannerisms,Verbigeration,Rigidity,Negativism,Waxy flexibility,Withdrawal | 99.3 | 98.4 | 100.0 |
| 10 | Immobility/stupor,Mutism,Grimacing,Echopraxia/echolalia,Stereotypy,Mannerisms,Rigidity,Negativism,Waxy flexibility,Withdrawal | 99.3 | 98.4 | 100.0 |
| 10 | Immobility/stupor,Mutism,Grimacing,Echopraxia/echolalia,Stereotypy,Mannerisms,Verbigeration,Rigidity,Negativism,Withdrawal | 99.3 | 98.4 | 100.0 |
| 10 | Immobility/stupor,Mutism,Grimacing,Stereotypy,Mannerisms,Verbigeration,Rigidity,Negativism,Waxy flexibility,Withdrawal | 99.3 | 98.4 | 100.0 |
| 10 | Immobility/stupor,Mutism,Posturing/catalepsy,Echopraxia/echolalia,Stereotypy,Mannerisms,Verbigeration,Rigidity,Negativism,Withdrawal | 99.3 | 98.4 | 100.0 |
| 10 | Immobility/stupor,Mutism,Posturing/catalepsy,Grimacing,Echopraxia/echolalia,Mannerisms,Rigidity,Negativism,Waxy flexibility,Withdrawal | 99.3 | 98.4 | 100.0 |
| 10 | Immobility/stupor,Mutism,Posturing/catalepsy,Grimacing,Echopraxia/echolalia,Mannerisms,Verbigeration,Rigidity,Negativism,Withdrawal | 99.3 | 98.4 | 100.0 |
| 10 | Immobility/stupor,Mutism,Posturing/catalepsy,Grimacing,Echopraxia/echolalia,Stereotypy,Mannerisms,Rigidity,Negativism,Withdrawal | 99.3 | 98.4 | 100.0 |
| 10 | Immobility/stupor,Mutism,Posturing/catalepsy,Grimacing,Stereotypy,Mannerisms,Verbigeration,Rigidity,Negativism,Withdrawal | 99.3 | 98.4 | 100.0 |
| 10 | Immobility/stupor,Mutism,Posturing/catalepsy,Stereotypy,Mannerisms,Verbigeration,Rigidity,Negativism,Waxy flexibility,Withdrawal | 99.3 | 98.4 | 100.0 |
| 10 | Immobility/stupor,Mutism,Staring,Echopraxia/echolalia,Mannerisms,Verbigeration,Rigidity,Negativism,Waxy flexibility,Withdrawal | 99.3 | 98.4 | 100.0 |
| 10 | Immobility/stupor,Mutism,Staring,Grimacing,Echopraxia/echolalia,Mannerisms,Verbigeration,Rigidity,Negativism,Waxy flexibility | 99.3 | 98.4 | 100.0 |
| 10 | Immobility/stupor,Mutism,Staring,Grimacing,Echopraxia/echolalia,Stereotypy,Mannerisms,Verbigeration,Negativism,Waxy flexibility | 99.3 | 98.4 | 100.0 |
| 10 | Immobility/stupor,Mutism,Staring,Grimacing,Echopraxia/echolalia,Stereotypy,Mannerisms,Verbigeration,Rigidity,Withdrawal | 99.3 | 98.4 | 100.0 |
| 10 | Immobility/stupor,Mutism,Staring,Grimacing,Echopraxia/echolalia,Stereotypy,Verbigeration,Rigidity,Negativism,Withdrawal | 99.3 | 98.4 | 100.0 |
| 10 | Immobility/stupor,Mutism,Staring,Grimacing,Mannerisms,Verbigeration,Rigidity,Negativism,Waxy flexibility,Withdrawal | 99.3 | 98.4 | 100.0 |
| 10 | Immobility/stupor,Mutism,Staring,Grimacing,Stereotypy,Mannerisms,Verbigeration,Negativism,Waxy flexibility,Withdrawal | 99.3 | 98.4 | 100.0 |
| 10 | Immobility/stupor,Mutism,Staring,Grimacing,Stereotypy,Verbigeration,Rigidity,Negativism,Waxy flexibility,Withdrawal | 99.3 | 98.4 | 100.0 |
| 10 | Immobility/stupor,Mutism,Staring,Posturing/catalepsy,Echopraxia/echolalia,Mannerisms,Verbigeration,Negativism,Waxy flexibility,Withdrawal | 99.3 | 98.4 | 100.0 |
| 10 | Immobility/stupor,Mutism,Staring,Posturing/catalepsy,Echopraxia/echolalia,Mannerisms,Verbigeration,Rigidity,Negativism,Waxy flexibility | 99.3 | 98.4 | 100.0 |
| 10 | Immobility/stupor,Mutism,Staring,Posturing/catalepsy,Echopraxia/echolalia,Stereotypy,Mannerisms,Rigidity,Negativism,Waxy flexibility | 99.3 | 98.4 | 100.0 |
| 10 | Immobility/stupor,Mutism,Staring,Posturing/catalepsy,Echopraxia/echolalia,Stereotypy,Mannerisms,Verbigeration,Negativism,Waxy flexibility | 99.3 | 98.4 | 100.0 |
| 10 | Immobility/stupor,Mutism,Staring,Posturing/catalepsy,Grimacing,Echopraxia/echolalia,Mannerisms,Rigidity,Negativism,Waxy flexibility | 99.3 | 98.4 | 100.0 |
| 10 | Immobility/stupor,Mutism,Staring,Posturing/catalepsy,Grimacing,Echopraxia/echolalia,Mannerisms,Verbigeration,Rigidity,Withdrawal | 99.3 | 98.4 | 100.0 |
| 10 | Immobility/stupor,Mutism,Staring,Posturing/catalepsy,Grimacing,Mannerisms,Rigidity,Negativism,Waxy flexibility,Withdrawal | 99.3 | 98.4 | 100.0 |
| 10 | Immobility/stupor,Mutism,Staring,Posturing/catalepsy,Grimacing,Stereotypy,Mannerisms,Rigidity,Negativism,Waxy flexibility | 99.3 | 98.4 | 100.0 |
| 10 | Immobility/stupor,Mutism,Staring,Posturing/catalepsy,Grimacing,Stereotypy,Mannerisms,Rigidity,Waxy flexibility,Withdrawal | 99.3 | 98.4 | 100.0 |
| 10 | Immobility/stupor,Mutism,Staring,Posturing/catalepsy,Grimacing,Stereotypy,Verbigeration,Rigidity,Negativism,Withdrawal | 99.3 | 98.4 | 100.0 |
| 10 | Immobility/stupor,Mutism,Staring,Posturing/catalepsy,Mannerisms,Verbigeration,Rigidity,Negativism,Waxy flexibility,Withdrawal | 99.3 | 98.4 | 100.0 |
| 10 | Immobility/stupor,Posturing/catalepsy,Echopraxia/echolalia,Stereotypy,Mannerisms,Verbigeration,Rigidity,Negativism,Waxy flexibility,Withdrawal | 99.3 | 98.4 | 100.0 |
| 10 | Immobility/stupor,Posturing/catalepsy,Grimacing,Echopraxia/echolalia,Mannerisms,Verbigeration,Rigidity,Negativism,Waxy flexibility,Withdrawal | 99.3 | 98.4 | 100.0 |
| 10 | Immobility/stupor,Posturing/catalepsy,Grimacing,Echopraxia/echolalia,Stereotypy,Mannerisms,Rigidity,Negativism,Waxy flexibility,Withdrawal | 99.3 | 98.4 | 100.0 |
| 10 | Immobility/stupor,Posturing/catalepsy,Grimacing,Echopraxia/echolalia,Stereotypy,Mannerisms,Verbigeration,Rigidity,Negativism,Withdrawal | 99.3 | 98.4 | 100.0 |
| 10 | Immobility/stupor,Posturing/catalepsy,Grimacing,Stereotypy,Mannerisms,Verbigeration,Rigidity,Negativism,Waxy flexibility,Withdrawal | 99.3 | 98.4 | 100.0 |
| 10 | Immobility/stupor,Staring,Grimacing,Echopraxia/echolalia,Stereotypy,Mannerisms,Verbigeration,Negativism,Waxy flexibility,Withdrawal | 99.3 | 98.4 | 100.0 |
| 10 | Immobility/stupor,Staring,Posturing/catalepsy,Echopraxia/echolalia,Stereotypy,Mannerisms,Verbigeration,Negativism,Waxy flexibility,Withdrawal | 99.3 | 98.4 | 100.0 |
| 10 | Immobility/stupor,Staring,Posturing/catalepsy,Echopraxia/echolalia,Stereotypy,Mannerisms,Verbigeration,Rigidity,Negativism,Waxy flexibility | 99.3 | 98.4 | 100.0 |
| 10 | Immobility/stupor,Staring,Posturing/catalepsy,Grimacing,Echopraxia/echolalia,Mannerisms,Verbigeration,Negativism,Waxy flexibility,Withdrawal | 99.3 | 98.4 | 100.0 |
| 10 | Immobility/stupor,Staring,Posturing/catalepsy,Grimacing,Echopraxia/echolalia,Mannerisms,Verbigeration,Rigidity,Negativism,Waxy flexibility | 99.3 | 98.4 | 100.0 |
| 10 | Immobility/stupor,Staring,Posturing/catalepsy,Grimacing,Echopraxia/echolalia,Stereotypy,Mannerisms,Rigidity,Negativism,Waxy flexibility | 99.3 | 98.4 | 100.0 |
| 10 | Immobility/stupor,Staring,Posturing/catalepsy,Grimacing,Echopraxia/echolalia,Stereotypy,Mannerisms,Verbigeration,Negativism,Waxy flexibility | 99.3 | 98.4 | 100.0 |
| 10 | Mutism,Staring,Grimacing,Echopraxia/echolalia,Stereotypy,Verbigeration,Rigidity,Negativism,Waxy flexibility,Withdrawal | 99.3 | 98.4 | 100.0 |
| 10 | Mutism,Staring,Posturing/catalepsy,Grimacing,Echopraxia/echolalia,Mannerisms,Verbigeration,Rigidity,Waxy flexibility,Withdrawal | 99.3 | 98.4 | 100.0 |
| 10 | Mutism,Staring,Posturing/catalepsy,Grimacing,Echopraxia/echolalia,Stereotypy,Verbigeration,Rigidity,Negativism,Withdrawal | 99.3 | 98.4 | 100.0 |
| 10 | Mutism,Staring,Posturing/catalepsy,Grimacing,Stereotypy,Verbigeration,Rigidity,Negativism,Waxy flexibility,Withdrawal | 99.3 | 98.4 | 100.0 |
| 10 | Staring,Posturing/catalepsy,Echopraxia/echolalia,Stereotypy,Mannerisms,Verbigeration,Rigidity,Negativism,Waxy flexibility,Withdrawal | 99.3 | 98.4 | 100.0 |
| 10 | Staring,Posturing/catalepsy,Grimacing,Echopraxia/echolalia,Mannerisms,Verbigeration,Rigidity,Negativism,Waxy flexibility,Withdrawal | 99.3 | 98.4 | 100.0 |
| 10 | Excitement,Immobility/stupor,Grimacing,Echopraxia/echolalia,Stereotypy,Mannerisms,Verbigeration,Rigidity,Negativism,Waxy flexibility | 99.1 | 98.2 | 99.8 |
| 10 | Excitement,Immobility/stupor,Grimacing,Stereotypy,Mannerisms,Verbigeration,Rigidity,Negativism,Waxy flexibility,Withdrawal | 99.1 | 98.2 | 99.8 |
| 10 | Excitement,Immobility/stupor,Mutism,Grimacing,Echopraxia/echolalia,Mannerisms,Verbigeration,Rigidity,Waxy flexibility,Withdrawal | 99.1 | 98.2 | 99.8 |
| 10 | Excitement,Immobility/stupor,Mutism,Grimacing,Echopraxia/echolalia,Stereotypy,Mannerisms,Rigidity,Negativism,Waxy flexibility | 99.1 | 98.2 | 99.8 |
| 10 | Excitement,Immobility/stupor,Mutism,Grimacing,Echopraxia/echolalia,Stereotypy,Mannerisms,Rigidity,Waxy flexibility,Withdrawal | 99.1 | 98.2 | 99.8 |
| 10 | Excitement,Immobility/stupor,Mutism,Grimacing,Echopraxia/echolalia,Stereotypy,Mannerisms,Verbigeration,Rigidity,Withdrawal | 99.1 | 98.2 | 99.8 |
| 10 | Excitement,Immobility/stupor,Mutism,Grimacing,Stereotypy,Mannerisms,Rigidity,Negativism,Waxy flexibility,Withdrawal | 99.1 | 98.2 | 99.8 |
| 10 | Excitement,Immobility/stupor,Mutism,Grimacing,Stereotypy,Mannerisms,Verbigeration,Rigidity,Waxy flexibility,Withdrawal | 99.1 | 98.2 | 99.8 |
| 10 | Excitement,Immobility/stupor,Mutism,Posturing/catalepsy,Echopraxia/echolalia,Stereotypy,Mannerisms,Rigidity,Negativism,Waxy flexibility | 99.1 | 98.2 | 99.8 |
| 10 | Excitement,Immobility/stupor,Mutism,Posturing/catalepsy,Echopraxia/echolalia,Stereotypy,Mannerisms,Rigidity,Waxy flexibility,Withdrawal | 99.1 | 98.2 | 99.8 |
| 10 | Excitement,Immobility/stupor,Mutism,Posturing/catalepsy,Echopraxia/echolalia,Stereotypy,Mannerisms,Verbigeration,Negativism,Withdrawal | 99.1 | 98.2 | 99.8 |
| 10 | Excitement,Immobility/stupor,Mutism,Posturing/catalepsy,Echopraxia/echolalia,Stereotypy,Mannerisms,Verbigeration,Rigidity,Waxy flexibility | 99.1 | 98.2 | 99.8 |
| 10 | Excitement,Immobility/stupor,Mutism,Posturing/catalepsy,Echopraxia/echolalia,Stereotypy,Verbigeration,Negativism,Waxy flexibility,Withdrawal | 99.1 | 98.2 | 99.8 |
| 10 | Excitement,Immobility/stupor,Mutism,Posturing/catalepsy,Grimacing,Echopraxia/echolalia,Mannerisms,Verbigeration,Negativism,Withdrawal | 99.1 | 98.2 | 99.8 |
| 10 | Excitement,Immobility/stupor,Mutism,Posturing/catalepsy,Grimacing,Echopraxia/echolalia,Mannerisms,Verbigeration,Rigidity,Waxy flexibility | 99.1 | 98.2 | 99.8 |
| 10 | Excitement,Immobility/stupor,Mutism,Posturing/catalepsy,Grimacing,Echopraxia/echolalia,Stereotypy,Mannerisms,Verbigeration,Rigidity | 99.1 | 98.2 | 99.8 |
| 10 | Excitement,Immobility/stupor,Mutism,Posturing/catalepsy,Grimacing,Echopraxia/echolalia,Stereotypy,Verbigeration,Negativism,Withdrawal | 99.1 | 98.2 | 99.8 |
| 10 | Excitement,Immobility/stupor,Mutism,Posturing/catalepsy,Grimacing,Echopraxia/echolalia,Stereotypy,Verbigeration,Rigidity,Waxy flexibility | 99.1 | 98.2 | 99.8 |
| 10 | Excitement,Immobility/stupor,Mutism,Posturing/catalepsy,Grimacing,Echopraxia/echolalia,Verbigeration,Negativism,Waxy flexibility,Withdrawal | 99.1 | 98.2 | 99.8 |
| 10 | Excitement,Immobility/stupor,Mutism,Posturing/catalepsy,Stereotypy,Mannerisms,Rigidity,Negativism,Waxy flexibility,Withdrawal | 99.1 | 98.2 | 99.8 |
| 10 | Excitement,Immobility/stupor,Mutism,Staring,Grimacing,Echopraxia/echolalia,Mannerisms,Verbigeration,Negativism,Withdrawal | 99.1 | 98.2 | 99.8 |
| 10 | Excitement,Immobility/stupor,Mutism,Staring,Grimacing,Echopraxia/echolalia,Stereotypy,Mannerisms,Verbigeration,Negativism | 99.1 | 98.2 | 99.8 |
| 10 | Excitement,Immobility/stupor,Mutism,Staring,Grimacing,Echopraxia/echolalia,Stereotypy,Mannerisms,Waxy flexibility,Withdrawal | 99.1 | 98.2 | 99.8 |
| 10 | Excitement,Immobility/stupor,Mutism,Staring,Grimacing,Echopraxia/echolalia,Stereotypy,Verbigeration,Waxy flexibility,Withdrawal | 99.1 | 98.2 | 99.8 |
| 10 | Excitement,Immobility/stupor,Mutism,Staring,Grimacing,Mannerisms,Verbigeration,Negativism,Waxy flexibility,Withdrawal | 99.1 | 98.2 | 99.8 |
| 10 | Excitement,Immobility/stupor,Mutism,Staring,Grimacing,Stereotypy,Mannerisms,Negativism,Waxy flexibility,Withdrawal | 99.1 | 98.2 | 99.8 |
| 10 | Excitement,Immobility/stupor,Mutism,Staring,Posturing/catalepsy,Echopraxia/echolalia,Stereotypy,Mannerisms,Verbigeration,Waxy flexibility | 99.1 | 98.2 | 99.8 |
| 10 | Excitement,Immobility/stupor,Mutism,Staring,Posturing/catalepsy,Echopraxia/echolalia,Stereotypy,Mannerisms,Verbigeration,Withdrawal | 99.1 | 98.2 | 99.8 |
| 10 | Excitement,Immobility/stupor,Mutism,Staring,Posturing/catalepsy,Grimacing,Echopraxia/echolalia,Stereotypy,Mannerisms,Waxy flexibility | 99.1 | 98.2 | 99.8 |
| 10 | Excitement,Immobility/stupor,Mutism,Staring,Posturing/catalepsy,Grimacing,Stereotypy,Mannerisms,Verbigeration,Waxy flexibility | 99.1 | 98.2 | 99.8 |
| 10 | Excitement,Immobility/stupor,Mutism,Staring,Posturing/catalepsy,Grimacing,Stereotypy,Mannerisms,Verbigeration,Withdrawal | 99.1 | 98.2 | 99.8 |
| 10 | Excitement,Immobility/stupor,Mutism,Staring,Posturing/catalepsy,Grimacing,Stereotypy,Mannerisms,Waxy flexibility,Withdrawal | 99.1 | 98.2 | 99.8 |
| 10 | Excitement,Immobility/stupor,Mutism,Staring,Posturing/catalepsy,Stereotypy,Mannerisms,Verbigeration,Waxy flexibility,Withdrawal | 99.1 | 98.2 | 99.8 |
| 10 | Excitement,Immobility/stupor,Posturing/catalepsy,Echopraxia/echolalia,Stereotypy,Mannerisms,Verbigeration,Rigidity,Negativism,Waxy flexibility | 99.1 | 98.2 | 99.8 |
| 10 | Excitement,Immobility/stupor,Posturing/catalepsy,Echopraxia/echolalia,Stereotypy,Mannerisms,Verbigeration,Rigidity,Waxy flexibility,Withdrawal | 99.1 | 98.2 | 99.8 |
| 10 | Excitement,Immobility/stupor,Posturing/catalepsy,Grimacing,Echopraxia/echolalia,Mannerisms,Verbigeration,Rigidity,Waxy flexibility,Withdrawal | 99.1 | 98.2 | 99.8 |
| 10 | Excitement,Immobility/stupor,Posturing/catalepsy,Grimacing,Echopraxia/echolalia,Stereotypy,Mannerisms,Verbigeration,Rigidity,Negativism | 99.1 | 98.2 | 99.8 |
| 10 | Excitement,Immobility/stupor,Posturing/catalepsy,Grimacing,Echopraxia/echolalia,Stereotypy,Mannerisms,Verbigeration,Rigidity,Withdrawal | 99.1 | 98.2 | 99.8 |
| 10 | Excitement,Immobility/stupor,Posturing/catalepsy,Grimacing,Echopraxia/echolalia,Stereotypy,Verbigeration,Rigidity,Negativism,Waxy flexibility | 99.1 | 98.2 | 99.8 |
| 10 | Excitement,Immobility/stupor,Posturing/catalepsy,Grimacing,Stereotypy,Mannerisms,Rigidity,Negativism,Waxy flexibility,Withdrawal | 99.1 | 98.2 | 99.8 |
| 10 | Excitement,Immobility/stupor,Posturing/catalepsy,Grimacing,Stereotypy,Mannerisms,Verbigeration,Rigidity,Negativism,Waxy flexibility | 99.1 | 98.2 | 99.8 |
| 10 | Excitement,Immobility/stupor,Staring,Grimacing,Echopraxia/echolalia,Mannerisms,Verbigeration,Negativism,Waxy flexibility,Withdrawal | 99.1 | 98.2 | 99.8 |
| 10 | Excitement,Immobility/stupor,Staring,Grimacing,Echopraxia/echolalia,Stereotypy,Mannerisms,Verbigeration,Negativism,Waxy flexibility | 99.1 | 98.2 | 99.8 |
| 10 | Excitement,Immobility/stupor,Staring,Grimacing,Echopraxia/echolalia,Stereotypy,Verbigeration,Negativism,Waxy flexibility,Withdrawal | 99.1 | 98.2 | 99.8 |
| 10 | Excitement,Immobility/stupor,Staring,Grimacing,Stereotypy,Mannerisms,Verbigeration,Negativism,Waxy flexibility,Withdrawal | 99.1 | 98.2 | 99.8 |
| 10 | Excitement,Immobility/stupor,Staring,Posturing/catalepsy,Grimacing,Echopraxia/echolalia,Mannerisms,Verbigeration,Waxy flexibility,Withdrawal | 99.1 | 98.2 | 99.8 |
| 10 | Excitement,Immobility/stupor,Staring,Posturing/catalepsy,Grimacing,Echopraxia/echolalia,Stereotypy,Mannerisms,Rigidity,Waxy flexibility | 99.1 | 98.2 | 99.8 |
| 10 | Excitement,Immobility/stupor,Staring,Posturing/catalepsy,Grimacing,Echopraxia/echolalia,Stereotypy,Mannerisms,Waxy flexibility,Withdrawal | 99.1 | 98.2 | 99.8 |
| 10 | Excitement,Immobility/stupor,Staring,Posturing/catalepsy,Grimacing,Echopraxia/echolalia,Stereotypy,Verbigeration,Rigidity,Waxy flexibility | 99.1 | 98.2 | 99.8 |
| 10 | Excitement,Mutism,Staring,Grimacing,Echopraxia/echolalia,Stereotypy,Mannerisms,Verbigeration,Waxy flexibility,Withdrawal | 99.1 | 98.2 | 99.8 |
| 10 | Excitement,Staring,Grimacing,Echopraxia/echolalia,Stereotypy,Verbigeration,Rigidity,Negativism,Waxy flexibility,Withdrawal | 99.1 | 98.2 | 99.8 |
| 10 | Excitement,Staring,Posturing/catalepsy,Echopraxia/echolalia,Stereotypy,Mannerisms,Verbigeration,Negativism,Waxy flexibility,Withdrawal | 99.1 | 98.2 | 99.8 |
| 10 | Excitement,Staring,Posturing/catalepsy,Echopraxia/echolalia,Stereotypy,Mannerisms,Verbigeration,Rigidity,Waxy flexibility,Withdrawal | 99.1 | 98.2 | 99.8 |
| 10 | Excitement,Staring,Posturing/catalepsy,Echopraxia/echolalia,Stereotypy,Verbigeration,Rigidity,Negativism,Waxy flexibility,Withdrawal | 99.1 | 98.2 | 99.8 |
| 10 | Excitement,Staring,Posturing/catalepsy,Grimacing,Echopraxia/echolalia,Mannerisms,Verbigeration,Negativism,Waxy flexibility,Withdrawal | 99.1 | 98.2 | 99.8 |
| 10 | Excitement,Staring,Posturing/catalepsy,Grimacing,Echopraxia/echolalia,Stereotypy,Mannerisms,Negativism,Waxy flexibility,Withdrawal | 99.1 | 98.2 | 99.8 |
| 10 | Excitement,Staring,Posturing/catalepsy,Grimacing,Echopraxia/echolalia,Stereotypy,Mannerisms,Verbigeration,Negativism,Withdrawal | 99.1 | 98.2 | 99.8 |
| 10 | Excitement,Staring,Posturing/catalepsy,Grimacing,Echopraxia/echolalia,Stereotypy,Rigidity,Negativism,Waxy flexibility,Withdrawal | 99.1 | 98.2 | 99.8 |
| 10 | Excitement,Staring,Posturing/catalepsy,Grimacing,Echopraxia/echolalia,Stereotypy,Verbigeration,Rigidity,Negativism,Withdrawal | 99.1 | 98.2 | 99.8 |
| 10 | Excitement,Staring,Posturing/catalepsy,Grimacing,Echopraxia/echolalia,Verbigeration,Rigidity,Negativism,Waxy flexibility,Withdrawal | 99.1 | 98.2 | 99.8 |
| 10 | Excitement,Staring,Posturing/catalepsy,Grimacing,Stereotypy,Mannerisms,Verbigeration,Negativism,Waxy flexibility,Withdrawal | 99.1 | 98.2 | 99.8 |
| 10 | Excitement,Staring,Posturing/catalepsy,Grimacing,Stereotypy,Verbigeration,Rigidity,Negativism,Waxy flexibility,Withdrawal | 99.1 | 98.2 | 99.8 |
| 10 | Immobility/stupor,Mutism,Grimacing,Echopraxia/echolalia,Mannerisms,Verbigeration,Rigidity,Negativism,Waxy flexibility,Withdrawal | 99.1 | 98.2 | 99.8 |
| 10 | Immobility/stupor,Mutism,Grimacing,Echopraxia/echolalia,Stereotypy,Mannerisms,Verbigeration,Rigidity,Negativism,Waxy flexibility | 99.1 | 98.2 | 99.8 |
| 10 | Immobility/stupor,Mutism,Posturing/catalepsy,Echopraxia/echolalia,Mannerisms,Verbigeration,Rigidity,Negativism,Waxy flexibility,Withdrawal | 99.1 | 98.2 | 99.8 |
| 10 | Immobility/stupor,Mutism,Posturing/catalepsy,Echopraxia/echolalia,Stereotypy,Mannerisms,Rigidity,Negativism,Waxy flexibility,Withdrawal | 99.1 | 98.2 | 99.8 |
| 10 | Immobility/stupor,Mutism,Posturing/catalepsy,Echopraxia/echolalia,Stereotypy,Mannerisms,Verbigeration,Rigidity,Negativism,Waxy flexibility | 99.1 | 98.2 | 99.8 |
| 10 | Immobility/stupor,Mutism,Posturing/catalepsy,Grimacing,Echopraxia/echolalia,Mannerisms,Verbigeration,Rigidity,Negativism,Waxy flexibility | 99.1 | 98.2 | 99.8 |
| 10 | Immobility/stupor,Mutism,Posturing/catalepsy,Grimacing,Echopraxia/echolalia,Stereotypy,Mannerisms,Verbigeration,Rigidity,Negativism | 99.1 | 98.2 | 99.8 |
| 10 | Immobility/stupor,Mutism,Posturing/catalepsy,Grimacing,Mannerisms,Verbigeration,Rigidity,Negativism,Waxy flexibility,Withdrawal | 99.1 | 98.2 | 99.8 |
| 10 | Immobility/stupor,Mutism,Posturing/catalepsy,Grimacing,Stereotypy,Mannerisms,Rigidity,Negativism,Waxy flexibility,Withdrawal | 99.1 | 98.2 | 99.8 |
| 10 | Immobility/stupor,Mutism,Posturing/catalepsy,Grimacing,Stereotypy,Mannerisms,Verbigeration,Rigidity,Negativism,Waxy flexibility | 99.1 | 98.2 | 99.8 |
| 10 | Immobility/stupor,Mutism,Staring,Echopraxia/echolalia,Stereotypy,Mannerisms,Verbigeration,Negativism,Waxy flexibility,Withdrawal | 99.1 | 98.2 | 99.8 |
| 10 | Immobility/stupor,Mutism,Staring,Echopraxia/echolalia,Stereotypy,Verbigeration,Rigidity,Negativism,Waxy flexibility,Withdrawal | 99.1 | 98.2 | 99.8 |
| 10 | Immobility/stupor,Mutism,Staring,Grimacing,Echopraxia/echolalia,Mannerisms,Verbigeration,Negativism,Waxy flexibility,Withdrawal | 99.1 | 98.2 | 99.8 |
| 10 | Immobility/stupor,Mutism,Staring,Grimacing,Echopraxia/echolalia,Mannerisms,Verbigeration,Rigidity,Waxy flexibility,Withdrawal | 99.1 | 98.2 | 99.8 |
| 10 | Immobility/stupor,Mutism,Staring,Grimacing,Echopraxia/echolalia,Stereotypy,Mannerisms,Verbigeration,Rigidity,Waxy flexibility | 99.1 | 98.2 | 99.8 |
| 10 | Immobility/stupor,Mutism,Staring,Grimacing,Echopraxia/echolalia,Stereotypy,Verbigeration,Rigidity,Negativism,Waxy flexibility | 99.1 | 98.2 | 99.8 |
| 10 | Immobility/stupor,Mutism,Staring,Posturing/catalepsy,Echopraxia/echolalia,Stereotypy,Mannerisms,Rigidity,Waxy flexibility,Withdrawal | 99.1 | 98.2 | 99.8 |
| 10 | Immobility/stupor,Mutism,Staring,Posturing/catalepsy,Echopraxia/echolalia,Stereotypy,Mannerisms,Verbigeration,Rigidity,Withdrawal | 99.1 | 98.2 | 99.8 |
| 10 | Immobility/stupor,Mutism,Staring,Posturing/catalepsy,Echopraxia/echolalia,Stereotypy,Verbigeration,Rigidity,Negativism,Withdrawal | 99.1 | 98.2 | 99.8 |
| 10 | Immobility/stupor,Mutism,Staring,Posturing/catalepsy,Grimacing,Echopraxia/echolalia,Mannerisms,Rigidity,Waxy flexibility,Withdrawal | 99.1 | 98.2 | 99.8 |
| 10 | Immobility/stupor,Mutism,Staring,Posturing/catalepsy,Grimacing,Echopraxia/echolalia,Stereotypy,Mannerisms,Verbigeration,Rigidity | 99.1 | 98.2 | 99.8 |
| 10 | Immobility/stupor,Mutism,Staring,Posturing/catalepsy,Grimacing,Echopraxia/echolalia,Stereotypy,Mannerisms,Waxy flexibility,Withdrawal | 99.1 | 98.2 | 99.8 |
| 10 | Immobility/stupor,Mutism,Staring,Posturing/catalepsy,Grimacing,Echopraxia/echolalia,Stereotypy,Rigidity,Negativism,Withdrawal | 99.1 | 98.2 | 99.8 |
| 10 | Immobility/stupor,Mutism,Staring,Posturing/catalepsy,Grimacing,Echopraxia/echolalia,Stereotypy,Verbigeration,Negativism,Withdrawal | 99.1 | 98.2 | 99.8 |
| 10 | Immobility/stupor,Mutism,Staring,Posturing/catalepsy,Grimacing,Echopraxia/echolalia,Stereotypy,Verbigeration,Rigidity,Negativism | 99.1 | 98.2 | 99.8 |
| 10 | Immobility/stupor,Mutism,Staring,Posturing/catalepsy,Grimacing,Mannerisms,Verbigeration,Rigidity,Negativism,Waxy flexibility | 99.1 | 98.2 | 99.8 |
| 10 | Immobility/stupor,Mutism,Staring,Posturing/catalepsy,Grimacing,Mannerisms,Verbigeration,Rigidity,Waxy flexibility,Withdrawal | 99.1 | 98.2 | 99.8 |
| 10 | Immobility/stupor,Mutism,Staring,Posturing/catalepsy,Grimacing,Stereotypy,Mannerisms,Verbigeration,Rigidity,Waxy flexibility | 99.1 | 98.2 | 99.8 |
| 10 | Immobility/stupor,Mutism,Staring,Posturing/catalepsy,Grimacing,Stereotypy,Verbigeration,Negativism,Waxy flexibility,Withdrawal | 99.1 | 98.2 | 99.8 |
| 10 | Immobility/stupor,Mutism,Staring,Posturing/catalepsy,Grimacing,Stereotypy,Verbigeration,Rigidity,Negativism,Waxy flexibility | 99.1 | 98.2 | 99.8 |
| 10 | Immobility/stupor,Mutism,Staring,Posturing/catalepsy,Stereotypy,Mannerisms,Verbigeration,Rigidity,Waxy flexibility,Withdrawal | 99.1 | 98.2 | 99.8 |
| 10 | Immobility/stupor,Mutism,Staring,Posturing/catalepsy,Stereotypy,Verbigeration,Rigidity,Negativism,Waxy flexibility,Withdrawal | 99.1 | 98.2 | 99.8 |
| 10 | Immobility/stupor,Staring,Grimacing,Echopraxia/echolalia,Stereotypy,Mannerisms,Verbigeration,Rigidity,Waxy flexibility,Withdrawal | 99.1 | 98.2 | 99.8 |
| 10 | Immobility/stupor,Staring,Grimacing,Echopraxia/echolalia,Stereotypy,Verbigeration,Rigidity,Negativism,Waxy flexibility,Withdrawal | 99.1 | 98.2 | 99.8 |
| 10 | Immobility/stupor,Staring,Posturing/catalepsy,Grimacing,Echopraxia/echolalia,Stereotypy,Mannerisms,Rigidity,Waxy flexibility,Withdrawal | 99.1 | 98.2 | 99.8 |
| 10 | Immobility/stupor,Staring,Posturing/catalepsy,Grimacing,Echopraxia/echolalia,Stereotypy,Mannerisms,Verbigeration,Rigidity,Withdrawal | 99.1 | 98.2 | 99.8 |
| 10 | Immobility/stupor,Staring,Posturing/catalepsy,Grimacing,Echopraxia/echolalia,Stereotypy,Verbigeration,Rigidity,Negativism,Withdrawal | 99.1 | 98.2 | 99.8 |
| 10 | Immobility/stupor,Staring,Posturing/catalepsy,Grimacing,Stereotypy,Mannerisms,Verbigeration,Rigidity,Waxy flexibility,Withdrawal | 99.1 | 98.2 | 99.8 |
| 10 | Immobility/stupor,Staring,Posturing/catalepsy,Grimacing,Stereotypy,Verbigeration,Rigidity,Negativism,Waxy flexibility,Withdrawal | 99.1 | 98.2 | 99.8 |
| 10 | Mutism,Staring,Posturing/catalepsy,Echopraxia/echolalia,Stereotypy,Mannerisms,Verbigeration,Rigidity,Waxy flexibility,Withdrawal | 99.1 | 98.2 | 99.8 |
| 10 | Mutism,Staring,Posturing/catalepsy,Echopraxia/echolalia,Stereotypy,Verbigeration,Rigidity,Negativism,Waxy flexibility,Withdrawal | 99.1 | 98.2 | 99.8 |
| 10 | Mutism,Staring,Posturing/catalepsy,Grimacing,Echopraxia/echolalia,Stereotypy,Mannerisms,Verbigeration,Rigidity,Waxy flexibility | 99.1 | 98.2 | 99.8 |
| 10 | Mutism,Staring,Posturing/catalepsy,Grimacing,Echopraxia/echolalia,Stereotypy,Mannerisms,Verbigeration,Waxy flexibility,Withdrawal | 99.1 | 98.2 | 99.8 |
| 10 | Mutism,Staring,Posturing/catalepsy,Grimacing,Echopraxia/echolalia,Stereotypy,Rigidity,Negativism,Waxy flexibility,Withdrawal | 99.1 | 98.2 | 99.8 |
| 10 | Mutism,Staring,Posturing/catalepsy,Grimacing,Echopraxia/echolalia,Stereotypy,Verbigeration,Negativism,Waxy flexibility,Withdrawal | 99.1 | 98.2 | 99.8 |
| 10 | Mutism,Staring,Posturing/catalepsy,Grimacing,Echopraxia/echolalia,Stereotypy,Verbigeration,Rigidity,Negativism,Waxy flexibility | 99.1 | 98.2 | 99.8 |
| 10 | Staring,Posturing/catalepsy,Grimacing,Echopraxia/echolalia,Stereotypy,Mannerisms,Verbigeration,Negativism,Waxy flexibility,Withdrawal | 99.1 | 98.2 | 99.8 |
| 10 | Excitement,Immobility/stupor,Grimacing,Echopraxia/echolalia,Stereotypy,Mannerisms,Verbigeration,Rigidity,Waxy flexibility,Withdrawal | 98.9 | 97.8 | 99.8 |
| 10 | Excitement,Immobility/stupor,Mutism,Posturing/catalepsy,Echopraxia/echolalia,Mannerisms,Verbigeration,Negativism,Waxy flexibility,Withdrawal | 98.9 | 97.8 | 99.8 |
| 10 | Excitement,Immobility/stupor,Mutism,Posturing/catalepsy,Grimacing,Echopraxia/echolalia,Mannerisms,Negativism,Waxy flexibility,Withdrawal | 98.9 | 97.8 | 99.8 |
| 10 | Excitement,Immobility/stupor,Mutism,Posturing/catalepsy,Grimacing,Echopraxia/echolalia,Stereotypy,Mannerisms,Negativism,Withdrawal | 98.9 | 97.8 | 99.8 |
| 10 | Excitement,Immobility/stupor,Mutism,Posturing/catalepsy,Grimacing,Echopraxia/echolalia,Stereotypy,Negativism,Waxy flexibility,Withdrawal | 98.9 | 97.8 | 99.8 |
| 10 | Excitement,Immobility/stupor,Mutism,Posturing/catalepsy,Grimacing,Mannerisms,Verbigeration,Negativism,Waxy flexibility,Withdrawal | 98.9 | 97.8 | 99.8 |
| 10 | Excitement,Immobility/stupor,Mutism,Posturing/catalepsy,Grimacing,Stereotypy,Mannerisms,Rigidity,Negativism,Waxy flexibility | 98.9 | 97.8 | 99.8 |
| 10 | Excitement,Immobility/stupor,Mutism,Posturing/catalepsy,Grimacing,Stereotypy,Mannerisms,Rigidity,Waxy flexibility,Withdrawal | 98.9 | 97.8 | 99.8 |
| 10 | Excitement,Immobility/stupor,Mutism,Posturing/catalepsy,Grimacing,Stereotypy,Mannerisms,Verbigeration,Negativism,Withdrawal | 98.9 | 97.8 | 99.8 |
| 10 | Excitement,Immobility/stupor,Mutism,Posturing/catalepsy,Grimacing,Stereotypy,Mannerisms,Verbigeration,Rigidity,Waxy flexibility | 98.9 | 97.8 | 99.8 |
| 10 | Excitement,Immobility/stupor,Mutism,Posturing/catalepsy,Grimacing,Stereotypy,Verbigeration,Negativism,Waxy flexibility,Withdrawal | 98.9 | 97.8 | 99.8 |
| 10 | Excitement,Immobility/stupor,Mutism,Posturing/catalepsy,Stereotypy,Mannerisms,Verbigeration,Negativism,Waxy flexibility,Withdrawal | 98.9 | 97.8 | 99.8 |
| 10 | Excitement,Immobility/stupor,Mutism,Staring,Echopraxia/echolalia,Stereotypy,Mannerisms,Verbigeration,Waxy flexibility,Withdrawal | 98.9 | 97.8 | 99.8 |
| 10 | Excitement,Immobility/stupor,Mutism,Staring,Grimacing,Echopraxia/echolalia,Mannerisms,Verbigeration,Waxy flexibility,Withdrawal | 98.9 | 97.8 | 99.8 |
| 10 | Excitement,Immobility/stupor,Mutism,Staring,Grimacing,Echopraxia/echolalia,Stereotypy,Mannerisms,Verbigeration,Waxy flexibility | 98.9 | 97.8 | 99.8 |
| 10 | Excitement,Immobility/stupor,Mutism,Staring,Posturing/catalepsy,Grimacing,Echopraxia/echolalia,Stereotypy,Mannerisms,Verbigeration | 98.9 | 97.8 | 99.8 |
| 10 | Excitement,Immobility/stupor,Posturing/catalepsy,Grimacing,Echopraxia/echolalia,Stereotypy,Mannerisms,Rigidity,Negativism,Waxy flexibility | 98.9 | 97.8 | 99.8 |
| 10 | Excitement,Immobility/stupor,Posturing/catalepsy,Grimacing,Echopraxia/echolalia,Stereotypy,Mannerisms,Rigidity,Waxy flexibility,Withdrawal | 98.9 | 97.8 | 99.8 |
| 10 | Excitement,Immobility/stupor,Posturing/catalepsy,Grimacing,Echopraxia/echolalia,Stereotypy,Verbigeration,Rigidity,Waxy flexibility,Withdrawal | 98.9 | 97.8 | 99.8 |
| 10 | Excitement,Immobility/stupor,Staring,Posturing/catalepsy,Echopraxia/echolalia,Stereotypy,Mannerisms,Verbigeration,Waxy flexibility,Withdrawal | 98.9 | 97.8 | 99.8 |
| 10 | Excitement,Immobility/stupor,Staring,Posturing/catalepsy,Grimacing,Echopraxia/echolalia,Stereotypy,Mannerisms,Verbigeration,Withdrawal | 98.9 | 97.8 | 99.8 |
| 10 | Excitement,Immobility/stupor,Staring,Posturing/catalepsy,Grimacing,Stereotypy,Mannerisms,Verbigeration,Waxy flexibility,Withdrawal | 98.9 | 97.8 | 99.8 |
| 10 | Excitement,Staring,Posturing/catalepsy,Grimacing,Echopraxia/echolalia,Stereotypy,Verbigeration,Rigidity,Waxy flexibility,Withdrawal | 98.9 | 97.8 | 99.8 |
| 10 | Immobility/stupor,Mutism,Grimacing,Echopraxia/echolalia,Stereotypy,Verbigeration,Rigidity,Negativism,Waxy flexibility,Withdrawal | 98.9 | 97.8 | 99.8 |
| 10 | Immobility/stupor,Mutism,Posturing/catalepsy,Grimacing,Echopraxia/echolalia,Stereotypy,Mannerisms,Rigidity,Negativism,Waxy flexibility | 98.9 | 97.8 | 99.8 |
| 10 | Immobility/stupor,Mutism,Posturing/catalepsy,Grimacing,Echopraxia/echolalia,Stereotypy,Verbigeration,Rigidity,Negativism,Withdrawal | 98.9 | 97.8 | 99.8 |
| 10 | Immobility/stupor,Mutism,Posturing/catalepsy,Grimacing,Stereotypy,Verbigeration,Rigidity,Negativism,Waxy flexibility,Withdrawal | 98.9 | 97.8 | 99.8 |
| 10 | Immobility/stupor,Mutism,Staring,Echopraxia/echolalia,Stereotypy,Mannerisms,Verbigeration,Rigidity,Waxy flexibility,Withdrawal | 98.9 | 97.8 | 99.8 |
| 10 | Immobility/stupor,Mutism,Staring,Grimacing,Echopraxia/echolalia,Stereotypy,Rigidity,Negativism,Waxy flexibility,Withdrawal | 98.9 | 97.8 | 99.8 |
| 10 | Immobility/stupor,Mutism,Staring,Grimacing,Echopraxia/echolalia,Stereotypy,Verbigeration,Rigidity,Waxy flexibility,Withdrawal | 98.9 | 97.8 | 99.8 |
| 10 | Immobility/stupor,Mutism,Staring,Posturing/catalepsy,Echopraxia/echolalia,Mannerisms,Verbigeration,Rigidity,Waxy flexibility,Withdrawal | 98.9 | 97.8 | 99.8 |
| 10 | Immobility/stupor,Mutism,Staring,Posturing/catalepsy,Echopraxia/echolalia,Stereotypy,Rigidity,Negativism,Waxy flexibility,Withdrawal | 98.9 | 97.8 | 99.8 |
| 10 | Immobility/stupor,Mutism,Staring,Posturing/catalepsy,Echopraxia/echolalia,Stereotypy,Verbigeration,Rigidity,Negativism,Waxy flexibility | 98.9 | 97.8 | 99.8 |
| 10 | Immobility/stupor,Mutism,Staring,Posturing/catalepsy,Grimacing,Echopraxia/echolalia,Mannerisms,Verbigeration,Rigidity,Waxy flexibility | 98.9 | 97.8 | 99.8 |
| 10 | Immobility/stupor,Mutism,Staring,Posturing/catalepsy,Grimacing,Echopraxia/echolalia,Mannerisms,Verbigeration,Waxy flexibility,Withdrawal | 98.9 | 97.8 | 99.8 |
| 10 | Immobility/stupor,Mutism,Staring,Posturing/catalepsy,Grimacing,Echopraxia/echolalia,Stereotypy,Mannerisms,Verbigeration,Withdrawal | 98.9 | 97.8 | 99.8 |
| 10 | Immobility/stupor,Mutism,Staring,Posturing/catalepsy,Grimacing,Echopraxia/echolalia,Stereotypy,Negativism,Waxy flexibility,Withdrawal | 98.9 | 97.8 | 99.8 |
| 10 | Immobility/stupor,Mutism,Staring,Posturing/catalepsy,Grimacing,Echopraxia/echolalia,Stereotypy,Verbigeration,Negativism,Waxy flexibility | 98.9 | 97.8 | 99.8 |
| 10 | Immobility/stupor,Mutism,Staring,Posturing/catalepsy,Grimacing,Echopraxia/echolalia,Stereotypy,Verbigeration,Rigidity,Withdrawal | 98.9 | 97.8 | 99.8 |
| 10 | Immobility/stupor,Mutism,Staring,Posturing/catalepsy,Grimacing,Echopraxia/echolalia,Verbigeration,Rigidity,Negativism,Withdrawal | 98.9 | 97.8 | 99.8 |
| 10 | Immobility/stupor,Mutism,Staring,Posturing/catalepsy,Grimacing,Stereotypy,Mannerisms,Verbigeration,Waxy flexibility,Withdrawal | 98.9 | 97.8 | 99.8 |
| 10 | Immobility/stupor,Mutism,Staring,Posturing/catalepsy,Grimacing,Stereotypy,Verbigeration,Rigidity,Waxy flexibility,Withdrawal | 98.9 | 97.8 | 99.8 |
| 10 | Immobility/stupor,Posturing/catalepsy,Grimacing,Echopraxia/echolalia,Stereotypy,Mannerisms,Verbigeration,Rigidity,Negativism,Waxy flexibility | 98.9 | 97.8 | 99.8 |
| 10 | Immobility/stupor,Staring,Posturing/catalepsy,Echopraxia/echolalia,Stereotypy,Verbigeration,Rigidity,Negativism,Waxy flexibility,Withdrawal | 98.9 | 97.8 | 99.8 |
| 10 | Immobility/stupor,Staring,Posturing/catalepsy,Grimacing,Echopraxia/echolalia,Stereotypy,Rigidity,Negativism,Waxy flexibility,Withdrawal | 98.9 | 97.8 | 99.8 |
| 10 | Mutism,Staring,Posturing/catalepsy,Grimacing,Echopraxia/echolalia,Stereotypy,Verbigeration,Rigidity,Waxy flexibility,Withdrawal | 98.9 | 97.8 | 99.8 |
| 10 | Mutism,Staring,Posturing/catalepsy,Grimacing,Echopraxia/echolalia,Verbigeration,Rigidity,Negativism,Waxy flexibility,Withdrawal | 98.9 | 97.8 | 99.8 |
| 10 | Excitement,Immobility/stupor,Mutism,Grimacing,Echopraxia/echolalia,Stereotypy,Mannerisms,Verbigeration,Rigidity,Waxy flexibility | 98.7 | 97.5 | 99.6 |
| 10 | Excitement,Immobility/stupor,Mutism,Posturing/catalepsy,Echopraxia/echolalia,Stereotypy,Mannerisms,Negativism,Waxy flexibility,Withdrawal | 98.7 | 97.5 | 99.6 |
| 10 | Excitement,Immobility/stupor,Mutism,Staring,Grimacing,Echopraxia/echolalia,Stereotypy,Mannerisms,Verbigeration,Withdrawal | 98.7 | 97.5 | 99.6 |
| 10 | Excitement,Immobility/stupor,Mutism,Staring,Grimacing,Stereotypy,Mannerisms,Verbigeration,Waxy flexibility,Withdrawal | 98.7 | 97.5 | 99.6 |
| 10 | Excitement,Immobility/stupor,Posturing/catalepsy,Echopraxia/echolalia,Stereotypy,Mannerisms,Verbigeration,Negativism,Waxy flexibility,Withdrawal | 98.7 | 97.5 | 99.6 |
| 10 | Excitement,Immobility/stupor,Posturing/catalepsy,Grimacing,Echopraxia/echolalia,Mannerisms,Verbigeration,Negativism,Waxy flexibility,Withdrawal | 98.7 | 97.5 | 99.6 |
| 10 | Excitement,Immobility/stupor,Posturing/catalepsy,Grimacing,Echopraxia/echolalia,Stereotypy,Mannerisms,Verbigeration,Negativism,Withdrawal | 98.7 | 97.5 | 99.6 |
| 10 | Excitement,Immobility/stupor,Posturing/catalepsy,Grimacing,Stereotypy,Mannerisms,Verbigeration,Rigidity,Waxy flexibility,Withdrawal | 98.7 | 97.5 | 99.6 |
| 10 | Excitement,Immobility/stupor,Staring,Grimacing,Echopraxia/echolalia,Stereotypy,Mannerisms,Verbigeration,Waxy flexibility,Withdrawal | 98.7 | 97.5 | 99.6 |
| 10 | Excitement,Staring,Grimacing,Echopraxia/echolalia,Stereotypy,Mannerisms,Verbigeration,Negativism,Waxy flexibility,Withdrawal | 98.7 | 97.5 | 99.6 |
| 10 | Excitement,Staring,Grimacing,Echopraxia/echolalia,Stereotypy,Mannerisms,Verbigeration,Rigidity,Negativism,Waxy flexibility | 98.7 | 97.5 | 99.6 |
| 10 | Excitement,Staring,Posturing/catalepsy,Echopraxia/echolalia,Stereotypy,Mannerisms,Verbigeration,Rigidity,Negativism,Waxy flexibility | 98.7 | 97.5 | 99.6 |
| 10 | Immobility/stupor,Mutism,Posturing/catalepsy,Grimacing,Echopraxia/echolalia,Mannerisms,Verbigeration,Negativism,Waxy flexibility,Withdrawal | 98.7 | 97.5 | 99.6 |
| 10 | Immobility/stupor,Mutism,Posturing/catalepsy,Grimacing,Echopraxia/echolalia,Stereotypy,Mannerisms,Negativism,Waxy flexibility,Withdrawal | 98.7 | 97.5 | 99.6 |
| 10 | Immobility/stupor,Mutism,Posturing/catalepsy,Grimacing,Echopraxia/echolalia,Verbigeration,Rigidity,Negativism,Waxy flexibility,Withdrawal | 98.7 | 97.5 | 99.6 |
| 10 | Immobility/stupor,Mutism,Staring,Grimacing,Echopraxia/echolalia,Stereotypy,Mannerisms,Verbigeration,Waxy flexibility,Withdrawal | 98.7 | 97.5 | 99.6 |
| 10 | Immobility/stupor,Mutism,Staring,Grimacing,Echopraxia/echolalia,Stereotypy,Verbigeration,Negativism,Waxy flexibility,Withdrawal | 98.7 | 97.5 | 99.6 |
| 10 | Immobility/stupor,Mutism,Staring,Posturing/catalepsy,Echopraxia/echolalia,Stereotypy,Mannerisms,Verbigeration,Rigidity,Waxy flexibility | 98.7 | 97.5 | 99.6 |
| 10 | Immobility/stupor,Mutism,Staring,Posturing/catalepsy,Echopraxia/echolalia,Stereotypy,Verbigeration,Negativism,Waxy flexibility,Withdrawal | 98.7 | 97.5 | 99.6 |
| 10 | Immobility/stupor,Mutism,Staring,Posturing/catalepsy,Echopraxia/echolalia,Verbigeration,Rigidity,Negativism,Waxy flexibility,Withdrawal | 98.7 | 97.5 | 99.6 |
| 10 | Immobility/stupor,Mutism,Staring,Posturing/catalepsy,Grimacing,Echopraxia/echolalia,Rigidity,Negativism,Waxy flexibility,Withdrawal | 98.7 | 97.5 | 99.6 |
| 10 | Immobility/stupor,Mutism,Staring,Posturing/catalepsy,Grimacing,Echopraxia/echolalia,Stereotypy,Mannerisms,Rigidity,Waxy flexibility | 98.7 | 97.5 | 99.6 |
| 10 | Immobility/stupor,Mutism,Staring,Posturing/catalepsy,Grimacing,Echopraxia/echolalia,Stereotypy,Mannerisms,Verbigeration,Waxy flexibility | 98.7 | 97.5 | 99.6 |
| 10 | Immobility/stupor,Mutism,Staring,Posturing/catalepsy,Grimacing,Echopraxia/echolalia,Verbigeration,Negativism,Waxy flexibility,Withdrawal | 98.7 | 97.5 | 99.6 |
| 10 | Immobility/stupor,Mutism,Staring,Posturing/catalepsy,Grimacing,Echopraxia/echolalia,Verbigeration,Rigidity,Negativism,Waxy flexibility | 98.7 | 97.5 | 99.6 |
| 10 | Immobility/stupor,Mutism,Staring,Posturing/catalepsy,Grimacing,Stereotypy,Rigidity,Negativism,Waxy flexibility,Withdrawal | 98.7 | 97.5 | 99.6 |
| 10 | Immobility/stupor,Posturing/catalepsy,Grimacing,Echopraxia/echolalia,Stereotypy,Verbigeration,Rigidity,Negativism,Waxy flexibility,Withdrawal | 98.7 | 97.5 | 99.6 |
| 10 | Immobility/stupor,Staring,Posturing/catalepsy,Grimacing,Echopraxia/echolalia,Mannerisms,Verbigeration,Rigidity,Waxy flexibility,Withdrawal | 98.7 | 97.5 | 99.6 |
| 10 | Immobility/stupor,Staring,Posturing/catalepsy,Grimacing,Echopraxia/echolalia,Stereotypy,Verbigeration,Negativism,Waxy flexibility,Withdrawal | 98.7 | 97.5 | 99.6 |
| 10 | Immobility/stupor,Staring,Posturing/catalepsy,Grimacing,Echopraxia/echolalia,Stereotypy,Verbigeration,Rigidity,Negativism,Waxy flexibility | 98.7 | 97.5 | 99.6 |
| 10 | Immobility/stupor,Staring,Posturing/catalepsy,Grimacing,Echopraxia/echolalia,Verbigeration,Rigidity,Negativism,Waxy flexibility,Withdrawal | 98.7 | 97.5 | 99.6 |
| 10 | Staring,Posturing/catalepsy,Grimacing,Echopraxia/echolalia,Stereotypy,Mannerisms,Verbigeration,Rigidity,Waxy flexibility,Withdrawal | 98.7 | 97.5 | 99.6 |
| 10 | Excitement,Immobility/stupor,Mutism,Posturing/catalepsy,Grimacing,Echopraxia/echolalia,Mannerisms,Verbigeration,Negativism,Waxy flexibility | 98.6 | 97.5 | 99.6 |
| 10 | Excitement,Immobility/stupor,Mutism,Posturing/catalepsy,Grimacing,Echopraxia/echolalia,Stereotypy,Mannerisms,Rigidity,Waxy flexibility | 98.6 | 97.5 | 99.6 |
| 10 | Excitement,Immobility/stupor,Mutism,Posturing/catalepsy,Grimacing,Echopraxia/echolalia,Stereotypy,Verbigeration,Negativism,Waxy flexibility | 98.6 | 97.5 | 99.6 |
| 10 | Excitement,Immobility/stupor,Staring,Posturing/catalepsy,Grimacing,Echopraxia/echolalia,Stereotypy,Mannerisms,Verbigeration,Waxy flexibility | 98.6 | 97.5 | 99.6 |
| 10 | Excitement,Immobility/stupor,Staring,Posturing/catalepsy,Grimacing,Echopraxia/echolalia,Stereotypy,Verbigeration,Waxy flexibility,Withdrawal | 98.6 | 97.5 | 99.6 |
| 10 | Excitement,Staring,Posturing/catalepsy,Grimacing,Echopraxia/echolalia,Stereotypy,Mannerisms,Verbigeration,Rigidity,Negativism | 98.6 | 97.5 | 99.6 |
| 10 | Excitement,Staring,Posturing/catalepsy,Grimacing,Echopraxia/echolalia,Stereotypy,Verbigeration,Negativism,Waxy flexibility,Withdrawal | 98.6 | 97.5 | 99.6 |
| 10 | Immobility/stupor,Mutism,Posturing/catalepsy,Echopraxia/echolalia,Stereotypy,Verbigeration,Rigidity,Negativism,Waxy flexibility,Withdrawal | 98.6 | 97.5 | 99.6 |
| 10 | Immobility/stupor,Mutism,Posturing/catalepsy,Grimacing,Echopraxia/echolalia,Stereotypy,Mannerisms,Verbigeration,Negativism,Withdrawal | 98.6 | 97.5 | 99.6 |
| 10 | Immobility/stupor,Mutism,Posturing/catalepsy,Grimacing,Echopraxia/echolalia,Stereotypy,Rigidity,Negativism,Waxy flexibility,Withdrawal | 98.6 | 97.5 | 99.6 |
| 10 | Immobility/stupor,Mutism,Posturing/catalepsy,Grimacing,Echopraxia/echolalia,Stereotypy,Verbigeration,Rigidity,Negativism,Waxy flexibility | 98.6 | 97.5 | 99.6 |
| 10 | Immobility/stupor,Mutism,Staring,Grimacing,Echopraxia/echolalia,Verbigeration,Rigidity,Negativism,Waxy flexibility,Withdrawal | 98.6 | 97.5 | 99.6 |
| 10 | Immobility/stupor,Mutism,Staring,Posturing/catalepsy,Grimacing,Echopraxia/echolalia,Stereotypy,Rigidity,Waxy flexibility,Withdrawal | 98.6 | 97.5 | 99.6 |
| 10 | Excitement,Immobility/stupor,Mutism,Echopraxia/echolalia,Stereotypy,Mannerisms,Verbigeration,Negativism,Waxy flexibility,Withdrawal | 98.4 | 97.3 | 99.6 |
| 10 | Excitement,Immobility/stupor,Mutism,Grimacing,Echopraxia/echolalia,Stereotypy,Mannerisms,Verbigeration,Negativism,Withdrawal | 98.4 | 97.1 | 99.6 |
| 10 | Excitement,Immobility/stupor,Mutism,Grimacing,Echopraxia/echolalia,Stereotypy,Verbigeration,Negativism,Waxy flexibility,Withdrawal | 98.4 | 97.1 | 99.6 |
| 10 | Excitement,Immobility/stupor,Mutism,Posturing/catalepsy,Echopraxia/echolalia,Stereotypy,Mannerisms,Verbigeration,Negativism,Waxy flexibility | 98.4 | 97.1 | 99.6 |
| 10 | Excitement,Immobility/stupor,Mutism,Posturing/catalepsy,Grimacing,Echopraxia/echolalia,Stereotypy,Mannerisms,Verbigeration,Negativism | 98.4 | 97.1 | 99.6 |
| 10 | Excitement,Immobility/stupor,Mutism,Posturing/catalepsy,Grimacing,Stereotypy,Mannerisms,Negativism,Waxy flexibility,Withdrawal | 98.4 | 97.1 | 99.6 |
| 10 | Excitement,Immobility/stupor,Mutism,Posturing/catalepsy,Grimacing,Stereotypy,Mannerisms,Verbigeration,Negativism,Waxy flexibility | 98.4 | 97.1 | 99.6 |
| 10 | Excitement,Immobility/stupor,Posturing/catalepsy,Grimacing,Echopraxia/echolalia,Stereotypy,Mannerisms,Negativism,Waxy flexibility,Withdrawal | 98.4 | 97.1 | 99.6 |
| 10 | Excitement,Immobility/stupor,Posturing/catalepsy,Grimacing,Echopraxia/echolalia,Stereotypy,Verbigeration,Negativism,Waxy flexibility,Withdrawal | 98.4 | 97.1 | 99.6 |
| 10 | Excitement,Immobility/stupor,Posturing/catalepsy,Grimacing,Stereotypy,Mannerisms,Verbigeration,Negativism,Waxy flexibility,Withdrawal | 98.4 | 97.3 | 99.6 |
| 10 | Excitement,Staring,Posturing/catalepsy,Grimacing,Echopraxia/echolalia,Mannerisms,Verbigeration,Rigidity,Negativism,Waxy flexibility | 98.4 | 97.1 | 99.6 |
| 10 | Excitement,Staring,Posturing/catalepsy,Grimacing,Echopraxia/echolalia,Stereotypy,Mannerisms,Rigidity,Negativism,Waxy flexibility | 98.4 | 97.3 | 99.6 |
| 10 | Excitement,Staring,Posturing/catalepsy,Grimacing,Echopraxia/echolalia,Stereotypy,Mannerisms,Verbigeration,Waxy flexibility,Withdrawal | 98.4 | 97.3 | 99.6 |
| 10 | Excitement,Staring,Posturing/catalepsy,Grimacing,Stereotypy,Mannerisms,Verbigeration,Rigidity,Negativism,Waxy flexibility | 98.4 | 97.1 | 99.6 |
| 10 | Immobility/stupor,Mutism,Grimacing,Echopraxia/echolalia,Stereotypy,Mannerisms,Verbigeration,Rigidity,Waxy flexibility,Withdrawal | 98.4 | 97.3 | 99.6 |
| 10 | Immobility/stupor,Mutism,Posturing/catalepsy,Echopraxia/echolalia,Stereotypy,Mannerisms,Verbigeration,Negativism,Waxy flexibility,Withdrawal | 98.4 | 97.1 | 99.6 |
| 10 | Immobility/stupor,Mutism,Posturing/catalepsy,Grimacing,Echopraxia/echolalia,Mannerisms,Verbigeration,Rigidity,Waxy flexibility,Withdrawal | 98.4 | 97.1 | 99.6 |
| 10 | Immobility/stupor,Mutism,Posturing/catalepsy,Grimacing,Echopraxia/echolalia,Stereotypy,Mannerisms,Rigidity,Waxy flexibility,Withdrawal | 98.4 | 97.1 | 99.6 |
| 10 | Immobility/stupor,Mutism,Posturing/catalepsy,Grimacing,Echopraxia/echolalia,Stereotypy,Mannerisms,Verbigeration,Rigidity,Withdrawal | 98.4 | 97.3 | 99.6 |
| 10 | Immobility/stupor,Mutism,Posturing/catalepsy,Grimacing,Stereotypy,Mannerisms,Verbigeration,Negativism,Waxy flexibility,Withdrawal | 98.4 | 97.1 | 99.6 |
| 10 | Immobility/stupor,Mutism,Staring,Posturing/catalepsy,Echopraxia/echolalia,Stereotypy,Verbigeration,Rigidity,Waxy flexibility,Withdrawal | 98.4 | 97.1 | 99.6 |
| 10 | Immobility/stupor,Mutism,Staring,Posturing/catalepsy,Grimacing,Echopraxia/echolalia,Stereotypy,Rigidity,Negativism,Waxy flexibility | 98.4 | 97.1 | 99.6 |
| 10 | Immobility/stupor,Mutism,Staring,Posturing/catalepsy,Grimacing,Echopraxia/echolalia,Stereotypy,Verbigeration,Rigidity,Waxy flexibility | 98.4 | 97.3 | 99.6 |
| 10 | Immobility/stupor,Mutism,Staring,Posturing/catalepsy,Grimacing,Verbigeration,Rigidity,Negativism,Waxy flexibility,Withdrawal | 98.4 | 97.1 | 99.6 |
| 10 | Immobility/stupor,Posturing/catalepsy,Grimacing,Echopraxia/echolalia,Stereotypy,Mannerisms,Verbigeration,Negativism,Waxy flexibility,Withdrawal | 98.4 | 97.3 | 99.6 |
| 10 | Immobility/stupor,Staring,Posturing/catalepsy,Echopraxia/echolalia,Stereotypy,Mannerisms,Verbigeration,Rigidity,Waxy flexibility,Withdrawal | 98.4 | 97.1 | 99.6 |
| 10 | Immobility/stupor,Staring,Posturing/catalepsy,Grimacing,Echopraxia/echolalia,Stereotypy,Mannerisms,Verbigeration,Rigidity,Waxy flexibility | 98.4 | 97.1 | 99.6 |
| 10 | Immobility/stupor,Staring,Posturing/catalepsy,Grimacing,Echopraxia/echolalia,Stereotypy,Mannerisms,Verbigeration,Waxy flexibility,Withdrawal | 98.4 | 97.1 | 99.6 |
| 10 | Staring,Posturing/catalepsy,Grimacing,Echopraxia/echolalia,Stereotypy,Verbigeration,Rigidity,Negativism,Waxy flexibility,Withdrawal | 98.4 | 97.1 | 99.6 |
| 10 | Excitement,Immobility/stupor,Mutism,Grimacing,Echopraxia/echolalia,Stereotypy,Mannerisms,Negativism,Waxy flexibility,Withdrawal | 98.2 | 96.9 | 99.3 |
| 10 | Excitement,Immobility/stupor,Mutism,Posturing/catalepsy,Grimacing,Echopraxia/echolalia,Mannerisms,Verbigeration,Waxy flexibility,Withdrawal | 98.2 | 96.9 | 99.3 |
| 10 | Excitement,Immobility/stupor,Mutism,Posturing/catalepsy,Grimacing,Echopraxia/echolalia,Stereotypy,Mannerisms,Negativism,Waxy flexibility | 98.2 | 96.9 | 99.3 |
| 10 | Excitement,Immobility/stupor,Mutism,Posturing/catalepsy,Grimacing,Echopraxia/echolalia,Stereotypy,Verbigeration,Waxy flexibility,Withdrawal | 98.2 | 96.9 | 99.3 |
| 10 | Excitement,Immobility/stupor,Posturing/catalepsy,Grimacing,Echopraxia/echolalia,Stereotypy,Mannerisms,Verbigeration,Rigidity,Waxy flexibility | 98.2 | 96.9 | 99.3 |
| 10 | Excitement,Staring,Posturing/catalepsy,Grimacing,Echopraxia/echolalia,Stereotypy,Mannerisms,Verbigeration,Rigidity,Waxy flexibility | 98.2 | 96.9 | 99.3 |
| 10 | Excitement,Staring,Posturing/catalepsy,Grimacing,Echopraxia/echolalia,Stereotypy,Verbigeration,Rigidity,Negativism,Waxy flexibility | 98.2 | 96.9 | 99.3 |
| 10 | Immobility/stupor,Mutism,Grimacing,Echopraxia/echolalia,Stereotypy,Mannerisms,Verbigeration,Negativism,Waxy flexibility,Withdrawal | 98.2 | 96.9 | 99.3 |
| 10 | Immobility/stupor,Mutism,Posturing/catalepsy,Grimacing,Echopraxia/echolalia,Stereotypy,Mannerisms,Verbigeration,Negativism,Waxy flexibility | 98.2 | 96.9 | 99.3 |
| 10 | Immobility/stupor,Mutism,Posturing/catalepsy,Grimacing,Stereotypy,Mannerisms,Verbigeration,Rigidity,Waxy flexibility,Withdrawal | 98.2 | 96.9 | 99.3 |
| 10 | Immobility/stupor,Mutism,Staring,Posturing/catalepsy,Echopraxia/echolalia,Stereotypy,Mannerisms,Verbigeration,Waxy flexibility,Withdrawal | 98.2 | 96.9 | 99.3 |
| 10 | Immobility/stupor,Mutism,Staring,Posturing/catalepsy,Grimacing,Echopraxia/echolalia,Stereotypy,Verbigeration,Waxy flexibility,Withdrawal | 98.2 | 96.9 | 99.3 |
| 10 | Immobility/stupor,Mutism,Staring,Posturing/catalepsy,Grimacing,Echopraxia/echolalia,Verbigeration,Rigidity,Waxy flexibility,Withdrawal | 98.2 | 96.9 | 99.3 |
| 10 | Immobility/stupor,Staring,Posturing/catalepsy,Grimacing,Echopraxia/echolalia,Stereotypy,Verbigeration,Rigidity,Waxy flexibility,Withdrawal | 98.2 | 96.9 | 99.3 |
| 10 | Staring,Posturing/catalepsy,Grimacing,Echopraxia/echolalia,Stereotypy,Mannerisms,Verbigeration,Rigidity,Negativism,Waxy flexibility | 98.2 | 96.9 | 99.3 |
| 10 | Excitement,Immobility/stupor,Grimacing,Echopraxia/echolalia,Stereotypy,Mannerisms,Verbigeration,Negativism,Waxy flexibility,Withdrawal | 98.0 | 96.6 | 99.1 |
| 10 | Excitement,Immobility/stupor,Mutism,Grimacing,Echopraxia/echolalia,Mannerisms,Verbigeration,Negativism,Waxy flexibility,Withdrawal | 98.0 | 96.6 | 99.1 |
| 10 | Excitement,Immobility/stupor,Mutism,Grimacing,Echopraxia/echolalia,Stereotypy,Mannerisms,Verbigeration,Negativism,Waxy flexibility | 98.0 | 96.6 | 99.1 |
| 10 | Excitement,Immobility/stupor,Mutism,Grimacing,Stereotypy,Mannerisms,Verbigeration,Negativism,Waxy flexibility,Withdrawal | 98.0 | 96.6 | 99.1 |
| 10 | Excitement,Immobility/stupor,Mutism,Posturing/catalepsy,Echopraxia/echolalia,Stereotypy,Mannerisms,Verbigeration,Waxy flexibility,Withdrawal | 98.0 | 96.6 | 99.1 |
| 10 | Excitement,Immobility/stupor,Mutism,Posturing/catalepsy,Grimacing,Echopraxia/echolalia,Stereotypy,Mannerisms,Verbigeration,Withdrawal | 98.0 | 96.6 | 99.1 |
| 10 | Excitement,Immobility/stupor,Mutism,Posturing/catalepsy,Grimacing,Echopraxia/echolalia,Stereotypy,Mannerisms,Waxy flexibility,Withdrawal | 98.0 | 96.6 | 99.1 |
| 10 | Excitement,Immobility/stupor,Posturing/catalepsy,Grimacing,Echopraxia/echolalia,Stereotypy,Mannerisms,Verbigeration,Negativism,Waxy flexibility | 98.0 | 96.6 | 99.1 |
| 10 | Excitement,Staring,Posturing/catalepsy,Grimacing,Echopraxia/echolalia,Stereotypy,Mannerisms,Verbigeration,Negativism,Waxy flexibility | 98.0 | 96.6 | 99.1 |
| 10 | Immobility/stupor,Mutism,Posturing/catalepsy,Grimacing,Echopraxia/echolalia,Stereotypy,Verbigeration,Negativism,Waxy flexibility,Withdrawal | 98.0 | 96.6 | 99.1 |
| 10 | Immobility/stupor,Mutism,Posturing/catalepsy,Grimacing,Echopraxia/echolalia,Stereotypy,Verbigeration,Rigidity,Waxy flexibility,Withdrawal | 98.0 | 96.6 | 99.1 |
| 10 | Excitement,Immobility/stupor,Mutism,Posturing/catalepsy,Grimacing,Stereotypy,Mannerisms,Verbigeration,Waxy flexibility,Withdrawal | 97.8 | 96.2 | 99.1 |
| 10 | Immobility/stupor,Mutism,Posturing/catalepsy,Grimacing,Echopraxia/echolalia,Stereotypy,Mannerisms,Verbigeration,Rigidity,Waxy flexibility | 97.8 | 96.2 | 99.1 |
| 10 | Immobility/stupor,Posturing/catalepsy,Grimacing,Echopraxia/echolalia,Stereotypy,Mannerisms,Verbigeration,Rigidity,Waxy flexibility,Withdrawal | 97.8 | 96.2 | 99.1 |
| 10 | Immobility/stupor,Mutism,Posturing/catalepsy,Echopraxia/echolalia,Stereotypy,Mannerisms,Verbigeration,Rigidity,Waxy flexibility,Withdrawal | 97.7 | 96.2 | 99.1 |
| 10 | Excitement,Immobility/stupor,Mutism,Grimacing,Echopraxia/echolalia,Stereotypy,Mannerisms,Verbigeration,Waxy flexibility,Withdrawal | 97.5 | 96.0 | 98.9 |
| 10 | Excitement,Immobility/stupor,Mutism,Posturing/catalepsy,Grimacing,Echopraxia/echolalia,Stereotypy,Mannerisms,Verbigeration,Waxy flexibility | 97.5 | 96.0 | 98.9 |
| 10 | Excitement,Immobility/stupor,Posturing/catalepsy,Grimacing,Echopraxia/echolalia,Stereotypy,Mannerisms,Verbigeration,Waxy flexibility,Withdrawal | 97.5 | 96.0 | 98.9 |
| 10 | Excitement,Mutism,Posturing/catalepsy,Echopraxia/echolalia,Mannerisms,Verbigeration,Rigidity,Negativism,Waxy flexibility,Withdrawal | 97.3 | 95.7 | 98.7 |
| 10 | Excitement,Mutism,Posturing/catalepsy,Echopraxia/echolalia,Stereotypy,Mannerisms,Verbigeration,Rigidity,Negativism,Withdrawal | 97.3 | 95.7 | 98.7 |
| 10 | Excitement,Mutism,Posturing/catalepsy,Echopraxia/echolalia,Stereotypy,Verbigeration,Rigidity,Negativism,Waxy flexibility,Withdrawal | 97.3 | 95.7 | 98.7 |
| 10 | Excitement,Mutism,Posturing/catalepsy,Grimacing,Echopraxia/echolalia,Mannerisms,Verbigeration,Rigidity,Negativism,Withdrawal | 97.3 | 95.7 | 98.7 |
| 10 | Excitement,Mutism,Posturing/catalepsy,Grimacing,Echopraxia/echolalia,Verbigeration,Rigidity,Negativism,Waxy flexibility,Withdrawal | 97.3 | 95.7 | 98.7 |
| 10 | Excitement,Mutism,Posturing/catalepsy,Grimacing,Mannerisms,Verbigeration,Rigidity,Negativism,Waxy flexibility,Withdrawal | 97.3 | 95.7 | 98.7 |
| 10 | Excitement,Mutism,Posturing/catalepsy,Stereotypy,Mannerisms,Verbigeration,Rigidity,Negativism,Waxy flexibility,Withdrawal | 97.3 | 95.7 | 98.7 |
| 10 | Immobility/stupor,Mutism,Posturing/catalepsy,Grimacing,Echopraxia/echolalia,Stereotypy,Mannerisms,Verbigeration,Waxy flexibility,Withdrawal | 97.3 | 95.7 | 98.7 |
| 10 | Excitement,Mutism,Echopraxia/echolalia,Stereotypy,Mannerisms,Verbigeration,Rigidity,Negativism,Waxy flexibility,Withdrawal | 97.1 | 95.5 | 98.4 |
| 10 | Excitement,Mutism,Posturing/catalepsy,Grimacing,Echopraxia/echolalia,Mannerisms,Rigidity,Negativism,Waxy flexibility,Withdrawal | 97.1 | 95.5 | 98.7 |
| 10 | Excitement,Mutism,Posturing/catalepsy,Grimacing,Echopraxia/echolalia,Stereotypy,Verbigeration,Rigidity,Negativism,Withdrawal | 97.1 | 95.5 | 98.4 |
| 10 | Excitement,Mutism,Posturing/catalepsy,Grimacing,Stereotypy,Mannerisms,Verbigeration,Rigidity,Negativism,Withdrawal | 97.1 | 95.5 | 98.4 |
| 10 | Excitement,Mutism,Posturing/catalepsy,Grimacing,Stereotypy,Verbigeration,Rigidity,Negativism,Waxy flexibility,Withdrawal | 97.1 | 95.5 | 98.4 |
| 10 | Mutism,Posturing/catalepsy,Echopraxia/echolalia,Stereotypy,Mannerisms,Verbigeration,Rigidity,Negativism,Waxy flexibility,Withdrawal | 97.1 | 95.5 | 98.7 |
| 10 | Mutism,Posturing/catalepsy,Grimacing,Echopraxia/echolalia,Mannerisms,Verbigeration,Rigidity,Negativism,Waxy flexibility,Withdrawal | 97.1 | 95.5 | 98.4 |
| 10 | Mutism,Posturing/catalepsy,Grimacing,Stereotypy,Mannerisms,Verbigeration,Rigidity,Negativism,Waxy flexibility,Withdrawal | 97.1 | 95.5 | 98.4 |
| 10 | Excitement,Mutism,Grimacing,Echopraxia/echolalia,Stereotypy,Verbigeration,Rigidity,Negativism,Waxy flexibility,Withdrawal | 96.9 | 95.1 | 98.4 |
| 10 | Excitement,Mutism,Grimacing,Stereotypy,Mannerisms,Verbigeration,Rigidity,Negativism,Waxy flexibility,Withdrawal | 96.9 | 95.1 | 98.4 |
| 10 | Excitement,Mutism,Posturing/catalepsy,Echopraxia/echolalia,Stereotypy,Mannerisms,Rigidity,Negativism,Waxy flexibility,Withdrawal | 96.9 | 95.1 | 98.4 |
| 10 | Excitement,Mutism,Posturing/catalepsy,Grimacing,Echopraxia/echolalia,Stereotypy,Mannerisms,Rigidity,Negativism,Withdrawal | 96.9 | 95.1 | 98.4 |
| 10 | Excitement,Mutism,Posturing/catalepsy,Grimacing,Echopraxia/echolalia,Stereotypy,Rigidity,Negativism,Waxy flexibility,Withdrawal | 96.9 | 95.1 | 98.4 |
| 10 | Mutism,Grimacing,Echopraxia/echolalia,Stereotypy,Mannerisms,Verbigeration,Rigidity,Negativism,Waxy flexibility,Withdrawal | 96.9 | 95.1 | 98.4 |
| 10 | Mutism,Posturing/catalepsy,Grimacing,Echopraxia/echolalia,Stereotypy,Mannerisms,Rigidity,Negativism,Waxy flexibility,Withdrawal | 96.9 | 95.1 | 98.4 |
| 10 | Mutism,Posturing/catalepsy,Grimacing,Echopraxia/echolalia,Stereotypy,Mannerisms,Verbigeration,Rigidity,Negativism,Withdrawal | 96.9 | 95.1 | 98.4 |
| 10 | Excitement,Mutism,Grimacing,Echopraxia/echolalia,Mannerisms,Verbigeration,Rigidity,Negativism,Waxy flexibility,Withdrawal | 96.7 | 94.8 | 98.2 |
| 10 | Excitement,Mutism,Grimacing,Echopraxia/echolalia,Stereotypy,Mannerisms,Rigidity,Negativism,Waxy flexibility,Withdrawal | 96.7 | 94.8 | 98.2 |
| 10 | Excitement,Mutism,Grimacing,Echopraxia/echolalia,Stereotypy,Mannerisms,Verbigeration,Rigidity,Negativism,Withdrawal | 96.6 | 94.8 | 98.2 |
| 10 | Excitement,Mutism,Posturing/catalepsy,Echopraxia/echolalia,Stereotypy,Mannerisms,Verbigeration,Rigidity,Waxy flexibility,Withdrawal | 96.6 | 94.8 | 98.2 |
| 10 | Excitement,Mutism,Posturing/catalepsy,Grimacing,Echopraxia/echolalia,Mannerisms,Verbigeration,Rigidity,Negativism,Waxy flexibility | 96.6 | 94.8 | 98.2 |
| 10 | Excitement,Mutism,Posturing/catalepsy,Grimacing,Echopraxia/echolalia,Mannerisms,Verbigeration,Rigidity,Waxy flexibility,Withdrawal | 96.6 | 94.8 | 98.2 |
| 10 | Excitement,Mutism,Posturing/catalepsy,Grimacing,Stereotypy,Mannerisms,Rigidity,Negativism,Waxy flexibility,Withdrawal | 96.6 | 94.8 | 98.2 |
| 10 | Excitement,Mutism,Posturing/catalepsy,Echopraxia/echolalia,Stereotypy,Mannerisms,Verbigeration,Rigidity,Negativism,Waxy flexibility | 96.4 | 94.6 | 98.0 |
| 10 | Excitement,Mutism,Posturing/catalepsy,Grimacing,Echopraxia/echolalia,Stereotypy,Mannerisms,Verbigeration,Rigidity,Negativism | 96.4 | 94.6 | 98.0 |
| 10 | Excitement,Mutism,Posturing/catalepsy,Grimacing,Echopraxia/echolalia,Stereotypy,Verbigeration,Rigidity,Negativism,Waxy flexibility | 96.4 | 94.6 | 98.0 |
| 10 | Excitement,Mutism,Posturing/catalepsy,Grimacing,Echopraxia/echolalia,Stereotypy,Verbigeration,Rigidity,Waxy flexibility,Withdrawal | 96.4 | 94.6 | 98.0 |
| 10 | Excitement,Mutism,Posturing/catalepsy,Grimacing,Stereotypy,Mannerisms,Verbigeration,Rigidity,Waxy flexibility,Withdrawal | 96.4 | 94.6 | 98.0 |
| 10 | Mutism,Posturing/catalepsy,Grimacing,Echopraxia/echolalia,Stereotypy,Verbigeration,Rigidity,Negativism,Waxy flexibility,Withdrawal | 96.4 | 94.6 | 98.0 |
| 10 | Excitement,Mutism,Posturing/catalepsy,Grimacing,Echopraxia/echolalia,Stereotypy,Mannerisms,Rigidity,Waxy flexibility,Withdrawal | 96.2 | 94.4 | 97.8 |
| 10 | Excitement,Mutism,Posturing/catalepsy,Grimacing,Stereotypy,Mannerisms,Verbigeration,Rigidity,Negativism,Waxy flexibility | 96.2 | 94.4 | 97.8 |
| 10 | Mutism,Posturing/catalepsy,Grimacing,Echopraxia/echolalia,Stereotypy,Mannerisms,Verbigeration,Rigidity,Negativism,Waxy flexibility | 96.2 | 94.4 | 97.8 |
| 10 | Excitement,Mutism,Grimacing,Echopraxia/echolalia,Stereotypy,Mannerisms,Verbigeration,Rigidity,Negativism,Waxy flexibility | 96.0 | 94.2 | 97.8 |
| 10 | Excitement,Mutism,Posturing/catalepsy,Grimacing,Echopraxia/echolalia,Stereotypy,Mannerisms,Rigidity,Negativism,Waxy flexibility | 96.0 | 94.2 | 97.8 |
| 10 | Excitement,Posturing/catalepsy,Echopraxia/echolalia,Stereotypy,Mannerisms,Verbigeration,Rigidity,Negativism,Waxy flexibility,Withdrawal | 96.0 | 94.2 | 97.8 |
| 10 | Excitement,Posturing/catalepsy,Grimacing,Echopraxia/echolalia,Mannerisms,Verbigeration,Rigidity,Negativism,Waxy flexibility,Withdrawal | 96.0 | 93.9 | 97.8 |
| 10 | Posturing/catalepsy,Grimacing,Echopraxia/echolalia,Stereotypy,Mannerisms,Verbigeration,Rigidity,Negativism,Waxy flexibility,Withdrawal | 96.0 | 93.9 | 97.8 |
| 10 | Excitement,Mutism,Grimacing,Echopraxia/echolalia,Stereotypy,Mannerisms,Verbigeration,Rigidity,Waxy flexibility,Withdrawal | 95.7 | 93.7 | 97.5 |
| 10 | Excitement,Mutism,Posturing/catalepsy,Grimacing,Echopraxia/echolalia,Stereotypy,Mannerisms,Verbigeration,Rigidity,Withdrawal | 95.7 | 93.7 | 97.5 |
| 10 | Excitement,Posturing/catalepsy,Grimacing,Echopraxia/echolalia,Stereotypy,Mannerisms,Verbigeration,Rigidity,Negativism,Withdrawal | 95.7 | 93.7 | 97.5 |
| 10 | Excitement,Posturing/catalepsy,Grimacing,Stereotypy,Mannerisms,Verbigeration,Rigidity,Negativism,Waxy flexibility,Withdrawal | 95.7 | 93.7 | 97.5 |
| 10 | Mutism,Posturing/catalepsy,Grimacing,Echopraxia/echolalia,Stereotypy,Mannerisms,Verbigeration,Rigidity,Waxy flexibility,Withdrawal | 95.7 | 93.7 | 97.5 |
| 10 | Excitement,Mutism,Posturing/catalepsy,Echopraxia/echolalia,Stereotypy,Mannerisms,Verbigeration,Negativism,Waxy flexibility,Withdrawal | 95.5 | 93.5 | 97.3 |
| 10 | Excitement,Posturing/catalepsy,Grimacing,Echopraxia/echolalia,Stereotypy,Mannerisms,Rigidity,Negativism,Waxy flexibility,Withdrawal | 95.5 | 93.5 | 97.3 |
| 10 | Excitement,Posturing/catalepsy,Grimacing,Echopraxia/echolalia,Stereotypy,Verbigeration,Rigidity,Negativism,Waxy flexibility,Withdrawal | 95.5 | 93.5 | 97.3 |
| 10 | Excitement,Mutism,Posturing/catalepsy,Grimacing,Echopraxia/echolalia,Mannerisms,Verbigeration,Negativism,Waxy flexibility,Withdrawal | 95.3 | 93.3 | 97.1 |
| 10 | Excitement,Mutism,Posturing/catalepsy,Grimacing,Echopraxia/echolalia,Stereotypy,Mannerisms,Verbigeration,Negativism,Withdrawal | 95.3 | 93.3 | 97.1 |
| 10 | Excitement,Mutism,Posturing/catalepsy,Grimacing,Echopraxia/echolalia,Stereotypy,Mannerisms,Verbigeration,Rigidity,Waxy flexibility | 95.3 | 93.3 | 97.1 |
| 10 | Excitement,Mutism,Posturing/catalepsy,Grimacing,Echopraxia/echolalia,Stereotypy,Verbigeration,Negativism,Waxy flexibility,Withdrawal | 95.3 | 93.3 | 97.1 |
| 10 | Excitement,Mutism,Posturing/catalepsy,Grimacing,Stereotypy,Mannerisms,Verbigeration,Negativism,Waxy flexibility,Withdrawal | 95.3 | 93.3 | 97.1 |
| 10 | Mutism,Posturing/catalepsy,Grimacing,Echopraxia/echolalia,Stereotypy,Mannerisms,Verbigeration,Negativism,Waxy flexibility,Withdrawal | 95.3 | 93.3 | 97.1 |
| 10 | Excitement,Mutism,Posturing/catalepsy,Grimacing,Echopraxia/echolalia,Stereotypy,Mannerisms,Negativism,Waxy flexibility,Withdrawal | 95.1 | 93.0 | 96.9 |
| 10 | Excitement,Posturing/catalepsy,Grimacing,Echopraxia/echolalia,Stereotypy,Mannerisms,Verbigeration,Rigidity,Waxy flexibility,Withdrawal | 95.1 | 93.0 | 96.9 |
| 10 | Excitement,Grimacing,Echopraxia/echolalia,Stereotypy,Mannerisms,Verbigeration,Rigidity,Negativism,Waxy flexibility,Withdrawal | 94.2 | 91.9 | 96.2 |
| 10 | Excitement,Mutism,Posturing/catalepsy,Grimacing,Echopraxia/echolalia,Stereotypy,Mannerisms,Verbigeration,Negativism,Waxy flexibility | 94.2 | 91.9 | 96.2 |
| 10 | Excitement,Mutism,Posturing/catalepsy,Grimacing,Echopraxia/echolalia,Stereotypy,Mannerisms,Verbigeration,Waxy flexibility,Withdrawal | 94.2 | 91.9 | 96.2 |
| 10 | Excitement,Mutism,Grimacing,Echopraxia/echolalia,Stereotypy,Mannerisms,Verbigeration,Negativism,Waxy flexibility,Withdrawal | 93.9 | 91.7 | 96.0 |
| 10 | Excitement,Posturing/catalepsy,Grimacing,Echopraxia/echolalia,Stereotypy,Mannerisms,Verbigeration,Negativism,Waxy flexibility,Withdrawal | 93.7 | 91.5 | 96.0 |
| 10 | Excitement,Posturing/catalepsy,Grimacing,Echopraxia/echolalia,Stereotypy,Mannerisms,Verbigeration,Rigidity,Negativism,Waxy flexibility | 91.7 | 89.0 | 94.2 |

Table S11: sensitivity (relative to the full BFCSI) of all eleven question combinations, and the 95% confidence interval derived from bootstrapping, sorted by decreasing sensitivity.

|  | |  |  | 95% CI | |
| --- | --- | --- | --- | --- | --- |
| # of Qs | Combination | | Sensitivity | Lower | Upper |
| 11 | | Excitement,Immobility/stupor,Mutism,Posturing/catalepsy,Echopraxia/echolalia,Stereotypy,Mannerisms,Verbigeration,Rigidity,Negativism,Withdrawal | 100.0 | 100.0 | 100.0 |
| 11 | | Excitement,Immobility/stupor,Mutism,Posturing/catalepsy,Echopraxia/echolalia,Stereotypy,Verbigeration,Rigidity,Negativism,Waxy flexibility,Withdrawal | 100.0 | 100.0 | 100.0 |
| 11 | | Excitement,Immobility/stupor,Mutism,Posturing/catalepsy,Grimacing,Echopraxia/echolalia,Mannerisms,Verbigeration,Rigidity,Negativism,Withdrawal | 100.0 | 100.0 | 100.0 |
| 11 | | Excitement,Immobility/stupor,Mutism,Posturing/catalepsy,Grimacing,Echopraxia/echolalia,Stereotypy,Verbigeration,Rigidity,Negativism,Withdrawal | 100.0 | 100.0 | 100.0 |
| 11 | | Excitement,Immobility/stupor,Mutism,Posturing/catalepsy,Grimacing,Echopraxia/echolalia,Verbigeration,Rigidity,Negativism,Waxy flexibility,Withdrawal | 100.0 | 100.0 | 100.0 |
| 11 | | Excitement,Immobility/stupor,Mutism,Staring,Echopraxia/echolalia,Stereotypy,Mannerisms,Rigidity,Negativism,Waxy flexibility,Withdrawal | 100.0 | 100.0 | 100.0 |
| 11 | | Excitement,Immobility/stupor,Mutism,Staring,Echopraxia/echolalia,Stereotypy,Mannerisms,Verbigeration,Rigidity,Negativism,Waxy flexibility | 100.0 | 100.0 | 100.0 |
| 11 | | Excitement,Immobility/stupor,Mutism,Staring,Echopraxia/echolalia,Stereotypy,Mannerisms,Verbigeration,Rigidity,Negativism,Withdrawal | 100.0 | 100.0 | 100.0 |
| 11 | | Excitement,Immobility/stupor,Mutism,Staring,Echopraxia/echolalia,Stereotypy,Mannerisms,Verbigeration,Rigidity,Waxy flexibility,Withdrawal | 100.0 | 100.0 | 100.0 |
| 11 | | Excitement,Immobility/stupor,Mutism,Staring,Echopraxia/echolalia,Stereotypy,Verbigeration,Rigidity,Negativism,Waxy flexibility,Withdrawal | 100.0 | 100.0 | 100.0 |
| 11 | | Excitement,Immobility/stupor,Mutism,Staring,Grimacing,Echopraxia/echolalia,Stereotypy,Mannerisms,Rigidity,Negativism,Withdrawal | 100.0 | 100.0 | 100.0 |
| 11 | | Excitement,Immobility/stupor,Mutism,Staring,Grimacing,Echopraxia/echolalia,Stereotypy,Mannerisms,Rigidity,Waxy flexibility,Withdrawal | 100.0 | 100.0 | 100.0 |
| 11 | | Excitement,Immobility/stupor,Mutism,Staring,Grimacing,Echopraxia/echolalia,Stereotypy,Rigidity,Negativism,Waxy flexibility,Withdrawal | 100.0 | 100.0 | 100.0 |
| 11 | | Excitement,Immobility/stupor,Mutism,Staring,Grimacing,Echopraxia/echolalia,Stereotypy,Verbigeration,Rigidity,Negativism,Waxy flexibility | 100.0 | 100.0 | 100.0 |
| 11 | | Excitement,Immobility/stupor,Mutism,Staring,Grimacing,Echopraxia/echolalia,Stereotypy,Verbigeration,Rigidity,Negativism,Withdrawal | 100.0 | 100.0 | 100.0 |
| 11 | | Excitement,Immobility/stupor,Mutism,Staring,Grimacing,Echopraxia/echolalia,Stereotypy,Verbigeration,Rigidity,Waxy flexibility,Withdrawal | 100.0 | 100.0 | 100.0 |
| 11 | | Excitement,Immobility/stupor,Mutism,Staring,Grimacing,Stereotypy,Mannerisms,Verbigeration,Rigidity,Negativism,Waxy flexibility | 100.0 | 100.0 | 100.0 |
| 11 | | Excitement,Immobility/stupor,Mutism,Staring,Grimacing,Stereotypy,Mannerisms,Verbigeration,Rigidity,Negativism,Withdrawal | 100.0 | 100.0 | 100.0 |
| 11 | | Excitement,Immobility/stupor,Mutism,Staring,Grimacing,Stereotypy,Mannerisms,Verbigeration,Rigidity,Waxy flexibility,Withdrawal | 100.0 | 100.0 | 100.0 |
| 11 | | Excitement,Immobility/stupor,Mutism,Staring,Grimacing,Stereotypy,Verbigeration,Rigidity,Negativism,Waxy flexibility,Withdrawal | 100.0 | 100.0 | 100.0 |
| 11 | | Excitement,Immobility/stupor,Mutism,Staring,Posturing/catalepsy,Echopraxia/echolalia,Mannerisms,Rigidity,Negativism,Waxy flexibility,Withdrawal | 100.0 | 100.0 | 100.0 |
| 11 | | Excitement,Immobility/stupor,Mutism,Staring,Posturing/catalepsy,Echopraxia/echolalia,Mannerisms,Verbigeration,Negativism,Waxy flexibility,Withdrawal | 100.0 | 100.0 | 100.0 |
| 11 | | Excitement,Immobility/stupor,Mutism,Staring,Posturing/catalepsy,Echopraxia/echolalia,Mannerisms,Verbigeration,Rigidity,Negativism,Waxy flexibility | 100.0 | 100.0 | 100.0 |
| 11 | | Excitement,Immobility/stupor,Mutism,Staring,Posturing/catalepsy,Echopraxia/echolalia,Mannerisms,Verbigeration,Rigidity,Negativism,Withdrawal | 100.0 | 100.0 | 100.0 |
| 11 | | Excitement,Immobility/stupor,Mutism,Staring,Posturing/catalepsy,Echopraxia/echolalia,Mannerisms,Verbigeration,Rigidity,Waxy flexibility,Withdrawal | 100.0 | 100.0 | 100.0 |
| 11 | | Excitement,Immobility/stupor,Mutism,Staring,Posturing/catalepsy,Echopraxia/echolalia,Stereotypy,Mannerisms,Negativism,Waxy flexibility,Withdrawal | 100.0 | 100.0 | 100.0 |
| 11 | | Excitement,Immobility/stupor,Mutism,Staring,Posturing/catalepsy,Echopraxia/echolalia,Stereotypy,Mannerisms,Rigidity,Negativism,Withdrawal | 100.0 | 100.0 | 100.0 |
| 11 | | Excitement,Immobility/stupor,Mutism,Staring,Posturing/catalepsy,Echopraxia/echolalia,Stereotypy,Mannerisms,Rigidity,Waxy flexibility,Withdrawal | 100.0 | 100.0 | 100.0 |
| 11 | | Excitement,Immobility/stupor,Mutism,Staring,Posturing/catalepsy,Echopraxia/echolalia,Stereotypy,Mannerisms,Verbigeration,Negativism,Waxy flexibility | 100.0 | 100.0 | 100.0 |
| 11 | | Excitement,Immobility/stupor,Mutism,Staring,Posturing/catalepsy,Echopraxia/echolalia,Stereotypy,Mannerisms,Verbigeration,Negativism,Withdrawal | 100.0 | 100.0 | 100.0 |
| 11 | | Excitement,Immobility/stupor,Mutism,Staring,Posturing/catalepsy,Echopraxia/echolalia,Stereotypy,Mannerisms,Verbigeration,Rigidity,Negativism | 100.0 | 100.0 | 100.0 |
| 11 | | Excitement,Immobility/stupor,Mutism,Staring,Posturing/catalepsy,Echopraxia/echolalia,Stereotypy,Mannerisms,Verbigeration,Rigidity,Withdrawal | 100.0 | 100.0 | 100.0 |
| 11 | | Excitement,Immobility/stupor,Mutism,Staring,Posturing/catalepsy,Echopraxia/echolalia,Stereotypy,Rigidity,Negativism,Waxy flexibility,Withdrawal | 100.0 | 100.0 | 100.0 |
| 11 | | Excitement,Immobility/stupor,Mutism,Staring,Posturing/catalepsy,Echopraxia/echolalia,Stereotypy,Verbigeration,Negativism,Waxy flexibility,Withdrawal | 100.0 | 100.0 | 100.0 |
| 11 | | Excitement,Immobility/stupor,Mutism,Staring,Posturing/catalepsy,Echopraxia/echolalia,Stereotypy,Verbigeration,Rigidity,Negativism,Waxy flexibility | 100.0 | 100.0 | 100.0 |
| 11 | | Excitement,Immobility/stupor,Mutism,Staring,Posturing/catalepsy,Echopraxia/echolalia,Stereotypy,Verbigeration,Rigidity,Negativism,Withdrawal | 100.0 | 100.0 | 100.0 |
| 11 | | Excitement,Immobility/stupor,Mutism,Staring,Posturing/catalepsy,Echopraxia/echolalia,Stereotypy,Verbigeration,Rigidity,Waxy flexibility,Withdrawal | 100.0 | 100.0 | 100.0 |
| 11 | | Excitement,Immobility/stupor,Mutism,Staring,Posturing/catalepsy,Echopraxia/echolalia,Verbigeration,Rigidity,Negativism,Waxy flexibility,Withdrawal | 100.0 | 100.0 | 100.0 |
| 11 | | Excitement,Immobility/stupor,Mutism,Staring,Posturing/catalepsy,Grimacing,Echopraxia/echolalia,Mannerisms,Negativism,Waxy flexibility,Withdrawal | 100.0 | 100.0 | 100.0 |
| 11 | | Excitement,Immobility/stupor,Mutism,Staring,Posturing/catalepsy,Grimacing,Echopraxia/echolalia,Mannerisms,Rigidity,Negativism,Withdrawal | 100.0 | 100.0 | 100.0 |
| 11 | | Excitement,Immobility/stupor,Mutism,Staring,Posturing/catalepsy,Grimacing,Echopraxia/echolalia,Mannerisms,Rigidity,Waxy flexibility,Withdrawal | 100.0 | 100.0 | 100.0 |
| 11 | | Excitement,Immobility/stupor,Mutism,Staring,Posturing/catalepsy,Grimacing,Echopraxia/echolalia,Mannerisms,Verbigeration,Negativism,Waxy flexibility | 100.0 | 100.0 | 100.0 |
| 11 | | Excitement,Immobility/stupor,Mutism,Staring,Posturing/catalepsy,Grimacing,Echopraxia/echolalia,Mannerisms,Verbigeration,Negativism,Withdrawal | 100.0 | 100.0 | 100.0 |
| 11 | | Excitement,Immobility/stupor,Mutism,Staring,Posturing/catalepsy,Grimacing,Echopraxia/echolalia,Mannerisms,Verbigeration,Rigidity,Negativism | 100.0 | 100.0 | 100.0 |
| 11 | | Excitement,Immobility/stupor,Mutism,Staring,Posturing/catalepsy,Grimacing,Echopraxia/echolalia,Mannerisms,Verbigeration,Rigidity,Withdrawal | 100.0 | 100.0 | 100.0 |
| 11 | | Excitement,Immobility/stupor,Mutism,Staring,Posturing/catalepsy,Grimacing,Echopraxia/echolalia,Rigidity,Negativism,Waxy flexibility,Withdrawal | 100.0 | 100.0 | 100.0 |
| 11 | | Excitement,Immobility/stupor,Mutism,Staring,Posturing/catalepsy,Grimacing,Echopraxia/echolalia,Stereotypy,Mannerisms,Negativism,Withdrawal | 100.0 | 100.0 | 100.0 |
| 11 | | Excitement,Immobility/stupor,Mutism,Staring,Posturing/catalepsy,Grimacing,Echopraxia/echolalia,Stereotypy,Mannerisms,Rigidity,Withdrawal | 100.0 | 100.0 | 100.0 |
| 11 | | Excitement,Immobility/stupor,Mutism,Staring,Posturing/catalepsy,Grimacing,Echopraxia/echolalia,Stereotypy,Negativism,Waxy flexibility,Withdrawal | 100.0 | 100.0 | 100.0 |
| 11 | | Excitement,Immobility/stupor,Mutism,Staring,Posturing/catalepsy,Grimacing,Echopraxia/echolalia,Stereotypy,Rigidity,Negativism,Withdrawal | 100.0 | 100.0 | 100.0 |
| 11 | | Excitement,Immobility/stupor,Mutism,Staring,Posturing/catalepsy,Grimacing,Echopraxia/echolalia,Stereotypy,Rigidity,Waxy flexibility,Withdrawal | 100.0 | 100.0 | 100.0 |
| 11 | | Excitement,Immobility/stupor,Mutism,Staring,Posturing/catalepsy,Grimacing,Echopraxia/echolalia,Stereotypy,Verbigeration,Negativism,Waxy flexibility | 100.0 | 100.0 | 100.0 |
| 11 | | Excitement,Immobility/stupor,Mutism,Staring,Posturing/catalepsy,Grimacing,Echopraxia/echolalia,Stereotypy,Verbigeration,Negativism,Withdrawal | 100.0 | 100.0 | 100.0 |
| 11 | | Excitement,Immobility/stupor,Mutism,Staring,Posturing/catalepsy,Grimacing,Echopraxia/echolalia,Stereotypy,Verbigeration,Rigidity,Negativism | 100.0 | 100.0 | 100.0 |
| 11 | | Excitement,Immobility/stupor,Mutism,Staring,Posturing/catalepsy,Grimacing,Echopraxia/echolalia,Stereotypy,Verbigeration,Rigidity,Withdrawal | 100.0 | 100.0 | 100.0 |
| 11 | | Excitement,Immobility/stupor,Mutism,Staring,Posturing/catalepsy,Grimacing,Echopraxia/echolalia,Verbigeration,Negativism,Waxy flexibility,Withdrawal | 100.0 | 100.0 | 100.0 |
| 11 | | Excitement,Immobility/stupor,Mutism,Staring,Posturing/catalepsy,Grimacing,Echopraxia/echolalia,Verbigeration,Rigidity,Negativism,Waxy flexibility | 100.0 | 100.0 | 100.0 |
| 11 | | Excitement,Immobility/stupor,Mutism,Staring,Posturing/catalepsy,Grimacing,Echopraxia/echolalia,Verbigeration,Rigidity,Negativism,Withdrawal | 100.0 | 100.0 | 100.0 |
| 11 | | Excitement,Immobility/stupor,Mutism,Staring,Posturing/catalepsy,Grimacing,Echopraxia/echolalia,Verbigeration,Rigidity,Waxy flexibility,Withdrawal | 100.0 | 100.0 | 100.0 |
| 11 | | Excitement,Immobility/stupor,Mutism,Staring,Posturing/catalepsy,Grimacing,Mannerisms,Verbigeration,Negativism,Waxy flexibility,Withdrawal | 100.0 | 100.0 | 100.0 |
| 11 | | Excitement,Immobility/stupor,Mutism,Staring,Posturing/catalepsy,Grimacing,Mannerisms,Verbigeration,Rigidity,Negativism,Waxy flexibility | 100.0 | 100.0 | 100.0 |
| 11 | | Excitement,Immobility/stupor,Mutism,Staring,Posturing/catalepsy,Grimacing,Mannerisms,Verbigeration,Rigidity,Negativism,Withdrawal | 100.0 | 100.0 | 100.0 |
| 11 | | Excitement,Immobility/stupor,Mutism,Staring,Posturing/catalepsy,Grimacing,Mannerisms,Verbigeration,Rigidity,Waxy flexibility,Withdrawal | 100.0 | 100.0 | 100.0 |
| 11 | | Excitement,Immobility/stupor,Mutism,Staring,Posturing/catalepsy,Grimacing,Stereotypy,Mannerisms,Verbigeration,Negativism,Waxy flexibility | 100.0 | 100.0 | 100.0 |
| 11 | | Excitement,Immobility/stupor,Mutism,Staring,Posturing/catalepsy,Grimacing,Stereotypy,Mannerisms,Verbigeration,Negativism,Withdrawal | 100.0 | 100.0 | 100.0 |
| 11 | | Excitement,Immobility/stupor,Mutism,Staring,Posturing/catalepsy,Grimacing,Stereotypy,Mannerisms,Verbigeration,Rigidity,Negativism | 100.0 | 100.0 | 100.0 |
| 11 | | Excitement,Immobility/stupor,Mutism,Staring,Posturing/catalepsy,Grimacing,Stereotypy,Mannerisms,Verbigeration,Rigidity,Withdrawal | 100.0 | 100.0 | 100.0 |
| 11 | | Excitement,Immobility/stupor,Mutism,Staring,Posturing/catalepsy,Grimacing,Stereotypy,Verbigeration,Negativism,Waxy flexibility,Withdrawal | 100.0 | 100.0 | 100.0 |
| 11 | | Excitement,Immobility/stupor,Mutism,Staring,Posturing/catalepsy,Grimacing,Stereotypy,Verbigeration,Rigidity,Negativism,Waxy flexibility | 100.0 | 100.0 | 100.0 |
| 11 | | Excitement,Immobility/stupor,Mutism,Staring,Posturing/catalepsy,Grimacing,Stereotypy,Verbigeration,Rigidity,Negativism,Withdrawal | 100.0 | 100.0 | 100.0 |
| 11 | | Excitement,Immobility/stupor,Mutism,Staring,Posturing/catalepsy,Grimacing,Stereotypy,Verbigeration,Rigidity,Waxy flexibility,Withdrawal | 100.0 | 100.0 | 100.0 |
| 11 | | Excitement,Immobility/stupor,Mutism,Staring,Posturing/catalepsy,Mannerisms,Verbigeration,Rigidity,Negativism,Waxy flexibility,Withdrawal | 100.0 | 100.0 | 100.0 |
| 11 | | Excitement,Immobility/stupor,Mutism,Staring,Posturing/catalepsy,Stereotypy,Mannerisms,Verbigeration,Negativism,Waxy flexibility,Withdrawal | 100.0 | 100.0 | 100.0 |
| 11 | | Excitement,Immobility/stupor,Mutism,Staring,Posturing/catalepsy,Stereotypy,Mannerisms,Verbigeration,Rigidity,Negativism,Waxy flexibility | 100.0 | 100.0 | 100.0 |
| 11 | | Excitement,Immobility/stupor,Mutism,Staring,Posturing/catalepsy,Stereotypy,Mannerisms,Verbigeration,Rigidity,Negativism,Withdrawal | 100.0 | 100.0 | 100.0 |
| 11 | | Excitement,Immobility/stupor,Mutism,Staring,Posturing/catalepsy,Stereotypy,Mannerisms,Verbigeration,Rigidity,Waxy flexibility,Withdrawal | 100.0 | 100.0 | 100.0 |
| 11 | | Excitement,Immobility/stupor,Mutism,Staring,Posturing/catalepsy,Stereotypy,Verbigeration,Rigidity,Negativism,Waxy flexibility,Withdrawal | 100.0 | 100.0 | 100.0 |
| 11 | | Excitement,Immobility/stupor,Mutism,Staring,Stereotypy,Mannerisms,Verbigeration,Rigidity,Negativism,Waxy flexibility,Withdrawal | 100.0 | 100.0 | 100.0 |
| 11 | | Excitement,Immobility/stupor,Staring,Echopraxia/echolalia,Stereotypy,Mannerisms,Verbigeration,Rigidity,Negativism,Waxy flexibility,Withdrawal | 100.0 | 100.0 | 100.0 |
| 11 | | Excitement,Immobility/stupor,Staring,Grimacing,Echopraxia/echolalia,Stereotypy,Mannerisms,Rigidity,Negativism,Waxy flexibility,Withdrawal | 100.0 | 100.0 | 100.0 |
| 11 | | Excitement,Immobility/stupor,Staring,Grimacing,Echopraxia/echolalia,Stereotypy,Mannerisms,Verbigeration,Rigidity,Negativism,Withdrawal | 100.0 | 100.0 | 100.0 |
| 11 | | Excitement,Immobility/stupor,Staring,Grimacing,Stereotypy,Mannerisms,Verbigeration,Rigidity,Negativism,Waxy flexibility,Withdrawal | 100.0 | 100.0 | 100.0 |
| 11 | | Excitement,Immobility/stupor,Staring,Posturing/catalepsy,Echopraxia/echolalia,Mannerisms,Verbigeration,Rigidity,Negativism,Waxy flexibility,Withdrawal | 100.0 | 100.0 | 100.0 |
| 11 | | Excitement,Immobility/stupor,Staring,Posturing/catalepsy,Echopraxia/echolalia,Stereotypy,Mannerisms,Rigidity,Negativism,Waxy flexibility,Withdrawal | 100.0 | 100.0 | 100.0 |
| 11 | | Excitement,Immobility/stupor,Staring,Posturing/catalepsy,Echopraxia/echolalia,Stereotypy,Mannerisms,Verbigeration,Rigidity,Negativism,Withdrawal | 100.0 | 100.0 | 100.0 |
| 11 | | Excitement,Immobility/stupor,Staring,Posturing/catalepsy,Grimacing,Echopraxia/echolalia,Mannerisms,Rigidity,Negativism,Waxy flexibility,Withdrawal | 100.0 | 100.0 | 100.0 |
| 11 | | Excitement,Immobility/stupor,Staring,Posturing/catalepsy,Grimacing,Echopraxia/echolalia,Mannerisms,Verbigeration,Rigidity,Negativism,Withdrawal | 100.0 | 100.0 | 100.0 |
| 11 | | Excitement,Immobility/stupor,Staring,Posturing/catalepsy,Grimacing,Echopraxia/echolalia,Stereotypy,Mannerisms,Rigidity,Negativism,Withdrawal | 100.0 | 100.0 | 100.0 |
| 11 | | Excitement,Immobility/stupor,Staring,Posturing/catalepsy,Grimacing,Mannerisms,Verbigeration,Rigidity,Negativism,Waxy flexibility,Withdrawal | 100.0 | 100.0 | 100.0 |
| 11 | | Excitement,Immobility/stupor,Staring,Posturing/catalepsy,Grimacing,Stereotypy,Mannerisms,Verbigeration,Rigidity,Negativism,Withdrawal | 100.0 | 100.0 | 100.0 |
| 11 | | Excitement,Immobility/stupor,Staring,Posturing/catalepsy,Stereotypy,Mannerisms,Verbigeration,Rigidity,Negativism,Waxy flexibility,Withdrawal | 100.0 | 100.0 | 100.0 |
| 11 | | Excitement,Mutism,Staring,Echopraxia/echolalia,Stereotypy,Mannerisms,Verbigeration,Rigidity,Negativism,Waxy flexibility,Withdrawal | 100.0 | 100.0 | 100.0 |
| 11 | | Excitement,Mutism,Staring,Grimacing,Echopraxia/echolalia,Stereotypy,Mannerisms,Rigidity,Negativism,Waxy flexibility,Withdrawal | 100.0 | 100.0 | 100.0 |
| 11 | | Excitement,Mutism,Staring,Grimacing,Echopraxia/echolalia,Stereotypy,Mannerisms,Verbigeration,Rigidity,Negativism,Waxy flexibility | 100.0 | 100.0 | 100.0 |
| 11 | | Excitement,Mutism,Staring,Grimacing,Echopraxia/echolalia,Stereotypy,Mannerisms,Verbigeration,Rigidity,Negativism,Withdrawal | 100.0 | 100.0 | 100.0 |
| 11 | | Excitement,Mutism,Staring,Grimacing,Echopraxia/echolalia,Stereotypy,Mannerisms,Verbigeration,Rigidity,Waxy flexibility,Withdrawal | 100.0 | 100.0 | 100.0 |
| 11 | | Excitement,Mutism,Staring,Grimacing,Echopraxia/echolalia,Stereotypy,Verbigeration,Rigidity,Negativism,Waxy flexibility,Withdrawal | 100.0 | 100.0 | 100.0 |
| 11 | | Excitement,Mutism,Staring,Grimacing,Stereotypy,Mannerisms,Verbigeration,Rigidity,Negativism,Waxy flexibility,Withdrawal | 100.0 | 100.0 | 100.0 |
| 11 | | Excitement,Mutism,Staring,Posturing/catalepsy,Echopraxia/echolalia,Mannerisms,Verbigeration,Rigidity,Negativism,Waxy flexibility,Withdrawal | 100.0 | 100.0 | 100.0 |
| 11 | | Excitement,Mutism,Staring,Posturing/catalepsy,Echopraxia/echolalia,Stereotypy,Mannerisms,Rigidity,Negativism,Waxy flexibility,Withdrawal | 100.0 | 100.0 | 100.0 |
| 11 | | Excitement,Mutism,Staring,Posturing/catalepsy,Echopraxia/echolalia,Stereotypy,Mannerisms,Verbigeration,Negativism,Waxy flexibility,Withdrawal | 100.0 | 100.0 | 100.0 |
| 11 | | Excitement,Mutism,Staring,Posturing/catalepsy,Echopraxia/echolalia,Stereotypy,Mannerisms,Verbigeration,Rigidity,Negativism,Waxy flexibility | 100.0 | 100.0 | 100.0 |
| 11 | | Excitement,Mutism,Staring,Posturing/catalepsy,Echopraxia/echolalia,Stereotypy,Mannerisms,Verbigeration,Rigidity,Negativism,Withdrawal | 100.0 | 100.0 | 100.0 |
| 11 | | Excitement,Mutism,Staring,Posturing/catalepsy,Echopraxia/echolalia,Stereotypy,Mannerisms,Verbigeration,Rigidity,Waxy flexibility,Withdrawal | 100.0 | 100.0 | 100.0 |
| 11 | | Excitement,Mutism,Staring,Posturing/catalepsy,Echopraxia/echolalia,Stereotypy,Verbigeration,Rigidity,Negativism,Waxy flexibility,Withdrawal | 100.0 | 100.0 | 100.0 |
| 11 | | Excitement,Mutism,Staring,Posturing/catalepsy,Grimacing,Echopraxia/echolalia,Mannerisms,Rigidity,Negativism,Waxy flexibility,Withdrawal | 100.0 | 100.0 | 100.0 |
| 11 | | Excitement,Mutism,Staring,Posturing/catalepsy,Grimacing,Echopraxia/echolalia,Mannerisms,Verbigeration,Negativism,Waxy flexibility,Withdrawal | 100.0 | 100.0 | 100.0 |
| 11 | | Excitement,Mutism,Staring,Posturing/catalepsy,Grimacing,Echopraxia/echolalia,Mannerisms,Verbigeration,Rigidity,Negativism,Waxy flexibility | 100.0 | 100.0 | 100.0 |
| 11 | | Excitement,Mutism,Staring,Posturing/catalepsy,Grimacing,Echopraxia/echolalia,Mannerisms,Verbigeration,Rigidity,Negativism,Withdrawal | 100.0 | 100.0 | 100.0 |
| 11 | | Excitement,Mutism,Staring,Posturing/catalepsy,Grimacing,Echopraxia/echolalia,Mannerisms,Verbigeration,Rigidity,Waxy flexibility,Withdrawal | 100.0 | 100.0 | 100.0 |
| 11 | | Excitement,Mutism,Staring,Posturing/catalepsy,Grimacing,Echopraxia/echolalia,Stereotypy,Mannerisms,Negativism,Waxy flexibility,Withdrawal | 100.0 | 100.0 | 100.0 |
| 11 | | Excitement,Mutism,Staring,Posturing/catalepsy,Grimacing,Echopraxia/echolalia,Stereotypy,Mannerisms,Rigidity,Negativism,Withdrawal | 100.0 | 100.0 | 100.0 |
| 11 | | Excitement,Mutism,Staring,Posturing/catalepsy,Grimacing,Echopraxia/echolalia,Stereotypy,Mannerisms,Rigidity,Waxy flexibility,Withdrawal | 100.0 | 100.0 | 100.0 |
| 11 | | Excitement,Mutism,Staring,Posturing/catalepsy,Grimacing,Echopraxia/echolalia,Stereotypy,Mannerisms,Verbigeration,Negativism,Waxy flexibility | 100.0 | 100.0 | 100.0 |
| 11 | | Excitement,Mutism,Staring,Posturing/catalepsy,Grimacing,Echopraxia/echolalia,Stereotypy,Mannerisms,Verbigeration,Negativism,Withdrawal | 100.0 | 100.0 | 100.0 |
| 11 | | Excitement,Mutism,Staring,Posturing/catalepsy,Grimacing,Echopraxia/echolalia,Stereotypy,Mannerisms,Verbigeration,Rigidity,Negativism | 100.0 | 100.0 | 100.0 |
| 11 | | Excitement,Mutism,Staring,Posturing/catalepsy,Grimacing,Echopraxia/echolalia,Stereotypy,Mannerisms,Verbigeration,Rigidity,Withdrawal | 100.0 | 100.0 | 100.0 |
| 11 | | Excitement,Mutism,Staring,Posturing/catalepsy,Grimacing,Echopraxia/echolalia,Stereotypy,Rigidity,Negativism,Waxy flexibility,Withdrawal | 100.0 | 100.0 | 100.0 |
| 11 | | Excitement,Mutism,Staring,Posturing/catalepsy,Grimacing,Echopraxia/echolalia,Stereotypy,Verbigeration,Negativism,Waxy flexibility,Withdrawal | 100.0 | 100.0 | 100.0 |
| 11 | | Excitement,Mutism,Staring,Posturing/catalepsy,Grimacing,Echopraxia/echolalia,Stereotypy,Verbigeration,Rigidity,Negativism,Waxy flexibility | 100.0 | 100.0 | 100.0 |
| 11 | | Excitement,Mutism,Staring,Posturing/catalepsy,Grimacing,Echopraxia/echolalia,Stereotypy,Verbigeration,Rigidity,Negativism,Withdrawal | 100.0 | 100.0 | 100.0 |
| 11 | | Excitement,Mutism,Staring,Posturing/catalepsy,Grimacing,Echopraxia/echolalia,Stereotypy,Verbigeration,Rigidity,Waxy flexibility,Withdrawal | 100.0 | 100.0 | 100.0 |
| 11 | | Excitement,Mutism,Staring,Posturing/catalepsy,Grimacing,Echopraxia/echolalia,Verbigeration,Rigidity,Negativism,Waxy flexibility,Withdrawal | 100.0 | 100.0 | 100.0 |
| 11 | | Excitement,Mutism,Staring,Posturing/catalepsy,Grimacing,Mannerisms,Verbigeration,Rigidity,Negativism,Waxy flexibility,Withdrawal | 100.0 | 100.0 | 100.0 |
| 11 | | Excitement,Mutism,Staring,Posturing/catalepsy,Grimacing,Stereotypy,Mannerisms,Verbigeration,Negativism,Waxy flexibility,Withdrawal | 100.0 | 100.0 | 100.0 |
| 11 | | Excitement,Mutism,Staring,Posturing/catalepsy,Grimacing,Stereotypy,Mannerisms,Verbigeration,Rigidity,Negativism,Withdrawal | 100.0 | 100.0 | 100.0 |
| 11 | | Excitement,Mutism,Staring,Posturing/catalepsy,Grimacing,Stereotypy,Mannerisms,Verbigeration,Rigidity,Waxy flexibility,Withdrawal | 100.0 | 100.0 | 100.0 |
| 11 | | Excitement,Mutism,Staring,Posturing/catalepsy,Grimacing,Stereotypy,Verbigeration,Rigidity,Negativism,Waxy flexibility,Withdrawal | 100.0 | 100.0 | 100.0 |
| 11 | | Excitement,Mutism,Staring,Posturing/catalepsy,Stereotypy,Mannerisms,Verbigeration,Rigidity,Negativism,Waxy flexibility,Withdrawal | 100.0 | 100.0 | 100.0 |
| 11 | | Immobility/stupor,Mutism,Staring,Grimacing,Echopraxia/echolalia,Stereotypy,Mannerisms,Rigidity,Negativism,Waxy flexibility,Withdrawal | 100.0 | 100.0 | 100.0 |
| 11 | | Immobility/stupor,Mutism,Staring,Grimacing,Echopraxia/echolalia,Stereotypy,Mannerisms,Verbigeration,Rigidity,Negativism,Withdrawal | 100.0 | 100.0 | 100.0 |
| 11 | | Immobility/stupor,Mutism,Staring,Grimacing,Stereotypy,Mannerisms,Verbigeration,Rigidity,Negativism,Waxy flexibility,Withdrawal | 100.0 | 100.0 | 100.0 |
| 11 | | Immobility/stupor,Mutism,Staring,Posturing/catalepsy,Grimacing,Echopraxia/echolalia,Stereotypy,Mannerisms,Negativism,Waxy flexibility,Withdrawal | 100.0 | 100.0 | 100.0 |
| 11 | | Immobility/stupor,Mutism,Staring,Posturing/catalepsy,Grimacing,Echopraxia/echolalia,Stereotypy,Mannerisms,Rigidity,Negativism,Withdrawal | 100.0 | 100.0 | 100.0 |
| 11 | | Immobility/stupor,Mutism,Staring,Posturing/catalepsy,Grimacing,Echopraxia/echolalia,Stereotypy,Mannerisms,Verbigeration,Negativism,Withdrawal | 100.0 | 100.0 | 100.0 |
| 11 | | Immobility/stupor,Mutism,Staring,Posturing/catalepsy,Grimacing,Stereotypy,Mannerisms,Verbigeration,Negativism,Waxy flexibility,Withdrawal | 100.0 | 100.0 | 100.0 |
| 11 | | Immobility/stupor,Mutism,Staring,Posturing/catalepsy,Grimacing,Stereotypy,Mannerisms,Verbigeration,Rigidity,Negativism,Withdrawal | 100.0 | 100.0 | 100.0 |
| 11 | | Immobility/stupor,Staring,Grimacing,Echopraxia/echolalia,Stereotypy,Mannerisms,Verbigeration,Rigidity,Negativism,Waxy flexibility,Withdrawal | 100.0 | 100.0 | 100.0 |
| 11 | | Immobility/stupor,Staring,Posturing/catalepsy,Grimacing,Echopraxia/echolalia,Stereotypy,Mannerisms,Rigidity,Negativism,Waxy flexibility,Withdrawal | 100.0 | 100.0 | 100.0 |
| 11 | | Immobility/stupor,Staring,Posturing/catalepsy,Grimacing,Echopraxia/echolalia,Stereotypy,Mannerisms,Verbigeration,Rigidity,Negativism,Withdrawal | 100.0 | 100.0 | 100.0 |
| 11 | | Immobility/stupor,Staring,Posturing/catalepsy,Grimacing,Stereotypy,Mannerisms,Verbigeration,Rigidity,Negativism,Waxy flexibility,Withdrawal | 100.0 | 100.0 | 100.0 |
| 11 | | Mutism,Staring,Grimacing,Echopraxia/echolalia,Stereotypy,Mannerisms,Verbigeration,Rigidity,Negativism,Waxy flexibility,Withdrawal | 100.0 | 100.0 | 100.0 |
| 11 | | Mutism,Staring,Posturing/catalepsy,Grimacing,Echopraxia/echolalia,Stereotypy,Mannerisms,Rigidity,Negativism,Waxy flexibility,Withdrawal | 100.0 | 100.0 | 100.0 |
| 11 | | Mutism,Staring,Posturing/catalepsy,Grimacing,Echopraxia/echolalia,Stereotypy,Mannerisms,Verbigeration,Negativism,Waxy flexibility,Withdrawal | 100.0 | 100.0 | 100.0 |
| 11 | | Mutism,Staring,Posturing/catalepsy,Grimacing,Echopraxia/echolalia,Stereotypy,Mannerisms,Verbigeration,Rigidity,Negativism,Withdrawal | 100.0 | 100.0 | 100.0 |
| 11 | | Mutism,Staring,Posturing/catalepsy,Grimacing,Stereotypy,Mannerisms,Verbigeration,Rigidity,Negativism,Waxy flexibility,Withdrawal | 100.0 | 100.0 | 100.0 |
| 11 | | Excitement,Immobility/stupor,Mutism,Echopraxia/echolalia,Stereotypy,Mannerisms,Verbigeration,Rigidity,Negativism,Waxy flexibility,Withdrawal | 99.8 | 99.3 | 100.0 |
| 11 | | Excitement,Immobility/stupor,Mutism,Grimacing,Echopraxia/echolalia,Stereotypy,Mannerisms,Verbigeration,Rigidity,Negativism,Withdrawal | 99.8 | 99.3 | 100.0 |
| 11 | | Excitement,Immobility/stupor,Mutism,Grimacing,Echopraxia/echolalia,Stereotypy,Verbigeration,Rigidity,Negativism,Waxy flexibility,Withdrawal | 99.8 | 99.3 | 100.0 |
| 11 | | Excitement,Immobility/stupor,Mutism,Posturing/catalepsy,Echopraxia/echolalia,Mannerisms,Verbigeration,Rigidity,Negativism,Waxy flexibility,Withdrawal | 99.8 | 99.3 | 100.0 |
| 11 | | Excitement,Immobility/stupor,Mutism,Posturing/catalepsy,Echopraxia/echolalia,Stereotypy,Mannerisms,Verbigeration,Rigidity,Negativism,Waxy flexibility | 99.8 | 99.3 | 100.0 |
| 11 | | Excitement,Immobility/stupor,Mutism,Posturing/catalepsy,Grimacing,Echopraxia/echolalia,Mannerisms,Rigidity,Negativism,Waxy flexibility,Withdrawal | 99.8 | 99.3 | 100.0 |
| 11 | | Excitement,Immobility/stupor,Mutism,Posturing/catalepsy,Grimacing,Echopraxia/echolalia,Mannerisms,Verbigeration,Rigidity,Negativism,Waxy flexibility | 99.8 | 99.3 | 100.0 |
| 11 | | Excitement,Immobility/stupor,Mutism,Posturing/catalepsy,Grimacing,Echopraxia/echolalia,Stereotypy,Mannerisms,Rigidity,Negativism,Withdrawal | 99.8 | 99.3 | 100.0 |
| 11 | | Excitement,Immobility/stupor,Mutism,Posturing/catalepsy,Grimacing,Echopraxia/echolalia,Stereotypy,Mannerisms,Verbigeration,Rigidity,Negativism | 99.8 | 99.3 | 100.0 |
| 11 | | Excitement,Immobility/stupor,Mutism,Posturing/catalepsy,Grimacing,Echopraxia/echolalia,Stereotypy,Rigidity,Negativism,Waxy flexibility,Withdrawal | 99.8 | 99.3 | 100.0 |
| 11 | | Excitement,Immobility/stupor,Mutism,Posturing/catalepsy,Grimacing,Echopraxia/echolalia,Stereotypy,Verbigeration,Rigidity,Negativism,Waxy flexibility | 99.8 | 99.3 | 100.0 |
| 11 | | Excitement,Immobility/stupor,Mutism,Posturing/catalepsy,Grimacing,Mannerisms,Verbigeration,Rigidity,Negativism,Waxy flexibility,Withdrawal | 99.8 | 99.3 | 100.0 |
| 11 | | Excitement,Immobility/stupor,Mutism,Posturing/catalepsy,Grimacing,Stereotypy,Mannerisms,Verbigeration,Rigidity,Negativism,Withdrawal | 99.8 | 99.3 | 100.0 |
| 11 | | Excitement,Immobility/stupor,Mutism,Posturing/catalepsy,Grimacing,Stereotypy,Verbigeration,Rigidity,Negativism,Waxy flexibility,Withdrawal | 99.8 | 99.3 | 100.0 |
| 11 | | Excitement,Immobility/stupor,Mutism,Posturing/catalepsy,Stereotypy,Mannerisms,Verbigeration,Rigidity,Negativism,Waxy flexibility,Withdrawal | 99.8 | 99.3 | 100.0 |
| 11 | | Excitement,Immobility/stupor,Mutism,Staring,Echopraxia/echolalia,Mannerisms,Verbigeration,Rigidity,Negativism,Waxy flexibility,Withdrawal | 99.8 | 99.3 | 100.0 |
| 11 | | Excitement,Immobility/stupor,Mutism,Staring,Grimacing,Echopraxia/echolalia,Mannerisms,Rigidity,Negativism,Waxy flexibility,Withdrawal | 99.8 | 99.3 | 100.0 |
| 11 | | Excitement,Immobility/stupor,Mutism,Staring,Grimacing,Echopraxia/echolalia,Mannerisms,Verbigeration,Rigidity,Negativism,Waxy flexibility | 99.8 | 99.3 | 100.0 |
| 11 | | Excitement,Immobility/stupor,Mutism,Staring,Grimacing,Echopraxia/echolalia,Mannerisms,Verbigeration,Rigidity,Negativism,Withdrawal | 99.8 | 99.3 | 100.0 |
| 11 | | Excitement,Immobility/stupor,Mutism,Staring,Grimacing,Echopraxia/echolalia,Mannerisms,Verbigeration,Rigidity,Waxy flexibility,Withdrawal | 99.8 | 99.3 | 100.0 |
| 11 | | Excitement,Immobility/stupor,Mutism,Staring,Grimacing,Echopraxia/echolalia,Stereotypy,Mannerisms,Rigidity,Negativism,Waxy flexibility | 99.8 | 99.3 | 100.0 |
| 11 | | Excitement,Immobility/stupor,Mutism,Staring,Grimacing,Echopraxia/echolalia,Stereotypy,Mannerisms,Verbigeration,Rigidity,Negativism | 99.8 | 99.3 | 100.0 |
| 11 | | Excitement,Immobility/stupor,Mutism,Staring,Grimacing,Echopraxia/echolalia,Stereotypy,Mannerisms,Verbigeration,Rigidity,Waxy flexibility | 99.8 | 99.3 | 100.0 |
| 11 | | Excitement,Immobility/stupor,Mutism,Staring,Grimacing,Echopraxia/echolalia,Stereotypy,Mannerisms,Verbigeration,Rigidity,Withdrawal | 99.8 | 99.3 | 100.0 |
| 11 | | Excitement,Immobility/stupor,Mutism,Staring,Grimacing,Echopraxia/echolalia,Verbigeration,Rigidity,Negativism,Waxy flexibility,Withdrawal | 99.8 | 99.3 | 100.0 |
| 11 | | Excitement,Immobility/stupor,Mutism,Staring,Grimacing,Mannerisms,Verbigeration,Rigidity,Negativism,Waxy flexibility,Withdrawal | 99.8 | 99.3 | 100.0 |
| 11 | | Excitement,Immobility/stupor,Mutism,Staring,Grimacing,Stereotypy,Mannerisms,Rigidity,Negativism,Waxy flexibility,Withdrawal | 99.8 | 99.3 | 100.0 |
| 11 | | Excitement,Immobility/stupor,Mutism,Staring,Posturing/catalepsy,Echopraxia/echolalia,Stereotypy,Mannerisms,Rigidity,Negativism,Waxy flexibility | 99.8 | 99.3 | 100.0 |
| 11 | | Excitement,Immobility/stupor,Mutism,Staring,Posturing/catalepsy,Echopraxia/echolalia,Stereotypy,Mannerisms,Verbigeration,Rigidity,Waxy flexibility | 99.8 | 99.3 | 100.0 |
| 11 | | Excitement,Immobility/stupor,Mutism,Staring,Posturing/catalepsy,Grimacing,Echopraxia/echolalia,Mannerisms,Rigidity,Negativism,Waxy flexibility | 99.8 | 99.3 | 100.0 |
| 11 | | Excitement,Immobility/stupor,Mutism,Staring,Posturing/catalepsy,Grimacing,Echopraxia/echolalia,Mannerisms,Verbigeration,Rigidity,Waxy flexibility | 99.8 | 99.3 | 100.0 |
| 11 | | Excitement,Immobility/stupor,Mutism,Staring,Posturing/catalepsy,Grimacing,Echopraxia/echolalia,Stereotypy,Mannerisms,Negativism,Waxy flexibility | 99.8 | 99.3 | 100.0 |
| 11 | | Excitement,Immobility/stupor,Mutism,Staring,Posturing/catalepsy,Grimacing,Echopraxia/echolalia,Stereotypy,Mannerisms,Rigidity,Negativism | 99.8 | 99.3 | 100.0 |
| 11 | | Excitement,Immobility/stupor,Mutism,Staring,Posturing/catalepsy,Grimacing,Echopraxia/echolalia,Stereotypy,Mannerisms,Verbigeration,Negativism | 99.8 | 99.3 | 100.0 |
| 11 | | Excitement,Immobility/stupor,Mutism,Staring,Posturing/catalepsy,Grimacing,Echopraxia/echolalia,Stereotypy,Mannerisms,Verbigeration,Rigidity | 99.8 | 99.3 | 100.0 |
| 11 | | Excitement,Immobility/stupor,Mutism,Staring,Posturing/catalepsy,Grimacing,Echopraxia/echolalia,Stereotypy,Rigidity,Negativism,Waxy flexibility | 99.8 | 99.3 | 100.0 |
| 11 | | Excitement,Immobility/stupor,Mutism,Staring,Posturing/catalepsy,Grimacing,Echopraxia/echolalia,Stereotypy,Verbigeration,Rigidity,Waxy flexibility | 99.8 | 99.3 | 100.0 |
| 11 | | Excitement,Immobility/stupor,Mutism,Staring,Posturing/catalepsy,Grimacing,Mannerisms,Rigidity,Negativism,Waxy flexibility,Withdrawal | 99.8 | 99.3 | 100.0 |
| 11 | | Excitement,Immobility/stupor,Mutism,Staring,Posturing/catalepsy,Grimacing,Stereotypy,Mannerisms,Negativism,Waxy flexibility,Withdrawal | 99.8 | 99.3 | 100.0 |
| 11 | | Excitement,Immobility/stupor,Mutism,Staring,Posturing/catalepsy,Grimacing,Stereotypy,Mannerisms,Rigidity,Negativism,Withdrawal | 99.8 | 99.3 | 100.0 |
| 11 | | Excitement,Immobility/stupor,Mutism,Staring,Posturing/catalepsy,Grimacing,Stereotypy,Mannerisms,Rigidity,Waxy flexibility,Withdrawal | 99.8 | 99.3 | 100.0 |
| 11 | | Excitement,Immobility/stupor,Mutism,Staring,Posturing/catalepsy,Grimacing,Stereotypy,Mannerisms,Verbigeration,Rigidity,Waxy flexibility | 99.8 | 99.3 | 100.0 |
| 11 | | Excitement,Immobility/stupor,Mutism,Staring,Posturing/catalepsy,Grimacing,Stereotypy,Rigidity,Negativism,Waxy flexibility,Withdrawal | 99.8 | 99.3 | 100.0 |
| 11 | | Excitement,Immobility/stupor,Mutism,Staring,Posturing/catalepsy,Grimacing,Verbigeration,Rigidity,Negativism,Waxy flexibility,Withdrawal | 99.8 | 99.3 | 100.0 |
| 11 | | Excitement,Immobility/stupor,Mutism,Staring,Posturing/catalepsy,Stereotypy,Mannerisms,Rigidity,Negativism,Waxy flexibility,Withdrawal | 99.8 | 99.3 | 100.0 |
| 11 | | Excitement,Immobility/stupor,Posturing/catalepsy,Echopraxia/echolalia,Stereotypy,Mannerisms,Verbigeration,Rigidity,Negativism,Waxy flexibility,Withdrawal | 99.8 | 99.3 | 100.0 |
| 11 | | Excitement,Immobility/stupor,Posturing/catalepsy,Grimacing,Echopraxia/echolalia,Mannerisms,Verbigeration,Rigidity,Negativism,Waxy flexibility,Withdrawal | 99.8 | 99.3 | 100.0 |
| 11 | | Excitement,Immobility/stupor,Posturing/catalepsy,Grimacing,Echopraxia/echolalia,Stereotypy,Mannerisms,Verbigeration,Rigidity,Negativism,Withdrawal | 99.8 | 99.3 | 100.0 |
| 11 | | Excitement,Immobility/stupor,Staring,Grimacing,Echopraxia/echolalia,Mannerisms,Verbigeration,Rigidity,Negativism,Waxy flexibility,Withdrawal | 99.8 | 99.3 | 100.0 |
| 11 | | Excitement,Immobility/stupor,Staring,Grimacing,Echopraxia/echolalia,Stereotypy,Mannerisms,Verbigeration,Rigidity,Negativism,Waxy flexibility | 99.8 | 99.3 | 100.0 |
| 11 | | Excitement,Immobility/stupor,Staring,Grimacing,Echopraxia/echolalia,Stereotypy,Mannerisms,Verbigeration,Rigidity,Waxy flexibility,Withdrawal | 99.8 | 99.3 | 100.0 |
| 11 | | Excitement,Immobility/stupor,Staring,Grimacing,Echopraxia/echolalia,Stereotypy,Verbigeration,Rigidity,Negativism,Waxy flexibility,Withdrawal | 99.8 | 99.3 | 100.0 |
| 11 | | Excitement,Immobility/stupor,Staring,Posturing/catalepsy,Echopraxia/echolalia,Stereotypy,Mannerisms,Verbigeration,Negativism,Waxy flexibility,Withdrawal | 99.8 | 99.3 | 100.0 |
| 11 | | Excitement,Immobility/stupor,Staring,Posturing/catalepsy,Echopraxia/echolalia,Stereotypy,Mannerisms,Verbigeration,Rigidity,Negativism,Waxy flexibility | 99.8 | 99.3 | 100.0 |
| 11 | | Excitement,Immobility/stupor,Staring,Posturing/catalepsy,Echopraxia/echolalia,Stereotypy,Mannerisms,Verbigeration,Rigidity,Waxy flexibility,Withdrawal | 99.8 | 99.3 | 100.0 |
| 11 | | Excitement,Immobility/stupor,Staring,Posturing/catalepsy,Echopraxia/echolalia,Stereotypy,Verbigeration,Rigidity,Negativism,Waxy flexibility,Withdrawal | 99.8 | 99.3 | 100.0 |
| 11 | | Excitement,Immobility/stupor,Staring,Posturing/catalepsy,Grimacing,Echopraxia/echolalia,Mannerisms,Verbigeration,Negativism,Waxy flexibility,Withdrawal | 99.8 | 99.3 | 100.0 |
| 11 | | Excitement,Immobility/stupor,Staring,Posturing/catalepsy,Grimacing,Echopraxia/echolalia,Mannerisms,Verbigeration,Rigidity,Negativism,Waxy flexibility | 99.8 | 99.3 | 100.0 |
| 11 | | Excitement,Immobility/stupor,Staring,Posturing/catalepsy,Grimacing,Echopraxia/echolalia,Mannerisms,Verbigeration,Rigidity,Waxy flexibility,Withdrawal | 99.8 | 99.3 | 100.0 |
| 11 | | Excitement,Immobility/stupor,Staring,Posturing/catalepsy,Grimacing,Echopraxia/echolalia,Stereotypy,Mannerisms,Negativism,Waxy flexibility,Withdrawal | 99.8 | 99.3 | 100.0 |
| 11 | | Excitement,Immobility/stupor,Staring,Posturing/catalepsy,Grimacing,Echopraxia/echolalia,Stereotypy,Mannerisms,Rigidity,Waxy flexibility,Withdrawal | 99.8 | 99.3 | 100.0 |
| 11 | | Excitement,Immobility/stupor,Staring,Posturing/catalepsy,Grimacing,Echopraxia/echolalia,Stereotypy,Mannerisms,Verbigeration,Negativism,Withdrawal | 99.8 | 99.3 | 100.0 |
| 11 | | Excitement,Immobility/stupor,Staring,Posturing/catalepsy,Grimacing,Echopraxia/echolalia,Stereotypy,Mannerisms,Verbigeration,Rigidity,Negativism | 99.8 | 99.3 | 100.0 |
| 11 | | Excitement,Immobility/stupor,Staring,Posturing/catalepsy,Grimacing,Echopraxia/echolalia,Stereotypy,Mannerisms,Verbigeration,Rigidity,Withdrawal | 99.8 | 99.3 | 100.0 |
| 11 | | Excitement,Immobility/stupor,Staring,Posturing/catalepsy,Grimacing,Echopraxia/echolalia,Stereotypy,Rigidity,Negativism,Waxy flexibility,Withdrawal | 99.8 | 99.3 | 100.0 |
| 11 | | Excitement,Immobility/stupor,Staring,Posturing/catalepsy,Grimacing,Echopraxia/echolalia,Stereotypy,Verbigeration,Rigidity,Negativism,Withdrawal | 99.8 | 99.3 | 100.0 |
| 11 | | Excitement,Immobility/stupor,Staring,Posturing/catalepsy,Grimacing,Echopraxia/echolalia,Verbigeration,Rigidity,Negativism,Waxy flexibility,Withdrawal | 99.8 | 99.3 | 100.0 |
| 11 | | Excitement,Immobility/stupor,Staring,Posturing/catalepsy,Grimacing,Stereotypy,Mannerisms,Rigidity,Negativism,Waxy flexibility,Withdrawal | 99.8 | 99.3 | 100.0 |
| 11 | | Excitement,Immobility/stupor,Staring,Posturing/catalepsy,Grimacing,Stereotypy,Mannerisms,Verbigeration,Negativism,Waxy flexibility,Withdrawal | 99.8 | 99.3 | 100.0 |
| 11 | | Excitement,Immobility/stupor,Staring,Posturing/catalepsy,Grimacing,Stereotypy,Mannerisms,Verbigeration,Rigidity,Negativism,Waxy flexibility | 99.8 | 99.3 | 100.0 |
| 11 | | Excitement,Immobility/stupor,Staring,Posturing/catalepsy,Grimacing,Stereotypy,Mannerisms,Verbigeration,Rigidity,Waxy flexibility,Withdrawal | 99.8 | 99.3 | 100.0 |
| 11 | | Excitement,Immobility/stupor,Staring,Posturing/catalepsy,Grimacing,Stereotypy,Verbigeration,Rigidity,Negativism,Waxy flexibility,Withdrawal | 99.8 | 99.3 | 100.0 |
| 11 | | Excitement,Mutism,Staring,Grimacing,Echopraxia/echolalia,Mannerisms,Verbigeration,Rigidity,Negativism,Waxy flexibility,Withdrawal | 99.8 | 99.3 | 100.0 |
| 11 | | Excitement,Mutism,Staring,Posturing/catalepsy,Grimacing,Echopraxia/echolalia,Stereotypy,Mannerisms,Rigidity,Negativism,Waxy flexibility | 99.8 | 99.3 | 100.0 |
| 11 | | Excitement,Mutism,Staring,Posturing/catalepsy,Grimacing,Echopraxia/echolalia,Stereotypy,Mannerisms,Verbigeration,Rigidity,Waxy flexibility | 99.8 | 99.3 | 100.0 |
| 11 | | Excitement,Mutism,Staring,Posturing/catalepsy,Grimacing,Stereotypy,Mannerisms,Rigidity,Negativism,Waxy flexibility,Withdrawal | 99.8 | 99.3 | 100.0 |
| 11 | | Excitement,Mutism,Staring,Posturing/catalepsy,Grimacing,Stereotypy,Mannerisms,Verbigeration,Rigidity,Negativism,Waxy flexibility | 99.8 | 99.3 | 100.0 |
| 11 | | Immobility/stupor,Mutism,Staring,Echopraxia/echolalia,Stereotypy,Mannerisms,Verbigeration,Rigidity,Negativism,Waxy flexibility,Withdrawal | 99.8 | 99.3 | 100.0 |
| 11 | | Immobility/stupor,Mutism,Staring,Grimacing,Echopraxia/echolalia,Stereotypy,Mannerisms,Verbigeration,Rigidity,Negativism,Waxy flexibility | 99.8 | 99.3 | 100.0 |
| 11 | | Immobility/stupor,Mutism,Staring,Posturing/catalepsy,Echopraxia/echolalia,Stereotypy,Mannerisms,Rigidity,Negativism,Waxy flexibility,Withdrawal | 99.8 | 99.3 | 100.0 |
| 11 | | Immobility/stupor,Mutism,Staring,Posturing/catalepsy,Echopraxia/echolalia,Stereotypy,Mannerisms,Verbigeration,Rigidity,Negativism,Withdrawal | 99.8 | 99.3 | 100.0 |
| 11 | | Immobility/stupor,Mutism,Staring,Posturing/catalepsy,Grimacing,Echopraxia/echolalia,Mannerisms,Rigidity,Negativism,Waxy flexibility,Withdrawal | 99.8 | 99.3 | 100.0 |
| 11 | | Immobility/stupor,Mutism,Staring,Posturing/catalepsy,Grimacing,Echopraxia/echolalia,Mannerisms,Verbigeration,Negativism,Waxy flexibility,Withdrawal | 99.8 | 99.3 | 100.0 |
| 11 | | Immobility/stupor,Mutism,Staring,Posturing/catalepsy,Grimacing,Echopraxia/echolalia,Mannerisms,Verbigeration,Rigidity,Negativism,Withdrawal | 99.8 | 99.3 | 100.0 |
| 11 | | Immobility/stupor,Mutism,Staring,Posturing/catalepsy,Grimacing,Echopraxia/echolalia,Stereotypy,Mannerisms,Verbigeration,Negativism,Waxy flexibility | 99.8 | 99.3 | 100.0 |
| 11 | | Immobility/stupor,Mutism,Staring,Posturing/catalepsy,Grimacing,Echopraxia/echolalia,Stereotypy,Mannerisms,Verbigeration,Rigidity,Negativism | 99.8 | 99.3 | 100.0 |
| 11 | | Immobility/stupor,Mutism,Staring,Posturing/catalepsy,Grimacing,Stereotypy,Mannerisms,Rigidity,Negativism,Waxy flexibility,Withdrawal | 99.8 | 99.3 | 100.0 |
| 11 | | Immobility/stupor,Mutism,Staring,Posturing/catalepsy,Grimacing,Stereotypy,Mannerisms,Verbigeration,Rigidity,Negativism,Waxy flexibility | 99.8 | 99.3 | 100.0 |
| 11 | | Immobility/stupor,Mutism,Staring,Posturing/catalepsy,Stereotypy,Mannerisms,Verbigeration,Rigidity,Negativism,Waxy flexibility,Withdrawal | 99.8 | 99.3 | 100.0 |
| 11 | | Immobility/stupor,Staring,Posturing/catalepsy,Echopraxia/echolalia,Stereotypy,Mannerisms,Verbigeration,Rigidity,Negativism,Waxy flexibility,Withdrawal | 99.8 | 99.3 | 100.0 |
| 11 | | Immobility/stupor,Staring,Posturing/catalepsy,Grimacing,Echopraxia/echolalia,Mannerisms,Verbigeration,Rigidity,Negativism,Waxy flexibility,Withdrawal | 99.8 | 99.3 | 100.0 |
| 11 | | Immobility/stupor,Staring,Posturing/catalepsy,Grimacing,Echopraxia/echolalia,Stereotypy,Mannerisms,Verbigeration,Negativism,Waxy flexibility,Withdrawal | 99.8 | 99.3 | 100.0 |
| 11 | | Mutism,Staring,Posturing/catalepsy,Echopraxia/echolalia,Stereotypy,Mannerisms,Verbigeration,Rigidity,Negativism,Waxy flexibility,Withdrawal | 99.8 | 99.3 | 100.0 |
| 11 | | Mutism,Staring,Posturing/catalepsy,Grimacing,Echopraxia/echolalia,Mannerisms,Verbigeration,Rigidity,Negativism,Waxy flexibility,Withdrawal | 99.8 | 99.3 | 100.0 |
| 11 | | Mutism,Staring,Posturing/catalepsy,Grimacing,Echopraxia/echolalia,Stereotypy,Mannerisms,Verbigeration,Rigidity,Negativism,Waxy flexibility | 99.8 | 99.3 | 100.0 |
| 11 | | Excitement,Immobility/stupor,Grimacing,Echopraxia/echolalia,Stereotypy,Mannerisms,Verbigeration,Rigidity,Negativism,Waxy flexibility,Withdrawal | 99.6 | 98.9 | 100.0 |
| 11 | | Excitement,Immobility/stupor,Mutism,Grimacing,Echopraxia/echolalia,Mannerisms,Verbigeration,Rigidity,Negativism,Waxy flexibility,Withdrawal | 99.6 | 98.9 | 100.0 |
| 11 | | Excitement,Immobility/stupor,Mutism,Grimacing,Echopraxia/echolalia,Stereotypy,Mannerisms,Rigidity,Negativism,Waxy flexibility,Withdrawal | 99.6 | 98.9 | 100.0 |
| 11 | | Excitement,Immobility/stupor,Mutism,Grimacing,Echopraxia/echolalia,Stereotypy,Mannerisms,Verbigeration,Rigidity,Negativism,Waxy flexibility | 99.6 | 98.9 | 100.0 |
| 11 | | Excitement,Immobility/stupor,Mutism,Grimacing,Stereotypy,Mannerisms,Verbigeration,Rigidity,Negativism,Waxy flexibility,Withdrawal | 99.6 | 98.9 | 100.0 |
| 11 | | Excitement,Immobility/stupor,Mutism,Posturing/catalepsy,Echopraxia/echolalia,Stereotypy,Mannerisms,Rigidity,Negativism,Waxy flexibility,Withdrawal | 99.6 | 98.9 | 100.0 |
| 11 | | Excitement,Immobility/stupor,Mutism,Posturing/catalepsy,Grimacing,Echopraxia/echolalia,Mannerisms,Verbigeration,Rigidity,Waxy flexibility,Withdrawal | 99.6 | 98.9 | 100.0 |
| 11 | | Excitement,Immobility/stupor,Mutism,Posturing/catalepsy,Grimacing,Echopraxia/echolalia,Stereotypy,Mannerisms,Verbigeration,Rigidity,Withdrawal | 99.6 | 98.9 | 100.0 |
| 11 | | Excitement,Immobility/stupor,Mutism,Posturing/catalepsy,Grimacing,Echopraxia/echolalia,Stereotypy,Verbigeration,Rigidity,Waxy flexibility,Withdrawal | 99.6 | 98.9 | 100.0 |
| 11 | | Excitement,Immobility/stupor,Mutism,Posturing/catalepsy,Grimacing,Stereotypy,Mannerisms,Verbigeration,Rigidity,Negativism,Waxy flexibility | 99.6 | 98.9 | 100.0 |
| 11 | | Excitement,Immobility/stupor,Mutism,Staring,Echopraxia/echolalia,Stereotypy,Mannerisms,Verbigeration,Negativism,Waxy flexibility,Withdrawal | 99.6 | 98.9 | 100.0 |
| 11 | | Excitement,Immobility/stupor,Mutism,Staring,Grimacing,Echopraxia/echolalia,Stereotypy,Mannerisms,Negativism,Waxy flexibility,Withdrawal | 99.6 | 98.9 | 100.0 |
| 11 | | Excitement,Immobility/stupor,Mutism,Staring,Grimacing,Echopraxia/echolalia,Stereotypy,Mannerisms,Verbigeration,Negativism,Waxy flexibility | 99.6 | 98.9 | 100.0 |
| 11 | | Excitement,Immobility/stupor,Mutism,Staring,Grimacing,Echopraxia/echolalia,Stereotypy,Mannerisms,Verbigeration,Negativism,Withdrawal | 99.6 | 98.9 | 100.0 |
| 11 | | Excitement,Immobility/stupor,Mutism,Staring,Posturing/catalepsy,Grimacing,Echopraxia/echolalia,Mannerisms,Verbigeration,Waxy flexibility,Withdrawal | 99.6 | 98.9 | 100.0 |
| 11 | | Excitement,Immobility/stupor,Mutism,Staring,Posturing/catalepsy,Grimacing,Echopraxia/echolalia,Stereotypy,Mannerisms,Waxy flexibility,Withdrawal | 99.6 | 98.9 | 100.0 |
| 11 | | Excitement,Immobility/stupor,Mutism,Staring,Posturing/catalepsy,Grimacing,Echopraxia/echolalia,Stereotypy,Verbigeration,Waxy flexibility,Withdrawal | 99.6 | 98.9 | 100.0 |
| 11 | | Excitement,Immobility/stupor,Posturing/catalepsy,Grimacing,Echopraxia/echolalia,Stereotypy,Mannerisms,Rigidity,Negativism,Waxy flexibility,Withdrawal | 99.6 | 98.9 | 100.0 |
| 11 | | Excitement,Immobility/stupor,Posturing/catalepsy,Grimacing,Echopraxia/echolalia,Stereotypy,Verbigeration,Rigidity,Negativism,Waxy flexibility,Withdrawal | 99.6 | 98.9 | 100.0 |
| 11 | | Excitement,Immobility/stupor,Posturing/catalepsy,Grimacing,Stereotypy,Mannerisms,Verbigeration,Rigidity,Negativism,Waxy flexibility,Withdrawal | 99.6 | 98.9 | 100.0 |
| 11 | | Excitement,Immobility/stupor,Staring,Posturing/catalepsy,Grimacing,Echopraxia/echolalia,Stereotypy,Mannerisms,Verbigeration,Negativism,Waxy flexibility | 99.6 | 98.9 | 100.0 |
| 11 | | Excitement,Immobility/stupor,Staring,Posturing/catalepsy,Grimacing,Echopraxia/echolalia,Stereotypy,Verbigeration,Rigidity,Waxy flexibility,Withdrawal | 99.6 | 98.9 | 100.0 |
| 11 | | Excitement,Mutism,Staring,Grimacing,Echopraxia/echolalia,Stereotypy,Mannerisms,Verbigeration,Negativism,Waxy flexibility,Withdrawal | 99.6 | 98.9 | 100.0 |
| 11 | | Excitement,Mutism,Staring,Posturing/catalepsy,Grimacing,Echopraxia/echolalia,Stereotypy,Mannerisms,Verbigeration,Waxy flexibility,Withdrawal | 99.6 | 98.9 | 100.0 |
| 11 | | Excitement,Staring,Posturing/catalepsy,Echopraxia/echolalia,Stereotypy,Mannerisms,Verbigeration,Rigidity,Negativism,Waxy flexibility,Withdrawal | 99.6 | 98.9 | 100.0 |
| 11 | | Excitement,Staring,Posturing/catalepsy,Grimacing,Echopraxia/echolalia,Mannerisms,Verbigeration,Rigidity,Negativism,Waxy flexibility,Withdrawal | 99.6 | 98.9 | 100.0 |
| 11 | | Excitement,Staring,Posturing/catalepsy,Grimacing,Stereotypy,Mannerisms,Verbigeration,Rigidity,Negativism,Waxy flexibility,Withdrawal | 99.6 | 98.9 | 100.0 |
| 11 | | Immobility/stupor,Mutism,Grimacing,Echopraxia/echolalia,Stereotypy,Mannerisms,Verbigeration,Rigidity,Negativism,Waxy flexibility,Withdrawal | 99.6 | 98.9 | 100.0 |
| 11 | | Immobility/stupor,Mutism,Posturing/catalepsy,Echopraxia/echolalia,Stereotypy,Mannerisms,Verbigeration,Rigidity,Negativism,Waxy flexibility,Withdrawal | 99.6 | 98.9 | 100.0 |
| 11 | | Immobility/stupor,Mutism,Posturing/catalepsy,Grimacing,Echopraxia/echolalia,Mannerisms,Verbigeration,Rigidity,Negativism,Waxy flexibility,Withdrawal | 99.6 | 98.9 | 100.0 |
| 11 | | Immobility/stupor,Mutism,Posturing/catalepsy,Grimacing,Echopraxia/echolalia,Stereotypy,Mannerisms,Verbigeration,Rigidity,Negativism,Withdrawal | 99.6 | 98.9 | 100.0 |
| 11 | | Immobility/stupor,Mutism,Posturing/catalepsy,Grimacing,Stereotypy,Mannerisms,Verbigeration,Rigidity,Negativism,Waxy flexibility,Withdrawal | 99.6 | 98.9 | 100.0 |
| 11 | | Immobility/stupor,Mutism,Staring,Grimacing,Echopraxia/echolalia,Mannerisms,Verbigeration,Rigidity,Negativism,Waxy flexibility,Withdrawal | 99.6 | 98.9 | 100.0 |
| 11 | | Immobility/stupor,Mutism,Staring,Grimacing,Echopraxia/echolalia,Stereotypy,Mannerisms,Verbigeration,Rigidity,Waxy flexibility,Withdrawal | 99.6 | 98.9 | 100.0 |
| 11 | | Immobility/stupor,Mutism,Staring,Posturing/catalepsy,Echopraxia/echolalia,Stereotypy,Mannerisms,Verbigeration,Negativism,Waxy flexibility,Withdrawal | 99.6 | 98.9 | 100.0 |
| 11 | | Immobility/stupor,Mutism,Staring,Posturing/catalepsy,Grimacing,Echopraxia/echolalia,Stereotypy,Mannerisms,Rigidity,Negativism,Waxy flexibility | 99.6 | 98.9 | 100.0 |
| 11 | | Immobility/stupor,Mutism,Staring,Posturing/catalepsy,Grimacing,Echopraxia/echolalia,Stereotypy,Mannerisms,Verbigeration,Rigidity,Withdrawal | 99.6 | 98.9 | 100.0 |
| 11 | | Immobility/stupor,Mutism,Staring,Posturing/catalepsy,Grimacing,Mannerisms,Verbigeration,Rigidity,Negativism,Waxy flexibility,Withdrawal | 99.6 | 98.9 | 100.0 |
| 11 | | Immobility/stupor,Mutism,Staring,Posturing/catalepsy,Grimacing,Stereotypy,Mannerisms,Verbigeration,Rigidity,Waxy flexibility,Withdrawal | 99.6 | 98.9 | 100.0 |
| 11 | | Immobility/stupor,Staring,Posturing/catalepsy,Grimacing,Echopraxia/echolalia,Stereotypy,Mannerisms,Verbigeration,Rigidity,Negativism,Waxy flexibility | 99.6 | 98.9 | 100.0 |
| 11 | | Mutism,Staring,Posturing/catalepsy,Grimacing,Echopraxia/echolalia,Stereotypy,Mannerisms,Verbigeration,Rigidity,Waxy flexibility,Withdrawal | 99.6 | 98.9 | 100.0 |
| 11 | | Excitement,Immobility/stupor,Mutism,Posturing/catalepsy,Echopraxia/echolalia,Stereotypy,Mannerisms,Verbigeration,Rigidity,Waxy flexibility,Withdrawal | 99.5 | 98.9 | 100.0 |
| 11 | | Excitement,Immobility/stupor,Mutism,Staring,Grimacing,Echopraxia/echolalia,Stereotypy,Verbigeration,Negativism,Waxy flexibility,Withdrawal | 99.5 | 98.9 | 100.0 |
| 11 | | Excitement,Immobility/stupor,Mutism,Staring,Posturing/catalepsy,Grimacing,Echopraxia/echolalia,Stereotypy,Mannerisms,Rigidity,Waxy flexibility | 99.5 | 98.9 | 100.0 |
| 11 | | Excitement,Immobility/stupor,Mutism,Staring,Posturing/catalepsy,Grimacing,Stereotypy,Mannerisms,Rigidity,Negativism,Waxy flexibility | 99.5 | 98.9 | 100.0 |
| 11 | | Excitement,Immobility/stupor,Staring,Posturing/catalepsy,Grimacing,Echopraxia/echolalia,Stereotypy,Mannerisms,Rigidity,Negativism,Waxy flexibility | 99.5 | 98.9 | 100.0 |
| 11 | | Excitement,Immobility/stupor,Staring,Posturing/catalepsy,Grimacing,Echopraxia/echolalia,Stereotypy,Verbigeration,Negativism,Waxy flexibility,Withdrawal | 99.5 | 98.9 | 100.0 |
| 11 | | Excitement,Immobility/stupor,Staring,Posturing/catalepsy,Grimacing,Echopraxia/echolalia,Stereotypy,Verbigeration,Rigidity,Negativism,Waxy flexibility | 99.5 | 98.9 | 100.0 |
| 11 | | Excitement,Staring,Grimacing,Echopraxia/echolalia,Stereotypy,Mannerisms,Verbigeration,Rigidity,Negativism,Waxy flexibility,Withdrawal | 99.5 | 98.9 | 100.0 |
| 11 | | Excitement,Staring,Posturing/catalepsy,Grimacing,Echopraxia/echolalia,Stereotypy,Mannerisms,Rigidity,Negativism,Waxy flexibility,Withdrawal | 99.5 | 98.9 | 100.0 |
| 11 | | Excitement,Staring,Posturing/catalepsy,Grimacing,Echopraxia/echolalia,Stereotypy,Mannerisms,Verbigeration,Rigidity,Negativism,Withdrawal | 99.5 | 98.9 | 100.0 |
| 11 | | Immobility/stupor,Mutism,Posturing/catalepsy,Grimacing,Echopraxia/echolalia,Stereotypy,Mannerisms,Rigidity,Negativism,Waxy flexibility,Withdrawal | 99.5 | 98.9 | 100.0 |
| 11 | | Immobility/stupor,Mutism,Staring,Grimacing,Echopraxia/echolalia,Stereotypy,Mannerisms,Verbigeration,Negativism,Waxy flexibility,Withdrawal | 99.5 | 98.9 | 100.0 |
| 11 | | Immobility/stupor,Mutism,Staring,Posturing/catalepsy,Echopraxia/echolalia,Mannerisms,Verbigeration,Rigidity,Negativism,Waxy flexibility,Withdrawal | 99.5 | 98.9 | 100.0 |
| 11 | | Immobility/stupor,Mutism,Staring,Posturing/catalepsy,Echopraxia/echolalia,Stereotypy,Mannerisms,Verbigeration,Rigidity,Negativism,Waxy flexibility | 99.5 | 98.9 | 100.0 |
| 11 | | Immobility/stupor,Mutism,Staring,Posturing/catalepsy,Grimacing,Echopraxia/echolalia,Mannerisms,Verbigeration,Rigidity,Negativism,Waxy flexibility | 99.5 | 98.9 | 100.0 |
| 11 | | Immobility/stupor,Mutism,Staring,Posturing/catalepsy,Grimacing,Echopraxia/echolalia,Stereotypy,Mannerisms,Rigidity,Waxy flexibility,Withdrawal | 99.5 | 98.9 | 100.0 |
| 11 | | Immobility/stupor,Posturing/catalepsy,Grimacing,Echopraxia/echolalia,Stereotypy,Mannerisms,Verbigeration,Rigidity,Negativism,Waxy flexibility,Withdrawal | 99.5 | 98.9 | 100.0 |
| 11 | | Staring,Posturing/catalepsy,Grimacing,Echopraxia/echolalia,Stereotypy,Mannerisms,Verbigeration,Rigidity,Negativism,Waxy flexibility,Withdrawal | 99.5 | 98.9 | 100.0 |
| 11 | | Excitement,Immobility/stupor,Mutism,Grimacing,Echopraxia/echolalia,Stereotypy,Mannerisms,Verbigeration,Rigidity,Waxy flexibility,Withdrawal | 99.3 | 98.4 | 100.0 |
| 11 | | Excitement,Immobility/stupor,Mutism,Posturing/catalepsy,Grimacing,Echopraxia/echolalia,Stereotypy,Mannerisms,Rigidity,Negativism,Waxy flexibility | 99.3 | 98.4 | 100.0 |
| 11 | | Excitement,Immobility/stupor,Mutism,Posturing/catalepsy,Grimacing,Echopraxia/echolalia,Stereotypy,Mannerisms,Rigidity,Waxy flexibility,Withdrawal | 99.3 | 98.4 | 100.0 |
| 11 | | Excitement,Immobility/stupor,Mutism,Posturing/catalepsy,Grimacing,Stereotypy,Mannerisms,Rigidity,Negativism,Waxy flexibility,Withdrawal | 99.3 | 98.4 | 100.0 |
| 11 | | Excitement,Immobility/stupor,Mutism,Posturing/catalepsy,Grimacing,Stereotypy,Mannerisms,Verbigeration,Rigidity,Waxy flexibility,Withdrawal | 99.3 | 98.4 | 100.0 |
| 11 | | Excitement,Immobility/stupor,Mutism,Staring,Grimacing,Echopraxia/echolalia,Mannerisms,Verbigeration,Negativism,Waxy flexibility,Withdrawal | 99.3 | 98.4 | 100.0 |
| 11 | | Excitement,Immobility/stupor,Mutism,Staring,Grimacing,Stereotypy,Mannerisms,Verbigeration,Negativism,Waxy flexibility,Withdrawal | 99.3 | 98.4 | 100.0 |
| 11 | | Excitement,Immobility/stupor,Mutism,Staring,Posturing/catalepsy,Echopraxia/echolalia,Stereotypy,Mannerisms,Verbigeration,Waxy flexibility,Withdrawal | 99.3 | 98.4 | 100.0 |
| 11 | | Excitement,Immobility/stupor,Mutism,Staring,Posturing/catalepsy,Grimacing,Echopraxia/echolalia,Stereotypy,Mannerisms,Verbigeration,Waxy flexibility | 99.3 | 98.4 | 100.0 |
| 11 | | Excitement,Immobility/stupor,Mutism,Staring,Posturing/catalepsy,Grimacing,Echopraxia/echolalia,Stereotypy,Mannerisms,Verbigeration,Withdrawal | 99.3 | 98.4 | 100.0 |
| 11 | | Excitement,Immobility/stupor,Mutism,Staring,Posturing/catalepsy,Grimacing,Stereotypy,Mannerisms,Verbigeration,Waxy flexibility,Withdrawal | 99.3 | 98.4 | 100.0 |
| 11 | | Excitement,Immobility/stupor,Posturing/catalepsy,Grimacing,Echopraxia/echolalia,Stereotypy,Mannerisms,Verbigeration,Rigidity,Negativism,Waxy flexibility | 99.3 | 98.4 | 100.0 |
| 11 | | Excitement,Immobility/stupor,Staring,Grimacing,Echopraxia/echolalia,Stereotypy,Mannerisms,Verbigeration,Negativism,Waxy flexibility,Withdrawal | 99.3 | 98.4 | 100.0 |
| 11 | | Excitement,Immobility/stupor,Staring,Posturing/catalepsy,Grimacing,Echopraxia/echolalia,Stereotypy,Mannerisms,Verbigeration,Rigidity,Waxy flexibility | 99.3 | 98.4 | 100.0 |
| 11 | | Excitement,Staring,Posturing/catalepsy,Grimacing,Echopraxia/echolalia,Stereotypy,Mannerisms,Verbigeration,Rigidity,Waxy flexibility,Withdrawal | 99.3 | 98.4 | 100.0 |
| 11 | | Immobility/stupor,Mutism,Posturing/catalepsy,Grimacing,Echopraxia/echolalia,Stereotypy,Mannerisms,Verbigeration,Rigidity,Negativism,Waxy flexibility | 99.3 | 98.4 | 100.0 |
| 11 | | Immobility/stupor,Mutism,Staring,Grimacing,Echopraxia/echolalia,Stereotypy,Verbigeration,Rigidity,Negativism,Waxy flexibility,Withdrawal | 99.3 | 98.4 | 100.0 |
| 11 | | Immobility/stupor,Mutism,Staring,Posturing/catalepsy,Grimacing,Echopraxia/echolalia,Mannerisms,Verbigeration,Rigidity,Waxy flexibility,Withdrawal | 99.3 | 98.4 | 100.0 |
| 11 | | Immobility/stupor,Mutism,Staring,Posturing/catalepsy,Grimacing,Echopraxia/echolalia,Stereotypy,Verbigeration,Rigidity,Negativism,Withdrawal | 99.3 | 98.4 | 100.0 |
| 11 | | Immobility/stupor,Mutism,Staring,Posturing/catalepsy,Grimacing,Stereotypy,Verbigeration,Rigidity,Negativism,Waxy flexibility,Withdrawal | 99.3 | 98.4 | 100.0 |
| 11 | | Mutism,Staring,Posturing/catalepsy,Grimacing,Echopraxia/echolalia,Stereotypy,Verbigeration,Rigidity,Negativism,Waxy flexibility,Withdrawal | 99.3 | 98.4 | 100.0 |
| 11 | | Excitement,Immobility/stupor,Mutism,Posturing/catalepsy,Echopraxia/echolalia,Stereotypy,Mannerisms,Verbigeration,Negativism,Waxy flexibility,Withdrawal | 99.1 | 98.2 | 99.8 |
| 11 | | Excitement,Immobility/stupor,Mutism,Posturing/catalepsy,Grimacing,Echopraxia/echolalia,Mannerisms,Verbigeration,Negativism,Waxy flexibility,Withdrawal | 99.1 | 98.2 | 99.8 |
| 11 | | Excitement,Immobility/stupor,Mutism,Posturing/catalepsy,Grimacing,Echopraxia/echolalia,Stereotypy,Mannerisms,Verbigeration,Negativism,Withdrawal | 99.1 | 98.2 | 99.8 |
| 11 | | Excitement,Immobility/stupor,Mutism,Posturing/catalepsy,Grimacing,Echopraxia/echolalia,Stereotypy,Mannerisms,Verbigeration,Rigidity,Waxy flexibility | 99.1 | 98.2 | 99.8 |
| 11 | | Excitement,Immobility/stupor,Mutism,Posturing/catalepsy,Grimacing,Echopraxia/echolalia,Stereotypy,Verbigeration,Negativism,Waxy flexibility,Withdrawal | 99.1 | 98.2 | 99.8 |
| 11 | | Excitement,Immobility/stupor,Mutism,Staring,Grimacing,Echopraxia/echolalia,Stereotypy,Mannerisms,Verbigeration,Waxy flexibility,Withdrawal | 99.1 | 98.2 | 99.8 |
| 11 | | Excitement,Immobility/stupor,Posturing/catalepsy,Grimacing,Echopraxia/echolalia,Stereotypy,Mannerisms,Verbigeration,Rigidity,Waxy flexibility,Withdrawal | 99.1 | 98.2 | 99.8 |
| 11 | | Excitement,Immobility/stupor,Staring,Posturing/catalepsy,Grimacing,Echopraxia/echolalia,Stereotypy,Mannerisms,Verbigeration,Waxy flexibility,Withdrawal | 99.1 | 98.2 | 99.8 |
| 11 | | Excitement,Staring,Posturing/catalepsy,Grimacing,Echopraxia/echolalia,Stereotypy,Mannerisms,Verbigeration,Negativism,Waxy flexibility,Withdrawal | 99.1 | 98.2 | 99.8 |
| 11 | | Excitement,Staring,Posturing/catalepsy,Grimacing,Echopraxia/echolalia,Stereotypy,Verbigeration,Rigidity,Negativism,Waxy flexibility,Withdrawal | 99.1 | 98.2 | 99.8 |
| 11 | | Immobility/stupor,Mutism,Posturing/catalepsy,Grimacing,Echopraxia/echolalia,Stereotypy,Verbigeration,Rigidity,Negativism,Waxy flexibility,Withdrawal | 99.1 | 98.2 | 99.8 |
| 11 | | Immobility/stupor,Mutism,Staring,Posturing/catalepsy,Echopraxia/echolalia,Stereotypy,Mannerisms,Verbigeration,Rigidity,Waxy flexibility,Withdrawal | 99.1 | 98.2 | 99.8 |
| 11 | | Immobility/stupor,Mutism,Staring,Posturing/catalepsy,Echopraxia/echolalia,Stereotypy,Verbigeration,Rigidity,Negativism,Waxy flexibility,Withdrawal | 99.1 | 98.2 | 99.8 |
| 11 | | Immobility/stupor,Mutism,Staring,Posturing/catalepsy,Grimacing,Echopraxia/echolalia,Stereotypy,Mannerisms,Verbigeration,Rigidity,Waxy flexibility | 99.1 | 98.2 | 99.8 |
| 11 | | Immobility/stupor,Mutism,Staring,Posturing/catalepsy,Grimacing,Echopraxia/echolalia,Stereotypy,Mannerisms,Verbigeration,Waxy flexibility,Withdrawal | 99.1 | 98.2 | 99.8 |
| 11 | | Immobility/stupor,Mutism,Staring,Posturing/catalepsy,Grimacing,Echopraxia/echolalia,Stereotypy,Rigidity,Negativism,Waxy flexibility,Withdrawal | 99.1 | 98.2 | 99.8 |
| 11 | | Immobility/stupor,Mutism,Staring,Posturing/catalepsy,Grimacing,Echopraxia/echolalia,Stereotypy,Verbigeration,Negativism,Waxy flexibility,Withdrawal | 99.1 | 98.2 | 99.8 |
| 11 | | Immobility/stupor,Mutism,Staring,Posturing/catalepsy,Grimacing,Echopraxia/echolalia,Stereotypy,Verbigeration,Rigidity,Negativism,Waxy flexibility | 99.1 | 98.2 | 99.8 |
| 11 | | Immobility/stupor,Staring,Posturing/catalepsy,Grimacing,Echopraxia/echolalia,Stereotypy,Mannerisms,Verbigeration,Rigidity,Waxy flexibility,Withdrawal | 99.1 | 98.2 | 99.8 |
| 11 | | Immobility/stupor,Staring,Posturing/catalepsy,Grimacing,Echopraxia/echolalia,Stereotypy,Verbigeration,Rigidity,Negativism,Waxy flexibility,Withdrawal | 99.1 | 98.2 | 99.8 |
| 11 | | Excitement,Immobility/stupor,Mutism,Posturing/catalepsy,Grimacing,Echopraxia/echolalia,Stereotypy,Mannerisms,Negativism,Waxy flexibility,Withdrawal | 98.9 | 97.8 | 99.8 |
| 11 | | Excitement,Immobility/stupor,Mutism,Posturing/catalepsy,Grimacing,Stereotypy,Mannerisms,Verbigeration,Negativism,Waxy flexibility,Withdrawal | 98.9 | 97.8 | 99.8 |
| 11 | | Immobility/stupor,Mutism,Posturing/catalepsy,Grimacing,Echopraxia/echolalia,Stereotypy,Mannerisms,Verbigeration,Negativism,Waxy flexibility,Withdrawal | 98.9 | 97.8 | 99.8 |
| 11 | | Immobility/stupor,Mutism,Staring,Posturing/catalepsy,Grimacing,Echopraxia/echolalia,Stereotypy,Verbigeration,Rigidity,Waxy flexibility,Withdrawal | 98.9 | 97.8 | 99.8 |
| 11 | | Immobility/stupor,Mutism,Staring,Posturing/catalepsy,Grimacing,Echopraxia/echolalia,Verbigeration,Rigidity,Negativism,Waxy flexibility,Withdrawal | 98.9 | 97.8 | 99.8 |
| 11 | | Excitement,Immobility/stupor,Mutism,Posturing/catalepsy,Grimacing,Echopraxia/echolalia,Stereotypy,Mannerisms,Verbigeration,Negativism,Waxy flexibility | 98.7 | 97.5 | 99.6 |
| 11 | | Excitement,Immobility/stupor,Posturing/catalepsy,Grimacing,Echopraxia/echolalia,Stereotypy,Mannerisms,Verbigeration,Negativism,Waxy flexibility,Withdrawal | 98.7 | 97.5 | 99.6 |
| 11 | | Excitement,Staring,Posturing/catalepsy,Grimacing,Echopraxia/echolalia,Stereotypy,Mannerisms,Verbigeration,Rigidity,Negativism,Waxy flexibility | 98.7 | 97.5 | 99.6 |
| 11 | | Immobility/stupor,Mutism,Posturing/catalepsy,Grimacing,Echopraxia/echolalia,Stereotypy,Mannerisms,Verbigeration,Rigidity,Waxy flexibility,Withdrawal | 98.7 | 97.5 | 99.6 |
| 11 | | Excitement,Immobility/stupor,Mutism,Grimacing,Echopraxia/echolalia,Stereotypy,Mannerisms,Verbigeration,Negativism,Waxy flexibility,Withdrawal | 98.4 | 97.1 | 99.6 |
| 11 | | Excitement,Immobility/stupor,Mutism,Posturing/catalepsy,Grimacing,Echopraxia/echolalia,Stereotypy,Mannerisms,Verbigeration,Waxy flexibility,Withdrawal | 98.2 | 96.9 | 99.3 |
| 11 | | Excitement,Mutism,Posturing/catalepsy,Echopraxia/echolalia,Stereotypy,Mannerisms,Verbigeration,Rigidity,Negativism,Waxy flexibility,Withdrawal | 97.5 | 96.0 | 98.9 |
| 11 | | Excitement,Mutism,Posturing/catalepsy,Grimacing,Echopraxia/echolalia,Mannerisms,Verbigeration,Rigidity,Negativism,Waxy flexibility,Withdrawal | 97.5 | 96.0 | 98.9 |
| 11 | | Excitement,Mutism,Posturing/catalepsy,Grimacing,Echopraxia/echolalia,Stereotypy,Mannerisms,Verbigeration,Rigidity,Negativism,Withdrawal | 97.3 | 95.7 | 98.7 |
| 11 | | Excitement,Mutism,Posturing/catalepsy,Grimacing,Echopraxia/echolalia,Stereotypy,Verbigeration,Rigidity,Negativism,Waxy flexibility,Withdrawal | 97.3 | 95.7 | 98.7 |
| 11 | | Excitement,Mutism,Posturing/catalepsy,Grimacing,Stereotypy,Mannerisms,Verbigeration,Rigidity,Negativism,Waxy flexibility,Withdrawal | 97.3 | 95.7 | 98.7 |
| 11 | | Mutism,Posturing/catalepsy,Grimacing,Echopraxia/echolalia,Stereotypy,Mannerisms,Verbigeration,Rigidity,Negativism,Waxy flexibility,Withdrawal | 97.3 | 95.7 | 98.7 |
| 11 | | Excitement,Mutism,Grimacing,Echopraxia/echolalia,Stereotypy,Mannerisms,Verbigeration,Rigidity,Negativism,Waxy flexibility,Withdrawal | 97.1 | 95.5 | 98.4 |
| 11 | | Excitement,Mutism,Posturing/catalepsy,Grimacing,Echopraxia/echolalia,Stereotypy,Mannerisms,Rigidity,Negativism,Waxy flexibility,Withdrawal | 97.1 | 95.5 | 98.7 |
| 11 | | Excitement,Mutism,Posturing/catalepsy,Grimacing,Echopraxia/echolalia,Stereotypy,Mannerisms,Verbigeration,Rigidity,Negativism,Waxy flexibility | 96.6 | 94.8 | 98.2 |
| 11 | | Excitement,Mutism,Posturing/catalepsy,Grimacing,Echopraxia/echolalia,Stereotypy,Mannerisms,Verbigeration,Rigidity,Waxy flexibility,Withdrawal | 96.6 | 94.8 | 98.2 |
| 11 | | Excitement,Posturing/catalepsy,Grimacing,Echopraxia/echolalia,Stereotypy,Mannerisms,Verbigeration,Rigidity,Negativism,Waxy flexibility,Withdrawal | 96.2 | 94.4 | 98.0 |
| 11 | | Excitement,Mutism,Posturing/catalepsy,Grimacing,Echopraxia/echolalia,Stereotypy,Mannerisms,Verbigeration,Negativism,Waxy flexibility,Withdrawal | 95.5 | 93.5 | 97.3 |

Table S12: sensitivity (relative to the full BFCSI) of all twelve question combinations, and the 95% confidence interval derived from bootstrapping, sorted by decreasing sensitivity.

|  |  |  | 95% CI | |
| --- | --- | --- | --- | --- |
| # of Qs | Combination | Sensitivity | Lower | Upper |
| 12 | Excitement,Immobility/stupor,Mutism,Posturing/catalepsy,Echopraxia/echolalia,Stereotypy,Mannerisms,Verbigeration,Rigidity,Negativism,Waxy flexibility,Withdrawal | 100.0 | 100.0 | 100.0 |
| 12 | Excitement,Immobility/stupor,Mutism,Posturing/catalepsy,Grimacing,Echopraxia/echolalia,Mannerisms,Verbigeration,Rigidity,Negativism,Waxy flexibility,Withdrawal | 100.0 | 100.0 | 100.0 |
| 12 | Excitement,Immobility/stupor,Mutism,Posturing/catalepsy,Grimacing,Echopraxia/echolalia,Stereotypy,Mannerisms,Verbigeration,Rigidity,Negativism,Withdrawal | 100.0 | 100.0 | 100.0 |
| 12 | Excitement,Immobility/stupor,Mutism,Posturing/catalepsy,Grimacing,Echopraxia/echolalia,Stereotypy,Verbigeration,Rigidity,Negativism,Waxy flexibility,Withdrawal | 100.0 | 100.0 | 100.0 |
| 12 | Excitement,Immobility/stupor,Mutism,Staring,Echopraxia/echolalia,Stereotypy,Mannerisms,Verbigeration,Rigidity,Negativism,Waxy flexibility,Withdrawal | 100.0 | 100.0 | 100.0 |
| 12 | Excitement,Immobility/stupor,Mutism,Staring,Grimacing,Echopraxia/echolalia,Stereotypy,Mannerisms,Rigidity,Negativism,Waxy flexibility,Withdrawal | 100.0 | 100.0 | 100.0 |
| 12 | Excitement,Immobility/stupor,Mutism,Staring,Grimacing,Echopraxia/echolalia,Stereotypy,Mannerisms,Verbigeration,Rigidity,Negativism,Waxy flexibility | 100.0 | 100.0 | 100.0 |
| 12 | Excitement,Immobility/stupor,Mutism,Staring,Grimacing,Echopraxia/echolalia,Stereotypy,Mannerisms,Verbigeration,Rigidity,Negativism,Withdrawal | 100.0 | 100.0 | 100.0 |
| 12 | Excitement,Immobility/stupor,Mutism,Staring,Grimacing,Echopraxia/echolalia,Stereotypy,Mannerisms,Verbigeration,Rigidity,Waxy flexibility,Withdrawal | 100.0 | 100.0 | 100.0 |
| 12 | Excitement,Immobility/stupor,Mutism,Staring,Grimacing,Echopraxia/echolalia,Stereotypy,Verbigeration,Rigidity,Negativism,Waxy flexibility,Withdrawal | 100.0 | 100.0 | 100.0 |
| 12 | Excitement,Immobility/stupor,Mutism,Staring,Grimacing,Stereotypy,Mannerisms,Verbigeration,Rigidity,Negativism,Waxy flexibility,Withdrawal | 100.0 | 100.0 | 100.0 |
| 12 | Excitement,Immobility/stupor,Mutism,Staring,Posturing/catalepsy,Echopraxia/echolalia,Mannerisms,Verbigeration,Rigidity,Negativism,Waxy flexibility,Withdrawal | 100.0 | 100.0 | 100.0 |
| 12 | Excitement,Immobility/stupor,Mutism,Staring,Posturing/catalepsy,Echopraxia/echolalia,Stereotypy,Mannerisms,Rigidity,Negativism,Waxy flexibility,Withdrawal | 100.0 | 100.0 | 100.0 |
| 12 | Excitement,Immobility/stupor,Mutism,Staring,Posturing/catalepsy,Echopraxia/echolalia,Stereotypy,Mannerisms,Verbigeration,Negativism,Waxy flexibility,Withdrawal | 100.0 | 100.0 | 100.0 |
| 12 | Excitement,Immobility/stupor,Mutism,Staring,Posturing/catalepsy,Echopraxia/echolalia,Stereotypy,Mannerisms,Verbigeration,Rigidity,Negativism,Waxy flexibility | 100.0 | 100.0 | 100.0 |
| 12 | Excitement,Immobility/stupor,Mutism,Staring,Posturing/catalepsy,Echopraxia/echolalia,Stereotypy,Mannerisms,Verbigeration,Rigidity,Negativism,Withdrawal | 100.0 | 100.0 | 100.0 |
| 12 | Excitement,Immobility/stupor,Mutism,Staring,Posturing/catalepsy,Echopraxia/echolalia,Stereotypy,Mannerisms,Verbigeration,Rigidity,Waxy flexibility,Withdrawal | 100.0 | 100.0 | 100.0 |
| 12 | Excitement,Immobility/stupor,Mutism,Staring,Posturing/catalepsy,Echopraxia/echolalia,Stereotypy,Verbigeration,Rigidity,Negativism,Waxy flexibility,Withdrawal | 100.0 | 100.0 | 100.0 |
| 12 | Excitement,Immobility/stupor,Mutism,Staring,Posturing/catalepsy,Grimacing,Echopraxia/echolalia,Mannerisms,Rigidity,Negativism,Waxy flexibility,Withdrawal | 100.0 | 100.0 | 100.0 |
| 12 | Excitement,Immobility/stupor,Mutism,Staring,Posturing/catalepsy,Grimacing,Echopraxia/echolalia,Mannerisms,Verbigeration,Negativism,Waxy flexibility,Withdrawal | 100.0 | 100.0 | 100.0 |
| 12 | Excitement,Immobility/stupor,Mutism,Staring,Posturing/catalepsy,Grimacing,Echopraxia/echolalia,Mannerisms,Verbigeration,Rigidity,Negativism,Waxy flexibility | 100.0 | 100.0 | 100.0 |
| 12 | Excitement,Immobility/stupor,Mutism,Staring,Posturing/catalepsy,Grimacing,Echopraxia/echolalia,Mannerisms,Verbigeration,Rigidity,Negativism,Withdrawal | 100.0 | 100.0 | 100.0 |
| 12 | Excitement,Immobility/stupor,Mutism,Staring,Posturing/catalepsy,Grimacing,Echopraxia/echolalia,Mannerisms,Verbigeration,Rigidity,Waxy flexibility,Withdrawal | 100.0 | 100.0 | 100.0 |
| 12 | Excitement,Immobility/stupor,Mutism,Staring,Posturing/catalepsy,Grimacing,Echopraxia/echolalia,Stereotypy,Mannerisms,Negativism,Waxy flexibility,Withdrawal | 100.0 | 100.0 | 100.0 |
| 12 | Excitement,Immobility/stupor,Mutism,Staring,Posturing/catalepsy,Grimacing,Echopraxia/echolalia,Stereotypy,Mannerisms,Rigidity,Negativism,Withdrawal | 100.0 | 100.0 | 100.0 |
| 12 | Excitement,Immobility/stupor,Mutism,Staring,Posturing/catalepsy,Grimacing,Echopraxia/echolalia,Stereotypy,Mannerisms,Rigidity,Waxy flexibility,Withdrawal | 100.0 | 100.0 | 100.0 |
| 12 | Excitement,Immobility/stupor,Mutism,Staring,Posturing/catalepsy,Grimacing,Echopraxia/echolalia,Stereotypy,Mannerisms,Verbigeration,Negativism,Waxy flexibility | 100.0 | 100.0 | 100.0 |
| 12 | Excitement,Immobility/stupor,Mutism,Staring,Posturing/catalepsy,Grimacing,Echopraxia/echolalia,Stereotypy,Mannerisms,Verbigeration,Negativism,Withdrawal | 100.0 | 100.0 | 100.0 |
| 12 | Excitement,Immobility/stupor,Mutism,Staring,Posturing/catalepsy,Grimacing,Echopraxia/echolalia,Stereotypy,Mannerisms,Verbigeration,Rigidity,Negativism | 100.0 | 100.0 | 100.0 |
| 12 | Excitement,Immobility/stupor,Mutism,Staring,Posturing/catalepsy,Grimacing,Echopraxia/echolalia,Stereotypy,Mannerisms,Verbigeration,Rigidity,Withdrawal | 100.0 | 100.0 | 100.0 |
| 12 | Excitement,Immobility/stupor,Mutism,Staring,Posturing/catalepsy,Grimacing,Echopraxia/echolalia,Stereotypy,Rigidity,Negativism,Waxy flexibility,Withdrawal | 100.0 | 100.0 | 100.0 |
| 12 | Excitement,Immobility/stupor,Mutism,Staring,Posturing/catalepsy,Grimacing,Echopraxia/echolalia,Stereotypy,Verbigeration,Negativism,Waxy flexibility,Withdrawal | 100.0 | 100.0 | 100.0 |
| 12 | Excitement,Immobility/stupor,Mutism,Staring,Posturing/catalepsy,Grimacing,Echopraxia/echolalia,Stereotypy,Verbigeration,Rigidity,Negativism,Waxy flexibility | 100.0 | 100.0 | 100.0 |
| 12 | Excitement,Immobility/stupor,Mutism,Staring,Posturing/catalepsy,Grimacing,Echopraxia/echolalia,Stereotypy,Verbigeration,Rigidity,Negativism,Withdrawal | 100.0 | 100.0 | 100.0 |
| 12 | Excitement,Immobility/stupor,Mutism,Staring,Posturing/catalepsy,Grimacing,Echopraxia/echolalia,Stereotypy,Verbigeration,Rigidity,Waxy flexibility,Withdrawal | 100.0 | 100.0 | 100.0 |
| 12 | Excitement,Immobility/stupor,Mutism,Staring,Posturing/catalepsy,Grimacing,Echopraxia/echolalia,Verbigeration,Rigidity,Negativism,Waxy flexibility,Withdrawal | 100.0 | 100.0 | 100.0 |
| 12 | Excitement,Immobility/stupor,Mutism,Staring,Posturing/catalepsy,Grimacing,Mannerisms,Verbigeration,Rigidity,Negativism,Waxy flexibility,Withdrawal | 100.0 | 100.0 | 100.0 |
| 12 | Excitement,Immobility/stupor,Mutism,Staring,Posturing/catalepsy,Grimacing,Stereotypy,Mannerisms,Verbigeration,Negativism,Waxy flexibility,Withdrawal | 100.0 | 100.0 | 100.0 |
| 12 | Excitement,Immobility/stupor,Mutism,Staring,Posturing/catalepsy,Grimacing,Stereotypy,Mannerisms,Verbigeration,Rigidity,Negativism,Waxy flexibility | 100.0 | 100.0 | 100.0 |
| 12 | Excitement,Immobility/stupor,Mutism,Staring,Posturing/catalepsy,Grimacing,Stereotypy,Mannerisms,Verbigeration,Rigidity,Negativism,Withdrawal | 100.0 | 100.0 | 100.0 |
| 12 | Excitement,Immobility/stupor,Mutism,Staring,Posturing/catalepsy,Grimacing,Stereotypy,Mannerisms,Verbigeration,Rigidity,Waxy flexibility,Withdrawal | 100.0 | 100.0 | 100.0 |
| 12 | Excitement,Immobility/stupor,Mutism,Staring,Posturing/catalepsy,Grimacing,Stereotypy,Verbigeration,Rigidity,Negativism,Waxy flexibility,Withdrawal | 100.0 | 100.0 | 100.0 |
| 12 | Excitement,Immobility/stupor,Mutism,Staring,Posturing/catalepsy,Stereotypy,Mannerisms,Verbigeration,Rigidity,Negativism,Waxy flexibility,Withdrawal | 100.0 | 100.0 | 100.0 |
| 12 | Excitement,Immobility/stupor,Staring,Grimacing,Echopraxia/echolalia,Stereotypy,Mannerisms,Verbigeration,Rigidity,Negativism,Waxy flexibility,Withdrawal | 100.0 | 100.0 | 100.0 |
| 12 | Excitement,Immobility/stupor,Staring,Posturing/catalepsy,Echopraxia/echolalia,Stereotypy,Mannerisms,Verbigeration,Rigidity,Negativism,Waxy flexibility,Withdrawal | 100.0 | 100.0 | 100.0 |
| 12 | Excitement,Immobility/stupor,Staring,Posturing/catalepsy,Grimacing,Echopraxia/echolalia,Mannerisms,Verbigeration,Rigidity,Negativism,Waxy flexibility,Withdrawal | 100.0 | 100.0 | 100.0 |
| 12 | Excitement,Immobility/stupor,Staring,Posturing/catalepsy,Grimacing,Echopraxia/echolalia,Stereotypy,Mannerisms,Rigidity,Negativism,Waxy flexibility,Withdrawal | 100.0 | 100.0 | 100.0 |
| 12 | Excitement,Immobility/stupor,Staring,Posturing/catalepsy,Grimacing,Echopraxia/echolalia,Stereotypy,Mannerisms,Verbigeration,Rigidity,Negativism,Withdrawal | 100.0 | 100.0 | 100.0 |
| 12 | Excitement,Immobility/stupor,Staring,Posturing/catalepsy,Grimacing,Stereotypy,Mannerisms,Verbigeration,Rigidity,Negativism,Waxy flexibility,Withdrawal | 100.0 | 100.0 | 100.0 |
| 12 | Excitement,Mutism,Staring,Grimacing,Echopraxia/echolalia,Stereotypy,Mannerisms,Verbigeration,Rigidity,Negativism,Waxy flexibility,Withdrawal | 100.0 | 100.0 | 100.0 |
| 12 | Excitement,Mutism,Staring,Posturing/catalepsy,Echopraxia/echolalia,Stereotypy,Mannerisms,Verbigeration,Rigidity,Negativism,Waxy flexibility,Withdrawal | 100.0 | 100.0 | 100.0 |
| 12 | Excitement,Mutism,Staring,Posturing/catalepsy,Grimacing,Echopraxia/echolalia,Mannerisms,Verbigeration,Rigidity,Negativism,Waxy flexibility,Withdrawal | 100.0 | 100.0 | 100.0 |
| 12 | Excitement,Mutism,Staring,Posturing/catalepsy,Grimacing,Echopraxia/echolalia,Stereotypy,Mannerisms,Rigidity,Negativism,Waxy flexibility,Withdrawal | 100.0 | 100.0 | 100.0 |
| 12 | Excitement,Mutism,Staring,Posturing/catalepsy,Grimacing,Echopraxia/echolalia,Stereotypy,Mannerisms,Verbigeration,Negativism,Waxy flexibility,Withdrawal | 100.0 | 100.0 | 100.0 |
| 12 | Excitement,Mutism,Staring,Posturing/catalepsy,Grimacing,Echopraxia/echolalia,Stereotypy,Mannerisms,Verbigeration,Rigidity,Negativism,Waxy flexibility | 100.0 | 100.0 | 100.0 |
| 12 | Excitement,Mutism,Staring,Posturing/catalepsy,Grimacing,Echopraxia/echolalia,Stereotypy,Mannerisms,Verbigeration,Rigidity,Negativism,Withdrawal | 100.0 | 100.0 | 100.0 |
| 12 | Excitement,Mutism,Staring,Posturing/catalepsy,Grimacing,Echopraxia/echolalia,Stereotypy,Mannerisms,Verbigeration,Rigidity,Waxy flexibility,Withdrawal | 100.0 | 100.0 | 100.0 |
| 12 | Excitement,Mutism,Staring,Posturing/catalepsy,Grimacing,Echopraxia/echolalia,Stereotypy,Verbigeration,Rigidity,Negativism,Waxy flexibility,Withdrawal | 100.0 | 100.0 | 100.0 |
| 12 | Excitement,Mutism,Staring,Posturing/catalepsy,Grimacing,Stereotypy,Mannerisms,Verbigeration,Rigidity,Negativism,Waxy flexibility,Withdrawal | 100.0 | 100.0 | 100.0 |
| 12 | Immobility/stupor,Mutism,Staring,Grimacing,Echopraxia/echolalia,Stereotypy,Mannerisms,Verbigeration,Rigidity,Negativism,Waxy flexibility,Withdrawal | 100.0 | 100.0 | 100.0 |
| 12 | Immobility/stupor,Mutism,Staring,Posturing/catalepsy,Grimacing,Echopraxia/echolalia,Stereotypy,Mannerisms,Rigidity,Negativism,Waxy flexibility,Withdrawal | 100.0 | 100.0 | 100.0 |
| 12 | Immobility/stupor,Mutism,Staring,Posturing/catalepsy,Grimacing,Echopraxia/echolalia,Stereotypy,Mannerisms,Verbigeration,Negativism,Waxy flexibility,Withdrawal | 100.0 | 100.0 | 100.0 |
| 12 | Immobility/stupor,Mutism,Staring,Posturing/catalepsy,Grimacing,Echopraxia/echolalia,Stereotypy,Mannerisms,Verbigeration,Rigidity,Negativism,Withdrawal | 100.0 | 100.0 | 100.0 |
| 12 | Immobility/stupor,Mutism,Staring,Posturing/catalepsy,Grimacing,Stereotypy,Mannerisms,Verbigeration,Rigidity,Negativism,Waxy flexibility,Withdrawal | 100.0 | 100.0 | 100.0 |
| 12 | Immobility/stupor,Staring,Posturing/catalepsy,Grimacing,Echopraxia/echolalia,Stereotypy,Mannerisms,Verbigeration,Rigidity,Negativism,Waxy flexibility,Withdrawal | 100.0 | 100.0 | 100.0 |
| 12 | Mutism,Staring,Posturing/catalepsy,Grimacing,Echopraxia/echolalia,Stereotypy,Mannerisms,Verbigeration,Rigidity,Negativism,Waxy flexibility,Withdrawal | 100.0 | 100.0 | 100.0 |
| 12 | Excitement,Immobility/stupor,Mutism,Grimacing,Echopraxia/echolalia,Stereotypy,Mannerisms,Verbigeration,Rigidity,Negativism,Waxy flexibility,Withdrawal | 99.8 | 99.3 | 100.0 |
| 12 | Excitement,Immobility/stupor,Mutism,Posturing/catalepsy,Grimacing,Echopraxia/echolalia,Stereotypy,Mannerisms,Rigidity,Negativism,Waxy flexibility,Withdrawal | 99.8 | 99.3 | 100.0 |
| 12 | Excitement,Immobility/stupor,Mutism,Posturing/catalepsy,Grimacing,Echopraxia/echolalia,Stereotypy,Mannerisms,Verbigeration,Rigidity,Negativism,Waxy flexibility | 99.8 | 99.3 | 100.0 |
| 12 | Excitement,Immobility/stupor,Mutism,Posturing/catalepsy,Grimacing,Stereotypy,Mannerisms,Verbigeration,Rigidity,Negativism,Waxy flexibility,Withdrawal | 99.8 | 99.3 | 100.0 |
| 12 | Excitement,Immobility/stupor,Mutism,Staring,Grimacing,Echopraxia/echolalia,Mannerisms,Verbigeration,Rigidity,Negativism,Waxy flexibility,Withdrawal | 99.8 | 99.3 | 100.0 |
| 12 | Excitement,Immobility/stupor,Mutism,Staring,Posturing/catalepsy,Grimacing,Echopraxia/echolalia,Stereotypy,Mannerisms,Rigidity,Negativism,Waxy flexibility | 99.8 | 99.3 | 100.0 |
| 12 | Excitement,Immobility/stupor,Mutism,Staring,Posturing/catalepsy,Grimacing,Echopraxia/echolalia,Stereotypy,Mannerisms,Verbigeration,Rigidity,Waxy flexibility | 99.8 | 99.3 | 100.0 |
| 12 | Excitement,Immobility/stupor,Mutism,Staring,Posturing/catalepsy,Grimacing,Stereotypy,Mannerisms,Rigidity,Negativism,Waxy flexibility,Withdrawal | 99.8 | 99.3 | 100.0 |
| 12 | Excitement,Immobility/stupor,Posturing/catalepsy,Grimacing,Echopraxia/echolalia,Stereotypy,Mannerisms,Verbigeration,Rigidity,Negativism,Waxy flexibility,Withdrawal | 99.8 | 99.3 | 100.0 |
| 12 | Excitement,Immobility/stupor,Staring,Posturing/catalepsy,Grimacing,Echopraxia/echolalia,Stereotypy,Mannerisms,Verbigeration,Negativism,Waxy flexibility,Withdrawal | 99.8 | 99.3 | 100.0 |
| 12 | Excitement,Immobility/stupor,Staring,Posturing/catalepsy,Grimacing,Echopraxia/echolalia,Stereotypy,Mannerisms,Verbigeration,Rigidity,Negativism,Waxy flexibility | 99.8 | 99.3 | 100.0 |
| 12 | Excitement,Immobility/stupor,Staring,Posturing/catalepsy,Grimacing,Echopraxia/echolalia,Stereotypy,Mannerisms,Verbigeration,Rigidity,Waxy flexibility,Withdrawal | 99.8 | 99.3 | 100.0 |
| 12 | Excitement,Immobility/stupor,Staring,Posturing/catalepsy,Grimacing,Echopraxia/echolalia,Stereotypy,Verbigeration,Rigidity,Negativism,Waxy flexibility,Withdrawal | 99.8 | 99.3 | 100.0 |
| 12 | Immobility/stupor,Mutism,Posturing/catalepsy,Grimacing,Echopraxia/echolalia,Stereotypy,Mannerisms,Verbigeration,Rigidity,Negativism,Waxy flexibility,Withdrawal | 99.8 | 99.3 | 100.0 |
| 12 | Immobility/stupor,Mutism,Staring,Posturing/catalepsy,Echopraxia/echolalia,Stereotypy,Mannerisms,Verbigeration,Rigidity,Negativism,Waxy flexibility,Withdrawal | 99.8 | 99.3 | 100.0 |
| 12 | Immobility/stupor,Mutism,Staring,Posturing/catalepsy,Grimacing,Echopraxia/echolalia,Mannerisms,Verbigeration,Rigidity,Negativism,Waxy flexibility,Withdrawal | 99.8 | 99.3 | 100.0 |
| 12 | Immobility/stupor,Mutism,Staring,Posturing/catalepsy,Grimacing,Echopraxia/echolalia,Stereotypy,Mannerisms,Verbigeration,Rigidity,Negativism,Waxy flexibility | 99.8 | 99.3 | 100.0 |
| 12 | Excitement,Immobility/stupor,Mutism,Posturing/catalepsy,Grimacing,Echopraxia/echolalia,Stereotypy,Mannerisms,Verbigeration,Rigidity,Waxy flexibility,Withdrawal | 99.6 | 98.9 | 100.0 |
| 12 | Excitement,Immobility/stupor,Mutism,Staring,Posturing/catalepsy,Grimacing,Echopraxia/echolalia,Stereotypy,Mannerisms,Verbigeration,Waxy flexibility,Withdrawal | 99.6 | 98.9 | 100.0 |
| 12 | Excitement,Staring,Posturing/catalepsy,Grimacing,Echopraxia/echolalia,Stereotypy,Mannerisms,Verbigeration,Rigidity,Negativism,Waxy flexibility,Withdrawal | 99.6 | 98.9 | 100.0 |
| 12 | Immobility/stupor,Mutism,Staring,Posturing/catalepsy,Grimacing,Echopraxia/echolalia,Stereotypy,Mannerisms,Verbigeration,Rigidity,Waxy flexibility,Withdrawal | 99.6 | 98.9 | 100.0 |
| 12 | Excitement,Immobility/stupor,Mutism,Staring,Grimacing,Echopraxia/echolalia,Stereotypy,Mannerisms,Verbigeration,Negativism,Waxy flexibility,Withdrawal | 99.5 | 98.9 | 100.0 |
| 12 | Immobility/stupor,Mutism,Staring,Posturing/catalepsy,Grimacing,Echopraxia/echolalia,Stereotypy,Verbigeration,Rigidity,Negativism,Waxy flexibility,Withdrawal | 99.3 | 98.4 | 100.0 |
| 12 | Excitement,Immobility/stupor,Mutism,Posturing/catalepsy,Grimacing,Echopraxia/echolalia,Stereotypy,Mannerisms,Verbigeration,Negativism,Waxy flexibility,Withdrawal | 99.1 | 98.2 | 99.8 |
| 12 | Excitement,Mutism,Posturing/catalepsy,Grimacing,Echopraxia/echolalia,Stereotypy,Mannerisms,Verbigeration,Rigidity,Negativism,Waxy flexibility,Withdrawal | 97.5 | 96.0 | 98.9 |

Table S13: sensitivity (relative to the full BFCSI) of all thirteen question combinations, and the 95% confidence interval derived from bootstrapping, sorted by decreasing sensitivity.

|  |  |  | 95% CI | |
| --- | --- | --- | --- | --- |
| # of Qs | Combination | Sensitivity | Lower | Upper |
| 13 | Excitement,Immobility/stupor,Mutism,Posturing/catalepsy,Grimacing,Echopraxia/echolalia,Stereotypy,Mannerisms,Verbigeration,Rigidity,Negativism,Waxy flexibility,Withdrawal | 100.0 | 100.0 | 100.0 |
| 13 | Excitement,Immobility/stupor,Mutism,Staring,Grimacing,Echopraxia/echolalia,Stereotypy,Mannerisms,Verbigeration,Rigidity,Negativism,Waxy flexibility,Withdrawal | 100.0 | 100.0 | 100.0 |
| 13 | Excitement,Immobility/stupor,Mutism,Staring,Posturing/catalepsy,Echopraxia/echolalia,Stereotypy,Mannerisms,Verbigeration,Rigidity,Negativism,Waxy flexibility,Withdrawal | 100.0 | 100.0 | 100.0 |
| 13 | Excitement,Immobility/stupor,Mutism,Staring,Posturing/catalepsy,Grimacing,Echopraxia/echolalia,Mannerisms,Verbigeration,Rigidity,Negativism,Waxy flexibility,Withdrawal | 100.0 | 100.0 | 100.0 |
| 13 | Excitement,Immobility/stupor,Mutism,Staring,Posturing/catalepsy,Grimacing,Echopraxia/echolalia,Stereotypy,Mannerisms,Rigidity,Negativism,Waxy flexibility,Withdrawal | 100.0 | 100.0 | 100.0 |
| 13 | Excitement,Immobility/stupor,Mutism,Staring,Posturing/catalepsy,Grimacing,Echopraxia/echolalia,Stereotypy,Mannerisms,Verbigeration,Negativism,Waxy flexibility,Withdrawal | 100.0 | 100.0 | 100.0 |
| 13 | Excitement,Immobility/stupor,Mutism,Staring,Posturing/catalepsy,Grimacing,Echopraxia/echolalia,Stereotypy,Mannerisms,Verbigeration,Rigidity,Negativism,Waxy flexibility | 100.0 | 100.0 | 100.0 |
| 13 | Excitement,Immobility/stupor,Mutism,Staring,Posturing/catalepsy,Grimacing,Echopraxia/echolalia,Stereotypy,Mannerisms,Verbigeration,Rigidity,Negativism,Withdrawal | 100.0 | 100.0 | 100.0 |
| 13 | Excitement,Immobility/stupor,Mutism,Staring,Posturing/catalepsy,Grimacing,Echopraxia/echolalia,Stereotypy,Mannerisms,Verbigeration,Rigidity,Waxy flexibility,Withdrawal | 100.0 | 100.0 | 100.0 |
| 13 | Excitement,Immobility/stupor,Mutism,Staring,Posturing/catalepsy,Grimacing,Echopraxia/echolalia,Stereotypy,Verbigeration,Rigidity,Negativism,Waxy flexibility,Withdrawal | 100.0 | 100.0 | 100.0 |
| 13 | Excitement,Immobility/stupor,Mutism,Staring,Posturing/catalepsy,Grimacing,Stereotypy,Mannerisms,Verbigeration,Rigidity,Negativism,Waxy flexibility,Withdrawal | 100.0 | 100.0 | 100.0 |
| 13 | Excitement,Immobility/stupor,Staring,Posturing/catalepsy,Grimacing,Echopraxia/echolalia,Stereotypy,Mannerisms,Verbigeration,Rigidity,Negativism,Waxy flexibility,Withdrawal | 100.0 | 100.0 | 100.0 |
| 13 | Excitement,Mutism,Staring,Posturing/catalepsy,Grimacing,Echopraxia/echolalia,Stereotypy,Mannerisms,Verbigeration,Rigidity,Negativism,Waxy flexibility,Withdrawal | 100.0 | 100.0 | 100.0 |
| 13 | Immobility/stupor,Mutism,Staring,Posturing/catalepsy,Grimacing,Echopraxia/echolalia,Stereotypy,Mannerisms,Verbigeration,Rigidity,Negativism,Waxy flexibility,Withdrawal | 100.0 | 100.0 | 100.0 |
